# Proteomic and genetic insights into ancestry-specific associations in Parkinson’s disease

**DOI:** 10.1101/2025.06.04.25329006

**Authors:** Amanda Wei-Yin Lim, Jia-Nee Foo, Elaine Guo-Yan Chew, Shen-Yang Lim, Ai Huey Tan, Stuart MacGregor, Miguel E. Rentería, Jue-Sheng Ong

**Affiliations:** Population Health Program, QIMR Berghofer, Brisbane, Queensland 4006, Australia; Faculty of Health, Medicine and Behavioural Sciences, The University of Queensland, Brisbane, Queensland 4006, Australia; Lee Kong Chian School of Medicine, Nanyang Technological University Singapore, Singapore 308232; Human Genetics, Genome Institute of Singapore, A*STAR, Singapore 138632; Division of Neurology, Department of Medicine, Faculty of Medicine, University of Malaya, 50603 Kuala Lumpur, Malaysia; The Mah Pooi Soo & Tan Chin Nam Centre for Parkinson’s & Related Disorders, University of Malaya, 50603 Kuala Lumpur, Malaysia; Brain & Mental Health Program, QIMR Berghofer, Brisbane, Queensland 4006, Australia

## Abstract

Genome-wide association studies of Parkinson’s disease (PD) have identified multiple genetic variants across populations, but the biological mechanisms remain largely unknown. To investigate ancestry-specific disease pathways, we performed a series of Mendelian randomisation analyses, integrating GWAS results with ancestry-specific protein quantitative trait loci data. For Europeans, we utilised the UK Biobank plasma proteomics (UKB-PPP) dataset alongside a European PD GWAS. For East Asians, we combined plasma proteomic data from Han Chinese and the East Asian subset of UKB-PPP with an East Asian PD GWAS. We identified 21 protein-encoding genes causally associated with PD in Europeans and 8 in East Asians, with *BST1* common to both (Europeans: OR=1.04, 95% CI 1.02-1.06; East Asians: OR=1.18, 95% CI 1.10-1.27). Ancestry-specific associations included *PRSS53* and *TXNDC15* in Europeans, and *HDGF*, *PM20D1*, and *TOP1* in East Asians. These findings highlighted ancestry-related differences in gene-protein-PD associations and have implications for biomarker development and targeted therapeutic strategies.

## INTRODUCTION

Parkinson’s disease (PD) is a chronic, progressively debilitating neurodegenerative disorder^1^ posing escalating global health concerns.^2^ As of 2019, the World Health Organization reported over 8.5 million individuals living with PD globally,^3^ resulting in 5.8 million disability-adjusted life years–an 81% increase since 2000.^3^ A 2023 meta-analysis of 83 studies across 37 countries revealed an all-age prevalence of 151 PD cases per 100,000 individuals worldwide,^3,4^ with notable regional variations. The 2021 age-standardised PD incidence and prevalence in Asia exceeded global averages, disproportionately affecting middle-aged and elderly males in countries with middle to low socio-demographic indices, despite earlier studies suggesting lower prevalence rates in certain Asian populations compared to European counterparts.^5–7^

Clinically, PD is characterised by bradykinesia, resting tremor, and muscle rigidity^1,8^ from the degeneration of dopaminergic neurons in the *substantia nigra*.^9,10^ Beyond these motor manifestations, PD encompasses a range of non-motor symptoms including cognitive impairment, autonomic dysfunction, and sleep disorders. These non-motor symptoms can precede motor symptoms by years and involve non-dopaminergic pathways.^11^

Current approved treatments for PD focus on symptomatic management through dopamine replacement therapies and, to a lesser extent, targeting non-dopaminergic pathways.^12,13^ However, none of these existing treatments are curative or disease-modifying.^1,9^ Recent drug trials have targeted alpha-synuclein aggregates, molecular pathways involved in neuronal survival (e.g. glucagon-like peptide-1 (*GLP-1*) agonists), and genetically defined subpopulations with pathogenic mutations (e.g. leucine-rich repeat kinase 2 (*LRRK2*) and beta-glucocerebrosidase (*GBA1*)).^13–17^

Biomarker development has been instrumental in understanding PD progression and informing clinical trials.^18,19^ Although the lack of reliable and sensitive progression biomarkers remains a challenge for developing neuroprotective therapies, ongoing research is addressing this gap,^18,20,21^ with major advances in alpha-synuclein assays^22^ and assays evaluating the biological impact of *LRRK2*-targeted drugs.^19,23^

The aetiology of PD involves environmental influences and genetic predisposition.^24–26^ While monogenic forms account for approximately 5-15% of PD cases,^24,27^ the majority are polygenic or idiopathic, involving rare and common variants.^28^ Whole-genome sequencing and genome-wide association studies (GWAS) reveal heterogeneous genetic architecture across populations, reflecting phenotypic and genotypic variability.^29–32^ Established PD genes such as *SNCA*, *LRRK2*, and *PINK1*—implicated in monogenic forms—exhibit population-specific distributions.^17,33^

European GWAS identified more than 90 common genetic variants linked to PD susceptibility, accounting for 16-36% of its heritability.^30^ In contrast, East Asian GWAS identified 11 loci, nine overlapping with European findings.^31^ Two novel population-specific loci, *SV2C* and *WBSCR17*, highlight unique genetic contributors in Asian populations. Multi-ancestry GWAS meta-analysis further expanded our understanding of the PD genetic architecture, which identified 78 distinct genomic regions associated with PD, including 12 previously undiscovered loci.^29^

Proteomics provide complementary insights into PD pathogenesis through protein-level gene expression analysis.^34^ Circulating proteins serve as functional intermediates mediating genetic disease effects while functioning as biomarkers for disease risk and progression and potential therapeutic targets.^35–37^ Integrating proteomic with genomic data allows for the identification of proteins causally linked to PD, revealing functional consequences of genetic variation for biomarker discovery and therapeutic development. Cross-ancestry protein-PD associations will provide insights into shared and ancestry-specific biological pathways.

This study aims to identify and compare proteins causally associated with PD risk in European and East Asian populations using Mendelian randomisation (MR). By leveraging the best available ancestry-specific proteomic and PD GWAS data at the time of this study, we aim to uncover differences in gene-protein-PD associations that could enhance understanding of population-specific disease mechanisms and potentially inform future biomarker development for PD diagnosis and treatment.

## MATERIALS AND METHODS

### Study design

We performed two-sample MR analyses to evaluate the causal effects of human circulating proteins on PD risk in European and East Asian populations. This design allows for the examination of causal relationships between exposures and outcomes of interest while leveraging large-scale GWAS data for both protein levels and PD risk, thereby maximising statistical power while maintaining ancestry-specific insights.^38^

Protein quantitative trait loci (pQTLs) serve as robust instrumental variables (IVs) in MR due to their strong associations with circulating protein levels.^39^ pQTLs are genetic variants that influence the expression levels of proteins in plasma, providing a direct link between genotype and phenotype.^36^ Cis-pQTLs, in particular, have large effect sizes and direct biological relevance to their target proteins, reducing concerns about horizontal pleiotropy.^40^ Furthermore, pQTLs typically explain a relatively large proportion of variance in protein levels, which enhances their utility in inferring causal relationships.

The MR framework relies on three core assumptions (Fig. 1): 1) Relevance: pQTLs must be strongly associated with the circulating protein of interest; 2) Independence: pQTLs must not be associated with any confounders of the protein-PD relationship; 3) Exclusion restriction: pQTLs must influence PD risk solely through their effects on the circulating protein, with no alternative pathways (horizontal pleiotropy).

**Figure 1:**
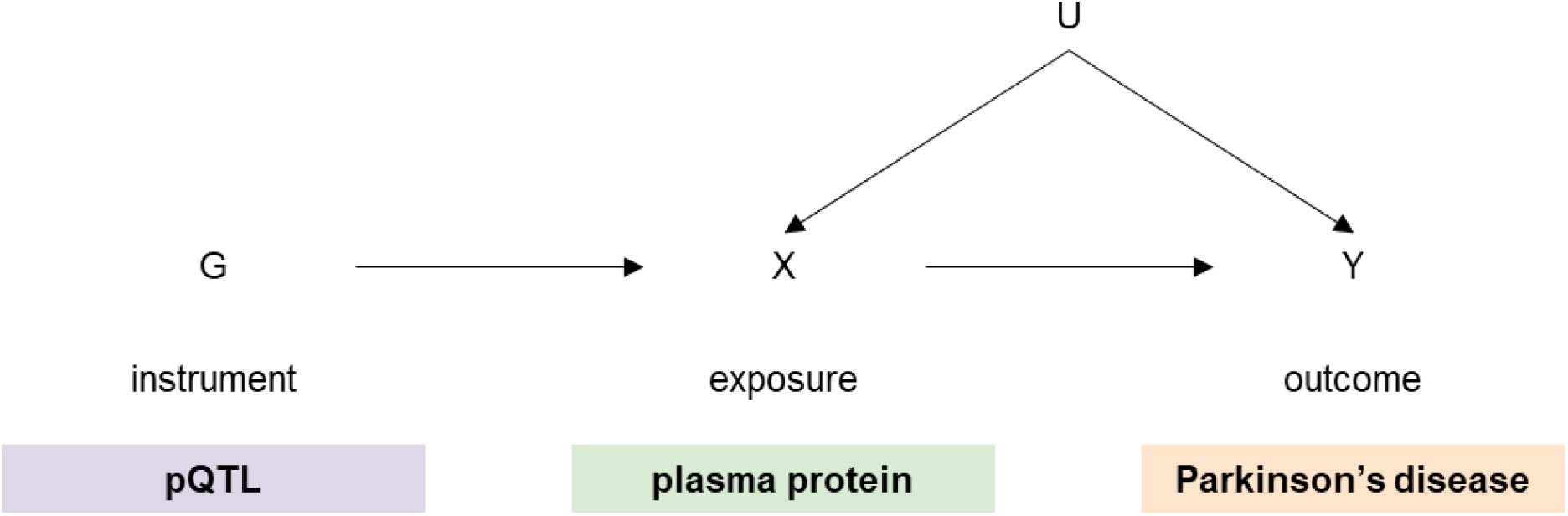
Directed acyclic graph illustrating the Mendelian randomisation framework. The diagram depicts the core assumption of a two-sample Mendelian randomisation analysis. Genetic variants associated with circulating protein levels (pQTLs) are used as instrumental variables to estimate the causal effect of plasma proteins (exposure) on Parkinson’s disease (outcome).

An additional assumption for two-sample MR is that the populations used for exposure and outcome GWAS must be comparable,^41^ i.e. drawn from similar underlying populations. Ancestry-specific validation of pQTLs ensures their relevance within the target populations.

### Exposure data: genetic associations of plasma proteins

#### European population

For Europeans, we accessed pQTL data from the UK Biobank Pharma Proteomics Project (UKB-PPP),^42^ which included 54,219 participants and measured 2,923 unique proteins using the Olink Explore 3072 PEA platform. Discovery and replication pQTL analyses were conducted in 34,557 and 17,806 individuals of European ancestry, respectively. Detailed methods for the UKB-PPP, including cohort selection and proteomics profiling, are described in the published paper.^42^

#### East Asian population

For East Asians, we combined pQTL data from two distinct sources–a subset of 262 individuals with East Asian ancestry from the UKB-PPP,^42^ and the Xu *et al*.^43^ cohort, which comprised 2,958 Han Chinese participants from the Guangzhou Nutrition and Health Study and Westlake Precision Birth Cohort, with 304 unique plasma proteins measured using data-independent acquisition mass spectrometry. We manually combined these datasets as they contained no overlapping single-nucleotide polymorphism (SNP)-protein associations. Although five proteins were common between datasets, there were no shared SNPs associated with these proteins. Similarly, one SNP appeared in both datasets but was associated with different proteins in each. From the combined data, we selected only proteins with genome-wide significant SNP associations, resulting in a final East Asian dataset of 3,220 participants with genetic associations for 229 unique proteins (including 74 unique plasma proteins from the Xu *et al*.^43^ cohort).

### Instrument selection for MR

IVs were carefully selected from genetic variants associated with plasma proteins (pQTLs). Selection followed a rigorous multi-step process. Initially, SNPs were identified using a genome-wide significance threshold (*P* < 5 × 10^-8^). To ensure independence, linkage disequilibrium (LD) clumping was performed with an r^2^ threshold of 0.01 and a physical distance threshold of 1000kb across each ancestry. For the European population, an in-house UK Biobank reference panel consisting of 40,510,321 variants from 4,990 individuals (2,341 males, 2,649 females) was used. For the East Asian population, we used the 1000 Genomes Phase 3 reference panel consisting of 23,117,731 variants from 504 individuals (244 males, 260 females). The strength of these IVs was assessed by calculating the proportion of variance (R^2^) explained by each protein’s IVs and evaluating the corresponding F-statistic. Only IVs with F-statistics exceeding 10 were retained to ensure sufficient statistical power for reliable causal inference. Both cis-acting and trans-acting pQTLs were included in our analyses.

### Outcome data: Genetic variants associated with Parkinson’s disease

#### European population

For Europeans, genetic association estimates for PD were obtained from publicly available GWAS summary statistics from Nalls *et al*.^30^ A comprehensive description of the study design and participant characteristics can be found in the original publication. This meta-analysis included data from 17 datasets involving European ancestry samples comprising 37,688 cases, 18,618 proxy cases, and 1.4 million controls. Ninety independent genome-wide significant loci associated with PD were identified.

#### East Asian population

For East Asians, we utilised PD GWAS summary statistics from Foo *et al*.^31^ which involved a meta-analysis in a cohort of 6,724 individuals with PD and 24,851 controls across six regions in East Asia (China, Taiwan, Hong Kong, South Korea, Singapore, Malaysia). Eleven genome-wide significant loci linked to PD were identified, including nine previously identified in Europeans.

### Mendelian randomisation analyses

MR analyses were performed using the TwoSampleMR package (version 0.4.22) in R (version 3.5.1). For each protein-PD association, we applied multiple MR methods to assess consistency and robustness. The Wald ratio was applied for single-SNP instruments and the inverse-variance weighted (IVW) method was the primary method for multi-SNP instruments. MR-Egger regression was employed to evaluate potential directional pleiotropy. To account for multiple testing, only associations passing a Benjamini-Hochberg false discovery rate (FDR) threshold of 0.05 were considered statistically significant. Effect estimates for PD risk were reported as odds ratios (ORs) with 95% confidence intervals, scaled to one standard deviation increase in protein levels. We reported protein-encoding genes in the results rather than protein names as these genes represent the functional genomic targets that encode the proteins of interest, facilitating direct translation to potential therapeutic targets while maintaining consistency with the genomic annotation in the datasets used for our analyses.

### Identification of ancestry-specific and cross-ancestry protein associations

To identify proteins associated with PD that are either shared between populations or specific to an ancestry, we conducted two sets of analyses. First, we performed ancestry-specific analysis, evaluating associations between all available proteins and PD within each population separately (Fig. 2). Second, we performed a shared protein analysis, restricting the evaluation to proteins that were present in both populations across the pQTL datasets for direct cross-population comparisons (Fig. 2). This dual approach allowed us to compare protein-PD associations across ancestries while capturing potential population-specific effects.

**Figure 2:**
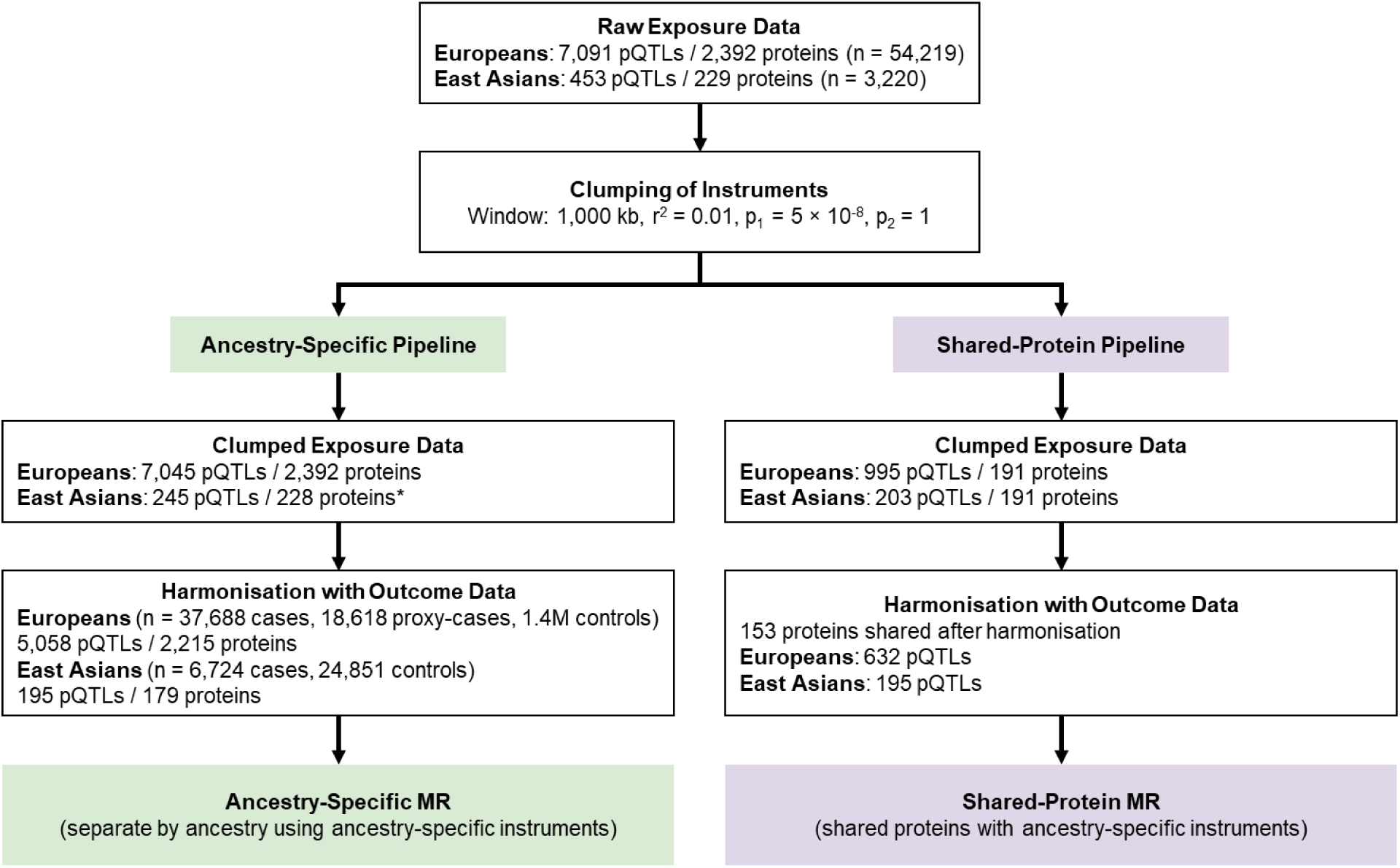
Overview of ancestry-specific and shared-protein Mendelian randomisation analyses. Summary characteristics of the European and East Asian datasets used in the ancestry-specific and shared-protein Mendelian randomisation analyses.

### Sensitivity analyses

To evaluate the robustness of our findings against potential violations of MR assumptions, several sensitivity analyses were conducted. Between-instrument heterogeneity was assessed using Cochran’s Q and I^2^ statistics to evaluate the consistency of causal estimates across individual IVs. Potential horizontal pleiotropy was examined using MR-Egger intercepts to detect directional pleiotropy, while the MR-PRESSO method was applied to identify and adjust for influential outlier IVs that might be influenced by horizontal pleiotropic effects.

### Triangulation with drug databases

We cross-referenced the causal proteins with therapeutic targets listed in multiple reference databases, including DrugBank^44^, ChEMBL,^44,45^ GeneCards,^46^ and UniProt.^46,47^ This systematic comparison aimed to identify proteins that are currently targeted by existing drugs or compounds, thereby highlighting potential candidates for repurposing in PD treatment. For proteins without known drug associations, we explored biological pathways and interactions relevant to PD pathogenesis to identify novel therapeutic opportunities. This triangulation approach^48–50^ not only facilitated the identification of existing therapeutic agents but also supported the discovery of novel drug targets based on protein functions implicated in PD pathogenesis. Findings from this triangulation were used to contextualise the potential clinical relevance of our results.

## RESULTS

There were notable differences in sample characteristics and IVs between European and East Asian populations. The European exposure dataset (Supplementary Table 1) had a higher average number of IVs per protein compared to the East Asian exposure dataset (Supplementary Table 2), reflecting differences in the availability and resolution of pQTL data. Table 1 summarises the key characteristics and outcomes of the MR analysis.

**Table 1:**
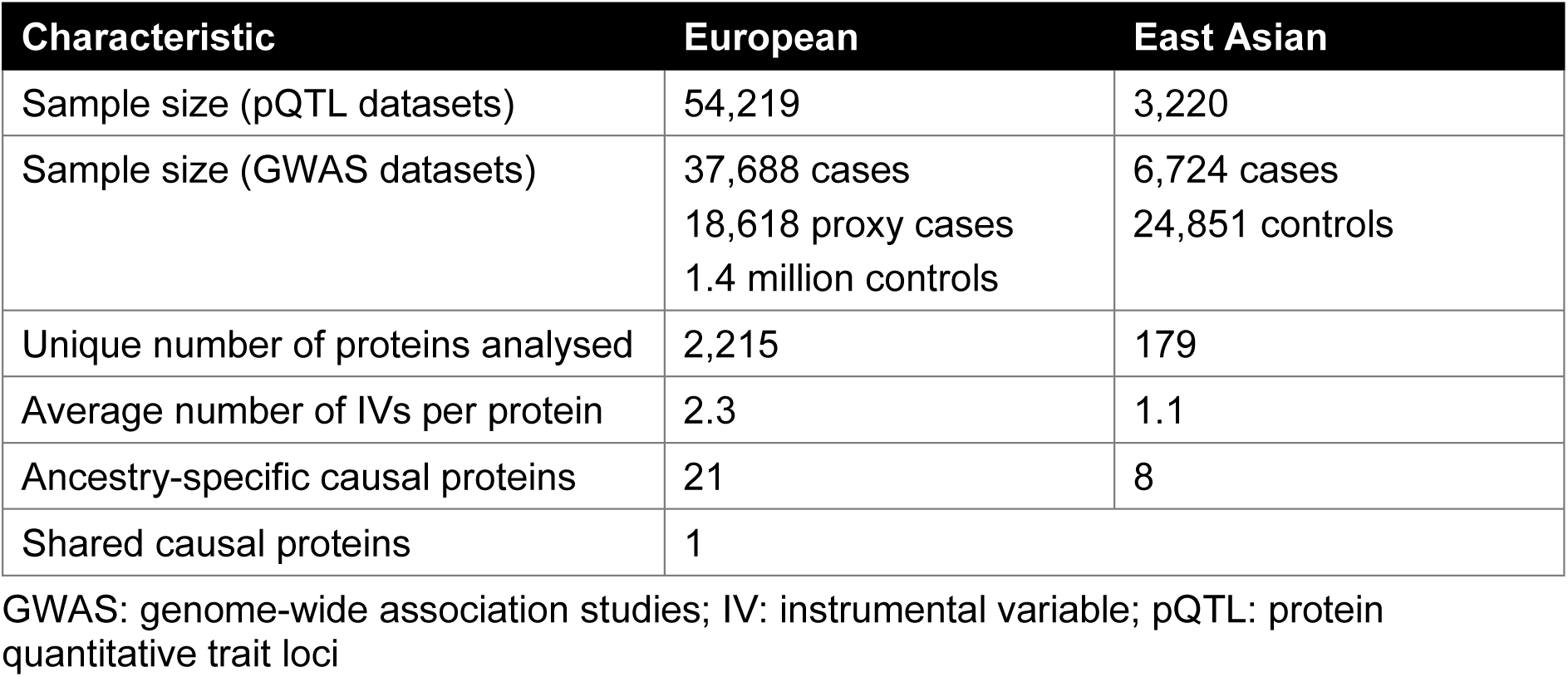
Overview of the Mendelian randomisation analysis.

### Ancestry-specific protein associations

The MR analysis identified distinct sets of blood-circulating proteins associated with PD risk in each ancestry. In Europeans, 21 proteins demonstrated causal associations with PD risk (Table 2 & Supplementary Table 3), whereas eight proteins were identified in East Asians (Table 3 & Supplementary Table 4). Only one protein-encoding gene, *BST1*, showed a shared association. The direction of effect for *BST1* was consistent across ancestries although East Asians exhibited a slightly greater magnitude (OR = 1.18, 95% CI 1.10, 1.27) compared to Europeans (OR = 1.04, 95% CI 1.02, 1.06).

**Table 2:** Associations of circulating proteins with Parkinson’s disease in Europeans.

| Protein-encoding gene | Number of IVs | Method<sup>a</sup> | OR (95% CI) | FDR | Egger intercept *P* |
| --- | --- | --- | --- | --- | --- |
| *AKT3* | 1 | Wald ratio | 2.12 (1.49, 3.01) | 0.010 | NA |
| *BST1* | 3 | IVW | 1.04 (1.02, 1.06) | 0.015 | NA |
| *C2CD2L* | 1 | Wald ratio | 2.45 (1.72, 3.48) | <0.001 | NA |
| *C5* | 1 | Wald ratio | 0.78 (0.69, 0.89) | 0.050 | NA |
| *CCN5* | 1 | Wald ratio | 2.24 (1.52, 3.29) | 0.014 | NA |
| *CD160* | 10 | IVW | 0.91 (0.87, 0.95) | 0.017 | 0.08 |
| *CDHR1* | 3 | IVW | 1.50 (1.27, 1.77) | 0.001 | NA |
| *EFCAB14* | 1 | Wald ratio | 4.89 (3.25, 7.36) | <0.001 | NA |
| *ENTPD2* | 1 | Wald ratio | 2.96 (2.04, 4.29) | <0.001 | NA |
| *FCGR2A* | 4 | IVW | 1.05 (1.03, 1.07) | <0.001 | 0.84 |
| *FCGR2B* | 2 | IVW | 0.87 (0.83, 0.90) | <0.001 | NA |
| *HIP1R* | 1 | Wald ratio | 4.74 (3.74, 6.01) | <0.001 | NA |
| *IDUA* | 10 | IVW | 0.92 (0.89, 0.95) | <0.001 | 0.19 |
| *INPP5J* | 1 | Wald ratio | 2.34 (1.72, 3.18) | <0.001 | NA |
| *MYLPF* | 1 | Wald ratio | 0.51 (0.36, 0.73) | 0.048 | NA |
| *PRSS53* | 7 | IVW | 0.92 (0.90, 0.94) | <0.001 | 0.59 |
| *TEF* | 1 | Wald ratio | 0.58 (0.45, 0.76) | 0.021 | NA |
| *TPM3* | 1 | Wald ratio | 0.53 (0.38, 0.74) | 0.042 | NA |
| *TRIM40* | 1 | Wald ratio | 1.24 (1.14, 1.34) | <0.001 | NA |
| *TSPAN7* | 1 | Wald ratio | 2.33 (1.62, 3.37) | 0.003 | NA |
| *TXNDC15* | 6 | IVW | 0.95 (0.92, 0.97) | 0.011 | 0.25 |

CI: confidence interval; FDR: false discovery rate; IV: instrumental variable; IVW: inverse-variance weighted; NA: not applicable; OR: odds ratio
<sup>a</sup>Results presented are limited to Wald ratio and inverse-variance weighted estimates. Results for other methods are available in the supplementary tables.

**Table 3:** Associations of circulating proteins with Parkinson’s disease in East Asians.

| Protein-encoding gene | Number of IVs | Method<sup>a</sup> | OR (95% CI) | FDR |
| --- | --- | --- | --- | --- |
| *APOM* | 1 | Wald ratio | 1.30 (1.10, 1.54) | 0.048 |
| *BST1* | 1 | Wald ratio | 1.18 (1.10, 1.27) | <0.001 |
| *C4A* | 3 | IVW | 0.84 (0.75, 0.93) | 0.045 |
| *C9orf131* | 2 | IVW | 0.80 (0.69, 0.92) | 0.045 |
| *HDGF* | 1 | Wald ratio | 1.11 (1.04, 1.19) | 0.044 |
| *MRPS34* | 2 | IVW | 0.80 (0.70, 0.93) | 0.048 |
| *PM20D1* | 1 | Wald ratio | 0.92 (0.88, 0.96) | 0.003 |
| *TOP1* | 1 | Wald ratio | 0.90 (0.85, 0.95) | 0.003 |

CI: confidence interval; FDR: false discovery rate; IV: instrumental variable; IVW: inverse-variance weighted; OR: odds ratio
<sup>a</sup>Results presented are limited to Wald ratio and inverse-variance weighted estimates. Results for other methods are available in the supplementary tables.

### Cross-ancestry analysis of shared protein associations

To enable direct comparisons of protein-PD associations across ancestries, we restricted the analysis to 191 proteins that were present in both the European and East Asian pQTL datasets. Following clumping and harmonisation, 153 shared proteins remained and were included in the MR analysis. On average, the European exposure dataset included 4.1 IVs per protein, whereas the East Asian dataset included 1.3 IVs per protein.

In Europeans, MR analysis identified *PRSS53* and *TXNDC15* as putatively causal proteins. In contrast, *HDGF*, *PM20D1*, and *TOP1* emerged as causal in East Asians (Supplementary Table 5). *BST1* was consistently associated with PD risk across both populations. Although *TXNDC15* exhibited a consistent direction of effect between ancestries, it did not reach statistical significance in East Asians after correction for multiple testing (FDR=0.09). Figure 3 illustrates the magnitude and direction of protein-PD associations across the two populations.

**Figure 3:**
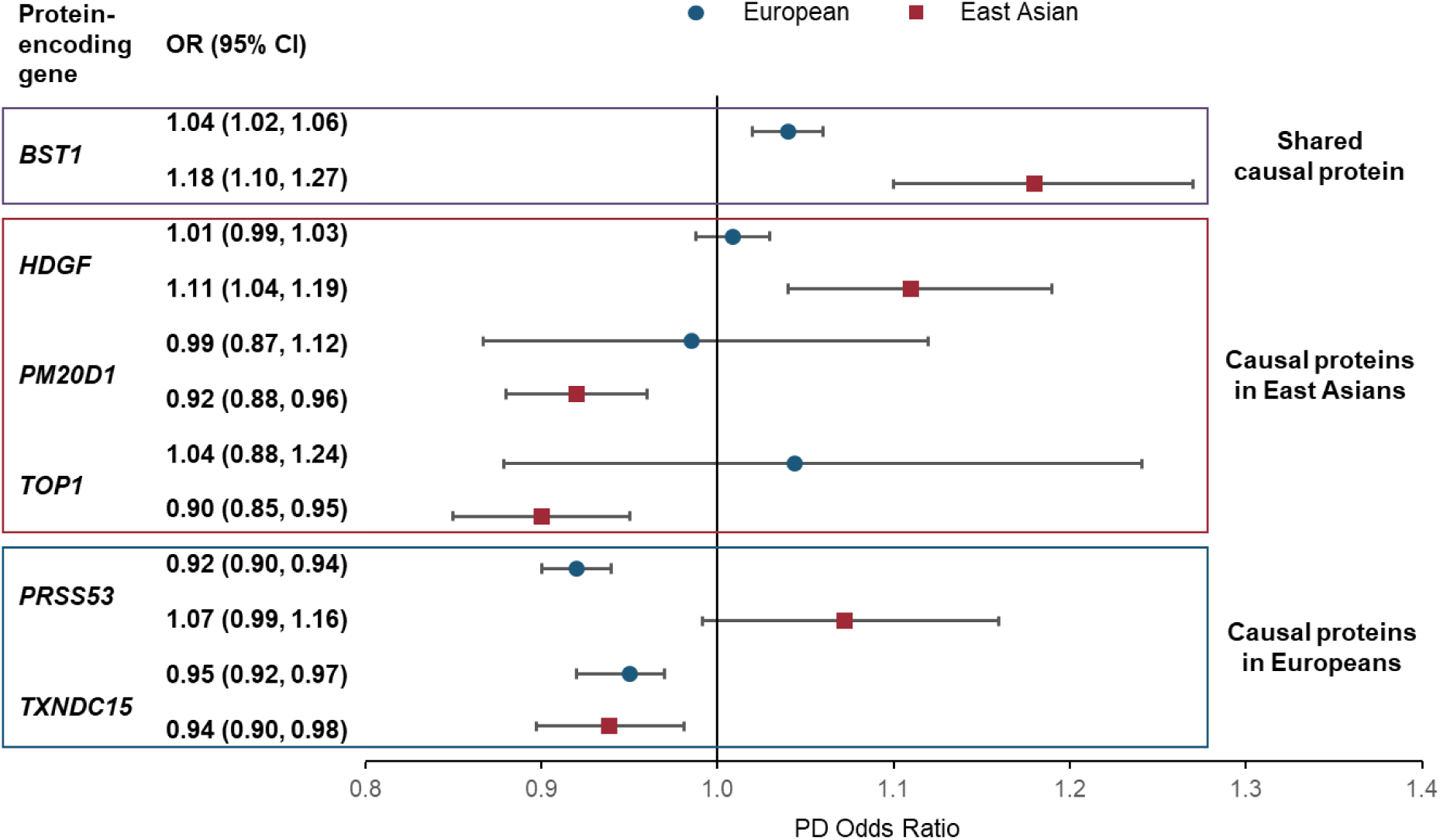
Forest plot of causal associations between plasma protein levels and Parkinson’s disease in the cross-ancestry Mendelian randomisation analyses. Point estimates represent the odds ratios for Parkinson’s disease per one standard deviation increase in circulating protein levels, with 95% confidence intervals. Results are stratified by ancestry to highlight cross-population consistency or heterogeneity.

### Sensitivity analyses

There was no evidence of substantial heterogeneity among the genetic instruments based on Cochran’s Q test across all evaluated exposures (Supplementary Table 6). Similarly, MR-Egger regression indicated no evidence of directional horizontal pleiotropy, particularly among exposures with a larger number of IVs (Supplementary Table 7). Given these findings, additional sensitivity analyses such as MR-PRESSO were not pursued. Overall, the results support the validity of the IVs and strengthen the causal interpretation of associations between the circulating proteins and PD risk. Due to the limited number of IVs per exposure, sensitivity analyses were not conducted in the East Asian populations.

### Triangulation with drug databases

Six protein-encoding genes identified in the shared protein analysis were evaluated for their therapeutic relevance (Table 4 & Supplementary Table 8). Several proteins are targets of drugs approved for non-PD indications or are under investigation in clinical trials for PD. *BST1*, which facilitates NAD+ nucleosidase activity, is targeted by nicotinamide, approved as an adjunctive therapy for macrocytic and secondary anaemia. Additionally, nicotinamide riboside, a precursor involved in NAD metabolism regulated by *BST1*, is being investigated in a phase 2 trial^46,47,51^ for its potential to delay the progression of early PD. *PM20D1*, involved in lipid and fatty acid metabolism, has been reported to show neuroprotective potential. Genetic studies suggest that increased *PM20D1* expression reduces Alzheimer’s disease pathology, while decreased expression exacerbates it,^52^ highlighting its potential as a therapeutic target for neurodegenerative diseases. Although no drugs currently target *PM20D1* specifically for PD, an investigational drug, JNJ-42165279, is undergoing a phase 2 trial^53^ for the potential treatment of major depressive disorder and anxiety—symptoms that are relatively common in individuals with PD and also recognised to be part of the PD prodrome in a substantial proportion of patients.^54,55^

**Table 4:** Drug triangulation of proteins identified with causal impact on Parkinson’s disease.

| Protein-encoding gene<sup>a</sup> | Protein | Drug name<sup>a,b</sup> | Clinical phase<sup>b</sup> |
| --- | --- | --- | --- |
| *BST1* | bone marrow stromal cell antigen 1 | a. nicotinamide<br>b. nicotinamide | a. Approved, investigational<br>b. Investigational (NCT03568968) |
| *HDGF* | hepatoma derived growth factor | a. copper | a. Approved, investigational<br>b. |
| *PM20D1* | Peptidase M20 Domain Containing 1 | a. cannabidiol<br>b. fostamatinib<br>c. JNJ-42165279 | a. Approved, investigational<br>b. Approved, investigational<br>c. Investigational (NCT02498392) |
| *PRSS53* | Serine Protease 53 | a. warfarin | a. Approved |
| *TOP1* | DNA Topoisomerase 1 | a. irinotecan<br>b. trastuzumab deruxtecan | a. Approved, investigational<br>b. Approved, investigational |
| *TXNDC15* | Thioredoxin Domain-Containing Protein 15 | None | NA |

NA: not applicable
<sup>a</sup>See Supplementary Tables for known function/gene ontology annotations of the protein-encoding genes, and indications of the related drugs.
<sup>b</sup>Data sources: DrugBank, EMBL-EBI, GeneCards, UniProt

## DISCUSSION

In this study, we integrated ancestry-specific proteomic data with GWAS data to investigate causal roles of circulating proteins in PD across European and East Asian populations using MR. We analysed 2,392 and 229 proteins in Europeans and East Asians respectively, identifying 21 and 8 proteins as causally associated with PD. Ancestry-common protein analysis revealed two shared causal protein-encoding genes: *BST1* and *TXNDC15* (the latter significant only nominally). Two additional protein-encoding genes showed European-specific effects, while three were East Asian-specific. Integration with clinical trial data and pharmacogenomic databases revealed several drug-repurposing candidates targeting identified protein pathways. Our results emphasise the value of ancestry-specific MR in understanding the genetic architecture of PD and the potential to inform the generalisability of therapeutic targets across diverse populations. These findings align with the recommendations from previous studies for prioritising diverse ancestry cohorts in genomics and other omics studies.

This study represents the first direct comparison of causal PD proteins between Europeans and East Asians. Most existing research, including pQTL or expression QTL (eQTL) studies, has focused on Europeans, limiting direct comparisons for East Asian populations. Our findings align and expand upon previous European studies.

A key finding in our study is the identification of *BST1* as a shared causal protein-encoding gene in both European and East Asian populations. Previous European MR studies^40,56^ reported an increased risk of PD with higher *BST1* levels, with comparable effect sizes to ours. However, studies evaluating both pQTL and eQTL effects^30,56^ indicated higher *BST1* expression associated with reduced PD risk and later age at onset, suggesting complex and context-dependent functional mechanisms. The pQTL-eQTL discrepancy highlights the need for additional research to better understand how *BST1* influences PD risk at genetic and protein expression levels. Limited eQTL-pQTL overlap indicates distinct mechanisms influencing protein levels and transcript abundance.^40,57^ This divergence, as shown in previous studies, suggests multi-level gene expression regulation, necessitating a direct proteomic analysis to fully capture these regulatory complexities.^40^ Given these findings, further exploration into the functional role of *BST1* in PD is important, particularly regarding immune-inflammatory processes.^58,59^ As an immune-associated gene implicated in neutrophil adhesion and migration, *BST1* may contribute to PD pathogenesis through neuroinflammatory mechanisms that affect dopaminergic neuron vulnerability.^58,59^

In our study, we also evaluated other previously reported causal PD protein-encoding genes in Europeans,^30,56^ including *CD38*, *CTSB*, *LGALS3* using blood pQTL data,^56^ and *GPNMB* and *PAM* using blood eQTL data (Supplementary Table 8).^30,56^ However, these protein-encoding genes did not meet the statistical significance threshold in our MR analysis. *CD38*, a *BST1* paralog, possesses similar ribosyl cyclase and cyclic ADP ribose hydrolase activities. Both influence calcium homeostasis by generating cyclic ADP-ribose, inducing calcium release from intracellular stores crucial for neurotransmitter release. Disruptions in calcium regulation could lead to dopaminergic neuronal death, a PD hallmark. While *CD38* and *BST1* share enzymatic functions, *BST1* appears to have distinct substrate preferences, highlighting potential differences in their roles in PD pathogenesis.

*TXNDC15* emerged as a potential shared causal protein-encoding gene between European and East Asian populations, although East Asian association did not reach nominal significance, likely due to reduced statistical power. Enhancer activity at rs11950533 locus tagging *TXNDC15* suggests regulatory potential with strong evidence of enhancer effects.^60^ *TXNDC15* was prioritised based on correlation with PD GWAS signals in cortical brain eQTL data,^61^ indicating possible regulatory relevance. While FOUNDIN-PD single-cell RNA sequencing data did not detect eQTL associations in dopaminergic neurons,^61^ its expression was observed, suggesting neuronal biological roles. The functional presence of *TXNDC15* in the brain was validated through glycopeptide detection in human brain tissue.^61,62^ The potential role of *TXNDC15* in PD pathogenesis likely involves thioredoxin domain-mediated cellular redox homeostasis maintenance.^63^ Given that oxidative stress is a well-established contributor to PD, the role of *TXNDC15* in mitigating this stress could be neuroprotective. These observations highlight the need for studies to explore spatial-temporal expression patterns in dopaminergic neurons and functional studies to elucidate its role in oxidative stress pathways.^63,64^

In East Asians, the identification of *PM20D1* as a causal protein-encoding gene adds new PD risk insights. Previous studies suggested that the *PM20D1*-N-Arachidonoyl Dopamine (NADA) pathway may play a protective role in PD dopamine metabolism regulation.^65^ *PM20D1* is situated within the PARK16 locus, and is associated with PD risk through SNPs within this locus that modulate its expression.^65,66^ *PM20D1* catalyses dopamine conversion to NADA, which inhibits alpha-synuclein aggregation, a hallmark of PD pathology. Furthermore, NADA regulates TRPV4-mediated calcium influx, potentially alleviating alpha-synuclein phosphorylation.^65^ Age-related declines in *PM20D1* expression and its therapeutic potential in synucleinopathy mouse models highlight its therapeutic value, especially since some loci linked to *PM20D1* are associated with ancestry-specific monogenic PD.^65^ Future studies should explore *PM20D1* activity enhancement for neuroprotection PD treatment development.

This study represents initial comprehensive protein-level PD risk factor comparisons across European and East Asian populations. We integrated large-scale proteomic and GWAS data, leveraging the most current and extensive datasets. MR provided robust causal estimates, and ancestry-specific analysis offers insights into population differences in PD pathogenesis. However, limitations must be acknowledged. First, East Asian pQTL and PD GWAS datasets were significantly smaller than European counterparts, limiting statistical power and the ability to detect associations. For instance, the number of causal protein-encoding genes identified in East Asians was three times lower than in Europeans, likely reflecting sample size disparities. Second, different proteomic platforms (antibody-based methods for Europeans and mass spectrometry for East Asians) may introduce technical variability that could influence cross-ancestry comparisons. Third, MR assumes genetic variants influence PD risk exclusively through their effects on protein levels, which is the exclusion restriction in MR. Violations of this assumption due to pleiotropy could potentially bias our results. Fourth, key PD-relevant gene products (e.g. α-synuclein, glucocerebrosidase) were underrepresented in the proteomic datasets utilised in our analysis, potentially explaining limited yield of casual proteins despite comprehensive approaches. Although we employed recent and largest available pQTL datasets, the technical limitations of current proteomic platforms may have constrained the detection of pathophysiologically significant protein associations. Finally, while plasma proteome reflects normal physiology and pathogenic processes, tissue-specific pQTL data (brain pQTLs) would improve the accuracy of our findings, but were not available for East Asians, limiting our analyses to blood pQTL data.

This study offers insights into ancestry-varied PD genetic factors, but important questions remain. Future research should understand specific biological mechanisms by which proteins influence PD risk through comprehensive functional studies incorporating in vitro and in vivo models. Exploring environmental factor interactions with protein-level risk factors is crucial for a more comprehensive understanding of PD aetiology, particularly as environmental exposures may differ across ancestries.

Replication in independent cohorts, particularly East Asians, is crucial for validating the generalisability of our results. Multi-ancestry proteomic and GWAS meta-analyses would increase statistical power and identify shared and ancestry-specific risk factors. Integration of protein-level discoveries with gene expression analysis and metabolite profiling could offer a more comprehensive understanding underlying PD pathogenesis. Laboratory investigations are essential to confirm the causal roles of identified proteins and assess their therapeutic target potential.

This study highlights the potential of circulating proteins as therapeutic targets and biomarkers for PD. The identification of *BST1* as a shared causal protein-encoding gene reinforces its value as a therapeutic target, supported by ongoing clinical investigation. Population-specific protein-encoding genes like *PM20D1* highlight the importance of genetic ancestry in risk assessment frameworks and may contribute to inclusive precision medicine approaches. Our findings emphasise the need for diversity in genetic and proteomic studies to ensure precision medicine advances benefit all populations. Such inclusivity is particularly important for diseases like PD where ancestry-specific factors significantly influence risk and treatment response.

In summary, our study provides valuable insights on common and population-specific protein-level PD risk factors in European and East Asian populations. These findings enhance our knowledge of PD pathogenesis and lay the groundwork for developing precise and ancestry-aware approaches to PD prevention, diagnosis, and treatment. As precision medicine advances in neurology, such ancestry-specific insights will be crucial for ensuring advances in PD management benefit patients across all populations.

## Data Availability

The data used in this study are provided in the supplementary tables. PD GWAS summary statistics for European and East Asian populations are available upon request to the respective authors. Plasma protein GWAS summary statistics for European and East Asian populations can be obtained from the previously published studies as referenced in the Methods section, of which the subsets are provided in the supplementary tables.

