## Supplementary Tables for "Proteomic and genetic insights into ancestry-specific associations in Parkinson’s disease"

**Supplementary Table 1: Exposure dataset in European populations**

| Protein-encoding gene | SNP | Other allele | Effect allele | Effect allele frequency | Beta | SE | p-value | F-statistic |
|---|---|---|---|---|---|---|---|---|
| *A1BG* | rs1260326 | T | C | 0.6084 | -0.137 | 0.007 | 6.3096E-80 | 383.04 |
| *A1BG* | rs35332062 | G | A | 0.1265 | -0.125 | 0.011 | 3.1623E-32 | 129.13 |
| *A1BG* | rs2723889 | T | C | 0.6728 | 0.053 | 0.007 | 1.5849E-12 | 57.33 |
| *A1BG* | rs145144275 | A | G | 0.009 | -2.134 | 0.038 | 1E-200 | 3153.71 |
| *AAMDC* | rs1354034 | T | C | 0.6045 | 0.055 | 0.007 | 2.5119E-15 | 61.73 |
| *AAMDC* | rs2186564 | G | A | 0.1062 | -1.064 | 0.012 | 1E-200 | 7861.78 |
| *AARSD1* | rs1354034 | T | C | 0.6046 | 0.067 | 0.007 | 7.9433E-20 | 91.61 |
| *AARSD1* | rs11170874 | A | G | 0.3309 | 0.053 | 0.008 | 1E-11 | 43.89 |
| *AARSD1* | rs323500 | C | T | 0.2899 | 0.132 | 0.008 | 2.5119E-61 | 272.25 |
| *ABHD14B* | rs373373105 | C | CG | 0.012 | -1.45 | 0.036 | 1E-200 | 1622.30 |
| *ABHD14B* | rs1354034 | T | C | 0.6039 | 0.056 | 0.008 | 3.1623E-13 | 49.00 |
| *ABHD14B* | rs6993770 | A | T | 0.2865 | -0.059 | 0.008 | 7.9433E-13 | 54.39 |
| *ABHD14B* | rs7080386 | C | A | 0.4119 | 0.058 | 0.008 | 1E-14 | 52.56 |
| *ABL1* | rs1354034 | T | C | 0.6046 | 0.082 | 0.008 | 3.1623E-27 | 105.06 |
| *ABO* | rs505922 | T | C | 0.3169 | 1.171 | 0.008 | 1E-200 | 21425.64 |
| *ABO* | rs17855739 | C | T | 0.0411 | 0.087 | 0.012 | 1E-12 | 52.56 |
| *ACAA1* | rs156265 | C | G | 0.1443 | -0.125 | 0.011 | 5.0119E-31 | 129.13 |
| *ACAA1* | rs4835265 | C | A | 0.1561 | 0.085 | 0.01 | 7.9433E-16 | 72.25 |
| *ACAA1* | rs11456266 | G | GT | 0.7556 | -0.072 | 0.009 | 7.9433E-16 | 64.00 |
| *ACADM* | rs61797339 | G | T | 0.3081 | 0.142 | 0.008 | 6.3096E-66 | 315.06 |
| *ACADSB* | rs1354034 | T | C | 0.6045 | 0.059 | 0.008 | 7.9433E-15 | 54.39 |
| *ACADSB* | rs11242109 | G | T | 0.4813 | -0.097 | 0.008 | 3.1623E-38 | 147.02 |
| *ACADSB* | rs342293 | C | G | 0.4592 | -0.059 | 0.007 | 2.5119E-15 | 71.04 |
| *ACADSB* | rs6993770 | A | T | 0.2866 | -0.09 | 0.008 | 1.2589E-27 | 126.56 |
| *ACADSB* | rs7896518 | A | G | 0.424 | 0.052 | 0.008 | 1.5849E-11 | 42.25 |
| *ACADSB* | rs7913063 | A | T | 0.0847 | 0.374 | 0.014 | 3.162E-167 | 713.65 |
| *ACAN* | rs1337526 | A | G | 0.8033 | 0.061 | 0.009 | 3.9811E-12 | 45.94 |
| *ACAN* | rs828602 | G | A | 0.4144 | 0.07 | 0.007 | 7.9433E-23 | 100.00 |
| *ACAN* | rs6461354 | C | T | 0.3935 | -0.05 | 0.007 | 1.9953E-12 | 51.02 |
| *ACAN* | rs12684083 | G | A | 0.2376 | -0.106 | 0.008 | 1.2589E-37 | 175.56 |
| *ACAN* | rs2519093 | C | T | 0.1845 | 0.092 | 0.009 | 2.5119E-24 | 104.49 |
| *ACAN* | rs56278466 | T | G | 0.6612 | 0.064 | 0.007 | 6.3096E-18 | 83.59 |
| *ACAN* | rs11000138 | A | T | 0.394 | -0.196 | 0.007 | 5.012E-160 | 784.00 |
| *ACAN* | rs895734 | C | T | 0.5884 | 0.09 | 0.007 | 7.9433E-37 | 165.31 |
| *ACAN* | rs59379014 | C | T | 0.0723 | 0.102 | 0.014 | 5.0119E-14 | 53.08 |
| *ACAN* | rs200201338 | TGAA | T | 0.0228 | 0.207 | 0.024 | 1.2589E-18 | 74.39 |
| *ACAN* | rs4932196 | T | C | 0.1911 | 0.134 | 0.009 | 1E-51 | 221.68 |
| *ACAN* | rs186021206 | G | A | 0.0065 | 0.355 | 0.048 | 1.2589E-13 | 54.70 |
| *ACAN* | rs55639531 | C | G | 0.285 | 0.075 | 0.008 | 6.3096E-22 | 87.89 |
| *ACAN* | rs17549173 | G | A | 0.0725 | 0.108 | 0.014 | 2.5119E-15 | 59.51 |
| *ACE* | rs34434834 | G | A | 0.0333 | 0.169 | 0.02 | 3.1623E-17 | 71.40 |
| *ACE* | rs186021206 | G | A | 0.0064 | 0.4 | 0.052 | 1.2589E-14 | 59.17 |
| *ACE* | rs4362 | T | C | 0.482 | -0.448 | 0.008 | 1E-200 | 3136.00 |
| *ACE2* | rs1047891 | C | A | 0.3133 | 0.052 | 0.007 | 1E-12 | 55.18 |
| *ACE2* | rs439651 | A | G | 0.5121 | 0.053 | 0.007 | 5.0119E-14 | 57.33 |
| *ACE2* | rs112875651 | G | A | 0.3906 | -0.078 | 0.007 | 1.2589E-28 | 124.16 |
| *ACE2* | rs11065384 | T | C | 0.6926 | 0.144 | 0.007 | 2.5119E-85 | 423.18 |
| *ACE2* | rs72694393 | G | T | 0.4794 | 0.06 | 0.007 | 1.9953E-18 | 73.47 |
| *ACE2* | rs28929474 | C | T | 0.0208 | 0.219 | 0.024 | 3.1623E-20 | 83.27 |
| *ACE2* | rs550999044 | A | AT | 0.2433 | 0.067 | 0.008 | 3.1623E-17 | 70.14 |
| *ACE2* | rs72787359 | A | C | 0.0734 | -0.096 | 0.013 | 3.1623E-13 | 54.53 |
| *ACHE* | rs2269615 | C | T | 0.5599 | -0.07 | 0.007 | 3.1623E-23 | 100.00 |
| *ACHE* | rs7636227 | G | A | 0.5532 | 0.051 | 0.007 | 3.9811E-13 | 53.08 |
| *ACHE* | rs35762167 | T | C | 0.5827 | -0.107 | 0.007 | 3.1623E-51 | 233.65 |
| *ACHE* | rs6976053 | C | T | 0.4706 | 0.313 | 0.007 | 1E-200 | 1999.37 |
| *ACHE* | rs2519093 | C | T | 0.1846 | -0.173 | 0.009 | 3.1623E-81 | 369.49 |
| *ACHE* | rs186021206 | G | A | 0.0064 | 0.496 | 0.048 | 3.9811E-25 | 106.78 |
| *ACOT13* | rs1354034 | T | C | 0.6046 | 0.068 | 0.008 | 7.9433E-19 | 72.25 |
| *ACOT13* | rs10058074 | G | A | 0.4751 | -0.056 | 0.008 | 1.5849E-13 | 49.00 |
| *ACOT13* | rs143667676 | G | T | 0.0021 | -1.055 | 0.082 | 1.9953E-37 | 165.53 |
| *ACOT13* | rs6993770 | A | T | 0.2866 | -0.072 | 0.008 | 6.3096E-18 | 81.00 |
| *ACOT13* | rs6580981 | G | A | 0.459 | 0.056 | 0.008 | 1.9953E-13 | 49.00 |
| *ACOX1* | rs10852766 | T | C | 0.6591 | 0.078 | 0.008 | 7.9433E-22 | 95.06 |
| *ACP1* | rs11553746 | C | T | 0.3549 | -0.379 | 0.008 | 1E-200 | 2244.39 |

| | | | | | | | | |
|---|---|---|---|---|---|---|---|---|
| *ACP5* | rs61801010 | A | G | 0.1061 | 0.105 | 0.012 | 1.9953E-19 | 76.56 |
| *ACP5* | rs9693404 | G | C | 0.1883 | 0.079 | 0.009 | 3.1623E-19 | 77.05 |
| *ACP5* | rs10813949 | G | A | 0.3103 | -0.057 | 0.007 | 1.5849E-14 | 66.31 |
| *ACP5* | rs35382467 | G | A | 0.0985 | -0.093 | 0.011 | 3.1623E-16 | 71.48 |
| *ACP5* | rs117566084 | C | T | 0.0099 | 0.364 | 0.034 | 5.0119E-26 | 114.62 |
| *ACP5* | rs1958559 | C | T | 0.602 | 0.055 | 0.007 | 3.1623E-15 | 61.73 |
| *ACP5* | rs2073333 | C | T | 0.2528 | 0.109 | 0.008 | 1E-43 | 185.64 |
| *ACP5* | rs78755089 | G | T | 0.0845 | 0.108 | 0.012 | 1.5849E-18 | 81.00 |
| *ACP5* | rs186021206 | G | A | 0.0063 | 0.332 | 0.047 | 1.9953E-12 | 49.90 |
| *ACP5* | rs2305799 | C | T | 0.1233 | -0.504 | 0.011 | 1E-200 | 2099.31 |
| *ACP6* | rs2999719 | T | C | 0.9291 | 0.249 | 0.015 | 3.9811E-60 | 275.56 |
| *ACP6* | rs77542178 | T | C | 0.0207 | -0.181 | 0.027 | 1E-11 | 44.94 |
| *ACP6* | rs2153463 | T | G | 0.7017 | 1.014 | 0.008 | 1E-200 | 16065.56 |
| *ACRBP* | rs342298 | C | T | 0.455 | -0.053 | 0.008 | 2.5119E-12 | 43.89 |
| *ACRBP* | rs6993770 | A | T | 0.2864 | -0.065 | 0.008 | 3.9811E-15 | 66.02 |
| *ACRBP* | rs150230220 | A | AT | 0.3111 | 0.077 | 0.008 | 5.0119E-21 | 92.64 |
| *ACRBP* | rs7959658 | A | G | 0.3611 | 0.161 | 0.008 | 1.2589E-93 | 405.02 |
| *ACRBP* | rs892090 | T | G | 0.8342 | 0.069 | 0.01 | 1E-11 | 47.61 |
| *ACRV1* | rs113811036 | CTT | C | 0.1931 | 0.066 | 0.006 | 1.2589E-25 | 121.00 |
| *ACRV1* | rs369963387 | G | A | 0.7288 | -0.042 | 0.006 | 1.5849E-11 | 49.00 |
| *ACSL1* | rs56295533 | G | A | 0.2064 | 0.096 | 0.011 | 1.2589E-18 | 76.17 |
| *ACTA2* | rs1324551 | C | T | 0.4988 | -0.06 | 0.007 | 1E-18 | 73.47 |
| *ACVRL1* | rs75581187 | C | T | 0.0718 | -0.094 | 0.014 | 5.0119E-12 | 45.08 |
| *ACVRL1* | rs9264623 | T | C | 0.6391 | -0.057 | 0.007 | 6.3096E-15 | 66.31 |
| *ACVRL1* | rs11244079 | G | A | 0.0503 | 0.11 | 0.016 | 6.3096E-12 | 47.27 |
| *ACVRL1* | rs2277382 | C | T | 0.0801 | 0.568 | 0.013 | 1E-200 | 1909.02 |
| *ACVRL1* | rs2837988 | C | A | 0.3617 | 0.082 | 0.007 | 1.5849E-28 | 137.22 |
| *ACY1* | rs1260326 | T | C | 0.6083 | -0.071 | 0.007 | 2.5119E-22 | 102.88 |
| *ACY1* | rs121912698 | C | T | 0.0044 | -1.692 | 0.054 | 1E-200 | 981.78 |
| *ACY1* | rs3747207 | G | A | 0.2153 | 0.077 | 0.009 | 5.0119E-19 | 73.20 |
| *ACY3* | rs542733750 | C | CT | 0.2985 | -0.058 | 0.008 | 1.5849E-12 | 52.56 |
| *ACY3* | rs6894249 | A | G | 0.387 | -0.063 | 0.008 | 6.3096E-17 | 62.02 |
| *ACY3* | rs1964528 | C | T | 0.1132 | -0.091 | 0.012 | 6.3096E-15 | 57.51 |
| *ACY3* | rs112875651 | G | A | 0.3916 | -0.053 | 0.008 | 5.0119E-12 | 43.89 |
| *ACY3* | rs3758938 | T | G | 0.3124 | 0.274 | 0.008 | 1E-200 | 1173.06 |
| *ACY3* | rs6057570 | A | T | 0.8278 | 0.073 | 0.01 | 7.9433E-14 | 53.29 |
| *ACY3* | rs738409 | C | G | 0.217 | 0.064 | 0.009 | 5.0119E-13 | 50.57 |
| *ACYP1* | rs1354034 | T | C | 0.6044 | 0.066 | 0.007 | 6.3096E-19 | 88.90 |
| *ACYP1* | rs6993770 | A | T | 0.2871 | -0.058 | 0.008 | 6.3096E-13 | 52.56 |
| *ACYP1* | rs2393967 | A | C | 0.314 | 0.056 | 0.008 | 1.5849E-12 | 49.00 |
| *ACYP1* | rs75948138 | C | G | 0.011 | -1.263 | 0.036 | 1E-200 | 1230.84 |
| *ADA* | rs140373387 | G | C | 0.0048 | 0.441 | 0.049 | 3.9811E-19 | 81.00 |
| *ADA* | rs35350651 | A | AC | 0.504 | -0.049 | 0.007 | 3.1623E-14 | 49.00 |
| *ADA* | rs11555566 | T | C | 0.0626 | 1.28 | 0.015 | 1E-200 | 7281.78 |
| *ADA2* | rs72930772 | C | T | 0.3351 | 0.079 | 0.008 | 7.9433E-25 | 97.52 |
| *ADA2* | rs148819155 | A | G | 0.0033 | -0.431 | 0.064 | 1.2589E-11 | 45.35 |
| *ADA2* | rs2903386 | T | C | 0.3713 | 0.058 | 0.007 | 7.9433E-15 | 68.65 |
| *ADA2* | rs2548999 | G | T | 0.6621 | 0.051 | 0.008 | 1.2589E-11 | 40.64 |
| *ADA2* | rs535777 | G | C | 0.1431 | 0.156 | 0.01 | 2.5119E-50 | 243.36 |
| *ADA2* | rs56278466 | T | G | 0.661 | 0.076 | 0.008 | 1E-23 | 90.25 |
| *ADA2* | rs12575367 | A | G | 0.2992 | 0.062 | 0.008 | 2.5119E-15 | 60.06 |
| *ADA2* | rs226379 | T | C | 0.355 | -0.051 | 0.008 | 1.5849E-11 | 40.64 |
| *ADA2* | rs117566084 | C | T | 0.0098 | 0.314 | 0.036 | 6.3096E-18 | 76.08 |
| *ADA2* | rs3184504 | T | C | 0.517 | -0.129 | 0.007 | 1.2589E-72 | 339.61 |
| *ADA2* | rs76428106 | T | C | 0.0128 | 0.224 | 0.033 | 1.2589E-11 | 46.08 |
| *ADA2* | rs145078947 | G | T | 0.0025 | 0.554 | 0.072 | 1E-14 | 59.20 |
| *ADA2* | rs186021206 | G | A | 0.0064 | 0.669 | 0.049 | 7.9433E-42 | 186.41 |
| *ADA2* | rs485186 | A | G | 0.5381 | 0.05 | 0.007 | 5.0119E-12 | 51.02 |
| *ADA2* | rs737963 | T | C | 0.6116 | 0.207 | 0.008 | 3.981E-164 | 669.52 |
| *ADA2* | rs738409 | C | G | 0.2166 | 0.078 | 0.009 | 1.9953E-18 | 75.11 |
| *ADAM12* | rs11244909 | T | C | 0.6094 | 0.129 | 0.008 | 1E-61 | 260.02 |
| *ADAM15* | rs11589479 | G | A | 0.1625 | 1.024 | 0.01 | 1E-200 | 10485.76 |
| *ADAM15* | rs146142570 | G | A | 0.0024 | 0.513 | 0.065 | 1.9953E-15 | 62.29 |
| *ADAM15* | rs77468121 | G | A | 0.0442 | 0.117 | 0.015 | 1.9953E-15 | 60.84 |
| *ADAM15* | rs62571868 | A | T | 0.2322 | 0.049 | 0.007 | 1E-11 | 49.00 |
| *ADAM15* | rs8176759 | G | A | 0.0614 | 0.507 | 0.013 | 1E-200 | 1521.00 |
| *ADAM15* | rs56278466 | T | G | 0.661 | 0.043 | 0.006 | 7.9433E-12 | 51.36 |
| *ADAM15* | rs3184504 | T | C | 0.5173 | -0.061 | 0.006 | 3.1623E-24 | 103.36 |

| | | | | | | | | |
|---|---|---|---|---|---|---|---|---|
| *ADAM15* | rs186021206 | G | A | 0.0063 | 0.687 | 0.042 | 3.1623E-61 | 267.56 |
| *ADAM22* | rs61747728 | C | T | 0.0379 | 0.169 | 0.018 | 6.3096E-22 | 88.15 |
| *ADAM22* | rs75166367 | G | A | 0.0604 | -0.112 | 0.014 | 2.5119E-15 | 64.00 |
| *ADAM22* | rs2279542 | C | G | 0.5051 | -0.565 | 0.008 | 1E-200 | 4987.89 |
| *ADAM22* | rs186021206 | G | A | 0.0064 | 0.332 | 0.046 | 7.9433E-13 | 52.09 |
| *ADAM22* | rs2659005 | C | T | 0.4411 | 0.051 | 0.007 | 1E-13 | 53.08 |
| *ADAM23* | rs61747728 | C | T | 0.038 | 0.121 | 0.016 | 1E-13 | 57.19 |
| *ADAM23* | rs72933203 | T | A | 0.103 | 0.878 | 0.012 | 1E-200 | 5353.36 |
| *ADAM23* | rs1294438 | C | T | 0.3591 | 0.056 | 0.007 | 2.5119E-17 | 64.00 |
| *ADAM23* | rs8176741 | G | A | 0.0613 | 0.222 | 0.013 | 7.9433E-64 | 291.62 |
| *ADAM23* | rs186021206 | G | A | 0.0064 | 0.482 | 0.043 | 6.3096E-29 | 125.65 |
| *ADAM8* | rs11806326 | T | G | 0.4776 | 0.07 | 0.007 | 6.3096E-22 | 100.00 |
| *ADAM8* | rs2596530 | G | A | 0.4936 | -0.059 | 0.007 | 1.5849E-16 | 71.04 |
| *ADAM8* | rs2995310 | C | T | 0.8941 | -0.709 | 0.012 | 1E-200 | 3490.84 |
| *ADAM8* | rs4055121 | C | T | 0.135 | -0.086 | 0.011 | 2.5119E-16 | 61.12 |
| *ADAM8* | rs3184504 | T | C | 0.5172 | -0.106 | 0.007 | 5.0119E-50 | 229.31 |
| *ADAM8* | rs76428106 | T | C | 0.0129 | 0.255 | 0.033 | 7.9433E-15 | 59.71 |
| *ADAM8* | rs186021206 | G | A | 0.0064 | 0.567 | 0.049 | 5.0119E-31 | 133.90 |
| *ADAM8* | rs2227319 | G | A | 0.3709 | -0.076 | 0.007 | 5.0119E-25 | 117.88 |
| *ADAM9* | rs9267123 | G | C | 0.1584 | 0.137 | 0.01 | 5.0119E-41 | 187.69 |
| *ADAM9* | rs61753672 | G | A | 0.0049 | 0.923 | 0.053 | 2.5119E-67 | 303.29 |
| *ADAM9* | rs507666 | G | A | 0.1844 | -0.08 | 0.01 | 1.2589E-16 | 64.00 |
| *ADAM9* | rs34307716 | G | A | 0.2758 | 0.132 | 0.008 | 3.9811E-56 | 272.25 |
| *ADAM9* | rs877081 | G | A | 0.1376 | -0.083 | 0.011 | 1.2589E-14 | 56.93 |
| *ADAM9* | rs186021206 | G | A | 0.0065 | 0.545 | 0.051 | 7.9433E-27 | 114.20 |
| *ADAMTS13* | rs4282054 | T | C | 0.559 | 0.049 | 0.007 | 1.2589E-11 | 49.00 |
| *ADAMTS13* | rs5023357 | T | G | 0.1083 | 0.103 | 0.011 | 1.9953E-19 | 87.68 |
| *ADAMTS13* | rs2910686 | T | C | 0.4498 | 0.075 | 0.007 | 1.5849E-25 | 114.80 |
| *ADAMTS13* | rs9266217 | A | G | 0.6084 | 0.052 | 0.008 | 6.3096E-12 | 42.25 |
| *ADAMTS13* | rs143073120 | C | CACTT | 0.5619 | -0.049 | 0.007 | 1E-11 | 49.00 |
| *ADAMTS13* | rs10253113 | A | G | 0.3074 | 0.066 | 0.008 | 1.2589E-17 | 68.06 |
| *ADAMTS13* | rs9987289 | A | G | 0.9079 | -0.096 | 0.012 | 6.3096E-15 | 64.00 |
| *ADAMTS13* | rs74715985 | A | G | 0.017 | -1.459 | 0.029 | 1E-200 | 2531.13 |
| *ADAMTS13* | rs1389065 | G | A | 0.7637 | -0.059 | 0.008 | 3.9811E-12 | 54.39 |
| *ADAMTS13* | rs746775574 | TACTAAG | T | 0.0926 | -0.102 | 0.012 | 1.5849E-16 | 72.25 |
| *ADAMTS13* | rs768453105 | GTTATGGT/ | G | 0.1823 | 0.069 | 0.009 | 7.9433E-14 | 58.78 |
| *ADAMTS13* | rs738408 | C | T | 0.2171 | 0.064 | 0.009 | 1.2589E-13 | 50.57 |
| *ADAMTS15* | rs5030062 | A | C | 0.3729 | -0.067 | 0.008 | 1.2589E-17 | 70.14 |
| *ADAMTS15* | rs4861708 | G | A | 0.5118 | -0.124 | 0.008 | 2.5119E-60 | 240.25 |
| *ADAMTS15* | rs2545801 | T | C | 0.7449 | -0.097 | 0.009 | 3.1623E-29 | 116.16 |
| *ADAMTS15* | rs73044885 | G | A | 0.0549 | 0.193 | 0.017 | 1E-28 | 128.89 |
| *ADAMTS15* | rs186021206 | G | A | 0.0063 | 0.522 | 0.052 | 1.5849E-23 | 100.77 |
| *ADAMTS16* | rs547625232 | T | TA | 0.4535 | -0.051 | 0.007 | 1.2589E-12 | 53.08 |
| *ADAMTS16* | rs40468 | A | G | 0.3383 | -0.123 | 0.007 | 5.0119E-61 | 308.76 |
| *ADAMTS4* | rs4656993 | A | G | 0.5857 | -0.118 | 0.008 | 7.9433E-54 | 217.56 |
| *ADAMTS4* | rs61658003 | C | A | 0.8424 | 0.07 | 0.01 | 1.5849E-11 | 49.00 |
| *ADAMTS8* | rs7529464 | G | C | 0.2494 | 0.065 | 0.008 | 3.9811E-16 | 66.02 |
| *ADAMTS8* | rs13107325 | C | T | 0.0749 | -0.144 | 0.013 | 3.1623E-28 | 122.70 |
| *ADAMTS8* | rs2131535 | G | A | 0.7313 | 0.493 | 0.008 | 1E-200 | 3797.64 |
| *ADAMTS8* | rs7209484 | T | C | 0.2421 | 0.06 | 0.008 | 1E-13 | 56.25 |
| *ADAMTSL2* | rs61830291 | A | C | 0.0973 | 0.141 | 0.013 | 3.1623E-29 | 117.64 |
| *ADAMTSL2* | rs34466290 | T | TA | 0.6687 | -0.061 | 0.008 | 1.9953E-14 | 58.14 |
| *ADAMTSL2* | rs9987289 | A | G | 0.9078 | -0.09 | 0.013 | 1.9953E-12 | 47.93 |
| *ADAMTSL2* | rs1008250 | A | G | 0.2821 | 0.106 | 0.009 | 1.5849E-35 | 138.72 |
| *ADAMTSL2* | rs168960 | A | G | 0.9196 | -0.104 | 0.013 | 1.5849E-14 | 64.00 |
| *ADAMTSL2* | rs28929474 | C | T | 0.0208 | 0.242 | 0.026 | 1E-20 | 86.63 |
| *ADAMTSL2* | rs1801689 | A | C | 0.0301 | 0.148 | 0.022 | 7.9433E-12 | 45.26 |
| *ADAMTSL2* | rs77542162 | A | G | 0.0229 | 0.167 | 0.025 | 1.5849E-11 | 44.62 |
| *ADAMTSL2* | rs73045269 | C | T | 0.1713 | 0.096 | 0.01 | 1.9953E-22 | 92.16 |
| *ADAMTSL4* | rs758689051 | G | T | 0.0018 | -2.569 | 0.098 | 3.162E-150 | 687.19 |
| *ADAMTSL4* | rs6746339 | C | T | 0.0106 | -0.309 | 0.036 | 1E-17 | 73.67 |
| *ADAMTSL4* | rs13107325 | C | T | 0.075 | -0.105 | 0.014 | 6.3096E-14 | 56.25 |
| *ADAMTSL4* | rs2954017 | T | C | 0.5192 | -0.064 | 0.007 | 5.0119E-18 | 83.59 |
| *ADAMTSL5* | rs773726167 | GC | G | 0.1775 | -0.152 | 0.01 | 5.0119E-57 | 231.04 |
| *ADAMTSL5* | rs791461 | T | C | 0.532 | 0.406 | 0.008 | 1E-200 | 2575.56 |
| *ADD1* | rs1354034 | T | C | 0.6044 | 0.074 | 0.007 | 1.2589E-23 | 111.76 |
| *ADD1* | rs1263339 | G | C | 0.1865 | -0.177 | 0.009 | 6.3096E-80 | 386.78 |
| *ADD1* | rs2501577 | A | G | 0.1689 | -0.135 | 0.01 | 7.9433E-44 | 182.25 |

| | | | | | | | | |
|---|---|---|---|---|---|---|---|---|
| *ADGRB3* | rs3798969 | A | G | 0.6235 | -0.941 | 0.008 | 1E-200 | 13835.64 |
| *ADGRB3* | rs200489612 | G | A | 0.0052 | 0.294 | 0.038 | 7.9433E-15 | 59.86 |
| *ADGRB3* | rs573431210 | T | G | 0.0266 | 0.117 | 0.016 | 7.9433E-14 | 53.47 |
| *ADGRD1* | rs1260326 | T | C | 0.6081 | -0.064 | 0.006 | 3.9811E-23 | 113.78 |
| *ADGRD1* | rs6706968 | A | C | 0.5809 | -0.051 | 0.006 | 1.5849E-15 | 72.25 |
| *ADGRD1* | rs687339 | C | T | 0.7713 | 0.086 | 0.008 | 3.9811E-30 | 115.56 |
| *ADGRD1* | rs35332062 | G | A | 0.1269 | -0.085 | 0.009 | 2.5119E-19 | 89.20 |
| *ADGRD1* | rs28601761 | C | G | 0.419 | -0.057 | 0.006 | 1.9953E-18 | 90.25 |
| *ADGRD1* | rs1870138 | A | G | 0.7961 | 0.07 | 0.008 | 1.9953E-19 | 76.56 |
| *ADGRD1* | rs174533 | G | A | 0.3468 | 0.069 | 0.007 | 1.9953E-25 | 97.16 |
| *ADGRD1* | rs11045856 | T | G | 0.2407 | 0.07 | 0.008 | 7.9433E-18 | 76.56 |
| *ADGRD1* | rs1169296 | A | G | 0.3256 | 0.058 | 0.007 | 3.9811E-15 | 68.65 |
| *ADGRD1* | rs4759533 | G | A | 0.9466 | 0.824 | 0.015 | 1E-200 | 3017.67 |
| *ADGRD1* | rs2239222 | A | G | 0.3482 | 0.047 | 0.007 | 1.5849E-12 | 45.08 |
| *ADGRD1* | rs186021206 | G | A | 0.0064 | 0.43 | 0.043 | 3.9811E-23 | 100.00 |
| *ADGRD1* | rs1801689 | A | C | 0.0299 | 0.144 | 0.018 | 6.3096E-15 | 64.00 |
| *ADGRD1* | rs1800961 | C | T | 0.0321 | -0.165 | 0.018 | 2.5119E-20 | 84.03 |
| *ADGRE1* | rs12616639 | A | G | 0.611 | 0.088 | 0.007 | 3.1623E-32 | 158.04 |
| *ADGRE1* | rs1257169 | C | A | 0.499 | -0.113 | 0.007 | 1.5849E-54 | 260.59 |
| *ADGRE1* | rs6440016 | C | A | 0.1491 | -0.083 | 0.01 | 3.9811E-16 | 68.89 |
| *ADGRE1* | rs35005076 | G | GC | 0.198 | 0.066 | 0.009 | 3.9811E-13 | 53.78 |
| *ADGRE1* | rs115216147 | T | A | 0.0665 | 0.118 | 0.015 | 1.5849E-15 | 61.88 |
| *ADGRE1* | rs4934470 | T | C | 0.7257 | -0.059 | 0.008 | 3.9811E-13 | 54.39 |
| *ADGRE1* | rs4601794 | A | G | 0.0588 | -0.184 | 0.015 | 3.9811E-33 | 150.47 |
| *ADGRE1* | rs3184504 | T | C | 0.5172 | -0.104 | 0.007 | 3.9811E-47 | 220.73 |
| *ADGRE1* | rs186021206 | G | A | 0.0065 | 1.096 | 0.05 | 5.012E-108 | 480.49 |
| *ADGRE1* | rs2305482 | A | C | 0.5394 | -0.05 | 0.007 | 3.9811E-12 | 51.02 |
| *ADGRE1* | rs1292061 | A | G | 0.5537 | 0.076 | 0.007 | 7.9433E-26 | 117.88 |
| *ADGRE1* | rs2725405 | G | C | 0.441 | 0.054 | 0.007 | 1.5849E-13 | 59.51 |
| *ADGRE1* | rs59055395 | T | C | 0.1125 | 0.444 | 0.012 | 1E-200 | 1369.00 |
| *ADGRE2* | rs61801010 | A | G | 0.106 | 0.1 | 0.012 | 1.2589E-16 | 69.44 |
| *ADGRE2* | rs1260326 | T | C | 0.6086 | 0.054 | 0.007 | 1E-13 | 59.51 |
| *ADGRE2* | rs78776915 | C | T | 0.4154 | 0.057 | 0.008 | 1.2589E-13 | 50.77 |
| *ADGRE2* | rs56278466 | T | G | 0.6615 | 0.119 | 0.007 | 1.2589E-57 | 289.00 |
| *ADGRE2* | rs950802 | G | A | 0.3064 | -0.06 | 0.008 | 6.3096E-15 | 56.25 |
| *ADGRE2* | rs3184504 | T | C | 0.5173 | -0.097 | 0.007 | 3.1623E-43 | 192.02 |
| *ADGRE2* | rs186021206 | G | A | 0.0064 | 1.09 | 0.048 | 3.981E-112 | 515.67 |
| *ADGRE2* | rs7251607 | A | C | 0.7529 | 0.287 | 0.008 | 1E-200 | 1287.02 |
| *ADGRE5* | rs61801010 | A | G | 0.1061 | 0.102 | 0.012 | 5.0119E-17 | 72.25 |
| *ADGRE5* | rs1260326 | T | C | 0.6085 | 0.056 | 0.007 | 2.5119E-14 | 64.00 |
| *ADGRE5* | rs34458031 | G | A | 0.0858 | 0.087 | 0.013 | 7.9433E-12 | 44.79 |
| *ADGRE5* | rs10093 | C | G | 0.3766 | 0.065 | 0.008 | 1E-17 | 66.02 |
| *ADGRE5* | rs56278466 | T | G | 0.6616 | 0.138 | 0.008 | 6.3096E-75 | 297.56 |
| *ADGRE5* | rs950802 | G | A | 0.3067 | -0.069 | 0.008 | 6.3096E-19 | 74.39 |
| *ADGRE5* | rs3184504 | T | C | 0.517 | -0.102 | 0.007 | 1E-46 | 212.33 |
| *ADGRE5* | rs186021206 | G | A | 0.0064 | 1.178 | 0.049 | 1.995E-126 | 577.96 |
| *ADGRE5* | rs139113505 | C | T | 0.0169 | -0.715 | 0.028 | 1.995E-143 | 652.07 |
| *ADGRE5* | rs6065904 | G | A | 0.2152 | 0.065 | 0.009 | 7.9433E-14 | 52.16 |
| *ADGRF5* | rs76183323 | G | A | 0.0105 | -0.844 | 0.038 | 6.31E-108 | 493.31 |
| *ADGRF5* | rs13412535 | G | A | 0.2311 | -0.06 | 0.009 | 3.9811E-12 | 44.44 |
| *ADGRF5* | rs11719450 | C | T | 0.3696 | 0.074 | 0.008 | 1.9953E-22 | 85.56 |
| *ADGRF5* | rs34716199 | A | AT | 0.5356 | 0.052 | 0.007 | 1.2589E-12 | 55.18 |
| *ADGRF5* | rs4098923 | A | G | 0.4478 | -0.079 | 0.008 | 7.9433E-25 | 97.52 |
| *ADGRF5* | rs342299 | C | T | 0.4546 | -0.063 | 0.007 | 6.3096E-18 | 81.00 |
| *ADGRF5* | rs927826 | G | T | 0.7052 | 0.187 | 0.008 | 6.31E-121 | 546.39 |
| *ADGRF5* | rs7896518 | A | G | 0.424 | 0.089 | 0.007 | 1.2589E-32 | 161.65 |
| *ADGRF5* | rs111338191 | A | AAATT | 0.5207 | -0.061 | 0.007 | 5.0119E-17 | 75.94 |
| *ADGRF5* | rs12445050 | C | T | 0.1383 | 0.088 | 0.01 | 3.9811E-17 | 77.44 |
| *ADGRF5* | rs892090 | T | G | 0.8342 | 0.123 | 0.01 | 2.5119E-36 | 151.29 |
| *ADGRF5* | rs1033549 | G | T | 0.3779 | 0.053 | 0.007 | 1.9953E-12 | 57.33 |
| *ADGRG1* | rs1047891 | C | A | 0.3125 | 0.063 | 0.008 | 5.0119E-16 | 62.02 |
| *ADGRG1* | rs10935473 | G | T | 0.4404 | -0.079 | 0.007 | 1E-27 | 127.37 |
| *ADGRG1* | rs28393425 | A | G | 0.1807 | -0.071 | 0.009 | 6.3096E-14 | 62.23 |
| *ADGRG1* | rs2519093 | C | T | 0.1845 | -0.113 | 0.009 | 6.3096E-34 | 157.64 |
| *ADGRG1* | rs56278466 | T | G | 0.6611 | 0.133 | 0.008 | 3.1623E-69 | 276.39 |
| *ADGRG1* | rs376888155 | G | GA | 0.3075 | -0.065 | 0.008 | 1E-16 | 66.02 |
| *ADGRG1* | rs3184504 | T | C | 0.5168 | -0.08 | 0.007 | 2.5119E-29 | 130.61 |
| *ADGRG1* | rs733464 | C | T | 0.4066 | 0.071 | 0.007 | 3.9811E-22 | 102.88 |

| | | | | | | | | |
|---|---|---|---|---|---|---|---|---|
| *ADGRG1* | rs186021206 | G | A | 0.0063 | 0.473 | 0.05 | 1.5849E-21 | 89.49 |
| *ADGRG1* | rs738409 | C | G | 0.2172 | 0.06 | 0.009 | 3.9811E-12 | 44.44 |
| *ADGRG2* | rs755740174 | AT | A | 0.291 | 0.073 | 0.008 | 7.9433E-22 | 83.27 |
| *ADGRG2* | rs13107325 | C | T | 0.0751 | 0.098 | 0.013 | 7.9433E-14 | 56.83 |
| *ADGRG2* | rs115912456 | A | G | 0.0407 | -0.148 | 0.017 | 1.9953E-17 | 75.79 |
| *ADGRG2* | rs8176719 | T | TC | 0.3401 | 0.051 | 0.007 | 1.5849E-12 | 53.08 |
| *ADGRG2* | rs74612335 | T | C | 0.1331 | -0.09 | 0.01 | 1E-18 | 81.00 |
| *ADGRG2* | rs186021206 | G | A | 0.0064 | 0.52 | 0.047 | 2.5119E-28 | 122.41 |
| *ADGRV1* | rs1031101 | A | G | 0.1422 | -0.165 | 0.011 | 3.9811E-50 | 225.00 |
| *ADH1B* | rs41275699 | A | G | 0.0022 | -1.293 | 0.09 | 3.1623E-47 | 206.40 |
| *ADH1B* | rs4979373 | C | T | 0.4905 | -0.071 | 0.007 | 1E-21 | 102.88 |
| *ADH1B* | rs10883451 | T | C | 0.5018 | -0.053 | 0.007 | 6.3096E-13 | 57.33 |
| *ADH1B* | rs10405357 | T | C | 0.4353 | -0.069 | 0.007 | 5.0119E-20 | 97.16 |
| *ADH1B* | rs3747207 | G | A | 0.2151 | 0.098 | 0.009 | 5.0119E-28 | 118.57 |
| *ADH4* | rs1800759 | T | G | 0.6131 | 0.13 | 0.007 | 5.0119E-68 | 344.90 |
| *ADH4* | rs112875651 | G | A | 0.3906 | -0.056 | 0.008 | 1.2589E-13 | 49.00 |
| *ADH4* | rs10733608 | G | T | 0.4889 | -0.063 | 0.007 | 6.3096E-18 | 81.00 |
| *ADH4* | rs10883451 | T | C | 0.5019 | -0.051 | 0.007 | 2.5119E-12 | 53.08 |
| *ADH4* | rs3747207 | G | A | 0.2157 | 0.102 | 0.009 | 3.1623E-31 | 128.44 |
| *ADIPOQ* | rs72657272 | C | T | 0.1526 | 0.063 | 0.009 | 1.9953E-12 | 49.00 |
| *ADIPOQ* | rs66782572 | A | G | 0.5388 | 0.059 | 0.007 | 1.5849E-19 | 71.04 |
| *ADIPOQ* | rs143257534 | C | T | 0.026 | -0.615 | 0.02 | 1.995E-199 | 945.56 |
| *ADIPOQ* | rs7859344 | T | A | 0.7148 | -0.048 | 0.007 | 6.3096E-12 | 47.02 |
| *ADIPOQ* | rs150090666 | C | T | 0.0009 | 0.872 | 0.116 | 6.3096E-14 | 56.51 |
| *ADIPOQ* | rs7134375 | C | A | 0.4298 | 0.065 | 0.006 | 7.9433E-24 | 117.36 |
| *ADIPOQ* | rs825453 | A | T | 0.6081 | -0.066 | 0.007 | 6.3096E-24 | 88.90 |
| *ADIPOQ* | rs2925979 | T | C | 0.698 | 0.092 | 0.007 | 6.3096E-40 | 172.73 |
| *ADIPOQ* | rs10422861 | C | T | 0.6668 | 0.065 | 0.007 | 5.0119E-22 | 86.22 |
| *ADM* | rs150777506 | T | TTATACTGA | 0.0102 | -0.505 | 0.035 | 3.9811E-47 | 208.18 |
| *ADM* | rs2403310 | G | C | 0.4723 | 0.16 | 0.007 | 5.012E-113 | 522.45 |
| *AFAP1* | rs41264705 | C | T | 0.2976 | 1.083 | 0.008 | 1E-200 | 18326.39 |
| *AFAP1* | rs3134935 | T | C | 0.2798 | 0.046 | 0.007 | 5.0119E-12 | 43.18 |
| *AFAP1* | rs36051450 | T | C | 0.1955 | -0.048 | 0.007 | 7.9433E-12 | 47.02 |
| *AFM* | rs553977556 | G | A | 0.0013 | -2.142 | 0.108 | 1.9953E-87 | 393.36 |
| *AFM* | rs10083143 | A | C | 0.0358 | 0.157 | 0.02 | 3.9811E-15 | 61.62 |
| *AFM* | rs200293726 | A | T | 0.306 | -0.07 | 0.008 | 6.3096E-18 | 76.56 |
| *AFP* | rs12059956 | G | A | 0.4218 | -0.047 | 0.007 | 1.2589E-11 | 45.08 |
| *AFP* | rs4149909 | A | G | 0.032 | -0.144 | 0.02 | 2.5119E-13 | 51.84 |
| *AFP* | rs11900031 | A | G | 0.1055 | -0.414 | 0.012 | 1E-200 | 1190.25 |
| *AFP* | rs62280667 | T | C | 0.3408 | -0.129 | 0.007 | 2.5119E-69 | 339.61 |
| *AFP* | rs59950280 | G | A | 0.3295 | 0.092 | 0.008 | 5.0119E-34 | 132.25 |
| *AFP* | rs12506899 | T | G | 0.3969 | -0.186 | 0.007 | 5.012E-150 | 706.04 |
| *AFP* | rs562329399 | T | TGG | 0.5769 | -0.072 | 0.007 | 1.5849E-24 | 105.80 |
| *AFP* | rs2045028 | A | G | 0.4808 | -0.047 | 0.007 | 1.5849E-11 | 45.08 |
| *AFP* | rs62576492 | A | G | 0.1127 | 0.134 | 0.011 | 7.9433E-33 | 148.40 |
| *AFP* | rs35751397 | T | TA | 0.4686 | -0.049 | 0.007 | 7.9433E-13 | 49.00 |
| *AFP* | rs10883567 | T | C | 0.4911 | -0.06 | 0.007 | 3.1623E-18 | 73.47 |
| *AFP* | rs11418587 | T | TA | 0.3342 | 0.064 | 0.008 | 1.2589E-14 | 64.00 |
| *AFP* | rs7310409 | A | G | 0.6235 | 0.105 | 0.007 | 2.5119E-49 | 225.00 |
| *AFP* | rs560191 | G | C | 0.2989 | 0.122 | 0.008 | 5.0119E-59 | 232.56 |
| *AFP* | rs144268704 | A | AGGTTTT | 0.3359 | 0.056 | 0.007 | 1.9953E-14 | 64.00 |
| *AFP* | rs17689024 | G | C | 0.3136 | -0.053 | 0.007 | 1.2589E-12 | 57.33 |
| *AFP* | rs11642010 | T | C | 0.0887 | -0.084 | 0.012 | 5.0119E-12 | 49.00 |
| *AFP* | rs4794048 | A | C | 0.481 | 0.071 | 0.007 | 1.2589E-24 | 102.88 |
| *AFP* | rs34349423 | A | AC | 0.6253 | -0.058 | 0.007 | 2.5119E-15 | 68.65 |
| *AFP* | rs1801689 | A | C | 0.0301 | -0.17 | 0.02 | 3.9811E-17 | 72.25 |
| *AFP* | rs35103294 | C | T | 0.6311 | 0.05 | 0.007 | 3.9811E-12 | 51.02 |
| *AFP* | rs2548458 | C | T | 0.5389 | -0.049 | 0.007 | 2.5119E-12 | 49.00 |
| *AFP* | rs6116934 | T | C | 0.2148 | 0.095 | 0.008 | 1.2589E-29 | 141.02 |
| *AFP* | rs113844904 | C | T | 0.1566 | -0.069 | 0.009 | 2.5119E-13 | 58.78 |
| *AFP* | rs6007594 | G | A | 0.2574 | 0.131 | 0.008 | 5.0119E-62 | 268.14 |
| *AGER* | rs66906568 | T | C | 0.2989 | -0.078 | 0.008 | 6.3096E-22 | 95.06 |
| *AGER* | rs10753239 | T | C | 0.4603 | -0.055 | 0.007 | 2.5119E-13 | 61.73 |
| *AGER* | rs6664840 | A | G | 0.4713 | -0.077 | 0.007 | 7.9433E-26 | 121.00 |
| *AGER* | rs2137715 | A | C | 0.3872 | 0.065 | 0.007 | 3.9811E-18 | 86.22 |
| *AGER* | rs13186378 | C | G | 0.3184 | 0.077 | 0.008 | 1.2589E-22 | 92.64 |
| *AGER* | rs204993 | A | G | 0.3199 | -0.17 | 0.008 | 2.512E-103 | 451.56 |
| *AGER* | rs7823498 | C | T | 0.7795 | 0.073 | 0.009 | 1E-16 | 65.79 |

| | | | | | | | | |
|---|---|---|---|---|---|---|---|---|
| *AGER* | rs11243421 | T | C | 0.562 | -0.131 | 0.007 | 1.2589E-70 | 350.22 |
| *AGER* | rs2487928 | G | A | 0.4482 | -0.056 | 0.007 | 3.1623E-14 | 64.00 |
| *AGER* | rs3028371 | CAATAATA. | C | 0.5318 | -0.053 | 0.007 | 1.2589E-12 | 57.33 |
| *AGER* | rs35393359 | C | T | 0.1701 | -0.092 | 0.01 | 2.5119E-21 | 84.64 |
| *AGER* | rs11407068 | T | TA | 0.78 | 0.064 | 0.009 | 1.9953E-12 | 50.57 |
| *AGER* | rs1896354 | A | G | 0.429 | 0.067 | 0.007 | 1.2589E-19 | 91.61 |
| *AGER* | rs55703124 | T | C | 0.7319 | -0.064 | 0.008 | 1.2589E-14 | 64.00 |
| *AGER* | rs10152355 | C | A | 0.4531 | 0.065 | 0.008 | 1.2589E-17 | 66.02 |
| *AGER* | rs34262842 | A | G | 0.1664 | -0.068 | 0.01 | 5.0119E-12 | 46.24 |
| *AGR2* | rs115317719 | G | T | 0.1874 | 0.078 | 0.01 | 1.9953E-15 | 60.84 |
| *AGR2* | rs7807123 | C | T | 0.5664 | 0.107 | 0.008 | 7.9433E-44 | 178.89 |
| *AGR2* | rs1580180 | T | C | 0.666 | 0.06 | 0.008 | 7.9433E-14 | 56.25 |
| *AGR2* | rs28633254 | T | C | 0.4174 | -0.057 | 0.008 | 2.5119E-13 | 50.77 |
| *AGR2* | rs601338 | G | A | 0.5092 | -0.059 | 0.008 | 1.9953E-14 | 54.39 |
| *AGR3* | rs58198139 | C | T | 0.6382 | 0.079 | 0.008 | 3.1623E-23 | 97.52 |
| *AGR3* | rs9271956 | T | C | 0.8336 | 0.072 | 0.01 | 7.9433E-12 | 51.84 |
| *AGR3* | rs34954997 | C | CTTCG | 0.2375 | 0.079 | 0.009 | 3.1623E-18 | 77.05 |
| *AGR3* | rs12983058 | G | T | 0.3266 | -0.058 | 0.008 | 1E-12 | 52.56 |
| *AGRN* | rs3121560 | C | A | 0.3405 | -0.378 | 0.008 | 1E-200 | 2232.56 |
| *AGRN* | rs551319785 | CA | C | 0.1397 | 0.099 | 0.012 | 3.9811E-16 | 68.06 |
| *AGRN* | rs13107325 | C | T | 0.0748 | 0.098 | 0.014 | 7.9433E-13 | 49.00 |
| *AGRN* | rs7654228 | T | C | 0.4099 | -0.087 | 0.007 | 1.5849E-32 | 154.47 |
| *AGRN* | rs4841133 | A | G | 0.908 | -0.086 | 0.012 | 2.5119E-12 | 51.36 |
| *AGRN* | rs881930 | G | A | 0.1719 | -0.085 | 0.01 | 2.5119E-19 | 72.25 |
| *AGRN* | rs8668 | A | G | 0.3032 | 0.12 | 0.008 | 3.9811E-53 | 225.00 |
| *AGRP* | rs12239737 | T | A | 0.3373 | -0.053 | 0.008 | 1.2589E-11 | 43.89 |
| *AGRP* | rs1260326 | T | C | 0.6085 | -0.053 | 0.007 | 1.5849E-12 | 57.33 |
| *AGRP* | rs778393561 | A | G | 0.3539 | -0.081 | 0.009 | 1.2589E-20 | 81.00 |
| *AGRP* | rs56393506 | C | T | 0.1721 | 0.092 | 0.01 | 1.2589E-20 | 84.64 |
| *AGRP* | rs80026582 | C | T | 0.0843 | -0.112 | 0.013 | 1.5849E-17 | 74.22 |
| *AGRP* | rs7850767 | C | T | 0.4871 | 0.052 | 0.007 | 1.9953E-12 | 55.18 |
| *AGRP* | rs964184 | G | C | 0.8668 | -0.148 | 0.011 | 1.9953E-43 | 181.02 |
| *AGRP* | rs188707867 | C | G | 0.0071 | -0.469 | 0.046 | 1E-24 | 103.95 |
| *AGRP* | rs740516 | C | G | 0.1484 | -0.077 | 0.01 | 1E-13 | 59.29 |
| *AGRP* | rs34954997 | C | CTTCG | 0.2374 | 0.122 | 0.009 | 6.3096E-46 | 183.75 |
| *AGT* | rs4762 | G | A | 0.1168 | 0.744 | 0.012 | 1E-200 | 3844.00 |
| *AGT* | rs62466318 | C | T | 0.2016 | -0.085 | 0.009 | 6.3096E-22 | 89.20 |
| *AGXT* | rs1497406 | A | G | 0.5796 | 0.064 | 0.007 | 1.9953E-18 | 83.59 |
| *AGXT* | rs4361977 | C | T | 0.4338 | -0.063 | 0.007 | 7.9433E-18 | 81.00 |
| *AGXT* | rs55649245 | G | A | 0.31 | -0.288 | 0.008 | 1E-200 | 1296.00 |
| *AGXT* | rs4835265 | C | A | 0.1562 | 0.13 | 0.01 | 7.9433E-39 | 169.00 |
| *AGXT* | rs112875651 | G | A | 0.391 | -0.053 | 0.008 | 3.1623E-12 | 43.89 |
| *AGXT* | rs10883451 | T | C | 0.5018 | -0.051 | 0.007 | 1.9953E-12 | 53.08 |
| *AGXT* | rs28929474 | C | T | 0.0208 | 0.198 | 0.025 | 6.3096E-15 | 62.73 |
| *AGXT* | rs4503880 | T | C | 0.7993 | -0.102 | 0.009 | 6.3096E-29 | 128.44 |
| *AGXT* | rs10405357 | T | C | 0.4354 | -0.057 | 0.007 | 1.5849E-14 | 66.31 |
| *AGXT* | rs3747207 | G | A | 0.2153 | 0.08 | 0.009 | 7.9433E-20 | 79.01 |
| *AHCY* | rs142714816 | A | G | 0.0008 | 0.918 | 0.133 | 5.0119E-12 | 47.64 |
| *AHCY* | rs13043752 | G | A | 0.0159 | -1.375 | 0.03 | 1E-200 | 2100.69 |
| *AHNAK* | rs74853209 | T | C | 0.028 | -0.473 | 0.022 | 6.31E-99 | 462.25 |
| *AHNAK2* | rs9805957 | A | C | 0.6425 | 0.216 | 0.008 | 1.585E-163 | 729.00 |
| *AHSG* | rs1900618 | C | T | 0.6401 | 0.676 | 0.008 | 1E-200 | 7140.25 |
| *AHSG* | rs28929474 | C | T | 0.0209 | 0.164 | 0.023 | 6.3096E-13 | 50.84 |
| *AHSP* | rs875741 | G | A | 0.4073 | -0.052 | 0.007 | 5.0119E-13 | 55.18 |
| *AHSP* | rs80215559 | T | C | 0.0769 | 0.133 | 0.013 | 1.2589E-23 | 104.67 |
| *AHSP* | rs11759553 | A | T | 0.2718 | -0.06 | 0.008 | 1E-13 | 56.25 |
| *AHSP* | rs590856 | G | A | 0.5528 | -0.079 | 0.007 | 7.9433E-28 | 127.37 |
| *AHSP* | rs6592965 | G | A | 0.4538 | -0.067 | 0.007 | 1E-20 | 91.61 |
| *AHSP* | rs10742583 | G | A | 0.8473 | 0.117 | 0.01 | 1.9953E-30 | 136.89 |
| *AHSP* | rs570013781 | G | A | 0.0024 | -0.814 | 0.078 | 1E-25 | 108.91 |
| *AHSP* | rs771133280 | TC | T | 0.2022 | 0.084 | 0.009 | 1.5849E-20 | 87.11 |
| *AHSP* | rs1000821 | C | T | 0.4654 | -0.112 | 0.007 | 1.2589E-55 | 256.00 |
| *AHSP* | rs390408 | A | G | 0.821 | 0.088 | 0.009 | 1E-20 | 95.60 |
| *AHSP* | rs855791 | A | G | 0.5606 | 0.079 | 0.007 | 1E-28 | 127.37 |
| *AIDA* | rs607929 | C | G | 0.4485 | -0.073 | 0.008 | 1.2589E-20 | 83.27 |
| *AIF1* | rs2261033 | A | G | 0.4307 | -0.111 | 0.008 | 7.9433E-47 | 192.52 |
| *AIF1L* | rs9275036 | T | C | 0.6697 | -0.059 | 0.008 | 1.9953E-12 | 54.39 |
| *AIF1L* | rs7856365 | G | A | 0.6845 | -0.311 | 0.008 | 1E-200 | 1511.27 |

| | | | | | | | | |
|---|---|---|---|---|---|---|---|---|
| *AIFM1* | rs1354034 | T | C | 0.6046 | 0.053 | 0.008 | 3.9811E-12 | 43.89 |
| *AIFM1* | rs6993770 | A | T | 0.2864 | -0.058 | 0.008 | 3.9811E-12 | 52.56 |
| *AK2* | rs138577419 | G | C | 0.0026 | -0.771 | 0.074 | 1.2589E-25 | 108.55 |
| *AKR1B1* | rs2229542 | T | C | 0.0135 | -0.587 | 0.033 | 3.1623E-69 | 316.41 |
| *AKR1B1* | rs6993770 | A | T | 0.2869 | -0.068 | 0.008 | 1.5849E-15 | 72.25 |
| *AKR1B1* | rs72481564 | C | T | 0.2266 | 0.083 | 0.01 | 5.0119E-16 | 68.89 |
| *AKR1C4* | rs11253048 | A | G | 0.177 | -0.068 | 0.01 | 1.5849E-11 | 46.24 |
| *AKR7L* | rs59622577 | C | T | 0.0974 | -0.317 | 0.013 | 5.012E-133 | 594.61 |
| *AKR7L* | rs58534292 | T | C | 0.65 | -0.066 | 0.008 | 1.9953E-16 | 68.06 |
| *AKR7L* | rs34160075 | G | A | 0.6278 | -0.074 | 0.008 | 1.2589E-18 | 85.56 |
| *AKR7L* | rs3747207 | G | A | 0.2152 | 0.076 | 0.009 | 1.2589E-16 | 71.31 |
| *AKT1S1* | rs1354034 | T | C | 0.6035 | 0.067 | 0.008 | 3.1623E-18 | 70.14 |
| *AKT1S1* | rs342298 | C | T | 0.4544 | -0.056 | 0.008 | 1.5849E-13 | 49.00 |
| *AKT1S1* | rs10733789 | T | C | 0.3133 | 0.066 | 0.008 | 3.9811E-16 | 68.06 |
| *AKT2* | rs1354034 | T | C | 0.6043 | 0.057 | 0.008 | 1.2589E-13 | 50.77 |
| *AKT2* | rs2706396 | A | C | 0.571 | 0.056 | 0.008 | 2.5119E-13 | 49.00 |
| *AKT2* | rs6993770 | A | T | 0.2864 | -0.068 | 0.008 | 1.5849E-16 | 72.25 |
| *AKT3* | rs4515770 | T | C | 0.2179 | 0.065 | 0.009 | 3.1623E-12 | 52.16 |
| *AKT3* | rs7916868 | A | T | 0.5019 | 0.055 | 0.008 | 7.9433E-13 | 47.27 |
| *ALCAM* | rs34926152 | G | T | 0.0485 | -0.303 | 0.016 | 5.0119E-80 | 358.63 |
| *ALCAM* | rs9274626 | T | C | 0.679 | -0.058 | 0.008 | 1.9953E-14 | 52.56 |
| *ALCAM* | rs199922514 | A | AAGAAGAA | 0.9076 | -0.111 | 0.012 | 6.3096E-21 | 85.56 |
| *ALCAM* | rs10901252 | G | C | 0.0615 | 0.102 | 0.014 | 1.2589E-12 | 53.08 |
| *ALCAM* | rs59822672 | G | C | 0.076 | -0.094 | 0.013 | 6.3096E-13 | 52.28 |
| *ALCAM* | rs11045856 | T | G | 0.2403 | 0.078 | 0.008 | 5.0119E-22 | 95.06 |
| *ALCAM* | rs904628 | G | A | 0.4763 | -0.059 | 0.007 | 1.2589E-17 | 71.04 |
| *ALCAM* | rs12448760 | G | A | 0.2861 | 0.096 | 0.008 | 1.9953E-36 | 144.00 |
| *ALCAM* | rs186021206 | G | A | 0.0064 | 0.85 | 0.047 | 3.9811E-72 | 327.07 |
| *ALCAM* | rs7224668 | T | C | 0.5574 | -0.051 | 0.007 | 3.1623E-13 | 53.08 |
| *ALCAM* | rs2638282 | G | A | 0.5328 | 0.059 | 0.007 | 1.2589E-17 | 71.04 |
| *ALCAM* | rs2294915 | C | T | 0.2317 | 0.078 | 0.008 | 7.9433E-22 | 95.06 |
| *ALDH1A1* | rs112596047 | A | T | 0.2655 | 0.091 | 0.008 | 1E-28 | 129.39 |
| *ALDH1A1* | rs3747207 | G | A | 0.2152 | 0.074 | 0.009 | 1.2589E-17 | 67.60 |
| *ALDH3A1* | rs887241 | A | C | 0.6575 | 0.484 | 0.008 | 1E-200 | 3660.25 |
| *ALDH5A1* | rs2744581 | A | G | 0.5189 | -0.141 | 0.008 | 5.0119E-77 | 310.64 |
| *ALPI* | rs550057 | C | T | 0.2561 | -0.726 | 0.008 | 1E-200 | 8235.56 |
| *ALPI* | rs11216930 | A | C | 0.2262 | -0.056 | 0.007 | 2.5119E-14 | 64.00 |
| *ALPI* | rs186021206 | G | A | 0.0063 | 0.751 | 0.043 | 1.2589E-69 | 305.03 |
| *ALPI* | rs79008529 | G | A | 0.0459 | 0.119 | 0.017 | 1.5849E-12 | 49.00 |
| *ALPI* | rs681343 | C | T | 0.5086 | -0.611 | 0.007 | 1E-200 | 7618.80 |
| *ALPP* | rs761164 | A | G | 0.149 | 0.069 | 0.009 | 1.5849E-13 | 58.78 |
| *ALPP* | rs12743084 | C | G | 0.5421 | 0.063 | 0.007 | 7.9433E-21 | 81.00 |
| *ALPP* | rs12478529 | C | T | 0.2347 | 0.506 | 0.009 | 1E-200 | 3160.94 |
| *ALPP* | rs71626052 | A | AAAAT | 0.4735 | -0.048 | 0.007 | 1.5849E-12 | 47.02 |
| *ALPP* | rs2076295 | T | G | 0.4485 | 0.046 | 0.007 | 1.2589E-11 | 43.18 |
| *ALPP* | rs55792059 | C | A | 0.1907 | 0.075 | 0.009 | 1.2589E-17 | 69.44 |
| *ALPP* | rs117847945 | A | G | 0.019 | 0.175 | 0.024 | 7.9433E-13 | 53.17 |
| *ALPP* | rs3936578 | A | C | 0.4898 | -0.08 | 0.007 | 5.0119E-32 | 130.61 |
| *ALPP* | rs782134971 | GAAACTGC( | G | 0.2543 | -0.245 | 0.008 | 1E-200 | 937.89 |
| *ALPP* | rs7167092 | T | C | 0.351 | -0.052 | 0.007 | 1E-13 | 55.18 |
| *ALPP* | rs186021206 | G | A | 0.0063 | 0.463 | 0.046 | 1.2589E-23 | 101.31 |
| *ALPP* | rs516246 | C | T | 0.5088 | -0.117 | 0.007 | 1.2589E-67 | 279.37 |
| *AMBP* | rs61747728 | C | T | 0.0379 | 0.149 | 0.018 | 6.3096E-16 | 68.52 |
| *AMBP* | rs75166367 | G | A | 0.0602 | -0.102 | 0.015 | 7.9433E-12 | 46.24 |
| *AMBP* | rs406658 | C | A | 0.1085 | 0.152 | 0.012 | 3.9811E-38 | 160.44 |
| *AMBP* | rs10265221 | T | C | 0.287 | 0.053 | 0.008 | 6.3096E-12 | 43.89 |
| *AMBP* | rs147941577 | A | G | 0.0028 | -1.69 | 0.067 | 2.512E-142 | 636.24 |
| *AMBP* | rs1169294 | G | A | 0.3081 | 0.058 | 0.008 | 6.3096E-14 | 52.56 |
| *AMBP* | rs28929474 | C | T | 0.0208 | 0.18 | 0.025 | 3.9811E-13 | 51.84 |
| *AMBP* | rs34262842 | A | G | 0.1663 | -0.064 | 0.01 | 1.5849E-11 | 40.96 |
| *AMBP* | rs704 | G | A | 0.473 | -0.109 | 0.007 | 7.9433E-54 | 242.47 |
| *AMIGO2* | rs3791101 | G | A | 0.3077 | 0.061 | 0.008 | 3.9811E-14 | 58.14 |
| *AMIGO2* | rs1524668 | A | C | 0.6668 | 0.062 | 0.008 | 3.1623E-15 | 60.06 |
| *AMIGO2* | rs1354034 | T | C | 0.6046 | -0.146 | 0.008 | 5.0119E-83 | 333.06 |
| *AMIGO2* | rs10935473 | G | T | 0.4404 | -0.178 | 0.008 | 2.512E-123 | 495.06 |
| *AMIGO2* | rs1281937 | T | C | 0.2752 | -0.062 | 0.008 | 7.9433E-14 | 60.06 |
| *AMIGO2* | rs385893 | T | C | 0.5239 | 0.061 | 0.007 | 3.1623E-16 | 75.94 |
| *AMIGO2* | rs2519093 | C | T | 0.1845 | -0.169 | 0.01 | 1.9953E-69 | 285.61 |

| | | | | | | | | |
|---|---|---|---|---|---|---|---|---|
| *AMIGO2* | rs174546 | C | T | 0.3442 | 0.063 | 0.008 | 3.9811E-16 | 62.02 |
| *AMIGO2* | rs4055121 | C | T | 0.1347 | -0.118 | 0.011 | 2.5119E-27 | 115.07 |
| *AMIGO2* | rs215384 | C | G | 0.411 | 0.157 | 0.008 | 2.5119E-94 | 385.14 |
| *AMIGO2* | rs2242497 | C | G | 0.3619 | -0.06 | 0.008 | 2.5119E-14 | 56.25 |
| *AMIGO2* | rs10774625 | A | G | 0.5036 | -0.083 | 0.007 | 1E-28 | 140.59 |
| *AMIGO2* | rs682619 | T | C | 0.3588 | 0.052 | 0.008 | 1.2589E-11 | 42.25 |
| *AMIGO2* | rs186021206 | G | A | 0.0063 | 0.981 | 0.051 | 3.1623E-81 | 370.00 |
| *AMN* | rs12740374 | G | T | 0.2214 | 0.07 | 0.008 | 1.5849E-16 | 76.56 |
| *AMN* | rs7979473 | A | G | 0.6163 | 0.051 | 0.007 | 1.9953E-12 | 53.08 |
| *AMN* | rs28929474 | C | T | 0.0206 | 0.682 | 0.027 | 3.162E-140 | 638.03 |
| *AMN* | rs2295829 | G | C | 0.3902 | 0.77 | 0.009 | 1E-200 | 7319.75 |
| *AMN* | rs1801689 | A | C | 0.03 | 0.238 | 0.021 | 6.3096E-31 | 128.44 |
| *AMOT* | rs4846913 | C | A | 0.6066 | 0.049 | 0.007 | 1.5849E-11 | 49.00 |
| *AMOT* | rs35719237 | C | CGA | 0.7444 | -0.064 | 0.008 | 3.1623E-15 | 64.00 |
| *AMOT* | rs2954021 | A | G | 0.5036 | 0.054 | 0.007 | 1E-14 | 59.51 |
| *AMOT* | rs533333382 | G | A | 0.1901 | 0.251 | 0.009 | 1.585E-168 | 777.79 |
| *AMOT* | rs2393791 | C | T | 0.6234 | 0.088 | 0.007 | 3.1623E-34 | 158.04 |
| *AMOT* | rs340005 | G | A | 0.6241 | 0.049 | 0.007 | 7.9433E-12 | 49.00 |
| *AMOTL2* | rs1353778 | A | T | 0.6127 | 0.061 | 0.008 | 6.3096E-15 | 58.14 |
| *AMOTL2* | rs3780420 | T | C | 0.3643 | 0.058 | 0.008 | 1.5849E-13 | 52.56 |
| *AMPD3* | rs117706710 | G | T | 0.0091 | -1.717 | 0.038 | 1E-200 | 2041.61 |
| *AMY1A_AMY1B_AMY1* | rs72694438 | G | A | 0.2068 | 0.403 | 0.01 | 1E-200 | 1624.09 |
| *AMY1A_AMY1B_AMY1* | rs291091 | T | C | 0.4531 | 0.065 | 0.008 | 1E-16 | 66.02 |
| *AMY1A_AMY1B_AMY1* | rs13159800 | T | C | 0.669 | -0.073 | 0.008 | 1.5849E-21 | 83.27 |
| *AMY1A_AMY1B_AMY1* | rs888686 | A | T | 0.5922 | -0.052 | 0.007 | 1.5849E-12 | 55.18 |
| *AMY1A_AMY1B_AMY1* | rs782134971 | GAAACTGC( | G | 0.2551 | -0.108 | 0.008 | 5.0119E-39 | 182.25 |
| *AMY1A_AMY1B_AMY1* | rs56278466 | T | G | 0.6602 | 0.051 | 0.008 | 1.2589E-11 | 40.64 |
| *AMY1A_AMY1B_AMY1* | rs3851583 | A | G | 0.2566 | -0.094 | 0.008 | 5.0119E-30 | 138.06 |
| *AMY1A_AMY1B_AMY1* | rs751116426 | TTA | T | 0.1922 | -0.076 | 0.009 | 7.9433E-17 | 71.31 |
| *AMY1A_AMY1B_AMY1* | rs2638282 | G | A | 0.5326 | 0.072 | 0.007 | 1.5849E-23 | 105.80 |
| *AMY2A* | rs78245241 | T | G | 0.0366 | -0.964 | 0.021 | 1E-200 | 2107.25 |
| *AMY2A* | rs55901622 | A | G | 0.2692 | 0.056 | 0.008 | 3.1623E-12 | 49.00 |
| *AMY2A* | rs1561929 | C | T | 0.8832 | 0.076 | 0.011 | 1.5849E-11 | 47.74 |
| *AMY2A* | rs550057 | C | T | 0.2551 | -0.151 | 0.008 | 1E-75 | 356.27 |
| *AMY2A* | rs11054115 | T | C | 0.2413 | -0.072 | 0.008 | 1.5849E-17 | 81.00 |
| *AMY2A* | rs533406 | A | G | 0.3825 | -0.071 | 0.007 | 1.2589E-22 | 102.88 |
| *AMY2A* | rs2638282 | G | A | 0.5326 | 0.052 | 0.007 | 3.9811E-13 | 55.18 |
| *AMY2B* | rs12080068 | C | A | 0.0922 | 0.659 | 0.014 | 1E-200 | 2215.72 |
| *AMY2B* | rs1561929 | C | T | 0.8829 | 0.081 | 0.011 | 1.9953E-12 | 54.22 |
| *AMY2B* | rs550057 | C | T | 0.255 | -0.152 | 0.008 | 1E-74 | 361.00 |
| *AMY2B* | rs56278466 | T | G | 0.6618 | 0.055 | 0.008 | 5.0119E-13 | 47.27 |
| *AMY2B* | rs28456 | A | G | 0.3139 | 0.054 | 0.008 | 3.1623E-12 | 45.56 |
| *AMY2B* | rs11054115 | T | C | 0.241 | -0.066 | 0.009 | 1.9953E-14 | 53.78 |
| *AMY2B* | rs533406 | A | G | 0.3826 | -0.078 | 0.007 | 1.5849E-25 | 124.16 |
| *AMY2B* | rs2638282 | G | A | 0.5324 | 0.053 | 0.007 | 6.3096E-13 | 57.33 |
| *ANG* | rs61791757 | G | A | 0.231 | -0.06 | 0.008 | 1.5849E-14 | 56.25 |
| *ANG* | rs11047261 | A | G | 0.0412 | 0.12 | 0.016 | 2.5119E-13 | 56.25 |
| *ANG* | rs8008227 | G | A | 0.1169 | 0.667 | 0.011 | 1E-200 | 3676.77 |
| *ANGPT1* | rs12041331 | G | A | 0.0863 | -0.103 | 0.013 | 1.2589E-14 | 62.78 |
| *ANGPT1* | rs13412535 | G | A | 0.2311 | -0.078 | 0.009 | 1.2589E-18 | 75.11 |
| *ANGPT1* | rs7618405 | C | A | 0.205 | -0.071 | 0.009 | 1.9953E-14 | 62.23 |
| *ANGPT1* | rs1354034 | T | C | 0.604 | -0.087 | 0.008 | 1.2589E-30 | 118.27 |
| *ANGPT1* | rs3792366 | G | A | 0.5928 | -0.055 | 0.008 | 6.3096E-13 | 47.27 |
| *ANGPT1* | rs755492124 | TAAAG | T | 0.3937 | -0.075 | 0.008 | 3.1623E-23 | 87.89 |
| *ANGPT1* | rs10058074 | G | A | 0.4757 | -0.065 | 0.007 | 1.9953E-18 | 86.22 |
| *ANGPT1* | rs2516471 | G | A | 0.0636 | -0.138 | 0.015 | 3.1623E-19 | 84.64 |
| *ANGPT1* | rs146842698 | T | TAAATA | 0.3053 | -0.069 | 0.008 | 3.9811E-17 | 74.39 |
| *ANGPT1* | rs6961069 | C | T | 0.4023 | 0.077 | 0.008 | 2.5119E-24 | 92.64 |
| *ANGPT1* | rs6993770 | A | T | 0.287 | -0.195 | 0.008 | 1.259E-122 | 594.14 |
| *ANGPT1* | rs2047792 | G | A | 0.0617 | -0.105 | 0.016 | 1E-11 | 43.07 |
| *ANGPT1* | rs61751937 | G | C | 0.0284 | 0.154 | 0.022 | 5.0119E-12 | 49.00 |
| *ANGPT1* | rs113344952 | G | C | 0.3534 | -0.058 | 0.008 | 6.3096E-13 | 52.56 |
| *ANGPT1* | rs7080386 | C | A | 0.412 | 0.098 | 0.008 | 1.2589E-38 | 150.06 |
| *ANGPT1* | rs71474568 | T | TTG | 0.3399 | 0.073 | 0.008 | 1.2589E-20 | 83.27 |
| *ANGPT1* | rs4759076 | T | C | 0.457 | 0.064 | 0.008 | 1.9953E-17 | 64.00 |
| *ANGPT1* | rs61978213 | G | A | 0.0415 | 0.141 | 0.019 | 3.1623E-14 | 55.07 |
| *ANGPT1* | rs59001897 | T | A | 0.1744 | 0.081 | 0.01 | 1.9953E-16 | 65.61 |
| *ANGPT1* | rs151234 | G | C | 0.1293 | 0.076 | 0.011 | 7.9433E-12 | 47.74 |

| | | | | | | | | |
|---|---|---|---|---|---|---|---|---|
| *ANGPT1* | rs12445050 | C | T | 0.1381 | 0.121 | 0.011 | 1.9953E-29 | 121.00 |
| *ANGPT1* | rs1654425 | T | C | 0.8345 | 0.186 | 0.01 | 1E-75 | 345.96 |
| *ANGPT1* | rs3827978 | C | T | 0.3535 | 0.081 | 0.008 | 3.1623E-25 | 102.52 |
| *ANGPT2* | rs13107325 | C | T | 0.0748 | 0.121 | 0.014 | 3.9811E-18 | 74.70 |
| *ANGPT2* | rs1968586 | C | T | 0.3554 | -0.157 | 0.008 | 1.2589E-89 | 385.14 |
| *ANGPT2* | rs4841133 | A | G | 0.908 | -0.108 | 0.013 | 3.9811E-17 | 69.02 |
| *ANGPT2* | rs34434834 | G | A | 0.0334 | 0.238 | 0.021 | 3.9811E-31 | 128.44 |
| *ANGPT2* | rs186021206 | G | A | 0.0064 | 0.862 | 0.05 | 1.9953E-65 | 297.22 |
| *ANGPTL1* | rs12753251 | G | A | 0.5341 | -0.284 | 0.008 | 1E-200 | 1260.25 |
| *ANGPTL1* | rs1137654 | A | T | 0.0942 | -0.093 | 0.013 | 5.0119E-13 | 51.18 |
| *ANGPTL1* | rs28929474 | C | T | 0.0209 | 0.195 | 0.025 | 1.5849E-14 | 60.84 |
| *ANGPTL1* | rs149394327 | G | C | 0.0301 | 0.245 | 0.021 | 7.9433E-31 | 136.11 |
| *ANGPTL1* | rs8103017 | C | G | 0.2908 | 0.061 | 0.008 | 5.0119E-14 | 58.14 |
| *ANGPTL2* | rs1130389 | C | T | 0.1475 | -0.099 | 0.011 | 1.2589E-18 | 81.00 |
| *ANGPTL2* | rs4141100 | G | A | 0.6137 | 0.064 | 0.008 | 2.5119E-17 | 64.00 |
| *ANGPTL2* | rs138260315 | G | GAT | 0.1779 | -0.118 | 0.01 | 2.5119E-33 | 139.24 |
| *ANGPTL2* | rs55799523 | C | A | 0.2653 | 0.264 | 0.009 | 1E-200 | 860.44 |
| *ANGPTL2* | rs181210490 | G | A | 0.0118 | 0.573 | 0.034 | 5.0119E-62 | 284.02 |
| *ANGPTL3* | rs763015008 | TTTAC | T | 0.3528 | -0.29 | 0.007 | 1E-200 | 1716.33 |
| *ANGPTL3* | rs3916027 | G | A | 0.2635 | 0.085 | 0.008 | 1.2589E-28 | 112.89 |
| *ANGPTL3* | rs10808546 | C | T | 0.4461 | 0.051 | 0.007 | 6.3096E-14 | 53.08 |
| *ANGPTL3* | rs964184 | G | C | 0.8667 | -0.118 | 0.01 | 3.9811E-32 | 139.24 |
| *ANGPTL3* | rs28929474 | C | T | 0.0208 | 0.239 | 0.024 | 1E-23 | 99.17 |
| *ANGPTL3* | rs72786786 | G | A | 0.3134 | 0.051 | 0.007 | 1E-11 | 53.08 |
| *ANGPTL3* | rs1065853 | G | T | 0.0812 | 0.131 | 0.012 | 1.2589E-25 | 119.17 |
| *ANGPTL4* | rs2278236 | G | A | 0.5143 | 0.28 | 0.008 | 1E-200 | 1225.00 |
| *ANGPTL7* | rs28991009 | G | T | 0.0085 | -2.155 | 0.04 | 1E-200 | 2902.52 |
| *ANGPTL7* | rs6764769 | A | G | 0.444 | -0.06 | 0.007 | 2.5119E-17 | 73.47 |
| *ANGPTL7* | rs7019896 | G | T | 0.9328 | 0.108 | 0.014 | 2.5119E-14 | 59.51 |
| *ANGPTL7* | rs529700 | T | C | 0.5148 | 0.072 | 0.008 | 7.9433E-18 | 81.00 |
| *ANGPTL7* | rs112771035 | C | G | 0.0713 | -0.118 | 0.014 | 1E-17 | 71.04 |
| *ANGPTL7* | rs186021206 | G | A | 0.0064 | 0.759 | 0.049 | 1.5849E-54 | 239.93 |
| *ANGPTL7* | rs704 | G | A | 0.4729 | 0.058 | 0.007 | 2.5119E-16 | 68.65 |
| *ANK2* | rs144646268 | G | T | 0.0387 | 0.16 | 0.02 | 1.9953E-15 | 64.00 |
| *ANKMY2* | rs1354034 | T | C | 0.6039 | 0.085 | 0.008 | 1.2589E-28 | 112.89 |
| *ANKMY2* | rs4719474 | C | A | 0.3761 | 0.082 | 0.008 | 1.5849E-24 | 105.06 |
| *ANKMY2* | rs7080386 | C | A | 0.4124 | 0.054 | 0.008 | 3.1623E-12 | 45.56 |
| *ANKMY2* | rs11502185 | T | C | 0.2597 | 0.072 | 0.01 | 6.3096E-14 | 51.84 |
| *ANKRD54* | rs35496925 | A | G | 0.0594 | 0.143 | 0.016 | 5.0119E-19 | 79.88 |
| *ANPEP* | rs13107325 | C | T | 0.0749 | 0.094 | 0.013 | 1.5849E-12 | 52.28 |
| *ANPEP* | rs4835265 | C | A | 0.1565 | 0.071 | 0.01 | 1.2589E-13 | 50.41 |
| *ANPEP* | rs68055275 | C | CT | 0.1449 | -0.103 | 0.01 | 3.9811E-24 | 106.09 |
| *ANPEP* | rs10142200 | A | G | 0.4088 | 0.053 | 0.007 | 3.9811E-14 | 57.33 |
| *ANPEP* | rs1042499 | T | C | 0.0571 | 0.819 | 0.016 | 1E-200 | 2620.16 |
| *ANPEP* | rs186021206 | G | A | 0.0064 | 0.639 | 0.048 | 1E-40 | 177.22 |
| *ANXA1* | rs35923643 | A | G | 0.196 | -0.093 | 0.009 | 6.3096E-23 | 106.78 |
| *ANXA1* | rs58534292 | T | C | 0.6497 | 0.053 | 0.008 | 1.5849E-11 | 43.89 |
| *ANXA1* | rs12885254 | T | A | 0.7048 | 0.064 | 0.009 | 2.5119E-13 | 50.57 |
| *ANXA1* | rs34436714 | C | A | 0.2089 | 0.129 | 0.009 | 1.9953E-44 | 205.44 |
| *ANXA11* | rs9281986 | A | AG | 0.166 | -0.104 | 0.01 | 3.1623E-23 | 108.16 |
| *ANXA11* | rs2784773 | T | C | 0.6442 | 0.066 | 0.008 | 3.9811E-17 | 68.06 |
| *ANXA2* | rs342298 | C | T | 0.4547 | -0.056 | 0.007 | 1.9953E-15 | 64.00 |
| *ANXA2* | rs8033800 | T | A | 0.6016 | -0.461 | 0.008 | 1E-200 | 3320.64 |
| *ANXA2* | rs4632248 | G | T | 0.2091 | 0.119 | 0.009 | 3.1623E-42 | 174.83 |
| *ANXA3* | rs146728751 | G | A | 0.0016 | -1.222 | 0.093 | 2.5119E-39 | 172.65 |
| *ANXA3* | rs6993770 | A | T | 0.2865 | -0.067 | 0.008 | 2.5119E-16 | 70.14 |
| *ANXA3* | rs7896518 | A | G | 0.424 | 0.059 | 0.008 | 7.9433E-15 | 54.39 |
| *ANXA3* | rs10418046 | T | G | 0.211 | 0.065 | 0.009 | 1E-12 | 52.16 |
| *ANXA4* | rs1354034 | T | C | 0.6037 | 0.053 | 0.008 | 3.9811E-12 | 43.89 |
| *ANXA4* | rs7896518 | A | G | 0.4235 | 0.069 | 0.008 | 2.5119E-19 | 74.39 |
| *ANXA5* | rs2306411 | A | G | 0.2914 | -0.117 | 0.008 | 1.9953E-43 | 213.89 |
| *AOC1* | rs1049742 | C | T | 0.081 | 0.495 | 0.014 | 1E-200 | 1250.13 |
| *AOC3* | rs2270894 | C | G | 0.203 | 0.081 | 0.008 | 1.5849E-21 | 102.52 |
| *AOC3* | rs13135092 | A | G | 0.0836 | 0.086 | 0.012 | 7.9433E-13 | 51.36 |
| *AOC3* | rs75901433 | A | G | 0.5578 | 0.054 | 0.007 | 2.5119E-15 | 59.51 |
| *AOC3* | rs767298 | G | T | 0.6339 | -0.065 | 0.007 | 3.9811E-21 | 86.22 |
| *AOC3* | rs9411365 | C | G | 0.0717 | 0.098 | 0.013 | 2.5119E-14 | 56.83 |
| *AOC3* | rs56278466 | T | G | 0.6612 | 0.067 | 0.007 | 3.9811E-22 | 91.61 |

| | | | | | | | | |
|---|---|---|---|---|---|---|---|---|
| *AOC3* | rs3967200 | C | T | 0.1348 | -0.101 | 0.01 | 1.5849E-25 | 102.01 |
| *AOC3* | rs1042704 | G | A | 0.2151 | -0.092 | 0.008 | 7.9433E-31 | 132.25 |
| *AOC3* | rs12913657 | T | C | 0.1466 | 0.079 | 0.009 | 1.5849E-17 | 77.05 |
| *AOC3* | rs186021206 | G | A | 0.0064 | 0.591 | 0.05 | 1.2589E-32 | 139.71 |
| *AOC3* | rs33986943 | G | A | 0.111 | -0.867 | 0.012 | 1E-200 | 5220.06 |
| *AP1G2* | rs28403575 | C | T | 0.0885 | -0.192 | 0.013 | 1.9953E-48 | 218.13 |
| *AP2B1* | rs3811444 | C | T | 0.3321 | -0.082 | 0.008 | 3.9811E-24 | 105.06 |
| *AP2B1* | rs1354034 | T | C | 0.604 | 0.058 | 0.008 | 6.3096E-14 | 52.56 |
| *AP3B1* | rs1354034 | T | C | 0.6046 | 0.077 | 0.008 | 5.0119E-24 | 92.64 |
| *AP3B1* | rs6993770 | A | T | 0.2866 | -0.058 | 0.008 | 1.5849E-12 | 52.56 |
| *AP3S2* | rs1354034 | T | C | 0.6043 | 0.063 | 0.008 | 1.9953E-16 | 62.02 |
| *APBB1IP* | rs7461753 | A | C | 0.4204 | 0.057 | 0.007 | 2.5119E-14 | 66.31 |
| *APBB1IP* | rs2992335 | C | G | 0.6009 | -0.217 | 0.008 | 3.981E-179 | 735.77 |
| *APBB1IP* | rs11671654 | C | G | 0.4945 | 0.15 | 0.007 | 1E-89 | 459.18 |
| *APBB1IP* | rs10418046 | T | G | 0.2114 | 0.113 | 0.009 | 2.5119E-35 | 157.64 |
| *APCS* | rs36126250 | T | G | 0.023 | -0.535 | 0.024 | 7.943E-114 | 496.92 |
| *APCS* | rs2842700 | C | A | 0.0535 | 0.155 | 0.016 | 5.0119E-23 | 93.85 |
| *APCS* | rs1260326 | T | C | 0.6083 | -0.095 | 0.007 | 1.5849E-40 | 184.18 |
| *APEX1* | rs1354034 | T | C | 0.6042 | 0.057 | 0.007 | 1.2589E-15 | 66.31 |
| *APEX1* | rs111338191 | A | AAATT | 0.5205 | -0.051 | 0.007 | 5.0119E-13 | 53.08 |
| *APEX1* | rs1130409 | T | G | 0.4801 | -0.389 | 0.007 | 1E-200 | 3088.18 |
| *APEX1* | rs4632248 | G | T | 0.2098 | 0.067 | 0.009 | 1.5849E-14 | 55.42 |
| *APLP1* | rs13412535 | G | A | 0.231 | 0.074 | 0.009 | 1E-16 | 67.60 |
| *APLP1* | rs4602861 | A | G | 0.2739 | 0.097 | 0.008 | 1.5849E-30 | 147.02 |
| *APLP1* | rs2229498 | G | A | 0.8358 | -0.112 | 0.01 | 7.9433E-29 | 125.44 |
| *APLP1* | rs12609736 | A | G | 0.8322 | -0.104 | 0.01 | 3.9811E-25 | 108.16 |
| *APLP1* | rs6045989 | G | A | 0.3555 | -0.057 | 0.008 | 3.1623E-13 | 50.77 |
| *APOA1* | rs62568183 | T | G | 0.1166 | -0.083 | 0.011 | 6.3096E-14 | 56.93 |
| *APOA1* | rs2727784 | C | T | 0.6497 | -0.064 | 0.007 | 1.9953E-17 | 83.59 |
| *APOA1* | rs1077835 | A | G | 0.222 | 0.135 | 0.009 | 1.2589E-56 | 225.00 |
| *APOA1* | rs3764261 | C | A | 0.3252 | 0.161 | 0.008 | 1E-99 | 405.02 |
| *APOA1* | rs77960347 | A | G | 0.0131 | 0.307 | 0.031 | 6.3096E-23 | 98.07 |
| *APOA1* | rs4420638 | A | G | 0.1902 | -0.083 | 0.009 | 3.9811E-20 | 85.05 |
| *APOA2* | rs4073054 | C | A | 0.6263 | 0.067 | 0.008 | 5.0119E-19 | 70.14 |
| *APOA2* | rs6682862 | G | A | 0.1673 | -0.076 | 0.01 | 7.9433E-15 | 57.76 |
| *APOA2* | rs11096641 | G | A | 0.4712 | -0.089 | 0.007 | 1.5849E-34 | 161.65 |
| *APOA2* | rs13107325 | C | T | 0.0748 | -0.145 | 0.014 | 1.5849E-25 | 107.27 |
| *APOA2* | rs4240624 | G | A | 0.908 | 0.121 | 0.013 | 1.2589E-21 | 86.63 |
| *APOA2* | rs28601761 | C | G | 0.4195 | 0.055 | 0.007 | 1.2589E-13 | 61.73 |
| *APOA2* | rs62568182 | T | G | 0.1211 | -0.086 | 0.011 | 1.9953E-14 | 61.12 |
| *APOA2* | rs12555241 | G | A | 0.2776 | -0.059 | 0.008 | 7.9433E-13 | 54.39 |
| *APOA2* | rs2727784 | C | T | 0.6503 | -0.057 | 0.008 | 1.2589E-13 | 50.77 |
| *APOA2* | rs339969 | C | A | 0.617 | 0.052 | 0.007 | 3.1623E-12 | 55.18 |
| *APOA2* | rs56228609 | C | T | 0.316 | 0.095 | 0.008 | 1.5849E-33 | 141.02 |
| *APOA2* | rs58542926 | C | T | 0.0754 | 0.103 | 0.014 | 7.9433E-14 | 54.13 |
| *APOA2* | rs429358 | T | C | 0.1557 | 0.127 | 0.01 | 1.9953E-36 | 161.29 |
| *APOA4* | rs150816167 | T | C | 0.0417 | 0.167 | 0.019 | 1E-18 | 77.25 |
| *APOA4* | rs9273206 | T | C | 0.8347 | 0.087 | 0.01 | 1.9953E-17 | 75.69 |
| *APOA4* | rs1729407 | C | G | 0.5488 | 0.141 | 0.008 | 6.3096E-77 | 310.64 |
| *APOBR* | rs12132412 | A | G | 0.3904 | -0.079 | 0.005 | 6.3096E-54 | 249.64 |
| *APOBR* | rs550057 | C | T | 0.2555 | -0.051 | 0.006 | 3.1623E-19 | 72.25 |
| *APOBR* | rs149271 | A | G | 0.41 | -1.123 | 0.008 | 1E-200 | 19705.14 |
| *APOC1* | rs12124281 | G | A | 0.352 | -0.055 | 0.007 | 7.9433E-14 | 61.73 |
| *APOC1* | rs55776147 | A | G | 0.6383 | 0.063 | 0.007 | 3.9811E-18 | 81.00 |
| *APOC1* | rs17489373 | G | A | 0.2647 | 0.082 | 0.008 | 3.9811E-25 | 105.06 |
| *APOC1* | rs3764261 | C | A | 0.3251 | 0.145 | 0.008 | 1.5849E-82 | 328.52 |
| *APOC1* | rs77960347 | A | G | 0.0131 | 0.282 | 0.031 | 7.9433E-20 | 82.75 |
| *APOC1* | rs8107974 | A | T | 0.0764 | -0.103 | 0.013 | 3.1623E-14 | 62.78 |
| *APOC1* | rs5112 | C | G | 0.5333 | 0.345 | 0.008 | 1E-200 | 1859.77 |
| *APOD* | rs139828053 | T | C | 0.0313 | -0.537 | 0.022 | 2.512E-130 | 595.80 |
| *APOD* | rs894211 | C | T | 0.2649 | 0.075 | 0.008 | 1.2589E-18 | 87.89 |
| *APOD* | rs10644928 | C | CTT | 0.3885 | 0.063 | 0.008 | 6.3096E-16 | 62.02 |
| *APOD* | rs1800775 | C | A | 0.4867 | 0.077 | 0.008 | 7.9433E-25 | 92.64 |
| *APOD* | rs5167 | T | G | 0.3524 | 0.058 | 0.008 | 7.9433E-14 | 52.56 |
| *APOE* | rs2740488 | A | C | 0.267 | -0.058 | 0.007 | 1.5849E-16 | 68.65 |
| *APOE* | rs99780 | C | T | 0.3506 | -0.068 | 0.007 | 7.9433E-26 | 94.37 |
| *APOE* | rs429358 | T | C | 0.1557 | -1.012 | 0.01 | 1E-200 | 10241.44 |
| *APOF* | rs112875651 | G | A | 0.3914 | 0.063 | 0.008 | 5.0119E-17 | 62.02 |

| | | | | | | | | |
|---|---|---|---|---|---|---|---|---|
| *APOF* | rs2020854 | T | C | 0.0697 | -0.351 | 0.014 | 6.31E-133 | 628.58 |
| *APOF* | rs147233090 | C | T | 0.0236 | -0.202 | 0.024 | 5.0119E-17 | 70.84 |
| *APOF* | rs11076175 | A | G | 0.1764 | -0.077 | 0.01 | 1E-15 | 59.29 |
| *APOF* | rs3809868 | G | A | 0.5184 | -0.066 | 0.007 | 1.2589E-19 | 88.90 |
| *APOF* | rs112001035 | G | A | 0.0612 | 0.126 | 0.016 | 6.3096E-16 | 62.02 |
| *APOF* | rs429358 | T | C | 0.1557 | 0.117 | 0.01 | 5.0119E-31 | 136.89 |
| *APOH* | rs1801689 | A | C | 0.0301 | -1.462 | 0.022 | 1E-200 | 4416.21 |
| *APOL1* | rs5030062 | A | C | 0.3733 | 0.063 | 0.007 | 1.2589E-19 | 81.00 |
| *APOL1* | rs4861708 | G | A | 0.5118 | 0.201 | 0.007 | 1.995E-190 | 824.51 |
| *APOL1* | rs1801020 | A | G | 0.7448 | 0.202 | 0.008 | 3.981E-149 | 637.56 |
| *APOL1* | rs9275282 | C | T | 0.4881 | 0.047 | 0.007 | 3.1623E-12 | 45.08 |
| *APOL1* | rs550478926 | C | G | 0.0835 | -0.092 | 0.013 | 6.3096E-13 | 50.08 |
| *APOL1* | rs751229834 | CA | C | 0.3071 | 0.054 | 0.007 | 1.9953E-13 | 59.51 |
| *APOL1* | rs9930957 | C | T | 0.1601 | 0.509 | 0.01 | 1E-200 | 2590.81 |
| *APOL1* | rs1800961 | C | T | 0.0323 | 0.13 | 0.019 | 7.9433E-12 | 46.81 |
| *APOL1* | rs136168 | G | A | 0.8133 | -0.318 | 0.009 | 1E-200 | 1248.44 |
| *APOM* | rs1260326 | T | C | 0.6081 | -0.093 | 0.007 | 3.1623E-36 | 176.51 |
| *APOM* | rs805258 | C | T | 0.0296 | -0.784 | 0.022 | 1E-200 | 1269.95 |
| *APOM* | rs115849089 | G | A | 0.1161 | 0.085 | 0.011 | 3.9811E-14 | 59.71 |
| *APOM* | rs56332871 | C | A | 0.2701 | 0.055 | 0.008 | 1.5849E-11 | 47.27 |
| *APOM* | rs247616 | C | T | 0.3252 | 0.131 | 0.008 | 3.1623E-63 | 268.14 |
| *APOM* | rs112001035 | G | A | 0.061 | 0.134 | 0.015 | 5.0119E-18 | 79.80 |
| *APOM* | rs1800961 | C | T | 0.0323 | -0.244 | 0.02 | 6.3096E-33 | 148.84 |
| *APOM* | rs738408 | C | T | 0.2167 | -0.061 | 0.009 | 1.9953E-12 | 45.94 |
| *APP* | rs12086222 | G | C | 0.1109 | 0.087 | 0.012 | 3.9811E-13 | 52.56 |
| *APP* | rs13412535 | G | A | 0.231 | -0.062 | 0.009 | 2.5119E-12 | 47.46 |
| *APP* | rs1354034 | T | C | 0.6041 | -0.101 | 0.008 | 2.5119E-40 | 159.39 |
| *APP* | rs755492124 | TAAAG | T | 0.3934 | -0.072 | 0.008 | 1E-20 | 81.00 |
| *APP* | rs274555 | C | T | 0.3986 | 0.066 | 0.008 | 1.2589E-17 | 68.06 |
| *APP* | rs138614581 | A | AT | 0.08 | -0.117 | 0.014 | 3.9811E-17 | 69.84 |
| *APP* | rs1917342 | A | G | 0.3032 | -0.06 | 0.008 | 3.1623E-13 | 56.25 |
| *APP* | rs6961069 | C | T | 0.4022 | 0.069 | 0.008 | 2.5119E-19 | 74.39 |
| *APP* | rs6993770 | A | T | 0.2864 | -0.084 | 0.008 | 2.5119E-24 | 110.25 |
| *APP* | rs11794772 | G | A | 0.2588 | -0.068 | 0.009 | 2.5119E-15 | 57.09 |
| *APP* | rs7896518 | A | G | 0.4243 | 0.104 | 0.008 | 7.9433E-42 | 169.00 |
| *APP* | rs2229498 | G | A | 0.8355 | 0.113 | 0.01 | 6.3096E-29 | 127.69 |
| *APP* | rs61978213 | G | A | 0.0416 | 0.129 | 0.019 | 6.3096E-12 | 46.10 |
| *APP* | rs59001897 | T | A | 0.1746 | 0.079 | 0.01 | 1.9953E-15 | 62.41 |
| *APP* | rs12445050 | C | T | 0.1383 | 0.112 | 0.011 | 6.3096E-25 | 103.67 |
| *APP* | rs892090 | T | G | 0.8342 | 0.17 | 0.01 | 5.0119E-63 | 289.00 |
| *APP* | rs6081565 | G | A | 0.3525 | 0.079 | 0.008 | 7.9433E-24 | 97.52 |
| *APP* | rs8131895 | C | A | 0.6541 | -0.073 | 0.008 | 3.9811E-20 | 83.27 |
| *APPL2* | rs10778371 | G | A | 0.3722 | 0.129 | 0.008 | 3.1623E-61 | 260.02 |
| *APRT* | rs1354034 | T | C | 0.6041 | 0.098 | 0.008 | 1.5849E-38 | 150.06 |
| *APRT* | rs854572 | C | G | 0.527 | 0.051 | 0.008 | 1E-11 | 40.64 |
| *APRT* | rs10876550 | G | A | 0.5582 | 0.061 | 0.007 | 3.9811E-16 | 75.94 |
| *APRT* | rs74403919 | T | A | 0.1719 | 0.067 | 0.01 | 1.2589E-11 | 44.89 |
| *APRT* | rs111433410 | C | T | 0.0097 | -0.351 | 0.038 | 3.9811E-20 | 85.32 |
| *APRT* | rs704 | G | A | 0.4733 | -0.081 | 0.007 | 5.0119E-28 | 133.90 |
| *AREG* | rs1691273 | C | T | 0.6921 | -0.177 | 0.008 | 1E-104 | 489.52 |
| *AREG* | rs712831 | T | C | 0.7852 | 0.071 | 0.009 | 5.0119E-15 | 62.23 |
| *AREG* | rs492602 | A | G | 0.5088 | -0.069 | 0.007 | 5.0119E-20 | 97.16 |
| *ARF6* | rs1354034 | T | C | 0.6046 | 0.059 | 0.008 | 1E-14 | 54.39 |
| *ARFIP1* | rs1354034 | T | C | 0.6043 | 0.059 | 0.008 | 7.9433E-15 | 54.39 |
| *ARFIP1* | rs4619875 | C | T | 0.4035 | 0.159 | 0.008 | 2.5119E-95 | 395.02 |
| *ARG1* | rs2781668 | C | T | 0.1668 | -0.237 | 0.01 | 6.31E-128 | 561.69 |
| *ARG1* | rs590856 | G | A | 0.5531 | 0.094 | 0.007 | 1.5849E-36 | 180.33 |
| *ARG1* | rs6592965 | G | A | 0.4542 | 0.057 | 0.007 | 3.9811E-15 | 66.31 |
| *ARG1* | rs62108438 | T | C | 0.392 | 0.094 | 0.007 | 1.5849E-36 | 180.33 |
| *ARG2* | rs13257521 | C | G | 0.2111 | 0.084 | 0.009 | 2.5119E-19 | 87.11 |
| *ARG2* | rs11056779 | A | G | 0.5705 | 0.058 | 0.008 | 3.9811E-14 | 52.56 |
| *ARG2* | rs35785237 | C | CA | 0.4305 | 0.056 | 0.008 | 1.2589E-12 | 49.00 |
| *ARG2* | rs140881784 | G | C | 0.0079 | -0.404 | 0.047 | 3.9811E-18 | 73.89 |
| *ARHGAP1* | rs6993770 | A | T | 0.2869 | -0.092 | 0.008 | 7.9433E-28 | 132.25 |
| *ARHGAP1* | rs12245149 | C | A | 0.4881 | -0.062 | 0.008 | 1.9953E-16 | 60.06 |
| *ARHGAP25* | rs13413887 | A | G | 0.7582 | 0.106 | 0.009 | 1.2589E-32 | 138.72 |
| *ARHGAP25* | rs4602861 | A | G | 0.274 | -0.067 | 0.009 | 6.3096E-15 | 55.42 |
| *ARHGAP45* | rs1354034 | T | C | 0.6044 | 0.075 | 0.008 | 7.9433E-23 | 87.89 |

| | | | | | | | | |
|---|---|---|---|---|---|---|---|---|
| *ARHGAP45* | rs34593562 | A | G | 0.4494 | -0.071 | 0.008 | 3.9811E-21 | 78.77 |
| *ARHGAP5* | rs529763782 | C | T | 0.6665 | -0.072 | 0.009 | 2.5119E-14 | 64.00 |
| *ARHGAP5* | rs8009570 | G | A | 0.3773 | -0.257 | 0.008 | 1E-200 | 1032.02 |
| *ARHGEF1* | rs1354034 | T | C | 0.6043 | 0.064 | 0.008 | 3.9811E-17 | 64.00 |
| *ARHGEF1* | rs6993770 | A | T | 0.2865 | -0.056 | 0.008 | 1.5849E-11 | 49.00 |
| *ARHGEF10* | rs1354034 | T | C | 0.6043 | 0.074 | 0.007 | 2.5119E-24 | 111.76 |
| *ARHGEF10* | rs11428326 | G | GT | 0.7468 | -0.066 | 0.008 | 1.9953E-15 | 68.06 |
| *ARHGEF10* | rs342293 | C | G | 0.4596 | -0.077 | 0.007 | 6.3096E-27 | 121.00 |
| *ARHGEF10* | rs13281104 | A | G | 0.2435 | 0.317 | 0.009 | 1E-200 | 1240.60 |
| *ARHGEF10* | rs6993770 | A | T | 0.2864 | -0.079 | 0.008 | 5.0119E-22 | 97.52 |
| *ARHGEF10* | rs4759076 | T | C | 0.457 | 0.074 | 0.007 | 1.5849E-24 | 111.76 |
| *ARHGEF5* | rs17881210 | G | A | 0.0707 | -0.106 | 0.014 | 3.9811E-14 | 57.33 |
| *ARHGEF5* | rs1635893 | A | G | 0.2092 | -0.522 | 0.01 | 1E-200 | 2724.84 |
| *ARL13B* | rs13412535 | G | A | 0.2313 | 0.068 | 0.009 | 1E-13 | 57.09 |
| *ARL13B* | rs1354034 | T | C | 0.6037 | 0.06 | 0.008 | 7.9433E-15 | 56.25 |
| *ARL2BP* | rs1354034 | T | C | 0.6037 | 0.11 | 0.008 | 1.9953E-46 | 189.06 |
| *ARL2BP* | rs72767003 | C | T | 0.1455 | 0.075 | 0.011 | 2.5119E-12 | 46.49 |
| *ARL2BP* | rs7198865 | G | A | 0.0352 | -0.746 | 0.021 | 1E-200 | 1261.94 |
| *ARSA* | rs114165349 | G | C | 0.0233 | 0.168 | 0.022 | 6.3096E-15 | 58.31 |
| *ARSA* | rs687339 | C | T | 0.7713 | 0.063 | 0.008 | 3.9811E-16 | 62.02 |
| *ARSA* | rs10745925 | T | C | 0.2877 | -0.267 | 0.007 | 1E-200 | 1454.88 |
| *ARSA* | rs145078947 | G | T | 0.0026 | 0.634 | 0.064 | 3.1623E-23 | 98.13 |
| *ARSA* | rs58542926 | C | T | 0.0755 | 0.184 | 0.012 | 3.1623E-50 | 235.11 |
| *ARSA* | rs429358 | T | C | 0.1551 | -0.062 | 0.009 | 5.0119E-12 | 47.46 |
| *ARSA* | rs873697 | G | A | 0.0451 | 1.499 | 0.018 | 1E-200 | 6935.19 |
| *ARSB* | rs1065757 | C | T | 0.4395 | -0.294 | 0.008 | 1E-200 | 1350.56 |
| *ARSB* | rs368914743 | G | GCACA | 0.4952 | -0.06 | 0.008 | 6.3096E-14 | 56.25 |
| *ARSB* | rs1293303 | G | C | 0.4456 | 0.073 | 0.007 | 5.0119E-23 | 108.76 |
| *ARSB* | rs6993770 | A | T | 0.2864 | -0.086 | 0.008 | 1.9953E-26 | 115.56 |
| *ARSB* | rs10968020 | T | C | 0.2644 | -0.067 | 0.008 | 1E-15 | 70.14 |
| *ARSB* | rs7896518 | A | G | 0.4236 | 0.083 | 0.007 | 3.1623E-28 | 140.59 |
| *ARSB* | rs149871778 | G | GAAC | 0.1046 | 0.087 | 0.012 | 3.1623E-13 | 52.56 |
| *ARSB* | rs10778152 | A | G | 0.2877 | -0.087 | 0.008 | 1.5849E-26 | 118.27 |
| *ARSB* | rs145078947 | G | T | 0.0026 | 0.59 | 0.072 | 2.5119E-16 | 67.15 |
| *ARSB* | rs8072896 | T | G | 0.4846 | -0.051 | 0.007 | 7.9433E-12 | 53.08 |
| *ARSB* | rs892090 | T | G | 0.8341 | 0.099 | 0.01 | 1.9953E-23 | 98.01 |
| *ART3* | rs150816167 | T | C | 0.0419 | 0.151 | 0.017 | 5.0119E-19 | 78.90 |
| *ART3* | rs1868164 | G | A | 0.6306 | -0.061 | 0.007 | 1.9953E-18 | 75.94 |
| *ART3* | rs11649031 | T | C | 0.5874 | -0.053 | 0.007 | 5.0119E-15 | 57.33 |
| *ART3* | rs12934455 | C | T | 0.1664 | -0.08 | 0.009 | 6.3096E-19 | 79.01 |
| *ART5* | rs75902876 | G | A | 0.0813 | 0.35 | 0.015 | 1.259E-115 | 544.44 |
| *ARTN* | rs112597211 | T | C | 0.1359 | 0.092 | 0.011 | 6.3096E-16 | 69.95 |
| *ASAH1* | rs2472205 | A | T | 0.041 | -1.099 | 0.019 | 1E-200 | 3345.71 |
| *ASAH1* | rs4734879 | A | G | 0.2742 | -0.062 | 0.008 | 3.1623E-13 | 60.06 |
| *ASAH1* | rs2559850 | G | A | 0.5876 | 0.067 | 0.007 | 1.5849E-20 | 91.61 |
| *ASAH1* | rs3184504 | T | C | 0.5173 | -0.05 | 0.007 | 1.2589E-12 | 51.02 |
| *ASAH1* | rs2127868 | G | A | 0.7947 | 0.078 | 0.009 | 7.9433E-19 | 75.11 |
| *ASAH1* | rs72701845 | G | A | 0.0375 | -0.214 | 0.019 | 1E-28 | 126.86 |
| *ASAH2* | rs12032634 | T | A | 0.5156 | -0.081 | 0.007 | 1.2589E-33 | 133.90 |
| *ASAH2* | rs7518301 | G | T | 0.7876 | -0.056 | 0.008 | 1E-11 | 49.00 |
| *ASAH2* | rs3732356 | G | T | 0.9352 | -0.112 | 0.014 | 3.1623E-16 | 64.00 |
| *ASAH2* | rs890793 | C | T | 0.3816 | -0.048 | 0.007 | 1.9953E-12 | 47.02 |
| *ASAH2* | rs804267 | G | A | 0.6833 | -0.054 | 0.007 | 7.9433E-14 | 59.51 |
| *ASAH2* | rs12056768 | T | G | 0.5833 | 0.065 | 0.007 | 6.3096E-22 | 86.22 |
| *ASAH2* | rs56278466 | T | G | 0.6614 | 0.068 | 0.008 | 6.3096E-18 | 72.25 |
| *ASAH2* | rs2842126 | A | G | 0.8045 | 0.691 | 0.009 | 1E-200 | 5894.83 |
| *ASAH2* | rs1519125 | G | C | 0.3862 | 0.053 | 0.007 | 1E-14 | 57.33 |
| *ASAH2* | rs35764600 | G | C | 0.3996 | -0.085 | 0.007 | 5.0119E-35 | 147.45 |
| *ASAH2* | rs7979473 | A | G | 0.616 | -0.057 | 0.007 | 1.9953E-16 | 66.31 |
| *ASAH2* | rs28929474 | C | T | 0.0208 | 0.162 | 0.023 | 3.1623E-12 | 49.61 |
| *ASAH2* | rs201947946 | G | GTT | 0.0233 | 0.188 | 0.026 | 1.5849E-13 | 52.28 |
| *ASAH2* | rs76975491 | C | CT | 0.5143 | -0.072 | 0.007 | 3.9811E-27 | 105.80 |
| *ASAH2* | rs77542162 | A | G | 0.023 | 0.29 | 0.022 | 5.0119E-39 | 173.76 |
| *ASGR1* | rs12740374 | G | T | 0.2212 | 0.127 | 0.009 | 2.5119E-46 | 199.12 |
| *ASGR1* | rs11405616 | C | CA | 0.6058 | 0.053 | 0.008 | 5.0119E-12 | 43.89 |
| *ASGR1* | rs7012814 | G | A | 0.4723 | -0.054 | 0.007 | 5.0119E-13 | 59.51 |
| *ASGR1* | rs2622935 | A | G | 0.8075 | -0.141 | 0.009 | 3.9811E-51 | 245.44 |
| *ASGR1* | rs28929474 | C | T | 0.0208 | 0.313 | 0.026 | 7.9433E-34 | 144.92 |

| | | | | | | | | |
|---|---|---|---|---|---|---|---|---|
| *ASGR1* | rs55714927 | C | T | 0.195 | -0.193 | 0.009 | 5.0119E-94 | 459.86 |
| *ASGR1* | rs1801689 | A | C | 0.0301 | 0.19 | 0.022 | 2.5119E-18 | 74.59 |
| *ASGR2* | rs61747728 | C | T | 0.038 | 0.128 | 0.019 | 6.3096E-12 | 45.39 |
| *ASGR2* | rs529541 | A | G | 0.1731 | 0.088 | 0.01 | 1.9953E-20 | 77.44 |
| *ASGR2* | rs12621059 | T | C | 0.2935 | 0.079 | 0.008 | 1.5849E-23 | 97.52 |
| *ASGR2* | rs35457250 | C | T | 0.0088 | 0.346 | 0.038 | 2.5119E-19 | 82.91 |
| *ASGR2* | rs1611185 | T | C | 0.4174 | -0.05 | 0.007 | 6.3096E-12 | 51.02 |
| *ASGR2* | rs4240624 | G | A | 0.908 | 0.107 | 0.012 | 1E-17 | 79.51 |
| *ASGR2* | rs28929474 | C | T | 0.0209 | 0.74 | 0.025 | 3.162E-188 | 876.16 |
| *ASGR2* | rs2304978 | C | T | 0.2898 | 0.325 | 0.008 | 1E-200 | 1650.39 |
| *ASGR2* | rs1801689 | A | C | 0.0301 | 0.841 | 0.022 | 1E-200 | 1461.32 |
| *ASGR2* | rs2733739 | A | G | 0.6275 | -0.052 | 0.007 | 1.5849E-12 | 55.18 |
| *ASPN* | rs74617384 | A | T | 0.0812 | -0.114 | 0.013 | 5.0119E-18 | 76.90 |
| *ASPN* | rs2295941 | A | G | 0.3272 | -0.257 | 0.008 | 1E-200 | 1032.02 |
| *ASPN* | rs67852477 | C | G | 0.0101 | 0.243 | 0.036 | 1.5849E-11 | 45.56 |
| *ASPN* | rs35887873 | T | C | 0.2283 | 0.076 | 0.009 | 5.0119E-19 | 71.31 |
| *ASPN* | rs1303 | T | G | 0.2552 | 0.083 | 0.008 | 1.9953E-23 | 107.64 |
| *ASPN* | rs186021206 | G | A | 0.0064 | 0.617 | 0.05 | 1.5849E-35 | 152.28 |
| *ASPSCR1* | rs1354034 | T | C | 0.6043 | 0.072 | 0.007 | 3.9811E-22 | 105.80 |
| *ASPSCR1* | rs9271358 | A | T | 0.7537 | -0.065 | 0.009 | 3.9811E-14 | 52.16 |
| *ASPSCR1* | rs342293 | C | G | 0.4596 | -0.057 | 0.007 | 7.9433E-15 | 66.31 |
| *ASPSCR1* | rs8074498 | T | A | 0.5811 | -0.213 | 0.008 | 1.259E-174 | 708.89 |
| *ASRGL1* | rs7896518 | A | G | 0.4236 | 0.069 | 0.007 | 6.3096E-21 | 97.16 |
| *ASRGL1* | rs1043785 | C | T | 0.2846 | -0.372 | 0.008 | 1E-200 | 2162.25 |
| *ASRGL1* | rs11671654 | C | G | 0.4953 | 0.288 | 0.007 | 1E-200 | 1692.73 |
| *ASS1* | rs1047891 | C | A | 0.3129 | 0.078 | 0.008 | 5.0119E-23 | 95.06 |
| *ASS1* | rs71633359 | T | C | 0.3206 | -0.055 | 0.008 | 1E-11 | 47.27 |
| *ASS1* | rs112875651 | G | A | 0.3916 | -0.056 | 0.008 | 1.2589E-13 | 49.00 |
| *ASS1* | rs10883451 | T | C | 0.5021 | -0.057 | 0.007 | 3.1623E-15 | 66.31 |
| *ASS1* | rs7117339 | C | T | 0.1173 | -0.084 | 0.011 | 1.2589E-13 | 58.31 |
| *ASS1* | rs3747207 | G | A | 0.2155 | 0.143 | 0.009 | 1.9953E-58 | 252.46 |
| *ATF2* | rs58534292 | T | C | 0.6496 | -0.07 | 0.008 | 2.5119E-17 | 76.56 |
| *ATF2* | rs2857339 | C | T | 0.4979 | 0.135 | 0.009 | 1.9953E-51 | 225.00 |
| *ATG16L1* | rs1354034 | T | C | 0.6046 | 0.052 | 0.008 | 6.3096E-12 | 42.25 |
| *ATG16L1* | rs342293 | C | G | 0.4598 | -0.058 | 0.007 | 1E-14 | 68.65 |
| *ATG16L1* | rs7896518 | A | G | 0.4237 | 0.056 | 0.008 | 3.1623E-13 | 49.00 |
| *ATG4A* | rs1354034 | T | C | 0.6038 | 0.053 | 0.008 | 1.5849E-12 | 43.89 |
| *ATG4A* | rs749679 | A | G | 0.2974 | 0.064 | 0.008 | 3.1623E-15 | 64.00 |
| *ATG4A* | rs8887 | T | C | 0.5738 | 0.064 | 0.008 | 3.9811E-17 | 64.00 |
| *ATOX1* | rs1354034 | T | C | 0.6041 | 0.06 | 0.008 | 6.3096E-15 | 56.25 |
| *ATOX1* | rs1050152 | C | T | 0.4341 | -0.052 | 0.008 | 5.0119E-12 | 42.25 |
| *ATOX1* | rs371549947 | C | T | 0.0008 | -1.447 | 0.138 | 1.2589E-25 | 109.95 |
| *ATOX1* | rs139130389 | C | CTA | 0.0726 | -0.121 | 0.014 | 5.0119E-17 | 74.70 |
| *ATOX1* | rs58534292 | T | C | 0.6494 | -0.11 | 0.008 | 1E-43 | 189.06 |
| *ATOX1* | rs73365419 | T | C | 0.6163 | -0.117 | 0.008 | 6.3096E-49 | 213.89 |
| *ATOX1* | rs892090 | T | G | 0.8343 | 0.079 | 0.01 | 6.3096E-15 | 62.41 |
| *ATP2B4* | rs607929 | C | G | 0.449 | -0.065 | 0.008 | 6.3096E-17 | 66.02 |
| *ATP5IF1* | rs510379 | T | C | 0.3598 | 0.112 | 0.008 | 1.2589E-45 | 196.00 |
| *ATP5IF1* | rs1354034 | T | C | 0.6041 | 0.061 | 0.008 | 1.2589E-15 | 58.14 |
| *ATP5IF1* | rs10733789 | T | C | 0.3132 | 0.06 | 0.008 | 1.9953E-13 | 56.25 |
| *ATP6AP2* | rs76415507 | T | G | 0.56 | -0.06 | 0.008 | 1.2589E-12 | 56.25 |
| *ATP6V1D* | rs189049184 | G | A | 0.8236 | -0.091 | 0.01 | 1.9953E-18 | 82.81 |
| *ATP6V1F* | rs1354034 | T | C | 0.6034 | 0.063 | 0.008 | 2.5119E-16 | 62.02 |
| *ATP6V1G1* | rs1354034 | T | C | 0.6045 | 0.052 | 0.008 | 1.5849E-11 | 42.25 |
| *ATP6V1G2* | rs2239705 | G | A | 0.1572 | 0.204 | 0.011 | 7.9433E-82 | 343.93 |
| *ATRAID* | rs61747728 | C | T | 0.038 | 0.155 | 0.019 | 3.1623E-16 | 66.55 |
| *ATRAID* | rs78444298 | G | A | 0.0195 | 0.434 | 0.027 | 5.0119E-60 | 258.38 |
| *ATRAID* | rs1275530 | A | G | 0.7071 | -0.279 | 0.008 | 1E-200 | 1216.27 |
| *ATRAID* | rs111285796 | T | TTCTGCCAC | 0.1826 | -0.083 | 0.009 | 1.5849E-18 | 85.05 |
| *ATRN* | rs9427104 | C | T | 0.4799 | 0.047 | 0.007 | 2.5119E-12 | 45.08 |
| *ATRN* | rs1260326 | T | C | 0.6084 | -0.06 | 0.007 | 6.3096E-18 | 73.47 |
| *ATRN* | rs751229834 | CA | C | 0.3074 | 0.053 | 0.007 | 5.0119E-13 | 57.33 |
| *ATRN* | rs11845606 | C | G | 0.061 | -0.106 | 0.014 | 1E-13 | 57.33 |
| *ATRN* | rs78971233 | T | A | 0.0534 | -1.293 | 0.016 | 1E-200 | 6530.66 |
| *ATXN10* | rs1354034 | T | C | 0.6035 | 0.074 | 0.008 | 1E-21 | 85.56 |
| *ATXN10* | rs342298 | C | T | 0.4544 | -0.054 | 0.008 | 1.2589E-12 | 45.56 |
| *ATXN10* | rs6993770 | A | T | 0.2869 | -0.064 | 0.008 | 3.1623E-14 | 64.00 |
| *ATXN10* | rs7080386 | C | A | 0.4124 | 0.056 | 0.008 | 5.0119E-13 | 49.00 |

| | | | | | | | | |
|---|---|---|---|---|---|---|---|---|
| *ATXN10* | rs138153 | G | C | 0.7067 | -0.106 | 0.008 | 1E-36 | 175.56 |
| *ATXN2L* | rs1354034 | T | C | 0.6043 | 0.073 | 0.008 | 1.2589E-21 | 83.27 |
| *ATXN3* | rs1354034 | T | C | 0.6043 | 0.051 | 0.007 | 3.1623E-12 | 53.08 |
| *ATXN3* | rs55656743 | A | G | 0.2447 | -0.307 | 0.008 | 1E-200 | 1472.64 |
| *AXIN1* | rs1354034 | T | C | 0.6043 | 0.057 | 0.008 | 3.9811E-14 | 50.77 |
| *AXIN1* | rs6993770 | A | T | 0.2869 | -0.062 | 0.008 | 1.5849E-14 | 60.06 |
| *AXL* | rs79412885 | G | A | 0.0742 | -0.164 | 0.014 | 1.2589E-30 | 137.22 |
| *AXL* | rs2470855 | G | A | 0.5743 | -0.054 | 0.007 | 2.5119E-13 | 59.51 |
| *AXL* | rs35350651 | A | AC | 0.5038 | -0.052 | 0.007 | 7.9433E-13 | 55.18 |
| *AXL* | rs7140110 | T | C | 0.2957 | 0.127 | 0.008 | 5.0119E-56 | 252.02 |
| *AXL* | rs186021206 | G | A | 0.0064 | 0.991 | 0.05 | 1.9953E-86 | 392.83 |
| *AXL* | rs7224668 | T | C | 0.5575 | -0.053 | 0.007 | 7.9433E-13 | 57.33 |
| *AXL* | rs66841352 | G | C | 0.4026 | -0.174 | 0.008 | 1.995E-118 | 473.06 |
| *AZI2* | rs1354034 | T | C | 0.6043 | 0.085 | 0.008 | 6.3096E-30 | 112.89 |
| *AZU1* | rs2210918 | T | C | 0.4715 | 0.051 | 0.007 | 7.9433E-12 | 53.08 |
| *AZU1* | rs2951842 | C | G | 0.5037 | -0.062 | 0.008 | 6.3096E-16 | 60.06 |
| *AZU1* | rs744781 | G | C | 0.4738 | -0.059 | 0.007 | 2.5119E-15 | 71.04 |
| *B2M* | rs7137828 | C | T | 0.516 | -0.099 | 0.007 | 2.5119E-44 | 200.02 |
| *B2M* | rs74439742 | C | T | 0.172 | -0.087 | 0.009 | 3.9811E-20 | 93.44 |
| *B3GNT7* | rs583104 | G | T | 0.7733 | 0.069 | 0.008 | 2.5119E-19 | 74.39 |
| *B3GNT7* | rs2290130 | G | A | 0.2296 | -0.747 | 0.009 | 1E-200 | 6889.00 |
| *B3GNT7* | rs13107325 | C | T | 0.075 | 0.114 | 0.012 | 2.5119E-20 | 90.25 |
| *B3GNT7* | rs6908045 | C | T | 0.5292 | -0.053 | 0.006 | 5.0119E-16 | 78.03 |
| *B3GNT7* | rs111630162 | T | C | 0.0078 | -0.441 | 0.037 | 5.0119E-33 | 142.06 |
| *B3GNT7* | rs79578725 | G | A | 0.0423 | 0.17 | 0.016 | 3.9811E-25 | 112.89 |
| *B3GNT7* | rs7865362 | C | T | 0.3625 | -0.07 | 0.007 | 1.5849E-24 | 100.00 |
| *B3GNT7* | rs56278466 | T | G | 0.6614 | 0.096 | 0.007 | 1.9953E-44 | 188.08 |
| *B3GNT7* | rs1955512 | G | A | 0.5723 | 0.148 | 0.007 | 1.995E-108 | 447.02 |
| *B3GNT7* | rs12881755 | A | G | 0.6851 | -0.05 | 0.007 | 7.9433E-13 | 51.02 |
| *B3GNT7* | rs201796794 | A | AG | 0.2502 | -0.064 | 0.007 | 7.9433E-18 | 83.59 |
| *B3GNT7* | rs200489612 | G | A | 0.0053 | 0.413 | 0.048 | 1.5849E-17 | 74.03 |
| *B3GNT7* | rs34321000 | C | T | 0.6714 | -0.054 | 0.007 | 3.1623E-15 | 59.51 |
| *B3GNT7* | rs516246 | C | T | 0.5094 | -0.044 | 0.007 | 7.9433E-12 | 39.51 |
| *B4GALT1* | rs61747728 | C | T | 0.0379 | 0.164 | 0.019 | 3.1623E-18 | 74.50 |
| *B4GALT1* | rs17138646 | T | G | 0.1155 | 0.1 | 0.011 | 1E-18 | 82.64 |
| *B4GALT1* | rs56249455 | T | A | 0.114 | -0.097 | 0.012 | 1.9953E-16 | 65.34 |
| *B4GALT1* | rs7019909 | C | T | 0.1023 | 0.332 | 0.012 | 3.162E-167 | 765.44 |
| *B4GALT1* | rs550057 | C | T | 0.2552 | 0.1 | 0.008 | 1.5849E-32 | 156.25 |
| *B4GALT1* | rs7247412 | T | C | 0.2669 | -0.323 | 0.008 | 1E-200 | 1630.14 |
| *B4GAT1* | rs10168551 | T | C | 0.2046 | -0.06 | 0.009 | 1E-11 | 44.44 |
| *B4GAT1* | rs6800514 | C | T | 0.3712 | 0.057 | 0.007 | 1.5849E-14 | 66.31 |
| *B4GAT1* | rs9274678 | A | G | 0.1531 | -0.094 | 0.01 | 1.2589E-21 | 88.36 |
| *B4GAT1* | rs10758194 | G | T | 0.4214 | -0.078 | 0.007 | 2.5119E-27 | 124.16 |
| *B4GAT1* | rs56278466 | T | G | 0.6612 | 0.1 | 0.008 | 1.9953E-40 | 156.25 |
| *B4GAT1* | rs10896113 | C | T | 0.2216 | -0.316 | 0.009 | 1E-200 | 1232.79 |
| *B4GAT1* | rs74611001 | C | A | 0.0236 | -0.205 | 0.024 | 1.2589E-17 | 72.96 |
| *B4GAT1* | rs28873836 | G | C | 0.266 | -0.198 | 0.008 | 1.259E-130 | 612.56 |
| *B4GAT1* | rs12975366 | T | C | 0.3954 | -0.054 | 0.007 | 2.5119E-13 | 59.51 |
| *BACH1* | rs342293 | C | G | 0.4594 | -0.05 | 0.007 | 1.5849E-11 | 51.02 |
| *BACH1* | rs6993770 | A | T | 0.2869 | -0.061 | 0.008 | 6.3096E-14 | 58.14 |
| *BAG3* | rs710446 | T | C | 0.4092 | -0.066 | 0.008 | 6.3096E-18 | 68.06 |
| *BAG3* | rs3733402 | G | A | 0.5124 | -0.091 | 0.008 | 3.1623E-33 | 129.39 |
| *BAG3* | rs2545801 | T | C | 0.7442 | -0.069 | 0.009 | 1E-15 | 58.78 |
| *BAG3* | rs35434411 | G | A | 0.0311 | 0.178 | 0.022 | 1.9953E-16 | 65.46 |
| *BAG3* | rs55714647 | G | GA | 0.934 | -0.107 | 0.015 | 3.9811E-12 | 50.88 |
| *BAG4* | rs1354034 | T | C | 0.6039 | 0.065 | 0.008 | 3.9811E-17 | 66.02 |
| *BAG4* | rs4632248 | G | T | 0.2097 | 0.079 | 0.009 | 2.5119E-17 | 77.05 |
| *BAG6* | rs7916868 | A | T | 0.5016 | 0.052 | 0.007 | 1.9953E-12 | 55.18 |
| *BAIAP2* | rs28456 | A | G | 0.3146 | 0.055 | 0.008 | 1.2589E-11 | 47.27 |
| *BAIAP2* | rs3741513 | T | A | 0.6107 | 0.079 | 0.008 | 3.9811E-24 | 97.52 |
| *BAIAP2* | rs147121761 | C | T | 0.282 | 0.078 | 0.009 | 1.9953E-19 | 75.11 |
| *BAIAP2* | rs9989483 | C | T | 0.1728 | 0.095 | 0.01 | 1.9953E-21 | 90.25 |
| *BAIAP2* | rs189448562 | G | C | 0.0113 | 0.406 | 0.037 | 1E-28 | 120.41 |
| *BAMBI* | rs1354034 | T | C | 0.6038 | -0.064 | 0.008 | 1.2589E-16 | 64.00 |
| *BANK1* | rs1354034 | T | C | 0.6043 | 0.073 | 0.008 | 2.5119E-22 | 83.27 |
| *BANK1* | rs3113676 | T | C | 0.985 | 0.478 | 0.03 | 6.3096E-56 | 253.87 |
| *BANK1* | rs342293 | C | G | 0.4594 | -0.057 | 0.007 | 1.5849E-14 | 66.31 |
| *BAP18* | rs1354034 | T | C | 0.6043 | 0.083 | 0.008 | 5.0119E-28 | 107.64 |

| | | | | | | | | |
|---|---|---|---|---|---|---|---|---|
| *BAP18* | rs76483802 | A | G | 0.3393 | -0.075 | 0.008 | 3.9811E-20 | 87.89 |
| *BAP18* | rs7137828 | C | T | 0.5163 | -0.05 | 0.007 | 1.2589E-11 | 51.02 |
| *BAP18* | rs8065014 | G | A | 0.0788 | -0.112 | 0.014 | 3.1623E-16 | 64.00 |
| *BAX* | rs1354034 | T | C | 0.6036 | 0.059 | 0.008 | 2.5119E-14 | 54.39 |
| *BAX* | rs12245367 | C | T | 0.4812 | -0.052 | 0.008 | 1.2589E-11 | 42.25 |
| *BCAM* | rs539657009 | C | T | 0.1298 | 0.078 | 0.011 | 5.0119E-13 | 50.28 |
| *BCAM* | rs4681846 | T | C | 0.629 | -0.07 | 0.007 | 3.9811E-22 | 100.00 |
| *BCAM* | rs11100831 | T | C | 0.4407 | 0.124 | 0.007 | 3.9811E-67 | 313.80 |
| *BCAM* | rs73080552 | G | A | 0.1105 | -0.081 | 0.011 | 5.0119E-13 | 54.22 |
| *BCAM* | rs12522289 | A | G | 0.0798 | -0.091 | 0.013 | 5.0119E-12 | 49.00 |
| *BCAM* | rs9270591 | C | T | 0.5298 | -0.065 | 0.007 | 2.5119E-20 | 86.22 |
| *BCAM* | rs778641135 | TTAC | T | 0.2573 | -0.063 | 0.008 | 1E-14 | 62.02 |
| *BCAM* | rs6592965 | G | A | 0.4539 | -0.083 | 0.007 | 5.0119E-32 | 140.59 |
| *BCAM* | rs8176672 | C | T | 0.0612 | 0.663 | 0.015 | 1E-200 | 1953.64 |
| *BCAM* | rs56278466 | T | G | 0.6612 | 0.159 | 0.007 | 1.585E-101 | 515.94 |
| *BCAM* | rs150158762 | A | AT | 0.1001 | 0.096 | 0.012 | 1.2589E-15 | 64.00 |
| *BCAM* | rs2137537 | T | C | 0.5482 | -0.048 | 0.007 | 1E-11 | 47.02 |
| *BCAM* | rs186021206 | G | A | 0.0064 | 0.378 | 0.048 | 3.1623E-15 | 62.02 |
| *BCAM* | rs11085824 | A | G | 0.3784 | 0.117 | 0.007 | 2.5119E-56 | 279.37 |
| *BCAM* | rs28399654 | G | A | 0.0332 | -0.654 | 0.02 | 1E-200 | 1069.29 |
| *BCAM* | rs2638282 | G | A | 0.5337 | 0.051 | 0.007 | 1.5849E-12 | 53.08 |
| *BCAM* | rs2410405 | C | T | 0.3776 | 0.111 | 0.007 | 1E-52 | 251.45 |
| *BCAN* | rs2365715 | A | G | 0.3903 | -0.231 | 0.008 | 1.585E-191 | 833.77 |
| *BCAN* | rs13086758 | T | A | 0.2142 | -0.066 | 0.009 | 1.2589E-13 | 53.78 |
| *BCAN* | rs2071592 | T | A | 0.3469 | -0.061 | 0.008 | 1.2589E-15 | 58.14 |
| *BCAN* | rs73086586 | C | T | 0.0612 | 0.154 | 0.015 | 3.9811E-24 | 105.40 |
| *BCAN* | rs61738649 | C | T | 0.0713 | -0.13 | 0.014 | 2.5119E-20 | 86.22 |
| *BCAN* | rs56278466 | T | G | 0.6614 | 0.128 | 0.008 | 1.9953E-62 | 256.00 |
| *BCAT1* | rs111466273 | A | G | 0.0059 | 0.479 | 0.047 | 3.1623E-24 | 103.87 |
| *BCAT1* | rs1354034 | T | C | 0.6045 | 0.055 | 0.007 | 7.9433E-14 | 61.73 |
| *BCAT1* | rs6993770 | A | T | 0.2866 | -0.067 | 0.008 | 1.9953E-17 | 70.14 |
| *BCAT1* | rs11047639 | G | T | 0.2952 | 0.282 | 0.008 | 1E-200 | 1242.56 |
| *BCHE* | rs70620 | G | A | 0.1722 | 0.067 | 0.009 | 6.3096E-14 | 55.42 |
| *BCHE* | rs1260326 | T | C | 0.6083 | -0.066 | 0.007 | 5.0119E-21 | 88.90 |
| *BCHE* | rs11687186 | C | G | 0.5517 | 0.161 | 0.007 | 5.012E-121 | 529.00 |
| *BCHE* | rs111696008 | G | A | 0.0173 | 0.65 | 0.027 | 7.943E-133 | 579.56 |
| *BCHE* | rs887829 | C | T | 0.3136 | -0.075 | 0.007 | 1.2589E-24 | 114.80 |
| *BCHE* | rs73165060 | C | A | 0.2095 | -0.585 | 0.009 | 1E-200 | 4225.00 |
| *BCHE* | rs183545662 | C | T | 0.0057 | 0.413 | 0.045 | 6.3096E-20 | 84.23 |
| *BCHE* | rs2516448 | T | C | 0.4937 | 0.057 | 0.007 | 3.1623E-17 | 66.31 |
| *BCHE* | rs10502 | G | C | 0.3808 | -0.053 | 0.007 | 2.5119E-14 | 57.33 |
| *BCHE* | rs72941804 | A | G | 0.0492 | 0.118 | 0.016 | 5.0119E-14 | 54.39 |
| *BCHE* | rs35493868 | C | G | 0.2012 | -0.078 | 0.008 | 1.9953E-20 | 95.06 |
| *BCHE* | rs4841132 | A | G | 0.908 | 0.155 | 0.012 | 6.3096E-40 | 166.84 |
| *BCHE* | rs7896518 | A | G | 0.4236 | -0.065 | 0.007 | 5.0119E-21 | 86.22 |
| *BCHE* | rs174568 | C | T | 0.3435 | 0.07 | 0.007 | 6.3096E-23 | 100.00 |
| *BCL2* | rs72700845 | C | T | 0.0171 | 0.225 | 0.029 | 7.9433E-15 | 60.20 |
| *BCL2* | rs445925 | G | A | 0.1095 | 0.084 | 0.012 | 2.5119E-12 | 49.00 |
| *BCL2L1* | rs1354034 | T | C | 0.6043 | 0.06 | 0.008 | 3.9811E-15 | 56.25 |
| *BCL2L1* | rs6993770 | A | T | 0.2865 | -0.065 | 0.008 | 6.3096E-15 | 66.02 |
| *BCL2L11* | rs13412535 | G | A | 0.2307 | -0.064 | 0.009 | 1.9953E-13 | 50.57 |
| *BCL2L11* | rs1354034 | T | C | 0.6043 | -0.054 | 0.007 | 5.0119E-13 | 59.51 |
| *BCL2L11* | rs5030062 | A | C | 0.3728 | -0.157 | 0.008 | 1.2589E-94 | 385.14 |
| *BCL2L11* | rs66530140 | T | C | 0.5137 | -0.293 | 0.007 | 1E-200 | 1752.02 |
| *BCL2L11* | rs1801020 | A | G | 0.745 | -0.154 | 0.008 | 6.3096E-76 | 370.56 |
| *BCL2L11* | rs1050328 | G | A | 0.2656 | -0.056 | 0.008 | 1.2589E-11 | 49.00 |
| *BCL2L11* | rs6993770 | A | T | 0.2869 | -0.104 | 0.008 | 3.1623E-38 | 169.00 |
| *BCL2L11* | rs10761737 | T | C | 0.4131 | 0.081 | 0.007 | 5.0119E-28 | 133.90 |
| *BCL2L11* | rs71354995 | A | G | 0.2564 | -0.212 | 0.008 | 3.981E-142 | 702.25 |
| *BCL2L11* | rs892090 | T | G | 0.8337 | 0.108 | 0.01 | 6.3096E-28 | 116.64 |
| *BCL2L15* | rs1230715 | A | C | 0.6731 | 0.177 | 0.008 | 1.585E-108 | 489.52 |
| *BCL2L15* | rs35440906 | C | CA | 0.1856 | 0.087 | 0.01 | 1.2589E-19 | 75.69 |
| *BCL2L15* | rs34436714 | C | A | 0.2092 | 0.09 | 0.009 | 7.9433E-23 | 100.00 |
| *BCL7B* | rs607929 | C | G | 0.4492 | -0.055 | 0.008 | 1.5849E-12 | 47.27 |
| *BCR* | rs149005797 | T | C | 0.0042 | -0.541 | 0.066 | 3.1623E-16 | 67.19 |
| *BDNF* | rs6671171 | G | A | 0.1112 | 0.084 | 0.012 | 1.2589E-12 | 49.00 |
| *BDNF* | rs13412535 | G | A | 0.2311 | -0.061 | 0.009 | 3.9811E-12 | 45.94 |
| *BDNF* | rs1354034 | T | C | 0.6046 | -0.114 | 0.008 | 3.1623E-51 | 203.06 |

| | | | | | | | | |
|---|---|---|---|---|---|---|---|---|
| *BDNF* | rs2278668 | T | C | 0.5933 | -0.052 | 0.008 | 1.2589E-11 | 42.25 |
| *BDNF* | rs755492124 | TAAAG | T | 0.3936 | -0.068 | 0.008 | 3.9811E-19 | 72.25 |
| *BDNF* | rs274555 | C | T | 0.3989 | 0.057 | 0.008 | 5.0119E-14 | 50.77 |
| *BDNF* | rs2516471 | G | A | 0.0636 | -0.121 | 0.015 | 3.1623E-15 | 65.07 |
| *BDNF* | rs146842698 | T | TAAATA | 0.305 | -0.059 | 0.008 | 6.3096E-13 | 54.39 |
| *BDNF* | rs6961069 | C | T | 0.4024 | 0.063 | 0.008 | 1E-16 | 62.02 |
| *BDNF* | rs6993770 | A | T | 0.2863 | -0.132 | 0.008 | 2.5119E-58 | 272.25 |
| *BDNF* | rs9410207 | T | C | 0.0652 | -0.104 | 0.015 | 6.3096E-12 | 48.07 |
| *BDNF* | rs61751937 | G | C | 0.0283 | 0.158 | 0.022 | 1.2589E-12 | 51.58 |
| *BDNF* | rs7080386 | C | A | 0.4122 | 0.114 | 0.008 | 3.1623E-51 | 203.06 |
| *BDNF* | rs1984541 | G | C | 0.4916 | -0.056 | 0.007 | 3.9811E-14 | 64.00 |
| *BDNF* | rs12445050 | C | T | 0.1383 | 0.105 | 0.011 | 1.2589E-22 | 91.12 |
| *BDNF* | rs1654425 | T | C | 0.8344 | 0.16 | 0.01 | 1.9953E-57 | 256.00 |
| *BDNF* | rs3827978 | C | T | 0.3533 | 0.075 | 0.008 | 6.3096E-22 | 87.89 |
| *BECN1* | rs1354034 | T | C | 0.6045 | 0.071 | 0.008 | 1.5849E-20 | 78.77 |
| *BECN1* | rs142201367 | T | TTTTCTAAC | 0.4961 | 0.07 | 0.008 | 3.9811E-20 | 76.56 |
| *BECN1* | rs1801020 | A | G | 0.7445 | 0.062 | 0.009 | 3.9811E-13 | 47.46 |
| *BECN1* | rs72829144 | C | A | 0.0334 | 0.216 | 0.021 | 2.5119E-25 | 105.80 |
| *BGLAP* | rs369453512 | G | GA | 0.5544 | 0.136 | 0.008 | 7.9433E-74 | 289.00 |
| *BGLAP* | rs541618 | A | G | 0.4586 | 0.052 | 0.007 | 3.1623E-12 | 55.18 |
| *BGLAP* | rs12331051 | C | T | 0.4283 | 0.052 | 0.008 | 1.5849E-11 | 42.25 |
| *BGLAP* | rs12209125 | A | C | 0.4051 | -0.052 | 0.008 | 3.1623E-12 | 42.25 |
| *BGLAP* | rs13264172 | T | A | 0.528 | 0.072 | 0.007 | 2.5119E-22 | 105.80 |
| *BGLAP* | rs548589580 | T | TA | 0.4838 | 0.079 | 0.007 | 6.3096E-27 | 127.37 |
| *BGLAP* | rs12597511 | C | T | 0.3528 | -0.069 | 0.008 | 3.9811E-19 | 74.39 |
| *BGLAP* | rs7412 | C | T | 0.0814 | -0.109 | 0.014 | 6.3096E-16 | 60.62 |
| *BGN* | rs744233 | G | A | 0.2947 | 0.065 | 0.008 | 3.9811E-15 | 66.02 |
| *BGN* | rs3756074 | G | C | 0.0495 | 0.153 | 0.018 | 6.3096E-18 | 72.25 |
| *BGN* | rs4463068 | A | T | 0.3979 | 0.06 | 0.008 | 5.0119E-15 | 56.25 |
| *BGN* | rs9266095 | A | G | 0.2119 | 0.09 | 0.009 | 3.9811E-22 | 100.00 |
| *BGN* | rs6993770 | A | T | 0.2869 | 0.096 | 0.008 | 1E-30 | 144.00 |
| *BGN* | rs7080386 | C | A | 0.4119 | -0.068 | 0.008 | 7.9433E-19 | 72.25 |
| *BGN* | rs2511241 | C | T | 0.9296 | -0.232 | 0.015 | 1.5849E-55 | 239.22 |
| *BGN* | rs77242536 | T | C | 0.1437 | 0.075 | 0.011 | 5.0119E-12 | 46.49 |
| *BGN* | rs780989226 | TA | T | 0.1099 | 0.122 | 0.012 | 6.3096E-24 | 103.36 |
| *BGN* | rs61758388 | C | A | 0.032 | -0.368 | 0.021 | 1.2589E-65 | 307.08 |
| *BGN* | rs1654425 | T | C | 0.8339 | -0.115 | 0.01 | 2.5119E-29 | 132.25 |
| *BGN* | rs6081555 | G | T | 0.3429 | -0.057 | 0.008 | 1.5849E-12 | 50.77 |
| *BID* | rs142201367 | T | TTTTCTAAC | 0.4958 | 0.083 | 0.008 | 3.9811E-27 | 107.64 |
| *BID* | rs10418046 | T | G | 0.2112 | 0.096 | 0.009 | 3.1623E-25 | 113.78 |
| *BID* | rs183393610 | G | A | 0.007 | -0.413 | 0.048 | 1E-17 | 74.03 |
| *BIN2* | rs1354034 | T | C | 0.6041 | 0.063 | 0.008 | 7.9433E-17 | 62.02 |
| *BIN2* | rs2631367 | C | G | 0.5188 | 0.054 | 0.007 | 6.3096E-13 | 59.51 |
| *BIN2* | rs6993770 | A | T | 0.2864 | -0.063 | 0.008 | 1.2589E-14 | 62.02 |
| *BIN2* | rs149960700 | C | CTTATTTA | 0.1077 | -0.184 | 0.012 | 1.9953E-52 | 235.11 |
| *BIRC2* | rs1354034 | T | C | 0.6038 | 0.053 | 0.008 | 5.0119E-12 | 43.89 |
| *BIRC2* | rs10761785 | G | T | 0.5105 | 0.051 | 0.008 | 1.2589E-11 | 40.64 |
| *BIRC2* | rs61754131 | A | G | 0.002 | -0.677 | 0.085 | 1.2589E-15 | 63.44 |
| *BLMH* | rs1250258 | C | T | 0.7368 | 0.062 | 0.008 | 7.9433E-16 | 60.06 |
| *BLMH* | rs915651 | G | A | 0.1306 | -0.083 | 0.01 | 1.5849E-16 | 68.89 |
| *BLMH* | rs1050565 | T | C | 0.3319 | 0.579 | 0.008 | 1E-200 | 5238.14 |
| *BLNK* | rs4954569 | C | T | 0.2522 | -0.09 | 0.009 | 5.0119E-24 | 100.00 |
| *BLNK* | rs4566648 | T | G | 0.3383 | -0.107 | 0.008 | 7.9433E-40 | 178.89 |
| *BLNK* | rs55769428 | A | C | 0.0381 | -0.149 | 0.02 | 1.2589E-13 | 55.50 |
| *BLNK* | rs76428106 | T | C | 0.0129 | 0.24 | 0.035 | 1E-11 | 47.02 |
| *BLVRB* | rs11759553 | A | T | 0.2719 | -0.078 | 0.008 | 1.5849E-21 | 95.06 |
| *BLVRB* | rs149698066 | G | A | 0.006 | -1.466 | 0.052 | 7.943E-172 | 794.81 |
| *BMP10* | rs34008398 | G | A | 0.0128 | -2.019 | 0.032 | 1E-200 | 3980.82 |
| *BMP10* | rs7848055 | A | C | 0.3291 | -0.153 | 0.007 | 5.0119E-97 | 477.73 |
| *BMP10* | rs10740131 | A | T | 0.4707 | 0.057 | 0.007 | 1E-16 | 66.31 |
| *BMP10* | rs67852477 | C | G | 0.0101 | 0.298 | 0.034 | 3.1623E-18 | 76.82 |
| *BMP10* | rs8022092 | C | A | 0.5025 | -0.051 | 0.007 | 1E-13 | 53.08 |
| *BMP10* | rs1956713 | G | A | 0.8513 | -0.13 | 0.01 | 3.9811E-41 | 169.00 |
| *BMP10* | rs4702 | G | A | 0.5577 | 0.056 | 0.007 | 3.1623E-16 | 64.00 |
| *BMP10* | rs186021206 | G | A | 0.0064 | 0.458 | 0.047 | 1.9953E-22 | 94.96 |
| *BMP4* | rs535025308 | G | C | 0.0411 | -0.351 | 0.019 | 1.5849E-75 | 341.28 |
| *BMP4* | rs12284180 | A | C | 0.2459 | 0.067 | 0.009 | 3.1623E-14 | 55.42 |
| *BMP4* | rs11111765 | G | C | 0.2578 | 0.062 | 0.009 | 5.0119E-13 | 47.46 |

| | | | | | | | | |
|---|---|---|---|---|---|---|---|---|
| *BMP4* | rs186021206 | G | A | 0.0064 | 0.754 | 0.052 | 2.5119E-48 | 210.25 |
| *BMP4* | rs704 | G | A | 0.4733 | -0.056 | 0.008 | 1.2589E-13 | 49.00 |
| *BMP6* | rs13412535 | G | A | 0.2312 | -0.063 | 0.009 | 1.2589E-12 | 49.00 |
| *BMP6* | rs1354034 | T | C | 0.6039 | -0.209 | 0.008 | 5.012E-164 | 682.52 |
| *BMP6* | rs3792386 | G | A | 0.5866 | -0.053 | 0.008 | 1E-11 | 43.89 |
| *BMP6* | rs5030062 | A | C | 0.3737 | -0.085 | 0.008 | 2.5119E-27 | 112.89 |
| *BMP6* | rs71640034 | G | A | 0.5137 | -0.148 | 0.008 | 1.2589E-85 | 342.25 |
| *BMP6* | rs2545801 | T | C | 0.7445 | -0.095 | 0.009 | 1.5849E-28 | 111.42 |
| *BMP6* | rs71542879 | C | CT | 0.4672 | 0.135 | 0.008 | 5.0119E-71 | 284.77 |
| *BMP6* | rs6993770 | A | T | 0.2866 | -0.08 | 0.008 | 3.1623E-22 | 100.00 |
| *BMP6* | rs10820606 | A | C | 0.2298 | -0.109 | 0.009 | 3.9811E-33 | 146.68 |
| *BMP6* | rs12762934 | C | T | 0.3249 | 0.102 | 0.008 | 1.9953E-37 | 162.56 |
| *BMP6* | rs535025308 | G | C | 0.0412 | -0.183 | 0.019 | 7.9433E-22 | 92.77 |
| *BMP6* | rs6580981 | G | A | 0.4584 | 0.072 | 0.008 | 1.2589E-21 | 81.00 |
| *BMP6* | rs186021206 | G | A | 0.0064 | 0.441 | 0.052 | 1.5849E-17 | 71.92 |
| *BMP6* | rs892090 | T | G | 0.8339 | 0.096 | 0.01 | 2.5119E-21 | 92.16 |
| *BMP6* | rs2834720 | G | A | 0.2731 | 0.062 | 0.008 | 1.9953E-13 | 60.06 |
| *BMPER* | rs79419884 | A | G | 0.1101 | 0.085 | 0.012 | 7.9433E-13 | 50.17 |
| *BMPER* | rs201897458 | C | T | 0.0781 | -0.324 | 0.014 | 5.012E-114 | 535.59 |
| *BMPER* | rs4418728 | G | T | 0.4513 | 0.064 | 0.007 | 1.9953E-17 | 83.59 |
| *BMPER* | rs186021206 | G | A | 0.0064 | 0.444 | 0.051 | 3.9811E-18 | 75.79 |
| *BNIP2* | rs6993770 | A | T | 0.2866 | -0.057 | 0.008 | 1E-11 | 50.77 |
| *BNIP3L* | rs201672609 | C | CAAAAT | 0.6377 | 0.054 | 0.008 | 1E-11 | 45.56 |
| *BNIP3L* | rs7080536 | G | A | 0.0437 | -0.154 | 0.019 | 1.2589E-16 | 65.70 |
| *BOC* | rs1260326 | T | C | 0.6088 | 0.064 | 0.008 | 7.9433E-17 | 64.00 |
| *BOC* | rs3856718 | G | A | 0.1708 | 0.207 | 0.01 | 1.5849E-94 | 428.49 |
| *BOC* | rs34437827 | A | G | 0.1947 | 0.066 | 0.009 | 2.5119E-12 | 53.78 |
| *BOC* | rs56408342 | G | A | 0.0535 | -0.144 | 0.017 | 6.3096E-18 | 71.75 |
| *BOC* | rs8176749 | C | T | 0.0614 | 0.129 | 0.016 | 1E-16 | 65.00 |
| *BOC* | rs145878042 | A | G | 0.0109 | 0.242 | 0.036 | 1.5849E-11 | 45.19 |
| *BOC* | rs1037117 | G | A | 0.2523 | -0.063 | 0.009 | 2.5119E-13 | 49.00 |
| *BOC* | rs186021206 | G | A | 0.0063 | 0.555 | 0.052 | 5.0119E-27 | 113.91 |
| *BOLA1* | rs72708157 | G | A | 0.0818 | -0.192 | 0.014 | 1.5849E-45 | 188.08 |
| *BOLA1* | rs1354034 | T | C | 0.6045 | 0.06 | 0.008 | 1.9953E-15 | 56.25 |
| *BOLA2_BOLA2B* | rs1354034 | T | C | 0.6045 | 0.057 | 0.008 | 3.1623E-14 | 50.77 |
| *BOLA2_BOLA2B* | rs111894927 | A | C | 0.0097 | -0.383 | 0.041 | 3.1623E-21 | 87.26 |
| *BPIFA2* | rs188261292 | G | T | 0.0034 | 0.822 | 0.067 | 1.2589E-34 | 150.52 |
| *BPIFA2* | rs6427943 | G | A | 0.4126 | 0.06 | 0.007 | 3.1623E-16 | 73.47 |
| *BPIFA2* | rs1624793 | A | G | 0.4646 | 0.067 | 0.007 | 3.9811E-20 | 91.61 |
| *BPIFA2* | rs103855 | G | A | 0.8999 | -0.081 | 0.012 | 1.5849E-11 | 45.56 |
| *BPIFA2* | rs1839091 | T | C | 0.6693 | -0.072 | 0.008 | 6.3096E-21 | 81.00 |
| *BPIFA2* | rs10435096 | T | C | 0.0951 | 0.091 | 0.012 | 1.9953E-13 | 57.51 |
| *BPIFA2* | rs67740686 | G | A | 0.1149 | -0.078 | 0.011 | 7.9433E-12 | 50.28 |
| *BPIFA2* | rs505922 | T | C | 0.3171 | -0.061 | 0.008 | 2.5119E-15 | 58.14 |
| *BPIFA2* | rs1013311 | G | T | 0.2709 | -0.182 | 0.008 | 1E-108 | 517.56 |
| *BPIFA2* | rs247617 | C | A | 0.3238 | 0.114 | 0.008 | 6.3096E-49 | 203.06 |
| *BPIFA2* | rs34691084 | G | GT | 0.1303 | 0.091 | 0.011 | 3.9811E-16 | 68.44 |
| *BPIFA2* | rs4843719 | C | A | 0.3089 | 0.072 | 0.008 | 6.3096E-20 | 81.00 |
| *BPIFA2* | rs17855739 | C | T | 0.0414 | 0.146 | 0.019 | 1.5849E-14 | 59.05 |
| *BPIFA2* | rs395908 | G | A | 0.1699 | 0.075 | 0.01 | 1.5849E-13 | 56.25 |
| *BPIFA2* | rs516316 | G | C | 0.5094 | 0.434 | 0.008 | 1E-200 | 2943.06 |
| *BPIFA2* | rs6059134 | T | A | 0.0367 | -0.35 | 0.019 | 2.5119E-73 | 339.34 |
| *BPIFA2* | rs4810479 | C | T | 0.7497 | -0.073 | 0.008 | 1.9953E-18 | 83.27 |
| *BPIFB1* | rs1619599 | G | A | 0.583 | 0.084 | 0.007 | 1.9953E-34 | 144.00 |
| *BPIFB1* | rs7429705 | C | T | 0.7037 | 0.064 | 0.007 | 1E-17 | 83.59 |
| *BPIFB1* | rs791361 | A | C | 0.773 | 0.069 | 0.008 | 1.5849E-17 | 74.39 |
| *BPIFB1* | rs34978289 | C | CT | 0.5168 | 0.084 | 0.007 | 1.9953E-35 | 144.00 |
| *BPIFB1* | rs9399137 | T | C | 0.258 | 0.104 | 0.008 | 5.0119E-41 | 169.00 |
| *BPIFB1* | rs1822200 | G | T | 0.2619 | 0.079 | 0.008 | 5.0119E-25 | 97.52 |
| *BPIFB1* | rs2721961 | T | G | 0.2831 | -0.058 | 0.008 | 3.1623E-14 | 52.56 |
| *BPIFB1* | rs2980888 | T | C | 0.6978 | 0.053 | 0.007 | 7.9433E-13 | 57.33 |
| *BPIFB1* | rs7040302 | T | G | 0.4069 | -0.054 | 0.007 | 1.9953E-15 | 59.51 |
| *BPIFB1* | rs569255485 | G | GT | 0.3366 | -0.053 | 0.007 | 3.1623E-13 | 57.33 |
| *BPIFB1* | rs247617 | C | A | 0.3243 | 0.111 | 0.007 | 7.9433E-53 | 251.45 |
| *BPIFB1* | rs12597477 | A | G | 0.1962 | -0.068 | 0.009 | 2.5119E-15 | 57.09 |
| *BPIFB1* | rs116843064 | G | A | 0.0196 | 0.199 | 0.024 | 2.5119E-16 | 68.75 |
| *BPIFB1* | rs681343 | C | T | 0.5091 | 0.084 | 0.007 | 1.9953E-35 | 144.00 |
| *BPIFB1* | rs2424963 | T | C | 0.5822 | -0.422 | 0.007 | 1E-200 | 3634.37 |

| | | | | | | | | |
|---|---|---|---|---|---|---|---|---|
| *BPIFB2* | rs469864 | T | C | 0.2095 | -0.062 | 0.009 | 1.2589E-12 | 47.46 |
| *BPIFB2* | rs553427 | C | T | 0.5205 | 0.053 | 0.007 | 5.0119E-14 | 57.33 |
| *BPIFB2* | rs11709077 | G | A | 0.1198 | -0.074 | 0.011 | 1.2589E-11 | 45.26 |
| *BPIFB2* | rs547113723 | G | T | 0.0081 | -0.469 | 0.04 | 3.9811E-31 | 137.48 |
| *BPIFB2* | rs17145750 | C | T | 0.1601 | 0.073 | 0.01 | 3.9811E-14 | 53.29 |
| *BPIFB2* | rs174535 | T | C | 0.3507 | 0.064 | 0.007 | 3.9811E-18 | 83.59 |
| *BPIFB2* | rs1169294 | G | A | 0.3085 | 0.133 | 0.008 | 3.1623E-67 | 276.39 |
| *BPIFB2* | rs11147230 | G | C | 0.5083 | 0.053 | 0.007 | 1E-13 | 57.33 |
| *BPIFB2* | rs62011287 | A | G | 0.3424 | 0.06 | 0.007 | 1E-15 | 73.47 |
| *BPIFB2* | rs11444334 | A | AC | 0.5023 | 0.08 | 0.007 | 1.5849E-29 | 130.61 |
| *BPIFB2* | rs1801689 | A | C | 0.0301 | 0.155 | 0.021 | 6.3096E-14 | 54.48 |
| *BPIFB2* | rs429358 | T | C | 0.1553 | -0.149 | 0.01 | 1.2589E-51 | 222.01 |
| *BPIFB2* | rs2377736 | T | C | 0.4027 | 0.265 | 0.008 | 1E-200 | 1097.27 |
| *BRAP* | rs1354034 | T | C | 0.6044 | 0.076 | 0.008 | 1.2589E-23 | 90.25 |
| *BRAP* | rs342298 | C | T | 0.4553 | -0.064 | 0.007 | 1E-17 | 83.59 |
| *BRAP* | rs7896518 | A | G | 0.4238 | 0.052 | 0.008 | 7.9433E-12 | 42.25 |
| *BRDT* | rs25640 | G | A | 0.4564 | -0.167 | 0.008 | 5.012E-104 | 435.77 |
| *BRDT* | rs11974227 | C | T | 0.0627 | 0.115 | 0.016 | 5.0119E-13 | 51.66 |
| *BRDT* | rs6993770 | A | T | 0.2865 | -0.077 | 0.008 | 5.0119E-20 | 92.64 |
| *BRDT* | rs59602812 | A | AT | 0.3182 | 0.11 | 0.008 | 1E-40 | 189.06 |
| *BRDT* | rs10822145 | C | T | 0.4731 | 0.056 | 0.008 | 3.9811E-13 | 49.00 |
| *BRK1* | rs1354034 | T | C | 0.6035 | 0.131 | 0.008 | 1.5849E-65 | 268.14 |
| *BRK1* | rs6054 | C | T | 0.0059 | 0.398 | 0.051 | 3.1623E-15 | 60.90 |
| *BRK1* | rs342293 | C | G | 0.4592 | -0.075 | 0.008 | 1.5849E-23 | 87.89 |
| *BRK1* | rs7896518 | A | G | 0.4244 | 0.071 | 0.008 | 3.1623E-20 | 78.77 |
| *BRK1* | rs11605246 | C | G | 0.2293 | 0.095 | 0.009 | 1.2589E-25 | 111.42 |
| *BRK1* | rs707236 | T | A | 0.346 | -0.148 | 0.008 | 1E-77 | 342.25 |
| *BRK1* | rs9897552 | G | A | 0.177 | 0.072 | 0.01 | 1.9953E-13 | 51.84 |
| *BRK1* | rs1065853 | G | T | 0.081 | -0.251 | 0.014 | 1.5849E-72 | 321.43 |
| *BRSK2* | rs7395567 | T | C | 0.7351 | -0.606 | 0.009 | 1E-200 | 4533.78 |
| *BSG* | rs61747728 | C | T | 0.038 | 0.184 | 0.019 | 1.5849E-22 | 93.78 |
| *BSG* | rs78444298 | G | A | 0.0194 | 0.208 | 0.026 | 5.0119E-15 | 64.00 |
| *BSG* | rs1260326 | T | C | 0.6085 | 0.059 | 0.007 | 3.1623E-15 | 71.04 |
| *BSG* | rs700753 | C | G | 0.6535 | 0.06 | 0.008 | 1.9953E-15 | 56.25 |
| *BSG* | rs2519093 | C | T | 0.1844 | -0.144 | 0.009 | 7.9433E-53 | 256.00 |
| *BSG* | rs77924615 | G | A | 0.1967 | -0.072 | 0.009 | 6.3096E-15 | 64.00 |
| *BSG* | rs200489612 | G | A | 0.0052 | 0.417 | 0.054 | 1.9953E-14 | 59.63 |
| *BSG* | rs56101188 | T | C | 0.0905 | 0.241 | 0.013 | 6.3096E-79 | 343.67 |
| *BST1* | rs2302465 | G | A | 0.1302 | -1.492 | 0.011 | 1E-200 | 18397.22 |
| *BST1* | rs199922514 | A | AAGAAGAA | 0.9076 | -0.058 | 0.008 | 2.5119E-12 | 52.56 |
| *BST1* | rs7952602 | G | C | 0.1349 | -0.103 | 0.007 | 3.9811E-49 | 216.51 |
| *BST1* | rs186021206 | G | A | 0.0064 | 0.529 | 0.033 | 5.0119E-59 | 256.97 |
| *BST1* | rs4760 | A | G | 0.1561 | -0.214 | 0.007 | 1E-200 | 934.61 |
| *BST2* | rs1063412 | G | A | 0.5592 | -0.052 | 0.007 | 1.9953E-12 | 55.18 |
| *BST2* | rs2111485 | A | G | 0.6104 | 0.061 | 0.007 | 3.1623E-16 | 75.94 |
| *BST2* | rs4835265 | C | A | 0.1564 | 0.073 | 0.01 | 5.0119E-13 | 53.29 |
| *BST2* | rs6926219 | G | A | 0.5453 | -0.057 | 0.007 | 7.9433E-15 | 66.31 |
| *BST2* | rs505922 | T | C | 0.3172 | 0.107 | 0.008 | 6.3096E-42 | 178.89 |
| *BST2* | rs17157266 | T | C | 0.1796 | -0.067 | 0.01 | 2.5119E-12 | 44.89 |
| *BST2* | rs139269536 | C | CTG | 0.2457 | 0.087 | 0.009 | 1E-23 | 93.44 |
| *BST2* | rs12373325 | T | C | 0.7977 | -0.062 | 0.009 | 1.2589E-11 | 47.46 |
| *BST2* | rs35251378 | G | A | 0.2938 | -0.082 | 0.008 | 1.9953E-24 | 105.06 |
| *BST2* | rs8101243 | T | C | 0.9567 | -0.344 | 0.018 | 7.9433E-79 | 365.23 |
| *BST2* | rs5992933 | G | T | 0.243 | -0.06 | 0.009 | 1.9953E-12 | 44.44 |
| *BTC* | rs1354034 | T | C | 0.6038 | 0.053 | 0.007 | 7.9433E-13 | 57.33 |
| *BTC* | rs28549760 | A | C | 0.1323 | -0.598 | 0.011 | 1E-200 | 2955.40 |
| *BTC* | rs342296 | G | A | 0.4566 | -0.05 | 0.007 | 6.3096E-12 | 51.02 |
| *BTC* | rs7044478 | C | G | 0.3334 | -0.057 | 0.008 | 6.3096E-13 | 50.77 |
| *BTC* | rs60822569 | T | TTTCTC | 0.554 | 0.062 | 0.007 | 5.0119E-17 | 78.45 |
| *BTD* | rs1260326 | T | C | 0.6084 | -0.076 | 0.007 | 1.9953E-30 | 117.88 |
| *BTD* | rs13078881 | G | C | 0.0433 | -1.618 | 0.018 | 1E-200 | 8080.01 |
| *BTD* | rs200917013 | CA | C | 0.6978 | 0.05 | 0.007 | 7.9433E-13 | 51.02 |
| *BTD* | rs73015965 | A | G | 0.0065 | 0.394 | 0.044 | 1.5849E-19 | 80.18 |
| *BTD* | rs10761760 | A | C | 0.4292 | -0.055 | 0.007 | 1.9953E-16 | 61.73 |
| *BTD* | rs751229834 | CA | C | 0.3074 | 0.052 | 0.007 | 1.5849E-13 | 55.18 |
| *BTD* | rs10860794 | C | A | 0.2879 | -0.056 | 0.007 | 6.3096E-15 | 64.00 |
| *BTD* | rs148480921 | C | CA | 0.0749 | 0.085 | 0.012 | 1E-11 | 50.17 |
| *BTD* | rs4788460 | C | T | 0.2944 | 0.064 | 0.007 | 1.5849E-19 | 83.59 |

| | | | | | | | | |
|---|---|---|---|---|---|---|---|---|
| *BTD* | rs1801689 | A | C | 0.0301 | 0.328 | 0.019 | 5.0119E-67 | 298.02 |
| *BTD* | rs12019136 | G | A | 0.0412 | 0.129 | 0.016 | 3.9811E-15 | 65.00 |
| *BTN1A1* | rs12211201 | A | G | 0.1058 | 0.3 | 0.012 | 3.162E-130 | 625.00 |
| *BTN2A1* | rs61747728 | C | T | 0.0379 | 0.192 | 0.019 | 5.0119E-25 | 102.12 |
| *BTN2A1* | rs143048713 | G | GGT | 0.1003 | 0.083 | 0.012 | 3.9811E-12 | 47.84 |
| *BTN2A1* | rs72843784 | G | T | 0.1105 | 0.472 | 0.012 | 1E-200 | 1547.11 |
| *BTN2A1* | rs1151851 | A | T | 0.4141 | 0.053 | 0.007 | 1.9953E-13 | 57.33 |
| *BTN2A1* | rs77924615 | G | A | 0.1967 | -0.076 | 0.009 | 3.9811E-17 | 71.31 |
| *BTN2A1* | rs186021206 | G | A | 0.0064 | 0.527 | 0.049 | 3.9811E-27 | 115.67 |
| *BTN2A1* | rs2659005 | C | T | 0.4407 | 0.065 | 0.007 | 1.9953E-19 | 86.22 |
| *BTN3A2* | rs9393710 | A | G | 0.1336 | -0.956 | 0.011 | 1E-200 | 7553.19 |
| *BTN3A2* | rs3184504 | T | C | 0.5174 | -0.078 | 0.007 | 2.5119E-33 | 124.16 |
| *BTN3A2* | rs74439742 | C | T | 0.172 | -0.094 | 0.009 | 3.9811E-27 | 109.09 |
| *C19orf12* | rs142552223 | G | A | 0.0376 | -0.136 | 0.02 | 1.2589E-11 | 46.24 |
| *C19orf12* | rs705379 | G | A | 0.4766 | 0.087 | 0.008 | 3.9811E-30 | 118.27 |
| *C19orf12* | rs16919942 | C | T | 0.1064 | -0.085 | 0.012 | 5.0119E-12 | 50.17 |
| *C19orf12* | rs28929474 | C | T | 0.0207 | 0.195 | 0.027 | 1.9953E-13 | 52.16 |
| *C19orf12* | rs146170087 | T | C | 0.001 | -1.001 | 0.122 | 2.5119E-16 | 67.32 |
| *C19orf12* | rs738409 | C | G | 0.2169 | 0.097 | 0.009 | 3.9811E-26 | 116.16 |
| *C1QA* | rs12058824 | G | A | 0.0826 | -0.631 | 0.013 | 1E-200 | 2355.98 |
| *C1QA* | rs188468174 | C | T | 0.0143 | 0.215 | 0.032 | 1E-11 | 45.14 |
| *C1QA* | rs2765501 | G | A | 0.4013 | 0.066 | 0.008 | 2.5119E-18 | 68.06 |
| *C1QA* | rs6532770 | C | A | 0.2438 | 0.151 | 0.008 | 6.3096E-73 | 356.27 |
| *C1QA* | rs4868218 | C | A | 0.7524 | -0.078 | 0.008 | 2.5119E-21 | 95.06 |
| *C1QA* | rs2734975 | C | A | 0.4602 | -0.062 | 0.007 | 3.1623E-18 | 78.45 |
| *C1QA* | rs1727 | C | A | 0.7245 | -0.111 | 0.008 | 1.5849E-44 | 192.52 |
| *C1QA* | rs573898146 | C | CT | 0.3732 | -0.052 | 0.007 | 3.9811E-12 | 55.18 |
| *C1QA* | rs4766578 | T | A | 0.5045 | -0.058 | 0.007 | 1.5849E-16 | 68.65 |
| *C1QA* | rs28372834 | T | TA | 0.164 | -0.066 | 0.01 | 6.3096E-12 | 43.56 |
| *C1QA* | rs2665399 | A | C | 0.8414 | -0.09 | 0.01 | 1E-20 | 81.00 |
| *C1QA* | rs117981338 | C | T | 0.0143 | 0.387 | 0.03 | 5.0119E-37 | 166.41 |
| *C1QA* | rs55841020 | T | A | 0.1739 | -0.074 | 0.009 | 1.9953E-15 | 67.60 |
| *C1QA* | rs28659908 | T | C | 0.4049 | 0.064 | 0.007 | 1.5849E-18 | 83.59 |
| *C1QBP* | rs74839303 | C | T | 0.1172 | -0.121 | 0.013 | 1.2589E-19 | 86.63 |
| *C1QL2* | rs4844573 | T | C | 0.3487 | -0.26 | 0.007 | 1E-200 | 1379.59 |
| *C1QL2* | rs34475538 | G | A | 0.2025 | -0.279 | 0.009 | 1E-200 | 961.00 |
| *C1QL2* | rs112125244 | T | TTTTG | 0.285 | 0.064 | 0.008 | 6.3096E-16 | 64.00 |
| *C1QL2* | rs876036 | T | C | 0.3187 | -0.052 | 0.008 | 5.0119E-12 | 42.25 |
| *C1QL2* | rs530255848 | C | CTT | 0.1693 | 0.071 | 0.01 | 1.5849E-13 | 50.41 |
| *C1QL2* | rs7955585 | C | T | 0.6254 | 0.085 | 0.007 | 5.0119E-31 | 147.45 |
| *C1QL2* | rs1420299 | T | C | 0.6552 | -0.096 | 0.007 | 1.5849E-37 | 188.08 |
| *C1QL2* | rs11149803 | C | G | 0.3146 | 0.15 | 0.008 | 3.9811E-86 | 351.56 |
| *C1QL2* | rs2957150 | A | G | 0.6438 | -0.062 | 0.007 | 3.9811E-17 | 78.45 |
| *C1QL2* | rs571497 | G | A | 0.1529 | -0.092 | 0.01 | 1E-20 | 84.64 |
| *C1QTNF1* | rs2498323 | G | A | 0.1006 | 0.134 | 0.012 | 7.9433E-28 | 124.69 |
| *C1QTNF1* | rs12359178 | G | A | 0.1445 | 0.104 | 0.011 | 5.0119E-23 | 89.39 |
| *C1QTNF1* | rs116539064 | C | T | 0.0973 | 0.398 | 0.013 | 1E-200 | 937.30 |
| *C1QTNF1* | rs2838952 | C | T | 0.0347 | -0.252 | 0.02 | 1.5849E-35 | 158.76 |
| *C1QTNF5* | rs12756819 | T | C | 0.0231 | -0.178 | 0.024 | 2.5119E-13 | 55.01 |
| *C1QTNF5* | rs35677470 | G | A | 0.0827 | -0.098 | 0.013 | 1.5849E-13 | 56.83 |
| *C1QTNF5* | rs10513546 | G | A | 0.2967 | 0.063 | 0.008 | 3.1623E-15 | 62.02 |
| *C1QTNF5* | rs674980 | C | T | 0.5338 | 0.062 | 0.007 | 1.9953E-17 | 78.45 |
| *C1QTNF5* | rs7720230 | T | G | 0.4635 | -0.051 | 0.007 | 2.5119E-12 | 53.08 |
| *C1QTNF5* | rs2856694 | T | C | 0.1859 | -0.076 | 0.01 | 1.9953E-14 | 57.76 |
| *C1QTNF5* | rs41341748 | G | A | 0.011 | 1.759 | 0.037 | 1E-200 | 2260.10 |
| *C1QTNF5* | rs35184771 | G | T | 0.3515 | -0.06 | 0.008 | 1E-14 | 56.25 |
| *C1QTNF5* | rs7232 | T | A | 0.3731 | -0.063 | 0.008 | 1E-16 | 62.02 |
| *C1QTNF5* | rs2509656 | G | A | 0.1434 | 0.163 | 0.01 | 3.9811E-55 | 265.69 |
| *C1QTNF6* | rs535685845 | A | AGGAGGGA | 0.0608 | -0.147 | 0.017 | 2.5119E-18 | 74.77 |
| *C1QTNF9* | rs72550870 | T | C | 0.0303 | -0.124 | 0.017 | 6.3096E-14 | 53.20 |
| *C1QTNF9* | rs10510410 | A | C | 0.2428 | -0.045 | 0.007 | 1E-11 | 41.33 |
| *C1QTNF9* | rs142021145 | C | A | 0.0035 | -0.356 | 0.048 | 1.5849E-13 | 55.01 |
| *C1QTNF9* | rs27289 | G | T | 0.553 | -0.061 | 0.006 | 2.5119E-26 | 103.36 |
| *C1QTNF9* | rs2524096 | G | T | 0.4196 | -0.07 | 0.006 | 1.5849E-33 | 136.11 |
| *C1QTNF9* | rs587611953 | C | A | 0.1619 | 0.116 | 0.009 | 6.3096E-42 | 166.12 |
| *C1QTNF9* | rs11259539 | T | C | 0.4087 | 0.067 | 0.006 | 7.9433E-31 | 124.69 |
| *C1QTNF9* | rs1961993 | C | A | 0.8027 | 1.151 | 0.01 | 1E-200 | 13248.01 |
| *C1QTNF9* | rs13333582 | T | C | 0.0427 | 0.097 | 0.014 | 5.0119E-12 | 48.01 |

| | | | | | | | | |
|---|---|---|---|---|---|---|---|---|
| *C1QTNF9* | rs12975366 | T | C | 0.3952 | -0.042 | 0.006 | 3.1623E-13 | 49.00 |
| *C1R* | rs12722051 | A | T | 0.2039 | 0.064 | 0.009 | 3.9811E-12 | 50.57 |
| *C1R* | rs10849546 | G | A | 0.1353 | -0.459 | 0.011 | 1E-200 | 1741.17 |
| *C1R* | rs12433460 | C | A | 0.2686 | -0.191 | 0.009 | 1.2589E-94 | 450.38 |
| *C1RL* | rs35447916 | C | CA | 0.4696 | -0.065 | 0.007 | 3.9811E-20 | 86.22 |
| *C1RL* | rs28362944 | T | C | 0.049 | 0.124 | 0.016 | 2.5119E-14 | 60.06 |
| *C1RL* | rs174566 | A | G | 0.3502 | 0.071 | 0.007 | 3.9811E-22 | 102.88 |
| *C1RL* | rs191448232 | G | C | 0.0122 | -2.152 | 0.034 | 1E-200 | 4006.15 |
| *C1RL* | rs149394327 | G | C | 0.03 | 0.35 | 0.021 | 1E-63 | 277.78 |
| *C1S* | rs73480556 | G | C | 0.2712 | -0.08 | 0.008 | 5.0119E-23 | 100.00 |
| *C1S* | rs12146727 | G | A | 0.135 | -0.668 | 0.011 | 1E-200 | 3687.80 |
| *C1S* | rs12433460 | C | A | 0.2686 | -0.199 | 0.009 | 5.012E-108 | 488.90 |
| *C2* | rs1260326 | T | C | 0.6081 | -0.069 | 0.007 | 1.5849E-21 | 97.16 |
| *C2* | rs6984305 | A | T | 0.8851 | 0.075 | 0.011 | 1.5849E-11 | 46.49 |
| *C2* | rs56278466 | T | G | 0.6611 | 0.104 | 0.007 | 5.0119E-44 | 220.73 |
| *C2CD2L* | rs142201367 | T | TTTTCTAAC | 0.4949 | 0.059 | 0.008 | 7.9433E-14 | 54.39 |
| *C2CD2L* | rs640603 | G | A | 0.3517 | -0.056 | 0.008 | 5.0119E-12 | 49.00 |
| *C2orf69* | rs17629007 | T | C | 0.0322 | 0.253 | 0.022 | 3.1623E-31 | 132.25 |
| *C2orf69* | rs1354034 | T | C | 0.604 | 0.063 | 0.008 | 1E-15 | 62.02 |
| *C3* | rs3753396 | A | G | 0.1717 | -0.128 | 0.01 | 3.1623E-36 | 163.84 |
| *C3* | rs1260326 | T | C | 0.6084 | -0.065 | 0.008 | 1.5849E-16 | 66.02 |
| *C3* | rs556679 | C | T | 0.1212 | 0.107 | 0.012 | 1.2589E-19 | 79.51 |
| *C3* | rs11569466 | G | T | 0.1365 | -0.167 | 0.011 | 7.9433E-50 | 230.49 |
| *C3* | rs12975366 | T | C | 0.396 | 0.072 | 0.008 | 7.9433E-20 | 81.00 |
| *C4BPB* | rs140584594 | A | G | 0.7319 | 0.06 | 0.008 | 6.3096E-14 | 56.25 |
| *C4BPB* | rs2316653 | A | G | 0.4992 | -0.06 | 0.007 | 3.1623E-17 | 73.47 |
| *C4BPB* | rs9822589 | G | A | 0.468 | 0.06 | 0.007 | 2.5119E-17 | 73.47 |
| *C4BPB* | rs687339 | C | T | 0.7711 | -0.06 | 0.008 | 1.5849E-12 | 56.25 |
| *C4BPB* | rs28507491 | G | A | 0.377 | -0.055 | 0.007 | 5.0119E-14 | 61.73 |
| *C4BPB* | rs757795770 | CTTTA | C | 0.7144 | 0.068 | 0.008 | 3.9811E-18 | 72.25 |
| *C4BPB* | rs601020 | G | A | 0.2448 | -0.064 | 0.008 | 3.1623E-14 | 64.00 |
| *C4BPB* | rs2068888 | G | A | 0.4501 | -0.049 | 0.007 | 6.3096E-12 | 49.00 |
| *C4BPB* | rs139130389 | C | CTA | 0.0726 | -0.153 | 0.014 | 7.9433E-30 | 119.43 |
| *C4BPB* | rs11621792 | C | T | 0.4525 | -0.049 | 0.007 | 3.9811E-12 | 49.00 |
| *C4BPB* | rs28929474 | C | T | 0.0206 | 0.364 | 0.025 | 1E-48 | 211.99 |
| *C4BPB* | rs9302635 | T | C | 0.182 | -0.137 | 0.009 | 2.5119E-50 | 231.72 |
| *C4BPB* | rs8178824 | C | T | 0.03 | 0.815 | 0.021 | 1E-200 | 1506.18 |
| *C5* | rs780094 | T | C | 0.6194 | -0.062 | 0.008 | 7.9433E-16 | 60.06 |
| *C5* | rs4837810 | T | C | 0.5772 | -0.143 | 0.008 | 6.3096E-78 | 319.52 |
| *C7* | rs74480769 | A | G | 0.0289 | -1.498 | 0.023 | 1E-200 | 4241.97 |
| *C7* | rs56278466 | T | G | 0.6607 | 0.077 | 0.007 | 6.3096E-28 | 121.00 |
| *C7* | rs738409 | C | G | 0.2167 | 0.069 | 0.008 | 1.5849E-17 | 74.39 |
| *C7orf50* | rs200169409 | GC | G | 0.0284 | -0.948 | 0.023 | 1E-200 | 1698.87 |
| *C8B* | rs626457 | C | T | 0.3297 | 0.519 | 0.008 | 1E-200 | 4208.77 |
| *C8B* | rs1260326 | T | C | 0.6084 | -0.055 | 0.007 | 1E-15 | 61.73 |
| *C9* | rs6734238 | A | G | 0.4031 | 0.053 | 0.008 | 3.9811E-12 | 43.89 |
| *C9* | rs34000044 | G | T | 0.0015 | -1.405 | 0.098 | 7.9433E-47 | 205.54 |
| *C9* | rs34954997 | C | CTTCG | 0.2376 | -0.064 | 0.009 | 5.0119E-13 | 50.57 |
| *C9orf40* | rs11759553 | A | T | 0.2723 | -0.066 | 0.008 | 1.5849E-15 | 68.06 |
| *C9orf40* | rs1879154 | T | C | 0.3949 | -0.06 | 0.007 | 1E-15 | 73.47 |
| *C9orf40* | rs75372982 | A | G | 0.2877 | -0.065 | 0.008 | 1.9953E-15 | 66.02 |
| *CA1* | rs3811444 | C | T | 0.3324 | 0.08 | 0.008 | 7.9433E-25 | 100.00 |
| *CA1* | rs11759553 | A | T | 0.2722 | -0.075 | 0.008 | 6.3096E-20 | 87.89 |
| *CA1* | rs12544332 | C | A | 0.589 | -0.089 | 0.007 | 6.3096E-33 | 161.65 |
| *CA1* | rs67544533 | A | G | 0.1834 | -0.132 | 0.009 | 3.1623E-44 | 215.11 |
| *CA11* | rs646776 | C | T | 0.7783 | 0.186 | 0.009 | 1.5849E-89 | 427.11 |
| *CA11* | rs4747198 | C | G | 0.2855 | -0.059 | 0.009 | 3.1623E-12 | 42.98 |
| *CA12* | rs61747728 | C | T | 0.038 | 0.169 | 0.019 | 7.9433E-19 | 79.12 |
| *CA12* | rs1260326 | T | C | 0.6081 | 0.075 | 0.008 | 3.1623E-23 | 87.89 |
| *CA12* | rs868399694 | TA | T | 0.4469 | 0.054 | 0.007 | 3.9811E-13 | 59.51 |
| *CA12* | rs13107325 | C | T | 0.0748 | 0.134 | 0.014 | 1.2589E-21 | 91.61 |
| *CA12* | rs12341245 | G | A | 0.3976 | 0.056 | 0.007 | 6.3096E-14 | 64.00 |
| *CA12* | rs12902855 | G | A | 0.483 | 0.062 | 0.008 | 1.9953E-16 | 60.06 |
| *CA12* | rs12916532 | T | C | 0.2741 | -0.294 | 0.008 | 1E-200 | 1350.56 |
| *CA12* | rs186021206 | G | A | 0.0063 | 0.639 | 0.051 | 3.1623E-36 | 156.99 |
| *CA12* | rs796389476 | TA | T | 0.1 | 0.125 | 0.012 | 3.9811E-24 | 108.51 |
| *CA12* | rs708686 | C | T | 0.2673 | 0.112 | 0.008 | 7.9433E-42 | 196.00 |
| *CA13* | rs1354034 | T | C | 0.6036 | 0.053 | 0.008 | 1.2589E-12 | 43.89 |

| | | | | | | | | |
|---|---|---|---|---|---|---|---|---|
| *CA13* | rs56072918 | C | T | 0.0683 | -0.526 | 0.015 | 1E-200 | 1229.67 |
| *CA14* | rs1122967 | C | G | 0.5103 | 0.174 | 0.007 | 2.512E-129 | 617.88 |
| *CA14* | rs61431557 | C | T | 0.1371 | -0.071 | 0.01 | 3.9811E-12 | 50.41 |
| *CA14* | rs2954021 | A | G | 0.504 | 0.075 | 0.007 | 2.5119E-26 | 114.80 |
| *CA14* | rs7954584 | G | A | 0.5543 | 0.058 | 0.007 | 1.9953E-16 | 68.65 |
| *CA2* | rs2980370 | A | G | 0.5207 | 0.102 | 0.007 | 6.3096E-45 | 212.33 |
| *CA3* | rs3811444 | C | T | 0.3323 | 0.112 | 0.008 | 6.3096E-50 | 196.00 |
| *CA3* | rs1047891 | C | A | 0.3131 | 0.086 | 0.008 | 5.0119E-29 | 115.56 |
| *CA3* | rs11759553 | A | T | 0.2725 | -0.114 | 0.008 | 1E-45 | 203.06 |
| *CA3* | rs2072696 | G | C | 0.2431 | -0.174 | 0.008 | 7.9433E-97 | 473.06 |
| *CA3* | rs1867146 | C | G | 0.1823 | -0.089 | 0.009 | 3.1623E-22 | 97.79 |
| *CA3* | rs1799918 | G | C | 0.3788 | -0.067 | 0.007 | 1.2589E-19 | 91.61 |
| *CA4* | rs184775562 | T | C | 0.0017 | 0.818 | 0.092 | 7.9433E-19 | 79.06 |
| *CA4* | rs78058190 | G | A | 0.0509 | -0.189 | 0.019 | 3.1623E-24 | 98.95 |
| *CA4* | rs115007843 | A | C | 0.1763 | -0.072 | 0.01 | 7.9433E-14 | 51.84 |
| *CA4* | rs9270588 | C | T | 0.5309 | 0.059 | 0.007 | 1.2589E-15 | 71.04 |
| *CA4* | rs582118 | A | G | 0.318 | 0.164 | 0.008 | 3.1623E-97 | 420.25 |
| *CA4* | rs61729512 | G | A | 0.1177 | 0.085 | 0.011 | 7.9433E-14 | 59.71 |
| *CA4* | rs117154637 | A | T | 0.0349 | -0.825 | 0.02 | 1E-200 | 1701.56 |
| *CA4* | rs10642898 | C | CCCG | 0.403 | 0.055 | 0.008 | 7.9433E-13 | 47.27 |
| *CA4* | rs4760 | A | G | 0.156 | -0.17 | 0.01 | 1.2589E-64 | 289.00 |
| *CA4* | rs12975366 | T | C | 0.3952 | -0.073 | 0.007 | 1.2589E-22 | 108.76 |
| *CA5A* | rs4835265 | C | A | 0.1559 | 0.071 | 0.009 | 2.5119E-15 | 62.23 |
| *CA5A* | rs4979373 | C | T | 0.4906 | -0.048 | 0.007 | 3.1623E-13 | 47.02 |
| *CA5A* | rs10883451 | T | C | 0.5019 | -0.045 | 0.006 | 5.0119E-12 | 56.25 |
| *CA5A* | rs55870502 | T | C | 0.1926 | 0.812 | 0.009 | 1E-200 | 8140.05 |
| *CA5A* | rs738409 | C | G | 0.2171 | 0.054 | 0.008 | 7.9433E-12 | 45.56 |
| *CA6* | rs3765963 | A | G | 0.4117 | 0.683 | 0.007 | 1E-200 | 9520.18 |
| *CA6* | rs946527 | T | C | 0.7078 | -0.056 | 0.008 | 5.0119E-13 | 49.00 |
| *CA6* | rs351388 | C | G | 0.592 | -0.051 | 0.007 | 1.9953E-12 | 53.08 |
| *CA6* | rs891499 | G | C | 0.3275 | 0.065 | 0.007 | 1E-23 | 86.22 |
| *CA6* | rs557296878 | CT | C | 0.5919 | 0.067 | 0.006 | 1.9953E-26 | 124.69 |
| *CA6* | rs60800828 | G | GT | 0.1938 | -0.065 | 0.008 | 1.9953E-15 | 66.02 |
| *CA6* | rs836143 | G | C | 0.6719 | -0.054 | 0.007 | 2.5119E-16 | 59.51 |
| *CA6* | rs4256938 | G | A | 0.0949 | -0.123 | 0.01 | 5.0119E-32 | 151.29 |
| *CA6* | rs290371 | G | A | 0.3555 | -0.071 | 0.006 | 3.9811E-28 | 140.03 |
| *CA6* | rs173539 | C | T | 0.3318 | 0.057 | 0.007 | 1.5849E-18 | 66.31 |
| *CA6* | rs1523135 | C | G | 0.7159 | 0.055 | 0.007 | 1E-15 | 61.73 |
| *CA6* | rs7250028 | T | G | 0.2371 | 0.058 | 0.007 | 1E-15 | 68.65 |
| *CA6* | rs601338 | G | A | 0.5098 | 0.063 | 0.006 | 1.9953E-24 | 110.25 |
| *CA9* | rs111709193 | CT | C | 0.1897 | 0.075 | 0.01 | 1.9953E-13 | 56.25 |
| *CA9* | rs183985235 | C | A | 0.0024 | -1.479 | 0.085 | 2.5119E-67 | 302.76 |
| *CA9* | rs56278466 | T | G | 0.6612 | 0.142 | 0.008 | 2.5119E-74 | 315.06 |
| *CA9* | rs616114 | C | T | 0.3896 | 0.082 | 0.008 | 1.5849E-27 | 105.06 |
| *CA9* | rs2608894 | T | C | 0.7648 | -0.063 | 0.009 | 6.3096E-13 | 49.00 |
| *CA9* | rs492602 | A | G | 0.5092 | -0.052 | 0.007 | 3.1623E-12 | 55.18 |
| *CABP2* | rs607929 | C | G | 0.4485 | -0.059 | 0.008 | 1.9953E-14 | 54.39 |
| *CABP2* | rs79034686 | C | T | 0.2523 | 0.07 | 0.009 | 3.1623E-15 | 60.49 |
| *CABP2* | rs62129286 | C | T | 0.2312 | 0.161 | 0.009 | 1.9953E-69 | 320.01 |
| *CACNB1* | rs58534292 | T | C | 0.6504 | -0.091 | 0.008 | 7.9433E-29 | 129.39 |
| *CACNB1* | rs57523015 | C | T | 0.6618 | -0.094 | 0.008 | 1.5849E-29 | 138.06 |
| *CACNB3* | rs1354034 | T | C | 0.6043 | 0.096 | 0.008 | 7.9433E-36 | 144.00 |
| *CACNB3* | rs11502185 | T | C | 0.2598 | 0.072 | 0.01 | 7.9433E-14 | 51.84 |
| *CACNB3* | rs7975385 | T | C | 0.2333 | 0.119 | 0.009 | 3.9811E-41 | 174.83 |
| *CACYBP* | rs1354034 | T | C | 0.6044 | 0.097 | 0.008 | 1.5849E-36 | 147.02 |
| *CACYBP* | rs342298 | C | T | 0.4547 | -0.057 | 0.008 | 6.3096E-14 | 50.77 |
| *CACYBP* | rs73396521 | G | A | 0.2451 | 0.06 | 0.009 | 1.2589E-11 | 44.44 |
| *CALB1* | rs143241372 | A | G | 0.0093 | 0.655 | 0.04 | 5.0119E-60 | 268.14 |
| *CALB1* | rs9529913 | C | T | 0.603 | -0.085 | 0.008 | 3.9811E-29 | 112.89 |
| *CALB1* | rs6127099 | A | T | 0.2776 | -0.063 | 0.008 | 1E-13 | 62.02 |
| *CALB2* | rs1476089 | G | A | 0.2877 | -0.069 | 0.008 | 1.5849E-16 | 74.39 |
| *CALCA* | rs3748916 | G | A | 0.4719 | 0.067 | 0.006 | 7.9433E-28 | 124.69 |
| *CALCA* | rs73995235 | G | T | 0.0601 | -0.112 | 0.013 | 1E-17 | 74.22 |
| *CALCA* | rs7008914 | T | C | 0.2552 | -0.054 | 0.007 | 2.5119E-14 | 59.51 |
| *CALCA* | rs782744846 | TTCTAAG | T | 0.4526 | -0.102 | 0.006 | 3.1623E-61 | 289.00 |
| *CALCA* | rs10841569 | A | G | 0.1454 | 0.088 | 0.009 | 3.9811E-24 | 95.60 |
| *CALCA* | rs138903858 | G | T | 0.0279 | -0.14 | 0.019 | 1.9953E-13 | 54.29 |
| *CALCA* | rs3790163 | A | G | 0.7928 | 0.069 | 0.008 | 7.9433E-19 | 74.39 |

| | | | | | | | | |
|---|---|---|---|---|---|---|---|---|
| *CALCB* | rs4757282 | G | T | 0.7599 | -0.127 | 0.009 | 1E-46 | 199.12 |
| *CALCOCO1* | rs1354034 | T | C | 0.6041 | 0.062 | 0.007 | 3.9811E-17 | 78.45 |
| *CALCOCO1* | rs342293 | C | G | 0.4594 | -0.054 | 0.007 | 7.9433E-14 | 59.51 |
| *CALCOCO1* | rs6993770 | A | T | 0.2863 | -0.072 | 0.008 | 5.0119E-19 | 81.00 |
| *CALCOCO1* | rs17102261 | T | G | 0.1502 | 0.139 | 0.01 | 3.1623E-41 | 193.21 |
| *CALCOCO2* | rs1354034 | T | C | 0.6045 | 0.195 | 0.008 | 6.31E-143 | 594.14 |
| *CALCOCO2* | rs3792382 | T | C | 0.5933 | -0.055 | 0.008 | 1E-12 | 47.27 |
| *CALCOCO2* | rs11242109 | G | T | 0.4812 | -0.057 | 0.007 | 3.9811E-14 | 66.31 |
| *CALCOCO2* | rs342298 | C | T | 0.4548 | -0.083 | 0.008 | 3.1623E-28 | 107.64 |
| *CALCOCO2* | rs11605246 | C | G | 0.2289 | 0.088 | 0.009 | 7.9433E-23 | 95.60 |
| *CALCOCO2* | rs200309755 | C | T | 0.537 | 0.054 | 0.008 | 2.5119E-12 | 45.56 |
| *CALCOCO2* | rs606911 | G | A | 0.1588 | 0.075 | 0.01 | 1.2589E-13 | 56.25 |
| *CAMKK1* | rs342296 | G | A | 0.4566 | -0.063 | 0.008 | 1.5849E-16 | 62.02 |
| *CAMKK1* | rs4759076 | T | C | 0.4569 | 0.076 | 0.008 | 3.1623E-23 | 90.25 |
| *CAMKK1* | rs753513 | C | T | 0.2867 | -0.297 | 0.008 | 1E-200 | 1378.27 |
| *CAMSAP1* | rs1354034 | T | C | 0.6043 | 0.077 | 0.007 | 6.3096E-25 | 121.00 |
| *CANT1* | rs61747728 | C | T | 0.038 | 0.142 | 0.019 | 3.1623E-13 | 55.86 |
| *CANT1* | rs1863622 | T | C | 0.2224 | -0.079 | 0.009 | 2.5119E-18 | 77.05 |
| *CANT1* | rs56278466 | T | G | 0.6615 | 0.088 | 0.008 | 7.9433E-29 | 121.00 |
| *CANT1* | rs423135 | G | A | 0.5177 | 0.059 | 0.008 | 2.5119E-15 | 54.39 |
| *CANT1* | rs17739056 | T | C | 0.1156 | -0.215 | 0.012 | 3.1623E-75 | 321.01 |
| *CANT1* | rs1654425 | T | C | 0.834 | 0.104 | 0.01 | 1E-24 | 108.16 |
| *CAPG* | rs6886 | T | C | 0.6674 | -0.872 | 0.008 | 1E-200 | 11881.00 |
| *CAPG* | rs9268925 | A | C | 0.8546 | -0.08 | 0.008 | 1.2589E-22 | 100.00 |
| *CAPG* | rs4632248 | G | T | 0.2096 | 0.186 | 0.007 | 6.31E-151 | 706.04 |
| *CAPN3* | rs710446 | T | C | 0.4088 | 0.074 | 0.008 | 1.2589E-21 | 85.56 |
| *CAPS* | rs113534515 | G | C | 0.2912 | 0.273 | 0.008 | 1E-200 | 1164.52 |
| *CAPS* | rs147363394 | C | A | 0.0014 | -2.202 | 0.096 | 1.995E-116 | 526.13 |
| *CARHSP1* | rs12597479 | G | T | 0.5259 | -0.078 | 0.007 | 1E-25 | 124.16 |
| *CASC3* | rs1354034 | T | C | 0.604 | 0.109 | 0.008 | 1.9953E-44 | 185.64 |
| *CASC3* | rs11502185 | T | C | 0.26 | 0.071 | 0.01 | 5.0119E-13 | 50.41 |
| *CASP1* | rs141551019 | A | G | 0.2259 | -0.066 | 0.009 | 3.9811E-13 | 53.78 |
| *CASP1* | rs34436714 | C | A | 0.209 | 0.325 | 0.009 | 1E-200 | 1304.01 |
| *CASP10* | rs7576306 | A | G | 0.4944 | 0.182 | 0.008 | 3.162E-126 | 517.56 |
| *CASP10* | rs6993770 | A | T | 0.2869 | -0.093 | 0.008 | 3.9811E-29 | 135.14 |
| *CASP2* | rs1354034 | T | C | 0.6043 | 0.097 | 0.008 | 3.9811E-37 | 147.02 |
| *CASP2* | rs139141690 | G | A | 0.0048 | 0.382 | 0.055 | 5.0119E-12 | 48.24 |
| *CASP2* | rs342299 | C | T | 0.4547 | -0.051 | 0.008 | 1.2589E-11 | 40.64 |
| *CASP2* | rs11502185 | T | C | 0.2598 | 0.068 | 0.01 | 1.5849E-12 | 46.24 |
| *CASP3* | rs62339863 | A | C | 0.2607 | -0.128 | 0.008 | 2.5119E-51 | 256.00 |
| *CASP3* | rs6993770 | A | T | 0.2865 | -0.078 | 0.008 | 1E-21 | 95.06 |
| *CASP4* | rs117001788 | C | T | 0.0319 | 0.348 | 0.023 | 3.1623E-51 | 228.93 |
| *CASP7* | rs1354034 | T | C | 0.6043 | 0.074 | 0.008 | 3.9811E-22 | 85.56 |
| *CASP7* | rs6993770 | A | T | 0.2865 | -0.057 | 0.008 | 6.3096E-12 | 50.77 |
| *CASP7* | rs141266925 | T | C | 0.0029 | -1.151 | 0.07 | 1.2589E-61 | 270.37 |
| *CASP7* | rs60822569 | T | TTTCTC | 0.5543 | 0.054 | 0.008 | 7.9433E-13 | 45.56 |
| *CASP8* | rs34841024 | T | G | 0.1363 | 0.31 | 0.011 | 5.012E-183 | 794.21 |
| *CASP8* | rs1354034 | T | C | 0.6042 | 0.065 | 0.007 | 1.5849E-18 | 86.22 |
| *CASP8* | rs3847326 | A | G | 0.4819 | -0.051 | 0.007 | 1.5849E-12 | 53.08 |
| *CASP8* | rs60822569 | T | TTTCTC | 0.5543 | 0.065 | 0.007 | 3.1623E-18 | 86.22 |
| *CASP9* | rs12024833 | A | G | 0.5248 | -0.081 | 0.008 | 2.5119E-26 | 102.52 |
| *CAT* | rs2284368 | T | C | 0.2206 | -0.184 | 0.009 | 1.259E-100 | 417.98 |
| *CBLIF* | rs333947 | G | A | 0.1483 | -0.071 | 0.01 | 6.3096E-13 | 50.41 |
| *CBLIF* | rs2348194 | A | T | 0.381 | 0.05 | 0.007 | 6.3096E-12 | 51.02 |
| *CBLIF* | rs13088531 | T | A | 0.2453 | -0.06 | 0.008 | 2.5119E-13 | 56.25 |
| *CBLIF* | rs1290790 | A | T | 0.4475 | -0.06 | 0.007 | 1.2589E-17 | 73.47 |
| *CBLIF* | rs72704793 | G | A | 0.3299 | 0.065 | 0.007 | 1.9953E-18 | 86.22 |
| *CBLIF* | rs7718236 | G | A | 0.8257 | -0.088 | 0.01 | 1.5849E-20 | 77.44 |
| *CBLIF* | rs1634716 | G | A | 0.1503 | -0.111 | 0.01 | 5.0119E-27 | 123.21 |
| *CBLIF* | rs807309 | C | T | 0.7376 | -0.465 | 0.008 | 1E-200 | 3378.52 |
| *CBLIF* | rs2976397 | G | T | 0.419 | -0.054 | 0.007 | 1.5849E-14 | 59.51 |
| *CBLIF* | rs532436 | G | A | 0.1848 | -0.211 | 0.009 | 1E-118 | 549.64 |
| *CBLIF* | rs56278466 | T | G | 0.6614 | 0.069 | 0.007 | 1E-20 | 97.16 |
| *CBLIF* | rs2472632 | C | A | 0.3599 | -0.062 | 0.007 | 3.1623E-17 | 78.45 |
| *CBLIF* | rs1787909 | G | C | 0.3446 | -0.22 | 0.007 | 1.585E-190 | 987.76 |
| *CBLIF* | rs7929203 | C | T | 0.2812 | 0.054 | 0.008 | 6.3096E-12 | 45.56 |
| *CBLIF* | rs4925105 | T | C | 0.5267 | -0.054 | 0.007 | 1E-14 | 59.51 |
| *CBLIF* | rs61580404 | G | T | 0.1012 | 0.085 | 0.012 | 6.3096E-13 | 50.17 |

| | | | | | | | | |
|---|---|---|---|---|---|---|---|---|
| *CBLIF* | rs584768 | G | A | 0.5332 | 0.052 | 0.007 | 1.2589E-13 | 55.18 |
| *CBLN4* | rs7684253 | C | T | 0.5533 | -0.095 | 0.007 | 7.9433E-38 | 184.18 |
| *CBLN4* | rs7116008 | G | A | 0.4331 | 0.067 | 0.007 | 1.2589E-19 | 91.61 |
| *CBLN4* | rs215226 | A | G | 0.4006 | 0.058 | 0.007 | 1E-14 | 68.65 |
| *CBLN4* | rs186021206 | G | A | 0.0064 | 0.492 | 0.05 | 1.2589E-22 | 96.83 |
| *CBLN4* | rs12624596 | G | A | 0.2863 | 0.188 | 0.008 | 3.162E-113 | 552.25 |
| *CBS* | rs4835265 | C | A | 0.1561 | 0.082 | 0.01 | 3.9811E-15 | 67.24 |
| *CBS* | rs6586281 | G | A | 0.176 | 0.156 | 0.01 | 3.9811E-55 | 243.36 |
| *CBS* | rs3747207 | G | A | 0.2156 | 0.105 | 0.009 | 5.0119E-30 | 136.11 |
| *CC2D1A* | rs61740117 | C | T | 0.0135 | -0.325 | 0.032 | 2.5119E-24 | 103.15 |
| *CCDC134* | rs10733789 | T | C | 0.3129 | 0.061 | 0.008 | 1.5849E-13 | 58.14 |
| *CCDC134* | rs78290111 | T | TG | 0.3573 | -0.06 | 0.008 | 3.9811E-14 | 56.25 |
| *CCDC50* | rs35380043 | A | T | 0.0482 | -0.419 | 0.018 | 1.585E-125 | 541.85 |
| *CCDC50* | rs5859867 | C | CT | 0.2865 | -0.102 | 0.008 | 3.1623E-34 | 162.56 |
| *CCDC50* | rs62445350 | A | T | 0.3081 | -0.071 | 0.008 | 1.9953E-18 | 78.77 |
| *CCDC50* | rs2583285 | G | A | 0.2854 | -0.184 | 0.009 | 3.981E-103 | 417.98 |
| *CCDC80* | rs6772337 | A | G | 0.6239 | -0.07 | 0.007 | 1E-22 | 100.00 |
| *CCDC80* | rs13107325 | C | T | 0.0751 | 0.138 | 0.013 | 5.0119E-26 | 112.69 |
| *CCDC80* | rs41265930 | T | C | 0.0739 | -0.146 | 0.013 | 7.9433E-29 | 126.13 |
| *CCDC80* | rs2142306 | T | C | 0.4122 | 0.131 | 0.007 | 2.5119E-78 | 350.22 |
| *CCDC80* | rs186021206 | G | A | 0.0064 | 0.908 | 0.047 | 1.9953E-83 | 373.23 |
| *CCER2* | rs905938 | T | C | 0.267 | 0.097 | 0.008 | 2.5119E-32 | 147.02 |
| *CCER2* | rs57060715 | G | A | 0.2709 | 0.058 | 0.009 | 1E-11 | 41.53 |
| *CCER2* | rs1561477 | T | C | 0.8923 | -0.109 | 0.012 | 1.9953E-20 | 82.51 |
| *CCER2* | rs17400325 | T | C | 0.0411 | 0.16 | 0.018 | 3.1623E-18 | 79.01 |
| *CCER2* | rs9290890 | A | C | 0.5829 | -0.061 | 0.007 | 1.9953E-16 | 75.94 |
| *CCER2* | rs2870749 | A | G | 0.4348 | -0.32 | 0.008 | 1E-200 | 1600.00 |
| *CCER2* | rs13168098 | G | A | 0.3805 | -0.054 | 0.008 | 1.2589E-12 | 45.56 |
| *CCER2* | rs9392840 | C | T | 0.2721 | 0.06 | 0.008 | 6.3096E-13 | 56.25 |
| *CCER2* | rs4837917 | T | C | 0.2928 | -0.096 | 0.008 | 3.9811E-33 | 144.00 |
| *CCER2* | rs7083373 | C | G | 0.4837 | -0.06 | 0.007 | 1.2589E-16 | 73.47 |
| *CCER2* | rs11832143 | A | G | 0.4565 | -0.05 | 0.007 | 7.9433E-12 | 51.02 |
| *CCER2* | rs11148502 | A | T | 0.4883 | 0.1 | 0.007 | 1.5849E-42 | 204.08 |
| *CCER2* | rs58932296 | C | CT | 0.5216 | -0.055 | 0.007 | 6.3096E-14 | 61.73 |
| *CCER2* | rs879627 | C | G | 0.4372 | 0.062 | 0.007 | 5.0119E-17 | 78.45 |
| *CCER2* | rs7236418 | G | C | 0.557 | 0.052 | 0.007 | 1E-12 | 55.18 |
| *CCER2* | rs2596606 | A | G | 0.1817 | 0.095 | 0.01 | 1.9953E-23 | 90.25 |
| *CCER2* | rs190921775 | G | T | 0.005 | -0.917 | 0.053 | 1E-67 | 299.36 |
| *CCER2* | rs5751435 | T | C | 0.8758 | -0.078 | 0.011 | 1.9953E-12 | 50.28 |
| *CCL11* | rs12075 | G | A | 0.5829 | 0.08 | 0.007 | 1.9953E-29 | 130.61 |
| *CCL11* | rs2228467 | T | C | 0.0611 | 0.41 | 0.015 | 6.31E-168 | 747.11 |
| *CCL11* | rs1979671 | C | T | 0.6867 | -0.079 | 0.008 | 1.2589E-24 | 97.52 |
| *CCL11* | rs1354034 | T | C | 0.6041 | -0.052 | 0.007 | 7.9433E-13 | 55.18 |
| *CCL11* | rs2516471 | G | A | 0.0635 | -0.116 | 0.015 | 1.5849E-15 | 59.80 |
| *CCL11* | rs2868169 | C | T | 0.8956 | -0.202 | 0.011 | 7.9433E-70 | 337.22 |
| *CCL11* | rs4602861 | A | G | 0.2739 | -0.056 | 0.008 | 1.9953E-12 | 49.00 |
| *CCL11* | rs7080386 | C | A | 0.412 | 0.066 | 0.007 | 1.9953E-20 | 88.90 |
| *CCL11* | rs5818270 | C | CT | 0.5832 | -0.071 | 0.007 | 1.2589E-22 | 102.88 |
| *CCL11* | rs140905292 | G | A | 0.2241 | -0.121 | 0.009 | 3.1623E-45 | 180.75 |
| *CCL11* | rs1613662 | G | A | 0.8341 | 0.092 | 0.009 | 3.1623E-22 | 104.49 |
| *CCL11* | rs55951234 | C | CCT | 0.5793 | 0.048 | 0.007 | 1.2589E-11 | 47.02 |
| *CCL13* | rs12075 | G | A | 0.583 | 0.223 | 0.007 | 1E-200 | 1014.88 |
| *CCL13* | rs13412535 | G | A | 0.231 | -0.07 | 0.008 | 1.2589E-16 | 76.56 |
| *CCL13* | rs142157305 | C | CTGTT | 0.383 | -0.106 | 0.007 | 1.5849E-46 | 229.31 |
| *CCL13* | rs12495098 | T | G | 0.0693 | 0.105 | 0.014 | 1.2589E-13 | 56.25 |
| *CCL13* | rs2522051 | T | C | 0.4521 | 0.052 | 0.007 | 3.9811E-13 | 55.18 |
| *CCL13* | rs3117110 | C | T | 0.2126 | 0.066 | 0.009 | 6.3096E-14 | 53.78 |
| *CCL13* | rs34231168 | C | CA | 0.2882 | -0.057 | 0.008 | 1.9953E-12 | 50.77 |
| *CCL13* | rs826732 | G | C | 0.4995 | 0.051 | 0.007 | 2.5119E-12 | 53.08 |
| *CCL13* | rs7080386 | C | A | 0.4119 | 0.073 | 0.007 | 1.2589E-23 | 108.76 |
| *CCL13* | rs59001897 | T | A | 0.1748 | 0.073 | 0.01 | 1E-14 | 53.29 |
| *CCL13* | rs12445050 | C | T | 0.1382 | 0.08 | 0.01 | 7.9433E-15 | 64.00 |
| *CCL13* | rs3136674 | T | C | 0.0537 | -0.462 | 0.016 | 1E-178 | 833.77 |
| *CCL13* | rs892090 | T | G | 0.8342 | 0.136 | 0.01 | 5.0119E-45 | 184.96 |
| *CCL14* | rs13962 | G | A | 0.1768 | 0.169 | 0.009 | 2.5119E-72 | 352.60 |
| *CCL14* | rs13412535 | G | A | 0.231 | -0.066 | 0.008 | 1.9953E-15 | 68.06 |
| *CCL14* | rs1354034 | T | C | 0.6038 | -0.065 | 0.007 | 1.2589E-19 | 86.22 |
| *CCL14* | rs6993770 | A | T | 0.2864 | -0.073 | 0.008 | 3.9811E-21 | 83.27 |

| | | | | | | | | |
|---|---|---|---|---|---|---|---|---|
| *CCL14* | rs774510679 | ACTTTGCC | A | 0.4124 | 0.061 | 0.007 | 1.5849E-17 | 75.94 |
| *CCL14* | rs9903158 | T | C | 0.0527 | -1.013 | 0.017 | 1E-200 | 3550.76 |
| *CCL14* | rs1671152 | T | G | 0.8411 | 0.103 | 0.01 | 1.9953E-26 | 106.09 |
| *CCL15* | rs782134971 | GAAACTGC( | G | 0.2543 | -0.139 | 0.007 | 1.9953E-84 | 394.31 |
| *CCL15* | rs28929474 | C | T | 0.0208 | 0.261 | 0.022 | 2.5119E-33 | 140.75 |
| *CCL15* | rs601338 | G | A | 0.5089 | -0.181 | 0.006 | 6.31E-184 | 910.03 |
| *CCL16* | rs11127048 | G | A | 0.6181 | -0.043 | 0.006 | 5.0119E-12 | 51.36 |
| *CCL16* | rs1047891 | C | A | 0.313 | -0.049 | 0.006 | 3.9811E-14 | 66.69 |
| *CCL16* | rs3134984 | T | A | 0.2098 | -0.074 | 0.008 | 2.5119E-21 | 85.56 |
| *CCL16* | rs34958196 | G | A | 0.1289 | -0.067 | 0.009 | 3.9811E-14 | 55.42 |
| *CCL16* | rs112689088 | T | C | 0.096 | -1.332 | 0.012 | 1E-200 | 12321.00 |
| *CCL16* | rs769925299 | TC | T | 0.6385 | 0.051 | 0.006 | 7.9433E-16 | 72.25 |
| *CCL17* | rs12075 | G | A | 0.5832 | 0.054 | 0.007 | 1.2589E-13 | 59.51 |
| *CCL17* | rs142289859 | T | C | 0.0334 | -0.557 | 0.021 | 3.162E-162 | 703.51 |
| *CCL17* | rs116135711 | T | C | 0.0361 | -0.171 | 0.02 | 1.9953E-17 | 73.10 |
| *CCL17* | rs2228467 | T | C | 0.0612 | 0.425 | 0.015 | 3.981E-168 | 802.78 |
| *CCL17* | rs11925560 | A | T | 0.3883 | 0.061 | 0.008 | 7.9433E-16 | 58.14 |
| *CCL17* | rs771327 | A | G | 0.5588 | 0.056 | 0.007 | 6.3096E-14 | 64.00 |
| *CCL17* | rs3804749 | C | T | 0.5923 | -0.053 | 0.008 | 3.9811E-12 | 43.89 |
| *CCL17* | rs2073643 | T | C | 0.5521 | -0.059 | 0.007 | 2.5119E-16 | 71.04 |
| *CCL17* | rs3763287 | C | A | 0.065 | -0.102 | 0.015 | 1.9953E-12 | 46.24 |
| *CCL17* | rs6993770 | A | T | 0.2866 | -0.132 | 0.008 | 3.9811E-62 | 272.25 |
| *CCL17* | rs61469632 | T | C | 0.0633 | -0.114 | 0.015 | 1E-14 | 57.76 |
| *CCL17* | rs10886430 | A | G | 0.1269 | -0.365 | 0.011 | 1E-200 | 1101.03 |
| *CCL17* | rs9302690 | G | A | 0.081 | 0.381 | 0.013 | 7.943E-179 | 858.94 |
| *CCL17* | rs1654425 | T | C | 0.8343 | 0.085 | 0.01 | 1E-18 | 72.25 |
| *CCL18* | rs6715862 | A | G | 0.5025 | -0.077 | 0.007 | 7.9433E-29 | 121.00 |
| *CCL18* | rs2015086 | A | G | 0.1328 | 0.706 | 0.011 | 1E-200 | 4119.31 |
| *CCL18* | rs145144275 | A | G | 0.0089 | -0.343 | 0.037 | 6.3096E-21 | 85.94 |
| *CCL18* | rs56298400 | G | A | 0.0829 | 0.089 | 0.012 | 7.9433E-13 | 55.01 |
| *CCL19* | rs2476601 | A | G | 0.8992 | -0.113 | 0.012 | 1.5849E-20 | 88.67 |
| *CCL19* | rs2305625 | C | T | 0.1118 | -0.148 | 0.012 | 3.9811E-37 | 152.11 |
| *CCL19* | rs201980192 | C | T | 0.0193 | 0.888 | 0.028 | 1E-200 | 1005.80 |
| *CCL19* | rs10972202 | C | T | 0.1227 | 0.105 | 0.011 | 7.9433E-21 | 91.12 |
| *CCL19* | rs35350651 | A | AC | 0.5038 | -0.065 | 0.007 | 3.1623E-19 | 86.22 |
| *CCL19* | rs2412973 | C | A | 0.4429 | -0.053 | 0.007 | 3.9811E-13 | 57.33 |
| *CCL2* | rs12075 | G | A | 0.583 | 0.247 | 0.007 | 1E-200 | 1245.08 |
| *CCL2* | rs2228467 | T | C | 0.0614 | 0.137 | 0.015 | 3.1623E-20 | 83.42 |
| *CCL2* | rs10510751 | T | G | 0.0709 | 0.281 | 0.014 | 1.2589E-89 | 402.86 |
| *CCL2* | rs4541868 | C | A | 0.2736 | -0.057 | 0.008 | 1.2589E-12 | 50.77 |
| *CCL2* | rs7080386 | C | A | 0.4118 | 0.067 | 0.007 | 1.5849E-20 | 91.61 |
| *CCL2* | rs76428106 | T | C | 0.0129 | -0.277 | 0.033 | 1.9953E-17 | 70.46 |
| *CCL2* | rs12601658 | T | A | 0.2287 | -0.068 | 0.008 | 7.9433E-16 | 72.25 |
| *CCL2* | rs1671152 | T | G | 0.8408 | 0.096 | 0.01 | 1E-22 | 92.16 |
| *CCL20* | rs1395338 | G | A | 0.6005 | 0.093 | 0.008 | 1.5849E-34 | 135.14 |
| *CCL20* | rs3129754 | A | G | 0.42 | 0.082 | 0.008 | 5.0119E-27 | 105.06 |
| *CCL20* | rs742493 | T | C | 0.1133 | -0.118 | 0.012 | 1.2589E-23 | 96.69 |
| *CCL20* | rs77542162 | A | G | 0.0229 | 0.27 | 0.025 | 1.9953E-27 | 116.64 |
| *CCL20* | rs117900576 | C | G | 0.0337 | 0.172 | 0.021 | 5.0119E-17 | 67.08 |
| *CCL21* | rs4671127 | C | G | 0.335 | -0.054 | 0.008 | 3.9811E-12 | 45.56 |
| *CCL21* | rs1354034 | T | C | 0.6041 | -0.065 | 0.008 | 5.0119E-18 | 66.02 |
| *CCL21* | rs11924792 | A | C | 0.1123 | -0.177 | 0.012 | 1E-51 | 217.56 |
| *CCL21* | rs885122 | A | G | 0.3949 | -0.062 | 0.008 | 6.3096E-16 | 60.06 |
| *CCL21* | rs17764730 | C | T | 0.2486 | -0.087 | 0.009 | 3.1623E-24 | 93.44 |
| *CCL21* | rs12722013 | T | C | 0.0474 | 0.315 | 0.017 | 3.1623E-73 | 343.34 |
| *CCL21* | rs10814138 | C | A | 0.3171 | -0.103 | 0.008 | 1.5849E-38 | 165.77 |
| *CCL21* | rs3184504 | T | C | 0.5173 | -0.079 | 0.007 | 3.1623E-27 | 127.37 |
| *CCL21* | rs12935829 | T | G | 0.3435 | 0.053 | 0.008 | 1E-11 | 43.89 |
| *CCL21* | rs62054652 | C | T | 0.071 | -0.107 | 0.014 | 1.2589E-13 | 58.41 |
| *CCL21* | rs2412973 | C | A | 0.4432 | -0.07 | 0.007 | 3.1623E-21 | 100.00 |
| *CCL22* | rs13098877 | C | T | 0.4445 | -0.052 | 0.008 | 3.9811E-12 | 42.25 |
| *CCL22* | rs9296027 | C | G | 0.1219 | 0.091 | 0.011 | 1E-15 | 68.44 |
| *CCL22* | rs1504215 | G | A | 0.3507 | -0.065 | 0.008 | 1E-16 | 66.02 |
| *CCL22* | rs3184504 | T | C | 0.517 | -0.057 | 0.007 | 1.2589E-14 | 66.31 |
| *CCL22* | rs41398344 | T | C | 0.0098 | -0.824 | 0.038 | 1.585E-104 | 470.20 |
| *CCL22* | rs77542162 | A | G | 0.0228 | 0.512 | 0.025 | 1E-92 | 419.43 |
| *CCL22* | rs11670056 | C | T | 0.074 | 0.107 | 0.014 | 3.9811E-14 | 58.41 |
| *CCL23* | rs61804164 | G | C | 0.1239 | -0.082 | 0.011 | 3.9811E-14 | 55.57 |

| | | | | | | | | |
|---|---|---|---|---|---|---|---|---|
| *CCL23* | rs780093 | T | C | 0.6197 | -0.066 | 0.007 | 7.9433E-20 | 88.90 |
| *CCL23* | rs142021145 | C | A | 0.0035 | 0.58 | 0.059 | 1.2589E-22 | 96.64 |
| *CCL23* | rs9260620 | T | G | 0.3291 | -0.055 | 0.008 | 3.1623E-13 | 47.27 |
| *CCL23* | rs964184 | G | C | 0.8668 | -0.104 | 0.01 | 7.9433E-24 | 108.16 |
| *CCL23* | rs11545007 | C | A | 0.0086 | -0.353 | 0.04 | 1.9953E-18 | 77.88 |
| *CCL23* | rs712048 | A | C | 0.869 | 0.569 | 0.011 | 1E-200 | 2675.71 |
| *CCL23* | rs1065853 | G | T | 0.0813 | 0.113 | 0.013 | 3.1623E-18 | 75.56 |
| *CCL24* | rs12083692 | A | T | 0.2587 | -0.052 | 0.005 | 6.3096E-24 | 108.16 |
| *CCL24* | rs2228467 | T | C | 0.0612 | 0.173 | 0.009 | 1E-74 | 369.49 |
| *CCL24* | rs7441544 | G | C | 0.5466 | 0.034 | 0.005 | 2.5119E-12 | 46.24 |
| *CCL24* | rs115216147 | T | A | 0.0666 | -0.076 | 0.009 | 2.5119E-16 | 71.31 |
| *CCL24* | rs111872865 | G | A | 0.0127 | 0.152 | 0.022 | 5.0119E-12 | 47.74 |
| *CCL24* | rs2024050 | A | G | 0.8952 | -1.375 | 0.012 | 1E-200 | 13129.34 |
| *CCL24* | rs2737246 | G | C | 0.2823 | -0.037 | 0.005 | 1.5849E-13 | 54.76 |
| *CCL24* | rs13294683 | C | T | 0.1015 | 0.071 | 0.007 | 3.9811E-21 | 102.88 |
| *CCL24* | rs10769256 | C | T | 0.4002 | 0.06 | 0.005 | 1.2589E-37 | 144.00 |
| *CCL24* | rs201510256 | C | T | 0.4738 | -0.043 | 0.005 | 1.9953E-15 | 73.96 |
| *CCL24* | rs8012229 | T | C | 0.0876 | -0.056 | 0.008 | 3.9811E-12 | 49.00 |
| *CCL24* | rs2939849 | C | G | 0.3963 | 0.056 | 0.005 | 1.9953E-33 | 125.44 |
| *CCL24* | rs1121985 | A | C | 0.6217 | 0.048 | 0.005 | 5.0119E-24 | 92.16 |
| *CCL24* | rs113374757 | C | T | 0.1655 | -0.042 | 0.006 | 1.2589E-11 | 49.00 |
| *CCL24* | rs11480320 | A | AT | 0.5843 | 0.034 | 0.005 | 1.2589E-12 | 46.24 |
| *CCL25* | rs9275576 | C | T | 0.1441 | 0.062 | 0.008 | 2.5119E-13 | 60.06 |
| *CCL25* | rs78061563 | G | A | 0.0731 | 0.094 | 0.011 | 1.9953E-16 | 73.02 |
| *CCL25* | rs118062058 | A | G | 0.0562 | -0.15 | 0.013 | 5.0119E-31 | 133.14 |
| *CCL25* | rs7295693 | T | C | 0.5927 | 0.123 | 0.006 | 1.9953E-91 | 420.25 |
| *CCL25* | rs186021206 | G | A | 0.0064 | 0.387 | 0.041 | 3.1623E-21 | 89.10 |
| *CCL25* | rs7223866 | C | G | 0.3301 | 0.044 | 0.006 | 6.3096E-12 | 53.78 |
| *CCL25* | rs112001035 | G | A | 0.0612 | -0.1 | 0.013 | 2.5119E-15 | 59.17 |
| *CCL25* | rs2032887 | A | G | 0.2369 | 0.77 | 0.009 | 1E-200 | 7319.75 |
| *CCL25* | rs601338 | G | A | 0.5089 | -0.39 | 0.007 | 1E-200 | 3104.08 |
| *CCL25* | rs34710186 | T | TA | 0.811 | 0.067 | 0.008 | 1E-17 | 70.14 |
| *CCL26* | rs12075 | G | A | 0.5829 | 0.192 | 0.008 | 1.585E-144 | 576.00 |
| *CCL26* | rs13412535 | G | A | 0.2307 | -0.061 | 0.009 | 5.0119E-12 | 45.94 |
| *CCL26* | rs17622656 | G | A | 0.3887 | -0.053 | 0.008 | 3.9811E-12 | 43.89 |
| *CCL26* | rs3135383 | T | G | 0.1284 | -0.094 | 0.011 | 2.5119E-17 | 73.02 |
| *CCL26* | rs41463245 | C | T | 0.0089 | -1.014 | 0.039 | 6.31E-147 | 676.00 |
| *CCL26* | rs6993770 | A | T | 0.2869 | -0.067 | 0.008 | 1.9953E-16 | 70.14 |
| *CCL26* | rs10761731 | A | T | 0.4147 | 0.057 | 0.007 | 2.5119E-14 | 66.31 |
| *CCL26* | rs1654425 | T | C | 0.8339 | 0.096 | 0.01 | 6.3096E-22 | 92.16 |
| *CCL27* | rs1150756 | C | T | 0.1313 | -0.095 | 0.011 | 2.5119E-19 | 74.59 |
| *CCL27* | rs118039278 | G | A | 0.0817 | 0.108 | 0.013 | 1.5849E-16 | 69.02 |
| *CCL27* | rs2070074 | A | G | 0.097 | -0.328 | 0.012 | 1.585E-159 | 747.11 |
| *CCL27* | rs964184 | G | C | 0.8667 | -0.106 | 0.01 | 5.0119E-24 | 112.36 |
| *CCL27* | rs77542162 | A | G | 0.0226 | 0.306 | 0.024 | 1.9953E-37 | 162.56 |
| *CCL27* | rs7412 | C | T | 0.0816 | 0.148 | 0.013 | 1E-29 | 129.61 |
| *CCL27* | rs2569491 | G | A | 0.298 | -0.058 | 0.008 | 1.2589E-13 | 52.56 |
| *CCL27* | rs74278448 | C | CGGG | 0.704 | 0.099 | 0.008 | 3.1623E-36 | 153.14 |
| *CCL28* | rs7535275 | A | G | 0.1325 | -0.078 | 0.011 | 5.0119E-13 | 50.28 |
| *CCL28* | rs41267433 | C | G | 0.1108 | 0.085 | 0.012 | 3.1623E-13 | 50.17 |
| *CCL28* | rs3732083 | T | C | 0.43 | -0.182 | 0.007 | 1.259E-131 | 676.00 |
| *CCL28* | rs13412535 | G | A | 0.2307 | -0.15 | 0.009 | 2.5119E-65 | 277.78 |
| *CCL28* | rs2278668 | T | C | 0.5935 | -0.068 | 0.008 | 1.2589E-19 | 72.25 |
| *CCL28* | rs55874224 | G | C | 0.3922 | 0.067 | 0.007 | 3.1623E-19 | 91.61 |
| *CCL28* | rs11734132 | G | C | 0.1738 | -0.065 | 0.01 | 1.2589E-11 | 42.25 |
| *CCL28* | rs200708155 | G | GTT | 0.5006 | -0.057 | 0.007 | 1E-14 | 66.31 |
| *CCL28* | rs114694170 | T | C | 0.0598 | 0.122 | 0.016 | 5.0119E-15 | 58.14 |
| *CCL28* | rs112838960 | T | TAATA | 0.5154 | 0.067 | 0.007 | 5.0119E-20 | 91.61 |
| *CCL28* | rs77024756 | A | G | 0.7025 | -0.072 | 0.009 | 7.9433E-15 | 64.00 |
| *CCL28* | rs1331309 | T | G | 0.2606 | 0.066 | 0.008 | 3.9811E-15 | 68.06 |
| *CCL28* | rs139141690 | G | A | 0.0047 | -0.648 | 0.054 | 5.0119E-33 | 144.00 |
| *CCL28* | rs2519093 | C | T | 0.1842 | -0.173 | 0.009 | 1.2589E-74 | 369.49 |
| *CCL28* | rs73000929 | G | A | 0.0373 | -0.247 | 0.019 | 1.2589E-37 | 169.00 |
| *CCL28* | rs56990409 | G | GA | 0.5505 | 0.087 | 0.007 | 2.5119E-31 | 154.47 |
| *CCL28* | rs60822569 | T | TTTCTC | 0.5545 | 0.131 | 0.007 | 1.2589E-69 | 350.22 |
| *CCL28* | rs4965385 | A | G | 0.2844 | -0.057 | 0.008 | 1.5849E-12 | 50.77 |
| *CCL28* | rs892090 | T | G | 0.8336 | 0.08 | 0.01 | 3.9811E-16 | 64.00 |
| *CCL28* | rs71329093 | A | G | 0.056 | -0.13 | 0.016 | 3.1623E-16 | 66.02 |

| | | | | | | | | |
|---|---|---|---|---|---|---|---|---|
| *CCL3* | rs60939770 | A | G | 0.3162 | -0.054 | 0.008 | 1E-12 | 45.56 |
| *CCL3* | rs1354034 | T | C | 0.6043 | -0.051 | 0.007 | 7.9433E-13 | 53.08 |
| *CCL3* | rs756846124 | CT | C | 0.1787 | 0.064 | 0.009 | 3.9811E-12 | 50.57 |
| *CCL3* | rs13153461 | G | A | 0.7433 | 0.072 | 0.008 | 1.9953E-19 | 81.00 |
| *CCL3* | rs9268644 | A | C | 0.5472 | 0.055 | 0.007 | 3.9811E-15 | 61.73 |
| *CCL3* | rs3184504 | T | C | 0.5174 | -0.073 | 0.007 | 1E-25 | 108.76 |
| *CCL3* | rs7485656 | A | G | 0.1401 | 0.075 | 0.01 | 7.9433E-14 | 56.25 |
| *CCL3* | rs1634486 | A | C | 0.2239 | 0.42 | 0.009 | 1E-200 | 2177.78 |
| *CCL4* | rs113341849 | G | A | 0.1185 | 0.615 | 0.011 | 1E-200 | 3125.83 |
| *CCL4* | rs756846124 | CT | C | 0.1788 | 0.073 | 0.009 | 6.3096E-16 | 65.79 |
| *CCL4* | rs9405083 | C | T | 0.251 | -0.057 | 0.008 | 1E-12 | 50.77 |
| *CCL4* | rs3184504 | T | C | 0.5171 | -0.055 | 0.007 | 7.9433E-16 | 61.73 |
| *CCL4* | rs13338037 | C | G | 0.1408 | 0.079 | 0.01 | 1.2589E-15 | 62.41 |
| *CCL4* | rs1015673 | C | T | 0.4249 | -0.314 | 0.008 | 1E-200 | 1540.56 |
| *CCL4* | rs892090 | T | G | 0.8342 | 0.07 | 0.009 | 2.5119E-14 | 60.49 |
| *CCL5* | rs1354034 | T | C | 0.6037 | -0.071 | 0.007 | 1.9953E-21 | 102.88 |
| *CCL5* | rs6961069 | C | T | 0.4025 | 0.063 | 0.008 | 3.1623E-17 | 62.02 |
| *CCL5* | rs6993770 | A | T | 0.2865 | -0.093 | 0.008 | 5.0119E-30 | 135.14 |
| *CCL5* | rs35754228 | A | AT | 0.3241 | -0.077 | 0.008 | 6.3096E-22 | 92.64 |
| *CCL5* | rs10761731 | A | T | 0.4147 | 0.128 | 0.007 | 2.5119E-65 | 334.37 |
| *CCL5* | rs1362213 | C | T | 0.4797 | -0.055 | 0.007 | 1.5849E-13 | 61.73 |
| *CCL5* | rs12445050 | C | T | 0.1384 | 0.081 | 0.011 | 2.5119E-14 | 54.22 |
| *CCL5* | rs2107538 | C | T | 0.1802 | -0.22 | 0.01 | 6.31E-115 | 484.00 |
| *CCL5* | rs1654425 | T | C | 0.8347 | 0.126 | 0.01 | 6.3096E-37 | 158.76 |
| *CCL5* | rs3827978 | C | T | 0.3539 | 0.064 | 0.008 | 1E-16 | 64.00 |
| *CCL7* | rs12075 | G | A | 0.5829 | -0.272 | 0.007 | 1E-200 | 1509.88 |
| *CCL7* | rs2228467 | T | C | 0.0611 | 0.301 | 0.015 | 1E-87 | 402.67 |
| *CCL7* | rs28380853 | T | A | 0.1448 | 0.094 | 0.011 | 7.9433E-17 | 73.02 |
| *CCL7* | rs6505397 | T | A | 0.4229 | 0.105 | 0.007 | 5.0119E-47 | 225.00 |
| *CCL7* | rs1654425 | T | C | 0.8339 | 0.081 | 0.01 | 1E-16 | 65.61 |
| *CCL8* | rs12075 | G | A | 0.5828 | -0.2 | 0.006 | 1E-200 | 1111.11 |
| *CCL8* | rs35173808 | C | A | 0.0964 | -0.088 | 0.01 | 2.5119E-17 | 77.44 |
| *CCL8* | rs2190504 | T | C | 0.4438 | 0.043 | 0.006 | 3.9811E-12 | 51.36 |
| *CCL8* | rs2737245 | G | T | 0.2817 | -0.053 | 0.007 | 1.9953E-14 | 57.33 |
| *CCL8* | rs61469632 | T | C | 0.0634 | -0.086 | 0.013 | 1.2589E-11 | 43.76 |
| *CCL8* | rs3740688 | G | T | 0.5432 | -0.046 | 0.006 | 1.5849E-13 | 58.78 |
| *CCL8* | rs1133763 | A | C | 0.1542 | -1.074 | 0.01 | 1E-200 | 11534.76 |
| *CCL8* | rs62125968 | G | A | 0.2347 | 0.054 | 0.007 | 1.2589E-13 | 59.51 |
| *CCL8* | rs892090 | T | G | 0.8341 | 0.089 | 0.008 | 1E-26 | 123.77 |
| *CCN1* | rs4949896 | G | T | 0.262 | -0.118 | 0.008 | 1E-45 | 217.56 |
| *CCN1* | rs1354034 | T | C | 0.6047 | -0.058 | 0.007 | 6.3096E-15 | 68.65 |
| *CCN1* | rs55730499 | C | T | 0.0822 | 0.117 | 0.013 | 1.2589E-18 | 81.00 |
| *CCN1* | rs328 | C | G | 0.1007 | -0.126 | 0.012 | 1.2589E-25 | 110.25 |
| *CCN1* | rs7848055 | A | C | 0.3289 | -0.082 | 0.008 | 5.0119E-26 | 105.06 |
| *CCN1* | rs964184 | G | C | 0.8668 | -0.133 | 0.011 | 3.1623E-35 | 146.19 |
| *CCN1* | rs11846959 | G | A | 0.2533 | 0.08 | 0.008 | 1.2589E-21 | 100.00 |
| *CCN1* | rs186021206 | G | A | 0.0063 | 0.419 | 0.05 | 1E-16 | 70.22 |
| *CCN1* | rs72654473 | C | A | 0.1092 | 0.122 | 0.012 | 1.9953E-25 | 103.36 |
| *CCN2* | rs1434282 | C | T | 0.7265 | -0.078 | 0.008 | 6.3096E-21 | 95.06 |
| *CCN2* | rs13412535 | G | A | 0.231 | -0.06 | 0.009 | 1E-11 | 44.44 |
| *CCN2* | rs1354034 | T | C | 0.6039 | -0.097 | 0.007 | 3.1623E-38 | 192.02 |
| *CCN2* | rs28625045 | G | A | 0.3931 | -0.059 | 0.008 | 5.0119E-15 | 54.39 |
| *CCN2* | rs2631367 | C | G | 0.5183 | 0.055 | 0.007 | 6.3096E-14 | 61.73 |
| *CCN2* | rs2516471 | G | A | 0.0636 | -0.114 | 0.016 | 2.5119E-13 | 50.77 |
| *CCN2* | rs1917342 | A | G | 0.3038 | -0.057 | 0.008 | 3.9811E-12 | 50.77 |
| *CCN2* | rs7767139 | C | G | 0.216 | -0.355 | 0.009 | 1E-200 | 1555.86 |
| *CCN2* | rs13236689 | T | G | 0.4079 | 0.066 | 0.007 | 1E-18 | 88.90 |
| *CCN2* | rs4631576 | C | T | 0.361 | 0.058 | 0.008 | 2.5119E-14 | 52.56 |
| *CCN2* | rs10761731 | A | T | 0.4147 | 0.122 | 0.007 | 5.0119E-60 | 303.76 |
| *CCN2* | rs79977579 | C | A | 0.0992 | 0.086 | 0.012 | 3.9811E-12 | 51.36 |
| *CCN2* | rs59001897 | T | A | 0.1745 | 0.076 | 0.01 | 1E-14 | 57.76 |
| *CCN2* | rs11075045 | A | G | 0.1816 | 0.067 | 0.01 | 2.5119E-12 | 44.89 |
| *CCN2* | rs12445050 | C | T | 0.138 | 0.11 | 0.011 | 3.1623E-25 | 100.00 |
| *CCN2* | rs1654425 | T | C | 0.8344 | 0.152 | 0.01 | 1.2589E-52 | 231.04 |
| *CCN2* | rs6081565 | G | A | 0.3524 | 0.066 | 0.008 | 7.9433E-18 | 68.06 |
| *CCN2* | rs56045941 | T | C | 0.1701 | 0.073 | 0.01 | 1.5849E-13 | 53.29 |
| *CCN3* | rs2279112 | G | A | 0.2587 | -0.274 | 0.008 | 1E-200 | 1173.06 |
| *CCN3* | rs7827593 | G | A | 0.135 | 0.078 | 0.01 | 3.1623E-14 | 60.84 |

| | | | | | | | | |
|---|---|---|---|---|---|---|---|---|
| *CCN3* | rs11172113 | T | C | 0.4127 | 0.049 | 0.007 | 6.3096E-13 | 49.00 |
| *CCN4* | rs2274319 | T | C | 0.6531 | -0.051 | 0.007 | 6.3096E-13 | 53.08 |
| *CCN4* | rs68066031 | T | C | 0.2299 | -0.096 | 0.008 | 3.9811E-33 | 144.00 |
| *CCN4* | rs1354034 | T | C | 0.6041 | -0.079 | 0.007 | 3.9811E-31 | 127.37 |
| *CCN4* | rs10033707 | T | G | 0.3374 | -0.051 | 0.007 | 1.2589E-12 | 53.08 |
| *CCN4* | rs114694170 | T | C | 0.0599 | 0.129 | 0.014 | 1.5849E-19 | 84.90 |
| *CCN4* | rs139141690 | G | A | 0.0048 | 0.366 | 0.05 | 1.5849E-13 | 53.58 |
| *CCN4* | rs4541868 | C | A | 0.2736 | -0.063 | 0.008 | 5.0119E-14 | 62.02 |
| *CCN4* | rs12156037 | A | G | 0.3536 | -0.439 | 0.008 | 1E-200 | 3011.27 |
| *CCN4* | rs150813342 | C | T | 0.0085 | 0.294 | 0.038 | 6.3096E-15 | 59.86 |
| *CCN4* | rs774510679 | ACTTTGCC | A | 0.4125 | 0.064 | 0.007 | 1.2589E-20 | 83.59 |
| *CCN4* | rs11591571 | G | A | 0.3408 | 0.057 | 0.007 | 1E-15 | 66.31 |
| *CCN4* | rs562526200 | G | GGCCGCC | 0.7739 | 0.074 | 0.008 | 6.3096E-20 | 85.56 |
| *CCN4* | rs10424665 | A | C | 0.1715 | 0.069 | 0.009 | 7.9433E-15 | 58.78 |
| *CCN4* | rs1654425 | T | C | 0.8343 | 0.079 | 0.009 | 2.5119E-18 | 77.05 |
| *CCN5* | rs4859682 | C | A | 0.4568 | 0.047 | 0.007 | 3.1623E-12 | 45.08 |
| *CCNE1* | rs139299944 | C | CT | 0.3437 | 0.11 | 0.008 | 2.5119E-41 | 189.06 |
| *CCS* | rs498045 | A | G | 0.526 | 0.481 | 0.007 | 1E-200 | 4721.65 |
| *CD101* | rs12130298 | A | C | 0.0752 | -1.278 | 0.014 | 1E-200 | 8333.08 |
| *CD101* | rs9271374 | G | A | 0.8063 | -0.096 | 0.007 | 7.9433E-39 | 188.08 |
| *CD101* | rs12667978 | C | T | 0.5712 | -0.041 | 0.006 | 2.5119E-12 | 46.69 |
| *CD101* | rs2082209 | A | G | 0.4448 | 0.068 | 0.006 | 7.9433E-32 | 128.44 |
| *CD101* | rs653178 | C | T | 0.5166 | -0.06 | 0.006 | 1.9953E-25 | 100.00 |
| *CD101* | rs4893 | A | G | 0.0398 | -0.17 | 0.015 | 6.3096E-31 | 128.44 |
| *CD101* | rs150358287 | C | A | 0.0119 | 0.333 | 0.027 | 6.3096E-36 | 152.11 |
| *CD109* | rs1260326 | T | C | 0.6084 | 0.047 | 0.006 | 1E-15 | 61.36 |
| *CD109* | rs8176749 | C | T | 0.0612 | 0.367 | 0.012 | 1E-200 | 935.34 |
| *CD109* | rs56278466 | T | G | 0.6612 | 0.096 | 0.006 | 3.9811E-57 | 256.00 |
| *CD109* | rs186021206 | G | A | 0.0064 | 0.554 | 0.039 | 6.3096E-46 | 201.79 |
| *CD109* | rs868875 | A | G | 0.3013 | -0.049 | 0.006 | 3.9811E-15 | 66.69 |
| *CD14* | rs3213563 | T | C | 0.559 | 0.096 | 0.007 | 1E-38 | 188.08 |
| *CD14* | rs5744441 | G | A | 0.2331 | -0.321 | 0.009 | 1E-200 | 1272.11 |
| *CD14* | rs28601761 | C | G | 0.4191 | -0.075 | 0.008 | 3.1623E-23 | 87.89 |
| *CD14* | rs529565 | T | C | 0.3174 | -0.065 | 0.008 | 2.5119E-16 | 66.02 |
| *CD14* | rs56278466 | T | G | 0.6615 | 0.09 | 0.008 | 3.9811E-31 | 126.56 |
| *CD14* | rs9414801 | G | A | 0.5332 | -0.062 | 0.007 | 7.9433E-17 | 78.45 |
| *CD14* | rs149394327 | G | C | 0.0299 | 0.171 | 0.022 | 2.5119E-15 | 60.42 |
| *CD160* | rs6699680 | A | G | 0.4154 | -0.055 | 0.007 | 1.9953E-13 | 61.73 |
| *CD160* | rs12562475 | C | T | 0.1032 | -0.128 | 0.013 | 1.2589E-22 | 96.95 |
| *CD160* | rs9728526 | G | A | 0.4396 | -0.394 | 0.008 | 1E-200 | 2425.56 |
| *CD160* | rs370856537 | A | C | 0.0805 | -0.211 | 0.015 | 5.0119E-43 | 197.87 |
| *CD160* | rs2009581 | G | A | 0.2724 | -0.162 | 0.008 | 7.9433E-89 | 410.06 |
| *CD160* | rs186535521 | C | T | 0.0169 | -0.285 | 0.03 | 6.3096E-22 | 90.25 |
| *CD160* | rs777572865 | CCCTACTTC | C | 0.3273 | -0.071 | 0.008 | 5.0119E-20 | 78.77 |
| *CD160* | rs2844616 | T | C | 0.4144 | 0.109 | 0.007 | 3.9811E-50 | 242.47 |
| *CD160* | rs8177590 | A | G | 0.4317 | -0.06 | 0.007 | 1.5849E-16 | 73.47 |
| *CD160* | rs2497318 | C | T | 0.452 | 0.094 | 0.007 | 1.5849E-38 | 180.33 |
| *CD160* | rs3016275 | C | T | 0.3369 | -0.086 | 0.008 | 6.3096E-30 | 115.56 |
| *CD160* | rs3184504 | T | C | 0.5174 | -0.14 | 0.007 | 1.2589E-85 | 400.00 |
| *CD160* | rs58745116 | G | A | 0.3897 | -0.075 | 0.007 | 3.9811E-24 | 114.80 |
| *CD160* | rs601338 | G | A | 0.5089 | -0.066 | 0.007 | 3.9811E-20 | 88.90 |
| *CD163* | rs2099684 | A | G | 0.3154 | 0.06 | 0.008 | 6.3096E-15 | 56.25 |
| *CD163* | rs62165726 | C | A | 0.0369 | -0.689 | 0.019 | 1E-200 | 1315.02 |
| *CD163* | rs190303734 | A | T | 0.2965 | 0.062 | 0.008 | 7.9433E-15 | 60.06 |
| *CD163* | rs11923060 | C | T | 0.1569 | -0.077 | 0.01 | 1.9953E-15 | 59.29 |
| *CD163* | rs115216147 | T | A | 0.0665 | 0.166 | 0.015 | 3.9811E-30 | 122.47 |
| *CD163* | rs11381821 | C | CA | 0.4677 | -0.055 | 0.007 | 1.9953E-14 | 61.73 |
| *CD163* | rs10267707 | C | G | 0.454 | 0.053 | 0.007 | 1.2589E-13 | 57.33 |
| *CD163* | rs2737231 | A | G | 0.3053 | -0.058 | 0.008 | 5.0119E-14 | 52.56 |
| *CD163* | rs2031902 | C | T | 0.5875 | -0.091 | 0.007 | 1E-35 | 169.00 |
| *CD163* | rs635634 | C | T | 0.184 | 0.093 | 0.009 | 3.9811E-24 | 106.78 |
| *CD163* | rs768890137 | CT | C | 0.3865 | 0.067 | 0.007 | 5.0119E-20 | 91.61 |
| *CD163* | rs11220505 | C | T | 0.2226 | 0.13 | 0.009 | 1E-51 | 208.64 |
| *CD163* | rs11054859 | G | A | 0.1027 | -0.186 | 0.012 | 7.9433E-57 | 240.25 |
| *CD163* | rs10774625 | A | G | 0.5039 | -0.078 | 0.007 | 1.2589E-28 | 124.16 |
| *CD163* | rs217184 | T | C | 0.1961 | -0.08 | 0.009 | 2.5119E-19 | 79.01 |
| *CD163* | rs186021206 | G | A | 0.0064 | 1.793 | 0.049 | 1E-200 | 1338.96 |
| *CD163* | rs601338 | G | A | 0.5094 | 0.059 | 0.007 | 1.2589E-16 | 71.04 |

| | | | | | | | | |
|---|---|---|---|---|---|---|---|---|
| *CD164* | rs4950771 | T | G | 0.9342 | -0.166 | 0.015 | 5.0119E-28 | 122.47 |
| *CD164* | rs45492299 | G | A | 0.1754 | -0.075 | 0.01 | 2.5119E-14 | 56.25 |
| *CD164* | rs9857570 | A | G | 0.324 | -0.094 | 0.008 | 6.3096E-32 | 138.06 |
| *CD164* | rs12213313 | C | G | 0.3299 | -0.149 | 0.008 | 7.9433E-78 | 346.89 |
| *CD164* | rs4734879 | A | G | 0.2742 | -0.058 | 0.008 | 6.3096E-12 | 52.56 |
| *CD164* | rs10820606 | A | C | 0.2299 | -0.073 | 0.009 | 6.3096E-16 | 65.79 |
| *CD164* | rs201220977 | C | CCTCT | 0.4659 | 0.066 | 0.008 | 5.0119E-18 | 68.06 |
| *CD164* | rs72701845 | G | A | 0.038 | -0.202 | 0.02 | 1.5849E-23 | 102.01 |
| *CD164* | rs892090 | T | G | 0.8342 | 0.096 | 0.01 | 2.5119E-21 | 92.16 |
| *CD164L2* | rs734725 | G | T | 0.0862 | 0.792 | 0.014 | 1E-200 | 3200.33 |
| *CD177* | rs78530667 | T | C | 0.1283 | -0.74 | 0.012 | 1E-200 | 3802.78 |
| *CD1C* | rs61788913 | G | T | 0.128 | -0.1 | 0.011 | 5.0119E-20 | 82.64 |
| *CD1C* | rs115248892 | G | T | 0.0027 | -1.624 | 0.069 | 1.995E-121 | 553.95 |
| *CD1C* | rs4384720 | C | G | 0.9196 | 0.105 | 0.013 | 6.3096E-16 | 65.24 |
| *CD1C* | rs10189685 | G | A | 0.2969 | -0.059 | 0.008 | 5.0119E-14 | 54.39 |
| *CD1C* | rs4698932 | G | A | 0.4941 | 0.053 | 0.007 | 1.5849E-13 | 57.33 |
| *CD1C* | rs9265969 | G | A | 0.1632 | 0.229 | 0.01 | 2.512E-121 | 524.41 |
| *CD1C* | rs11963621 | T | C | 0.1037 | 0.115 | 0.012 | 2.5119E-22 | 91.84 |
| *CD1C* | rs1504215 | G | A | 0.3503 | -0.097 | 0.008 | 1E-37 | 147.02 |
| *CD1C* | rs2721934 | C | T | 0.5591 | -0.051 | 0.007 | 1.9953E-12 | 53.08 |
| *CD1C* | rs139130389 | C | CTA | 0.0726 | -0.419 | 0.014 | 1E-200 | 895.72 |
| *CD1C* | rs572608 | A | T | 0.547 | 0.059 | 0.007 | 2.5119E-16 | 71.04 |
| *CD1C* | rs11218738 | G | A | 0.2491 | 0.069 | 0.008 | 6.3096E-17 | 74.39 |
| *CD1C* | rs150926693 | C | CTT | 0.1026 | 0.081 | 0.012 | 6.3096E-12 | 45.56 |
| *CD1C* | rs653178 | C | T | 0.5167 | -0.106 | 0.007 | 1E-50 | 229.31 |
| *CD1C* | rs76428106 | T | C | 0.0129 | 0.376 | 0.033 | 1.2589E-30 | 129.82 |
| *CD1C* | rs12599288 | T | C | 0.2961 | -0.113 | 0.008 | 2.5119E-47 | 199.52 |
| *CD1C* | rs60969446 | T | C | 0.5697 | -0.056 | 0.007 | 7.9433E-15 | 64.00 |
| *CD1C* | rs17272847 | G | A | 0.0874 | 0.095 | 0.013 | 3.9811E-14 | 53.40 |
| *CD1C* | rs6063502 | A | G | 0.3475 | 0.067 | 0.007 | 3.1623E-19 | 91.61 |
| *CD200* | rs1260326 | T | C | 0.6085 | 0.058 | 0.007 | 3.1623E-15 | 68.65 |
| *CD200* | rs6775517 | G | T | 0.3674 | 0.07 | 0.008 | 7.9433E-20 | 76.56 |
| *CD200* | rs13318932 | C | T | 0.1052 | 0.084 | 0.012 | 3.1623E-12 | 49.00 |
| *CD200* | rs2399420 | C | A | 0.9025 | 0.274 | 0.012 | 1E-108 | 521.36 |
| *CD200* | rs8176747 | C | G | 0.0612 | 0.286 | 0.015 | 3.9811E-80 | 363.54 |
| *CD200* | rs56278466 | T | G | 0.6618 | 0.099 | 0.008 | 3.1623E-38 | 153.14 |
| *CD200* | rs35830321 | C | CA | 0.1667 | -0.067 | 0.01 | 6.3096E-12 | 44.89 |
| *CD200* | rs2008174 | C | T | 0.7393 | -0.086 | 0.008 | 1.9953E-24 | 115.56 |
| *CD200R1* | rs72899532 | G | T | 0.1943 | -0.058 | 0.007 | 1E-14 | 68.65 |
| *CD200R1* | rs10935473 | G | T | 0.4399 | -0.317 | 0.008 | 1E-200 | 1570.14 |
| *CD200R1* | rs16860233 | C | T | 0.0389 | 1.825 | 0.02 | 1E-200 | 8326.56 |
| *CD200R1* | rs3129782 | G | A | 0.6767 | -0.076 | 0.006 | 2.5119E-32 | 160.44 |
| *CD200R1* | rs544503442 | G | A | 0.1427 | 0.072 | 0.009 | 1E-15 | 64.00 |
| *CD200R1* | rs10823374 | A | G | 0.4449 | 0.057 | 0.006 | 5.0119E-22 | 90.25 |
| *CD200R1* | rs4938939 | G | A | 0.2894 | 0.047 | 0.006 | 5.0119E-13 | 61.36 |
| *CD200R1* | rs75071241 | G | A | 0.0596 | -0.116 | 0.012 | 1.5849E-20 | 93.44 |
| *CD200R1* | rs10774625 | A | G | 0.5041 | -0.055 | 0.006 | 6.3096E-21 | 84.03 |
| *CD200R1* | rs186021206 | G | A | 0.0063 | 0.916 | 0.041 | 1.259E-112 | 499.14 |
| *CD207* | rs35533397 | C | T | 0.2263 | 0.092 | 0.009 | 2.5119E-24 | 104.49 |
| *CD207* | rs17718987 | G | C | 0.3251 | 0.473 | 0.008 | 1E-200 | 3495.77 |
| *CD207* | rs7432658 | T | C | 0.7887 | -0.07 | 0.008 | 6.3096E-17 | 76.56 |
| *CD207* | rs3129843 | A | G | 0.1294 | -0.181 | 0.01 | 1.5849E-70 | 327.61 |
| *CD207* | rs62408218 | C | T | 0.3599 | -0.102 | 0.007 | 1.9953E-46 | 212.33 |
| *CD207* | rs989437 | A | G | 0.6176 | 0.077 | 0.007 | 7.9433E-28 | 121.00 |
| *CD207* | rs2721926 | T | G | 0.5581 | -0.082 | 0.007 | 7.9433E-33 | 137.22 |
| *CD207* | rs17450261 | T | C | 0.2004 | 0.061 | 0.008 | 6.3096E-13 | 58.14 |
| *CD207* | rs79755767 | G | A | 0.099 | 0.08 | 0.012 | 3.9811E-12 | 44.44 |
| *CD207* | rs530409268 | C | CA | 0.4848 | -0.125 | 0.007 | 6.3096E-75 | 318.88 |
| *CD207* | rs34593439 | G | A | 0.1064 | -0.091 | 0.011 | 1.9953E-16 | 68.44 |
| *CD207* | rs4985556 | C | A | 0.1214 | -0.082 | 0.01 | 2.5119E-15 | 67.24 |
| *CD207* | rs2139493 | T | C | 0.2672 | -0.053 | 0.008 | 6.3096E-12 | 43.89 |
| *CD207* | rs7247412 | T | C | 0.2662 | -0.065 | 0.008 | 2.5119E-17 | 66.02 |
| *CD209* | rs7528419 | A | G | 0.2222 | -0.102 | 0.007 | 1.5849E-47 | 212.33 |
| *CD209* | rs9677444 | T | C | 0.62 | -0.044 | 0.006 | 1.9953E-13 | 53.78 |
| *CD209* | rs505922 | T | C | 0.3169 | 0.834 | 0.008 | 1E-200 | 10868.06 |
| *CD209* | rs10769256 | C | T | 0.3998 | 0.057 | 0.006 | 5.0119E-21 | 90.25 |
| *CD209* | rs7309447 | C | A | 0.6681 | 0.042 | 0.006 | 1.5849E-11 | 49.00 |
| *CD209* | rs183130 | C | T | 0.3249 | 0.045 | 0.006 | 7.9433E-13 | 56.25 |

| | | | | | | | | |
|---|---|---|---|---|---|---|---|---|
| *CD209* | rs16950595 | G | C | 0.0949 | -0.069 | 0.01 | 1E-11 | 47.61 |
| *CD209* | rs77542162 | A | G | 0.0229 | 0.15 | 0.02 | 2.5119E-14 | 56.25 |
| *CD209* | rs4804774 | G | C | 0.2368 | -0.414 | 0.008 | 1E-200 | 2678.06 |
| *CD22* | rs4848370 | C | T | 0.2716 | -0.071 | 0.008 | 2.5119E-19 | 78.77 |
| *CD22* | rs967367 | G | A | 0.388 | -0.12 | 0.007 | 1.2589E-61 | 293.88 |
| *CD22* | rs304154 | T | C | 0.3085 | 0.053 | 0.008 | 1.9953E-12 | 43.89 |
| *CD22* | rs111672628 | A | G | 0.0764 | -0.123 | 0.013 | 1.5849E-20 | 89.52 |
| *CD22* | rs9271065 | C | G | 0.8544 | -0.247 | 0.01 | 3.162E-134 | 610.09 |
| *CD22* | rs12554596 | A | G | 0.2037 | -0.066 | 0.009 | 3.1623E-14 | 53.78 |
| *CD22* | rs12784975 | T | C | 0.192 | -0.062 | 0.009 | 3.9811E-12 | 47.46 |
| *CD22* | rs111338191 | A | AAATT | 0.5213 | -0.052 | 0.007 | 1.5849E-13 | 55.18 |
| *CD22* | rs3213566 | T | C | 0.5281 | -0.049 | 0.007 | 3.1623E-12 | 49.00 |
| *CD22* | rs76428106 | T | C | 0.0128 | 0.229 | 0.032 | 1.2589E-12 | 51.21 |
| *CD22* | rs374039502 | T | A | 0.0206 | 0.337 | 0.026 | 1E-37 | 168.00 |
| *CD22* | rs9939427 | G | A | 0.2311 | 0.074 | 0.008 | 1E-18 | 85.56 |
| *CD22* | rs186021206 | G | A | 0.0064 | 0.995 | 0.048 | 5.0119E-94 | 429.70 |
| *CD22* | rs34557412 | A | G | 0.0068 | 0.713 | 0.043 | 1.2589E-62 | 274.94 |
| *CD22* | rs35222145 | T | G | 0.4917 | -0.054 | 0.007 | 1.9953E-14 | 59.51 |
| *CD22* | rs10411704 | G | T | 0.7855 | -0.295 | 0.009 | 1E-200 | 1074.38 |
| *CD22* | rs492602 | A | G | 0.5089 | 0.062 | 0.007 | 2.5119E-18 | 78.45 |
| *CD22* | rs6065926 | A | G | 0.7536 | -0.072 | 0.008 | 1.9953E-18 | 81.00 |
| *CD22* | rs73167342 | C | G | 0.7416 | -0.061 | 0.008 | 3.9811E-14 | 58.14 |
| *CD22* | rs73165110 | G | A | 0.0787 | -0.124 | 0.013 | 2.5119E-21 | 90.98 |
| *CD226* | rs2278668 | T | C | 0.5936 | -0.062 | 0.008 | 1.2589E-15 | 60.06 |
| *CD226* | rs210135 | T | A | 0.6996 | 0.071 | 0.008 | 6.3096E-18 | 78.77 |
| *CD226* | rs778641135 | TTAC | T | 0.2576 | 0.08 | 0.009 | 3.1623E-20 | 79.01 |
| *CD226* | rs6993770 | A | T | 0.2866 | -0.102 | 0.008 | 6.3096E-35 | 162.56 |
| *CD226* | rs72850966 | C | T | 0.0865 | -0.197 | 0.013 | 2.5119E-49 | 229.64 |
| *CD226* | rs3184504 | T | C | 0.5171 | -0.07 | 0.007 | 6.3096E-21 | 100.00 |
| *CD226* | rs9635249 | A | G | 0.7941 | 0.109 | 0.009 | 1.9953E-31 | 146.68 |
| *CD226* | rs151234 | G | C | 0.1288 | 0.079 | 0.011 | 1.5849E-12 | 51.58 |
| *CD226* | rs13381239 | T | A | 0.4717 | -0.264 | 0.008 | 1E-200 | 1089.00 |
| *CD226* | rs1654425 | T | C | 0.8339 | 0.101 | 0.01 | 1.2589E-23 | 102.01 |
| *CD244* | rs37456 | C | T | 0.3526 | 0.071 | 0.008 | 2.5119E-20 | 78.77 |
| *CD244* | rs115868021 | C | G | 0.0057 | -2.164 | 0.049 | 1E-200 | 1950.39 |
| *CD244* | rs12616639 | A | G | 0.6107 | 0.11 | 0.007 | 1.5849E-50 | 246.94 |
| *CD244* | rs2009581 | G | A | 0.2724 | -0.089 | 0.008 | 7.9433E-28 | 123.77 |
| *CD244* | rs34458031 | G | A | 0.0856 | 0.09 | 0.013 | 2.5119E-12 | 47.93 |
| *CD244* | rs6993770 | A | T | 0.2866 | 0.077 | 0.008 | 3.1623E-22 | 92.64 |
| *CD244* | rs10815098 | C | T | 0.2083 | 0.074 | 0.009 | 6.3096E-17 | 67.60 |
| *CD244* | rs113315674 | G | A | 0.3647 | 0.082 | 0.007 | 6.3096E-28 | 137.22 |
| *CD244* | rs774510679 | ACTTTGCC | A | 0.4125 | 0.059 | 0.007 | 1E-15 | 71.04 |
| *CD244* | rs12574844 | G | A | 0.0706 | -0.131 | 0.014 | 1.2589E-20 | 87.56 |
| *CD244* | rs79977579 | C | A | 0.0992 | 0.118 | 0.012 | 3.1623E-22 | 96.69 |
| *CD244* | rs3184504 | T | C | 0.5174 | -0.15 | 0.007 | 2.5119E-96 | 459.18 |
| *CD244* | rs10146962 | T | C | 0.3385 | -0.055 | 0.008 | 6.3096E-13 | 47.27 |
| *CD244* | rs186021206 | G | A | 0.0064 | 1.292 | 0.05 | 1.995E-149 | 667.71 |
| *CD244* | rs2659005 | C | T | 0.4413 | 0.087 | 0.007 | 3.9811E-33 | 154.47 |
| *CD248* | rs333947 | G | A | 0.1482 | -0.069 | 0.01 | 6.3096E-12 | 47.61 |
| *CD248* | rs4040185 | T | A | 0.1387 | 0.1 | 0.011 | 2.5119E-20 | 82.64 |
| *CD248* | rs1260326 | T | C | 0.6083 | 0.057 | 0.007 | 7.9433E-15 | 66.31 |
| *CD248* | rs67409736 | G | T | 0.5581 | 0.101 | 0.007 | 5.0119E-44 | 208.18 |
| *CD248* | rs78526588 | T | G | 0.0881 | -0.109 | 0.013 | 6.3096E-18 | 70.30 |
| *CD248* | rs73080552 | G | A | 0.1103 | -0.087 | 0.011 | 1.9953E-14 | 62.55 |
| *CD248* | rs6913309 | T | A | 0.28 | 0.056 | 0.008 | 1.9953E-12 | 49.00 |
| *CD248* | rs11766489 | T | C | 0.5173 | 0.065 | 0.007 | 1.2589E-19 | 86.22 |
| *CD248* | rs11764584 | G | A | 0.5552 | 0.083 | 0.007 | 5.0119E-31 | 140.59 |
| *CD248* | rs17657502 | C | T | 0.1798 | 0.068 | 0.009 | 2.5119E-13 | 57.09 |
| *CD248* | rs182547559 | C | G | 0.0044 | -0.763 | 0.056 | 2.5119E-42 | 185.64 |
| *CD248* | rs3751198 | A | G | 0.6032 | 0.081 | 0.007 | 7.9433E-28 | 133.90 |
| *CD27* | rs61747728 | C | T | 0.0379 | 0.137 | 0.019 | 3.1623E-13 | 51.99 |
| *CD27* | rs4722557 | G | A | 0.2592 | -0.063 | 0.008 | 3.9811E-14 | 62.02 |
| *CD27* | rs25680 | G | A | 0.2395 | 0.294 | 0.009 | 1E-200 | 1067.11 |
| *CD27* | rs7137828 | C | T | 0.5162 | -0.074 | 0.007 | 3.9811E-24 | 111.76 |
| *CD27* | rs150861794 | C | T | 0.0176 | 0.21 | 0.029 | 1E-12 | 52.44 |
| *CD27* | rs34562254 | G | A | 0.0972 | 0.1 | 0.012 | 1.2589E-16 | 69.44 |
| *CD274* | rs4657041 | T | C | 0.5357 | 0.061 | 0.007 | 6.3096E-17 | 75.94 |
| *CD274* | rs78444298 | G | A | 0.0193 | 0.196 | 0.026 | 1E-13 | 56.83 |

| | | | | | | | | |
|---|---|---|---|---|---|---|---|---|
| *CD274* | rs6993770 | A | T | 0.2865 | -0.061 | 0.008 | 2.5119E-14 | 58.14 |
| *CD274* | rs822340 | A | G | 0.7451 | 0.341 | 0.009 | 1E-200 | 1435.57 |
| *CD274* | rs111338191 | A | AAATT | 0.5209 | -0.085 | 0.007 | 3.1623E-31 | 147.45 |
| *CD274* | rs112635299 | G | T | 0.0217 | 0.216 | 0.025 | 1.9953E-18 | 74.65 |
| *CD274* | rs8178824 | C | T | 0.0299 | 0.151 | 0.021 | 1.2589E-12 | 51.70 |
| *CD276* | rs4665972 | T | C | 0.6086 | 0.055 | 0.006 | 3.1623E-18 | 84.03 |
| *CD276* | rs1042704 | G | A | 0.2154 | 0.058 | 0.007 | 7.9433E-15 | 68.65 |
| *CD276* | rs2239651 | T | C | 0.2649 | -0.072 | 0.007 | 7.9433E-25 | 105.80 |
| *CD276* | rs2291014 | G | A | 0.0981 | -1.337 | 0.012 | 1E-200 | 12413.67 |
| *CD276* | rs186021206 | G | A | 0.0064 | 0.414 | 0.042 | 1E-22 | 97.16 |
| *CD276* | rs2837988 | C | A | 0.3622 | 0.068 | 0.006 | 7.9433E-26 | 128.44 |
| *CD28* | rs37459 | A | G | 0.3532 | 0.11 | 0.008 | 5.0119E-46 | 189.06 |
| *CD28* | rs13406632 | G | C | 0.1676 | -0.261 | 0.01 | 6.31E-147 | 681.21 |
| *CD28* | rs9838308 | T | C | 0.3653 | -0.115 | 0.008 | 1E-48 | 206.64 |
| *CD28* | rs6796 | T | C | 0.2788 | 0.059 | 0.008 | 1E-12 | 54.39 |
| *CD28* | rs80318131 | CT | C | 0.3054 | 0.108 | 0.008 | 3.9811E-37 | 182.25 |
| *CD28* | rs1461729 | A | G | 0.8974 | -0.09 | 0.012 | 1.9953E-13 | 56.25 |
| *CD28* | rs782455289 | TAAGAC | T | 0.3162 | 0.079 | 0.008 | 3.1623E-23 | 97.52 |
| *CD28* | rs2293579 | G | A | 0.3861 | -0.055 | 0.008 | 5.0119E-13 | 47.27 |
| *CD28* | rs78689694 | G | C | 0.1326 | -0.096 | 0.011 | 3.9811E-18 | 76.17 |
| *CD28* | rs35350651 | A | AC | 0.5035 | -0.093 | 0.007 | 3.9811E-36 | 176.51 |
| *CD28* | rs71126747 | C | CA | 0.6679 | -0.063 | 0.008 | 7.9433E-15 | 62.02 |
| *CD28* | rs7161799 | C | T | 0.0764 | 0.131 | 0.014 | 1.5849E-20 | 87.56 |
| *CD28* | rs186021206 | G | A | 0.0063 | 1.362 | 0.052 | 2.512E-152 | 686.04 |
| *CD28* | rs2725405 | G | C | 0.4408 | 0.065 | 0.008 | 5.0119E-18 | 66.02 |
| *CD2AP* | rs1354034 | T | C | 0.6037 | 0.063 | 0.007 | 6.3096E-18 | 81.00 |
| *CD2AP* | rs1385742 | A | T | 0.6446 | -0.286 | 0.008 | 1E-200 | 1278.06 |
| *CD300A* | rs11888817 | T | C | 0.643 | 0.107 | 0.007 | 2.5119E-47 | 233.65 |
| *CD300A* | rs113238004 | GT | G | 0.085 | 0.109 | 0.013 | 1.9953E-17 | 70.30 |
| *CD300A* | rs6532770 | C | A | 0.2439 | 0.059 | 0.008 | 1.2589E-12 | 54.39 |
| *CD300A* | rs13107325 | C | T | 0.075 | 0.214 | 0.014 | 3.9811E-56 | 233.65 |
| *CD300A* | rs509227 | A | C | 0.5227 | 0.051 | 0.007 | 7.9433E-13 | 53.08 |
| *CD300A* | rs950802 | G | A | 0.3061 | 0.065 | 0.008 | 2.5119E-17 | 66.02 |
| *CD300A* | rs774868556 | A | ACA | 0.3064 | -0.468 | 0.009 | 1E-200 | 2704.00 |
| *CD300A* | rs35350651 | A | AC | 0.5031 | -0.072 | 0.007 | 1E-21 | 105.80 |
| *CD300A* | rs186021206 | G | A | 0.0064 | 0.87 | 0.049 | 3.9811E-69 | 315.24 |
| *CD300A* | rs4794824 | G | A | 0.3914 | 0.06 | 0.007 | 6.3096E-16 | 73.47 |
| *CD300A* | rs62087202 | G | A | 0.0485 | -0.623 | 0.017 | 1E-200 | 1343.01 |
| *CD300C* | rs11888817 | T | C | 0.642 | 0.12 | 0.007 | 1.5849E-63 | 293.88 |
| *CD300C* | rs79935131 | G | T | 0.0856 | 0.144 | 0.012 | 1E-31 | 144.00 |
| *CD300C* | rs13107325 | C | T | 0.0747 | 0.143 | 0.013 | 7.9433E-28 | 121.00 |
| *CD300C* | rs509227 | A | C | 0.5227 | 0.065 | 0.007 | 1.9953E-21 | 86.22 |
| *CD300C* | rs17843620 | A | G | 0.4236 | 0.05 | 0.007 | 7.9433E-12 | 51.02 |
| *CD300C* | rs4413619 | T | C | 0.8928 | -0.078 | 0.011 | 1.9953E-12 | 50.28 |
| *CD300C* | rs143110592 | A | T | 0.1951 | -0.062 | 0.009 | 7.9433E-13 | 47.46 |
| *CD300C* | rs635634 | C | T | 0.1845 | 0.06 | 0.009 | 1.5849E-11 | 44.44 |
| *CD300C* | rs181897168 | G | A | 0.0101 | 0.257 | 0.037 | 1.9953E-12 | 48.25 |
| *CD300C* | rs774868556 | A | ACA | 0.3067 | -0.356 | 0.008 | 1E-200 | 1980.25 |
| *CD300C* | rs35350651 | A | AC | 0.5038 | -0.072 | 0.007 | 2.5119E-24 | 105.80 |
| *CD300C* | rs10849818 | A | T | 0.554 | -0.05 | 0.007 | 1.5849E-12 | 51.02 |
| *CD300C* | rs186021206 | G | A | 0.0064 | 1.07 | 0.05 | 1.995E-102 | 457.96 |
| *CD300C* | rs58440339 | C | T | 0.0924 | 0.682 | 0.013 | 1E-200 | 2752.21 |
| *CD300C* | rs2659005 | C | T | 0.4413 | 0.058 | 0.007 | 1.5849E-15 | 68.65 |
| *CD300E* | rs748544288 | CAAG | C | 0.2274 | -0.065 | 0.008 | 2.5119E-15 | 66.02 |
| *CD300E* | rs111344982 | T | A | 0.7338 | 0.075 | 0.009 | 1.5849E-17 | 69.44 |
| *CD300E* | rs3778752 | G | T | 0.4596 | 0.08 | 0.007 | 1.9953E-30 | 130.61 |
| *CD300E* | rs3184504 | T | C | 0.5167 | -0.096 | 0.007 | 3.1623E-44 | 188.08 |
| *CD300E* | rs76428106 | T | C | 0.0129 | 0.299 | 0.032 | 3.1623E-21 | 87.31 |
| *CD300E* | rs1987102 | T | C | 0.2603 | -0.06 | 0.008 | 1.2589E-14 | 56.25 |
| *CD300E* | rs1719465 | T | C | 0.6871 | -0.361 | 0.008 | 1E-200 | 2036.27 |
| *CD300E* | rs4760 | A | G | 0.156 | 0.133 | 0.009 | 7.9433E-45 | 218.38 |
| *CD300LF* | rs12667978 | C | T | 0.5715 | -0.039 | 0.005 | 3.9811E-14 | 60.84 |
| *CD300LF* | rs3809111 | C | T | 0.0639 | -0.084 | 0.011 | 1.2589E-15 | 58.31 |
| *CD300LF* | rs3184504 | T | C | 0.5166 | -0.064 | 0.005 | 3.9811E-35 | 163.84 |
| *CD300LF* | rs4997081 | G | C | 0.1925 | -0.051 | 0.007 | 1.5849E-14 | 53.08 |
| *CD300LF* | rs2241244 | G | C | 0.5366 | -0.06 | 0.007 | 1.5849E-15 | 73.47 |
| *CD300LF* | rs35489971 | A | G | 0.8123 | 1.295 | 0.01 | 1E-200 | 16770.25 |
| *CD300LG* | rs61747728 | C | T | 0.0379 | 0.145 | 0.018 | 6.3096E-16 | 64.89 |

| | | | | | | | | |
|---|---|---|---|---|---|---|---|---|
| *CD300LG* | rs78058190 | G | A | 0.0515 | -0.134 | 0.018 | 2.5119E-14 | 55.42 |
| *CD300LG* | rs7647481 | G | A | 0.1216 | 0.078 | 0.011 | 1.2589E-13 | 50.28 |
| *CD300LG* | rs13107325 | C | T | 0.0748 | 0.094 | 0.013 | 7.9433E-13 | 52.28 |
| *CD300LG* | rs11967262 | C | G | 0.4856 | -0.087 | 0.007 | 7.9433E-37 | 154.47 |
| *CD300LG* | rs28588745 | A | T | 0.2066 | -0.107 | 0.009 | 1.9953E-36 | 141.35 |
| *CD300LG* | rs10901252 | G | C | 0.0613 | 0.231 | 0.014 | 2.5119E-58 | 272.25 |
| *CD300LG* | rs2068888 | G | A | 0.4502 | 0.051 | 0.007 | 1.9953E-13 | 53.08 |
| *CD300LG* | rs71468663 | A | AC | 0.0466 | -0.135 | 0.016 | 1.5849E-16 | 71.19 |
| *CD300LG* | rs1716403 | T | C | 0.6798 | -0.053 | 0.007 | 7.9433E-13 | 57.33 |
| *CD300LG* | rs174418 | T | C | 0.5981 | -0.058 | 0.007 | 2.5119E-16 | 68.65 |
| *CD300LG* | rs777834943 | C | CT | 0.183 | -0.07 | 0.009 | 1E-14 | 60.49 |
| *CD300LG* | rs12446515 | C | T | 0.3239 | 0.062 | 0.007 | 5.0119E-17 | 78.45 |
| *CD300LG* | rs72836561 | C | T | 0.0318 | -1.072 | 0.02 | 1E-200 | 2872.96 |
| *CD302* | rs2556106 | G | T | 0.8727 | -0.419 | 0.011 | 1E-200 | 1450.92 |
| *CD302* | rs10073754 | A | G | 0.125 | 0.099 | 0.011 | 1E-20 | 81.00 |
| *CD302* | rs9260603 | C | T | 0.4411 | -0.096 | 0.007 | 2.5119E-42 | 188.08 |
| *CD302* | rs782134971 | GAAACTGC( | G | 0.2543 | -0.074 | 0.008 | 5.0119E-20 | 85.56 |
| *CD302* | rs7970695 | G | A | 0.6237 | -0.068 | 0.007 | 7.9433E-21 | 94.37 |
| *CD302* | rs9302635 | T | C | 0.1824 | -0.065 | 0.009 | 6.3096E-13 | 52.16 |
| *CD302* | rs8178824 | C | T | 0.03 | 0.3 | 0.021 | 1E-47 | 204.08 |
| *CD302* | rs11662351 | T | C | 0.3909 | 0.162 | 0.007 | 1.995E-110 | 535.59 |
| *CD33* | rs62165726 | C | A | 0.037 | -0.101 | 0.01 | 2.5119E-24 | 102.01 |
| *CD33* | rs113238004 | GT | G | 0.0856 | 0.074 | 0.007 | 1.9953E-28 | 111.76 |
| *CD33* | rs7865362 | C | T | 0.3632 | 0.029 | 0.004 | 1E-13 | 52.56 |
| *CD33* | rs10998781 | A | G | 0.4439 | 0.049 | 0.004 | 1E-38 | 150.06 |
| *CD33* | rs139130389 | C | CTA | 0.0726 | -0.05 | 0.007 | 3.9811E-12 | 51.02 |
| *CD33* | rs4055121 | C | T | 0.1344 | -0.037 | 0.006 | 1.5849E-11 | 38.03 |
| *CD33* | rs186021206 | G | A | 0.0064 | 0.633 | 0.026 | 1.585E-133 | 592.74 |
| *CD33* | rs2455069 | A | G | 0.4272 | 1.177 | 0.008 | 1E-200 | 21645.77 |
| *CD34* | rs6671850 | C | T | 0.064 | 0.123 | 0.015 | 3.9811E-16 | 67.24 |
| *CD34* | rs532436 | G | A | 0.1851 | -0.461 | 0.01 | 1E-200 | 2125.21 |
| *CD34* | rs34434834 | G | A | 0.0333 | 0.219 | 0.02 | 7.9433E-27 | 119.90 |
| *CD34* | rs186021206 | G | A | 0.0064 | 0.721 | 0.05 | 1E-46 | 207.94 |
| *CD36* | rs3811444 | C | T | 0.3324 | -0.055 | 0.008 | 1.2589E-12 | 47.27 |
| *CD36* | rs7618405 | C | A | 0.2051 | -0.177 | 0.009 | 3.1623E-83 | 386.78 |
| *CD36* | rs62248908 | A | G | 0.174 | -0.071 | 0.01 | 3.9811E-13 | 50.41 |
| *CD36* | rs1354034 | T | C | 0.6039 | -0.062 | 0.008 | 2.5119E-16 | 60.06 |
| *CD36* | rs139596721 | C | CT | 0.0326 | 0.144 | 0.021 | 5.0119E-12 | 47.02 |
| *CD36* | rs4705938 | T | C | 0.4777 | -0.054 | 0.007 | 1.9953E-13 | 59.51 |
| *CD36* | rs511515 | A | G | 0.6986 | 0.056 | 0.008 | 1.9953E-12 | 49.00 |
| *CD36* | rs6927569 | T | C | 0.5217 | 0.057 | 0.007 | 7.9433E-15 | 66.31 |
| *CD36* | rs778641135 | TTAC | T | 0.2576 | 0.061 | 0.008 | 7.9433E-13 | 58.14 |
| *CD36* | rs10224886 | T | A | 0.2515 | -0.105 | 0.009 | 3.9811E-34 | 136.11 |
| *CD36* | rs6961069 | C | T | 0.4031 | 0.273 | 0.008 | 1E-200 | 1164.52 |
| *CD36* | rs139141690 | G | A | 0.0049 | 0.494 | 0.055 | 1.9953E-19 | 80.67 |
| *CD36* | rs1293303 | G | C | 0.4461 | 0.063 | 0.007 | 3.1623E-17 | 81.00 |
| *CD36* | rs6993770 | A | T | 0.2866 | -0.209 | 0.008 | 2.512E-145 | 682.52 |
| *CD36* | rs10820606 | A | C | 0.2293 | -0.065 | 0.009 | 1.5849E-13 | 52.16 |
| *CD36* | rs7080386 | C | A | 0.4124 | 0.091 | 0.007 | 5.0119E-34 | 169.00 |
| *CD36* | rs116052829 | C | T | 0.1027 | 0.086 | 0.012 | 1E-12 | 51.36 |
| *CD36* | rs34377578 | A | C | 0.2526 | 0.064 | 0.008 | 6.3096E-14 | 64.00 |
| *CD36* | rs113473196 | A | G | 0.087 | -0.195 | 0.013 | 7.9433E-51 | 225.00 |
| *CD36* | rs73000929 | G | A | 0.0374 | -0.142 | 0.019 | 2.5119E-13 | 55.86 |
| *CD36* | rs780615646 | AC | A | 0.7829 | -0.077 | 0.009 | 6.3096E-17 | 73.20 |
| *CD36* | rs1362213 | C | T | 0.4796 | -0.052 | 0.007 | 5.0119E-12 | 55.18 |
| *CD36* | rs4759076 | T | C | 0.457 | 0.072 | 0.007 | 3.1623E-22 | 105.80 |
| *CD36* | rs2588833 | A | C | 0.8361 | 0.069 | 0.01 | 3.9811E-12 | 47.61 |
| *CD36* | rs12719735 | G | T | 0.7109 | -0.068 | 0.008 | 5.0119E-17 | 72.25 |
| *CD36* | rs1654425 | T | C | 0.8339 | 0.095 | 0.01 | 7.9433E-22 | 90.25 |
| *CD36* | rs111527738 | A | G | 0.0208 | 0.217 | 0.026 | 3.9811E-17 | 69.66 |
| *CD38* | rs6449182 | C | G | 0.224 | -0.423 | 0.007 | 1E-200 | 3651.61 |
| *CD38* | rs7137828 | C | T | 0.5157 | -0.058 | 0.006 | 1.5849E-23 | 93.44 |
| *CD4* | rs147599819 | G | GGAT | 0.398 | -0.122 | 0.008 | 3.1623E-53 | 232.56 |
| *CD4* | rs964184 | G | C | 0.8668 | -0.095 | 0.011 | 3.9811E-19 | 74.59 |
| *CD4* | rs73053728 | A | G | 0.3023 | -0.359 | 0.008 | 1E-200 | 2013.77 |
| *CD4* | rs3184504 | T | C | 0.5174 | -0.088 | 0.007 | 3.9811E-33 | 158.04 |
| *CD4* | rs72654473 | C | A | 0.1095 | 0.096 | 0.012 | 1E-16 | 64.00 |
| *CD40* | rs1354034 | T | C | 0.604 | 0.09 | 0.007 | 2.5119E-34 | 165.31 |

| | | | | | | | | |
|---|---|---|---|---|---|---|---|---|
| *CD40* | rs342293 | C | G | 0.4592 | -0.067 | 0.007 | 5.0119E-20 | 91.61 |
| *CD40* | rs6993770 | A | T | 0.2866 | -0.076 | 0.008 | 3.1623E-21 | 90.25 |
| *CD40* | rs7896518 | A | G | 0.4239 | 0.074 | 0.007 | 1.5849E-23 | 111.76 |
| *CD40* | rs11603942 | C | T | 0.2469 | 0.059 | 0.008 | 3.1623E-12 | 54.39 |
| *CD40* | rs4608137 | G | A | 0.4873 | -0.052 | 0.007 | 7.9433E-13 | 55.18 |
| *CD40* | rs3184504 | T | C | 0.5172 | -0.054 | 0.007 | 5.0119E-14 | 59.51 |
| *CD40* | rs892090 | T | G | 0.8341 | 0.074 | 0.01 | 3.9811E-14 | 54.76 |
| *CD40* | rs4810485 | T | G | 0.7518 | 0.424 | 0.009 | 1E-200 | 2219.46 |
| *CD40LG* | rs13084580 | C | T | 0.1138 | 0.087 | 0.012 | 1.2589E-13 | 52.56 |
| *CD40LG* | rs1354034 | T | C | 0.6041 | -0.069 | 0.008 | 1.9953E-19 | 74.39 |
| *CD40LG* | rs755492124 | TAAAG | T | 0.3938 | -0.063 | 0.008 | 2.5119E-16 | 62.02 |
| *CD40LG* | rs139596721 | C | CT | 0.0322 | 0.147 | 0.021 | 6.3096E-12 | 49.00 |
| *CD40LG* | rs2248116 | C | A | 0.5711 | 0.057 | 0.008 | 6.3096E-14 | 50.77 |
| *CD40LG* | rs6961069 | C | T | 0.4021 | 0.052 | 0.008 | 1E-11 | 42.25 |
| *CD40LG* | rs6993770 | A | T | 0.2866 | -0.131 | 0.008 | 2.5119E-56 | 268.14 |
| *CD40LG* | rs151234 | G | C | 0.129 | 0.076 | 0.011 | 1E-11 | 47.74 |
| *CD40LG* | rs12445050 | C | T | 0.1382 | 0.091 | 0.011 | 3.9811E-17 | 68.44 |
| *CD40LG* | rs1654425 | T | C | 0.8343 | 0.136 | 0.01 | 5.0119E-41 | 184.96 |
| *CD40LG* | rs6081565 | G | A | 0.3526 | 0.058 | 0.008 | 1.2589E-13 | 52.56 |
| *CD40LG* | rs4239702 | T | C | 0.7223 | -0.116 | 0.008 | 1.9953E-43 | 210.25 |
| *CD46* | rs61747728 | C | T | 0.0382 | 0.16 | 0.019 | 1.9953E-16 | 70.91 |
| *CD46* | rs1970530 | G | A | 0.5947 | -0.089 | 0.008 | 3.1623E-31 | 123.77 |
| *CD46* | rs6993770 | A | T | 0.2864 | -0.066 | 0.008 | 1E-15 | 68.06 |
| *CD46* | rs579459 | T | C | 0.2074 | -0.063 | 0.009 | 7.9433E-12 | 49.00 |
| *CD46* | rs10740131 | A | T | 0.4704 | 0.06 | 0.008 | 1E-15 | 56.25 |
| *CD46* | rs972570 | C | T | 0.4874 | 0.079 | 0.008 | 1E-25 | 97.52 |
| *CD46* | rs545303153 | G | GGGGGCA | 0.003 | 0.608 | 0.079 | 1.5849E-14 | 59.23 |
| *CD46* | rs1654425 | T | C | 0.8346 | 0.073 | 0.01 | 7.9433E-13 | 53.29 |
| *CD48* | rs2295615 | C | G | 0.1372 | -0.933 | 0.011 | 1E-200 | 7194.12 |
| *CD48* | rs6705658 | C | T | 0.6419 | 0.06 | 0.007 | 1.2589E-18 | 73.47 |
| *CD48* | rs9822939 | T | C | 0.366 | -0.075 | 0.007 | 1.9953E-27 | 114.80 |
| *CD48* | rs4012172 | C | T | 0.3684 | -0.055 | 0.007 | 6.3096E-16 | 61.73 |
| *CD48* | rs35467801 | G | GT | 0.272 | -0.05 | 0.007 | 1.2589E-11 | 51.02 |
| *CD48* | rs2844622 | A | G | 0.6593 | 0.072 | 0.007 | 3.9811E-25 | 105.80 |
| *CD48* | rs60843925 | T | C | 0.1359 | -0.084 | 0.01 | 3.9811E-18 | 70.56 |
| *CD48* | rs3184504 | T | C | 0.5174 | -0.138 | 0.007 | 5.0119E-98 | 388.65 |
| *CD48* | rs2393716 | C | G | 0.4754 | -0.063 | 0.007 | 2.5119E-21 | 81.00 |
| *CD48* | rs76428106 | T | C | 0.0129 | 0.258 | 0.03 | 1.2589E-17 | 73.96 |
| *CD48* | rs186021206 | G | A | 0.0064 | 1.178 | 0.045 | 3.981E-149 | 685.28 |
| *CD48* | rs2659005 | C | T | 0.4407 | 0.098 | 0.007 | 1.9953E-49 | 196.00 |
| *CD48* | rs78530667 | T | C | 0.1286 | -0.072 | 0.01 | 1E-12 | 51.84 |
| *CD5* | rs10214237 | T | C | 0.2734 | -0.077 | 0.008 | 7.9433E-21 | 92.64 |
| *CD5* | rs952579 | G | A | 0.1448 | 0.071 | 0.01 | 5.0119E-12 | 50.41 |
| *CD5* | rs2844541 | T | A | 0.7732 | -0.074 | 0.009 | 3.9811E-17 | 67.60 |
| *CD5* | rs4939490 | C | G | 0.3914 | -0.136 | 0.008 | 2.5119E-73 | 289.00 |
| *CD5* | rs3184504 | T | C | 0.5166 | -0.148 | 0.007 | 5.0119E-92 | 447.02 |
| *CD5* | rs7161799 | C | T | 0.0766 | 0.138 | 0.014 | 1.9953E-23 | 97.16 |
| *CD5* | rs7208422 | A | T | 0.4774 | -0.05 | 0.007 | 5.0119E-12 | 51.02 |
| *CD5* | rs4893 | A | G | 0.0396 | -0.13 | 0.019 | 3.9811E-12 | 46.81 |
| *CD55* | rs1583090 | T | C | 0.2231 | -0.487 | 0.009 | 1E-200 | 2928.01 |
| *CD55* | rs13107325 | C | T | 0.0748 | 0.135 | 0.014 | 1.2589E-22 | 92.98 |
| *CD55* | rs767298 | G | T | 0.6341 | 0.061 | 0.008 | 7.9433E-16 | 58.14 |
| *CD55* | rs2142306 | T | C | 0.4119 | 0.074 | 0.007 | 7.9433E-24 | 111.76 |
| *CD55* | rs774510679 | ACTTTGCC | A | 0.412 | 0.054 | 0.007 | 3.1623E-13 | 59.51 |
| *CD55* | rs186021206 | G | A | 0.0063 | 1.07 | 0.05 | 3.981E-101 | 457.96 |
| *CD55* | rs3814995 | C | T | 0.3151 | 0.057 | 0.008 | 1.2589E-13 | 50.77 |
| *CD58* | rs10801908 | C | T | 0.1202 | -0.258 | 0.011 | 2.512E-115 | 550.12 |
| *CD58* | rs3213563 | T | C | 0.559 | -0.178 | 0.007 | 2.512E-128 | 646.61 |
| *CD58* | rs1260326 | T | C | 0.6086 | 0.056 | 0.007 | 1E-13 | 64.00 |
| *CD58* | rs10935473 | G | T | 0.4398 | -0.062 | 0.007 | 2.5119E-17 | 78.45 |
| *CD58* | rs13064814 | A | G | 0.7142 | -0.064 | 0.008 | 1.5849E-15 | 64.00 |
| *CD58* | rs199922514 | A | AAGAAGAA | 0.9077 | -0.111 | 0.013 | 1E-18 | 72.91 |
| *CD58* | rs10640079 | G | GT | 0.4706 | 0.052 | 0.007 | 1E-12 | 55.18 |
| *CD58* | rs3967200 | C | T | 0.1345 | -0.103 | 0.011 | 5.0119E-22 | 87.68 |
| *CD58* | rs186021206 | G | A | 0.0064 | 1.295 | 0.05 | 6.31E-147 | 670.81 |
| *CD58* | rs2659005 | C | T | 0.4406 | 0.054 | 0.007 | 2.5119E-13 | 59.51 |
| *CD58* | rs4760 | A | G | 0.1561 | 0.085 | 0.01 | 2.5119E-17 | 72.25 |
| *CD59* | rs34331968 | T | C | 0.4664 | -0.07 | 0.007 | 1.9953E-23 | 100.00 |

| | | | | | | | | |
|---|---|---|---|---|---|---|---|---|
| *CD59* | rs1398018 | T | C | 0.4429 | 0.05 | 0.007 | 1.2589E-12 | 51.02 |
| *CD59* | rs704701 | T | G | 0.3208 | -0.246 | 0.008 | 1E-200 | 945.56 |
| *CD59* | rs77924615 | G | A | 0.1963 | -0.071 | 0.009 | 1E-15 | 62.23 |
| *CD59* | rs407307 | A | G | 0.6787 | 0.053 | 0.008 | 2.5119E-12 | 43.89 |
| *CD5L* | rs188468174 | C | T | 0.0141 | 0.441 | 0.032 | 1.5849E-43 | 189.92 |
| *CD5L* | rs1925678 | T | C | 0.6659 | 0.058 | 0.008 | 1E-13 | 52.56 |
| *CD5L* | rs2476601 | A | G | 0.8993 | 0.083 | 0.012 | 1.5849E-11 | 47.84 |
| *CD5L* | rs2765501 | G | A | 0.4015 | 0.2 | 0.008 | 3.162E-152 | 625.00 |
| *CD5L* | rs3766370 | C | T | 0.3286 | -0.07 | 0.008 | 1E-18 | 76.56 |
| *CD5L* | rs17031793 | C | T | 0.1609 | -0.069 | 0.01 | 3.9811E-12 | 47.61 |
| *CD5L* | rs1898846 | T | C | 0.8584 | -0.075 | 0.01 | 6.3096E-13 | 56.25 |
| *CD5L* | rs617119 | C | T | 0.5129 | 0.066 | 0.007 | 1.2589E-19 | 88.90 |
| *CD5L* | rs9307257 | A | G | 0.4978 | 0.061 | 0.007 | 1E-16 | 75.94 |
| *CD5L* | rs6891054 | A | G | 0.2402 | -0.061 | 0.009 | 1.5849E-12 | 45.94 |
| *CD5L* | rs116446171 | C | G | 0.0209 | 0.187 | 0.026 | 5.0119E-13 | 51.73 |
| *CD5L* | rs1264457 | G | A | 0.6065 | -0.107 | 0.008 | 2.5119E-45 | 178.89 |
| *CD5L* | rs114373434 | C | G | 0.2732 | -0.062 | 0.008 | 1E-13 | 60.06 |
| *CD5L* | rs112348182 | G | A | 0.1142 | 0.085 | 0.012 | 1.5849E-13 | 50.17 |
| *CD5L* | rs4840568 | G | A | 0.268 | 0.057 | 0.008 | 6.3096E-12 | 50.77 |
| *CD5L* | rs2436887 | C | T | 0.2644 | -0.087 | 0.008 | 7.9433E-26 | 118.27 |
| *CD5L* | rs7903926 | G | C | 0.4937 | -0.05 | 0.007 | 1.5849E-11 | 51.02 |
| *CD5L* | rs9658720 | T | TG | 0.5538 | -0.069 | 0.007 | 2.5119E-20 | 97.16 |
| *CD5L* | rs35923643 | A | G | 0.1965 | 0.089 | 0.009 | 7.9433E-22 | 97.79 |
| *CD5L* | rs117972357 | G | A | 0.0026 | 0.534 | 0.073 | 3.1623E-13 | 53.51 |
| *CD5L* | rs12880641 | T | G | 0.6608 | -0.056 | 0.008 | 6.3096E-13 | 49.00 |
| *CD5L* | rs200123836 | C | T | 0.2718 | -0.056 | 0.008 | 1.5849E-11 | 49.00 |
| *CD5L* | rs6497185 | C | A | 0.7263 | 0.063 | 0.008 | 1.5849E-14 | 62.02 |
| *CD5L* | rs12924236 | G | C | 0.2413 | 0.059 | 0.009 | 3.1623E-12 | 42.98 |
| *CD5L* | rs71374033 | A | AT | 0.7424 | -0.089 | 0.008 | 2.5119E-26 | 123.77 |
| *CD5L* | rs9901675 | G | A | 0.0501 | 0.147 | 0.017 | 1.2589E-18 | 74.77 |
| *CD5L* | rs34557412 | A | G | 0.0069 | -0.301 | 0.044 | 1.2589E-11 | 46.80 |
| *CD5L* | rs6037651 | C | T | 0.5841 | 0.094 | 0.007 | 1.5849E-36 | 180.33 |
| *CD6* | rs7578575 | T | A | 0.2903 | 0.048 | 0.007 | 1E-12 | 47.02 |
| *CD6* | rs4672497 | C | G | 0.2217 | 0.054 | 0.007 | 2.5119E-13 | 59.51 |
| *CD6* | rs62246406 | G | A | 0.1757 | -0.054 | 0.008 | 1.5849E-11 | 45.56 |
| *CD6* | rs62454712 | T | C | 0.2426 | 0.049 | 0.007 | 7.9433E-12 | 49.00 |
| *CD6* | rs1483573 | A | G | 0.7488 | -0.048 | 0.007 | 6.3096E-12 | 47.02 |
| *CD6* | rs11230563 | C | T | 0.3477 | -0.81 | 0.008 | 1E-200 | 10251.56 |
| *CD6* | rs3184504 | T | C | 0.5173 | -0.105 | 0.006 | 3.9811E-67 | 306.25 |
| *CD6* | rs7161799 | C | T | 0.0767 | 0.1 | 0.011 | 1.5849E-18 | 82.64 |
| *CD6* | rs71374020 | A | AT | 0.299 | -0.05 | 0.007 | 1E-12 | 51.02 |
| *CD6* | rs200489612 | G | A | 0.0052 | 0.522 | 0.046 | 1.9953E-30 | 128.77 |
| *CD6* | rs7208422 | A | T | 0.4777 | -0.076 | 0.006 | 1.5849E-35 | 160.44 |
| *CD6* | rs28413174 | C | T | 0.1233 | 0.11 | 0.009 | 1.5849E-32 | 149.38 |
| *CD63* | rs2278668 | T | C | 0.5937 | -0.06 | 0.008 | 5.0119E-15 | 56.25 |
| *CD63* | rs78217698 | G | C | 0.0334 | -0.143 | 0.021 | 1.2589E-11 | 46.37 |
| *CD63* | rs755492124 | TAAAG | T | 0.3938 | -0.072 | 0.008 | 6.3096E-21 | 81.00 |
| *CD63* | rs342299 | C | T | 0.4545 | -0.052 | 0.008 | 3.9811E-12 | 42.25 |
| *CD63* | rs6993770 | A | T | 0.2865 | -0.174 | 0.008 | 6.3096E-97 | 473.06 |
| *CD63* | rs10761756 | C | T | 0.4698 | 0.054 | 0.008 | 5.0119E-13 | 45.56 |
| *CD63* | rs71474568 | T | TTG | 0.3402 | 0.085 | 0.008 | 1.2589E-26 | 112.89 |
| *CD63* | rs187372065 | C | G | 0.0029 | -0.717 | 0.075 | 1.5849E-21 | 91.39 |
| *CD63* | rs12445050 | C | T | 0.1382 | 0.082 | 0.011 | 3.9811E-14 | 55.57 |
| *CD63* | rs35496032 | G | A | 0.0136 | 0.363 | 0.033 | 6.3096E-29 | 121.00 |
| *CD63* | rs892090 | T | G | 0.8341 | 0.144 | 0.01 | 1E-45 | 207.36 |
| *CD63* | rs4813637 | G | A | 0.5853 | -0.111 | 0.008 | 7.9433E-48 | 192.52 |
| *CD69* | rs1354034 | T | C | 0.6039 | 0.088 | 0.008 | 5.0119E-30 | 121.00 |
| *CD69* | rs3804749 | C | T | 0.5927 | -0.054 | 0.008 | 6.3096E-12 | 45.56 |
| *CD69* | rs7758845 | A | C | 0.272 | 0.058 | 0.008 | 7.9433E-12 | 52.56 |
| *CD69* | rs6993770 | A | T | 0.2867 | -0.124 | 0.008 | 1.5849E-49 | 240.25 |
| *CD69* | rs7896518 | A | G | 0.4233 | 0.093 | 0.008 | 2.5119E-33 | 135.14 |
| *CD69* | rs12819552 | A | G | 0.4219 | -0.055 | 0.008 | 5.0119E-13 | 47.27 |
| *CD7* | rs2548993 | G | A | 0.7224 | 0.061 | 0.008 | 3.1623E-14 | 58.14 |
| *CD7* | rs2516501 | C | T | 0.2628 | -0.057 | 0.008 | 2.5119E-12 | 50.77 |
| *CD7* | rs11105298 | T | C | 0.2419 | -0.131 | 0.008 | 5.0119E-54 | 268.14 |
| *CD7* | rs3184504 | T | C | 0.5171 | -0.119 | 0.007 | 5.0119E-62 | 289.00 |
| *CD7* | rs3176831 | C | T | 0.0428 | -1.132 | 0.019 | 1E-200 | 3549.65 |
| *CD70* | rs17118876 | C | G | 0.57 | -0.039 | 0.006 | 3.1623E-12 | 42.25 |

| | | | | | | | | |
|---|---|---|---|---|---|---|---|---|
| *CD70* | rs12740374 | G | T | 0.2215 | -0.303 | 0.007 | 1E-200 | 1873.65 |
| *CD70* | rs57894427 | T | C | 0.0934 | 0.068 | 0.009 | 1.9953E-13 | 57.09 |
| *CD70* | rs7254538 | G | C | 0.543 | 0.75 | 0.007 | 1E-200 | 11479.59 |
| *CD72* | rs4848370 | C | T | 0.2708 | -0.06 | 0.008 | 7.9433E-13 | 56.25 |
| *CD72* | rs56364901 | C | A | 0.0766 | -0.107 | 0.014 | 1.9953E-14 | 58.41 |
| *CD72* | rs9269573 | T | A | 0.8541 | -0.151 | 0.011 | 3.9811E-46 | 188.44 |
| *CD72* | rs869487 | C | G | 0.2199 | -0.166 | 0.009 | 6.3096E-75 | 340.20 |
| *CD72* | rs3184504 | T | C | 0.5165 | -0.093 | 0.007 | 5.0119E-36 | 176.51 |
| *CD72* | rs76428106 | T | C | 0.0129 | 0.231 | 0.034 | 1.2589E-11 | 46.16 |
| *CD72* | rs374039502 | T | A | 0.0207 | 0.211 | 0.028 | 3.1623E-14 | 56.79 |
| *CD72* | rs34557412 | A | G | 0.0069 | 0.42 | 0.045 | 7.9433E-21 | 87.11 |
| *CD72* | rs35222145 | T | G | 0.4927 | -0.06 | 0.008 | 1.9953E-15 | 56.25 |
| *CD74* | rs79078220 | G | A | 0.1103 | -0.087 | 0.011 | 6.3096E-15 | 62.55 |
| *CD74* | rs9273368 | G | A | 0.3044 | 0.177 | 0.008 | 1.995E-115 | 489.52 |
| *CD74* | rs3184504 | T | C | 0.5171 | -0.126 | 0.007 | 1.9953E-72 | 324.00 |
| *CD74* | rs7194862 | T | C | 0.2898 | 0.138 | 0.008 | 1E-70 | 297.56 |
| *CD74* | rs186021206 | G | A | 0.0064 | 0.386 | 0.048 | 1E-15 | 64.67 |
| *CD74* | rs4969145 | T | C | 0.5828 | -0.049 | 0.007 | 1.2589E-11 | 49.00 |
| *CD74* | rs3827211 | A | G | 0.4521 | -0.05 | 0.007 | 3.1623E-12 | 51.02 |
| *CD79B* | rs61747728 | C | T | 0.038 | 0.131 | 0.019 | 5.0119E-12 | 47.54 |
| *CD79B* | rs78444298 | G | A | 0.0194 | 0.308 | 0.027 | 2.5119E-31 | 130.13 |
| *CD79B* | rs112151347 | G | A | 0.0271 | -0.209 | 0.023 | 3.1623E-19 | 82.57 |
| *CD79B* | rs77397204 | A | C | 0.3855 | -0.066 | 0.008 | 1.5849E-18 | 68.06 |
| *CD79B* | rs78777785 | A | T | 0.0763 | -0.118 | 0.014 | 6.3096E-18 | 71.04 |
| *CD79B* | rs55654737 | G | A | 0.3433 | -0.085 | 0.008 | 1.5849E-28 | 112.89 |
| *CD79B* | rs2523583 | G | A | 0.7054 | -0.054 | 0.008 | 7.9433E-12 | 45.56 |
| *CD79B* | rs2395904 | T | G | 0.2744 | -0.057 | 0.008 | 1.5849E-12 | 50.77 |
| *CD79B* | rs507666 | G | A | 0.1846 | -0.246 | 0.009 | 5.012E-148 | 747.11 |
| *CD79B* | rs7087507 | A | G | 0.3488 | 0.059 | 0.008 | 5.0119E-15 | 54.39 |
| *CD79B* | rs111338191 | A | AAATT | 0.5213 | -0.07 | 0.007 | 1.5849E-21 | 100.00 |
| *CD79B* | rs76428106 | T | C | 0.0128 | 0.261 | 0.033 | 5.0119E-15 | 62.55 |
| *CD79B* | rs13333054 | C | T | 0.2275 | 0.067 | 0.009 | 1.2589E-14 | 55.42 |
| *CD79B* | rs186021206 | G | A | 0.0064 | 0.797 | 0.05 | 1.2589E-56 | 254.08 |
| *CD79B* | rs12946510 | C | T | 0.4761 | -0.059 | 0.007 | 7.9433E-16 | 71.04 |
| *CD79B* | rs12946669 | C | T | 0.3942 | -0.163 | 0.007 | 3.162E-107 | 542.22 |
| *CD79B* | rs7213540 | C | T | 0.5394 | -0.055 | 0.007 | 5.0119E-14 | 61.73 |
| *CD80* | rs547373038 | G | T | 0.4934 | -0.053 | 0.007 | 1.5849E-12 | 57.33 |
| *CD80* | rs56078908 | A | T | 0.106 | 0.605 | 0.012 | 1E-200 | 2541.84 |
| *CD80* | rs3936289 | T | C | 0.2382 | 0.068 | 0.008 | 1E-15 | 72.25 |
| *CD80* | rs9270747 | A | G | 0.658 | -0.086 | 0.007 | 1E-31 | 150.94 |
| *CD80* | rs75071241 | G | A | 0.0596 | -0.12 | 0.015 | 1.2589E-16 | 64.00 |
| *CD80* | rs3184504 | T | C | 0.517 | -0.116 | 0.007 | 5.0119E-64 | 274.61 |
| *CD80* | rs7137504 | C | T | 0.471 | -0.049 | 0.007 | 1E-12 | 49.00 |
| *CD80* | rs186021206 | G | A | 0.0064 | 1.069 | 0.047 | 1.585E-112 | 517.32 |
| *CD80* | rs738409 | C | G | 0.2172 | 0.073 | 0.008 | 1E-18 | 83.27 |
| *CD82* | rs34118068 | G | A | 0.1052 | 0.11 | 0.013 | 7.9433E-18 | 71.60 |
| *CD83* | rs35332487 | C | A | 0.3047 | -0.057 | 0.008 | 7.9433E-13 | 50.77 |
| *CD83* | rs12526086 | G | A | 0.2225 | 0.2 | 0.009 | 1.585E-113 | 493.83 |
| *CD83* | rs17843620 | A | G | 0.4235 | 0.062 | 0.008 | 3.1623E-15 | 60.06 |
| *CD83* | rs7801921 | C | G | 0.4578 | 0.05 | 0.007 | 1E-11 | 51.02 |
| *CD83* | rs8176644 | C | T | 0.0598 | 0.129 | 0.015 | 3.1623E-17 | 73.96 |
| *CD83* | rs7869 | C | T | 0.2913 | 0.1 | 0.008 | 1.5849E-35 | 156.25 |
| *CD83* | rs4753126 | C | T | 0.501 | 0.051 | 0.007 | 2.5119E-12 | 53.08 |
| *CD83* | rs3184504 | T | C | 0.5174 | -0.112 | 0.007 | 6.3096E-54 | 256.00 |
| *CD83* | rs534297892 | CA | C | 0.5028 | -0.072 | 0.007 | 7.9433E-22 | 105.80 |
| *CD83* | rs76428106 | T | C | 0.013 | 0.354 | 0.033 | 2.5119E-26 | 115.07 |
| *CD83* | rs186021206 | G | A | 0.0064 | 0.447 | 0.05 | 5.0119E-19 | 79.92 |
| *CD83* | rs2150879 | G | A | 0.5533 | 0.05 | 0.007 | 7.9433E-12 | 51.02 |
| *CD83* | rs34878396 | C | T | 0.0126 | 0.302 | 0.033 | 1.9953E-20 | 83.75 |
| *CD84* | rs11265440 | A | G | 0.918 | -0.206 | 0.014 | 1.5849E-51 | 216.51 |
| *CD84* | rs41315846 | T | C | 0.4767 | -0.055 | 0.008 | 2.5119E-13 | 47.27 |
| *CD84* | rs1354034 | T | C | 0.6043 | 0.084 | 0.008 | 2.5119E-28 | 110.25 |
| *CD84* | rs114694170 | T | C | 0.0598 | 0.132 | 0.016 | 1.2589E-16 | 68.06 |
| *CD84* | rs778641135 | TTAC | T | 0.2575 | 0.074 | 0.009 | 7.9433E-18 | 67.60 |
| *CD84* | rs6993770 | A | T | 0.2869 | -0.078 | 0.008 | 3.9811E-21 | 95.06 |
| *CD84* | rs409801 | T | C | 0.5072 | 0.066 | 0.007 | 6.3096E-19 | 88.90 |
| *CD84* | rs7080386 | C | A | 0.4121 | 0.079 | 0.008 | 1.9953E-25 | 97.52 |
| *CD84* | rs597808 | A | G | 0.5159 | -0.082 | 0.007 | 6.3096E-28 | 137.22 |

| | | | | | | | | |
|---|---|---|---|---|---|---|---|---|
| *CD84* | rs10131298 | T | A | 0.2455 | 0.215 | 0.009 | 6.31E-133 | 570.68 |
| *CD84* | rs151165225 | C | T | 0.2551 | -0.094 | 0.009 | 6.3096E-28 | 109.09 |
| *CD86* | rs535373070 | A | AC | 0.3118 | 0.065 | 0.008 | 3.9811E-15 | 66.02 |
| *CD86* | rs139795227 | A | C | 0.0149 | -0.341 | 0.031 | 3.9811E-28 | 121.00 |
| *CD86* | rs151088139 | C | T | 0.145 | -0.092 | 0.01 | 5.0119E-19 | 84.64 |
| *CD86* | rs13063578 | T | A | 0.3981 | -0.084 | 0.007 | 5.0119E-29 | 144.00 |
| *CD86* | rs192337168 | G | T | 0.0025 | -1.369 | 0.074 | 5.0119E-76 | 342.25 |
| *CD86* | rs6796 | T | C | 0.279 | 0.056 | 0.008 | 5.0119E-12 | 49.00 |
| *CD86* | rs199922514 | A | AAGAAGAA | 0.9073 | -0.097 | 0.013 | 1.5849E-14 | 55.67 |
| *CD86* | rs635634 | C | T | 0.1841 | 0.073 | 0.009 | 1.2589E-14 | 65.79 |
| *CD86* | rs60194243 | C | T | 0.0591 | -0.283 | 0.015 | 2.5119E-74 | 355.95 |
| *CD86* | rs3184504 | T | C | 0.5172 | -0.125 | 0.007 | 1.5849E-66 | 318.88 |
| *CD86* | rs76428106 | T | C | 0.0129 | 0.528 | 0.033 | 3.1623E-56 | 256.00 |
| *CD86* | rs186021206 | G | A | 0.0065 | 1.22 | 0.05 | 1E-131 | 595.36 |
| *CD8A* | rs3020726 | A | G | 0.161 | 0.362 | 0.01 | 1E-200 | 1310.44 |
| *CD8A* | rs35467801 | G | GT | 0.2718 | -0.069 | 0.008 | 1.5849E-17 | 74.39 |
| *CD8A* | rs12211087 | T | A | 0.0921 | 0.472 | 0.013 | 1E-200 | 1318.25 |
| *CD8A* | rs12360863 | C | T | 0.2505 | -0.057 | 0.008 | 7.9433E-12 | 50.77 |
| *CD8A* | rs3184504 | T | C | 0.5171 | -0.064 | 0.007 | 3.1623E-19 | 83.59 |
| *CD93* | rs61747728 | C | T | 0.0385 | 0.177 | 0.02 | 2.5119E-19 | 78.32 |
| *CD93* | rs1260326 | T | C | 0.6081 | 0.06 | 0.008 | 1.2589E-14 | 56.25 |
| *CD93* | rs9275180 | C | T | 0.4726 | -0.052 | 0.008 | 6.3096E-12 | 42.25 |
| *CD93* | rs7821812 | G | C | 0.2081 | -0.081 | 0.009 | 5.0119E-18 | 81.00 |
| *CD93* | rs10901252 | G | C | 0.0613 | 0.14 | 0.016 | 1.5849E-18 | 76.56 |
| *CD99* | rs1260326 | T | C | 0.6084 | 0.057 | 0.007 | 7.9433E-18 | 66.31 |
| *CD99* | rs12149545 | G | A | 0.3164 | 0.049 | 0.007 | 1.2589E-12 | 49.00 |
| *CD99* | rs704 | G | A | 0.4732 | 0.062 | 0.006 | 5.0119E-22 | 106.78 |
| *CD99* | rs673408 | A | G | 0.4558 | -0.107 | 0.006 | 6.3096E-61 | 318.03 |
| *CD99L2* | rs4665710 | A | C | 0.7934 | -0.066 | 0.009 | 7.9433E-13 | 53.78 |
| *CD99L2* | rs139039156 | G | A | 0.0085 | -0.294 | 0.042 | 3.9811E-12 | 49.00 |
| *CD99L2* | rs6054 | C | T | 0.0059 | 0.387 | 0.05 | 1.5849E-14 | 59.91 |
| *CD99L2* | rs73015965 | A | G | 0.0065 | 0.349 | 0.05 | 3.9811E-12 | 48.72 |
| *CD99L2* | rs896173 | G | A | 0.4222 | -0.058 | 0.008 | 1.9953E-14 | 52.56 |
| *CD99L2* | rs11438680 | G | GA | 0.461 | 0.053 | 0.008 | 1.5849E-12 | 43.89 |
| *CD99L2* | rs174536 | A | C | 0.3484 | 0.123 | 0.008 | 1.9953E-55 | 236.39 |
| *CD99L2* | rs1801689 | A | C | 0.0301 | 0.481 | 0.022 | 1.585E-106 | 478.02 |
| *CD99L2* | rs1065853 | G | T | 0.0815 | 0.132 | 0.014 | 7.9433E-22 | 88.90 |
| *CDA* | rs2072671 | A | C | 0.3435 | -0.489 | 0.008 | 1E-200 | 3736.27 |
| *CDA* | rs6762442 | C | G | 0.6763 | 0.052 | 0.007 | 1.9953E-12 | 55.18 |
| *CDA* | rs10770 | T | C | 0.1189 | -0.112 | 0.011 | 1.5849E-26 | 103.67 |
| *CDA* | rs11775560 | A | G | 0.1861 | 0.112 | 0.009 | 1E-36 | 154.86 |
| *CDA* | rs11993233 | A | G | 0.426 | 0.149 | 0.007 | 2.512E-101 | 453.08 |
| *CDA* | rs34433840 | C | T | 0.2396 | -0.066 | 0.008 | 1.2589E-16 | 68.06 |
| *CDA* | rs61744929 | T | C | 0.0189 | 0.172 | 0.025 | 6.3096E-12 | 47.33 |
| *CDA* | rs9303283 | C | T | 0.564 | -0.064 | 0.007 | 1.9953E-20 | 83.59 |
| *CDA* | rs4807399 | C | T | 0.5335 | 0.054 | 0.007 | 6.3096E-15 | 59.51 |
| *CDAN1* | rs575220 | A | G | 0.5397 | 0.055 | 0.008 | 7.9433E-12 | 47.27 |
| *CDC26* | rs1354034 | T | C | 0.6047 | 0.068 | 0.008 | 2.5119E-19 | 72.25 |
| *CDC26* | rs7916868 | A | T | 0.5016 | 0.051 | 0.007 | 1E-11 | 53.08 |
| *CDC26* | rs11502185 | T | C | 0.2597 | 0.068 | 0.01 | 1.5849E-12 | 46.24 |
| *CDC27* | rs79702286 | A | G | 0.1018 | 0.136 | 0.013 | 2.5119E-27 | 109.44 |
| *CDC37* | rs1354034 | T | C | 0.6041 | 0.059 | 0.008 | 5.0119E-15 | 54.39 |
| *CDC37* | rs342298 | C | T | 0.4549 | -0.061 | 0.008 | 3.9811E-16 | 58.14 |
| *CDC37* | rs10733789 | T | C | 0.3132 | 0.055 | 0.008 | 1.2589E-11 | 47.27 |
| *CDC42BPB* | rs1354034 | T | C | 0.6046 | 0.068 | 0.008 | 1.2589E-19 | 72.25 |
| *CDC42BPB* | rs342293 | C | G | 0.4598 | -0.05 | 0.007 | 1.2589E-11 | 51.02 |
| *CDCP1* | rs2276862 | C | G | 0.1718 | -0.303 | 0.009 | 1E-200 | 1133.44 |
| *CDCP1* | rs113787140 | T | A | 0.5037 | -0.084 | 0.006 | 2.5119E-39 | 196.00 |
| *CDCP1* | rs77029323 | G | T | 0.4041 | 0.055 | 0.007 | 1E-16 | 61.73 |
| *CDCP1* | rs683486 | C | A | 0.4026 | -0.127 | 0.007 | 3.1623E-80 | 329.16 |
| *CDCP1* | rs11048473 | C | T | 0.2686 | 0.061 | 0.007 | 5.0119E-17 | 75.94 |
| *CDCP1* | rs35350651 | A | AC | 0.5036 | -0.047 | 0.006 | 1.9953E-13 | 61.36 |
| *CDCP1* | rs7485656 | A | G | 0.14 | 0.083 | 0.009 | 3.9811E-19 | 85.05 |
| *CDCP1* | rs186021206 | G | A | 0.0064 | 0.561 | 0.044 | 3.9811E-37 | 162.56 |
| *CDCP1* | rs738409 | C | G | 0.2171 | 0.064 | 0.008 | 2.5119E-16 | 64.00 |
| *CDH1* | rs532086 | T | C | 0.162 | 0.066 | 0.01 | 1.2589E-11 | 43.56 |
| *CDH1* | rs2519093 | C | T | 0.1844 | -0.352 | 0.009 | 1E-200 | 1529.68 |
| *CDH1* | rs10749609 | G | A | 0.7943 | 0.068 | 0.009 | 1.2589E-14 | 57.09 |

| | | | | | | | | |
|---|---|---|---|---|---|---|---|---|
| *CDH1* | rs186021206 | G | A | 0.0064 | 0.439 | 0.049 | 3.9811E-19 | 80.27 |
| *CDH1* | rs708686 | C | T | 0.2683 | 0.097 | 0.008 | 3.9811E-30 | 147.02 |
| *CDH1* | rs3814995 | C | T | 0.315 | 0.059 | 0.008 | 1.5849E-13 | 54.39 |
| *CDH1* | rs681343 | C | T | 0.5104 | -0.433 | 0.008 | 1E-200 | 2929.52 |
| *CDH15* | rs10923344 | G | A | 0.6181 | 0.049 | 0.006 | 7.9433E-15 | 66.69 |
| *CDH15* | rs11646135 | G | A | 0.1415 | 0.438 | 0.009 | 1E-200 | 2368.44 |
| *CDH15* | rs2240715 | G | C | 0.2994 | 0.062 | 0.007 | 1.9953E-20 | 78.45 |
| *CDH17* | rs12084158 | A | G | 0.0982 | 0.093 | 0.012 | 1.9953E-15 | 60.06 |
| *CDH17* | rs533137864 | C | CA | 0.145 | -0.1 | 0.009 | 1.2589E-26 | 123.46 |
| *CDH17* | rs12479244 | A | T | 0.2878 | 0.146 | 0.007 | 3.1623E-93 | 435.02 |
| *CDH17* | rs9815073 | C | A | 0.3428 | -0.061 | 0.007 | 1.2589E-17 | 75.94 |
| *CDH17* | rs3128921 | C | A | 0.2586 | 0.091 | 0.007 | 1.2589E-34 | 169.00 |
| *CDH17* | rs4556017 | C | T | 0.8536 | -0.12 | 0.009 | 6.3096E-39 | 177.78 |
| *CDH17* | rs763526069 | AC | A | 0.4742 | -0.274 | 0.007 | 1E-200 | 1532.16 |
| *CDH17* | rs550057 | C | T | 0.2553 | -0.46 | 0.008 | 1E-200 | 3306.25 |
| *CDH17* | rs12801491 | G | A | 0.1954 | 0.061 | 0.008 | 7.9433E-14 | 58.14 |
| *CDH17* | rs2280381 | C | T | 0.6144 | -0.051 | 0.007 | 2.5119E-14 | 53.08 |
| *CDH17* | rs186021206 | G | A | 0.0064 | 0.543 | 0.044 | 1.9953E-34 | 152.30 |
| *CDH17* | rs708686 | C | T | 0.2677 | 0.154 | 0.008 | 7.9433E-90 | 370.56 |
| *CDH17* | rs492602 | A | G | 0.5092 | -0.382 | 0.007 | 1E-200 | 2978.04 |
| *CDH2* | rs772351164 | GTTT | G | 0.0789 | -0.112 | 0.013 | 6.3096E-17 | 74.22 |
| *CDH2* | rs4074793 | A | G | 0.0737 | 0.119 | 0.013 | 2.5119E-19 | 83.79 |
| *CDH2* | rs62383006 | A | G | 0.0541 | -0.187 | 0.015 | 2.5119E-34 | 155.42 |
| *CDH2* | rs112875651 | G | A | 0.391 | -0.059 | 0.007 | 2.5119E-16 | 71.04 |
| *CDH2* | rs181242111 | G | A | 0.1179 | 0.089 | 0.011 | 1.2589E-14 | 65.46 |
| *CDH2* | rs9633740 | A | G | 0.7959 | 0.107 | 0.009 | 1.2589E-35 | 141.35 |
| *CDH2* | rs7108216 | T | C | 0.5984 | 0.049 | 0.007 | 3.9811E-12 | 49.00 |
| *CDH2* | rs11045820 | C | T | 0.1608 | -0.064 | 0.009 | 1E-11 | 50.57 |
| *CDH2* | rs28929474 | C | T | 0.0208 | 0.281 | 0.024 | 3.1623E-31 | 137.09 |
| *CDH2* | rs3785307 | T | C | 0.1148 | -0.146 | 0.011 | 7.9433E-41 | 176.17 |
| *CDH2* | rs56192742 | A | G | 0.237 | 0.087 | 0.008 | 7.9433E-27 | 118.27 |
| *CDH2* | rs12454164 | G | A | 0.311 | -0.101 | 0.007 | 5.0119E-42 | 208.18 |
| *CDH23* | rs2166631 | C | T | 0.2141 | -0.168 | 0.009 | 5.0119E-72 | 348.44 |
| *CDH3* | rs12137270 | C | T | 0.2447 | 0.066 | 0.009 | 1.9953E-14 | 53.78 |
| *CDH3* | rs61747728 | C | T | 0.0379 | 0.141 | 0.019 | 3.1623E-13 | 55.07 |
| *CDH3* | rs1260326 | T | C | 0.6085 | 0.058 | 0.008 | 1.9953E-14 | 52.56 |
| *CDH3* | rs4934189 | C | T | 0.5362 | 0.09 | 0.007 | 1.5849E-33 | 165.31 |
| *CDH3* | rs28640218 | G | T | 0.1914 | -0.071 | 0.009 | 5.0119E-14 | 62.23 |
| *CDH3* | rs138190335 | T | C | 0.0039 | 0.736 | 0.059 | 7.9433E-36 | 155.62 |
| *CDH3* | rs3814995 | C | T | 0.3147 | 0.064 | 0.008 | 1.2589E-15 | 64.00 |
| *CDH5* | rs61802301 | T | C | 0.5665 | 0.052 | 0.007 | 1E-13 | 55.18 |
| *CDH5* | rs1260326 | T | C | 0.6084 | 0.06 | 0.007 | 1E-17 | 73.47 |
| *CDH5* | rs13107325 | C | T | 0.0749 | 0.127 | 0.013 | 6.3096E-23 | 95.44 |
| *CDH5* | rs10093 | C | G | 0.3765 | 0.059 | 0.007 | 3.9811E-16 | 71.04 |
| *CDH5* | rs8176743 | C | T | 0.0614 | 1.205 | 0.015 | 1E-200 | 6453.44 |
| *CDH5* | rs35166255 | G | A | 0.0338 | 0.295 | 0.019 | 1E-55 | 241.07 |
| *CDH5* | rs16956504 | T | C | 0.1044 | 0.308 | 0.011 | 7.943E-169 | 784.00 |
| *CDH5* | rs186021206 | G | A | 0.0064 | 0.582 | 0.047 | 7.9433E-36 | 153.34 |
| *CDH5* | rs3814995 | C | T | 0.3149 | 0.052 | 0.007 | 7.9433E-13 | 55.18 |
| *CDH5* | rs660745 | T | C | 0.545 | -0.051 | 0.007 | 5.0119E-14 | 53.08 |
| *CDH6* | rs4626350 | G | A | 0.5551 | 0.51 | 0.008 | 1E-200 | 4064.06 |
| *CDH6* | rs6993770 | A | T | 0.2866 | 0.073 | 0.007 | 2.5119E-24 | 108.76 |
| *CDH6* | rs60757417 | C | G | 0.061 | 0.095 | 0.014 | 3.9811E-12 | 46.05 |
| *CDH6* | rs11553699 | A | G | 0.1356 | 0.072 | 0.01 | 5.0119E-13 | 51.84 |
| *CDH6* | rs186021206 | G | A | 0.0064 | 0.55 | 0.045 | 1E-34 | 149.38 |
| *CDH6* | rs9306336 | A | T | 0.4875 | 0.052 | 0.007 | 1.9953E-15 | 55.18 |
| *CDHR1* | rs61816761 | G | A | 0.0234 | 0.2 | 0.026 | 3.9811E-14 | 59.17 |
| *CDHR1* | rs12330170 | A | C | 0.4495 | 0.074 | 0.008 | 3.1623E-22 | 85.56 |
| *CDHR1* | rs73186030 | C | T | 0.1275 | 0.105 | 0.011 | 1.5849E-20 | 91.12 |
| *CDHR1* | rs7916271 | C | T | 0.4991 | 0.226 | 0.008 | 1.585E-187 | 798.06 |
| *CDHR2* | rs36086195 | C | T | 0.58 | 0.053 | 0.006 | 1.9953E-16 | 78.03 |
| *CDHR2* | rs4835265 | C | A | 0.156 | 0.063 | 0.009 | 6.3096E-13 | 49.00 |
| *CDHR2* | rs2963680 | C | T | 0.5021 | -0.102 | 0.006 | 6.3096E-57 | 289.00 |
| *CDHR2* | rs73226270 | A | T | 0.0693 | -0.092 | 0.013 | 3.9811E-13 | 50.08 |
| *CDHR2* | rs11621792 | C | T | 0.4521 | 0.046 | 0.006 | 1E-12 | 58.78 |
| *CDHR2* | rs28929474 | C | T | 0.0207 | 0.155 | 0.023 | 6.3096E-12 | 45.42 |
| *CDHR2* | rs151184170 | C | T | 0.2449 | 0.09 | 0.007 | 1.5849E-33 | 165.31 |
| *CDHR5* | rs5993 | A | T | 0.1612 | 0.256 | 0.009 | 1.585E-171 | 809.09 |

| | | | | | | | | |
|---|---|---|---|---|---|---|---|---|
| *CDHR5* | rs35457250 | C | T | 0.0089 | 0.305 | 0.036 | 1.2589E-17 | 71.78 |
| *CDHR5* | rs3130297 | C | T | 0.133 | -0.071 | 0.01 | 7.9433E-13 | 50.41 |
| *CDHR5* | rs10264306 | A | G | 0.1957 | 0.073 | 0.008 | 3.9811E-18 | 83.27 |
| *CDHR5* | rs12360820 | A | C | 0.8288 | 0.69 | 0.01 | 1E-200 | 4761.00 |
| *CDHR5* | rs174566 | A | G | 0.3498 | 0.056 | 0.008 | 1.2589E-12 | 49.00 |
| *CDHR5* | rs28929474 | C | T | 0.0208 | 0.231 | 0.023 | 7.9433E-23 | 100.87 |
| *CDHR5* | rs8178824 | C | T | 0.0299 | 0.257 | 0.02 | 7.9433E-39 | 165.12 |
| *CDK1* | rs10139050 | T | G | 0.7163 | -0.09 | 0.009 | 1.5849E-25 | 100.00 |
| *CDK5RAP3* | rs1354034 | T | C | 0.6043 | 0.083 | 0.008 | 2.5119E-27 | 107.64 |
| *CDK5RAP3* | rs9897552 | G | A | 0.1766 | 0.086 | 0.01 | 2.5119E-18 | 73.96 |
| *CDKN1A* | rs1354034 | T | C | 0.6046 | -0.081 | 0.008 | 7.9433E-27 | 102.52 |
| *CDKN1A* | rs3176345 | G | C | 0.0647 | -0.268 | 0.015 | 7.9433E-70 | 319.22 |
| *CDKN1A* | rs139141690 | G | A | 0.0048 | -0.418 | 0.055 | 3.1623E-14 | 57.76 |
| *CDKN1A* | rs6993770 | A | T | 0.2864 | -0.066 | 0.008 | 1.2589E-15 | 68.06 |
| *CDKN1A* | rs7080386 | C | A | 0.4117 | 0.055 | 0.008 | 3.1623E-13 | 47.27 |
| *CDKN1A* | rs964184 | G | C | 0.8668 | -0.078 | 0.011 | 1.2589E-12 | 50.28 |
| *CDKN1A* | rs58345103 | G | A | 0.1875 | -0.077 | 0.01 | 1E-15 | 59.29 |
| *CDKN2D* | rs1354034 | T | C | 0.6046 | 0.109 | 0.008 | 3.9811E-46 | 185.64 |
| *CDKN2D* | rs17622656 | G | A | 0.3887 | -0.056 | 0.008 | 6.3096E-13 | 49.00 |
| *CDKN2D* | rs342298 | C | T | 0.4545 | -0.051 | 0.008 | 1.5849E-11 | 40.64 |
| *CDKN2D* | rs6993770 | A | T | 0.2864 | -0.084 | 0.008 | 7.9433E-24 | 110.25 |
| *CDKN2D* | rs544626513 | G | C | 0.229 | 0.067 | 0.009 | 6.3096E-13 | 55.42 |
| *CDNF* | rs61738953 | C | G | 0.0348 | -1.129 | 0.021 | 1E-200 | 2890.34 |
| *CDON* | rs607980 | G | A | 0.9051 | -0.181 | 0.013 | 5.0119E-47 | 193.85 |
| *CDON* | rs215224 | T | C | 0.401 | 0.052 | 0.007 | 3.9811E-12 | 55.18 |
| *CDON* | rs186021206 | G | A | 0.0064 | 0.631 | 0.05 | 1.9953E-36 | 159.26 |
| *CDSN* | rs72674184 | T | C | 0.0791 | -0.092 | 0.012 | 7.9433E-15 | 58.78 |
| *CDSN* | rs61816761 | G | A | 0.0236 | 0.554 | 0.022 | 1E-135 | 634.12 |
| *CDSN* | rs7545538 | C | G | 0.4265 | -0.046 | 0.007 | 1.9953E-12 | 43.18 |
| *CDSN* | rs1436662 | A | G | 0.5936 | 0.071 | 0.006 | 3.9811E-28 | 140.03 |
| *CDSN* | rs75278193 | A | C | 0.188 | -0.07 | 0.008 | 6.3096E-18 | 76.56 |
| *CDSN* | rs3095306 | T | A | 0.61 | 0.644 | 0.008 | 1E-200 | 6480.25 |
| *CDSN* | rs7815687 | C | T | 0.1632 | 0.086 | 0.009 | 2.5119E-23 | 91.31 |
| *CDSN* | rs3758294 | T | C | 0.2036 | -0.054 | 0.008 | 7.9433E-12 | 45.56 |
| *CDSN* | rs17658212 | C | T | 0.0599 | -0.268 | 0.014 | 5.0119E-85 | 366.45 |
| *CDSN* | rs77924615 | G | A | 0.1966 | -0.055 | 0.008 | 6.3096E-12 | 47.27 |
| *CDSN* | rs562177063 | C | T | 0.0031 | -0.864 | 0.062 | 1E-44 | 194.20 |
| *CDSN* | rs567144736 | G | A | 0.0023 | -0.513 | 0.076 | 1.5849E-11 | 45.56 |
| *CDSN* | rs561579427 | A | G | 0.0021 | -0.532 | 0.075 | 1.5849E-12 | 50.32 |
| *CDSN* | rs35187787 | G | A | 0.0058 | -0.387 | 0.044 | 1.9953E-18 | 77.36 |
| *CDSN* | rs729988 | G | C | 0.3689 | -0.05 | 0.007 | 1.9953E-14 | 51.02 |
| *CDSN* | rs7253823 | C | T | 0.1684 | -0.185 | 0.009 | 3.162E-105 | 422.53 |
| *CDSN* | rs16981329 | G | C | 0.2274 | 0.06 | 0.008 | 3.1623E-15 | 56.25 |
| *CDSN* | rs12625565 | A | C | 0.2238 | -0.051 | 0.008 | 1.5849E-11 | 40.64 |
| *CEACAM1* | rs36086195 | C | T | 0.5795 | 0.055 | 0.007 | 2.5119E-13 | 61.73 |
| *CEACAM1* | rs114165349 | G | C | 0.0233 | 0.169 | 0.025 | 6.3096E-12 | 45.70 |
| *CEACAM1* | rs4835265 | C | A | 0.1563 | 0.076 | 0.01 | 6.3096E-14 | 57.76 |
| *CEACAM1* | rs11045856 | T | G | 0.2401 | 0.076 | 0.009 | 1.9953E-18 | 71.31 |
| *CEACAM1* | rs56956502 | G | A | 0.2475 | 0.067 | 0.009 | 7.9433E-15 | 55.42 |
| *CEACAM1* | rs10416089 | C | G | 0.0851 | -0.483 | 0.013 | 1E-200 | 1380.41 |
| *CEACAM16* | rs12747765 | C | A | 0.1582 | -0.074 | 0.009 | 5.0119E-15 | 67.60 |
| *CEACAM16* | rs1260326 | T | C | 0.6084 | 0.085 | 0.007 | 3.1623E-33 | 147.45 |
| *CEACAM16* | rs772513171 | CTTTT | C | 0.6726 | 0.05 | 0.007 | 1E-11 | 51.02 |
| *CEACAM16* | rs6460047 | T | C | 0.2085 | 0.058 | 0.008 | 6.3096E-12 | 52.56 |
| *CEACAM16* | rs28601761 | C | G | 0.419 | 0.105 | 0.007 | 7.9433E-50 | 225.00 |
| *CEACAM16* | rs2068888 | G | A | 0.4507 | -0.051 | 0.007 | 2.5119E-13 | 53.08 |
| *CEACAM16* | rs17715069 | G | C | 0.3815 | -0.335 | 0.008 | 1E-200 | 1753.52 |
| *CEACAM19* | rs12738484 | C | G | 0.2309 | -0.067 | 0.009 | 1.9953E-14 | 55.42 |
| *CEACAM19* | rs61816761 | G | A | 0.0234 | 0.205 | 0.026 | 1.9953E-15 | 62.17 |
| *CEACAM19* | rs776021 | C | A | 0.7374 | 0.084 | 0.008 | 1.5849E-23 | 110.25 |
| *CEACAM19* | rs35673728 | T | C | 0.0632 | -0.106 | 0.015 | 1.9953E-12 | 49.94 |
| *CEACAM19* | rs150985681 | G | A | 0.026 | -0.159 | 0.023 | 7.9433E-12 | 47.79 |
| *CEACAM19* | rs62119322 | G | C | 0.0294 | 0.623 | 0.022 | 1.995E-176 | 801.92 |
| *CEACAM20* | rs782134971 | GAAACTGC( | G | 0.2545 | -0.22 | 0.009 | 2.512E-139 | 597.53 |
| *CEACAM20* | rs10414398 | G | A | 0.0798 | -0.283 | 0.014 | 3.9811E-91 | 408.62 |
| *CEACAM20* | rs681343 | C | T | 0.509 | -0.161 | 0.008 | 6.3096E-98 | 405.02 |
| *CEACAM21* | rs7137828 | C | T | 0.5163 | -0.041 | 0.005 | 3.9811E-17 | 67.24 |
| *CEACAM21* | rs140838345 | A | AGAATC | 0.7771 | -1.267 | 0.009 | 1E-200 | 19818.38 |

| | | | | | | | | |
|---|---|---|---|---|---|---|---|---|
| *CEACAM3* | rs139026664 | C | A | 0.0839 | 0.187 | 0.015 | 2.5119E-37 | 155.42 |
| *CEACAM3* | rs62148538 | T | C | 0.0445 | 0.183 | 0.02 | 1.2589E-20 | 83.72 |
| *CEACAM5* | rs12523278 | A | G | 0.5019 | 0.052 | 0.007 | 1.9953E-13 | 55.18 |
| *CEACAM5* | rs60750824 | A | G | 0.1845 | -0.065 | 0.009 | 3.1623E-12 | 52.16 |
| *CEACAM5* | rs4242701 | A | G | 0.4354 | -0.057 | 0.007 | 3.9811E-15 | 66.31 |
| *CEACAM5* | rs375524395 | A | G | 0.4561 | -0.104 | 0.007 | 1.2589E-46 | 220.73 |
| *CEACAM5* | rs12799864 | C | T | 0.1958 | 0.08 | 0.009 | 2.5119E-19 | 79.01 |
| *CEACAM5* | rs7332827 | G | A | 0.7111 | 0.056 | 0.008 | 6.3096E-13 | 49.00 |
| *CEACAM5* | rs186021206 | G | A | 0.0062 | 0.341 | 0.049 | 5.0119E-12 | 48.43 |
| *CEACAM5* | rs117990138 | G | A | 0.0697 | -0.099 | 0.014 | 6.3096E-13 | 50.01 |
| *CEACAM5* | rs708686 | C | T | 0.2674 | 0.121 | 0.009 | 3.1623E-45 | 180.75 |
| *CEACAM5* | rs9621 | G | A | 0.0426 | 0.963 | 0.019 | 1E-200 | 2568.89 |
| *CEACAM5* | rs681343 | C | T | 0.5091 | 0.302 | 0.008 | 1E-200 | 1425.06 |
| *CEACAM5* | rs441810 | A | G | 0.2148 | 0.096 | 0.009 | 6.3096E-29 | 113.78 |
| *CEACAM6* | rs11127048 | G | A | 0.618 | 0.055 | 0.008 | 1.5849E-12 | 47.27 |
| *CEACAM6* | rs113542380 | G | A | 0.0737 | 0.115 | 0.014 | 7.9433E-16 | 67.47 |
| *CEACAM6* | rs12619285 | A | G | 0.2655 | -0.07 | 0.008 | 5.0119E-17 | 76.56 |
| *CEACAM6* | rs73221104 | T | C | 0.2871 | 0.057 | 0.008 | 1E-11 | 50.77 |
| *CEACAM6* | rs55734389 | C | G | 0.1393 | 0.075 | 0.011 | 1.5849E-12 | 46.49 |
| *CEACAM6* | rs73404209 | T | C | 0.0257 | 0.192 | 0.023 | 2.5119E-16 | 69.69 |
| *CEACAM6* | rs145902394 | T | TA | 0.4254 | -0.051 | 0.007 | 1E-11 | 53.08 |
| *CEACAM6* | rs11775560 | A | G | 0.186 | 0.148 | 0.009 | 5.0119E-55 | 270.42 |
| *CEACAM6* | rs375524395 | A | G | 0.4557 | -0.061 | 0.007 | 2.5119E-16 | 75.94 |
| *CEACAM6* | rs73017385 | G | A | 0.2404 | -0.133 | 0.009 | 3.9811E-53 | 218.38 |
| *CEACAM6* | rs186021206 | G | A | 0.0064 | 0.684 | 0.051 | 3.9811E-41 | 179.88 |
| *CEACAM6* | rs12453334 | C | T | 0.3665 | -0.076 | 0.008 | 2.5119E-23 | 90.25 |
| *CEACAM6* | rs199726619 | G | GAA | 0.1259 | -0.093 | 0.013 | 5.0119E-13 | 51.18 |
| *CEACAM6* | rs56378716 | A | G | 0.0135 | 0.516 | 0.032 | 6.3096E-58 | 260.02 |
| *CEACAM6* | rs12459454 | A | G | 0.2918 | -0.12 | 0.008 | 6.3096E-50 | 225.00 |
| *CEACAM6* | rs492602 | A | G | 0.5093 | 0.107 | 0.007 | 3.9811E-47 | 233.65 |
| *CEACAM6* | rs28398212 | C | T | 0.574 | 0.052 | 0.008 | 7.9433E-12 | 42.25 |
| *CEACAM6* | rs7275212 | A | T | 0.0084 | 0.512 | 0.041 | 1.2589E-36 | 155.95 |
| *CEACAM8* | rs3917932 | C | G | 0.5754 | -0.056 | 0.007 | 7.9433E-14 | 64.00 |
| *CEACAM8* | rs112694524 | G | A | 0.0703 | 0.1 | 0.015 | 1E-11 | 44.44 |
| *CEACAM8* | rs2236947 | C | A | 0.4821 | 0.086 | 0.007 | 3.9811E-31 | 150.94 |
| *CEACAM8* | rs3774298 | G | A | 0.6873 | 0.071 | 0.008 | 6.3096E-18 | 78.77 |
| *CEACAM8* | rs2738144 | A | T | 0.3348 | -0.065 | 0.008 | 7.9433E-16 | 66.02 |
| *CEACAM8* | rs35440906 | C | CA | 0.1852 | 0.209 | 0.009 | 1.995E-107 | 539.27 |
| *CEACAM8* | rs1058900 | T | C | 0.5665 | 0.058 | 0.008 | 2.5119E-14 | 52.56 |
| *CEACAM8* | rs2511241 | C | T | 0.9292 | 0.165 | 0.014 | 1.5849E-30 | 138.90 |
| *CEACAM8* | rs6503533 | C | T | 0.6171 | -0.106 | 0.008 | 3.1623E-44 | 175.56 |
| *CEACAM8* | rs114641762 | C | A | 0.1044 | -0.248 | 0.012 | 1.5849E-88 | 427.11 |
| *CEACAM8* | rs119468010 | G | A | 0.0036 | 0.454 | 0.062 | 2.5119E-13 | 53.62 |
| *CEACAM8* | rs12459454 | A | G | 0.2921 | -0.131 | 0.008 | 3.9811E-59 | 268.14 |
| *CEBPA* | rs28929474 | C | T | 0.0208 | 0.194 | 0.027 | 7.9433E-13 | 51.63 |
| *CEBPB* | rs1045929 | C | T | 0.3875 | 0.066 | 0.008 | 5.0119E-18 | 68.06 |
| *CEBPB* | rs10404947 | A | G | 0.8023 | 0.07 | 0.009 | 1E-13 | 60.49 |
| *CELA2A* | rs188524036 | T | G | 0.0116 | -0.535 | 0.035 | 6.3096E-52 | 233.65 |
| *CELA2A* | rs187351243 | G | A | 0.0213 | 0.26 | 0.026 | 1.9953E-23 | 100.00 |
| *CELA2A* | rs35004807 | T | TGTAA | 0.3444 | 0.074 | 0.008 | 3.9811E-21 | 85.56 |
| *CELA2A* | rs17032925 | T | C | 0.0824 | -0.168 | 0.013 | 1.2589E-35 | 167.01 |
| *CELA2A* | rs2842895 | G | C | 0.5515 | -0.053 | 0.007 | 1E-12 | 57.33 |
| *CELA2A* | rs9388490 | C | T | 0.4399 | -0.066 | 0.007 | 7.9433E-19 | 88.90 |
| *CELA2A* | rs13221897 | G | A | 0.5638 | -0.055 | 0.007 | 1.9953E-13 | 61.73 |
| *CELA2A* | rs13226219 | T | C | 0.1288 | -0.11 | 0.011 | 3.9811E-23 | 100.00 |
| *CELA2A* | rs77643237 | G | A | 0.0549 | 0.242 | 0.017 | 1.9953E-48 | 202.64 |
| *CELA2A* | rs77958433 | T | C | 0.6343 | -0.067 | 0.008 | 1.2589E-16 | 70.14 |
| *CELA2A* | rs10994860 | C | T | 0.1774 | -0.067 | 0.01 | 3.1623E-12 | 44.89 |
| *CELA2A* | rs4246215 | G | T | 0.36 | 0.061 | 0.008 | 1.9953E-15 | 58.14 |
| *CELA2A* | rs75593565 | A | G | 0.0252 | -0.264 | 0.024 | 2.5119E-29 | 121.00 |
| *CELA2A* | rs28929474 | C | T | 0.0208 | -0.662 | 0.026 | 3.162E-144 | 648.29 |
| *CELA2A* | rs28365941 | T | C | 0.1362 | -0.109 | 0.011 | 7.9433E-23 | 98.19 |
| *CELA2A* | rs2287796 | G | T | 0.4068 | 0.054 | 0.008 | 1E-12 | 45.56 |
| *CELA2A* | rs72802342 | C | A | 0.0758 | -0.116 | 0.014 | 5.0119E-16 | 68.65 |
| *CELA2A* | rs529900 | G | C | 0.3787 | -0.169 | 0.008 | 1.995E-109 | 446.27 |
| *CELA2A* | rs8088688 | A | T | 0.077 | 0.108 | 0.014 | 6.3096E-15 | 59.51 |
| *CELA3A* | rs34262568 | G | A | 0.231 | 0.237 | 0.009 | 3.981E-167 | 693.44 |
| *CELA3A* | rs333947 | G | A | 0.1483 | -0.076 | 0.01 | 7.9433E-14 | 57.76 |

| | | | | | | | | |
|---|---|---|---|---|---|---|---|---|
| *CELA3A* | rs17032925 | T | C | 0.0829 | -0.099 | 0.013 | 3.1623E-14 | 57.99 |
| *CELA3A* | rs2395203 | A | G | 0.2164 | -0.064 | 0.009 | 7.9433E-13 | 50.57 |
| *CELA3A* | rs6571015 | G | A | 0.7905 | 0.157 | 0.009 | 1E-70 | 304.31 |
| *CELA3A* | rs373980643 | GT | G | 0.456 | -0.076 | 0.007 | 7.9433E-26 | 117.88 |
| *CELA3A* | rs11778310 | A | G | 0.2936 | 0.072 | 0.008 | 5.0119E-20 | 81.00 |
| *CELA3A* | rs8176747 | C | G | 0.0613 | 0.5 | 0.015 | 1E-200 | 1111.11 |
| *CELA3A* | rs56278466 | T | G | 0.6615 | 0.06 | 0.008 | 1.9953E-15 | 56.25 |
| *CELA3A* | rs61910478 | A | T | 0.2686 | -0.07 | 0.008 | 2.5119E-18 | 76.56 |
| *CELA3A* | rs6575449 | C | T | 0.1848 | -0.11 | 0.009 | 3.1623E-32 | 149.38 |
| *CELA3A* | rs28365941 | T | C | 0.1361 | -0.082 | 0.011 | 3.1623E-14 | 55.57 |
| *CELA3A* | rs72802342 | C | A | 0.0755 | -0.257 | 0.014 | 6.3096E-77 | 336.98 |
| *CELA3A* | rs529900 | G | C | 0.3782 | -0.101 | 0.007 | 1E-42 | 208.18 |
| *CELA3A* | rs681343 | C | T | 0.509 | 0.197 | 0.007 | 1.995E-163 | 792.02 |
| *CELSR2* | rs12740374 | G | T | 0.221 | 0.46 | 0.009 | 1E-200 | 2612.35 |
| *CEMIP2* | rs28601761 | C | G | 0.4189 | -0.054 | 0.008 | 5.0119E-12 | 45.56 |
| *CEMIP2* | rs191912523 | T | G | 0.0833 | 0.254 | 0.014 | 1E-72 | 329.16 |
| *CEND1* | rs429358 | T | C | 0.1557 | 0.078 | 0.011 | 3.1623E-13 | 50.28 |
| *CENPF* | rs140399039 | T | C | 0.0011 | -1.096 | 0.122 | 3.1623E-19 | 80.71 |
| *CEP112* | rs12938334 | A | C | 0.4327 | -0.229 | 0.008 | 3.981E-185 | 819.39 |
| *CEP152* | rs77379665 | C | T | 0.0333 | 0.157 | 0.022 | 3.9811E-13 | 50.93 |
| *CEP170* | rs12072595 | T | G | 0.3601 | 0.07 | 0.008 | 6.3096E-19 | 76.56 |
| *CEP170* | rs1354034 | T | C | 0.6045 | 0.062 | 0.008 | 3.9811E-16 | 60.06 |
| *CEP20* | rs1354034 | T | C | 0.6038 | 0.101 | 0.008 | 1.5849E-38 | 159.39 |
| *CEP20* | rs342298 | C | T | 0.4548 | -0.057 | 0.008 | 1E-13 | 50.77 |
| *CEP20* | rs73396521 | G | A | 0.2449 | 0.063 | 0.009 | 1.2589E-12 | 49.00 |
| *CEP20* | rs74009487 | C | T | 0.0564 | -0.216 | 0.017 | 1.2589E-38 | 161.44 |
| *CEP20* | rs200309755 | C | T | 0.5373 | 0.054 | 0.008 | 1E-11 | 45.56 |
| *CEP43* | rs2501620 | A | C | 0.6501 | -0.064 | 0.008 | 3.9811E-16 | 64.00 |
| *CEP43* | rs2237276 | C | T | 0.4547 | 0.065 | 0.008 | 7.9433E-18 | 66.02 |
| *CEP43* | rs342298 | C | T | 0.4549 | -0.063 | 0.008 | 7.9433E-17 | 62.02 |
| *CEP43* | rs11170874 | A | G | 0.3308 | 0.063 | 0.008 | 7.9433E-15 | 62.02 |
| *CEP85* | rs751511889 | TAAA | T | 0.9094 | 0.132 | 0.013 | 1.9953E-22 | 103.10 |
| *CEP85* | rs1354034 | T | C | 0.6038 | 0.117 | 0.008 | 3.9811E-53 | 213.89 |
| *CEP85* | rs342293 | C | G | 0.4596 | -0.064 | 0.007 | 1.9953E-17 | 83.59 |
| *CEP85* | rs11604127 | C | T | 0.2307 | 0.062 | 0.009 | 5.0119E-12 | 47.46 |
| *CERT* | rs1354034 | T | C | 0.6035 | 0.084 | 0.008 | 2.5119E-27 | 110.25 |
| *CERT* | rs10942737 | T | C | 0.082 | 0.101 | 0.014 | 5.0119E-13 | 52.05 |
| *CERT* | rs342298 | C | T | 0.4544 | -0.054 | 0.008 | 1.5849E-12 | 45.56 |
| *CERT* | rs7297265 | A | G | 0.4432 | 0.055 | 0.008 | 1E-12 | 47.27 |
| *CES1* | rs12129540 | C | T | 0.0431 | 0.117 | 0.017 | 5.0119E-12 | 47.37 |
| *CES1* | rs1260326 | T | C | 0.6081 | -0.097 | 0.007 | 3.9811E-43 | 192.02 |
| *CES1* | rs7640956 | A | G | 0.8217 | -0.126 | 0.009 | 3.9811E-45 | 196.00 |
| *CES1* | rs17259257 | C | T | 0.0556 | 0.136 | 0.015 | 1.2589E-19 | 82.20 |
| *CES1* | rs187515714 | G | T | 0.026 | 0.147 | 0.021 | 7.9433E-12 | 49.00 |
| *CES1* | rs13240994 | T | C | 0.1963 | -0.066 | 0.009 | 1.5849E-14 | 53.78 |
| *CES1* | rs75231243 | G | A | 0.0676 | -0.135 | 0.014 | 6.3096E-23 | 92.98 |
| *CES1* | rs199922514 | A | AAGAAGAA | 0.9076 | -0.16 | 0.012 | 1E-41 | 177.78 |
| *CES1* | rs112875651 | G | A | 0.3909 | -0.059 | 0.007 | 1E-16 | 71.04 |
| *CES1* | rs72775768 | C | T | 0.2887 | -0.065 | 0.008 | 1.2589E-17 | 66.02 |
| *CES1* | rs2393969 | A | C | 0.4681 | -0.062 | 0.007 | 1.5849E-19 | 78.45 |
| *CES1* | rs10845139 | G | A | 0.0895 | -0.085 | 0.012 | 1.2589E-12 | 50.17 |
| *CES1* | rs7979473 | A | G | 0.6168 | 0.056 | 0.007 | 3.9811E-15 | 64.00 |
| *CES1* | rs143168962 | T | G | 0.0565 | -1.152 | 0.016 | 1E-200 | 5184.00 |
| *CES1* | rs58542926 | C | T | 0.0755 | 0.191 | 0.013 | 3.1623E-49 | 215.86 |
| *CES1* | rs7256200 | G | T | 0.128 | -0.098 | 0.01 | 1.5849E-21 | 96.04 |
| *CES1* | rs641738 | C | T | 0.4415 | -0.05 | 0.007 | 5.0119E-13 | 51.02 |
| *CES2* | rs140461033 | G | A | 0.0036 | -1.05 | 0.062 | 1E-64 | 286.81 |
| *CES3* | rs193084249 | A | G | 0.0234 | 0.229 | 0.025 | 2.5119E-20 | 83.91 |
| *CES3* | rs62260502 | G | A | 0.1785 | 0.133 | 0.01 | 1.9953E-43 | 176.89 |
| *CES3* | rs73015965 | A | G | 0.0064 | 0.581 | 0.05 | 1E-31 | 135.02 |
| *CES3* | rs17715183 | G | C | 0.0666 | -0.15 | 0.015 | 5.0119E-24 | 100.00 |
| *CES3* | rs9987289 | A | G | 0.908 | -0.176 | 0.013 | 1.9953E-43 | 183.29 |
| *CES3* | rs61373955 | A | ATAAGT | 0.4763 | 0.053 | 0.007 | 6.3096E-13 | 57.33 |
| *CES3* | rs7970695 | G | A | 0.6237 | 0.078 | 0.008 | 1E-24 | 95.06 |
| *CES3* | rs71647892 | T | C | 0.0127 | -0.854 | 0.033 | 2.512E-145 | 669.71 |
| *CES3* | rs11668882 | T | C | 0.4408 | -0.06 | 0.007 | 6.3096E-16 | 73.47 |
| *CETN2* | rs1354034 | T | C | 0.6035 | 0.085 | 0.008 | 1.2589E-27 | 112.89 |
| *CETN2* | rs342296 | G | A | 0.4562 | -0.065 | 0.008 | 1E-17 | 66.02 |

| | | | | | | | | |
|---|---|---|---|---|---|---|---|---|
| *CETN2* | rs117564216 | G | A | 0.0164 | -0.248 | 0.031 | 3.9811E-16 | 64.00 |
| *CETN3* | rs1354034 | T | C | 0.6046 | 0.108 | 0.008 | 2.5119E-46 | 182.25 |
| *CETN3* | rs4873 | C | G | 0.2766 | -0.173 | 0.008 | 3.9811E-93 | 467.64 |
| *CETN3* | rs958976 | T | C | 0.5884 | 0.073 | 0.008 | 7.9433E-22 | 83.27 |
| *CETN3* | rs342298 | C | T | 0.455 | -0.069 | 0.007 | 3.1623E-20 | 97.16 |
| *CETN3* | rs113315674 | G | A | 0.3648 | 0.054 | 0.008 | 3.9811E-12 | 45.56 |
| *CETN3* | rs941207 | C | G | 0.2706 | 0.071 | 0.008 | 5.0119E-17 | 78.77 |
| *CETN3* | rs5998067 | G | A | 0.707 | -0.138 | 0.008 | 1.2589E-63 | 297.56 |
| *CFB* | rs2228145 | A | C | 0.4063 | -0.049 | 0.007 | 6.3096E-12 | 49.00 |
| *CFB* | rs10801552 | T | C | 0.229 | 0.185 | 0.008 | 1.995E-109 | 534.77 |
| *CFB* | rs1260326 | T | C | 0.6082 | -0.067 | 0.007 | 7.9433E-21 | 91.61 |
| *CFB* | rs10033900 | T | C | 0.5158 | 0.05 | 0.007 | 1E-12 | 51.02 |
| *CFB* | rs641153 | G | A | 0.0964 | 0.63 | 0.013 | 1E-200 | 2348.52 |
| *CFB* | rs28601761 | C | G | 0.4197 | 0.06 | 0.007 | 2.5119E-17 | 73.47 |
| *CFB* | rs12975366 | T | C | 0.3956 | -0.049 | 0.007 | 6.3096E-12 | 49.00 |
| *CFC1* | rs11708067 | A | G | 0.2448 | 0.068 | 0.009 | 1.2589E-14 | 57.09 |
| *CFC1* | rs6790951 | C | T | 0.4791 | 0.06 | 0.008 | 3.9811E-15 | 56.25 |
| *CFC1* | rs2517546 | C | T | 0.1707 | -0.075 | 0.01 | 1E-13 | 56.25 |
| *CFC1* | rs35510806 | T | TA | 0.4714 | -0.052 | 0.008 | 7.9433E-12 | 42.25 |
| *CFC1* | rs2168101 | C | A | 0.3106 | 0.062 | 0.008 | 3.1623E-13 | 60.06 |
| *CFC1* | rs8038040 | G | A | 0.4268 | 0.069 | 0.008 | 3.1623E-19 | 74.39 |
| *CFC1* | rs28461437 | C | G | 0.5962 | -0.098 | 0.008 | 1.9953E-36 | 150.06 |
| *CFC1* | rs34931250 | C | T | 0.0606 | 0.108 | 0.016 | 1.5849E-11 | 45.56 |
| *CFD* | rs698084 | C | T | 0.1819 | 0.069 | 0.009 | 2.5119E-13 | 58.78 |
| *CFD* | rs704 | G | A | 0.4737 | -0.065 | 0.007 | 3.1623E-19 | 86.22 |
| *CFD* | rs71335276 | G | T | 0.2272 | -0.185 | 0.009 | 1.5849E-91 | 422.53 |
| *CFH* | rs731557 | C | A | 0.194 | 0.421 | 0.01 | 1E-200 | 1772.41 |
| *CFH* | rs36019311 | G | GTA | 0.1295 | -0.076 | 0.011 | 1E-11 | 47.74 |
| *CFH* | rs73201506 | A | G | 0.0455 | -0.137 | 0.018 | 2.5119E-14 | 57.93 |
| *CFHR2* | rs4085749 | C | T | 0.2271 | -1.037 | 0.009 | 1E-200 | 13276.16 |
| *CFHR2* | rs1260326 | T | C | 0.6083 | -0.031 | 0.005 | 7.9433E-12 | 38.44 |
| *CFHR4* | rs4915363 | T | C | 0.2364 | -0.793 | 0.009 | 1E-200 | 7763.57 |
| *CFHR4* | rs141880390 | G | A | 0.0029 | -0.506 | 0.043 | 5.0119E-32 | 138.47 |
| *CFHR4* | rs1461729 | A | G | 0.8973 | 0.054 | 0.008 | 3.1623E-12 | 45.56 |
| *CFHR4* | rs41274050 | C | T | 0.0099 | 0.174 | 0.023 | 1.2589E-13 | 57.23 |
| *CFHR4* | rs664034 | C | T | 0.5764 | 0.04 | 0.005 | 3.1623E-17 | 64.00 |
| *CFHR4* | rs2464190 | T | C | 0.412 | 0.035 | 0.005 | 1.9953E-13 | 49.00 |
| *CFHR4* | rs28929474 | C | T | 0.0208 | 0.113 | 0.016 | 3.9811E-12 | 49.88 |
| *CFHR4* | rs149394327 | G | C | 0.03 | 0.145 | 0.014 | 5.0119E-26 | 107.27 |
| *CFHR5* | rs165316 | A | G | 0.1982 | 0.066 | 0.01 | 3.9811E-12 | 43.56 |
| *CFHR5* | rs72732234 | T | C | 0.0173 | -1.589 | 0.03 | 1E-200 | 2805.47 |
| *CFHR5* | rs55709272 | T | C | 0.439 | 0.051 | 0.007 | 3.1623E-13 | 53.08 |
| *CFHR5* | rs28601761 | C | G | 0.4198 | -0.06 | 0.007 | 2.5119E-17 | 73.47 |
| *CFHR5* | rs10740131 | A | T | 0.47 | 0.062 | 0.007 | 6.3096E-19 | 78.45 |
| *CFHR5* | rs7979478 | A | G | 0.6157 | -0.091 | 0.007 | 1.9953E-36 | 169.00 |
| *CFHR5* | rs72731579 | A | G | 0.1648 | -0.067 | 0.01 | 3.1623E-12 | 44.89 |
| *CFI* | rs1260326 | T | C | 0.6084 | -0.093 | 0.007 | 7.9433E-36 | 176.51 |
| *CFI* | rs10033900 | T | C | 0.5163 | 0.216 | 0.007 | 1.259E-188 | 952.16 |
| *CFI* | rs204886 | T | C | 0.5062 | 0.053 | 0.007 | 1.9953E-13 | 57.33 |
| *CFP* | rs10737680 | A | C | 0.403 | 0.203 | 0.007 | 1.995E-163 | 841.00 |
| *CFP* | rs777318032 | C | T | 0.0011 | -0.974 | 0.122 | 1.2589E-15 | 63.74 |
| *CFP* | rs4151667 | T | A | 0.0461 | 0.399 | 0.017 | 6.31E-119 | 550.87 |
| *CFP* | rs4752973 | T | C | 0.1892 | -0.07 | 0.009 | 3.1623E-14 | 60.49 |
| *CFP* | rs10774625 | A | G | 0.5039 | -0.066 | 0.007 | 3.1623E-20 | 88.90 |
| *CFP* | rs186021206 | G | A | 0.0064 | 0.424 | 0.049 | 1E-17 | 74.88 |
| *CFP* | rs2230199 | G | C | 0.2165 | -0.149 | 0.009 | 3.9811E-63 | 274.09 |
| *CGB3_CGB5_CGB8* | rs16854211 | G | T | 0.1578 | 0.077 | 0.01 | 1.2589E-15 | 59.29 |
| *CGB3_CGB5_CGB8* | rs7252920 | A | C | 0.4856 | -0.061 | 0.007 | 3.1623E-18 | 75.94 |
| *CGREF1* | rs2384572 | G | T | 0.6252 | -0.561 | 0.008 | 1E-200 | 4917.52 |
| *CGREF1* | rs9273206 | T | C | 0.8351 | 0.078 | 0.009 | 5.0119E-18 | 75.11 |
| *CGREF1* | rs782134971 | GAAACTGC | G | 0.2543 | -0.246 | 0.008 | 1E-200 | 945.56 |
| *CGREF1* | rs4940867 | C | T | 0.144 | 0.079 | 0.009 | 6.3096E-17 | 77.05 |
| *CGREF1* | rs492602 | A | G | 0.5092 | -0.281 | 0.007 | 1E-200 | 1611.45 |
| *CHAC2* | rs78694551 | A | G | 0.2696 | -0.149 | 0.009 | 7.9433E-69 | 274.09 |
| *CHAD* | rs2476601 | A | G | 0.8988 | 0.084 | 0.012 | 3.1623E-12 | 49.00 |
| *CHAD* | rs1386625 | A | G | 0.9019 | -0.102 | 0.012 | 2.5119E-16 | 72.25 |
| *CHAD* | rs1485307 | T | C | 0.5423 | 0.088 | 0.007 | 2.5119E-33 | 158.04 |
| *CHAD* | rs117692263 | T | C | 0.0851 | 0.109 | 0.013 | 7.9433E-17 | 70.30 |

| | | | | | | | | |
|---|---|---|---|---|---|---|---|---|
| *CHAD* | rs9594738 | C | T | 0.4878 | 0.125 | 0.007 | 1.9953E-65 | 318.88 |
| *CHAD* | rs11625758 | T | C | 0.4953 | -0.056 | 0.007 | 1.5849E-14 | 64.00 |
| *CHAD* | rs144030994 | G | T | 0.0059 | -1.63 | 0.048 | 1E-200 | 1153.17 |
| *CHAD* | rs884205 | A | C | 0.7465 | -0.061 | 0.008 | 5.0119E-13 | 58.14 |
| *CHAD* | rs12975366 | T | C | 0.3953 | -0.097 | 0.007 | 1.5849E-38 | 192.02 |
| *CHCHD10* | rs10502 | G | C | 0.381 | -0.05 | 0.007 | 1.2589E-12 | 51.02 |
| *CHCHD10* | rs62143206 | G | T | 0.2094 | 0.132 | 0.009 | 6.3096E-54 | 215.11 |
| *CHCHD10* | rs34284791 | G | A | 0.1066 | -0.118 | 0.011 | 7.9433E-26 | 115.07 |
| *CHCHD6* | rs34723687 | G | C | 0.5077 | -0.052 | 0.007 | 2.5119E-12 | 55.18 |
| *CHCHD6* | rs9863101 | T | C | 0.1727 | -0.306 | 0.01 | 1E-200 | 936.36 |
| *CHGA* | rs4253328 | C | CTTT | 0.5129 | -0.071 | 0.007 | 5.0119E-22 | 102.88 |
| *CHGA* | rs56092644 | G | T | 0.2447 | 0.097 | 0.009 | 1.9953E-29 | 116.16 |
| *CHGA* | rs1801020 | A | G | 0.7449 | -0.069 | 0.008 | 2.5119E-16 | 74.39 |
| *CHGA* | rs17489891 | G | C | 0.1115 | -0.16 | 0.012 | 1E-42 | 177.78 |
| *CHGA* | rs11786721 | T | C | 0.4103 | 0.054 | 0.007 | 5.0119E-13 | 59.51 |
| *CHGA* | rs9581957 | C | T | 0.3278 | 0.054 | 0.008 | 5.0119E-12 | 45.56 |
| *CHGA* | rs729940 | C | T | 0.1571 | -0.246 | 0.01 | 2.512E-129 | 605.16 |
| *CHGB* | rs2509902 | T | C | 0.8574 | -0.081 | 0.01 | 3.9811E-17 | 65.61 |
| *CHGB* | rs13159918 | C | A | 0.3004 | -0.05 | 0.007 | 7.9433E-12 | 51.02 |
| *CHGB* | rs386705092 | C | CTTGAG | 0.3201 | -0.078 | 0.007 | 3.1623E-27 | 124.16 |
| *CHGB* | rs11154902 | C | T | 0.4626 | -0.089 | 0.007 | 1E-39 | 161.65 |
| *CHGB* | rs17092104 | G | C | 0.1093 | 0.132 | 0.011 | 1.5849E-34 | 144.00 |
| *CHGB* | rs28688991 | C | T | 0.1923 | -0.062 | 0.009 | 3.1623E-13 | 47.46 |
| *CHGB* | rs236153 | A | G | 0.3721 | -0.611 | 0.008 | 1E-200 | 5833.14 |
| *CHI3L1* | rs4950928 | G | C | 0.7927 | 0.672 | 0.009 | 1E-200 | 5575.11 |
| *CHI3L1* | rs11127048 | G | A | 0.6181 | -0.059 | 0.007 | 1E-17 | 71.04 |
| *CHI3L1* | rs28601761 | C | G | 0.4189 | -0.119 | 0.007 | 1E-69 | 289.00 |
| *CHI3L1* | rs2464190 | T | C | 0.4125 | 0.055 | 0.007 | 3.9811E-16 | 61.73 |
| *CHI3L1* | rs339969 | C | A | 0.6166 | -0.056 | 0.007 | 6.3096E-17 | 64.00 |
| *CHI3L1* | rs62117298 | T | C | 0.4813 | -0.062 | 0.007 | 6.3096E-21 | 78.45 |
| *CHIT1* | rs872583 | T | C | 0.203 | -1.092 | 0.009 | 1E-200 | 14721.78 |
| *CHIT1* | rs9272672 | T | C | 0.3184 | -0.049 | 0.006 | 7.9433E-16 | 66.69 |
| *CHIT1* | rs763913092 | GCAGCT | G | 0.4118 | 0.101 | 0.006 | 6.3096E-74 | 283.36 |
| *CHIT1* | rs11048473 | C | T | 0.2682 | 0.042 | 0.006 | 7.9433E-12 | 49.00 |
| *CHIT1* | rs11057840 | A | C | 0.1383 | 0.079 | 0.008 | 6.3096E-24 | 97.52 |
| *CHIT1* | rs34743976 | T | TA | 0.2795 | -0.049 | 0.006 | 7.9433E-16 | 66.69 |
| *CHL1* | rs931679 | T | A | 0.087 | 0.536 | 0.013 | 1E-200 | 1699.98 |
| *CHL1* | rs12574844 | G | A | 0.0709 | -0.116 | 0.014 | 1E-16 | 68.65 |
| *CHL1* | rs1800574 | C | T | 0.028 | 0.258 | 0.022 | 6.3096E-33 | 137.53 |
| *CHL1* | rs186021206 | G | A | 0.0064 | 0.716 | 0.049 | 3.9811E-48 | 213.52 |
| *CHL1* | rs8105767 | A | G | 0.2872 | -0.054 | 0.008 | 5.0119E-12 | 45.56 |
| *CHM* | rs1354034 | T | C | 0.604 | 0.061 | 0.008 | 5.0119E-15 | 58.14 |
| *CHMP1A* | rs7080386 | C | A | 0.4118 | 0.055 | 0.008 | 1.5849E-12 | 47.27 |
| *CHMP1A* | rs460879 | C | T | 0.4271 | -0.095 | 0.008 | 5.0119E-35 | 141.02 |
| *CHMP1A* | rs1671152 | T | G | 0.8408 | 0.073 | 0.01 | 3.1623E-12 | 53.29 |
| *CHMP6* | rs1260326 | T | C | 0.6086 | -0.058 | 0.008 | 5.0119E-14 | 52.56 |
| *CHMP6* | rs1050152 | C | T | 0.4344 | -0.053 | 0.008 | 1.9953E-12 | 43.89 |
| *CHMP6* | rs342298 | C | T | 0.4545 | -0.055 | 0.008 | 1.9953E-13 | 47.27 |
| *CHMP6* | rs56239087 | C | T | 0.172 | 0.203 | 0.01 | 3.1623E-93 | 412.09 |
| *CHRDL1* | rs215225 | A | G | 0.4283 | 0.049 | 0.007 | 7.9433E-13 | 49.00 |
| *CHRDL1* | rs777834943 | C | CT | 0.1834 | -0.069 | 0.009 | 6.3096E-15 | 58.78 |
| *CHRDL1* | rs186021206 | G | A | 0.0064 | 0.432 | 0.046 | 6.3096E-21 | 88.20 |
| *CHRDL1* | rs1065853 | G | T | 0.0812 | 0.107 | 0.012 | 3.9811E-18 | 79.51 |
| *CHRDL1* | rs2899319 | A | C | 0.6164 | 0.058 | 0.007 | 1.2589E-16 | 68.65 |
| *CHRDL2* | rs9810189 | C | A | 0.2854 | 0.056 | 0.008 | 1.2589E-12 | 49.00 |
| *CHRDL2* | rs574672574 | G | A | 0.0412 | -0.667 | 0.018 | 1E-200 | 1373.11 |
| *CHRDL2* | rs6590199 | G | A | 0.158 | 0.085 | 0.01 | 2.5119E-17 | 72.25 |
| *CHRDL2* | rs138903858 | G | T | 0.0277 | 0.193 | 0.022 | 6.3096E-18 | 76.96 |
| *CHRDL2* | rs200489612 | G | A | 0.0053 | 0.649 | 0.053 | 2.5119E-34 | 149.95 |
| *CIAPIN1* | rs1354034 | T | C | 0.6041 | 0.072 | 0.008 | 3.1623E-21 | 81.00 |
| *CIAPIN1* | rs342298 | C | T | 0.4549 | -0.052 | 0.007 | 5.0119E-12 | 55.18 |
| *CIAPIN1* | rs10733789 | T | C | 0.3132 | 0.056 | 0.008 | 6.3096E-12 | 49.00 |
| *CIAPIN1* | rs11458669 | T | TC | 0.5276 | 0.056 | 0.008 | 7.9433E-14 | 49.00 |
| *CILP* | rs56278466 | T | G | 0.6608 | 0.085 | 0.008 | 1.2589E-29 | 112.89 |
| *CILP* | rs35887873 | T | C | 0.2285 | 0.076 | 0.008 | 3.1623E-19 | 90.25 |
| *CILP* | rs6489548 | A | G | 0.5986 | -0.08 | 0.007 | 6.3096E-28 | 130.61 |
| *CILP* | rs2585035 | T | G | 0.6331 | -0.292 | 0.008 | 1E-200 | 1332.25 |
| *CIRBP* | rs1354034 | T | C | 0.6046 | 0.109 | 0.008 | 1.9953E-46 | 185.64 |

| | | | | | | | | |
|---|---|---|---|---|---|---|---|---|
| *CIRBP* | rs342298 | C | T | 0.455 | -0.06 | 0.008 | 1.9953E-15 | 56.25 |
| *CIRBP* | rs6993770 | A | T | 0.2866 | -0.058 | 0.008 | 1.5849E-12 | 52.56 |
| *CIRBP* | rs60822569 | T | TTTCTC | 0.5543 | 0.058 | 0.008 | 1.5849E-14 | 52.56 |
| *CIT* | rs3811444 | C | T | 0.3321 | -0.058 | 0.008 | 5.0119E-13 | 52.56 |
| *CIT* | rs17527306 | T | C | 0.0172 | -0.204 | 0.029 | 1.5849E-12 | 49.48 |
| *CKAP4* | rs61747728 | C | T | 0.038 | 0.135 | 0.019 | 2.5119E-13 | 50.48 |
| *CKAP4* | rs2704895 | G | A | 0.2219 | 0.223 | 0.009 | 3.981E-145 | 613.94 |
| *CKAP4* | rs28929474 | C | T | 0.0208 | 1.318 | 0.026 | 1E-200 | 2569.71 |
| *CKAP4* | rs28640218 | G | T | 0.1916 | -0.064 | 0.009 | 1.5849E-12 | 50.57 |
| *CKB* | rs1260326 | T | C | 0.6083 | 0.064 | 0.008 | 3.9811E-17 | 64.00 |
| *CKB* | rs4674100 | G | A | 0.2926 | -0.072 | 0.009 | 7.9433E-17 | 64.00 |
| *CKB* | rs13108218 | A | G | 0.619 | -0.057 | 0.008 | 1.5849E-13 | 50.77 |
| *CKB* | rs10027182 | G | A | 0.7442 | 0.058 | 0.008 | 1E-11 | 52.56 |
| *CKMT1A_CKMT1B* | rs700882 | C | T | 0.1509 | 0.071 | 0.011 | 1.2589E-11 | 41.66 |
| *CKMT1A_CKMT1B* | rs10205067 | A | G | 0.073 | -0.185 | 0.014 | 1.9953E-37 | 174.62 |
| *CKMT1A_CKMT1B* | rs34248556 | T | C | 0.656 | -0.07 | 0.008 | 3.1623E-18 | 76.56 |
| *CKMT1A_CKMT1B* | rs73082019 | A | G | 0.1012 | -0.094 | 0.013 | 6.3096E-14 | 52.28 |
| *CKMT1A_CKMT1B* | rs573943463 | C | T | 0.5063 | -0.078 | 0.008 | 1.5849E-24 | 95.06 |
| *CKMT1A_CKMT1B* | rs13292932 | C | A | 0.3094 | 0.066 | 0.008 | 7.9433E-16 | 68.06 |
| *CKMT1A_CKMT1B* | rs690604 | G | T | 0.0809 | -0.291 | 0.014 | 1.5849E-90 | 432.05 |
| *CKMT1A_CKMT1B* | rs601338 | G | A | 0.5096 | -0.089 | 0.008 | 3.9811E-32 | 123.77 |
| *CLASP1* | rs371337007 | G | A | 0.6457 | -0.082 | 0.009 | 3.9811E-19 | 83.01 |
| *CLC* | rs1186706 | G | T | 0.4906 | -0.05 | 0.007 | 1E-11 | 51.02 |
| *CLC* | rs10208293 | G | A | 0.2796 | -0.065 | 0.008 | 1.9953E-15 | 66.02 |
| *CLC* | rs4328821 | A | G | 0.1065 | -0.15 | 0.012 | 1.9953E-36 | 156.25 |
| *CLC* | rs11242115 | G | C | 0.3888 | -0.066 | 0.008 | 3.9811E-18 | 68.06 |
| *CLC* | rs448277 | C | T | 0.2891 | -0.061 | 0.008 | 3.1623E-13 | 58.14 |
| *CLC* | rs6467885 | T | C | 0.8956 | -0.186 | 0.012 | 1.9953E-54 | 240.25 |
| *CLC* | rs55646585 | C | T | 0.4138 | -0.073 | 0.007 | 1.2589E-22 | 108.76 |
| *CLC* | rs1888909 | T | C | 0.749 | -0.067 | 0.008 | 3.1623E-15 | 70.14 |
| *CLC* | rs35350651 | A | AC | 0.5041 | -0.052 | 0.007 | 1.5849E-12 | 55.18 |
| *CLC* | rs34210653 | G | A | 0.021 | -0.181 | 0.026 | 1.5849E-12 | 48.46 |
| *CLC* | rs143121234 | C | G | 0.0019 | -1.357 | 0.091 | 1E-50 | 222.37 |
| *CLC* | rs34436714 | C | A | 0.2087 | 0.14 | 0.009 | 6.3096E-54 | 241.98 |
| *CLEC10A* | rs781651672 | TTAAG | T | 0.4479 | 0.176 | 0.007 | 3.981E-140 | 632.16 |
| *CLEC10A* | rs9271378 | A | G | 0.4904 | 0.069 | 0.007 | 1E-23 | 97.16 |
| *CLEC10A* | rs971481 | C | G | 0.6137 | -0.12 | 0.007 | 1.5849E-64 | 293.88 |
| *CLEC10A* | rs13294683 | C | T | 0.1016 | 0.203 | 0.011 | 1E-71 | 340.57 |
| *CLEC10A* | rs2519093 | C | T | 0.1843 | 0.13 | 0.009 | 2.5119E-48 | 208.64 |
| *CLEC10A* | rs12771902 | G | A | 0.3253 | -0.056 | 0.007 | 1.9953E-14 | 64.00 |
| *CLEC10A* | rs215223 | G | A | 0.4001 | -0.084 | 0.007 | 6.3096E-33 | 144.00 |
| *CLEC10A* | rs3184504 | T | C | 0.517 | -0.059 | 0.007 | 6.3096E-18 | 71.04 |
| *CLEC10A* | rs56156922 | T | C | 0.3249 | 0.061 | 0.007 | 6.3096E-17 | 75.94 |
| *CLEC10A* | rs90951 | A | G | 0.3331 | 0.544 | 0.008 | 1E-200 | 4624.00 |
| *CLEC10A* | rs12938273 | G | A | 0.5543 | 0.093 | 0.007 | 1.9953E-35 | 176.51 |
| *CLEC10A* | rs769669239 | CCTTT | C | 0.4713 | -0.065 | 0.007 | 2.5119E-21 | 86.22 |
| *CLEC11A* | rs13063578 | T | A | 0.3984 | -0.074 | 0.008 | 3.1623E-22 | 85.56 |
| *CLEC11A* | rs4407843 | A | C | 0.5682 | -0.061 | 0.008 | 7.9433E-16 | 58.14 |
| *CLEC11A* | rs2282683 | A | G | 0.2098 | -0.074 | 0.009 | 1.9953E-16 | 67.60 |
| *CLEC11A* | rs4981022 | G | A | 0.6809 | 0.073 | 0.008 | 1.9953E-19 | 83.27 |
| *CLEC11A* | rs12050709 | T | C | 0.2445 | -0.059 | 0.009 | 7.9433E-12 | 42.98 |
| *CLEC11A* | rs2179768 | G | A | 0.5126 | -0.065 | 0.007 | 1.9953E-18 | 86.22 |
| *CLEC11A* | rs141369989 | G | A | 0.0285 | 0.82 | 0.023 | 1E-200 | 1271.08 |
| *CLEC11A* | rs13036464 | G | C | 0.407 | 0.057 | 0.008 | 3.9811E-14 | 50.77 |
| *CLEC12A* | rs12230629 | T | C | 0.267 | 0.494 | 0.009 | 1E-200 | 3012.79 |
| *CLEC12A* | rs11260012 | T | C | 0.4874 | -0.081 | 0.007 | 1.5849E-28 | 133.90 |
| *CLEC14A* | rs6664906 | T | C | 0.4642 | -0.102 | 0.007 | 5.0119E-45 | 212.33 |
| *CLEC14A* | rs61747728 | C | T | 0.0379 | 0.169 | 0.019 | 7.9433E-20 | 79.12 |
| *CLEC14A* | rs75166367 | G | A | 0.0604 | -0.113 | 0.015 | 3.9811E-14 | 56.75 |
| *CLEC14A* | rs13107325 | C | T | 0.0748 | 0.161 | 0.014 | 5.0119E-32 | 132.25 |
| *CLEC14A* | rs507666 | G | A | 0.1846 | -0.343 | 0.009 | 1E-200 | 1452.46 |
| *CLEC14A* | rs4842659 | G | A | 0.7137 | -0.094 | 0.008 | 1.5849E-32 | 138.06 |
| *CLEC14A* | rs10151239 | C | A | 0.4035 | 0.084 | 0.007 | 1.2589E-30 | 144.00 |
| *CLEC14A* | rs777834943 | C | CT | 0.183 | -0.075 | 0.009 | 1E-15 | 69.44 |
| *CLEC14A* | rs186021206 | G | A | 0.0064 | 0.432 | 0.049 | 1E-18 | 77.73 |
| *CLEC14A* | rs33950747 | C | T | 0.075 | 0.152 | 0.014 | 3.1623E-29 | 117.88 |
| *CLEC1A* | rs7669607 | T | C | 0.791 | 0.059 | 0.009 | 3.9811E-12 | 42.98 |
| *CLEC1A* | rs10982156 | T | A | 0.0695 | 0.227 | 0.015 | 6.3096E-52 | 229.02 |

| | | | | | | | | |
|---|---|---|---|---|---|---|---|---|
| *CLEC1A* | rs10901252 | G | C | 0.0615 | 0.78 | 0.015 | 1E-200 | 2704.00 |
| *CLEC1A* | rs7961483 | C | T | 0.62 | -0.317 | 0.008 | 1E-200 | 1570.14 |
| *CLEC1A* | rs77924615 | G | A | 0.1966 | -0.066 | 0.009 | 6.3096E-14 | 53.78 |
| *CLEC1A* | rs186021206 | G | A | 0.0063 | 0.443 | 0.048 | 3.1623E-20 | 85.18 |
| *CLEC1B* | rs3804749 | C | T | 0.5926 | -0.081 | 0.008 | 6.3096E-26 | 102.52 |
| *CLEC1B* | rs11242109 | G | T | 0.4811 | -0.065 | 0.008 | 7.9433E-18 | 66.02 |
| *CLEC1B* | rs6961069 | C | T | 0.4021 | 0.057 | 0.008 | 6.3096E-14 | 50.77 |
| *CLEC1B* | rs6993770 | A | T | 0.2865 | -0.131 | 0.008 | 3.9811E-56 | 268.14 |
| *CLEC1B* | rs7896518 | A | G | 0.4241 | 0.092 | 0.008 | 7.9433E-33 | 132.25 |
| *CLEC1B* | rs659928 | G | A | 0.2539 | 0.167 | 0.009 | 7.9433E-84 | 344.31 |
| *CLEC1B* | rs1654425 | T | C | 0.8345 | 0.112 | 0.01 | 1.5849E-28 | 125.44 |
| *CLEC2L* | rs607929 | C | G | 0.4485 | -0.065 | 0.008 | 3.9811E-17 | 66.02 |
| *CLEC3B* | rs13065490 | A | T | 0.2691 | 0.238 | 0.008 | 6.31E-177 | 885.06 |
| *CLEC3B* | rs13107325 | C | T | 0.075 | 0.122 | 0.014 | 2.5119E-18 | 75.94 |
| *CLEC3B* | rs111338191 | A | AAATT | 0.5208 | 0.051 | 0.007 | 6.3096E-12 | 53.08 |
| *CLEC3B* | rs77542162 | A | G | 0.0229 | 0.209 | 0.025 | 1.5849E-17 | 69.89 |
| *CLEC4A* | rs200688856 | G | C | 0.3417 | 0.129 | 0.007 | 3.9811E-69 | 339.61 |
| *CLEC4A* | rs791357 | T | C | 0.7202 | -0.058 | 0.008 | 1.5849E-13 | 52.56 |
| *CLEC4A* | rs1634791 | A | G | 0.5419 | 0.072 | 0.007 | 6.3096E-24 | 105.80 |
| *CLEC4A* | rs11775560 | A | G | 0.1859 | 0.069 | 0.009 | 1E-14 | 58.78 |
| *CLEC4A* | rs7074896 | T | C | 0.3107 | -0.054 | 0.008 | 1E-12 | 45.56 |
| *CLEC4A* | rs11603123 | G | A | 0.0337 | 0.467 | 0.019 | 1E-127 | 604.12 |
| *CLEC4A* | rs117213717 | G | A | 0.0141 | -2.006 | 0.032 | 1E-200 | 3929.72 |
| *CLEC4A* | rs10774624 | G | A | 0.5136 | 0.064 | 0.007 | 1.5849E-17 | 83.59 |
| *CLEC4A* | rs174418 | T | C | 0.5981 | -0.088 | 0.007 | 1.2589E-34 | 158.04 |
| *CLEC4A* | rs247617 | C | A | 0.3243 | 0.066 | 0.007 | 1E-18 | 88.90 |
| *CLEC4A* | rs11654208 | G | A | 0.3672 | -0.049 | 0.007 | 1.2589E-11 | 49.00 |
| *CLEC4A* | rs77960347 | A | G | 0.0132 | 0.215 | 0.031 | 1.9953E-12 | 48.10 |
| *CLEC4A* | rs4760 | A | G | 0.1562 | -0.108 | 0.01 | 2.5119E-29 | 116.64 |
| *CLEC4C* | rs7573683 | T | C | 0.542 | -0.054 | 0.006 | 7.9433E-20 | 81.00 |
| *CLEC4C* | rs174847 | C | T | 0.455 | 0.067 | 0.006 | 1.9953E-29 | 124.69 |
| *CLEC4C* | rs11899659 | A | G | 0.4071 | 0.045 | 0.006 | 7.9433E-14 | 56.25 |
| *CLEC4C* | rs1439287 | G | A | 0.4837 | 0.098 | 0.006 | 2.5119E-60 | 266.78 |
| *CLEC4C* | rs7595770 | G | T | 0.078 | -0.104 | 0.011 | 3.1623E-21 | 89.39 |
| *CLEC4C* | rs7658518 | G | A | 0.2826 | -0.079 | 0.007 | 1E-32 | 127.37 |
| *CLEC4C* | rs6818511 | T | C | 0.6592 | 0.117 | 0.006 | 2.5119E-77 | 380.25 |
| *CLEC4C* | rs17329170 | G | A | 0.0978 | -0.071 | 0.01 | 7.9433E-13 | 50.41 |
| *CLEC4C* | rs4713570 | C | T | 0.2662 | 0.065 | 0.007 | 3.9811E-22 | 86.22 |
| *CLEC4C* | rs74551305 | T | C | 0.0565 | 0.095 | 0.013 | 1.9953E-12 | 53.40 |
| *CLEC4C* | rs2347784 | C | G | 0.2717 | -0.072 | 0.007 | 6.3096E-27 | 105.80 |
| *CLEC4C* | rs876038 | C | T | 0.3156 | -0.206 | 0.006 | 1E-200 | 1178.78 |
| *CLEC4C* | rs10107630 | C | T | 0.574 | 0.062 | 0.006 | 7.9433E-25 | 106.78 |
| *CLEC4C* | rs10971417 | C | T | 0.2533 | -0.056 | 0.007 | 1.2589E-16 | 64.00 |
| *CLEC4C* | rs10441775 | A | G | 0.6917 | 0.046 | 0.006 | 1.2589E-12 | 58.78 |
| *CLEC4C* | rs7873862 | C | T | 0.1858 | 0.083 | 0.008 | 7.9433E-28 | 107.64 |
| *CLEC4C* | rs11789604 | T | G | 0.453 | 0.04 | 0.006 | 1E-11 | 44.44 |
| *CLEC4C* | rs10838702 | G | T | 0.3858 | 0.058 | 0.006 | 1.9953E-21 | 93.44 |
| *CLEC4C* | rs7129402 | C | A | 0.7202 | -0.138 | 0.007 | 1.5849E-94 | 388.65 |
| *CLEC4C* | rs10845821 | C | T | 0.4054 | 0.852 | 0.008 | 1E-200 | 11342.25 |
| *CLEC4C* | rs76428106 | T | C | 0.013 | 0.507 | 0.027 | 1.2589E-77 | 352.60 |
| *CLEC4C* | rs62045817 | C | T | 0.1504 | -0.083 | 0.008 | 7.9433E-23 | 107.64 |
| *CLEC4C* | rs62057151 | C | T | 0.2326 | -0.054 | 0.007 | 7.9433E-15 | 59.51 |
| *CLEC4C* | rs16947425 | C | A | 0.1589 | -0.068 | 0.008 | 2.5119E-17 | 72.25 |
| *CLEC4D* | rs2070901 | G | T | 0.2703 | -0.205 | 0.008 | 1E-151 | 656.64 |
| *CLEC4D* | rs6429432 | A | C | 0.8937 | -0.092 | 0.011 | 1.5849E-16 | 69.95 |
| *CLEC4D* | rs149110519 | C | T | 0.0363 | 0.14 | 0.018 | 2.5119E-14 | 60.49 |
| *CLEC4D* | rs445 | C | T | 0.0953 | -0.083 | 0.012 | 1E-12 | 47.84 |
| *CLEC4D* | rs2511241 | C | T | 0.9295 | 0.188 | 0.013 | 1E-44 | 209.14 |
| *CLEC4D* | rs4242896 | A | C | 0.5718 | 0.531 | 0.008 | 1E-200 | 4405.64 |
| *CLEC4D* | rs3826331 | T | C | 0.5985 | -0.119 | 0.007 | 6.3096E-65 | 289.00 |
| *CLEC4D* | rs872629 | C | A | 0.1138 | -0.152 | 0.011 | 3.1623E-45 | 190.94 |
| *CLEC4G* | rs10935473 | G | T | 0.4399 | -0.107 | 0.007 | 1.2589E-48 | 233.65 |
| *CLEC4G* | rs371581946 | G | GA | 0.4237 | 0.098 | 0.008 | 5.0119E-38 | 150.06 |
| *CLEC4G* | rs7865362 | C | T | 0.3627 | 0.069 | 0.008 | 1E-18 | 74.39 |
| *CLEC4G* | rs505922 | T | C | 0.3166 | 0.294 | 0.008 | 1E-200 | 1350.56 |
| *CLEC4G* | rs56278466 | T | G | 0.6618 | -0.087 | 0.008 | 7.9433E-30 | 118.27 |
| *CLEC4G* | rs10830278 | T | A | 0.7986 | 0.086 | 0.009 | 3.9811E-21 | 91.31 |
| *CLEC4G* | rs11172147 | G | A | 0.2435 | -0.062 | 0.008 | 1.5849E-13 | 60.06 |

| | | | | | | | | |
|---|---|---|---|---|---|---|---|---|
| *CLEC4G* | rs12709103 | A | G | 0.6762 | -0.055 | 0.008 | 7.9433E-13 | 47.27 |
| *CLEC4G* | rs17701598 | G | A | 0.2507 | -0.061 | 0.008 | 2.5119E-13 | 58.14 |
| *CLEC4G* | rs76560987 | C | T | 0.0709 | 0.374 | 0.014 | 2.512E-151 | 713.65 |
| *CLEC4M* | rs7528419 | A | G | 0.2221 | -0.096 | 0.007 | 2.5119E-43 | 188.08 |
| *CLEC4M* | rs505922 | T | C | 0.3171 | 0.84 | 0.008 | 1E-200 | 11025.00 |
| *CLEC4M* | rs10769256 | C | T | 0.4003 | 0.049 | 0.006 | 7.9433E-17 | 66.69 |
| *CLEC4M* | rs56156922 | T | C | 0.3247 | 0.046 | 0.006 | 1.2589E-13 | 58.78 |
| *CLEC4M* | rs77542162 | A | G | 0.023 | 0.132 | 0.019 | 7.9433E-12 | 48.27 |
| *CLEC4M* | rs4804774 | G | C | 0.237 | -0.364 | 0.008 | 1E-200 | 2070.25 |
| *CLEC5A* | rs10801568 | T | G | 0.2481 | -0.071 | 0.009 | 1.2589E-16 | 62.23 |
| *CLEC5A* | rs13107325 | C | T | 0.0751 | 0.096 | 0.013 | 1.2589E-12 | 54.53 |
| *CLEC5A* | rs9273113 | A | G | 0.3314 | -0.074 | 0.009 | 7.9433E-18 | 67.60 |
| *CLEC5A* | rs11769630 | T | A | 0.0727 | 0.131 | 0.014 | 7.9433E-21 | 87.56 |
| *CLEC5A* | rs6962383 | A | C | 0.2971 | 0.286 | 0.008 | 1E-200 | 1278.06 |
| *CLEC5A* | rs10838702 | G | T | 0.3854 | -0.059 | 0.007 | 3.9811E-16 | 71.04 |
| *CLEC5A* | rs56136109 | A | G | 0.0281 | -0.148 | 0.021 | 5.0119E-12 | 49.67 |
| *CLEC5A* | rs588136 | C | T | 0.7845 | -0.071 | 0.009 | 1.9953E-16 | 62.23 |
| *CLEC5A* | rs186021206 | G | A | 0.0064 | 0.442 | 0.048 | 7.9433E-20 | 84.79 |
| *CLEC5A* | rs3826331 | T | C | 0.5983 | -0.071 | 0.007 | 3.1623E-23 | 102.88 |
| *CLEC5A* | rs143333049 | A | G | 0.0405 | 0.132 | 0.019 | 1E-12 | 48.27 |
| *CLEC5A* | rs2254448 | C | G | 0.7778 | 0.181 | 0.009 | 1.995E-99 | 404.46 |
| *CLEC6A* | rs188468174 | C | T | 0.0143 | 0.215 | 0.03 | 5.0119E-13 | 51.36 |
| *CLEC6A* | rs2070901 | G | T | 0.27 | -0.12 | 0.008 | 1.5849E-52 | 225.00 |
| *CLEC6A* | rs10186921 | C | T | 0.5697 | -0.053 | 0.007 | 3.9811E-14 | 57.33 |
| *CLEC6A* | rs9880192 | G | C | 0.4141 | -0.051 | 0.007 | 6.3096E-13 | 53.08 |
| *CLEC6A* | rs2158799 | C | G | 0.6096 | 0.05 | 0.007 | 5.0119E-12 | 51.02 |
| *CLEC6A* | rs13277237 | G | A | 0.5678 | -0.049 | 0.007 | 3.1623E-12 | 49.00 |
| *CLEC6A* | rs11045427 | C | A | 0.215 | -0.643 | 0.009 | 1E-200 | 5104.31 |
| *CLEC6A* | rs3184504 | T | C | 0.517 | -0.094 | 0.008 | 1.5849E-35 | 138.06 |
| *CLEC6A* | rs76428106 | T | C | 0.013 | 0.269 | 0.032 | 3.1623E-17 | 70.67 |
| *CLEC6A* | rs2239630 | A | G | 0.5625 | -0.049 | 0.007 | 3.1623E-12 | 49.00 |
| *CLEC6A* | rs1147128 | G | A | 0.5424 | -0.085 | 0.007 | 6.3096E-34 | 147.45 |
| *CLEC6A* | rs8081692 | A | G | 0.3929 | 0.055 | 0.007 | 1.5849E-14 | 61.73 |
| *CLEC6A* | rs872629 | C | A | 0.1134 | -0.132 | 0.011 | 3.9811E-33 | 144.00 |
| *CLEC7A* | rs2647011 | C | A | 0.5385 | -0.041 | 0.006 | 6.3096E-12 | 46.69 |
| *CLEC7A* | rs16910526 | A | C | 0.0854 | -1.562 | 0.013 | 1E-200 | 14436.95 |
| *CLEC7A* | rs7137828 | C | T | 0.5163 | -0.074 | 0.007 | 3.9811E-23 | 111.76 |
| *CLGN* | rs6852397 | G | T | 0.0937 | 1.175 | 0.013 | 1E-200 | 8169.38 |
| *CLIC5* | rs2022423 | A | G | 0.3586 | -0.121 | 0.008 | 7.9433E-51 | 228.77 |
| *CLIP2* | rs117471007 | C | T | 0.0629 | 0.25 | 0.015 | 7.9433E-59 | 277.78 |
| *CLIP2* | rs6993770 | A | T | 0.2869 | -0.067 | 0.008 | 1.9953E-16 | 70.14 |
| *CLMP* | rs1260326 | T | C | 0.6084 | 0.05 | 0.007 | 2.5119E-13 | 51.02 |
| *CLMP* | rs73017877 | C | G | 0.5524 | 0.336 | 0.007 | 1E-200 | 2304.00 |
| *CLMP* | rs77924615 | G | A | 0.1966 | -0.075 | 0.009 | 1.5849E-18 | 69.44 |
| *CLNS1A* | rs6695321 | A | G | 0.3814 | 0.06 | 0.008 | 1E-14 | 56.25 |
| *CLPS* | rs376333472 | G | A | 0.0247 | 0.197 | 0.022 | 7.9433E-20 | 80.18 |
| *CLPS* | rs2766588 | T | C | 0.6074 | 0.55 | 0.008 | 1E-200 | 4726.56 |
| *CLPS* | rs138435015 | C | CTTTG | 0.9601 | 0.132 | 0.017 | 1.5849E-14 | 60.29 |
| *CLPS* | rs2653414 | C | A | 0.0127 | 0.256 | 0.029 | 3.1623E-18 | 77.93 |
| *CLPS* | rs7015116 | T | G | 0.1431 | -0.072 | 0.01 | 3.1623E-14 | 51.84 |
| *CLPS* | rs7068821 | G | T | 0.259 | -0.053 | 0.008 | 1.2589E-12 | 43.89 |
| *CLPS* | rs2286779 | G | C | 0.519 | -0.048 | 0.007 | 1.9953E-13 | 47.02 |
| *CLPS* | rs9652674 | G | A | 0.6912 | 0.089 | 0.007 | 2.5119E-35 | 161.65 |
| *CLPS* | rs35866622 | C | T | 0.5005 | 0.051 | 0.007 | 3.1623E-14 | 53.08 |
| *CLSPN* | rs1008982 | T | C | 0.3729 | -0.104 | 0.008 | 6.3096E-38 | 169.00 |
| *CLSTN2* | rs41265459 | T | A | 0.0067 | 2.283 | 0.046 | 1E-200 | 2463.18 |
| *CLSTN2* | rs6984305 | A | T | 0.885 | -0.103 | 0.011 | 1.5849E-21 | 87.68 |
| *CLSTN2* | rs77542162 | A | G | 0.0229 | 0.202 | 0.023 | 1.2589E-18 | 77.13 |
| *CLSTN2* | rs329721 | C | T | 0.3354 | -0.086 | 0.007 | 7.9433E-31 | 150.94 |
| *CLSTN3* | rs6984305 | A | T | 0.8848 | -0.108 | 0.011 | 1E-22 | 96.40 |
| *CLSTN3* | rs2519093 | C | T | 0.1844 | -0.237 | 0.009 | 7.943E-149 | 693.44 |
| *CLSTN3* | rs2035675 | A | G | 0.245 | 0.059 | 0.008 | 3.9811E-13 | 54.39 |
| *CLSTN3* | rs2167285 | C | T | 0.5663 | -0.468 | 0.008 | 1E-200 | 3422.25 |
| *CLSTN3* | rs111338191 | A | AAATT | 0.5212 | -0.057 | 0.008 | 2.5119E-14 | 50.77 |
| *CLSTN3* | rs28929474 | C | T | 0.0208 | 0.198 | 0.025 | 6.3096E-16 | 62.73 |
| *CLSTN3* | rs11075921 | C | T | 0.1597 | 0.065 | 0.01 | 7.9433E-12 | 42.25 |
| *CLSTN3* | rs77542162 | A | G | 0.023 | 0.254 | 0.023 | 1.5849E-27 | 121.96 |
| *CLTA* | rs3811444 | C | T | 0.3332 | -0.118 | 0.008 | 1E-47 | 217.56 |

| | | | | | | | | |
|---|---|---|---|---|---|---|---|---|
| *CLU* | rs1260326 | T | C | 0.6084 | -0.054 | 0.008 | 1E-12 | 45.56 |
| *CLU* | rs553014 | C | T | 0.5129 | 0.094 | 0.008 | 5.0119E-36 | 138.06 |
| *CLU* | rs7247412 | T | C | 0.2667 | -0.063 | 0.008 | 5.0119E-14 | 62.02 |
| *CLUL1* | rs34881159 | T | A | 0.4907 | 0.042 | 0.006 | 1.2589E-11 | 49.00 |
| *CLUL1* | rs17482753 | G | T | 0.1009 | 0.079 | 0.01 | 1.5849E-15 | 62.41 |
| *CLUL1* | rs174553 | A | G | 0.3443 | -0.049 | 0.006 | 3.9811E-15 | 66.69 |
| *CLUL1* | rs3751198 | A | G | 0.6042 | 0.047 | 0.006 | 1.5849E-14 | 61.36 |
| *CLUL1* | rs1601935 | G | T | 0.6559 | -0.051 | 0.006 | 1E-15 | 72.25 |
| *CLUL1* | rs12149545 | G | A | 0.3159 | 0.074 | 0.006 | 3.1623E-30 | 152.11 |
| *CLUL1* | rs186021206 | G | A | 0.0064 | 0.347 | 0.041 | 2.5119E-17 | 71.63 |
| *CLUL1* | rs12957470 | G | C | 0.2864 | 0.614 | 0.008 | 1E-200 | 5890.56 |
| *CLUL1* | rs4465830 | A | G | 0.1871 | -0.118 | 0.008 | 1.5849E-53 | 217.56 |
| *CMC1* | rs370517451 | G | A | 0.0146 | -0.314 | 0.034 | 1E-20 | 85.29 |
| *CMC1* | rs1354034 | T | C | 0.6045 | 0.059 | 0.008 | 6.3096E-15 | 54.39 |
| *CMC1* | rs61915365 | C | G | 0.1687 | -0.076 | 0.01 | 2.5119E-14 | 57.76 |
| *CMIP* | rs342298 | C | T | 0.455 | -0.056 | 0.008 | 1E-13 | 49.00 |
| *CNDP1* | rs645040 | G | T | 0.7723 | 0.113 | 0.008 | 1E-40 | 199.52 |
| *CNDP1* | rs1030431 | A | G | 0.669 | -0.056 | 0.008 | 1.2589E-13 | 49.00 |
| *CNDP1* | rs28601761 | C | G | 0.4192 | -0.049 | 0.007 | 1E-11 | 49.00 |
| *CNDP1* | rs139974673 | T | C | 0.0244 | -0.214 | 0.023 | 7.9433E-21 | 86.57 |
| *CNDP1* | rs17817077 | G | A | 0.3931 | 0.294 | 0.008 | 1E-200 | 1350.56 |
| *CNDP1* | rs8102710 | G | T | 0.2455 | -0.364 | 0.008 | 1E-200 | 2070.25 |
| *CNDP1* | rs12609475 | C | A | 0.1695 | -0.244 | 0.01 | 2.512E-134 | 595.36 |
| *CNDP1* | rs2294915 | C | T | 0.2315 | -0.06 | 0.008 | 1E-12 | 56.25 |
| *CNP* | rs1354034 | T | C | 0.6046 | 0.061 | 0.008 | 1E-15 | 58.14 |
| *CNP* | rs342298 | C | T | 0.4547 | -0.065 | 0.008 | 1E-17 | 66.02 |
| *CNPY2* | rs28929474 | C | T | 0.0208 | 0.475 | 0.027 | 2.5119E-69 | 309.50 |
| *CNPY4* | rs144305120 | T | G | 0.0706 | -0.335 | 0.015 | 3.162E-113 | 498.78 |
| *CNPY4* | rs342298 | C | T | 0.4545 | -0.053 | 0.008 | 3.1623E-12 | 43.89 |
| *CNST* | rs10740118 | G | C | 0.4139 | 0.055 | 0.008 | 3.1623E-13 | 47.27 |
| *CNTN1* | rs1814170 | A | T | 0.1003 | -0.152 | 0.011 | 3.1623E-43 | 190.94 |
| *CNTN1* | rs56278466 | T | G | 0.6614 | 0.059 | 0.007 | 1.2589E-17 | 71.04 |
| *CNTN1* | rs184621187 | G | A | 0.025 | 0.171 | 0.024 | 1.9953E-12 | 50.77 |
| *CNTN1* | rs11177604 | C | A | 0.3835 | 0.199 | 0.007 | 5.012E-175 | 808.18 |
| *CNTN1* | rs2071916 | T | C | 0.5857 | -0.301 | 0.007 | 1E-200 | 1849.00 |
| *CNTN1* | rs186021206 | G | A | 0.0064 | 0.481 | 0.045 | 2.5119E-26 | 114.25 |
| *CNTN2* | rs2071533 | T | G | 0.866 | -1.175 | 0.011 | 1E-200 | 11410.12 |
| *CNTN2* | rs912057 | A | G | 0.6104 | -0.074 | 0.006 | 1.5849E-32 | 152.11 |
| *CNTN2* | rs782455289 | TAAGAC | T | 0.3165 | -0.06 | 0.007 | 5.0119E-20 | 73.47 |
| *CNTN2* | rs56278466 | T | G | 0.6612 | 0.105 | 0.006 | 3.1623E-60 | 306.25 |
| *CNTN2* | rs74612335 | T | C | 0.1332 | -0.077 | 0.009 | 6.3096E-18 | 73.20 |
| *CNTN2* | rs186021206 | G | A | 0.0064 | 0.462 | 0.042 | 1.5849E-28 | 121.00 |
| *CNTN3* | rs114165349 | G | C | 0.0233 | 0.261 | 0.022 | 1.5849E-31 | 140.75 |
| *CNTN3* | rs3213563 | T | C | 0.559 | 0.053 | 0.007 | 3.1623E-15 | 57.33 |
| *CNTN3* | rs1260326 | T | C | 0.6084 | -0.086 | 0.007 | 6.3096E-36 | 150.94 |
| *CNTN3* | rs4677414 | A | G | 0.9291 | -0.512 | 0.014 | 1E-200 | 1337.47 |
| *CNTN3* | rs1154988 | T | A | 0.7729 | -0.084 | 0.008 | 7.9433E-24 | 110.25 |
| *CNTN3* | rs33951980 | C | T | 0.1288 | -0.117 | 0.01 | 1E-31 | 136.89 |
| *CNTN3* | rs4921914 | C | T | 0.779 | 0.071 | 0.008 | 1.2589E-18 | 78.77 |
| *CNTN3* | rs10822145 | C | T | 0.4723 | -0.057 | 0.007 | 2.5119E-17 | 66.31 |
| *CNTN3* | rs2030291 | A | T | 0.3866 | 0.048 | 0.007 | 5.0119E-12 | 47.02 |
| *CNTN3* | rs36124182 | A | G | 0.0364 | 0.132 | 0.018 | 1.5849E-13 | 53.78 |
| *CNTN3* | rs147233090 | C | T | 0.0235 | 0.305 | 0.022 | 7.9433E-43 | 192.20 |
| *CNTN3* | rs144268704 | A | AGGTTTTG( | 0.337 | 0.053 | 0.007 | 1.5849E-13 | 57.33 |
| *CNTN3* | rs11646097 | T | C | 0.592 | -0.074 | 0.007 | 1.2589E-27 | 111.76 |
| *CNTN3* | rs11647695 | A | C | 0.5535 | 0.058 | 0.007 | 6.3096E-18 | 68.65 |
| *CNTN3* | rs34088055 | C | T | 0.3161 | 0.053 | 0.007 | 6.3096E-13 | 57.33 |
| *CNTN3* | rs1292066 | T | A | 0.2146 | 0.061 | 0.008 | 5.0119E-14 | 58.14 |
| *CNTN3* | rs77542162 | A | G | 0.0229 | 0.21 | 0.022 | 1E-20 | 91.12 |
| *CNTN3* | rs55714572 | C | T | 0.147 | 0.065 | 0.01 | 1.2589E-11 | 42.25 |
| *CNTN4* | rs163352 | G | C | 0.8149 | -0.316 | 0.01 | 1E-200 | 998.56 |
| *CNTN4* | rs28366036 | A | C | 0.3442 | -0.055 | 0.008 | 7.9433E-13 | 47.27 |
| *CNTN4* | rs11730129 | T | C | 0.7825 | -0.079 | 0.008 | 1.9953E-21 | 97.52 |
| *CNTN4* | rs9270487 | A | G | 0.3363 | -0.053 | 0.008 | 1.5849E-12 | 43.89 |
| *CNTN4* | rs9987289 | A | G | 0.9081 | -0.091 | 0.012 | 1.9953E-14 | 57.51 |
| *CNTN4* | rs174583 | C | T | 0.3539 | 0.053 | 0.007 | 2.5119E-13 | 57.33 |
| *CNTN4* | rs4055121 | C | T | 0.1351 | -0.185 | 0.01 | 1.9953E-73 | 342.25 |
| *CNTN4* | rs7138520 | C | G | 0.4969 | -0.055 | 0.007 | 1E-15 | 61.73 |

| | | | | | | | | |
|---|---|---|---|---|---|---|---|---|
| *CNTN4* | rs186021206 | G | A | 0.0064 | 0.888 | 0.047 | 1.2589E-78 | 356.97 |
| *CNTN4* | rs2899319 | A | C | 0.6166 | 0.049 | 0.007 | 1.2589E-11 | 49.00 |
| *CNTN5* | rs780094 | T | C | 0.6197 | 0.044 | 0.006 | 1.9953E-12 | 53.78 |
| *CNTN5* | rs13133886 | A | C | 0.7822 | -0.126 | 0.007 | 3.1623E-65 | 324.00 |
| *CNTN5* | rs141744022 | A | AAATGTAC | 0.8307 | 0.068 | 0.008 | 1E-15 | 72.25 |
| *CNTN5* | rs10993170 | G | C | 0.0445 | 0.229 | 0.015 | 1.5849E-50 | 233.07 |
| *CNTN5* | rs72767003 | C | T | 0.1456 | -0.065 | 0.009 | 1E-13 | 52.16 |
| *CNTN5* | rs898776 | G | A | 0.9222 | -0.792 | 0.013 | 1E-200 | 3711.62 |
| *CNTN5* | rs186021206 | G | A | 0.0064 | 0.421 | 0.042 | 1.2589E-23 | 100.48 |
| *CNTN5* | rs134077 | A | G | 0.562 | 0.071 | 0.006 | 1E-29 | 140.03 |
| *CNTNAP2* | rs7810370 | A | G | 0.7144 | -0.554 | 0.008 | 1E-200 | 4795.56 |
| *CNTNAP2* | rs56278466 | T | G | 0.6618 | 0.122 | 0.007 | 1.5849E-65 | 303.76 |
| *CNTNAP2* | rs1033415 | A | G | 0.6457 | 0.059 | 0.007 | 1.2589E-16 | 71.04 |
| *CNTNAP4* | rs71534539 | G | A | 0.4646 | -0.107 | 0.009 | 6.3096E-36 | 141.35 |
| *COCH* | rs13412535 | G | A | 0.2311 | -0.237 | 0.008 | 1.259E-178 | 877.64 |
| *COCH* | rs7618405 | C | A | 0.2047 | -0.077 | 0.009 | 5.0119E-19 | 73.20 |
| *COCH* | rs1354034 | T | C | 0.6045 | -0.056 | 0.007 | 1.9953E-15 | 64.00 |
| *COCH* | rs545956457 | G | GT | 0.3678 | -0.051 | 0.007 | 6.3096E-12 | 53.08 |
| *COCH* | rs139141690 | G | A | 0.0048 | 0.654 | 0.051 | 3.1623E-37 | 164.44 |
| *COCH* | rs342292 | C | G | 0.4568 | 0.053 | 0.007 | 1.9953E-14 | 57.33 |
| *COCH* | rs576233947 | GT | G | 0.9087 | -0.14 | 0.012 | 3.9811E-30 | 136.11 |
| *COCH* | rs6993770 | A | T | 0.2866 | -0.193 | 0.008 | 1.995E-137 | 582.02 |
| *COCH* | rs663939 | A | G | 0.6457 | -0.051 | 0.007 | 1.9953E-12 | 53.08 |
| *COCH* | rs6580981 | G | A | 0.4582 | 0.08 | 0.007 | 1E-29 | 130.61 |
| *COCH* | rs28400019 | G | A | 0.115 | 0.51 | 0.012 | 1E-200 | 1806.25 |
| *COCH* | rs1552948 | C | T | 0.2952 | -0.115 | 0.008 | 7.9433E-51 | 206.64 |
| *COCH* | rs892090 | T | G | 0.8342 | 0.103 | 0.009 | 6.3096E-28 | 130.98 |
| *COL15A1* | rs1398018 | T | C | 0.4424 | 0.056 | 0.007 | 1E-14 | 64.00 |
| *COL15A1* | rs10819566 | T | C | 0.3647 | 0.189 | 0.008 | 1.259E-135 | 558.14 |
| *COL15A1* | rs77924615 | G | A | 0.1971 | -0.073 | 0.009 | 1.5849E-15 | 65.79 |
| *COL18A1* | rs1260326 | T | C | 0.6086 | -0.065 | 0.007 | 3.1623E-19 | 86.22 |
| *COL18A1* | rs28817415 | C | T | 0.4569 | 0.059 | 0.007 | 1.2589E-16 | 71.04 |
| *COL18A1* | rs116539064 | C | T | 0.0974 | 0.115 | 0.012 | 3.1623E-21 | 91.84 |
| *COL18A1* | rs9976834 | C | T | 0.1839 | -0.197 | 0.009 | 6.3096E-98 | 479.12 |
| *COL1A1* | rs1386625 | A | G | 0.902 | -0.128 | 0.013 | 1.5849E-24 | 96.95 |
| *COL1A1* | rs2023472 | A | G | 0.5875 | 0.05 | 0.007 | 1.2589E-11 | 51.02 |
| *COL1A1* | rs10955915 | A | G | 0.5403 | 0.098 | 0.007 | 6.3096E-40 | 196.00 |
| *COL1A1* | rs58973023 | A | T | 0.4872 | 0.131 | 0.007 | 1.5849E-70 | 350.22 |
| *COL1A1* | rs12879725 | C | T | 0.4948 | -0.056 | 0.007 | 3.1623E-14 | 64.00 |
| *COL1A1* | rs117068593 | C | T | 0.1871 | -0.074 | 0.009 | 3.1623E-15 | 67.60 |
| *COL1A1* | rs147266928 | T | C | 0.0012 | 1.425 | 0.111 | 7.9433E-38 | 164.81 |
| *COL1A1* | rs146385050 | C | A | 0.1831 | 0.133 | 0.01 | 7.9433E-44 | 176.89 |
| *COL1A1* | rs2957137 | A | T | 0.6395 | -0.059 | 0.008 | 1.9953E-14 | 54.39 |
| *COL24A1* | rs11161756 | T | C | 0.0922 | -0.257 | 0.013 | 7.9433E-86 | 390.82 |
| *COL24A1* | rs774510679 | ACTTTGCC | A | 0.4122 | 0.078 | 0.008 | 1.5849E-24 | 95.06 |
| *COL24A1* | rs62059251 | G | A | 0.3091 | 0.076 | 0.008 | 2.5119E-20 | 90.25 |
| *COL28A1* | rs6775517 | G | T | 0.3669 | 0.055 | 0.007 | 2.5119E-16 | 61.73 |
| *COL28A1* | rs7810134 | G | A | 0.232 | 0.905 | 0.009 | 1E-200 | 10111.42 |
| *COL28A1* | rs804289 | A | C | 0.328 | -0.047 | 0.007 | 7.9433E-12 | 45.08 |
| *COL28A1* | rs62509471 | A | G | 0.1767 | -0.081 | 0.009 | 6.3096E-20 | 81.00 |
| *COL28A1* | rs56278466 | T | G | 0.6614 | 0.092 | 0.007 | 1.5849E-41 | 172.73 |
| *COL28A1* | rs35772006 | A | G | 0.5867 | -0.066 | 0.007 | 1E-23 | 88.90 |
| *COL28A1* | rs55682587 | C | T | 0.022 | 0.164 | 0.022 | 6.3096E-14 | 55.57 |
| *COL2A1* | rs755492124 | TAAAG | T | 0.3937 | -0.053 | 0.006 | 3.9811E-16 | 78.03 |
| *COL2A1* | rs154507 | G | C | 0.5561 | -0.066 | 0.006 | 3.9811E-25 | 121.00 |
| *COL2A1* | rs10075801 | A | G | 0.4879 | -0.056 | 0.006 | 2.5119E-18 | 87.11 |
| *COL2A1* | rs3823764 | C | T | 0.8419 | 0.07 | 0.009 | 7.9433E-16 | 60.49 |
| *COL2A1* | rs6993770 | A | T | 0.2866 | -0.104 | 0.007 | 1.9953E-50 | 220.73 |
| *COL2A1* | rs139521338 | G | GA | 0.2857 | -0.048 | 0.007 | 1E-11 | 47.02 |
| *COL2A1* | rs7970830 | G | A | 0.4791 | -0.06 | 0.008 | 3.9811E-15 | 56.25 |
| *COL2A1* | rs10875723 | A | G | 0.3831 | -0.744 | 0.008 | 1E-200 | 8649.00 |
| *COL2A1* | rs61269268 | A | C | 0.1732 | -0.105 | 0.008 | 1.5849E-36 | 172.27 |
| *COL2A1* | rs11382616 | G | GC | 0.703 | -0.089 | 0.007 | 1E-37 | 161.65 |
| *COL2A1* | rs12445050 | C | T | 0.1387 | 0.074 | 0.009 | 3.1623E-16 | 67.60 |
| *COL2A1* | rs35005573 | G | T | 0.4803 | -0.064 | 0.006 | 2.5119E-24 | 113.78 |
| *COL2A1* | rs892090 | T | G | 0.8344 | 0.116 | 0.009 | 1E-41 | 166.12 |
| *COL2A1* | rs1033549 | G | T | 0.3778 | 0.05 | 0.007 | 1.2589E-14 | 51.02 |
| *COL3A1* | rs540608250 | A | G | 0.0028 | 1.335 | 0.079 | 1.5849E-64 | 285.57 |

| | | | | | | | | |
|---|---|---|---|---|---|---|---|---|
| *COL3A1* | rs146385050 | C | A | 0.1835 | 0.239 | 0.01 | 7.943E-131 | 571.21 |
| *COL4A1* | rs2498323 | G | A | 0.1006 | -0.145 | 0.012 | 3.9811E-33 | 146.01 |
| *COL4A1* | rs72793280 | C | T | 0.462 | 0.052 | 0.007 | 1.2589E-12 | 55.18 |
| *COL4A1* | rs35493868 | C | G | 0.201 | 0.076 | 0.009 | 5.0119E-17 | 71.31 |
| *COL4A1* | rs28600710 | G | A | 0.2087 | 0.082 | 0.009 | 5.0119E-20 | 83.01 |
| *COL4A1* | rs3741414 | C | T | 0.2463 | 0.072 | 0.008 | 2.5119E-17 | 81.00 |
| *COL4A1* | rs652572 | A | G | 0.6606 | 0.184 | 0.008 | 3.162E-123 | 529.00 |
| *COL4A1* | rs116539064 | C | T | 0.0973 | 0.189 | 0.013 | 1.9953E-51 | 211.37 |
| *COL4A1* | rs12483377 | G | A | 0.0886 | 0.415 | 0.013 | 1E-200 | 1019.08 |
| *COL4A1* | rs738408 | C | T | 0.2169 | 0.074 | 0.009 | 6.3096E-17 | 67.60 |
| *COL5A1* | rs9821691 | G | T | 0.4291 | 0.056 | 0.008 | 1.2589E-13 | 49.00 |
| *COL5A1* | rs114694170 | T | C | 0.0598 | 0.115 | 0.016 | 2.5119E-13 | 51.66 |
| *COL5A1* | rs4541868 | C | A | 0.2741 | -0.072 | 0.008 | 6.3096E-18 | 81.00 |
| *COL5A1* | rs11995824 | C | G | 0.5434 | 0.058 | 0.007 | 1E-14 | 68.65 |
| *COL5A1* | rs12002679 | C | G | 0.2815 | 0.236 | 0.009 | 2.512E-168 | 687.60 |
| *COL5A1* | rs56278466 | T | G | 0.661 | 0.103 | 0.008 | 1E-39 | 165.77 |
| *COL5A1* | rs12358789 | A | G | 0.4362 | 0.058 | 0.008 | 3.9811E-14 | 52.56 |
| *COL5A1* | rs2509582 | T | C | 0.7442 | 0.073 | 0.009 | 1E-17 | 65.79 |
| *COL5A1* | rs9594738 | C | T | 0.4878 | 0.089 | 0.007 | 3.1623E-33 | 161.65 |
| *COL5A1* | rs892090 | T | G | 0.8339 | 0.072 | 0.01 | 6.3096E-13 | 51.84 |
| *COL6A3* | rs1050785 | C | A | 0.6008 | 0.092 | 0.008 | 2.5119E-33 | 132.25 |
| *COL6A3* | rs186021206 | G | A | 0.0064 | 0.389 | 0.05 | 1E-14 | 60.53 |
| *COL6A3* | rs3088026 | C | T | 0.0889 | -0.113 | 0.013 | 1.5849E-18 | 75.56 |
| *COL9A1* | rs79220007 | T | C | 0.0772 | 0.113 | 0.014 | 1.5849E-16 | 65.15 |
| *COL9A1* | rs543674622 | A | G | 0.0028 | 1.526 | 0.077 | 3.9811E-88 | 392.76 |
| *COL9A1* | rs34072915 | T | G | 0.1151 | -0.101 | 0.011 | 3.1623E-19 | 84.31 |
| *COL9A1* | rs11831278 | C | T | 0.1614 | 0.085 | 0.01 | 5.0119E-18 | 72.25 |
| *COLEC12* | rs11606287 | C | A | 0.3636 | -0.053 | 0.007 | 1E-13 | 57.33 |
| *COLEC12* | rs34262842 | A | G | 0.1664 | -0.069 | 0.009 | 7.9433E-14 | 58.78 |
| *COLEC12* | rs149622251 | C | T | 0.006 | -0.849 | 0.045 | 5.0119E-78 | 355.95 |
| *COMMD1* | rs55677935 | C | T | 0.0174 | -0.673 | 0.03 | 3.981E-114 | 503.25 |
| *COMMD1* | rs1354034 | T | C | 0.6045 | 0.08 | 0.008 | 1.9953E-25 | 100.00 |
| *COMMD1* | rs150753801 | T | C | 0.0074 | -0.447 | 0.046 | 3.1623E-22 | 94.43 |
| *COMMD1* | rs10876550 | G | A | 0.5566 | 0.055 | 0.008 | 3.1623E-13 | 47.27 |
| *COMMD9* | rs1354034 | T | C | 0.6044 | 0.063 | 0.008 | 3.1623E-16 | 62.02 |
| *COMMD9* | rs2868346 | C | T | 0.7593 | -0.074 | 0.009 | 2.5119E-17 | 67.60 |
| *COMP* | rs4233367 | T | C | 0.6085 | -0.066 | 0.007 | 1.5849E-19 | 88.90 |
| *COMP* | rs56278466 | T | G | 0.6609 | 0.124 | 0.008 | 1E-60 | 240.25 |
| *COMP* | rs60648773 | G | A | 0.2279 | -0.067 | 0.009 | 5.0119E-15 | 55.42 |
| *COMP* | rs7294354 | T | G | 0.5679 | -0.051 | 0.007 | 1E-12 | 53.08 |
| *COMP* | rs7310615 | C | G | 0.5171 | 0.049 | 0.007 | 5.0119E-12 | 49.00 |
| *COMP* | rs186021206 | G | A | 0.0064 | 0.987 | 0.049 | 1.5849E-90 | 405.73 |
| *COMP* | rs12974746 | A | G | 0.0302 | -0.579 | 0.022 | 3.981E-157 | 692.65 |
| *COMT* | rs1354034 | T | C | 0.6037 | 0.051 | 0.007 | 3.9811E-12 | 53.08 |
| *COMT* | rs4680 | G | A | 0.5176 | -0.445 | 0.008 | 1E-200 | 3094.14 |
| *COQ7* | rs4665972 | T | C | 0.6084 | -0.062 | 0.008 | 1.5849E-15 | 60.06 |
| *COQ7* | rs4835265 | C | A | 0.1561 | 0.132 | 0.01 | 7.9433E-37 | 174.24 |
| *COQ7* | rs11074359 | C | T | 0.639 | -0.121 | 0.008 | 6.3096E-54 | 228.77 |
| *COQ7* | rs7189522 | T | C | 0.7274 | -0.057 | 0.008 | 1.2589E-11 | 50.77 |
| *COQ7* | rs9959832 | C | T | 0.7977 | -0.106 | 0.009 | 3.1623E-29 | 138.72 |
| *COQ7* | rs7256200 | G | T | 0.1279 | -0.081 | 0.011 | 1E-12 | 54.22 |
| *CORO1A* | rs342299 | C | T | 0.4549 | -0.063 | 0.008 | 7.9433E-17 | 62.02 |
| *CORO1A* | rs10432210 | C | T | 0.209 | 0.064 | 0.009 | 3.1623E-12 | 50.57 |
| *COX5B* | rs112327139 | C | T | 0.0073 | -0.689 | 0.045 | 1.5849E-53 | 234.43 |
| *COX5B* | rs201327644 | C | T | 0.0067 | -0.617 | 0.05 | 7.9433E-35 | 152.28 |
| *COX5B* | rs17569603 | T | C | 0.1744 | 0.069 | 0.01 | 7.9433E-12 | 47.61 |
| *COX5B* | rs35366140 | G | GA | 0.1674 | -0.079 | 0.01 | 1E-14 | 62.41 |
| *CPA1* | rs35004807 | T | TGTAA | 0.3449 | 0.067 | 0.008 | 1.2589E-17 | 70.14 |
| *CPA1* | rs2641348 | A | G | 0.1113 | -0.095 | 0.012 | 5.0119E-16 | 62.67 |
| *CPA1* | rs13103023 | G | A | 0.3281 | -0.073 | 0.008 | 1.5849E-20 | 83.27 |
| *CPA1* | rs11724862 | T | C | 0.2361 | -0.063 | 0.009 | 1.5849E-12 | 49.00 |
| *CPA1* | rs56762787 | T | TC | 0.1771 | -0.07 | 0.01 | 3.9811E-13 | 49.00 |
| *CPA1* | rs11759956 | C | T | 0.5489 | 0.058 | 0.007 | 5.0119E-15 | 68.65 |
| *CPA1* | rs3215532 | G | GA | 0.2526 | 0.07 | 0.008 | 1.9953E-16 | 76.56 |
| *CPA1* | rs373980643 | GT | G | 0.456 | -0.075 | 0.007 | 3.1623E-24 | 114.80 |
| *CPA1* | rs572393412 | C | CA | 0.3096 | 0.073 | 0.009 | 3.1623E-17 | 65.79 |
| *CPA1* | rs13226219 | T | C | 0.1285 | 0.256 | 0.011 | 1E-117 | 541.62 |
| *CPA1* | rs4921967 | G | T | 0.6866 | -0.073 | 0.008 | 5.0119E-20 | 83.27 |

| | | | | | | | | |
|---|---|---|---|---|---|---|---|---|
| *CPA1* | rs10901250 | G | A | 0.1214 | 0.091 | 0.011 | 6.3096E-16 | 68.44 |
| *CPA1* | rs56278466 | T | G | 0.6614 | 0.055 | 0.008 | 7.9433E-13 | 47.27 |
| *CPA1* | rs174548 | C | G | 0.3144 | 0.074 | 0.008 | 3.9811E-21 | 85.56 |
| *CPA1* | rs686056 | G | A | 0.3224 | -0.093 | 0.008 | 6.3096E-33 | 135.14 |
| *CPA1* | rs1492237 | T | C | 0.656 | -0.059 | 0.008 | 2.5119E-14 | 54.39 |
| *CPA1* | rs76734539 | G | A | 0.1434 | -0.085 | 0.01 | 3.1623E-16 | 72.25 |
| *CPA1* | rs72802342 | C | A | 0.0755 | -0.367 | 0.014 | 3.981E-146 | 687.19 |
| *CPA1* | rs1673931 | T | C | 0.3783 | -0.115 | 0.008 | 5.0119E-51 | 206.64 |
| *CPA1* | rs17138478 | C | A | 0.1309 | 0.082 | 0.011 | 3.1623E-14 | 55.57 |
| *CPA1* | rs1698113 | T | C | 0.4844 | 0.06 | 0.008 | 1.9953E-14 | 56.25 |
| *CPA1* | rs4817932 | C | G | 0.3633 | -0.06 | 0.008 | 7.9433E-14 | 56.25 |
| *CPA1* | rs80184311 | G | C | 0.1056 | 0.08 | 0.012 | 1.5849E-11 | 44.44 |
| *CPA2* | rs6952090 | T | C | 0.3482 | -0.369 | 0.008 | 1E-200 | 2127.52 |
| *CPA2* | rs56057779 | G | C | 0.5639 | 0.063 | 0.007 | 3.1623E-18 | 81.00 |
| *CPA2* | rs2519093 | C | T | 0.1847 | -0.111 | 0.009 | 7.9433E-33 | 152.11 |
| *CPA2* | rs56278466 | T | G | 0.6613 | 0.085 | 0.008 | 6.3096E-29 | 112.89 |
| *CPA2* | rs72802342 | C | A | 0.0757 | -0.229 | 0.014 | 1.2589E-61 | 267.56 |
| *CPA4* | rs80293268 | G | C | 0.0483 | -0.157 | 0.017 | 1.2589E-20 | 85.29 |
| *CPA4* | rs11384422 | A | AG | 0.5824 | 0.047 | 0.007 | 1.5849E-11 | 45.08 |
| *CPA4* | rs34587586 | G | T | 0.3973 | -0.535 | 0.008 | 1E-200 | 4472.27 |
| *CPA4* | rs529700 | T | C | 0.5145 | 0.079 | 0.008 | 1.9953E-22 | 97.52 |
| *CPA4* | rs7310615 | C | G | 0.5165 | 0.051 | 0.007 | 7.9433E-14 | 53.08 |
| *CPA4* | rs186021206 | G | A | 0.0065 | 0.343 | 0.047 | 2.5119E-13 | 53.26 |
| *CPA4* | rs12986064 | T | C | 0.5075 | -0.082 | 0.007 | 5.0119E-31 | 137.22 |
| *CPB1* | rs333947 | G | A | 0.1481 | -0.076 | 0.01 | 1.9953E-13 | 57.76 |
| *CPB1* | rs2027447 | T | C | 0.1113 | -0.097 | 0.012 | 1E-16 | 65.34 |
| *CPB1* | rs17032925 | T | C | 0.0828 | -0.101 | 0.013 | 3.9811E-14 | 60.36 |
| *CPB1* | rs13318853 | G | A | 0.2373 | 0.129 | 0.009 | 3.9811E-50 | 205.44 |
| *CPB1* | rs35400735 | T | TG | 0.2671 | -0.064 | 0.009 | 5.0119E-13 | 50.57 |
| *CPB1* | rs3215532 | G | GA | 0.2526 | 0.067 | 0.008 | 3.1623E-15 | 70.14 |
| *CPB1* | rs9482772 | T | C | 0.4509 | 0.075 | 0.007 | 5.0119E-24 | 114.80 |
| *CPB1* | rs572393412 | C | CA | 0.3096 | 0.072 | 0.009 | 5.0119E-17 | 64.00 |
| *CPB1* | rs4921967 | G | T | 0.6866 | -0.079 | 0.008 | 3.1623E-23 | 97.52 |
| *CPB1* | rs56278466 | T | G | 0.6614 | 0.055 | 0.008 | 1.2589E-12 | 47.27 |
| *CPB1* | rs174548 | C | G | 0.3144 | 0.072 | 0.008 | 7.9433E-20 | 81.00 |
| *CPB1* | rs686056 | G | A | 0.3224 | -0.089 | 0.008 | 3.9811E-30 | 123.77 |
| *CPB1* | rs72802342 | C | A | 0.0755 | -0.407 | 0.014 | 5.012E-178 | 845.15 |
| *CPB1* | rs1673931 | T | C | 0.3783 | -0.16 | 0.008 | 1.2589E-96 | 400.00 |
| *CPB1* | rs2211864 | T | A | 0.5071 | 0.058 | 0.007 | 2.5119E-15 | 68.65 |
| *CPB1* | rs80184311 | G | C | 0.1056 | 0.08 | 0.012 | 1.5849E-11 | 44.44 |
| *CPB2* | rs1260326 | T | C | 0.6082 | -0.088 | 0.007 | 7.9433E-38 | 158.04 |
| *CPB2* | rs451637 | C | T | 0.3099 | -0.056 | 0.007 | 5.0119E-14 | 64.00 |
| *CPB2* | rs192519670 | T | C | 0.0041 | -0.398 | 0.058 | 1E-11 | 47.09 |
| *CPB2* | rs2126263 | G | A | 0.8985 | 0.084 | 0.011 | 6.3096E-14 | 58.31 |
| *CPB2* | rs78689694 | G | C | 0.1327 | 0.079 | 0.01 | 1.2589E-15 | 62.41 |
| *CPB2* | rs6561283 | A | G | 0.7353 | 0.674 | 0.008 | 1E-200 | 7098.06 |
| *CPB2* | rs28929474 | C | T | 0.0209 | 0.161 | 0.023 | 6.3096E-12 | 49.00 |
| *CPE* | rs1550270 | T | C | 0.3009 | 0.256 | 0.008 | 1E-200 | 1024.00 |
| *CPE* | rs34958196 | G | A | 0.1292 | 0.083 | 0.011 | 1E-13 | 56.93 |
| *CPE* | rs532436 | G | A | 0.1847 | -0.083 | 0.01 | 1.2589E-17 | 68.89 |
| *CPE* | rs56278466 | T | G | 0.6611 | 0.063 | 0.008 | 1.9953E-15 | 62.02 |
| *CPE* | rs597808 | A | G | 0.5155 | 0.073 | 0.007 | 1E-22 | 108.76 |
| *CPE* | rs2393775 | G | A | 0.6236 | 0.1 | 0.008 | 3.1623E-38 | 156.25 |
| *CPE* | rs184458383 | C | T | 0.0764 | -0.125 | 0.014 | 7.9433E-19 | 79.72 |
| *CPM* | rs11118611 | A | G | 0.2911 | 0.089 | 0.008 | 3.9811E-29 | 123.77 |
| *CPM* | rs1862069 | G | A | 0.5375 | -0.054 | 0.007 | 1.5849E-13 | 59.51 |
| *CPM* | rs28601761 | C | G | 0.4193 | -0.083 | 0.007 | 5.0119E-29 | 140.59 |
| *CPM* | rs56278466 | T | G | 0.661 | 0.066 | 0.008 | 3.1623E-18 | 68.06 |
| *CPM* | rs10751647 | T | C | 0.4018 | 0.052 | 0.007 | 2.5119E-12 | 55.18 |
| *CPM* | rs7978197 | C | T | 0.0141 | -0.475 | 0.031 | 6.3096E-54 | 234.78 |
| *CPM* | rs1601935 | G | T | 0.6562 | 0.077 | 0.008 | 5.0119E-24 | 92.64 |
| *CPM* | rs200293726 | A | T | 0.3065 | 0.057 | 0.008 | 6.3096E-13 | 50.77 |
| *CPM* | rs186021206 | G | A | 0.0064 | 0.347 | 0.05 | 2.5119E-12 | 48.16 |
| *CPM* | rs5112 | C | G | 0.5335 | -0.058 | 0.008 | 6.3096E-14 | 52.56 |
| *CPM* | rs11668882 | T | C | 0.4407 | 0.054 | 0.007 | 1.2589E-13 | 59.51 |
| *CPM* | rs6115156 | G | C | 0.5223 | 0.078 | 0.007 | 2.5119E-27 | 124.16 |
| *CPOX* | rs35393423 | A | AT | 0.3238 | -0.062 | 0.008 | 1.9953E-14 | 60.06 |
| *CPOX* | rs1729988 | C | T | 0.131 | -0.157 | 0.011 | 5.0119E-44 | 203.71 |

| | | | | | | | | |
|---|---|---|---|---|---|---|---|---|
| *CPOX* | rs1800562 | G | A | 0.0777 | 0.193 | 0.014 | 3.9811E-42 | 190.05 |
| *CPOX* | rs855791 | A | G | 0.5609 | 0.063 | 0.008 | 1E-16 | 62.02 |
| *CPPED1* | rs8062313 | T | C | 0.1733 | 0.828 | 0.01 | 1E-200 | 6855.84 |
| *CPQ* | rs687339 | C | T | 0.771 | 0.078 | 0.008 | 2.5119E-22 | 95.06 |
| *CPQ* | rs13107325 | C | T | 0.075 | -0.244 | 0.013 | 1E-80 | 352.28 |
| *CPQ* | rs56055826 | T | A | 0.0607 | -0.105 | 0.014 | 1.9953E-13 | 56.25 |
| *CPQ* | rs148684432 | G | A | 0.0088 | -1.949 | 0.038 | 1E-200 | 2630.61 |
| *CPQ* | rs2272662 | T | C | 0.5778 | -0.054 | 0.007 | 7.9433E-14 | 59.51 |
| *CPQ* | rs10745925 | T | C | 0.2877 | -0.417 | 0.008 | 1E-200 | 2717.02 |
| *CPQ* | rs11622951 | G | A | 0.4618 | 0.055 | 0.007 | 3.1623E-16 | 61.73 |
| *CPQ* | rs145078947 | G | T | 0.0025 | 1.185 | 0.067 | 1.5849E-70 | 312.81 |
| *CPQ* | rs55707100 | C | T | 0.0242 | 0.158 | 0.022 | 3.9811E-13 | 51.58 |
| *CPQ* | rs58542926 | C | T | 0.0756 | 0.196 | 0.013 | 2.5119E-53 | 227.31 |
| *CPQ* | rs34954997 | C | CTTCG | 0.2376 | -0.077 | 0.008 | 3.9811E-22 | 92.64 |
| *CPVL* | rs710446 | T | C | 0.4088 | -0.053 | 0.006 | 1.5849E-18 | 78.03 |
| *CPVL* | rs2545801 | T | C | 0.7444 | -0.05 | 0.007 | 1.9953E-13 | 51.02 |
| *CPVL* | rs6914950 | C | T | 0.1473 | 0.509 | 0.009 | 1E-200 | 3198.53 |
| *CPVL* | rs993632 | C | T | 0.6431 | -0.05 | 0.007 | 1.9953E-14 | 51.02 |
| *CPVL* | rs34219043 | C | T | 0.1996 | -0.842 | 0.009 | 1E-200 | 8752.64 |
| *CPVL* | rs56278466 | T | G | 0.661 | 0.063 | 0.006 | 5.0119E-24 | 110.25 |
| *CPVL* | rs769611959 | GTGA | G | 0.3008 | 0.073 | 0.007 | 1.2589E-27 | 108.76 |
| *CPVL* | rs4883201 | A | G | 0.104 | 0.101 | 0.01 | 6.3096E-25 | 102.01 |
| *CPVL* | rs111338191 | A | AAATT | 0.5204 | -0.05 | 0.006 | 3.1623E-17 | 69.44 |
| *CPVL* | rs145078947 | G | T | 0.0026 | 1.492 | 0.059 | 7.943E-143 | 639.49 |
| *CPVL* | rs15951 | G | A | 0.5334 | -0.088 | 0.006 | 3.1623E-48 | 215.11 |
| *CPVL* | rs186021206 | G | A | 0.0064 | 0.325 | 0.041 | 1.5849E-15 | 62.83 |
| *CPVL* | rs74181667 | A | G | 0.3665 | 0.055 | 0.006 | 5.0119E-19 | 84.03 |
| *CPXM1* | rs1434282 | C | T | 0.7262 | -0.084 | 0.008 | 1E-24 | 110.25 |
| *CPXM1* | rs13412535 | G | A | 0.2311 | -0.058 | 0.009 | 1.5849E-11 | 41.53 |
| *CPXM1* | rs7618405 | C | A | 0.2047 | -0.064 | 0.009 | 1E-12 | 50.57 |
| *CPXM1* | rs1354034 | T | C | 0.6041 | -0.065 | 0.007 | 1.2589E-18 | 86.22 |
| *CPXM1* | rs755492124 | TAAAG | T | 0.3938 | -0.057 | 0.007 | 7.9433E-15 | 66.31 |
| *CPXM1* | rs3016013 | G | A | 0.7207 | 0.094 | 0.008 | 3.9811E-31 | 138.06 |
| *CPXM1* | rs6961069 | C | T | 0.4023 | 0.055 | 0.007 | 1.2589E-13 | 61.73 |
| *CPXM1* | rs6972167 | G | T | 0.1983 | -0.063 | 0.009 | 3.9811E-12 | 49.00 |
| *CPXM1* | rs6993770 | A | T | 0.2861 | -0.164 | 0.008 | 7.9433E-94 | 420.25 |
| *CPXM1* | rs692502 | G | A | 0.5562 | 0.051 | 0.007 | 3.9811E-12 | 53.08 |
| *CPXM1* | rs61469632 | T | C | 0.0635 | -0.115 | 0.015 | 1.2589E-14 | 58.78 |
| *CPXM1* | rs10740118 | G | C | 0.4144 | 0.069 | 0.007 | 6.3096E-21 | 97.16 |
| *CPXM1* | rs71474568 | T | TTG | 0.3401 | 0.083 | 0.008 | 7.9433E-28 | 107.64 |
| *CPXM1* | rs139130389 | C | CTA | 0.0726 | -0.095 | 0.014 | 7.9433E-12 | 46.05 |
| *CPXM1* | rs7970830 | G | A | 0.4792 | -0.058 | 0.007 | 1E-15 | 68.65 |
| *CPXM1* | rs4759076 | T | C | 0.4574 | 0.052 | 0.007 | 6.3096E-13 | 55.18 |
| *CPXM1* | rs12445050 | C | T | 0.1384 | 0.109 | 0.01 | 1.5849E-25 | 118.81 |
| *CPXM1* | rs57839578 | A | T | 0.4792 | -0.052 | 0.007 | 1E-12 | 55.18 |
| *CPXM1* | rs892090 | T | G | 0.8343 | 0.162 | 0.01 | 3.9811E-62 | 262.44 |
| *CPXM1* | rs215545 | T | C | 0.7645 | 0.363 | 0.009 | 1E-200 | 1626.78 |
| *CPXM1* | rs4814837 | C | T | 0.3431 | 0.07 | 0.008 | 7.9433E-19 | 76.56 |
| *CPXM1* | rs16989483 | T | C | 0.2721 | -0.059 | 0.008 | 1.2589E-12 | 54.39 |
| *CPXM2* | rs28741041 | G | C | 0.3871 | -0.477 | 0.008 | 1E-200 | 3555.14 |
| *CR1* | rs679515 | T | C | 0.8255 | -0.54 | 0.01 | 1E-200 | 2916.00 |
| *CR1* | rs1257169 | C | A | 0.4993 | -0.096 | 0.007 | 7.9433E-45 | 188.08 |
| *CR1* | rs13064814 | A | G | 0.7141 | -0.127 | 0.008 | 1.2589E-62 | 252.02 |
| *CR1* | rs1800759 | T | G | 0.6134 | -0.048 | 0.007 | 6.3096E-12 | 47.02 |
| *CR1* | rs6910861 | A | G | 0.4548 | -0.048 | 0.007 | 3.1623E-12 | 47.02 |
| *CR1* | rs13204265 | G | C | 0.1609 | 0.082 | 0.009 | 1E-18 | 83.01 |
| *CR1* | rs4841132 | A | G | 0.9082 | -0.085 | 0.012 | 7.9433E-13 | 50.17 |
| *CR1* | rs2031902 | C | T | 0.5876 | -0.05 | 0.007 | 5.0119E-13 | 51.02 |
| *CR1* | rs59602812 | A | AT | 0.3181 | 0.064 | 0.007 | 3.1623E-18 | 83.59 |
| *CR1* | rs68055275 | C | CT | 0.1447 | -0.136 | 0.01 | 1.5849E-42 | 184.96 |
| *CR1* | rs74622686 | A | G | 0.147 | 0.078 | 0.01 | 7.9433E-16 | 60.84 |
| *CR1* | rs10774625 | A | G | 0.5039 | -0.061 | 0.007 | 3.1623E-19 | 75.94 |
| *CR1* | rs186021206 | G | A | 0.0064 | 1.661 | 0.047 | 1E-200 | 1248.95 |
| *CR1* | rs34557412 | A | G | 0.007 | 0.358 | 0.041 | 3.9811E-18 | 76.24 |
| *CR1* | rs77542162 | A | G | 0.0228 | 0.175 | 0.023 | 2.5119E-14 | 57.89 |
| *CR1* | rs35106244 | C | T | 0.4634 | 0.074 | 0.007 | 1.2589E-25 | 111.76 |
| *CR2* | rs4656100 | T | C | 0.5452 | 0.065 | 0.007 | 7.9433E-19 | 86.22 |
| *CR2* | rs61821113 | C | T | 0.1137 | -0.283 | 0.011 | 2.512E-134 | 661.89 |

| | | | | | | | | |
|---|---|---|---|---|---|---|---|---|
| *CR2* | rs11585945 | A | T | 0.5961 | 0.078 | 0.007 | 1.5849E-25 | 124.16 |
| *CR2* | rs7634389 | T | C | 0.3898 | -0.068 | 0.007 | 3.1623E-20 | 94.37 |
| *CR2* | rs10012161 | A | G | 0.8161 | -0.093 | 0.009 | 1.9953E-23 | 106.78 |
| *CR2* | rs1047437 | C | G | 0.1695 | -0.068 | 0.01 | 1.9953E-12 | 46.24 |
| *CR2* | rs2760984 | G | C | 0.1844 | -0.188 | 0.01 | 3.1623E-85 | 353.44 |
| *CR2* | rs9460100 | T | C | 0.5532 | 0.059 | 0.007 | 7.9433E-16 | 71.04 |
| *CR2* | rs2389581 | C | G | 0.348 | 0.093 | 0.008 | 2.5119E-34 | 135.14 |
| *CR2* | rs7798316 | A | T | 0.4277 | 0.051 | 0.007 | 2.5119E-12 | 53.08 |
| *CR2* | rs12535578 | G | A | 0.2786 | 0.08 | 0.008 | 3.9811E-23 | 100.00 |
| *CR2* | rs12554848 | T | C | 0.272 | -0.062 | 0.008 | 1.9953E-14 | 60.06 |
| *CR2* | rs4055121 | C | T | 0.1348 | -0.081 | 0.011 | 1.9953E-14 | 54.22 |
| *CR2* | rs114697502 | C | T | 0.0858 | 0.11 | 0.013 | 1.2589E-17 | 71.60 |
| *CR2* | rs10774624 | G | A | 0.5132 | -0.069 | 0.007 | 1.2589E-21 | 97.16 |
| *CR2* | rs3751437 | A | G | 0.2383 | 0.061 | 0.008 | 5.0119E-13 | 58.14 |
| *CR2* | rs374039502 | T | A | 0.0205 | 0.243 | 0.027 | 1.9953E-19 | 81.00 |
| *CR2* | rs186021206 | G | A | 0.0064 | 1.021 | 0.05 | 1E-93 | 416.98 |
| *CR2* | rs34557412 | A | G | 0.0068 | 0.805 | 0.044 | 3.9811E-75 | 334.72 |
| *CR2* | rs12950377 | T | C | 0.1568 | -0.108 | 0.01 | 1.2589E-27 | 116.64 |
| *CR2* | rs660637 | C | G | 0.5839 | 0.072 | 0.007 | 1E-22 | 105.80 |
| *CR2* | rs200437269 | C | CTG | 0.1098 | 0.093 | 0.012 | 1.5849E-14 | 60.06 |
| *CR2* | rs73165110 | G | A | 0.0787 | -0.098 | 0.013 | 3.1623E-13 | 56.83 |
| *CRACR2A* | rs1354034 | T | C | 0.6038 | 0.052 | 0.008 | 5.0119E-12 | 42.25 |
| *CRACR2A* | rs6993770 | A | T | 0.2869 | -0.059 | 0.008 | 1E-12 | 54.39 |
| *CRACR2A* | rs11227136 | G | A | 0.2433 | -0.063 | 0.009 | 6.3096E-13 | 49.00 |
| *CRACR2A* | rs6489498 | A | G | 0.3426 | -0.134 | 0.008 | 5.0119E-65 | 280.56 |
| *CRADD* | rs1354034 | T | C | 0.6039 | 0.055 | 0.008 | 7.9433E-13 | 47.27 |
| *CRADD* | rs342298 | C | T | 0.4545 | -0.052 | 0.008 | 6.3096E-12 | 42.25 |
| *CRADD* | rs7896518 | A | G | 0.4238 | 0.054 | 0.008 | 2.5119E-12 | 45.56 |
| *CRADD* | rs56944668 | C | T | 0.157 | -0.1 | 0.01 | 7.9433E-22 | 100.00 |
| *CREG1* | rs7513428 | T | C | 0.8409 | -0.254 | 0.01 | 3.162E-146 | 645.16 |
| *CREG1* | rs687339 | C | T | 0.771 | 0.068 | 0.009 | 1.2589E-15 | 57.09 |
| *CREG1* | rs13107325 | C | T | 0.0749 | 0.122 | 0.014 | 3.1623E-19 | 75.94 |
| *CREG1* | rs77945361 | G | A | 0.1482 | -0.069 | 0.01 | 7.9433E-12 | 47.61 |
| *CREG1* | rs4594967 | A | G | 0.6752 | 0.06 | 0.008 | 5.0119E-15 | 56.25 |
| *CREG1* | rs12359178 | G | A | 0.1444 | 0.102 | 0.01 | 1.9953E-23 | 104.04 |
| *CREG1* | rs9414801 | G | A | 0.5332 | -0.049 | 0.007 | 1.2589E-11 | 49.00 |
| *CREG1* | rs139130389 | C | CTA | 0.0725 | -0.11 | 0.014 | 1E-15 | 61.73 |
| *CREG1* | rs150641790 | T | TG | 0.0748 | 0.104 | 0.014 | 1.9953E-14 | 55.18 |
| *CREG1* | rs145078947 | G | T | 0.0025 | 1.005 | 0.071 | 3.9811E-45 | 200.36 |
| *CREG1* | rs58542926 | C | T | 0.0752 | 0.211 | 0.014 | 2.5119E-54 | 227.15 |
| *CREG1* | rs429358 | T | C | 0.1558 | -0.121 | 0.01 | 2.5119E-34 | 146.41 |
| *CREG1* | rs3747207 | G | A | 0.2148 | 0.068 | 0.009 | 3.9811E-15 | 57.09 |
| *CRELD1* | rs61747728 | C | T | 0.0381 | 0.114 | 0.016 | 1.5849E-12 | 50.77 |
| *CRELD1* | rs9878047 | T | C | 0.249 | 0.832 | 0.009 | 1E-200 | 8545.98 |
| *CRELD1* | rs204893 | T | C | 0.4311 | 0.045 | 0.006 | 6.3096E-13 | 56.25 |
| *CRELD1* | rs28929474 | C | T | 0.0207 | 0.209 | 0.022 | 1E-21 | 90.25 |
| *CRELD2* | rs1295638 | A | G | 0.5159 | 0.063 | 0.007 | 1E-18 | 81.00 |
| *CRELD2* | rs9852409 | T | A | 0.8216 | -0.185 | 0.009 | 1.5849E-87 | 422.53 |
| *CRELD2* | rs6796 | T | C | 0.2788 | 0.083 | 0.008 | 2.5119E-25 | 107.64 |
| *CRELD2* | rs3752617 | G | T | 0.0678 | -0.15 | 0.014 | 7.9433E-26 | 114.80 |
| *CRELD2* | rs9987289 | A | G | 0.9079 | -0.172 | 0.012 | 2.5119E-44 | 205.44 |
| *CRELD2* | rs10488686 | G | A | 0.3659 | 0.056 | 0.007 | 3.9811E-14 | 64.00 |
| *CRELD2* | rs1805214 | G | A | 0.1904 | -0.07 | 0.009 | 1E-14 | 60.49 |
| *CRELD2* | rs79091287 | G | A | 0.0855 | -0.088 | 0.013 | 5.0119E-12 | 45.82 |
| *CRELD2* | rs1801689 | A | C | 0.0302 | 0.155 | 0.021 | 6.3096E-14 | 54.48 |
| *CRELD2* | rs2659005 | C | T | 0.4407 | 0.049 | 0.007 | 5.0119E-12 | 49.00 |
| *CRELD2* | rs74510325 | C | G | 0.0348 | -0.813 | 0.02 | 1E-200 | 1652.42 |
| *CRH* | rs12740374 | G | T | 0.2209 | -0.106 | 0.009 | 7.9433E-34 | 138.72 |
| *CRH* | rs10183338 | G | T | 0.2918 | -0.058 | 0.008 | 3.9811E-13 | 52.56 |
| *CRH* | rs4561605 | C | G | 0.461 | 0.071 | 0.007 | 5.0119E-22 | 102.88 |
| *CRH* | rs13017862 | T | C | 0.1827 | -0.076 | 0.009 | 7.9433E-16 | 71.31 |
| *CRH* | rs7687767 | A | G | 0.1841 | 0.084 | 0.009 | 3.1623E-19 | 87.11 |
| *CRH* | rs767653779 | TCTTA | T | 0.7854 | 0.065 | 0.009 | 1.9953E-13 | 52.16 |
| *CRH* | rs6887216 | T | A | 0.6377 | 0.065 | 0.008 | 1E-17 | 66.02 |
| *CRH* | rs331700 | G | A | 0.4548 | 0.05 | 0.007 | 7.9433E-12 | 51.02 |
| *CRH* | rs12518248 | T | C | 0.4172 | 0.059 | 0.007 | 7.9433E-16 | 71.04 |
| *CRH* | rs9503212 | A | G | 0.5216 | 0.109 | 0.007 | 2.5119E-50 | 242.47 |
| *CRH* | rs9264839 | C | T | 0.1671 | -0.096 | 0.01 | 6.3096E-23 | 92.16 |

| | | | | | | | | |
|---|---|---|---|---|---|---|---|---|
| *CRH* | rs1229027 | G | A | 0.2428 | -0.09 | 0.008 | 5.0119E-26 | 126.56 |
| *CRH* | rs111632177 | G | A | 0.114 | 0.092 | 0.011 | 7.9433E-16 | 69.95 |
| *CRH* | rs59104426 | T | A | 0.0334 | -0.681 | 0.021 | 1E-200 | 1051.61 |
| *CRH* | rs4263799 | C | T | 0.48 | 0.164 | 0.007 | 1.259E-105 | 548.90 |
| *CRH* | rs7387462 | C | T | 0.0434 | 0.44 | 0.018 | 5.012E-125 | 597.53 |
| *CRH* | rs5898348 | G | GA | 0.2728 | 0.08 | 0.008 | 1.9953E-22 | 100.00 |
| *CRH* | rs59585065 | G | GT | 0.7118 | 0.094 | 0.008 | 5.0119E-29 | 138.06 |
| *CRH* | rs1463597 | A | G | 0.4587 | -0.067 | 0.007 | 2.5119E-20 | 91.61 |
| *CRH* | rs12296430 | G | C | 0.1868 | -0.174 | 0.009 | 1.5849E-77 | 373.78 |
| *CRH* | rs10774624 | G | A | 0.5131 | -0.058 | 0.007 | 3.1623E-15 | 68.65 |
| *CRH* | rs7192602 | G | A | 0.2971 | -0.058 | 0.008 | 5.0119E-13 | 52.56 |
| *CRH* | rs77542162 | A | G | 0.023 | 0.164 | 0.024 | 1.5849E-11 | 46.69 |
| *CRH* | rs7412 | C | T | 0.081 | 0.094 | 0.013 | 1.9953E-12 | 52.28 |
| *CRHBP* | rs11162598 | G | A | 0.3568 | 0.053 | 0.007 | 1.5849E-13 | 57.33 |
| *CRHBP* | rs6798915 | C | G | 0.3022 | 0.052 | 0.007 | 1.9953E-12 | 55.18 |
| *CRHBP* | rs6896257 | G | A | 0.6385 | -0.588 | 0.008 | 1E-200 | 5402.25 |
| *CRHBP* | rs6924387 | A | G | 0.4058 | -0.058 | 0.007 | 6.3096E-17 | 68.65 |
| *CRHBP* | rs10104003 | C | T | 0.2101 | 0.151 | 0.008 | 1.5849E-73 | 356.27 |
| *CRIM1* | rs201258494 | T | TGCG | 0.1919 | -0.153 | 0.009 | 3.1623E-61 | 289.00 |
| *CRIM1* | rs6474527 | G | C | 0.6514 | -0.102 | 0.008 | 1E-39 | 162.56 |
| *CRIM1* | rs11244061 | C | T | 0.1155 | -0.094 | 0.011 | 1.5849E-16 | 73.02 |
| *CRIM1* | rs3751198 | A | G | 0.6037 | 0.057 | 0.007 | 1.9953E-14 | 66.31 |
| *CRIP2* | rs705379 | G | A | 0.477 | 0.09 | 0.007 | 7.9433E-36 | 165.31 |
| *CRIP2* | rs495203 | C | T | 0.3178 | 0.073 | 0.008 | 1.9953E-21 | 83.27 |
| *CRIP2* | rs80324969 | G | C | 0.171 | 0.183 | 0.01 | 1E-74 | 334.89 |
| *CRISP2* | rs9287979 | A | G | 0.6248 | -0.066 | 0.006 | 1.5849E-32 | 121.00 |
| *CRISP2* | rs35303892 | A | AT | 0.1958 | -0.083 | 0.007 | 6.3096E-34 | 140.59 |
| *CRISP2* | rs3087450 | G | A | 0.0129 | -0.167 | 0.024 | 1.9953E-12 | 48.42 |
| *CRISP2* | rs34687149 | T | TA | 0.0521 | 0.103 | 0.012 | 2.5119E-17 | 73.67 |
| *CRISP2* | rs555247 | G | A | 0.4556 | 0.489 | 0.007 | 1E-200 | 4880.02 |
| *CRISP2* | rs72558770 | A | ATAG | 0.5198 | -0.049 | 0.007 | 5.0119E-14 | 49.00 |
| *CRISP2* | rs6460105 | A | C | 0.6968 | -0.135 | 0.006 | 3.162E-116 | 506.25 |
| *CRISP2* | rs58248492 | A | AAGT | 0.2196 | -0.067 | 0.007 | 6.3096E-25 | 91.61 |
| *CRISP2* | rs10993994 | T | C | 0.6079 | -0.041 | 0.005 | 1.2589E-13 | 67.24 |
| *CRISP2* | rs74973131 | G | A | 0.0321 | -0.169 | 0.015 | 1E-28 | 126.94 |
| *CRISP2* | rs1766138 | G | C | 0.325 | -0.04 | 0.006 | 3.1623E-12 | 44.44 |
| *CRISP2* | rs8030179 | A | T | 0.7991 | -0.045 | 0.007 | 1E-11 | 41.33 |
| *CRISP2* | rs145144275 | A | G | 0.0088 | -0.479 | 0.029 | 1.2589E-62 | 272.82 |
| *CRISP2* | rs4911257 | T | C | 0.396 | -0.049 | 0.005 | 7.9433E-19 | 96.04 |
| *CRISP2* | rs67610389 | G | A | 0.2883 | 0.045 | 0.006 | 3.1623E-14 | 56.25 |
| *CRISP3* | rs3917932 | C | G | 0.5754 | -0.084 | 0.007 | 3.1623E-38 | 144.00 |
| *CRISP3* | rs149290349 | G | A | 0.0735 | 0.087 | 0.012 | 2.5119E-12 | 52.56 |
| *CRISP3* | rs778139 | G | A | 0.4887 | -0.052 | 0.006 | 2.5119E-16 | 75.11 |
| *CRISP3* | rs749520716 | AT | A | 0.4307 | 0.048 | 0.007 | 2.5119E-13 | 47.02 |
| *CRISP3* | rs35987226 | T | TA | 0.2215 | 0.066 | 0.008 | 5.0119E-18 | 68.06 |
| *CRISP3* | rs62246445 | G | A | 0.1752 | 0.065 | 0.008 | 1E-14 | 66.02 |
| *CRISP3* | rs2734031 | T | C | 0.9102 | -0.101 | 0.011 | 2.5119E-19 | 84.31 |
| *CRISP3* | rs218264 | A | T | 0.2475 | 0.08 | 0.007 | 6.3096E-27 | 130.61 |
| *CRISP3* | rs7705526 | C | A | 0.3246 | 0.064 | 0.007 | 1.2589E-20 | 83.59 |
| *CRISP3* | rs114300451 | G | A | 0.0675 | -0.147 | 0.015 | 2.5119E-23 | 96.04 |
| *CRISP3* | rs1025347 | G | A | 0.4962 | -0.571 | 0.007 | 1E-200 | 6653.90 |
| *CRISP3* | rs149110519 | C | T | 0.0364 | 0.25 | 0.02 | 5.0119E-36 | 156.25 |
| *CRISP3* | rs2158799 | C | G | 0.6094 | 0.049 | 0.007 | 6.3096E-14 | 49.00 |
| *CRISP3* | rs11769630 | T | A | 0.0729 | 0.094 | 0.012 | 2.5119E-14 | 61.36 |
| *CRISP3* | rs773462635 | TGACTA | T | 0.6415 | -0.051 | 0.007 | 3.1623E-14 | 53.08 |
| *CRISP3* | rs45577137 | A | G | 0.0452 | 0.127 | 0.016 | 1E-14 | 63.00 |
| *CRISP3* | rs10107630 | C | T | 0.5743 | -0.065 | 0.006 | 6.3096E-24 | 117.36 |
| *CRISP3* | rs150813342 | C | T | 0.0086 | -0.264 | 0.036 | 1.5849E-13 | 53.78 |
| *CRISP3* | rs55742421 | A | C | 0.2051 | -0.057 | 0.008 | 6.3096E-13 | 50.77 |
| *CRISP3* | rs2074038 | G | T | 0.1002 | 0.083 | 0.011 | 7.9433E-15 | 56.93 |
| *CRISP3* | rs771695343 | CTTGGTTGT | C | 0.3302 | 0.06 | 0.007 | 1.5849E-18 | 73.47 |
| *CRISP3* | rs7989654 | G | A | 0.6288 | 0.049 | 0.007 | 1.5849E-13 | 49.00 |
| *CRISP3* | rs34460966 | C | CA | 0.6243 | -0.06 | 0.007 | 2.5119E-19 | 73.47 |
| *CRISP3* | rs391855 | A | T | 0.4238 | 0.047 | 0.006 | 6.3096E-13 | 61.36 |
| *CRISP3* | rs186021206 | G | A | 0.0064 | 0.364 | 0.044 | 1.5849E-16 | 68.44 |
| *CRISP3* | rs11655264 | G | C | 0.4507 | -0.108 | 0.006 | 7.9433E-64 | 324.00 |
| *CRISP3* | rs893182 | T | C | 0.9511 | 0.411 | 0.015 | 6.31E-163 | 750.76 |
| *CRISP3* | rs243587 | A | C | 0.4792 | -0.06 | 0.006 | 1.5849E-20 | 100.00 |

| | | | | | | | | |
|---|---|---|---|---|---|---|---|---|
| *CRKL* | rs6993770 | A | T | 0.2866 | -0.071 | 0.008 | 1.2589E-17 | 78.77 |
| *CRLF1* | rs72538440 | G | GC | 0.4243 | -0.05 | 0.007 | 1.2589E-11 | 51.02 |
| *CRLF1* | rs11676298 | C | G | 0.192 | -0.069 | 0.009 | 1E-13 | 58.78 |
| *CRLF1* | rs11362821 | G | A | 0.9753 | -0.17 | 0.024 | 5.0119E-13 | 50.17 |
| *CRLF1* | rs180408 | C | G | 0.7785 | 0.059 | 0.009 | 1.2589E-11 | 42.98 |
| *CRLF1* | rs10774624 | G | A | 0.5134 | -0.064 | 0.007 | 1E-18 | 83.59 |
| *CRLF1* | rs1126464 | G | C | 0.2438 | -0.065 | 0.008 | 1.5849E-14 | 66.02 |
| *CRLF1* | rs12986064 | T | C | 0.5079 | -0.073 | 0.008 | 2.5119E-22 | 83.27 |
| *CRNN* | rs10494275 | G | A | 0.0712 | -1.187 | 0.014 | 1E-200 | 7188.62 |
| *CRNN* | rs9274369 | A | G | 0.7825 | 0.074 | 0.008 | 1.5849E-19 | 85.56 |
| *CRNN* | rs34357864 | G | GA | 0.3171 | 0.054 | 0.007 | 1.5849E-14 | 59.51 |
| *CRNN* | rs704 | G | A | 0.4727 | 0.092 | 0.007 | 2.5119E-45 | 172.73 |
| *CRTAC1* | rs1042445 | C | T | 0.2222 | 0.492 | 0.008 | 1E-200 | 3782.25 |
| *CRTAC1* | rs56007204 | C | T | 0.212 | -0.778 | 0.008 | 1E-200 | 9457.56 |
| *CRTAM* | rs13401811 | G | A | 0.1817 | -0.068 | 0.009 | 1.2589E-13 | 57.09 |
| *CRTAM* | rs1047891 | C | A | 0.3129 | 0.06 | 0.008 | 1E-14 | 56.25 |
| *CRTAM* | rs13063578 | T | A | 0.3976 | -0.1 | 0.007 | 1.5849E-43 | 204.08 |
| *CRTAM* | rs35829610 | C | CTG | 0.3385 | -0.057 | 0.008 | 3.1623E-14 | 50.77 |
| *CRTAM* | rs9267084 | G | A | 0.1536 | 0.125 | 0.01 | 1.5849E-36 | 156.25 |
| *CRTAM* | rs2327832 | A | G | 0.2226 | 0.061 | 0.009 | 1E-12 | 45.94 |
| *CRTAM* | rs3181356 | C | T | 0.2291 | 0.062 | 0.009 | 1.5849E-12 | 47.46 |
| *CRTAM* | rs11215416 | A | G | 0.2574 | -0.066 | 0.008 | 1E-14 | 68.06 |
| *CRTAM* | rs2370794 | A | G | 0.3366 | 0.313 | 0.008 | 1E-200 | 1530.77 |
| *CRTAM* | rs3184504 | T | C | 0.517 | -0.086 | 0.007 | 3.1623E-34 | 150.94 |
| *CRTAM* | rs111714972 | T | C | 0.3681 | 0.052 | 0.008 | 1.5849E-11 | 42.25 |
| *CRTAM* | rs186021206 | G | A | 0.0064 | 0.627 | 0.049 | 1.2589E-37 | 163.74 |
| *CRTAM* | rs2725405 | G | C | 0.441 | 0.05 | 0.007 | 2.5119E-12 | 51.02 |
| *CRTAP* | rs607929 | C | G | 0.4485 | -0.087 | 0.008 | 2.5119E-29 | 118.27 |
| *CRYBB1* | rs1354034 | T | C | 0.6045 | 0.063 | 0.008 | 1E-16 | 62.02 |
| *CRYBB1* | rs9402686 | G | A | 0.2713 | 0.07 | 0.008 | 6.3096E-17 | 76.56 |
| *CRYBB1* | rs342298 | C | T | 0.4545 | -0.081 | 0.007 | 7.9433E-28 | 133.90 |
| *CRYBB1* | rs6993770 | A | T | 0.2866 | -0.199 | 0.008 | 3.981E-129 | 618.77 |
| *CRYBB1* | rs4631576 | C | T | 0.3607 | 0.066 | 0.008 | 1.2589E-17 | 68.06 |
| *CRYBB1* | rs4783183 | C | G | 0.8741 | -0.088 | 0.011 | 1.9953E-15 | 64.00 |
| *CRYBB1* | rs4822754 | A | G | 0.5283 | -0.157 | 0.007 | 1.2589E-98 | 503.04 |
| *CRYBB2* | rs327189 | T | C | 0.1178 | -0.083 | 0.012 | 6.3096E-12 | 47.84 |
| *CRYBB2* | rs11586604 | G | T | 0.4715 | 0.059 | 0.007 | 7.9433E-16 | 71.04 |
| *CRYBB2* | rs56257618 | C | T | 0.1045 | 0.088 | 0.012 | 1.9953E-13 | 53.78 |
| *CRYBB2* | rs78247684 | G | A | 0.2462 | 0.071 | 0.009 | 1E-16 | 62.23 |
| *CRYBB2* | rs34481144 | C | T | 0.5019 | -0.076 | 0.008 | 6.3096E-24 | 90.25 |
| *CRYBB2* | rs6578313 | T | C | 0.7949 | 0.063 | 0.009 | 5.0119E-12 | 49.00 |
| *CRYBB2* | rs7136079 | T | C | 0.59 | -0.077 | 0.007 | 2.5119E-25 | 121.00 |
| *CRYBB2* | rs523175 | G | A | 0.5659 | 0.058 | 0.007 | 5.0119E-15 | 68.65 |
| *CRYBB2* | rs12050132 | A | T | 0.2403 | 0.112 | 0.009 | 7.9433E-39 | 154.86 |
| *CRYBB2* | rs5742915 | T | C | 0.4624 | -0.052 | 0.007 | 1.2589E-12 | 55.18 |
| *CRYBB2* | rs7242807 | G | C | 0.4925 | -0.081 | 0.007 | 7.9433E-29 | 133.90 |
| *CRYBB2* | rs45506792 | C | T | 0.007 | -0.391 | 0.051 | 1.9953E-14 | 58.78 |
| *CRYBB2* | rs56084239 | C | T | 0.1725 | 0.358 | 0.01 | 1E-200 | 1281.64 |
| *CRYGD* | rs201408120 | G | C | 0.021 | -0.154 | 0.023 | 1E-11 | 44.83 |
| *CRYGD* | rs1434282 | C | T | 0.7265 | 0.133 | 0.007 | 5.0119E-79 | 361.00 |
| *CRYGD* | rs34570682 | G | A | 0.3097 | -0.71 | 0.008 | 1E-200 | 7876.56 |
| *CRYGD* | rs72627509 | C | G | 0.1841 | 0.075 | 0.008 | 1.9953E-20 | 87.89 |
| *CRYGD* | rs9374080 | T | C | 0.457 | 0.058 | 0.006 | 3.1623E-20 | 93.44 |
| *CRYGD* | rs60757417 | C | G | 0.0605 | -0.134 | 0.013 | 3.1623E-24 | 106.25 |
| *CRYGD* | rs10883359 | A | G | 0.2851 | 0.049 | 0.007 | 1.9953E-12 | 49.00 |
| *CRYGD* | rs2295125 | A | G | 0.7098 | -0.053 | 0.007 | 1.2589E-14 | 57.33 |
| *CRYGD* | rs7256400 | G | A | 0.2178 | -0.084 | 0.008 | 6.3096E-29 | 110.25 |
| *CRYM* | rs2733910 | C | T | 0.2566 | -0.415 | 0.009 | 1E-200 | 2126.23 |
| *CRYZL1* | rs1354034 | T | C | 0.6045 | 0.087 | 0.008 | 7.9433E-30 | 118.27 |
| *CRYZL1* | rs6993770 | A | T | 0.2866 | -0.058 | 0.008 | 3.1623E-12 | 52.56 |
| *CRYZL1* | rs7896518 | A | G | 0.4236 | 0.067 | 0.008 | 5.0119E-18 | 70.14 |
| *CRYZL1* | rs11502185 | T | C | 0.2596 | 0.068 | 0.01 | 1.9953E-12 | 46.24 |
| *CRYZL1* | rs13050238 | C | T | 0.0316 | -0.37 | 0.021 | 1.5849E-66 | 310.43 |
| *CSDE1* | rs1354034 | T | C | 0.6037 | 0.073 | 0.008 | 3.9811E-21 | 83.27 |
| *CSDE1* | rs607929 | C | G | 0.449 | -0.096 | 0.008 | 3.1623E-35 | 144.00 |
| *CSF1* | rs11577194 | T | C | 0.5241 | -0.187 | 0.007 | 7.943E-138 | 713.65 |
| *CSF1* | rs17315646 | C | G | 0.6062 | 0.065 | 0.008 | 1.5849E-17 | 66.02 |
| *CSF1* | rs3132455 | G | C | 0.3476 | 0.06 | 0.008 | 6.3096E-15 | 56.25 |

| | | | | | | | | |
|---|---|---|---|---|---|---|---|---|
| *CSF1* | rs423135 | G | A | 0.5185 | 0.055 | 0.007 | 1.2589E-13 | 61.73 |
| *CSF1R* | rs79412885 | G | A | 0.0743 | -0.13 | 0.014 | 7.9433E-21 | 86.22 |
| *CSF1R* | rs11899659 | A | G | 0.4065 | -0.052 | 0.007 | 1.2589E-12 | 55.18 |
| *CSF1R* | rs62165726 | C | A | 0.0369 | -0.381 | 0.019 | 1.5849E-89 | 402.11 |
| *CSF1R* | rs111772691 | C | CAAAT | 0.3483 | 0.056 | 0.008 | 2.5119E-13 | 49.00 |
| *CSF1R* | rs2470750 | A | T | 0.3927 | -0.131 | 0.007 | 5.0119E-71 | 350.22 |
| *CSF1R* | rs2293329 | G | A | 0.8258 | 0.087 | 0.009 | 3.1623E-20 | 93.44 |
| *CSF1R* | rs35829610 | C | CTG | 0.3383 | -0.053 | 0.008 | 6.3096E-12 | 43.89 |
| *CSF1R* | rs56048668 | A | C | 0.0067 | -1.538 | 0.044 | 1E-200 | 1221.82 |
| *CSF1R* | rs1281944 | T | C | 0.1296 | 0.09 | 0.011 | 1E-15 | 66.94 |
| *CSF1R* | rs4841132 | A | G | 0.9081 | -0.093 | 0.012 | 3.9811E-14 | 60.06 |
| *CSF1R* | rs7865362 | C | T | 0.3631 | 0.086 | 0.008 | 1.5849E-30 | 115.56 |
| *CSF1R* | rs635634 | C | T | 0.1843 | 0.122 | 0.009 | 1.2589E-39 | 183.75 |
| *CSF1R* | rs75071241 | G | A | 0.0597 | -0.168 | 0.015 | 5.0119E-29 | 125.44 |
| *CSF1R* | rs111338191 | A | AAATT | 0.521 | -0.098 | 0.007 | 5.0119E-42 | 196.00 |
| *CSF1R* | rs766232355 | ATAAAG | A | 0.4907 | 0.056 | 0.007 | 3.9811E-15 | 64.00 |
| *CSF1R* | rs186021206 | G | A | 0.0064 | 1.563 | 0.049 | 1E-200 | 1017.48 |
| *CSF1R* | rs61267327 | C | T | 0.0248 | -0.161 | 0.023 | 1.9953E-12 | 49.00 |
| *CSF1R* | rs516316 | G | C | 0.5087 | 0.055 | 0.007 | 2.5119E-14 | 61.73 |
| *CSF1R* | rs738409 | C | G | 0.2171 | 0.066 | 0.009 | 1.9953E-14 | 53.78 |
| *CSF2* | rs2009581 | G | A | 0.2725 | -0.086 | 0.009 | 6.3096E-24 | 91.31 |
| *CSF2* | rs25882 | T | C | 0.2066 | 0.244 | 0.009 | 7.943E-149 | 735.01 |
| *CSF2* | rs9270604 | T | C | 0.3187 | 0.063 | 0.008 | 1.9953E-14 | 62.02 |
| *CSF2* | rs2967683 | T | C | 0.2365 | -0.071 | 0.009 | 1E-15 | 62.23 |
| *CSF2RA* | rs9394107 | A | G | 0.2301 | 0.069 | 0.01 | 1.2589E-11 | 47.61 |
| *CSF2RA* | rs200489612 | G | A | 0.0052 | 0.541 | 0.058 | 6.3096E-21 | 87.00 |
| *CSF2RB* | rs8176741 | G | A | 0.061 | 0.098 | 0.01 | 1.5849E-22 | 96.04 |
| *CSF2RB* | rs7310615 | C | G | 0.5175 | -0.036 | 0.005 | 6.3096E-14 | 51.84 |
| *CSF2RB* | rs200489612 | G | A | 0.0052 | 0.302 | 0.036 | 6.3096E-17 | 70.37 |
| *CSF2RB* | rs11655264 | G | C | 0.4509 | -0.041 | 0.005 | 1E-17 | 67.24 |
| *CSF2RB* | rs2239749 | A | G | 0.4586 | -1.122 | 0.008 | 1E-200 | 19670.06 |
| *CSF3* | rs3917932 | C | G | 0.5754 | -0.174 | 0.008 | 7.943E-116 | 473.06 |
| *CSF3* | rs41348 | A | G | 0.4178 | -0.053 | 0.008 | 5.0119E-12 | 43.89 |
| *CSF3* | rs14408 | T | C | 0.3661 | -0.069 | 0.008 | 1.2589E-18 | 74.39 |
| *CSF3* | rs748908389 | C | CT | 0.4709 | 0.076 | 0.008 | 3.9811E-24 | 90.25 |
| *CSF3R* | rs3917999 | C | T | 0.0106 | 1.132 | 0.038 | 1.995E-199 | 887.41 |
| *CSF3R* | rs11888817 | T | C | 0.6422 | 0.056 | 0.008 | 6.3096E-13 | 49.00 |
| *CSF3R* | rs2853951 | C | T | 0.5886 | 0.066 | 0.008 | 6.3096E-18 | 68.06 |
| *CSF3R* | rs445 | C | T | 0.0954 | -0.093 | 0.013 | 2.5119E-13 | 51.18 |
| *CSF3R* | rs10107630 | C | T | 0.5743 | -0.062 | 0.008 | 3.1623E-16 | 60.06 |
| *CSF3R* | rs240556 | C | T | 0.2395 | 0.146 | 0.009 | 6.3096E-61 | 263.16 |
| *CSF3R* | rs10774625 | A | G | 0.5041 | -0.08 | 0.007 | 3.9811E-27 | 130.61 |
| *CSF3R* | rs186021206 | G | A | 0.0063 | 0.657 | 0.052 | 1E-36 | 159.63 |
| *CSF3R* | rs3826331 | T | C | 0.5976 | -0.128 | 0.008 | 1E-62 | 256.00 |
| *CSNK1D* | rs9273369 | T | C | 0.1459 | -0.095 | 0.011 | 3.1623E-18 | 74.59 |
| *CSNK2A1* | rs1354034 | T | C | 0.6043 | 0.053 | 0.008 | 3.1623E-12 | 43.89 |
| *CSNK2A1* | rs7896518 | A | G | 0.4237 | 0.055 | 0.008 | 5.0119E-13 | 47.27 |
| *CSNK2A1* | rs61269268 | A | C | 0.1729 | 0.135 | 0.01 | 2.5119E-42 | 182.25 |
| *CSNK2A1* | rs429358 | T | C | 0.1556 | 0.214 | 0.01 | 1.5849E-94 | 457.96 |
| *CSPG4* | rs333947 | G | A | 0.1482 | -0.085 | 0.01 | 1E-17 | 72.25 |
| *CSPG4* | rs61801010 | A | G | 0.1063 | 0.084 | 0.012 | 1.9953E-12 | 49.00 |
| *CSPG4* | rs13303 | T | C | 0.5598 | 0.103 | 0.007 | 3.1623E-47 | 216.51 |
| *CSPG4* | rs76967933 | C | A | 0.0867 | -0.09 | 0.013 | 6.3096E-13 | 47.93 |
| *CSPG4* | rs1188865 | G | C | 0.8192 | -0.062 | 0.009 | 1.5849E-11 | 47.46 |
| *CSPG4* | rs2389858 | T | A | 0.4903 | -0.073 | 0.007 | 5.0119E-25 | 108.76 |
| *CSPG4* | rs56278466 | T | G | 0.6608 | 0.061 | 0.007 | 1.5849E-16 | 75.94 |
| *CSPG4* | rs3751198 | A | G | 0.6032 | 0.054 | 0.007 | 1E-13 | 59.51 |
| *CSPG4* | rs7318906 | G | A | 0.4293 | 0.056 | 0.007 | 7.9433E-15 | 64.00 |
| *CSPG4* | rs11854097 | A | G | 0.2557 | -0.134 | 0.008 | 1.2589E-61 | 280.56 |
| *CSPG4* | rs200489612 | G | A | 0.0052 | 0.472 | 0.053 | 5.0119E-19 | 79.31 |
| *CSRP3* | rs4408310 | C | A | 0.5398 | 0.057 | 0.008 | 1.5849E-13 | 50.77 |
| *CST1* | rs72694438 | G | A | 0.2075 | -0.075 | 0.009 | 3.1623E-17 | 69.44 |
| *CST1* | rs61685000 | G | C | 0.409 | -0.102 | 0.007 | 6.3096E-45 | 212.33 |
| *CST1* | rs4976271 | G | T | 0.5966 | -0.078 | 0.007 | 1.9953E-26 | 124.16 |
| *CST1* | rs745377 | G | T | 0.5228 | 0.069 | 0.007 | 3.9811E-22 | 97.16 |
| *CST1* | rs146306173 | G | GC | 0.2529 | -0.149 | 0.008 | 1.2589E-73 | 346.89 |
| *CST1* | rs1998764 | C | T | 0.5594 | 0.064 | 0.007 | 5.0119E-19 | 83.59 |
| *CST1* | rs67020211 | T | G | 0.5544 | -0.085 | 0.007 | 1.2589E-32 | 147.45 |

| | | | | | | | | |
|---|---|---|---|---|---|---|---|---|
| *CST1* | rs601338 | G | A | 0.5085 | 0.067 | 0.007 | 1E-20 | 91.61 |
| *CST1* | rs4549181 | C | T | 0.2998 | -0.583 | 0.009 | 1E-200 | 4196.16 |
| *CST3* | rs13146355 | G | A | 0.4567 | 0.065 | 0.007 | 3.9811E-21 | 86.22 |
| *CST3* | rs756700851 | CT | C | 0.7967 | 0.058 | 0.009 | 1.5849E-11 | 41.53 |
| *CST3* | rs3184504 | T | C | 0.5173 | -0.049 | 0.007 | 6.3096E-13 | 49.00 |
| *CST3* | rs77924615 | G | A | 0.197 | -0.062 | 0.009 | 1.9953E-12 | 47.46 |
| *CST3* | rs911119 | C | T | 0.7765 | 0.328 | 0.008 | 1E-200 | 1681.00 |
| *CST5* | rs72694438 | G | A | 0.207 | -0.052 | 0.007 | 3.9811E-12 | 55.18 |
| *CST5* | rs4950775 | C | T | 0.4182 | -0.05 | 0.006 | 1.9953E-16 | 69.44 |
| *CST5* | rs34639880 | G | GT | 0.4549 | 0.06 | 0.006 | 5.0119E-23 | 100.00 |
| *CST5* | rs2576233 | G | T | 0.2438 | 0.051 | 0.007 | 6.3096E-13 | 53.08 |
| *CST5* | rs3820640 | T | C | 0.159 | 0.058 | 0.008 | 1.5849E-12 | 52.56 |
| *CST5* | rs7428376 | T | C | 0.23 | -0.052 | 0.007 | 5.0119E-13 | 55.18 |
| *CST5* | rs17637318 | T | C | 0.2923 | -0.055 | 0.007 | 6.3096E-17 | 61.73 |
| *CST5* | rs4976271 | G | T | 0.5972 | -0.066 | 0.006 | 1.5849E-26 | 121.00 |
| *CST5* | rs745377 | G | T | 0.5241 | 0.053 | 0.006 | 2.5119E-18 | 78.03 |
| *CST5* | rs11054082 | G | C | 0.2512 | -0.182 | 0.007 | 6.31E-149 | 676.00 |
| *CST5* | rs67020211 | T | G | 0.5537 | -0.056 | 0.006 | 2.5119E-20 | 87.11 |
| *CST5* | rs601338 | G | A | 0.5094 | 0.097 | 0.006 | 6.3096E-58 | 261.36 |
| *CST5* | rs1799841 | A | G | 0.4357 | 0.731 | 0.007 | 1E-200 | 10905.33 |
| *CST5* | rs545804263 | C | T | 0.0072 | 0.325 | 0.045 | 3.9811E-13 | 52.16 |
| *CST6* | rs80293268 | G | C | 0.0483 | -0.387 | 0.018 | 5.012E-107 | 462.25 |
| *CST6* | rs28635651 | A | G | 0.3301 | 0.067 | 0.008 | 1.2589E-18 | 70.14 |
| *CST6* | rs1260326 | T | C | 0.6084 | 0.059 | 0.007 | 1E-15 | 71.04 |
| *CST6* | rs13015258 | T | G | 0.6211 | 0.056 | 0.007 | 6.3096E-14 | 64.00 |
| *CST6* | rs121908120 | T | A | 0.0275 | -0.191 | 0.022 | 6.3096E-18 | 75.37 |
| *CST6* | rs833270 | T | C | 0.6172 | 0.05 | 0.007 | 1E-11 | 51.02 |
| *CST6* | rs75990859 | A | T | 0.4292 | 0.049 | 0.007 | 7.9433E-12 | 49.00 |
| *CST6* | rs75680898 | A | G | 0.145 | 0.071 | 0.01 | 3.9811E-12 | 50.41 |
| *CST6* | rs114964313 | G | A | 0.3193 | 0.062 | 0.008 | 1.2589E-14 | 60.06 |
| *CST6* | rs155942 | T | C | 0.7017 | -0.059 | 0.008 | 3.9811E-14 | 54.39 |
| *CST6* | rs7905367 | G | C | 0.7976 | 0.086 | 0.009 | 1.5849E-22 | 91.31 |
| *CST6* | rs12577165 | T | C | 0.024 | 0.613 | 0.024 | 1.259E-148 | 652.38 |
| *CST6* | rs36034702 | C | T | 0.1687 | -0.114 | 0.01 | 6.3096E-32 | 129.96 |
| *CST7* | rs114694170 | T | C | 0.0602 | 0.123 | 0.012 | 3.9811E-23 | 105.06 |
| *CST7* | rs6993770 | A | T | 0.2867 | -0.152 | 0.006 | 7.943E-122 | 641.78 |
| *CST7* | rs7098181 | G | T | 0.4138 | -0.065 | 0.006 | 3.1623E-28 | 117.36 |
| *CST7* | rs12767683 | C | A | 0.3249 | 0.086 | 0.006 | 1.2589E-43 | 205.44 |
| *CST7* | rs2511241 | C | T | 0.9294 | 0.089 | 0.011 | 3.9811E-15 | 65.46 |
| *CST7* | rs35979828 | C | T | 0.0692 | -0.09 | 0.012 | 7.9433E-15 | 56.25 |
| *CST7* | rs117566084 | C | T | 0.01 | 0.295 | 0.029 | 1E-23 | 103.48 |
| *CST7* | rs3184504 | T | C | 0.5171 | -0.068 | 0.006 | 1E-31 | 128.44 |
| *CST7* | rs145078947 | G | T | 0.0026 | 0.42 | 0.058 | 3.1623E-13 | 52.44 |
| *CST7* | rs4783183 | C | G | 0.8743 | -0.079 | 0.009 | 1.5849E-19 | 77.05 |
| *CST7* | rs1045929 | C | T | 0.3882 | 0.077 | 0.006 | 3.9811E-38 | 164.69 |
| *CST7* | rs227651 | A | G | 0.231 | -0.836 | 0.009 | 1E-200 | 8628.35 |
| *CST7* | rs574174001 | C | T | 0.0059 | 1.444 | 0.05 | 1E-181 | 834.05 |
| *CTBS* | rs114165349 | G | C | 0.0234 | 0.226 | 0.024 | 7.9433E-21 | 88.67 |
| *CTBS* | rs79723172 | G | A | 0.03 | 1.378 | 0.021 | 1E-200 | 4305.86 |
| *CTBS* | rs687339 | C | T | 0.7714 | 0.155 | 0.008 | 1E-83 | 375.39 |
| *CTBS* | rs9857914 | G | A | 0.2787 | 0.051 | 0.007 | 7.9433E-12 | 53.08 |
| *CTBS* | rs72729623 | C | T | 0.1467 | -0.074 | 0.009 | 3.9811E-15 | 67.60 |
| *CTBS* | rs2169387 | A | G | 0.8989 | -0.085 | 0.011 | 1.9953E-14 | 59.71 |
| *CTBS* | rs4764823 | G | A | 0.2878 | -0.324 | 0.008 | 1E-200 | 1640.25 |
| *CTBS* | rs145078947 | G | T | 0.0025 | 0.687 | 0.066 | 5.0119E-25 | 108.35 |
| *CTBS* | rs139974673 | T | C | 0.0243 | 0.276 | 0.022 | 1.5849E-37 | 157.39 |
| *CTBS* | rs3865331 | C | T | 0.7043 | -0.06 | 0.008 | 1E-13 | 56.25 |
| *CTBS* | rs58542926 | C | T | 0.0753 | 0.111 | 0.013 | 1E-18 | 72.91 |
| *CTBS* | rs429358 | T | C | 0.1556 | -0.062 | 0.009 | 1.5849E-11 | 47.46 |
| *CTBS* | rs1800961 | C | T | 0.0324 | 0.165 | 0.019 | 1E-18 | 75.42 |
| *CTF1* | rs342298 | C | T | 0.455 | -0.054 | 0.008 | 1.9953E-12 | 45.56 |
| *CTF1* | rs11170874 | A | G | 0.3309 | 0.056 | 0.008 | 1.2589E-11 | 49.00 |
| *CTHRC1* | rs827592 | C | G | 0.5439 | -0.162 | 0.007 | 1E-124 | 535.59 |
| *CTHRC1* | rs1562782 | A | G | 0.4092 | -0.046 | 0.007 | 1.5849E-11 | 43.18 |
| *CTRB1* | rs11759956 | C | T | 0.5492 | 0.05 | 0.007 | 6.3096E-13 | 51.02 |
| *CTRB1* | rs3132568 | A | G | 0.2557 | -0.057 | 0.008 | 1.2589E-12 | 50.77 |
| *CTRB1* | rs7766106 | C | T | 0.4951 | 0.074 | 0.007 | 1.5849E-26 | 111.76 |
| *CTRB1* | rs2329566 | G | A | 0.9537 | 0.151 | 0.017 | 7.9433E-20 | 78.90 |

| | | | | | | | | |
|---|---|---|---|---|---|---|---|---|
| *CTRB1* | rs6666 | T | C | 0.579 | -0.054 | 0.007 | 1.5849E-14 | 59.51 |
| *CTRB1* | rs4921967 | G | T | 0.6866 | -0.068 | 0.007 | 1E-19 | 94.37 |
| *CTRB1* | rs2519093 | C | T | 0.1843 | -0.1 | 0.009 | 5.0119E-29 | 123.46 |
| *CTRB1* | rs4751995 | A | G | 0.5191 | -0.047 | 0.007 | 1E-11 | 45.08 |
| *CTRB1* | rs174544 | C | A | 0.3087 | 0.067 | 0.007 | 2.5119E-19 | 91.61 |
| *CTRB1* | rs61910478 | A | T | 0.2686 | -0.057 | 0.008 | 1.9953E-13 | 50.77 |
| *CTRB1* | rs8051363 | A | G | 0.7145 | 0.574 | 0.008 | 1E-200 | 5148.06 |
| *CTRB1* | rs1126464 | G | C | 0.2438 | -0.086 | 0.009 | 1.5849E-22 | 91.31 |
| *CTRB1* | rs17138478 | C | A | 0.1304 | 0.07 | 0.01 | 1E-11 | 49.00 |
| *CTRC* | rs497078 | C | T | 0.0982 | -0.569 | 0.012 | 1E-200 | 2248.34 |
| *CTRC* | rs6571015 | G | A | 0.7905 | 0.205 | 0.009 | 1.585E-127 | 518.83 |
| *CTRC* | rs112166936 | A | C | 0.4499 | -0.078 | 0.007 | 5.0119E-29 | 124.16 |
| *CTRC* | rs11778310 | A | G | 0.2937 | 0.064 | 0.008 | 3.9811E-17 | 64.00 |
| *CTRC* | rs8176743 | C | T | 0.0614 | 0.885 | 0.015 | 1E-200 | 3481.00 |
| *CTRC* | rs56278466 | T | G | 0.6615 | 0.06 | 0.007 | 1.9953E-16 | 73.47 |
| *CTRC* | rs9581943 | G | A | 0.4042 | 0.049 | 0.007 | 2.5119E-12 | 49.00 |
| *CTRC* | rs35775091 | G | A | 0.2742 | 0.061 | 0.008 | 1.9953E-15 | 58.14 |
| *CTRC* | rs72802342 | C | A | 0.0757 | -0.238 | 0.013 | 1E-71 | 335.17 |
| *CTRC* | rs529900 | G | C | 0.378 | -0.099 | 0.007 | 6.3096E-44 | 200.02 |
| *CTRC* | rs200489612 | G | A | 0.0052 | 0.521 | 0.052 | 6.3096E-24 | 100.38 |
| *CTRC* | rs601338 | G | A | 0.5097 | 0.11 | 0.007 | 1E-56 | 246.94 |
| *CTRC* | rs530223046 | A | ATTT | 0.1295 | -0.081 | 0.011 | 6.3096E-14 | 54.22 |
| *CTRL* | rs61200828 | A | C | 0.8985 | -0.133 | 0.012 | 3.1623E-28 | 122.84 |
| *CTRL* | rs2816941 | A | G | 0.2275 | 0.094 | 0.009 | 1.5849E-27 | 109.09 |
| *CTRL* | rs1260326 | T | C | 0.6083 | 0.062 | 0.007 | 3.9811E-17 | 78.45 |
| *CTRL* | rs376333472 | G | A | 0.0248 | 0.163 | 0.024 | 6.3096E-12 | 46.13 |
| *CTRL* | rs11967891 | A | T | 0.1843 | -0.075 | 0.01 | 2.5119E-14 | 56.25 |
| *CTRL* | rs6571015 | G | A | 0.7905 | 0.132 | 0.009 | 6.3096E-50 | 215.11 |
| *CTRL* | rs1578060 | G | C | 0.4562 | -0.062 | 0.007 | 1.5849E-17 | 78.45 |
| *CTRL* | rs3812304 | G | A | 0.2323 | -0.068 | 0.009 | 1.5849E-15 | 57.09 |
| *CTRL* | rs608337 | G | A | 0.9309 | -0.17 | 0.014 | 1.2589E-32 | 147.45 |
| *CTRL* | rs8176743 | C | T | 0.0616 | 0.493 | 0.015 | 1E-200 | 1080.22 |
| *CTRL* | rs56278466 | T | G | 0.661 | 0.06 | 0.008 | 2.5119E-15 | 56.25 |
| *CTRL* | rs1946515 | A | T | 0.6239 | -0.062 | 0.008 | 1.9953E-13 | 60.06 |
| *CTRL* | rs7142320 | A | C | 0.7272 | -0.057 | 0.008 | 3.1623E-12 | 50.77 |
| *CTRL* | rs28929474 | C | T | 0.0208 | -0.943 | 0.026 | 1E-200 | 1315.46 |
| *CTRL* | rs4983549 | G | A | 0.3209 | 0.106 | 0.008 | 6.3096E-42 | 175.56 |
| *CTRL* | rs20549 | A | G | 0.167 | 0.089 | 0.01 | 1E-19 | 79.21 |
| *CTRL* | rs8051363 | A | G | 0.7145 | 0.224 | 0.008 | 1.995E-166 | 784.00 |
| *CTRL* | rs529900 | G | C | 0.3788 | -0.082 | 0.008 | 5.0119E-28 | 105.06 |
| *CTRL* | rs6501457 | C | T | 0.7747 | 0.09 | 0.009 | 2.5119E-25 | 100.00 |
| *CTRL* | rs681343 | C | T | 0.5091 | 0.137 | 0.007 | 1.9953E-78 | 383.04 |
| *CTRL* | rs6020369 | T | C | 0.4656 | -0.05 | 0.007 | 3.1623E-12 | 51.02 |
| *CTSB* | rs4713570 | C | T | 0.2664 | 0.068 | 0.008 | 3.9811E-18 | 72.25 |
| *CTSB* | rs709821 | G | C | 0.2618 | 0.591 | 0.009 | 1E-200 | 4312.11 |
| *CTSB* | rs10740131 | A | T | 0.4699 | 0.048 | 0.007 | 2.5119E-12 | 47.02 |
| *CTSB* | rs41281340 | C | G | 0.0944 | -0.128 | 0.012 | 1.2589E-27 | 113.78 |
| *CTSB* | rs4764824 | C | G | 0.2885 | -0.088 | 0.008 | 1.2589E-30 | 121.00 |
| *CTSB* | rs145078947 | G | T | 0.0025 | 0.531 | 0.069 | 1E-14 | 59.22 |
| *CTSB* | rs8107974 | A | T | 0.0766 | 0.127 | 0.013 | 1.5849E-22 | 95.44 |
| *CTSC* | rs2854275 | C | A | 0.1466 | 0.118 | 0.009 | 1.5849E-41 | 171.90 |
| *CTSC* | rs9457800 | C | T | 0.1347 | -0.072 | 0.009 | 1.9953E-15 | 64.00 |
| *CTSC* | rs10822143 | C | T | 0.5015 | 0.045 | 0.006 | 1.9953E-13 | 56.25 |
| *CTSC* | rs11600158 | A | G | 0.0984 | 0.989 | 0.013 | 1E-200 | 5787.70 |
| *CTSC* | rs4764823 | G | A | 0.2878 | -0.148 | 0.007 | 1.995E-102 | 447.02 |
| *CTSC* | rs145078947 | G | T | 0.0026 | 0.855 | 0.061 | 6.3096E-45 | 196.46 |
| *CTSC* | rs139974673 | T | C | 0.0244 | 0.136 | 0.02 | 1E-11 | 46.24 |
| *CTSD* | rs2298632 | C | T | 0.5 | -0.051 | 0.007 | 1.9953E-12 | 53.08 |
| *CTSD* | rs1260326 | T | C | 0.6085 | -0.07 | 0.007 | 2.5119E-21 | 100.00 |
| *CTSD* | rs55861089 | A | G | 0.1042 | -0.507 | 0.012 | 1E-200 | 1785.06 |
| *CTSD* | rs10778152 | A | G | 0.2887 | -0.157 | 0.008 | 1.5849E-84 | 385.14 |
| *CTSD* | rs145078947 | G | T | 0.0026 | 0.491 | 0.071 | 5.0119E-12 | 47.82 |
| *CTSD* | rs58542926 | C | T | 0.0751 | 0.105 | 0.014 | 1.5849E-14 | 56.25 |
| *CTSE* | rs2422141 | C | G | 0.4887 | -0.076 | 0.008 | 6.3096E-24 | 90.25 |
| *CTSE* | rs41302235 | T | C | 0.1876 | -0.643 | 0.01 | 1E-200 | 4134.49 |
| *CTSE* | rs7606173 | G | C | 0.4324 | -0.111 | 0.007 | 1.2589E-54 | 251.45 |
| *CTSE* | rs36124964 | T | C | 0.6565 | 0.056 | 0.008 | 7.9433E-14 | 49.00 |
| *CTSE* | rs535777 | G | C | 0.1432 | -0.117 | 0.01 | 6.3096E-30 | 136.89 |

| | | | | | | | | |
|---|---|---|---|---|---|---|---|---|
| *CTSE* | rs2519093 | C | T | 0.1846 | -0.269 | 0.009 | 1E-185 | 893.35 |
| *CTSE* | rs56278466 | T | G | 0.661 | 0.065 | 0.007 | 5.0119E-18 | 86.22 |
| *CTSE* | rs681343 | C | T | 0.5089 | -0.092 | 0.007 | 3.1623E-38 | 172.73 |
| *CTSF* | rs143480126 | C | CAAAAAAA | 0.7046 | 0.056 | 0.008 | 7.9433E-12 | 49.00 |
| *CTSF* | rs1260326 | T | C | 0.6087 | -0.103 | 0.007 | 1E-44 | 216.51 |
| *CTSF* | rs59916403 | G | T | 0.3494 | -0.054 | 0.007 | 5.0119E-13 | 59.51 |
| *CTSF* | rs187325356 | T | A | 0.0076 | 0.336 | 0.042 | 1.5849E-15 | 64.00 |
| *CTSF* | rs115850161 | T | C | 0.0606 | -0.178 | 0.015 | 5.0119E-32 | 140.82 |
| *CTSF* | rs41280463 | G | A | 0.1683 | -0.066 | 0.01 | 5.0119E-12 | 43.56 |
| *CTSF* | rs17145750 | C | T | 0.16 | -0.071 | 0.01 | 3.1623E-13 | 50.41 |
| *CTSF* | rs2001945 | G | C | 0.5191 | -0.051 | 0.007 | 1.2589E-12 | 53.08 |
| *CTSF* | rs102275 | T | C | 0.3513 | -0.057 | 0.008 | 1.9953E-14 | 50.77 |
| *CTSF* | rs1044522 | G | A | 0.2407 | 0.217 | 0.008 | 1.995E-146 | 735.77 |
| *CTSF* | rs10128858 | A | G | 0.2877 | -0.281 | 0.008 | 1E-200 | 1233.77 |
| *CTSF* | rs2393791 | C | T | 0.6239 | -0.059 | 0.008 | 3.1623E-15 | 54.39 |
| *CTSF* | rs71435622 | A | AT | 0.2868 | 0.059 | 0.008 | 7.9433E-13 | 54.39 |
| *CTSF* | rs145078947 | G | T | 0.0025 | 0.78 | 0.071 | 5.0119E-28 | 120.69 |
| *CTSF* | rs139974673 | T | C | 0.0245 | 0.242 | 0.023 | 7.9433E-26 | 110.71 |
| *CTSF* | rs429358 | T | C | 0.1556 | -0.077 | 0.01 | 5.0119E-15 | 59.29 |
| *CTSH* | rs4764822 | C | T | 0.2872 | -0.096 | 0.005 | 7.9433E-73 | 368.64 |
| *CTSH* | rs60250730 | C | G | 0.1658 | 1.158 | 0.01 | 1E-200 | 13409.64 |
| *CTSL* | rs144274836 | C | T | 0.1137 | -0.092 | 0.011 | 1.5849E-15 | 69.95 |
| *CTSL* | rs3129938 | T | A | 0.8336 | -0.085 | 0.01 | 3.9811E-18 | 72.25 |
| *CTSL* | rs9497576 | G | T | 0.0746 | 0.106 | 0.014 | 1.9953E-14 | 57.33 |
| *CTSL* | rs17151689 | C | A | 0.0462 | -0.13 | 0.017 | 3.9811E-14 | 58.48 |
| *CTSL* | rs2282240 | C | T | 0.2523 | 0.091 | 0.008 | 5.0119E-27 | 129.39 |
| *CTSL* | rs868561 | G | C | 0.5039 | -0.148 | 0.007 | 2.5119E-92 | 447.02 |
| *CTSL* | rs150370599 | C | T | 0.0808 | 0.21 | 0.013 | 1.9953E-56 | 260.95 |
| *CTSL* | rs76904798 | C | T | 0.1416 | 0.104 | 0.01 | 7.9433E-24 | 108.16 |
| *CTSL* | rs10846744 | G | C | 0.142 | 0.073 | 0.01 | 1.9953E-12 | 53.29 |
| *CTSL* | rs140735783 | T | TC | 0.1844 | -0.107 | 0.01 | 3.9811E-29 | 114.49 |
| *CTSL* | rs8176396 | C | T | 0.0238 | -0.161 | 0.024 | 1.2589E-11 | 45.00 |
| *CTSL* | rs9901675 | G | A | 0.0502 | -0.14 | 0.016 | 1.9953E-17 | 76.56 |
| *CTSL* | rs77733715 | A | G | 0.009 | 0.303 | 0.038 | 3.1623E-15 | 63.58 |
| *CTSL* | rs10469365 | A | G | 0.109 | -0.138 | 0.012 | 2.5119E-32 | 132.25 |
| *CTSL* | rs200373 | T | A | 0.4887 | 0.13 | 0.007 | 1E-70 | 344.90 |
| *CTSL* | rs1065853 | G | T | 0.0814 | 0.095 | 0.013 | 1.2589E-12 | 53.40 |
| *CTSL* | rs121434529 | C | T | 0.005 | 0.405 | 0.051 | 1.2589E-15 | 63.06 |
| *CTSO* | rs583104 | G | T | 0.7731 | 0.074 | 0.009 | 1.5849E-17 | 67.60 |
| *CTSO* | rs687339 | C | T | 0.7714 | 0.088 | 0.009 | 6.3096E-24 | 95.60 |
| *CTSO* | rs2334114 | C | A | 0.2133 | -0.218 | 0.009 | 1.585E-128 | 586.72 |
| *CTSO* | rs116105128 | G | C | 0.1187 | -0.101 | 0.012 | 5.0119E-16 | 70.84 |
| *CTSO* | rs75474551 | G | T | 0.0929 | -0.244 | 0.013 | 1E-83 | 352.28 |
| *CTSO* | rs56278466 | T | G | 0.6613 | 0.055 | 0.008 | 6.3096E-13 | 47.27 |
| *CTSO* | rs174560 | T | C | 0.3146 | 0.067 | 0.008 | 1.2589E-17 | 70.14 |
| *CTSO* | rs35221426 | G | T | 0.1471 | 0.133 | 0.01 | 7.9433E-38 | 176.89 |
| *CTSO* | rs146326139 | G | C | 0.0139 | -0.237 | 0.032 | 1E-13 | 54.85 |
| *CTSO* | rs58542926 | C | T | 0.0754 | 0.254 | 0.014 | 1.5849E-75 | 329.16 |
| *CTSO* | rs738409 | C | G | 0.2172 | 0.063 | 0.009 | 7.9433E-13 | 49.00 |
| *CTSS* | rs771911677 | AAT | A | 0.475 | -0.441 | 0.008 | 1E-200 | 3038.77 |
| *CTSS* | rs9365125 | T | C | 0.1414 | 0.098 | 0.011 | 1.5849E-18 | 79.37 |
| *CTSS* | rs59602812 | A | AT | 0.3196 | -0.063 | 0.008 | 3.9811E-14 | 62.02 |
| *CTSS* | rs79751758 | C | A | 0.1063 | 0.094 | 0.013 | 6.3096E-14 | 52.28 |
| *CTSS* | rs3829241 | G | A | 0.4057 | -0.06 | 0.008 | 2.5119E-14 | 56.25 |
| *CTSS* | rs4883201 | A | G | 0.1057 | 0.167 | 0.013 | 5.0119E-40 | 165.02 |
| *CTSS* | rs3184504 | T | C | 0.5166 | -0.059 | 0.008 | 3.1623E-14 | 54.39 |
| *CTSV* | rs113292219 | C | T | 0.0526 | 0.106 | 0.015 | 7.9433E-12 | 49.94 |
| *CTSV* | rs13407583 | G | A | 0.2097 | 0.063 | 0.009 | 1.9953E-13 | 49.00 |
| *CTSV* | rs28493806 | A | G | 0.3713 | 0.05 | 0.007 | 3.1623E-12 | 51.02 |
| *CTSV* | rs35716097 | C | T | 0.296 | 0.073 | 0.008 | 6.3096E-22 | 83.27 |
| *CTSV* | rs260237 | C | T | 0.1117 | -0.09 | 0.011 | 3.1623E-16 | 66.94 |
| *CTSV* | rs9382146 | G | A | 0.5741 | -0.112 | 0.007 | 3.9811E-58 | 256.00 |
| *CTSV* | rs12686511 | C | T | 0.2068 | 0.257 | 0.009 | 3.981E-187 | 815.42 |
| *CTSV* | rs532436 | G | A | 0.1847 | -0.112 | 0.009 | 5.0119E-35 | 154.86 |
| *CTSV* | rs1427204 | A | C | 0.4542 | 0.073 | 0.007 | 3.1623E-26 | 108.76 |
| *CTSV* | rs2559850 | G | A | 0.5867 | 0.12 | 0.007 | 1.9953E-63 | 293.88 |
| *CTSV* | rs6490117 | C | G | 0.4532 | -0.051 | 0.007 | 2.5119E-13 | 53.08 |
| *CTSV* | rs1169288 | A | C | 0.3139 | -0.114 | 0.008 | 7.9433E-52 | 203.06 |

| | | | | | | | | |
|---|---|---|---|---|---|---|---|---|
| *CTSV* | rs112746916 | C | CTGTT | 0.2178 | 0.064 | 0.009 | 1E-12 | 50.57 |
| *CTSV* | rs4077451 | T | A | 0.8545 | 0.1 | 0.01 | 1.9953E-24 | 100.00 |
| *CTSV* | rs123698 | G | C | 0.605 | 0.049 | 0.007 | 2.5119E-12 | 49.00 |
| *CTSV* | rs12019136 | G | A | 0.0413 | 0.134 | 0.017 | 1E-14 | 62.13 |
| *CTSV* | rs78581838 | C | T | 0.0896 | 0.094 | 0.012 | 1.2589E-14 | 61.36 |
| *CTSZ* | rs41271951 | A | G | 0.0851 | -0.11 | 0.013 | 7.9433E-18 | 71.60 |
| *CTSZ* | rs687339 | C | T | 0.7707 | 0.075 | 0.009 | 1.9953E-18 | 69.44 |
| *CTSZ* | rs3021302 | T | C | 0.1684 | 0.069 | 0.009 | 3.9811E-13 | 58.78 |
| *CTSZ* | rs76778371 | G | A | 0.0932 | -0.083 | 0.012 | 1.2589E-11 | 47.84 |
| *CTSZ* | rs149871778 | G | GAAC | 0.1041 | 0.094 | 0.012 | 3.1623E-15 | 61.36 |
| *CTSZ* | rs10745925 | T | C | 0.2883 | -0.321 | 0.008 | 1E-200 | 1610.02 |
| *CTSZ* | rs150641790 | T | TG | 0.0747 | 0.097 | 0.014 | 1.2589E-12 | 48.01 |
| *CTSZ* | rs145078947 | G | T | 0.0025 | 0.773 | 0.071 | 2.5119E-27 | 118.53 |
| *CTSZ* | rs200210321 | A | AG | 0.0739 | 0.169 | 0.014 | 6.3096E-35 | 145.72 |
| *CTSZ* | rs429358 | T | C | 0.1558 | -0.087 | 0.01 | 7.9433E-19 | 75.69 |
| *CTSZ* | rs199825459 | A | G | 0.0084 | -1.954 | 0.04 | 1E-200 | 2386.32 |
| *CUZD1* | rs6571015 | G | A | 0.7905 | 0.123 | 0.009 | 3.1623E-41 | 186.78 |
| *CUZD1* | rs505922 | T | C | 0.3169 | 0.375 | 0.008 | 1E-200 | 2197.27 |
| *CUZD1* | rs708686 | C | T | 0.268 | 0.08 | 0.008 | 2.5119E-21 | 100.00 |
| *CWC15* | rs16921241 | A | G | 0.6825 | -0.058 | 0.008 | 7.9433E-13 | 52.56 |
| *CX3CL1* | rs114921587 | G | A | 0.0968 | 0.093 | 0.013 | 2.5119E-13 | 51.18 |
| *CX3CL1* | rs3732378 | G | A | 0.1747 | 0.084 | 0.01 | 1E-17 | 70.56 |
| *CX3CL1* | rs9264666 | G | C | 0.5549 | -0.089 | 0.007 | 1E-32 | 161.65 |
| *CX3CL1* | rs502417 | T | C | 0.7397 | 0.059 | 0.008 | 3.1623E-12 | 54.39 |
| *CX3CL1* | rs507666 | G | A | 0.1846 | -0.124 | 0.01 | 3.9811E-38 | 153.76 |
| *CX3CL1* | rs9414801 | G | A | 0.5334 | -0.055 | 0.007 | 1.9953E-13 | 61.73 |
| *CX3CL1* | rs781264602 | AAAG | A | 0.6189 | -0.232 | 0.008 | 3.981E-196 | 841.00 |
| *CX3CL1* | rs602662 | G | A | 0.5369 | 0.055 | 0.007 | 1.9953E-13 | 61.73 |
| *CXADR* | rs6705147 | C | T | 0.33 | -0.058 | 0.008 | 1.5849E-13 | 52.56 |
| *CXADR* | rs56398830 | G | A | 0.0112 | -0.414 | 0.035 | 1.2589E-32 | 139.92 |
| *CXADR* | rs12938714 | G | A | 0.3299 | 0.086 | 0.008 | 1.9953E-28 | 115.56 |
| *CXADR* | rs681343 | C | T | 0.509 | -0.177 | 0.007 | 5.012E-126 | 639.37 |
| *CXADR* | rs2849900 | T | C | 0.6681 | -0.124 | 0.008 | 7.9433E-56 | 240.25 |
| *CXCL1* | rs12075 | G | A | 0.5834 | 0.063 | 0.007 | 5.0119E-20 | 81.00 |
| *CXCL1* | rs3097411 | C | G | 0.2357 | 0.674 | 0.009 | 1E-200 | 5608.35 |
| *CXCL1* | rs16870187 | C | A | 0.738 | -0.108 | 0.009 | 2.5119E-33 | 144.00 |
| *CXCL1* | rs7080386 | C | A | 0.412 | 0.09 | 0.007 | 3.1623E-38 | 165.31 |
| *CXCL1* | rs111338191 | A | AAATT | 0.5208 | -0.048 | 0.007 | 3.1623E-12 | 47.02 |
| *CXCL10* | rs113010081 | T | C | 0.1186 | -0.081 | 0.011 | 6.3096E-13 | 54.22 |
| *CXCL10* | rs11548618 | G | A | 0.0068 | 1.73 | 0.044 | 1E-200 | 1545.92 |
| *CXCL10* | rs28366364 | C | G | 0.4017 | -0.064 | 0.007 | 1E-17 | 83.59 |
| *CXCL10* | rs3184504 | T | C | 0.5173 | -0.103 | 0.007 | 1.9953E-46 | 216.51 |
| *CXCL11* | rs76036957 | G | A | 0.0094 | 0.278 | 0.038 | 5.0119E-13 | 53.52 |
| *CXCL11* | rs35170645 | A | G | 0.5136 | -0.224 | 0.007 | 3.162E-197 | 1024.00 |
| *CXCL11* | rs13181561 | G | A | 0.7452 | 0.063 | 0.008 | 1.2589E-13 | 62.02 |
| *CXCL11* | rs11962818 | C | T | 0.1587 | -0.083 | 0.01 | 5.0119E-16 | 68.89 |
| *CXCL11* | rs139141690 | G | A | 0.0048 | 1.157 | 0.054 | 6.31E-102 | 459.07 |
| *CXCL11* | rs7080386 | C | A | 0.4122 | 0.101 | 0.007 | 1E-41 | 208.18 |
| *CXCL11* | rs11596235 | C | T | 0.3116 | -0.09 | 0.008 | 3.9811E-30 | 126.56 |
| *CXCL11* | rs3184504 | T | C | 0.5171 | -0.124 | 0.007 | 1.9953E-64 | 313.80 |
| *CXCL11* | rs1654425 | T | C | 0.8345 | 0.099 | 0.01 | 1E-23 | 98.01 |
| *CXCL12* | rs6542680 | C | T | 0.8212 | -0.073 | 0.01 | 1.9953E-13 | 53.29 |
| *CXCL12* | rs6768977 | A | G | 0.5335 | 0.052 | 0.008 | 7.9433E-12 | 42.25 |
| *CXCL12* | rs113760175 | G | A | 0.0708 | 0.133 | 0.015 | 1.9953E-19 | 78.62 |
| *CXCL12* | rs3131626 | A | G | 0.3252 | 0.061 | 0.008 | 5.0119E-13 | 58.14 |
| *CXCL12* | rs7028191 | C | T | 0.4374 | -0.053 | 0.008 | 3.1623E-12 | 43.89 |
| *CXCL12* | rs11239028 | C | T | 0.3385 | 0.195 | 0.008 | 3.162E-128 | 594.14 |
| *CXCL12* | rs7896518 | A | G | 0.4238 | 0.07 | 0.008 | 5.0119E-19 | 76.56 |
| *CXCL12* | rs3184504 | T | C | 0.5177 | -0.055 | 0.008 | 1.9953E-13 | 47.27 |
| *CXCL12* | rs892090 | T | G | 0.8336 | 0.073 | 0.01 | 6.3096E-13 | 53.29 |
| *CXCL13* | rs2053526 | A | G | 0.3974 | -0.055 | 0.008 | 1.5849E-12 | 47.27 |
| *CXCL13* | rs13204736 | G | A | 0.3482 | 0.061 | 0.008 | 6.3096E-14 | 58.14 |
| *CXCL13* | rs55730499 | C | T | 0.0823 | 0.106 | 0.014 | 1E-14 | 57.33 |
| *CXCL13* | rs111338191 | A | AAATT | 0.5209 | -0.077 | 0.008 | 2.5119E-24 | 92.64 |
| *CXCL13* | rs34557412 | A | G | 0.0069 | 0.36 | 0.046 | 3.1623E-15 | 61.25 |
| *CXCL13* | rs77542162 | A | G | 0.0229 | 0.305 | 0.025 | 7.9433E-34 | 148.84 |
| *CXCL13* | rs190712692 | G | A | 0.0538 | 0.142 | 0.017 | 1.2589E-16 | 69.77 |
| *CXCL13* | rs2412973 | C | A | 0.4429 | -0.069 | 0.008 | 3.9811E-20 | 74.39 |

| | | | | | | | | |
|---|---|---|---|---|---|---|---|---|
| *CXCL14* | rs4744 | G | A | 0.2274 | -0.07 | 0.008 | 1E-16 | 76.56 |
| *CXCL14* | rs13171268 | C | G | 0.3572 | -0.072 | 0.007 | 3.1623E-22 | 105.80 |
| *CXCL14* | rs766802806 | CTTATTGCC | C | 0.2913 | -0.067 | 0.008 | 1.2589E-17 | 70.14 |
| *CXCL16* | rs2009581 | G | A | 0.2725 | 0.176 | 0.008 | 1.585E-105 | 484.00 |
| *CXCL16* | rs4347833 | G | T | 0.528 | 0.05 | 0.007 | 3.9811E-12 | 51.02 |
| *CXCL16* | rs79806033 | G | A | 0.1948 | -0.122 | 0.009 | 5.0119E-42 | 183.75 |
| *CXCL16* | rs4449583 | C | T | 0.3268 | -0.054 | 0.008 | 1.9953E-12 | 45.56 |
| *CXCL16* | rs117030384 | A | T | 0.0223 | -0.292 | 0.024 | 6.3096E-33 | 148.03 |
| *CXCL16* | rs748113 | T | C | 0.4346 | -0.103 | 0.007 | 1E-46 | 216.51 |
| *CXCL16* | rs1250567 | T | C | 0.4531 | -0.078 | 0.007 | 1E-27 | 124.16 |
| *CXCL16* | rs3184504 | T | C | 0.5169 | 0.088 | 0.007 | 7.9433E-36 | 158.04 |
| *CXCL16* | rs748988 | A | G | 0.614 | 0.058 | 0.007 | 3.1623E-15 | 68.65 |
| *CXCL16* | rs78086290 | A | T | 0.5292 | 0.189 | 0.007 | 1.585E-148 | 729.00 |
| *CXCL16* | rs77542162 | A | G | 0.0229 | 0.304 | 0.024 | 1.9953E-36 | 160.44 |
| *CXCL16* | rs57143961 | C | CT | 0.1545 | 0.102 | 0.01 | 2.5119E-25 | 104.04 |
| *CXCL16* | rs6031302 | C | G | 0.7255 | -0.085 | 0.008 | 1.5849E-26 | 112.89 |
| *CXCL17* | rs1124 | G | A | 0.3229 | -0.07 | 0.007 | 3.1623E-23 | 100.00 |
| *CXCL17* | rs9987332 | G | A | 0.4248 | -0.048 | 0.007 | 1E-12 | 47.02 |
| *CXCL17* | rs1366050 | C | T | 0.3416 | -0.049 | 0.007 | 1.9953E-12 | 49.00 |
| *CXCL17* | rs117753190 | C | G | 0.037 | 0.171 | 0.018 | 6.3096E-22 | 90.25 |
| *CXCL17* | rs4807612 | T | C | 0.3218 | -0.064 | 0.007 | 3.9811E-19 | 83.59 |
| *CXCL17* | rs11668599 | A | G | 0.0781 | 0.118 | 0.012 | 1.5849E-21 | 96.69 |
| *CXCL3* | rs12640189 | C | T | 0.1734 | 0.068 | 0.01 | 1.5849E-11 | 46.24 |
| *CXCL3* | rs3104369 | T | C | 0.7703 | -0.101 | 0.009 | 1.9953E-29 | 125.94 |
| *CXCL3* | rs7896518 | A | G | 0.4239 | 0.07 | 0.008 | 6.3096E-20 | 76.56 |
| *CXCL5* | rs62191340 | G | A | 0.5428 | 0.052 | 0.007 | 1.2589E-13 | 55.18 |
| *CXCL5* | rs753826255 | TTTTTA | T | 0.3888 | 0.05 | 0.007 | 5.0119E-12 | 51.02 |
| *CXCL5* | rs1354034 | T | C | 0.6036 | -0.12 | 0.007 | 3.9811E-62 | 293.88 |
| *CXCL5* | rs12640189 | C | T | 0.173 | 0.084 | 0.01 | 3.9811E-18 | 70.56 |
| *CXCL5* | rs352045 | T | G | 0.8834 | -0.588 | 0.011 | 1E-200 | 2857.39 |
| *CXCL5* | rs6993770 | A | T | 0.2866 | 0.081 | 0.008 | 6.3096E-25 | 102.52 |
| *CXCL5* | rs4631576 | C | T | 0.3612 | 0.064 | 0.007 | 3.1623E-18 | 83.59 |
| *CXCL5* | rs10821545 | T | G | 0.3178 | -0.091 | 0.008 | 7.9433E-33 | 129.39 |
| *CXCL5* | rs10761731 | A | T | 0.414 | 0.216 | 0.007 | 6.31E-196 | 952.16 |
| *CXCL5* | rs1654425 | T | C | 0.834 | 0.09 | 0.01 | 3.1623E-21 | 81.00 |
| *CXCL6* | rs12075 | G | A | 0.5829 | 0.086 | 0.007 | 7.9433E-34 | 150.94 |
| *CXCL6* | rs1260326 | T | C | 0.6085 | -0.052 | 0.007 | 3.1623E-13 | 55.18 |
| *CXCL6* | rs16858768 | A | C | 0.0031 | 0.441 | 0.064 | 5.0119E-12 | 47.48 |
| *CXCL6* | rs776170649 | CT | C | 0.2053 | -0.061 | 0.009 | 3.1623E-12 | 45.94 |
| *CXCL6* | rs16850073 | C | T | 0.3755 | 0.56 | 0.008 | 1E-200 | 4900.00 |
| *CXCL6* | rs10761737 | T | C | 0.413 | 0.066 | 0.007 | 1.5849E-20 | 88.90 |
| *CXCL6* | rs111338191 | A | AAATT | 0.5213 | -0.061 | 0.007 | 7.9433E-18 | 75.94 |
| *CXCL6* | rs892090 | T | G | 0.8341 | 0.107 | 0.009 | 1E-29 | 141.35 |
| *CXCL8* | rs2241244 | G | C | 0.537 | 0.078 | 0.007 | 5.0119E-26 | 124.16 |
| *CXCL8* | rs2308604 | T | C | 0.6643 | -0.061 | 0.008 | 1.9953E-14 | 58.14 |
| *CXCL8* | rs892090 | T | G | 0.834 | 0.109 | 0.01 | 3.1623E-27 | 118.81 |
| *CXCL8* | rs12075 | G | A | 0.5831 | 0.138 | 0.007 | 6.3096E-76 | 388.65 |
| *CXCL8* | rs16858768 | A | C | 0.0031 | 0.962 | 0.068 | 3.9811E-46 | 200.14 |
| *CXCL8* | rs74468557 | G | A | 0.0544 | -0.18 | 0.017 | 1.9953E-27 | 112.11 |
| *CXCL8* | rs4541868 | C | A | 0.2736 | -0.062 | 0.008 | 7.9433E-14 | 60.06 |
| *CXCL8* | rs7080386 | C | A | 0.412 | 0.084 | 0.007 | 5.0119E-29 | 144.00 |
| *CXCL9* | rs2476601 | A | G | 0.8994 | -0.091 | 0.011 | 1.9953E-15 | 68.44 |
| *CXCL9* | rs13118503 | C | T | 0.241 | -0.149 | 0.008 | 5.0119E-77 | 346.89 |
| *CXCL9* | rs2858864 | C | T | 0.218 | 0.073 | 0.009 | 6.3096E-17 | 65.79 |
| *CXCL9* | rs4252129 | C | T | 0.0165 | -0.312 | 0.027 | 1.2589E-30 | 133.53 |
| *CXCL9* | rs3184504 | T | C | 0.5173 | -0.108 | 0.007 | 5.0119E-56 | 238.04 |
| *CYB5R2* | rs342293 | C | G | 0.4597 | -0.071 | 0.007 | 3.1623E-22 | 102.88 |
| *CYB5R2* | rs7080386 | C | A | 0.4117 | 0.052 | 0.007 | 3.1623E-12 | 55.18 |
| *CYB5R2* | rs61683338 | C | G | 0.1119 | -0.385 | 0.012 | 1E-200 | 1029.34 |
| *CYTL1* | rs11722554 | G | A | 0.0379 | -1.317 | 0.019 | 1E-200 | 4804.68 |
| *CYTL1* | rs7193058 | G | A | 0.7579 | 0.056 | 0.008 | 2.5119E-13 | 49.00 |
| *DAAM1* | rs11242109 | G | T | 0.4811 | -0.057 | 0.008 | 7.9433E-14 | 50.77 |
| *DAAM1* | rs342293 | C | G | 0.4596 | -0.057 | 0.008 | 3.9811E-14 | 50.77 |
| *DAAM1* | rs1151519 | G | C | 0.7971 | 0.106 | 0.009 | 1.5849E-29 | 138.72 |
| *DAAM1* | rs11553699 | A | G | 0.1353 | -0.1 | 0.012 | 3.9811E-18 | 69.44 |
| *DAAM1* | rs7160523 | G | T | 0.4402 | 0.056 | 0.008 | 3.9811E-13 | 49.00 |
| *DAAM1* | rs55656227 | G | A | 0.8112 | 0.067 | 0.01 | 3.9811E-12 | 44.89 |
| *DAB2* | rs6993770 | A | T | 0.2863 | -0.076 | 0.008 | 3.1623E-20 | 90.25 |

| | | | | | | | | |
|---|---|---|---|---|---|---|---|---|
| *DAB2* | rs7916868 | A | T | 0.5013 | 0.058 | 0.007 | 1.2589E-14 | 68.65 |
| *DAG1* | rs1354034 | T | C | 0.6041 | 0.053 | 0.008 | 2.5119E-12 | 43.89 |
| *DAG1* | rs111338191 | A | AAATT | 0.5211 | -0.059 | 0.008 | 5.0119E-15 | 54.39 |
| *DAG1* | rs892090 | T | G | 0.8342 | 0.074 | 0.01 | 3.1623E-13 | 54.76 |
| *DAPK2* | rs4759076 | T | C | 0.457 | 0.068 | 0.007 | 1E-19 | 94.37 |
| *DAPK2* | rs34772505 | C | T | 0.43 | 0.387 | 0.008 | 1E-200 | 2340.14 |
| *DAPP1* | rs1354034 | T | C | 0.6043 | 0.059 | 0.008 | 1E-14 | 54.39 |
| *DAPP1* | rs7688733 | A | G | 0.4777 | -0.1 | 0.008 | 6.3096E-40 | 156.25 |
| *DARS1* | rs149170955 | G | A | 0.0019 | -0.954 | 0.089 | 1E-26 | 114.90 |
| *DARS1* | rs78909033 | G | A | 0.1364 | -0.1 | 0.011 | 6.3096E-20 | 82.64 |
| *DARS1* | rs1354034 | T | C | 0.6039 | 0.068 | 0.008 | 6.3096E-19 | 72.25 |
| *DARS1* | rs342298 | C | T | 0.4545 | -0.057 | 0.008 | 5.0119E-14 | 50.77 |
| *DBH* | rs768375575 | C | CCTT | 0.0963 | 0.072 | 0.008 | 3.1623E-19 | 81.00 |
| *DBH* | rs61830291 | A | C | 0.0974 | 0.054 | 0.008 | 6.3096E-12 | 45.56 |
| *DBH* | rs201286730 | AAC | A | 0.1083 | 0.075 | 0.007 | 3.1623E-24 | 114.80 |
| *DBH* | rs1611115 | T | C | 0.794 | 1.17 | 0.009 | 1E-200 | 16900.00 |
| *DBH* | rs2420915 | G | A | 0.0859 | -0.064 | 0.008 | 7.9433E-15 | 64.00 |
| *DBH* | rs7944748 | T | G | 0.3758 | 0.039 | 0.005 | 2.5119E-16 | 60.84 |
| *DBH* | rs2160725 | A | C | 0.485 | 0.04 | 0.005 | 2.5119E-18 | 64.00 |
| *DBH* | rs56254331 | A | C | 0.1715 | 0.072 | 0.006 | 2.5119E-32 | 144.00 |
| *DBH* | rs754623 | C | T | 0.883 | 0.054 | 0.007 | 3.9811E-14 | 59.51 |
| *DBI* | rs72833212 | T | G | 0.205 | -0.326 | 0.009 | 1E-200 | 1312.05 |
| *DBI* | rs1354034 | T | C | 0.6042 | 0.051 | 0.008 | 1.5849E-11 | 40.64 |
| *DBI* | rs1671152 | T | G | 0.8409 | 0.07 | 0.01 | 5.0119E-12 | 49.00 |
| *DBN1* | rs1354034 | T | C | 0.6039 | 0.063 | 0.008 | 3.1623E-16 | 62.02 |
| *DBN1* | rs342293 | C | G | 0.4593 | -0.052 | 0.008 | 5.0119E-12 | 42.25 |
| *DBNL* | rs1354034 | T | C | 0.6041 | 0.055 | 0.008 | 3.1623E-13 | 47.27 |
| *DBNL* | rs7799945 | A | G | 0.3271 | 0.058 | 0.008 | 3.9811E-13 | 52.56 |
| *DBNL* | rs342298 | C | T | 0.4545 | -0.055 | 0.007 | 3.1623E-13 | 61.73 |
| *DBNL* | rs6993770 | A | T | 0.2867 | -0.064 | 0.008 | 6.3096E-15 | 64.00 |
| *DCBLD2* | rs12487717 | G | A | 0.0131 | -1.091 | 0.032 | 1E-200 | 1162.38 |
| *DCBLD2* | rs7356034 | G | A | 0.2817 | -0.062 | 0.008 | 7.9433E-15 | 60.06 |
| *DCBLD2* | rs149999508 | C | CG | 0.2316 | -0.073 | 0.008 | 6.3096E-18 | 83.27 |
| *DCBLD2* | rs28601761 | C | G | 0.4194 | -0.063 | 0.007 | 1.9953E-18 | 81.00 |
| *DCBLD2* | rs115478735 | A | T | 0.1839 | -0.063 | 0.009 | 5.0119E-12 | 49.00 |
| *DCBLD2* | rs56278466 | T | G | 0.6612 | 0.085 | 0.007 | 1.9953E-30 | 147.45 |
| *DCBLD2* | rs174581 | G | A | 0.3541 | 0.056 | 0.007 | 2.5119E-14 | 64.00 |
| *DCBLD2* | rs2464190 | T | C | 0.4119 | 0.048 | 0.007 | 1.5849E-11 | 47.02 |
| *DCBLD2* | rs28929474 | C | T | 0.0206 | 0.501 | 0.025 | 3.1623E-90 | 401.60 |
| *DCBLD2* | rs11078597 | T | C | 0.1879 | 0.077 | 0.009 | 1.2589E-17 | 73.20 |
| *DCBLD2* | rs9906593 | A | C | 0.2366 | 0.062 | 0.008 | 5.0119E-14 | 60.06 |
| *DCBLD2* | rs77542162 | A | G | 0.0228 | -0.199 | 0.024 | 2.5119E-17 | 68.75 |
| *DCBLD2* | rs58542926 | C | T | 0.0754 | -0.114 | 0.013 | 1E-17 | 76.90 |
| *DCBLD2* | rs1672992 | C | T | 0.8606 | 0.144 | 0.01 | 6.3096E-46 | 207.36 |
| *DCBLD2* | rs9980195 | T | C | 0.5263 | -0.048 | 0.007 | 6.3096E-12 | 47.02 |
| *DCC* | rs9807459 | G | A | 0.3069 | 0.247 | 0.008 | 3.981E-196 | 953.27 |
| *DCN* | rs1260326 | T | C | 0.6088 | 0.059 | 0.007 | 7.9433E-16 | 71.04 |
| *DCTD* | rs1354034 | T | C | 0.6044 | 0.062 | 0.008 | 6.3096E-16 | 60.06 |
| *DCTN1* | rs1339847 | G | A | 0.1093 | -0.107 | 0.012 | 1.9953E-19 | 79.51 |
| *DCTN1* | rs35505243 | A | AT | 0.0843 | -0.1 | 0.014 | 3.1623E-13 | 51.02 |
| *DCTPP1* | rs61801010 | A | G | 0.106 | 0.094 | 0.012 | 3.1623E-14 | 61.36 |
| *DCTPP1* | rs1354034 | T | C | 0.6037 | 0.055 | 0.007 | 1E-13 | 61.73 |
| *DCTPP1* | rs12342201 | G | A | 0.4767 | -0.054 | 0.007 | 1E-13 | 59.51 |
| *DCTPP1* | rs56278466 | T | G | 0.6615 | 0.095 | 0.008 | 3.9811E-35 | 141.02 |
| *DCTPP1* | rs3751198 | A | G | 0.6042 | 0.057 | 0.008 | 3.9811E-14 | 50.77 |
| *DCTPP1* | rs146094440 | C | G | 0.0132 | -0.52 | 0.035 | 7.9433E-50 | 220.73 |
| *DCUN1D1* | rs607929 | C | G | 0.4487 | -0.074 | 0.008 | 2.5119E-21 | 85.56 |
| *DCXR* | rs4979373 | C | T | 0.4907 | -0.074 | 0.007 | 7.9433E-25 | 111.76 |
| *DCXR* | rs10883451 | T | C | 0.5012 | -0.058 | 0.007 | 1.2589E-16 | 68.65 |
| *DCXR* | rs139130389 | C | CTA | 0.0726 | -0.125 | 0.014 | 3.1623E-20 | 79.72 |
| *DCXR* | rs11604580 | G | A | 0.1072 | -0.091 | 0.011 | 1.5849E-15 | 68.44 |
| *DCXR* | rs61746217 | C | T | 0.003 | -0.767 | 0.065 | 1.5849E-32 | 139.24 |
| *DCXR* | rs3747207 | G | A | 0.2152 | 0.081 | 0.009 | 3.1623E-21 | 81.00 |
| *DDAH1* | rs1498374 | C | T | 0.2067 | -0.186 | 0.009 | 6.3096E-90 | 427.11 |
| *DDAH1* | rs1434282 | C | T | 0.7262 | 0.088 | 0.009 | 5.0119E-25 | 95.60 |
| *DDAH1* | rs342293 | C | G | 0.4591 | -0.052 | 0.007 | 3.9811E-12 | 55.18 |
| *DDAH1* | rs61905116 | A | G | 0.0642 | 0.171 | 0.015 | 3.1623E-29 | 129.96 |
| *DDC* | rs34103191 | A | G | 0.6565 | -0.053 | 0.008 | 7.9433E-12 | 43.89 |

| | | | | | | | | |
|---|---|---|---|---|---|---|---|---|
| *DDC* | rs138453460 | G | A | 0.0558 | 0.153 | 0.019 | 6.3096E-16 | 64.84 |
| *DDC* | rs12112308 | T | C | 0.0161 | -1.435 | 0.03 | 1E-200 | 2288.03 |
| *DDC* | rs12342201 | G | A | 0.4766 | -0.057 | 0.007 | 3.9811E-15 | 66.31 |
| *DDC* | rs601338 | G | A | 0.5092 | -0.087 | 0.007 | 6.3096E-33 | 154.47 |
| *DDC* | rs3747207 | G | A | 0.2153 | 0.077 | 0.009 | 1.5849E-18 | 73.20 |
| *DDHD2* | rs342298 | C | T | 0.4549 | -0.061 | 0.007 | 2.5119E-16 | 75.94 |
| *DDHD2* | rs571633916 | C | CA | 0.2441 | -0.109 | 0.009 | 7.9433E-37 | 146.68 |
| *DDI2* | rs41270291 | G | A | 0.0113 | -0.264 | 0.035 | 2.5119E-14 | 56.89 |
| *DDI2* | rs34315295 | G | A | 0.1482 | 0.086 | 0.01 | 1.5849E-16 | 73.96 |
| *DDR1* | rs3095353 | T | A | 0.5864 | -0.347 | 0.007 | 1E-200 | 2457.33 |
| *DDR1* | rs4841132 | A | G | 0.9081 | -0.132 | 0.012 | 1.9953E-27 | 121.00 |
| *DDR1* | rs8176644 | C | T | 0.06 | 0.106 | 0.015 | 5.0119E-13 | 49.94 |
| *DDR1* | rs56278466 | T | G | 0.6611 | 0.061 | 0.007 | 1.2589E-16 | 75.94 |
| *DDR1* | rs78689694 | G | C | 0.1329 | -0.087 | 0.01 | 5.0119E-17 | 75.69 |
| *DDR1* | rs60309576 | G | A | 0.2578 | -0.065 | 0.008 | 3.9811E-16 | 66.02 |
| *DDR1* | rs139097404 | T | C | 0.0225 | -0.163 | 0.024 | 1E-11 | 46.13 |
| *DDR1* | rs186021206 | G | A | 0.0064 | 0.841 | 0.048 | 3.9811E-68 | 306.98 |
| *DDR1* | rs111913125 | A | AATAGGTT. | 0.1004 | 0.124 | 0.012 | 3.1623E-26 | 106.78 |
| *DDR1* | rs708686 | C | T | 0.2678 | 0.055 | 0.008 | 6.3096E-12 | 47.27 |
| *DDR1* | rs73930562 | G | A | 0.152 | -0.072 | 0.01 | 1.9953E-13 | 51.84 |
| *DDR1* | rs681343 | C | T | 0.5095 | 0.128 | 0.007 | 2.5119E-73 | 334.37 |
| *DDR1* | rs55953905 | C | T | 0.2472 | 0.115 | 0.008 | 3.9811E-45 | 206.64 |
| *DDT* | rs1006771 | G | T | 0.645 | -0.206 | 0.008 | 1.259E-158 | 663.06 |
| *DDX58* | rs1133071 | A | G | 0.2988 | 0.289 | 0.008 | 1E-200 | 1305.02 |
| *DECR1* | rs189045259 | G | A | 0.0045 | -0.953 | 0.056 | 6.3096E-64 | 289.61 |
| *DECR1* | rs6993770 | A | T | 0.2869 | -0.065 | 0.008 | 5.0119E-15 | 66.02 |
| *DECR1* | rs1671152 | T | G | 0.8405 | 0.075 | 0.01 | 5.0119E-13 | 56.25 |
| *DEFA1_DEFA1B* | rs3917932 | C | G | 0.5757 | -0.078 | 0.007 | 3.1623E-25 | 124.16 |
| *DEFA1_DEFA1B* | rs4927110 | A | G | 0.4781 | -0.051 | 0.007 | 3.9811E-12 | 53.08 |
| *DEFA1_DEFA1B* | rs61740288 | G | A | 0.0218 | 0.216 | 0.026 | 3.9811E-17 | 69.02 |
| *DEFA1_DEFA1B* | rs13063578 | T | A | 0.3975 | -0.056 | 0.007 | 7.9433E-14 | 64.00 |
| *DEFA1_DEFA1B* | rs77547572 | G | A | 0.0813 | 0.099 | 0.014 | 7.9433E-13 | 50.01 |
| *DEFA1_DEFA1B* | rs3128959 | G | A | 0.1225 | 0.192 | 0.011 | 2.5119E-64 | 304.66 |
| *DEFA1_DEFA1B* | rs915125 | C | T | 0.278 | 0.065 | 0.008 | 3.9811E-15 | 66.02 |
| *DEFA1_DEFA1B* | rs210962 | C | T | 0.2376 | -0.099 | 0.009 | 1.9953E-30 | 121.00 |
| *DEFA1_DEFA1B* | rs116236945 | C | T | 0.3865 | -0.274 | 0.009 | 1E-200 | 926.86 |
| *DEFA1_DEFA1B* | rs35440906 | C | CA | 0.1859 | 0.13 | 0.01 | 5.0119E-41 | 169.00 |
| *DEFA1_DEFA1B* | rs10103048 | A | C | 0.5947 | -0.058 | 0.008 | 6.3096E-14 | 52.56 |
| *DEFA1_DEFA1B* | rs59974297 | G | GT | 0.89 | -0.094 | 0.012 | 1.2589E-15 | 61.36 |
| *DEFA1_DEFA1B* | rs2241244 | G | C | 0.5368 | -0.113 | 0.007 | 7.9433E-54 | 260.59 |
| *DEFA1_DEFA1B* | rs145144275 | A | G | 0.009 | -0.281 | 0.039 | 5.0119E-13 | 51.91 |
| *DEFB103A_DEFB103E* | rs34931250 | C | T | 0.0606 | 0.135 | 0.016 | 1.2589E-16 | 71.19 |
| *DEFB104A_DEFB104E* | rs183772362 | C | T | 0.0306 | 0.4 | 0.021 | 2.5119E-80 | 362.81 |
| *DEFB104A_DEFB104E* | rs144072245 | G | C | 0.0125 | 0.819 | 0.035 | 2.512E-122 | 547.56 |
| *DEFB118* | rs77587796 | C | T | 0.1173 | 0.105 | 0.014 | 1E-14 | 56.25 |
| *DEFB4A_DEFB4B* | rs117797877 | C | T | 0.0243 | 0.233 | 0.026 | 3.9811E-19 | 80.31 |
| *DEFB4A_DEFB4B* | rs4831643 | G | A | 0.28 | -0.126 | 0.008 | 3.1623E-50 | 248.06 |
| *DEFB4A_DEFB4B* | rs34954997 | C | CTTCG | 0.2373 | 0.068 | 0.009 | 1.5849E-15 | 57.09 |
| *DFFA* | rs183281838 | G | T | 0.0027 | -0.649 | 0.08 | 5.0119E-16 | 65.81 |
| *DFFA* | rs1354034 | T | C | 0.6041 | 0.065 | 0.008 | 6.3096E-18 | 66.02 |
| *DFFA* | rs10733789 | T | C | 0.3132 | 0.057 | 0.008 | 1.2589E-12 | 50.77 |
| *DGCR6* | rs62143198 | G | A | 0.2099 | 0.066 | 0.01 | 3.9811E-12 | 43.56 |
| *DGCR6* | rs430899 | A | C | 0.6279 | 0.131 | 0.008 | 1.2589E-60 | 268.14 |
| *DHRS4L2* | rs146691898 | A | G | 0.0523 | -0.651 | 0.017 | 1E-200 | 1466.44 |
| *DIABLO* | rs1354034 | T | C | 0.6038 | 0.053 | 0.008 | 5.0119E-12 | 43.89 |
| *DIABLO* | rs10733789 | T | C | 0.3121 | 0.061 | 0.008 | 1.2589E-13 | 58.14 |
| *DIPK2B* | rs77515410 | C | T | 0.0415 | 0.14 | 0.018 | 3.9811E-15 | 60.49 |
| *DIPK2B* | rs56278466 | T | G | 0.661 | 0.139 | 0.008 | 1E-75 | 301.89 |
| *DIPK2B* | rs12975366 | T | C | 0.3953 | -0.055 | 0.007 | 2.5119E-14 | 61.73 |
| *DKK1* | rs12407843 | G | A | 0.1102 | 0.081 | 0.012 | 7.9433E-12 | 45.56 |
| *DKK1* | rs7618405 | C | A | 0.2049 | -0.075 | 0.009 | 5.0119E-16 | 69.44 |
| *DKK1* | rs1354034 | T | C | 0.6042 | -0.087 | 0.008 | 1E-30 | 118.27 |
| *DKK1* | rs3804753 | A | G | 0.5934 | -0.052 | 0.008 | 5.0119E-12 | 42.25 |
| *DKK1* | rs142178438 | G | GTA | 0.8127 | -0.067 | 0.01 | 2.5119E-12 | 44.89 |
| *DKK1* | rs755492124 | TAAAG | T | 0.3932 | -0.06 | 0.008 | 1.5849E-15 | 56.25 |
| *DKK1* | rs35173808 | C | A | 0.0967 | -0.087 | 0.012 | 3.9811E-12 | 52.56 |
| *DKK1* | rs6961069 | C | T | 0.4021 | 0.067 | 0.008 | 3.9811E-19 | 70.14 |
| *DKK1* | rs6993770 | A | T | 0.2865 | -0.185 | 0.008 | 7.943E-113 | 534.77 |

| | | | | | | | | |
|---|---|---|---|---|---|---|---|---|
| *DKK1* | rs7097068 | T | G | 0.4347 | -0.103 | 0.008 | 5.0119E-42 | 165.77 |
| *DKK1* | rs774510679 | ACTTTGCC | A | 0.4127 | 0.142 | 0.008 | 6.3096E-79 | 315.06 |
| *DKK1* | rs71474568 | T | TTG | 0.3398 | 0.076 | 0.008 | 2.5119E-22 | 90.25 |
| *DKK1* | rs59001897 | T | A | 0.1746 | 0.072 | 0.01 | 1.5849E-13 | 51.84 |
| *DKK1* | rs12445050 | C | T | 0.1382 | 0.101 | 0.011 | 5.0119E-21 | 84.31 |
| *DKK1* | rs34416903 | T | C | 0.2798 | 0.072 | 0.008 | 1.2589E-17 | 81.00 |
| *DKK1* | rs892090 | T | G | 0.8343 | 0.165 | 0.01 | 1E-61 | 272.25 |
| *DKK1* | rs6081565 | G | A | 0.3523 | 0.068 | 0.008 | 1.5849E-18 | 72.25 |
| *DKK3* | rs539657009 | C | T | 0.1299 | 0.1 | 0.011 | 3.1623E-21 | 82.64 |
| *DKK3* | rs2389858 | T | A | 0.4905 | -0.065 | 0.007 | 1.2589E-21 | 86.22 |
| *DKK3* | rs507666 | G | A | 0.1845 | -0.103 | 0.009 | 6.3096E-31 | 130.98 |
| *DKK3* | rs196255 | C | T | 0.4132 | -0.065 | 0.007 | 6.3096E-21 | 86.22 |
| *DKK3* | rs11022114 | G | A | 0.3153 | 0.372 | 0.008 | 1E-200 | 2162.25 |
| *DKK3* | rs1609860 | C | A | 0.0815 | 0.306 | 0.013 | 1.995E-129 | 554.06 |
| *DKK3* | rs186021206 | G | A | 0.0064 | 0.443 | 0.047 | 6.3096E-21 | 88.84 |
| *DKK4* | rs77203322 | T | C | 0.0498 | -0.55 | 0.017 | 1E-200 | 1046.71 |
| *DKK4* | rs1964528 | C | T | 0.1132 | -0.08 | 0.012 | 7.9433E-12 | 44.44 |
| *DKK4* | rs782134971 | GAAACTGC( | G | 0.2543 | -0.224 | 0.008 | 1.259E-153 | 784.00 |
| *DKK4* | rs7232 | T | A | 0.3733 | 0.073 | 0.008 | 6.3096E-22 | 83.27 |
| *DKK4* | rs77542162 | A | G | 0.023 | 0.545 | 0.025 | 1.995E-108 | 475.24 |
| *DKKL1* | rs2740488 | A | C | 0.2664 | -0.037 | 0.004 | 3.9811E-19 | 85.56 |
| *DKKL1* | rs12149545 | G | A | 0.3163 | 0.048 | 0.004 | 3.1623E-34 | 144.00 |
| *DKKL1* | rs77542162 | A | G | 0.023 | 0.39 | 0.012 | 1E-200 | 1056.25 |
| *DKKL1* | rs149615216 | C | T | 0.011 | 0.132 | 0.018 | 6.3096E-14 | 53.78 |
| *DKKL1* | rs2303759 | T | G | 0.2525 | -1.26 | 0.009 | 1E-200 | 19600.00 |
| *DLG4* | rs1354034 | T | C | 0.6043 | 0.059 | 0.008 | 1.2589E-14 | 54.39 |
| *DLG4* | rs9271344 | T | C | 0.3578 | -0.058 | 0.008 | 1.5849E-13 | 52.56 |
| *DLK1* | rs6772323 | A | C | 0.3744 | 0.05 | 0.007 | 1.5849E-12 | 51.02 |
| *DLK1* | rs2282802 | G | A | 0.56 | -0.049 | 0.007 | 7.9433E-13 | 49.00 |
| *DLK1* | rs732194 | C | T | 0.4773 | 0.056 | 0.007 | 1.9953E-16 | 64.00 |
| *DLK1* | rs10823391 | C | T | 0.4674 | -0.053 | 0.007 | 7.9433E-15 | 57.33 |
| *DLK1* | rs10893501 | A | G | 0.1329 | 0.07 | 0.01 | 3.1623E-12 | 49.00 |
| *DLK1* | rs12881545 | G | C | 0.6691 | 0.492 | 0.008 | 1E-200 | 3782.25 |
| *DLK1* | rs112001035 | G | A | 0.061 | -0.223 | 0.015 | 1.2589E-52 | 221.02 |
| *DLL1* | rs61747728 | C | T | 0.0379 | 0.178 | 0.019 | 2.5119E-21 | 87.77 |
| *DLL1* | rs9264625 | G | C | 0.6319 | -0.081 | 0.008 | 5.0119E-26 | 102.52 |
| *DLL1* | rs9356631 | A | G | 0.4089 | 0.157 | 0.007 | 1E-99 | 503.04 |
| *DLL1* | rs8176672 | C | T | 0.0612 | 0.181 | 0.015 | 6.3096E-34 | 145.60 |
| *DLL1* | rs139130389 | C | CTA | 0.0726 | -0.27 | 0.014 | 2.5119E-84 | 371.94 |
| *DLL1* | rs10850034 | T | A | 0.3624 | 0.055 | 0.008 | 3.1623E-12 | 47.27 |
| *DLL1* | rs3814995 | C | T | 0.3148 | 0.061 | 0.008 | 1.5849E-15 | 58.14 |
| *DLL1* | rs2837988 | C | A | 0.3616 | 0.092 | 0.008 | 3.9811E-34 | 132.25 |
| *DMP1* | rs3001032 | T | C | 0.3199 | 0.064 | 0.008 | 2.5119E-16 | 64.00 |
| *DMP1* | rs13303 | T | C | 0.5599 | 0.062 | 0.007 | 3.9811E-17 | 78.45 |
| *DMP1* | rs187930321 | A | C | 0.0015 | 0.715 | 0.098 | 2.5119E-13 | 53.23 |
| *DMP1* | rs139276089 | G | C | 0.5133 | -0.083 | 0.007 | 3.1623E-29 | 140.59 |
| *DMP1* | rs2731672 | T | C | 0.7437 | -0.071 | 0.008 | 1.9953E-17 | 78.77 |
| *DMP1* | rs11995824 | C | G | 0.5438 | 0.068 | 0.007 | 2.5119E-20 | 94.37 |
| *DMP1* | rs2519093 | C | T | 0.1842 | -0.072 | 0.01 | 3.1623E-14 | 51.84 |
| *DMP1* | rs12789028 | G | A | 0.2002 | -0.072 | 0.009 | 6.3096E-15 | 64.00 |
| *DMP1* | rs7137828 | C | T | 0.5155 | -0.05 | 0.007 | 7.9433E-12 | 51.02 |
| *DMP1* | rs9594738 | C | T | 0.4875 | 0.105 | 0.007 | 3.1623E-46 | 225.00 |
| *DMP1* | rs806295 | G | A | 0.5313 | -0.053 | 0.007 | 5.0119E-13 | 57.33 |
| *DMP1* | rs117068593 | C | T | 0.1869 | -0.066 | 0.009 | 2.5119E-12 | 53.78 |
| *DMP1* | rs8085237 | T | C | 0.3126 | -0.058 | 0.008 | 2.5119E-13 | 52.56 |
| *DMP1* | rs913678 | T | C | 0.3303 | -0.057 | 0.008 | 3.1623E-13 | 50.77 |
| *DNAJA2* | rs342298 | C | T | 0.4547 | -0.062 | 0.008 | 1.9953E-16 | 60.06 |
| *DNAJA4* | rs11759553 | A | T | 0.2727 | -0.086 | 0.008 | 7.9433E-26 | 115.56 |
| *DNAJA4* | rs11635166 | C | T | 0.1786 | 0.157 | 0.009 | 3.1623E-61 | 304.31 |
| *DNAJB1* | rs3811444 | C | T | 0.3323 | -0.07 | 0.008 | 1.2589E-18 | 76.56 |
| *DNAJB1* | rs1354034 | T | C | 0.6046 | 0.082 | 0.008 | 1.5849E-27 | 105.06 |
| *DNAJB1* | rs342298 | C | T | 0.4545 | -0.053 | 0.007 | 1.9953E-12 | 57.33 |
| *DNAJB1* | rs10733789 | T | C | 0.3131 | 0.057 | 0.008 | 1E-12 | 50.77 |
| *DNAJB1* | rs9612021 | T | C | 0.6694 | -0.086 | 0.008 | 3.9811E-27 | 115.56 |
| *DNAJB14* | rs1354034 | T | C | 0.6046 | 0.09 | 0.008 | 6.3096E-32 | 126.56 |
| *DNAJB14* | rs342299 | C | T | 0.4551 | -0.052 | 0.008 | 5.0119E-12 | 42.25 |
| *DNAJB14* | rs6993770 | A | T | 0.2866 | -0.057 | 0.008 | 1E-11 | 50.77 |
| *DNAJB6* | rs342298 | C | T | 0.4545 | -0.053 | 0.008 | 3.9811E-12 | 43.89 |

| | | | | | | | | |
|---|---|---|---|---|---|---|---|---|
| *DNAJB6* | rs12668458 | T | C | 0.4583 | 0.191 | 0.008 | 5.012E-134 | 570.02 |
| *DNAJB6* | rs6993770 | A | T | 0.2872 | -0.061 | 0.008 | 1.9953E-13 | 58.14 |
| *DNAJB8* | rs12668458 | T | C | 0.4572 | 0.082 | 0.008 | 1.2589E-25 | 105.06 |
| *DNAJB8* | rs112988467 | C | T | 0.2695 | 0.063 | 0.009 | 1E-12 | 49.00 |
| *DNAJC21* | rs1354034 | T | C | 0.6043 | 0.055 | 0.008 | 6.3096E-13 | 47.27 |
| *DNAJC21* | rs35176895 | T | C | 0.4419 | -0.107 | 0.008 | 6.3096E-46 | 178.89 |
| *DNAJC21* | rs9271344 | T | C | 0.3578 | -0.056 | 0.008 | 7.9433E-13 | 49.00 |
| *DNAJC21* | rs342296 | G | A | 0.4571 | -0.056 | 0.007 | 6.3096E-14 | 64.00 |
| *DNAJC21* | rs60822569 | T | TTTCTC | 0.5543 | 0.052 | 0.008 | 7.9433E-12 | 42.25 |
| *DNAJC6* | rs7527768 | A | G | 0.5849 | 0.079 | 0.008 | 1.2589E-25 | 97.52 |
| *DNAJC6* | rs342293 | C | G | 0.4595 | -0.09 | 0.008 | 1E-32 | 126.56 |
| *DNAJC6* | rs7080386 | C | A | 0.4117 | 0.089 | 0.008 | 3.9811E-31 | 123.77 |
| *DNAJC6* | rs34416903 | T | C | 0.2794 | 0.071 | 0.009 | 7.9433E-17 | 62.23 |
| *DNAJC6* | rs10418205 | A | G | 0.3733 | -0.108 | 0.008 | 1E-43 | 182.25 |
| *DNAJC9* | rs7681303 | T | C | 0.8204 | -0.066 | 0.01 | 5.0119E-12 | 43.56 |
| *DNER* | rs34412673 | A | T | 0.3318 | -0.28 | 0.008 | 1E-200 | 1225.00 |
| *DNER* | rs1713987 | A | T | 0.1853 | 0.081 | 0.009 | 3.1623E-18 | 81.00 |
| *DNER* | rs9379084 | G | A | 0.1143 | 0.085 | 0.012 | 3.9811E-13 | 50.17 |
| *DNER* | rs748543364 | AG | A | 0.3402 | 0.053 | 0.008 | 1.2589E-11 | 43.89 |
| *DNER* | rs4741849 | G | T | 0.5276 | -0.05 | 0.007 | 5.0119E-12 | 51.02 |
| *DNER* | rs62625031 | A | T | 0.0387 | 0.145 | 0.019 | 1E-14 | 58.24 |
| *DNER* | rs7298766 | A | G | 0.2945 | 0.054 | 0.008 | 5.0119E-12 | 45.56 |
| *DNER* | rs186021206 | G | A | 0.0064 | 0.455 | 0.049 | 3.1623E-20 | 86.22 |
| *DNER* | rs8127156 | C | G | 0.2555 | 0.102 | 0.008 | 1.2589E-35 | 162.56 |
| *DNM1* | rs6993770 | A | T | 0.2866 | 0.056 | 0.008 | 6.3096E-12 | 49.00 |
| *DNM1* | rs3003599 | A | G | 0.8029 | 0.321 | 0.009 | 1E-200 | 1272.11 |
| *DNM1* | rs7896518 | A | G | 0.4237 | 0.079 | 0.008 | 1.2589E-25 | 97.52 |
| *DNM3* | rs6427756 | A | G | 0.735 | -0.072 | 0.008 | 1E-17 | 81.00 |
| *DNM3* | rs13062095 | T | C | 0.3383 | -0.053 | 0.008 | 1.2589E-11 | 43.89 |
| *DNM3* | rs707388 | T | A | 0.1584 | 0.075 | 0.01 | 5.0119E-13 | 56.25 |
| *DNM3* | rs342296 | G | A | 0.4567 | -0.14 | 0.007 | 5.0119E-80 | 400.00 |
| *DNM3* | rs7896518 | A | G | 0.4237 | 0.065 | 0.008 | 5.0119E-18 | 66.02 |
| *DNM3* | rs12957249 | A | G | 0.2769 | -0.263 | 0.008 | 1E-200 | 1080.77 |
| *DNMBP* | rs114694170 | T | C | 0.0598 | 0.124 | 0.016 | 5.0119E-15 | 60.06 |
| *DNMBP* | rs7069556 | C | G | 0.078 | -0.211 | 0.014 | 1.2589E-50 | 227.15 |
| *DNMBP* | rs10876550 | G | A | 0.5582 | 0.054 | 0.007 | 3.1623E-13 | 59.51 |
| *DNPEP* | rs115897075 | G | C | 0.0182 | 0.33 | 0.028 | 3.9811E-32 | 138.90 |
| *DNPEP* | rs71309976 | C | T | 0.0107 | 0.291 | 0.039 | 3.9811E-14 | 55.67 |
| *DNPEP* | rs7974006 | A | G | 0.1027 | 0.204 | 0.013 | 3.9811E-59 | 246.25 |
| *DNPH1* | rs77321231 | C | T | 0.1043 | -0.46 | 0.012 | 1E-200 | 1469.44 |
| *DOC2B* | rs8069133 | A | G | 0.6741 | 0.067 | 0.008 | 1E-15 | 70.14 |
| *DOCK9* | rs61997961 | C | T | 0.1962 | -0.117 | 0.011 | 5.0119E-27 | 113.13 |
| *DOCK9* | rs10427551 | T | A | 0.168 | -0.165 | 0.01 | 7.9433E-57 | 272.25 |
| *DOK2* | rs342298 | C | T | 0.4549 | -0.054 | 0.007 | 6.3096E-13 | 59.51 |
| *DOK2* | rs67385415 | T | C | 0.4459 | 0.062 | 0.008 | 1.9953E-16 | 60.06 |
| *DOK2* | rs6993770 | A | T | 0.2866 | -0.09 | 0.008 | 1.5849E-27 | 126.56 |
| *DPEP1* | rs12508709 | C | T | 0.5139 | 0.044 | 0.006 | 1E-13 | 53.78 |
| *DPEP1* | rs10807578 | G | C | 0.8032 | 0.048 | 0.007 | 5.0119E-12 | 47.02 |
| *DPEP1* | rs79137699 | T | C | 0.0328 | -0.129 | 0.016 | 3.1623E-16 | 65.00 |
| *DPEP1* | rs115478735 | A | T | 0.1842 | 0.053 | 0.007 | 3.1623E-13 | 57.33 |
| *DPEP1* | rs78689694 | G | C | 0.1328 | -0.097 | 0.008 | 5.0119E-32 | 147.02 |
| *DPEP1* | rs409170 | A | G | 0.5967 | 0.799 | 0.008 | 1E-200 | 9975.02 |
| *DPEP1* | rs186021206 | G | A | 0.0064 | 0.99 | 0.038 | 1.585E-146 | 678.74 |
| *DPEP1* | rs8111600 | C | T | 0.0398 | 0.11 | 0.014 | 1.9953E-14 | 61.73 |
| *DPEP2* | rs148866368 | G | A | 0.5503 | -0.054 | 0.008 | 1.5849E-11 | 45.56 |
| *DPEP2* | rs3184504 | T | C | 0.5168 | -0.061 | 0.007 | 3.1623E-17 | 75.94 |
| *DPEP2* | rs144339907 | A | C | 0.0036 | -2.28 | 0.061 | 1E-200 | 1397.04 |
| *DPEP2* | rs186021206 | G | A | 0.0063 | 0.661 | 0.05 | 2.5119E-39 | 174.77 |
| *DPP10* | rs17355498 | G | T | 0.4956 | -0.261 | 0.007 | 1E-200 | 1390.22 |
| *DPP10* | rs73165969 | G | A | 0.1813 | 0.064 | 0.009 | 1E-11 | 50.57 |
| *DPP10* | rs4552882 | G | A | 0.2014 | -0.092 | 0.009 | 1E-24 | 104.49 |
| *DPP10* | rs2519093 | C | T | 0.1842 | -0.122 | 0.009 | 1.2589E-38 | 183.75 |
| *DPP10* | rs56278466 | T | G | 0.6618 | 0.119 | 0.008 | 3.9811E-55 | 221.27 |
| *DPP4* | rs13015258 | T | G | 0.6212 | -0.229 | 0.008 | 1E-200 | 819.39 |
| *DPP4* | rs3807865 | G | A | 0.4119 | 0.076 | 0.007 | 1.2589E-25 | 117.88 |
| *DPP4* | rs1617484 | G | A | 0.5451 | 0.059 | 0.007 | 1.5849E-16 | 71.04 |
| *DPP4* | rs17674290 | T | G | 0.1476 | 0.077 | 0.01 | 3.1623E-14 | 59.29 |
| *DPP4* | rs2682725 | T | C | 0.6006 | 0.066 | 0.007 | 5.0119E-19 | 88.90 |

| | | | | | | | | |
|---|---|---|---|---|---|---|---|---|
| *DPP6* | rs2071592 | T | A | 0.3471 | -0.06 | 0.007 | 6.3096E-16 | 73.47 |
| *DPP6* | rs2240046 | C | G | 0.0624 | 0.108 | 0.015 | 1E-12 | 51.84 |
| *DPP6* | rs3734960 | T | C | 0.2621 | 0.261 | 0.008 | 1E-200 | 1064.39 |
| *DPP6* | rs507666 | G | A | 0.1847 | -0.122 | 0.009 | 5.0119E-40 | 183.75 |
| *DPP6* | rs56278466 | T | G | 0.6611 | 0.179 | 0.008 | 6.31E-124 | 500.64 |
| *DPP6* | rs2289702 | C | T | 0.1064 | 0.093 | 0.012 | 1E-15 | 60.06 |
| *DPP7* | rs35457250 | C | T | 0.0089 | 0.374 | 0.036 | 2.5119E-25 | 107.93 |
| *DPP7* | rs13253029 | C | T | 0.6911 | 0.088 | 0.007 | 2.5119E-33 | 158.04 |
| *DPP7* | rs9414801 | G | A | 0.5336 | -0.062 | 0.007 | 7.9433E-20 | 78.45 |
| *DPP7* | rs10778152 | A | G | 0.2883 | -0.223 | 0.008 | 1E-188 | 777.02 |
| *DPP7* | rs145078947 | G | T | 0.0026 | 0.54 | 0.066 | 3.1623E-16 | 66.94 |
| *DPP7* | rs147233090 | C | T | 0.0235 | 0.164 | 0.022 | 2.5119E-13 | 55.57 |
| *DPP7* | rs58542926 | C | T | 0.0753 | 0.148 | 0.013 | 7.9433E-31 | 129.61 |
| *DPT* | rs1018454 | A | C | 0.5791 | 0.33 | 0.007 | 1E-200 | 2222.45 |
| *DPT* | rs7639267 | G | T | 0.5599 | 0.051 | 0.007 | 1.5849E-14 | 53.08 |
| *DPY30* | rs72794684 | A | G | 0.0485 | 0.133 | 0.018 | 3.1623E-14 | 54.60 |
| *DPY30* | rs6993770 | A | T | 0.2865 | -0.059 | 0.008 | 7.9433E-13 | 54.39 |
| *DPY30* | rs111338191 | A | AAATT | 0.5205 | -0.051 | 0.008 | 1E-11 | 40.64 |
| *DRAXIN* | rs6679089 | G | A | 0.0559 | -0.605 | 0.016 | 1E-200 | 1429.79 |
| *DRAXIN* | rs76529516 | A | G | 0.0273 | 0.225 | 0.021 | 5.0119E-26 | 114.80 |
| *DRAXIN* | rs13063578 | T | A | 0.3976 | -0.073 | 0.007 | 1.9953E-24 | 108.76 |
| *DRAXIN* | rs10002742 | G | A | 0.4418 | -0.049 | 0.007 | 6.3096E-12 | 49.00 |
| *DRAXIN* | rs17361097 | G | A | 0.4378 | 0.057 | 0.007 | 5.0119E-16 | 66.31 |
| *DRAXIN* | rs304153 | T | A | 0.3085 | 0.068 | 0.008 | 3.1623E-19 | 72.25 |
| *DRAXIN* | rs180798958 | G | A | 0.0177 | -0.194 | 0.027 | 3.9811E-13 | 51.63 |
| *DRAXIN* | rs4704963 | T | C | 0.0762 | -0.192 | 0.013 | 1.9953E-47 | 218.13 |
| *DRAXIN* | rs652951 | C | A | 0.4922 | -0.054 | 0.007 | 1.5849E-14 | 59.51 |
| *DRAXIN* | rs9483788 | T | C | 0.2572 | -0.058 | 0.008 | 6.3096E-13 | 52.56 |
| *DRAXIN* | rs60160896 | C | T | 0.4129 | 0.061 | 0.007 | 7.9433E-18 | 75.94 |
| *DRAXIN* | rs62621812 | G | A | 0.0211 | 0.272 | 0.025 | 1.2589E-27 | 118.37 |
| *DRAXIN* | rs2217715 | C | A | 0.5706 | 0.08 | 0.007 | 7.9433E-30 | 130.61 |
| *DRAXIN* | rs1250546 | A | G | 0.4388 | 0.048 | 0.007 | 6.3096E-12 | 47.02 |
| *DRAXIN* | rs11410806 | T | TA | 0.186 | 0.072 | 0.009 | 2.5119E-15 | 64.00 |
| *DRAXIN* | rs9927316 | C | G | 0.2387 | 0.088 | 0.008 | 7.9433E-27 | 121.00 |
| *DRAXIN* | rs7508294 | G | A | 0.3365 | 0.061 | 0.007 | 2.5119E-16 | 75.94 |
| *DRAXIN* | rs681343 | C | T | 0.5095 | 0.063 | 0.007 | 5.0119E-19 | 81.00 |
| *DRG2* | rs1354034 | T | C | 0.6046 | 0.065 | 0.008 | 1.5849E-17 | 66.02 |
| *DRG2* | rs147899431 | G | C | 0.0437 | 0.27 | 0.018 | 1.2589E-48 | 225.00 |
| *DSC2* | rs61747728 | C | T | 0.0378 | 0.188 | 0.019 | 2.5119E-23 | 97.91 |
| *DSC2* | rs8176741 | G | A | 0.0613 | 0.13 | 0.015 | 6.3096E-18 | 75.11 |
| *DSC2* | rs12359178 | G | A | 0.1447 | 0.089 | 0.01 | 5.0119E-18 | 79.21 |
| *DSC2* | rs745680717 | CCTG | C | 0.4036 | -0.055 | 0.007 | 1.2589E-13 | 61.73 |
| *DSC2* | rs111338191 | A | AAATT | 0.5209 | -0.061 | 0.007 | 5.0119E-17 | 75.94 |
| *DSC2* | rs186021206 | G | A | 0.0064 | 0.416 | 0.05 | 7.9433E-17 | 69.22 |
| *DSC2* | rs2665405 | G | A | 0.5484 | 0.103 | 0.007 | 1E-45 | 216.51 |
| *DSC2* | rs4799305 | G | A | 0.8843 | -0.342 | 0.012 | 3.981E-193 | 812.25 |
| *DSC2* | rs3814995 | C | T | 0.315 | 0.089 | 0.008 | 3.9811E-30 | 123.77 |
| *DSC2* | rs516316 | G | C | 0.51 | -0.092 | 0.007 | 1.2589E-36 | 172.73 |
| *DSCAM* | rs148701924 | T | A | 0.003 | -0.453 | 0.067 | 1.2589E-11 | 45.71 |
| *DSCAM* | rs779206978 | ATTTTTT | A | 0.2515 | 0.061 | 0.008 | 3.9811E-13 | 58.14 |
| *DSCAM* | rs9274454 | T | G | 0.1865 | -0.107 | 0.01 | 1.5849E-28 | 114.49 |
| *DSCAM* | rs144643811 | T | TCA | 0.5664 | 0.063 | 0.007 | 5.0119E-18 | 81.00 |
| *DSCAM* | rs56278466 | T | G | 0.6612 | 0.177 | 0.008 | 3.162E-120 | 489.52 |
| *DSCAM* | rs7136229 | G | A | 0.8915 | -0.089 | 0.012 | 1.2589E-14 | 55.01 |
| *DSCAM* | rs186021206 | G | A | 0.0064 | 0.497 | 0.049 | 6.3096E-24 | 102.88 |
| *DSCAM* | rs78111814 | T | C | 0.038 | 0.95 | 0.02 | 1E-200 | 2256.25 |
| *DSG2* | rs61747728 | C | T | 0.0379 | 0.155 | 0.018 | 5.0119E-17 | 74.15 |
| *DSG2* | rs13127398 | T | A | 0.0717 | 0.111 | 0.014 | 1.2589E-15 | 62.86 |
| *DSG2* | rs998584 | C | A | 0.4799 | -0.084 | 0.007 | 2.5119E-32 | 144.00 |
| *DSG2* | rs648802 | G | C | 0.5399 | -0.138 | 0.007 | 3.9811E-83 | 388.65 |
| *DSG2* | rs8176741 | G | A | 0.0611 | 0.432 | 0.015 | 3.981E-182 | 829.44 |
| *DSG2* | rs2848680 | A | G | 0.4759 | -0.218 | 0.007 | 1E-198 | 969.88 |
| *DSG2* | rs708686 | C | T | 0.2679 | 0.112 | 0.008 | 3.9811E-44 | 196.00 |
| *DSG2* | rs3814995 | C | T | 0.3153 | 0.061 | 0.008 | 1.5849E-15 | 58.14 |
| *DSG3* | rs80293268 | G | C | 0.0484 | -0.183 | 0.017 | 3.9811E-27 | 115.88 |
| *DSG3* | rs61816761 | G | A | 0.0236 | 0.243 | 0.024 | 5.0119E-24 | 102.52 |
| *DSG3* | rs1260326 | T | C | 0.6084 | 0.078 | 0.007 | 2.5119E-28 | 124.16 |
| *DSG3* | rs833270 | T | C | 0.6173 | 0.06 | 0.007 | 3.9811E-17 | 73.47 |

| | | | | | | | | |
|---|---|---|---|---|---|---|---|---|
| *DSG3* | rs13107325 | C | T | 0.0749 | 0.166 | 0.013 | 1.5849E-36 | 163.05 |
| *DSG3* | rs7905367 | G | C | 0.7973 | 0.073 | 0.009 | 6.3096E-18 | 65.79 |
| *DSG3* | rs683486 | C | A | 0.4026 | 0.064 | 0.007 | 3.9811E-19 | 83.59 |
| *DSG3* | rs2286782 | C | A | 0.2187 | -0.087 | 0.009 | 1.2589E-24 | 93.44 |
| *DSG3* | rs12306780 | A | T | 0.3479 | 0.18 | 0.007 | 3.162E-135 | 661.22 |
| *DSG3* | rs34177108 | C | A | 0.2727 | 0.058 | 0.008 | 1E-13 | 52.56 |
| *DSG3* | rs186021206 | G | A | 0.0064 | 0.795 | 0.047 | 2.5119E-63 | 286.11 |
| *DSG3* | rs7234091 | C | T | 0.1809 | -0.308 | 0.009 | 1E-200 | 1171.16 |
| *DSG3* | rs12457893 | A | C | 0.4578 | -0.049 | 0.007 | 5.0119E-12 | 49.00 |
| *DSG3* | rs3814995 | C | T | 0.3149 | 0.066 | 0.007 | 6.3096E-19 | 88.90 |
| *DSG3* | rs2682581 | C | G | 0.5291 | -0.071 | 0.007 | 5.0119E-25 | 102.88 |
| *DSG4* | rs80293268 | G | C | 0.0484 | -0.209 | 0.018 | 1E-31 | 134.82 |
| *DSG4* | rs1707115 | T | C | 0.2483 | -0.058 | 0.008 | 7.9433E-12 | 52.56 |
| *DSG4* | rs11589479 | G | A | 0.1628 | -0.078 | 0.01 | 3.9811E-15 | 60.84 |
| *DSG4* | rs1260326 | T | C | 0.6081 | 0.062 | 0.007 | 1.2589E-16 | 78.45 |
| *DSG4* | rs1345417 | C | G | 0.6037 | 0.082 | 0.008 | 3.9811E-27 | 105.06 |
| *DSG4* | rs13107325 | C | T | 0.0748 | 0.121 | 0.014 | 2.5119E-18 | 74.70 |
| *DSG4* | rs190638675 | C | T | 0.1224 | -0.083 | 0.011 | 1.5849E-13 | 56.93 |
| *DSG4* | rs2436396 | A | G | 0.7014 | -0.087 | 0.008 | 1.9953E-27 | 118.27 |
| *DSG4* | rs12203592 | C | T | 0.2177 | -0.069 | 0.009 | 1.5849E-14 | 58.78 |
| *DSG4* | rs9358325 | A | T | 0.831 | -0.08 | 0.01 | 1.9953E-16 | 64.00 |
| *DSG4* | rs1918719 | T | C | 0.1959 | -0.082 | 0.009 | 2.5119E-19 | 83.01 |
| *DSG4* | rs7905367 | G | C | 0.7972 | 0.102 | 0.009 | 1.2589E-29 | 128.44 |
| *DSG4* | rs2286782 | C | A | 0.2186 | -0.171 | 0.009 | 6.3096E-81 | 361.00 |
| *DSG4* | rs12426301 | C | T | 0.2028 | 0.122 | 0.009 | 1E-40 | 183.75 |
| *DSG4* | rs1800176 | C | T | 0.3129 | 0.071 | 0.008 | 2.5119E-19 | 78.77 |
| *DSG4* | rs7973165 | T | C | 0.2729 | 0.073 | 0.008 | 6.3096E-19 | 83.27 |
| *DSG4* | rs9896243 | C | G | 0.2162 | 0.096 | 0.009 | 1.9953E-27 | 113.78 |
| *DSG4* | rs55842605 | C | T | 0.5532 | 0.071 | 0.007 | 3.1623E-22 | 102.88 |
| *DSG4* | rs34620697 | C | T | 0.006 | -0.368 | 0.047 | 6.3096E-15 | 61.31 |
| *DSG4* | rs12457893 | A | C | 0.4578 | -0.078 | 0.007 | 5.0119E-26 | 124.16 |
| *DSG4* | rs10417997 | G | A | 0.605 | -0.051 | 0.007 | 6.3096E-12 | 53.08 |
| *DTD1* | rs1354034 | T | C | 0.6044 | 0.079 | 0.008 | 1.2589E-24 | 97.52 |
| *DTD1* | rs6993770 | A | T | 0.287 | -0.061 | 0.008 | 1.5849E-13 | 58.14 |
| *DTD1* | rs60822569 | T | TTTCTC | 0.5542 | 0.06 | 0.008 | 2.5119E-15 | 56.25 |
| *DTD1* | rs6081238 | G | A | 0.376 | -0.166 | 0.008 | 2.512E-100 | 430.56 |
| *DTX3* | rs542212699 | G | A | 0.0088 | -0.68 | 0.037 | 1.5849E-73 | 337.76 |
| *DTX3* | rs151828 | T | C | 0.547 | 0.157 | 0.007 | 1.259E-111 | 503.04 |
| *DTX3* | rs2232659 | C | T | 0.0174 | 0.184 | 0.027 | 5.0119E-12 | 46.44 |
| *DTX3* | rs2277323 | G | A | 0.2483 | -0.109 | 0.008 | 5.0119E-42 | 185.64 |
| *DTYMK* | rs28630677 | C | T | 0.0421 | -0.138 | 0.019 | 7.9433E-14 | 52.75 |
| *DTYMK* | rs1354034 | T | C | 0.6045 | 0.066 | 0.008 | 2.5119E-18 | 68.06 |
| *DTYMK* | rs6993770 | A | T | 0.2866 | -0.06 | 0.008 | 3.1623E-13 | 56.25 |
| *DTYMK* | rs4759076 | T | C | 0.4567 | 0.063 | 0.007 | 6.3096E-17 | 81.00 |
| *DUSP13* | rs6480771 | T | C | 0.4109 | -0.265 | 0.008 | 1E-200 | 1097.27 |
| *DUSP29* | rs4745762 | C | A | 0.3388 | -0.14 | 0.008 | 1.5849E-67 | 306.25 |
| *DUSP3* | rs7080386 | C | A | 0.4124 | 0.066 | 0.008 | 5.0119E-17 | 68.06 |
| *DUSP3* | rs141728140 | G | A | 0.1605 | 0.093 | 0.011 | 1.2589E-17 | 71.48 |
| *DUT* | rs1354034 | T | C | 0.6046 | 0.066 | 0.008 | 6.3096E-18 | 68.06 |
| *DUT* | rs10075801 | A | G | 0.4879 | -0.058 | 0.008 | 3.1623E-14 | 52.56 |
| *DUT* | rs6993770 | A | T | 0.2866 | -0.069 | 0.008 | 1E-16 | 74.39 |
| *DUT* | rs6580981 | G | A | 0.459 | 0.057 | 0.008 | 1E-13 | 50.77 |
| *DUT* | rs247617 | C | A | 0.3244 | -0.057 | 0.008 | 1.5849E-12 | 50.77 |
| *DUT* | rs892090 | T | G | 0.8343 | 0.071 | 0.01 | 3.9811E-12 | 50.41 |
| *DXO* | rs45531831 | C | T | 0.0289 | -1.087 | 0.022 | 1E-200 | 2441.26 |
| *DXO* | rs342298 | C | T | 0.4553 | -0.053 | 0.007 | 2.5119E-13 | 57.33 |
| *DXO* | rs6993770 | A | T | 0.2865 | -0.068 | 0.008 | 1.5849E-17 | 72.25 |
| *DYNC1H1* | rs1339847 | G | A | 0.1092 | 0.371 | 0.012 | 1E-200 | 955.84 |
| *DYNC1H1* | rs1354034 | T | C | 0.6044 | 0.072 | 0.008 | 1E-20 | 81.00 |
| *DYNLT1* | rs1339847 | G | A | 0.1091 | 1.041 | 0.012 | 1E-200 | 7525.56 |
| *DYNLT1* | rs78909033 | G | A | 0.1358 | -0.065 | 0.01 | 7.9433E-12 | 42.25 |
| *DYNLT1* | rs1354034 | T | C | 0.604 | 0.225 | 0.007 | 1E-200 | 1033.16 |
| *DYNLT1* | rs2306751 | C | G | 0.5792 | 0.081 | 0.007 | 5.0119E-34 | 133.90 |
| *DYNLT1* | rs1824872 | T | C | 0.7071 | -0.05 | 0.007 | 1.9953E-12 | 51.02 |
| *DYNLT1* | rs139141690 | G | A | 0.0047 | 0.356 | 0.048 | 1.9953E-13 | 55.01 |
| *DYNLT1* | rs342296 | G | A | 0.4566 | -0.063 | 0.007 | 6.3096E-22 | 81.00 |
| *DYNLT1* | rs11604127 | C | T | 0.2309 | 0.097 | 0.008 | 1.2589E-35 | 147.02 |
| *DYNLT1* | rs79880068 | C | T | 0.1028 | 0.089 | 0.011 | 1.2589E-16 | 65.46 |

| | | | | | | | | |
|---|---|---|---|---|---|---|---|---|
| *DYNLT1* | rs73135265 | C | A | 0.145 | -0.07 | 0.009 | 6.3096E-14 | 60.49 |
| *DYNLT3* | rs1339847 | G | A | 0.1092 | 0.387 | 0.012 | 1E-200 | 1040.06 |
| *DYNLT3* | rs1354034 | T | C | 0.6044 | 0.108 | 0.008 | 3.9811E-46 | 182.25 |
| *EBAG9* | rs1354034 | T | C | 0.6041 | 0.08 | 0.008 | 1E-25 | 100.00 |
| *EBAG9* | rs11950562 | A | C | 0.4786 | -0.052 | 0.008 | 6.3096E-12 | 42.25 |
| *EBAG9* | rs342293 | C | G | 0.459 | -0.052 | 0.008 | 3.9811E-12 | 42.25 |
| *EBAG9* | rs6993770 | A | T | 0.2865 | -0.07 | 0.008 | 5.0119E-17 | 76.56 |
| *EBAG9* | rs2022934 | G | A | 0.3857 | -0.069 | 0.008 | 6.3096E-19 | 74.39 |
| *EBI3_IL27* | rs4970836 | G | A | 0.7733 | 0.071 | 0.008 | 7.9433E-18 | 78.77 |
| *EBI3_IL27* | rs10183338 | G | T | 0.2916 | -0.054 | 0.008 | 1.5849E-12 | 45.56 |
| *EBI3_IL27* | rs62165726 | C | A | 0.037 | -0.277 | 0.018 | 1.9953E-51 | 236.82 |
| *EBI3_IL27* | rs11456863 | A | AT | 0.3515 | 0.051 | 0.007 | 3.1623E-12 | 53.08 |
| *EBI3_IL27* | rs4234589 | A | G | 0.1386 | 0.07 | 0.01 | 3.9811E-12 | 49.00 |
| *EBI3_IL27* | rs11713634 | G | A | 0.2787 | -0.131 | 0.008 | 6.3096E-65 | 268.14 |
| *EBI3_IL27* | rs5743618 | C | A | 0.2239 | 0.116 | 0.008 | 1.9953E-43 | 210.25 |
| *EBI3_IL27* | rs4713572 | T | C | 0.4002 | 0.066 | 0.007 | 1.2589E-20 | 88.90 |
| *EBI3_IL27* | rs2529440 | C | T | 0.4442 | 0.051 | 0.007 | 3.1623E-13 | 53.08 |
| *EBI3_IL27* | rs339060 | A | G | 0.5531 | 0.062 | 0.007 | 5.0119E-19 | 78.45 |
| *EBI3_IL27* | rs4986790 | A | G | 0.0596 | -0.118 | 0.015 | 6.3096E-16 | 61.88 |
| *EBI3_IL27* | rs4486555 | G | T | 0.4097 | -0.194 | 0.007 | 3.162E-164 | 768.08 |
| *EBI3_IL27* | rs6589939 | A | G | 0.378 | -0.058 | 0.007 | 3.1623E-16 | 68.65 |
| *EBI3_IL27* | rs12832263 | C | T | 0.144 | 0.072 | 0.01 | 3.9811E-13 | 51.84 |
| *EBI3_IL27* | rs58298943 | C | T | 0.0846 | 0.089 | 0.012 | 1E-12 | 55.01 |
| *EBI3_IL27* | rs3184504 | T | C | 0.5166 | -0.08 | 0.007 | 3.9811E-31 | 130.61 |
| *EBI3_IL27* | rs186021206 | G | A | 0.0064 | 0.826 | 0.048 | 1.9953E-67 | 296.13 |
| *EBI3_IL27* | rs704 | G | A | 0.473 | -0.098 | 0.007 | 2.5119E-45 | 196.00 |
| *EBI3_IL27* | rs8109160 | C | T | 0.2899 | -0.413 | 0.008 | 1E-200 | 2665.14 |
| *EBI3_IL27* | rs4804669 | A | G | 0.7803 | -0.099 | 0.009 | 7.9433E-30 | 121.00 |
| *EBI3_IL27* | rs7264079 | G | C | 0.1367 | 0.098 | 0.01 | 1.9953E-22 | 96.04 |
| *ECE1* | rs181637959 | A | T | 0.0064 | -1.028 | 0.048 | 6.31E-101 | 458.67 |
| *ECE1* | rs147953260 | G | A | 0.0026 | 0.566 | 0.075 | 5.0119E-14 | 56.95 |
| *ECE1* | rs181242111 | G | A | 0.1176 | 0.086 | 0.013 | 7.9433E-12 | 43.76 |
| *ECE1* | rs35166255 | G | A | 0.0336 | 0.227 | 0.021 | 2.5119E-27 | 116.85 |
| *ECE1* | rs186021206 | G | A | 0.0064 | 0.538 | 0.052 | 3.1623E-25 | 107.04 |
| *ECHDC3* | rs1497406 | A | G | 0.5797 | 0.047 | 0.007 | 7.9433E-12 | 45.08 |
| *ECHDC3* | rs1260326 | T | C | 0.6086 | -0.051 | 0.007 | 3.1623E-13 | 53.08 |
| *ECHDC3* | rs4835265 | C | A | 0.1562 | 0.116 | 0.009 | 1.9953E-34 | 166.12 |
| *ECHDC3* | rs72781910 | G | A | 0.0922 | -0.955 | 0.013 | 1E-200 | 5396.60 |
| *ECHDC3* | rs12373325 | T | C | 0.798 | -0.086 | 0.009 | 1E-23 | 91.31 |
| *ECHS1* | rs6993770 | A | T | 0.2866 | -0.057 | 0.008 | 7.9433E-12 | 50.77 |
| *ECHS1* | rs1049951 | G | A | 0.9438 | -0.447 | 0.017 | 2.512E-160 | 691.38 |
| *ECI2* | rs660560 | A | G | 0.3252 | -0.278 | 0.008 | 1E-200 | 1207.56 |
| *ECM1* | rs78448142 | C | T | 0.0325 | -0.148 | 0.022 | 1E-11 | 45.26 |
| *ECM1* | rs3737240 | C | T | 0.3944 | -0.677 | 0.008 | 1E-200 | 7161.39 |
| *ECM1* | rs111856038 | G | A | 0.1078 | 0.075 | 0.011 | 1E-11 | 46.49 |
| *ECM1* | rs451637 | C | T | 0.3097 | -0.052 | 0.008 | 3.9811E-12 | 42.25 |
| *EDA2R* | rs4335411 | G | A | 0.7599 | -0.062 | 0.007 | 2.5119E-17 | 78.45 |
| *EDA2R* | rs10069690 | C | T | 0.2592 | -0.05 | 0.007 | 7.9433E-14 | 51.02 |
| *EDA2R* | rs78378222 | T | G | 0.0128 | -0.26 | 0.027 | 2.5119E-22 | 92.73 |
| *EDAR* | rs72665955 | G | A | 0.119 | 0.129 | 0.011 | 1E-31 | 137.53 |
| *EDAR* | rs12712870 | A | G | 0.6905 | 0.092 | 0.008 | 5.0119E-31 | 132.25 |
| *EDAR* | rs140661471 | G | A | 0.0137 | 1.66 | 0.032 | 1E-200 | 2691.02 |
| *EDAR* | rs28362642 | A | G | 0.1124 | 0.078 | 0.011 | 7.9433E-12 | 50.28 |
| *EDAR* | rs1420476 | T | A | 0.9554 | -0.141 | 0.017 | 3.1623E-16 | 68.79 |
| *EDAR* | rs10446711 | A | G | 0.1737 | 0.065 | 0.009 | 5.0119E-12 | 52.16 |
| *EDAR* | rs114694170 | T | C | 0.0598 | -0.206 | 0.015 | 6.3096E-42 | 188.60 |
| *EDAR* | rs139141690 | G | A | 0.0048 | -0.379 | 0.053 | 6.3096E-13 | 51.14 |
| *EDAR* | rs67036916 | G | A | 0.4557 | 0.072 | 0.007 | 1.2589E-23 | 105.80 |
| *EDAR* | rs6993770 | A | T | 0.2869 | 0.227 | 0.008 | 5.012E-180 | 805.14 |
| *EDAR* | rs4742045 | T | G | 0.4134 | -0.053 | 0.007 | 2.5119E-13 | 57.33 |
| *EDAR* | rs10820606 | A | C | 0.2292 | -0.06 | 0.009 | 3.9811E-12 | 44.44 |
| *EDAR* | rs10821548 | C | T | 0.3242 | -0.106 | 0.008 | 2.5119E-44 | 175.56 |
| *EDAR* | rs7080386 | C | A | 0.4121 | 0.117 | 0.007 | 3.1623E-58 | 279.37 |
| *EDAR* | rs12762934 | C | T | 0.3248 | -0.147 | 0.008 | 1.2589E-82 | 337.64 |
| *EDAR* | rs652963 | C | T | 0.8434 | 0.084 | 0.01 | 1E-17 | 70.56 |
| *EDAR* | rs780615646 | AC | A | 0.7831 | -0.063 | 0.009 | 3.1623E-12 | 49.00 |
| *EDAR* | rs11553699 | A | G | 0.1355 | -0.098 | 0.011 | 2.5119E-19 | 79.37 |
| *EDAR* | rs551672892 | G | GA | 0.2121 | -0.064 | 0.009 | 3.1623E-13 | 50.57 |

| | | | | | | | | |
|---|---|---|---|---|---|---|---|---|
| *EDAR* | rs1820994 | A | G | 0.7871 | 0.104 | 0.009 | 6.3096E-33 | 133.53 |
| *EDAR* | rs8057254 | T | A | 0.1897 | 0.077 | 0.009 | 5.0119E-17 | 73.20 |
| *EDAR* | rs74035509 | C | T | 0.0779 | -0.205 | 0.013 | 1.2589E-52 | 248.67 |
| *EDAR* | rs111527738 | A | G | 0.0206 | -0.235 | 0.025 | 7.9433E-21 | 88.36 |
| *EDDM3B* | rs3827905 | G | A | 0.6685 | -0.16 | 0.005 | 1E-200 | 1024.00 |
| *EDF1* | rs111338191 | A | AAATT | 0.5211 | -0.053 | 0.008 | 6.3096E-12 | 43.89 |
| *EDIL3* | rs56210800 | C | G | 0.1304 | -0.161 | 0.011 | 1.5849E-47 | 214.22 |
| *EDIL3* | rs111338191 | A | AAATT | 0.5212 | 0.052 | 0.008 | 7.9433E-12 | 42.25 |
| *EDIL3* | rs78606665 | G | A | 0.0607 | 0.108 | 0.016 | 6.3096E-12 | 45.56 |
| *EDIL3* | rs112001035 | G | A | 0.0609 | 0.259 | 0.016 | 1.9953E-59 | 262.04 |
| *EDN1* | rs5370 | G | T | 0.2202 | 0.272 | 0.009 | 5.012E-198 | 913.38 |
| *EEF1D* | rs3811444 | C | T | 0.3324 | -0.123 | 0.008 | 2.5119E-55 | 236.39 |
| *EEF1D* | rs1354034 | T | C | 0.6045 | 0.08 | 0.008 | 1.9953E-26 | 100.00 |
| *EEF1D* | rs342298 | C | T | 0.4545 | -0.05 | 0.007 | 1E-11 | 51.02 |
| *EFCAB14* | rs562442365 | G | A | 0.0084 | -0.529 | 0.043 | 2.5119E-34 | 151.35 |
| *EFCAB14* | rs35117964 | A | G | 0.1909 | -0.102 | 0.01 | 3.9811E-26 | 104.04 |
| *EFCAB14* | rs7166599 | A | G | 0.3341 | 0.056 | 0.008 | 5.0119E-13 | 49.00 |
| *EFCAB2* | rs61844237 | G | C | 0.4694 | -0.384 | 0.008 | 1E-200 | 2304.00 |
| *EFCAB2* | rs1354034 | T | C | 0.6041 | 0.106 | 0.007 | 2.5119E-45 | 229.31 |
| *EFCAB2* | rs342298 | C | T | 0.4552 | -0.056 | 0.007 | 5.0119E-14 | 64.00 |
| *EFCAB2* | rs11502185 | T | C | 0.2596 | 0.076 | 0.009 | 1E-15 | 71.31 |
| *EFEMP1* | rs188468174 | C | T | 0.0143 | -0.244 | 0.029 | 6.3096E-17 | 70.79 |
| *EFEMP1* | rs1260326 | T | C | 0.6084 | 0.053 | 0.007 | 3.9811E-14 | 57.33 |
| *EFEMP1* | rs3791679 | A | G | 0.2263 | 0.226 | 0.008 | 1.259E-166 | 798.06 |
| *EFEMP1* | rs2070634 | T | G | 0.5148 | 0.05 | 0.007 | 1.5849E-13 | 51.02 |
| *EFHD1* | rs146096867 | C | T | 0.084 | 0.325 | 0.013 | 3.981E-137 | 625.00 |
| *EFHD1* | rs777233686 | AG | A | 0.4123 | 0.128 | 0.007 | 7.9433E-71 | 334.37 |
| *EFNA1* | rs4390169 | A | G | 0.5187 | 0.317 | 0.008 | 1E-200 | 1570.14 |
| *EFNA1* | rs28817415 | C | T | 0.4569 | 0.06 | 0.007 | 3.1623E-16 | 73.47 |
| *EFNA1* | rs174578 | T | A | 0.3529 | 0.055 | 0.008 | 3.9811E-13 | 47.27 |
| *EFNA1* | rs28929474 | C | T | 0.0207 | 0.184 | 0.026 | 6.3096E-13 | 50.08 |
| *EFNA4* | rs148289726 | C | T | 0.0025 | -1.304 | 0.078 | 1.5849E-62 | 279.49 |
| *EFNA4* | rs6737671 | T | C | 0.5131 | 0.059 | 0.007 | 1.2589E-15 | 71.04 |
| *EFNA4* | rs3184504 | T | C | 0.5169 | -0.063 | 0.007 | 2.5119E-17 | 81.00 |
| *EFNA4* | rs7187875 | A | G | 0.541 | -0.06 | 0.007 | 3.9811E-16 | 73.47 |
| *EFNA4* | rs12934455 | C | T | 0.1665 | -0.078 | 0.01 | 7.9433E-15 | 60.84 |
| *EFNA4* | rs4760 | A | G | 0.156 | 0.089 | 0.01 | 1.5849E-18 | 79.21 |
| *EFNB2* | rs2845705 | A | T | 0.7967 | 0.186 | 0.01 | 3.1623E-83 | 345.96 |
| *EGF* | rs755492124 | TAAAG | T | 0.3934 | -0.07 | 0.008 | 1.9953E-19 | 76.56 |
| *EGF* | rs3831508 | G | GT | 0.4018 | 0.134 | 0.008 | 3.9811E-67 | 280.56 |
| *EGF* | rs2631367 | C | G | 0.5184 | 0.063 | 0.008 | 7.9433E-17 | 62.02 |
| *EGF* | rs6961069 | C | T | 0.4023 | 0.065 | 0.008 | 3.1623E-17 | 66.02 |
| *EGF* | rs6993770 | A | T | 0.2867 | -0.095 | 0.008 | 3.9811E-30 | 141.02 |
| *EGF* | rs10820606 | A | C | 0.2298 | -0.069 | 0.009 | 3.1623E-14 | 58.78 |
| *EGF* | rs7896518 | A | G | 0.4243 | 0.089 | 0.008 | 1E-30 | 123.77 |
| *EGF* | rs1362213 | C | T | 0.4796 | -0.058 | 0.008 | 5.0119E-14 | 52.56 |
| *EGF* | rs12445050 | C | T | 0.1381 | 0.091 | 0.011 | 6.3096E-17 | 68.44 |
| *EGF* | rs892090 | T | G | 0.8342 | 0.147 | 0.01 | 3.9811E-47 | 216.09 |
| *EGF* | rs6081565 | G | A | 0.3525 | 0.061 | 0.008 | 1.5849E-14 | 58.14 |
| *EGFL7* | rs78945826 | A | C | 0.0313 | -0.152 | 0.022 | 6.3096E-12 | 47.74 |
| *EGFL7* | rs823066 | A | G | 0.9487 | -0.115 | 0.017 | 3.1623E-12 | 45.76 |
| *EGFL7* | rs78909033 | G | A | 0.1362 | -0.074 | 0.011 | 3.1623E-12 | 45.26 |
| *EGFL7* | rs114694170 | T | C | 0.06 | 0.108 | 0.015 | 3.1623E-12 | 51.84 |
| *EGFL7* | rs3131622 | T | G | 0.4498 | 0.066 | 0.007 | 1.2589E-19 | 88.90 |
| *EGFL7* | rs6796 | T | C | 0.2788 | -0.166 | 0.008 | 7.9433E-92 | 430.56 |
| *EGFL7* | rs10259992 | G | A | 0.374 | -0.083 | 0.008 | 1.9953E-28 | 107.64 |
| *EGFL7* | rs17154155 | G | T | 0.4075 | 0.051 | 0.007 | 3.9811E-12 | 53.08 |
| *EGFL7* | rs339061 | T | C | 0.5548 | -0.056 | 0.007 | 3.1623E-14 | 64.00 |
| *EGFL7* | rs883006 | C | G | 0.5624 | -0.069 | 0.007 | 7.9433E-21 | 97.16 |
| *EGFL7* | rs8193003 | T | C | 0.6081 | 0.058 | 0.008 | 1.9953E-14 | 52.56 |
| *EGFL7* | rs74557797 | G | T | 0.0934 | -0.285 | 0.014 | 6.31E-99 | 414.41 |
| *EGFL7* | rs10761731 | A | T | 0.4146 | 0.068 | 0.007 | 5.0119E-20 | 94.37 |
| *EGFL7* | rs55646096 | C | T | 0.1775 | -0.163 | 0.01 | 5.0119E-65 | 265.69 |
| *EGFL7* | rs9635249 | A | G | 0.794 | 0.076 | 0.009 | 1E-16 | 71.31 |
| *EGFL7* | rs74035509 | C | T | 0.0784 | 0.107 | 0.014 | 5.0119E-15 | 58.41 |
| *EGFL7* | rs892090 | T | G | 0.8338 | 0.115 | 0.01 | 1E-31 | 132.25 |
| *EGFLAM* | rs113400125 | C | T | 0.0263 | 1.456 | 0.023 | 1E-200 | 4007.44 |
| *EGFLAM* | rs628064 | A | T | 0.4573 | -0.05 | 0.007 | 7.9433E-14 | 51.02 |

| | | | | | | | | |
|---|---|---|---|---|---|---|---|---|
| *EGFLAM* | rs12975366 | T | C | 0.3955 | 0.089 | 0.007 | 3.9811E-39 | 161.65 |
| *EGFR* | rs10081281 | C | T | 0.4534 | 0.171 | 0.007 | 7.943E-121 | 596.76 |
| *EGFR* | rs199922514 | A | AAGAAGAA | 0.9076 | -0.113 | 0.012 | 1E-19 | 88.67 |
| *EGFR* | rs3967200 | C | T | 0.1346 | -0.09 | 0.011 | 2.5119E-17 | 66.94 |
| *EGFR* | rs540639423 | C | T | 0.0087 | -0.285 | 0.041 | 1.9953E-12 | 48.32 |
| *EGFR* | rs186021206 | G | A | 0.0064 | 0.622 | 0.049 | 2.5119E-36 | 161.13 |
| *EGLN1* | rs1434282 | C | T | 0.7261 | 0.083 | 0.008 | 1.5849E-23 | 107.64 |
| *EGLN1* | rs77466488 | A | G | 0.0355 | 0.337 | 0.02 | 6.3096E-65 | 283.92 |
| *EGLN1* | rs915652 | G | A | 0.1306 | -0.075 | 0.011 | 3.9811E-12 | 46.49 |
| *EGLN1* | rs342296 | G | A | 0.4565 | -0.076 | 0.007 | 3.9811E-25 | 117.88 |
| *EHBP1* | rs1354034 | T | C | 0.6045 | 0.055 | 0.008 | 2.5119E-13 | 47.27 |
| *EHBP1* | rs6993770 | A | T | 0.2866 | -0.071 | 0.008 | 1.9953E-18 | 78.77 |
| *EHBP1* | rs4759076 | T | C | 0.4567 | 0.064 | 0.007 | 1E-17 | 83.59 |
| *EHD3* | rs592039 | G | A | 0.7087 | -0.193 | 0.009 | 1.259E-113 | 459.86 |
| *EHD3* | rs10820606 | A | C | 0.2288 | -0.078 | 0.009 | 3.1623E-17 | 75.11 |
| *EIF1AX* | rs9271459 | T | C | 0.5016 | -0.074 | 0.008 | 6.3096E-20 | 85.56 |
| *EIF1AX* | rs12895651 | T | C | 0.7005 | -0.064 | 0.009 | 2.5119E-12 | 50.57 |
| *EIF2AK2* | rs10187017 | T | C | 0.4727 | 0.15 | 0.008 | 1.9953E-88 | 351.56 |
| *EIF2AK2* | rs1354034 | T | C | 0.6047 | 0.083 | 0.008 | 1.2589E-27 | 107.64 |
| *EIF2AK3* | rs6547787 | T | G | 0.3401 | -0.121 | 0.008 | 7.9433E-52 | 228.77 |
| *EIF2AK3* | rs1354034 | T | C | 0.6037 | 0.071 | 0.008 | 3.9811E-20 | 78.77 |
| *EIF2AK3* | rs6993770 | A | T | 0.2865 | -0.062 | 0.008 | 7.9433E-14 | 60.06 |
| *EIF2S2* | rs607929 | C | G | 0.4487 | -0.073 | 0.008 | 6.3096E-21 | 83.27 |
| *EIF4B* | rs1354034 | T | C | 0.6041 | 0.084 | 0.008 | 5.0119E-28 | 110.25 |
| *EIF4B* | rs342298 | C | T | 0.4544 | -0.059 | 0.008 | 7.9433E-15 | 54.39 |
| *EIF4B* | rs10733789 | T | C | 0.3128 | 0.06 | 0.008 | 1.5849E-13 | 56.25 |
| *EIF4B* | rs11604127 | C | T | 0.2309 | 0.069 | 0.009 | 1.9953E-14 | 58.78 |
| *EIF4B* | rs60822569 | T | TTTCTC | 0.5547 | 0.053 | 0.008 | 3.1623E-12 | 43.89 |
| *EIF4E* | rs1354034 | T | C | 0.6039 | 0.065 | 0.008 | 3.1623E-17 | 66.02 |
| *EIF4E* | rs342298 | C | T | 0.4545 | -0.056 | 0.008 | 1.2589E-13 | 49.00 |
| *EIF4E* | rs6993770 | A | T | 0.2866 | -0.063 | 0.008 | 6.3096E-14 | 62.02 |
| *EIF4E* | rs60822569 | T | TTTCTC | 0.5541 | 0.052 | 0.008 | 1E-11 | 42.25 |
| *EIF4G1* | rs1354034 | T | C | 0.6041 | 0.091 | 0.008 | 1.5849E-33 | 129.39 |
| *EIF4G1* | rs342298 | C | T | 0.4547 | -0.058 | 0.007 | 1E-14 | 68.65 |
| *EIF4G1* | rs6993770 | A | T | 0.2866 | -0.057 | 0.008 | 5.0119E-12 | 50.77 |
| *EIF4G3* | rs11444926 | G | GA | 0.5432 | 0.072 | 0.007 | 6.3096E-22 | 105.80 |
| *EIF4G3* | rs6993770 | A | T | 0.2864 | -0.062 | 0.008 | 5.0119E-14 | 60.06 |
| *EIF5* | rs138711694 | T | C | 0.0176 | -0.25 | 0.031 | 3.9811E-16 | 65.04 |
| *ELAC1* | rs1354034 | T | C | 0.6043 | 0.1 | 0.008 | 1.2589E-39 | 156.25 |
| *ELAC1* | rs6993770 | A | T | 0.2864 | -0.08 | 0.008 | 6.3096E-22 | 100.00 |
| *ELAC1* | rs11502185 | T | C | 0.2598 | 0.081 | 0.01 | 3.1623E-17 | 65.61 |
| *ELAC1* | rs60822569 | T | TTTCTC | 0.5541 | 0.056 | 0.008 | 1E-13 | 49.00 |
| *ELAC1* | rs142132015 | T | C | 0.0048 | -0.861 | 0.054 | 1E-57 | 254.23 |
| *ELAC1* | rs892090 | T | G | 0.8343 | 0.069 | 0.01 | 1E-11 | 47.61 |
| *ELN* | rs4252125 | G | A | 0.2945 | 0.061 | 0.006 | 2.5119E-23 | 103.36 |
| *ELN* | rs17855988 | G | C | 0.1001 | 0.144 | 0.009 | 1.5849E-52 | 256.00 |
| *ELN* | rs529565 | T | C | 0.3176 | -0.05 | 0.006 | 1E-16 | 69.44 |
| *ELN* | rs964184 | G | C | 0.8664 | -0.084 | 0.008 | 1E-24 | 110.25 |
| *ELN* | rs10843390 | C | T | 0.2759 | 0.044 | 0.006 | 2.5119E-12 | 53.78 |
| *ELN* | rs28929474 | C | T | 0.0209 | 0.21 | 0.019 | 3.9811E-27 | 122.16 |
| *ELN* | rs72802342 | C | A | 0.0757 | 0.159 | 0.011 | 1E-49 | 208.93 |
| *ELN* | rs584007 | A | G | 0.642 | 0.043 | 0.006 | 7.9433E-14 | 51.36 |
| *ELN* | rs2569488 | T | C | 0.3168 | -0.064 | 0.006 | 6.3096E-26 | 113.78 |
| *ELOA* | rs35863927 | T | C | 0.5966 | 0.086 | 0.008 | 3.9811E-30 | 115.56 |
| *ELOA* | rs10418046 | T | G | 0.2114 | 0.143 | 0.009 | 5.0119E-57 | 252.46 |
| *ENAH* | rs10799316 | G | A | 0.7547 | 0.124 | 0.009 | 1.9953E-44 | 189.83 |
| *ENDOU* | rs78924983 | C | T | 0.0512 | -0.129 | 0.016 | 6.3096E-16 | 65.00 |
| *ENDOU* | rs61816761 | G | A | 0.0234 | 0.247 | 0.023 | 1.5849E-26 | 115.33 |
| *ENDOU* | rs12029741 | G | A | 0.3532 | 0.053 | 0.007 | 5.0119E-14 | 57.33 |
| *ENDOU* | rs955658 | T | C | 0.5881 | 0.12 | 0.007 | 2.5119E-70 | 293.88 |
| *ENDOU* | rs121908120 | T | A | 0.0275 | -0.168 | 0.021 | 2.5119E-16 | 64.00 |
| *ENDOU* | rs766504877 | TG | T | 0.1611 | 0.082 | 0.009 | 1.2589E-19 | 83.01 |
| *ENDOU* | rs12203057 | C | T | 0.3939 | -0.055 | 0.007 | 3.1623E-16 | 61.73 |
| *ENDOU* | rs7808585 | C | T | 0.7578 | 0.068 | 0.008 | 7.9433E-19 | 72.25 |
| *ENDOU* | rs752941245 | CAT | C | 0.5649 | 0.077 | 0.007 | 5.0119E-31 | 121.00 |
| *ENDOU* | rs11168215 | A | C | 0.3069 | 0.327 | 0.007 | 1E-200 | 2182.22 |
| *ENDOU* | rs768780444 | TA | T | 0.6312 | 0.054 | 0.007 | 1.9953E-14 | 59.51 |
| *ENDOU* | rs35673728 | T | C | 0.0628 | -0.103 | 0.014 | 5.0119E-14 | 54.13 |

| | | | | | | | | |
|---|---|---|---|---|---|---|---|---|
| *ENDOU* | rs34560261 | C | T | 0.1691 | -0.113 | 0.009 | 6.3096E-36 | 157.64 |
| *ENDOU* | rs9939286 | T | G | 0.6344 | -0.057 | 0.007 | 3.1623E-16 | 66.31 |
| *ENG* | rs61762319 | A | G | 0.03 | -0.213 | 0.02 | 1.5849E-25 | 113.42 |
| *ENG* | rs6437412 | T | C | 0.2856 | 0.065 | 0.008 | 1E-16 | 66.02 |
| *ENG* | rs9987289 | A | G | 0.9079 | -0.082 | 0.012 | 1.2589E-11 | 46.69 |
| *ENG* | rs11794565 | C | T | 0.4634 | 0.259 | 0.008 | 1E-200 | 1048.14 |
| *ENG* | rs2519093 | C | T | 0.1843 | -0.504 | 0.01 | 1E-200 | 2540.16 |
| *ENG* | rs35166255 | G | A | 0.0338 | 0.293 | 0.019 | 7.9433E-52 | 237.81 |
| *ENG* | rs186021206 | G | A | 0.0064 | 0.732 | 0.048 | 2.5119E-52 | 232.56 |
| *ENG* | rs4760 | A | G | 0.156 | 0.235 | 0.01 | 3.162E-131 | 552.25 |
| *ENG* | rs558698413 | C | A | 0.0009 | -0.979 | 0.124 | 3.9811E-15 | 62.33 |
| *ENO1* | rs11544513 | G | T | 0.0033 | -1.322 | 0.066 | 3.9811E-90 | 401.21 |
| *ENO1* | rs1354034 | T | C | 0.6043 | 0.064 | 0.008 | 6.3096E-17 | 64.00 |
| *ENO2* | rs1354034 | T | C | 0.6042 | 0.057 | 0.008 | 3.9811E-14 | 50.77 |
| *ENO2* | rs6993770 | A | T | 0.2865 | -0.147 | 0.008 | 3.1623E-71 | 337.64 |
| *ENO2* | rs60757417 | C | G | 0.0606 | -0.107 | 0.016 | 7.9433E-12 | 44.72 |
| *ENO2* | rs7896518 | A | G | 0.4241 | 0.106 | 0.008 | 6.3096E-44 | 175.56 |
| *ENO2* | rs11502185 | T | C | 0.2598 | 0.065 | 0.009 | 7.9433E-12 | 52.16 |
| *ENO2* | rs11446182 | A | AG | 0.274 | 0.145 | 0.008 | 2.5119E-68 | 328.52 |
| *ENO3* | rs74434374 | C | A | 0.0435 | 0.12 | 0.018 | 1E-11 | 44.44 |
| *ENO3* | rs7305678 | T | G | 0.8648 | -0.128 | 0.011 | 1.9953E-33 | 135.40 |
| *ENO3* | rs2243090 | T | C | 0.6702 | -0.09 | 0.008 | 1.5849E-32 | 126.56 |
| *ENO3* | rs12975366 | T | C | 0.395 | -0.094 | 0.007 | 1.5849E-37 | 180.33 |
| *ENOX2* | rs1354034 | T | C | 0.6037 | 0.07 | 0.008 | 2.5119E-19 | 76.56 |
| *ENPEP* | rs33966350 | G | A | 0.0128 | -0.323 | 0.034 | 3.9811E-21 | 90.25 |
| *ENPP2* | rs34707604 | T | C | 0.2585 | 0.057 | 0.007 | 1.9953E-16 | 66.31 |
| *ENPP2* | rs35570672 | T | C | 0.7809 | 0.068 | 0.007 | 1.9953E-20 | 94.37 |
| *ENPP2* | rs13267597 | T | G | 0.3269 | -0.262 | 0.007 | 1E-200 | 1400.90 |
| *ENPP2* | rs10901252 | G | C | 0.0615 | 0.101 | 0.012 | 1E-16 | 70.84 |
| *ENPP2* | rs11830764 | G | C | 0.0681 | -0.1 | 0.012 | 7.9433E-18 | 69.44 |
| *ENPP2* | rs7970695 | G | A | 0.6242 | 0.095 | 0.006 | 1.9953E-55 | 250.69 |
| *ENPP2* | rs186021206 | G | A | 0.0064 | 0.338 | 0.04 | 5.0119E-17 | 71.40 |
| *ENPP2* | rs77542162 | A | G | 0.0229 | 0.149 | 0.02 | 2.5119E-14 | 55.50 |
| *ENPP5* | rs4665972 | T | C | 0.6086 | 0.038 | 0.005 | 3.9811E-14 | 57.76 |
| *ENPP5* | rs9472701 | G | A | 0.6428 | -0.778 | 0.007 | 1E-200 | 12352.73 |
| *ENPP5* | rs1299525 | C | T | 0.4307 | 0.039 | 0.005 | 1.9953E-15 | 60.84 |
| *ENPP5* | rs75491173 | G | A | 0.0556 | 0.074 | 0.011 | 3.9811E-12 | 45.26 |
| *ENPP5* | rs11072817 | G | A | 0.7008 | 0.051 | 0.005 | 7.9433E-22 | 104.04 |
| *ENPP5* | rs12903256 | G | T | 0.3428 | -0.042 | 0.005 | 1.5849E-16 | 70.56 |
| *ENPP5* | rs200489612 | G | A | 0.0052 | 0.349 | 0.037 | 1.5849E-21 | 88.97 |
| *ENPP5* | rs141947687 | A | AG | 0.6367 | -0.035 | 0.005 | 2.5119E-12 | 49.00 |
| *ENPP6* | rs317900 | C | T | 0.3313 | 0.137 | 0.008 | 6.3096E-70 | 293.27 |
| *ENPP6* | rs78293932 | C | T | 0.0239 | 0.953 | 0.024 | 1E-200 | 1576.75 |
| *ENPP6* | rs9266366 | T | C | 0.8508 | 0.33 | 0.01 | 1E-200 | 1089.00 |
| *ENPP6* | rs17256042 | G | A | 0.0519 | -0.114 | 0.016 | 3.1623E-13 | 50.77 |
| *ENPP6* | rs186021206 | G | A | 0.0065 | 0.564 | 0.047 | 1.2589E-32 | 144.00 |
| *ENPP7* | rs114165349 | G | C | 0.0233 | 0.151 | 0.019 | 3.1623E-15 | 63.16 |
| *ENPP7* | rs199900492 | CA | C | 0.5189 | -0.046 | 0.006 | 3.9811E-15 | 58.78 |
| *ENPP7* | rs28601761 | C | G | 0.4199 | -0.042 | 0.006 | 6.3096E-13 | 49.00 |
| *ENPP7* | rs762439986 | CATA | C | 0.4685 | 0.063 | 0.006 | 5.0119E-28 | 110.25 |
| *ENPP7* | rs1169294 | G | A | 0.3082 | 0.058 | 0.006 | 1E-20 | 93.44 |
| *ENPP7* | rs28929474 | C | T | 0.0207 | 0.141 | 0.02 | 2.5119E-12 | 49.70 |
| *ENPP7* | rs147233090 | C | T | 0.0235 | 0.143 | 0.019 | 7.9433E-14 | 56.65 |
| *ENPP7* | rs8076533 | C | T | 0.3666 | 0.95 | 0.008 | 1E-200 | 14101.56 |
| *ENPP7* | rs6050555 | A | G | 0.5403 | 0.04 | 0.006 | 1E-11 | 44.44 |
| *ENSA* | rs190018150 | G | A | 0.041 | 0.131 | 0.019 | 1.2589E-11 | 47.54 |
| *ENSA* | rs1354034 | T | C | 0.6044 | 0.061 | 0.008 | 3.9811E-15 | 58.14 |
| *ENTPD2* | rs56278466 | T | G | 0.661 | 0.05 | 0.007 | 3.9811E-12 | 51.02 |
| *ENTPD2* | rs5848 | C | T | 0.274 | 0.062 | 0.008 | 2.5119E-15 | 60.06 |
| *ENTPD5* | rs35457250 | C | T | 0.0089 | 0.759 | 0.038 | 3.1623E-88 | 398.95 |
| *ENTPD5* | rs6054 | C | T | 0.0059 | 0.344 | 0.048 | 1E-12 | 51.36 |
| *ENTPD5* | rs174574 | A | C | 0.6474 | -0.066 | 0.007 | 1.9953E-18 | 88.90 |
| *ENTPD5* | rs10843403 | T | C | 0.6094 | 0.075 | 0.007 | 2.5119E-24 | 114.80 |
| *ENTPD5* | rs146086623 | G | A | 0.0534 | -0.973 | 0.017 | 1E-200 | 3275.88 |
| *ENTPD5* | rs28929474 | C | T | 0.0208 | 0.552 | 0.027 | 1.2589E-95 | 417.98 |
| *ENTPD5* | rs763665 | C | T | 0.1599 | 0.08 | 0.01 | 3.9811E-16 | 64.00 |
| *ENTPD5* | rs149394327 | G | C | 0.0301 | 0.412 | 0.021 | 5.0119E-85 | 384.91 |
| *ENTPD5* | rs7236989 | T | C | 0.6279 | -0.052 | 0.007 | 1.9953E-12 | 55.18 |

| | | | | | | | | |
|---|---|---|---|---|---|---|---|---|
| *ENTPD6* | rs61804164 | G | C | 0.1238 | 0.069 | 0.01 | 7.9433E-12 | 47.61 |
| *ENTPD6* | rs66824127 | T | C | 0.557 | 0.045 | 0.007 | 1.2589E-11 | 41.33 |
| *ENTPD6* | rs782134971 | GAAACTGC( | G | 0.2539 | 0.148 | 0.008 | 1E-85 | 342.25 |
| *ENTPD6* | rs139196635 | G | A | 0.0221 | 0.164 | 0.023 | 1.2589E-12 | 50.84 |
| *ENTPD6* | rs28929474 | C | T | 0.0208 | 0.187 | 0.023 | 2.5119E-16 | 66.10 |
| *ENTPD6* | rs704 | G | A | 0.4738 | -0.051 | 0.007 | 5.0119E-15 | 53.08 |
| *ENTPD6* | rs17695224 | G | A | 0.2664 | -0.096 | 0.007 | 3.1623E-38 | 188.08 |
| *ENTPD6* | rs6050446 | A | G | 0.9665 | 1.864 | 0.021 | 1E-200 | 7878.68 |
| *ENTPD6* | rs6061054 | T | G | 0.9648 | 1.572 | 0.021 | 1E-200 | 5603.59 |
| *ENTR1* | rs11145904 | A | C | 0.1725 | -0.297 | 0.01 | 5.012E-195 | 882.09 |
| *ENTR1* | rs3747207 | G | A | 0.2156 | 0.071 | 0.009 | 2.5119E-15 | 62.23 |
| *EPCAM* | rs2241764 | C | T | 0.3382 | -0.084 | 0.008 | 6.3096E-28 | 110.25 |
| *EPCAM* | rs35853577 | T | C | 0.4254 | -0.067 | 0.007 | 1.5849E-19 | 91.61 |
| *EPCAM* | rs1713810 | A | G | 0.852 | -0.084 | 0.01 | 3.1623E-16 | 70.56 |
| *EPCAM* | rs939885 | G | A | 0.4857 | 0.06 | 0.007 | 1.2589E-16 | 73.47 |
| *EPCAM* | rs10066308 | G | A | 0.3094 | -0.053 | 0.008 | 1.5849E-11 | 43.89 |
| *EPCAM* | rs6911827 | C | T | 0.4524 | 0.049 | 0.007 | 1E-11 | 49.00 |
| *EPCAM* | rs9478784 | C | T | 0.0952 | 0.12 | 0.012 | 1.9953E-22 | 100.00 |
| *EPCAM* | rs931493 | G | A | 0.0317 | -0.185 | 0.021 | 1.9953E-18 | 77.61 |
| *EPCAM* | rs2291428 | G | C | 0.2441 | -0.096 | 0.008 | 6.3096E-30 | 144.00 |
| *EPCAM* | rs56398830 | G | A | 0.0112 | -0.845 | 0.035 | 7.943E-132 | 582.88 |
| *EPCAM* | rs35193388 | G | GC | 0.735 | -0.072 | 0.008 | 6.3096E-18 | 81.00 |
| *EPCAM* | rs12938714 | G | A | 0.3302 | 0.164 | 0.008 | 3.162E-100 | 420.25 |
| *EPCAM* | rs12453447 | A | T | 0.5939 | -0.07 | 0.008 | 1E-16 | 76.56 |
| *EPCAM* | rs2318997 | T | G | 0.4333 | -0.054 | 0.007 | 1.5849E-13 | 59.51 |
| *EPCAM* | rs681343 | C | T | 0.5096 | -0.234 | 0.007 | 1E-200 | 1117.47 |
| *EPCAM* | rs3746778 | G | A | 0.4206 | 0.06 | 0.008 | 1.5849E-15 | 56.25 |
| *EPHA1* | rs1497406 | A | G | 0.5792 | 0.061 | 0.007 | 1E-18 | 75.94 |
| *EPHA1* | rs61747728 | C | T | 0.038 | 0.133 | 0.018 | 1E-13 | 54.60 |
| *EPHA1* | rs75045569 | T | G | 0.1827 | 0.254 | 0.009 | 1.995E-172 | 796.49 |
| *EPHA1* | rs10740131 | A | T | 0.4707 | 0.096 | 0.007 | 2.5119E-44 | 188.08 |
| *EPHA1* | rs28929474 | C | T | 0.0208 | 0.29 | 0.024 | 1.9953E-33 | 146.01 |
| *EPHA1* | rs3814995 | C | T | 0.3146 | 0.057 | 0.007 | 7.9433E-15 | 66.31 |
| *EPHA2* | rs924204 | A | G | 0.5901 | -0.133 | 0.007 | 3.1623E-72 | 361.00 |
| *EPHA2* | rs61747728 | C | T | 0.038 | 0.151 | 0.019 | 1.2589E-15 | 63.16 |
| *EPHA2* | rs11689257 | G | A | 0.0691 | -0.111 | 0.015 | 3.9811E-14 | 54.76 |
| *EPHA2* | rs8176746 | G | T | 0.0617 | 0.141 | 0.015 | 6.3096E-21 | 88.36 |
| *EPHA2* | rs12934455 | C | T | 0.1661 | -0.066 | 0.01 | 1.2589E-11 | 43.56 |
| *EPHA2* | rs679574 | C | G | 0.5091 | -0.069 | 0.007 | 1.9953E-21 | 97.16 |
| *EPHA4* | rs113651398 | CT | C | 0.6488 | -0.062 | 0.008 | 3.1623E-16 | 60.06 |
| *EPHA4* | rs1260326 | T | C | 0.6081 | 0.063 | 0.007 | 1.2589E-17 | 81.00 |
| *EPHA4* | rs10932916 | G | A | 0.5141 | 0.139 | 0.007 | 1.5849E-84 | 394.31 |
| *EPHA4* | rs2519093 | C | T | 0.1846 | -0.612 | 0.01 | 1E-200 | 3745.44 |
| *EPHA4* | rs186021206 | G | A | 0.0064 | 0.655 | 0.049 | 3.1623E-41 | 178.69 |
| *EPHA4* | rs284662 | T | C | 0.6216 | 0.054 | 0.007 | 1E-13 | 59.51 |
| *EPHB4* | rs61747728 | C | T | 0.0381 | 0.154 | 0.019 | 2.5119E-16 | 65.70 |
| *EPHB4* | rs314361 | A | G | 0.5204 | 0.154 | 0.007 | 5.012E-100 | 484.00 |
| *EPHB4* | rs11782073 | A | G | 0.742 | 0.056 | 0.008 | 1.5849E-11 | 49.00 |
| *EPHB4* | rs507666 | G | A | 0.1845 | -0.368 | 0.01 | 1E-200 | 1354.24 |
| *EPHB4* | rs7099071 | T | C | 0.1688 | 0.067 | 0.01 | 5.0119E-12 | 44.89 |
| *EPHB4* | rs7909516 | C | T | 0.2244 | -0.093 | 0.009 | 5.0119E-27 | 106.78 |
| *EPHB4* | rs9558661 | C | T | 0.2015 | -0.133 | 0.009 | 1.9953E-48 | 218.38 |
| *EPHB4* | rs28640218 | G | T | 0.1917 | -0.079 | 0.009 | 1.2589E-17 | 77.05 |
| *EPHB4* | rs186021206 | G | A | 0.0064 | 0.395 | 0.05 | 1.9953E-15 | 62.41 |
| *EPHB4* | rs1137844 | C | G | 0.3154 | 0.064 | 0.008 | 2.5119E-16 | 64.00 |
| *EPHB6* | rs61747728 | C | T | 0.0379 | 0.199 | 0.019 | 5.0119E-25 | 109.70 |
| *EPHB6* | rs7789303 | A | G | 0.2659 | -0.318 | 0.009 | 1E-200 | 1248.44 |
| *EPHB6* | rs13329952 | T | C | 0.1918 | -0.081 | 0.009 | 6.3096E-18 | 81.00 |
| *EPHX2* | rs71553864 | A | AGTC | 0.0576 | 0.26 | 0.017 | 7.9433E-56 | 233.91 |
| *EPO* | rs218264 | A | T | 0.2476 | 0.078 | 0.009 | 6.3096E-19 | 75.11 |
| *EPO* | rs2032444 | T | G | 0.3519 | -0.074 | 0.008 | 1.2589E-20 | 85.56 |
| *EPO* | rs1546723 | G | A | 0.4495 | 0.058 | 0.008 | 3.1623E-14 | 52.56 |
| *EPO* | rs34164109 | C | T | 0.2602 | 0.208 | 0.009 | 3.981E-126 | 534.12 |
| *EPO* | rs11976235 | C | T | 0.0059 | 0.547 | 0.049 | 1E-28 | 124.62 |
| *EPO* | rs78744187 | C | T | 0.0815 | -0.114 | 0.014 | 1.2589E-16 | 66.31 |
| *EPO* | rs4811073 | C | T | 0.5429 | -0.053 | 0.008 | 1.9953E-12 | 43.89 |
| *EPO* | rs855791 | A | G | 0.5609 | -0.078 | 0.008 | 1E-24 | 95.06 |
| *EPPK1* | rs1434282 | C | T | 0.7262 | -0.1 | 0.008 | 2.5119E-37 | 156.25 |

| | | | | | | | | |
|---|---|---|---|---|---|---|---|---|
| *EPPK1* | rs342293 | C | G | 0.4595 | -0.052 | 0.007 | 6.3096E-14 | 55.18 |
| *EPPK1* | rs73375129 | A | G | 0.1366 | -0.544 | 0.011 | 1E-200 | 2445.75 |
| *EPPK1* | rs60757417 | C | G | 0.0606 | -0.11 | 0.015 | 6.3096E-14 | 53.78 |
| *EPPK1* | rs695113 | C | T | 0.6949 | 0.057 | 0.008 | 3.1623E-14 | 50.77 |
| *EPS8L2* | rs9271657 | T | C | 0.2139 | 0.065 | 0.009 | 1.5849E-12 | 52.16 |
| *EPS8L2* | rs1106192 | C | G | 0.0457 | -0.118 | 0.017 | 1E-11 | 48.18 |
| *EPS8L2* | rs11246276 | G | T | 0.1588 | -0.394 | 0.01 | 1E-200 | 1552.36 |
| *EPS8L2* | rs3747207 | G | A | 0.2155 | 0.062 | 0.009 | 1.5849E-12 | 47.46 |
| *ERBB2* | rs9270464 | A | G | 0.3389 | -0.068 | 0.008 | 3.9811E-18 | 72.25 |
| *ERBB2* | rs1461729 | A | G | 0.8972 | -0.09 | 0.012 | 7.9433E-15 | 56.25 |
| *ERBB2* | rs8176749 | C | T | 0.0612 | 0.11 | 0.015 | 3.9811E-14 | 53.78 |
| *ERBB2* | rs11045856 | T | G | 0.2401 | 0.071 | 0.008 | 3.9811E-18 | 78.77 |
| *ERBB2* | rs28929474 | C | T | 0.0206 | 0.244 | 0.025 | 3.9811E-23 | 95.26 |
| *ERBB2* | rs4788460 | C | T | 0.294 | 0.052 | 0.008 | 1.2589E-11 | 42.25 |
| *ERBB2* | rs3764354 | C | T | 0.1461 | 0.2 | 0.01 | 1.2589E-88 | 400.00 |
| *ERBB2* | rs1801689 | A | C | 0.0302 | 0.309 | 0.021 | 3.1623E-50 | 216.51 |
| *ERBB3* | rs1260326 | T | C | 0.6086 | -0.131 | 0.007 | 1.2589E-74 | 350.22 |
| *ERBB3* | rs13234131 | A | G | 0.1271 | -0.085 | 0.01 | 3.9811E-16 | 72.25 |
| *ERBB3* | rs4731549 | C | T | 0.5502 | -0.102 | 0.007 | 1.5849E-47 | 212.33 |
| *ERBB3* | rs28601761 | C | G | 0.4195 | -0.056 | 0.007 | 3.1623E-15 | 64.00 |
| *ERBB3* | rs10761737 | T | C | 0.4131 | -0.049 | 0.007 | 7.9433E-12 | 49.00 |
| *ERBB3* | rs75526830 | A | G | 0.0049 | 2.485 | 0.053 | 1E-200 | 2198.37 |
| *ERBB3* | rs150844304 | A | C | 0.0241 | 0.205 | 0.023 | 1.9953E-19 | 79.44 |
| *ERBB3* | rs149394327 | G | C | 0.0301 | 0.19 | 0.02 | 1.5849E-20 | 90.25 |
| *ERBB4* | rs9787076 | A | C | 0.3295 | 0.064 | 0.008 | 6.3096E-17 | 64.00 |
| *ERBB4* | rs35383942 | C | T | 0.0617 | 0.127 | 0.015 | 1E-17 | 71.68 |
| *ERBB4* | rs12692385 | C | T | 0.6714 | 0.082 | 0.008 | 2.5119E-26 | 105.06 |
| *ERBB4* | rs4665972 | T | C | 0.6087 | 0.064 | 0.008 | 1.2589E-17 | 64.00 |
| *ERBB4* | rs6735267 | C | T | 0.2688 | 0.219 | 0.008 | 2.512E-153 | 749.39 |
| *ERBB4* | rs11757137 | C | T | 0.44 | 0.058 | 0.007 | 1E-14 | 68.65 |
| *ERBB4* | rs34673027 | C | A | 0.4471 | -0.076 | 0.007 | 5.0119E-26 | 117.88 |
| *ERBB4* | rs2519093 | C | T | 0.1842 | -0.136 | 0.009 | 7.9433E-49 | 228.35 |
| *ERBB4* | rs56278466 | T | G | 0.6612 | 0.11 | 0.008 | 2.5119E-48 | 189.06 |
| *ERBB4* | rs11227279 | G | A | 0.3437 | -0.051 | 0.008 | 1E-11 | 40.64 |
| *ERBB4* | rs11830324 | T | A | 0.1694 | -0.078 | 0.01 | 3.9811E-16 | 60.84 |
| *ERBB4* | rs9567991 | C | T | 0.6558 | 0.052 | 0.007 | 3.9811E-12 | 55.18 |
| *ERBB4* | rs2541593 | C | A | 0.8104 | 0.068 | 0.009 | 1.2589E-13 | 57.09 |
| *ERBB4* | rs186021206 | G | A | 0.0064 | 0.975 | 0.049 | 1.5849E-87 | 395.93 |
| *ERBB4* | rs72835417 | G | A | 0.11 | 0.152 | 0.011 | 1.9953E-40 | 190.94 |
| *ERBB4* | rs485186 | A | G | 0.5378 | 0.097 | 0.007 | 1E-41 | 192.02 |
| *ERBIN* | rs342296 | G | A | 0.4563 | -0.052 | 0.008 | 5.0119E-12 | 42.25 |
| *ERBIN* | rs10822145 | C | T | 0.4719 | 0.055 | 0.008 | 1.9953E-13 | 47.27 |
| *ERC2* | rs79784038 | G | C | 0.0256 | -0.17 | 0.025 | 5.0119E-12 | 46.24 |
| *EREG* | rs78803121 | G | T | 0.058 | -0.188 | 0.016 | 3.9811E-31 | 138.06 |
| *EREG* | rs12655145 | A | G | 0.6159 | 0.075 | 0.008 | 3.9811E-22 | 87.89 |
| *EREG* | rs2516471 | G | A | 0.0636 | -0.11 | 0.016 | 2.5119E-12 | 47.27 |
| *EREG* | rs6993770 | A | T | 0.2865 | -0.093 | 0.008 | 1.9953E-28 | 135.14 |
| *EREG* | rs10820606 | A | C | 0.2296 | -0.11 | 0.009 | 1.9953E-33 | 149.38 |
| *EREG* | rs7080386 | C | A | 0.4119 | 0.07 | 0.008 | 1.2589E-19 | 76.56 |
| *EREG* | rs892090 | T | G | 0.834 | 0.119 | 0.01 | 3.1623E-31 | 141.61 |
| *EREG* | rs6081569 | G | C | 0.3689 | 0.055 | 0.008 | 1.9953E-12 | 47.27 |
| *ERI1* | rs1354034 | T | C | 0.6041 | 0.052 | 0.008 | 1.5849E-11 | 42.25 |
| *ERI1* | rs5985 | C | A | 0.2582 | -0.084 | 0.009 | 3.9811E-22 | 87.11 |
| *ERI1* | rs7814328 | T | C | 0.4608 | -0.071 | 0.008 | 1E-20 | 78.77 |
| *ERMAP* | rs28594470 | A | T | 0.4195 | -0.087 | 0.007 | 3.9811E-34 | 154.47 |
| *ERMAP* | rs11210710 | G | T | 0.4761 | -0.091 | 0.007 | 1.5849E-38 | 169.00 |
| *ERMAP* | rs4672497 | C | G | 0.2213 | -0.07 | 0.008 | 5.0119E-17 | 76.56 |
| *ERMAP* | rs11736498 | A | G | 0.4479 | 0.27 | 0.007 | 1E-200 | 1487.76 |
| *ERMAP* | rs28749166 | C | T | 0.3672 | -0.058 | 0.007 | 1.9953E-15 | 68.65 |
| *ERMAP* | rs6592965 | G | A | 0.4536 | -0.099 | 0.007 | 2.5119E-46 | 200.02 |
| *ERMAP* | rs6993770 | A | T | 0.2866 | -0.061 | 0.008 | 1E-15 | 58.14 |
| *ERMAP* | rs746064 | G | A | 0.3797 | 0.449 | 0.008 | 1E-200 | 3150.02 |
| *ERN1* | rs1260326 | T | C | 0.6084 | -0.059 | 0.008 | 1.2589E-14 | 54.39 |
| *ERN1* | rs6054 | C | T | 0.006 | 0.382 | 0.05 | 1.5849E-14 | 58.37 |
| *ERN1* | rs8176746 | G | T | 0.0614 | 0.204 | 0.015 | 5.0119E-40 | 184.96 |
| *ERN1* | rs5787948 | C | CA | 0.7327 | -0.06 | 0.009 | 1.9953E-12 | 44.44 |
| *ERN1* | rs174566 | A | G | 0.3503 | 0.104 | 0.008 | 3.9811E-41 | 169.00 |
| *ERN1* | rs28929474 | C | T | 0.0208 | 0.222 | 0.026 | 1.2589E-17 | 72.91 |

| | | | | | | | | |
|---|---|---|---|---|---|---|---|---|
| *ERN1* | rs217184 | T | C | 0.1963 | -0.085 | 0.009 | 1.9953E-19 | 89.20 |
| *ERN1* | rs1801689 | A | C | 0.0301 | 0.526 | 0.022 | 1.995E-128 | 571.64 |
| *ERN1* | rs429358 | T | C | 0.1555 | -0.147 | 0.01 | 6.3096E-46 | 216.09 |
| *ERN1* | rs681343 | C | T | 0.509 | -0.124 | 0.007 | 1.5849E-61 | 313.80 |
| *ERP29* | rs1354034 | T | C | 0.6044 | 0.068 | 0.008 | 7.9433E-19 | 72.25 |
| *ERP29* | rs342298 | C | T | 0.4549 | -0.058 | 0.008 | 1.5849E-14 | 52.56 |
| *ERP29* | rs6993770 | A | T | 0.287 | -0.065 | 0.008 | 5.0119E-15 | 66.02 |
| *ERP44* | rs4846923 | T | G | 0.7042 | -0.059 | 0.008 | 1.9953E-13 | 54.39 |
| *ERP44* | rs1260326 | T | C | 0.608 | -0.055 | 0.007 | 1.9953E-13 | 61.73 |
| *ERP44* | rs7614709 | G | A | 0.2224 | -0.25 | 0.009 | 2.512E-176 | 771.60 |
| *ERP44* | rs6993770 | A | T | 0.2864 | -0.061 | 0.008 | 1.9953E-14 | 58.14 |
| *ERP44* | rs188976415 | C | A | 0.0036 | -0.594 | 0.061 | 1.5849E-22 | 94.82 |
| *ERP44* | rs36115375 | C | A | 0.0631 | 0.723 | 0.015 | 1E-200 | 2323.24 |
| *ERP44* | rs17580 | T | A | 0.0495 | -0.132 | 0.017 | 3.1623E-15 | 60.29 |
| *ESAM* | rs6961069 | C | T | 0.4026 | 0.06 | 0.007 | 1.9953E-15 | 73.47 |
| *ESAM* | rs6993770 | A | T | 0.2865 | -0.105 | 0.008 | 3.9811E-38 | 172.27 |
| *ESAM* | rs8176672 | C | T | 0.0613 | 0.205 | 0.015 | 3.9811E-41 | 186.78 |
| *ESAM* | rs7080386 | C | A | 0.4115 | 0.086 | 0.007 | 1.5849E-30 | 150.94 |
| *ESAM* | rs61753651 | G | A | 0.0039 | -1.89 | 0.059 | 1E-200 | 1026.17 |
| *ESAM* | rs2127870 | G | C | 0.7948 | 0.087 | 0.009 | 2.5119E-21 | 93.44 |
| *ESAM* | rs188942006 | G | C | 0.0651 | 0.109 | 0.015 | 6.3096E-13 | 52.80 |
| *ESAM* | rs1654425 | T | C | 0.8343 | 0.133 | 0.01 | 1E-40 | 176.89 |
| *ESM1* | rs61801010 | A | G | 0.106 | 0.086 | 0.012 | 6.3096E-12 | 51.36 |
| *ESM1* | rs557470468 | T | C | 0.005 | 0.45 | 0.053 | 2.5119E-17 | 72.09 |
| *ESM1* | rs4242051 | T | C | 0.7457 | 0.192 | 0.009 | 5.012E-111 | 455.11 |
| *ESM1* | rs224916 | A | G | 0.7259 | -0.077 | 0.008 | 3.1623E-20 | 92.64 |
| *ESM1* | rs113760175 | G | A | 0.0707 | 0.136 | 0.014 | 2.5119E-21 | 94.37 |
| *ESM1* | rs9269891 | C | T | 0.4612 | -0.068 | 0.008 | 1.2589E-16 | 72.25 |
| *ESM1* | rs1475718 | G | A | 0.4368 | -0.081 | 0.007 | 1E-27 | 133.90 |
| *ESM1* | rs10761782 | A | G | 0.475 | 0.06 | 0.007 | 1.5849E-16 | 73.47 |
| *ESM1* | rs7310615 | C | G | 0.5176 | -0.067 | 0.007 | 7.9433E-20 | 91.61 |
| *ESR1* | rs373801706 | A | G | 0.0535 | 0.149 | 0.017 | 1.2589E-17 | 76.82 |
| *ESYT2* | rs1354034 | T | C | 0.6043 | 0.078 | 0.008 | 1E-24 | 95.06 |
| *ESYT2* | rs2709865 | T | C | 0.9422 | -0.291 | 0.016 | 1.9953E-73 | 330.79 |
| *EVI5* | rs11808092 | C | A | 0.2553 | -0.145 | 0.009 | 1.5849E-63 | 259.57 |
| *EXOSC10* | rs79874727 | G | A | 0.1061 | 0.107 | 0.014 | 3.1623E-14 | 58.41 |
| *EXTL1* | rs2736831 | C | A | 0.6926 | -0.412 | 0.008 | 1E-200 | 2652.25 |
| *EZR* | rs3734470 | A | T | 0.6968 | 0.097 | 0.008 | 3.1623E-32 | 147.02 |
| *EZR* | rs7208422 | A | T | 0.4772 | -0.056 | 0.007 | 7.9433E-14 | 64.00 |
| *EZR* | rs10405357 | T | C | 0.4356 | -0.062 | 0.008 | 2.5119E-16 | 60.06 |
| *F10* | rs35797675 | T | G | 0.2146 | -0.075 | 0.009 | 1.2589E-16 | 69.44 |
| *F10* | rs28601761 | C | G | 0.4195 | -0.053 | 0.008 | 1.2589E-12 | 43.89 |
| *F10* | rs10982156 | T | A | 0.0693 | -0.454 | 0.015 | 1.585E-195 | 916.07 |
| *F10* | rs6479877 | C | G | 0.4403 | -0.08 | 0.007 | 3.9811E-27 | 130.61 |
| *F10* | rs547138 | T | A | 0.6051 | 0.179 | 0.008 | 1E-118 | 500.64 |
| *F10* | rs924135 | A | T | 0.6105 | 0.057 | 0.008 | 6.3096E-14 | 50.77 |
| *F10* | rs2330795 | G | A | 0.3382 | -0.073 | 0.008 | 5.0119E-21 | 83.27 |
| *F11* | rs1260326 | T | C | 0.6079 | -0.066 | 0.007 | 1.2589E-22 | 88.90 |
| *F11* | rs710446 | T | C | 0.409 | 0.474 | 0.007 | 1E-200 | 4585.22 |
| *F11* | rs2289252 | C | T | 0.3995 | 0.385 | 0.007 | 1E-200 | 3025.00 |
| *F11* | rs451637 | C | T | 0.3097 | -0.052 | 0.007 | 7.9433E-13 | 55.18 |
| *F11* | rs77542162 | A | G | 0.0229 | 0.158 | 0.022 | 5.0119E-13 | 51.58 |
| *F11R* | rs60315407 | T | C | 0.2912 | 0.063 | 0.008 | 6.3096E-14 | 62.02 |
| *F11R* | rs1354034 | T | C | 0.6041 | 0.075 | 0.008 | 2.5119E-22 | 87.89 |
| *F11R* | rs3804753 | A | G | 0.5935 | -0.055 | 0.008 | 7.9433E-13 | 47.27 |
| *F11R* | rs11242109 | G | T | 0.4814 | -0.064 | 0.008 | 3.1623E-17 | 64.00 |
| *F11R* | rs6993770 | A | T | 0.2864 | -0.069 | 0.008 | 1.5849E-16 | 74.39 |
| *F11R* | rs409801 | T | C | 0.5069 | 0.053 | 0.008 | 1.9953E-12 | 43.89 |
| *F11R* | rs10733789 | T | C | 0.3134 | 0.059 | 0.008 | 5.0119E-13 | 54.39 |
| *F11R* | rs4608137 | G | A | 0.4875 | -0.063 | 0.008 | 1.2589E-16 | 62.02 |
| *F11R* | rs60822569 | T | TTTCTC | 0.5545 | 0.055 | 0.008 | 3.1623E-13 | 47.27 |
| *F11R* | rs1654425 | T | C | 0.8344 | 0.088 | 0.01 | 6.3096E-18 | 77.44 |
| *F12* | rs1260326 | T | C | 0.6084 | -0.068 | 0.005 | 7.9433E-37 | 184.96 |
| *F12* | rs1801020 | A | G | 0.7449 | 1.182 | 0.009 | 1E-200 | 17248.44 |
| *F12* | rs204886 | T | C | 0.5062 | 0.037 | 0.005 | 5.0119E-13 | 54.76 |
| *F12* | rs13234378 | A | T | 0.1271 | -0.061 | 0.008 | 7.9433E-15 | 58.14 |
| *F12* | rs7924036 | G | T | 0.501 | -0.047 | 0.005 | 1.5849E-19 | 88.36 |
| *F12* | rs12975366 | T | C | 0.3952 | 0.047 | 0.005 | 1.9953E-18 | 88.36 |

| | | | | | | | | |
|---|---|---|---|---|---|---|---|---|
| *F13B* | rs12134960 | G | C | 0.1591 | -0.559 | 0.01 | 1E-200 | 3124.81 |
| *F13B* | rs1047891 | C | A | 0.3129 | -0.07 | 0.008 | 2.5119E-20 | 76.56 |
| *F13B* | rs12204494 | T | G | 0.3823 | 0.236 | 0.007 | 1E-200 | 1136.65 |
| *F13B* | rs570572562 | A | AT | 0.2649 | -0.085 | 0.008 | 1.9953E-26 | 112.89 |
| *F2* | rs10982156 | T | A | 0.0693 | -0.111 | 0.015 | 6.3096E-13 | 54.76 |
| *F2* | rs1799963 | G | A | 0.0109 | 0.731 | 0.037 | 7.9433E-86 | 390.33 |
| *F2* | rs151130188 | G | T | 0.0083 | 0.49 | 0.049 | 7.9433E-24 | 100.00 |
| *F2R* | rs13088560 | G | T | 0.1761 | -0.08 | 0.01 | 7.9433E-15 | 64.00 |
| *F2R* | rs1354034 | T | C | 0.6039 | 0.094 | 0.008 | 1.9953E-34 | 138.06 |
| *F2R* | rs168753 | A | T | 0.1726 | -0.152 | 0.01 | 6.3096E-52 | 231.04 |
| *F2R* | rs11759553 | A | T | 0.2722 | 0.061 | 0.008 | 5.0119E-13 | 58.14 |
| *F2R* | rs6993770 | A | T | 0.2867 | -0.082 | 0.008 | 3.1623E-23 | 105.06 |
| *F2R* | rs579459 | T | C | 0.2077 | -0.063 | 0.009 | 1.2589E-11 | 49.00 |
| *F2R* | rs7080386 | C | A | 0.4122 | 0.112 | 0.008 | 1.2589E-48 | 196.00 |
| *F2R* | rs17655730 | T | C | 0.2454 | 0.06 | 0.009 | 1E-11 | 44.44 |
| *F2R* | rs892090 | T | G | 0.8342 | 0.085 | 0.01 | 5.0119E-17 | 72.25 |
| *F3* | rs6666213 | A | G | 0.3729 | 0.185 | 0.007 | 7.943E-145 | 698.47 |
| *F3* | rs780094 | T | C | 0.6199 | 0.051 | 0.007 | 5.0119E-13 | 53.08 |
| *F3* | rs2519093 | C | T | 0.1843 | -0.276 | 0.009 | 1E-200 | 940.44 |
| *F3* | rs199881213 | C | CA | 0.0737 | -0.109 | 0.013 | 1.5849E-16 | 70.30 |
| *F3* | rs488703 | G | A | 0.1014 | 0.097 | 0.011 | 6.3096E-18 | 77.76 |
| *F3* | rs200489612 | G | A | 0.0053 | 0.485 | 0.051 | 2.5119E-21 | 90.44 |
| *F3* | rs35218105 | C | CA | 0.3148 | 0.086 | 0.008 | 3.9811E-30 | 115.56 |
| *F3* | rs2659005 | C | T | 0.4414 | 0.048 | 0.007 | 3.9811E-12 | 47.02 |
| *F3* | rs516316 | G | C | 0.509 | -0.283 | 0.007 | 1E-200 | 1634.47 |
| *F7* | rs1260326 | T | C | 0.6083 | -0.126 | 0.007 | 5.0119E-73 | 324.00 |
| *F7* | rs772513171 | CTTTT | C | 0.6724 | 0.053 | 0.007 | 1.5849E-13 | 57.33 |
| *F7* | rs35732917 | T | C | 0.2851 | -0.066 | 0.008 | 1.2589E-18 | 68.06 |
| *F7* | rs7075901 | C | A | 0.4685 | -0.057 | 0.007 | 3.9811E-17 | 66.31 |
| *F7* | rs7232 | T | A | 0.3738 | 0.059 | 0.007 | 5.0119E-17 | 71.04 |
| *F7* | rs510335 | G | T | 0.1122 | -0.779 | 0.012 | 1E-200 | 4214.17 |
| *F7* | rs28929474 | C | T | 0.0208 | 0.162 | 0.024 | 7.9433E-12 | 45.56 |
| *F7* | rs540574262 | G | GT | 0.0865 | 0.221 | 0.012 | 3.1623E-71 | 339.17 |
| *F9* | rs1260326 | T | C | 0.6081 | -0.11 | 0.008 | 2.5119E-47 | 189.06 |
| *F9* | rs56383788 | A | G | 0.1994 | -0.073 | 0.009 | 3.9811E-15 | 65.79 |
| *FABP1* | rs2241883 | T | C | 0.3068 | -0.237 | 0.008 | 6.31E-195 | 877.64 |
| *FABP1* | rs17655730 | T | C | 0.2454 | 0.06 | 0.009 | 2.5119E-12 | 44.44 |
| *FABP2* | rs1375131 | T | C | 0.2658 | 0.058 | 0.009 | 1.5849E-11 | 41.53 |
| *FABP2* | rs17009129 | C | T | 0.3251 | -0.213 | 0.008 | 3.162E-156 | 708.89 |
| *FABP2* | rs6472539 | G | C | 0.5158 | -0.071 | 0.007 | 1.2589E-21 | 102.88 |
| *FABP2* | rs17655730 | T | C | 0.2454 | 0.086 | 0.009 | 5.0119E-23 | 91.31 |
| *FABP2* | rs8111874 | G | A | 0.4646 | -0.057 | 0.008 | 1E-13 | 50.77 |
| *FABP3* | rs61780805 | C | T | 0.0478 | -0.192 | 0.016 | 3.1623E-32 | 144.00 |
| *FABP3* | rs12463674 | A | G | 0.3157 | 0.064 | 0.007 | 1E-17 | 83.59 |
| *FABP3* | rs7481951 | A | T | 0.5876 | 0.061 | 0.007 | 3.9811E-18 | 75.94 |
| *FABP4* | rs7647481 | G | A | 0.1215 | 0.104 | 0.011 | 7.9433E-23 | 89.39 |
| *FABP4* | rs112313579 | T | TG | 0.1591 | -0.118 | 0.009 | 1.5849E-35 | 171.90 |
| *FABP5* | rs118019635 | C | A | 0.0736 | -0.2 | 0.014 | 7.9433E-44 | 204.08 |
| *FABP5* | rs6993770 | A | T | 0.2863 | -0.058 | 0.008 | 3.1623E-12 | 52.56 |
| *FABP6* | rs1130435 | C | T | 0.4012 | -0.131 | 0.008 | 1.5849E-62 | 268.14 |
| *FABP6* | rs5875434 | C | CT | 0.547 | -0.153 | 0.008 | 1.2589E-90 | 365.77 |
| *FABP6* | rs6472539 | G | C | 0.5152 | -0.209 | 0.008 | 1E-167 | 682.52 |
| *FABP6* | rs35866622 | C | T | 0.5001 | -0.075 | 0.008 | 5.0119E-23 | 87.89 |
| *FABP9* | rs72646785 | G | T | 0.166 | 0.156 | 0.01 | 1.5849E-58 | 243.36 |
| *FABP9* | rs36010924 | A | G | 0.1844 | 0.371 | 0.009 | 1E-200 | 1699.27 |
| *FABP9* | rs774004 | G | A | 0.7405 | 0.076 | 0.008 | 1.2589E-21 | 90.25 |
| *FABP9* | rs1344815 | A | G | 0.6538 | -0.05 | 0.007 | 7.9433E-12 | 51.02 |
| *FABP9* | rs1345417 | C | G | 0.603 | 0.072 | 0.007 | 1.2589E-22 | 105.80 |
| *FABP9* | rs57866025 | C | T | 0.2664 | 0.055 | 0.008 | 6.3096E-12 | 47.27 |
| *FABP9* | rs2436396 | A | G | 0.7012 | -0.064 | 0.008 | 7.9433E-17 | 64.00 |
| *FABP9* | rs28497692 | G | T | 0.0384 | -0.224 | 0.019 | 1.5849E-33 | 138.99 |
| *FABP9* | rs541551288 | G | GA | 0.1964 | -0.091 | 0.009 | 2.5119E-24 | 102.23 |
| *FABP9* | rs7905367 | G | C | 0.7976 | 0.081 | 0.009 | 7.9433E-21 | 81.00 |
| *FABP9* | rs3809185 | G | A | 0.0615 | -0.203 | 0.015 | 5.0119E-44 | 183.15 |
| *FABP9* | rs7971732 | T | G | 0.2727 | 0.061 | 0.008 | 1.5849E-14 | 58.14 |
| *FABP9* | rs150743616 | C | T | 0.0015 | 0.87 | 0.093 | 7.9433E-21 | 87.51 |
| *FABP9* | rs765892649 | AC | A | 0.4662 | -0.308 | 0.007 | 1E-200 | 1936.00 |
| *FABP9* | rs12457893 | A | C | 0.4575 | -0.068 | 0.007 | 1.2589E-21 | 94.37 |

| | | | | | | | | |
|---|---|---|---|---|---|---|---|---|
| *FABP9* | rs867348 | A | T | 0.6781 | -0.091 | 0.008 | 1.9953E-32 | 129.39 |
| *FADD* | rs342298 | C | T | 0.4549 | -0.051 | 0.008 | 1.2589E-11 | 40.64 |
| *FADD* | rs3847326 | A | G | 0.4817 | -0.051 | 0.007 | 6.3096E-12 | 53.08 |
| *FADD* | rs10898847 | G | A | 0.408 | 0.08 | 0.008 | 1.9953E-25 | 100.00 |
| *FADD* | rs60822569 | T | TTTCTC | 0.5541 | 0.064 | 0.008 | 3.1623E-17 | 64.00 |
| *FAM13A* | rs1354034 | T | C | 0.6046 | 0.072 | 0.008 | 7.9433E-22 | 81.00 |
| *FAM13A* | rs34262577 | A | AAGGG | 0.1691 | -0.167 | 0.01 | 7.9433E-63 | 278.89 |
| *FAM13A* | rs342293 | C | G | 0.4598 | 0.113 | 0.007 | 1.2589E-52 | 260.59 |
| *FAM13A* | rs60757417 | C | G | 0.0605 | -0.132 | 0.016 | 3.1623E-17 | 68.06 |
| *FAM13A* | rs1054852 | A | G | 0.3752 | 0.059 | 0.008 | 1E-13 | 54.39 |
| *FAM171A2* | rs139026664 | C | A | 0.084 | 0.129 | 0.014 | 3.9811E-19 | 84.90 |
| *FAM171B* | rs3886170 | C | T | 0.2787 | -0.377 | 0.008 | 1E-200 | 2220.77 |
| *FAM171B* | rs7635884 | C | T | 0.6405 | 0.056 | 0.008 | 2.5119E-13 | 49.00 |
| *FAM171B* | rs8176741 | G | A | 0.0612 | 0.11 | 0.015 | 7.9433E-13 | 53.78 |
| *FAM172A* | rs1354034 | T | C | 0.6045 | 0.053 | 0.008 | 5.0119E-12 | 43.89 |
| *FAM172A* | rs1470150 | T | C | 0.1318 | 0.191 | 0.011 | 1E-65 | 301.50 |
| *FAM172A* | rs113708600 | A | C | 0.0666 | 0.119 | 0.015 | 5.0119E-15 | 62.94 |
| *FAM20A* | rs1260326 | T | C | 0.6086 | -0.096 | 0.007 | 1.2589E-40 | 188.08 |
| *FAM20A* | rs249641 | A | G | 0.6176 | 0.052 | 0.007 | 1E-12 | 55.18 |
| *FAM20A* | rs451637 | C | T | 0.3096 | -0.058 | 0.008 | 1.5849E-13 | 52.56 |
| *FAM20A* | rs150231592 | G | A | 0.0438 | -0.18 | 0.019 | 3.9811E-21 | 89.75 |
| *FAM20A* | rs35332062 | G | A | 0.1267 | -0.099 | 0.011 | 1.2589E-20 | 81.00 |
| *FAM20A* | rs2001846 | T | C | 0.5197 | -0.055 | 0.007 | 1E-14 | 61.73 |
| *FAM20A* | rs3841602 | A | AGGCGGC | 0.4684 | 0.051 | 0.007 | 5.0119E-13 | 53.08 |
| *FAM20A* | rs2622935 | A | G | 0.8071 | -0.138 | 0.009 | 1.2589E-53 | 235.11 |
| *FAM20A* | rs10778152 | A | G | 0.2876 | -0.12 | 0.008 | 1.5849E-52 | 225.00 |
| *FAM20A* | rs28929474 | C | T | 0.0209 | 0.167 | 0.025 | 1E-11 | 44.62 |
| *FAM20A* | rs704 | G | A | 0.4733 | -0.084 | 0.007 | 2.5119E-31 | 144.00 |
| *FAM20A* | rs2952271 | G | A | 0.6531 | -0.239 | 0.008 | 1E-200 | 892.52 |
| *FAM20A* | rs7247412 | T | C | 0.2666 | -0.068 | 0.008 | 1.2589E-17 | 72.25 |
| *FAM3B* | rs3129777 | C | A | 0.2152 | -0.115 | 0.009 | 6.3096E-37 | 163.27 |
| *FAM3B* | rs2519093 | C | T | 0.1845 | -0.108 | 0.009 | 5.0119E-34 | 144.00 |
| *FAM3B* | rs2559850 | G | A | 0.587 | 0.056 | 0.007 | 1.9953E-15 | 64.00 |
| *FAM3B* | rs548921462 | C | CT | 0.544 | -0.052 | 0.007 | 1.9953E-12 | 55.18 |
| *FAM3B* | rs17743980 | G | A | 0.3144 | 0.058 | 0.007 | 3.9811E-15 | 68.65 |
| *FAM3B* | rs708686 | C | T | 0.2673 | 0.253 | 0.008 | 1E-200 | 1000.14 |
| *FAM3B* | rs602662 | G | A | 0.5362 | 0.25 | 0.007 | 1E-200 | 1275.51 |
| *FAM3B* | rs57529409 | C | T | 0.0472 | -1.153 | 0.017 | 1E-200 | 4600.03 |
| *FAM3C* | rs36198735 | A | AAAC | 0.733 | -0.114 | 0.008 | 5.0119E-43 | 203.06 |
| *FAM3C* | rs36060036 | C | T | 0.1678 | -0.073 | 0.01 | 6.3096E-14 | 53.29 |
| *FAM3C* | rs9889902 | T | C | 0.3421 | 0.059 | 0.008 | 1.5849E-14 | 54.39 |
| *FAM3D* | rs12493131 | T | G | 0.9037 | 0.492 | 0.011 | 1E-200 | 2000.53 |
| *FAM3D* | rs4556017 | C | T | 0.8533 | -0.089 | 0.009 | 2.5119E-23 | 97.79 |
| *FAM3D* | rs550057 | C | T | 0.2554 | 0.713 | 0.008 | 1E-200 | 7943.27 |
| *FAM3D* | rs708686 | C | T | 0.2677 | 0.122 | 0.008 | 1.2589E-57 | 232.56 |
| *FAM3D* | rs681343 | C | T | 0.5092 | -0.478 | 0.007 | 1E-200 | 4662.94 |
| *FAP* | rs1751479 | G | A | 0.6457 | -0.064 | 0.008 | 5.0119E-17 | 64.00 |
| *FAP* | rs4664454 | C | G | 0.0101 | 1.075 | 0.037 | 3.981E-187 | 844.14 |
| *FAP* | rs9266252 | G | A | 0.7009 | 0.118 | 0.008 | 2.5119E-49 | 217.56 |
| *FAP* | rs74044565 | G | C | 0.1385 | 0.087 | 0.011 | 2.5119E-16 | 62.55 |
| *FAP* | rs34885055 | C | G | 0.1195 | 0.16 | 0.011 | 1.2589E-46 | 211.57 |
| *FAP* | rs740516 | C | G | 0.1486 | -0.149 | 0.01 | 1.9953E-47 | 222.01 |
| *FARSA* | rs2576005 | A | T | 0.85 | 0.081 | 0.011 | 7.9433E-14 | 54.22 |
| *FAS* | rs113705304 | C | G | 0.0888 | 0.152 | 0.012 | 2.5119E-36 | 160.44 |
| *FAS* | rs10887882 | T | A | 0.5849 | -0.257 | 0.007 | 1E-200 | 1347.94 |
| *FAS* | rs123698 | G | C | 0.6043 | 0.051 | 0.007 | 3.1623E-13 | 53.08 |
| *FAS* | rs738409 | C | G | 0.217 | 0.06 | 0.008 | 3.1623E-13 | 56.25 |
| *FASLG* | rs79419452 | T | A | 0.1562 | -0.099 | 0.009 | 1.5849E-26 | 121.00 |
| *FASLG* | rs11127048 | G | A | 0.6182 | -0.051 | 0.007 | 3.1623E-13 | 53.08 |
| *FASLG* | rs13063578 | T | A | 0.3978 | -0.169 | 0.007 | 1E-132 | 582.88 |
| *FASLG* | rs13111860 | A | G | 0.7123 | -0.054 | 0.007 | 5.0119E-13 | 59.51 |
| *FASLG* | rs9259888 | T | C | 0.647 | -0.079 | 0.008 | 1E-21 | 97.52 |
| *FASLG* | rs117343599 | G | C | 0.0229 | -0.174 | 0.023 | 3.1623E-14 | 57.23 |
| *FASLG* | rs3927680 | T | A | 0.6197 | 0.054 | 0.007 | 1E-14 | 59.51 |
| *FASLG* | rs8177655 | G | A | 0.3293 | 0.075 | 0.007 | 3.1623E-24 | 114.80 |
| *FASLG* | rs2646421 | G | C | 0.3793 | 0.052 | 0.007 | 5.0119E-13 | 55.18 |
| *FASLG* | rs978522 | T | C | 0.4051 | 0.291 | 0.007 | 1E-200 | 1728.18 |
| *FASLG* | rs745932882 | AG | A | 0.456 | 0.106 | 0.007 | 1.2589E-51 | 229.31 |

| | | | | | | | | |
|---|---|---|---|---|---|---|---|---|
| *FASLG* | rs648710 | A | T | 0.7434 | -0.109 | 0.008 | 5.0119E-45 | 185.64 |
| *FASLG* | rs11053802 | C | T | 0.6866 | -0.067 | 0.007 | 5.0119E-20 | 91.61 |
| *FASLG* | rs3184504 | T | C | 0.5174 | -0.164 | 0.007 | 7.943E-131 | 548.90 |
| *FASLG* | rs748988 | A | G | 0.6141 | 0.065 | 0.007 | 1.2589E-20 | 86.22 |
| *FASLG* | rs186021206 | G | A | 0.0064 | 0.611 | 0.047 | 1.9953E-39 | 169.00 |
| *FASLG* | rs9916257 | G | T | 0.4392 | -0.123 | 0.007 | 1.9953E-73 | 308.76 |
| *FASLG* | rs7221605 | C | T | 0.4987 | 0.053 | 0.007 | 3.9811E-15 | 57.33 |
| *FASLG* | rs1205340 | G | A | 0.5026 | 0.073 | 0.007 | 5.0119E-27 | 108.76 |
| *FASLG* | rs35477491 | C | T | 0.2238 | -0.091 | 0.01 | 1.9953E-21 | 82.81 |
| *FASLG* | rs12628239 | G | A | 0.3211 | -0.051 | 0.007 | 1.9953E-12 | 53.08 |
| *FASLG* | rs739141 | C | T | 0.6624 | -0.062 | 0.007 | 3.9811E-18 | 78.45 |
| *FBLN2* | rs333947 | G | A | 0.1482 | -0.07 | 0.01 | 2.5119E-12 | 49.00 |
| *FBLN2* | rs1260326 | T | C | 0.6083 | 0.054 | 0.007 | 1.2589E-13 | 59.51 |
| *FBLN2* | rs7623097 | A | G | 0.0063 | -1.563 | 0.046 | 1E-200 | 1154.52 |
| *FBLN2* | rs1010554 | T | C | 0.471 | -0.096 | 0.007 | 2.5119E-39 | 188.08 |
| *FBLN2* | rs10769256 | C | T | 0.4003 | 0.05 | 0.007 | 7.9433E-12 | 51.02 |
| *FBN2* | rs78821460 | G | C | 0.0835 | 0.545 | 0.014 | 1E-200 | 1515.43 |
| *FBP1* | rs499206 | A | G | 0.925 | 0.575 | 0.014 | 1E-200 | 1686.86 |
| *FBP1* | rs10883451 | T | C | 0.5018 | -0.057 | 0.007 | 7.9433E-15 | 66.31 |
| *FBP1* | rs79894691 | G | A | 0.085 | 0.095 | 0.013 | 3.9811E-13 | 53.40 |
| *FBP1* | rs10405357 | T | C | 0.4354 | -0.074 | 0.007 | 2.5119E-23 | 111.76 |
| *FBP1* | rs738409 | C | G | 0.2169 | 0.072 | 0.009 | 3.9811E-16 | 64.00 |
| *FCAMR* | rs188468174 | C | T | 0.0142 | 0.238 | 0.031 | 2.5119E-14 | 58.94 |
| *FCAMR* | rs646776 | C | T | 0.778 | 0.163 | 0.009 | 1.2589E-77 | 328.01 |
| *FCAMR* | rs11119933 | C | T | 0.1784 | 0.447 | 0.01 | 1E-200 | 1998.09 |
| *FCAMR* | rs1047891 | C | A | 0.3127 | 0.058 | 0.008 | 2.5119E-14 | 52.56 |
| *FCAMR* | rs56819515 | T | A | 0.1249 | -0.083 | 0.011 | 7.9433E-15 | 56.93 |
| *FCAMR* | rs2884401 | G | C | 0.7441 | -0.055 | 0.008 | 7.9433E-12 | 47.27 |
| *FCAMR* | rs11154519 | G | A | 0.5741 | 0.056 | 0.007 | 6.3096E-15 | 64.00 |
| *FCAMR* | rs4418728 | G | T | 0.4511 | -0.07 | 0.007 | 2.5119E-23 | 100.00 |
| *FCAMR* | rs118088990 | G | A | 0.0608 | -0.119 | 0.015 | 6.3096E-16 | 62.94 |
| *FCAMR* | rs61755050 | T | C | 0.006 | -0.405 | 0.046 | 7.9433E-19 | 77.52 |
| *FCAMR* | rs111338191 | A | AAATT | 0.5207 | -0.057 | 0.007 | 1E-15 | 66.31 |
| *FCAMR* | rs28929474 | C | T | 0.0209 | 0.203 | 0.025 | 1.5849E-16 | 65.93 |
| *FCAMR* | rs78357146 | A | G | 0.0303 | 0.143 | 0.02 | 2.5119E-12 | 51.12 |
| *FCAMR* | rs344560 | T | C | 0.9454 | 0.121 | 0.016 | 7.9433E-15 | 57.19 |
| *FCAMR* | rs2207132 | G | A | 0.0326 | -0.159 | 0.02 | 1E-15 | 63.20 |
| *FCAMR* | rs1800961 | C | T | 0.0323 | 0.155 | 0.02 | 5.0119E-15 | 60.06 |
| *FCAMR* | rs2266959 | G | T | 0.1874 | 0.086 | 0.009 | 1.2589E-21 | 91.31 |
| *FCAMR* | rs738409 | C | G | 0.2172 | 0.074 | 0.008 | 1.9953E-18 | 85.56 |
| *FCAR* | rs188468174 | C | T | 0.0143 | -0.579 | 0.027 | 1.259E-105 | 459.86 |
| *FCAR* | rs2070901 | G | T | 0.2703 | 0.052 | 0.007 | 7.9433E-14 | 55.18 |
| *FCAR* | rs9862795 | A | T | 0.5299 | 0.045 | 0.006 | 5.0119E-13 | 56.25 |
| *FCAR* | rs2853950 | C | T | 0.4365 | 0.06 | 0.006 | 6.3096E-22 | 100.00 |
| *FCAR* | rs11155297 | G | T | 0.2635 | -0.048 | 0.007 | 1.5849E-11 | 47.02 |
| *FCAR* | rs6796 | T | C | 0.2788 | 0.093 | 0.007 | 1E-40 | 176.51 |
| *FCAR* | rs73017385 | G | A | 0.2403 | 0.093 | 0.007 | 1.9953E-37 | 176.51 |
| *FCAR* | rs3184504 | T | C | 0.5174 | -0.081 | 0.006 | 2.5119E-39 | 182.25 |
| *FCAR* | rs186021206 | G | A | 0.0064 | 0.648 | 0.043 | 1.2589E-51 | 227.10 |
| *FCAR* | rs3826331 | T | C | 0.5985 | -0.081 | 0.006 | 3.9811E-37 | 182.25 |
| *FCAR* | rs12608573 | C | G | 0.2176 | 0.844 | 0.009 | 1E-200 | 8794.27 |
| *FCER1A* | rs61828219 | C | A | 0.1656 | 0.174 | 0.01 | 7.9433E-65 | 302.76 |
| *FCER2* | rs1432296 | C | T | 0.1419 | 0.074 | 0.01 | 5.0119E-13 | 54.76 |
| *FCER2* | rs112824187 | G | A | 0.0764 | -0.149 | 0.014 | 1.9953E-28 | 113.27 |
| *FCER2* | rs2844606 | A | G | 0.4179 | -0.089 | 0.007 | 2.5119E-34 | 161.65 |
| *FCER2* | rs9488914 | C | T | 0.3568 | 0.095 | 0.008 | 7.9433E-37 | 141.02 |
| *FCER2* | rs17494629 | T | G | 0.2017 | -0.06 | 0.009 | 1.2589E-11 | 44.44 |
| *FCER2* | rs62502726 | T | C | 0.1512 | -0.077 | 0.01 | 1E-14 | 59.29 |
| *FCER2* | rs562240450 | AC | A | 0.3178 | -0.058 | 0.008 | 7.9433E-14 | 52.56 |
| *FCER2* | rs7087507 | A | G | 0.3494 | 0.053 | 0.007 | 1.2589E-12 | 57.33 |
| *FCER2* | rs12784975 | T | C | 0.1919 | -0.078 | 0.009 | 3.9811E-18 | 75.11 |
| *FCER2* | rs111338191 | A | AAATT | 0.5212 | -0.05 | 0.007 | 3.1623E-12 | 51.02 |
| *FCER2* | rs374039502 | T | A | 0.0206 | 0.193 | 0.027 | 5.0119E-13 | 51.10 |
| *FCER2* | rs9939427 | G | A | 0.2308 | 0.069 | 0.009 | 6.3096E-16 | 58.78 |
| *FCER2* | rs35222145 | T | G | 0.4921 | -0.092 | 0.007 | 6.3096E-37 | 172.73 |
| *FCER2* | rs2277989 | A | G | 0.4874 | -0.292 | 0.007 | 1E-200 | 1740.08 |
| *FCGR2A* | rs11810143 | A | G | 0.1174 | -1.474 | 0.012 | 1E-200 | 15088.03 |
| *FCGR2A* | rs62165726 | C | A | 0.0369 | -0.116 | 0.015 | 3.1623E-14 | 59.80 |

| | | | | | | | | |
|---|---|---|---|---|---|---|---|---|
| *FCGR2A* | rs782819119 | AAGAAGGG | A | 0.3102 | 0.048 | 0.006 | 2.5119E-14 | 64.00 |
| *FCGR2A* | rs75071241 | G | A | 0.0598 | -0.086 | 0.012 | 1.2589E-12 | 51.36 |
| *FCGR2A* | rs7137828 | C | T | 0.5156 | -0.055 | 0.006 | 7.9433E-22 | 84.03 |
| *FCGR2A* | rs186021206 | G | A | 0.0064 | 1.087 | 0.04 | 1E-164 | 738.48 |
| *FCGR2B* | rs1801274 | A | G | 0.5344 | -0.451 | 0.008 | 1E-200 | 3178.14 |
| *FCGR2B* | rs4240356 | C | G | 0.548 | -0.049 | 0.007 | 1.2589E-11 | 49.00 |
| *FCGR2B* | rs505922 | T | C | 0.3164 | -0.176 | 0.008 | 1E-112 | 484.00 |
| *FCGR2B* | rs6115999 | C | T | 0.0119 | 0.298 | 0.034 | 1.2589E-18 | 76.82 |
| *FCGR3B* | rs3762281 | A | G | 0.5047 | 0.08 | 0.007 | 3.1623E-27 | 130.61 |
| *FCGR3B* | rs200688856 | G | C | 0.341 | -0.862 | 0.008 | 1E-200 | 11610.06 |
| *FCGR3B* | rs34860591 | T | C | 0.0848 | 0.087 | 0.012 | 5.0119E-14 | 52.56 |
| *FCGR3B* | rs28371261 | G | A | 0.1134 | -0.076 | 0.01 | 6.3096E-14 | 57.76 |
| *FCGR3B* | rs6796 | T | C | 0.2788 | 0.061 | 0.007 | 1.2589E-17 | 75.94 |
| *FCGR3B* | rs7795319 | C | T | 0.3448 | -0.058 | 0.007 | 1.2589E-17 | 68.65 |
| *FCGR3B* | rs73017399 | G | C | 0.2392 | 0.2 | 0.008 | 1.585E-152 | 625.00 |
| *FCGR3B* | rs7137828 | C | T | 0.5158 | -0.073 | 0.006 | 5.0119E-30 | 148.03 |
| *FCGR3B* | rs10138570 | A | G | 0.5317 | -0.07 | 0.006 | 3.1623E-27 | 136.11 |
| *FCGR3B* | rs36082322 | T | C | 0.1096 | 0.079 | 0.01 | 1.2589E-14 | 62.41 |
| *FCGR3B* | rs186021206 | G | A | 0.0064 | 0.612 | 0.044 | 1.2589E-43 | 193.46 |
| *FCGR3B* | rs2241244 | G | C | 0.537 | -0.084 | 0.006 | 6.3096E-39 | 196.00 |
| *FCGR3B* | rs4760 | A | G | 0.1564 | -0.279 | 0.009 | 1E-200 | 961.00 |
| *FCN1* | rs72550870 | T | C | 0.0303 | 0.173 | 0.02 | 1.2589E-17 | 74.82 |
| *FCN1* | rs6542680 | C | T | 0.8214 | 0.158 | 0.009 | 5.0119E-67 | 308.20 |
| *FCN1* | rs1486236 | C | A | 0.3733 | 0.14 | 0.007 | 6.3096E-81 | 400.00 |
| *FCN1* | rs2304456 | T | G | 0.111 | 0.147 | 0.011 | 1.9953E-39 | 178.59 |
| *FCN1* | rs115216147 | T | A | 0.0666 | -0.105 | 0.014 | 1.9953E-13 | 56.25 |
| *FCN1* | rs4585607 | A | G | 0.4979 | 0.064 | 0.008 | 7.9433E-17 | 64.00 |
| *FCN1* | rs199980439 | C | CA | 0.1224 | -0.075 | 0.011 | 1.5849E-12 | 46.49 |
| *FCN1* | rs41341748 | G | A | 0.011 | 0.335 | 0.033 | 1E-23 | 103.05 |
| *FCN1* | rs11103604 | G | T | 0.2661 | 0.442 | 0.008 | 1E-200 | 3052.56 |
| *FCN1* | rs2511241 | C | T | 0.9296 | 0.105 | 0.014 | 1.9953E-14 | 56.25 |
| *FCN1* | rs112771035 | C | G | 0.0713 | -0.248 | 0.014 | 6.3096E-74 | 313.80 |
| *FCN1* | rs6503533 | C | T | 0.616 | -0.052 | 0.007 | 6.3096E-13 | 55.18 |
| *FCN1* | rs12986064 | T | C | 0.508 | -0.055 | 0.007 | 2.5119E-14 | 61.73 |
| *FCN1* | rs6115999 | C | T | 0.0118 | 0.293 | 0.033 | 3.1623E-19 | 78.83 |
| *FCN2* | rs72550870 | T | C | 0.0304 | -0.216 | 0.018 | 2.5119E-32 | 144.00 |
| *FCN2* | rs1260326 | T | C | 0.6081 | -0.045 | 0.006 | 5.0119E-12 | 56.25 |
| *FCN2* | rs35677470 | G | A | 0.0828 | 0.128 | 0.011 | 5.0119E-29 | 135.40 |
| *FCN2* | rs62276455 | T | A | 0.4425 | -0.047 | 0.006 | 3.1623E-13 | 61.36 |
| *FCN2* | rs11386832 | T | TA | 0.7376 | 0.14 | 0.007 | 5.0119E-86 | 400.00 |
| *FCN2* | rs111331197 | C | T | 0.0111 | 0.278 | 0.03 | 3.1623E-20 | 85.87 |
| *FCN2* | rs7041446 | G | A | 0.4595 | -0.62 | 0.007 | 1E-200 | 7844.90 |
| *FCN2* | rs174533 | G | A | 0.3463 | 0.047 | 0.007 | 3.9811E-12 | 45.08 |
| *FCN2* | rs7122830 | G | A | 0.1329 | -0.473 | 0.01 | 1E-200 | 2237.29 |
| *FCN2* | rs7295693 | T | C | 0.5928 | 0.044 | 0.006 | 1.2589E-11 | 53.78 |
| *FCN2* | rs56205943 | G | A | 0.2436 | -0.055 | 0.007 | 6.3096E-14 | 61.73 |
| *FCRL1* | rs4971154 | C | T | 0.5031 | -0.358 | 0.007 | 1E-200 | 2615.59 |
| *FCRL1* | rs2009581 | G | A | 0.2726 | -0.055 | 0.008 | 5.0119E-12 | 47.27 |
| *FCRL1* | rs62292644 | A | C | 0.3855 | -0.068 | 0.007 | 2.5119E-20 | 94.37 |
| *FCRL1* | rs17361097 | G | A | 0.4383 | 0.056 | 0.007 | 1E-14 | 64.00 |
| *FCRL1* | rs112824187 | G | A | 0.0762 | -0.16 | 0.013 | 1.5849E-32 | 151.48 |
| *FCRL1* | rs1226013 | T | G | 0.551 | 0.097 | 0.007 | 7.9433E-42 | 192.02 |
| *FCRL1* | rs9280782 | C | CA | 0.5075 | -0.068 | 0.007 | 1E-19 | 94.37 |
| *FCRL1* | rs10692222 | C | CATT | 0.2755 | -0.083 | 0.008 | 1.9953E-25 | 107.64 |
| *FCRL1* | rs4245597 | G | A | 0.6542 | -0.111 | 0.007 | 2.5119E-50 | 251.45 |
| *FCRL1* | rs12784975 | T | C | 0.1921 | -0.072 | 0.009 | 1.2589E-15 | 64.00 |
| *FCRL1* | rs1035606 | C | T | 0.2838 | 0.054 | 0.008 | 6.3096E-12 | 45.56 |
| *FCRL1* | rs76428106 | T | C | 0.013 | 0.298 | 0.033 | 5.0119E-20 | 81.55 |
| *FCRL1* | rs374039502 | T | A | 0.0205 | 0.311 | 0.027 | 1.5849E-31 | 132.68 |
| *FCRL1* | rs9927316 | C | G | 0.2388 | 0.092 | 0.008 | 7.9433E-28 | 132.25 |
| *FCRL1* | rs186021206 | G | A | 0.0064 | 0.711 | 0.049 | 3.9811E-48 | 210.55 |
| *FCRL1* | rs34557412 | A | G | 0.0069 | 0.662 | 0.043 | 1.9953E-53 | 237.02 |
| *FCRL1* | rs35222145 | T | G | 0.4917 | -0.082 | 0.007 | 2.5119E-30 | 137.22 |
| *FCRL1* | rs77542162 | A | G | 0.0226 | 0.174 | 0.024 | 3.9811E-13 | 52.56 |
| *FCRL1* | rs79966604 | G | GGT | 0.3373 | 0.07 | 0.008 | 3.1623E-20 | 76.56 |
| *FCRL1* | rs73165110 | G | A | 0.0788 | -0.123 | 0.013 | 1E-20 | 89.52 |
| *FCRL2* | rs188468174 | C | T | 0.0143 | -0.298 | 0.031 | 1E-21 | 92.41 |
| *FCRL2* | rs12568320 | A | G | 0.1677 | 0.767 | 0.01 | 1E-200 | 5882.89 |

| | | | | | | | | |
|---|---|---|---|---|---|---|---|---|
| *FCRL2* | rs72832854 | T | G | 0.0249 | -0.15 | 0.021 | 1.5849E-12 | 51.02 |
| *FCRL2* | rs61696487 | C | CA | 0.1642 | -0.071 | 0.009 | 3.1623E-15 | 62.23 |
| *FCRL2* | rs5001409 | A | C | 0.3906 | -0.071 | 0.007 | 6.3096E-27 | 102.88 |
| *FCRL2* | rs9295574 | G | A | 0.4339 | 0.067 | 0.007 | 6.3096E-24 | 91.61 |
| *FCRL2* | rs114619738 | T | C | 0.0097 | 1.111 | 0.034 | 1E-200 | 1067.75 |
| *FCRL2* | rs778224678 | GACT | G | 0.114 | 0.1 | 0.01 | 1.2589E-22 | 100.00 |
| *FCRL2* | rs12554596 | A | G | 0.2038 | -0.071 | 0.008 | 1.2589E-18 | 78.77 |
| *FCRL2* | rs10791824 | A | G | 0.5661 | 0.061 | 0.007 | 7.9433E-21 | 75.94 |
| *FCRL2* | rs4766578 | T | A | 0.5047 | -0.053 | 0.006 | 1E-16 | 78.03 |
| *FCRL2* | rs374039502 | T | A | 0.0206 | 0.408 | 0.024 | 1.5849E-63 | 289.00 |
| *FCRL2* | rs186021206 | G | A | 0.0064 | 0.605 | 0.045 | 3.9811E-41 | 180.75 |
| *FCRL2* | rs34557412 | A | G | 0.0068 | 0.82 | 0.04 | 1.5849E-94 | 420.25 |
| *FCRL2* | rs753788823 | CAA | C | 0.2324 | -0.06 | 0.008 | 1.2589E-14 | 56.25 |
| *FCRL2* | rs34931250 | C | T | 0.0607 | 0.404 | 0.014 | 1E-189 | 832.73 |
| *FCRL2* | rs11085015 | T | G | 0.8006 | 0.095 | 0.008 | 1E-29 | 141.02 |
| *FCRL2* | rs73165110 | G | A | 0.0787 | -0.102 | 0.012 | 2.5119E-17 | 72.25 |
| *FCRL3* | rs7522061 | T | C | 0.4831 | 0.965 | 0.008 | 1E-200 | 14550.39 |
| *FCRL3* | rs2009581 | G | A | 0.2723 | -0.044 | 0.006 | 3.1623E-12 | 53.78 |
| *FCRL3* | rs7137828 | C | T | 0.5166 | -0.043 | 0.006 | 1.5849E-14 | 51.36 |
| *FCRL3* | rs186021206 | G | A | 0.0064 | 0.535 | 0.039 | 1.9953E-43 | 188.18 |
| *FCRL3* | rs34557412 | A | G | 0.0068 | 0.329 | 0.034 | 6.3096E-22 | 93.63 |
| *FCRL5* | rs188468174 | C | T | 0.0143 | -0.534 | 0.031 | 1.9953E-67 | 296.73 |
| *FCRL5* | rs61804164 | G | C | 0.1241 | -0.078 | 0.011 | 2.5119E-12 | 50.28 |
| *FCRL5* | rs7452226 | A | G | 0.5286 | -0.065 | 0.007 | 3.9811E-19 | 86.22 |
| *FCRL5* | rs6583437 | G | A | 0.3773 | -0.057 | 0.007 | 1.5849E-16 | 66.31 |
| *FCRL5* | rs111338191 | A | AAATT | 0.5212 | -0.056 | 0.007 | 1.5849E-16 | 64.00 |
| *FCRL5* | rs76428106 | T | C | 0.0128 | 0.233 | 0.031 | 7.9433E-14 | 56.49 |
| *FCRL5* | rs3803286 | A | G | 0.6659 | -0.066 | 0.007 | 3.9811E-20 | 88.90 |
| *FCRL5* | rs11624491 | C | T | 0.6564 | 0.168 | 0.007 | 3.162E-118 | 576.00 |
| *FCRL5* | rs3803800 | A | G | 0.7891 | -0.075 | 0.008 | 1.2589E-19 | 87.89 |
| *FCRL5* | rs34562254 | G | A | 0.097 | 0.164 | 0.011 | 1.9953E-47 | 222.28 |
| *FCRL5* | rs754292 | C | T | 0.5454 | -0.046 | 0.007 | 7.9433E-12 | 43.18 |
| *FCRL6* | rs55650803 | C | T | 0.1924 | -0.891 | 0.009 | 1E-200 | 9801.00 |
| *FCRL6* | rs2549006 | A | G | 0.6667 | 0.059 | 0.007 | 2.5119E-18 | 71.04 |
| *FCRL6* | rs9270747 | A | G | 0.6578 | 0.126 | 0.007 | 5.0119E-75 | 324.00 |
| *FCRL6* | rs482246 | A | G | 0.3388 | -0.074 | 0.007 | 1E-28 | 111.76 |
| *FCRL6* | rs3184504 | T | C | 0.5174 | -0.079 | 0.006 | 3.1623E-36 | 173.36 |
| *FCRL6* | rs186021206 | G | A | 0.0064 | 0.312 | 0.044 | 1E-12 | 50.28 |
| *FCRL6* | rs58745116 | G | A | 0.3901 | -0.085 | 0.007 | 1E-38 | 147.45 |
| *FCRL6* | rs2255166 | T | C | 0.4371 | 0.044 | 0.006 | 3.9811E-12 | 53.78 |
| *FCRLB* | rs188468174 | C | T | 0.0143 | -0.322 | 0.032 | 2.5119E-23 | 101.25 |
| *FCRLB* | rs61801180 | G | C | 0.0614 | 1.363 | 0.016 | 1E-200 | 7256.91 |
| *FCRLB* | rs1973612 | C | T | 0.5134 | -0.06 | 0.006 | 1E-22 | 100.00 |
| *FCRLB* | rs7789913 | C | T | 0.401 | 0.063 | 0.006 | 3.1623E-24 | 110.25 |
| *FCRLB* | rs73169662 | T | C | 0.1146 | 0.102 | 0.01 | 1.2589E-26 | 104.04 |
| *FCRLB* | rs7232 | T | A | 0.3729 | 0.044 | 0.006 | 3.1623E-12 | 53.78 |
| *FCRLB* | rs186021206 | G | A | 0.0063 | 0.34 | 0.042 | 1E-15 | 65.53 |
| *FEN1* | rs10418046 | T | G | 0.2122 | 0.064 | 0.009 | 2.5119E-12 | 50.57 |
| *FES* | rs1894401 | G | A | 0.523 | 0.133 | 0.008 | 1E-67 | 276.39 |
| *FETUB* | rs1862069 | G | A | 0.5376 | -0.05 | 0.007 | 1.2589E-12 | 51.02 |
| *FETUB* | rs114780909 | T | A | 0.0089 | -2.139 | 0.04 | 1E-200 | 2859.58 |
| *FETUB* | rs28601761 | C | G | 0.4193 | -0.118 | 0.007 | 1.2589E-60 | 284.16 |
| *FETUB* | rs11438680 | G | GA | 0.461 | 0.08 | 0.007 | 7.9433E-30 | 130.61 |
| *FETUB* | rs150844304 | A | C | 0.0241 | -0.265 | 0.023 | 1.9953E-31 | 132.75 |
| *FGA* | rs2228145 | A | C | 0.4064 | -0.051 | 0.007 | 3.9811E-12 | 53.08 |
| *FGA* | rs55709272 | T | C | 0.439 | 0.055 | 0.007 | 5.0119E-14 | 61.73 |
| *FGA* | rs140473879 | C | T | 0.0038 | -1.307 | 0.061 | 7.943E-101 | 459.08 |
| *FGA* | rs10900807 | G | C | 0.1835 | -0.079 | 0.009 | 5.0119E-17 | 77.05 |
| *FGD3* | rs6993770 | A | T | 0.2864 | -0.056 | 0.008 | 1E-11 | 49.00 |
| *FGF19* | rs2276890 | C | G | 0.3278 | 0.106 | 0.008 | 3.9811E-40 | 175.56 |
| *FGF19* | rs10504255 | G | A | 0.6654 | 0.098 | 0.008 | 3.9811E-35 | 150.06 |
| *FGF19* | rs8176741 | G | A | 0.0613 | 0.166 | 0.016 | 1.5849E-26 | 107.64 |
| *FGF19* | rs708686 | C | T | 0.268 | 0.075 | 0.009 | 7.9433E-19 | 69.44 |
| *FGF19* | rs492602 | A | G | 0.5096 | -0.225 | 0.008 | 3.162E-193 | 791.02 |
| *FGF2* | rs1434282 | C | T | 0.7261 | 0.058 | 0.008 | 1.2589E-14 | 52.56 |
| *FGF2* | rs1354034 | T | C | 0.6043 | 0.056 | 0.007 | 1.2589E-16 | 64.00 |
| *FGF2* | rs2922979 | C | G | 0.3095 | -0.711 | 0.008 | 1E-200 | 7898.77 |
| *FGF2* | rs6993770 | A | T | 0.2869 | -0.124 | 0.007 | 2.5119E-62 | 313.80 |

| | | | | | | | | |
|---|---|---|---|---|---|---|---|---|
| *FGF2* | rs3780420 | T | C | 0.3644 | 0.053 | 0.007 | 5.0119E-14 | 57.33 |
| *FGF2* | rs34416903 | T | C | 0.2799 | 0.053 | 0.008 | 1.9953E-12 | 43.89 |
| *FGF21* | rs1260326 | T | C | 0.6086 | -0.136 | 0.008 | 7.9433E-71 | 289.00 |
| *FGF21* | rs1047891 | C | A | 0.3126 | -0.064 | 0.008 | 2.5119E-15 | 64.00 |
| *FGF21* | rs13234378 | A | T | 0.1272 | -0.157 | 0.011 | 3.9811E-45 | 203.71 |
| *FGF21* | rs3779195 | T | A | 0.1846 | -0.079 | 0.01 | 2.5119E-16 | 62.41 |
| *FGF21* | rs2954021 | A | G | 0.504 | -0.055 | 0.007 | 1.2589E-13 | 61.73 |
| *FGF21* | rs62074055 | G | C | 0.5031 | -0.054 | 0.007 | 3.1623E-13 | 59.51 |
| *FGF21* | rs838131 | A | C | 0.4566 | -0.123 | 0.008 | 5.0119E-58 | 236.39 |
| *FGF23* | rs17199004 | T | G | 0.2644 | 0.077 | 0.008 | 1E-19 | 92.64 |
| *FGF23* | rs6862195 | G | T | 0.3109 | -0.07 | 0.008 | 3.9811E-18 | 76.56 |
| *FGF23* | rs34357864 | G | GA | 0.3175 | 0.086 | 0.008 | 5.0119E-27 | 115.56 |
| *FGF23* | rs6489536 | G | C | 0.3385 | 0.108 | 0.008 | 1.2589E-43 | 182.25 |
| *FGF23* | rs6561643 | A | T | 0.3468 | 0.077 | 0.008 | 3.9811E-22 | 92.64 |
| *FGF23* | rs6127099 | A | T | 0.2777 | -0.096 | 0.008 | 7.9433E-30 | 144.00 |
| *FGF5* | rs12509595 | T | C | 0.2934 | 0.682 | 0.008 | 1E-200 | 7267.56 |
| *FGF5* | rs17580 | T | A | 0.0496 | 0.112 | 0.014 | 5.0119E-15 | 64.00 |
| *FGFBP1* | rs7528419 | A | G | 0.2221 | -0.106 | 0.009 | 1.9953E-34 | 138.72 |
| *FGFBP1* | rs61816761 | G | A | 0.0236 | 0.175 | 0.025 | 3.1623E-12 | 49.00 |
| *FGFBP1* | rs1260326 | T | C | 0.6087 | 0.051 | 0.007 | 7.9433E-12 | 53.08 |
| *FGFBP1* | rs6800909 | T | G | 0.2947 | 0.064 | 0.008 | 6.3096E-16 | 64.00 |
| *FGFBP1* | rs9968276 | G | A | 0.2024 | -0.121 | 0.009 | 6.3096E-41 | 180.75 |
| *FGFBP1* | rs61738275 | C | T | 0.1515 | 0.071 | 0.01 | 1.9953E-12 | 50.41 |
| *FGFBP1* | rs3184504 | T | C | 0.5173 | 0.056 | 0.007 | 3.9811E-15 | 64.00 |
| *FGFBP1* | rs1532085 | A | G | 0.6168 | -0.051 | 0.007 | 7.9433E-12 | 53.08 |
| *FGFBP1* | rs77542162 | A | G | 0.0228 | 0.179 | 0.024 | 1E-13 | 55.63 |
| *FGFBP1* | rs151113958 | A | AG | 0.1196 | -0.083 | 0.011 | 1E-13 | 56.93 |
| *FGFBP1* | rs1065853 | G | T | 0.0815 | -0.282 | 0.013 | 6.31E-100 | 470.56 |
| *FGFBP1* | rs2074735 | G | C | 0.0636 | 0.145 | 0.015 | 7.9433E-23 | 93.44 |
| *FGFBP2* | rs900400 | T | C | 0.398 | -0.056 | 0.007 | 1.2589E-17 | 64.00 |
| *FGFBP2* | rs35700679 | A | G | 0.1483 | 0.719 | 0.01 | 1E-200 | 5169.61 |
| *FGFBP2* | rs9272302 | C | T | 0.1653 | -0.081 | 0.009 | 1E-19 | 81.00 |
| *FGFBP2* | rs836468 | T | A | 0.3118 | 0.048 | 0.007 | 1.5849E-12 | 47.02 |
| *FGFBP2* | rs2740488 | A | C | 0.2668 | -0.058 | 0.007 | 7.9433E-16 | 68.65 |
| *FGFBP2* | rs2070895 | G | A | 0.2214 | 0.076 | 0.008 | 3.9811E-23 | 90.25 |
| *FGFBP2* | rs36229491 | T | TA | 0.324 | 0.092 | 0.007 | 6.3096E-42 | 172.73 |
| *FGFBP2* | rs34931250 | C | T | 0.0606 | 1.176 | 0.015 | 1E-200 | 6146.56 |
| *FGFBP2* | rs77960347 | A | G | 0.0131 | 0.209 | 0.028 | 7.9433E-14 | 55.72 |
| *FGFBP2* | rs1065853 | G | T | 0.0812 | 0.099 | 0.012 | 3.1623E-17 | 68.06 |
| *FGFBP3* | rs12740374 | G | T | 0.2217 | -0.097 | 0.008 | 3.9811E-31 | 147.02 |
| *FGFBP3* | rs1434282 | C | T | 0.7265 | -0.055 | 0.008 | 2.5119E-12 | 47.27 |
| *FGFBP3* | rs11711157 | C | T | 0.2782 | -0.068 | 0.008 | 1.9953E-18 | 72.25 |
| *FGFBP3* | rs11734099 | G | A | 0.1742 | 0.065 | 0.009 | 1.9953E-12 | 52.16 |
| *FGFBP3* | rs6923313 | C | T | 0.6562 | -0.055 | 0.008 | 3.1623E-13 | 47.27 |
| *FGFBP3* | rs4252117 | A | G | 0.2952 | -0.345 | 0.008 | 1E-200 | 1859.77 |
| *FGFBP3* | rs13269700 | A | G | 0.3232 | 0.054 | 0.007 | 5.0119E-13 | 59.51 |
| *FGFBP3* | rs6993770 | A | T | 0.2865 | -0.08 | 0.008 | 2.5119E-25 | 100.00 |
| *FGFBP3* | rs555601744 | C | CA | 0.3115 | -0.39 | 0.008 | 1E-200 | 2376.56 |
| *FGFBP3* | rs61751507 | C | T | 0.0435 | -0.252 | 0.018 | 3.9811E-46 | 196.00 |
| *FGFBP3* | rs17127600 | T | G | 0.1704 | -0.078 | 0.009 | 2.5119E-17 | 75.11 |
| *FGFBP3* | rs61658003 | C | A | 0.8428 | 0.087 | 0.01 | 3.9811E-19 | 75.69 |
| *FGFR2* | rs924204 | A | G | 0.5895 | 0.059 | 0.007 | 2.5119E-16 | 71.04 |
| *FGFR2* | rs6968170 | G | A | 0.2834 | -0.058 | 0.008 | 1.5849E-13 | 52.56 |
| *FGFR2* | rs7826120 | T | C | 0.67 | 0.058 | 0.008 | 1E-14 | 52.56 |
| *FGFR2* | rs2001945 | G | C | 0.5191 | -0.057 | 0.007 | 6.3096E-16 | 66.31 |
| *FGFR2* | rs2981430 | G | A | 0.5803 | -0.113 | 0.007 | 5.0119E-56 | 260.59 |
| *FGFR2* | rs111632263 | T | A | 0.1277 | 0.074 | 0.011 | 3.1623E-12 | 45.26 |
| *FGFR2* | rs186021206 | G | A | 0.0064 | 0.519 | 0.048 | 7.9433E-27 | 116.91 |
| *FGFR4* | rs10753239 | T | C | 0.4601 | -0.043 | 0.005 | 1.5849E-16 | 73.96 |
| *FGFR4* | rs5402 | T | A | 0.1197 | -0.086 | 0.008 | 1.5849E-27 | 115.56 |
| *FGFR4* | rs6855868 | G | A | 0.3279 | 0.397 | 0.006 | 1E-200 | 4378.03 |
| *FGFR4* | rs351855 | G | A | 0.296 | 1.009 | 0.008 | 1E-200 | 15907.52 |
| *FGFR4* | rs2253491 | G | A | 0.205 | 0.089 | 0.006 | 6.3096E-44 | 220.03 |
| *FGFR4* | rs796332416 | TA | T | 0.2484 | -0.04 | 0.006 | 1.5849E-11 | 44.44 |
| *FGFR4* | rs7799998 | A | G | 0.45 | -0.054 | 0.005 | 1E-25 | 116.64 |
| *FGFR4* | rs11783515 | A | G | 0.6795 | 0.043 | 0.006 | 5.0119E-15 | 51.36 |
| *FGFR4* | rs1131773 | A | G | 0.1443 | 0.058 | 0.007 | 1.5849E-15 | 68.65 |
| *FGFR4* | rs9633740 | A | G | 0.7961 | 0.068 | 0.006 | 6.3096E-27 | 128.44 |

| | | | | | | | | |
|---|---|---|---|---|---|---|---|---|
| *FGFR4* | rs174581 | G | A | 0.3543 | 0.055 | 0.005 | 6.3096E-25 | 121.00 |
| *FGFR4* | rs11045819 | C | A | 0.161 | 0.1 | 0.007 | 3.1623E-47 | 204.08 |
| *FGFR4* | rs1169288 | A | C | 0.3143 | 0.056 | 0.006 | 1.2589E-23 | 87.11 |
| *FGFR4* | rs112001035 | G | A | 0.0614 | -0.105 | 0.011 | 5.0119E-22 | 91.12 |
| *FGFR4* | rs41292412 | C | T | 0.0127 | 0.506 | 0.023 | 5.012E-104 | 484.00 |
| *FGFR4* | rs33950747 | C | T | 0.0753 | 0.078 | 0.01 | 1.2589E-15 | 60.84 |
| *FGL1* | rs1260326 | T | C | 0.6086 | -0.093 | 0.006 | 1.9953E-53 | 240.25 |
| *FGL1* | rs3739404 | C | A | 0.691 | 0.657 | 0.008 | 1E-200 | 6744.52 |
| *FGL1* | rs2351958 | C | A | 0.3867 | 0.048 | 0.006 | 2.5119E-15 | 64.00 |
| *FGL1* | rs10083137 | A | G | 0.0345 | 0.111 | 0.016 | 7.9433E-12 | 48.13 |
| *FGL1* | rs12444166 | G | A | 0.3222 | 0.046 | 0.006 | 5.0119E-13 | 58.78 |
| *FGL1* | rs186021206 | G | A | 0.0064 | -0.276 | 0.041 | 1.2589E-11 | 45.32 |
| *FGL1* | rs731170 | A | G | 0.6865 | -0.043 | 0.006 | 1E-11 | 51.36 |
| *FGR* | rs3737803 | G | A | 0.0735 | -0.179 | 0.014 | 5.0119E-36 | 163.47 |
| *FH* | rs141575023 | C | T | 0.1383 | 0.087 | 0.011 | 1.2589E-14 | 62.55 |
| *FHIT* | rs1354034 | T | C | 0.6035 | 0.063 | 0.008 | 3.1623E-16 | 62.02 |
| *FHIT* | rs13070475 | G | A | 0.6442 | 0.089 | 0.008 | 5.0119E-29 | 123.77 |
| *FIS1* | rs1354034 | T | C | 0.6042 | 0.063 | 0.008 | 1.9953E-16 | 62.02 |
| *FIS1* | rs7778271 | T | C | 0.0543 | -0.346 | 0.017 | 1.5849E-97 | 414.24 |
| *FIS1* | rs342293 | C | G | 0.4594 | -0.051 | 0.008 | 1.2589E-11 | 40.64 |
| *FIS1* | rs6993770 | A | T | 0.2866 | -0.056 | 0.008 | 1.5849E-11 | 49.00 |
| *FIS1* | rs10733789 | T | C | 0.3132 | 0.06 | 0.008 | 1.9953E-13 | 56.25 |
| *FIS1* | rs58807415 | A | G | 0.2438 | 0.059 | 0.009 | 1.5849E-11 | 42.98 |
| *FKBP14* | rs1354034 | T | C | 0.6043 | 0.111 | 0.008 | 1E-47 | 192.52 |
| *FKBP14* | rs114694170 | T | C | 0.06 | 0.113 | 0.016 | 1.2589E-12 | 49.88 |
| *FKBP14* | rs181397928 | G | C | 0.0008 | -1.018 | 0.142 | 6.3096E-13 | 51.39 |
| *FKBP14* | rs6993770 | A | T | 0.2864 | -0.068 | 0.008 | 3.9811E-16 | 72.25 |
| *FKBP14* | rs10820606 | A | C | 0.229 | -0.068 | 0.009 | 7.9433E-14 | 57.09 |
| *FKBP14* | rs11502185 | T | C | 0.2598 | 0.073 | 0.01 | 3.9811E-14 | 53.29 |
| *FKBP14* | rs351978 | A | G | 0.5831 | 0.052 | 0.008 | 1.2589E-11 | 42.25 |
| *FKBP1B* | rs872169 | C | G | 0.2049 | -0.255 | 0.009 | 5.012E-164 | 802.78 |
| *FKBP1B* | rs1354034 | T | C | 0.6042 | 0.081 | 0.008 | 1E-26 | 102.52 |
| *FKBP1B* | rs342296 | G | A | 0.4566 | -0.069 | 0.007 | 1.9953E-20 | 97.16 |
| *FKBP1B* | rs7916868 | A | T | 0.5016 | 0.072 | 0.007 | 6.3096E-22 | 105.80 |
| *FKBP1B* | rs11502185 | T | C | 0.2596 | 0.073 | 0.01 | 2.5119E-14 | 53.29 |
| *FKBP1B* | rs200309755 | C | T | 0.5372 | 0.066 | 0.008 | 1.5849E-17 | 68.06 |
| *FKBP4* | rs1354034 | T | C | 0.6041 | 0.084 | 0.008 | 1E-27 | 110.25 |
| *FKBP4* | rs342298 | C | T | 0.4544 | -0.057 | 0.008 | 6.3096E-14 | 50.77 |
| *FKBP4* | rs6993770 | A | T | 0.2865 | -0.062 | 0.008 | 7.9433E-14 | 60.06 |
| *FKBP4* | rs201265508 | A | G | 0.2935 | 0.071 | 0.009 | 5.0119E-14 | 62.23 |
| *FKBP4* | rs3814251 | T | C | 0.0386 | -0.134 | 0.02 | 1.2589E-11 | 44.89 |
| *FKBP4* | rs6580980 | A | G | 0.5651 | 0.062 | 0.008 | 7.9433E-16 | 60.06 |
| *FKBP4* | rs10774624 | G | A | 0.5133 | -0.058 | 0.008 | 3.9811E-14 | 52.56 |
| *FKBP5* | rs1354034 | T | C | 0.6041 | 0.052 | 0.008 | 7.9433E-12 | 42.25 |
| *FKBP5* | rs17622656 | G | A | 0.3887 | -0.052 | 0.008 | 7.9433E-12 | 42.25 |
| *FKBP5* | rs2817032 | T | C | 0.2869 | 0.209 | 0.008 | 5.012E-138 | 682.52 |
| *FKBP5* | rs342293 | C | G | 0.459 | -0.051 | 0.007 | 7.9433E-12 | 53.08 |
| *FKBPL* | rs1354034 | T | C | 0.6046 | 0.055 | 0.008 | 5.0119E-13 | 47.27 |
| *FKBPL* | rs9469087 | A | G | 0.0037 | -0.62 | 0.061 | 6.3096E-24 | 103.31 |
| *FLI1* | rs1354034 | T | C | 0.6038 | 0.06 | 0.008 | 1.2589E-14 | 56.25 |
| *FLI1* | rs6993770 | A | T | 0.2869 | -0.075 | 0.008 | 5.0119E-19 | 87.89 |
| *FLRT2* | rs3748770 | G | A | 0.2671 | 0.249 | 0.008 | 1E-200 | 968.77 |
| *FLRT2* | rs1260326 | T | C | 0.6084 | 0.052 | 0.007 | 1.9953E-13 | 55.18 |
| *FLRT2* | rs10935473 | G | T | 0.4404 | -0.056 | 0.007 | 3.9811E-16 | 64.00 |
| *FLRT2* | rs1363490 | A | C | 0.2841 | -0.055 | 0.008 | 3.9811E-13 | 47.27 |
| *FLRT2* | rs4841132 | A | G | 0.9081 | -0.117 | 0.012 | 1.2589E-22 | 95.06 |
| *FLRT2* | rs10901252 | G | C | 0.0614 | 0.103 | 0.014 | 5.0119E-13 | 54.13 |
| *FLRT2* | rs141826184 | C | T | 0.0045 | 0.429 | 0.051 | 5.0119E-17 | 70.76 |
| *FLRT2* | rs17646457 | G | A | 0.1506 | -0.699 | 0.01 | 1E-200 | 4886.01 |
| *FLRT2* | rs77542162 | A | G | 0.023 | 0.213 | 0.023 | 1.2589E-20 | 85.76 |
| *FLRT2* | rs6135225 | G | T | 0.305 | -0.195 | 0.008 | 3.162E-148 | 594.14 |
| *FLT1* | rs13088856 | T | G | 0.2976 | 0.065 | 0.008 | 2.5119E-16 | 66.02 |
| *FLT1* | rs13107325 | C | T | 0.0749 | 0.106 | 0.014 | 2.5119E-14 | 57.33 |
| *FLT1* | rs6921438 | G | A | 0.4906 | 0.345 | 0.008 | 1E-200 | 1859.77 |
| *FLT1* | rs6993770 | A | T | 0.2864 | 0.102 | 0.008 | 3.9811E-36 | 162.56 |
| *FLT1* | rs34881325 | C | T | 0.3763 | 0.059 | 0.008 | 1.9953E-14 | 54.39 |
| *FLT1* | rs2519093 | C | T | 0.1846 | -0.183 | 0.01 | 1.5849E-82 | 334.89 |
| *FLT1* | rs552252905 | C | G | 0.0012 | -1.145 | 0.113 | 3.1623E-24 | 102.67 |

| | | | | | | | | |
|---|---|---|---|---|---|---|---|---|
| *FLT1* | rs200489612 | G | A | 0.0052 | 0.508 | 0.055 | 3.9811E-20 | 85.31 |
| *FLT3* | rs2736100 | C | A | 0.4988 | -0.053 | 0.008 | 3.9811E-12 | 43.89 |
| *FLT3* | rs1847472 | C | A | 0.3542 | -0.059 | 0.008 | 6.3096E-14 | 54.39 |
| *FLT3* | rs4766578 | T | A | 0.5041 | -0.066 | 0.008 | 2.5119E-18 | 68.06 |
| *FLT3* | rs12579868 | C | T | 0.4699 | -0.073 | 0.008 | 1.2589E-21 | 83.27 |
| *FLT3* | rs9554228 | A | G | 0.6272 | -0.084 | 0.008 | 7.9433E-27 | 110.25 |
| *FLT3* | rs186021206 | G | A | 0.0063 | 0.639 | 0.053 | 1E-33 | 145.36 |
| *FLT3LG* | rs17534670 | G | A | 0.4927 | 0.093 | 0.007 | 5.0119E-40 | 176.51 |
| *FLT3LG* | rs2001703 | A | G | 0.619 | -0.117 | 0.007 | 1.5849E-59 | 279.37 |
| *FLT3LG* | rs6784404 | G | A | 0.4175 | -0.058 | 0.007 | 3.1623E-16 | 68.65 |
| *FLT3LG* | rs218265 | T | C | 0.1535 | -0.076 | 0.01 | 7.9433E-15 | 57.76 |
| *FLT3LG* | rs7705526 | C | A | 0.3249 | -0.123 | 0.008 | 3.1623E-59 | 236.39 |
| *FLT3LG* | rs56353819 | C | T | 0.3511 | 0.071 | 0.007 | 6.3096E-22 | 102.88 |
| *FLT3LG* | rs13219432 | T | C | 0.4568 | 0.065 | 0.007 | 5.0119E-20 | 86.22 |
| *FLT3LG* | rs42032 | G | A | 0.2617 | -0.057 | 0.008 | 1E-12 | 50.77 |
| *FLT3LG* | rs2256187 | G | A | 0.1537 | -0.068 | 0.01 | 2.5119E-12 | 46.24 |
| *FLT3LG* | rs1800056 | T | C | 0.014 | -0.245 | 0.03 | 1.9953E-16 | 66.69 |
| *FLT3LG* | rs4936290 | A | C | 0.3478 | 0.062 | 0.007 | 5.0119E-17 | 78.45 |
| *FLT3LG* | rs7310615 | C | G | 0.5177 | 0.055 | 0.007 | 5.0119E-15 | 61.73 |
| *FLT3LG* | rs76428106 | T | C | 0.0128 | 1.309 | 0.033 | 1E-200 | 1573.44 |
| *FLT3LG* | rs78378222 | T | G | 0.0128 | -0.22 | 0.032 | 5.0119E-12 | 47.27 |
| *FLT3LG* | rs80277818 | A | G | 0.1251 | -0.073 | 0.011 | 6.3096E-12 | 44.04 |
| *FLT3LG* | rs17758695 | C | T | 0.0292 | 0.194 | 0.021 | 7.9433E-21 | 85.34 |
| *FLT3LG* | rs117955557 | G | T | 0.2554 | 0.057 | 0.008 | 1.5849E-12 | 50.77 |
| *FLT3LG* | rs62237617 | C | T | 0.0027 | -0.586 | 0.075 | 6.3096E-15 | 61.05 |
| *FLT4* | rs11162623 | C | T | 0.3396 | 0.066 | 0.007 | 7.9433E-23 | 88.90 |
| *FLT4* | rs10935473 | G | T | 0.44 | -0.694 | 0.007 | 1E-200 | 9829.31 |
| *FLT4* | rs34221241 | T | C | 0.1083 | -0.443 | 0.01 | 1E-200 | 1962.49 |
| *FLT4* | rs6913664 | C | T | 0.4727 | -0.063 | 0.006 | 1.5849E-23 | 110.25 |
| *FLT4* | rs7019909 | C | T | 0.1025 | 0.083 | 0.011 | 1E-14 | 56.93 |
| *FLT4* | rs10981199 | T | C | 0.3559 | 0.053 | 0.007 | 7.9433E-15 | 57.33 |
| *FLT4* | rs2519093 | C | T | 0.1842 | -0.38 | 0.008 | 1E-200 | 2256.25 |
| *FLT4* | rs139130389 | C | CTA | 0.0726 | -0.137 | 0.012 | 3.1623E-29 | 130.34 |
| *FLT4* | rs186021206 | G | A | 0.0064 | 0.51 | 0.044 | 1E-31 | 134.35 |
| *FLT4* | rs1012383 | G | T | 0.8265 | -0.057 | 0.008 | 1.2589E-11 | 50.77 |
| *FLT4* | rs67525224 | G | C | 0.1853 | 0.08 | 0.008 | 1E-22 | 100.00 |
| *FLT4* | rs4611747 | C | T | 0.2884 | 0.062 | 0.007 | 7.9433E-19 | 78.45 |
| *FLT4* | rs132653 | G | T | 0.8177 | 0.056 | 0.008 | 7.9433E-12 | 49.00 |
| *FN1* | rs1250259 | T | A | 0.7365 | 0.448 | 0.008 | 1E-200 | 3136.00 |
| *FN1* | rs13108218 | A | G | 0.6192 | -0.051 | 0.008 | 1.2589E-11 | 40.64 |
| *FN1* | rs41341748 | G | A | 0.011 | 0.265 | 0.035 | 3.1623E-14 | 57.33 |
| *FNDC1* | rs755740174 | AT | A | 0.2912 | 0.058 | 0.008 | 5.0119E-14 | 52.56 |
| *FNDC1* | rs73588939 | A | G | 0.2667 | -0.428 | 0.008 | 1E-200 | 2862.25 |
| *FNDC1* | rs7047089 | C | T | 0.3707 | 0.076 | 0.007 | 6.3096E-26 | 117.88 |
| *FNDC1* | rs7178458 | T | C | 0.7158 | 0.138 | 0.008 | 1.9953E-70 | 297.56 |
| *FNTA* | rs12614 | C | T | 0.0847 | 0.144 | 0.014 | 3.1623E-26 | 105.80 |
| *FOLH1* | rs139299944 | C | CT | 0.3437 | 0.113 | 0.008 | 3.9811E-44 | 199.52 |
| *FOLH1* | rs61886488 | G | T | 0.0575 | 0.191 | 0.017 | 7.9433E-29 | 126.23 |
| *FOLH1* | rs139654989 | G | C | 0.0623 | 0.166 | 0.016 | 5.0119E-25 | 107.64 |
| *FOLR1* | rs423144 | G | T | 0.4257 | 0.057 | 0.007 | 3.1623E-15 | 66.31 |
| *FOLR1* | rs61747728 | C | T | 0.0379 | 0.139 | 0.019 | 6.3096E-14 | 53.52 |
| *FOLR1* | rs13107325 | C | T | 0.075 | 0.106 | 0.014 | 7.9433E-15 | 57.33 |
| *FOLR1* | rs10740131 | A | T | 0.4704 | 0.049 | 0.007 | 3.9811E-12 | 49.00 |
| *FOLR1* | rs115974400 | T | C | 0.0681 | -0.181 | 0.014 | 6.3096E-37 | 167.15 |
| *FOLR1* | rs777834943 | C | CT | 0.1833 | -0.064 | 0.009 | 5.0119E-12 | 50.57 |
| *FOLR1* | rs186021206 | G | A | 0.0064 | 0.725 | 0.049 | 1.2589E-49 | 218.92 |
| *FOLR1* | rs78060698 | G | A | 0.0408 | 0.208 | 0.018 | 1E-30 | 133.53 |
| *FOLR1* | rs681343 | C | T | 0.5092 | 0.116 | 0.007 | 1E-58 | 274.61 |
| *FOLR2* | rs333947 | G | A | 0.1484 | 0.082 | 0.01 | 2.5119E-16 | 67.24 |
| *FOLR2* | rs1260326 | T | C | 0.6085 | 0.058 | 0.007 | 3.9811E-15 | 68.65 |
| *FOLR2* | rs2470853 | A | G | 0.5746 | -0.068 | 0.007 | 6.3096E-21 | 94.37 |
| *FOLR2* | rs509227 | A | C | 0.5227 | 0.058 | 0.007 | 7.9433E-16 | 68.65 |
| *FOLR2* | rs80177780 | C | A | 0.7523 | 0.112 | 0.009 | 3.1623E-34 | 154.86 |
| *FOLR2* | rs17138358 | G | C | 0.3938 | -0.066 | 0.007 | 2.5119E-19 | 88.90 |
| *FOLR2* | rs7865362 | C | T | 0.3632 | 0.084 | 0.008 | 7.9433E-29 | 110.25 |
| *FOLR2* | rs2519093 | C | T | 0.1846 | 0.065 | 0.009 | 2.5119E-12 | 52.16 |
| *FOLR2* | rs603424 | G | A | 0.1665 | -0.107 | 0.01 | 3.9811E-29 | 114.49 |
| *FOLR2* | rs950803 | T | A | 0.6294 | 0.106 | 0.007 | 3.1623E-46 | 229.31 |

| | | | | | | | | |
|---|---|---|---|---|---|---|---|---|
| *FOLR2* | rs146893981 | C | T | 0.0294 | 0.321 | 0.021 | 1.2589E-51 | 233.65 |
| *FOLR2* | rs61729512 | G | A | 0.1178 | 0.261 | 0.011 | 2.512E-119 | 562.98 |
| *FOLR2* | rs551125 | C | T | 0.5813 | -0.049 | 0.007 | 1E-11 | 49.00 |
| *FOLR2* | rs4760 | A | G | 0.1561 | 0.144 | 0.01 | 2.5119E-48 | 207.36 |
| *FOLR2* | rs12975366 | T | C | 0.3955 | -0.143 | 0.007 | 1.5849E-84 | 417.33 |
| *FOLR3* | rs333947 | G | A | 0.1484 | 0.055 | 0.008 | 1.2589E-11 | 47.27 |
| *FOLR3* | rs62235167 | C | G | 0.3058 | 0.044 | 0.006 | 3.9811E-12 | 53.78 |
| *FOLR3* | rs2470872 | A | T | 0.5743 | -0.053 | 0.006 | 7.9433E-20 | 78.03 |
| *FOLR3* | rs70993902 | T | TCC | 0.7768 | 0.075 | 0.007 | 6.3096E-25 | 114.80 |
| *FOLR3* | rs10242866 | C | T | 0.3935 | -0.05 | 0.006 | 3.1623E-17 | 69.44 |
| *FOLR3* | rs2031902 | C | T | 0.5872 | -0.06 | 0.006 | 1.9953E-24 | 100.00 |
| *FOLR3* | rs603424 | G | A | 0.1664 | -0.071 | 0.008 | 6.3096E-20 | 78.77 |
| *FOLR3* | rs950802 | G | A | 0.3061 | 0.077 | 0.008 | 1.5849E-21 | 92.64 |
| *FOLR3* | rs139130389 | C | CTA | 0.0726 | 1.7 | 0.014 | 1E-200 | 14744.90 |
| *FOLR3* | rs61729512 | G | A | 0.118 | 0.16 | 0.009 | 6.3096E-70 | 316.05 |
| *FOLR3* | rs4760 | A | G | 0.156 | 0.08 | 0.008 | 1.2589E-23 | 100.00 |
| *FOLR3* | rs12975366 | T | C | 0.3953 | -0.094 | 0.006 | 7.9433E-57 | 245.44 |
| *FOXJ3* | rs1139978 | T | G | 0.1388 | -0.103 | 0.011 | 1.2589E-20 | 87.68 |
| *FOXJ3* | rs1354034 | T | C | 0.6039 | 0.06 | 0.008 | 6.3096E-15 | 56.25 |
| *FOXO1* | rs342298 | C | T | 0.4546 | -0.084 | 0.008 | 3.9811E-29 | 110.25 |
| *FOXO1* | rs10820606 | A | C | 0.2292 | -0.126 | 0.009 | 3.9811E-44 | 196.00 |
| *FOXO1* | rs7297265 | A | G | 0.4427 | 0.066 | 0.008 | 3.1623E-18 | 68.06 |
| *FOXO1* | rs17061453 | A | T | 0.0431 | 0.28 | 0.018 | 5.0119E-52 | 241.98 |
| *FOXO3* | rs34136174 | A | C | 0.4252 | 0.055 | 0.008 | 1.5849E-12 | 47.27 |
| *FRMD7* | rs147252207 | T | C | 0.0026 | -0.931 | 0.08 | 2.5119E-31 | 135.43 |
| *FRZB* | rs13009 | C | T | 0.0815 | -0.817 | 0.014 | 1E-200 | 3405.56 |
| *FRZB* | rs11452310 | T | TA | 0.5695 | 0.048 | 0.007 | 1.2589E-12 | 47.02 |
| *FRZB* | rs2856694 | T | C | 0.1855 | -0.153 | 0.009 | 6.3096E-63 | 289.00 |
| *FRZB* | rs3184504 | T | C | 0.5168 | -0.055 | 0.007 | 2.5119E-16 | 61.73 |
| *FRZB* | rs6018652 | G | A | 0.7913 | -0.058 | 0.008 | 1.5849E-12 | 52.56 |
| *FSHB* | rs10429910 | A | C | 0.1407 | 0.09 | 0.007 | 3.9811E-34 | 165.31 |
| *FSHB* | rs9362387 | C | T | 0.2612 | -0.049 | 0.006 | 5.0119E-17 | 66.69 |
| *FSHB* | rs11031006 | G | A | 0.143 | -0.147 | 0.007 | 6.3096E-91 | 441.00 |
| *FST* | rs1260326 | T | C | 0.6086 | -0.11 | 0.008 | 7.9433E-48 | 189.06 |
| *FST* | rs31226 | T | C | 0.6025 | -0.142 | 0.008 | 1.9953E-77 | 315.06 |
| *FST* | rs139974673 | T | C | 0.0244 | 0.178 | 0.024 | 7.9433E-14 | 55.01 |
| *FSTL1* | rs1260326 | T | C | 0.6084 | 0.066 | 0.008 | 1.9953E-17 | 68.06 |
| *FSTL1* | rs1147707 | C | T | 0.3948 | -0.149 | 0.008 | 7.9433E-80 | 346.89 |
| *FSTL1* | rs7042335 | C | T | 0.3286 | -0.059 | 0.008 | 2.5119E-13 | 54.39 |
| *FSTL1* | rs2519093 | C | T | 0.1842 | -0.076 | 0.01 | 1.2589E-14 | 57.76 |
| *FSTL1* | rs11075921 | C | T | 0.1597 | 0.082 | 0.01 | 2.5119E-15 | 67.24 |
| *FSTL1* | rs200489612 | G | A | 0.0053 | 0.476 | 0.057 | 5.0119E-17 | 69.74 |
| *FSTL1* | rs10418205 | A | G | 0.3733 | -0.057 | 0.008 | 6.3096E-13 | 50.77 |
| *FSTL3* | rs77924615 | G | A | 0.1967 | -0.077 | 0.009 | 3.9811E-17 | 73.20 |
| *FSTL3* | rs916904 | A | G | 0.6335 | 0.087 | 0.008 | 1.9953E-30 | 118.27 |
| *FTCD* | rs7041363 | C | G | 0.4901 | -0.065 | 0.007 | 2.5119E-18 | 86.22 |
| *FTCD* | rs10883451 | T | C | 0.5015 | -0.052 | 0.007 | 1.9953E-12 | 55.18 |
| *FTCD* | rs58542926 | C | T | 0.0754 | 0.111 | 0.014 | 1.5849E-15 | 62.86 |
| *FTCD* | rs550999731 | C | G | 0.0035 | -0.948 | 0.072 | 1.9953E-39 | 173.36 |
| *FTCD* | rs3747207 | G | A | 0.2156 | 0.111 | 0.009 | 1.2589E-35 | 152.11 |
| *FUCA1* | rs13551 | T | C | 0.339 | -0.382 | 0.005 | 1E-200 | 5836.96 |
| *FUCA1* | rs11155297 | G | T | 0.264 | -1.3 | 0.008 | 1E-200 | 26406.25 |
| *FUCA1* | rs4764823 | G | A | 0.2883 | -0.091 | 0.005 | 1.2589E-90 | 331.24 |
| *FUCA1* | rs148815576 | T | G | 0.0019 | 0.391 | 0.05 | 5.0119E-15 | 61.15 |
| *FUCA1* | rs339969 | C | A | 0.617 | 0.028 | 0.004 | 1E-11 | 49.00 |
| *FUOM* | rs7041363 | C | G | 0.4899 | -0.068 | 0.007 | 3.9811E-20 | 94.37 |
| *FUOM* | rs10883451 | T | C | 0.5018 | -0.06 | 0.007 | 1.9953E-16 | 73.47 |
| *FUOM* | rs11816588 | T | C | 0.0326 | 0.503 | 0.021 | 1.995E-130 | 573.72 |
| *FUOM* | rs4753124 | C | G | 0.1072 | -0.082 | 0.012 | 3.9811E-12 | 46.69 |
| *FUOM* | rs28929474 | C | T | 0.0207 | 0.19 | 0.026 | 1.2589E-13 | 53.40 |
| *FUOM* | rs200210321 | A | AG | 0.0741 | 0.109 | 0.014 | 5.0119E-15 | 60.62 |
| *FUOM* | rs3747207 | G | A | 0.2151 | 0.111 | 0.009 | 5.0119E-36 | 152.11 |
| *FURIN* | rs1260326 | T | C | 0.6087 | -0.102 | 0.008 | 1.5849E-41 | 162.56 |
| *FURIN* | rs34430945 | T | C | 0.129 | -0.08 | 0.011 | 3.1623E-13 | 52.89 |
| *FURIN* | rs2071410 | C | G | 0.3258 | 0.194 | 0.008 | 3.162E-135 | 588.06 |
| *FUS* | rs10418046 | T | G | 0.2121 | 0.103 | 0.009 | 1.9953E-28 | 130.98 |
| *FUT1* | rs782134971 | GAAACTGC | G | 0.2546 | 0.31 | 0.009 | 1E-200 | 1186.42 |
| *FUT3_FUT5* | rs1800759 | T | G | 0.6134 | -0.061 | 0.006 | 7.9433E-21 | 103.36 |

| | | | | | | | | |
|---|---|---|---|---|---|---|---|---|
| *FUT3_FUT5* | rs2106854 | C | T | 0.1854 | -0.056 | 0.008 | 1E-11 | 49.00 |
| *FUT3_FUT5* | rs34434863 | T | G | 0.3086 | 0.053 | 0.007 | 7.9433E-14 | 57.33 |
| *FUT3_FUT5* | rs28601761 | C | G | 0.4192 | 0.071 | 0.006 | 6.3096E-28 | 140.03 |
| *FUT3_FUT5* | rs62542743 | C | A | 0.0534 | 0.101 | 0.014 | 2.5119E-12 | 52.05 |
| *FUT3_FUT5* | rs140107293 | A | G | 0.1556 | 0.061 | 0.009 | 2.5119E-12 | 45.94 |
| *FUT3_FUT5* | rs56278466 | T | G | 0.6611 | 0.074 | 0.007 | 3.1623E-28 | 111.76 |
| *FUT3_FUT5* | rs9738365 | C | A | 0.2646 | 0.089 | 0.007 | 1.9953E-34 | 161.65 |
| *FUT3_FUT5* | rs2393791 | C | T | 0.6237 | 0.138 | 0.007 | 1E-96 | 388.65 |
| *FUT3_FUT5* | rs178765 | G | A | 0.2831 | 0.068 | 0.007 | 3.1623E-22 | 94.37 |
| *FUT3_FUT5* | rs3760775 | G | T | 0.0603 | -1.068 | 0.015 | 1E-200 | 5069.44 |
| *FUT3_FUT5* | rs58542926 | C | T | 0.0754 | 0.114 | 0.014 | 6.3096E-17 | 66.31 |
| *FUT3_FUT5* | rs492602 | A | G | 0.5096 | -0.081 | 0.007 | 7.9433E-29 | 133.90 |
| *FUT3_FUT5* | rs6073431 | C | T | 0.5319 | 0.047 | 0.006 | 3.9811E-13 | 61.36 |
| *FUT8* | rs7618405 | C | A | 0.2053 | -0.055 | 0.008 | 7.9433E-12 | 47.27 |
| *FUT8* | rs1354034 | T | C | 0.6041 | -0.065 | 0.007 | 1.5849E-22 | 86.22 |
| *FUT8* | rs3792386 | G | A | 0.5871 | -0.047 | 0.007 | 1.9953E-12 | 45.08 |
| *FUT8* | rs10016018 | A | T | 0.3974 | -0.053 | 0.007 | 1.9953E-15 | 57.33 |
| *FUT8* | rs10075801 | A | G | 0.4887 | -0.049 | 0.007 | 1.5849E-13 | 49.00 |
| *FUT8* | rs138614581 | A | AT | 0.0803 | -0.083 | 0.012 | 3.1623E-12 | 47.84 |
| *FUT8* | rs6961069 | C | T | 0.4022 | 0.052 | 0.007 | 5.0119E-15 | 55.18 |
| *FUT8* | rs6993770 | A | T | 0.2865 | -0.147 | 0.007 | 2.5119E-93 | 441.00 |
| *FUT8* | rs11591571 | G | A | 0.3409 | 0.055 | 0.007 | 1E-15 | 61.73 |
| *FUT8* | rs10774625 | A | G | 0.5038 | -0.045 | 0.006 | 3.9811E-12 | 56.25 |
| *FUT8* | rs2127870 | G | C | 0.7952 | -0.915 | 0.009 | 1E-200 | 10336.11 |
| *FUT8* | rs12445050 | C | T | 0.1381 | 0.089 | 0.009 | 3.9811E-21 | 97.79 |
| *FUT8* | rs892090 | T | G | 0.8341 | 0.125 | 0.009 | 1.9953E-46 | 192.90 |
| *FUT8* | rs6112319 | G | A | 0.3529 | 0.054 | 0.007 | 1.5849E-15 | 59.51 |
| *FXN* | rs1354034 | T | C | 0.6041 | 0.055 | 0.008 | 5.0119E-13 | 47.27 |
| *FXN* | rs6993770 | A | T | 0.2863 | -0.064 | 0.008 | 1E-14 | 64.00 |
| *FXN* | rs112450466 | T | TAAAATA | 0.3791 | 0.079 | 0.008 | 6.3096E-24 | 97.52 |
| *FXN* | rs10733789 | T | C | 0.3131 | 0.06 | 0.008 | 3.1623E-13 | 56.25 |
| *FXN* | rs60822569 | T | TTTCTC | 0.5546 | 0.055 | 0.008 | 3.1623E-13 | 47.27 |
| *FXYD5* | rs1354034 | T | C | 0.6035 | 0.078 | 0.008 | 1E-24 | 95.06 |
| *FXYD5* | rs7896518 | A | G | 0.4238 | 0.055 | 0.008 | 1E-12 | 47.27 |
| *FXYD5* | rs756845 | A | G | 0.7799 | -0.38 | 0.009 | 1E-200 | 1782.72 |
| *FYB1* | rs1354034 | T | C | 0.604 | 0.074 | 0.008 | 5.0119E-22 | 85.56 |
| *FYB1* | rs112838960 | T | TAATA | 0.5158 | 0.065 | 0.008 | 1E-17 | 66.02 |
| *FYB1* | rs8100773 | T | C | 0.801 | 0.069 | 0.01 | 6.3096E-13 | 47.61 |
| *GABARAP* | rs11967437 | C | T | 0.0072 | 0.793 | 0.046 | 2.5119E-67 | 297.19 |
| *GABARAP* | rs773111 | C | A | 0.3358 | -0.065 | 0.008 | 6.3096E-16 | 66.02 |
| *GADD45GIP1* | rs1354034 | T | C | 0.6045 | 0.063 | 0.008 | 3.9811E-16 | 62.02 |
| *GADD45GIP1* | rs10058074 | G | A | 0.4752 | -0.056 | 0.008 | 2.5119E-13 | 49.00 |
| *GADD45GIP1* | rs342299 | C | T | 0.4549 | -0.055 | 0.008 | 7.9433E-13 | 47.27 |
| *GADD45GIP1* | rs6993770 | A | T | 0.2866 | -0.068 | 0.008 | 7.9433E-16 | 72.25 |
| *GADD45GIP1* | rs11085147 | C | T | 0.0991 | 0.15 | 0.013 | 7.9433E-30 | 133.14 |
| *GAL* | rs320366 | C | T | 0.7491 | -0.059 | 0.008 | 6.3096E-13 | 54.39 |
| *GAL* | rs760077 | A | T | 0.6049 | -0.08 | 0.007 | 6.3096E-29 | 130.61 |
| *GAL* | rs28631481 | C | T | 0.3834 | 0.053 | 0.007 | 1.5849E-13 | 57.33 |
| *GAL* | rs11680123 | A | G | 0.5934 | 0.196 | 0.007 | 2.512E-165 | 784.00 |
| *GAL* | rs121908120 | T | A | 0.0275 | -0.243 | 0.022 | 5.0119E-29 | 122.00 |
| *GAL* | rs1530018 | A | G | 0.233 | -0.078 | 0.008 | 7.9433E-21 | 95.06 |
| *GAL* | rs3773364 | A | G | 0.1605 | 0.196 | 0.01 | 2.5119E-94 | 384.16 |
| *GAL* | rs34826710 | C | T | 0.1055 | 0.146 | 0.011 | 1.5849E-37 | 176.17 |
| *GAL* | rs560623757 | C | CTT | 0.1041 | 0.12 | 0.011 | 7.9433E-26 | 119.01 |
| *GAL* | rs565427 | G | A | 0.3579 | 0.063 | 0.007 | 3.9811E-18 | 81.00 |
| *GAL* | rs76634175 | T | A | 0.0778 | 0.152 | 0.013 | 1.9953E-31 | 136.71 |
| *GAL* | rs2540119 | T | C | 0.759 | -0.057 | 0.008 | 2.5119E-12 | 50.77 |
| *GAL* | rs9601479 | G | A | 0.5089 | 0.051 | 0.007 | 3.1623E-13 | 53.08 |
| *GAL* | rs36034702 | C | T | 0.1688 | -0.088 | 0.009 | 7.9433E-21 | 95.60 |
| *GAL* | rs720130 | G | T | 0.4077 | -0.055 | 0.007 | 5.0119E-15 | 61.73 |
| *GAL* | rs56228609 | C | T | 0.3158 | 0.075 | 0.007 | 1.9953E-23 | 114.80 |
| *GAL* | rs77542162 | A | G | 0.0229 | 0.738 | 0.024 | 1E-200 | 945.56 |
| *GAL* | rs55949690 | G | A | 0.5233 | 0.051 | 0.007 | 6.3096E-13 | 53.08 |
| *GAL* | rs6028484 | T | C | 0.5443 | 0.055 | 0.007 | 1.9953E-15 | 61.73 |
| *GAL* | rs6024978 | A | C | 0.2259 | -0.066 | 0.008 | 3.9811E-15 | 68.06 |
| *GALNT10* | rs4672497 | C | G | 0.2218 | 0.07 | 0.008 | 1.9953E-16 | 76.56 |
| *GALNT10* | rs890793 | C | T | 0.382 | 0.384 | 0.008 | 1E-200 | 2304.00 |
| *GALNT10* | rs635634 | C | T | 0.1844 | 0.26 | 0.009 | 5.012E-176 | 834.57 |

| | | | | | | | | |
|---|---|---|---|---|---|---|---|---|
| *GALNT10* | rs56278466 | T | G | 0.6611 | 0.057 | 0.007 | 2.5119E-14 | 66.31 |
| *GALNT10* | rs2695294 | G | A | 0.5832 | 0.06 | 0.007 | 3.9811E-17 | 73.47 |
| *GALNT10* | rs3184504 | T | C | 0.5168 | -0.057 | 0.007 | 6.3096E-16 | 66.31 |
| *GALNT10* | rs145078947 | G | T | 0.0026 | 0.57 | 0.069 | 1.5849E-16 | 68.24 |
| *GALNT10* | rs704 | G | A | 0.4733 | 0.086 | 0.007 | 1E-34 | 150.94 |
| *GALNT10* | rs601338 | G | A | 0.5088 | 0.052 | 0.007 | 1.9953E-13 | 55.18 |
| *GALNT2* | rs4846914 | G | A | 0.6061 | 0.293 | 0.007 | 1E-200 | 1752.02 |
| *GALNT2* | rs9273216 | C | A | 0.5546 | -0.059 | 0.008 | 7.9433E-15 | 54.39 |
| *GALNT2* | rs9987289 | A | G | 0.9078 | -0.085 | 0.012 | 1.5849E-12 | 50.17 |
| *GALNT2* | rs12359178 | G | A | 0.1445 | 0.072 | 0.01 | 3.9811E-13 | 51.84 |
| *GALNT2* | rs8181612 | T | A | 0.7058 | -0.128 | 0.008 | 3.9811E-61 | 256.00 |
| *GALNT2* | rs704 | G | A | 0.4729 | 0.083 | 0.007 | 1E-32 | 140.59 |
| *GALNT2* | rs12975366 | T | C | 0.3954 | -0.145 | 0.007 | 3.1623E-91 | 429.08 |
| *GALNT3* | rs2116546 | T | C | 0.2979 | -0.389 | 0.008 | 1E-200 | 2364.39 |
| *GALNT3* | rs9310400 | C | T | 0.3282 | 0.055 | 0.008 | 7.9433E-13 | 47.27 |
| *GALNT3* | rs13107325 | C | T | 0.0747 | -0.121 | 0.014 | 1.2589E-18 | 74.70 |
| *GALNT3* | rs9391787 | T | C | 0.5935 | 0.062 | 0.008 | 1E-14 | 60.06 |
| *GALNT3* | rs7791957 | T | C | 0.4313 | 0.096 | 0.007 | 3.9811E-40 | 188.08 |
| *GALNT3* | rs782134971 | GAAACTGC( | G | 0.2542 | 0.31 | 0.008 | 1E-200 | 1501.56 |
| *GALNT3* | rs3184504 | T | C | 0.5177 | -0.08 | 0.007 | 6.3096E-29 | 130.61 |
| *GALNT3* | rs145078947 | G | T | 0.0026 | 0.509 | 0.071 | 6.3096E-13 | 51.39 |
| *GALNT3* | rs146261845 | C | T | 0.0069 | 0.334 | 0.047 | 7.9433E-13 | 50.50 |
| *GALNT3* | rs492602 | A | G | 0.5092 | -0.183 | 0.007 | 1E-139 | 683.45 |
| *GALNT5* | rs1427407 | T | G | 0.8479 | -0.12 | 0.01 | 5.0119E-31 | 144.00 |
| *GALNT5* | rs714649 | G | T | 0.0182 | -0.941 | 0.028 | 1E-200 | 1129.44 |
| *GALNT5* | rs3864106 | G | C | 0.5843 | -0.117 | 0.007 | 2.5119E-56 | 279.37 |
| *GALNT5* | rs552268504 | G | GA | 0.3884 | -0.056 | 0.008 | 2.5119E-13 | 49.00 |
| *GALNT5* | rs6592965 | G | A | 0.454 | -0.089 | 0.007 | 1.2589E-34 | 161.65 |
| *GALNT5* | rs507666 | G | A | 0.1846 | -0.155 | 0.009 | 7.9433E-61 | 296.60 |
| *GALNT5* | rs10876169 | T | A | 0.4222 | 0.06 | 0.007 | 1.9953E-16 | 73.47 |
| *GALNT5* | rs74035509 | C | T | 0.0781 | -0.142 | 0.014 | 1.5849E-25 | 102.88 |
| *GALNT5* | rs186021206 | G | A | 0.0064 | 0.582 | 0.05 | 1.9953E-31 | 135.49 |
| *GALNT7* | rs13107325 | C | T | 0.0748 | -0.14 | 0.014 | 1E-22 | 100.00 |
| *GALNT7* | rs144873913 | C | A | 0.0023 | -0.855 | 0.077 | 7.9433E-29 | 123.30 |
| *GALNT7* | rs11751024 | C | A | 0.3982 | 0.072 | 0.008 | 7.9433E-21 | 81.00 |
| *GALNT7* | rs2691582 | C | T | 0.5058 | -0.057 | 0.008 | 1.9953E-14 | 50.77 |
| *GALNT7* | rs117905808 | G | A | 0.02 | 0.727 | 0.027 | 5.012E-157 | 725.01 |
| *GALNT7* | rs139130389 | C | CTA | 0.0726 | 0.116 | 0.014 | 5.0119E-16 | 68.65 |
| *GALNT7* | rs74972303 | C | G | 0.0849 | 0.166 | 0.013 | 3.1623E-35 | 163.05 |
| *GALNT7* | rs534297892 | CA | C | 0.5032 | 0.054 | 0.008 | 1.5849E-12 | 45.56 |
| *GALNT7* | rs9534300 | A | T | 0.7081 | -0.12 | 0.008 | 6.3096E-49 | 225.00 |
| *GALNT7* | rs12975366 | T | C | 0.3953 | -0.217 | 0.008 | 5.012E-175 | 735.77 |
| *GAPDH* | rs1354034 | T | C | 0.6042 | 0.061 | 0.008 | 5.0119E-15 | 58.14 |
| *GART* | rs35923643 | A | G | 0.1965 | -0.071 | 0.01 | 1.5849E-13 | 50.41 |
| *GAS2* | rs2279240 | T | C | 0.7069 | 0.114 | 0.008 | 6.3096E-42 | 203.06 |
| *GAS6* | rs143634417 | C | CTTATT | 0.3391 | 0.051 | 0.007 | 1.2589E-11 | 53.08 |
| *GAS6* | rs56073752 | G | A | 0.5128 | 0.049 | 0.007 | 6.3096E-12 | 49.00 |
| *GAS6* | rs1260326 | T | C | 0.6085 | 0.057 | 0.007 | 3.9811E-15 | 66.31 |
| *GAS6* | rs62165726 | C | A | 0.037 | -0.129 | 0.019 | 7.9433E-12 | 46.10 |
| *GAS6* | rs1983009 | C | T | 0.1012 | 0.099 | 0.012 | 7.9433E-16 | 68.06 |
| *GAS6* | rs13108218 | A | G | 0.6189 | 0.05 | 0.007 | 1E-11 | 51.02 |
| *GAS6* | rs6840282 | C | T | 0.6143 | -0.051 | 0.007 | 5.0119E-12 | 53.08 |
| *GAS6* | rs112840324 | A | G | 0.7089 | 0.076 | 0.009 | 1E-17 | 71.31 |
| *GAS6* | rs17138358 | G | C | 0.3938 | -0.054 | 0.007 | 1.2589E-13 | 59.51 |
| *GAS6* | rs17145750 | C | T | 0.1595 | 0.074 | 0.01 | 3.1623E-14 | 54.76 |
| *GAS6* | rs12667978 | C | T | 0.5706 | 0.06 | 0.007 | 7.9433E-17 | 73.47 |
| *GAS6* | rs4841133 | A | G | 0.908 | -0.131 | 0.012 | 1.2589E-26 | 119.17 |
| *GAS6* | rs4876611 | A | G | 0.718 | 0.074 | 0.008 | 1.5849E-20 | 85.56 |
| *GAS6* | rs11230313 | A | C | 0.3281 | -0.056 | 0.008 | 5.0119E-13 | 49.00 |
| *GAS6* | rs6602909 | T | C | 0.3242 | -0.338 | 0.008 | 1E-200 | 1785.06 |
| *GAS6* | rs150844304 | A | C | 0.024 | -0.157 | 0.023 | 1E-11 | 46.60 |
| *GAS6* | rs186021206 | G | A | 0.0064 | 1.078 | 0.049 | 5.012E-107 | 484.00 |
| *GAS6* | rs1688030 | T | C | 0.934 | -0.106 | 0.014 | 1.5849E-13 | 57.33 |
| *GAS6* | rs74816457 | G | C | 0.4028 | -0.221 | 0.007 | 1E-200 | 996.76 |
| *GAS6* | rs3747207 | G | A | 0.2155 | 0.08 | 0.009 | 1.2589E-20 | 79.01 |
| *GASK1A* | rs493412 | C | T | 0.6295 | 0.73 | 0.008 | 1E-200 | 8326.56 |
| *GAST* | rs144268065 | A | ATTCTCTTA | 0.2862 | -0.068 | 0.008 | 6.3096E-17 | 72.25 |
| *GAST* | rs72704793 | G | A | 0.3296 | 0.071 | 0.008 | 1E-19 | 78.77 |

| | | | | | | | | |
|---|---|---|---|---|---|---|---|---|
| *GAST* | rs56092644 | G | T | 0.2446 | 0.13 | 0.009 | 2.5119E-52 | 208.64 |
| *GAST* | rs2395184 | G | A | 0.1669 | -0.125 | 0.01 | 2.5119E-37 | 156.25 |
| *GAST* | rs2472632 | C | A | 0.3599 | 0.073 | 0.008 | 1.5849E-21 | 83.27 |
| *GAST* | rs749161312 | CCCCTCCTC | C | 0.2154 | 0.086 | 0.009 | 1.5849E-21 | 91.31 |
| *GATD3* | rs11242109 | G | T | 0.4812 | -0.052 | 0.008 | 1.2589E-11 | 42.25 |
| *GATD3* | rs342298 | C | T | 0.4548 | -0.059 | 0.008 | 1.9953E-14 | 54.39 |
| *GATD3* | rs6993770 | A | T | 0.2866 | -0.065 | 0.008 | 1E-14 | 66.02 |
| *GATD3* | rs6580980 | A | G | 0.5635 | 0.052 | 0.008 | 1.5849E-11 | 42.25 |
| *GATD3* | rs2187313 | T | C | 0.8177 | -0.124 | 0.01 | 3.9811E-36 | 153.76 |
| *GBP1* | rs148526074 | G | T | 0.0276 | -1.314 | 0.023 | 1E-200 | 3263.89 |
| *GBP1* | rs3184504 | T | C | 0.5176 | -0.113 | 0.007 | 3.9811E-56 | 260.59 |
| *GBP2* | rs7531439 | A | C | 0.7905 | 0.124 | 0.009 | 1.5849E-39 | 189.83 |
| *GBP4* | rs608339 | A | T | 0.5077 | -0.805 | 0.008 | 1E-200 | 10125.39 |
| *GC* | rs1260326 | T | C | 0.6084 | -0.033 | 0.005 | 1E-11 | 43.56 |
| *GC* | rs1352846 | A | G | 0.2923 | 1.202 | 0.008 | 1E-200 | 22575.06 |
| *GC* | rs28601761 | C | G | 0.4192 | -0.042 | 0.005 | 1.2589E-17 | 70.56 |
| *GC* | rs7080386 | C | A | 0.4118 | -0.033 | 0.005 | 5.0119E-12 | 43.56 |
| *GCC1* | rs1354034 | T | C | 0.6043 | 0.071 | 0.008 | 6.3096E-21 | 78.77 |
| *GCC1* | rs342298 | C | T | 0.4553 | -0.057 | 0.007 | 1.9953E-14 | 66.31 |
| *GCC1* | rs6993770 | A | T | 0.2865 | -0.064 | 0.008 | 1E-14 | 64.00 |
| *GCC1* | rs11502185 | T | C | 0.2597 | 0.069 | 0.01 | 3.9811E-13 | 47.61 |
| *GCG* | rs17145750 | C | T | 0.1602 | -0.084 | 0.01 | 3.9811E-16 | 70.56 |
| *GCG* | rs39281 | A | G | 0.527 | 0.055 | 0.008 | 3.9811E-13 | 47.27 |
| *GCG* | rs964184 | G | C | 0.8667 | -0.082 | 0.011 | 3.1623E-13 | 55.57 |
| *GCG* | rs72654473 | C | A | 0.1089 | 0.089 | 0.012 | 3.1623E-13 | 55.01 |
| *GCHFR* | rs1427407 | T | G | 0.8479 | -0.112 | 0.01 | 3.9811E-28 | 125.44 |
| *GCHFR* | rs9399137 | T | C | 0.2577 | 0.071 | 0.008 | 7.9433E-17 | 78.77 |
| *GCHFR* | rs2289218 | T | A | 0.4578 | 0.111 | 0.007 | 5.0119E-51 | 251.45 |
| *GCLM* | rs12563495 | G | A | 0.5087 | -0.06 | 0.008 | 3.9811E-15 | 56.25 |
| *GCNT1* | rs2009581 | G | A | 0.2732 | -0.061 | 0.008 | 3.9811E-15 | 58.14 |
| *GCNT1* | rs35829610 | C | CTG | 0.3381 | -0.065 | 0.007 | 6.3096E-19 | 86.22 |
| *GCNT1* | rs2282683 | A | G | 0.21 | -0.496 | 0.009 | 1E-200 | 3037.23 |
| *GCNT1* | rs505922 | T | C | 0.3169 | 0.073 | 0.008 | 5.0119E-20 | 83.27 |
| *GCNT1* | rs117163080 | T | C | 0.1378 | 0.076 | 0.01 | 2.5119E-14 | 57.76 |
| *GCNT1* | rs3184504 | T | C | 0.5168 | -0.129 | 0.007 | 2.5119E-77 | 339.61 |
| *GCNT1* | rs492602 | A | G | 0.5088 | -0.072 | 0.007 | 1E-24 | 105.80 |
| *GDF15* | rs1054221 | T | C | 0.1459 | 0.392 | 0.009 | 1E-200 | 1897.09 |
| *GDF2* | rs34008398 | G | A | 0.0128 | -0.225 | 0.031 | 2.5119E-13 | 52.68 |
| *GDF2* | rs68066031 | T | C | 0.2298 | 0.167 | 0.008 | 2.5119E-90 | 435.77 |
| *GDF2* | rs1912826 | G | A | 0.5128 | 0.092 | 0.007 | 1.2589E-40 | 172.73 |
| *GDF2* | rs78408340 | C | G | 0.0097 | -0.309 | 0.035 | 1.2589E-18 | 77.94 |
| *GDF2* | rs10820966 | A | T | 0.3322 | -0.181 | 0.007 | 1.995E-131 | 668.59 |
| *GDF2* | rs9414801 | G | A | 0.5335 | -0.062 | 0.007 | 3.9811E-19 | 78.45 |
| *GDF2* | rs67852477 | C | G | 0.0103 | 0.328 | 0.034 | 1.2589E-21 | 93.07 |
| *GDF2* | rs3184504 | T | C | 0.5174 | 0.047 | 0.007 | 7.9433E-12 | 45.08 |
| *GDF2* | rs1956713 | G | A | 0.8518 | -0.152 | 0.01 | 3.1623E-54 | 231.04 |
| *GDF2* | rs4702 | G | A | 0.5577 | 0.056 | 0.007 | 1E-15 | 64.00 |
| *GDF2* | rs12441560 | G | A | 0.2749 | 0.062 | 0.008 | 1.2589E-15 | 60.06 |
| *GDF2* | rs77303550 | C | T | 0.1967 | -0.073 | 0.009 | 7.9433E-17 | 65.79 |
| *GDF2* | rs186021206 | G | A | 0.0064 | 0.479 | 0.047 | 3.9811E-24 | 103.87 |
| *GDNF* | rs62360373 | G | A | 0.0952 | -0.393 | 0.013 | 1E-200 | 913.90 |
| *GDNF* | rs4808305 | T | C | 0.3231 | 0.066 | 0.008 | 6.3096E-17 | 68.06 |
| *GET3* | rs58534292 | T | C | 0.6503 | 0.064 | 0.008 | 1.5849E-15 | 64.00 |
| *GET3* | rs8009574 | T | G | 0.628 | 0.072 | 0.008 | 2.5119E-19 | 81.00 |
| *GFAP* | rs150394890 | G | T | 0.0086 | -0.469 | 0.038 | 6.3096E-35 | 152.33 |
| *GFAP* | rs6713456 | T | A | 0.4349 | 0.051 | 0.007 | 3.9811E-13 | 53.08 |
| *GFAP* | rs147795230 | T | C | 0.0012 | 1.305 | 0.111 | 7.9433E-32 | 138.22 |
| *GFAP* | rs5011436 | A | C | 0.4151 | -0.059 | 0.007 | 3.9811E-17 | 71.04 |
| *GFAP* | rs3132880 | C | T | 0.4598 | 0.058 | 0.007 | 2.5119E-16 | 68.65 |
| *GFAP* | rs7114754 | T | C | 0.0781 | -0.145 | 0.013 | 3.1623E-29 | 124.41 |
| *GFAP* | rs59291670 | C | T | 0.0215 | 0.278 | 0.024 | 5.0119E-31 | 134.17 |
| *GFAP* | rs429358 | T | C | 0.1558 | 0.103 | 0.01 | 7.9433E-27 | 106.09 |
| *GFER* | rs61516948 | C | T | 0.126 | -0.104 | 0.011 | 1.2589E-19 | 89.39 |
| *GFRA1* | rs10982156 | T | A | 0.0694 | 0.185 | 0.014 | 1.9953E-37 | 174.62 |
| *GFRA1* | rs11197603 | T | C | 0.148 | -0.303 | 0.01 | 1.995E-197 | 918.09 |
| *GFRA1* | rs6041 | G | A | 0.1002 | -0.216 | 0.012 | 1.5849E-75 | 324.00 |
| *GFRA1* | rs12934455 | C | T | 0.1663 | -0.07 | 0.01 | 1.9953E-13 | 49.00 |
| *GFRA2* | rs13107325 | C | T | 0.0748 | 0.108 | 0.014 | 7.9433E-15 | 59.51 |

| | | | | | | | | |
|---|---|---|---|---|---|---|---|---|
| *GFRA2* | rs28478097 | C | G | 0.616 | -0.055 | 0.007 | 1.9953E-13 | 61.73 |
| *GFRA2* | rs9275580 | A | G | 0.2281 | -0.09 | 0.009 | 1.5849E-25 | 100.00 |
| *GFRA2* | rs1480999 | G | A | 0.5621 | 0.309 | 0.008 | 1E-200 | 1491.89 |
| *GFRA2* | rs603424 | G | A | 0.1663 | -0.067 | 0.01 | 6.3096E-12 | 44.89 |
| *GFRA2* | rs186021206 | G | A | 0.0063 | 0.834 | 0.05 | 1.9953E-61 | 278.22 |
| *GFRA3* | rs12027119 | A | G | 0.6672 | -0.053 | 0.008 | 5.0119E-12 | 43.89 |
| *GFRA3* | rs35383942 | C | T | 0.0617 | 0.111 | 0.015 | 1.2589E-13 | 54.76 |
| *GFRA3* | rs12710696 | T | C | 0.6396 | -0.061 | 0.007 | 3.1623E-16 | 75.94 |
| *GFRA3* | rs73158489 | C | T | 0.1602 | 0.075 | 0.01 | 2.5119E-14 | 56.25 |
| *GFRA3* | rs13107325 | C | T | 0.0746 | 0.14 | 0.014 | 2.5119E-24 | 100.00 |
| *GFRA3* | rs3756599 | G | C | 0.3253 | -0.064 | 0.008 | 1E-16 | 64.00 |
| *GFRA3* | rs56259296 | T | A | 0.3655 | 0.092 | 0.008 | 1.9953E-29 | 132.25 |
| *GFRA3* | rs62405538 | A | T | 0.2124 | -0.063 | 0.009 | 7.9433E-13 | 49.00 |
| *GFRA3* | rs10956755 | A | G | 0.2998 | -0.054 | 0.008 | 7.9433E-12 | 45.56 |
| *GFRA3* | rs56278466 | T | G | 0.6614 | 0.081 | 0.008 | 1.2589E-26 | 102.52 |
| *GFRA3* | rs10887793 | G | T | 0.4491 | 0.065 | 0.007 | 3.9811E-19 | 86.22 |
| *GFRA3* | rs10742826 | C | T | 0.6835 | 0.06 | 0.008 | 7.9433E-15 | 56.25 |
| *GFRA3* | rs112771035 | C | G | 0.0713 | -0.097 | 0.014 | 3.9811E-12 | 48.01 |
| *GFRA3* | rs11830324 | T | A | 0.1696 | -0.135 | 0.01 | 3.9811E-44 | 182.25 |
| *GFRA3* | rs1161106 | C | A | 0.1231 | -0.075 | 0.011 | 7.9433E-12 | 46.49 |
| *GFRA3* | rs1618875 | C | T | 0.4928 | 0.208 | 0.007 | 3.981E-179 | 882.94 |
| *GFRA3* | rs12901860 | G | A | 0.1658 | 0.071 | 0.01 | 1.5849E-13 | 50.41 |
| *GFRA3* | rs186021206 | G | A | 0.0064 | 0.536 | 0.049 | 1.9953E-27 | 119.66 |
| *GFRA3* | rs11655511 | C | T | 0.7665 | 0.104 | 0.009 | 2.5119E-33 | 133.53 |
| *GFRA3* | rs3744052 | C | T | 0.2054 | 0.074 | 0.009 | 1.2589E-16 | 67.60 |
| *GFRAL* | rs11695471 | T | A | 0.3334 | -0.052 | 0.007 | 6.3096E-13 | 55.18 |
| *GFRAL* | rs893832 | A | G | 0.452 | -0.072 | 0.007 | 2.5119E-26 | 105.80 |
| *GFRAL* | rs4710973 | T | C | 0.6276 | -0.068 | 0.008 | 2.5119E-19 | 72.25 |
| *GFRAL* | rs72988065 | A | G | 0.1536 | 0.37 | 0.01 | 1E-200 | 1369.00 |
| *GFRAL* | rs4263799 | C | T | 0.4801 | 0.058 | 0.007 | 6.3096E-18 | 68.65 |
| *GFRAL* | rs8176747 | C | G | 0.0617 | 0.506 | 0.014 | 1E-200 | 1306.31 |
| *GFRAL* | rs11603123 | G | A | 0.0334 | 0.439 | 0.019 | 7.943E-122 | 533.85 |
| *GFRAL* | rs7147346 | C | T | 0.4645 | -0.046 | 0.007 | 6.3096E-12 | 43.18 |
| *GFRAL* | rs186021206 | G | A | 0.0066 | 0.371 | 0.046 | 5.0119E-16 | 65.05 |
| *GFRAL* | rs17265513 | T | C | 0.1998 | 0.078 | 0.008 | 1.5849E-20 | 95.06 |
| *GGACT* | rs1354034 | T | C | 0.6045 | 0.055 | 0.007 | 7.9433E-14 | 61.73 |
| *GGACT* | rs6993770 | A | T | 0.2866 | -0.09 | 0.008 | 7.9433E-30 | 126.56 |
| *GGACT* | rs7896518 | A | G | 0.424 | 0.055 | 0.007 | 5.0119E-14 | 61.73 |
| *GGACT* | rs4772304 | G | A | 0.5128 | -0.445 | 0.008 | 1E-200 | 3094.14 |
| *GGCT* | rs1354034 | T | C | 0.6045 | 0.069 | 0.008 | 2.5119E-19 | 74.39 |
| *GGCT* | rs574628490 | G | GT | 0.1255 | 0.083 | 0.012 | 1.2589E-11 | 47.84 |
| *GGCT* | rs4259 | G | A | 0.7268 | -0.119 | 0.008 | 3.1623E-45 | 221.27 |
| *GGH* | rs114165349 | G | C | 0.0231 | 0.203 | 0.024 | 7.9433E-18 | 71.54 |
| *GGH* | rs451637 | C | T | 0.3096 | -0.062 | 0.008 | 3.1623E-15 | 60.06 |
| *GGH* | rs1513275 | T | C | 0.1917 | 0.07 | 0.009 | 2.5119E-14 | 60.49 |
| *GGH* | rs74329104 | G | A | 0.0659 | 0.723 | 0.015 | 1E-200 | 2323.24 |
| *GGH* | rs28601761 | C | G | 0.4187 | 0.052 | 0.008 | 1E-11 | 42.25 |
| *GGH* | rs6488368 | A | G | 0.973 | -0.243 | 0.022 | 5.0119E-28 | 122.00 |
| *GGH* | rs10778152 | A | G | 0.2886 | -0.26 | 0.008 | 1E-200 | 1056.25 |
| *GGH* | rs117230571 | A | G | 0.0764 | 0.099 | 0.013 | 1.5849E-13 | 57.99 |
| *GGH* | rs139974673 | T | C | 0.0243 | 0.169 | 0.023 | 1.5849E-13 | 53.99 |
| *GGH* | rs768736886 | AT | A | 0.2977 | 0.068 | 0.008 | 1.9953E-18 | 72.25 |
| *GGH* | rs12970396 | G | A | 0.3013 | 0.056 | 0.008 | 6.3096E-13 | 49.00 |
| *GGH* | rs58542926 | C | T | 0.0753 | 0.102 | 0.013 | 3.1623E-14 | 61.56 |
| *GGH* | rs12975366 | T | C | 0.3956 | -0.097 | 0.007 | 3.1623E-41 | 192.02 |
| *GGT1* | rs924204 | A | G | 0.5894 | 0.064 | 0.007 | 1.5849E-20 | 83.59 |
| *GGT1* | rs4665972 | T | C | 0.6084 | -0.048 | 0.007 | 1E-11 | 47.02 |
| *GGT1* | rs4835265 | C | A | 0.1562 | 0.09 | 0.009 | 7.9433E-22 | 100.00 |
| *GGT1* | rs7970695 | G | A | 0.6244 | 0.132 | 0.007 | 6.3096E-78 | 355.59 |
| *GGT1* | rs550999044 | A | AT | 0.244 | 0.077 | 0.008 | 7.9433E-22 | 92.64 |
| *GGT1* | rs4998040 | G | A | 0.7883 | -0.061 | 0.008 | 3.1623E-13 | 58.14 |
| *GGT1* | rs28509371 | T | C | 0.3374 | 0.254 | 0.007 | 1E-200 | 1316.65 |
| *GGT5* | rs74612335 | T | C | 0.1334 | -0.104 | 0.011 | 1.2589E-21 | 89.39 |
| *GGT5* | rs186021206 | G | A | 0.0064 | 0.635 | 0.051 | 3.9811E-36 | 155.03 |
| *GGT5* | rs573431210 | T | G | 0.0267 | 0.157 | 0.023 | 7.9433E-12 | 46.60 |
| *GGT5* | rs5760275 | C | G | 0.2074 | 0.329 | 0.009 | 1E-200 | 1336.31 |
| *GHR* | rs1260326 | T | C | 0.6081 | -0.049 | 0.007 | 3.1623E-12 | 49.00 |
| *GHR* | rs4073476 | A | C | 0.2733 | -0.685 | 0.009 | 1E-200 | 5792.90 |

| | | | | | | | | |
|---|---|---|---|---|---|---|---|---|
| *GHR* | rs112316267 | A | T | 0.9185 | 0.111 | 0.015 | 5.0119E-14 | 54.76 |
| *GHR* | rs28601761 | C | G | 0.419 | -0.05 | 0.007 | 6.3096E-13 | 51.02 |
| *GHR* | rs174568 | C | T | 0.3444 | 0.049 | 0.007 | 1E-11 | 49.00 |
| *GHRL* | rs751198981 | TG | T | 0.195 | -0.077 | 0.009 | 1.5849E-16 | 73.20 |
| *GHRL* | rs34911341 | C | T | 0.006 | 1.419 | 0.047 | 1E-200 | 911.53 |
| *GHRL* | rs7438696 | G | T | 0.2815 | 0.07 | 0.008 | 3.9811E-18 | 76.56 |
| *GHRL* | rs2087038 | G | A | 0.7098 | 0.067 | 0.008 | 3.1623E-17 | 70.14 |
| *GHRL* | rs11745870 | C | T | 0.314 | -0.076 | 0.008 | 1.5849E-22 | 90.25 |
| *GHRL* | rs11786721 | T | C | 0.4104 | 0.071 | 0.007 | 5.0119E-22 | 102.88 |
| *GHRL* | rs4500149 | G | A | 0.4248 | 0.064 | 0.007 | 2.5119E-18 | 83.59 |
| *GHRL* | rs8176746 | G | T | 0.0613 | -0.158 | 0.015 | 3.1623E-26 | 110.95 |
| *GHRL* | rs558760274 | A | C | 0.7921 | 0.063 | 0.009 | 3.1623E-12 | 49.00 |
| *GHRL* | rs17138478 | C | A | 0.131 | 0.078 | 0.011 | 1.9953E-13 | 50.28 |
| *GHRL* | rs34954997 | C | CTTCG | 0.238 | 0.063 | 0.009 | 1E-13 | 49.00 |
| *GHRL* | rs6129534 | G | T | 0.2958 | 0.074 | 0.008 | 6.3096E-21 | 85.56 |
| *GIGYF2* | rs1354034 | T | C | 0.6043 | 0.072 | 0.008 | 1.2589E-21 | 81.00 |
| *GIGYF2* | rs5021727 | A | G | 0.4605 | 0.061 | 0.008 | 5.0119E-16 | 58.14 |
| *GIMAP7* | rs3735080 | C | T | 0.2314 | -0.607 | 0.009 | 1E-200 | 4548.75 |
| *GIMAP7* | rs56156922 | T | C | 0.3246 | 0.066 | 0.007 | 1.9953E-18 | 88.90 |
| *GIMAP7* | rs4632248 | G | T | 0.2094 | 0.059 | 0.009 | 6.3096E-12 | 42.98 |
| *GIP* | rs6879048 | A | T | 0.1664 | -0.138 | 0.01 | 1.9953E-45 | 190.44 |
| *GIP* | rs612169 | A | G | 0.3178 | -0.103 | 0.008 | 1E-39 | 165.77 |
| *GIP* | rs766253041 | C | CCTCCT | 0.1995 | 0.127 | 0.009 | 2.5119E-41 | 199.12 |
| *GIPC2* | rs7537908 | A | G | 0.2061 | -0.314 | 0.009 | 1E-200 | 1217.23 |
| *GIPC3* | rs6993770 | A | T | 0.2866 | -0.065 | 0.008 | 3.9811E-15 | 66.02 |
| *GIPC3* | rs34722692 | G | A | 0.1119 | -0.434 | 0.012 | 1E-200 | 1308.03 |
| *GIT1* | rs1354034 | T | C | 0.6039 | 0.061 | 0.008 | 1.5849E-15 | 58.14 |
| *GIT1* | rs342296 | G | A | 0.4564 | -0.051 | 0.007 | 1E-11 | 53.08 |
| *GIT1* | rs142511067 | C | G | 0.0217 | 0.245 | 0.027 | 3.1623E-20 | 82.34 |
| *GKN1* | rs71469854 | C | T | 0.1752 | 0.088 | 0.01 | 2.5119E-18 | 77.44 |
| *GKN1* | rs601338 | G | A | 0.5092 | -0.086 | 0.008 | 5.0119E-29 | 115.56 |
| *GKN1* | rs4816636 | T | G | 0.7489 | 0.064 | 0.009 | 3.9811E-13 | 50.57 |
| *GLA* | rs1260326 | T | C | 0.6081 | -0.068 | 0.008 | 3.9811E-19 | 72.25 |
| *GLA* | rs2559851 | C | A | 0.5868 | 0.057 | 0.008 | 1E-13 | 50.77 |
| *GLA* | rs11650885 | C | T | 0.4608 | -0.063 | 0.008 | 1.2589E-16 | 62.02 |
| *GLB1* | rs114165349 | G | C | 0.0232 | 0.177 | 0.024 | 1E-13 | 54.39 |
| *GLB1* | rs7637133 | C | A | 0.616 | 0.228 | 0.007 | 1E-200 | 1060.90 |
| *GLB1* | rs687339 | C | T | 0.7708 | 0.069 | 0.009 | 1.5849E-15 | 58.78 |
| *GLB1* | rs9457800 | C | T | 0.1351 | -0.071 | 0.01 | 1E-11 | 50.41 |
| *GLB1* | rs1293303 | G | C | 0.4456 | 0.074 | 0.007 | 6.3096E-25 | 111.76 |
| *GLB1* | rs10778152 | A | G | 0.2882 | -0.288 | 0.008 | 1E-200 | 1296.00 |
| *GLB1* | rs150641790 | T | TG | 0.0748 | 0.107 | 0.014 | 3.1623E-15 | 58.41 |
| *GLB1* | rs145078947 | G | T | 0.0026 | 0.729 | 0.07 | 1.9953E-25 | 108.46 |
| *GLB1* | rs58542926 | C | T | 0.0755 | 0.198 | 0.014 | 1E-48 | 200.02 |
| *GLB1* | rs142042446 | G | GTAA | 0.1496 | -0.07 | 0.01 | 5.0119E-12 | 49.00 |
| *GLO1* | rs4746 | T | G | 0.4638 | -0.331 | 0.008 | 1E-200 | 1711.89 |
| *GLOD4* | rs1354034 | T | C | 0.604 | 0.058 | 0.008 | 1.9953E-14 | 52.56 |
| *GLOD4* | rs150230220 | A | AT | 0.3114 | 0.064 | 0.008 | 3.9811E-15 | 64.00 |
| *GLRX* | rs6556886 | G | A | 0.1291 | -0.753 | 0.011 | 1E-200 | 4686.02 |
| *GLRX* | rs11759553 | A | T | 0.2728 | -0.065 | 0.007 | 3.1623E-18 | 86.22 |
| *GLRX5* | rs342299 | C | T | 0.4549 | -0.058 | 0.008 | 2.5119E-14 | 52.56 |
| *GLRX5* | rs7080386 | C | A | 0.412 | 0.063 | 0.008 | 7.9433E-16 | 62.02 |
| *GLRX5* | rs60822569 | T | TTTCTC | 0.5542 | 0.052 | 0.008 | 1.5849E-11 | 42.25 |
| *GLRX5* | rs179133 | C | T | 0.1826 | 0.156 | 0.01 | 1.5849E-54 | 243.36 |
| *GLT8D2* | rs117801489 | T | C | 0.0173 | 0.675 | 0.03 | 1E-114 | 506.25 |
| *GLYR1* | rs7080536 | G | A | 0.0437 | -0.229 | 0.019 | 5.0119E-34 | 145.27 |
| *GM2A* | rs153478 | A | G | 0.6428 | -0.695 | 0.008 | 1E-200 | 7547.27 |
| *GM2A* | rs77924615 | G | A | 0.1967 | -0.064 | 0.008 | 5.0119E-15 | 64.00 |
| *GMFG* | rs1354034 | T | C | 0.6045 | 0.054 | 0.008 | 1.9953E-12 | 45.56 |
| *GMFG* | rs342298 | C | T | 0.4545 | -0.063 | 0.008 | 7.9433E-17 | 62.02 |
| *GMFG* | rs194016 | A | G | 0.1098 | -0.104 | 0.012 | 5.0119E-18 | 75.11 |
| *GMPR* | rs1042391 | T | A | 0.3798 | 0.252 | 0.008 | 1E-200 | 992.25 |
| *GMPR* | rs342293 | C | G | 0.4594 | -0.049 | 0.007 | 7.9433E-12 | 49.00 |
| *GMPR2* | rs1354034 | T | C | 0.6044 | 0.068 | 0.008 | 3.9811E-19 | 72.25 |
| *GMPR2* | rs7765828 | C | G | 0.381 | -0.094 | 0.008 | 6.3096E-34 | 138.06 |
| *GMPR2* | rs7080386 | C | A | 0.412 | 0.061 | 0.008 | 1.5849E-15 | 58.14 |
| *GMPR2* | rs11502185 | T | C | 0.2597 | 0.065 | 0.01 | 1E-11 | 42.25 |
| *GMPR2* | rs199867463 | T | C | 0.0017 | -1.599 | 0.099 | 1.5849E-58 | 260.87 |

| | | | | | | | | |
|---|---|---|---|---|---|---|---|---|
| *GNAS* | rs188468174 | C | T | 0.0142 | 0.241 | 0.032 | 5.0119E-14 | 56.72 |
| *GNAS* | rs41263822 | C | A | 0.3435 | -0.081 | 0.008 | 1E-22 | 102.52 |
| *GNAS* | rs6026450 | A | G | 0.632 | 0.075 | 0.008 | 1.5849E-22 | 87.89 |
| *GNE* | rs1354034 | T | C | 0.6038 | 0.128 | 0.008 | 6.3096E-62 | 256.00 |
| *GNE* | rs6993770 | A | T | 0.2869 | -0.061 | 0.008 | 3.9811E-13 | 58.14 |
| *GNE* | rs7090758 | T | C | 0.4762 | -0.053 | 0.008 | 3.1623E-12 | 43.89 |
| *GNE* | rs11502185 | T | C | 0.2597 | 0.069 | 0.01 | 1.2589E-12 | 47.61 |
| *GNLY* | rs144015106 | A | G | 0.0068 | -0.503 | 0.042 | 5.0119E-33 | 143.43 |
| *GNLY* | rs628745 | T | C | 0.33 | 0.057 | 0.007 | 6.3096E-16 | 66.31 |
| *GNLY* | rs7545591 | C | G | 0.3219 | 0.058 | 0.007 | 3.9811E-16 | 68.65 |
| *GNLY* | rs7577293 | C | T | 0.2307 | 0.708 | 0.009 | 1E-200 | 6188.44 |
| *GNLY* | rs73069104 | T | C | 0.3627 | 0.067 | 0.007 | 3.9811E-22 | 91.61 |
| *GNLY* | rs11711157 | C | T | 0.2782 | -0.056 | 0.007 | 2.5119E-14 | 64.00 |
| *GNLY* | rs2927608 | G | A | 0.4498 | 0.072 | 0.007 | 2.5119E-27 | 105.80 |
| *GNLY* | rs9368661 | C | T | 0.2598 | 0.126 | 0.008 | 7.9433E-61 | 248.06 |
| *GNLY* | rs8177651 | T | A | 0.3233 | 0.055 | 0.007 | 3.9811E-14 | 61.73 |
| *GNLY* | rs745932882 | AG | A | 0.4558 | 0.056 | 0.007 | 1.2589E-16 | 64.00 |
| *GNLY* | rs4919420 | A | G | 0.4162 | -0.066 | 0.007 | 7.9433E-23 | 88.90 |
| *GNLY* | rs7137828 | C | T | 0.5164 | -0.045 | 0.007 | 1.5849E-11 | 41.33 |
| *GNLY* | rs9916257 | G | T | 0.4397 | -0.056 | 0.007 | 5.0119E-17 | 64.00 |
| *GNPDA1* | rs2228243 | A | G | 0.2003 | 0.087 | 0.009 | 3.1623E-20 | 93.44 |
| *GNPDA1* | rs11949303 | T | C | 0.1486 | 0.128 | 0.011 | 7.9433E-33 | 135.40 |
| *GNPDA1* | rs55761427 | G | A | 0.0265 | -0.285 | 0.024 | 1.9953E-33 | 141.02 |
| *GNPDA2* | rs1354034 | T | C | 0.6044 | 0.069 | 0.008 | 5.0119E-20 | 74.39 |
| *GNPDA2* | rs769805766 | CTATTA | C | 0.6402 | 0.236 | 0.008 | 1.259E-197 | 870.25 |
| *GNPDA2* | rs9324861 | C | T | 0.1493 | -0.248 | 0.01 | 1E-124 | 615.04 |
| *GNPDA2* | rs7974006 | A | G | 0.1026 | 0.105 | 0.012 | 3.9811E-17 | 76.56 |
| *GOLGA3* | rs3811444 | C | T | 0.3327 | -0.187 | 0.008 | 1.585E-122 | 546.39 |
| *GOLGA3* | rs1354034 | T | C | 0.6043 | 0.096 | 0.008 | 3.9811E-37 | 144.00 |
| *GOLGA3* | rs342299 | C | T | 0.455 | -0.055 | 0.007 | 2.5119E-13 | 61.73 |
| *GOLGA3* | rs11147139 | G | A | 0.2221 | 0.118 | 0.009 | 1E-39 | 171.90 |
| *GOLM2* | rs10804823 | A | T | 0.4936 | 0.052 | 0.007 | 1.9953E-12 | 55.18 |
| *GOLM2* | rs13107325 | C | T | 0.0747 | -0.106 | 0.014 | 5.0119E-14 | 57.33 |
| *GOLM2* | rs1265905 | A | G | 0.1298 | -0.111 | 0.011 | 3.1623E-24 | 101.83 |
| *GOLM2* | rs41266341 | A | C | 0.0089 | 0.409 | 0.039 | 1E-25 | 109.98 |
| *GOLM2* | rs550057 | C | T | 0.2557 | 0.262 | 0.009 | 1E-200 | 847.46 |
| *GOLM2* | rs488962 | C | T | 0.226 | 0.214 | 0.009 | 3.162E-129 | 565.38 |
| *GOLM2* | rs10777172 | T | C | 0.7123 | 0.199 | 0.008 | 3.981E-131 | 618.77 |
| *GOLM2* | rs550761239 | C | T | 0.0042 | 0.557 | 0.064 | 3.9811E-18 | 75.74 |
| *GOLM2* | rs77542162 | A | G | 0.0229 | 0.235 | 0.025 | 7.9433E-22 | 88.36 |
| *GOPC* | rs1354034 | T | C | 0.6043 | 0.077 | 0.008 | 3.1623E-24 | 92.64 |
| *GOPC* | rs6993770 | A | T | 0.2869 | -0.057 | 0.008 | 7.9433E-12 | 50.77 |
| *GOPC* | rs11502185 | T | C | 0.2598 | 0.077 | 0.01 | 7.9433E-16 | 59.29 |
| *GOPC* | rs892090 | T | G | 0.8337 | 0.069 | 0.01 | 1E-11 | 47.61 |
| *GORASP2* | rs753954550 | TAAAGG | T | 0.2925 | 0.058 | 0.008 | 3.9811E-12 | 52.56 |
| *GORASP2* | rs1354034 | T | C | 0.6046 | 0.072 | 0.008 | 7.9433E-21 | 81.00 |
| *GORASP2* | rs6993770 | A | T | 0.2866 | -0.065 | 0.008 | 5.0119E-15 | 66.02 |
| *GOT1* | rs56278466 | T | G | 0.6608 | 0.109 | 0.008 | 3.1623E-45 | 185.64 |
| *GOT1* | rs3747207 | G | A | 0.2155 | 0.083 | 0.009 | 5.0119E-21 | 85.05 |
| *GP1BA* | rs3767809 | T | C | 0.224 | 0.067 | 0.009 | 7.9433E-14 | 55.42 |
| *GP1BA* | rs3804749 | C | T | 0.5929 | -0.053 | 0.008 | 3.1623E-12 | 43.89 |
| *GP1BA* | rs78565404 | C | T | 0.0548 | 0.132 | 0.017 | 1.9953E-15 | 60.29 |
| *GP1BA* | rs2853677 | G | A | 0.5775 | -0.052 | 0.008 | 3.1623E-12 | 42.25 |
| *GP1BA* | rs35786788 | G | A | 0.2524 | 0.104 | 0.009 | 5.0119E-33 | 133.53 |
| *GP1BA* | rs6961069 | C | T | 0.4025 | 0.061 | 0.008 | 1.5849E-15 | 58.14 |
| *GP1BA* | rs6993770 | A | T | 0.2865 | -0.126 | 0.008 | 7.9433E-53 | 248.06 |
| *GP1BA* | rs385893 | T | C | 0.5243 | 0.108 | 0.007 | 3.1623E-47 | 238.04 |
| *GP1BA* | rs12762934 | C | T | 0.3254 | 0.054 | 0.008 | 7.9433E-12 | 45.56 |
| *GP1BA* | rs11217176 | G | A | 0.2685 | 0.068 | 0.008 | 7.9433E-16 | 72.25 |
| *GP1BA* | rs7944186 | A | G | 0.5259 | -0.087 | 0.007 | 3.9811E-31 | 154.47 |
| *GP1BA* | rs3184504 | T | C | 0.5171 | -0.135 | 0.007 | 1.5849E-73 | 371.94 |
| *GP1BA* | rs2127870 | G | C | 0.7951 | 0.064 | 0.009 | 5.0119E-12 | 50.57 |
| *GP1BA* | rs1555405 | G | A | 0.2506 | -0.072 | 0.009 | 7.9433E-17 | 64.00 |
| *GP1BA* | rs7176023 | T | C | 0.0655 | 0.11 | 0.015 | 3.9811E-13 | 53.78 |
| *GP1BA* | rs553749201 | C | A | 0.0528 | -0.256 | 0.017 | 3.1623E-51 | 226.77 |
| *GP1BB* | rs826404 | T | C | 0.5862 | 0.19 | 0.008 | 1E-132 | 564.06 |
| *GP1BB* | rs3811444 | C | T | 0.3322 | -0.063 | 0.008 | 5.0119E-15 | 62.02 |
| *GP1BB* | rs854572 | C | G | 0.5282 | 0.067 | 0.008 | 7.9433E-19 | 70.14 |

| | | | | | | | | |
|---|---|---|---|---|---|---|---|---|
| *GP1BB* | rs6993770 | A | T | 0.2865 | -0.105 | 0.008 | 1.9953E-36 | 172.27 |
| *GP1BB* | rs7896518 | A | G | 0.4248 | 0.111 | 0.008 | 3.1623E-47 | 192.52 |
| *GP1BB* | rs6580981 | G | A | 0.4581 | 0.076 | 0.008 | 1.2589E-23 | 90.25 |
| *GP1BB* | rs77639164 | A | C | 0.2782 | 0.098 | 0.009 | 1E-30 | 118.57 |
| *GP1BB* | rs56337033 | C | T | 0.0037 | 0.52 | 0.067 | 7.9433E-15 | 60.24 |
| *GP1BB* | rs547921381 | C | T | 0.0045 | -1.018 | 0.061 | 6.3096E-63 | 278.51 |
| *GP2* | rs6571015 | G | A | 0.7904 | 0.148 | 0.008 | 3.1623E-71 | 342.25 |
| *GP2* | rs6459924 | T | C | 0.2411 | -0.062 | 0.008 | 2.5119E-15 | 60.06 |
| *GP2* | rs505922 | T | C | 0.3162 | 0.556 | 0.008 | 1E-200 | 4830.25 |
| *GP2* | rs1401981 | G | A | 0.1692 | -0.066 | 0.009 | 3.1623E-13 | 53.78 |
| *GP2* | rs4430753 | A | G | 0.4878 | -0.233 | 0.007 | 1E-200 | 1107.94 |
| *GP2* | rs200489612 | G | A | 0.0052 | 0.373 | 0.051 | 1.5849E-13 | 53.49 |
| *GP2* | rs708686 | C | T | 0.2678 | 0.063 | 0.008 | 3.1623E-16 | 62.02 |
| *GP2* | rs492602 | A | G | 0.5088 | 0.169 | 0.007 | 1E-135 | 582.88 |
| *GP5* | rs56221835 | G | C | 0.0927 | 0.093 | 0.013 | 1.5849E-12 | 51.18 |
| *GP5* | rs3804749 | C | T | 0.593 | -0.111 | 0.008 | 7.9433E-45 | 192.52 |
| *GP5* | rs10638925 | C | CCATAAA | 0.2288 | -0.137 | 0.009 | 2.5119E-50 | 231.72 |
| *GP5* | rs6993770 | A | T | 0.287 | -0.081 | 0.008 | 3.9811E-22 | 102.52 |
| *GP5* | rs12005199 | G | A | 0.3119 | 0.058 | 0.008 | 5.0119E-12 | 52.56 |
| *GP5* | rs8099942 | C | G | 0.8006 | 0.07 | 0.01 | 7.9433E-13 | 49.00 |
| *GP6* | rs1354034 | T | C | 0.604 | 0.059 | 0.008 | 3.9811E-15 | 54.39 |
| *GP6* | rs17622656 | G | A | 0.3886 | -0.053 | 0.008 | 2.5119E-12 | 43.89 |
| *GP6* | rs6993770 | A | T | 0.2863 | -0.078 | 0.008 | 1E-21 | 95.06 |
| *GP6* | rs7896518 | A | G | 0.4243 | 0.059 | 0.008 | 7.9433E-15 | 54.39 |
| *GP6* | rs10774624 | G | A | 0.5136 | -0.051 | 0.007 | 6.3096E-12 | 53.08 |
| *GP6* | rs892090 | T | G | 0.8343 | 0.413 | 0.01 | 1E-200 | 1705.69 |
| *GPA33* | rs72689400 | C | A | 0.0162 | -0.679 | 0.029 | 7.943E-124 | 548.21 |
| *GPA33* | rs2241764 | C | T | 0.3386 | -0.094 | 0.008 | 7.9433E-35 | 138.06 |
| *GPA33* | rs35853577 | T | C | 0.4255 | -0.074 | 0.007 | 1E-23 | 111.76 |
| *GPA33* | rs2077610 | T | A | 0.5518 | 0.051 | 0.007 | 2.5119E-12 | 53.08 |
| *GPA33* | rs9816553 | G | A | 0.8521 | -0.09 | 0.01 | 1.9953E-18 | 81.00 |
| *GPA33* | rs939885 | G | A | 0.4848 | 0.072 | 0.007 | 3.1623E-23 | 105.80 |
| *GPA33* | rs533067298 | CT | C | 0.5111 | -0.049 | 0.007 | 1E-11 | 49.00 |
| *GPA33* | rs6911827 | C | T | 0.4531 | 0.051 | 0.007 | 1.5849E-12 | 53.08 |
| *GPA33* | rs9478784 | C | T | 0.0955 | 0.123 | 0.012 | 7.9433E-24 | 105.06 |
| *GPA33* | rs931493 | G | A | 0.0318 | -0.176 | 0.021 | 5.0119E-17 | 70.24 |
| *GPA33* | rs2291428 | G | C | 0.244 | -0.085 | 0.008 | 3.9811E-24 | 112.89 |
| *GPA33* | rs56398830 | G | A | 0.0111 | -0.889 | 0.035 | 2.512E-146 | 645.16 |
| *GPA33* | rs35193388 | G | GC | 0.7354 | -0.074 | 0.008 | 5.0119E-19 | 85.56 |
| *GPA33* | rs6505173 | A | G | 0.4397 | 0.162 | 0.007 | 3.981E-109 | 535.59 |
| *GPA33* | rs681343 | C | T | 0.5088 | -0.207 | 0.007 | 5.012E-177 | 874.47 |
| *GPA33* | rs3746778 | G | A | 0.4202 | 0.072 | 0.007 | 7.9433E-22 | 105.80 |
| *GPC1* | rs1535701 | T | G | 0.6225 | 0.079 | 0.007 | 1.5849E-27 | 127.37 |
| *GPC1* | rs12758626 | A | G | 0.3491 | -0.05 | 0.007 | 1.5849E-11 | 51.02 |
| *GPC1* | rs150816167 | T | C | 0.0417 | 0.125 | 0.018 | 1.9953E-12 | 48.23 |
| *GPC1* | rs2273967 | C | T | 0.2491 | 0.072 | 0.008 | 1E-18 | 81.00 |
| *GPC1* | rs71414736 | C | G | 0.0215 | -0.192 | 0.025 | 1.5849E-14 | 58.98 |
| *GPC1* | rs147895889 | A | G | 0.0108 | -0.272 | 0.035 | 1.5849E-14 | 60.40 |
| *GPC1* | rs77078946 | T | G | 0.1141 | 0.506 | 0.011 | 1E-200 | 2116.00 |
| *GPC1* | rs200409727 | A | AT | 0.1966 | -0.062 | 0.009 | 3.1623E-12 | 47.46 |
| *GPC1* | rs3965330 | G | A | 0.4687 | 0.189 | 0.007 | 1E-155 | 729.00 |
| *GPC1* | rs34236411 | C | CA | 0.7659 | -0.082 | 0.008 | 5.0119E-23 | 105.06 |
| *GPC1* | rs2625445 | G | A | 0.2435 | -0.074 | 0.008 | 2.5119E-19 | 85.56 |
| *GPC1* | rs7835610 | C | T | 0.5046 | -0.055 | 0.007 | 2.5119E-15 | 61.73 |
| *GPC1* | rs11414746 | T | TA | 0.4233 | 0.079 | 0.007 | 3.9811E-28 | 127.37 |
| *GPC1* | rs56278466 | T | G | 0.6612 | 0.062 | 0.007 | 7.9433E-17 | 78.45 |
| *GPC1* | rs7971751 | A | C | 0.569 | -0.052 | 0.007 | 3.1623E-13 | 55.18 |
| *GPC1* | rs9318186 | A | G | 0.555 | -0.052 | 0.007 | 2.5119E-13 | 55.18 |
| *GPC1* | rs7147722 | G | T | 0.5383 | 0.103 | 0.007 | 1.5849E-48 | 216.51 |
| *GPC1* | rs1801689 | A | C | 0.03 | 0.191 | 0.021 | 1E-20 | 82.72 |
| *GPC1* | rs5763646 | T | C | 0.4116 | 0.058 | 0.007 | 6.3096E-16 | 68.65 |
| *GPC5* | rs1260326 | T | C | 0.6084 | -0.058 | 0.006 | 6.3096E-20 | 93.44 |
| *GPC5* | rs1354034 | T | C | 0.6041 | -0.048 | 0.006 | 1E-14 | 64.00 |
| *GPC5* | rs4098923 | A | G | 0.4477 | -0.044 | 0.006 | 1E-11 | 53.78 |
| *GPC5* | rs6993770 | A | T | 0.2865 | -0.062 | 0.007 | 1E-19 | 78.45 |
| *GPC5* | rs28601761 | C | G | 0.4191 | -0.051 | 0.006 | 1E-15 | 72.25 |
| *GPC5* | rs7979478 | A | G | 0.6161 | 0.052 | 0.006 | 5.0119E-16 | 75.11 |
| *GPC5* | rs342706 | T | C | 0.2717 | -0.878 | 0.009 | 1E-200 | 9517.09 |

| | | | | | | | | |
|---|---|---|---|---|---|---|---|---|
| *GPC5* | rs1654425 | T | C | 0.8343 | 0.092 | 0.008 | 1.2589E-28 | 132.25 |
| *GPD1* | rs35428353 | G | A | 0.0027 | -1.239 | 0.072 | 2.5119E-66 | 296.13 |
| *GPHA2* | rs114391421 | C | A | 0.0461 | 0.499 | 0.018 | 3.981E-174 | 768.52 |
| *GPHA2* | rs215225 | A | G | 0.4277 | 0.086 | 0.007 | 1.2589E-30 | 150.94 |
| *GPHA2* | rs1002136 | C | A | 0.4002 | -0.101 | 0.008 | 1.9953E-40 | 159.39 |
| *GPI* | rs1354034 | T | C | 0.6042 | 0.061 | 0.008 | 1.9953E-15 | 58.14 |
| *GPI* | rs342290 | A | C | 0.4555 | -0.055 | 0.008 | 1.9953E-13 | 47.27 |
| *GPI* | rs763990408 | A | ATG | 0.0286 | -0.222 | 0.023 | 7.9433E-23 | 93.16 |
| *GPIHBP1* | rs12664131 | T | A | 0.1519 | -0.072 | 0.01 | 5.0119E-12 | 51.84 |
| *GPIHBP1* | rs9657541 | C | T | 0.2055 | -0.072 | 0.009 | 2.5119E-14 | 64.00 |
| *GPIHBP1* | rs6420185 | G | A | 0.388 | -0.272 | 0.008 | 1E-200 | 1156.00 |
| *GPIHBP1* | rs8176741 | G | A | 0.0614 | 0.149 | 0.016 | 1E-21 | 86.72 |
| *GPNMB* | rs11721064 | G | T | 0.6054 | -0.063 | 0.007 | 3.1623E-17 | 81.00 |
| *GPNMB* | rs12203592 | C | T | 0.2163 | -0.076 | 0.009 | 2.5119E-17 | 71.31 |
| *GPNMB* | rs75801644 | G | A | 0.0227 | -1.118 | 0.026 | 1E-200 | 1849.00 |
| *GPNMB* | rs74612335 | T | C | 0.1331 | -0.089 | 0.011 | 1.2589E-16 | 65.46 |
| *GPNMB* | rs186021206 | G | A | 0.0064 | 0.738 | 0.05 | 1.2589E-49 | 217.86 |
| *GPR158* | rs1931291 | A | T | 0.6948 | -0.064 | 0.008 | 3.1623E-15 | 64.00 |
| *GPR15L* | rs61816761 | G | A | 0.0234 | 0.236 | 0.025 | 1.5849E-21 | 89.11 |
| *GPR15L* | rs9326982 | T | C | 0.394 | 0.053 | 0.007 | 3.9811E-13 | 57.33 |
| *GPR15L* | rs4399275 | T | G | 0.4214 | 0.221 | 0.008 | 2.512E-187 | 763.14 |
| *GPR15L* | rs117268623 | C | T | 0.0325 | 0.141 | 0.02 | 1.9953E-12 | 49.70 |
| *GPR37* | rs780094 | T | C | 0.62 | -0.083 | 0.007 | 2.5119E-37 | 140.59 |
| *GPR37* | rs511154 | A | G | 0.7663 | 0.071 | 0.007 | 3.9811E-21 | 102.88 |
| *GPR37* | rs782470771 | AAG | A | 0.1279 | -0.108 | 0.011 | 2.5119E-22 | 96.40 |
| *GPR37* | rs60598740 | G | A | 0.2618 | -0.785 | 0.009 | 1E-200 | 7607.72 |
| *GPR37* | rs28601761 | C | G | 0.4189 | -0.09 | 0.006 | 7.9433E-44 | 225.00 |
| *GPR37* | rs11023881 | T | A | 0.3874 | 0.059 | 0.006 | 3.9811E-20 | 96.69 |
| *GPR37* | rs964184 | G | C | 0.8669 | -0.063 | 0.009 | 7.9433E-12 | 49.00 |
| *GPR37* | rs10083137 | A | G | 0.0343 | 0.211 | 0.017 | 3.9811E-34 | 154.05 |
| *GPR37* | rs1292066 | T | A | 0.2145 | 0.06 | 0.008 | 3.9811E-15 | 56.25 |
| *GPR37* | rs390082 | T | G | 0.1094 | 0.083 | 0.01 | 1.5849E-16 | 68.89 |
| *GPRC5C* | rs4835265 | C | A | 0.1562 | 0.089 | 0.01 | 1.9953E-17 | 79.21 |
| *GPRC5C* | rs149425950 | A | AAT | 0.1511 | 0.081 | 0.011 | 2.5119E-14 | 54.22 |
| *GPRC5C* | rs3741513 | T | A | 0.6107 | 0.058 | 0.008 | 1.2589E-13 | 52.56 |
| *GPRC5C* | rs550999044 | A | AT | 0.2438 | 0.097 | 0.009 | 5.0119E-28 | 116.16 |
| *GPRC5C* | rs2670824 | C | G | 0.9236 | -0.213 | 0.014 | 1.9953E-49 | 231.47 |
| *GPRC5C* | rs9959832 | C | T | 0.7977 | -0.073 | 0.009 | 7.9433E-15 | 65.79 |
| *GPRC5C* | rs189448562 | G | C | 0.0112 | 0.431 | 0.037 | 6.3096E-32 | 135.69 |
| *GRAP2* | rs1354034 | T | C | 0.6037 | 0.053 | 0.008 | 3.9811E-12 | 43.89 |
| *GRAP2* | rs6993770 | A | T | 0.2866 | -0.063 | 0.008 | 2.5119E-14 | 62.02 |
| *GRAP2* | rs148328786 | A | G | 0.0135 | 0.322 | 0.034 | 3.1623E-21 | 89.69 |
| *GRHPR* | rs1354034 | T | C | 0.6045 | 0.057 | 0.007 | 3.1623E-14 | 66.31 |
| *GRHPR* | rs342298 | C | T | 0.4545 | -0.051 | 0.007 | 6.3096E-12 | 53.08 |
| *GRHPR* | rs6993770 | A | T | 0.2866 | -0.059 | 0.008 | 3.9811E-13 | 54.39 |
| *GRHPR* | rs4878690 | C | T | 0.3263 | 0.269 | 0.008 | 1E-200 | 1130.64 |
| *GRIK2* | rs59321237 | C | T | 0.0292 | 0.738 | 0.023 | 1E-200 | 1029.57 |
| *GRIK2* | rs35106244 | C | T | 0.4633 | 0.061 | 0.008 | 3.9811E-15 | 58.14 |
| *GRK5* | rs10886430 | A | G | 0.1275 | -0.407 | 0.012 | 1E-200 | 1150.34 |
| *GRN* | rs12740374 | G | T | 0.2216 | -0.676 | 0.009 | 1E-200 | 5641.68 |
| *GRN* | rs75029704 | C | T | 0.0957 | 0.161 | 0.012 | 3.1623E-43 | 180.01 |
| *GRN* | rs1137827 | C | T | 0.0613 | 0.109 | 0.014 | 7.9433E-15 | 60.62 |
| *GRN* | rs885828 | G | A | 0.2915 | -0.237 | 0.008 | 1E-200 | 877.64 |
| *GRN* | rs2298475 | T | C | 0.0755 | -0.102 | 0.013 | 7.9433E-16 | 61.56 |
| *GRN* | rs558314 | C | G | 0.5456 | -0.046 | 0.007 | 1E-11 | 43.18 |
| *GRN* | rs72802342 | C | A | 0.0758 | -0.197 | 0.013 | 7.9433E-52 | 229.64 |
| *GRN* | rs186021206 | G | A | 0.0064 | 0.684 | 0.047 | 2.5119E-48 | 211.80 |
| *GRN* | rs5848 | C | T | 0.2745 | -0.25 | 0.008 | 1E-200 | 976.56 |
| *GRN* | rs1065853 | G | T | 0.0814 | 0.097 | 0.012 | 6.3096E-15 | 65.34 |
| *GRP* | rs964184 | G | C | 0.8669 | -0.085 | 0.011 | 3.9811E-15 | 59.71 |
| *GRP* | rs76895963 | T | G | 0.0207 | 0.222 | 0.028 | 3.9811E-15 | 62.86 |
| *GRP* | rs11105337 | A | T | 0.1705 | -0.074 | 0.01 | 3.1623E-14 | 54.76 |
| *GRP* | rs529361264 | A | AT | 0.2586 | 0.065 | 0.008 | 1.2589E-14 | 66.02 |
| *GRP* | rs7216766 | C | G | 0.8155 | -0.075 | 0.009 | 1.5849E-15 | 69.44 |
| *GRP* | rs17835713 | G | A | 0.1653 | 0.462 | 0.01 | 1E-200 | 2134.44 |
| *GRP* | rs141700858 | T | TCGC | 0.1616 | -0.122 | 0.011 | 1.2589E-29 | 123.01 |
| *GRP* | rs72654473 | C | A | 0.1094 | 0.114 | 0.012 | 2.5119E-22 | 90.25 |
| *GRPEL1* | rs4361977 | C | T | 0.4338 | -0.053 | 0.008 | 1.9953E-12 | 43.89 |

| | | | | | | | | |
|---|---|---|---|---|---|---|---|---|
| *GRPEL1* | rs4835265 | C | A | 0.156 | 0.107 | 0.01 | 5.0119E-25 | 114.49 |
| *GRPEL1* | rs9959832 | C | T | 0.7982 | -0.081 | 0.009 | 7.9433E-18 | 81.00 |
| *GRSF1* | rs342298 | C | T | 0.4547 | -0.055 | 0.008 | 1.9953E-12 | 47.27 |
| *GSAP* | rs73333648 | C | G | 0.0538 | -0.141 | 0.017 | 2.5119E-16 | 68.79 |
| *GSN* | rs116185403 | C | T | 0.007 | 1.026 | 0.045 | 1E-116 | 519.84 |
| *GSN* | rs7305932 | T | A | 0.779 | -0.062 | 0.009 | 1E-11 | 47.46 |
| *GSN* | rs3184504 | T | C | 0.517 | 0.079 | 0.007 | 1.5849E-26 | 127.37 |
| *GSN* | rs9936550 | C | T | 0.0771 | -0.098 | 0.014 | 3.9811E-12 | 49.00 |
| *GSN* | rs12986064 | T | C | 0.5081 | -0.082 | 0.008 | 1.2589E-26 | 105.06 |
| *GSR* | rs34744948 | G | C | 0.0486 | 0.128 | 0.017 | 6.3096E-14 | 56.69 |
| *GSR* | rs8190982 | T | TCC | 0.3254 | -0.125 | 0.008 | 1E-53 | 244.14 |
| *GSR* | rs12342201 | G | A | 0.4762 | -0.062 | 0.007 | 7.9433E-17 | 78.45 |
| *GSR* | rs7974006 | A | G | 0.1025 | 0.151 | 0.012 | 6.3096E-34 | 158.34 |
| *GSTA1* | rs28383314 | T | C | 0.6258 | 0.053 | 0.008 | 5.0119E-12 | 43.89 |
| *GSTA1* | rs6458871 | C | T | 0.5737 | 0.315 | 0.007 | 1E-200 | 2025.00 |
| *GSTA1* | rs112875651 | G | A | 0.391 | -0.059 | 0.007 | 3.9811E-15 | 71.04 |
| *GSTA1* | rs10733608 | G | T | 0.4881 | -0.068 | 0.007 | 1.2589E-20 | 94.37 |
| *GSTA1* | rs10883451 | T | C | 0.5018 | -0.053 | 0.007 | 2.5119E-13 | 57.33 |
| *GSTA1* | rs28929474 | C | T | 0.0208 | 0.216 | 0.025 | 1.2589E-17 | 74.65 |
| *GSTA1* | rs10405357 | T | C | 0.4354 | -0.058 | 0.007 | 1.5849E-15 | 68.65 |
| *GSTA1* | rs3747207 | G | A | 0.2153 | 0.11 | 0.009 | 1.9953E-36 | 149.38 |
| *GSTA3* | rs2749005 | T | G | 0.6478 | 0.325 | 0.008 | 1E-200 | 1650.39 |
| *GSTA3* | rs11370710 | C | CA | 0.4939 | -0.052 | 0.007 | 5.0119E-12 | 55.18 |
| *GSTA3* | rs28929474 | C | T | 0.0207 | 0.218 | 0.026 | 6.3096E-17 | 70.30 |
| *GSTA3* | rs738409 | C | G | 0.2167 | 0.097 | 0.009 | 2.5119E-27 | 116.16 |
| *GSTM4* | rs140584594 | A | G | 0.7321 | -0.43 | 0.009 | 1E-200 | 2282.72 |
| *GSTP1* | rs762803 | C | A | 0.4489 | 0.205 | 0.008 | 3.981E-152 | 656.64 |
| *GSTT2B* | rs5751777 | C | T | 0.5883 | -1.037 | 0.008 | 1E-200 | 16802.64 |
| *GTF2IRD1* | rs149375627 | T | TG | 0.4351 | 0.092 | 0.009 | 1E-25 | 104.49 |
| *GTPBP2* | rs342298 | C | T | 0.4547 | -0.054 | 0.008 | 1E-12 | 45.56 |
| *GTPBP2* | rs6993770 | A | T | 0.2866 | -0.08 | 0.008 | 6.3096E-22 | 100.00 |
| *GUCA2A* | rs142157365 | A | T | 0.0023 | -2.109 | 0.078 | 5.012E-162 | 731.08 |
| *GUCA2A* | rs3044077 | T | TGT | 0.6558 | -0.08 | 0.008 | 2.5119E-24 | 100.00 |
| *GUCA2A* | rs10455394 | C | G | 0.4905 | -0.088 | 0.007 | 5.0119E-33 | 158.04 |
| *GUCA2A* | rs782819119 | AAGAAGGG | A | 0.31 | 0.064 | 0.008 | 1.2589E-15 | 64.00 |
| *GUCA2A* | rs2511241 | C | T | 0.9296 | 0.11 | 0.014 | 1.9953E-14 | 61.73 |
| *GUCA2A* | rs4982408 | A | C | 0.8372 | 0.086 | 0.01 | 3.9811E-18 | 73.96 |
| *GUCA2A* | rs1424231 | T | C | 0.3365 | 0.052 | 0.008 | 1.5849E-11 | 42.25 |
| *GUCA2A* | rs17743980 | G | A | 0.3151 | 0.056 | 0.008 | 1.5849E-12 | 49.00 |
| *GUCY2C* | rs2516979 | C | G | 0.3144 | -0.112 | 0.01 | 7.9433E-31 | 125.44 |
| *GUSB* | rs6779146 | T | C | 0.2309 | -0.1 | 0.008 | 1.9953E-33 | 156.25 |
| *GUSB* | rs115850161 | T | C | 0.0606 | 0.178 | 0.015 | 1E-33 | 140.82 |
| *GUSB* | rs72814178 | A | G | 0.1323 | 0.074 | 0.01 | 7.9433E-13 | 54.76 |
| *GUSB* | rs111331197 | C | T | 0.011 | -0.256 | 0.033 | 1.9953E-14 | 60.18 |
| *GUSB* | rs28538349 | G | A | 0.5186 | -0.052 | 0.007 | 3.1623E-13 | 55.18 |
| *GUSB* | rs149606212 | C | T | 0.0021 | -1.405 | 0.076 | 1.2589E-76 | 341.76 |
| *GUSB* | rs11003134 | C | A | 0.1435 | 0.069 | 0.01 | 3.9811E-12 | 47.61 |
| *GUSB* | rs99780 | C | T | 0.3501 | 0.083 | 0.007 | 5.0119E-30 | 140.59 |
| *GUSB* | rs10778152 | A | G | 0.2884 | -0.257 | 0.008 | 1E-200 | 1032.02 |
| *GUSB* | rs150641790 | T | TG | 0.0747 | 0.095 | 0.013 | 1E-12 | 53.40 |
| *GUSB* | rs139974673 | T | C | 0.0243 | 0.275 | 0.022 | 1.9953E-34 | 156.25 |
| *GUSB* | rs1801690 | C | G | 0.0566 | -0.274 | 0.015 | 6.3096E-74 | 333.67 |
| *GUSB* | rs5826313 | C | CA | 0.3213 | 0.063 | 0.007 | 6.3096E-17 | 81.00 |
| *GUSB* | rs2974755 | C | T | 0.6021 | 0.048 | 0.007 | 7.9433E-12 | 47.02 |
| *GUSB* | rs58542926 | C | T | 0.0755 | 0.193 | 0.013 | 1.5849E-48 | 220.41 |
| *GYS1* | rs3748136 | G | A | 0.2126 | -0.31 | 0.009 | 1E-200 | 1186.42 |
| *GYS1* | rs59305841 | A | T | 0.0423 | 0.126 | 0.019 | 1E-11 | 43.98 |
| *GZMA* | rs76036957 | G | A | 0.0093 | 0.303 | 0.038 | 1.5849E-15 | 63.58 |
| *GZMA* | rs13063578 | T | A | 0.3978 | -0.065 | 0.007 | 1.9953E-18 | 86.22 |
| *GZMA* | rs17033014 | A | G | 0.3328 | -0.057 | 0.008 | 1.5849E-13 | 50.77 |
| *GZMA* | rs2047745 | T | C | 0.2087 | 0.07 | 0.009 | 7.9433E-15 | 60.49 |
| *GZMA* | rs3846730 | C | T | 0.2383 | -0.06 | 0.009 | 3.9811E-12 | 44.44 |
| *GZMA* | rs9264645 | G | A | 0.6415 | 0.266 | 0.008 | 1E-200 | 1105.56 |
| *GZMA* | rs2347784 | C | G | 0.2721 | -0.074 | 0.008 | 2.5119E-19 | 85.56 |
| *GZMA* | rs6476398 | C | T | 0.3099 | -0.087 | 0.008 | 3.9811E-28 | 118.27 |
| *GZMA* | rs41280225 | A | C | 0.1314 | -0.089 | 0.011 | 1.2589E-16 | 65.46 |
| *GZMA* | rs113430316 | T | TA | 0.2815 | -0.056 | 0.008 | 6.3096E-12 | 49.00 |
| *GZMA* | rs75587749 | C | T | 0.0853 | 0.093 | 0.013 | 1E-12 | 51.18 |

| | | | | | | | | |
|---|---|---|---|---|---|---|---|---|
| *GZMA* | rs4764809 | A | T | 0.4934 | 0.06 | 0.007 | 3.1623E-16 | 73.47 |
| *GZMA* | rs3184504 | T | C | 0.5174 | -0.114 | 0.007 | 6.3096E-56 | 265.22 |
| *GZMA* | rs145078947 | G | T | 0.0026 | 0.656 | 0.071 | 3.9811E-20 | 85.37 |
| *GZMA* | rs2034892 | G | A | 0.6856 | -0.054 | 0.008 | 5.0119E-12 | 45.56 |
| *GZMA* | rs12986064 | T | C | 0.5076 | -0.079 | 0.008 | 1E-25 | 97.52 |
| *GZMB* | rs2843153 | C | T | 0.7879 | -0.066 | 0.009 | 3.1623E-14 | 53.78 |
| *GZMB* | rs1984520 | C | T | 0.3684 | -0.059 | 0.007 | 1.5849E-15 | 71.04 |
| *GZMB* | rs2725124 | C | A | 0.6179 | 0.07 | 0.007 | 1E-21 | 100.00 |
| *GZMB* | rs41563615 | G | T | 0.0543 | 0.413 | 0.016 | 3.162E-148 | 666.29 |
| *GZMB* | rs745932882 | AG | A | 0.4562 | 0.061 | 0.007 | 7.9433E-18 | 75.94 |
| *GZMB* | rs576785 | T | C | 0.2939 | -0.089 | 0.008 | 1E-30 | 123.77 |
| *GZMB* | rs2460740 | T | G | 0.6842 | -0.08 | 0.008 | 1E-25 | 100.00 |
| *GZMB* | rs35350651 | A | AC | 0.5041 | -0.055 | 0.007 | 3.9811E-15 | 61.73 |
| *GZMB* | rs8192918 | C | CT | 0.224 | -0.354 | 0.009 | 1E-200 | 1547.11 |
| *GZMB* | rs12451746 | G | A | 0.4296 | -0.133 | 0.007 | 6.3096E-77 | 361.00 |
| *GZMB* | rs56133626 | G | A | 0.326 | -0.06 | 0.008 | 1.5849E-15 | 56.25 |
| *GZMH* | rs10732976 | G | A | 0.6944 | -0.087 | 0.008 | 3.1623E-27 | 118.27 |
| *GZMH* | rs2524080 | A | T | 0.6765 | 0.123 | 0.008 | 5.0119E-55 | 236.39 |
| *GZMH* | rs2347784 | C | G | 0.272 | -0.063 | 0.008 | 5.0119E-14 | 62.02 |
| *GZMH* | rs78325861 | C | G | 0.0419 | 0.159 | 0.018 | 3.9811E-18 | 78.03 |
| *GZMH* | rs576785 | T | C | 0.2939 | -0.108 | 0.008 | 1E-40 | 182.25 |
| *GZMH* | rs2859660 | C | T | 0.8419 | -0.093 | 0.011 | 1.9953E-18 | 71.48 |
| *GZMH* | rs190749747 | C | G | 0.0112 | -0.897 | 0.037 | 3.162E-132 | 587.73 |
| *GZMH* | rs56133626 | G | A | 0.3265 | -0.053 | 0.008 | 1.5849E-11 | 43.89 |
| *H2AP* | rs35835517 | G | T | 0.1484 | 0.145 | 0.011 | 1E-39 | 173.76 |
| *HADH* | rs17038246 | C | T | 0.081 | 0.106 | 0.014 | 3.9811E-14 | 57.33 |
| *HADH* | rs190627972 | T | C | 0.1608 | 0.147 | 0.011 | 1E-40 | 178.59 |
| *HAGH* | rs115389912 | G | A | 0.0051 | -0.809 | 0.05 | 3.1623E-59 | 261.79 |
| *HAO1* | rs6656611 | T | C | 0.6018 | 0.062 | 0.007 | 5.0119E-17 | 78.45 |
| *HAO1* | rs12904 | G | A | 0.4115 | -0.058 | 0.007 | 2.5119E-15 | 68.65 |
| *HAO1* | rs7608141 | A | C | 0.3322 | 0.059 | 0.008 | 7.9433E-14 | 54.39 |
| *HAO1* | rs4835265 | C | A | 0.1565 | 0.119 | 0.01 | 6.3096E-33 | 141.61 |
| *HAO1* | rs112875651 | G | A | 0.3911 | -0.051 | 0.007 | 6.3096E-12 | 53.08 |
| *HAO1* | rs151068477 | G | T | 0.0083 | 0.285 | 0.042 | 1.5849E-11 | 46.05 |
| *HAO1* | rs12373325 | T | C | 0.798 | -0.098 | 0.009 | 1.5849E-27 | 118.57 |
| *HAO1* | rs738409 | C | G | 0.217 | 0.065 | 0.009 | 1E-13 | 52.16 |
| *HARS1* | rs1354034 | T | C | 0.6041 | 0.073 | 0.008 | 1.9953E-21 | 83.27 |
| *HARS1* | rs342298 | C | T | 0.4545 | -0.053 | 0.008 | 1.2589E-12 | 43.89 |
| *HARS1* | rs60822569 | T | TTTCTC | 0.5545 | 0.052 | 0.008 | 5.0119E-12 | 42.25 |
| *HAVCR1* | rs2070803 | A | G | 0.426 | 0.049 | 0.006 | 1E-15 | 66.69 |
| *HAVCR1* | rs434215 | A | G | 0.7238 | -0.061 | 0.008 | 1.9953E-13 | 58.14 |
| *HAVCR1* | rs6555820 | C | A | 0.4796 | -0.476 | 0.007 | 1E-200 | 4624.00 |
| *HAVCR1* | rs112875651 | G | A | 0.391 | -0.061 | 0.006 | 1.5849E-22 | 103.36 |
| *HAVCR1* | rs2070895 | G | A | 0.2214 | 0.078 | 0.007 | 1.2589E-26 | 124.16 |
| *HAVCR1* | rs77924615 | G | A | 0.1967 | -0.12 | 0.008 | 1.9953E-54 | 225.00 |
| *HAVCR1* | rs112001035 | G | A | 0.0612 | 0.221 | 0.013 | 3.1623E-65 | 289.00 |
| *HAVCR2* | rs61747728 | C | T | 0.0379 | 0.156 | 0.019 | 2.5119E-16 | 67.41 |
| *HAVCR2* | rs7569939 | A | G | 0.52 | -0.061 | 0.007 | 6.3096E-17 | 75.94 |
| *HAVCR2* | rs7630058 | C | T | 0.2209 | -0.083 | 0.009 | 7.9433E-21 | 85.05 |
| *HAVCR2* | rs116363290 | C | T | 0.0057 | -1.704 | 0.052 | 1E-200 | 1073.82 |
| *HAVCR2* | rs509227 | A | C | 0.5228 | 0.054 | 0.007 | 1.5849E-13 | 59.51 |
| *HAVCR2* | rs950802 | G | A | 0.3063 | 0.114 | 0.008 | 3.1623E-47 | 203.06 |
| *HAVCR2* | rs77924615 | G | A | 0.1966 | -0.065 | 0.009 | 1.9953E-12 | 52.16 |
| *HAVCR2* | rs186021206 | G | A | 0.0064 | 0.479 | 0.05 | 7.9433E-22 | 91.78 |
| *HBEGF* | rs3811444 | C | T | 0.3323 | -0.061 | 0.008 | 6.3096E-15 | 58.14 |
| *HBEGF* | rs78909033 | G | A | 0.1359 | -0.086 | 0.011 | 1.5849E-15 | 61.12 |
| *HBEGF* | rs540210247 | C | CA | 0.5269 | -0.057 | 0.008 | 1.5849E-13 | 50.77 |
| *HBEGF* | rs1354034 | T | C | 0.6042 | -0.158 | 0.008 | 1.2589E-97 | 390.06 |
| *HBEGF* | rs274555 | C | T | 0.3986 | 0.054 | 0.008 | 1.2589E-12 | 45.56 |
| *HBEGF* | rs4098923 | A | G | 0.4477 | -0.066 | 0.008 | 1.9953E-17 | 68.06 |
| *HBEGF* | rs62396356 | A | G | 0.1344 | 0.085 | 0.011 | 5.0119E-15 | 59.71 |
| *HBEGF* | rs342299 | C | T | 0.455 | -0.052 | 0.007 | 3.9811E-12 | 55.18 |
| *HBEGF* | rs447124 | C | T | 0.4942 | 0.055 | 0.007 | 7.9433E-14 | 61.73 |
| *HBEGF* | rs10820606 | A | C | 0.2291 | -0.115 | 0.009 | 1E-37 | 163.27 |
| *HBEGF* | rs467369 | C | T | 0.5272 | 0.063 | 0.007 | 2.5119E-17 | 81.00 |
| *HBEGF* | rs10761737 | T | C | 0.4128 | 0.174 | 0.008 | 3.162E-117 | 473.06 |
| *HBEGF* | rs17490626 | G | C | 0.1297 | 0.078 | 0.011 | 1.5849E-12 | 50.28 |
| *HBEGF* | rs71474568 | T | TTG | 0.34 | 0.057 | 0.008 | 3.1623E-13 | 50.77 |

| | | | | | | | | |
|---|---|---|---|---|---|---|---|---|
| *HBEGF* | rs489298 | C | T | 0.3953 | -0.052 | 0.008 | 6.3096E-12 | 42.25 |
| *HBEGF* | rs1716505 | C | G | 0.316 | 0.056 | 0.008 | 3.1623E-12 | 49.00 |
| *HBEGF* | rs11553699 | A | G | 0.1351 | 0.164 | 0.011 | 7.9433E-48 | 222.28 |
| *HBEGF* | rs12445050 | C | T | 0.1383 | 0.075 | 0.011 | 1.9953E-12 | 46.49 |
| *HBEGF* | rs516051 | A | C | 0.5188 | -0.068 | 0.007 | 3.1623E-20 | 94.37 |
| *HBEGF* | rs58635160 | CTT | C | 0.4963 | -0.056 | 0.008 | 3.1623E-13 | 49.00 |
| *HBEGF* | rs1654425 | T | C | 0.8343 | 0.15 | 0.01 | 7.9433E-51 | 225.00 |
| *HBEGF* | rs3790176 | G | A | 0.3423 | 0.058 | 0.008 | 1.2589E-13 | 52.56 |
| *HBEGF* | rs116905185 | T | C | 0.0108 | 0.302 | 0.036 | 3.1623E-17 | 70.37 |
| *HBQ1* | rs11759553 | A | T | 0.272 | -0.055 | 0.008 | 3.1623E-12 | 47.27 |
| *HBQ1* | rs556207545 | C | CAAAAAAA | 0.8105 | 0.472 | 0.009 | 1E-200 | 2750.42 |
| *HBZ* | rs17625587 | G | A | 0.2614 | 0.044 | 0.006 | 1.5849E-13 | 53.78 |
| *HBZ* | rs7606173 | G | C | 0.4318 | -0.136 | 0.005 | 3.981E-143 | 739.84 |
| *HBZ* | rs9992667 | C | T | 0.216 | 0.061 | 0.006 | 6.3096E-22 | 103.36 |
| *HBZ* | rs592423 | A | C | 0.5532 | -0.063 | 0.005 | 5.0119E-33 | 158.76 |
| *HBZ* | rs2461286 | A | G | 0.6485 | -1.031 | 0.008 | 1E-200 | 16608.77 |
| *HBZ* | rs9909979 | A | C | 0.2023 | 0.06 | 0.007 | 1.9953E-19 | 73.47 |
| *HBZ* | rs56397034 | G | C | 0.3908 | -0.06 | 0.005 | 7.9433E-29 | 144.00 |
| *HCG22* | rs2905757 | C | T | 0.323 | 0.397 | 0.008 | 1E-200 | 2462.64 |
| *HCLS1* | rs1354034 | T | C | 0.6041 | 0.052 | 0.008 | 3.9811E-12 | 42.25 |
| *HCLS1* | rs3732410 | T | C | 0.2251 | 0.095 | 0.009 | 1E-26 | 111.42 |
| *HCLS1* | rs11170874 | A | G | 0.3307 | 0.056 | 0.008 | 1.9953E-12 | 49.00 |
| *HCLS1* | rs34436714 | C | A | 0.2091 | 0.108 | 0.009 | 1.9953E-32 | 144.00 |
| *HDAC8* | rs28672722 | G | T | 0.2224 | 0.081 | 0.009 | 2.5119E-18 | 81.00 |
| *HDDC2* | rs73581685 | C | T | 0.1804 | -0.874 | 0.01 | 1E-200 | 7638.76 |
| *HDGF* | rs3806417 | C | A | 0.3346 | 1.146 | 0.008 | 1E-200 | 20520.56 |
| *HDGF* | rs6993770 | A | T | 0.2863 | -0.051 | 0.005 | 1.5849E-21 | 104.04 |
| *HDGFL2* | rs1354034 | T | C | 0.6043 | 0.117 | 0.008 | 1.2589E-53 | 213.89 |
| *HDGFL2* | rs6993770 | A | T | 0.2865 | -0.059 | 0.008 | 7.9433E-13 | 54.39 |
| *HEBP1* | rs1941 | C | G | 0.0377 | -0.32 | 0.02 | 7.9433E-57 | 256.00 |
| *HEG1* | rs6438868 | C | G | 0.6173 | 0.109 | 0.007 | 6.3096E-51 | 242.47 |
| *HEG1* | rs9272779 | T | C | 0.4042 | -0.062 | 0.007 | 5.0119E-18 | 78.45 |
| *HEG1* | rs2519093 | C | T | 0.1846 | -0.429 | 0.009 | 1E-200 | 2272.11 |
| *HEG1* | rs964184 | G | C | 0.8667 | -0.084 | 0.01 | 6.3096E-16 | 70.56 |
| *HEG1* | rs34434834 | G | A | 0.0333 | 0.274 | 0.02 | 1.2589E-43 | 187.69 |
| *HEG1* | rs11057606 | G | A | 0.564 | 0.085 | 0.007 | 6.3096E-33 | 147.45 |
| *HEG1* | rs6602911 | C | T | 0.3547 | 0.07 | 0.007 | 1.2589E-21 | 100.00 |
| *HEG1* | rs186021206 | G | A | 0.0064 | 0.83 | 0.049 | 1.5849E-65 | 286.92 |
| *HEPACAM2* | rs304303 | T | G | 0.7305 | -0.069 | 0.007 | 5.0119E-21 | 97.16 |
| *HEPACAM2* | rs7548219 | A | G | 0.2328 | -0.074 | 0.008 | 7.9433E-22 | 85.56 |
| *HEPACAM2* | rs1260326 | T | C | 0.6083 | 0.053 | 0.007 | 3.9811E-15 | 57.33 |
| *HEPACAM2* | rs4672497 | C | G | 0.2218 | -0.209 | 0.008 | 2.512E-152 | 682.52 |
| *HEPACAM2* | rs7560701 | G | C | 0.4989 | -0.075 | 0.007 | 1.9953E-29 | 114.80 |
| *HEPACAM2* | rs3864106 | G | C | 0.5838 | -0.503 | 0.007 | 1E-200 | 5163.45 |
| *HEPACAM2* | rs13107325 | C | T | 0.0749 | 0.104 | 0.012 | 7.9433E-17 | 75.11 |
| *HEPACAM2* | rs2479016 | A | G | 0.4271 | -0.055 | 0.007 | 3.9811E-16 | 61.73 |
| *HEPACAM2* | rs884366 | G | A | 0.3101 | -0.058 | 0.007 | 1.9953E-16 | 68.65 |
| *HEPACAM2* | rs9402685 | T | C | 0.2614 | 0.073 | 0.008 | 3.1623E-22 | 83.27 |
| *HEPACAM2* | rs592423 | A | C | 0.5531 | 0.117 | 0.007 | 1E-69 | 279.37 |
| *HEPACAM2* | rs6796 | T | C | 0.2791 | 0.053 | 0.007 | 6.3096E-13 | 57.33 |
| *HEPACAM2* | rs6592965 | G | A | 0.4545 | -0.133 | 0.007 | 7.9433E-90 | 361.00 |
| *HEPACAM2* | rs183332045 | G | T | 0.0073 | -0.318 | 0.04 | 7.9433E-16 | 63.20 |
| *HEPACAM2* | rs2519093 | C | T | 0.1844 | -0.42 | 0.009 | 1E-200 | 2177.78 |
| *HEPACAM2* | rs17476364 | T | C | 0.1099 | 0.072 | 0.011 | 7.9433E-12 | 42.84 |
| *HEPACAM2* | rs67372909 | T | C | 0.2407 | 0.081 | 0.008 | 3.9811E-26 | 102.52 |
| *HEPACAM2* | rs10891245 | T | G | 0.2962 | -0.054 | 0.007 | 3.9811E-14 | 59.51 |
| *HEPACAM2* | rs3967200 | C | T | 0.1345 | -0.089 | 0.01 | 2.5119E-20 | 79.21 |
| *HEPACAM2* | rs552960 | T | C | 0.493 | 0.048 | 0.007 | 3.1623E-13 | 47.02 |
| *HEPACAM2* | rs186021206 | G | A | 0.0065 | 1.238 | 0.045 | 7.943E-164 | 756.86 |
| *HEPACAM2* | rs72835417 | G | A | 0.1098 | 0.238 | 0.011 | 5.012E-112 | 468.13 |
| *HEPACAM2* | rs8887 | T | C | 0.5736 | -0.081 | 0.007 | 2.5119E-33 | 133.90 |
| *HEPACAM2* | rs485186 | A | G | 0.5376 | 0.115 | 0.007 | 3.9811E-68 | 269.90 |
| *HEPACAM2* | rs746064 | G | A | 0.3802 | 0.232 | 0.007 | 1E-200 | 1098.45 |
| *HEPH* | rs13107325 | C | T | 0.0751 | 0.181 | 0.014 | 1.9953E-38 | 167.15 |
| *HEPH* | rs782134971 | GAAACTGC( | G | 0.2546 | -0.25 | 0.009 | 1.995E-189 | 771.60 |
| *HEPH* | rs186021206 | G | A | 0.0064 | 0.667 | 0.05 | 7.9433E-40 | 177.96 |
| *HEPH* | rs681343 | C | T | 0.5092 | -0.18 | 0.007 | 1E-129 | 661.22 |
| *HEPH* | rs928873 | C | T | 0.2868 | -0.079 | 0.008 | 3.1623E-22 | 97.52 |

| | | | | | | | | |
|---|---|---|---|---|---|---|---|---|
| *HEXIM1* | rs1354034 | T | C | 0.6043 | 0.136 | 0.008 | 2.5119E-72 | 289.00 |
| *HEXIM1* | rs114694170 | T | C | 0.0598 | 0.124 | 0.016 | 2.5119E-15 | 60.06 |
| *HEXIM1* | rs7080386 | C | A | 0.4121 | 0.057 | 0.008 | 1.9953E-14 | 50.77 |
| *HEXIM1* | rs11502185 | T | C | 0.2598 | 0.082 | 0.009 | 3.1623E-18 | 83.01 |
| *HEXIM1* | rs60822569 | T | TTTCTC | 0.5547 | 0.062 | 0.007 | 1.2589E-16 | 78.45 |
| *HEXIM1* | rs36160143 | C | T | 0.1975 | 0.084 | 0.009 | 6.3096E-19 | 87.11 |
| *HEXIM1* | rs1671152 | T | G | 0.8405 | 0.069 | 0.01 | 7.9433E-12 | 47.61 |
| *HGF* | rs59950280 | G | A | 0.3296 | 0.139 | 0.008 | 6.3096E-70 | 301.89 |
| *HGF* | rs5745687 | C | T | 0.0652 | -0.185 | 0.015 | 2.5119E-36 | 152.11 |
| *HGF* | rs2511241 | C | T | 0.9294 | 0.151 | 0.014 | 1.2589E-26 | 116.33 |
| *HGF* | rs7310615 | C | G | 0.5177 | -0.058 | 0.007 | 1.9953E-15 | 68.65 |
| *HGF* | rs704 | G | A | 0.4733 | -0.059 | 0.007 | 3.9811E-16 | 71.04 |
| *HGFAC* | rs11127048 | G | A | 0.6182 | -0.048 | 0.007 | 6.3096E-12 | 47.02 |
| *HGFAC* | rs1047891 | C | A | 0.3132 | 0.096 | 0.007 | 3.1623E-40 | 188.08 |
| *HGFAC* | rs4690097 | C | T | 0.6845 | -0.483 | 0.008 | 1E-200 | 3645.14 |
| *HGFAC* | rs4861708 | G | A | 0.5118 | 0.062 | 0.008 | 2.5119E-16 | 60.06 |
| *HGFAC* | rs2731672 | T | C | 0.7439 | 0.061 | 0.008 | 6.3096E-16 | 58.14 |
| *HGFAC* | rs4895864 | C | T | 0.5746 | 0.047 | 0.007 | 3.1623E-12 | 45.08 |
| *HGFAC* | rs9987289 | A | G | 0.9081 | 0.15 | 0.012 | 3.1623E-38 | 156.25 |
| *HGFAC* | rs28929474 | C | T | 0.021 | 0.214 | 0.023 | 5.0119E-20 | 86.57 |
| *HGFAC* | rs7212366 | C | T | 0.4916 | 0.062 | 0.007 | 1.2589E-17 | 78.45 |
| *HGS* | rs3811444 | C | T | 0.3326 | -0.062 | 0.008 | 1.2589E-14 | 60.06 |
| *HGS* | rs1354034 | T | C | 0.6038 | 0.058 | 0.008 | 6.3096E-14 | 52.56 |
| *HGS* | rs342293 | C | G | 0.4596 | -0.053 | 0.008 | 3.9811E-12 | 43.89 |
| *HHEX* | rs35457250 | C | T | 0.0089 | 0.271 | 0.04 | 1E-11 | 45.90 |
| *HHEX* | rs6993770 | A | T | 0.2865 | -0.06 | 0.008 | 3.9811E-13 | 56.25 |
| *HHEX* | rs200309755 | C | T | 0.5369 | 0.053 | 0.008 | 1.2589E-11 | 43.89 |
| *HHEX* | rs8178824 | C | T | 0.0299 | 0.204 | 0.022 | 1.9953E-20 | 85.98 |
| *HIP1R* | rs10847864 | G | T | 0.3591 | 0.095 | 0.008 | 6.3096E-35 | 141.02 |
| *HJV* | rs12740374 | G | T | 0.2212 | -0.189 | 0.009 | 6.3096E-95 | 441.00 |
| *HJV* | rs41302089 | G | T | 0.0159 | -0.611 | 0.031 | 5.0119E-88 | 388.47 |
| *HJV* | rs2954021 | A | G | 0.5038 | -0.073 | 0.008 | 1E-21 | 83.27 |
| *HJV* | rs77542162 | A | G | 0.023 | 0.201 | 0.025 | 1.5849E-15 | 64.64 |
| *HLA-A* | rs2041385 | C | T | 0.2831 | -0.044 | 0.005 | 1.2589E-19 | 77.44 |
| *HLA-A* | rs7137828 | C | T | 0.5161 | -0.04 | 0.004 | 6.3096E-20 | 100.00 |
| *HLA-A* | rs74439742 | C | T | 0.1719 | -0.063 | 0.006 | 7.9433E-28 | 110.25 |
| *HLA-DRA* | rs36096565 | A | G | 0.2122 | 0.681 | 0.01 | 1E-200 | 4637.61 |
| *HLA-DRA* | rs76904798 | C | T | 0.1416 | 0.088 | 0.009 | 1.2589E-20 | 95.60 |
| *HLA-DRA* | rs3184504 | T | C | 0.5174 | -0.064 | 0.007 | 1E-22 | 83.59 |
| *HLA-E* | rs2927608 | G | A | 0.4494 | -0.188 | 0.007 | 1.995E-155 | 721.31 |
| *HLA-E* | rs2523594 | C | T | 0.6217 | -0.321 | 0.008 | 1E-200 | 1610.02 |
| *HLA-E* | rs3184504 | T | C | 0.5174 | -0.091 | 0.007 | 5.0119E-39 | 169.00 |
| *HLA-E* | rs74439742 | C | T | 0.172 | -0.093 | 0.009 | 7.9433E-24 | 106.78 |
| *HMBS* | rs72997349 | T | C | 0.0309 | 0.38 | 0.021 | 2.5119E-73 | 327.44 |
| *HMCN2* | rs6415851 | A | G | 0.5119 | 0.624 | 0.008 | 1E-200 | 6084.00 |
| *HMCN2* | rs56278466 | T | G | 0.6611 | 0.082 | 0.007 | 5.0119E-32 | 137.22 |
| *HMCN2* | rs35887873 | T | C | 0.2278 | 0.086 | 0.008 | 3.1623E-28 | 115.56 |
| *HMCN2* | rs34042070 | C | G | 0.1853 | -0.108 | 0.009 | 1.5849E-36 | 144.00 |
| *HMCN2* | rs200489612 | G | A | 0.0052 | 0.448 | 0.049 | 1.5849E-19 | 83.59 |
| *HMOX1* | rs371562288 | CT | C | 0.6276 | 0.057 | 0.008 | 1.2589E-12 | 50.77 |
| *HMOX1* | rs385076 | T | C | 0.6443 | 0.17 | 0.008 | 3.162E-107 | 451.56 |
| *HMOX1* | rs6786207 | G | T | 0.6505 | -0.078 | 0.008 | 5.0119E-24 | 95.06 |
| *HMOX1* | rs17229943 | A | C | 0.0505 | 0.171 | 0.017 | 1E-24 | 101.18 |
| *HMOX1* | rs13167071 | G | C | 0.6421 | -0.057 | 0.008 | 1.2589E-13 | 50.77 |
| *HMOX1* | rs2394941 | T | A | 0.2689 | -0.065 | 0.008 | 6.3096E-15 | 66.02 |
| *HMOX1* | rs2637678 | T | C | 0.4086 | 0.073 | 0.007 | 1.2589E-22 | 108.76 |
| *HMOX1* | rs17476364 | T | C | 0.1102 | 0.085 | 0.012 | 6.3096E-13 | 50.17 |
| *HMOX1* | rs7080536 | G | A | 0.0436 | 0.356 | 0.018 | 1.9953E-84 | 391.16 |
| *HMOX1* | rs10846744 | G | C | 0.1421 | 0.091 | 0.01 | 3.1623E-18 | 82.81 |
| *HMOX1* | rs796677417 | GA | G | 0.355 | 0.063 | 0.008 | 3.9811E-16 | 62.02 |
| *HMOX1* | rs12149545 | G | A | 0.316 | 0.069 | 0.008 | 1.2589E-18 | 74.39 |
| *HMOX1* | rs34931250 | C | T | 0.0603 | 0.122 | 0.015 | 3.1623E-15 | 66.15 |
| *HMOX1* | rs1065853 | G | T | 0.0816 | 0.188 | 0.013 | 3.1623E-44 | 209.14 |
| *HMOX1* | rs34632751 | T | C | 0.0269 | 0.324 | 0.023 | 1.5849E-46 | 198.44 |
| *HMOX1* | rs112229561 | C | T | 0.1199 | -0.091 | 0.011 | 1.5849E-15 | 68.44 |
| *HMOX1* | rs113460913 | T | TGA | 0.1537 | -0.2 | 0.01 | 2.5119E-85 | 400.00 |
| *HMOX1* | rs2071747 | G | C | 0.0584 | -0.155 | 0.016 | 6.3096E-23 | 93.85 |
| *HMOX2* | rs4835265 | C | A | 0.1562 | 0.101 | 0.01 | 3.9811E-22 | 102.01 |

| | | | | | | | | |
|---|---|---|---|---|---|---|---|---|
| *HMOX2* | rs4785965 | G | A | 0.7346 | -0.121 | 0.009 | 7.9433E-45 | 180.75 |
| *HMOX2* | rs4503880 | T | C | 0.7993 | -0.087 | 0.009 | 5.0119E-20 | 93.44 |
| *HMOX2* | rs429358 | T | C | 0.1554 | 0.107 | 0.01 | 1.9953E-24 | 114.49 |
| *HNMT* | rs11558538 | C | T | 0.1082 | -1.117 | 0.012 | 1E-200 | 8664.51 |
| *HNMT* | rs7032525 | T | C | 0.2002 | -0.06 | 0.008 | 7.9433E-14 | 56.25 |
| *HNMT* | rs10418046 | T | G | 0.2112 | 0.06 | 0.008 | 1.5849E-14 | 56.25 |
| *HNMT* | rs738409 | C | G | 0.2172 | 0.067 | 0.008 | 2.5119E-18 | 70.14 |
| *HNRNPK* | rs6993770 | A | T | 0.2867 | -0.066 | 0.008 | 3.1623E-15 | 68.06 |
| *HNRNPUL1* | rs1354034 | T | C | 0.6043 | 0.123 | 0.007 | 1.5849E-60 | 308.76 |
| *HNRNPUL1* | rs11604127 | C | T | 0.2308 | 0.075 | 0.009 | 1.2589E-17 | 69.44 |
| *HNRNPUL1* | rs10774624 | G | A | 0.5134 | -0.063 | 0.007 | 7.9433E-18 | 81.00 |
| *HPCAL1* | rs375285744 | T | C | 0.149 | 0.219 | 0.011 | 1E-94 | 396.37 |
| *HPGDS* | rs763228358 | AG | A | 0.2455 | -0.063 | 0.009 | 3.1623E-13 | 49.00 |
| *HPGDS* | rs2865353 | T | C | 0.6119 | 0.463 | 0.008 | 1E-200 | 3349.52 |
| *HPGDS* | rs2398827 | T | C | 0.6016 | 0.066 | 0.007 | 1E-19 | 88.90 |
| *HPSE* | rs7618405 | C | A | 0.2047 | -0.074 | 0.009 | 3.9811E-15 | 67.60 |
| *HPSE* | rs61751211 | T | C | 0.0107 | -0.805 | 0.04 | 1.5849E-88 | 405.02 |
| *HPSE* | rs755492124 | TAAAG | T | 0.3938 | -0.053 | 0.008 | 1E-11 | 43.89 |
| *HPSE* | rs3734208 | C | G | 0.6428 | -0.073 | 0.008 | 3.1623E-20 | 83.27 |
| *HPSE* | rs6961069 | C | T | 0.4025 | 0.058 | 0.008 | 7.9433E-14 | 52.56 |
| *HPSE* | rs6993770 | A | T | 0.2866 | -0.158 | 0.008 | 3.1623E-79 | 390.06 |
| *HPSE* | rs149871778 | G | GAAC | 0.1048 | 0.178 | 0.012 | 6.3096E-47 | 220.03 |
| *HPSE* | rs15951 | G | A | 0.5339 | -0.058 | 0.008 | 1.9953E-14 | 52.56 |
| *HPSE* | rs892090 | T | G | 0.8343 | 0.124 | 0.01 | 3.9811E-34 | 153.76 |
| *HPSE* | rs3827978 | C | T | 0.3533 | 0.065 | 0.008 | 1.5849E-16 | 66.02 |
| *HRC* | rs1260326 | T | C | 0.6084 | 0.065 | 0.008 | 1E-16 | 66.02 |
| *HRC* | rs9853056 | T | C | 0.5594 | 0.086 | 0.008 | 5.0119E-29 | 115.56 |
| *HRC* | rs163192 | C | A | 0.4637 | 0.084 | 0.008 | 5.0119E-28 | 110.25 |
| *HRC* | rs28361032 | A | G | 0.3905 | -0.057 | 0.008 | 7.9433E-12 | 50.77 |
| *HRC* | rs71475909 | A | G | 0.3573 | 0.057 | 0.008 | 1E-12 | 50.77 |
| *HRC* | rs148966785 | G | A | 0.0083 | -0.68 | 0.042 | 5.0119E-59 | 262.13 |
| *HRG* | rs12406245 | G | A | 0.6323 | 0.058 | 0.007 | 1.5849E-15 | 68.65 |
| *HRG* | rs1260326 | T | C | 0.6082 | -0.051 | 0.007 | 1.5849E-12 | 53.08 |
| *HRG* | rs9898 | C | T | 0.329 | 0.501 | 0.008 | 1E-200 | 3921.89 |
| *HRG* | rs59950280 | G | A | 0.3302 | 0.052 | 0.008 | 5.0119E-12 | 42.25 |
| *HRG* | rs4934470 | T | C | 0.7262 | -0.119 | 0.008 | 1E-51 | 221.27 |
| *HS1BP3* | rs77551133 | G | A | 0.0067 | -0.866 | 0.047 | 6.3096E-77 | 339.50 |
| *HS1BP3* | rs11502185 | T | C | 0.2596 | 0.073 | 0.01 | 3.9811E-14 | 53.29 |
| *HS3ST3B1* | rs4970836 | G | A | 0.773 | 0.074 | 0.009 | 1.5849E-17 | 67.60 |
| *HS3ST3B1* | rs4846914 | G | A | 0.6064 | 0.14 | 0.007 | 6.3096E-80 | 400.00 |
| *HS3ST3B1* | rs11096641 | G | A | 0.4713 | -0.057 | 0.007 | 5.0119E-15 | 66.31 |
| *HS3ST3B1* | rs5030062 | A | C | 0.3729 | -0.064 | 0.007 | 1.2589E-17 | 83.59 |
| *HS3ST3B1* | rs13107325 | C | T | 0.0748 | -0.107 | 0.014 | 1E-14 | 58.41 |
| *HS3ST3B1* | rs4241819 | C | T | 0.5093 | -0.092 | 0.007 | 3.1623E-37 | 172.73 |
| *HS3ST3B1* | rs1801020 | A | G | 0.7451 | -0.064 | 0.008 | 1.5849E-14 | 64.00 |
| *HS3ST3B1* | rs28929474 | C | T | 0.0207 | 0.229 | 0.025 | 1.5849E-19 | 83.91 |
| *HS3ST3B1* | rs9302635 | T | C | 0.1824 | 0.07 | 0.009 | 6.3096E-14 | 60.49 |
| *HS3ST3B1* | rs17669311 | G | A | 0.3921 | 0.329 | 0.008 | 1E-200 | 1691.27 |
| *HS6ST1* | rs12740374 | G | T | 0.2209 | -0.075 | 0.009 | 1.5849E-17 | 69.44 |
| *HS6ST1* | rs907866 | G | A | 0.4478 | -0.076 | 0.007 | 2.5119E-24 | 117.88 |
| *HS6ST1* | rs6761320 | C | T | 0.4284 | -0.21 | 0.008 | 5.012E-172 | 689.06 |
| *HS6ST1* | rs17282078 | T | A | 0.13 | -0.096 | 0.011 | 6.3096E-19 | 76.17 |
| *HS6ST1* | rs13107325 | C | T | 0.0748 | 0.17 | 0.014 | 1.2589E-34 | 147.45 |
| *HS6ST1* | rs1290147 | G | C | 0.6485 | -0.052 | 0.008 | 7.9433E-12 | 42.25 |
| *HS6ST1* | rs573098 | A | T | 0.6745 | 0.064 | 0.008 | 3.9811E-16 | 64.00 |
| *HS6ST1* | rs33951980 | C | T | 0.1288 | -0.078 | 0.011 | 5.0119E-13 | 50.28 |
| *HS6ST1* | rs6952090 | T | C | 0.3483 | -0.061 | 0.008 | 1.5849E-15 | 58.14 |
| *HS6ST1* | rs56278466 | T | G | 0.6612 | 0.068 | 0.008 | 1E-18 | 72.25 |
| *HS6ST1* | rs1149597 | C | T | 0.1692 | -0.079 | 0.01 | 3.1623E-16 | 62.41 |
| *HS6ST1* | rs770578737 | GGA | G | 0.7871 | -0.061 | 0.009 | 1.2589E-11 | 45.94 |
| *HS6ST1* | rs35350651 | A | AC | 0.5033 | -0.054 | 0.007 | 1E-13 | 59.51 |
| *HS6ST1* | rs12903915 | T | C | 0.3411 | -0.064 | 0.008 | 6.3096E-17 | 64.00 |
| *HS6ST1* | rs72802342 | C | A | 0.0759 | -0.19 | 0.014 | 3.9811E-42 | 184.18 |
| *HS6ST1* | rs704 | G | A | 0.4729 | -0.152 | 0.007 | 2.5119E-96 | 471.51 |
| *HS6ST1* | rs77542162 | A | G | 0.0229 | 0.296 | 0.024 | 7.9433E-34 | 152.11 |
| *HSBP1* | rs12038396 | C | T | 0.6528 | 0.062 | 0.007 | 2.5119E-17 | 78.45 |
| *HSBP1* | rs3811444 | C | T | 0.3327 | -0.078 | 0.007 | 2.5119E-26 | 124.16 |
| *HSBP1* | rs72799293 | G | A | 0.0239 | 0.278 | 0.023 | 3.1623E-34 | 146.09 |

| | | | | | | | | |
|---|---|---|---|---|---|---|---|---|
| *HSBP1* | rs847803 | A | G | 0.6764 | -0.056 | 0.007 | 3.1623E-14 | 64.00 |
| *HSBP1* | rs13084580 | C | T | 0.1136 | -0.148 | 0.011 | 1.2589E-41 | 181.02 |
| *HSBP1* | rs1354034 | T | C | 0.6045 | 0.056 | 0.007 | 1.9953E-15 | 64.00 |
| *HSBP1* | rs114694170 | T | C | 0.0599 | -0.127 | 0.015 | 1E-17 | 71.68 |
| *HSBP1* | rs6938130 | A | T | 0.4077 | -0.048 | 0.007 | 1E-11 | 47.02 |
| *HSBP1* | rs753413362 | C | CA | 0.209 | 0.072 | 0.009 | 2.5119E-16 | 64.00 |
| *HSBP1* | rs342293 | C | G | 0.4595 | -0.086 | 0.007 | 3.1623E-35 | 150.94 |
| *HSBP1* | rs7832219 | T | C | 0.2739 | 0.056 | 0.008 | 6.3096E-13 | 49.00 |
| *HSBP1* | rs10820606 | A | C | 0.2288 | 0.176 | 0.008 | 5.0119E-97 | 484.00 |
| *HSBP1* | rs61916664 | A | G | 0.5307 | -0.049 | 0.007 | 7.9433E-12 | 49.00 |
| *HSBP1* | rs12321467 | C | T | 0.3638 | -0.059 | 0.008 | 1E-14 | 54.39 |
| *HSBP1* | rs60822569 | T | TTTCTC | 0.553 | 0.09 | 0.007 | 7.9433E-38 | 165.31 |
| *HSBP1* | rs707236 | T | A | 0.3454 | -0.568 | 0.008 | 1E-200 | 5041.00 |
| *HSBP1* | rs767170860 | CA | C | 0.5021 | 0.056 | 0.007 | 1.5849E-14 | 64.00 |
| *HSBP1* | rs9951002 | T | A | 0.2329 | 0.075 | 0.008 | 1E-19 | 87.89 |
| *HSD11B1* | rs78444298 | G | A | 0.0194 | 0.249 | 0.027 | 7.9433E-20 | 85.05 |
| *HSD11B1* | rs7542235 | A | G | 0.1948 | 0.156 | 0.009 | 5.0119E-62 | 300.44 |
| *HSD11B1* | rs4240624 | G | A | 0.9079 | 0.169 | 0.013 | 2.5119E-39 | 169.00 |
| *HSD11B1* | rs174576 | C | A | 0.3523 | 0.075 | 0.008 | 6.3096E-22 | 87.89 |
| *HSD11B1* | rs695110 | T | C | 0.1542 | 0.078 | 0.01 | 5.0119E-14 | 60.84 |
| *HSD11B1* | rs1076485 | C | T | 0.1236 | 0.109 | 0.011 | 1E-21 | 98.19 |
| *HSD11B1* | rs28929474 | C | T | 0.0207 | 0.656 | 0.026 | 2.512E-137 | 636.59 |
| *HSD11B1* | rs56156922 | T | C | 0.3249 | 0.054 | 0.008 | 1.5849E-11 | 45.56 |
| *HSD11B1* | rs2532351 | C | G | 0.4207 | 0.054 | 0.008 | 3.9811E-12 | 45.56 |
| *HSD11B1* | rs1801689 | A | C | 0.0301 | 0.444 | 0.022 | 7.9433E-92 | 407.31 |
| *HSD11B1* | rs58542926 | C | T | 0.0752 | 0.228 | 0.014 | 6.3096E-58 | 265.22 |
| *HSD11B1* | rs141887102 | A | C | 0.1826 | 0.087 | 0.01 | 3.1623E-19 | 75.69 |
| *HSD17B14* | rs55730499 | C | T | 0.0822 | 0.086 | 0.011 | 7.9433E-16 | 61.12 |
| *HSD17B14* | rs112875651 | G | A | 0.3912 | -0.061 | 0.006 | 7.9433E-24 | 103.36 |
| *HSD17B14* | rs35299026 | G | A | 0.0471 | -1.523 | 0.018 | 1E-200 | 7159.04 |
| *HSDL2* | rs62180180 | C | T | 0.3052 | -0.074 | 0.007 | 1.5849E-23 | 111.76 |
| *HSDL2* | rs10817342 | C | T | 0.3801 | 0.601 | 0.008 | 1E-200 | 5643.77 |
| *HSPA1A* | rs1354034 | T | C | 0.6041 | 0.052 | 0.008 | 7.9433E-12 | 42.25 |
| *HSPA1A* | rs181412562 | C | T | 0.0018 | -1.266 | 0.089 | 3.1623E-46 | 202.34 |
| *HSPA1A* | rs342293 | C | G | 0.4593 | -0.056 | 0.007 | 3.9811E-14 | 64.00 |
| *HSPA1A* | rs11218968 | A | G | 0.0419 | -0.147 | 0.019 | 2.5119E-15 | 59.86 |
| *HSPA2* | rs10498515 | C | T | 0.799 | 0.127 | 0.009 | 3.9811E-42 | 199.12 |
| *HSPB1* | rs1354034 | T | C | 0.6037 | 0.052 | 0.007 | 6.3096E-13 | 55.18 |
| *HSPB1* | rs6926219 | G | A | 0.5458 | -0.092 | 0.007 | 1E-37 | 172.73 |
| *HSPB1* | rs111642997 | C | CTTTTA | 0.2448 | -0.553 | 0.009 | 1E-200 | 3775.42 |
| *HSPB1* | rs342298 | C | T | 0.455 | -0.077 | 0.008 | 3.9811E-24 | 92.64 |
| *HSPB1* | rs7896518 | A | G | 0.4236 | 0.078 | 0.007 | 2.5119E-26 | 124.16 |
| *HSPB1* | rs11553699 | A | G | 0.1357 | -0.08 | 0.011 | 1.9953E-13 | 52.89 |
| *HSPB6* | rs12463674 | A | G | 0.3155 | 0.058 | 0.007 | 5.0119E-16 | 68.65 |
| *HSPB6* | rs12331051 | C | T | 0.428 | 0.059 | 0.007 | 5.0119E-18 | 71.04 |
| *HSPB6* | rs151828 | T | C | 0.5475 | 0.048 | 0.007 | 1.9953E-13 | 47.02 |
| *HSPB6* | rs13437444 | C | T | 0.2153 | 0.062 | 0.008 | 3.9811E-15 | 60.06 |
| *HSPB6* | rs2234962 | T | C | 0.2159 | 0.058 | 0.008 | 3.9811E-13 | 52.56 |
| *HSPB6* | rs1337298 | C | T | 0.4394 | -0.05 | 0.007 | 3.1623E-14 | 51.02 |
| *HSPG2* | rs6684152 | A | G | 0.2938 | -0.175 | 0.008 | 1.995E-103 | 478.52 |
| *HSPG2* | rs13146355 | G | A | 0.4564 | 0.05 | 0.007 | 6.3096E-12 | 51.02 |
| *HSPG2* | rs774259378 | CT | C | 0.3772 | -0.052 | 0.008 | 3.9811E-12 | 42.25 |
| *HSPG2* | rs77924615 | G | A | 0.1969 | -0.078 | 0.009 | 5.0119E-17 | 75.11 |
| *HTRA2* | rs6993770 | A | T | 0.2861 | -0.062 | 0.008 | 7.9433E-14 | 60.06 |
| *HYAL1* | rs12371 | A | G | 0.0878 | -0.09 | 0.013 | 6.3096E-12 | 47.93 |
| *HYAL1* | rs116482870 | C | T | 0.0623 | -0.549 | 0.016 | 1E-200 | 1177.35 |
| *HYAL1* | rs12321232 | G | T | 0.2336 | -0.064 | 0.009 | 2.5119E-13 | 50.57 |
| *HYAL1* | rs142650905 | C | T | 0.0014 | -1.586 | 0.1 | 5.0119E-57 | 251.54 |
| *HYOU1* | rs7532110 | T | A | 0.8034 | 0.09 | 0.009 | 6.3096E-22 | 100.00 |
| *HYOU1* | rs61747728 | C | T | 0.0381 | 0.226 | 0.019 | 5.0119E-32 | 141.48 |
| *HYOU1* | rs59484402 | C | CT | 0.6114 | -0.052 | 0.008 | 1E-11 | 42.25 |
| *HYOU1* | rs75166367 | G | A | 0.0603 | -0.137 | 0.016 | 1.2589E-18 | 73.32 |
| *HYOU1* | rs149547076 | T | C | 0.0107 | -0.33 | 0.036 | 1.2589E-19 | 84.03 |
| *HYOU1* | rs17715183 | G | C | 0.0666 | -0.111 | 0.015 | 1E-13 | 54.76 |
| *HYOU1* | rs2509121 | C | T | 0.4007 | 0.183 | 0.008 | 7.943E-129 | 523.27 |
| *HYOU1* | rs28929474 | C | T | 0.0208 | 0.302 | 0.026 | 1.5849E-31 | 134.92 |
| *HYOU1* | rs11636578 | T | C | 0.3243 | 0.06 | 0.008 | 3.9811E-14 | 56.25 |
| *HYOU1* | rs1801689 | A | C | 0.0302 | 0.288 | 0.022 | 1.2589E-40 | 171.37 |

| | | | | | | | | |
|---|---|---|---|---|---|---|---|---|
| *HYOU1* | rs2659005 | C | T | 0.441 | 0.082 | 0.007 | 1.5849E-28 | 137.22 |
| *HYOU1* | rs33950747 | C | T | 0.075 | 0.111 | 0.014 | 1.9953E-15 | 62.86 |
| *HYOU1* | rs10419198 | C | T | 0.2503 | 0.059 | 0.009 | 3.1623E-12 | 42.98 |
| *HYOU1* | rs136205 | C | G | 0.7483 | -0.061 | 0.009 | 1.5849E-12 | 45.94 |
| *ICA1* | rs1354034 | T | C | 0.6035 | 0.053 | 0.008 | 3.1623E-12 | 43.89 |
| *ICA1* | rs3952 | A | G | 0.3136 | -0.218 | 0.008 | 1.995E-158 | 742.56 |
| *ICAM1* | rs1049728 | G | C | 0.0593 | -0.167 | 0.015 | 2.5119E-29 | 123.95 |
| *ICAM1* | rs34434834 | G | A | 0.0333 | 0.148 | 0.019 | 2.5119E-14 | 60.68 |
| *ICAM1* | rs111338191 | A | AAATT | 0.5206 | -0.082 | 0.007 | 6.3096E-32 | 137.22 |
| *ICAM1* | rs186021206 | G | A | 0.0064 | 0.636 | 0.047 | 6.3096E-41 | 183.11 |
| *ICAM1* | rs5030377 | A | G | 0.378 | 0.415 | 0.008 | 1E-200 | 2691.02 |
| *ICAM1* | rs3136642 | A | G | 0.3675 | -0.056 | 0.008 | 6.3096E-13 | 49.00 |
| *ICAM2* | rs11682865 | G | C | 0.1946 | -0.051 | 0.007 | 5.0119E-13 | 53.08 |
| *ICAM2* | rs10935473 | G | T | 0.4399 | -0.717 | 0.007 | 1E-200 | 10491.61 |
| *ICAM2* | rs9399137 | T | C | 0.2579 | 0.057 | 0.006 | 6.3096E-19 | 90.25 |
| *ICAM2* | rs6993770 | A | T | 0.2865 | -0.064 | 0.006 | 5.0119E-25 | 113.78 |
| *ICAM2* | rs2519093 | C | T | 0.1843 | -0.859 | 0.009 | 1E-200 | 9109.64 |
| *ICAM2* | rs78689694 | G | C | 0.1325 | -0.083 | 0.008 | 1E-23 | 107.64 |
| *ICAM2* | rs111626763 | T | TACTC | 0.5883 | 0.039 | 0.006 | 6.3096E-12 | 42.25 |
| *ICAM2* | rs111338191 | A | AAATT | 0.5209 | -0.079 | 0.006 | 3.9811E-45 | 173.36 |
| *ICAM2* | rs186021206 | G | A | 0.0064 | 1.269 | 0.038 | 1E-200 | 1115.21 |
| *ICAM2* | rs574698129 | CA | C | 0.1879 | 0.101 | 0.007 | 1E-41 | 208.18 |
| *ICAM2* | rs368565 | C | T | 0.4317 | -0.044 | 0.006 | 7.9433E-15 | 53.78 |
| *ICAM2* | rs2837988 | C | A | 0.3616 | 0.049 | 0.006 | 6.3096E-17 | 66.69 |
| *ICAM3* | rs188468174 | C | T | 0.0143 | -0.372 | 0.03 | 1.2589E-34 | 153.76 |
| *ICAM3* | rs778683816 | T | TGC | 0.354 | 0.051 | 0.007 | 6.3096E-12 | 53.08 |
| *ICAM3* | rs2289235 | G | A | 0.4688 | -0.149 | 0.007 | 1.9953E-98 | 453.08 |
| *ICAM3* | rs6764279 | C | A | 0.2775 | -0.088 | 0.008 | 2.5119E-28 | 121.00 |
| *ICAM3* | rs2524078 | A | G | 0.1736 | -0.195 | 0.01 | 6.3096E-92 | 380.25 |
| *ICAM3* | rs7758383 | A | G | 0.4871 | 0.083 | 0.007 | 7.9433E-31 | 140.59 |
| *ICAM3* | rs2278467 | T | C | 0.155 | -0.216 | 0.01 | 3.162E-106 | 466.56 |
| *ICAM3* | rs12574844 | G | A | 0.0706 | -0.156 | 0.014 | 2.5119E-29 | 124.16 |
| *ICAM3* | rs3184504 | T | C | 0.5171 | -0.138 | 0.007 | 1.9953E-84 | 388.65 |
| *ICAM3* | rs494632 | C | T | 0.4739 | -0.058 | 0.007 | 6.3096E-16 | 68.65 |
| *ICAM3* | rs76428106 | T | C | 0.0128 | 0.304 | 0.033 | 1.5849E-20 | 84.86 |
| *ICAM3* | rs9316367 | C | T | 0.2515 | -0.113 | 0.008 | 3.1623E-43 | 199.52 |
| *ICAM3* | rs186021206 | G | A | 0.0064 | 1.198 | 0.049 | 1.995E-131 | 597.75 |
| *ICAM3* | rs34562254 | G | A | 0.097 | 0.088 | 0.012 | 1.9953E-13 | 53.78 |
| *ICAM3* | rs2305482 | A | C | 0.5401 | -0.062 | 0.007 | 3.9811E-18 | 78.45 |
| *ICAM3* | rs2696459 | G | C | 0.2299 | -0.063 | 0.009 | 1.2589E-13 | 49.00 |
| *ICAM3* | rs56378716 | A | G | 0.0134 | 0.298 | 0.031 | 6.3096E-22 | 92.41 |
| *ICAM3* | rs2725405 | G | C | 0.4409 | 0.097 | 0.007 | 1.5849E-41 | 192.02 |
| *ICAM3* | rs28378712 | T | G | 0.3128 | 0.318 | 0.008 | 1E-200 | 1580.06 |
| *ICAM3* | rs602662 | G | A | 0.5362 | 0.053 | 0.007 | 3.9811E-13 | 57.33 |
| *ICAM3* | rs6517655 | G | C | 0.2725 | 0.063 | 0.008 | 3.9811E-15 | 62.02 |
| *ICAM4* | rs139898146 | C | A | 0.4154 | -0.678 | 0.008 | 1E-200 | 7182.56 |
| *ICAM4* | rs11576522 | A | G | 0.6552 | 0.076 | 0.008 | 1E-22 | 90.25 |
| *ICAM4* | rs7606173 | G | C | 0.4323 | -0.065 | 0.006 | 3.1623E-24 | 117.36 |
| *ICAM4* | rs13103731 | C | T | 0.4491 | 0.215 | 0.006 | 1E-200 | 1284.03 |
| *ICAM4* | rs2523990 | A | G | 0.5397 | -0.053 | 0.006 | 1.2589E-16 | 78.03 |
| *ICAM4* | rs62427982 | C | T | 0.3255 | 0.057 | 0.007 | 1.2589E-16 | 66.31 |
| *ICAM4* | rs9376091 | C | T | 0.2591 | -0.095 | 0.007 | 3.1623E-38 | 184.18 |
| *ICAM4* | rs6592965 | G | A | 0.4543 | -0.057 | 0.006 | 1E-18 | 90.25 |
| *ICAM4* | rs1595608 | T | G | 0.2408 | 0.054 | 0.007 | 7.9433E-13 | 59.51 |
| *ICAM4* | rs11627485 | T | C | 0.4494 | 0.048 | 0.006 | 6.3096E-14 | 64.00 |
| *ICAM4* | rs17580 | T | A | 0.0496 | 0.159 | 0.015 | 1.5849E-27 | 112.36 |
| *ICAM4* | rs186021206 | G | A | 0.0064 | 0.515 | 0.044 | 1E-31 | 137.00 |
| *ICAM4* | rs73425119 | G | C | 0.2597 | 0.072 | 0.007 | 6.3096E-23 | 105.80 |
| *ICAM4* | rs5030377 | A | G | 0.3783 | -0.115 | 0.007 | 1.2589E-67 | 269.90 |
| *ICAM4* | rs746064 | G | A | 0.38 | 0.094 | 0.007 | 3.9811E-46 | 180.33 |
| *ICAM5* | rs3864106 | G | C | 0.5841 | -0.045 | 0.006 | 1E-13 | 56.25 |
| *ICAM5* | rs2049865 | C | A | 0.5783 | 0.048 | 0.006 | 2.5119E-15 | 64.00 |
| *ICAM5* | rs2519093 | C | T | 0.1843 | -0.408 | 0.008 | 1E-200 | 2601.00 |
| *ICAM5* | rs186021206 | G | A | 0.0064 | 0.59 | 0.041 | 2.5119E-46 | 207.08 |
| *ICAM5* | rs77542162 | A | G | 0.0226 | 0.523 | 0.02 | 5.012E-147 | 683.82 |
| *ICAM5* | rs281439 | G | C | 0.7799 | 0.844 | 0.009 | 1E-200 | 8794.27 |
| *ICOSLG* | rs74152937 | G | A | 0.0457 | 0.122 | 0.017 | 1.2589E-12 | 51.50 |
| *ICOSLG* | rs186021206 | G | A | 0.0063 | 0.782 | 0.05 | 1.9953E-55 | 244.61 |

| | | | | | | | | |
|---|---|---|---|---|---|---|---|---|
| *ICOSLG* | rs6518352 | A | G | 0.7209 | -0.402 | 0.009 | 1E-200 | 1995.11 |
| *IDI2* | rs1044261 | C | T | 0.0782 | -0.616 | 0.014 | 1E-200 | 1936.00 |
| *IDO1* | rs2637681 | T | G | 0.4125 | 0.073 | 0.007 | 1.2589E-22 | 108.76 |
| *IDO1* | rs12342201 | G | A | 0.4766 | -0.058 | 0.007 | 5.0119E-15 | 68.65 |
| *IDO1* | rs13215699 | G | A | 0.1851 | 0.078 | 0.01 | 3.9811E-15 | 60.84 |
| *IDO1* | rs7010461 | C | T | 0.3341 | 0.276 | 0.008 | 1E-200 | 1190.25 |
| *IDO1* | rs11993233 | A | G | 0.4262 | -0.054 | 0.007 | 5.0119E-13 | 59.51 |
| *IDO1* | rs12363646 | T | C | 0.2914 | -0.06 | 0.008 | 7.9433E-14 | 56.25 |
| *IDO1* | rs597808 | A | G | 0.5158 | -0.124 | 0.007 | 2.5119E-65 | 313.80 |
| *IDO1* | rs4632248 | G | T | 0.2097 | 0.082 | 0.009 | 3.9811E-20 | 83.01 |
| *IDS* | rs529565 | T | C | 0.3175 | -0.057 | 0.008 | 1.5849E-12 | 50.77 |
| *IDS* | rs10128858 | A | G | 0.2879 | -0.21 | 0.008 | 6.31E-140 | 689.06 |
| *IDS* | rs704 | G | A | 0.4733 | -0.087 | 0.007 | 2.5119E-31 | 154.47 |
| *IDUA* | rs687339 | C | T | 0.7712 | 0.058 | 0.008 | 5.0119E-14 | 52.56 |
| *IDUA* | rs3796622 | T | C | 0.6398 | 0.543 | 0.008 | 1E-200 | 4607.02 |
| *IDUA* | rs114816312 | C | T | 0.0078 | 0.288 | 0.042 | 1.2589E-11 | 47.02 |
| *IDUA* | rs2389688 | A | C | 0.4417 | -0.052 | 0.008 | 3.9811E-12 | 42.25 |
| *IDUA* | rs10778152 | A | G | 0.2878 | -0.239 | 0.007 | 1E-200 | 1165.73 |
| *IDUA* | rs2254971 | G | C | 0.3719 | 0.052 | 0.007 | 1.9953E-14 | 55.18 |
| *IDUA* | rs71435622 | A | AT | 0.2869 | 0.054 | 0.007 | 6.3096E-13 | 59.51 |
| *IDUA* | rs28929474 | C | T | 0.0208 | 0.261 | 0.023 | 1.5849E-30 | 128.77 |
| *IDUA* | rs147233090 | C | T | 0.0235 | 0.208 | 0.021 | 3.9811E-22 | 98.10 |
| *IDUA* | rs58542926 | C | T | 0.0754 | 0.115 | 0.012 | 7.9433E-21 | 91.84 |
| *IDUA* | rs429358 | T | C | 0.1555 | -0.077 | 0.009 | 1.2589E-17 | 73.20 |
| *IDUA* | rs1883711 | G | C | 0.0307 | 0.131 | 0.019 | 6.3096E-12 | 47.54 |
| *IFI30* | rs71640036 | G | T | 0.5056 | 0.05 | 0.007 | 1.5849E-12 | 51.02 |
| *IFI30* | rs2394172 | C | T | 0.0665 | 0.111 | 0.014 | 1.9953E-15 | 62.86 |
| *IFI30* | rs3184504 | T | C | 0.5172 | -0.084 | 0.007 | 1.9953E-33 | 144.00 |
| *IFI30* | rs10846742 | G | A | 0.8555 | -0.089 | 0.01 | 5.0119E-19 | 79.21 |
| *IFI30* | rs7259445 | A | C | 0.2184 | -0.507 | 0.009 | 1E-200 | 3173.44 |
| *IFIT3* | rs6753170 | T | C | 0.6276 | 0.07 | 0.008 | 1.2589E-18 | 76.56 |
| *IFIT3* | rs76036957 | G | A | 0.0094 | 0.382 | 0.04 | 1E-21 | 91.20 |
| *IFIT3* | rs55792153 | A | C | 0.7437 | 0.2 | 0.009 | 1.585E-114 | 493.83 |
| *IFIT3* | rs610913 | G | T | 0.6181 | -0.084 | 0.008 | 6.3096E-27 | 110.25 |
| *IFIT3* | rs2274878 | C | G | 0.2461 | -0.108 | 0.009 | 5.0119E-35 | 144.00 |
| *IFNAR1* | rs186021206 | G | A | 0.0065 | 0.515 | 0.049 | 3.9811E-26 | 110.46 |
| *IFNAR1* | rs12720356 | A | C | 0.0986 | -0.211 | 0.012 | 5.0119E-70 | 309.17 |
| *IFNAR1* | rs914142 | G | A | 0.7287 | -0.45 | 0.008 | 1E-200 | 3164.06 |
| *IFNG* | rs76830965 | C | A | 0.1179 | 0.083 | 0.012 | 3.1623E-12 | 47.84 |
| *IFNG* | rs9264259 | T | C | 0.7133 | 0.064 | 0.009 | 1.9953E-12 | 50.57 |
| *IFNGR1* | rs61747728 | C | T | 0.0379 | 0.195 | 0.019 | 1.5849E-24 | 105.33 |
| *IFNGR1* | rs1260326 | T | C | 0.6086 | 0.055 | 0.007 | 2.5119E-13 | 61.73 |
| *IFNGR1* | rs73065135 | G | C | 0.0992 | 0.087 | 0.012 | 1.5849E-12 | 52.56 |
| *IFNGR1* | rs9853056 | T | C | 0.5592 | 0.05 | 0.007 | 1E-11 | 51.02 |
| *IFNGR1* | rs2394177 | A | G | 0.075 | 0.125 | 0.014 | 3.9811E-19 | 79.72 |
| *IFNGR1* | rs9376267 | C | T | 0.2374 | -0.202 | 0.009 | 6.31E-122 | 503.75 |
| *IFNGR1* | rs8176672 | C | T | 0.0613 | 0.36 | 0.015 | 3.981E-123 | 576.00 |
| *IFNGR1* | rs777834943 | C | CT | 0.1833 | -0.081 | 0.01 | 3.1623E-17 | 65.61 |
| *IFNGR1* | rs186021206 | G | A | 0.0064 | 0.848 | 0.05 | 3.9811E-64 | 287.64 |
| *IFNGR1* | rs2659005 | C | T | 0.4407 | 0.055 | 0.007 | 6.3096E-14 | 61.73 |
| *IFNGR2* | rs12740374 | G | T | 0.2212 | -0.075 | 0.006 | 6.3096E-31 | 156.25 |
| *IFNGR2* | rs10901252 | G | C | 0.0614 | 0.127 | 0.011 | 3.1623E-29 | 133.30 |
| *IFNGR2* | rs56278466 | T | G | 0.6614 | 0.072 | 0.006 | 1.2589E-36 | 144.00 |
| *IFNGR2* | rs174564 | A | G | 0.3488 | 0.095 | 0.006 | 7.9433E-64 | 250.69 |
| *IFNGR2* | rs12149545 | G | A | 0.3164 | 0.055 | 0.006 | 2.5119E-21 | 84.03 |
| *IFNGR2* | rs72802342 | C | A | 0.0759 | -0.071 | 0.01 | 1E-11 | 50.41 |
| *IFNGR2* | rs1065853 | G | T | 0.0811 | 0.106 | 0.01 | 1.5849E-26 | 112.36 |
| *IFNGR2* | rs492602 | A | G | 0.5096 | -0.054 | 0.005 | 1.5849E-23 | 116.64 |
| *IFNGR2* | rs9808753 | A | G | 0.1404 | 1.433 | 0.011 | 1E-200 | 16970.98 |
| *IFNL1* | rs35667974 | T | C | 0.0217 | -0.335 | 0.026 | 2.5119E-37 | 166.01 |
| *IFNL1* | rs3775291 | C | T | 0.2968 | -0.092 | 0.008 | 3.1623E-28 | 132.25 |
| *IFNL1* | rs9270747 | A | G | 0.6574 | 0.074 | 0.008 | 2.5119E-19 | 85.56 |
| *IFNL1* | rs4346838 | G | A | 0.5238 | 0.056 | 0.008 | 3.1623E-13 | 49.00 |
| *IFNL1* | rs10424978 | C | A | 0.5999 | -0.059 | 0.008 | 6.3096E-14 | 54.39 |
| *IFNLR1* | rs139958347 | G | C | 0.0396 | -0.942 | 0.021 | 1E-200 | 2012.16 |
| *IFNLR1* | rs10753239 | T | C | 0.4604 | -0.056 | 0.008 | 3.9811E-13 | 49.00 |
| *IFNLR1* | rs1801689 | A | C | 0.0301 | 0.23 | 0.022 | 1.2589E-26 | 109.30 |
| *IGBP1* | rs6993770 | A | T | 0.2864 | -0.063 | 0.008 | 1E-14 | 62.02 |

| | | | | | | | | |
|---|---|---|---|---|---|---|---|---|
| *IGBP1* | rs10733789 | T | C | 0.3128 | 0.055 | 0.008 | 1E-11 | 47.27 |
| *IGDCC4* | rs3184504 | T | C | 0.5174 | 0.05 | 0.007 | 5.0119E-13 | 51.02 |
| *IGDCC4* | rs7147722 | G | T | 0.5386 | 0.054 | 0.007 | 1E-14 | 59.51 |
| *IGDCC4* | rs56362513 | T | C | 0.0203 | 0.964 | 0.026 | 1E-200 | 1374.70 |
| *IGDCC4* | rs77542162 | A | G | 0.0229 | 0.174 | 0.023 | 3.9811E-14 | 57.23 |
| *IGF1R* | rs9271421 | T | C | 0.8549 | -0.12 | 0.01 | 3.9811E-33 | 144.00 |
| *IGF1R* | rs2519093 | C | T | 0.1845 | -0.481 | 0.009 | 1E-200 | 2856.31 |
| *IGF1R* | rs3184504 | T | C | 0.5168 | -0.058 | 0.007 | 1.5849E-16 | 68.65 |
| *IGF1R* | rs35115159 | A | G | 0.203 | 0.13 | 0.009 | 3.1623E-49 | 208.64 |
| *IGF1R* | rs11574938 | G | C | 0.5164 | 0.093 | 0.007 | 5.0119E-40 | 176.51 |
| *IGF1R* | rs200489612 | G | A | 0.0052 | 0.882 | 0.053 | 3.9811E-62 | 276.94 |
| *IGF1R* | rs9916257 | G | T | 0.4394 | -0.065 | 0.007 | 5.0119E-20 | 86.22 |
| *IGF1R* | rs56133626 | G | A | 0.3261 | -0.068 | 0.008 | 1E-19 | 72.25 |
| *IGF1R* | rs760462 | T | C | 0.818 | 0.082 | 0.009 | 1.5849E-19 | 83.01 |
| *IGF2R* | rs6727484 | G | A | 0.1709 | 0.073 | 0.009 | 1.9953E-15 | 65.79 |
| *IGF2R* | rs3131623 | T | A | 0.1836 | 0.111 | 0.01 | 1.2589E-30 | 123.21 |
| *IGF2R* | rs2282140 | C | T | 0.1312 | 0.556 | 0.011 | 1E-200 | 2554.84 |
| *IGF2R* | rs9987289 | A | G | 0.9081 | -0.087 | 0.012 | 3.1623E-13 | 52.56 |
| *IGF2R* | rs72752535 | A | C | 0.005 | 0.422 | 0.054 | 3.9811E-15 | 61.07 |
| *IGF2R* | rs7974486 | A | G | 0.8356 | 0.08 | 0.009 | 1.5849E-17 | 79.01 |
| *IGF2R* | rs11111000 | C | T | 0.0925 | -0.125 | 0.012 | 1.2589E-25 | 108.51 |
| *IGF2R* | rs10144207 | C | T | 0.3541 | -0.056 | 0.007 | 1.2589E-14 | 64.00 |
| *IGF2R* | rs145078947 | G | T | 0.0026 | 0.989 | 0.068 | 1.9953E-47 | 211.53 |
| *IGF2R* | rs186021206 | G | A | 0.0064 | 0.37 | 0.048 | 7.9433E-15 | 59.42 |
| *IGF2R* | rs1838105 | A | G | 0.649 | -0.049 | 0.007 | 1.2589E-11 | 49.00 |
| *IGF2R* | rs1801689 | A | C | 0.0301 | 0.149 | 0.02 | 1.9953E-13 | 55.50 |
| *IGF2R* | rs200378 | T | C | 0.5052 | 0.062 | 0.007 | 5.0119E-19 | 78.45 |
| *IGFBP1* | rs12740374 | G | T | 0.2213 | -0.073 | 0.009 | 1.2589E-16 | 65.79 |
| *IGFBP1* | rs1260326 | T | C | 0.6084 | -0.081 | 0.008 | 6.3096E-27 | 102.52 |
| *IGFBP2* | rs1260326 | T | C | 0.6084 | 0.065 | 0.008 | 1.2589E-17 | 66.02 |
| *IGFBP2* | rs4674100 | G | A | 0.2928 | -0.07 | 0.009 | 3.1623E-16 | 60.49 |
| *IGFBP2* | rs13108218 | A | G | 0.6189 | -0.057 | 0.008 | 6.3096E-14 | 50.77 |
| *IGFBP2* | rs34690971 | A | AT | 0.7441 | 0.061 | 0.008 | 7.9433E-13 | 58.14 |
| *IGFBP3* | rs4234798 | T | G | 0.6161 | 0.105 | 0.007 | 3.9811E-51 | 225.00 |
| *IGFBP3* | rs2854746 | G | C | 0.3974 | 0.461 | 0.007 | 1E-200 | 4337.16 |
| *IGFBP3* | rs9987289 | A | G | 0.908 | -0.08 | 0.012 | 1E-11 | 44.44 |
| *IGFBP3* | rs1065656 | C | G | 0.6939 | 0.12 | 0.007 | 3.9811E-58 | 293.88 |
| *IGFBP3* | rs738409 | C | G | 0.2167 | -0.064 | 0.008 | 6.3096E-15 | 64.00 |
| *IGFBP4* | rs4859682 | C | A | 0.4567 | 0.053 | 0.007 | 6.3096E-14 | 57.33 |
| *IGFBP4* | rs28640218 | G | T | 0.1914 | -0.069 | 0.009 | 5.0119E-14 | 58.78 |
| *IGFBP6* | rs13146355 | G | A | 0.4564 | 0.048 | 0.007 | 1.9953E-13 | 47.02 |
| *IGFBP6* | rs10265221 | T | C | 0.2867 | 0.052 | 0.007 | 6.3096E-13 | 55.18 |
| *IGFBP6* | rs822688 | C | T | 0.1222 | -0.126 | 0.01 | 3.1623E-35 | 158.76 |
| *IGFBP6* | rs4997081 | G | C | 0.1923 | -0.075 | 0.008 | 1E-19 | 87.89 |
| *IGFBP7* | rs1718860 | G | A | 0.7332 | -0.372 | 0.008 | 1E-200 | 2162.25 |
| *IGFBP7* | rs3104394 | G | A | 0.8603 | -0.124 | 0.01 | 3.1623E-35 | 153.76 |
| *IGFBP7* | rs7282172 | C | G | 0.1727 | 0.065 | 0.009 | 6.3096E-13 | 52.16 |
| *IGFBP7* | rs738409 | C | G | 0.217 | 0.06 | 0.008 | 2.5119E-13 | 56.25 |
| *IGFBPL1* | rs1260326 | T | C | 0.6083 | -0.07 | 0.007 | 1.9953E-25 | 100.00 |
| *IGFBPL1* | rs7857243 | G | A | 0.8069 | 0.495 | 0.009 | 1E-200 | 3025.00 |
| *IGHMBP2* | rs607929 | C | G | 0.449 | -0.054 | 0.008 | 2.5119E-12 | 45.56 |
| *IGHMBP2* | rs11228377 | T | C | 0.5984 | -0.079 | 0.008 | 7.9433E-24 | 97.52 |
| *IGLC2* | rs188468174 | C | T | 0.0141 | -0.284 | 0.032 | 3.9811E-19 | 78.77 |
| *IGLC2* | rs12741847 | C | T | 0.3904 | -0.054 | 0.008 | 1.2589E-12 | 45.56 |
| *IGLC2* | rs115975731 | G | A | 0.0318 | -0.396 | 0.021 | 5.0119E-76 | 355.59 |
| *IGLC2* | rs111632177 | G | A | 0.1138 | 0.08 | 0.012 | 7.9433E-12 | 44.44 |
| *IGLC2* | rs12880641 | T | G | 0.6609 | -0.058 | 0.008 | 1.2589E-13 | 52.56 |
| *IGLC2* | rs1407 | C | G | 0.2838 | -0.071 | 0.009 | 3.9811E-16 | 62.23 |
| *IGLC2* | rs34562254 | G | A | 0.0973 | 0.109 | 0.012 | 1.2589E-18 | 82.51 |
| *IGLC2* | rs2856876 | C | A | 0.1162 | 0.229 | 0.012 | 7.9433E-87 | 364.17 |
| *IGSF21* | rs11261050 | C | T | 0.319 | -0.152 | 0.008 | 3.1623E-82 | 361.00 |
| *IGSF21* | rs2395204 | A | G | 0.1774 | -0.083 | 0.01 | 3.1623E-18 | 68.89 |
| *IGSF21* | rs13292932 | C | A | 0.3093 | 0.055 | 0.008 | 3.9811E-12 | 47.27 |
| *IGSF21* | rs186021206 | G | A | 0.0065 | 0.683 | 0.049 | 3.1623E-43 | 194.29 |
| *IGSF3* | rs2284860 | T | C | 0.2166 | 0.11 | 0.009 | 6.3096E-34 | 149.38 |
| *IGSF3* | rs8176749 | C | T | 0.0614 | 0.14 | 0.015 | 6.3096E-20 | 87.11 |
| *IGSF3* | rs56278466 | T | G | 0.6611 | 0.153 | 0.008 | 1.2589E-85 | 365.77 |
| *IGSF3* | rs3747207 | G | A | 0.2157 | 0.064 | 0.009 | 6.3096E-13 | 50.57 |

| | | | | | | | | |
|---|---|---|---|---|---|---|---|---|
| *IGSF8* | rs12408242 | T | C | 0.2147 | 0.156 | 0.009 | 1.2589E-65 | 300.44 |
| *IGSF8* | rs6967348 | G | A | 0.9123 | -0.092 | 0.013 | 3.1623E-12 | 50.08 |
| *IGSF8* | rs6993770 | A | T | 0.2866 | -0.086 | 0.008 | 1.9953E-25 | 115.56 |
| *IGSF8* | rs28365809 | T | C | 0.1461 | -0.135 | 0.011 | 1.5849E-35 | 150.62 |
| *IGSF8* | rs892090 | T | G | 0.8338 | 0.073 | 0.01 | 3.9811E-13 | 53.29 |
| *IGSF9* | rs3747617 | C | T | 0.0533 | -0.479 | 0.017 | 2.512E-181 | 793.91 |
| *IGSF9* | rs1260326 | T | C | 0.6086 | -0.136 | 0.008 | 7.9433E-74 | 289.00 |
| *IGSF9* | rs1047891 | C | A | 0.3127 | -0.067 | 0.008 | 1.9953E-17 | 70.14 |
| *IGSF9* | rs71556736 | C | T | 0.1292 | -0.125 | 0.011 | 1.2589E-30 | 129.13 |
| *IGSF9* | rs139974673 | T | C | 0.0244 | 0.165 | 0.024 | 2.5119E-12 | 47.27 |
| *IKZF2* | rs34136174 | A | C | 0.4252 | 0.079 | 0.008 | 2.5119E-23 | 97.52 |
| *IKZF2* | rs762465 | C | T | 0.6351 | 0.106 | 0.008 | 1E-39 | 175.56 |
| *IL10* | rs2476601 | A | G | 0.8995 | -0.088 | 0.013 | 3.9811E-12 | 45.82 |
| *IL10* | rs3024495 | C | T | 0.1537 | -0.141 | 0.011 | 3.9811E-40 | 164.31 |
| *IL10* | rs67619842 | A | T | 0.1237 | 0.181 | 0.012 | 6.3096E-54 | 227.51 |
| *IL10* | rs3135932 | A | G | 0.1655 | 0.265 | 0.01 | 2.512E-147 | 702.25 |
| *IL10* | rs7203793 | C | G | 0.3573 | -0.067 | 0.008 | 3.9811E-17 | 70.14 |
| *IL10RA* | rs3135932 | A | G | 0.1655 | 0.189 | 0.01 | 1.2589E-74 | 357.21 |
| *IL10RB* | rs7532110 | T | A | 0.8035 | 0.06 | 0.009 | 3.1623E-12 | 44.44 |
| *IL10RB* | rs61747728 | C | T | 0.0379 | 0.192 | 0.018 | 1.9953E-27 | 113.78 |
| *IL10RB* | rs56387622 | T | C | 0.0992 | 0.079 | 0.011 | 6.3096E-12 | 51.58 |
| *IL10RB* | rs73632737 | G | C | 0.0721 | 0.089 | 0.013 | 1.2589E-11 | 46.87 |
| *IL10RB* | rs186021206 | G | A | 0.0064 | 0.413 | 0.047 | 6.3096E-19 | 77.22 |
| *IL10RB* | rs2659005 | C | T | 0.4407 | 0.07 | 0.007 | 1E-24 | 100.00 |
| *IL10RB* | rs2515717 | G | A | 0.4495 | 0.532 | 0.007 | 1E-200 | 5776.00 |
| *IL11* | rs188468174 | C | T | 0.0142 | 0.242 | 0.033 | 1.9953E-13 | 53.78 |
| *IL11* | rs28479853 | A | G | 0.8566 | -0.076 | 0.011 | 1E-11 | 47.74 |
| *IL12A_IL12B* | rs12471768 | T | C | 0.705 | 0.05 | 0.007 | 1.5849E-11 | 51.02 |
| *IL12A_IL12B* | rs13025330 | C | T | 0.2172 | -0.06 | 0.008 | 3.9811E-13 | 56.25 |
| *IL12A_IL12B* | rs72852162 | A | C | 0.1125 | 0.077 | 0.011 | 1.2589E-12 | 49.00 |
| *IL12A_IL12B* | rs11130215 | A | G | 0.1732 | 0.098 | 0.009 | 1E-26 | 118.57 |
| *IL12A_IL12B* | rs9815073 | C | A | 0.3422 | 0.303 | 0.008 | 1E-200 | 1434.52 |
| *IL12A_IL12B* | rs4244437 | G | A | 0.6767 | 0.543 | 0.008 | 1E-200 | 4607.02 |
| *IL12A_IL12B* | rs9265987 | T | C | 0.3224 | 0.098 | 0.007 | 1.9953E-41 | 196.00 |
| *IL12A_IL12B* | rs836559 | G | C | 0.4587 | 0.068 | 0.007 | 2.5119E-23 | 94.37 |
| *IL12A_IL12B* | rs4287251 | T | C | 0.2009 | -0.064 | 0.008 | 3.1623E-14 | 64.00 |
| *IL12A_IL12B* | rs11039216 | C | T | 0.5304 | 0.046 | 0.007 | 1.5849E-11 | 43.18 |
| *IL12A_IL12B* | rs561877 | G | A | 0.3179 | -0.056 | 0.007 | 1.5849E-14 | 64.00 |
| *IL12A_IL12B* | rs10876864 | G | A | 0.5727 | -0.07 | 0.007 | 3.9811E-24 | 100.00 |
| *IL12A_IL12B* | rs3184504 | T | C | 0.5168 | -0.129 | 0.007 | 5.0119E-81 | 339.61 |
| *IL12A_IL12B* | rs76428106 | T | C | 0.0129 | 0.39 | 0.031 | 7.9433E-36 | 158.27 |
| *IL12A_IL12B* | rs423575 | C | G | 0.4844 | -0.052 | 0.007 | 2.5119E-14 | 55.18 |
| *IL12A_IL12B* | rs12588969 | G | C | 0.6841 | -0.107 | 0.007 | 1.2589E-48 | 233.65 |
| *IL12A_IL12B* | rs1987102 | T | C | 0.26 | -0.075 | 0.008 | 1.9953E-22 | 87.89 |
| *IL12B* | rs58394161 | T | C | 0.1521 | 0.067 | 0.009 | 7.9433E-13 | 55.42 |
| *IL12B* | rs12471768 | T | C | 0.7049 | 0.05 | 0.007 | 1.2589E-11 | 51.02 |
| *IL12B* | rs13025330 | C | T | 0.2167 | -0.057 | 0.008 | 3.9811E-12 | 50.77 |
| *IL12B* | rs72852162 | A | C | 0.1124 | 0.082 | 0.011 | 1.5849E-14 | 55.57 |
| *IL12B* | rs11130215 | A | G | 0.1735 | 0.09 | 0.009 | 7.9433E-24 | 100.00 |
| *IL12B* | rs9815073 | C | A | 0.3428 | 0.304 | 0.007 | 1E-200 | 1886.04 |
| *IL12B* | rs4244437 | G | A | 0.6773 | 0.56 | 0.008 | 1E-200 | 4900.00 |
| *IL12B* | rs3130459 | T | C | 0.2569 | 0.108 | 0.008 | 1.9953E-44 | 182.25 |
| *IL12B* | rs836558 | C | A | 0.4589 | 0.067 | 0.007 | 1.2589E-23 | 91.61 |
| *IL12B* | rs4287251 | T | C | 0.2006 | -0.062 | 0.008 | 7.9433E-14 | 60.06 |
| *IL12B* | rs11039216 | C | T | 0.5302 | 0.048 | 0.007 | 1.2589E-12 | 47.02 |
| *IL12B* | rs502822 | C | T | 0.3026 | -0.061 | 0.007 | 7.9433E-17 | 75.94 |
| *IL12B* | rs10876864 | G | A | 0.573 | -0.07 | 0.007 | 6.3096E-25 | 100.00 |
| *IL12B* | rs3184504 | T | C | 0.5174 | -0.129 | 0.007 | 6.3096E-84 | 339.61 |
| *IL12B* | rs76428106 | T | C | 0.0128 | 0.398 | 0.031 | 2.5119E-38 | 164.83 |
| *IL12B* | rs374173 | G | C | 0.4838 | -0.052 | 0.007 | 1.9953E-14 | 55.18 |
| *IL12B* | rs12588969 | G | C | 0.6842 | -0.108 | 0.007 | 1.2589E-50 | 238.04 |
| *IL12B* | rs1987102 | T | C | 0.26 | -0.077 | 0.008 | 3.9811E-24 | 92.64 |
| *IL12B* | rs4843855 | T | C | 0.7277 | -0.051 | 0.008 | 1.5849E-11 | 40.64 |
| *IL12B* | rs73068668 | G | A | 0.0754 | 0.09 | 0.013 | 1.2589E-12 | 47.93 |
| *IL12RB1* | rs2844619 | C | G | 0.6074 | 0.084 | 0.007 | 6.3096E-30 | 144.00 |
| *IL12RB1* | rs950802 | G | A | 0.3065 | 0.279 | 0.008 | 1E-200 | 1216.27 |
| *IL12RB1* | rs3184504 | T | C | 0.5177 | -0.081 | 0.007 | 1.2589E-29 | 133.90 |
| *IL12RB1* | rs12720356 | A | C | 0.0986 | -0.133 | 0.013 | 2.5119E-26 | 104.67 |

| | | | | | | | | |
|---|---|---|---|---|---|---|---|---|
| *IL12RB1* | rs369378 | G | A | 0.2131 | -0.456 | 0.009 | 1E-200 | 2567.11 |
| *IL12RB2* | rs511874 | G | C | 0.2833 | -0.074 | 0.009 | 5.0119E-18 | 67.60 |
| *IL13* | rs34118068 | G | A | 0.1054 | 0.148 | 0.013 | 1E-30 | 129.61 |
| *IL13RA1* | rs139130389 | C | CTA | 0.0725 | -0.142 | 0.015 | 2.5119E-22 | 89.62 |
| *IL13RA1* | rs351973 | A | G | 0.6411 | 0.079 | 0.008 | 7.9433E-23 | 97.52 |
| *IL13RA1* | rs73201506 | A | G | 0.045 | -0.142 | 0.018 | 1E-14 | 62.23 |
| *IL15* | rs149968614 | C | T | 0.0018 | 0.649 | 0.088 | 1.5849E-13 | 54.39 |
| *IL15* | rs6693121 | C | A | 0.4075 | 0.052 | 0.008 | 6.3096E-12 | 42.25 |
| *IL15* | rs4848370 | C | T | 0.2716 | 0.145 | 0.008 | 1.9953E-67 | 328.52 |
| *IL15* | rs1058315 | G | C | 0.2269 | -0.067 | 0.009 | 6.3096E-14 | 55.42 |
| *IL15* | rs4683324 | T | C | 0.5775 | -0.081 | 0.008 | 3.9811E-27 | 102.52 |
| *IL15* | rs9844706 | G | A | 0.8901 | -0.085 | 0.012 | 7.9433E-13 | 50.17 |
| *IL15* | rs2523594 | C | T | 0.6217 | -0.123 | 0.008 | 3.9811E-57 | 236.39 |
| *IL15* | rs6924776 | A | G | 0.4867 | 0.052 | 0.007 | 2.5119E-12 | 55.18 |
| *IL15* | rs1378901 | T | C | 0.437 | 0.051 | 0.007 | 1.2589E-11 | 53.08 |
| *IL15* | rs8177636 | T | C | 0.4318 | -0.056 | 0.007 | 5.0119E-14 | 64.00 |
| *IL15* | rs2646420 | C | T | 0.3793 | -0.063 | 0.008 | 1.5849E-16 | 62.02 |
| *IL15* | rs1926197 | A | G | 0.4547 | -0.058 | 0.007 | 5.0119E-15 | 68.65 |
| *IL15* | rs12363646 | T | C | 0.2914 | -0.063 | 0.008 | 1.2589E-14 | 62.02 |
| *IL15* | rs7310615 | C | G | 0.5177 | 0.09 | 0.007 | 7.9433E-34 | 165.31 |
| *IL15* | rs76428106 | T | C | 0.013 | -0.302 | 0.034 | 6.3096E-19 | 78.90 |
| *IL15* | rs11632933 | G | A | 0.618 | 0.052 | 0.008 | 1E-11 | 42.25 |
| *IL15* | rs58745116 | G | A | 0.3901 | 0.147 | 0.008 | 3.1623E-82 | 337.64 |
| *IL15* | rs3212802 | G | T | 0.1838 | 0.089 | 0.01 | 3.9811E-20 | 79.21 |
| *IL15* | rs7268811 | C | A | 0.1671 | 0.07 | 0.01 | 2.5119E-12 | 49.00 |
| *IL15* | rs35477491 | C | T | 0.2239 | 0.072 | 0.011 | 1E-11 | 42.84 |
| *IL15* | rs228951 | A | G | 0.3663 | 0.074 | 0.008 | 2.5119E-21 | 85.56 |
| *IL15RA* | rs539115936 | C | CT | 0.4707 | -0.066 | 0.008 | 1E-17 | 68.06 |
| *IL15RA* | rs1050486 | C | T | 0.6159 | 0.063 | 0.008 | 7.9433E-15 | 62.02 |
| *IL15RA* | rs3136630 | C | T | 0.2957 | -0.259 | 0.008 | 1E-200 | 1048.14 |
| *IL15RA* | rs111338191 | A | AAATT | 0.5215 | -0.053 | 0.007 | 7.9433E-13 | 57.33 |
| *IL16* | rs12342201 | G | A | 0.4766 | -0.061 | 0.007 | 1.2589E-18 | 75.94 |
| *IL16* | rs58754882 | G | A | 0.0904 | -0.968 | 0.013 | 1E-200 | 5544.52 |
| *IL17C* | rs144937970 | G | A | 0.0088 | 0.356 | 0.04 | 7.9433E-19 | 79.21 |
| *IL17C* | rs8177833 | GC | G | 0.0552 | 0.117 | 0.016 | 1E-12 | 53.47 |
| *IL17C* | rs17700884 | G | T | 0.362 | -0.12 | 0.008 | 7.9433E-53 | 225.00 |
| *IL17D* | rs4770059 | T | C | 0.7367 | -0.398 | 0.008 | 1E-200 | 2475.06 |
| *IL17D* | rs77542162 | A | G | 0.023 | 0.248 | 0.022 | 7.9433E-29 | 127.07 |
| *IL17F* | rs66834286 | C | CGTTTGTTT | 0.1074 | -0.256 | 0.012 | 6.3096E-94 | 455.11 |
| *IL17F* | rs10152051 | A | G | 0.4466 | -0.068 | 0.008 | 5.0119E-17 | 72.25 |
| *IL17RA* | rs78689694 | G | C | 0.133 | -0.064 | 0.008 | 1.2589E-15 | 64.00 |
| *IL17RA* | rs186021206 | G | A | 0.0064 | 0.474 | 0.037 | 3.1623E-37 | 164.12 |
| *IL17RA* | rs4819959 | G | A | 0.495 | 0.857 | 0.008 | 1E-200 | 11475.77 |
| *IL17RB* | rs1260326 | T | C | 0.6086 | 0.056 | 0.006 | 3.9811E-20 | 87.11 |
| *IL17RB* | rs6445607 | G | T | 0.6247 | -0.807 | 0.008 | 1E-200 | 10175.77 |
| *IL17RB* | rs57413756 | C | G | 0.1113 | -0.085 | 0.012 | 1.5849E-12 | 50.17 |
| *IL17RB* | rs2894204 | C | T | 0.6326 | -0.065 | 0.006 | 1.5849E-26 | 117.36 |
| *IL17RB* | rs34121855 | T | G | 0.2011 | 0.053 | 0.007 | 6.3096E-13 | 57.33 |
| *IL17RB* | rs1870148 | G | A | 0.7891 | 0.057 | 0.007 | 5.0119E-15 | 66.31 |
| *IL17RB* | rs10841753 | T | C | 0.1862 | 0.064 | 0.008 | 1.9953E-17 | 64.00 |
| *IL17RB* | rs1169284 | T | C | 0.3129 | -0.055 | 0.006 | 5.0119E-18 | 84.03 |
| *IL17RB* | rs28929474 | C | T | 0.0208 | 0.229 | 0.021 | 1.9953E-28 | 118.91 |
| *IL17RB* | rs77542162 | A | G | 0.0228 | 0.154 | 0.02 | 1E-14 | 59.29 |
| *IL18* | rs385076 | T | C | 0.6443 | 0.22 | 0.008 | 2.512E-187 | 756.25 |
| *IL18* | rs77762937 | G | T | 0.2878 | 0.06 | 0.008 | 5.0119E-14 | 56.25 |
| *IL18* | rs130066 | G | C | 0.4581 | -0.053 | 0.007 | 1.2589E-13 | 57.33 |
| *IL18* | rs2290414 | G | A | 0.4408 | -0.072 | 0.007 | 5.0119E-24 | 105.80 |
| *IL18* | rs117558678 | A | T | 0.2403 | 0.072 | 0.009 | 1.2589E-16 | 64.00 |
| *IL18* | rs17103763 | T | A | 0.2606 | 0.108 | 0.008 | 1.5849E-38 | 182.25 |
| *IL18* | rs2250417 | T | C | 0.5405 | -0.295 | 0.007 | 1E-200 | 1776.02 |
| *IL18* | rs10675507 | C | CTT | 0.5168 | -0.053 | 0.007 | 1.2589E-13 | 57.33 |
| *IL18* | rs7137828 | C | T | 0.5159 | -0.066 | 0.007 | 1.9953E-20 | 88.90 |
| *IL18* | rs10418046 | T | G | 0.2112 | 0.066 | 0.009 | 3.1623E-14 | 53.78 |
| *IL18BP* | rs61747728 | C | T | 0.0382 | 0.164 | 0.019 | 1.5849E-18 | 74.50 |
| *IL18BP* | rs10901252 | G | C | 0.0614 | 0.155 | 0.015 | 7.9433E-25 | 106.78 |
| *IL18BP* | rs5743659 | G | A | 0.0061 | -0.547 | 0.048 | 1.2589E-30 | 129.87 |
| *IL18BP* | rs112771035 | C | G | 0.0714 | 0.144 | 0.014 | 1.5849E-24 | 105.80 |
| *IL18BP* | rs7137828 | C | T | 0.516 | -0.117 | 0.007 | 5.0119E-60 | 279.37 |

| | | | | | | | | |
|---|---|---|---|---|---|---|---|---|
| *IL18BP* | rs186021206 | G | A | 0.0064 | 0.865 | 0.05 | 3.1623E-68 | 299.29 |
| *IL18BP* | rs10445407 | C | A | 0.4791 | -0.051 | 0.007 | 1.5849E-12 | 53.08 |
| *IL18R1* | rs10190555 | A | G | 0.7741 | -0.898 | 0.009 | 1E-200 | 9955.60 |
| *IL18R1* | rs407258 | T | C | 0.25 | -0.067 | 0.007 | 6.3096E-22 | 91.61 |
| *IL18R1* | rs28636815 | A | G | 0.3767 | -0.044 | 0.006 | 1E-12 | 53.78 |
| *IL18R1* | rs56278466 | T | G | 0.6613 | 0.111 | 0.006 | 3.1623E-68 | 342.25 |
| *IL18R1* | rs111338191 | A | AAATT | 0.5213 | -0.043 | 0.006 | 1.2589E-12 | 51.36 |
| *IL18R1* | rs7970695 | G | A | 0.6238 | -0.047 | 0.006 | 7.9433E-14 | 61.36 |
| *IL18R1* | rs28929474 | C | T | 0.0208 | 0.264 | 0.021 | 7.9433E-36 | 158.04 |
| *IL18R1* | rs147233090 | C | T | 0.0236 | 0.149 | 0.02 | 7.9433E-14 | 55.50 |
| *IL18R1* | rs77303550 | C | T | 0.1959 | -0.067 | 0.008 | 2.5119E-18 | 70.14 |
| *IL18R1* | rs1801689 | A | C | 0.0302 | 0.429 | 0.018 | 1E-130 | 568.03 |
| *IL18R1* | rs12019136 | G | A | 0.0413 | 0.113 | 0.015 | 7.9433E-14 | 56.75 |
| *IL18RAP* | rs72490060 | C | T | 0.1948 | 0.118 | 0.011 | 6.3096E-27 | 115.07 |
| *IL18RAP* | rs62146110 | C | T | 0.2268 | 0.074 | 0.009 | 6.3096E-16 | 67.60 |
| *IL18RAP* | rs1655901 | T | C | 0.5587 | -0.072 | 0.008 | 1.5849E-20 | 81.00 |
| *IL18RAP* | rs4774021 | G | T | 0.137 | 0.157 | 0.011 | 7.9433E-46 | 203.71 |
| *IL19* | rs2243191 | T | C | 0.7795 | 0.356 | 0.009 | 1E-200 | 1564.64 |
| *IL19* | rs80235202 | A | G | 0.2823 | 0.056 | 0.008 | 1E-11 | 49.00 |
| *IL19* | rs34819998 | G | C | 0.1457 | 0.076 | 0.011 | 1.2589E-12 | 47.74 |
| *IL19* | rs2608894 | T | C | 0.7649 | -0.069 | 0.009 | 5.0119E-15 | 58.78 |
| *IL19* | rs492602 | A | G | 0.5093 | 0.378 | 0.007 | 1E-200 | 2916.00 |
| *IL1A* | rs28464365 | T | C | 0.0438 | -0.267 | 0.019 | 1.5849E-45 | 197.48 |
| *IL1B* | rs6993770 | A | T | 0.2873 | -0.077 | 0.008 | 3.1623E-20 | 92.64 |
| *IL1B* | rs34436714 | C | A | 0.2089 | 0.297 | 0.009 | 1E-200 | 1089.00 |
| *IL1R1* | rs61830291 | A | C | 0.0975 | -0.126 | 0.012 | 1E-24 | 110.25 |
| *IL1R1* | rs6545929 | G | A | 0.1947 | -0.084 | 0.009 | 1.9953E-19 | 87.11 |
| *IL1R1* | rs3917238 | C | T | 0.3007 | -0.184 | 0.008 | 1.585E-114 | 529.00 |
| *IL1R1* | rs55709272 | T | C | 0.4393 | 0.135 | 0.007 | 5.0119E-74 | 371.94 |
| *IL1R1* | rs10935473 | G | T | 0.4401 | -0.289 | 0.007 | 1E-200 | 1704.51 |
| *IL1R1* | rs12505880 | C | A | 0.344 | 0.052 | 0.008 | 1.5849E-11 | 42.25 |
| *IL1R1* | rs1634775 | A | G | 0.6225 | -0.075 | 0.007 | 1.2589E-23 | 114.80 |
| *IL1R1* | rs4841133 | A | G | 0.9082 | -0.088 | 0.013 | 1.5849E-12 | 45.82 |
| *IL1R1* | rs8176672 | C | T | 0.0612 | 0.214 | 0.015 | 5.0119E-46 | 203.54 |
| *IL1R1* | rs56278466 | T | G | 0.6612 | 0.064 | 0.008 | 3.9811E-17 | 64.00 |
| *IL1R1* | rs28929474 | C | T | 0.0207 | 0.25 | 0.025 | 6.3096E-23 | 100.00 |
| *IL1R1* | rs186021206 | G | A | 0.0064 | 0.605 | 0.05 | 3.1623E-34 | 146.41 |
| *IL1R1* | rs2292642 | C | T | 0.6043 | -0.07 | 0.007 | 3.9811E-21 | 100.00 |
| *IL1R1* | rs2837988 | C | A | 0.3617 | 0.089 | 0.008 | 7.9433E-32 | 123.77 |
| *IL1R2* | rs165316 | A | G | 0.1981 | -0.063 | 0.009 | 1.9953E-13 | 49.00 |
| *IL1R2* | rs629301 | G | T | 0.7777 | -0.07 | 0.008 | 2.5119E-17 | 76.56 |
| *IL1R2* | rs2310170 | T | G | 0.3995 | -0.517 | 0.008 | 1E-200 | 4176.39 |
| *IL1R2* | rs3129853 | G | A | 0.3065 | -0.071 | 0.007 | 1E-21 | 102.88 |
| *IL1R2* | rs6973520 | C | T | 0.5067 | 0.067 | 0.007 | 2.5119E-22 | 91.61 |
| *IL1R2* | rs28601761 | C | G | 0.4196 | -0.125 | 0.007 | 3.9811E-71 | 318.88 |
| *IL1R2* | rs1870138 | A | G | 0.7962 | 0.084 | 0.008 | 5.0119E-23 | 110.25 |
| *IL1R2* | rs10831245 | G | A | 0.3866 | 0.066 | 0.007 | 1.2589E-20 | 88.90 |
| *IL1R2* | rs11045856 | T | G | 0.24 | 0.064 | 0.008 | 1E-15 | 64.00 |
| *IL1R2* | rs1169296 | A | G | 0.3252 | 0.073 | 0.007 | 3.9811E-23 | 108.76 |
| *IL1R2* | rs186021206 | G | A | 0.0064 | 0.497 | 0.047 | 6.3096E-26 | 111.82 |
| *IL1R2* | rs77542162 | A | G | 0.0229 | 0.179 | 0.023 | 7.9433E-15 | 60.57 |
| *IL1RAP* | rs6444442 | A | G | 0.8458 | -1.401 | 0.01 | 1E-200 | 19628.01 |
| *IL1RAP* | rs753950345 | AATCC | A | 0.163 | 0.059 | 0.007 | 2.5119E-19 | 71.04 |
| *IL1RAP* | rs4921914 | C | T | 0.779 | -0.055 | 0.006 | 7.9433E-21 | 84.03 |
| *IL1RAP* | rs7310409 | A | G | 0.6242 | 0.041 | 0.005 | 2.5119E-16 | 67.24 |
| *IL1RAP* | rs139974673 | T | C | 0.0244 | 0.196 | 0.016 | 3.1623E-36 | 150.06 |
| *IL1RAP* | rs273510 | A | G | 0.293 | -0.044 | 0.005 | 1.9953E-16 | 77.44 |
| *IL1RL1* | rs13028833 | C | G | 0.5168 | 0.063 | 0.007 | 6.3096E-20 | 81.00 |
| *IL1RL1* | rs12470864 | G | A | 0.3906 | -0.689 | 0.007 | 1E-200 | 9688.18 |
| *IL1RL1* | rs7616330 | C | A | 0.1752 | -0.051 | 0.007 | 5.0119E-12 | 53.08 |
| *IL1RL1* | rs9829963 | T | C | 0.2135 | 0.07 | 0.007 | 1.5849E-24 | 100.00 |
| *IL1RL1* | rs13107325 | C | T | 0.0749 | 0.113 | 0.011 | 1.2589E-26 | 105.53 |
| *IL1RL1* | rs2519093 | C | T | 0.1844 | -0.091 | 0.007 | 2.5119E-36 | 169.00 |
| *IL1RL1* | rs11603123 | G | A | 0.0336 | 0.381 | 0.016 | 1.995E-132 | 567.04 |
| *IL1RL1* | rs1378942 | C | A | 0.6813 | 0.053 | 0.006 | 3.9811E-19 | 78.03 |
| *IL1RL1* | rs12445547 | G | T | 0.3122 | -0.06 | 0.006 | 1.2589E-23 | 100.00 |
| *IL1RL1* | rs186021206 | G | A | 0.0064 | 0.963 | 0.038 | 1E-138 | 642.22 |
| *IL1RL1* | rs142016905 | T | TAGG | 0.1496 | -0.063 | 0.008 | 1E-15 | 62.02 |

| | | | | | | | | |
|---|---|---|---|---|---|---|---|---|
| *IL1RL1* | rs2899319 | A | C | 0.6169 | 0.044 | 0.006 | 3.9811E-14 | 53.78 |
| *IL1RL2* | rs10779835 | T | C | 0.6125 | -0.049 | 0.007 | 1.2589E-12 | 49.00 |
| *IL1RL2* | rs3917265 | T | C | 0.5434 | -0.437 | 0.007 | 1E-200 | 3897.33 |
| *IL1RL2* | rs11928797 | C | A | 0.116 | 0.075 | 0.01 | 7.9433E-13 | 56.25 |
| *IL1RL2* | rs13203302 | C | T | 0.0838 | 0.106 | 0.012 | 1.2589E-18 | 78.03 |
| *IL1RL2* | rs10815254 | C | T | 0.2806 | 0.051 | 0.007 | 1E-11 | 53.08 |
| *IL1RL2* | rs56278466 | T | G | 0.661 | 0.067 | 0.007 | 2.5119E-21 | 91.61 |
| *IL1RL2* | rs2291075 | C | T | 0.3863 | 0.049 | 0.007 | 1.5849E-12 | 49.00 |
| *IL1RL2* | rs11065385 | A | G | 0.6929 | -0.162 | 0.007 | 6.31E-109 | 535.59 |
| *IL1RL2* | rs3007036 | A | G | 0.5394 | 0.058 | 0.007 | 5.0119E-18 | 68.65 |
| *IL1RL2* | rs28929474 | C | T | 0.0206 | 0.216 | 0.024 | 3.9811E-20 | 81.00 |
| *IL1RL2* | rs62011287 | A | G | 0.3419 | -0.054 | 0.007 | 2.5119E-14 | 59.51 |
| *IL1RL2* | rs7599 | A | G | 0.6344 | -0.056 | 0.007 | 3.9811E-16 | 64.00 |
| *IL1RN* | rs55709272 | T | C | 0.439 | -0.213 | 0.008 | 1.585E-173 | 708.89 |
| *IL1RN* | rs12941811 | T | C | 0.5747 | -0.055 | 0.007 | 2.5119E-13 | 61.73 |
| *IL20RA* | rs143005532 | C | T | 0.0015 | 0.815 | 0.099 | 1.9953E-16 | 67.77 |
| *IL22* | rs6689 | A | G | 0.189 | -0.072 | 0.009 | 6.3096E-15 | 64.00 |
| *IL22* | rs760407193 | AAGAC | A | 0.2756 | 0.106 | 0.008 | 1.2589E-38 | 175.56 |
| *IL22* | rs4794067 | T | C | 0.2586 | -0.06 | 0.008 | 3.1623E-13 | 56.25 |
| *IL22RA1* | rs7355101 | A | T | 0.1954 | -0.318 | 0.01 | 1E-200 | 1011.24 |
| *IL22RA1* | rs681343 | C | T | 0.5092 | -0.149 | 0.008 | 5.0119E-86 | 346.89 |
| *IL2RA* | rs61747728 | C | T | 0.0381 | 0.128 | 0.017 | 2.5119E-13 | 56.69 |
| *IL2RA* | rs734866 | A | C | 0.1948 | 0.087 | 0.008 | 6.3096E-25 | 118.27 |
| *IL2RA* | rs3094005 | G | T | 0.1409 | 0.12 | 0.01 | 1.9953E-35 | 144.00 |
| *IL2RA* | rs12722497 | C | A | 0.0887 | 0.956 | 0.013 | 1E-200 | 5407.91 |
| *IL2RA* | rs7137828 | C | T | 0.5157 | -0.083 | 0.007 | 1E-35 | 140.59 |
| *IL2RA* | rs1955512 | G | A | 0.5721 | 0.063 | 0.007 | 1E-19 | 81.00 |
| *IL2RA* | rs924138 | T | C | 0.6109 | -0.064 | 0.007 | 3.1623E-20 | 83.59 |
| *IL2RA* | rs2659007 | G | A | 0.45 | 0.054 | 0.007 | 1.2589E-15 | 59.51 |
| *IL2RG* | rs186021206 | G | A | 0.0064 | 0.413 | 0.052 | 2.5119E-15 | 63.08 |
| *IL31* | rs4520243 | T | C | 0.347 | 0.082 | 0.008 | 7.9433E-24 | 105.06 |
| *IL31RA* | rs161651 | C | T | 0.3052 | -0.631 | 0.008 | 1E-200 | 6221.27 |
| *IL31RA* | rs1570350 | A | G | 0.4387 | 0.047 | 0.007 | 1.9953E-12 | 45.08 |
| *IL32* | rs117199990 | C | T | 0.101 | 0.108 | 0.012 | 1.9953E-19 | 81.00 |
| *IL32* | rs145920606 | A | G | 0.0848 | 0.098 | 0.013 | 5.0119E-14 | 56.83 |
| *IL32* | rs12905732 | G | C | 0.2 | 0.096 | 0.009 | 1.9953E-26 | 113.78 |
| *IL32* | rs45499297 | T | C | 0.0943 | -0.395 | 0.013 | 1E-200 | 923.22 |
| *IL32* | rs247617 | C | A | 0.3243 | 0.104 | 0.008 | 1.5849E-40 | 169.00 |
| *IL32* | rs12975366 | T | C | 0.3954 | -0.07 | 0.007 | 1.9953E-21 | 100.00 |
| *IL33* | rs185705622 | G | C | 0.0046 | -0.388 | 0.057 | 1.2589E-11 | 46.34 |
| *IL34* | rs4985556 | C | A | 0.1217 | -1.07 | 0.012 | 1E-200 | 7950.69 |
| *IL36A* | rs895497 | A | G | 0.7035 | -0.063 | 0.008 | 1.5849E-13 | 62.02 |
| *IL36G* | rs61816761 | G | A | 0.0236 | 0.256 | 0.027 | 1E-21 | 89.90 |
| *IL36G* | rs13410389 | G | A | 0.1617 | -0.107 | 0.01 | 1E-24 | 114.49 |
| *IL3RA* | rs876038 | C | T | 0.3159 | -0.055 | 0.007 | 1.5849E-13 | 61.73 |
| *IL3RA* | rs2519093 | C | T | 0.1843 | -0.592 | 0.01 | 1E-200 | 3504.64 |
| *IL3RA* | rs35458154 | G | A | 0.0256 | 0.245 | 0.022 | 1.9953E-28 | 124.02 |
| *IL3RA* | rs186021206 | G | A | 0.0064 | 0.748 | 0.048 | 2.5119E-55 | 242.84 |
| *IL4R* | rs73133996 | G | C | 0.4393 | -0.054 | 0.007 | 7.9433E-14 | 59.51 |
| *IL4R* | rs541020703 | T | TA | 0.3228 | 0.131 | 0.008 | 6.3096E-63 | 268.14 |
| *IL4R* | rs2519093 | C | T | 0.1842 | -0.086 | 0.009 | 1.9953E-20 | 91.31 |
| *IL4R* | rs149425950 | A | AAT | 0.151 | 0.081 | 0.01 | 1.2589E-15 | 65.61 |
| *IL4R* | rs28929474 | C | T | 0.0207 | 0.482 | 0.025 | 1.9953E-80 | 371.72 |
| *IL4R* | rs8060025 | T | G | 0.6122 | 0.314 | 0.008 | 1E-200 | 1540.56 |
| *IL4R* | rs186021206 | G | A | 0.0064 | 0.631 | 0.05 | 3.9811E-37 | 159.26 |
| *IL5RA* | rs188468174 | C | T | 0.0143 | -0.716 | 0.029 | 5.012E-137 | 609.58 |
| *IL5RA* | rs11385062 | G | GA | 0.2373 | -0.054 | 0.008 | 1.2589E-11 | 45.56 |
| *IL5RA* | rs77400868 | A | G | 0.1333 | 0.517 | 0.011 | 1E-200 | 2209.00 |
| *IL5RA* | rs9819371 | C | T | 0.0654 | -0.106 | 0.015 | 1E-12 | 49.94 |
| *IL5RA* | rs9869437 | C | A | 0.3505 | 0.076 | 0.008 | 1E-22 | 90.25 |
| *IL5RA* | rs93059 | G | A | 0.437 | -0.056 | 0.007 | 2.5119E-16 | 64.00 |
| *IL5RA* | rs6810926 | A | G | 0.1707 | 0.075 | 0.009 | 3.1623E-17 | 69.44 |
| *IL5RA* | rs72541008 | A | ACT | 0.2755 | -0.064 | 0.008 | 1.9953E-17 | 64.00 |
| *IL5RA* | rs112838960 | T | TAATA | 0.5156 | 0.068 | 0.007 | 6.3096E-24 | 94.37 |
| *IL5RA* | rs3134745 | C | T | 0.3268 | -0.124 | 0.007 | 3.1623E-66 | 313.80 |
| *IL5RA* | rs66867810 | G | A | 0.3224 | -0.056 | 0.007 | 6.3096E-15 | 64.00 |
| *IL5RA* | rs7758383 | A | G | 0.4878 | -0.053 | 0.007 | 3.1623E-15 | 57.33 |
| *IL5RA* | rs7797255 | A | G | 0.3999 | -0.068 | 0.007 | 3.9811E-23 | 94.37 |

| | | | | | | | | |
|---|---|---|---|---|---|---|---|---|
| *IL5RA* | rs78740585 | G | A | 0.1138 | 0.194 | 0.011 | 1E-75 | 311.04 |
| *IL5RA* | rs2511713 | A | G | 0.2649 | -0.065 | 0.008 | 7.9433E-18 | 66.02 |
| *IL5RA* | rs12342831 | T | C | 0.2538 | -0.054 | 0.008 | 1.5849E-12 | 45.56 |
| *IL5RA* | rs10896045 | A | G | 0.7029 | -0.05 | 0.007 | 6.3096E-12 | 51.02 |
| *IL5RA* | rs12880641 | T | G | 0.6604 | -0.06 | 0.007 | 1.9953E-17 | 73.47 |
| *IL5RA* | rs71374030 | A | ATTAT | 0.7394 | -0.086 | 0.008 | 3.9811E-29 | 115.56 |
| *IL5RA* | rs3803800 | A | G | 0.7896 | -0.072 | 0.008 | 1.9953E-18 | 81.00 |
| *IL5RA* | rs34562254 | G | A | 0.0971 | 0.194 | 0.011 | 3.1623E-66 | 311.04 |
| *IL5RA* | rs4794067 | T | C | 0.259 | -0.052 | 0.008 | 1.5849E-11 | 42.25 |
| *IL5RA* | rs1005980 | C | G | 0.4622 | -0.074 | 0.007 | 7.9433E-28 | 111.76 |
| *IL5RA* | rs386812017 | G | GT | 0.3898 | 0.055 | 0.007 | 1.9953E-15 | 61.73 |
| *IL6* | rs55709272 | T | C | 0.4386 | 0.053 | 0.007 | 6.3096E-13 | 57.33 |
| *IL6* | rs12133641 | A | G | 0.4061 | 0.231 | 0.008 | 1E-200 | 833.77 |
| *IL6* | rs7449585 | G | C | 0.2676 | 0.08 | 0.008 | 5.0119E-22 | 100.00 |
| *IL6* | rs9977672 | G | A | 0.2587 | -0.06 | 0.008 | 1.9953E-12 | 56.25 |
| *IL6R* | rs4129267 | C | T | 0.4043 | 1.105 | 0.008 | 1E-200 | 19078.52 |
| *IL6R* | rs9272460 | A | G | 0.3593 | 0.037 | 0.005 | 1E-13 | 54.76 |
| *IL6R* | rs4055121 | C | T | 0.1347 | -0.052 | 0.007 | 1E-13 | 55.18 |
| *IL6R* | rs186021206 | G | A | 0.0064 | 0.426 | 0.032 | 1.5849E-39 | 177.22 |
| *IL6R* | rs760482467 | CTTTTTTT | C | 0.5832 | 0.04 | 0.005 | 7.9433E-17 | 64.00 |
| *IL6R* | rs123698 | G | C | 0.6046 | 0.034 | 0.005 | 1E-12 | 46.24 |
| *IL6ST* | rs3791101 | G | A | 0.308 | 0.059 | 0.008 | 6.3096E-14 | 54.39 |
| *IL6ST* | rs10935473 | G | T | 0.4403 | -0.174 | 0.007 | 5.012E-122 | 617.88 |
| *IL6ST* | rs138283229 | C | T | 0.0055 | 0.381 | 0.05 | 1.9953E-14 | 58.06 |
| *IL6ST* | rs11739016 | C | T | 0.1188 | 0.261 | 0.011 | 3.981E-117 | 562.98 |
| *IL6ST* | rs4721064 | G | A | 0.4018 | 0.051 | 0.007 | 6.3096E-12 | 53.08 |
| *IL6ST* | rs507666 | G | A | 0.1845 | -0.215 | 0.009 | 3.162E-114 | 570.68 |
| *IL6ST* | rs11011672 | G | T | 0.7028 | 0.061 | 0.008 | 6.3096E-14 | 58.14 |
| *IL6ST* | rs4055121 | C | T | 0.1344 | -0.149 | 0.011 | 1.5849E-43 | 183.48 |
| *IL6ST* | rs186021206 | G | A | 0.0064 | 1.21 | 0.05 | 3.981E-129 | 585.64 |
| *IL6ST* | rs11444782 | A | AT | 0.5578 | -0.055 | 0.007 | 1E-13 | 61.73 |
| *IL6ST* | rs2008174 | C | T | 0.7395 | -0.075 | 0.009 | 1E-18 | 69.44 |
| *IL7* | rs3804749 | C | T | 0.5925 | -0.065 | 0.008 | 3.9811E-17 | 66.02 |
| *IL7* | rs3207561 | A | G | 0.3752 | 0.054 | 0.008 | 3.9811E-12 | 45.56 |
| *IL7* | rs146201210 | C | T | 0.0952 | -0.183 | 0.013 | 3.9811E-45 | 198.16 |
| *IL7* | rs6993770 | A | T | 0.2869 | -0.105 | 0.008 | 6.3096E-36 | 172.27 |
| *IL7R* | rs34556682 | A | G | 0.0806 | -0.082 | 0.011 | 1E-14 | 55.57 |
| *IL7R* | rs3791101 | G | A | 0.3081 | 0.054 | 0.006 | 1E-17 | 81.00 |
| *IL7R* | rs9850919 | C | T | 0.5943 | 0.049 | 0.006 | 2.5119E-16 | 66.69 |
| *IL7R* | rs13071610 | G | A | 0.1165 | -0.061 | 0.009 | 1.5849E-11 | 45.94 |
| *IL7R* | rs6897932 | C | T | 0.2712 | -0.889 | 0.008 | 1E-200 | 12348.77 |
| *IL7R* | rs12189871 | C | T | 0.092 | 0.16 | 0.01 | 3.1623E-57 | 256.00 |
| *IL7R* | rs170622 | C | T | 0.2076 | -0.053 | 0.007 | 7.9433E-14 | 57.33 |
| *IL7R* | rs2519093 | C | T | 0.1843 | -0.593 | 0.008 | 1E-200 | 5494.52 |
| *IL7R* | rs1059091 | A | G | 0.3184 | -0.042 | 0.006 | 1.2589E-11 | 49.00 |
| *IL7R* | rs35166255 | G | A | 0.0336 | 0.464 | 0.016 | 3.162E-181 | 841.00 |
| *IL7R* | rs186021206 | G | A | 0.0064 | 1.125 | 0.04 | 2.512E-175 | 791.02 |
| *ILKAP* | rs6993770 | A | T | 0.287 | -0.062 | 0.008 | 1.9953E-13 | 60.06 |
| *IMMT* | rs547695713 | CAAAAA | C | 0.5451 | -0.116 | 0.007 | 5.0119E-55 | 274.61 |
| *IMMT* | rs35835517 | G | T | 0.148 | -0.08 | 0.01 | 2.5119E-14 | 64.00 |
| *IMPA1* | rs342298 | C | T | 0.4544 | -0.051 | 0.008 | 1.5849E-11 | 40.64 |
| *IMPA1* | rs2400589 | C | T | 0.2972 | -0.212 | 0.008 | 7.943E-145 | 702.25 |
| *IMPACT* | rs1354034 | T | C | 0.6043 | 0.067 | 0.007 | 3.9811E-19 | 91.61 |
| *IMPG1* | rs12368869 | T | C | 0.2283 | 0.076 | 0.009 | 1.5849E-16 | 71.31 |
| *IMPG1* | rs58534292 | T | C | 0.6504 | 0.057 | 0.008 | 3.1623E-12 | 50.77 |
| *IMPG1* | rs112765826 | A | G | 0.6375 | 0.065 | 0.008 | 3.1623E-14 | 66.02 |
| *ING1* | rs6993770 | A | T | 0.2869 | -0.059 | 0.008 | 3.1623E-12 | 54.39 |
| *INHBB* | rs17050272 | G | A | 0.4119 | 0.365 | 0.007 | 1E-200 | 2718.88 |
| *INHBB* | rs9375702 | C | T | 0.6934 | -0.054 | 0.008 | 1.9953E-12 | 45.56 |
| *INHBB* | rs7826120 | T | C | 0.6698 | 0.076 | 0.008 | 1.5849E-23 | 90.25 |
| *INHBB* | rs771043500 | AATT | A | 0.4838 | 0.052 | 0.007 | 3.9811E-13 | 55.18 |
| *INHBB* | rs7970695 | G | A | 0.6237 | -0.05 | 0.007 | 1.2589E-11 | 51.02 |
| *INHBB* | rs28929474 | C | T | 0.0207 | 0.269 | 0.025 | 1E-26 | 115.78 |
| *INHBB* | rs56325564 | G | A | 0.483 | 0.064 | 0.007 | 1E-18 | 83.59 |
| *INHBC* | rs1730858 | T | C | 0.6513 | 0.054 | 0.007 | 1.2589E-13 | 59.51 |
| *INHBC* | rs1260326 | T | C | 0.6084 | -0.062 | 0.007 | 1E-18 | 78.45 |
| *INHBC* | rs204893 | T | C | 0.4309 | 0.05 | 0.007 | 6.3096E-13 | 51.02 |
| *INHBC* | rs10822145 | C | T | 0.4723 | -0.054 | 0.007 | 6.3096E-15 | 59.51 |

| | | | | | | | | |
|---|---|---|---|---|---|---|---|---|
| *INHBC* | rs3741414 | C | T | 0.2466 | -0.659 | 0.009 | 1E-200 | 5361.49 |
| *INHBC* | rs111349624 | CAA | C | 0.0845 | -0.092 | 0.014 | 1E-11 | 43.18 |
| *INHBC* | rs28929474 | C | T | 0.0207 | 0.19 | 0.024 | 3.9811E-15 | 62.67 |
| *INPP1* | rs141472596 | A | G | 0.0015 | -1.553 | 0.104 | 1.2589E-50 | 222.99 |
| *INPP1* | rs342298 | C | T | 0.4545 | -0.053 | 0.008 | 1.5849E-12 | 43.89 |
| *INPP5D* | rs776060558 | CT | C | 0.1916 | -0.212 | 0.009 | 1.995E-114 | 554.86 |
| *INPP5J* | rs13306780 | A | C | 0.702 | 0.068 | 0.008 | 1.2589E-16 | 72.25 |
| *INSL3* | rs2011425 | T | G | 0.0789 | -0.062 | 0.009 | 1.5849E-12 | 47.46 |
| *INSL3* | rs7763473 | C | T | 0.691 | 0.049 | 0.005 | 1.5849E-21 | 96.04 |
| *INSL4* | rs80293268 | G | C | 0.0484 | -0.133 | 0.018 | 5.0119E-13 | 54.60 |
| *INSL4* | rs7806596 | G | A | 0.4416 | -0.097 | 0.008 | 7.9433E-38 | 147.02 |
| *INSL4* | rs41313766 | C | T | 0.0173 | -0.652 | 0.03 | 6.31E-107 | 472.34 |
| *INSL4* | rs144359915 | G | A | 0.0026 | -0.536 | 0.078 | 6.3096E-12 | 47.22 |
| *INSL5* | rs11809759 | A | G | 0.3986 | 0.18 | 0.007 | 3.981E-130 | 661.22 |
| *INSL5* | rs4849328 | C | G | 0.554 | -0.074 | 0.007 | 6.3096E-24 | 111.76 |
| *INSL5* | rs771100389 | CTCCGT | C | 0.0827 | 0.109 | 0.013 | 7.9433E-16 | 70.30 |
| *INSL5* | rs12365875 | T | C | 0.1438 | -0.146 | 0.01 | 2.5119E-45 | 213.16 |
| *INSL5* | rs10423928 | T | A | 0.1899 | 0.069 | 0.009 | 5.0119E-14 | 58.78 |
| *INSR* | rs61804210 | C | T | 0.1049 | 0.085 | 0.013 | 1.5849E-11 | 42.75 |
| *INSR* | rs35350651 | A | AC | 0.504 | -0.061 | 0.008 | 1.5849E-15 | 58.14 |
| *INSR* | rs112635299 | G | T | 0.0217 | 0.184 | 0.026 | 3.1623E-12 | 50.08 |
| *INSR* | rs7197010 | T | C | 0.5409 | 0.055 | 0.008 | 1E-12 | 47.27 |
| *INSR* | rs704 | G | A | 0.4735 | -0.06 | 0.008 | 7.9433E-15 | 56.25 |
| *INSR* | rs11658693 | A | G | 0.0803 | 0.098 | 0.014 | 3.9811E-12 | 49.00 |
| *INSR* | rs1801689 | A | C | 0.0304 | 0.195 | 0.022 | 2.5119E-18 | 78.56 |
| *INSR* | rs760459 | A | T | 0.7905 | 0.064 | 0.009 | 1.5849E-11 | 50.57 |
| *IPCEF1* | rs1354034 | T | C | 0.6041 | 0.064 | 0.008 | 5.0119E-17 | 64.00 |
| *IPCEF1* | rs883273 | C | A | 0.3217 | 0.084 | 0.008 | 1E-25 | 110.25 |
| *IQGAP2* | rs10037254 | G | A | 0.6044 | 0.145 | 0.008 | 5.0119E-80 | 328.52 |
| *IRAG2* | rs7080536 | G | A | 0.0437 | -0.128 | 0.019 | 6.3096E-12 | 45.39 |
| *IRAG2* | rs72932837 | C | T | 0.2406 | -0.076 | 0.009 | 3.9811E-18 | 71.31 |
| *IRAK1* | rs1354034 | T | C | 0.6035 | 0.058 | 0.008 | 1.5849E-14 | 52.56 |
| *IRAK4* | rs183549317 | G | A | 0.0008 | -1.085 | 0.129 | 3.9811E-17 | 70.74 |
| *ISLR2* | rs13135092 | A | G | 0.0833 | 0.089 | 0.013 | 1.5849E-12 | 46.87 |
| *ISLR2* | rs9263451 | C | A | 0.1047 | -0.08 | 0.011 | 1.9953E-12 | 52.89 |
| *ISLR2* | rs2519093 | C | T | 0.1839 | -0.649 | 0.01 | 1E-200 | 4212.01 |
| *ISLR2* | rs60843925 | T | C | 0.136 | -0.13 | 0.01 | 7.9433E-37 | 169.00 |
| *ISLR2* | rs34868798 | T | C | 0.2235 | 0.246 | 0.008 | 5.012E-185 | 945.56 |
| *ISLR2* | rs186021206 | G | A | 0.0064 | 0.953 | 0.048 | 1.2589E-88 | 394.19 |
| *ISLR2* | rs4803462 | T | A | 0.6073 | 0.061 | 0.007 | 7.9433E-18 | 75.94 |
| *ISM1* | rs539657009 | C | T | 0.1293 | 0.077 | 0.011 | 5.0119E-12 | 49.00 |
| *ISM1* | rs7641117 | C | G | 0.6694 | 0.059 | 0.008 | 3.1623E-14 | 54.39 |
| *ISM1* | rs1998123 | A | G | 0.697 | -0.187 | 0.008 | 1.995E-119 | 546.39 |
| *IST1* | rs1354034 | T | C | 0.6045 | 0.053 | 0.008 | 3.9811E-12 | 43.89 |
| *IST1* | rs342298 | C | T | 0.4555 | -0.053 | 0.008 | 2.5119E-12 | 43.89 |
| *IST1* | rs10733789 | T | C | 0.313 | 0.061 | 0.008 | 1E-13 | 58.14 |
| *IST1* | rs79258602 | A | C | 0.0233 | -0.257 | 0.025 | 6.3096E-25 | 105.68 |
| *ITGA11* | rs1260326 | T | C | 0.6085 | 0.062 | 0.007 | 1.2589E-16 | 78.45 |
| *ITGA11* | rs7111242 | A | G | 0.8816 | -0.12 | 0.011 | 7.9433E-26 | 119.01 |
| *ITGA11* | rs35887873 | T | C | 0.2283 | 0.12 | 0.009 | 1E-43 | 177.78 |
| *ITGA11* | rs753768034 | G | GT | 0.6183 | -0.073 | 0.008 | 1.9953E-21 | 83.27 |
| *ITGA11* | rs2306022 | C | T | 0.09 | -0.359 | 0.013 | 2.512E-169 | 762.61 |
| *ITGA11* | rs186021206 | G | A | 0.0064 | 0.794 | 0.05 | 1.5849E-56 | 252.17 |
| *ITGA11* | rs3092635 | A | AT | 0.8483 | 0.087 | 0.01 | 2.5119E-17 | 75.69 |
| *ITGA2* | rs1354034 | T | C | 0.6044 | 0.047 | 0.007 | 7.9433E-13 | 45.08 |
| *ITGA2* | rs755331637 | C | CA | 0.7016 | -0.62 | 0.008 | 1E-200 | 6006.25 |
| *ITGA2* | rs12245149 | C | A | 0.4882 | -0.049 | 0.006 | 1.5849E-14 | 66.69 |
| *ITGA2* | rs6589572 | G | A | 0.8759 | -0.142 | 0.01 | 1.5849E-48 | 201.64 |
| *ITGA2* | rs17580 | T | A | 0.0496 | 0.263 | 0.015 | 7.9433E-71 | 307.42 |
| *ITGA2* | rs438811 | C | T | 0.2373 | 0.091 | 0.008 | 1.2589E-33 | 129.39 |
| *ITGA2* | rs2868346 | C | T | 0.7598 | -0.093 | 0.007 | 1.2589E-35 | 176.51 |
| *ITGA5* | rs539657009 | C | T | 0.1298 | 0.095 | 0.011 | 2.5119E-17 | 74.59 |
| *ITGA5* | rs2124440 | G | A | 0.5559 | 0.059 | 0.007 | 1E-15 | 71.04 |
| *ITGA5* | rs139078629 | G | A | 0.0096 | -0.693 | 0.037 | 3.9811E-77 | 350.80 |
| *ITGA5* | rs56185965 | T | A | 0.1609 | -0.069 | 0.01 | 1E-11 | 47.61 |
| *ITGA5* | rs78776915 | C | T | 0.4156 | 0.065 | 0.008 | 3.9811E-16 | 66.02 |
| *ITGA5* | rs41341748 | G | A | 0.011 | 0.24 | 0.035 | 6.3096E-12 | 47.02 |
| *ITGA5* | rs8176746 | G | T | 0.0614 | 0.228 | 0.015 | 1.9953E-51 | 231.04 |

| | | | | | | | | |
|---|---|---|---|---|---|---|---|---|
| *ITGA5* | rs10748526 | T | C | 0.7946 | 0.118 | 0.009 | 1.5849E-39 | 171.90 |
| *ITGA5* | rs199991349 | AC | A | 0.0134 | 0.216 | 0.032 | 7.9433E-12 | 45.56 |
| *ITGA5* | rs3184504 | T | C | 0.5175 | -0.106 | 0.007 | 3.9811E-48 | 229.31 |
| *ITGA5* | rs7161799 | C | T | 0.0767 | 0.092 | 0.014 | 1.5849E-11 | 43.18 |
| *ITGA5* | rs186021206 | G | A | 0.0064 | 0.707 | 0.05 | 1.2589E-45 | 199.94 |
| *ITGA5* | rs7288265 | G | A | 0.3288 | -0.076 | 0.008 | 1.5849E-22 | 90.25 |
| *ITGA6* | rs6714597 | T | C | 0.3082 | 0.231 | 0.008 | 1.995E-176 | 833.77 |
| *ITGA6* | rs5030062 | A | C | 0.3728 | 0.076 | 0.008 | 6.3096E-23 | 90.25 |
| *ITGA6* | rs4861708 | G | A | 0.5119 | 0.17 | 0.007 | 7.943E-114 | 589.80 |
| *ITGA6* | rs2897457 | G | T | 0.4913 | -0.122 | 0.007 | 7.9433E-61 | 303.76 |
| *ITGA6* | rs2731673 | T | C | 0.7448 | 0.086 | 0.008 | 3.9811E-24 | 115.56 |
| *ITGA6* | rs579459 | T | C | 0.2075 | -0.104 | 0.009 | 1E-29 | 133.53 |
| *ITGA6* | rs7896518 | A | G | 0.4241 | 0.063 | 0.008 | 1.2589E-16 | 62.02 |
| *ITGA6* | rs543259046 | G | GTA | 0.0822 | -0.108 | 0.014 | 1.9953E-15 | 59.51 |
| *ITGAL* | rs3856240 | T | C | 0.4642 | 0.057 | 0.008 | 5.0119E-14 | 50.77 |
| *ITGAL* | rs597808 | A | G | 0.5158 | -0.097 | 0.008 | 3.9811E-38 | 147.02 |
| *ITGAL* | rs11574938 | G | C | 0.5157 | -0.156 | 0.008 | 3.1623E-94 | 380.25 |
| *ITGAL* | rs760462 | T | C | 0.8177 | 0.067 | 0.01 | 5.0119E-12 | 44.89 |
| *ITGAM* | rs10164604 | T | A | 0.9003 | 0.083 | 0.012 | 1.5849E-11 | 47.84 |
| *ITGAM* | rs118162691 | C | A | 0.0436 | 0.143 | 0.018 | 3.1623E-15 | 63.11 |
| *ITGAM* | rs7865362 | C | T | 0.3633 | 0.11 | 0.008 | 2.5119E-45 | 189.06 |
| *ITGAM* | rs4601794 | A | G | 0.0593 | -0.112 | 0.016 | 3.9811E-13 | 49.00 |
| *ITGAM* | rs3184504 | T | C | 0.5175 | -0.054 | 0.007 | 1E-13 | 59.51 |
| *ITGAM* | rs11150613 | C | T | 0.676 | 0.164 | 0.008 | 2.5119E-95 | 420.25 |
| *ITGAM* | rs776419906 | G | A | 0.001 | -0.992 | 0.124 | 1.5849E-15 | 64.00 |
| *ITGAM* | rs186021206 | G | A | 0.0064 | 0.996 | 0.05 | 1E-86 | 396.81 |
| *ITGAM* | rs4804774 | G | C | 0.2366 | 0.084 | 0.009 | 7.9433E-21 | 87.11 |
| *ITGAM* | rs2230531 | C | T | 0.0083 | 0.487 | 0.042 | 6.3096E-31 | 134.45 |
| *ITGAV* | rs2595391 | T | G | 0.1081 | 0.229 | 0.011 | 5.0119E-88 | 433.40 |
| *ITGAV* | rs60648773 | G | A | 0.2273 | -0.067 | 0.009 | 2.5119E-15 | 55.42 |
| *ITGAV* | rs111338191 | A | AAATT | 0.5204 | 0.056 | 0.007 | 3.9811E-15 | 64.00 |
| *ITGAV* | rs17580 | T | A | 0.0495 | 0.12 | 0.016 | 1.9953E-13 | 56.25 |
| *ITGAV* | rs186021206 | G | A | 0.0064 | 0.863 | 0.049 | 3.9811E-70 | 310.19 |
| *ITGB1* | rs246411 | T | C | 0.319 | 0.099 | 0.008 | 1E-37 | 153.14 |
| *ITGB1* | rs7070761 | A | T | 0.4876 | -0.049 | 0.007 | 6.3096E-12 | 49.00 |
| *ITGB1* | rs11220462 | G | A | 0.1326 | -0.128 | 0.011 | 3.1623E-33 | 135.40 |
| *ITGB1* | rs186021206 | G | A | 0.0064 | 1.094 | 0.05 | 1.259E-107 | 478.73 |
| *ITGB1* | rs12484278 | G | A | 0.1952 | 0.064 | 0.009 | 2.5119E-12 | 50.57 |
| *ITGB1BP2* | rs2631367 | C | G | 0.5187 | 0.052 | 0.008 | 7.9433E-12 | 42.25 |
| *ITGB1BP2* | rs342298 | C | T | 0.455 | -0.098 | 0.008 | 5.0119E-38 | 150.06 |
| *ITGB1BP2* | rs11553699 | A | G | 0.1351 | -0.102 | 0.012 | 1E-18 | 72.25 |
| *ITGB1BP2* | rs13338372 | A | G | 0.1896 | -0.067 | 0.01 | 5.0119E-12 | 44.89 |
| *ITGB2* | rs7433300 | G | A | 0.6154 | -0.051 | 0.007 | 6.3096E-12 | 53.08 |
| *ITGB2* | rs118162691 | C | A | 0.0435 | 0.142 | 0.018 | 1.5849E-15 | 62.23 |
| *ITGB2* | rs7865362 | C | T | 0.363 | 0.089 | 0.008 | 1.5849E-31 | 123.77 |
| *ITGB2* | rs56278466 | T | G | 0.6616 | 0.066 | 0.008 | 5.0119E-18 | 68.06 |
| *ITGB2* | rs4601794 | A | G | 0.0588 | -0.119 | 0.015 | 7.9433E-15 | 62.94 |
| *ITGB2* | rs3184504 | T | C | 0.5174 | -0.122 | 0.007 | 1.2589E-64 | 303.76 |
| *ITGB2* | rs11574639 | G | A | 0.6839 | 0.123 | 0.008 | 3.9811E-56 | 236.39 |
| *ITGB2* | rs776419906 | G | A | 0.001 | -0.994 | 0.121 | 2.5119E-16 | 67.48 |
| *ITGB2* | rs186021206 | G | A | 0.0063 | 1.112 | 0.05 | 1.585E-109 | 494.62 |
| *ITGB2* | rs57543961 | C | CAT | 0.4161 | -0.051 | 0.007 | 1E-11 | 53.08 |
| *ITGB2* | rs760462 | T | C | 0.818 | 0.149 | 0.009 | 5.0119E-57 | 274.09 |
| *ITGB5* | rs35986780 | A | G | 0.2971 | -0.06 | 0.008 | 7.9433E-14 | 56.25 |
| *ITGB5* | rs1354034 | T | C | 0.6041 | 0.077 | 0.007 | 3.9811E-25 | 121.00 |
| *ITGB5* | rs185447638 | G | A | 0.0187 | 0.297 | 0.027 | 3.1623E-28 | 121.00 |
| *ITGB5* | rs56399423 | T | C | 0.4348 | -0.05 | 0.007 | 7.9433E-12 | 51.02 |
| *ITGB5* | rs7896518 | A | G | 0.4241 | 0.092 | 0.007 | 3.1623E-35 | 172.73 |
| *ITGB5* | rs139130389 | C | CTA | 0.0726 | -0.428 | 0.014 | 1E-200 | 934.61 |
| *ITGB5* | rs186021206 | G | A | 0.0064 | 0.524 | 0.05 | 1.2589E-25 | 109.83 |
| *ITGB6* | rs6432601 | A | G | 0.6925 | 0.289 | 0.008 | 1E-200 | 1305.02 |
| *ITGB6* | rs13107325 | C | T | 0.0747 | -0.136 | 0.013 | 6.3096E-24 | 109.44 |
| *ITGB6* | rs6471893 | C | G | 0.2408 | 0.068 | 0.008 | 3.9811E-16 | 72.25 |
| *ITGB6* | rs11414746 | T | TA | 0.4227 | 0.059 | 0.007 | 3.9811E-16 | 71.04 |
| *ITGB6* | rs4382584 | G | A | 0.2751 | 0.08 | 0.008 | 3.9811E-24 | 100.00 |
| *ITGB6* | rs56278466 | T | G | 0.661 | 0.053 | 0.007 | 7.9433E-13 | 57.33 |
| *ITGB6* | rs7310615 | C | G | 0.518 | 0.053 | 0.007 | 5.0119E-14 | 57.33 |
| *ITGB6* | rs75348693 | G | A | 0.5464 | 0.064 | 0.007 | 1.2589E-19 | 83.59 |

| | | | | | | | | |
|---|---|---|---|---|---|---|---|---|
| *ITGB6* | rs77542162 | A | G | 0.023 | 0.203 | 0.024 | 6.3096E-18 | 71.54 |
| *ITGB7* | rs37454 | C | A | 0.3527 | 0.171 | 0.007 | 7.943E-121 | 596.76 |
| *ITGB7* | rs343808 | C | T | 0.2421 | 0.056 | 0.008 | 1.2589E-11 | 49.00 |
| *ITGB7* | rs6759003 | T | C | 0.6395 | 0.077 | 0.007 | 1E-25 | 121.00 |
| *ITGB7* | rs62168051 | C | G | 0.1569 | 0.21 | 0.01 | 3.162E-106 | 441.00 |
| *ITGB7* | rs6771051 | G | T | 0.4565 | -0.108 | 0.007 | 1.5849E-51 | 238.04 |
| *ITGB7* | rs4686445 | T | C | 0.3179 | -0.129 | 0.008 | 1E-64 | 260.02 |
| *ITGB7* | rs1130389 | C | T | 0.147 | 0.107 | 0.011 | 2.5119E-23 | 94.62 |
| *ITGB7* | rs6796 | T | C | 0.279 | 0.067 | 0.008 | 5.0119E-18 | 70.14 |
| *ITGB7* | rs6955795 | C | G | 0.4535 | 0.055 | 0.007 | 6.3096E-15 | 61.73 |
| *ITGB7* | rs7865745 | G | A | 0.3106 | 0.07 | 0.008 | 2.5119E-20 | 76.56 |
| *ITGB7* | rs635634 | C | T | 0.1841 | 0.106 | 0.009 | 3.1623E-31 | 138.72 |
| *ITGB7* | rs78689694 | G | C | 0.1325 | -0.144 | 0.01 | 5.0119E-44 | 207.36 |
| *ITGB7* | rs12232003 | T | C | 0.0744 | 0.181 | 0.013 | 5.0119E-42 | 193.85 |
| *ITGB7* | rs10774625 | A | G | 0.5034 | -0.053 | 0.007 | 1.9953E-14 | 57.33 |
| *ITGB7* | rs7161799 | C | T | 0.0766 | 0.138 | 0.013 | 1.2589E-25 | 112.69 |
| *ITGB7* | rs9896869 | A | G | 0.5472 | -0.354 | 0.007 | 1E-200 | 2557.47 |
| *ITGB7* | rs186021206 | G | A | 0.0064 | 1.815 | 0.051 | 1E-200 | 1266.52 |
| *ITGB7* | rs2659005 | C | T | 0.4416 | 0.086 | 0.007 | 3.1623E-31 | 150.94 |
| *ITGBL1* | rs41341748 | G | A | 0.011 | 0.351 | 0.032 | 1E-27 | 120.31 |
| *ITGBL1* | rs116867595 | G | C | 0.0403 | 0.116 | 0.017 | 1E-11 | 46.56 |
| *ITGBL1* | rs3751198 | A | G | 0.6032 | 0.049 | 0.007 | 1.2589E-12 | 49.00 |
| *ITGBL1* | rs12018094 | G | T | 0.1878 | -0.363 | 0.009 | 1E-200 | 1626.78 |
| *ITGBL1* | rs73045269 | C | T | 0.1713 | 0.072 | 0.009 | 6.3096E-16 | 64.00 |
| *ITGBL1* | rs738409 | C | G | 0.217 | 0.071 | 0.008 | 1.2589E-18 | 78.77 |
| *ITIH1* | rs678 | A | T | 0.3572 | -0.14 | 0.008 | 1.9953E-69 | 306.25 |
| *ITIH3* | rs1260326 | T | C | 0.6083 | -0.055 | 0.007 | 3.1623E-15 | 61.73 |
| *ITIH3* | rs9881468 | C | T | 0.3431 | 0.31 | 0.007 | 1E-200 | 1961.22 |
| *ITIH3* | rs35656703 | C | A | 0.2513 | -0.203 | 0.008 | 1.995E-138 | 643.89 |
| *ITIH3* | rs244451 | T | C | 0.6809 | -0.081 | 0.007 | 1.5849E-28 | 133.90 |
| *ITIH3* | rs35246381 | C | T | 0.7799 | 0.057 | 0.008 | 3.1623E-12 | 50.77 |
| *ITIH3* | rs28601761 | C | G | 0.4192 | 0.055 | 0.007 | 1.9953E-15 | 61.73 |
| *ITIH3* | rs4981022 | G | A | 0.6812 | 0.056 | 0.007 | 3.1623E-14 | 64.00 |
| *ITIH4* | rs1260326 | T | C | 0.6084 | -0.094 | 0.007 | 1.9953E-38 | 180.33 |
| *ITIH4* | rs2710331 | T | C | 0.342 | 0.248 | 0.008 | 1E-200 | 961.00 |
| *ITIH4* | rs2731673 | T | C | 0.7447 | 0.114 | 0.008 | 6.3096E-45 | 203.06 |
| *ITIH4* | rs535696392 | TA | T | 0.4958 | 0.049 | 0.007 | 6.3096E-12 | 49.00 |
| *ITIH5* | rs41298373 | G | A | 0.0995 | 0.169 | 0.013 | 6.3096E-40 | 169.00 |
| *ITM2A* | rs9264683 | A | T | 0.1688 | -0.1 | 0.01 | 1.2589E-23 | 100.00 |
| *ITM2A* | rs709932 | C | T | 0.1537 | -0.298 | 0.01 | 7.943E-177 | 888.04 |
| *ITM2A* | rs1046294 | T | C | 0.3966 | -0.063 | 0.008 | 1.5849E-16 | 62.02 |
| *ITPA* | rs1354034 | T | C | 0.6037 | 0.047 | 0.007 | 1.5849E-12 | 45.08 |
| *ITPA* | rs1127354 | C | A | 0.0745 | -1.185 | 0.015 | 1E-200 | 6241.00 |
| *ITPR1* | rs68066031 | T | C | 0.2299 | -0.138 | 0.009 | 1.5849E-51 | 235.11 |
| *ITPR1* | rs76604555 | G | A | 0.1221 | -0.138 | 0.012 | 7.9433E-32 | 132.25 |
| *ITPR1* | rs1354034 | T | C | 0.6045 | -0.059 | 0.008 | 1.5849E-14 | 54.39 |
| *ITPR1* | rs6993770 | A | T | 0.2866 | -0.095 | 0.008 | 2.5119E-29 | 141.02 |
| *ITPR1* | rs7080386 | C | A | 0.4117 | 0.077 | 0.008 | 3.9811E-23 | 92.64 |
| *ITPR1* | rs7172696 | A | G | 0.7165 | -0.124 | 0.008 | 3.1623E-48 | 240.25 |
| *ITPR1* | rs12445050 | C | T | 0.1387 | 0.08 | 0.011 | 3.1623E-13 | 52.89 |
| *ITPR1* | rs1654425 | T | C | 0.8346 | 0.12 | 0.01 | 1.5849E-31 | 144.00 |
| *ITPRIP* | rs139662037 | T | C | 0.002 | 0.671 | 0.09 | 6.3096E-14 | 55.59 |
| *JAM2* | rs3811444 | C | T | 0.3325 | 0.154 | 0.008 | 1E-90 | 370.56 |
| *JAM2* | rs1260326 | T | C | 0.6085 | 0.051 | 0.007 | 2.5119E-12 | 53.08 |
| *JAM2* | rs8176741 | G | A | 0.0612 | 0.248 | 0.015 | 1.2589E-61 | 273.35 |
| *JAM2* | rs12277151 | C | T | 0.0771 | -0.106 | 0.014 | 5.0119E-15 | 57.33 |
| *JAM2* | rs3184504 | T | C | 0.5171 | 0.053 | 0.007 | 5.0119E-14 | 57.33 |
| *JAM2* | rs36060036 | C | T | 0.1674 | -0.077 | 0.01 | 7.9433E-16 | 59.29 |
| *JAM3* | rs3811444 | C | T | 0.3325 | -0.291 | 0.008 | 1E-200 | 1323.14 |
| *JAM3* | rs1354034 | T | C | 0.6044 | 0.087 | 0.008 | 3.9811E-30 | 118.27 |
| *JAM3* | rs3804749 | C | T | 0.593 | -0.058 | 0.008 | 6.3096E-14 | 52.56 |
| *JAM3* | rs6993770 | A | T | 0.287 | -0.115 | 0.008 | 1E-43 | 206.64 |
| *JAM3* | rs7896518 | A | G | 0.4239 | 0.067 | 0.008 | 3.9811E-18 | 70.14 |
| *JAM3* | rs71474568 | T | TTG | 0.3401 | 0.059 | 0.008 | 1E-13 | 54.39 |
| *JAM3* | rs94499 | A | G | 0.2188 | 0.123 | 0.009 | 7.9433E-42 | 186.78 |
| *JAM3* | rs892090 | T | G | 0.8339 | 0.086 | 0.01 | 1.5849E-17 | 73.96 |
| *JCHAIN* | rs188468174 | C | T | 0.0142 | 0.358 | 0.032 | 1.2589E-28 | 125.16 |
| *JCHAIN* | rs139015452 | C | CACTG | 0.2295 | -0.069 | 0.009 | 5.0119E-14 | 58.78 |

| | | | | | | | | |
|---|---|---|---|---|---|---|---|---|
| *JCHAIN* | rs617119 | C | T | 0.5129 | 0.059 | 0.008 | 6.3096E-15 | 54.39 |
| *JCHAIN* | rs116446171 | C | G | 0.0214 | 0.211 | 0.026 | 1E-15 | 65.86 |
| *JCHAIN* | rs1264457 | G | A | 0.6069 | -0.082 | 0.008 | 7.9433E-26 | 105.06 |
| *JCHAIN* | rs62445869 | G | A | 0.2748 | -0.061 | 0.008 | 1E-12 | 58.14 |
| *JCHAIN* | rs2511710 | G | T | 0.2645 | -0.087 | 0.009 | 5.0119E-24 | 93.44 |
| *JCHAIN* | rs9658720 | T | TG | 0.5541 | -0.059 | 0.008 | 1.5849E-14 | 54.39 |
| *JCHAIN* | rs36051450 | T | C | 0.1959 | 0.084 | 0.01 | 3.9811E-18 | 70.56 |
| *JCHAIN* | rs8051050 | A | G | 0.2602 | 0.084 | 0.009 | 1.2589E-22 | 87.11 |
| *JCHAIN* | rs4815598 | G | C | 0.5435 | 0.058 | 0.008 | 1E-14 | 52.56 |
| *JPT2* | rs1354034 | T | C | 0.6046 | 0.069 | 0.008 | 1.9953E-19 | 74.39 |
| *JPT2* | rs342298 | C | T | 0.455 | -0.056 | 0.008 | 1.5849E-13 | 49.00 |
| *KAZALD1* | rs7639267 | G | T | 0.5603 | 0.062 | 0.007 | 5.0119E-19 | 78.45 |
| *KAZALD1* | rs2228243 | A | G | 0.2001 | -0.218 | 0.009 | 5.012E-139 | 586.72 |
| *KAZALD1* | rs3731211 | T | A | 0.7212 | -0.062 | 0.008 | 1.5849E-15 | 60.06 |
| *KAZALD1* | rs2519093 | C | T | 0.1845 | -0.068 | 0.009 | 2.5119E-14 | 57.09 |
| *KAZALD1* | rs11190812 | G | A | 0.0398 | -1.696 | 0.019 | 1E-200 | 7967.91 |
| *KAZALD1* | rs3751198 | A | G | 0.6034 | 0.054 | 0.007 | 3.9811E-14 | 59.51 |
| *KAZALD1* | rs35350651 | A | AC | 0.5035 | -0.069 | 0.007 | 7.9433E-24 | 97.16 |
| *KAZN* | rs1354034 | T | C | 0.6046 | 0.101 | 0.008 | 3.1623E-41 | 159.39 |
| *KAZN* | rs342293 | C | G | 0.4598 | -0.062 | 0.007 | 7.9433E-17 | 78.45 |
| *KDM3A* | rs4548048 | C | T | 0.1033 | 0.141 | 0.013 | 7.9433E-27 | 117.64 |
| *KDM3A* | rs10957978 | G | T | 0.6481 | 0.055 | 0.008 | 6.3096E-12 | 47.27 |
| *KDM3A* | rs12588969 | G | C | 0.684 | -0.059 | 0.008 | 6.3096E-13 | 54.39 |
| *KDR* | rs11682865 | G | C | 0.1946 | -0.063 | 0.008 | 3.1623E-14 | 62.02 |
| *KDR* | rs10935473 | G | T | 0.44 | -0.158 | 0.007 | 5.012E-124 | 509.47 |
| *KDR* | rs34231037 | A | G | 0.0347 | -1.325 | 0.02 | 1E-200 | 4389.06 |
| *KDR* | rs9267947 | A | G | 0.4777 | -0.07 | 0.007 | 3.9811E-26 | 100.00 |
| *KDR* | rs932589 | C | T | 0.4044 | -0.056 | 0.007 | 1.5849E-16 | 64.00 |
| *KDR* | rs7012814 | G | A | 0.4724 | -0.049 | 0.007 | 2.5119E-13 | 49.00 |
| *KDR* | rs13273990 | T | C | 0.4133 | 0.046 | 0.007 | 5.0119E-12 | 43.18 |
| *KDR* | rs3110291 | A | C | 0.2013 | 0.058 | 0.008 | 3.9811E-12 | 52.56 |
| *KDR* | rs2031610 | T | C | 0.3612 | -0.05 | 0.007 | 1.9953E-13 | 51.02 |
| *KDR* | rs115974400 | T | C | 0.0681 | -0.142 | 0.013 | 6.3096E-27 | 119.31 |
| *KDR* | rs17625540 | G | A | 0.2952 | -0.05 | 0.007 | 1E-11 | 51.02 |
| *KDR* | rs35166255 | G | A | 0.0338 | 0.249 | 0.018 | 1.5849E-42 | 191.36 |
| *KDR* | rs215225 | A | G | 0.4279 | -0.062 | 0.007 | 1.2589E-20 | 78.45 |
| *KDR* | rs186021206 | G | A | 0.0064 | 0.437 | 0.045 | 5.0119E-22 | 94.31 |
| *KEL* | rs2281179 | C | T | 0.5508 | 0.047 | 0.007 | 1.2589E-12 | 45.08 |
| *KEL* | rs1427407 | T | G | 0.8477 | -0.152 | 0.009 | 7.9433E-62 | 285.23 |
| *KEL* | rs35176351 | C | A | 0.5834 | -0.314 | 0.007 | 1E-200 | 2012.16 |
| *KEL* | rs11752051 | G | A | 0.4854 | 0.069 | 0.007 | 1.9953E-25 | 97.16 |
| *KEL* | rs8176059 | G | A | 0.0115 | -1.247 | 0.032 | 1E-200 | 1518.56 |
| *KEL* | rs34009363 | T | TAA | 0.0649 | -0.093 | 0.013 | 3.1623E-12 | 51.18 |
| *KEL* | rs2519093 | C | T | 0.1838 | -0.419 | 0.009 | 1E-200 | 2167.42 |
| *KEL* | rs17476364 | T | C | 0.11 | 0.075 | 0.011 | 1.2589E-12 | 46.49 |
| *KEL* | rs11023872 | C | A | 0.0843 | 0.092 | 0.012 | 1.2589E-14 | 58.78 |
| *KEL* | rs4055121 | C | T | 0.1346 | -0.096 | 0.01 | 5.0119E-23 | 92.16 |
| *KEL* | rs35350651 | A | AC | 0.5038 | -0.05 | 0.007 | 2.5119E-14 | 51.02 |
| *KEL* | rs11630124 | T | A | 0.0228 | 0.221 | 0.022 | 1.5849E-23 | 100.91 |
| *KEL* | rs186021206 | G | A | 0.0064 | 1.337 | 0.045 | 1.259E-191 | 882.75 |
| *KEL* | rs8887 | T | C | 0.5743 | -0.065 | 0.007 | 6.3096E-22 | 86.22 |
| *KEL* | rs492602 | A | G | 0.5092 | 0.062 | 0.007 | 1.5849E-20 | 78.45 |
| *KEL* | rs6014993 | A | G | 0.4873 | 0.05 | 0.007 | 1.9953E-14 | 51.02 |
| *KHK* | rs2304681 | G | A | 0.3706 | -0.428 | 0.008 | 1E-200 | 2862.25 |
| *KHK* | rs3747207 | G | A | 0.2156 | 0.067 | 0.009 | 3.1623E-14 | 55.42 |
| *KIAA0319* | rs3181238 | C | T | 0.6398 | 0.308 | 0.008 | 1E-200 | 1482.25 |
| *KIAA2013* | rs71534539 | G | A | 0.4645 | -0.064 | 0.008 | 3.9811E-14 | 64.00 |
| *KIF1C* | rs148934699 | C | T | 0.0062 | -0.543 | 0.049 | 5.0119E-29 | 122.80 |
| *KIF22* | rs1354034 | T | C | 0.6037 | 0.059 | 0.008 | 3.9811E-14 | 54.39 |
| *KIF22* | rs368914743 | G | GCACA | 0.4952 | -0.056 | 0.008 | 2.5119E-12 | 49.00 |
| *KIF22* | rs7041446 | G | A | 0.4594 | 0.062 | 0.008 | 6.3096E-16 | 60.06 |
| *KIF22* | rs138828400 | C | T | 0.0029 | -1.05 | 0.07 | 1E-50 | 225.00 |
| *KIFBP* | rs1354034 | T | C | 0.6046 | 0.107 | 0.008 | 2.5119E-44 | 178.89 |
| *KIFBP* | rs342293 | C | G | 0.4591 | -0.081 | 0.008 | 7.9433E-27 | 102.52 |
| *KIFBP* | rs11502185 | T | C | 0.2596 | 0.07 | 0.01 | 3.9811E-13 | 49.00 |
| *KIFBP* | rs58603603 | A | G | 0.0769 | -0.102 | 0.014 | 5.0119E-13 | 53.08 |
| *KIR2DL2* | rs30378 | A | G | 0.6673 | -0.048 | 0.007 | 5.0119E-13 | 47.02 |
| *KIR2DL2* | rs9264665 | T | C | 0.6473 | -0.557 | 0.007 | 1E-200 | 6331.61 |

| | | | | | | | | |
|---|---|---|---|---|---|---|---|---|
| *KIR2DL2* | rs34992608 | G | A | 0.1234 | -0.087 | 0.009 | 3.9811E-20 | 93.44 |
| *KIR2DL2* | rs10774625 | A | G | 0.504 | -0.07 | 0.006 | 1E-29 | 136.11 |
| *KIR2DL2* | rs60955532 | C | A | 0.5597 | 0.62 | 0.007 | 1E-200 | 7844.90 |
| *KIR2DL3* | rs1050451 | C | G | 0.6476 | -0.543 | 0.008 | 1E-200 | 4607.02 |
| *KIR2DL3* | rs35522829 | G | T | 0.1235 | -0.096 | 0.01 | 3.9811E-21 | 92.16 |
| *KIR2DL3* | rs7137828 | C | T | 0.5158 | -0.073 | 0.007 | 1E-27 | 108.76 |
| *KIR2DL3* | rs16985907 | G | C | 0.038 | -0.879 | 0.018 | 1E-200 | 2384.69 |
| *KIR2DS4* | rs9264261 | T | C | 0.7146 | -0.054 | 0.006 | 1.5849E-20 | 81.00 |
| *KIR2DS4* | rs11513729 | C | T | 0.4135 | 0.034 | 0.005 | 7.9433E-12 | 46.24 |
| *KIR2DS4* | rs7255036 | G | C | 0.2258 | 1.325 | 0.009 | 1E-200 | 21674.38 |
| *KIR3DL1* | rs9259983 | C | A | 0.035 | 0.528 | 0.014 | 1E-200 | 1422.37 |
| *KIR3DL1* | rs12150998 | A | G | 0.5643 | -0.926 | 0.008 | 1E-200 | 13398.06 |
| *KIR3DL2* | rs150240916 | G | A | 0.0338 | 0.341 | 0.022 | 7.9433E-55 | 240.25 |
| *KIRREL2* | rs6571015 | G | A | 0.791 | 0.152 | 0.009 | 1.5849E-63 | 285.23 |
| *KIRREL2* | rs8176743 | C | T | 0.0615 | 0.542 | 0.015 | 1E-200 | 1305.62 |
| *KIRREL2* | rs421195 | C | T | 0.1078 | -0.306 | 0.012 | 1.995E-143 | 650.25 |
| *KIRREL2* | rs8045196 | T | G | 0.731 | -0.109 | 0.008 | 2.5119E-38 | 185.64 |
| *KIRREL2* | rs1673931 | T | C | 0.3785 | 0.067 | 0.008 | 7.9433E-19 | 70.14 |
| *KIRREL2* | rs186021206 | G | A | 0.0064 | 0.492 | 0.05 | 1E-22 | 96.83 |
| *KIRREL2* | rs404299 | G | A | 0.0333 | -0.459 | 0.021 | 1E-110 | 477.73 |
| *KIRREL2* | rs2638280 | G | A | 0.5367 | -0.066 | 0.007 | 1E-18 | 88.90 |
| *KIT* | rs115340020 | G | A | 0.0284 | -0.174 | 0.021 | 3.9811E-16 | 68.65 |
| *KIT* | rs2884425 | C | T | 0.4937 | -0.07 | 0.007 | 1.2589E-22 | 100.00 |
| *KIT* | rs74227709 | G | A | 0.0735 | 0.16 | 0.014 | 1.2589E-31 | 130.61 |
| *KIT* | rs12470153 | G | C | 0.6602 | -0.054 | 0.008 | 1E-12 | 45.56 |
| *KIT* | rs6795463 | G | T | 0.4311 | -0.06 | 0.007 | 5.0119E-17 | 73.47 |
| *KIT* | rs6440573 | C | G | 0.9503 | 0.252 | 0.016 | 5.0119E-53 | 248.06 |
| *KIT* | rs6762074 | G | A | 0.3335 | -0.079 | 0.008 | 1.2589E-25 | 97.52 |
| *KIT* | rs218263 | C | T | 0.1238 | -0.08 | 0.011 | 1.2589E-13 | 52.89 |
| *KIT* | rs72704195 | A | G | 0.3773 | -0.054 | 0.008 | 6.3096E-13 | 45.56 |
| *KIT* | rs12203592 | C | T | 0.217 | -0.108 | 0.009 | 3.9811E-34 | 144.00 |
| *KIT* | rs2844572 | A | C | 0.3431 | -0.058 | 0.007 | 6.3096E-15 | 68.65 |
| *KIT* | rs12350739 | G | A | 0.6123 | -0.137 | 0.007 | 1.5849E-77 | 383.04 |
| *KIT* | rs10118718 | G | A | 0.0997 | 0.087 | 0.012 | 3.9811E-13 | 52.56 |
| *KIT* | rs7098111 | C | T | 0.1668 | 0.112 | 0.01 | 5.0119E-31 | 125.44 |
| *KIT* | rs3008336 | T | C | 0.8357 | -0.072 | 0.01 | 5.0119E-14 | 51.84 |
| *KIT* | rs35056404 | G | A | 0.0609 | 0.181 | 0.015 | 1E-33 | 145.60 |
| *KIT* | rs948055 | G | C | 0.5226 | -0.073 | 0.007 | 1.9953E-24 | 108.76 |
| *KIT* | rs12316107 | C | A | 0.2441 | 0.072 | 0.008 | 6.3096E-18 | 81.00 |
| *KIT* | rs138854007 | C | G | 0.0753 | -0.121 | 0.014 | 6.3096E-17 | 74.70 |
| *KIT* | rs746028214 | TATTA | T | 0.6308 | -0.055 | 0.007 | 5.0119E-14 | 61.73 |
| *KIT* | rs562342198 | A | G | 0.0405 | -0.171 | 0.019 | 1.2589E-19 | 81.00 |
| *KIT* | rs1805007 | C | T | 0.1059 | -0.151 | 0.012 | 2.5119E-39 | 158.34 |
| *KIT* | rs17758695 | C | T | 0.0293 | -0.238 | 0.021 | 1.2589E-29 | 128.44 |
| *KIT* | rs78744187 | C | T | 0.0815 | 0.324 | 0.013 | 5.012E-137 | 621.16 |
| *KIT* | rs2248555 | A | G | 0.5033 | 0.099 | 0.007 | 6.3096E-44 | 200.02 |
| *KITLG* | rs590820 | A | G | 0.417 | 0.052 | 0.007 | 3.9811E-12 | 55.18 |
| *KITLG* | rs705379 | G | A | 0.4776 | 0.081 | 0.007 | 1E-27 | 133.90 |
| *KITLG* | rs983309 | T | G | 0.8829 | 0.085 | 0.011 | 7.9433E-14 | 59.71 |
| *KITLG* | rs112875651 | G | A | 0.391 | 0.067 | 0.008 | 1E-18 | 70.14 |
| *KITLG* | rs2740488 | A | C | 0.2666 | -0.103 | 0.008 | 1.9953E-35 | 165.77 |
| *KITLG* | rs507666 | G | A | 0.1845 | -0.072 | 0.009 | 5.0119E-14 | 64.00 |
| *KITLG* | rs174564 | A | G | 0.3488 | 0.068 | 0.008 | 7.9433E-19 | 72.25 |
| *KITLG* | rs673335 | T | C | 0.1553 | 0.082 | 0.01 | 5.0119E-16 | 67.24 |
| *KITLG* | rs11105121 | C | T | 0.6625 | -0.054 | 0.008 | 3.9811E-12 | 45.56 |
| *KITLG* | rs150844304 | A | C | 0.0241 | -0.188 | 0.024 | 2.5119E-15 | 61.36 |
| *KITLG* | rs247617 | C | A | 0.3239 | 0.135 | 0.008 | 7.9433E-66 | 284.77 |
| *KITLG* | rs2292318 | C | T | 0.1237 | 0.157 | 0.011 | 1.9953E-44 | 203.71 |
| *KITLG* | rs34931250 | C | T | 0.0603 | -0.113 | 0.015 | 1.9953E-13 | 56.75 |
| *KITLG* | rs1645788 | A | G | 0.2325 | 0.105 | 0.009 | 1E-33 | 136.11 |
| *KITLG* | rs2868346 | C | T | 0.7594 | -0.31 | 0.009 | 1E-200 | 1186.42 |
| *KLB* | rs114165349 | G | C | 0.0233 | -0.147 | 0.021 | 2.5119E-12 | 49.00 |
| *KLB* | rs13108218 | A | G | 0.6183 | 0.052 | 0.008 | 1.5849E-11 | 42.25 |
| *KLB* | rs13103023 | G | A | 0.3276 | -0.762 | 0.008 | 1E-200 | 9072.56 |
| *KLB* | rs3135911 | C | A | 0.2907 | -0.166 | 0.007 | 7.943E-122 | 562.37 |
| *KLB* | rs35254320 | C | T | 0.1468 | 0.077 | 0.009 | 1E-17 | 73.20 |
| *KLB* | rs28929474 | C | T | 0.0207 | 0.325 | 0.022 | 3.9811E-48 | 218.23 |
| *KLB* | rs60134803 | A | G | 0.2352 | 0.053 | 0.008 | 7.9433E-12 | 43.89 |

| | | | | | | | | |
|---|---|---|---|---|---|---|---|---|
| *KLB* | rs112001035 | G | A | 0.0613 | -0.667 | 0.014 | 1E-200 | 2269.84 |
| *KLB* | rs41292412 | C | T | 0.0126 | -0.199 | 0.029 | 6.3096E-12 | 47.09 |
| *KLB* | rs149131600 | C | T | 0.1752 | 0.065 | 0.008 | 1E-14 | 66.02 |
| *KLF4* | rs7080536 | G | A | 0.0435 | -0.186 | 0.019 | 1E-22 | 95.83 |
| *KLF4* | rs10418046 | T | G | 0.2114 | 0.065 | 0.009 | 3.1623E-12 | 52.16 |
| *KLK1* | rs12758813 | G | A | 0.3987 | 0.041 | 0.006 | 3.9811E-13 | 46.69 |
| *KLK1* | rs2576570 | A | C | 0.5769 | -0.055 | 0.006 | 5.0119E-22 | 84.03 |
| *KLK1* | rs423144 | G | T | 0.4257 | 0.048 | 0.006 | 1.5849E-17 | 64.00 |
| *KLK1* | rs35275076 | T | C | 0.5464 | -0.047 | 0.006 | 3.1623E-16 | 61.36 |
| *KLK1* | rs11889674 | G | A | 0.3918 | 0.047 | 0.006 | 7.9433E-17 | 61.36 |
| *KLK1* | rs2170582 | A | G | 0.6705 | -0.041 | 0.006 | 3.9811E-12 | 46.69 |
| *KLK1* | rs3749751 | C | T | 0.5913 | 0.039 | 0.006 | 1E-11 | 42.25 |
| *KLK1* | rs8176693 | C | T | 0.0614 | 0.175 | 0.012 | 3.9811E-52 | 212.67 |
| *KLK1* | rs3862630 | C | T | 0.1225 | 0.217 | 0.009 | 1E-134 | 581.35 |
| *KLK1* | rs7397745 | T | C | 0.5845 | 0.049 | 0.006 | 3.9811E-18 | 66.69 |
| *KLK1* | rs9529913 | C | T | 0.6036 | 0.061 | 0.006 | 1.2589E-26 | 103.36 |
| *KLK1* | rs55714927 | C | T | 0.1949 | 0.066 | 0.007 | 6.3096E-21 | 88.90 |
| *KLK1* | rs601338 | G | A | 0.5092 | 0.889 | 0.008 | 1E-200 | 12348.77 |
| *KLK1* | rs66464836 | C | G | 0.1119 | -0.071 | 0.009 | 6.3096E-16 | 62.23 |
| *KLK10* | rs4576692 | C | T | 0.6761 | -0.058 | 0.007 | 1.5849E-15 | 68.65 |
| *KLK10* | rs2023472 | A | G | 0.588 | 0.047 | 0.007 | 7.9433E-12 | 45.08 |
| *KLK10* | rs375524395 | A | G | 0.4557 | 0.049 | 0.007 | 1E-12 | 49.00 |
| *KLK10* | rs139130389 | C | CTA | 0.0726 | -0.189 | 0.013 | 7.9433E-48 | 211.37 |
| *KLK10* | rs1065853 | G | T | 0.0815 | 0.094 | 0.014 | 1.2589E-11 | 45.08 |
| *KLK10* | rs2569454 | T | C | 0.5783 | 0.593 | 0.008 | 1E-200 | 5494.52 |
| *KLK11* | rs34255800 | G | C | 0.1541 | -0.23 | 0.01 | 1.995E-114 | 529.00 |
| *KLK11* | rs369705328 | G | C | 0.1283 | 0.078 | 0.011 | 6.3096E-13 | 50.28 |
| *KLK11* | rs139130389 | C | CTA | 0.0727 | -0.453 | 0.014 | 1E-200 | 1046.98 |
| *KLK11* | rs36060036 | C | T | 0.1671 | -0.069 | 0.01 | 1.2589E-12 | 47.61 |
| *KLK11* | rs186021206 | G | A | 0.0064 | 0.475 | 0.049 | 5.0119E-22 | 93.97 |
| *KLK11* | rs117268623 | C | T | 0.0325 | -1.147 | 0.021 | 1E-200 | 2983.24 |
| *KLK11* | rs12986064 | T | C | 0.5081 | -0.051 | 0.008 | 1.5849E-11 | 40.64 |
| *KLK12* | rs2479016 | A | G | 0.4274 | -0.068 | 0.005 | 1.9953E-50 | 184.96 |
| *KLK12* | rs11220477 | C | T | 0.0418 | -0.133 | 0.011 | 3.9811E-33 | 146.19 |
| *KLK12* | rs79744308 | G | A | 0.0415 | 0.141 | 0.019 | 3.9811E-13 | 55.07 |
| *KLK12* | rs3745540 | A | G | 0.5822 | -1.167 | 0.008 | 1E-200 | 21279.52 |
| *KLK13* | rs12995525 | T | G | 0.4961 | 0.061 | 0.007 | 3.9811E-18 | 75.94 |
| *KLK13* | rs2479016 | A | G | 0.4274 | -0.058 | 0.007 | 7.9433E-16 | 68.65 |
| *KLK13* | rs529700 | T | C | 0.5143 | 0.056 | 0.008 | 1.2589E-11 | 49.00 |
| *KLK13* | rs226379 | T | C | 0.3555 | 0.077 | 0.007 | 7.9433E-26 | 121.00 |
| *KLK13* | rs5511 | A | T | 0.2199 | 0.142 | 0.008 | 2.5119E-63 | 315.06 |
| *KLK13* | rs55707100 | C | T | 0.0242 | -0.168 | 0.023 | 1E-13 | 53.35 |
| *KLK13* | rs186021206 | G | A | 0.0063 | 0.64 | 0.048 | 7.9433E-40 | 177.78 |
| *KLK13* | rs75181977 | A | G | 0.1025 | 0.098 | 0.012 | 2.5119E-17 | 66.69 |
| *KLK13* | rs78060698 | G | A | 0.0408 | 0.187 | 0.019 | 5.0119E-23 | 96.87 |
| *KLK13* | rs681343 | C | T | 0.5086 | -0.074 | 0.008 | 7.9433E-23 | 85.56 |
| *KLK13* | rs3760739 | G | T | 0.3502 | -0.454 | 0.008 | 1E-200 | 3220.56 |
| *KLK14* | rs61816761 | G | A | 0.0236 | 0.169 | 0.024 | 1.5849E-12 | 49.59 |
| *KLK14* | rs58741371 | T | C | 0.1969 | 0.064 | 0.009 | 1.9953E-13 | 50.57 |
| *KLK14* | rs6880842 | A | G | 0.3847 | 0.048 | 0.007 | 1.5849E-11 | 47.02 |
| *KLK14* | rs11249893 | C | T | 0.4766 | 0.052 | 0.007 | 1.9953E-14 | 55.18 |
| *KLK14* | rs60978445 | T | C | 0.187 | -0.072 | 0.009 | 5.0119E-16 | 64.00 |
| *KLK14* | rs635634 | C | T | 0.1841 | -0.076 | 0.009 | 1.2589E-17 | 71.31 |
| *KLK14* | rs11599750 | C | T | 0.4092 | -0.062 | 0.007 | 2.5119E-19 | 78.45 |
| *KLK14* | rs61729512 | G | A | 0.1178 | 0.136 | 0.011 | 6.3096E-37 | 152.86 |
| *KLK14* | rs1863115 | C | A | 0.742 | -0.069 | 0.008 | 3.1623E-17 | 74.39 |
| *KLK14* | rs2569491 | G | A | 0.2985 | 0.416 | 0.008 | 1E-200 | 2704.00 |
| *KLK14* | rs12986064 | T | C | 0.5077 | -0.068 | 0.008 | 5.0119E-19 | 72.25 |
| *KLK14* | rs1276365 | G | A | 0.4422 | -0.047 | 0.007 | 1.5849E-11 | 45.08 |
| *KLK15* | rs7560701 | G | C | 0.4993 | -0.063 | 0.006 | 2.5119E-29 | 110.25 |
| *KLK15* | rs4536908 | C | T | 0.4606 | 0.039 | 0.006 | 2.5119E-12 | 42.25 |
| *KLK15* | rs72840032 | C | T | 0.043 | -0.104 | 0.014 | 6.3096E-14 | 55.18 |
| *KLK15* | rs626277 | A | C | 0.3963 | -0.044 | 0.006 | 1.2589E-14 | 53.78 |
| *KLK15* | rs73048483 | G | A | 0.0738 | 1.415 | 0.015 | 1E-200 | 8898.78 |
| *KLK3* | rs62113212 | C | T | 0.0745 | -0.105 | 0.009 | 1.2589E-31 | 136.11 |
| *KLK4* | rs1572993 | G | A | 0.4146 | -0.048 | 0.006 | 1.9953E-15 | 64.00 |
| *KLK4* | rs28929474 | C | T | 0.0208 | -0.441 | 0.021 | 2.512E-99 | 441.00 |
| *KLK4* | rs2664153 | G | A | 0.6388 | -0.418 | 0.007 | 1E-200 | 3565.80 |

| | | | | | | | | |
|---|---|---|---|---|---|---|---|---|
| *KLK6* | rs9933330 | C | T | 0.1816 | -0.066 | 0.01 | 5.0119E-12 | 43.56 |
| *KLK6* | rs268891 | A | C | 0.651 | -0.192 | 0.008 | 2.512E-131 | 576.00 |
| *KLK6* | rs12986064 | T | C | 0.5079 | -0.069 | 0.008 | 3.9811E-19 | 74.39 |
| *KLK7* | rs80293268 | G | C | 0.0484 | -0.157 | 0.017 | 1.9953E-19 | 85.29 |
| *KLK7* | rs61816761 | G | A | 0.0238 | 0.244 | 0.025 | 3.9811E-23 | 95.26 |
| *KLK7* | rs61729512 | G | A | 0.1182 | 0.217 | 0.011 | 1.2589E-84 | 389.17 |
| *KLK7* | rs2353482 | G | A | 0.7254 | -0.058 | 0.008 | 3.1623E-13 | 52.56 |
| *KLK7* | rs1991821 | A | G | 0.9101 | 0.693 | 0.013 | 1E-200 | 2841.71 |
| *KLK7* | rs12986064 | T | C | 0.5073 | -0.182 | 0.008 | 5.012E-120 | 517.56 |
| *KLK8* | rs61816761 | G | A | 0.0235 | 0.324 | 0.024 | 3.9811E-41 | 182.25 |
| *KLK8* | rs121908120 | T | A | 0.0276 | -0.155 | 0.021 | 3.9811E-13 | 54.48 |
| *KLK8* | rs186021206 | G | A | 0.0063 | 0.39 | 0.048 | 2.5119E-16 | 66.02 |
| *KLK8* | rs74705037 | G | A | 0.0455 | 1.051 | 0.018 | 1E-200 | 3409.26 |
| *KLK8* | rs12986064 | T | C | 0.5076 | -0.061 | 0.008 | 1.9953E-15 | 58.14 |
| *KLKB1* | rs1260326 | T | C | 0.608 | -0.102 | 0.007 | 1.5849E-49 | 212.33 |
| *KLKB1* | rs710446 | T | C | 0.4088 | 0.354 | 0.007 | 1E-200 | 2557.47 |
| *KLKB1* | rs4253306 | G | T | 0.1004 | -0.562 | 0.012 | 1E-200 | 2193.36 |
| *KLKB1* | rs2731674 | T | G | 0.7445 | -0.084 | 0.008 | 1.2589E-27 | 110.25 |
| *KLKB1* | rs451637 | C | T | 0.3097 | -0.056 | 0.007 | 6.3096E-14 | 64.00 |
| *KLKB1* | rs738408 | C | T | 0.2166 | -0.059 | 0.008 | 5.0119E-13 | 54.39 |
| *KLRB1* | rs4149909 | A | G | 0.0321 | 0.152 | 0.021 | 2.5119E-13 | 52.39 |
| *KLRB1* | rs6759003 | T | C | 0.6397 | 0.081 | 0.008 | 1.9953E-26 | 102.52 |
| *KLRB1* | rs2009581 | G | A | 0.2724 | -0.072 | 0.008 | 1.2589E-18 | 81.00 |
| *KLRB1* | rs4449583 | C | T | 0.3268 | 0.072 | 0.008 | 5.0119E-20 | 81.00 |
| *KLRB1* | rs746778623 | TGGGAACA | T | 0.3326 | -0.054 | 0.008 | 5.0119E-12 | 45.56 |
| *KLRB1* | rs2249932 | T | C | 0.2975 | 0.085 | 0.008 | 1.9953E-25 | 112.89 |
| *KLRB1* | rs900567 | C | T | 0.7525 | -0.064 | 0.009 | 3.9811E-14 | 50.57 |
| *KLRB1* | rs59602812 | A | AT | 0.3179 | -0.069 | 0.008 | 6.3096E-19 | 74.39 |
| *KLRB1* | rs7086174 | C | A | 0.3283 | 0.063 | 0.008 | 7.9433E-16 | 62.02 |
| *KLRB1* | rs563890415 | T | C | 0.0063 | -1.13 | 0.047 | 1.995E-130 | 578.04 |
| *KLRB1* | rs3184504 | T | C | 0.5174 | -0.126 | 0.007 | 6.3096E-67 | 324.00 |
| *KLRB1* | rs12437803 | G | A | 0.193 | -0.184 | 0.009 | 1.5849E-85 | 417.98 |
| *KLRB1* | rs11071528 | G | C | 0.8052 | 0.079 | 0.009 | 1E-17 | 77.05 |
| *KLRD1* | rs2009581 | G | A | 0.2724 | -0.111 | 0.008 | 1.9953E-47 | 192.52 |
| *KLRD1* | rs13063578 | T | A | 0.3978 | -0.079 | 0.007 | 6.3096E-30 | 127.37 |
| *KLRD1* | rs2706385 | T | G | 0.6616 | 0.055 | 0.007 | 3.1623E-14 | 61.73 |
| *KLRD1* | rs1655895 | T | C | 0.853 | -0.273 | 0.01 | 3.162E-173 | 745.29 |
| *KLRD1* | rs12684650 | C | T | 0.3192 | -0.054 | 0.007 | 1E-13 | 59.51 |
| *KLRD1* | rs2537752 | T | A | 0.5994 | -0.418 | 0.007 | 1E-200 | 3565.80 |
| *KLRD1* | rs3184504 | T | C | 0.5174 | -0.103 | 0.007 | 1.9953E-47 | 216.51 |
| *KLRD1* | rs11869875 | G | A | 0.3255 | -0.063 | 0.007 | 3.1623E-18 | 81.00 |
| *KLRD1* | rs3744283 | G | C | 0.1598 | -0.069 | 0.009 | 6.3096E-14 | 58.78 |
| *KLRD1* | rs875622 | A | G | 0.7682 | 0.055 | 0.008 | 1E-11 | 47.27 |
| *KLRD1* | rs28873836 | G | C | 0.2662 | -0.24 | 0.008 | 1E-200 | 900.00 |
| *KLRD1* | rs35545130 | C | G | 0.3223 | -0.064 | 0.008 | 3.9811E-15 | 64.00 |
| *KLRF1* | rs2643902 | T | C | 0.7879 | -0.065 | 0.009 | 3.9811E-13 | 52.16 |
| *KLRF1* | rs183053322 | C | T | 0.012 | -0.259 | 0.034 | 5.0119E-14 | 58.03 |
| *KLRF1* | rs2009581 | G | A | 0.2722 | -0.057 | 0.008 | 1.9953E-12 | 50.77 |
| *KLRF1* | rs13063578 | T | A | 0.3982 | -0.061 | 0.007 | 1.5849E-16 | 75.94 |
| *KLRF1* | rs2522051 | T | C | 0.4522 | -0.065 | 0.007 | 1.2589E-18 | 86.22 |
| *KLRF1* | rs8177653 | A | G | 0.3348 | 0.064 | 0.008 | 7.9433E-17 | 64.00 |
| *KLRF1* | rs513365 | A | G | 0.2882 | -0.073 | 0.008 | 5.0119E-20 | 83.27 |
| *KLRF1* | rs146460124 | G | A | 0.0017 | 2.67 | 0.088 | 3.162E-200 | 920.57 |
| *KLRF1* | rs3184504 | T | C | 0.5171 | -0.109 | 0.007 | 6.3096E-51 | 242.47 |
| *KLRF1* | rs9916257 | G | T | 0.4386 | -0.114 | 0.007 | 3.9811E-55 | 265.22 |
| *KLRF1* | rs4968607 | G | A | 0.1591 | -0.077 | 0.01 | 7.9433E-15 | 59.29 |
| *KLRF1* | rs150358287 | C | A | 0.0119 | 0.267 | 0.033 | 1.2589E-15 | 65.46 |
| *KLRF1* | rs35915710 | C | A | 0.1679 | -0.071 | 0.01 | 1.5849E-13 | 50.41 |
| *KLRK1* | rs4672505 | A | G | 0.6393 | 0.106 | 0.007 | 6.3096E-47 | 229.31 |
| *KLRK1* | rs2009581 | G | A | 0.2722 | -0.12 | 0.008 | 1E-51 | 225.00 |
| *KLRK1* | rs13063578 | T | A | 0.398 | -0.094 | 0.007 | 3.9811E-39 | 180.33 |
| *KLRK1* | rs2844512 | C | G | 0.6041 | 0.07 | 0.007 | 2.5119E-22 | 100.00 |
| *KLRK1* | rs756546977 | ACTAAT | A | 0.7155 | -0.533 | 0.008 | 1E-200 | 4438.89 |
| *KLRK1* | rs10774624 | G | A | 0.5135 | -0.085 | 0.008 | 3.9811E-29 | 112.89 |
| *KLRK1* | rs186021206 | G | A | 0.0065 | 0.4 | 0.048 | 7.9433E-17 | 69.44 |
| *KLRK1* | rs3803813 | T | C | 0.1589 | -0.074 | 0.01 | 1.9953E-14 | 54.76 |
| *KLRK1* | rs2659005 | C | T | 0.4408 | 0.094 | 0.007 | 3.9811E-40 | 180.33 |
| *KRT18* | rs142552223 | G | A | 0.0374 | -0.129 | 0.019 | 1.2589E-11 | 46.10 |

| | | | | | | | | |
|---|---|---|---|---|---|---|---|---|
| *KRT18* | rs7678352 | C | T | 0.1636 | 0.072 | 0.01 | 7.9433E-14 | 51.84 |
| *KRT18* | rs7043196 | C | T | 0.4968 | -0.052 | 0.007 | 6.3096E-13 | 55.18 |
| *KRT18* | rs35696875 | T | TCA | 0.6978 | 0.053 | 0.008 | 1.5849E-11 | 43.89 |
| *KRT18* | rs10883451 | T | C | 0.5017 | -0.054 | 0.007 | 3.9811E-14 | 59.51 |
| *KRT18* | rs11601507 | C | A | 0.0674 | 0.196 | 0.014 | 1.2589E-43 | 196.00 |
| *KRT18* | rs28929474 | C | T | 0.0207 | 0.201 | 0.025 | 1.5849E-15 | 64.64 |
| *KRT18* | rs132642 | A | T | 0.8301 | 0.071 | 0.01 | 7.9433E-14 | 50.41 |
| *KRT18* | rs738409 | C | G | 0.2172 | 0.102 | 0.009 | 6.3096E-32 | 128.44 |
| *KRT19* | rs7313347 | G | A | 0.594 | -0.051 | 0.008 | 1E-11 | 40.64 |
| *KRT19* | rs7405611 | G | A | 0.6228 | 0.147 | 0.008 | 1E-81 | 337.64 |
| *KRT19* | rs601338 | G | A | 0.5091 | -0.059 | 0.007 | 5.0119E-15 | 71.04 |
| *KRT5* | rs35661734 | T | C | 0.0387 | 0.278 | 0.019 | 3.1623E-49 | 214.08 |
| *KRT5* | rs210135 | T | A | 0.6999 | -0.077 | 0.008 | 3.9811E-22 | 92.64 |
| *KRT5* | rs73015965 | A | G | 0.0064 | -0.693 | 0.049 | 7.9433E-45 | 200.02 |
| *KRT5* | rs607860 | C | T | 0.4066 | -0.101 | 0.007 | 5.0119E-42 | 208.18 |
| *KRT5* | rs7240884 | A | G | 0.4223 | 0.085 | 0.007 | 1.2589E-30 | 147.45 |
| *KYAT1* | rs138429240 | C | T | 0.0049 | 0.671 | 0.053 | 3.1623E-36 | 160.29 |
| *KYAT1* | rs3811444 | C | T | 0.3321 | -0.054 | 0.008 | 7.9433E-12 | 45.56 |
| *KYAT1* | rs1354034 | T | C | 0.6038 | 0.062 | 0.008 | 5.0119E-16 | 60.06 |
| *KYAT1* | rs35072117 | T | TG | 0.4609 | 0.056 | 0.008 | 1.2589E-12 | 49.00 |
| *KYAT1* | rs6993770 | A | T | 0.2865 | -0.058 | 0.008 | 1.5849E-12 | 52.56 |
| *KYAT1* | rs60822569 | T | TTTCTC | 0.5541 | 0.061 | 0.008 | 1E-15 | 58.14 |
| *KYNU* | rs78189771 | A | G | 0.0477 | 0.57 | 0.017 | 1E-200 | 1124.22 |
| *KYNU* | rs201117812 | G | A | 0.171 | -0.077 | 0.011 | 3.1623E-13 | 49.00 |
| *KYNU* | rs113609637 | A | AT | 0.1414 | -0.071 | 0.01 | 7.9433E-12 | 50.41 |
| *KYNU* | rs10846740 | T | C | 0.859 | -0.085 | 0.011 | 3.9811E-16 | 59.71 |
| *KYNU* | rs200210321 | A | AG | 0.0741 | 0.105 | 0.014 | 1.9953E-14 | 56.25 |
| *KYNU* | rs3747207 | G | A | 0.2157 | 0.104 | 0.009 | 5.0119E-33 | 133.53 |
| *L1CAM* | rs3791101 | G | A | 0.3078 | 0.067 | 0.008 | 3.9811E-18 | 70.14 |
| *L1CAM* | rs1260326 | T | C | 0.6087 | 0.06 | 0.007 | 1.9953E-16 | 73.47 |
| *L1CAM* | rs56278466 | T | G | 0.6612 | 0.054 | 0.007 | 3.9811E-13 | 59.51 |
| *L1CAM* | rs139130389 | C | CTA | 0.0726 | -0.307 | 0.014 | 1E-111 | 480.86 |
| *L1CAM* | rs3781781 | T | C | 0.3162 | -0.104 | 0.008 | 1.9953E-42 | 169.00 |
| *L1CAM* | rs186021206 | G | A | 0.0064 | 1.298 | 0.049 | 1.995E-156 | 701.71 |
| *L1CAM* | rs35866622 | C | T | 0.5 | 0.049 | 0.007 | 1E-11 | 49.00 |
| *L1CAM* | rs5750830 | C | A | 0.7487 | -0.085 | 0.008 | 2.5119E-25 | 112.89 |
| *L3HYPDH* | rs1354034 | T | C | 0.6045 | 0.057 | 0.008 | 1E-13 | 50.77 |
| *L3HYPDH* | rs11950562 | A | C | 0.4779 | -0.053 | 0.008 | 3.1623E-12 | 43.89 |
| *L3HYPDH* | rs342293 | C | G | 0.4592 | -0.058 | 0.008 | 2.5119E-14 | 52.56 |
| *L3HYPDH* | rs6993770 | A | T | 0.2866 | -0.063 | 0.008 | 3.1623E-14 | 62.02 |
| *L3HYPDH* | rs35751397 | T | TA | 0.4689 | 0.054 | 0.008 | 7.9433E-13 | 45.56 |
| *L3HYPDH* | rs2838493 | A | G | 0.8176 | -0.116 | 0.01 | 1.2589E-32 | 134.56 |
| *LACRT* | rs10462072 | C | T | 0.7799 | -0.062 | 0.009 | 6.3096E-12 | 47.46 |
| *LACRT* | rs929542 | T | C | 0.8783 | -0.189 | 0.011 | 7.9433E-61 | 295.21 |
| *LACRT* | rs11825821 | T | G | 0.3706 | -0.156 | 0.008 | 5.0119E-88 | 380.25 |
| *LACRT* | rs113293194 | A | C | 0.0696 | 0.129 | 0.015 | 2.5119E-18 | 73.96 |
| *LACRT* | rs681343 | C | T | 0.509 | 0.075 | 0.007 | 1.5849E-23 | 114.80 |
| *LACTB2* | rs1354034 | T | C | 0.6037 | 0.07 | 0.008 | 6.3096E-20 | 76.56 |
| *LACTB2* | rs191588099 | G | A | 0.0071 | -1.11 | 0.047 | 7.943E-126 | 557.76 |
| *LAG3* | rs12740374 | G | T | 0.2212 | -0.19 | 0.009 | 1E-105 | 445.68 |
| *LAG3* | rs115216147 | T | A | 0.0666 | -0.158 | 0.015 | 1E-26 | 110.95 |
| *LAG3* | rs35829610 | C | CTG | 0.3381 | -0.074 | 0.008 | 2.5119E-22 | 85.56 |
| *LAG3* | rs9264419 | G | A | 0.1104 | 0.106 | 0.011 | 3.9811E-20 | 92.86 |
| *LAG3* | rs950802 | G | A | 0.3063 | 0.053 | 0.008 | 7.9433E-12 | 43.89 |
| *LAG3* | rs1056008 | T | C | 0.2611 | -0.06 | 0.008 | 3.9811E-13 | 56.25 |
| *LAG3* | rs3782735 | G | A | 0.6025 | 0.175 | 0.008 | 1E-117 | 478.52 |
| *LAG3* | rs3184504 | T | C | 0.5168 | -0.107 | 0.007 | 1E-49 | 233.65 |
| *LAG3* | rs139974673 | T | C | 0.0244 | -0.203 | 0.023 | 3.1623E-18 | 77.90 |
| *LAG3* | rs200489612 | G | A | 0.0052 | 0.586 | 0.054 | 3.9811E-27 | 117.76 |
| *LAG3* | rs77542162 | A | G | 0.023 | 0.286 | 0.024 | 1E-32 | 142.01 |
| *LAIR1* | rs333947 | G | A | 0.1483 | 0.065 | 0.009 | 1.5849E-12 | 52.16 |
| *LAIR1* | rs3184504 | T | C | 0.5172 | -0.049 | 0.006 | 6.3096E-14 | 66.69 |
| *LAIR1* | rs77924615 | G | A | 0.1967 | -0.064 | 0.008 | 1.9953E-14 | 64.00 |
| *LAIR1* | rs111949142 | T | A | 0.1842 | 0.723 | 0.01 | 1E-200 | 5227.29 |
| *LAIR2* | rs13063578 | T | A | 0.3976 | -0.045 | 0.006 | 6.3096E-15 | 56.25 |
| *LAIR2* | rs3184504 | T | C | 0.517 | -0.066 | 0.006 | 3.9811E-31 | 121.00 |
| *LAIR2* | rs35663614 | G | GC | 0.6255 | -0.064 | 0.008 | 3.1623E-16 | 64.00 |
| *LAIR2* | rs73070113 | A | T | 0.135 | -1.06 | 0.012 | 1E-200 | 7802.78 |

| | | | | | | | | |
|---|---|---|---|---|---|---|---|---|
| *LAMA4* | rs61809091 | T | C | 0.415 | 0.083 | 0.008 | 1.9953E-24 | 107.64 |
| *LAMA4* | rs1421929 | A | G | 0.4914 | -0.05 | 0.007 | 1.5849E-11 | 51.02 |
| *LAMA4* | rs1917342 | A | G | 0.3036 | -0.056 | 0.008 | 3.1623E-12 | 49.00 |
| *LAMA4* | rs7765769 | T | C | 0.4433 | 0.167 | 0.008 | 1.259E-109 | 435.77 |
| *LAMA4* | rs41281051 | T | C | 0.4589 | -0.055 | 0.007 | 1.5849E-13 | 61.73 |
| *LAMA4* | rs6993770 | A | T | 0.2867 | -0.102 | 0.008 | 1.2589E-35 | 162.56 |
| *LAMA4* | rs10761731 | A | T | 0.4148 | 0.088 | 0.008 | 1E-31 | 121.00 |
| *LAMA4* | rs61729512 | G | A | 0.1179 | 0.087 | 0.012 | 3.9811E-14 | 52.56 |
| *LAMA4* | rs13339274 | C | T | 0.1442 | 0.08 | 0.01 | 1.9953E-14 | 64.00 |
| *LAMA4* | rs143040759 | G | A | 0.0212 | 0.614 | 0.026 | 3.162E-125 | 557.69 |
| *LAMA4* | rs892090 | T | G | 0.8342 | 0.126 | 0.01 | 2.5119E-36 | 158.76 |
| *LAMA4* | rs4813351 | C | T | 0.3707 | 0.052 | 0.008 | 7.9433E-12 | 42.25 |
| *LAMB1* | rs11683120 | A | G | 0.5493 | -0.098 | 0.006 | 5.0119E-53 | 266.78 |
| *LAMB1* | rs13107325 | C | T | 0.075 | 0.085 | 0.012 | 2.5119E-12 | 50.17 |
| *LAMB1* | rs2910686 | T | C | 0.4495 | 0.134 | 0.006 | 1E-96 | 498.78 |
| *LAMB1* | rs9268833 | C | T | 0.3173 | 0.076 | 0.007 | 2.5119E-28 | 117.88 |
| *LAMB1* | rs436573 | A | T | 0.4253 | -0.637 | 0.008 | 1E-200 | 6340.14 |
| *LAMB1* | rs3124776 | T | C | 0.4788 | -0.047 | 0.006 | 1.5849E-13 | 61.36 |
| *LAMB1* | rs1366 | A | G | 0.4489 | 0.047 | 0.006 | 1.5849E-13 | 61.36 |
| *LAMB1* | rs1609860 | C | A | 0.0812 | 0.117 | 0.012 | 7.9433E-24 | 95.06 |
| *LAMB1* | rs112635299 | G | T | 0.0216 | 0.148 | 0.022 | 1.2589E-11 | 45.26 |
| *LAMB1* | rs11075921 | C | T | 0.1597 | 0.081 | 0.009 | 7.9433E-21 | 81.00 |
| *LAMB1* | rs55799523 | C | A | 0.2658 | 0.09 | 0.007 | 6.3096E-34 | 165.31 |
| *LAMP1* | rs9857570 | A | G | 0.3243 | -0.056 | 0.008 | 1E-11 | 49.00 |
| *LAMP1* | rs9270664 | G | A | 0.6362 | -0.056 | 0.008 | 1.5849E-12 | 49.00 |
| *LAMP1* | rs1293303 | G | C | 0.4458 | 0.1 | 0.008 | 3.9811E-38 | 156.25 |
| *LAMP1* | rs74612335 | T | C | 0.1333 | -0.088 | 0.011 | 6.3096E-15 | 64.00 |
| *LAMP1* | rs9549739 | G | A | 0.2922 | 0.067 | 0.009 | 1E-14 | 55.42 |
| *LAMP1* | rs34843303 | T | C | 0.1119 | -0.11 | 0.012 | 1.9953E-19 | 84.03 |
| *LAMP1* | rs186021206 | G | A | 0.0064 | 0.405 | 0.052 | 1E-14 | 60.66 |
| *LAMP2* | rs10908419 | G | A | 0.4917 | -0.053 | 0.007 | 1.5849E-12 | 57.33 |
| *LAMP2* | rs41274050 | C | T | 0.0099 | 0.267 | 0.038 | 1.2589E-12 | 49.37 |
| *LAMP3* | rs11921884 | G | T | 0.6491 | 0.233 | 0.008 | 1E-200 | 848.27 |
| *LAMP3* | rs13107325 | C | T | 0.075 | 0.101 | 0.013 | 6.3096E-14 | 60.36 |
| *LAMP3* | rs9263519 | G | A | 0.3268 | -0.061 | 0.008 | 6.3096E-16 | 58.14 |
| *LAMP3* | rs507666 | G | A | 0.1846 | -0.071 | 0.009 | 7.9433E-15 | 62.23 |
| *LAMP3* | rs35705950 | G | T | 0.1102 | 0.082 | 0.011 | 3.9811E-13 | 55.57 |
| *LAMP3* | rs7962469 | A | G | 0.6456 | 0.054 | 0.007 | 2.5119E-13 | 59.51 |
| *LAMP3* | rs7137828 | C | T | 0.5162 | -0.061 | 0.007 | 6.3096E-18 | 75.94 |
| *LAMP3* | rs117169628 | G | A | 0.1433 | -0.09 | 0.01 | 5.0119E-19 | 81.00 |
| *LAMP3* | rs186021206 | G | A | 0.0064 | 0.435 | 0.049 | 3.1623E-19 | 78.81 |
| *LAMP3* | rs892225 | A | G | 0.3805 | -0.066 | 0.007 | 1.5849E-19 | 88.90 |
| *LARP1* | rs1354034 | T | C | 0.6044 | 0.069 | 0.008 | 1.9953E-19 | 74.39 |
| *LAT* | rs3804749 | C | T | 0.5927 | -0.059 | 0.008 | 2.5119E-14 | 54.39 |
| *LAT* | rs17622656 | G | A | 0.3884 | -0.054 | 0.008 | 3.1623E-12 | 45.56 |
| *LAT* | rs6993770 | A | T | 0.2866 | -0.094 | 0.008 | 3.9811E-30 | 138.06 |
| *LAT* | rs7080386 | C | A | 0.412 | 0.066 | 0.008 | 3.1623E-18 | 68.06 |
| *LAT* | rs892090 | T | G | 0.8341 | 0.096 | 0.01 | 1.2589E-21 | 92.16 |
| *LAT2* | rs1354034 | T | C | 0.6046 | 0.061 | 0.008 | 1E-15 | 58.14 |
| *LATS1* | rs1354034 | T | C | 0.6039 | 0.094 | 0.008 | 1.2589E-33 | 138.06 |
| *LATS1* | rs6993770 | A | T | 0.2866 | -0.058 | 0.008 | 5.0119E-12 | 52.56 |
| *LAYN* | rs61747728 | C | T | 0.0379 | 0.125 | 0.018 | 3.9811E-12 | 48.23 |
| *LAYN* | rs539726870 | A | G | 0.1179 | 0.089 | 0.011 | 3.1623E-15 | 65.46 |
| *LAYN* | rs8176719 | T | TC | 0.34 | 0.069 | 0.007 | 1.9953E-21 | 97.16 |
| *LAYN* | rs542275 | C | T | 0.6162 | 0.389 | 0.007 | 1E-200 | 3088.18 |
| *LAYN* | rs7203642 | A | G | 0.1922 | -0.068 | 0.009 | 7.9433E-15 | 57.09 |
| *LBP* | rs112001035 | G | A | 0.061 | 0.109 | 0.014 | 2.5119E-14 | 60.62 |
| *LBP* | rs573329980 | G | A | 0.1947 | 0.09 | 0.01 | 3.9811E-18 | 81.00 |
| *LBP* | rs2232613 | C | T | 0.0823 | -1.188 | 0.014 | 1E-200 | 7200.73 |
| *LBR* | rs1354034 | T | C | 0.6042 | 0.165 | 0.008 | 1E-105 | 425.39 |
| *LBR* | rs342293 | C | G | 0.4596 | -0.06 | 0.007 | 6.3096E-16 | 73.47 |
| *LBR* | rs10733789 | T | C | 0.3133 | 0.062 | 0.008 | 7.9433E-15 | 60.06 |
| *LBR* | rs7080536 | G | A | 0.0435 | -0.223 | 0.018 | 6.3096E-34 | 153.48 |
| *LBR* | rs11604127 | C | T | 0.2309 | 0.087 | 0.009 | 6.3096E-23 | 93.44 |
| *LBR* | rs10876550 | G | A | 0.558 | 0.054 | 0.007 | 6.3096E-13 | 59.51 |
| *LCAT* | rs768234885 | CA | C | 0.356 | -0.061 | 0.008 | 2.5119E-15 | 58.14 |
| *LCAT* | rs1260326 | T | C | 0.6086 | -0.092 | 0.008 | 1.2589E-34 | 132.25 |
| *LCAT* | rs1047891 | C | A | 0.3131 | -0.056 | 0.008 | 2.5119E-12 | 49.00 |

| | | | | | | | | |
|---|---|---|---|---|---|---|---|---|
| *LCAT* | rs204886 | T | C | 0.5061 | 0.056 | 0.007 | 1.2589E-14 | 64.00 |
| *LCAT* | rs1513275 | T | C | 0.192 | 0.066 | 0.01 | 5.0119E-12 | 43.56 |
| *LCAT* | rs34121855 | T | G | 0.2011 | -0.088 | 0.009 | 5.0119E-22 | 95.60 |
| *LCAT* | rs9987289 | A | G | 0.908 | 0.098 | 0.013 | 1E-14 | 56.83 |
| *LCAT* | rs6589565 | A | G | 0.9328 | -0.123 | 0.015 | 6.3096E-17 | 67.24 |
| *LCAT* | rs7205167 | A | T | 0.488 | 0.053 | 0.007 | 5.0119E-13 | 57.33 |
| *LCAT* | rs4986970 | A | T | 0.0353 | -0.318 | 0.02 | 3.9811E-57 | 252.81 |
| *LCAT* | rs367070 | A | G | 0.2299 | -0.154 | 0.009 | 3.1623E-69 | 292.79 |
| *LCAT* | rs1800961 | C | T | 0.0323 | -0.172 | 0.021 | 7.9433E-17 | 67.08 |
| *LCAT* | rs3747207 | G | A | 0.2156 | -0.063 | 0.009 | 7.9433E-13 | 49.00 |
| *LCN15* | rs4880140 | G | C | 0.7792 | 0.657 | 0.009 | 1E-200 | 5329.00 |
| *LCN15* | rs80158569 | G | A | 0.0792 | -0.094 | 0.013 | 5.0119E-13 | 52.28 |
| *LCN15* | rs1291142 | A | G | 0.6726 | -0.06 | 0.007 | 2.5119E-16 | 73.47 |
| *LCN2* | rs3917932 | C | G | 0.5755 | -0.073 | 0.008 | 3.1623E-22 | 83.27 |
| *LCN2* | rs3014874 | G | A | 0.2638 | 0.075 | 0.008 | 5.0119E-19 | 87.89 |
| *LCN2* | rs113542380 | G | A | 0.0739 | 0.104 | 0.014 | 5.0119E-13 | 55.18 |
| *LCN2* | rs2305637 | C | T | 0.1758 | 0.083 | 0.01 | 2.5119E-17 | 68.89 |
| *LCN2* | rs218264 | A | T | 0.2464 | 0.07 | 0.009 | 5.0119E-16 | 60.49 |
| *LCN2* | rs5869700 | C | CT | 0.3277 | 0.076 | 0.008 | 7.9433E-22 | 90.25 |
| *LCN2* | rs12520928 | C | A | 0.1653 | 0.096 | 0.01 | 5.0119E-22 | 92.16 |
| *LCN2* | rs2158799 | C | G | 0.6088 | 0.062 | 0.008 | 6.3096E-16 | 60.06 |
| *LCN2* | rs1870028 | C | T | 0.0725 | 0.158 | 0.014 | 3.9811E-28 | 127.37 |
| *LCN2* | rs7846314 | A | T | 0.1859 | 0.123 | 0.01 | 5.0119E-38 | 151.29 |
| *LCN2* | rs4407843 | A | C | 0.5681 | -0.066 | 0.008 | 2.5119E-18 | 68.06 |
| *LCN2* | rs542243688 | C | A | 0.0285 | -0.236 | 0.025 | 1.2589E-20 | 89.11 |
| *LCN2* | rs181969679 | A | G | 0.009 | 0.331 | 0.041 | 3.1623E-16 | 65.18 |
| *LCN2* | rs3826331 | T | C | 0.5985 | -0.125 | 0.008 | 7.9433E-61 | 244.14 |
| *LCN2* | rs36058989 | C | T | 0.2179 | -0.067 | 0.009 | 1E-13 | 55.42 |
| *LCN2* | rs78030362 | A | G | 0.0727 | -0.149 | 0.014 | 5.0119E-25 | 113.27 |
| *LCN2* | rs243591 | A | C | 0.4791 | -0.058 | 0.007 | 7.9433E-15 | 68.65 |
| *LCP1* | rs47219 | A | T | 0.2324 | -0.057 | 0.007 | 1.2589E-15 | 66.31 |
| *LCP1* | rs78506734 | C | T | 0.1749 | 0.058 | 0.008 | 7.9433E-13 | 52.56 |
| *LCP1* | rs12342201 | G | A | 0.4761 | -0.062 | 0.006 | 1E-24 | 106.78 |
| *LCP1* | rs763990408 | A | ATG | 0.0287 | -0.207 | 0.018 | 3.9811E-30 | 132.25 |
| *LCP1* | rs7323373 | T | A | 0.9065 | -1.473 | 0.013 | 1E-200 | 12838.63 |
| *LDLR* | rs765974791 | TGATTTCAC | T | 0.6407 | 0.099 | 0.008 | 1E-39 | 153.14 |
| *LDLR* | rs1260326 | T | C | 0.6085 | -0.082 | 0.007 | 3.9811E-29 | 137.22 |
| *LDLR* | rs4704834 | A | G | 0.6482 | 0.082 | 0.008 | 1.9953E-27 | 105.06 |
| *LDLR* | rs451637 | C | T | 0.31 | -0.065 | 0.008 | 5.0119E-16 | 66.02 |
| *LDLR* | rs3812316 | C | G | 0.1266 | -0.095 | 0.011 | 7.9433E-19 | 74.59 |
| *LDLR* | rs13702 | T | C | 0.2867 | -0.093 | 0.008 | 3.1623E-31 | 135.14 |
| *LDLR* | rs2737246 | G | C | 0.2819 | -0.074 | 0.008 | 2.5119E-20 | 85.56 |
| *LDLR* | rs28601761 | C | G | 0.4194 | -0.095 | 0.007 | 2.5119E-38 | 184.18 |
| *LDLR* | rs2068888 | G | A | 0.4499 | -0.05 | 0.007 | 3.1623E-12 | 51.02 |
| *LDLR* | rs174533 | G | A | 0.3462 | 0.074 | 0.008 | 1.9953E-22 | 85.56 |
| *LDLR* | rs964184 | G | C | 0.8664 | -0.204 | 0.011 | 2.5119E-82 | 343.93 |
| *LDLR* | rs261342 | G | C | 0.7772 | -0.061 | 0.009 | 1.9953E-12 | 45.94 |
| *LDLR* | rs247617 | C | A | 0.3243 | -0.093 | 0.008 | 1.5849E-33 | 135.14 |
| *LDLR* | rs77542162 | A | G | 0.0229 | 0.21 | 0.024 | 2.5119E-18 | 76.56 |
| *LDLR* | rs116843064 | G | A | 0.0197 | -0.21 | 0.026 | 1E-15 | 65.24 |
| *LDLR* | rs8107974 | A | T | 0.0766 | -0.11 | 0.014 | 6.3096E-16 | 61.73 |
| *LDLR* | rs1065853 | G | T | 0.0815 | 0.43 | 0.013 | 1E-200 | 1094.08 |
| *LDLRAP1* | rs75446219 | C | T | 0.0358 | 0.179 | 0.02 | 1E-18 | 80.10 |
| *LDLRAP1* | rs6993770 | A | T | 0.2864 | -0.071 | 0.008 | 1E-17 | 78.77 |
| *LDLRAP1* | rs4759076 | T | C | 0.457 | 0.061 | 0.008 | 6.3096E-16 | 58.14 |
| *LECT2* | rs11573156 | G | C | 0.2281 | 0.053 | 0.006 | 1.5849E-18 | 78.03 |
| *LECT2* | rs10027182 | G | A | 0.7442 | -0.039 | 0.006 | 6.3096E-12 | 42.25 |
| *LECT2* | rs12331618 | A | G | 0.5127 | 0.037 | 0.005 | 1.5849E-13 | 54.76 |
| *LECT2* | rs31517 | T | C | 0.6292 | -0.8 | 0.007 | 1E-200 | 13061.22 |
| *LECT2* | rs4252117 | A | G | 0.2949 | 0.086 | 0.005 | 5.0119E-55 | 295.84 |
| *LECT2* | rs507666 | G | A | 0.1846 | -0.054 | 0.006 | 3.1623E-17 | 81.00 |
| *LECT2* | rs28929474 | C | T | 0.0209 | 0.121 | 0.017 | 3.9811E-12 | 50.66 |
| *LECT2* | rs2569491 | G | A | 0.2986 | -0.047 | 0.005 | 7.9433E-18 | 88.36 |
| *LEFTY2* | rs748667196 | GC | G | 0.3703 | -0.713 | 0.007 | 1E-200 | 10374.88 |
| *LEFTY2* | rs10027275 | G | C | 0.7415 | 0.055 | 0.007 | 1E-15 | 61.73 |
| *LEFTY2* | rs635634 | C | T | 0.1842 | -0.082 | 0.008 | 1.9953E-26 | 105.06 |
| *LEFTY2* | rs113878851 | C | CT | 0.1719 | -0.055 | 0.008 | 1E-11 | 47.27 |
| *LEFTY2* | rs7412 | C | T | 0.0814 | 0.11 | 0.011 | 7.9433E-24 | 100.00 |

| | | | | | | | | |
|---|---|---|---|---|---|---|---|---|
| *LEFTY2* | rs738408 | C | T | 0.217 | 0.055 | 0.007 | 2.5119E-14 | 61.73 |
| *LEG1* | rs908702 | C | T | 0.4476 | 0.1 | 0.007 | 1.5849E-44 | 204.08 |
| *LEG1* | rs12991732 | G | C | 0.2664 | -0.055 | 0.008 | 7.9433E-12 | 47.27 |
| *LEG1* | rs4362858 | T | C | 0.7173 | -0.065 | 0.008 | 1.9953E-16 | 66.02 |
| *LEG1* | rs12500456 | C | T | 0.3016 | -0.053 | 0.008 | 6.3096E-12 | 43.89 |
| *LEG1* | rs7447651 | C | T | 0.637 | -0.054 | 0.007 | 2.5119E-13 | 59.51 |
| *LEG1* | rs1196393 | G | A | 0.4667 | -0.111 | 0.007 | 7.9433E-51 | 251.45 |
| *LEG1* | rs1550942 | A | G | 0.5061 | -0.372 | 0.007 | 1E-200 | 2824.16 |
| *LEG1* | rs12533016 | T | C | 0.4123 | 0.078 | 0.007 | 7.9433E-27 | 124.16 |
| *LEG1* | rs56278466 | T | G | 0.661 | 0.074 | 0.007 | 3.1623E-23 | 111.76 |
| *LEG1* | rs2956074 | T | C | 0.4043 | -0.058 | 0.007 | 1.2589E-15 | 68.65 |
| *LEG1* | rs4937126 | G | A | 0.3154 | -0.079 | 0.008 | 3.1623E-25 | 97.52 |
| *LEG1* | rs838968 | A | G | 0.9624 | -0.164 | 0.019 | 1.9953E-18 | 74.50 |
| *LEG1* | rs4761529 | A | G | 0.7391 | -0.055 | 0.008 | 1E-11 | 47.27 |
| *LEG1* | rs848125 | T | G | 0.5583 | 0.103 | 0.007 | 5.0119E-47 | 216.51 |
| *LEG1* | rs2842329 | T | G | 0.2658 | 0.055 | 0.008 | 1.5849E-11 | 47.27 |
| *LEG1* | rs17855739 | C | T | 0.0411 | 0.966 | 0.019 | 1E-200 | 2584.92 |
| *LEG1* | rs34262244 | G | A | 0.2947 | -0.056 | 0.008 | 1.2589E-11 | 49.00 |
| *LEG1* | rs601338 | G | A | 0.5086 | 0.24 | 0.008 | 1E-200 | 900.00 |
| *LEP* | rs56094641 | A | G | 0.4024 | 0.047 | 0.007 | 7.9433E-13 | 45.08 |
| *LEPR* | rs10399687 | C | T | 0.1711 | -0.276 | 0.01 | 1.585E-173 | 761.76 |
| *LEPR* | rs1260326 | T | C | 0.6083 | -0.151 | 0.008 | 7.9433E-89 | 356.27 |
| *LEPR* | rs111269058 | C | T | 0.1217 | -0.082 | 0.011 | 2.5119E-13 | 55.57 |
| *LEPR* | rs2298475 | T | C | 0.0752 | -0.096 | 0.014 | 3.9811E-12 | 47.02 |
| *LEPR* | rs2464190 | T | C | 0.4122 | 0.063 | 0.007 | 6.3096E-17 | 81.00 |
| *LEPR* | rs28929474 | C | T | 0.0208 | 0.299 | 0.026 | 2.5119E-31 | 132.25 |
| *LEPR* | rs186021206 | G | A | 0.0064 | 0.414 | 0.05 | 1.5849E-16 | 68.56 |
| *LEPR* | rs1801689 | A | C | 0.0301 | 0.255 | 0.021 | 1.2589E-32 | 147.45 |
| *LGALS1* | rs10418046 | T | G | 0.2109 | 0.066 | 0.009 | 1.9953E-13 | 53.78 |
| *LGALS1* | rs4820294 | G | A | 0.3337 | -0.283 | 0.008 | 1E-200 | 1251.39 |
| *LGALS3* | rs78542815 | A | G | 0.2228 | 0.114 | 0.008 | 5.0119E-50 | 203.06 |
| *LGALS3* | rs6472539 | G | C | 0.5152 | -0.052 | 0.006 | 1.9953E-16 | 75.11 |
| *LGALS3* | rs751544923 | AG | A | 0.0569 | 0.166 | 0.014 | 1E-33 | 140.59 |
| *LGALS3* | rs7310409 | A | G | 0.6241 | -0.082 | 0.007 | 5.0119E-36 | 137.22 |
| *LGALS3* | rs3832943 | A | AATT | 0.4238 | -0.534 | 0.007 | 1E-200 | 5819.51 |
| *LGALS3* | rs17855739 | C | T | 0.041 | 0.181 | 0.016 | 7.9433E-30 | 127.97 |
| *LGALS3* | rs516246 | C | T | 0.5087 | 0.077 | 0.006 | 1.2589E-33 | 164.69 |
| *LGALS3* | rs12986064 | T | C | 0.5073 | -0.064 | 0.007 | 1.9953E-22 | 83.59 |
| *LGALS3BP* | rs115216147 | T | A | 0.0665 | 0.107 | 0.015 | 3.1623E-13 | 50.88 |
| *LGALS3BP* | rs11242113 | G | A | 0.186 | 0.089 | 0.009 | 7.9433E-22 | 97.79 |
| *LGALS3BP* | rs28753035 | G | C | 0.1656 | 0.068 | 0.01 | 7.9433E-12 | 46.24 |
| *LGALS3BP* | rs41341748 | G | A | 0.011 | 1.641 | 0.035 | 1E-200 | 2198.27 |
| *LGALS3BP* | rs8176741 | G | A | 0.0612 | 0.104 | 0.015 | 2.5119E-12 | 48.07 |
| *LGALS3BP* | rs3972616 | A | AT | 0.4947 | -0.053 | 0.007 | 3.1623E-13 | 57.33 |
| *LGALS3BP* | rs7232 | T | A | 0.3732 | -0.061 | 0.007 | 1.9953E-16 | 75.94 |
| *LGALS3BP* | rs4968402 | G | A | 0.8393 | 0.099 | 0.01 | 3.9811E-24 | 98.01 |
| *LGALS3BP* | rs111526614 | C | T | 0.0183 | -0.48 | 0.028 | 2.5119E-64 | 293.88 |
| *LGALS3BP* | rs6115999 | C | T | 0.0118 | 0.766 | 0.034 | 3.162E-114 | 507.57 |
| *LGALS4* | rs113323158 | A | ACTC | 0.5129 | -0.049 | 0.007 | 3.1623E-13 | 49.00 |
| *LGALS4* | rs505922 | T | C | 0.3167 | -0.526 | 0.008 | 1E-200 | 4323.06 |
| *LGALS4* | rs17655730 | T | C | 0.2454 | 0.058 | 0.008 | 2.5119E-13 | 52.56 |
| *LGALS4* | rs140695578 | A | T | 0.0319 | 0.28 | 0.019 | 3.9811E-47 | 217.17 |
| *LGALS7_LGALS7B* | rs61816761 | G | A | 0.0237 | 0.316 | 0.024 | 3.9811E-38 | 173.36 |
| *LGALS7_LGALS7B* | rs833270 | T | C | 0.6171 | 0.049 | 0.007 | 1.2589E-11 | 49.00 |
| *LGALS7_LGALS7B* | rs676457 | A | T | 0.318 | 0.318 | 0.008 | 1E-200 | 1580.06 |
| *LGALS7_LGALS7B* | rs17658212 | C | T | 0.0596 | -0.106 | 0.015 | 2.5119E-12 | 49.94 |
| *LGALS7_LGALS7B* | rs1549299 | G | A | 0.7055 | -0.057 | 0.008 | 1E-13 | 50.77 |
| *LGALS7_LGALS7B* | rs6508818 | A | G | 0.5784 | 0.225 | 0.007 | 1E-200 | 1033.16 |
| *LGALS7_LGALS7B* | rs584768 | G | A | 0.5332 | 0.048 | 0.007 | 1.5849E-11 | 47.02 |
| *LGALS8* | rs1475133 | G | A | 0.7617 | 0.306 | 0.009 | 1E-200 | 1156.00 |
| *LGALS8* | rs1354034 | T | C | 0.6042 | 0.074 | 0.007 | 2.5119E-24 | 111.76 |
| *LGALS8* | rs342293 | C | G | 0.4596 | -0.084 | 0.007 | 1E-31 | 144.00 |
| *LGALS8* | rs55646585 | C | T | 0.4135 | 0.063 | 0.007 | 3.9811E-18 | 81.00 |
| *LGALS8* | rs2519093 | C | T | 0.1846 | -0.482 | 0.01 | 1E-200 | 2323.24 |
| *LGALS8* | rs7080386 | C | A | 0.4122 | 0.056 | 0.007 | 1.5849E-14 | 64.00 |
| *LGALS8* | rs60822569 | T | TTTCTC | 0.5544 | 0.053 | 0.007 | 3.1623E-13 | 57.33 |
| *LGALS9* | rs3184504 | T | C | 0.5172 | -0.076 | 0.007 | 1.2589E-29 | 117.88 |
| *LGALS9* | rs12936577 | G | T | 0.0225 | -0.294 | 0.027 | 1E-26 | 118.57 |

| | | | | | | | | |
|---|---|---|---|---|---|---|---|---|
| *LGALS9* | rs79526289 | A | G | 0.2504 | -0.514 | 0.008 | 1E-200 | 4128.06 |
| *LGALS9* | rs601338 | G | A | 0.5097 | 0.084 | 0.007 | 2.5119E-35 | 144.00 |
| *LGMN* | rs3811444 | C | T | 0.3321 | -0.067 | 0.008 | 1.5849E-18 | 70.14 |
| *LGMN* | rs35677470 | G | A | 0.0829 | 0.129 | 0.013 | 7.9433E-23 | 98.47 |
| *LGMN* | rs6848819 | A | C | 0.0325 | 0.147 | 0.02 | 5.0119E-13 | 54.02 |
| *LGMN* | rs8191772 | A | G | 0.3357 | 0.061 | 0.008 | 1.5849E-15 | 58.14 |
| *LGMN* | rs17154130 | T | C | 0.0624 | 0.12 | 0.015 | 1.5849E-15 | 64.00 |
| *LGMN* | rs7832219 | T | C | 0.2742 | -0.118 | 0.008 | 1.9953E-48 | 217.56 |
| *LGMN* | rs2737246 | G | C | 0.2823 | -0.059 | 0.008 | 3.9811E-13 | 54.39 |
| *LGMN* | rs6986384 | G | A | 0.0806 | 0.093 | 0.013 | 1.9953E-12 | 51.18 |
| *LGMN* | rs600038 | T | C | 0.2073 | -0.072 | 0.009 | 1E-15 | 64.00 |
| *LGMN* | rs2250645 | C | T | 0.2999 | 0.072 | 0.008 | 6.3096E-20 | 81.00 |
| *LGMN* | rs12767683 | C | A | 0.325 | 0.061 | 0.008 | 3.9811E-15 | 58.14 |
| *LGMN* | rs149871778 | G | GAAC | 0.1049 | 0.373 | 0.012 | 1E-200 | 966.17 |
| *LGMN* | rs117566084 | C | T | 0.01 | 0.459 | 0.037 | 6.3096E-36 | 153.89 |
| *LGMN* | rs148659834 | G | A | 0.0121 | -1.055 | 0.034 | 1E-200 | 962.82 |
| *LGMN* | rs3026096 | G | A | 0.4928 | -0.056 | 0.007 | 1.2589E-14 | 64.00 |
| *LGMN* | rs55976836 | G | A | 0.2303 | -0.061 | 0.009 | 1.9953E-12 | 45.94 |
| *LGMN* | rs1654425 | T | C | 0.8344 | 0.105 | 0.01 | 3.9811E-27 | 110.25 |
| *LHB* | rs72481571 | G | A | 0.1063 | 0.157 | 0.013 | 3.1623E-33 | 145.85 |
| *LHB* | rs3795048 | A | T | 0.0612 | -0.12 | 0.016 | 3.1623E-14 | 56.25 |
| *LIFR* | rs61801010 | A | G | 0.1059 | 0.133 | 0.012 | 1.5849E-26 | 122.84 |
| *LIFR* | rs34579268 | C | T | 0.3711 | 0.063 | 0.008 | 1.2589E-16 | 62.02 |
| *LIFR* | rs34211178 | G | A | 0.4402 | -0.058 | 0.007 | 3.1623E-15 | 68.65 |
| *LIFR* | rs11923060 | C | T | 0.1566 | 0.087 | 0.01 | 6.3096E-18 | 75.69 |
| *LIFR* | rs62353655 | G | A | 0.0456 | -0.36 | 0.018 | 6.3096E-93 | 400.00 |
| *LIFR* | rs562289 | C | T | 0.23 | 0.066 | 0.009 | 3.1623E-14 | 53.78 |
| *LIFR* | rs2519093 | C | T | 0.1842 | -0.421 | 0.01 | 1E-200 | 1772.41 |
| *LIFR* | rs11596680 | A | G | 0.3274 | 0.057 | 0.008 | 5.0119E-13 | 50.77 |
| *LIFR* | rs78689694 | G | C | 0.1327 | -0.099 | 0.011 | 3.1623E-20 | 81.00 |
| *LIFR* | rs186021206 | G | A | 0.0064 | 1.01 | 0.05 | 7.9433E-90 | 408.04 |
| *LIFR* | rs2252576 | C | T | 0.2438 | 0.061 | 0.009 | 1E-12 | 45.94 |
| *LILRA2* | rs2070901 | G | T | 0.2701 | 0.068 | 0.007 | 1.5849E-20 | 94.37 |
| *LILRA2* | rs1257169 | C | A | 0.4986 | -0.078 | 0.006 | 7.9433E-33 | 169.00 |
| *LILRA2* | rs140473879 | C | T | 0.0038 | -1.007 | 0.054 | 1.2589E-76 | 347.75 |
| *LILRA2* | rs2723509 | G | A | 0.4086 | -0.049 | 0.007 | 7.9433E-14 | 49.00 |
| *LILRA2* | rs635634 | C | T | 0.1843 | 0.061 | 0.008 | 3.1623E-13 | 58.14 |
| *LILRA2* | rs4055121 | C | T | 0.1351 | -0.083 | 0.009 | 3.1623E-18 | 85.05 |
| *LILRA2* | rs3184504 | T | C | 0.5175 | -0.063 | 0.006 | 1E-22 | 110.25 |
| *LILRA2* | rs76428106 | T | C | 0.0128 | 0.234 | 0.03 | 3.9811E-15 | 60.84 |
| *LILRA2* | rs186021206 | G | A | 0.0064 | 0.892 | 0.044 | 3.9811E-90 | 410.98 |
| *LILRA2* | rs77542162 | A | G | 0.0229 | 0.152 | 0.022 | 1.9953E-12 | 47.74 |
| *LILRA2* | rs35866622 | C | T | 0.5005 | 0.052 | 0.007 | 1.9953E-12 | 55.18 |
| *LILRA2* | rs28469702 | A | G | 0.0906 | -1.036 | 0.013 | 1E-200 | 6350.86 |
| *LILRA3* | rs141094656 | T | C | 0.0148 | 0.167 | 0.02 | 1.5849E-16 | 69.72 |
| *LILRA3* | rs111338191 | A | AAATT | 0.5211 | -0.04 | 0.005 | 1.5849E-17 | 64.00 |
| *LILRA3* | rs186021206 | G | A | 0.0064 | 0.36 | 0.032 | 2.5119E-29 | 126.56 |
| *LILRA3* | rs367070 | A | G | 0.2304 | -1.219 | 0.009 | 1E-200 | 18345.20 |
| *LILRA5* | rs748544288 | CAAG | C | 0.2269 | -0.067 | 0.009 | 1E-14 | 55.42 |
| *LILRA5* | rs4233366 | C | T | 0.2683 | 0.126 | 0.008 | 3.9811E-54 | 248.06 |
| *LILRA5* | rs731557 | C | A | 0.1938 | 0.211 | 0.009 | 5.012E-119 | 549.64 |
| *LILRA5* | rs597697 | A | G | 0.5762 | 0.053 | 0.007 | 1.9953E-13 | 57.33 |
| *LILRA5* | rs10980802 | A | G | 0.4849 | -0.074 | 0.007 | 3.9811E-25 | 111.76 |
| *LILRA5* | rs1892548 | T | C | 0.6582 | -0.064 | 0.008 | 3.1623E-17 | 64.00 |
| *LILRA5* | rs8033923 | A | G | 0.2071 | -0.069 | 0.009 | 3.9811E-15 | 58.78 |
| *LILRA5* | rs186021206 | G | A | 0.0065 | 0.39 | 0.049 | 1.2589E-15 | 63.35 |
| *LILRA5* | rs34464703 | T | TG | 0.3299 | -0.36 | 0.008 | 1E-200 | 2025.00 |
| *LILRA5* | rs913672 | T | G | 0.3405 | 0.052 | 0.008 | 3.9811E-12 | 42.25 |
| *LILRA6* | rs111338191 | A | AAATT | 0.5207 | -0.039 | 0.006 | 1.5849E-12 | 42.25 |
| *LILRA6* | rs186021206 | G | A | 0.0064 | 0.34 | 0.038 | 1.9953E-19 | 80.06 |
| *LILRA6* | rs11668526 | C | T | 0.5935 | 0.796 | 0.008 | 1E-200 | 9900.25 |
| *LILRB1* | rs1257169 | C | A | 0.4987 | -0.07 | 0.006 | 2.5119E-29 | 136.11 |
| *LILRB1* | rs27295 | C | T | 0.5528 | -0.046 | 0.006 | 1.2589E-13 | 58.78 |
| *LILRB1* | rs9266271 | T | C | 0.3509 | 0.205 | 0.007 | 1E-200 | 857.65 |
| *LILRB1* | rs2031902 | C | T | 0.5873 | -0.047 | 0.006 | 1.9953E-13 | 61.36 |
| *LILRB1* | rs115478735 | A | T | 0.1841 | 0.083 | 0.008 | 5.0119E-25 | 107.64 |
| *LILRB1* | rs1072160 | G | A | 0.4451 | 0.078 | 0.006 | 1.2589E-35 | 169.00 |
| *LILRB1* | rs75071241 | G | A | 0.0596 | -0.09 | 0.013 | 5.0119E-12 | 47.93 |

| | | | | | | | | |
|---|---|---|---|---|---|---|---|---|
| *LILRB1* | rs75974417 | C | T | 0.2107 | 0.063 | 0.008 | 1.9953E-16 | 62.02 |
| *LILRB1* | rs186021206 | G | A | 0.0064 | 0.938 | 0.043 | 1E-106 | 475.85 |
| *LILRB1* | rs377056194 | T | TAC | 0.0901 | 0.104 | 0.013 | 3.1623E-16 | 64.00 |
| *LILRB1* | rs367070 | A | G | 0.2302 | -0.671 | 0.009 | 1E-200 | 5558.53 |
| *LILRB2* | rs10801559 | G | A | 0.4032 | -0.081 | 0.005 | 1.2589E-50 | 262.44 |
| *LILRB2* | rs9267084 | G | A | 0.1536 | 0.137 | 0.007 | 1.2589E-77 | 383.04 |
| *LILRB2* | rs2031902 | C | T | 0.5872 | -0.04 | 0.005 | 1E-13 | 64.00 |
| *LILRB2* | rs1072160 | G | A | 0.4452 | 0.059 | 0.005 | 1E-28 | 139.24 |
| *LILRB2* | rs3184504 | T | C | 0.517 | -0.05 | 0.005 | 1.5849E-21 | 100.00 |
| *LILRB2* | rs72744945 | A | C | 0.097 | 0.061 | 0.009 | 6.3096E-12 | 45.94 |
| *LILRB2* | rs186021206 | G | A | 0.0064 | 0.585 | 0.036 | 3.9811E-58 | 264.06 |
| *LILRB2* | rs77542162 | A | G | 0.0228 | 0.124 | 0.018 | 2.5119E-12 | 47.46 |
| *LILRB2* | rs383925 | C | T | 0.2055 | -1.161 | 0.009 | 1E-200 | 16641.00 |
| *LILRB2* | rs3747207 | G | A | 0.2154 | 0.045 | 0.006 | 1.5849E-12 | 56.25 |
| *LILRB4* | rs9270270 | T | C | 0.8391 | -0.732 | 0.01 | 1E-200 | 5358.24 |
| *LILRB4* | rs3184504 | T | C | 0.5177 | -0.054 | 0.006 | 1.5849E-17 | 81.00 |
| *LILRB4* | rs1065853 | G | T | 0.0811 | 0.084 | 0.012 | 5.0119E-12 | 49.00 |
| *LILRB4* | rs7247412 | T | C | 0.2663 | -0.072 | 0.008 | 1.9953E-21 | 81.00 |
| *LILRB4* | rs417477 | A | G | 0.1185 | 0.381 | 0.01 | 1E-200 | 1451.61 |
| *LILRB5* | rs2082206 | G | A | 0.4453 | 0.05 | 0.004 | 3.1623E-37 | 156.25 |
| *LILRB5* | rs186021206 | G | A | 0.0064 | 0.354 | 0.027 | 5.0119E-40 | 171.90 |
| *LILRB5* | rs12975366 | T | C | 0.3952 | -1.185 | 0.008 | 1E-200 | 21941.02 |
| *LIPF* | rs13195237 | A | G | 0.2339 | -0.084 | 0.009 | 2.5119E-22 | 87.11 |
| *LIPF* | rs746756189 | CA | C | 0.2798 | -0.31 | 0.008 | 1E-200 | 1501.56 |
| *LIPF* | rs12799829 | C | T | 0.341 | 0.141 | 0.008 | 1.9953E-74 | 310.64 |
| *LIPF* | rs392702 | G | A | 0.0338 | 0.255 | 0.02 | 3.1623E-36 | 162.56 |
| *LIPF* | rs4812975 | G | A | 0.7504 | -0.064 | 0.008 | 2.5119E-14 | 64.00 |
| *LMNB1* | rs2746507 | A | T | 0.6187 | 0.152 | 0.008 | 3.9811E-86 | 361.00 |
| *LMNB1* | rs7080536 | G | A | 0.0436 | 0.312 | 0.019 | 3.1623E-63 | 269.65 |
| *LMNB2* | rs2706762 | C | T | 0.1515 | -0.099 | 0.01 | 1.5849E-22 | 98.01 |
| *LMNB2* | rs4348853 | T | C | 0.5575 | 0.054 | 0.007 | 1.9953E-13 | 59.51 |
| *LMNB2* | rs6045561 | T | C | 0.276 | -0.056 | 0.008 | 6.3096E-12 | 49.00 |
| *LMOD1* | rs12342201 | G | A | 0.4766 | -0.049 | 0.007 | 7.9433E-13 | 49.00 |
| *LMOD1* | rs2341097 | C | T | 0.3597 | -0.05 | 0.007 | 3.9811E-12 | 51.02 |
| *LONP1* | rs342293 | C | G | 0.4595 | -0.055 | 0.008 | 5.0119E-13 | 47.27 |
| *LONP1* | rs6993770 | A | T | 0.2866 | -0.061 | 0.008 | 3.9811E-13 | 58.14 |
| *LONP1* | rs60822569 | T | TTTCTC | 0.553 | 0.054 | 0.008 | 1.2589E-12 | 45.56 |
| *LONP1* | rs74996080 | G | T | 0.1045 | -0.281 | 0.013 | 6.31E-107 | 467.22 |
| *LPA* | rs56393506 | C | T | 0.1721 | 0.761 | 0.01 | 1E-200 | 5791.21 |
| *LPA* | rs1065853 | G | T | 0.081 | -0.135 | 0.011 | 7.9433E-35 | 150.62 |
| *LPCAT2* | rs16955343 | C | T | 0.046 | -0.253 | 0.018 | 6.3096E-45 | 197.56 |
| *LPL* | rs1260326 | T | C | 0.6083 | 0.054 | 0.006 | 3.9811E-17 | 81.00 |
| *LPL* | rs5835988 | T | TG | 0.4034 | 0.047 | 0.006 | 3.1623E-13 | 61.36 |
| *LPL* | rs1515104 | T | A | 0.6498 | -0.052 | 0.007 | 2.5119E-15 | 55.18 |
| *LPL* | rs72959041 | G | A | 0.0497 | -0.101 | 0.015 | 6.3096E-12 | 45.34 |
| *LPL* | rs149522 | C | T | 0.3987 | -0.048 | 0.007 | 1.9953E-13 | 47.02 |
| *LPL* | rs1801177 | G | A | 0.0176 | -1.479 | 0.025 | 1E-200 | 3499.91 |
| *LPL* | rs7201247 | T | G | 0.3727 | 0.047 | 0.006 | 3.1623E-13 | 61.36 |
| *LPL* | rs116843064 | G | A | 0.0196 | 0.234 | 0.023 | 3.9811E-25 | 103.51 |
| *LPO* | rs61804164 | G | C | 0.1241 | 0.08 | 0.011 | 1.5849E-12 | 52.89 |
| *LPO* | rs6703244 | A | G | 0.4129 | 0.078 | 0.007 | 1.2589E-25 | 124.16 |
| *LPO* | rs908702 | C | T | 0.4477 | 0.053 | 0.007 | 6.3096E-13 | 57.33 |
| *LPO* | rs9274442 | G | C | 0.2827 | -0.079 | 0.009 | 3.1623E-18 | 77.05 |
| *LPO* | rs62418808 | G | A | 0.3315 | -0.175 | 0.008 | 1.995E-109 | 478.52 |
| *LPO* | rs7791463 | G | A | 0.5344 | -0.076 | 0.007 | 6.3096E-25 | 117.88 |
| *LPO* | rs56278466 | T | G | 0.6611 | 0.138 | 0.008 | 6.3096E-70 | 297.56 |
| *LPO* | rs1427751 | C | T | 0.8253 | -0.106 | 0.01 | 1.5849E-27 | 112.36 |
| *LPO* | rs7219860 | G | A | 0.2181 | -0.307 | 0.009 | 1E-200 | 1163.57 |
| *LPO* | rs12019136 | G | A | 0.0407 | 0.625 | 0.019 | 1E-200 | 1082.06 |
| *LPO* | rs34262244 | G | A | 0.2943 | -0.069 | 0.008 | 2.5119E-16 | 74.39 |
| *LPO* | rs516246 | C | T | 0.5094 | 0.155 | 0.008 | 5.0119E-94 | 375.39 |
| *LPO* | rs61739245 | A | T | 0.0189 | -0.253 | 0.027 | 7.9433E-21 | 87.80 |
| *LPP* | rs342296 | G | A | 0.4568 | -0.057 | 0.008 | 5.0119E-14 | 50.77 |
| *LRCH4* | rs56328569 | C | G | 0.2222 | 0.077 | 0.009 | 1.2589E-17 | 73.20 |
| *LRCH4* | rs34127158 | A | T | 0.3392 | -0.065 | 0.008 | 2.5119E-16 | 66.02 |
| *LRCH4* | rs11620465 | C | T | 0.5208 | 0.065 | 0.008 | 3.1623E-17 | 66.02 |
| *LRCH4* | rs10418046 | T | G | 0.2112 | 0.08 | 0.009 | 3.1623E-18 | 79.01 |
| *LRG1* | rs6700896 | C | T | 0.3774 | -0.047 | 0.007 | 7.9433E-12 | 45.08 |

| | | | | | | | | |
|---|---|---|---|---|---|---|---|---|
| *LRG1* | rs7968455 | A | T | 0.382 | -0.083 | 0.007 | 1E-31 | 140.59 |
| *LRG1* | rs147233090 | C | T | 0.0237 | -0.163 | 0.022 | 1.5849E-13 | 54.89 |
| *LRG1* | rs8105192 | C | G | 0.5 | -0.56 | 0.007 | 1E-200 | 6400.00 |
| *LRIG1* | rs4846922 | T | C | 0.6551 | 0.094 | 0.007 | 1.9953E-46 | 180.33 |
| *LRIG1* | rs59104589 | C | T | 0.357 | -0.046 | 0.006 | 6.3096E-13 | 58.78 |
| *LRIG1* | rs2306272 | T | C | 0.2882 | -0.858 | 0.008 | 1E-200 | 11502.56 |
| *LRIG1* | rs9372538 | C | T | 0.2829 | 0.075 | 0.007 | 1.9953E-27 | 114.80 |
| *LRIG1* | rs56278466 | T | G | 0.6611 | 0.074 | 0.007 | 5.0119E-30 | 111.76 |
| *LRIG1* | rs11045856 | T | G | 0.2401 | 0.081 | 0.007 | 1.2589E-28 | 133.90 |
| *LRIG1* | rs10083143 | A | C | 0.0358 | 0.138 | 0.017 | 1.2589E-16 | 65.90 |
| *LRIG1* | rs7979473 | A | G | 0.6163 | -0.046 | 0.006 | 7.9433E-13 | 58.78 |
| *LRIG1* | rs28929474 | C | T | 0.0207 | 0.335 | 0.022 | 1.5849E-53 | 231.87 |
| *LRIG1* | rs339969 | C | A | 0.6165 | 0.046 | 0.006 | 3.9811E-13 | 58.78 |
| *LRIG1* | rs774552666 | AATAACGG' | A | 0.9338 | 0.092 | 0.012 | 1.2589E-13 | 58.78 |
| *LRIG3* | rs11172841 | A | C | 0.2247 | 0.09 | 0.009 | 5.0119E-23 | 100.00 |
| *LRP1* | rs1010554 | T | C | 0.4706 | -0.047 | 0.007 | 7.9433E-13 | 45.08 |
| *LRP1* | rs140556958 | G | A | 0.0009 | 1.844 | 0.127 | 1.9953E-47 | 210.82 |
| *LRP1* | rs59674561 | G | T | 0.0253 | 1.844 | 0.023 | 1E-200 | 6427.86 |
| *LRP1* | rs328 | C | G | 0.1006 | -0.224 | 0.011 | 3.9811E-95 | 414.68 |
| *LRP1* | rs10820966 | A | T | 0.3323 | -0.115 | 0.007 | 1.5849E-61 | 269.90 |
| *LRP1* | rs772851187 | TGCTGTAG( | T | 0.4685 | 0.052 | 0.007 | 1.2589E-15 | 55.18 |
| *LRP1* | rs2229498 | G | A | 0.8355 | -0.115 | 0.009 | 3.9811E-39 | 163.27 |
| *LRP1* | rs71279465 | T | TAATAAAT/ | 0.3022 | 0.058 | 0.007 | 1.2589E-15 | 68.65 |
| *LRP1* | rs11846959 | G | A | 0.2533 | 0.118 | 0.007 | 3.9811E-56 | 284.16 |
| *LRP1* | rs200489612 | G | A | 0.0052 | 0.614 | 0.049 | 6.3096E-36 | 157.02 |
| *LRP1* | rs112467088 | A | T | 0.3091 | 0.056 | 0.007 | 3.1623E-15 | 64.00 |
| *LRP11* | rs61747728 | C | T | 0.038 | 0.17 | 0.018 | 1.9953E-22 | 89.20 |
| *LRP11* | rs2342858 | C | T | 0.3545 | 0.605 | 0.008 | 1E-200 | 5719.14 |
| *LRP11* | rs36060036 | C | T | 0.1677 | -0.066 | 0.009 | 3.9811E-13 | 53.78 |
| *LRP11* | rs186021206 | G | A | 0.0064 | 0.428 | 0.046 | 1.5849E-20 | 86.57 |
| *LRP11* | rs1801689 | A | C | 0.0301 | 0.182 | 0.02 | 1.9953E-20 | 82.81 |
| *LRPAP1* | rs143486729 | G | A | 0.0338 | -0.55 | 0.018 | 1E-200 | 933.64 |
| *LRPAP1* | rs13107325 | C | T | 0.0747 | 0.101 | 0.012 | 1.5849E-16 | 70.84 |
| *LRPAP1* | rs115881826 | A | G | 0.0369 | 1.789 | 0.019 | 1E-200 | 8865.71 |
| *LRPAP1* | rs7923609 | A | G | 0.4705 | 0.052 | 0.006 | 1E-16 | 75.11 |
| *LRPAP1* | rs55646096 | C | T | 0.1785 | -0.067 | 0.008 | 7.9433E-16 | 70.14 |
| *LRPAP1* | rs117654599 | T | A | 0.013 | 0.309 | 0.029 | 1.9953E-26 | 113.53 |
| *LRPAP1* | rs11846959 | G | A | 0.2531 | 0.074 | 0.007 | 3.1623E-24 | 111.76 |
| *LRRC25* | rs4672491 | T | G | 0.5212 | -0.071 | 0.007 | 3.1623E-24 | 102.88 |
| *LRRC25* | rs13388024 | A | G | 0.5034 | -0.048 | 0.007 | 3.1623E-12 | 47.02 |
| *LRRC25* | rs2470750 | A | T | 0.3926 | -0.208 | 0.007 | 5.012E-183 | 882.94 |
| *LRRC25* | rs13107325 | C | T | 0.0748 | -0.143 | 0.013 | 3.1623E-27 | 121.00 |
| *LRRC25* | rs509227 | A | C | 0.5225 | 0.288 | 0.007 | 1E-200 | 1692.73 |
| *LRRC25* | rs2858331 | A | G | 0.3682 | -0.081 | 0.007 | 1E-28 | 133.90 |
| *LRRC25* | rs12667978 | C | T | 0.5716 | -0.142 | 0.007 | 3.1623E-91 | 411.51 |
| *LRRC25* | rs199922514 | A | AAGAAGAA | 0.9077 | -0.091 | 0.012 | 2.5119E-14 | 57.51 |
| *LRRC25* | rs950802 | G | A | 0.3063 | 0.171 | 0.008 | 3.162E-113 | 456.89 |
| *LRRC25* | rs4055121 | C | T | 0.1347 | -0.084 | 0.01 | 2.5119E-16 | 70.56 |
| *LRRC25* | rs186021206 | G | A | 0.0063 | 1.049 | 0.048 | 2.512E-105 | 477.60 |
| *LRRC25* | rs2659005 | C | T | 0.4412 | 0.078 | 0.007 | 1E-28 | 124.16 |
| *LRRC25* | rs754032589 | TAGAAAAA | T | 0.4105 | -0.339 | 0.007 | 1E-200 | 2345.33 |
| *LRRC25* | rs4893 | A | G | 0.0397 | -0.453 | 0.018 | 2.512E-134 | 633.36 |
| *LRRC37A2* | rs13107325 | C | T | 0.0748 | 0.091 | 0.009 | 1.5849E-23 | 102.23 |
| *LRRC37A2* | rs2844566 | T | A | 0.3641 | -0.038 | 0.005 | 3.1623E-14 | 57.76 |
| *LRRC37A2* | rs7310238 | A | G | 0.7125 | -0.048 | 0.005 | 1.9953E-19 | 92.16 |
| *LRRC37A2* | rs62057151 | C | T | 0.2326 | 1.115 | 0.009 | 1E-200 | 15348.46 |
| *LRRC37A2* | rs41410147 | C | T | 0.0669 | 0.365 | 0.017 | 1.9953E-98 | 460.99 |
| *LRRC37A2* | rs1065853 | G | T | 0.0812 | 0.066 | 0.009 | 5.0119E-14 | 53.78 |
| *LRRC38* | rs35329337 | A | AAT | 0.3136 | 0.359 | 0.008 | 1E-200 | 2013.77 |
| *LRRC59* | rs1354034 | T | C | 0.6045 | 0.105 | 0.008 | 7.9433E-43 | 172.27 |
| *LRRC59* | rs342293 | C | G | 0.4592 | -0.064 | 0.008 | 1.9953E-17 | 64.00 |
| *LRRC59* | rs35751397 | T | TA | 0.4689 | 0.059 | 0.008 | 7.9433E-15 | 54.39 |
| *LRRC59* | rs73396521 | G | A | 0.245 | 0.067 | 0.009 | 3.1623E-14 | 55.42 |
| *LRRFIP1* | rs62183645 | T | A | 0.2184 | 0.071 | 0.009 | 2.5119E-15 | 62.23 |
| *LRRN1* | rs9839475 | G | A | 0.3485 | 0.811 | 0.008 | 1E-200 | 10276.89 |
| *LRRN1* | rs702634 | G | A | 0.6906 | 0.047 | 0.007 | 1E-11 | 45.08 |
| *LRRN1* | rs576529960 | C | CTT | 0.342 | -0.046 | 0.007 | 1.5849E-11 | 43.18 |
| *LRRN1* | rs1405212 | T | C | 0.6283 | -0.064 | 0.007 | 1.5849E-22 | 83.59 |

| | | | | | | | | |
|---|---|---|---|---|---|---|---|---|
| *LRRN1* | rs11414746 | T | TA | 0.4227 | 0.048 | 0.006 | 1.9953E-13 | 64.00 |
| *LRRN1* | rs55861089 | A | G | 0.1046 | -0.11 | 0.01 | 1.9953E-26 | 121.00 |
| *LRRN1* | rs594144 | C | T | 0.4664 | 0.049 | 0.006 | 1.5849E-14 | 66.69 |
| *LRRN1* | rs6496452 | A | T | 0.4503 | 0.045 | 0.006 | 2.5119E-12 | 56.25 |
| *LRTM2* | rs79473248 | G | A | 0.0316 | 0.322 | 0.021 | 1.9953E-52 | 235.11 |
| *LRTM2* | rs369705328 | G | C | 0.1282 | 0.078 | 0.011 | 1.9953E-12 | 50.28 |
| *LRTM2* | rs1870148 | G | A | 0.789 | 0.062 | 0.009 | 3.9811E-12 | 47.46 |
| *LRTM2* | rs41276696 | G | A | 0.04 | -0.37 | 0.019 | 1.2589E-85 | 379.22 |
| *LRTM2* | rs186021206 | G | A | 0.0064 | 0.643 | 0.05 | 1.5849E-37 | 165.38 |
| *LSM8* | rs232220 | T | C | 0.0493 | -0.155 | 0.021 | 1E-13 | 54.48 |
| *LSP1* | rs907612 | C | T | 0.3786 | 0.149 | 0.008 | 1.5849E-85 | 346.89 |
| *LSP1* | rs12941811 | T | C | 0.5751 | -0.053 | 0.007 | 6.3096E-13 | 57.33 |
| *LTA* | rs2009581 | G | A | 0.2725 | -0.071 | 0.007 | 1.5849E-24 | 102.88 |
| *LTA* | rs58106596 | G | A | 0.2209 | 0.06 | 0.008 | 1E-15 | 56.25 |
| *LTA* | rs2229092 | A | C | 0.0637 | -1.493 | 0.015 | 1E-200 | 9906.88 |
| *LTA* | rs764024035 | GT | G | 0.1692 | 0.181 | 0.008 | 1.585E-103 | 511.89 |
| *LTA* | rs3184504 | T | C | 0.5174 | -0.146 | 0.006 | 1E-122 | 592.11 |
| *LTA* | rs186021206 | G | A | 0.0064 | 0.305 | 0.043 | 7.9433E-13 | 50.31 |
| *LTA* | rs34557412 | A | G | 0.0068 | 0.438 | 0.038 | 1.9953E-31 | 132.86 |
| *LTA* | rs8071789 | C | T | 0.4881 | -0.043 | 0.006 | 1.2589E-11 | 51.36 |
| *LTA* | rs344560 | T | C | 0.9452 | 0.166 | 0.014 | 5.0119E-34 | 140.59 |
| *LTA* | rs738409 | C | G | 0.2175 | 0.058 | 0.007 | 7.9433E-15 | 68.65 |
| *LTA4H* | rs2540486 | C | G | 0.3467 | 0.102 | 0.008 | 1.2589E-39 | 162.56 |
| *LTB* | rs2395470 | G | A | 0.3304 | -0.167 | 0.008 | 7.9433E-96 | 435.77 |
| *LTB* | rs764024035 | GT | G | 0.1691 | 0.085 | 0.01 | 3.9811E-17 | 72.25 |
| *LTB* | rs10774624 | G | A | 0.5134 | -0.066 | 0.008 | 1.2589E-18 | 68.06 |
| *LTBP2* | rs4920605 | G | A | 0.568 | 0.07 | 0.006 | 1.2589E-28 | 136.11 |
| *LTBP2* | rs333947 | G | A | 0.1481 | -0.064 | 0.009 | 5.0119E-13 | 50.57 |
| *LTBP2* | rs61801010 | A | G | 0.1062 | 0.088 | 0.011 | 1E-16 | 64.00 |
| *LTBP2* | rs2462512 | C | A | 0.5079 | -0.052 | 0.006 | 1.5849E-16 | 75.11 |
| *LTBP2* | rs3751198 | A | G | 0.6039 | 0.085 | 0.006 | 1.9953E-39 | 200.69 |
| *LTBP2* | rs888414 | G | A | 0.3826 | -0.105 | 0.006 | 6.3096E-60 | 306.25 |
| *LTBP2* | rs2118181 | T | C | 0.1025 | -0.081 | 0.01 | 5.0119E-15 | 65.61 |
| *LTBP2* | rs186021206 | G | A | 0.0064 | 0.591 | 0.043 | 1E-42 | 188.90 |
| *LTBP2* | rs146385050 | C | A | 0.1831 | 0.056 | 0.008 | 7.9433E-12 | 49.00 |
| *LTBP3* | rs376096585 | C | CT | 0.2234 | 0.061 | 0.008 | 3.1623E-13 | 58.14 |
| *LTBP3* | rs6718159 | T | A | 0.5632 | -0.049 | 0.007 | 7.9433E-12 | 49.00 |
| *LTBP3* | rs140336797 | A | AT | 0.4336 | 0.059 | 0.007 | 2.5119E-17 | 71.04 |
| *LTBP3* | rs4099470 | C | T | 0.0548 | 0.896 | 0.016 | 1E-200 | 3136.00 |
| *LTBP3* | rs28929474 | C | T | 0.0207 | 0.26 | 0.024 | 1.2589E-26 | 117.36 |
| *LTBP3* | rs77303550 | C | T | 0.1959 | -0.068 | 0.009 | 7.9433E-15 | 57.09 |
| *LTBP3* | rs1801689 | A | C | 0.0298 | 0.726 | 0.021 | 1E-200 | 1195.18 |
| *LTBP3* | rs73045269 | C | T | 0.1715 | 0.146 | 0.009 | 3.1623E-57 | 263.16 |
| *LTBR* | rs61747728 | C | T | 0.038 | 0.202 | 0.019 | 3.1623E-27 | 113.03 |
| *LTBR* | rs1260326 | T | C | 0.6087 | 0.07 | 0.007 | 1.5849E-21 | 100.00 |
| *LTBR* | rs12613853 | A | G | 0.383 | 0.053 | 0.007 | 1.2589E-12 | 57.33 |
| *LTBR* | rs1434055 | G | A | 0.5555 | -0.063 | 0.007 | 5.0119E-18 | 81.00 |
| *LTBR* | rs27295 | C | T | 0.5526 | -0.05 | 0.007 | 3.1623E-12 | 51.02 |
| *LTBR* | rs10849449 | A | G | 0.655 | -0.311 | 0.008 | 1E-200 | 1511.27 |
| *LTBR* | rs12810719 | A | G | 0.2304 | 0.079 | 0.009 | 1E-19 | 77.05 |
| *LTBR* | rs28929474 | C | T | 0.0209 | 0.952 | 0.025 | 1E-200 | 1450.09 |
| *LTBR* | rs77924615 | G | A | 0.1966 | -0.073 | 0.009 | 7.9433E-16 | 65.79 |
| *LTBR* | rs1801689 | A | C | 0.0302 | 0.152 | 0.021 | 3.9811E-13 | 52.39 |
| *LTBR* | rs123698 | G | C | 0.605 | 0.068 | 0.007 | 3.1623E-20 | 94.37 |
| *LUZP2* | rs76794772 | T | A | 0.0367 | -0.591 | 0.021 | 3.981E-178 | 792.02 |
| *LXN* | rs6785524 | C | A | 0.5253 | -0.167 | 0.008 | 2.512E-109 | 435.77 |
| *LXN* | rs55636108 | T | C | 0.2063 | 0.075 | 0.009 | 1E-15 | 69.44 |
| *LY6D* | rs61816761 | G | A | 0.0236 | 0.32 | 0.025 | 1.2589E-38 | 163.84 |
| *LY6D* | rs2870238 | C | T | 0.503 | 0.049 | 0.007 | 1.9953E-12 | 49.00 |
| *LY6D* | rs2572925 | C | T | 0.4045 | -0.443 | 0.008 | 1E-200 | 3066.39 |
| *LY6D* | rs77924615 | G | A | 0.1967 | -0.068 | 0.009 | 1.9953E-14 | 57.09 |
| *LY6D* | rs56035820 | C | T | 0.2335 | 0.059 | 0.008 | 3.1623E-12 | 54.39 |
| *LY75* | rs72957586 | G | A | 0.5575 | 0.934 | 0.008 | 1E-200 | 13630.56 |
| *LY75* | rs34794906 | T | C | 0.2645 | 0.049 | 0.006 | 1E-16 | 66.69 |
| *LY75* | rs1137827 | C | T | 0.0612 | 0.094 | 0.011 | 3.1623E-18 | 73.02 |
| *LY75* | rs12574844 | G | A | 0.0706 | -0.101 | 0.01 | 3.1623E-23 | 102.01 |
| *LY75* | rs3184504 | T | C | 0.5174 | -0.055 | 0.005 | 3.1623E-26 | 121.00 |
| *LY75* | rs3213572 | G | A | 0.4702 | -0.035 | 0.005 | 1E-11 | 49.00 |

| | | | | | | | | |
|---|---|---|---|---|---|---|---|---|
| *LY75* | rs186021206 | G | A | 0.0064 | 0.923 | 0.036 | 1E-146 | 657.35 |
| *LY9* | rs188468174 | C | T | 0.0144 | -0.249 | 0.032 | 6.3096E-15 | 60.55 |
| *LY9* | rs37455 | A | G | 0.3521 | 0.07 | 0.008 | 3.9811E-19 | 76.56 |
| *LY9* | rs12128261 | T | G | 0.2173 | 0.6 | 0.009 | 1E-200 | 4444.44 |
| *LY9* | rs2009581 | G | A | 0.2725 | -0.063 | 0.008 | 1.2589E-15 | 62.02 |
| *LY9* | rs9824474 | A | G | 0.367 | -0.055 | 0.007 | 7.9433E-14 | 61.73 |
| *LY9* | rs7634389 | T | C | 0.3896 | -0.071 | 0.007 | 3.9811E-23 | 102.88 |
| *LY9* | rs35829610 | C | CTG | 0.3384 | -0.063 | 0.007 | 1.2589E-17 | 81.00 |
| *LY9* | rs11185602 | A | G | 0.3246 | -0.051 | 0.007 | 5.0119E-12 | 53.08 |
| *LY9* | rs778224678 | GACT | G | 0.1139 | 0.081 | 0.011 | 2.5119E-13 | 54.22 |
| *LY9* | rs8176719 | T | TC | 0.34 | 0.057 | 0.007 | 1E-14 | 66.31 |
| *LY9* | rs75071241 | G | A | 0.0599 | -0.103 | 0.015 | 3.1623E-12 | 47.15 |
| *LY9* | rs3184504 | T | C | 0.5168 | -0.124 | 0.007 | 5.0119E-71 | 313.80 |
| *LY9* | rs76428106 | T | C | 0.0128 | 0.268 | 0.032 | 1E-16 | 70.14 |
| *LY9* | rs186021206 | G | A | 0.0065 | 1.036 | 0.048 | 1E-103 | 465.84 |
| *LY9* | rs2725405 | G | C | 0.4415 | 0.083 | 0.007 | 3.9811E-32 | 140.59 |
| *LY96* | rs188468174 | C | T | 0.0142 | -0.487 | 0.032 | 2.5119E-53 | 231.61 |
| *LY96* | rs35668054 | C | T | 0.0759 | -0.116 | 0.014 | 6.3096E-16 | 68.65 |
| *LY96* | rs67612510 | G | A | 0.5364 | -0.076 | 0.009 | 1.2589E-18 | 71.31 |
| *LY96* | rs3804333 | C | T | 0.2036 | -0.062 | 0.009 | 1.5849E-11 | 47.46 |
| *LY96* | rs6472812 | A | G | 0.9563 | -0.202 | 0.018 | 3.1623E-29 | 125.94 |
| *LY96* | rs3181356 | C | T | 0.2291 | 0.062 | 0.009 | 1.5849E-11 | 47.46 |
| *LY96* | rs7943728 | G | A | 0.1936 | 0.078 | 0.009 | 1E-16 | 75.11 |
| *LY96* | rs3184504 | T | C | 0.517 | -0.061 | 0.007 | 1E-16 | 75.94 |
| *LY96* | rs117775520 | G | A | 0.0362 | -0.187 | 0.02 | 1.5849E-21 | 87.42 |
| *LY96* | rs3803800 | A | G | 0.7891 | -0.081 | 0.009 | 2.5119E-19 | 81.00 |
| *LY96* | rs34557412 | A | G | 0.0068 | -0.463 | 0.045 | 7.9433E-25 | 105.86 |
| *LY96* | rs5763821 | A | C | 0.3899 | -0.064 | 0.008 | 1.2589E-16 | 64.00 |
| *LYAR* | rs11723339 | C | T | 0.6207 | -0.055 | 0.008 | 5.0119E-12 | 47.27 |
| *LYN* | rs1354034 | T | C | 0.6046 | 0.056 | 0.008 | 2.5119E-13 | 49.00 |
| *LYN* | rs2055663 | T | C | 0.8639 | -0.091 | 0.011 | 1.2589E-16 | 68.44 |
| *LYN* | rs10733789 | T | C | 0.3131 | 0.055 | 0.008 | 1.2589E-11 | 47.27 |
| *LYN* | rs2186882 | G | A | 0.2701 | 0.072 | 0.008 | 3.1623E-17 | 81.00 |
| *LYN* | rs892090 | T | G | 0.8341 | 0.07 | 0.01 | 3.9811E-12 | 49.00 |
| *LYPD3* | rs1260326 | T | C | 0.6086 | 0.052 | 0.007 | 1E-13 | 55.18 |
| *LYPD3* | rs1047891 | C | A | 0.3125 | 0.055 | 0.007 | 5.0119E-14 | 61.73 |
| *LYPD3* | rs2479016 | A | G | 0.4271 | -0.078 | 0.007 | 3.1623E-29 | 124.16 |
| *LYPD3* | rs112155110 | C | T | 0.4018 | -0.054 | 0.007 | 3.9811E-14 | 59.51 |
| *LYPD3* | rs7842080 | A | G | 0.2169 | 0.179 | 0.008 | 1E-102 | 500.64 |
| *LYPD3* | rs2519093 | C | T | 0.1844 | -0.149 | 0.009 | 3.1623E-64 | 274.09 |
| *LYPD3* | rs139130389 | C | CTA | 0.0727 | -0.477 | 0.013 | 1E-200 | 1346.33 |
| *LYPD3* | rs10893502 | T | C | 0.5905 | -0.072 | 0.007 | 3.1623E-24 | 105.80 |
| *LYPD3* | rs2286783 | G | A | 0.4025 | 0.05 | 0.007 | 7.9433E-13 | 51.02 |
| *LYPD3* | rs56990244 | C | G | 0.5881 | -0.1 | 0.007 | 1.2589E-47 | 204.08 |
| *LYPD3* | rs62034713 | A | G | 0.2492 | 0.162 | 0.008 | 5.0119E-93 | 410.06 |
| *LYPD3* | rs186021206 | G | A | 0.0064 | 0.959 | 0.047 | 6.3096E-93 | 416.33 |
| *LYPD3* | rs276937 | A | T | 0.3199 | 0.133 | 0.007 | 1.5849E-73 | 361.00 |
| *LYPD3* | rs708686 | C | T | 0.2677 | 0.083 | 0.008 | 7.9433E-24 | 107.64 |
| *LYPD3* | rs11671471 | G | A | 0.1127 | -0.522 | 0.012 | 1E-200 | 1892.25 |
| *LYPD3* | rs601338 | G | A | 0.5093 | 0.26 | 0.007 | 1E-200 | 1379.59 |
| *LYPD3* | rs383510 | T | C | 0.5096 | -0.08 | 0.007 | 1.2589E-31 | 130.61 |
| *LYPD8* | rs12729620 | A | G | 0.0586 | 1.068 | 0.018 | 1E-200 | 3520.44 |
| *LYPD8* | rs12046713 | C | T | 0.0936 | 0.847 | 0.013 | 1E-200 | 4245.02 |
| *LYPD8* | rs12022008 | T | C | 0.0872 | 0.918 | 0.015 | 1E-200 | 3745.44 |
| *LYPD8* | rs12044252 | T | C | 0.0937 | 0.846 | 0.013 | 1E-200 | 4235.01 |
| *LYPD8* | rs4556017 | C | T | 0.8535 | -0.077 | 0.01 | 1.5849E-14 | 59.29 |
| *LYPD8* | rs11220520 | C | T | 0.195 | 0.106 | 0.009 | 1.5849E-32 | 138.72 |
| *LYPD8* | rs186021206 | G | A | 0.0063 | 0.55 | 0.049 | 1.5849E-29 | 125.99 |
| *LYPD8* | rs73338360 | G | C | 0.1007 | 0.128 | 0.012 | 1.2589E-27 | 113.78 |
| *LYPD8* | rs708686 | C | T | 0.2676 | 0.168 | 0.008 | 3.162E-99 | 441.00 |
| *LYPD8* | rs4760 | A | G | 0.1563 | -0.117 | 0.01 | 2.5119E-33 | 136.89 |
| *LYSMD3* | rs68066031 | T | C | 0.2291 | 0.095 | 0.009 | 5.0119E-25 | 111.42 |
| *LYSMD3* | rs1354034 | T | C | 0.604 | 0.053 | 0.008 | 1.2589E-11 | 43.89 |
| *LYSMD3* | rs10805870 | A | C | 0.405 | 0.055 | 0.008 | 3.9811E-12 | 47.27 |
| *LYSMD3* | rs6519245 | C | A | 0.4115 | -0.06 | 0.008 | 1.2589E-14 | 56.25 |
| *LYVE1* | rs7639097 | G | T | 0.6138 | -0.063 | 0.008 | 7.9433E-16 | 62.02 |
| *LYVE1* | rs7865362 | C | T | 0.3632 | 0.132 | 0.008 | 2.5119E-66 | 272.25 |
| *LYVE1* | rs147866228 | C | T | 0.008 | -0.378 | 0.041 | 2.5119E-20 | 85.00 |

| | | | | | | | | |
|---|---|---|---|---|---|---|---|---|
| *LYVE1* | rs781822643 | CA | C | 0.3371 | 0.061 | 0.008 | 2.5119E-15 | 58.14 |
| *LYVE1* | rs7077538 | T | G | 0.3234 | -0.054 | 0.008 | 2.5119E-12 | 45.56 |
| *LYVE1* | rs7119709 | G | A | 0.0398 | 0.35 | 0.019 | 2.5119E-75 | 339.34 |
| *LYVE1* | rs10769263 | A | C | 0.387 | -0.058 | 0.008 | 1.5849E-14 | 52.56 |
| *LYVE1* | rs6591649 | A | G | 0.3451 | 0.066 | 0.008 | 2.5119E-17 | 68.06 |
| *LYVE1* | rs112771035 | C | G | 0.0714 | 0.462 | 0.014 | 1E-200 | 1089.00 |
| *LYVE1* | rs61946383 | A | G | 0.3477 | -0.055 | 0.008 | 3.9811E-13 | 47.27 |
| *LYVE1* | rs1955512 | G | A | 0.5727 | 0.066 | 0.007 | 1.2589E-18 | 88.90 |
| *LYVE1* | rs186021206 | G | A | 0.0064 | 1.398 | 0.05 | 6.31E-172 | 781.76 |
| *LYVE1* | rs6112930 | G | A | 0.3909 | -0.069 | 0.007 | 1E-20 | 97.16 |
| *LYZL2* | rs138933538 | T | C | 0.0022 | -0.516 | 0.074 | 2.5119E-12 | 48.62 |
| *LYZL2* | rs116792212 | C | T | 0.0396 | 0.495 | 0.018 | 1.585E-167 | 756.25 |
| *LZTFL1* | rs1129183 | C | T | 0.0777 | -0.266 | 0.014 | 2.5119E-79 | 361.00 |
| *LZTFL1* | rs1354034 | T | C | 0.6044 | 0.072 | 0.008 | 7.9433E-21 | 81.00 |
| *LZTFL1* | rs574628490 | G | GT | 0.1252 | 0.087 | 0.012 | 3.1623E-12 | 52.56 |
| *LZTFL1* | rs61237993 | G | A | 0.1273 | 0.089 | 0.011 | 5.0119E-15 | 65.46 |
| *LZTFL1* | rs11502185 | T | C | 0.2597 | 0.067 | 0.01 | 3.9811E-12 | 44.89 |
| *LZTFL1* | rs60822569 | T | TTTCTC | 0.5542 | 0.053 | 0.008 | 3.9811E-12 | 43.89 |
| *M6PR* | rs4883201 | A | G | 0.104 | -0.28 | 0.013 | 3.162E-108 | 463.91 |
| *MAD1L1* | rs143579216 | G | A | 0.0028 | 1.275 | 0.075 | 3.9811E-64 | 289.00 |
| *MAGED1* | rs1354034 | T | C | 0.6038 | 0.06 | 0.008 | 1.5849E-14 | 56.25 |
| *MAGED1* | rs342293 | C | G | 0.4596 | -0.074 | 0.008 | 3.9811E-22 | 85.56 |
| *MAMDC2* | rs768375575 | C | CCTT | 0.0964 | 0.085 | 0.012 | 1E-11 | 50.17 |
| *MAMDC2* | rs13412535 | G | A | 0.2311 | 0.068 | 0.009 | 1.5849E-15 | 57.09 |
| *MAMDC2* | rs7580831 | C | A | 0.3261 | -0.109 | 0.008 | 5.0119E-45 | 185.64 |
| *MAMDC2* | rs12646270 | G | A | 0.0551 | 0.196 | 0.016 | 3.1623E-35 | 150.06 |
| *MAMDC2* | rs13107325 | C | T | 0.0751 | 0.196 | 0.014 | 7.9433E-46 | 196.00 |
| *MAMDC2* | rs62572585 | C | A | 0.2584 | 0.224 | 0.008 | 1.259E-161 | 784.00 |
| *MAMDC2* | rs68055275 | C | CT | 0.1449 | -0.089 | 0.01 | 1.9953E-17 | 79.21 |
| *MAMDC2* | rs4981022 | G | A | 0.6813 | 0.058 | 0.008 | 1E-13 | 52.56 |
| *MAMDC2* | rs147233090 | C | T | 0.0236 | -0.184 | 0.024 | 1.2589E-14 | 58.78 |
| *MAMDC2* | rs11854097 | A | G | 0.2557 | -0.058 | 0.008 | 2.5119E-12 | 52.56 |
| *MAMDC2* | rs186021206 | G | A | 0.0064 | 0.476 | 0.05 | 7.9433E-22 | 90.63 |
| *MAMDC2* | rs11701038 | C | T | 0.1619 | 0.068 | 0.01 | 6.3096E-12 | 46.24 |
| *MAMDC4* | rs12740374 | G | T | 0.2208 | -0.162 | 0.009 | 2.5119E-75 | 324.00 |
| *MAMDC4* | rs1260326 | T | C | 0.6084 | -0.135 | 0.008 | 2.5119E-72 | 284.77 |
| *MAMDC4* | rs13240065 | G | A | 0.1275 | -0.178 | 0.011 | 2.5119E-59 | 261.85 |
| *MAMDC4* | rs35523349 | C | CA | 0.4593 | -0.056 | 0.008 | 1.2589E-13 | 49.00 |
| *MAN1A2* | rs12740374 | G | T | 0.2212 | -0.232 | 0.009 | 1.585E-160 | 664.49 |
| *MAN1A2* | rs10923344 | G | A | 0.6177 | 0.193 | 0.007 | 2.512E-152 | 760.18 |
| *MAN1A2* | rs4846913 | C | A | 0.6071 | 0.052 | 0.007 | 1.9953E-12 | 55.18 |
| *MAN1A2* | rs13153570 | A | G | 0.4435 | -0.051 | 0.007 | 3.1623E-13 | 53.08 |
| *MAN1A2* | rs6908943 | C | T | 0.1765 | -0.103 | 0.009 | 2.5119E-27 | 130.98 |
| *MAN1A2* | rs10982154 | T | A | 0.0688 | 0.124 | 0.015 | 2.5119E-17 | 68.34 |
| *MAN1A2* | rs56278466 | T | G | 0.6614 | 0.133 | 0.007 | 3.9811E-72 | 361.00 |
| *MAN1A2* | rs77542162 | A | G | 0.0229 | 0.234 | 0.023 | 1.5849E-23 | 103.51 |
| *MAN1A2* | rs28873836 | G | C | 0.266 | -0.198 | 0.008 | 2.512E-135 | 612.56 |
| *MAN2B2* | rs114165349 | G | C | 0.0233 | 0.141 | 0.018 | 1.9953E-15 | 61.36 |
| *MAN2B2* | rs687339 | C | T | 0.7713 | 0.073 | 0.006 | 1.5849E-30 | 148.03 |
| *MAN2B2* | rs2301791 | T | A | 0.4782 | 0.824 | 0.007 | 1E-200 | 13856.65 |
| *MAN2B2* | rs77945361 | G | A | 0.1484 | -0.077 | 0.01 | 3.1623E-14 | 59.29 |
| *MAN2B2* | rs4895864 | C | T | 0.5746 | 0.055 | 0.005 | 1E-24 | 121.00 |
| *MAN2B2* | rs9414801 | G | A | 0.5332 | -0.048 | 0.005 | 5.0119E-19 | 92.16 |
| *MAN2B2* | rs41280378 | T | G | 0.0429 | -0.102 | 0.013 | 7.9433E-15 | 61.56 |
| *MAN2B2* | rs10745925 | T | C | 0.2876 | -0.361 | 0.006 | 1E-200 | 3620.03 |
| *MAN2B2* | rs71435622 | A | AT | 0.2868 | 0.049 | 0.006 | 1.9953E-15 | 66.69 |
| *MAN2B2* | rs145078947 | G | T | 0.0025 | 0.758 | 0.053 | 5.0119E-46 | 204.54 |
| *MAN2B2* | rs58542926 | C | T | 0.0755 | 0.193 | 0.01 | 3.1623E-81 | 372.49 |
| *MAN2B2* | rs429358 | T | C | 0.1553 | -0.091 | 0.007 | 2.5119E-34 | 169.00 |
| *MAN2B2* | rs738409 | C | G | 0.2172 | 0.066 | 0.006 | 1E-24 | 121.00 |
| *MANEAL* | rs11016919 | C | T | 0.137 | 0.102 | 0.011 | 3.1623E-19 | 85.98 |
| *MANF* | rs11721253 | A | G | 0.8648 | 0.098 | 0.011 | 3.9811E-19 | 79.37 |
| *MANF* | rs1354034 | T | C | 0.6041 | 0.065 | 0.008 | 1.9953E-17 | 66.02 |
| *MANF* | rs3804749 | C | T | 0.5926 | -0.052 | 0.008 | 1.5849E-11 | 42.25 |
| *MANF* | rs342298 | C | T | 0.4545 | -0.065 | 0.008 | 1E-17 | 66.02 |
| *MANF* | rs6993770 | A | T | 0.2867 | -0.059 | 0.008 | 1.2589E-12 | 54.39 |
| *MANF* | rs892090 | T | G | 0.8342 | 0.073 | 0.01 | 7.9433E-13 | 53.29 |
| *MANSC1* | rs3917932 | C | G | 0.576 | -0.057 | 0.008 | 6.3096E-14 | 50.77 |

| | | | | | | | | |
|---|---|---|---|---|---|---|---|---|
| *MANSC1* | rs61747728 | C | T | 0.0379 | 0.182 | 0.019 | 1.2589E-20 | 91.76 |
| *MANSC1* | rs1260326 | T | C | 0.6087 | 0.082 | 0.008 | 7.9433E-27 | 105.06 |
| *MANSC1* | rs6705658 | C | T | 0.6418 | 0.053 | 0.008 | 6.3096E-12 | 43.89 |
| *MANSC1* | rs75166367 | G | A | 0.0604 | -0.122 | 0.016 | 6.3096E-15 | 58.14 |
| *MANSC1* | rs11721168 | G | A | 0.376 | 0.056 | 0.008 | 3.9811E-13 | 49.00 |
| *MANSC1* | rs112926042 | C | T | 0.3092 | 0.056 | 0.008 | 5.0119E-12 | 49.00 |
| *MANSC1* | rs4861708 | G | A | 0.5119 | -0.073 | 0.007 | 1.9953E-22 | 108.76 |
| *MANSC1* | rs2143466 | C | T | 0.3857 | -0.063 | 0.008 | 1.5849E-16 | 62.02 |
| *MANSC1* | rs11137085 | G | C | 0.3848 | 0.059 | 0.008 | 5.0119E-14 | 54.39 |
| *MANSC1* | rs495828 | G | T | 0.2082 | -0.088 | 0.009 | 1.2589E-21 | 95.60 |
| *MANSC1* | rs1059091 | A | G | 0.3184 | -0.06 | 0.008 | 1E-13 | 56.25 |
| *MANSC1* | rs73017394 | C | A | 0.2392 | -0.139 | 0.009 | 6.3096E-56 | 238.53 |
| *MANSC1* | rs9506687 | G | A | 0.2685 | 0.061 | 0.008 | 5.0119E-13 | 58.14 |
| *MANSC1* | rs4997081 | G | C | 0.1925 | -0.068 | 0.01 | 7.9433E-13 | 46.24 |
| *MANSC1* | rs186021206 | G | A | 0.0064 | 0.735 | 0.051 | 3.1623E-46 | 207.70 |
| *MANSC1* | rs12941811 | T | C | 0.5746 | -0.063 | 0.008 | 5.0119E-17 | 62.02 |
| *MANSC1* | rs55715868 | A | G | 0.5366 | -0.059 | 0.008 | 3.1623E-14 | 54.39 |
| *MANSC4* | rs17482078 | C | T | 0.225 | 0.053 | 0.007 | 7.9433E-13 | 57.33 |
| *MANSC4* | rs9275109 | G | T | 0.6176 | -0.135 | 0.006 | 6.3096E-96 | 506.25 |
| *MANSC4* | rs10995299 | C | T | 0.4109 | 0.076 | 0.006 | 7.9433E-33 | 160.44 |
| *MANSC4* | rs34311349 | T | A | 0.2278 | 0.938 | 0.009 | 1E-200 | 10862.27 |
| *MAP1LC3B2* | rs58534292 | T | C | 0.6506 | -0.057 | 0.008 | 3.1623E-12 | 50.77 |
| *MAP1LC3B2* | rs2857338 | T | G | 0.4977 | 0.092 | 0.009 | 3.9811E-25 | 104.49 |
| *MAP2* | rs1342442 | G | A | 0.6528 | 0.073 | 0.008 | 1E-20 | 83.27 |
| *MAP2* | rs113770841 | G | C | 0.0151 | 0.435 | 0.031 | 6.3096E-46 | 196.90 |
| *MAP2* | rs78909033 | G | A | 0.1359 | 0.114 | 0.011 | 1.5849E-25 | 107.40 |
| *MAP2* | rs1354034 | T | C | 0.6043 | 0.071 | 0.008 | 1.2589E-20 | 78.77 |
| *MAP2* | rs342293 | C | G | 0.46 | -0.059 | 0.007 | 3.1623E-15 | 71.04 |
| *MAP2* | rs4759076 | T | C | 0.4571 | 0.085 | 0.008 | 3.1623E-29 | 112.89 |
| *MAP2* | rs10418205 | A | G | 0.3738 | -0.059 | 0.008 | 3.1623E-14 | 54.39 |
| *MAP2K1* | rs1354034 | T | C | 0.6046 | 0.076 | 0.008 | 6.3096E-24 | 90.25 |
| *MAP2K1* | rs6993770 | A | T | 0.2866 | -0.065 | 0.008 | 3.9811E-15 | 66.02 |
| *MAP2K1* | rs12245367 | C | T | 0.4813 | -0.054 | 0.007 | 3.9811E-13 | 59.51 |
| *MAP2K1* | rs77428206 | C | A | 0.0961 | 0.086 | 0.013 | 1.5849E-11 | 43.76 |
| *MAP2K6* | rs342298 | C | T | 0.4546 | -0.064 | 0.008 | 1.9953E-17 | 64.00 |
| *MAP2K6* | rs7896518 | A | G | 0.4241 | 0.06 | 0.008 | 6.3096E-15 | 56.25 |
| *MAP2K6* | rs60822569 | T | TTTCTC | 0.5547 | 0.053 | 0.008 | 3.1623E-12 | 43.89 |
| *MAP3K5* | rs1354034 | T | C | 0.6038 | 0.055 | 0.008 | 2.5119E-13 | 47.27 |
| *MAP4K5* | rs17780143 | G | A | 0.0575 | -0.373 | 0.016 | 7.943E-119 | 543.47 |
| *MAPK13* | rs12210904 | C | A | 0.2938 | 0.066 | 0.009 | 1.2589E-14 | 53.78 |
| *MAPK9* | rs3763131 | T | C | 0.1951 | 0.143 | 0.009 | 2.5119E-51 | 252.46 |
| *MAPKAPK2* | rs11119390 | C | G | 0.2986 | 0.147 | 0.008 | 1.5849E-71 | 337.64 |
| *MAPRE3* | rs71534539 | G | A | 0.464 | -0.125 | 0.009 | 3.1623E-48 | 192.90 |
| *MAPRE3* | rs952558 | A | T | 0.6256 | 0.061 | 0.008 | 1.2589E-14 | 58.14 |
| *MARCO* | rs12740374 | G | T | 0.221 | -0.13 | 0.009 | 3.9811E-51 | 208.64 |
| *MARCO* | rs4672487 | T | G | 0.5684 | -0.092 | 0.007 | 3.1623E-35 | 172.73 |
| *MARCO* | rs35288880 | C | T | 0.1632 | -0.34 | 0.01 | 1E-200 | 1156.00 |
| *MARCO* | rs62165726 | C | A | 0.037 | -0.365 | 0.019 | 6.3096E-79 | 369.04 |
| *MARCO* | rs59859734 | A | T | 0.4142 | -0.07 | 0.007 | 1.2589E-21 | 100.00 |
| *MARCO* | rs112125244 | T | TTTTG | 0.2851 | -0.059 | 0.008 | 2.5119E-13 | 54.39 |
| *MARCO* | rs12667978 | C | T | 0.5699 | 0.055 | 0.007 | 5.0119E-14 | 61.73 |
| *MARCO* | rs73338697 | G | A | 0.0073 | -0.309 | 0.043 | 1.2589E-12 | 51.64 |
| *MARCO* | rs11861563 | G | A | 0.5856 | -0.062 | 0.007 | 3.9811E-17 | 78.45 |
| *MARCO* | rs186021206 | G | A | 0.0064 | 1.09 | 0.049 | 1.995E-109 | 494.84 |
| *MARS1* | rs1354034 | T | C | 0.6044 | 0.076 | 0.008 | 1.9953E-23 | 90.25 |
| *MARS1* | rs9328454 | C | T | 0.2903 | -0.056 | 0.008 | 1E-11 | 49.00 |
| *MARS1* | rs342298 | C | T | 0.4547 | -0.058 | 0.008 | 1.9953E-14 | 52.56 |
| *MASP1* | rs3765901 | G | A | 0.7893 | 0.123 | 0.009 | 1.5849E-43 | 186.78 |
| *MASP1* | rs3820897 | T | C | 0.8215 | 0.192 | 0.01 | 6.3096E-90 | 368.64 |
| *MASP1* | rs1486236 | C | A | 0.3735 | 0.314 | 0.008 | 1E-200 | 1540.56 |
| *MASP1* | rs698090 | C | T | 0.6443 | 0.213 | 0.008 | 7.943E-171 | 708.89 |
| *MASP1* | rs9987289 | A | G | 0.908 | -0.091 | 0.012 | 1.5849E-13 | 57.51 |
| *MASP1* | rs41341748 | G | A | 0.0111 | 0.257 | 0.034 | 3.9811E-14 | 57.14 |
| *MASP1* | rs10104003 | C | T | 0.2095 | 0.099 | 0.009 | 3.1623E-29 | 121.00 |
| *MASP1* | rs7899547 | T | G | 0.6529 | 0.133 | 0.008 | 3.9811E-70 | 276.39 |
| *MASP1* | rs4926 | G | A | 0.2744 | -0.088 | 0.008 | 5.0119E-28 | 121.00 |
| *MASP1* | rs215223 | G | A | 0.4002 | -0.099 | 0.007 | 3.9811E-42 | 200.02 |
| *MASP1* | rs6118 | C | T | 0.0998 | 0.1 | 0.012 | 3.1623E-17 | 69.44 |

| | | | | | | | | |
|---|---|---|---|---|---|---|---|---|
| *MASP1* | rs704 | G | A | 0.4733 | -0.071 | 0.007 | 2.5119E-23 | 102.88 |
| *MASP1* | rs9944724 | T | G | 0.3773 | 0.06 | 0.007 | 2.5119E-16 | 73.47 |
| *MATN2* | rs13159365 | C | T | 0.5104 | 0.08 | 0.007 | 1.5849E-30 | 130.61 |
| *MATN2* | rs2685412 | C | G | 0.2706 | 0.257 | 0.008 | 1E-200 | 1032.02 |
| *MATN2* | rs1869609 | A | G | 0.6633 | 0.396 | 0.008 | 1E-200 | 2450.25 |
| *MATN2* | rs35157100 | G | GA | 0.2915 | -0.168 | 0.008 | 1.995E-106 | 441.00 |
| *MATN2* | rs2269434 | T | C | 0.3468 | 0.052 | 0.007 | 1.2589E-12 | 55.18 |
| *MATN2* | rs77542162 | A | G | 0.0229 | 0.161 | 0.023 | 3.9811E-12 | 49.00 |
| *MATN3* | rs11694716 | G | A | 0.6364 | -0.564 | 0.008 | 1E-200 | 4970.25 |
| *MATN3* | rs66530140 | T | C | 0.5134 | -0.058 | 0.007 | 6.3096E-18 | 68.65 |
| *MATN3* | rs2519093 | C | T | 0.1844 | -0.086 | 0.009 | 3.1623E-23 | 91.31 |
| *MATN3* | rs11657101 | G | A | 0.3622 | -0.119 | 0.007 | 1.9953E-64 | 289.00 |
| *MAVS* | rs7080386 | C | A | 0.4121 | 0.055 | 0.008 | 3.9811E-13 | 47.27 |
| *MAVS* | rs45437096 | C | T | 0.1454 | -0.135 | 0.011 | 7.9433E-34 | 150.62 |
| *MAX* | rs17622656 | G | A | 0.3883 | -0.059 | 0.008 | 7.9433E-14 | 54.39 |
| *MAX* | rs342298 | C | T | 0.4544 | -0.054 | 0.008 | 2.5119E-12 | 45.56 |
| *MAX* | rs13095 | G | C | 0.4694 | 0.059 | 0.008 | 1.5849E-14 | 54.39 |
| *MAX* | rs8005829 | C | T | 0.5831 | 0.078 | 0.008 | 5.0119E-24 | 95.06 |
| *MAX* | rs12588296 | A | C | 0.2512 | -0.067 | 0.009 | 6.3096E-14 | 55.42 |
| *MB* | rs12463674 | A | G | 0.3157 | 0.076 | 0.007 | 2.5119E-27 | 117.88 |
| *MB* | rs2151597 | G | A | 0.4388 | 0.05 | 0.007 | 1E-14 | 51.02 |
| *MB* | rs2234962 | T | C | 0.2156 | 0.056 | 0.008 | 1E-12 | 49.00 |
| *MB* | rs7481951 | A | T | 0.588 | 0.069 | 0.007 | 6.3096E-25 | 97.16 |
| *MB* | rs1028883 | T | G | 0.5708 | 0.065 | 0.007 | 1.5849E-22 | 86.22 |
| *MB* | rs4901541 | C | T | 0.5612 | -0.054 | 0.007 | 2.5119E-16 | 59.51 |
| *MB* | rs1799938 | G | A | 0.1141 | 0.071 | 0.01 | 3.1623E-12 | 50.41 |
| *MB* | rs5844883 | T | TA | 0.4227 | -0.048 | 0.007 | 6.3096E-13 | 47.02 |
| *MBL2* | rs1260326 | T | C | 0.608 | 0.065 | 0.005 | 1.2589E-37 | 169.00 |
| *MBL2* | rs9398803 | A | G | 0.4728 | 0.049 | 0.005 | 1E-22 | 96.04 |
| *MBL2* | rs112875651 | G | A | 0.3911 | 0.038 | 0.005 | 6.3096E-14 | 57.76 |
| *MBL2* | rs505922 | T | C | 0.3172 | 0.258 | 0.005 | 1E-200 | 2662.56 |
| *MBL2* | rs7899547 | T | G | 0.6536 | 0.915 | 0.008 | 1E-200 | 13081.64 |
| *MBL2* | rs147741557 | G | GA | 0.1606 | -0.054 | 0.007 | 1E-15 | 59.51 |
| *MCAM* | rs1260326 | T | C | 0.6083 | 0.069 | 0.007 | 2.5119E-21 | 97.16 |
| *MCAM* | rs13107325 | C | T | 0.075 | 0.105 | 0.014 | 1E-14 | 56.25 |
| *MCAM* | rs41289902 | C | T | 0.0143 | 0.484 | 0.03 | 7.9433E-59 | 260.28 |
| *MCAM* | rs8176693 | C | T | 0.0613 | 0.229 | 0.015 | 1E-53 | 233.07 |
| *MCAM* | rs34587557 | T | C | 0.051 | -0.324 | 0.016 | 6.3096E-89 | 410.06 |
| *MCAM* | rs11220462 | G | A | 0.1327 | -0.099 | 0.011 | 6.3096E-21 | 81.00 |
| *MCAM* | rs7310615 | C | G | 0.5175 | 0.056 | 0.007 | 2.5119E-15 | 64.00 |
| *MCAM* | rs186021206 | G | A | 0.0064 | 0.967 | 0.049 | 1E-86 | 389.46 |
| *MCAM* | rs7286917 | A | G | 0.7447 | -0.075 | 0.008 | 1E-18 | 87.89 |
| *MCEE* | rs6716795 | T | C | 0.536 | -0.188 | 0.008 | 3.981E-132 | 552.25 |
| *MCEMP1* | rs1342442 | G | A | 0.6529 | 0.113 | 0.008 | 3.9811E-47 | 199.52 |
| *MCEMP1* | rs78909033 | G | A | 0.1364 | 0.103 | 0.011 | 3.1623E-21 | 87.68 |
| *MCEMP1* | rs1354034 | T | C | 0.6039 | 0.053 | 0.008 | 1.9953E-12 | 43.89 |
| *MCEMP1* | rs114694170 | T | C | 0.0602 | -0.2 | 0.016 | 1.5849E-36 | 156.25 |
| *MCEMP1* | rs6993770 | A | T | 0.2866 | -0.121 | 0.008 | 3.1623E-49 | 228.77 |
| *MCEMP1* | rs10820606 | A | C | 0.2293 | 0.156 | 0.009 | 6.3096E-66 | 300.44 |
| *MCEMP1* | rs8193004 | C | T | 0.6022 | -0.083 | 0.008 | 1E-26 | 107.64 |
| *MCEMP1* | rs10776883 | C | T | 0.3121 | 0.278 | 0.008 | 1E-200 | 1207.56 |
| *MCEMP1* | rs71474568 | T | TTG | 0.3398 | 0.088 | 0.008 | 6.3096E-29 | 121.00 |
| *MCEMP1* | rs73000929 | G | A | 0.0374 | -0.168 | 0.02 | 1.2589E-17 | 70.56 |
| *MCEMP1* | rs4759076 | T | C | 0.457 | 0.07 | 0.008 | 2.5119E-20 | 76.56 |
| *MCEMP1* | rs4783183 | C | G | 0.8744 | -0.109 | 0.011 | 2.5119E-22 | 98.19 |
| *MCEMP1* | rs3848232 | G | C | 0.2952 | 0.059 | 0.008 | 7.9433E-13 | 54.39 |
| *MCEMP1* | rs141879681 | A | G | 0.0064 | 1.432 | 0.049 | 6.31E-187 | 854.07 |
| *MCFD2* | rs6419615 | G | C | 0.7071 | -0.146 | 0.008 | 3.1623E-68 | 333.06 |
| *MCFD2* | rs6993770 | A | T | 0.2866 | -0.079 | 0.008 | 3.1623E-21 | 97.52 |
| *MCFD2* | rs112635299 | G | T | 0.0216 | 0.346 | 0.026 | 1.5849E-40 | 177.09 |
| *MCFD2* | rs17696641 | T | A | 0.148 | 0.085 | 0.011 | 1.2589E-15 | 59.71 |
| *MCTS1* | rs72508413 | A | G | 0.3426 | -0.102 | 0.008 | 3.9811E-36 | 162.56 |
| *MDGA1* | rs9349050 | T | C | 0.5143 | -1.08 | 0.007 | 1E-200 | 23804.08 |
| *MDGA1* | rs9987289 | A | G | 0.908 | -0.062 | 0.008 | 6.3096E-14 | 60.06 |
| *MDGA1* | rs35887873 | T | C | 0.2283 | 0.068 | 0.006 | 1.9953E-33 | 128.44 |
| *MDGA1* | rs186021206 | G | A | 0.0064 | 0.508 | 0.033 | 1E-54 | 236.97 |
| *MDGA1* | rs77542162 | A | G | 0.023 | 0.139 | 0.016 | 1.5849E-18 | 75.47 |
| *MDGA1* | rs7287617 | G | A | 0.645 | 0.039 | 0.005 | 5.0119E-15 | 60.84 |

| | | | | | | | | |
|---|---|---|---|---|---|---|---|---|
| *MDH1* | rs71422399 | C | T | 0.2652 | 0.092 | 0.008 | 1.2589E-27 | 132.25 |
| *MDK* | rs6427756 | A | G | 0.735 | -0.081 | 0.008 | 1.2589E-21 | 102.52 |
| *MDK* | rs7618405 | C | A | 0.2047 | -0.067 | 0.009 | 5.0119E-13 | 55.42 |
| *MDK* | rs1354034 | T | C | 0.6041 | -0.059 | 0.008 | 6.3096E-15 | 54.39 |
| *MDK* | rs9358555 | A | G | 0.139 | -0.073 | 0.011 | 1.2589E-11 | 44.04 |
| *MDK* | rs6961069 | C | T | 0.4023 | 0.052 | 0.008 | 1E-11 | 42.25 |
| *MDK* | rs6993770 | A | T | 0.2861 | -0.081 | 0.008 | 7.9433E-23 | 102.52 |
| *MDK* | rs7080386 | C | A | 0.4122 | 0.069 | 0.008 | 3.9811E-20 | 74.39 |
| *MDK* | rs541295226 | A | G | 0.0019 | -1.689 | 0.102 | 3.9811E-61 | 274.19 |
| *MDK* | rs117125830 | C | T | 0.0059 | -0.515 | 0.049 | 3.9811E-26 | 110.46 |
| *MDK* | rs3184504 | T | C | 0.5173 | -0.067 | 0.007 | 1E-19 | 91.61 |
| *MDK* | rs12445050 | C | T | 0.1384 | 0.075 | 0.011 | 3.9811E-12 | 46.49 |
| *MDK* | rs57839578 | A | T | 0.4792 | -0.052 | 0.007 | 3.1623E-12 | 55.18 |
| *MDK* | rs892090 | T | G | 0.8343 | 0.123 | 0.01 | 1.5849E-34 | 151.29 |
| *MDK* | rs14142 | C | T | 0.3729 | -0.062 | 0.008 | 1.2589E-15 | 60.06 |
| *MDM1* | rs2870812 | G | A | 0.7128 | -0.202 | 0.008 | 3.981E-125 | 637.56 |
| *MECR* | rs144612117 | C | CCCTCTGG/ | 0.1352 | 0.107 | 0.011 | 2.5119E-21 | 94.62 |
| *MECR* | rs11242109 | G | T | 0.4812 | -0.057 | 0.008 | 1E-13 | 50.77 |
| *MECR* | rs6993770 | A | T | 0.2866 | -0.063 | 0.008 | 5.0119E-14 | 62.02 |
| *MECR* | rs7896518 | A | G | 0.4236 | 0.054 | 0.008 | 5.0119E-12 | 45.56 |
| *MED18* | rs12245149 | C | A | 0.4881 | -0.057 | 0.008 | 7.9433E-14 | 50.77 |
| *MED18* | rs200309755 | C | T | 0.5373 | 0.055 | 0.008 | 3.9811E-12 | 47.27 |
| *MEGF10* | rs62276885 | G | T | 0.1814 | 0.06 | 0.009 | 7.9433E-12 | 44.44 |
| *MEGF10* | rs112394041 | C | CT | 0.4583 | 0.398 | 0.007 | 1E-200 | 3232.73 |
| *MEGF10* | rs6938239 | G | A | 0.8607 | -0.066 | 0.01 | 1E-11 | 43.56 |
| *MEGF10* | rs200889152 | A | C | 0.3749 | -0.065 | 0.007 | 5.0119E-21 | 86.22 |
| *MEGF10* | rs10813959 | T | C | 0.4223 | 0.077 | 0.007 | 3.9811E-30 | 121.00 |
| *MEGF10* | rs508196 | C | T | 0.4358 | -0.053 | 0.007 | 7.9433E-15 | 57.33 |
| *MEGF10* | rs7232 | T | A | 0.3735 | 0.051 | 0.007 | 1.2589E-13 | 53.08 |
| *MEGF10* | rs2236653 | C | T | 0.4261 | 0.051 | 0.007 | 3.1623E-14 | 53.08 |
| *MEGF10* | rs10849082 | T | C | 0.4675 | -0.06 | 0.007 | 3.9811E-19 | 73.47 |
| *MEGF10* | rs632887 | A | G | 0.4108 | 0.08 | 0.007 | 1.9953E-31 | 130.61 |
| *MEGF10* | rs3007036 | A | G | 0.5392 | 0.057 | 0.007 | 3.1623E-17 | 66.31 |
| *MEGF10* | rs186021206 | G | A | 0.0064 | 0.668 | 0.047 | 3.9811E-46 | 202.00 |
| *MEGF10* | rs77542162 | A | G | 0.0229 | 0.799 | 0.023 | 1E-200 | 1206.81 |
| *MEGF10* | rs6068599 | T | C | 0.6818 | -0.057 | 0.007 | 3.1623E-15 | 66.31 |
| *MEGF11* | rs11630800 | C | T | 0.2107 | 0.164 | 0.009 | 5.0119E-68 | 332.05 |
| *MEGF11* | rs77542162 | A | G | 0.0229 | 0.411 | 0.026 | 7.9433E-58 | 249.88 |
| *MEGF9* | rs646776 | C | T | 0.7779 | -0.084 | 0.009 | 6.3096E-22 | 87.11 |
| *MEGF9* | rs4652308 | T | C | 0.4746 | 0.055 | 0.007 | 5.0119E-14 | 61.73 |
| *MEGF9* | rs13107325 | C | T | 0.0749 | 0.182 | 0.014 | 3.9811E-39 | 169.00 |
| *MEGF9* | rs80318131 | CT | C | 0.3058 | 0.065 | 0.008 | 7.9433E-15 | 66.02 |
| *MEGF9* | rs1461729 | A | G | 0.8974 | -0.089 | 0.012 | 1.2589E-13 | 55.01 |
| *MEGF9* | rs28601761 | C | G | 0.4194 | -0.081 | 0.007 | 1.2589E-27 | 133.90 |
| *MEGF9* | rs10984978 | T | C | 0.7008 | 0.341 | 0.008 | 1E-200 | 1816.89 |
| *MEGF9* | rs9633740 | A | G | 0.7954 | 0.079 | 0.009 | 2.5119E-18 | 77.05 |
| *MEGF9* | rs751229834 | CA | C | 0.3069 | 0.079 | 0.008 | 2.5119E-23 | 97.52 |
| *MEGF9* | rs11045819 | C | A | 0.1608 | 0.152 | 0.01 | 3.9811E-53 | 231.04 |
| *MEGF9* | rs186021206 | G | A | 0.0064 | 0.46 | 0.05 | 6.3096E-20 | 84.64 |
| *MEGF9* | rs77542162 | A | G | 0.0229 | 0.26 | 0.024 | 1.5849E-26 | 117.36 |
| *MEGF9* | rs73066226 | T | C | 0.1636 | 0.083 | 0.01 | 3.1623E-17 | 68.89 |
| *MELTF* | rs8177245 | A | G | 0.3419 | 0.054 | 0.008 | 5.0119E-12 | 45.56 |
| *MELTF* | rs3773862 | C | T | 0.4528 | -0.408 | 0.008 | 1E-200 | 2601.00 |
| *MELTF* | rs13135092 | A | G | 0.0836 | 0.101 | 0.012 | 3.9811E-17 | 70.84 |
| *MELTF* | rs55818129 | T | G | 0.1176 | -0.074 | 0.01 | 5.0119E-13 | 54.76 |
| *MELTF* | rs79220007 | T | C | 0.0773 | -0.267 | 0.012 | 1.995E-103 | 495.06 |
| *MELTF* | rs56278466 | T | G | 0.6612 | 0.107 | 0.007 | 1.9953E-53 | 233.65 |
| *MELTF* | rs174576 | C | A | 0.3523 | 0.054 | 0.007 | 5.0119E-15 | 59.51 |
| *MELTF* | rs61755050 | T | C | 0.006 | -0.32 | 0.043 | 5.0119E-14 | 55.38 |
| *MELTF* | rs2076085 | C | A | 0.5347 | -0.057 | 0.007 | 1.9953E-18 | 66.31 |
| *MENT* | rs12132412 | A | G | 0.3902 | -0.081 | 0.007 | 7.9433E-32 | 133.90 |
| *MENT* | rs13402561 | C | G | 0.8172 | -0.092 | 0.009 | 3.1623E-26 | 104.49 |
| *MENT* | rs1486236 | C | A | 0.3732 | -0.072 | 0.007 | 1E-24 | 105.80 |
| *MENT* | rs850313 | C | G | 0.7122 | -0.059 | 0.007 | 1.5849E-15 | 71.04 |
| *MENT* | rs1372344 | A | T | 0.2612 | 0.099 | 0.008 | 3.1623E-39 | 153.14 |
| *MENT* | rs964184 | G | C | 0.8667 | 0.131 | 0.01 | 1.2589E-40 | 171.61 |
| *MENT* | rs821840 | A | G | 0.3248 | 0.122 | 0.007 | 2.5119E-65 | 303.76 |
| *MENT* | rs116843064 | G | A | 0.0195 | 0.201 | 0.026 | 6.3096E-15 | 59.76 |

| | | | | | | | | |
|---|---|---|---|---|---|---|---|---|
| *MENT* | rs200210321 | A | AG | 0.0741 | -0.127 | 0.014 | 1.5849E-20 | 82.29 |
| *MENT* | rs483082 | G | T | 0.2372 | 0.567 | 0.008 | 1E-200 | 5023.27 |
| *MEP1A* | rs1047891 | C | A | 0.3128 | 0.065 | 0.008 | 3.1623E-18 | 66.02 |
| *MEP1A* | rs299284 | C | T | 0.1064 | 0.152 | 0.011 | 1.2589E-41 | 190.94 |
| *MEP1A* | rs2274656 | G | A | 0.3848 | -0.194 | 0.007 | 3.162E-160 | 768.08 |
| *MEP1A* | rs2954021 | A | G | 0.5037 | -0.047 | 0.007 | 1.2589E-11 | 45.08 |
| *MEP1A* | rs10733608 | G | T | 0.4888 | -0.15 | 0.007 | 1.9953E-98 | 459.18 |
| *MEP1A* | rs10793962 | A | T | 0.0615 | 0.377 | 0.015 | 6.31E-147 | 631.68 |
| *MEP1A* | rs2185756 | A | C | 0.4062 | -0.077 | 0.007 | 3.9811E-27 | 121.00 |
| *MEP1A* | rs2862954 | T | C | 0.5018 | -0.16 | 0.007 | 7.943E-117 | 522.45 |
| *MEP1A* | rs339969 | C | A | 0.6169 | 0.056 | 0.007 | 3.1623E-15 | 64.00 |
| *MEP1A* | rs545303153 | G | GGGGGCA | 0.003 | 0.517 | 0.073 | 1.2589E-12 | 50.16 |
| *MEP1A* | rs12950511 | C | T | 0.3406 | 0.052 | 0.007 | 1.9953E-12 | 55.18 |
| *MEP1A* | rs11662351 | T | C | 0.3907 | -0.216 | 0.007 | 3.981E-197 | 952.16 |
| *MEP1A* | rs708686 | C | T | 0.2677 | 0.181 | 0.008 | 6.31E-116 | 511.89 |
| *MEP1A* | rs681343 | C | T | 0.5092 | -0.156 | 0.007 | 1.259E-108 | 496.65 |
| *MEP1A* | rs1800961 | C | T | 0.0324 | -0.244 | 0.02 | 7.9433E-36 | 148.84 |
| *MEP1B* | rs193084249 | A | G | 0.0236 | -0.153 | 0.016 | 2.5119E-21 | 91.44 |
| *MEP1B* | rs1730862 | G | A | 0.6574 | 0.041 | 0.005 | 1.2589E-15 | 67.24 |
| *MEP1B* | rs1260326 | T | C | 0.6083 | 0.058 | 0.005 | 7.9433E-32 | 134.56 |
| *MEP1B* | rs370213063 | CT | C | 0.3076 | -0.041 | 0.005 | 6.3096E-15 | 67.24 |
| *MEP1B* | rs9884482 | T | C | 0.3725 | -0.036 | 0.005 | 3.1623E-13 | 51.84 |
| *MEP1B* | rs4895864 | C | T | 0.5746 | 0.039 | 0.005 | 7.9433E-16 | 60.84 |
| *MEP1B* | rs6973520 | C | T | 0.5061 | 0.036 | 0.005 | 7.9433E-14 | 51.84 |
| *MEP1B* | rs3735165 | T | C | 0.3091 | 0.072 | 0.005 | 2.5119E-44 | 207.36 |
| *MEP1B* | rs4292651 | C | A | 0.6087 | -0.033 | 0.005 | 1.5849E-11 | 43.56 |
| *MEP1B* | rs112875651 | G | A | 0.391 | 0.042 | 0.005 | 6.3096E-17 | 70.56 |
| *MEP1B* | rs174537 | G | T | 0.3452 | 0.052 | 0.005 | 6.3096E-25 | 108.16 |
| *MEP1B* | rs11045856 | T | G | 0.2404 | 0.044 | 0.006 | 3.9811E-15 | 53.78 |
| *MEP1B* | rs7979473 | A | G | 0.6162 | -0.051 | 0.005 | 6.3096E-25 | 104.04 |
| *MEP1B* | rs17616063 | A | G | 0.0747 | -0.104 | 0.009 | 5.0119E-30 | 133.53 |
| *MEP1B* | rs16940186 | T | C | 0.1488 | 0.135 | 0.007 | 1E-87 | 371.94 |
| *MEP1B* | rs7212366 | C | T | 0.4911 | -0.047 | 0.005 | 6.3096E-20 | 88.36 |
| *MEP1B* | rs680321 | T | C | 0.456 | -1.074 | 0.008 | 1E-200 | 18023.06 |
| *MEP1B* | rs1800961 | C | T | 0.0322 | 0.098 | 0.014 | 3.9811E-13 | 49.00 |
| *MEPE* | rs181163165 | C | T | 0.0048 | -0.431 | 0.057 | 2.5119E-14 | 57.17 |
| *MEPE* | rs1393691 | G | T | 0.5194 | -0.078 | 0.007 | 2.5119E-28 | 124.16 |
| *MEPE* | rs6547692 | G | A | 0.5568 | 0.051 | 0.007 | 1.2589E-12 | 53.08 |
| *MEPE* | rs77623237 | T | C | 0.1759 | -0.144 | 0.009 | 1.2589E-53 | 256.00 |
| *MEPE* | rs17013212 | C | T | 0.1366 | -0.182 | 0.01 | 1E-69 | 331.24 |
| *MEPE* | rs4861708 | G | A | 0.5115 | -0.064 | 0.007 | 2.5119E-19 | 83.59 |
| *MEPE* | rs55712380 | G | T | 0.2808 | -0.053 | 0.008 | 1.5849E-11 | 43.89 |
| *MEPE* | rs757980 | G | A | 0.7512 | 0.068 | 0.008 | 1.5849E-16 | 72.25 |
| *MEPE* | rs7842942 | T | C | 0.5664 | 0.066 | 0.007 | 2.5119E-20 | 88.90 |
| *MEPE* | rs544626513 | G | C | 0.2289 | 0.066 | 0.009 | 3.1623E-14 | 53.78 |
| *MEPE* | rs3008070 | T | G | 0.2193 | -0.061 | 0.009 | 7.9433E-13 | 45.94 |
| *MEPE* | rs806292 | A | G | 0.532 | -0.058 | 0.007 | 5.0119E-16 | 68.65 |
| *MEPE* | rs2614763 | C | A | 0.4546 | 0.1 | 0.007 | 1.9953E-45 | 204.08 |
| *MEPE* | rs4776350 | C | G | 0.2312 | 0.062 | 0.008 | 1E-13 | 60.06 |
| *MEPE* | rs10521142 | A | G | 0.5579 | 0.073 | 0.007 | 1.9953E-24 | 108.76 |
| *MERTK* | rs59484402 | C | CT | 0.6116 | -0.088 | 0.007 | 1.2589E-33 | 158.04 |
| *MERTK* | rs10188640 | G | A | 0.6257 | -0.266 | 0.007 | 1E-200 | 1444.00 |
| *MERTK* | rs10935473 | G | T | 0.44 | -0.077 | 0.007 | 7.9433E-28 | 121.00 |
| *MERTK* | rs139750425 | T | TCTGC | 0.2573 | -0.055 | 0.008 | 6.3096E-12 | 47.27 |
| *MERTK* | rs35371668 | C | T | 0.1814 | -0.107 | 0.009 | 1.9953E-30 | 141.35 |
| *MERTK* | rs11596680 | A | G | 0.3276 | 0.067 | 0.008 | 6.3096E-19 | 70.14 |
| *MERTK* | rs7088799 | T | G | 0.4139 | 0.072 | 0.007 | 1E-23 | 105.80 |
| *MERTK* | rs174546 | C | T | 0.3446 | 0.073 | 0.007 | 6.3096E-23 | 108.76 |
| *MERTK* | rs494144 | T | C | 0.5374 | -0.082 | 0.007 | 1.9953E-31 | 137.22 |
| *MERTK* | rs9738226 | A | G | 0.6234 | -0.062 | 0.007 | 1.5849E-17 | 78.45 |
| *MERTK* | rs28929474 | C | T | 0.0208 | 0.196 | 0.025 | 1.9953E-15 | 61.47 |
| *MERTK* | rs423135 | G | A | 0.5179 | 0.068 | 0.007 | 3.9811E-22 | 94.37 |
| *MERTK* | rs186021206 | G | A | 0.0064 | 0.61 | 0.048 | 1.9953E-36 | 161.50 |
| *MERTK* | rs12451766 | A | G | 0.5049 | 0.059 | 0.007 | 6.3096E-17 | 71.04 |
| *MERTK* | rs12609794 | G | A | 0.7485 | 0.061 | 0.008 | 5.0119E-14 | 58.14 |
| *MERTK* | rs111981233 | T | G | 0.0796 | 0.091 | 0.013 | 1.5849E-12 | 49.00 |
| *MESD* | rs1354034 | T | C | 0.6041 | 0.069 | 0.008 | 2.5119E-19 | 74.39 |
| *MESD* | rs6993770 | A | T | 0.2864 | -0.088 | 0.008 | 3.1623E-26 | 121.00 |

| | | | | | | | | |
|---|---|---|---|---|---|---|---|---|
| *MET* | rs3768321 | G | T | 0.1937 | -0.063 | 0.009 | 1.9953E-12 | 49.00 |
| *MET* | rs200287431 | T | TGGCGGAT( | 0.175 | 0.068 | 0.009 | 3.1623E-13 | 57.09 |
| *MET* | rs5398 | G | A | 0.2843 | -0.088 | 0.008 | 1.9953E-29 | 121.00 |
| *MET* | rs1858830 | C | G | 0.4426 | -0.16 | 0.008 | 1.5849E-98 | 400.00 |
| *MET* | rs7799998 | A | G | 0.4498 | -0.075 | 0.007 | 3.9811E-25 | 114.80 |
| *MET* | rs507666 | G | A | 0.1846 | -0.331 | 0.009 | 1E-200 | 1352.60 |
| *MET* | rs10740129 | G | A | 0.4696 | 0.068 | 0.007 | 7.9433E-22 | 94.37 |
| *MET* | rs1878036 | T | G | 0.7945 | 0.124 | 0.009 | 6.3096E-46 | 189.83 |
| *MET* | rs174547 | T | C | 0.3441 | 0.104 | 0.007 | 2.5119E-44 | 220.73 |
| *MET* | rs11045819 | C | A | 0.1607 | 0.12 | 0.01 | 1E-35 | 144.00 |
| *MET* | rs7310409 | A | G | 0.6239 | -0.054 | 0.007 | 1.9953E-13 | 59.51 |
| *MET* | rs1041316 | G | A | 0.7292 | 0.082 | 0.008 | 7.9433E-25 | 105.06 |
| *MET* | rs28929474 | C | T | 0.0208 | 0.315 | 0.025 | 6.3096E-37 | 158.76 |
| *MET* | rs1982784 | A | G | 0.8138 | 0.068 | 0.009 | 6.3096E-14 | 57.09 |
| *MET* | rs4850 | G | A | 0.0549 | 0.145 | 0.015 | 7.9433E-21 | 93.44 |
| *MET* | rs186021206 | G | A | 0.0064 | 0.572 | 0.049 | 1.5849E-31 | 136.27 |
| *MET* | rs77542162 | A | G | 0.0229 | 0.387 | 0.024 | 1E-59 | 260.02 |
| *MET* | rs10401969 | T | C | 0.0769 | -0.102 | 0.013 | 1.5849E-14 | 61.56 |
| *MET* | rs112167630 | G | A | 0.0289 | 0.26 | 0.021 | 1.5849E-34 | 153.29 |
| *MET* | rs914187 | T | C | 0.5997 | -0.057 | 0.007 | 3.9811E-15 | 66.31 |
| *METAP1D* | rs138869820 | G | A | 0.0017 | -0.812 | 0.093 | 1.9953E-18 | 76.23 |
| *METAP1D* | rs6993770 | A | T | 0.2869 | -0.065 | 0.008 | 5.0119E-15 | 66.02 |
| *METAP1D* | rs6580980 | A | G | 0.5651 | 0.061 | 0.008 | 1.2589E-15 | 58.14 |
| *METAP2* | rs1354034 | T | C | 0.6041 | 0.096 | 0.008 | 1.9953E-36 | 144.00 |
| *METAP2* | rs6993770 | A | T | 0.2863 | -0.069 | 0.008 | 3.9811E-17 | 74.39 |
| *METAP2* | rs11502185 | T | C | 0.2598 | 0.067 | 0.01 | 2.5119E-12 | 44.89 |
| *METAP2* | rs11170874 | A | G | 0.3311 | 0.059 | 0.008 | 1.9953E-13 | 54.39 |
| *METAP2* | rs10405535 | A | G | 0.685 | 0.096 | 0.008 | 3.9811E-32 | 144.00 |
| *MFAP4* | rs12733767 | C | T | 0.1036 | 0.095 | 0.012 | 5.0119E-15 | 62.67 |
| *MFAP4* | rs7636227 | G | A | 0.5532 | 0.11 | 0.007 | 3.9811E-50 | 246.94 |
| *MFAP4* | rs689122 | T | C | 0.5233 | 0.051 | 0.007 | 3.9811E-12 | 53.08 |
| *MFAP4* | rs36189266 | G | A | 0.6644 | -0.058 | 0.008 | 5.0119E-13 | 52.56 |
| *MFAP4* | rs507666 | G | A | 0.1848 | -0.079 | 0.01 | 7.9433E-17 | 62.41 |
| *MFAP4* | rs139356332 | G | C | 0.0232 | -0.722 | 0.025 | 1.995E-180 | 834.05 |
| *MFAP4* | rs2303729 | G | A | 0.4344 | -0.053 | 0.007 | 1E-12 | 57.33 |
| *MFAP4* | rs738409 | C | G | 0.2166 | 0.064 | 0.009 | 3.9811E-13 | 50.57 |
| *MFAP5* | rs4894803 | A | G | 0.4096 | 0.077 | 0.007 | 1.5849E-25 | 121.00 |
| *MFAP5* | rs1918352 | G | A | 0.552 | 0.051 | 0.007 | 5.0119E-12 | 53.08 |
| *MFAP5* | rs532436 | G | A | 0.1848 | -0.083 | 0.009 | 1E-18 | 85.05 |
| *MFAP5* | rs4132450 | C | T | 0.0045 | -1.77 | 0.06 | 3.981E-193 | 870.25 |
| *MFAP5* | rs1126464 | G | C | 0.2443 | -0.059 | 0.008 | 1.9953E-12 | 54.39 |
| *MFAP5* | rs7412 | C | T | 0.0815 | 0.096 | 0.013 | 5.0119E-13 | 54.53 |
| *MFGE8* | rs768234885 | CA | C | 0.3557 | -0.063 | 0.008 | 1.2589E-16 | 62.02 |
| *MFGE8* | rs1260326 | T | C | 0.6084 | -0.065 | 0.007 | 1E-18 | 86.22 |
| *MFGE8* | rs56210800 | C | G | 0.1308 | -0.078 | 0.011 | 2.5119E-13 | 50.28 |
| *MFGE8* | rs11134475 | A | G | 0.6392 | 0.057 | 0.008 | 5.0119E-14 | 50.77 |
| *MFGE8* | rs521977 | T | G | 0.6769 | 0.064 | 0.008 | 7.9433E-17 | 64.00 |
| *MFGE8* | rs17145750 | C | T | 0.16 | -0.08 | 0.01 | 7.9433E-16 | 64.00 |
| *MFGE8* | rs200646600 | ATTTTT | A | 0.2322 | -0.07 | 0.009 | 3.9811E-15 | 60.49 |
| *MFGE8* | rs28601761 | C | G | 0.4192 | -0.061 | 0.007 | 1.2589E-16 | 75.94 |
| *MFGE8* | rs7232 | T | A | 0.3734 | 0.188 | 0.008 | 2.512E-136 | 552.25 |
| *MFGE8* | rs964184 | G | C | 0.8669 | -0.163 | 0.011 | 2.5119E-52 | 219.58 |
| *MFGE8* | rs2141247 | C | A | 0.6133 | 0.08 | 0.007 | 7.9433E-27 | 130.61 |
| *MFGE8* | rs6602913 | A | C | 0.3656 | 0.063 | 0.007 | 3.9811E-17 | 81.00 |
| *MFGE8* | rs139566989 | C | CGCTGCTC( | 0.784 | -0.079 | 0.009 | 3.1623E-18 | 77.05 |
| *MFGE8* | rs34239095 | C | G | 0.2816 | 0.373 | 0.008 | 1E-200 | 2173.89 |
| *MFGE8* | rs34954997 | C | CTTCG | 0.2375 | 0.06 | 0.009 | 1.9953E-12 | 44.44 |
| *MGLL* | rs7578 | G | A | 0.5572 | 0.06 | 0.008 | 1.2589E-15 | 56.25 |
| *MGLL* | rs6993770 | A | T | 0.2867 | -0.091 | 0.008 | 7.9433E-28 | 129.39 |
| *MGLL* | rs77303550 | C | T | 0.1962 | 0.066 | 0.009 | 3.9811E-12 | 53.78 |
| *MGMT* | rs1354034 | T | C | 0.6043 | 0.055 | 0.007 | 1.5849E-14 | 61.73 |
| *MGMT* | rs342298 | C | T | 0.4546 | -0.055 | 0.007 | 1E-14 | 61.73 |
| *MGMT* | rs1008982 | T | C | 0.3732 | -0.414 | 0.008 | 1E-200 | 2678.06 |
| *MIA* | rs4917 | T | C | 0.6399 | 0.057 | 0.007 | 1E-17 | 66.31 |
| *MIA* | rs28817415 | C | T | 0.4568 | 0.056 | 0.006 | 5.0119E-18 | 87.11 |
| *MIA* | rs60028714 | C | T | 0.6218 | -0.055 | 0.007 | 3.1623E-16 | 61.73 |
| *MIA* | rs3132568 | A | G | 0.2556 | -0.052 | 0.007 | 1.2589E-12 | 55.18 |
| *MIA* | rs77542162 | A | G | 0.0228 | 0.271 | 0.022 | 3.1623E-36 | 151.74 |

| | | | | | | | | |
|---|---|---|---|---|---|---|---|---|
| *MIA* | rs2607421 | A | G | 0.073 | 1.383 | 0.014 | 1E-200 | 9758.62 |
| *MICALL2* | rs2289035 | G | A | 0.3978 | 0.069 | 0.008 | 5.0119E-19 | 74.39 |
| *MICB_MICA* | rs2009581 | G | A | 0.2725 | -0.03 | 0.004 | 1E-12 | 56.25 |
| *MICB_MICA* | rs10935473 | G | T | 0.4399 | -0.1 | 0.004 | 3.981E-157 | 625.00 |
| *MICB_MICA* | rs3132467 | G | A | 0.2568 | -1.102 | 0.009 | 1E-200 | 14992.64 |
| *MICB_MICA* | rs2682471 | T | G | 0.427 | -0.03 | 0.004 | 5.0119E-15 | 56.25 |
| *MICB_MICA* | rs3184504 | T | C | 0.5174 | -0.026 | 0.004 | 2.5119E-12 | 42.25 |
| *MICB_MICA* | rs35715239 | G | C | 0.3336 | -0.055 | 0.004 | 2.5119E-44 | 189.06 |
| *MICB_MICA* | rs186021206 | G | A | 0.0064 | 0.322 | 0.026 | 1.9953E-36 | 153.38 |
| *MICB_MICA* | rs4760 | A | G | 0.1562 | 0.037 | 0.005 | 3.1623E-13 | 54.76 |
| *MIF* | rs1354034 | T | C | 0.604 | 0.066 | 0.008 | 3.9811E-18 | 68.06 |
| *MIF* | rs60822569 | T | TTTCTC | 0.5545 | 0.053 | 0.008 | 1.5849E-12 | 43.89 |
| *MIF* | rs2330634 | C | G | 0.5962 | -0.11 | 0.008 | 1.9953E-46 | 189.06 |
| *MILR1* | rs140655352 | A | AT | 0.0854 | 0.2 | 0.013 | 1.5849E-56 | 236.69 |
| *MILR1* | rs2524119 | T | C | 0.5449 | 0.059 | 0.007 | 3.9811E-17 | 71.04 |
| *MILR1* | rs4055121 | C | T | 0.1344 | -0.1 | 0.01 | 1.9953E-22 | 100.00 |
| *MILR1* | rs904628 | G | A | 0.4762 | -0.055 | 0.007 | 6.3096E-15 | 61.73 |
| *MILR1* | rs186021206 | G | A | 0.0064 | 1.25 | 0.052 | 6.31E-127 | 577.85 |
| *MILR1* | rs704 | G | A | 0.4736 | -0.059 | 0.008 | 6.3096E-15 | 54.39 |
| *MILR1* | rs138176943 | G | A | 0.0578 | 0.796 | 0.016 | 1E-200 | 2475.06 |
| *MINDY1* | rs140386498 | A | T | 0.0117 | -0.628 | 0.035 | 1.5849E-73 | 321.95 |
| *MINDY1* | rs6993770 | A | T | 0.2864 | -0.064 | 0.008 | 5.0119E-15 | 64.00 |
| *MINDY1* | rs4759076 | T | C | 0.4567 | 0.059 | 0.008 | 2.5119E-15 | 54.39 |
| *MINK1* | rs342298 | C | T | 0.4553 | -0.059 | 0.008 | 3.9811E-15 | 54.39 |
| *MINK1* | rs6993770 | A | T | 0.2865 | -0.063 | 0.008 | 3.1623E-14 | 62.02 |
| *MINK1* | rs190385998 | T | A | 0.003 | -0.644 | 0.075 | 7.9433E-18 | 73.73 |
| *MINK1* | rs200309755 | C | T | 0.5369 | 0.054 | 0.008 | 3.1623E-12 | 45.56 |
| *MITD1* | rs13009147 | T | G | 0.6524 | -0.075 | 0.008 | 1.5849E-21 | 87.89 |
| *MITD1* | rs1354034 | T | C | 0.6041 | 0.056 | 0.008 | 2.5119E-13 | 49.00 |
| *MLN* | rs870465 | G | A | 0.1745 | 0.063 | 0.009 | 6.3096E-12 | 49.00 |
| *MLN* | rs826404 | T | C | 0.5865 | -0.061 | 0.007 | 7.9433E-20 | 75.94 |
| *MLN* | rs11240492 | G | C | 0.8661 | 0.1 | 0.01 | 2.5119E-24 | 100.00 |
| *MLN* | rs16862260 | G | A | 0.2083 | -0.226 | 0.008 | 1.259E-158 | 798.06 |
| *MLN* | rs147351977 | G | A | 0.0284 | 0.139 | 0.02 | 3.1623E-12 | 48.30 |
| *MLN* | rs12521989 | A | T | 0.3483 | -0.066 | 0.007 | 1.5849E-21 | 88.90 |
| *MLN* | rs56092644 | G | T | 0.2443 | 0.079 | 0.008 | 7.9433E-25 | 97.52 |
| *MLN* | rs67131976 | C | T | 0.1725 | 0.172 | 0.009 | 6.3096E-77 | 365.23 |
| *MLN* | rs1547671 | C | T | 0.5443 | -0.414 | 0.007 | 1E-200 | 3497.88 |
| *MLN* | rs13217983 | C | T | 0.1382 | 0.075 | 0.01 | 2.5119E-13 | 56.25 |
| *MLN* | rs776997164 | TAGAC | T | 0.1838 | 0.105 | 0.009 | 1.5849E-30 | 136.11 |
| *MLN* | rs12114906 | T | C | 0.2514 | -0.063 | 0.008 | 5.0119E-16 | 62.02 |
| *MLN* | rs3762055 | T | C | 0.0684 | 0.175 | 0.014 | 3.9811E-36 | 156.25 |
| *MLN* | rs782134971 | GAAACTGC( | G | 0.2542 | -0.073 | 0.008 | 7.9433E-22 | 83.27 |
| *MLN* | rs4309096 | T | C | 0.6459 | 0.065 | 0.007 | 1E-20 | 86.22 |
| *MLN* | rs5785928 | C | CT | 0.6923 | -0.058 | 0.007 | 1E-15 | 68.65 |
| *MLN* | rs11213809 | A | G | 0.6562 | -0.051 | 0.007 | 2.5119E-13 | 53.08 |
| *MLN* | rs2485287 | A | C | 0.4647 | 0.076 | 0.007 | 6.3096E-31 | 117.88 |
| *MLN* | rs28837202 | T | C | 0.4373 | 0.061 | 0.007 | 3.1623E-20 | 75.94 |
| *MLN* | rs566487891 | G | C | 0.7926 | 0.061 | 0.008 | 1.2589E-13 | 58.14 |
| *MLN* | rs17681738 | C | T | 0.3294 | 0.053 | 0.007 | 6.3096E-14 | 57.33 |
| *MLN* | rs74175746 | T | TTA | 0.176 | 0.069 | 0.009 | 1.9953E-15 | 58.78 |
| *MLN* | rs633372 | G | A | 0.5345 | -0.072 | 0.007 | 2.5119E-27 | 105.80 |
| *MLN* | rs2208150 | T | C | 0.5905 | 0.084 | 0.007 | 2.5119E-35 | 144.00 |
| *MME* | rs1497406 | A | G | 0.58 | 0.076 | 0.007 | 1E-28 | 117.88 |
| *MME* | rs4687657 | G | T | 0.2598 | 0.581 | 0.008 | 1E-200 | 5274.39 |
| *MME* | rs79837905 | A | G | 0.0799 | -0.107 | 0.013 | 1.2589E-15 | 67.75 |
| *MME* | rs4835265 | C | A | 0.156 | 0.118 | 0.009 | 2.5119E-36 | 171.90 |
| *MME* | rs112875651 | G | A | 0.3909 | -0.049 | 0.007 | 2.5119E-12 | 49.00 |
| *MME* | rs151068477 | G | T | 0.0083 | 0.275 | 0.04 | 3.9811E-12 | 47.27 |
| *MME* | rs1650146 | A | G | 0.6175 | 0.049 | 0.007 | 2.5119E-12 | 49.00 |
| *MME* | rs17674290 | T | G | 0.1475 | 0.065 | 0.01 | 1.2589E-11 | 42.25 |
| *MME* | rs61007561 | A | AG | 0.2456 | 0.11 | 0.008 | 3.9811E-44 | 189.06 |
| *MME* | rs12373325 | T | C | 0.7982 | -0.096 | 0.008 | 1.2589E-29 | 144.00 |
| *MME* | rs58659609 | C | A | 0.1852 | 0.074 | 0.009 | 3.9811E-17 | 67.60 |
| *MME* | rs189448562 | G | C | 0.0111 | 0.275 | 0.033 | 6.3096E-17 | 69.44 |
| *MME* | rs12157484 | C | T | 0.2429 | -0.056 | 0.008 | 1.2589E-12 | 49.00 |
| *MMP1* | rs12136856 | C | G | 0.6531 | 0.092 | 0.007 | 2.5119E-37 | 172.73 |
| *MMP1* | rs12622659 | G | A | 0.301 | -0.059 | 0.007 | 1.9953E-15 | 71.04 |

| | | | | | | | | |
|---|---|---|---|---|---|---|---|---|
| *MMP1* | rs76036957 | G | A | 0.0095 | 0.316 | 0.036 | 5.0119E-19 | 77.05 |
| *MMP1* | rs28706788 | G | A | 0.5415 | -0.055 | 0.007 | 1.2589E-15 | 61.73 |
| *MMP1* | rs5879021 | G | GC | 0.5202 | 0.071 | 0.007 | 6.3096E-25 | 102.88 |
| *MMP1* | rs6993770 | A | T | 0.2867 | -0.157 | 0.008 | 1E-95 | 385.14 |
| *MMP1* | rs10820606 | A | C | 0.2293 | 0.08 | 0.008 | 3.1623E-22 | 100.00 |
| *MMP1* | rs534191 | C | T | 0.4539 | 0.448 | 0.008 | 1E-200 | 3136.00 |
| *MMP1* | rs60822569 | T | TTTCTC | 0.5535 | 0.053 | 0.007 | 1.5849E-14 | 57.33 |
| *MMP1* | rs757848155 | CAAA | C | 0.6392 | 0.052 | 0.007 | 3.1623E-13 | 55.18 |
| *MMP1* | rs12445050 | C | T | 0.1384 | 0.073 | 0.01 | 1.2589E-13 | 53.29 |
| *MMP1* | rs1654425 | T | C | 0.8341 | 0.12 | 0.009 | 1E-38 | 177.78 |
| *MMP1* | rs6081569 | G | C | 0.3686 | 0.058 | 0.007 | 1.9953E-16 | 68.65 |
| *MMP10* | rs187265184 | C | G | 0.3563 | 0.064 | 0.009 | 1.9953E-13 | 50.57 |
| *MMP10* | rs486055 | C | T | 0.1622 | -0.396 | 0.01 | 1E-200 | 1568.16 |
| *MMP10* | rs681343 | C | T | 0.5095 | 0.137 | 0.007 | 1.2589E-81 | 383.04 |
| *MMP12* | rs2770 | G | A | 0.4475 | -0.055 | 0.007 | 7.9433E-14 | 61.73 |
| *MMP12* | rs17368814 | A | G | 0.1253 | -0.749 | 0.011 | 1E-200 | 4636.37 |
| *MMP12* | rs61729512 | G | A | 0.118 | 0.1 | 0.01 | 1.9953E-23 | 100.00 |
| *MMP12* | rs7310615 | C | G | 0.5168 | -0.045 | 0.006 | 3.9811E-12 | 56.25 |
| *MMP12* | rs12986064 | T | C | 0.5076 | -0.08 | 0.007 | 1.9953E-33 | 130.61 |
| *MMP15* | rs187793617 | C | G | 0.1604 | 0.123 | 0.011 | 1.2589E-29 | 125.03 |
| *MMP3* | rs41341748 | G | A | 0.011 | 0.245 | 0.025 | 3.1623E-22 | 96.04 |
| *MMP3* | rs678815 | G | C | 0.483 | 0.479 | 0.006 | 1E-200 | 6373.36 |
| *MMP3* | rs2267372 | A | G | 0.5998 | 0.05 | 0.005 | 7.9433E-21 | 100.00 |
| *MMP7* | rs6694034 | A | C | 0.4488 | 0.109 | 0.007 | 2.5119E-51 | 242.47 |
| *MMP7* | rs11568819 | G | A | 0.061 | 0.567 | 0.015 | 1E-200 | 1428.84 |
| *MMP7* | rs759947295 | CTGTT | C | 0.78 | -0.064 | 0.009 | 3.9811E-13 | 50.57 |
| *MMP7* | rs77924615 | G | A | 0.1967 | -0.092 | 0.009 | 1.5849E-24 | 104.49 |
| *MMP7* | rs12975366 | T | C | 0.3952 | -0.063 | 0.007 | 5.0119E-18 | 81.00 |
| *MMP8* | rs2511241 | C | T | 0.9294 | 0.189 | 0.015 | 1.5849E-38 | 158.76 |
| *MMP8* | rs35231465 | G | A | 0.0248 | -1.252 | 0.024 | 1E-200 | 2721.36 |
| *MMP8* | rs7208815 | C | T | 0.3929 | 0.087 | 0.007 | 1.2589E-33 | 154.47 |
| *MMP9* | rs1886654 | T | C | 0.8944 | -0.081 | 0.012 | 1.2589E-11 | 45.56 |
| *MMP9* | rs28498283 | A | T | 0.2547 | 0.059 | 0.009 | 6.3096E-12 | 42.98 |
| *MMP9* | rs4619875 | C | T | 0.4034 | -0.059 | 0.007 | 3.9811E-15 | 71.04 |
| *MMP9* | rs9267111 | A | G | 0.3485 | 0.07 | 0.008 | 1.5849E-19 | 76.56 |
| *MMP9* | rs2158799 | C | G | 0.609 | 0.054 | 0.008 | 1.5849E-12 | 45.56 |
| *MMP9* | rs2511241 | C | T | 0.9295 | 0.384 | 0.014 | 5.012E-155 | 752.33 |
| *MMP9* | rs2289511 | G | A | 0.581 | -0.059 | 0.007 | 3.9811E-15 | 71.04 |
| *MMP9* | rs749834305 | GA | G | 0.3927 | 0.083 | 0.007 | 1.5849E-28 | 140.59 |
| *MMP9* | rs17576 | A | G | 0.3514 | 0.145 | 0.008 | 5.0119E-79 | 328.52 |
| *MMUT* | rs775536954 | CTCGGCGA | C | 0.3722 | 0.103 | 0.008 | 5.0119E-37 | 165.77 |
| *MOCS2* | rs1354034 | T | C | 0.6044 | 0.084 | 0.007 | 1.9953E-32 | 144.00 |
| *MOCS2* | rs6880055 | A | G | 0.3422 | 0.449 | 0.008 | 1E-200 | 3150.02 |
| *MOCS2* | rs6967348 | G | A | 0.9118 | -0.092 | 0.012 | 6.3096E-14 | 58.78 |
| *MOCS2* | rs60822569 | T | TTTCTC | 0.5542 | 0.049 | 0.007 | 2.5119E-12 | 49.00 |
| *MOG* | rs9461541 | A | G | 0.0488 | -0.451 | 0.017 | 1.585E-160 | 703.81 |
| *MOG* | rs8021280 | C | G | 0.463 | 0.054 | 0.007 | 1.2589E-14 | 59.51 |
| *MORF4L2* | rs71534539 | G | A | 0.464 | -0.076 | 0.009 | 7.9433E-19 | 71.31 |
| *MPHOSPH8* | rs1354034 | T | C | 0.6038 | 0.068 | 0.008 | 2.5119E-18 | 72.25 |
| *MPHOSPH8* | rs35424555 | A | G | 0.1224 | 0.088 | 0.012 | 5.0119E-14 | 53.78 |
| *MPI* | rs1354034 | T | C | 0.6046 | 0.068 | 0.008 | 3.9811E-19 | 72.25 |
| *MPI* | rs7080386 | C | A | 0.4117 | 0.06 | 0.008 | 2.5119E-15 | 56.25 |
| *MPI* | rs4886632 | G | T | 0.5763 | 0.2 | 0.008 | 5.012E-152 | 625.00 |
| *MPIG6B* | rs6961069 | C | T | 0.4024 | 0.056 | 0.008 | 3.9811E-13 | 49.00 |
| *MPIG6B* | rs6993770 | A | T | 0.2867 | -0.101 | 0.008 | 1.2589E-33 | 159.39 |
| *MPIG6B* | rs7896518 | A | G | 0.4241 | 0.077 | 0.008 | 3.1623E-23 | 92.64 |
| *MPIG6B* | rs1362213 | C | T | 0.4795 | -0.054 | 0.008 | 2.5119E-12 | 45.56 |
| *MPIG6B* | rs892090 | T | G | 0.8342 | 0.102 | 0.01 | 1E-23 | 104.04 |
| *MPO* | rs2120782 | G | C | 0.2053 | 0.086 | 0.009 | 2.5119E-21 | 91.31 |
| *MPO* | rs13277237 | G | A | 0.5676 | -0.075 | 0.007 | 7.9433E-24 | 114.80 |
| *MPO* | rs757081 | C | G | 0.3322 | 0.055 | 0.008 | 1.9953E-12 | 47.27 |
| *MPO* | rs12941811 | T | C | 0.5745 | -0.056 | 0.008 | 1.5849E-13 | 49.00 |
| *MPO* | rs34097845 | C | T | 0.0574 | -0.507 | 0.017 | 1E-200 | 889.44 |
| *MPO* | rs6034875 | G | A | 0.5804 | -0.068 | 0.007 | 3.1623E-20 | 94.37 |
| *MPRIP* | rs3910516 | A | G | 0.7364 | 0.071 | 0.009 | 1.5849E-16 | 62.23 |
| *MRC1* | rs11125916 | C | A | 0.1948 | 0.062 | 0.009 | 1.9953E-12 | 47.46 |
| *MRC1* | rs2579651 | A | G | 0.8716 | 0.074 | 0.011 | 3.1623E-12 | 45.26 |
| *MRC1* | rs1257169 | C | A | 0.499 | -0.059 | 0.007 | 3.1623E-17 | 71.04 |

| | | | | | | | | |
|---|---|---|---|---|---|---|---|---|
| *MRC1* | rs35771982 | G | C | 0.5083 | 0.133 | 0.007 | 6.3096E-81 | 361.00 |
| *MRC1* | rs12515263 | G | A | 0.0429 | 0.23 | 0.017 | 1E-40 | 183.04 |
| *MRC1* | rs35139284 | C | T | 0.3213 | 0.083 | 0.008 | 3.1623E-28 | 107.64 |
| *MRC1* | rs12667978 | C | T | 0.5709 | 0.052 | 0.007 | 1.9953E-13 | 55.18 |
| *MRC1* | rs4876611 | A | G | 0.7181 | 0.069 | 0.008 | 1.2589E-18 | 74.39 |
| *MRC1* | rs74118031 | T | C | 0.2336 | -0.285 | 0.008 | 1E-200 | 1269.14 |
| *MRC1* | rs5030924 | A | T | 0.4446 | -0.072 | 0.007 | 2.5119E-23 | 105.80 |
| *MRC1* | rs174564 | A | G | 0.3489 | 0.077 | 0.007 | 5.0119E-26 | 121.00 |
| *MRC1* | rs186021206 | G | A | 0.0064 | 0.582 | 0.048 | 7.9433E-34 | 147.02 |
| *MRC1* | rs7246479 | T | G | 0.5098 | 0.054 | 0.007 | 6.3096E-15 | 59.51 |
| *MRC1* | rs4611747 | C | T | 0.2885 | 0.072 | 0.008 | 1.5849E-20 | 81.00 |
| *MRI1* | rs1354034 | T | C | 0.604 | 0.055 | 0.007 | 5.0119E-14 | 61.73 |
| *MRI1* | rs452960 | A | T | 0.4218 | -0.347 | 0.008 | 1E-200 | 1881.39 |
| *MRPL58* | rs1354034 | T | C | 0.6043 | 0.057 | 0.008 | 1.5849E-13 | 50.77 |
| *MRPL58* | rs342293 | C | G | 0.4596 | -0.058 | 0.008 | 2.5119E-14 | 52.56 |
| *MRPL58* | rs6993770 | A | T | 0.2864 | -0.066 | 0.008 | 3.1623E-15 | 68.06 |
| *MRPL58* | rs11085147 | C | T | 0.0991 | 0.174 | 0.013 | 2.5119E-40 | 179.15 |
| *MSLN* | rs72851462 | G | C | 0.1154 | -0.101 | 0.011 | 3.9811E-21 | 84.31 |
| *MSLN* | rs2517403 | T | C | 0.3319 | -0.068 | 0.007 | 6.3096E-21 | 94.37 |
| *MSLN* | rs1214759 | A | G | 0.6814 | -0.093 | 0.007 | 6.3096E-38 | 176.51 |
| *MSLN* | rs11788059 | T | C | 0.3395 | 0.071 | 0.007 | 3.9811E-23 | 102.88 |
| *MSLN* | rs2235503 | C | A | 0.193 | 0.754 | 0.009 | 1E-200 | 7018.72 |
| *MSLN* | rs4807630 | C | T | 0.309 | 0.056 | 0.007 | 3.9811E-14 | 64.00 |
| *MSLN* | rs34450188 | A | G | 0.5573 | 0.081 | 0.007 | 6.3096E-32 | 133.90 |
| *MSLN* | rs492602 | A | G | 0.5089 | 0.068 | 0.007 | 6.3096E-23 | 94.37 |
| *MSMB* | rs4687657 | G | T | 0.259 | 0.089 | 0.006 | 1E-52 | 220.03 |
| *MSMB* | rs62406524 | G | C | 0.0962 | -0.11 | 0.009 | 5.0119E-34 | 149.38 |
| *MSMB* | rs73671501 | C | T | 0.1703 | 0.079 | 0.007 | 6.3096E-32 | 127.37 |
| *MSMB* | rs10993994 | T | C | 0.6083 | 0.936 | 0.007 | 1E-200 | 17879.51 |
| *MSMB* | rs36120341 | C | T | 0.3638 | -0.049 | 0.005 | 2.5119E-20 | 96.04 |
| *MSR1* | rs41271951 | A | G | 0.085 | -0.101 | 0.012 | 3.9811E-17 | 70.84 |
| *MSR1* | rs547373038 | G | T | 0.494 | -0.076 | 0.007 | 7.9433E-28 | 117.88 |
| *MSR1* | rs2293330 | T | G | 0.8267 | 0.068 | 0.009 | 3.1623E-14 | 57.09 |
| *MSR1* | rs1471401 | A | G | 0.661 | 0.05 | 0.007 | 1.2589E-12 | 51.02 |
| *MSR1* | rs115216147 | T | A | 0.0666 | 0.094 | 0.014 | 7.9433E-12 | 45.08 |
| *MSR1* | rs509227 | A | C | 0.5227 | 0.054 | 0.007 | 1.2589E-15 | 59.51 |
| *MSR1* | rs139299944 | C | CT | 0.3436 | -0.097 | 0.007 | 5.0119E-43 | 192.02 |
| *MSR1* | rs10863 | A | G | 0.853 | 0.071 | 0.009 | 6.3096E-14 | 62.23 |
| *MSR1* | rs41341748 | G | A | 0.0111 | -1.547 | 0.033 | 1E-200 | 2197.62 |
| *MSR1* | rs2031902 | C | T | 0.5875 | -0.058 | 0.007 | 3.1623E-17 | 68.65 |
| *MSR1* | rs635634 | C | T | 0.1845 | 0.075 | 0.009 | 6.3096E-18 | 69.44 |
| *MSR1* | rs11220505 | C | T | 0.2229 | 0.143 | 0.008 | 1.9953E-69 | 319.52 |
| *MSR1* | rs186021206 | G | A | 0.0064 | 1.383 | 0.046 | 1E-198 | 903.92 |
| *MSRA* | rs1354034 | T | C | 0.6041 | 0.06 | 0.008 | 3.9811E-15 | 56.25 |
| *MSRA* | rs12546887 | C | T | 0.5902 | -0.057 | 0.008 | 6.3096E-14 | 50.77 |
| *MSRA* | rs139130389 | C | CTA | 0.0726 | -0.153 | 0.014 | 1.9953E-26 | 119.43 |
| *MST1* | rs9435340 | T | A | 0.6553 | -0.049 | 0.005 | 1.9953E-25 | 96.04 |
| *MST1* | rs1260326 | T | C | 0.6083 | -0.068 | 0.005 | 3.9811E-51 | 184.96 |
| *MST1* | rs11130213 | C | T | 0.2912 | -1.051 | 0.008 | 1E-200 | 17259.39 |
| *MST1* | rs1799895 | C | G | 0.0119 | -0.146 | 0.021 | 1.5849E-12 | 48.34 |
| *MST1* | rs8111 | C | T | 0.2538 | 0.056 | 0.005 | 1.2589E-28 | 125.44 |
| *MST1* | rs28929474 | C | T | 0.0209 | 0.175 | 0.015 | 1E-29 | 136.11 |
| *MST1* | rs149394327 | G | C | 0.03 | 0.18 | 0.013 | 7.9433E-44 | 191.72 |
| *MSTN* | rs143242500 | A | G | 0.002 | 0.786 | 0.087 | 1.2589E-19 | 81.62 |
| *MSTN* | rs34950020 | G | A | 0.3187 | -0.079 | 0.007 | 3.9811E-26 | 127.37 |
| *MSTN* | rs1815739 | T | C | 0.5574 | 0.061 | 0.007 | 1.9953E-18 | 75.94 |
| *MSTN* | rs8096658 | C | G | 0.4886 | 0.054 | 0.007 | 1.5849E-14 | 59.51 |
| *MTDH* | rs1354034 | T | C | 0.6039 | 0.147 | 0.008 | 7.9433E-80 | 337.64 |
| *MTDH* | rs342293 | C | G | 0.4593 | -0.056 | 0.008 | 1.5849E-13 | 49.00 |
| *MTDH* | rs6993770 | A | T | 0.2866 | -0.069 | 0.008 | 3.1623E-16 | 74.39 |
| *MTDH* | rs11502185 | T | C | 0.2597 | 0.088 | 0.01 | 2.5119E-19 | 77.44 |
| *MTDH* | rs60822569 | T | TTTCTC | 0.5541 | 0.053 | 0.008 | 6.3096E-12 | 43.89 |
| *MTHFD2* | rs183347927 | G | A | 0.0026 | 1.945 | 0.079 | 3.981E-133 | 606.16 |
| *MTHFSD* | rs1354034 | T | C | 0.6041 | 0.06 | 0.007 | 7.9433E-17 | 73.47 |
| *MTHFSD* | rs10900809 | G | A | 0.3887 | -0.05 | 0.007 | 1.2589E-11 | 51.02 |
| *MTHFSD* | rs9939348 | T | C | 0.5978 | 0.464 | 0.008 | 1E-200 | 3364.00 |
| *MTIF3* | rs10075801 | A | G | 0.4879 | -0.058 | 0.008 | 3.9811E-14 | 52.56 |
| *MTIF3* | rs6993770 | A | T | 0.2866 | -0.064 | 0.008 | 1.2589E-14 | 64.00 |

| | | | | | | | | |
|---|---|---|---|---|---|---|---|---|
| *MTIF3* | rs11170874 | A | G | 0.3311 | 0.06 | 0.008 | 2.5119E-13 | 56.25 |
| *MTIF3* | rs1218825 | G | C | 0.9561 | 0.218 | 0.019 | 7.9433E-32 | 131.65 |
| *MTSS2* | rs1354034 | T | C | 0.6044 | 0.057 | 0.008 | 6.3096E-14 | 50.77 |
| *MTSS2* | rs6993770 | A | T | 0.2864 | 0.064 | 0.008 | 1.2589E-14 | 64.00 |
| *MTSS2* | rs113315674 | G | A | 0.3649 | 0.076 | 0.008 | 7.9433E-23 | 90.25 |
| *MTSS2* | rs7297265 | A | G | 0.4424 | 0.074 | 0.008 | 2.5119E-22 | 85.56 |
| *MTSS2* | rs2487972 | T | A | 0.0124 | -0.348 | 0.036 | 1.2589E-21 | 93.44 |
| *MTUS1* | rs142323734 | G | A | 0.0406 | 0.432 | 0.019 | 3.162E-111 | 516.96 |
| *MUC13* | rs1127233 | T | G | 0.262 | 0.244 | 0.009 | 1E-177 | 735.01 |
| *MUC13* | rs4556017 | C | T | 0.8533 | -0.104 | 0.011 | 1E-22 | 89.39 |
| *MUC13* | rs2519093 | C | T | 0.1838 | -0.252 | 0.01 | 1.259E-147 | 635.04 |
| *MUC16* | rs62193070 | A | G | 0.1628 | -0.259 | 0.01 | 1.585E-135 | 670.81 |
| *MUC16* | rs7205754 | C | A | 0.0395 | -0.201 | 0.02 | 1E-24 | 101.00 |
| *MUC16* | rs10421318 | G | A | 0.3508 | -0.072 | 0.008 | 1.5849E-18 | 81.00 |
| *MUC2* | rs1933630 | C | A | 0.157 | 0.07 | 0.008 | 1E-16 | 76.56 |
| *MUC2* | rs505922 | T | C | 0.317 | 0.557 | 0.007 | 1E-200 | 6331.61 |
| *MUC2* | rs12416873 | G | A | 0.2078 | -0.265 | 0.008 | 1E-200 | 1097.27 |
| *MUC2* | rs10899345 | A | G | 0.3366 | -0.082 | 0.007 | 3.9811E-35 | 137.22 |
| *MUC2* | rs11215025 | C | T | 0.1573 | -0.065 | 0.009 | 2.5119E-14 | 52.16 |
| *MUC2* | rs11837407 | A | G | 0.5646 | 0.044 | 0.006 | 1E-12 | 53.78 |
| *MUC2* | rs2608894 | T | C | 0.7649 | -0.153 | 0.008 | 1E-76 | 365.77 |
| *MUC2* | rs681343 | C | T | 0.5093 | 0.573 | 0.007 | 1E-200 | 6700.59 |
| *MVK* | rs28934897 | G | A | 0.002 | -1.007 | 0.094 | 6.3096E-27 | 114.76 |
| *MXRA8* | rs200138692 | G | C | 0.0011 | -1.128 | 0.106 | 1.5849E-26 | 113.24 |
| *MXRA8* | rs1260326 | T | C | 0.6081 | 0.055 | 0.007 | 3.1623E-14 | 61.73 |
| *MXRA8* | rs368548301 | T | TTAA | 0.6183 | -0.067 | 0.007 | 1.2589E-19 | 91.61 |
| *MXRA8* | rs13167730 | G | T | 0.0955 | 0.093 | 0.012 | 6.3096E-15 | 60.06 |
| *MXRA8* | rs331 | G | A | 0.2642 | 0.056 | 0.008 | 1.9953E-12 | 49.00 |
| *MXRA8* | rs56278466 | T | G | 0.661 | 0.077 | 0.007 | 2.5119E-25 | 121.00 |
| *MXRA8* | rs200839648 | T | A | 0.868 | -0.113 | 0.011 | 2.5119E-23 | 105.53 |
| *MXRA8* | rs261290 | T | C | 0.6543 | -0.103 | 0.007 | 7.9433E-43 | 216.51 |
| *MXRA8* | rs247617 | C | A | 0.324 | 0.099 | 0.008 | 3.9811E-39 | 153.14 |
| *MXRA8* | rs58761784 | T | C | 0.2849 | -0.058 | 0.008 | 1.9953E-13 | 52.56 |
| *MXRA8* | rs1065853 | G | T | 0.0809 | 0.16 | 0.013 | 1E-34 | 151.48 |
| *MYBPC1* | rs6431221 | C | A | 0.5146 | 0.05 | 0.007 | 5.0119E-13 | 51.02 |
| *MYBPC1* | rs12463674 | A | G | 0.3157 | 0.075 | 0.008 | 3.9811E-23 | 87.89 |
| *MYBPC1* | rs2234962 | T | C | 0.216 | 0.065 | 0.008 | 1.9953E-14 | 66.02 |
| *MYBPC1* | rs7481951 | A | T | 0.5876 | 0.107 | 0.007 | 7.9433E-51 | 233.65 |
| *MYBPC1* | rs1034048 | A | G | 0.5593 | 0.065 | 0.007 | 5.0119E-20 | 86.22 |
| *MYBPC1* | rs1801690 | C | G | 0.0565 | 0.131 | 0.015 | 5.0119E-18 | 76.27 |
| *MYCBP2* | rs1354034 | T | C | 0.6043 | 0.063 | 0.008 | 7.9433E-17 | 62.02 |
| *MYDGF* | rs1354034 | T | C | 0.6045 | 0.063 | 0.008 | 2.5119E-16 | 62.02 |
| *MYDGF* | rs6993770 | A | T | 0.2866 | -0.074 | 0.008 | 7.9433E-19 | 85.56 |
| *MYDGF* | rs60893137 | A | C | 0.0213 | -0.307 | 0.026 | 5.0119E-32 | 139.42 |
| *MYH9* | rs1354034 | T | C | 0.6046 | 0.075 | 0.008 | 7.9433E-23 | 87.89 |
| *MYH9* | rs10075801 | A | G | 0.4878 | -0.058 | 0.008 | 2.5119E-14 | 52.56 |
| *MYH9* | rs342293 | C | G | 0.4597 | -0.056 | 0.008 | 1.2589E-13 | 49.00 |
| *MYH9* | rs6993770 | A | T | 0.2863 | -0.075 | 0.008 | 1E-19 | 87.89 |
| *MYH9* | rs7210399 | T | C | 0.5151 | -0.058 | 0.008 | 1.5849E-14 | 52.56 |
| *MYL3* | rs6431221 | C | A | 0.5153 | 0.063 | 0.007 | 2.5119E-19 | 81.00 |
| *MYL3* | rs12463674 | A | G | 0.3155 | 0.084 | 0.008 | 1.5849E-27 | 110.25 |
| *MYL3* | rs200564310 | T | G | 0.157 | -0.071 | 0.01 | 3.9811E-13 | 50.41 |
| *MYL3* | rs73015965 | A | G | 0.0065 | 0.34 | 0.048 | 7.9433E-13 | 50.17 |
| *MYL3* | rs2234962 | T | C | 0.2163 | 0.062 | 0.009 | 5.0119E-13 | 47.46 |
| *MYL3* | rs7481951 | A | T | 0.5874 | 0.123 | 0.007 | 3.9811E-64 | 308.76 |
| *MYL3* | rs1034048 | A | G | 0.5593 | 0.081 | 0.007 | 1.5849E-29 | 133.90 |
| *MYL3* | rs76452927 | G | A | 0.0536 | 0.121 | 0.016 | 1.5849E-14 | 57.19 |
| *MYL4* | rs111803166 | A | G | 0.009 | 0.331 | 0.039 | 1.9953E-17 | 72.03 |
| *MYL6B* | rs71534539 | G | A | 0.4646 | -0.075 | 0.009 | 1E-18 | 69.44 |
| *MYLPF* | rs10833712 | T | G | 0.5786 | 0.054 | 0.008 | 6.3096E-13 | 45.56 |
| *MYOC* | rs171003 | G | A | 0.5803 | 0.267 | 0.007 | 1E-200 | 1454.88 |
| *MYOC* | rs6888037 | G | T | 0.7508 | 0.084 | 0.008 | 1.9953E-25 | 110.25 |
| *MYOC* | rs56278466 | T | G | 0.6612 | 0.102 | 0.007 | 7.9433E-44 | 212.33 |
| *MYOC* | rs215225 | A | G | 0.4278 | 0.052 | 0.007 | 1.2589E-13 | 55.18 |
| *MYOC* | rs7310615 | C | G | 0.5178 | 0.058 | 0.007 | 6.3096E-17 | 68.65 |
| *MYOC* | rs11632195 | C | T | 0.6388 | -0.087 | 0.007 | 6.3096E-33 | 154.47 |
| *MYOC* | rs186021206 | G | A | 0.0064 | 0.511 | 0.048 | 7.9433E-27 | 113.33 |
| *MYOC* | rs61739916 | T | C | 0.055 | 0.14 | 0.015 | 5.0119E-20 | 87.11 |

| | | | | | | | | |
|---|---|---|---|---|---|---|---|---|
| *MYOM2* | rs117165873 | A | G | 0.0045 | 1.071 | 0.056 | 1.9953E-80 | 365.77 |
| *MYOM2* | rs10833711 | G | A | 0.5283 | 0.065 | 0.008 | 1.9953E-17 | 66.02 |
| *MYOM3* | rs6700245 | T | C | 0.1467 | -0.107 | 0.01 | 1.5849E-26 | 114.49 |
| *MYOM3* | rs6431221 | C | A | 0.5147 | 0.064 | 0.007 | 1.2589E-19 | 83.59 |
| *MYOM3* | rs12464787 | G | A | 0.3109 | 0.082 | 0.008 | 3.9811E-26 | 105.06 |
| *MYOM3* | rs663494 | A | T | 0.6674 | 0.057 | 0.008 | 1.9953E-12 | 50.77 |
| *MYOM3* | rs72842207 | C | T | 0.2154 | 0.078 | 0.009 | 1.2589E-19 | 75.11 |
| *MYOM3* | rs7481951 | A | T | 0.5876 | 0.127 | 0.007 | 7.9433E-69 | 329.16 |
| *MYOM3* | rs370844464 | ATTT | A | 0.5132 | 0.071 | 0.007 | 3.1623E-23 | 102.88 |
| *MYOM3* | rs1799938 | G | A | 0.1147 | 0.099 | 0.011 | 3.1623E-19 | 81.00 |
| *MZB1* | rs149547076 | T | C | 0.0107 | -0.547 | 0.037 | 1.5849E-50 | 218.56 |
| *MZB1* | rs11739135 | G | C | 0.4152 | 0.051 | 0.008 | 7.9433E-12 | 40.64 |
| *MZB1* | rs140248321 | A | AGAT | 0.0644 | 0.248 | 0.017 | 5.0119E-50 | 212.82 |
| *MZB1* | rs6796 | T | C | 0.2789 | 0.056 | 0.008 | 1.5849E-11 | 49.00 |
| *MZB1* | rs778224678 | GACT | G | 0.1142 | 0.104 | 0.012 | 5.0119E-19 | 75.11 |
| *MZB1* | rs61839660 | C | T | 0.0971 | 0.099 | 0.012 | 1.5849E-15 | 68.06 |
| *MZB1* | rs3803286 | A | G | 0.6657 | -0.062 | 0.008 | 1.9953E-15 | 60.06 |
| *MZB1* | rs10150206 | G | A | 0.6583 | 0.157 | 0.008 | 2.5119E-86 | 385.14 |
| *MZB1* | rs3803800 | A | G | 0.7886 | -0.092 | 0.009 | 1.2589E-24 | 104.49 |
| *MZB1* | rs34562254 | G | A | 0.0972 | 0.156 | 0.012 | 2.5119E-36 | 169.00 |
| *MZB1* | rs9624334 | G | C | 0.1511 | -0.085 | 0.01 | 1.5849E-16 | 72.25 |
| *MZT1* | rs1354034 | T | C | 0.6038 | 0.058 | 0.008 | 1E-13 | 52.56 |
| *NAA10* | rs1354034 | T | C | 0.6037 | 0.055 | 0.008 | 3.1623E-12 | 47.27 |
| *NAA80* | rs201512291 | G | A | 0.0065 | -0.719 | 0.047 | 5.0119E-52 | 234.02 |
| *NAA80* | rs1354034 | T | C | 0.604 | 0.066 | 0.008 | 7.9433E-18 | 68.06 |
| *NAA80* | rs6993770 | A | T | 0.2866 | -0.07 | 0.008 | 3.9811E-17 | 76.56 |
| *NAA80* | rs7896518 | A | G | 0.4237 | 0.066 | 0.008 | 2.5119E-17 | 68.06 |
| *NAA80* | rs892090 | T | G | 0.8342 | 0.071 | 0.01 | 3.1623E-12 | 50.41 |
| *NAAA* | rs5014539 | G | T | 0.5726 | 0.048 | 0.006 | 6.3096E-14 | 64.00 |
| *NAAA* | rs687339 | C | T | 0.7708 | 0.057 | 0.008 | 1.2589E-13 | 50.77 |
| *NAAA* | rs112197434 | A | T | 0.2337 | -0.669 | 0.009 | 1E-200 | 5525.44 |
| *NAAA* | rs77945361 | G | A | 0.1483 | -0.086 | 0.01 | 2.5119E-16 | 73.96 |
| *NAAA* | rs4471106 | G | A | 0.287 | -0.05 | 0.007 | 3.9811E-12 | 51.02 |
| *NAAA* | rs4878583 | C | T | 0.1734 | 0.072 | 0.009 | 3.1623E-17 | 64.00 |
| *NAAA* | rs772851187 | TGCTGTAG( | T | 0.4682 | 0.059 | 0.006 | 1E-19 | 96.69 |
| *NAAA* | rs7904738 | G | C | 0.088 | -0.087 | 0.011 | 1.5849E-14 | 62.55 |
| *NAAA* | rs2862954 | T | C | 0.5023 | -0.05 | 0.006 | 3.9811E-15 | 69.44 |
| *NAAA* | rs7096937 | T | C | 0.7301 | -0.063 | 0.007 | 5.0119E-18 | 81.00 |
| *NAAA* | rs10860794 | C | A | 0.2882 | -0.363 | 0.007 | 1E-200 | 2689.16 |
| *NAAA* | rs150641790 | T | TG | 0.0749 | 0.105 | 0.012 | 1.9953E-17 | 76.56 |
| *NAAA* | rs71435622 | A | AT | 0.2868 | 0.065 | 0.007 | 1.5849E-18 | 86.22 |
| *NAAA* | rs145078947 | G | T | 0.0026 | 0.807 | 0.063 | 1.5849E-37 | 164.08 |
| *NAAA* | rs200210321 | A | AG | 0.0742 | 0.125 | 0.012 | 6.3096E-24 | 108.51 |
| *NAAA* | rs429358 | T | C | 0.1553 | -0.118 | 0.009 | 1.5849E-39 | 171.90 |
| *NAAA* | rs626283 | G | C | 0.4416 | 0.061 | 0.007 | 6.3096E-21 | 75.94 |
| *NAAA* | rs738408 | C | T | 0.2173 | 0.056 | 0.008 | 7.9433E-13 | 49.00 |
| *NADK* | rs1014988 | G | A | 0.5095 | 0.252 | 0.007 | 1E-200 | 1296.00 |
| *NADK* | rs71309976 | C | T | 0.0107 | 0.293 | 0.037 | 3.9811E-15 | 62.71 |
| *NADK* | rs389883 | G | T | 0.6756 | 0.057 | 0.008 | 7.9433E-14 | 50.77 |
| *NADK* | rs56077246 | T | C | 0.4276 | 0.083 | 0.007 | 3.9811E-30 | 140.59 |
| *NADK* | rs59581534 | T | A | 0.0406 | 0.376 | 0.018 | 3.1623E-95 | 436.35 |
| *NAGA* | rs684773 | A | C | 0.7669 | 0.064 | 0.009 | 1E-12 | 50.57 |
| *NAGA* | rs28672722 | G | T | 0.2224 | 0.092 | 0.009 | 6.3096E-24 | 104.49 |
| *NAGA* | rs10778152 | A | G | 0.2882 | -0.211 | 0.008 | 7.943E-138 | 695.64 |
| *NAGA* | rs121434529 | C | T | 0.005 | -1.294 | 0.053 | 1.259E-129 | 596.10 |
| *NAGK* | rs11680831 | T | C | 0.3423 | -0.223 | 0.008 | 1E-173 | 777.02 |
| *NAGK* | rs60629714 | T | C | 0.2001 | -0.066 | 0.009 | 1.5849E-12 | 53.78 |
| *NAGPA* | rs1260326 | T | C | 0.6081 | -0.06 | 0.007 | 1E-15 | 73.47 |
| *NAGPA* | rs535777 | G | C | 0.1432 | -0.092 | 0.011 | 3.1623E-18 | 69.95 |
| *NAGPA* | rs28601761 | C | G | 0.419 | -0.053 | 0.007 | 7.9433E-13 | 57.33 |
| *NAGPA* | rs507666 | G | A | 0.1848 | -0.12 | 0.009 | 7.9433E-37 | 177.78 |
| *NAGPA* | rs77582897 | T | C | 0.5197 | 0.07 | 0.008 | 1.2589E-17 | 76.56 |
| *NAGPA* | rs12599777 | A | G | 0.1826 | -0.41 | 0.01 | 1E-200 | 1681.00 |
| *NAGPA* | rs186021206 | G | A | 0.0064 | 0.632 | 0.05 | 1.5849E-36 | 159.77 |
| *NAGPA* | rs1801689 | A | C | 0.0299 | 0.18 | 0.021 | 3.1623E-17 | 73.47 |
| *NAP1L4* | rs3811444 | C | T | 0.3327 | -0.053 | 0.008 | 6.3096E-12 | 43.89 |
| *NAP1L4* | rs1354034 | T | C | 0.6047 | 0.054 | 0.007 | 5.0119E-13 | 59.51 |
| *NAP1L4* | rs1018846 | G | A | 0.0639 | 0.104 | 0.015 | 2.5119E-12 | 48.07 |

| | | | | | | | | |
|---|---|---|---|---|---|---|---|---|
| *NAP1L4* | rs6993770 | A | T | 0.2865 | -0.066 | 0.008 | 1.9953E-16 | 68.06 |
| *NAP1L4* | rs57530484 | T | TAACAGAC | 0.767 | 0.263 | 0.009 | 1E-200 | 853.94 |
| *NAP1L4* | rs12449930 | C | T | 0.4664 | 0.071 | 0.007 | 1.9953E-22 | 102.88 |
| *NAPRT* | rs10282929 | G | T | 0.7959 | 0.41 | 0.009 | 1E-200 | 2075.31 |
| *NARS1* | rs1354034 | T | C | 0.6043 | 0.061 | 0.008 | 1.2589E-15 | 58.14 |
| *NARS1* | rs13257521 | C | G | 0.2109 | 0.091 | 0.009 | 6.3096E-23 | 102.23 |
| *NARS1* | rs60822569 | T | TTTCTC | 0.5541 | 0.053 | 0.008 | 3.1623E-12 | 43.89 |
| *NBL1* | rs12408663 | T | C | 0.2322 | -0.123 | 0.009 | 1.9953E-44 | 186.78 |
| *NBL1* | rs1260326 | T | C | 0.6087 | 0.055 | 0.008 | 3.1623E-13 | 47.27 |
| *NBL1* | rs36060036 | C | T | 0.1674 | -0.076 | 0.01 | 1.2589E-14 | 57.76 |
| *NBN* | rs1354034 | T | C | 0.6035 | 0.061 | 0.008 | 6.3096E-16 | 58.14 |
| *NBN* | rs143331722 | G | A | 0.0122 | -0.315 | 0.036 | 2.5119E-18 | 76.56 |
| *NBN* | rs74378230 | A | G | 0.0044 | -0.607 | 0.058 | 6.3096E-26 | 109.53 |
| *NCAM1* | rs333947 | G | A | 0.1479 | -0.065 | 0.01 | 1E-11 | 42.25 |
| *NCAM1* | rs2301455 | C | T | 0.5586 | 0.06 | 0.007 | 3.1623E-18 | 73.47 |
| *NCAM1* | rs66824127 | T | C | 0.5574 | 0.06 | 0.007 | 1.9953E-18 | 73.47 |
| *NCAM1* | rs28383282 | C | T | 0.2115 | 0.062 | 0.009 | 6.3096E-13 | 47.46 |
| *NCAM1* | rs11214489 | C | T | 0.1849 | -0.333 | 0.009 | 1E-200 | 1369.00 |
| *NCAM1* | rs705702 | A | G | 0.337 | 0.057 | 0.007 | 2.5119E-15 | 66.31 |
| *NCAM1* | rs9318188 | A | G | 0.5445 | -0.114 | 0.007 | 1E-60 | 265.22 |
| *NCAM1* | rs2035645 | T | G | 0.5619 | 0.107 | 0.007 | 5.0119E-54 | 233.65 |
| *NCAM1* | rs776023876 | CCCCA | C | 0.6939 | 0.085 | 0.007 | 3.9811E-30 | 147.45 |
| *NCAM2* | rs199922514 | A | AAGAAGAA | 0.9078 | -0.079 | 0.011 | 6.3096E-12 | 51.58 |
| *NCAM2* | rs2071916 | T | C | 0.5859 | -0.194 | 0.007 | 1.585E-182 | 768.08 |
| *NCAM2* | rs186021206 | G | A | 0.0064 | 0.427 | 0.045 | 3.1623E-21 | 90.04 |
| *NCAM2* | rs77542162 | A | G | 0.0229 | 0.188 | 0.022 | 1.5849E-17 | 73.02 |
| *NCAM2* | rs4760 | A | G | 0.156 | 0.116 | 0.009 | 3.1623E-37 | 166.12 |
| *NCAM2* | rs760715 | T | C | 0.2473 | 0.068 | 0.008 | 2.5119E-17 | 72.25 |
| *NCAN* | rs61804160 | T | A | 0.1424 | 0.067 | 0.01 | 1E-11 | 44.89 |
| *NCAN* | rs6786821 | C | G | 0.6218 | 0.095 | 0.007 | 1.2589E-40 | 184.18 |
| *NCAN* | rs12521652 | A | G | 0.3299 | -0.05 | 0.007 | 1.5849E-11 | 51.02 |
| *NCAN* | rs6913309 | T | A | 0.2798 | 0.053 | 0.008 | 1.2589E-11 | 43.89 |
| *NCAN* | rs12175504 | T | G | 0.3542 | 0.083 | 0.007 | 5.0119E-30 | 140.59 |
| *NCAN* | rs12801188 | G | A | 0.3614 | 0.057 | 0.007 | 2.5119E-15 | 66.31 |
| *NCAN* | rs3751198 | A | G | 0.6038 | 0.055 | 0.007 | 2.5119E-14 | 61.73 |
| *NCAN* | rs60111444 | A | G | 0.226 | 0.068 | 0.008 | 1.5849E-16 | 72.25 |
| *NCAN* | rs7191514 | G | A | 0.3425 | -0.075 | 0.007 | 2.5119E-24 | 114.80 |
| *NCAN* | rs6504649 | C | G | 0.3879 | -0.068 | 0.007 | 1.5849E-21 | 94.37 |
| *NCAN* | rs7234321 | T | G | 0.3776 | -0.062 | 0.007 | 5.0119E-18 | 78.45 |
| *NCAN* | rs3761077 | G | T | 0.1102 | -0.241 | 0.011 | 2.512E-102 | 480.01 |
| *NCF2* | rs35012521 | T | A | 0.0101 | -0.948 | 0.036 | 6.31E-154 | 693.44 |
| *NCF2* | rs149698681 | G | C | 0.0216 | 0.209 | 0.025 | 3.9811E-17 | 69.89 |
| *NCF2* | rs12941811 | T | C | 0.5748 | -0.055 | 0.007 | 1.9953E-14 | 61.73 |
| *NCF2* | rs4632248 | G | T | 0.2093 | 0.158 | 0.009 | 1.5849E-71 | 308.20 |
| *NCLN* | rs9268228 | G | A | 0.6261 | 0.06 | 0.008 | 7.9433E-13 | 56.25 |
| *NCR1* | rs181807963 | G | T | 0.0076 | -0.307 | 0.043 | 1.2589E-12 | 50.97 |
| *NCR1* | rs6693836 | G | A | 0.6384 | 0.065 | 0.008 | 1.2589E-17 | 66.02 |
| *NCR1* | rs11586197 | T | C | 0.2478 | 0.067 | 0.008 | 2.5119E-15 | 70.14 |
| *NCR1* | rs76529516 | A | G | 0.0276 | 0.155 | 0.022 | 2.5119E-12 | 49.64 |
| *NCR1* | rs2009581 | G | A | 0.2725 | -0.127 | 0.008 | 1.9953E-54 | 252.02 |
| *NCR1* | rs13104208 | T | C | 0.1595 | 0.082 | 0.01 | 1.2589E-16 | 67.24 |
| *NCR1* | rs2248116 | C | A | 0.5708 | -0.062 | 0.007 | 2.5119E-17 | 78.45 |
| *NCR1* | rs9260316 | G | T | 0.5653 | -0.064 | 0.007 | 7.9433E-18 | 83.59 |
| *NCR1* | rs78565598 | T | G | 0.1678 | 0.07 | 0.01 | 1E-12 | 49.00 |
| *NCR1* | rs7086174 | C | A | 0.3286 | 0.083 | 0.008 | 1.5849E-26 | 107.64 |
| *NCR1* | rs1250569 | T | C | 0.4451 | 0.064 | 0.007 | 3.9811E-18 | 83.59 |
| *NCR1* | rs6586167 | T | G | 0.4986 | 0.056 | 0.007 | 2.5119E-14 | 64.00 |
| *NCR1* | rs745932882 | AG | A | 0.4562 | 0.062 | 0.007 | 3.1623E-17 | 78.45 |
| *NCR1* | rs3003 | C | T | 0.8377 | -0.086 | 0.01 | 2.5119E-16 | 73.96 |
| *NCR1* | rs3782473 | T | C | 0.2371 | 0.076 | 0.009 | 1E-18 | 71.31 |
| *NCR1* | rs3184504 | T | C | 0.5174 | -0.185 | 0.007 | 6.31E-143 | 698.47 |
| *NCR1* | rs755343690 | TCTGGTGG | T | 0.6187 | 0.087 | 0.008 | 5.0119E-30 | 118.27 |
| *NCR1* | rs9916257 | G | T | 0.4393 | -0.082 | 0.007 | 3.1623E-29 | 137.22 |
| *NCR1* | rs4065275 | A | G | 0.5008 | 0.071 | 0.007 | 1.5849E-22 | 102.88 |
| *NCR1* | rs2659007 | G | A | 0.4505 | 0.056 | 0.007 | 1.9953E-14 | 64.00 |
| *NCR1* | rs7255591 | G | C | 0.0887 | -0.426 | 0.013 | 1E-200 | 1073.82 |
| *NCR1* | rs6103572 | T | C | 0.726 | 0.065 | 0.008 | 1E-15 | 66.02 |
| *NCR3LG1* | rs61747728 | C | T | 0.038 | 0.135 | 0.017 | 7.9433E-15 | 63.06 |

| | | | | | | | | |
|---|---|---|---|---|---|---|---|---|
| *NCR3LG1* | rs1047891 | C | A | 0.3128 | 0.05 | 0.007 | 3.9811E-12 | 51.02 |
| *NCR3LG1* | rs10935473 | G | T | 0.4404 | -0.301 | 0.007 | 1E-200 | 1849.00 |
| *NCR3LG1* | rs10058350 | T | C | 0.7024 | 0.062 | 0.007 | 1.9953E-17 | 78.45 |
| *NCR3LG1* | rs507666 | G | A | 0.1844 | -0.326 | 0.009 | 1E-200 | 1312.05 |
| *NCR3LG1* | rs12146443 | T | C | 0.6173 | 0.361 | 0.007 | 1E-200 | 2659.61 |
| *NCR3LG1* | rs36027301 | C | T | 0.057 | 0.107 | 0.015 | 6.3096E-13 | 50.88 |
| *NCR3LG1* | rs777834943 | C | CT | 0.1834 | -0.06 | 0.009 | 5.0119E-12 | 44.44 |
| *NCR3LG1* | rs186021206 | G | A | 0.0065 | 0.954 | 0.046 | 5.0119E-97 | 430.11 |
| *NCR3LG1* | rs12980852 | G | A | 0.6599 | -0.057 | 0.007 | 1E-15 | 66.31 |
| *NCS1* | rs66927685 | T | TGAACCTTA | 0.5268 | -0.049 | 0.007 | 1.5849E-12 | 49.00 |
| *NCS1* | rs4704825 | A | G | 0.6366 | 0.051 | 0.007 | 3.9811E-13 | 53.08 |
| *NCS1* | rs662 | T | C | 0.2875 | 0.061 | 0.008 | 3.9811E-16 | 58.14 |
| *NCS1* | rs12342201 | G | A | 0.4765 | -0.071 | 0.007 | 5.0119E-25 | 102.88 |
| *NCS1* | rs1054879 | G | A | 0.5078 | 0.118 | 0.007 | 1.5849E-66 | 284.16 |
| *NCS1* | rs9807819 | C | T | 0.3625 | -0.056 | 0.007 | 1.9953E-15 | 64.00 |
| *NCS1* | rs6065904 | G | A | 0.2153 | -0.056 | 0.008 | 1.2589E-11 | 49.00 |
| *NDUFB7* | rs6580981 | G | A | 0.459 | 0.054 | 0.008 | 1E-12 | 45.56 |
| *NEB* | rs41341748 | G | A | 0.011 | 0.266 | 0.037 | 6.3096E-13 | 51.68 |
| *NECAP2* | rs2393967 | A | C | 0.3142 | 0.062 | 0.008 | 7.9433E-14 | 60.06 |
| *NECTIN2* | rs61747728 | C | T | 0.038 | 0.183 | 0.019 | 2.5119E-21 | 92.77 |
| *NECTIN2* | rs553340154 | T | G | 0.3345 | 0.065 | 0.009 | 3.1623E-12 | 52.16 |
| *NECTIN2* | rs777834943 | C | CT | 0.1835 | -0.079 | 0.01 | 6.3096E-16 | 62.41 |
| *NECTIN2* | rs1137844 | C | G | 0.3153 | 0.06 | 0.008 | 1.5849E-13 | 56.25 |
| *NECTIN2* | rs440277 | G | A | 0.3104 | -0.183 | 0.008 | 3.162E-112 | 523.27 |
| *NECTIN2* | rs601338 | G | A | 0.5092 | -0.158 | 0.008 | 3.1623E-97 | 390.06 |
| *NECTIN4* | rs35434391 | G | T | 0.1171 | -0.447 | 0.012 | 1E-200 | 1387.56 |
| *NECTIN4* | rs61747728 | C | T | 0.0379 | 0.144 | 0.019 | 1.2589E-13 | 57.44 |
| *NECTIN4* | rs1260326 | T | C | 0.6087 | 0.068 | 0.007 | 1.5849E-19 | 94.37 |
| *NECTIN4* | rs79069746 | T | C | 0.1382 | 0.1 | 0.011 | 1.9953E-21 | 82.64 |
| *NECTIN4* | rs77924615 | G | A | 0.1968 | -0.079 | 0.009 | 1.9953E-17 | 77.05 |
| *NEDD4L* | rs77100396 | C | A | 0.016 | 0.216 | 0.03 | 3.9811E-13 | 51.84 |
| *NEDD4L* | rs74930558 | G | A | 0.2182 | 0.086 | 0.009 | 6.3096E-22 | 91.31 |
| *NEDD9* | rs34436714 | C | A | 0.2092 | 0.11 | 0.009 | 1.9953E-31 | 149.38 |
| *NEFL* | rs7197999 | C | T | 0.5236 | 0.046 | 0.007 | 1.9953E-12 | 43.18 |
| *NELL1* | rs10890482 | T | C | 0.8084 | 0.076 | 0.009 | 3.1623E-17 | 71.31 |
| *NELL1* | rs2990223 | A | G | 0.6053 | 0.061 | 0.007 | 3.9811E-17 | 75.94 |
| *NELL1* | rs4344946 | A | G | 0.8642 | -0.102 | 0.01 | 6.3096E-23 | 104.04 |
| *NELL1* | rs3903119 | T | C | 0.6603 | -0.059 | 0.007 | 2.5119E-15 | 71.04 |
| *NELL1* | rs13107325 | C | T | 0.0749 | 0.176 | 0.013 | 2.5119E-39 | 183.29 |
| *NELL1* | rs9501590 | A | G | 0.3401 | 0.054 | 0.007 | 5.0119E-13 | 59.51 |
| *NELL1* | rs2207136 | T | C | 0.4668 | -0.053 | 0.007 | 6.3096E-14 | 57.33 |
| *NELL1* | rs11458788 | A | AC | 0.5053 | 0.051 | 0.007 | 1E-12 | 53.08 |
| *NELL1* | rs2339228 | G | A | 0.5017 | 0.063 | 0.007 | 1.9953E-19 | 81.00 |
| *NELL1* | rs56278466 | T | G | 0.6612 | 0.067 | 0.007 | 1.2589E-19 | 91.61 |
| *NELL1* | rs7917848 | C | G | 0.6511 | -0.099 | 0.007 | 7.9433E-41 | 200.02 |
| *NELL1* | rs61652119 | G | A | 0.0486 | 0.794 | 0.017 | 1E-200 | 2181.44 |
| *NELL1* | rs542695580 | C | CT | 0.373 | 0.065 | 0.008 | 1.5849E-16 | 66.02 |
| *NELL1* | rs375125008 | T | TTTTTTC | 0.3779 | -0.067 | 0.008 | 3.1623E-18 | 70.14 |
| *NELL1* | rs2298475 | T | C | 0.0753 | -0.163 | 0.014 | 3.1623E-32 | 135.56 |
| *NELL1* | rs7298766 | A | G | 0.2942 | 0.235 | 0.008 | 7.943E-197 | 862.89 |
| *NELL1* | rs141826184 | C | T | 0.0045 | 0.825 | 0.054 | 1.9953E-53 | 233.41 |
| *NELL1* | rs75526993 | G | A | 0.1425 | 0.126 | 0.01 | 2.5119E-34 | 158.76 |
| *NELL1* | rs4541022 | C | T | 0.8812 | -0.085 | 0.011 | 6.3096E-15 | 59.71 |
| *NELL1* | rs186021206 | G | A | 0.0064 | 0.622 | 0.049 | 1.2589E-37 | 161.13 |
| *NELL1* | rs72835417 | G | A | 0.11 | 0.14 | 0.011 | 1.9953E-35 | 161.98 |
| *NELL2* | rs17285324 | A | T | 0.1747 | -0.07 | 0.009 | 1E-13 | 60.49 |
| *NELL2* | rs13303 | T | C | 0.5597 | 0.063 | 0.007 | 3.9811E-18 | 81.00 |
| *NELL2* | rs1126735 | A | G | 0.3688 | 0.056 | 0.007 | 5.0119E-14 | 64.00 |
| *NELL2* | rs55818129 | T | G | 0.1176 | -0.077 | 0.011 | 6.3096E-12 | 49.00 |
| *NELL2* | rs155628 | C | A | 0.3182 | -0.055 | 0.008 | 1E-12 | 47.27 |
| *NELL2* | rs113239947 | A | T | 0.2427 | -0.068 | 0.008 | 2.5119E-16 | 72.25 |
| *NELL2* | rs2519093 | C | T | 0.1843 | -0.198 | 0.009 | 1.585E-100 | 484.00 |
| *NELL2* | rs56278466 | T | G | 0.6615 | 0.062 | 0.008 | 1.2589E-16 | 60.06 |
| *NELL2* | rs76485191 | G | T | 0.3398 | 0.056 | 0.008 | 5.0119E-13 | 49.00 |
| *NELL2* | rs36027301 | C | T | 0.0569 | -0.125 | 0.015 | 6.3096E-16 | 69.44 |
| *NELL2* | rs382115 | T | C | 0.6037 | -0.174 | 0.007 | 1.259E-121 | 617.88 |
| *NELL2* | rs540730 | T | C | 0.751 | -0.066 | 0.008 | 3.1623E-15 | 68.06 |
| *NELL2* | rs186021206 | G | A | 0.0064 | 0.426 | 0.049 | 3.9811E-18 | 75.58 |

| | | | | | | | | |
|---|---|---|---|---|---|---|---|---|
| *NELL2* | rs4804790 | G | T | 0.0708 | -0.132 | 0.014 | 3.9811E-21 | 88.90 |
| *NENF* | rs607929 | C | G | 0.4492 | -0.089 | 0.008 | 7.9433E-31 | 123.77 |
| *NENF* | rs6473226 | T | C | 0.6259 | 0.06 | 0.008 | 3.9811E-14 | 56.25 |
| *NEO1* | rs11632433 | T | G | 0.0977 | 0.114 | 0.013 | 2.5119E-19 | 76.90 |
| *NEXN* | rs71590706 | AT | A | 0.0764 | -0.149 | 0.014 | 2.5119E-26 | 113.27 |
| *NEXN* | rs1354034 | T | C | 0.6042 | 0.055 | 0.008 | 3.1623E-13 | 47.27 |
| *NEXN* | rs342293 | C | G | 0.4596 | -0.052 | 0.007 | 3.1623E-12 | 55.18 |
| *NEXN* | rs7924036 | G | T | 0.5013 | 0.052 | 0.007 | 1.9953E-12 | 55.18 |
| *NFASC* | rs2519093 | C | T | 0.1843 | -0.09 | 0.009 | 3.9811E-25 | 100.00 |
| *NFASC* | rs56278466 | T | G | 0.6613 | 0.167 | 0.007 | 6.31E-121 | 569.16 |
| *NFASC* | rs58534292 | T | C | 0.65 | 0.054 | 0.007 | 2.5119E-14 | 59.51 |
| *NFASC* | rs12895651 | T | C | 0.7007 | 0.092 | 0.008 | 2.5119E-29 | 132.25 |
| *NFASC* | rs3747207 | G | A | 0.2157 | 0.066 | 0.008 | 1E-15 | 68.06 |
| *NFATC1* | rs657693 | A | G | 0.3602 | -0.11 | 0.008 | 1.5849E-46 | 189.06 |
| *NFATC3* | rs570919834 | C | T | 0.2555 | 0.108 | 0.01 | 7.9433E-30 | 116.64 |
| *NFATC3* | rs13263709 | T | C | 0.6481 | 0.06 | 0.008 | 7.9433E-14 | 56.25 |
| *NFE2* | rs1354034 | T | C | 0.6045 | 0.053 | 0.008 | 3.9811E-12 | 43.89 |
| *NFE2* | rs342298 | C | T | 0.4548 | -0.069 | 0.008 | 2.5119E-20 | 74.39 |
| *NFE2* | rs60822569 | T | TTTCTC | 0.553 | 0.066 | 0.008 | 1.9953E-18 | 68.06 |
| *NFKB1* | rs1354034 | T | C | 0.6037 | 0.063 | 0.008 | 2.5119E-16 | 62.02 |
| *NFKB1* | rs17032705 | G | A | 0.3982 | 0.115 | 0.008 | 1.2589E-50 | 206.64 |
| *NFKBIE* | rs1354034 | T | C | 0.6038 | 0.058 | 0.008 | 2.5119E-14 | 52.56 |
| *NFKBIE* | rs28362856 | G | C | 0.0431 | -0.523 | 0.019 | 3.981E-174 | 757.70 |
| *NFU1* | rs74637005 | G | A | 0.0366 | -0.328 | 0.021 | 5.0119E-57 | 243.95 |
| *NFU1* | rs1354034 | T | C | 0.6046 | 0.058 | 0.008 | 5.0119E-14 | 52.56 |
| *NFU1* | rs342299 | C | T | 0.4551 | -0.054 | 0.008 | 1.9953E-12 | 45.56 |
| *NFU1* | rs6993770 | A | T | 0.2866 | -0.073 | 0.008 | 1.9953E-18 | 83.27 |
| *NFU1* | rs6580981 | G | A | 0.459 | 0.059 | 0.008 | 1E-14 | 54.39 |
| *NFX1* | rs1354034 | T | C | 0.6037 | 0.057 | 0.008 | 6.3096E-14 | 50.77 |
| *NGRN* | rs3751488 | G | A | 0.2345 | 0.061 | 0.009 | 1E-11 | 45.94 |
| *NGRN* | rs11085147 | C | T | 0.099 | 0.119 | 0.013 | 6.3096E-20 | 83.79 |
| *NHLRC3* | rs9264645 | G | A | 0.6414 | 0.12 | 0.008 | 1.5849E-54 | 225.00 |
| *NHLRC3* | rs56278466 | T | G | 0.6605 | 0.057 | 0.008 | 1.5849E-13 | 50.77 |
| *NHLRC3* | rs111338191 | A | AAATT | 0.5199 | -0.056 | 0.007 | 3.1623E-14 | 64.00 |
| *NHLRC3* | rs149175958 | G | T | 0.0044 | -2.127 | 0.057 | 1E-200 | 1392.47 |
| *NHLRC3* | rs1801248 | C | T | 0.0432 | 0.206 | 0.018 | 3.1623E-29 | 130.98 |
| *NHLRC3* | rs2269335 | G | T | 0.3257 | 0.063 | 0.008 | 1.9953E-15 | 62.02 |
| *NHLRC3* | rs112001035 | G | A | 0.0613 | -0.107 | 0.016 | 7.9433E-12 | 44.72 |
| *NID1* | rs6671171 | G | A | 0.111 | 0.085 | 0.012 | 3.9811E-13 | 50.17 |
| *NID1* | rs76183323 | G | A | 0.0106 | -0.762 | 0.038 | 1E-87 | 402.11 |
| *NID1* | rs13412535 | G | A | 0.2309 | -0.069 | 0.009 | 1.2589E-15 | 58.78 |
| *NID1* | rs11719450 | C | T | 0.3702 | 0.064 | 0.008 | 2.5119E-17 | 64.00 |
| *NID1* | rs34716199 | A | AT | 0.5356 | 0.055 | 0.007 | 1.2589E-13 | 61.73 |
| *NID1* | rs4098923 | A | G | 0.4475 | -0.069 | 0.008 | 1.5849E-19 | 74.39 |
| *NID1* | rs342299 | C | T | 0.4551 | -0.056 | 0.007 | 3.1623E-14 | 64.00 |
| *NID1* | rs6993770 | A | T | 0.2865 | -0.057 | 0.008 | 1.5849E-12 | 50.77 |
| *NID1* | rs927826 | G | T | 0.7057 | 0.187 | 0.008 | 7.943E-120 | 546.39 |
| *NID1* | rs10761731 | A | T | 0.4147 | 0.089 | 0.007 | 3.1623E-33 | 161.65 |
| *NID1* | rs35350651 | A | AC | 0.5037 | -0.05 | 0.007 | 3.1623E-12 | 51.02 |
| *NID1* | rs12445050 | C | T | 0.1383 | 0.091 | 0.011 | 5.0119E-18 | 68.44 |
| *NID1* | rs892090 | T | G | 0.8345 | 0.129 | 0.01 | 5.0119E-39 | 166.41 |
| *NID2* | rs13412535 | G | A | 0.231 | -0.058 | 0.008 | 7.9433E-13 | 52.56 |
| *NID2* | rs7618405 | C | A | 0.2046 | -0.058 | 0.009 | 1E-11 | 41.53 |
| *NID2* | rs1354034 | T | C | 0.6041 | -0.056 | 0.007 | 5.0119E-16 | 64.00 |
| *NID2* | rs755492124 | TAAAG | T | 0.3934 | -0.059 | 0.007 | 2.5119E-17 | 71.04 |
| *NID2* | rs274555 | C | T | 0.3986 | 0.052 | 0.007 | 2.5119E-13 | 55.18 |
| *NID2* | rs35173808 | C | A | 0.0963 | -0.096 | 0.012 | 1.2589E-16 | 64.00 |
| *NID2* | rs1917342 | A | G | 0.3032 | -0.051 | 0.007 | 5.0119E-12 | 53.08 |
| *NID2* | rs6961069 | C | T | 0.4022 | 0.058 | 0.007 | 1.2589E-16 | 68.65 |
| *NID2* | rs6993770 | A | T | 0.2864 | -0.095 | 0.008 | 5.0119E-36 | 141.02 |
| *NID2* | rs10820606 | A | C | 0.2299 | -0.071 | 0.008 | 5.0119E-18 | 78.77 |
| *NID2* | rs10740118 | G | C | 0.4147 | 0.088 | 0.007 | 1.9953E-36 | 158.04 |
| *NID2* | rs12767683 | C | A | 0.3246 | 0.057 | 0.007 | 7.9433E-15 | 66.31 |
| *NID2* | rs35277580 | G | A | 0.492 | -0.048 | 0.007 | 1.9953E-12 | 47.02 |
| *NID2* | rs2516600 | T | C | 0.5437 | 0.576 | 0.008 | 1E-200 | 5184.00 |
| *NID2* | rs59001897 | T | A | 0.1746 | 0.065 | 0.009 | 6.3096E-13 | 52.16 |
| *NID2* | rs12445050 | C | T | 0.1383 | 0.1 | 0.01 | 3.1623E-24 | 100.00 |
| *NID2* | rs892090 | T | G | 0.8342 | 0.143 | 0.009 | 6.3096E-54 | 252.46 |

| | | | | | | | | |
|---|---|---|---|---|---|---|---|---|
| *NID2* | rs3827978 | C | T | 0.3535 | 0.07 | 0.007 | 2.5119E-22 | 100.00 |
| *NINJ1* | rs9274442 | G | C | 0.2824 | -0.084 | 0.009 | 7.9433E-20 | 87.11 |
| *NINJ1* | rs11379524 | A | AC | 0.6301 | 0.243 | 0.008 | 1E-200 | 922.64 |
| *NIT1* | rs76502631 | G | A | 0.0284 | -0.726 | 0.023 | 1E-200 | 996.36 |
| *NIT2* | rs200416332 | G | A | 0.0016 | -1.264 | 0.094 | 7.9433E-41 | 180.82 |
| *NME3* | rs188468174 | C | T | 0.0143 | -0.372 | 0.031 | 2.5119E-32 | 144.00 |
| *NME3* | rs57008601 | T | G | 0.5689 | -0.055 | 0.007 | 1.9953E-13 | 61.73 |
| *NME3* | rs796157456 | AC | A | 0.5 | 0.051 | 0.007 | 6.3096E-12 | 53.08 |
| *NME3* | rs373385028 | A | T | 0.5511 | -0.079 | 0.007 | 1.9953E-26 | 127.37 |
| *NME3* | rs1014651 | G | A | 0.5312 | 0.05 | 0.007 | 1.5849E-11 | 51.02 |
| *NME3* | rs2519093 | C | T | 0.1843 | -0.113 | 0.01 | 2.5119E-32 | 127.69 |
| *NME3* | rs11825456 | T | A | 0.0674 | 0.103 | 0.015 | 3.9811E-12 | 47.15 |
| *NME3* | rs2193005 | G | C | 0.3527 | -0.253 | 0.008 | 1E-200 | 1000.14 |
| *NME3* | rs2278093 | C | A | 0.2846 | -0.059 | 0.008 | 1.9953E-12 | 54.39 |
| *NME3* | rs12424638 | G | A | 0.0765 | -0.122 | 0.015 | 5.0119E-16 | 66.15 |
| *NME3* | rs55707100 | C | T | 0.0241 | -0.178 | 0.024 | 1E-13 | 55.01 |
| *NME3* | rs749285440 | G | A | 0.0014 | 3.052 | 0.108 | 1E-176 | 798.59 |
| *NME3* | rs1801689 | A | C | 0.0302 | 0.156 | 0.022 | 3.9811E-13 | 50.28 |
| *NME3* | rs312068 | T | C | 0.4533 | 0.093 | 0.007 | 2.5119E-35 | 176.51 |
| *NME3* | rs7599 | A | G | 0.6336 | -0.152 | 0.008 | 1.5849E-88 | 361.00 |
| *NME3* | rs12975366 | T | C | 0.3952 | -0.077 | 0.008 | 1.5849E-24 | 92.64 |
| *NME3* | rs5749077 | A | C | 0.2309 | 0.064 | 0.009 | 7.9433E-13 | 50.57 |
| *NMI* | rs1048135 | G | A | 0.4722 | -0.453 | 0.008 | 1E-200 | 3206.39 |
| *NMI* | rs1354034 | T | C | 0.6044 | 0.051 | 0.007 | 6.3096E-12 | 53.08 |
| *NMI* | rs528854 | G | A | 0.9077 | 0.092 | 0.013 | 1.9953E-13 | 50.08 |
| *NMNAT1* | rs180754466 | C | A | 0.0096 | 0.671 | 0.039 | 3.9811E-66 | 296.02 |
| *NMNAT1* | rs9271573 | A | C | 0.5975 | 0.054 | 0.007 | 2.5119E-13 | 59.51 |
| *NMNAT1* | rs58534292 | T | C | 0.6501 | -0.088 | 0.008 | 2.5119E-30 | 121.00 |
| *NMNAT1* | rs34799380 | T | C | 0.1515 | -0.19 | 0.01 | 1E-77 | 361.00 |
| *NMNAT1* | rs704 | G | A | 0.4733 | -0.2 | 0.007 | 1E-165 | 816.33 |
| *NMT1* | rs1354034 | T | C | 0.6043 | 0.055 | 0.008 | 7.9433E-13 | 47.27 |
| *NOMO1* | rs111779744 | C | CCA | 0.1512 | 0.084 | 0.01 | 1.5849E-17 | 70.56 |
| *NOMO1* | rs174544 | C | A | 0.3087 | 0.077 | 0.008 | 1E-24 | 92.64 |
| *NOMO1* | rs28929474 | C | T | 0.0207 | 0.396 | 0.025 | 1.2589E-58 | 250.91 |
| *NOMO1* | rs3891245 | T | G | 0.228 | -0.511 | 0.009 | 1E-200 | 3223.72 |
| *NOMO1* | rs1801689 | A | C | 0.0301 | 0.183 | 0.02 | 3.1623E-19 | 83.72 |
| *NOS1* | rs12574283 | G | A | 0.7099 | 0.068 | 0.008 | 1.5849E-17 | 72.25 |
| *NOS1* | rs4767532 | C | T | 0.2036 | -0.142 | 0.009 | 1E-56 | 248.94 |
| *NOS2* | rs2297518 | G | A | 0.1888 | -0.091 | 0.01 | 1E-20 | 82.81 |
| *NOS3* | rs7572209 | T | C | 0.1988 | -0.085 | 0.009 | 2.5119E-19 | 89.20 |
| *NOS3* | rs6415128 | G | A | 0.5589 | 0.051 | 0.008 | 1.2589E-11 | 40.64 |
| *NOTCH1* | rs1260326 | T | C | 0.6084 | 0.058 | 0.007 | 7.9433E-15 | 68.65 |
| *NOTCH1* | rs4240624 | G | A | 0.908 | -0.106 | 0.013 | 5.0119E-17 | 66.49 |
| *NOTCH1* | rs182330532 | G | A | 0.0012 | 1.22 | 0.107 | 5.0119E-30 | 130.00 |
| *NOTCH1* | rs56278466 | T | G | 0.6614 | 0.079 | 0.008 | 3.9811E-25 | 97.52 |
| *NOTCH1* | rs34434834 | G | A | 0.0334 | 0.142 | 0.02 | 3.1623E-12 | 50.41 |
| *NOTCH1* | rs111338191 | A | AAATT | 0.5209 | -0.051 | 0.007 | 3.9811E-12 | 53.08 |
| *NOTCH1* | rs186021206 | G | A | 0.0064 | 0.636 | 0.05 | 7.9433E-37 | 161.80 |
| *NOTCH2* | rs2641348 | A | G | 0.1111 | -0.172 | 0.012 | 1.9953E-47 | 205.44 |
| *NOTCH2* | rs12088164 | A | G | 0.0382 | 1.611 | 0.019 | 1E-200 | 7189.25 |
| *NOTCH2* | rs17580 | T | A | 0.0497 | 0.287 | 0.016 | 1.2589E-74 | 321.75 |
| *NOTCH2* | rs186021206 | G | A | 0.0064 | 0.331 | 0.047 | 1.9953E-12 | 49.60 |
| *NOTCH2* | rs77542162 | A | G | 0.0229 | 0.158 | 0.023 | 5.0119E-12 | 47.19 |
| *NOTCH3* | rs61801010 | A | G | 0.1062 | 0.103 | 0.012 | 5.0119E-18 | 73.67 |
| *NOTCH3* | rs1260326 | T | C | 0.6084 | 0.068 | 0.007 | 1.5849E-21 | 94.37 |
| *NOTCH3* | rs8192585 | G | A | 0.0467 | -0.115 | 0.017 | 3.9811E-12 | 45.76 |
| *NOTCH3* | rs34114302 | T | C | 0.6776 | 0.071 | 0.007 | 2.5119E-21 | 102.88 |
| *NOTCH3* | rs774272255 | TTA | T | 0.2768 | 0.064 | 0.008 | 6.3096E-16 | 64.00 |
| *NOTCH3* | rs507666 | G | A | 0.1845 | -0.073 | 0.009 | 1E-15 | 65.79 |
| *NOTCH3* | rs12359178 | G | A | 0.1447 | 0.081 | 0.01 | 3.1623E-16 | 65.61 |
| *NOTCH3* | rs58298943 | C | T | 0.0846 | 0.116 | 0.013 | 2.5119E-20 | 79.62 |
| *NOTCH3* | rs17580 | T | A | 0.0494 | -0.155 | 0.016 | 6.3096E-22 | 93.85 |
| *NOTCH3* | rs139974673 | T | C | 0.0243 | -0.198 | 0.023 | 2.5119E-18 | 74.11 |
| *NOTCH3* | rs186021206 | G | A | 0.0064 | 0.483 | 0.048 | 1.5849E-23 | 101.25 |
| *NOTCH3* | rs77542162 | A | G | 0.0229 | 0.189 | 0.023 | 7.9433E-16 | 67.53 |
| *NOTCH3* | rs147373451 | T | C | 0.003 | -1.324 | 0.07 | 7.9433E-80 | 357.75 |
| *NPC2* | rs28394165 | T | C | 0.4564 | 0.049 | 0.007 | 1.2589E-11 | 49.00 |
| *NPC2* | rs1050420 | C | T | 0.368 | -0.051 | 0.007 | 6.3096E-12 | 53.08 |

| | | | | | | | | |
|---|---|---|---|---|---|---|---|---|
| *NPC2* | rs773481775 | ATCTTT | A | 0.0963 | -0.139 | 0.012 | 3.9811E-30 | 134.17 |
| *NPC2* | rs3184504 | T | C | 0.5173 | -0.058 | 0.007 | 5.0119E-16 | 68.65 |
| *NPC2* | rs140130028 | C | T | 0.0091 | -2.022 | 0.038 | 1E-200 | 2831.36 |
| *NPDC1* | rs13146355 | G | A | 0.4565 | 0.051 | 0.007 | 1.5849E-12 | 53.08 |
| *NPDC1* | rs10265221 | T | C | 0.2869 | 0.059 | 0.008 | 1.5849E-13 | 54.39 |
| *NPDC1* | rs7953184 | T | G | 0.3301 | 0.053 | 0.008 | 5.0119E-12 | 43.89 |
| *NPDC1* | rs548328000 | C | CA | 0.5807 | -0.065 | 0.007 | 7.9433E-19 | 86.22 |
| *NPDC1* | rs36060036 | C | T | 0.1677 | -0.083 | 0.01 | 1.2589E-17 | 68.89 |
| *NPDC1* | rs11672900 | A | G | 0.5444 | 0.057 | 0.007 | 3.9811E-15 | 66.31 |
| *NPHS1* | rs1337526 | A | G | 0.8037 | 0.072 | 0.009 | 1.5849E-15 | 64.00 |
| *NPHS1* | rs9383023 | T | C | 0.2571 | -0.058 | 0.008 | 3.9811E-12 | 52.56 |
| *NPHS1* | rs2298475 | T | C | 0.0751 | -0.092 | 0.014 | 1.2589E-11 | 43.18 |
| *NPHS1* | rs200857931 | A | AT | 0.0751 | -0.801 | 0.015 | 1E-200 | 2851.56 |
| *NPL* | rs116448311 | C | T | 0.0055 | -1.708 | 0.045 | 1E-200 | 1440.62 |
| *NPL* | rs192695654 | C | T | 0.0059 | 0.412 | 0.043 | 2.5119E-21 | 91.80 |
| *NPL* | rs7675964 | C | T | 0.2739 | -0.093 | 0.007 | 1.2589E-36 | 176.51 |
| *NPL* | rs72492305 | C | T | 0.6749 | -0.052 | 0.007 | 7.9433E-14 | 55.18 |
| *NPL* | rs2398827 | T | C | 0.602 | 0.05 | 0.007 | 1.5849E-13 | 51.02 |
| *NPL* | rs11057841 | C | T | 0.14 | 0.132 | 0.01 | 2.5119E-42 | 174.24 |
| *NPL* | rs1065853 | G | T | 0.0812 | 0.088 | 0.012 | 3.9811E-13 | 53.78 |
| *NPPB* | rs198389 | A | G | 0.4186 | 0.183 | 0.008 | 1.259E-127 | 523.27 |
| *NPPB* | rs2234962 | T | C | 0.2156 | 0.068 | 0.009 | 3.9811E-14 | 57.09 |
| *NPPC* | rs77806605 | C | G | 0.266 | 0.052 | 0.008 | 3.9811E-12 | 42.25 |
| *NPPC* | rs4859682 | C | A | 0.4567 | 0.047 | 0.007 | 1E-12 | 45.08 |
| *NPR1* | rs9274278 | C | T | 0.4483 | -0.06 | 0.008 | 7.9433E-13 | 56.25 |
| *NPTX1* | rs78265569 | C | A | 0.0883 | 0.102 | 0.012 | 1.5849E-18 | 72.25 |
| *NPTX1* | rs12025349 | C | T | 0.6032 | 0.097 | 0.007 | 1E-47 | 192.02 |
| *NPTX1* | rs13064675 | T | C | 0.1946 | -0.059 | 0.008 | 7.9433E-13 | 54.39 |
| *NPTX1* | rs4693802 | A | G | 0.2459 | -0.063 | 0.008 | 1E-16 | 62.02 |
| *NPTX1* | rs66833621 | T | C | 0.1939 | 0.118 | 0.008 | 3.1623E-45 | 217.56 |
| *NPTX1* | rs16896742 | A | G | 0.3633 | -0.239 | 0.007 | 1E-200 | 1165.73 |
| *NPTX1* | rs2519093 | C | T | 0.1843 | -0.097 | 0.008 | 1.5849E-30 | 147.02 |
| *NPTX1* | rs10786548 | G | A | 0.891 | 0.074 | 0.01 | 1.2589E-12 | 54.76 |
| *NPTX1* | rs7937122 | C | G | 0.3903 | 0.078 | 0.007 | 1E-30 | 124.16 |
| *NPTX1* | rs186021206 | G | A | 0.0064 | 0.71 | 0.049 | 1.9953E-47 | 209.95 |
| *NPTX1* | rs62068268 | C | T | 0.0895 | 0.818 | 0.013 | 1E-200 | 3959.31 |
| *NPTX1* | rs4806469 | G | A | 0.1894 | 0.058 | 0.008 | 3.1623E-12 | 52.56 |
| *NPTX1* | rs7280064 | C | T | 0.1537 | 0.069 | 0.009 | 1.5849E-14 | 58.78 |
| *NPTX2* | rs13303 | T | C | 0.5598 | 0.055 | 0.006 | 6.3096E-19 | 84.03 |
| *NPTX2* | rs71309976 | C | T | 0.0106 | 0.281 | 0.032 | 6.3096E-19 | 77.11 |
| *NPTX2* | rs147023494 | C | T | 0.0273 | -0.171 | 0.019 | 1E-19 | 81.00 |
| *NPTX2* | rs61007246 | C | T | 0.7146 | 0.344 | 0.007 | 1E-200 | 2415.02 |
| *NPTX2* | rs144589499 | T | TA | 0.0886 | 0.177 | 0.011 | 1.2589E-61 | 258.92 |
| *NPTX2* | rs35444 | A | G | 0.3847 | -0.053 | 0.006 | 3.9811E-17 | 78.03 |
| *NPTX2* | rs36127550 | G | T | 0.178 | 0.062 | 0.008 | 5.0119E-15 | 60.06 |
| *NPTX2* | rs186021206 | G | A | 0.0064 | 0.295 | 0.042 | 1.5849E-12 | 49.33 |
| *NPTX2* | rs192164176 | C | G | 0.002 | 0.499 | 0.072 | 3.9811E-12 | 48.03 |
| *NPTXR* | rs759777208 | C | CT | 0.2001 | -0.066 | 0.009 | 2.5119E-14 | 53.78 |
| *NPTXR* | rs2240046 | C | G | 0.0627 | 0.097 | 0.014 | 1.5849E-11 | 48.01 |
| *NPTXR* | rs61007246 | C | T | 0.7143 | 0.098 | 0.008 | 3.1623E-38 | 150.06 |
| *NPTXR* | rs369705328 | G | C | 0.1283 | 0.089 | 0.01 | 7.9433E-18 | 79.21 |
| *NPTXR* | rs186021206 | G | A | 0.0064 | 0.316 | 0.047 | 1.5849E-11 | 45.20 |
| *NPTXR* | rs111577133 | A | G | 0.0193 | 2.022 | 0.027 | 1E-200 | 5608.35 |
| *NPY* | rs765974791 | TGATTTCAC | T | 0.6408 | 0.066 | 0.008 | 3.9811E-18 | 68.06 |
| *NPY* | rs5030062 | A | C | 0.3732 | -0.103 | 0.008 | 3.9811E-43 | 165.77 |
| *NPY* | rs4861708 | G | A | 0.5116 | -0.186 | 0.007 | 3.981E-143 | 706.04 |
| *NPY* | rs639933 | C | A | 0.6314 | 0.055 | 0.008 | 5.0119E-13 | 47.27 |
| *NPY* | rs1801020 | A | G | 0.7446 | -0.09 | 0.008 | 1.9953E-27 | 126.56 |
| *NPY* | rs2523675 | G | A | 0.3412 | 0.054 | 0.008 | 1.9953E-12 | 45.56 |
| *NPY* | rs56393506 | C | T | 0.1722 | 0.075 | 0.01 | 1.2589E-14 | 56.25 |
| *NPY* | rs9718966 | G | C | 0.5131 | 0.055 | 0.007 | 1.2589E-13 | 61.73 |
| *NPY* | rs4722337 | A | T | 0.5549 | -0.158 | 0.007 | 7.943E-103 | 509.47 |
| *NPY* | rs33951980 | C | T | 0.1288 | -0.077 | 0.011 | 1.2589E-12 | 49.00 |
| *NPY* | rs75802599 | G | C | 0.0899 | -0.089 | 0.013 | 2.5119E-12 | 46.87 |
| *NPY* | rs16888589 | A | G | 0.0768 | 0.176 | 0.014 | 5.0119E-38 | 158.04 |
| *NPY* | rs1809844 | C | T | 0.4971 | -0.057 | 0.007 | 6.3096E-15 | 66.31 |
| *NPY* | rs77542162 | A | G | 0.0229 | 0.301 | 0.024 | 1.9953E-35 | 157.29 |
| *NPY* | rs72654473 | C | A | 0.1093 | 0.143 | 0.012 | 6.3096E-35 | 142.01 |

| | | | | | | | | |
|---|---|---|---|---|---|---|---|---|
| *NRCAM* | rs61804210 | C | T | 0.1047 | 0.104 | 0.012 | 3.1623E-18 | 75.11 |
| *NRCAM* | rs1260326 | T | C | 0.6083 | 0.065 | 0.007 | 3.9811E-18 | 86.22 |
| *NRCAM* | rs1391373 | T | C | 0.359 | 0.057 | 0.008 | 3.9811E-14 | 50.77 |
| *NRCAM* | rs138528835 | T | TTAAA | 0.3082 | 0.298 | 0.008 | 1E-200 | 1387.56 |
| *NRCAM* | rs532436 | G | A | 0.1848 | -0.109 | 0.009 | 7.9433E-31 | 146.68 |
| *NRCAM* | rs56278466 | T | G | 0.6615 | 0.138 | 0.008 | 3.1623E-72 | 297.56 |
| *NRCAM* | rs1265564 | A | C | 0.4419 | 0.051 | 0.007 | 1E-11 | 53.08 |
| *NRCAM* | rs186021206 | G | A | 0.0064 | 0.377 | 0.05 | 3.9811E-14 | 56.85 |
| *NRGN* | rs11121012 | A | G | 0.6466 | 0.086 | 0.008 | 1.5849E-27 | 115.56 |
| *NRGN* | rs1354034 | T | C | 0.6042 | 0.053 | 0.008 | 5.0119E-12 | 43.89 |
| *NRGN* | rs342293 | C | G | 0.4596 | -0.054 | 0.007 | 5.0119E-13 | 59.51 |
| *NRGN* | rs6993770 | A | T | 0.2866 | -0.065 | 0.008 | 3.1623E-15 | 66.02 |
| *NRP1* | rs181835401 | G | A | 0.3543 | 0.092 | 0.008 | 7.9433E-34 | 132.25 |
| *NRP1* | rs764992666 | TCTTAAAG[illegible] | T | 0.6034 | 0.216 | 0.007 | 1.259E-184 | 952.16 |
| *NRP1* | rs2090034 | G | A | 0.4682 | -0.052 | 0.007 | 7.9433E-13 | 55.18 |
| *NRP1* | rs1260326 | T | C | 0.6085 | 0.066 | 0.007 | 5.0119E-19 | 88.90 |
| *NRP1* | rs71071936 | C | CT | 0.1217 | 0.088 | 0.011 | 7.9433E-16 | 64.00 |
| *NRP1* | rs2506150 | G | A | 0.3595 | -0.305 | 0.008 | 1E-200 | 1453.52 |
| *NRP1* | rs186021206 | G | A | 0.0064 | 0.603 | 0.049 | 3.1623E-34 | 151.44 |
| *NRP1* | rs77542162 | A | G | 0.0229 | 0.213 | 0.024 | 1E-18 | 78.77 |
| *NRP2* | rs61801010 | A | G | 0.106 | 0.086 | 0.012 | 1.5849E-12 | 51.36 |
| *NRP2* | rs4611630 | G | A | 0.1941 | 0.062 | 0.009 | 1.2589E-11 | 47.46 |
| *NRP2* | rs2289023 | A | G | 0.2268 | -0.202 | 0.009 | 1E-120 | 503.75 |
| *NRP2* | rs17285324 | A | T | 0.1749 | -0.075 | 0.009 | 1.9953E-15 | 69.44 |
| *NRP2* | rs13081028 | G | A | 0.557 | 0.059 | 0.007 | 1.5849E-16 | 71.04 |
| *NRP2* | rs55818129 | T | G | 0.1177 | -0.076 | 0.011 | 6.3096E-12 | 47.74 |
| *NRP2* | rs2844572 | A | C | 0.343 | -0.078 | 0.008 | 3.1623E-25 | 95.06 |
| *NRP2* | rs56287412 | T | C | 0.5111 | 0.062 | 0.007 | 6.3096E-18 | 78.45 |
| *NRP2* | rs1070073 | T | G | 0.6728 | 0.072 | 0.008 | 1.5849E-21 | 81.00 |
| *NRTN* | rs36055559 | G | A | 0.1725 | -0.076 | 0.011 | 1E-12 | 47.74 |
| *NSFL1C* | rs1354034 | T | C | 0.6042 | 0.063 | 0.007 | 3.1623E-17 | 81.00 |
| *NSFL1C* | rs6042675 | G | C | 0.3894 | -0.08 | 0.008 | 3.1623E-26 | 100.00 |
| *NT5C* | rs1354034 | T | C | 0.6045 | 0.056 | 0.007 | 3.1623E-15 | 64.00 |
| *NT5C* | rs7896518 | A | G | 0.424 | 0.059 | 0.007 | 7.9433E-17 | 71.04 |
| *NT5C* | rs60822569 | T | TTTCTC | 0.5538 | 0.05 | 0.007 | 1E-12 | 51.02 |
| *NT5C* | rs78625720 | G | A | 0.0315 | -1.539 | 0.021 | 1E-200 | 5370.80 |
| *NT5C1A* | rs10631043 | T | TCTGA | 0.878 | 0.159 | 0.011 | 2.5119E-48 | 208.93 |
| *NT5C3A* | rs1354034 | T | C | 0.6043 | 0.058 | 0.008 | 2.5119E-14 | 52.56 |
| *NT5C3A* | rs2706396 | A | C | 0.5704 | 0.051 | 0.008 | 1.2589E-11 | 40.64 |
| *NT5C3A* | rs10252880 | G | C | 0.3254 | -0.109 | 0.008 | 7.9433E-42 | 185.64 |
| *NT5C3A* | rs6993770 | A | T | 0.2869 | -0.064 | 0.008 | 1.2589E-14 | 64.00 |
| *NT5C3A* | rs7896518 | A | G | 0.4241 | 0.068 | 0.008 | 1E-18 | 72.25 |
| *NT5E* | rs1497406 | A | G | 0.5795 | 0.06 | 0.007 | 1.9953E-19 | 73.47 |
| *NT5E* | rs7429191 | G | A | 0.7072 | 0.054 | 0.007 | 5.0119E-14 | 59.51 |
| *NT5E* | rs13107325 | C | T | 0.0749 | -0.094 | 0.013 | 7.9433E-14 | 52.28 |
| *NT5E* | rs4835265 | C | A | 0.1563 | 0.118 | 0.009 | 1E-38 | 171.90 |
| *NT5E* | rs10075805 | A | G | 0.2755 | -0.055 | 0.007 | 7.9433E-14 | 61.73 |
| *NT5E* | rs2229523 | A | G | 0.6597 | -0.587 | 0.008 | 1E-200 | 5383.89 |
| *NT5E* | rs754466 | A | T | 0.2533 | 0.051 | 0.008 | 1.5849E-11 | 40.64 |
| *NT5E* | rs550999044 | A | AT | 0.2435 | 0.114 | 0.008 | 1.2589E-49 | 203.06 |
| *NT5E* | rs12373325 | T | C | 0.798 | -0.078 | 0.008 | 1.5849E-21 | 95.06 |
| *NT5E* | rs601338 | G | A | 0.5093 | 0.054 | 0.007 | 2.5119E-16 | 59.51 |
| *NT5E* | rs189448562 | G | C | 0.0112 | 0.324 | 0.032 | 1.5849E-24 | 102.52 |
| *NT5E* | rs6076335 | T | G | 0.481 | 0.05 | 0.007 | 2.5119E-14 | 51.02 |
| *NTF3* | rs13402475 | C | G | 0.8167 | -0.086 | 0.01 | 2.5119E-18 | 73.96 |
| *NTF3* | rs10868235 | C | T | 0.526 | -0.085 | 0.008 | 6.3096E-30 | 112.89 |
| *NTF3* | rs73039984 | C | T | 0.1125 | -0.232 | 0.012 | 1.5849E-83 | 373.78 |
| *NTF3* | rs117126605 | T | C | 0.0176 | -0.356 | 0.029 | 1.5849E-35 | 150.70 |
| *NTF3* | rs12454720 | G | T | 0.5294 | 0.087 | 0.008 | 1.5849E-30 | 118.27 |
| *NTF4* | rs6562775 | A | G | 0.4962 | -0.053 | 0.008 | 5.0119E-12 | 43.89 |
| *NTproBNP* | rs198379 | T | C | 0.413 | 0.261 | 0.007 | 1E-200 | 1390.22 |
| *NTproBNP* | rs3800560 | C | T | 0.0847 | 0.087 | 0.012 | 1.2589E-12 | 52.56 |
| *NTproBNP* | rs72671655 | T | A | 0.038 | 0.144 | 0.018 | 7.9433E-16 | 64.00 |
| *NTproBNP* | rs2234962 | T | C | 0.2155 | 0.087 | 0.008 | 1.5849E-25 | 118.27 |
| *NTproBNP* | rs17064679 | T | C | 0.3937 | -0.048 | 0.007 | 7.9433E-12 | 47.02 |
| *NTRK2* | rs4665972 | T | C | 0.6082 | 0.058 | 0.008 | 1.5849E-14 | 52.56 |
| *NTRK2* | rs13107325 | C | T | 0.0751 | 0.099 | 0.014 | 1.9953E-12 | 50.01 |
| *NTRK2* | rs77562929 | G | T | 0.0071 | 0.495 | 0.046 | 1.9953E-26 | 115.80 |

| | | | | | | | | |
|---|---|---|---|---|---|---|---|---|
| *NTRK2* | rs56278466 | T | G | 0.6615 | 0.084 | 0.008 | 3.1623E-27 | 110.25 |
| *NTRK2* | rs2298475 | T | C | 0.0752 | -0.101 | 0.014 | 2.5119E-13 | 52.05 |
| *NTRK2* | rs186021206 | G | A | 0.0064 | 0.516 | 0.05 | 7.9433E-25 | 106.50 |
| *NTRK2* | rs2008174 | C | T | 0.7395 | -0.068 | 0.009 | 1.5849E-15 | 57.09 |
| *NTRK3* | rs34324830 | C | T | 0.1141 | -0.098 | 0.011 | 1E-17 | 79.37 |
| *NTRK3* | rs7340151 | C | T | 0.6719 | 0.056 | 0.008 | 7.9433E-13 | 49.00 |
| *NTRK3* | rs35887873 | T | C | 0.2283 | 0.059 | 0.009 | 5.0119E-12 | 42.98 |
| *NTRK3* | rs111338191 | A | AAATT | 0.5209 | 0.08 | 0.007 | 1E-27 | 130.61 |
| *NTRK3* | rs79445108 | G | A | 0.0683 | -0.16 | 0.015 | 6.3096E-28 | 113.78 |
| *NTRK3* | rs28735437 | G | C | 0.1398 | 0.273 | 0.011 | 7.943E-146 | 615.94 |
| *NTRK3* | rs200489612 | G | A | 0.0052 | 0.524 | 0.054 | 5.0119E-22 | 94.16 |
| *NTRK3* | rs914187 | T | C | 0.5998 | -0.112 | 0.007 | 5.0119E-51 | 256.00 |
| *NTRK3* | rs2413593 | A | G | 0.4946 | 0.054 | 0.007 | 2.5119E-13 | 59.51 |
| *NUB1* | rs1354034 | T | C | 0.6043 | 0.084 | 0.008 | 3.1623E-28 | 110.25 |
| *NUB1* | rs342298 | C | T | 0.4546 | -0.057 | 0.008 | 3.9811E-14 | 50.77 |
| *NUB1* | rs113920358 | C | T | 0.129 | -0.266 | 0.012 | 1.259E-116 | 491.36 |
| *NUB1* | rs11502185 | T | C | 0.2598 | 0.072 | 0.01 | 3.9811E-14 | 51.84 |
| *NUBP1* | rs71534539 | G | A | 0.4645 | -0.084 | 0.008 | 1.5849E-23 | 110.25 |
| *NUCB2* | rs2524096 | G | T | 0.42 | 0.051 | 0.007 | 3.1623E-12 | 53.08 |
| *NUCB2* | rs6993770 | A | T | 0.2863 | -0.077 | 0.008 | 1.5849E-22 | 92.64 |
| *NUCB2* | rs214068 | T | C | 0.5769 | -0.406 | 0.008 | 1E-200 | 2575.56 |
| *NUCB2* | rs111338191 | A | AAATT | 0.5206 | -0.053 | 0.007 | 2.5119E-13 | 57.33 |
| *NUCB2* | rs892090 | T | G | 0.8342 | 0.082 | 0.01 | 3.1623E-17 | 67.24 |
| *NUDC* | rs1354034 | T | C | 0.6041 | 0.079 | 0.008 | 1.5849E-25 | 97.52 |
| *NUDC* | rs342298 | C | T | 0.4547 | -0.063 | 0.007 | 2.5119E-17 | 81.00 |
| *NUDT16* | rs1354034 | T | C | 0.6047 | 0.065 | 0.008 | 2.5119E-17 | 66.02 |
| *NUDT16* | rs150437674 | C | G | 0.0113 | -1.211 | 0.036 | 1E-200 | 1131.57 |
| *NUDT16* | rs10733789 | T | C | 0.3128 | 0.069 | 0.008 | 1.5849E-17 | 74.39 |
| *NUDT2* | rs7848667 | A | G | 0.1569 | 0.332 | 0.01 | 1E-200 | 1102.24 |
| *NUDT5* | rs1354034 | T | C | 0.6041 | 0.064 | 0.008 | 3.1623E-17 | 64.00 |
| *NUDT5* | rs10508438 | G | A | 0.3356 | 0.061 | 0.008 | 1E-14 | 58.14 |
| *NUDT5* | rs60822569 | T | TTTCTC | 0.5545 | 0.059 | 0.008 | 3.9811E-15 | 54.39 |
| *NUMB* | rs1354034 | T | C | 0.6039 | 0.081 | 0.008 | 2.5119E-26 | 102.52 |
| *NUMB* | rs6993770 | A | T | 0.2866 | -0.058 | 0.008 | 3.9811E-12 | 52.56 |
| *NUMB* | rs11502185 | T | C | 0.2597 | 0.07 | 0.01 | 2.5119E-13 | 49.00 |
| *NUMB* | rs7141139 | G | T | 0.7109 | 0.084 | 0.008 | 7.9433E-24 | 110.25 |
| *NXPH3* | rs60191984 | T | TTGGG | 0.4012 | -0.068 | 0.008 | 3.9811E-19 | 72.25 |
| *NXPH3* | rs39602 | C | G | 0.5555 | -0.086 | 0.007 | 5.0119E-31 | 150.94 |
| *NXPH3* | rs1558918 | A | T | 0.6718 | -0.062 | 0.008 | 1.2589E-15 | 60.06 |
| *NXPH3* | rs35887873 | T | C | 0.2282 | 0.078 | 0.009 | 5.0119E-19 | 75.11 |
| *NXPH3* | rs215225 | A | G | 0.4283 | 0.057 | 0.007 | 1.9953E-14 | 66.31 |
| *NXPH3* | rs966541 | A | G | 0.2758 | 0.255 | 0.008 | 1E-200 | 1016.02 |
| *NXPH3* | rs186021206 | G | A | 0.0063 | 0.574 | 0.051 | 3.9811E-29 | 126.67 |
| *NXPH3* | rs147028083 | C | G | 0.0071 | -2.1 | 0.045 | 1E-200 | 2177.78 |
| *OBP2B* | rs144623173 | T | TA | 0.4122 | -0.054 | 0.005 | 3.9811E-23 | 116.64 |
| *OBP2B* | rs61685000 | G | C | 0.4105 | -0.037 | 0.005 | 7.9433E-12 | 54.76 |
| *OBP2B* | rs12621139 | G | A | 0.1825 | 0.07 | 0.007 | 6.3096E-24 | 100.00 |
| *OBP2B* | rs463725 | G | C | 0.4243 | 0.036 | 0.005 | 1.5849E-11 | 51.84 |
| *OBP2B* | rs34320230 | C | T | 0.5353 | -0.06 | 0.005 | 3.1623E-29 | 144.00 |
| *OBP2B* | rs1825146 | A | C | 0.5729 | -0.048 | 0.005 | 2.5119E-19 | 92.16 |
| *OBP2B* | rs4962104 | T | C | 0.8001 | 0.704 | 0.008 | 1E-200 | 7744.00 |
| *OBP2B* | rs11516994 | C | A | 0.3558 | 0.042 | 0.006 | 3.1623E-14 | 49.00 |
| *OBP2B* | rs7305932 | T | A | 0.7794 | -0.046 | 0.006 | 1E-12 | 58.78 |
| *OBP2B* | rs9543287 | C | G | 0.3229 | -0.047 | 0.006 | 3.1623E-17 | 61.36 |
| *OBP2B* | rs8047587 | G | T | 0.4381 | -0.04 | 0.005 | 3.9811E-14 | 64.00 |
| *OBP2B* | rs12986064 | T | C | 0.5076 | -0.048 | 0.005 | 7.9433E-19 | 92.16 |
| *OBP2B* | rs11911496 | A | G | 0.4856 | -0.04 | 0.005 | 1.9953E-14 | 64.00 |
| *OBP2B* | rs1978060 | A | G | 0.6144 | 0.04 | 0.005 | 1.5849E-13 | 64.00 |
| *OCLN* | rs2583707 | T | C | 0.024 | -0.473 | 0.024 | 5.0119E-86 | 388.42 |
| *ODAM* | rs76153614 | T | C | 0.0523 | -0.587 | 0.017 | 1E-200 | 1192.28 |
| *OGA* | rs342293 | C | G | 0.4598 | -0.052 | 0.007 | 2.5119E-12 | 55.18 |
| *OGA* | rs6993770 | A | T | 0.2866 | -0.07 | 0.008 | 1E-17 | 76.56 |
| *OGA* | rs3781299 | C | A | 0.2914 | -0.134 | 0.008 | 1.5849E-60 | 280.56 |
| *OGFR* | rs6993770 | A | T | 0.2864 | -0.055 | 0.008 | 1E-11 | 47.27 |
| *OGFR* | rs200108407 | G | A | 0.0013 | -1.697 | 0.103 | 7.9433E-61 | 271.45 |
| *OGN* | rs13146355 | G | A | 0.4566 | 0.043 | 0.006 | 3.9811E-12 | 51.36 |
| *OGN* | rs7026361 | G | T | 0.3689 | 0.597 | 0.007 | 1E-200 | 7273.65 |
| *OGN* | rs145678014 | G | T | 0.0447 | -0.112 | 0.015 | 2.5119E-13 | 55.75 |

| | | | | | | | | |
|---|---|---|---|---|---|---|---|---|
| *OGN* | rs113878851 | C | CT | 0.1717 | -0.063 | 0.009 | 1E-13 | 49.00 |
| *OLFM4* | rs28498283 | A | T | 0.255 | 0.074 | 0.008 | 7.9433E-21 | 85.56 |
| *OLFM4* | rs72606404 | G | C | 0.1841 | -0.113 | 0.009 | 6.3096E-38 | 157.64 |
| *OLFM4* | rs1870026 | G | A | 0.0604 | 0.096 | 0.014 | 1.2589E-11 | 47.02 |
| *OLFM4* | rs457420 | C | T | 0.386 | 0.063 | 0.007 | 1.5849E-19 | 81.00 |
| *OLFM4* | rs4917766 | G | A | 0.5969 | 0.069 | 0.007 | 1.2589E-23 | 97.16 |
| *OLFM4* | rs17552047 | G | A | 0.2764 | -0.859 | 0.009 | 1E-200 | 9109.64 |
| *OLFM4* | rs1045929 | C | T | 0.3879 | 0.057 | 0.007 | 3.1623E-16 | 66.31 |
| *OLFM4* | rs138284624 | C | T | 0.0132 | 0.248 | 0.03 | 7.9433E-17 | 68.34 |
| *OLFM4* | rs10420726 | C | T | 0.2268 | 0.121 | 0.008 | 2.5119E-50 | 228.77 |
| *OLFM4* | rs113080783 | A | G | 0.0276 | 0.312 | 0.021 | 1.2589E-48 | 220.73 |
| *OLFM4* | rs9305635 | C | G | 0.3313 | -0.054 | 0.007 | 3.9811E-14 | 59.51 |
| *OLFM4* | rs577625804 | C | CT | 0.812 | -0.089 | 0.009 | 1.9953E-24 | 97.79 |
| *OLR1* | rs61740470 | C | T | 0.0103 | 0.281 | 0.037 | 2.5119E-14 | 57.68 |
| *OLR1* | rs2871960 | A | C | 0.4453 | -0.103 | 0.007 | 1.9953E-43 | 216.51 |
| *OLR1* | rs45489595 | C | T | 0.1133 | -0.162 | 0.012 | 3.9811E-43 | 182.25 |
| *OLR1* | rs2511241 | C | T | 0.9296 | 0.2 | 0.015 | 7.9433E-43 | 177.78 |
| *OLR1* | rs587862 | A | G | 0.2695 | 0.059 | 0.008 | 3.9811E-12 | 54.39 |
| *OLR1* | rs936964 | C | A | 0.2751 | 0.108 | 0.009 | 6.3096E-37 | 144.00 |
| *OLR1* | rs11871747 | T | C | 0.3936 | 0.067 | 0.008 | 1E-18 | 70.14 |
| *OMD* | rs13303 | T | C | 0.5597 | 0.097 | 0.007 | 3.1623E-41 | 192.02 |
| *OMD* | rs4253281 | G | A | 0.5131 | -0.061 | 0.007 | 3.1623E-17 | 75.94 |
| *OMD* | rs35209758 | A | AT | 0.3088 | 0.08 | 0.008 | 1E-23 | 100.00 |
| *OMD* | rs117801489 | T | C | 0.017 | -0.443 | 0.028 | 7.9433E-58 | 250.32 |
| *OMD* | rs548589580 | T | TA | 0.4838 | 0.064 | 0.007 | 6.3096E-19 | 83.59 |
| *OMD* | rs704 | G | A | 0.4733 | -0.088 | 0.007 | 1.2589E-34 | 158.04 |
| *OMD* | rs738408 | C | T | 0.217 | 0.062 | 0.009 | 6.3096E-13 | 47.46 |
| *OMG* | rs112168180 | C | T | 0.0278 | -0.163 | 0.022 | 1.9953E-13 | 54.89 |
| *OMG* | rs6457541 | G | C | 0.6576 | -0.061 | 0.008 | 1.5849E-15 | 58.14 |
| *OMG* | rs3757724 | C | T | 0.0613 | 0.182 | 0.015 | 1.5849E-32 | 147.22 |
| *OMG* | rs35107030 | C | G | 0.0437 | 0.18 | 0.018 | 1.9953E-23 | 100.00 |
| *OMG* | rs56278466 | T | G | 0.6611 | 0.115 | 0.008 | 1E-50 | 206.64 |
| *OMG* | rs200489612 | G | A | 0.0052 | 0.532 | 0.055 | 5.0119E-22 | 93.56 |
| *OMG* | rs72813607 | G | A | 0.1402 | -0.125 | 0.011 | 1.5849E-32 | 129.13 |
| *OMG* | rs17196752 | C | T | 0.189 | -0.069 | 0.009 | 1.9953E-13 | 58.78 |
| *OMP* | rs2233546 | C | T | 0.0531 | -0.387 | 0.017 | 3.162E-110 | 518.23 |
| *OPHN1* | rs1354034 | T | C | 0.6043 | 0.062 | 0.008 | 1.9953E-16 | 60.06 |
| *OPLAH* | rs6993770 | A | T | 0.2866 | -0.087 | 0.008 | 3.1623E-25 | 118.27 |
| *OPLAH* | rs55646585 | C | T | 0.4142 | 0.187 | 0.008 | 1.585E-131 | 546.39 |
| *OPTC* | rs139024490 | T | C | 0.0044 | -1.725 | 0.054 | 1E-200 | 1020.45 |
| *OPTC* | rs78011579 | G | A | 0.0312 | -0.243 | 0.022 | 3.9811E-28 | 122.00 |
| *OPTC* | rs67449961 | G | C | 0.8184 | 0.089 | 0.01 | 1.5849E-19 | 79.21 |
| *OPTC* | rs2108790 | C | T | 0.607 | 0.052 | 0.007 | 1.2589E-12 | 55.18 |
| *OPTC* | rs55851518 | G | A | 0.0561 | -0.158 | 0.016 | 2.5119E-24 | 97.52 |
| *OPTC* | rs10750291 | G | A | 0.3247 | -0.073 | 0.008 | 3.1623E-21 | 83.27 |
| *OPTC* | rs4436712 | G | T | 0.3853 | 0.06 | 0.007 | 3.9811E-16 | 73.47 |
| *OPTC* | rs35731193 | G | GA | 0.4446 | -0.054 | 0.008 | 7.9433E-13 | 45.56 |
| *OPTC* | rs186021206 | G | A | 0.0063 | 0.657 | 0.05 | 5.0119E-40 | 172.66 |
| *ORM1* | rs2017857 | C | G | 0.7486 | 0.059 | 0.009 | 6.3096E-12 | 42.98 |
| *ORM1* | rs10982156 | T | A | 0.0693 | 0.741 | 0.016 | 1E-200 | 2144.85 |
| *ORM1* | rs7896518 | A | G | 0.4236 | -0.059 | 0.007 | 3.1623E-15 | 71.04 |
| *ORM1* | rs11054386 | A | T | 0.57 | -0.052 | 0.007 | 1.9953E-12 | 55.18 |
| *OSCAR* | rs13063578 | T | A | 0.3981 | -0.045 | 0.007 | 7.9433E-12 | 41.33 |
| *OSCAR* | rs73401939 | A | G | 0.0057 | 0.618 | 0.043 | 2.5119E-46 | 206.56 |
| *OSCAR* | rs12544501 | T | C | 0.5686 | -0.06 | 0.007 | 5.0119E-20 | 73.47 |
| *OSCAR* | rs7137828 | C | T | 0.5162 | -0.082 | 0.006 | 1.2589E-37 | 186.78 |
| *OSCAR* | rs76428106 | T | C | 0.0129 | 0.348 | 0.03 | 7.9433E-32 | 134.56 |
| *OSCAR* | rs186021206 | G | A | 0.0064 | 0.624 | 0.044 | 5.0119E-45 | 201.12 |
| *OSCAR* | rs2305482 | A | C | 0.5395 | -0.044 | 0.006 | 6.3096E-12 | 53.78 |
| *OSCAR* | rs2659005 | C | T | 0.4407 | 0.044 | 0.007 | 1.2589E-11 | 39.51 |
| *OSCAR* | rs4442925 | C | T | 0.4488 | 0.521 | 0.008 | 1E-200 | 4241.27 |
| *OSM* | rs28498283 | A | T | 0.2552 | 0.066 | 0.009 | 3.1623E-14 | 53.78 |
| *OSM* | rs9325122 | C | T | 0.5521 | -0.051 | 0.007 | 7.9433E-12 | 53.08 |
| *OSM* | rs12264390 | T | C | 0.2401 | 0.06 | 0.009 | 3.9811E-12 | 44.44 |
| *OSM* | rs2511241 | C | T | 0.9294 | 0.362 | 0.015 | 3.981E-136 | 582.42 |
| *OSM* | rs7310615 | C | G | 0.5175 | -0.065 | 0.007 | 1.9953E-18 | 86.22 |
| *OSM* | rs10483987 | T | C | 0.1531 | -0.079 | 0.01 | 3.9811E-14 | 62.41 |
| *OSM* | rs11871747 | T | C | 0.3934 | 0.127 | 0.008 | 1.5849E-62 | 252.02 |

| | | | | | | | | |
|---|---|---|---|---|---|---|---|---|
| *OSM* | rs4823082 | G | C | 0.0229 | -0.188 | 0.025 | 3.1623E-14 | 56.55 |
| *OSMR* | rs2228145 | A | C | 0.4057 | -0.05 | 0.006 | 1.5849E-14 | 69.44 |
| *OSMR* | rs9292723 | C | T | 0.2767 | -0.596 | 0.008 | 1E-200 | 5550.25 |
| *OSMR* | rs8176693 | C | T | 0.0614 | 0.1 | 0.013 | 3.9811E-14 | 59.17 |
| *OSMR* | rs776516693 | TTCTG | T | 0.134 | -0.081 | 0.009 | 5.0119E-18 | 81.00 |
| *OSMR* | rs56187480 | G | A | 0.3428 | 0.063 | 0.007 | 5.0119E-21 | 81.00 |
| *OSMR* | rs186021206 | G | A | 0.0064 | 0.595 | 0.044 | 3.1623E-42 | 182.86 |
| *OSTN* | rs34711532 | C | CG | 0.7291 | -0.106 | 0.009 | 1E-33 | 138.72 |
| *OTOA* | rs8055493 | A | G | 0.4255 | -0.057 | 0.008 | 3.9811E-13 | 50.77 |
| *OTUD6B* | rs1354034 | T | C | 0.6046 | 0.066 | 0.008 | 1.9953E-18 | 68.06 |
| *OTUD6B* | rs342298 | C | T | 0.455 | -0.051 | 0.007 | 1.5849E-11 | 53.08 |
| *OTUD7B* | rs1354034 | T | C | 0.6043 | 0.075 | 0.008 | 1.2589E-22 | 87.89 |
| *OTUD7B* | rs6993770 | A | T | 0.2864 | -0.065 | 0.008 | 3.1623E-15 | 66.02 |
| *OXCT1* | rs10075801 | A | G | 0.4879 | -0.052 | 0.008 | 1.2589E-11 | 42.25 |
| *OXCT1* | rs342298 | C | T | 0.4548 | -0.059 | 0.008 | 7.9433E-15 | 54.39 |
| *OXCT1* | rs6993770 | A | T | 0.2866 | -0.064 | 0.008 | 3.1623E-14 | 64.00 |
| *OXCT1* | rs7896518 | A | G | 0.4236 | 0.054 | 0.008 | 6.3096E-12 | 45.56 |
| *OXT* | rs10929757 | A | C | 0.5758 | 0.056 | 0.007 | 2.5119E-15 | 64.00 |
| *OXT* | rs11127048 | G | A | 0.6181 | -0.056 | 0.007 | 1.2589E-14 | 64.00 |
| *OXT* | rs4895864 | C | T | 0.5742 | 0.049 | 0.007 | 2.5119E-12 | 49.00 |
| *OXT* | rs13229619 | G | A | 0.1279 | -0.098 | 0.01 | 5.0119E-21 | 96.04 |
| *OXT* | rs45446698 | T | G | 0.042 | -0.13 | 0.017 | 5.0119E-14 | 58.48 |
| *OXT* | rs112875651 | G | A | 0.391 | -0.067 | 0.007 | 2.5119E-20 | 91.61 |
| *OXT* | rs9414801 | G | A | 0.5331 | -0.047 | 0.007 | 1.2589E-11 | 45.08 |
| *OXT* | rs4782568 | C | G | 0.4508 | 0.056 | 0.007 | 6.3096E-16 | 64.00 |
| *OXT* | rs877172 | T | G | 0.3124 | 0.627 | 0.009 | 1E-200 | 4853.44 |
| *OXT* | rs1800961 | C | T | 0.0322 | -0.142 | 0.021 | 1.2589E-11 | 45.72 |
| *P4HB* | rs3910516 | A | G | 0.7369 | 0.065 | 0.009 | 3.1623E-14 | 52.16 |
| *P4HB* | rs1354034 | T | C | 0.6041 | 0.058 | 0.008 | 5.0119E-14 | 52.56 |
| *P4HB* | rs6993770 | A | T | 0.2861 | -0.058 | 0.008 | 3.1623E-12 | 52.56 |
| *P4HB* | rs112635299 | G | T | 0.0214 | 0.191 | 0.026 | 1.9953E-13 | 53.97 |
| *P4HB* | rs113708033 | G | T | 0.1918 | 0.087 | 0.01 | 1.5849E-19 | 75.69 |
| *PACS2* | rs13412535 | G | A | 0.2313 | 0.085 | 0.009 | 1.9953E-21 | 89.20 |
| *PACS2* | rs1354034 | T | C | 0.6037 | 0.076 | 0.008 | 5.0119E-23 | 90.25 |
| *PACS2* | rs7149013 | T | C | 0.2945 | 0.178 | 0.008 | 7.943E-103 | 495.06 |
| *PADI2* | rs2235913 | G | A | 0.6175 | 0.122 | 0.008 | 1E-53 | 232.56 |
| *PADI4* | rs2240336 | C | T | 0.4145 | -0.257 | 0.008 | 1E-200 | 1032.02 |
| *PAEP* | rs9409964 | G | A | 0.1878 | 0.871 | 0.009 | 1E-200 | 9365.94 |
| *PAEP* | rs4805162 | A | G | 0.5236 | -0.047 | 0.006 | 1.9953E-17 | 61.36 |
| *PAFAH1B3* | rs71534539 | G | A | 0.4643 | -0.137 | 0.008 | 7.9433E-60 | 293.27 |
| *PAFAH1B3* | rs10957979 | A | G | 0.6484 | 0.075 | 0.008 | 3.9811E-21 | 87.89 |
| *PAFAH2* | rs150853838 | G | A | 0.004 | -0.653 | 0.064 | 1.2589E-24 | 104.10 |
| *PAG1* | rs1354034 | T | C | 0.6038 | 0.054 | 0.008 | 3.9811E-12 | 45.56 |
| *PAG1* | rs4436084 | T | C | 0.3499 | -0.078 | 0.008 | 1.5849E-22 | 95.06 |
| *PAG1* | rs7310615 | C | G | 0.5176 | -0.076 | 0.008 | 1.2589E-23 | 90.25 |
| *PAG1* | rs1532085 | A | G | 0.6171 | 0.059 | 0.008 | 1E-13 | 54.39 |
| *PAG1* | rs247617 | C | A | 0.3247 | -0.081 | 0.008 | 5.0119E-23 | 102.52 |
| *PAG1* | rs7208422 | A | T | 0.4775 | -0.08 | 0.008 | 1.2589E-25 | 100.00 |
| *PAGR1* | rs13257521 | C | G | 0.2105 | 0.093 | 0.009 | 2.5119E-23 | 106.78 |
| *PAK4* | rs11170874 | A | G | 0.331 | 0.059 | 0.008 | 6.3096E-13 | 54.39 |
| *PALM* | rs150394890 | G | T | 0.0086 | 0.288 | 0.04 | 5.0119E-13 | 51.84 |
| *PALM* | rs12610821 | C | G | 0.0567 | 0.319 | 0.016 | 6.3096E-88 | 397.50 |
| *PALM* | rs429358 | T | C | 0.1555 | 0.197 | 0.01 | 1E-83 | 388.09 |
| *PALM2* | rs1023005 | C | T | 0.2305 | 0.234 | 0.009 | 1.995E-161 | 676.00 |
| *PALM3* | rs607929 | C | G | 0.449 | -0.084 | 0.008 | 3.1623E-27 | 110.25 |
| *PAM* | rs149802978 | C | G | 0.0539 | -1.099 | 0.017 | 1E-200 | 4179.24 |
| *PAM* | rs75901433 | A | G | 0.558 | 0.051 | 0.007 | 6.3096E-14 | 53.08 |
| *PAM* | rs56278466 | T | G | 0.6616 | 0.108 | 0.007 | 1.2589E-53 | 238.04 |
| *PAM* | rs12975366 | T | C | 0.3952 | -0.073 | 0.007 | 6.3096E-27 | 108.76 |
| *PAMR1* | rs1260326 | T | C | 0.6084 | -0.059 | 0.007 | 1E-17 | 71.04 |
| *PAMR1* | rs1150753 | A | G | 0.1311 | -0.222 | 0.01 | 1.995E-106 | 492.84 |
| *PAMR1* | rs61736408 | T | C | 0.1567 | -0.476 | 0.01 | 1E-200 | 2265.76 |
| *PAMR1* | rs4055121 | C | T | 0.1351 | -0.073 | 0.011 | 6.3096E-12 | 44.04 |
| *PAPPA* | rs967968 | C | G | 0.5114 | 0.077 | 0.007 | 1.9953E-26 | 121.00 |
| *PAPPA* | rs4328821 | A | G | 0.1067 | -0.119 | 0.012 | 1.5849E-24 | 98.34 |
| *PAPPA* | rs13107325 | C | T | 0.0747 | 0.103 | 0.014 | 6.3096E-14 | 54.13 |
| *PAPPA* | rs7808353 | A | T | 0.1668 | -0.084 | 0.01 | 6.3096E-18 | 70.56 |
| *PAPPA* | rs117448041 | T | C | 0.0196 | 0.187 | 0.026 | 1.2589E-12 | 51.73 |

| | | | | | | | | |
|---|---|---|---|---|---|---|---|---|
| *PAPPA* | rs13448 | T | C | 0.2653 | -0.058 | 0.008 | 2.5119E-12 | 52.56 |
| *PAPPA* | rs147496186 | G | C | 0.0418 | -0.129 | 0.018 | 1.2589E-12 | 51.36 |
| *PAPPA* | rs550729 | T | C | 0.729 | 0.263 | 0.008 | 1E-200 | 1080.77 |
| *PAPPA* | rs74035509 | C | T | 0.0778 | 0.107 | 0.014 | 1.9953E-15 | 58.41 |
| *PAPPA* | rs35617692 | C | A | 0.0111 | 0.299 | 0.034 | 3.9811E-18 | 77.34 |
| *PARD3* | rs9418120 | G | A | 0.2605 | 0.076 | 0.009 | 2.5119E-18 | 71.31 |
| *PARK7* | rs3766606 | G | T | 0.1739 | 0.209 | 0.01 | 2.512E-100 | 436.81 |
| *PARK7* | rs1354034 | T | C | 0.6041 | 0.057 | 0.008 | 2.5119E-14 | 50.77 |
| *PARK7* | rs11170874 | A | G | 0.3311 | 0.055 | 0.008 | 6.3096E-12 | 47.27 |
| *PARP1* | rs1433574 | A | C | 0.1607 | -0.159 | 0.01 | 1.2589E-55 | 252.81 |
| *PARP1* | rs4632248 | G | T | 0.2094 | 0.149 | 0.009 | 3.9811E-60 | 274.09 |
| *PAXX* | rs6993770 | A | T | 0.287 | -0.059 | 0.008 | 1E-12 | 54.39 |
| *PAXX* | rs10774624 | G | A | 0.5125 | -0.052 | 0.008 | 5.0119E-12 | 42.25 |
| *PBLD* | rs61854835 | A | G | 0.2369 | 0.576 | 0.009 | 1E-200 | 4096.00 |
| *PBXIP1* | rs390082 | T | G | 0.1096 | 0.081 | 0.012 | 7.9433E-12 | 45.56 |
| *PCARE* | rs141575023 | C | T | 0.1383 | 0.234 | 0.011 | 3.9811E-95 | 452.53 |
| *PCBD1* | rs7041363 | C | G | 0.4899 | -0.06 | 0.007 | 7.9433E-16 | 73.47 |
| *PCBD1* | rs2630336 | G | T | 0.3602 | -0.181 | 0.008 | 3.981E-121 | 511.89 |
| *PCBD1* | rs10883451 | T | C | 0.5018 | -0.053 | 0.007 | 6.3096E-13 | 57.33 |
| *PCBD1* | rs59581534 | T | A | 0.0406 | 0.26 | 0.018 | 5.0119E-45 | 208.64 |
| *PCBD1* | rs217184 | T | C | 0.1963 | -0.073 | 0.009 | 2.5119E-15 | 65.79 |
| *PCBD1* | rs58542926 | C | T | 0.0755 | 0.098 | 0.014 | 1.9953E-12 | 49.00 |
| *PCBD1* | rs3747207 | G | A | 0.2151 | 0.093 | 0.009 | 1.2589E-25 | 106.78 |
| *PCBP2* | rs1354034 | T | C | 0.6043 | 0.057 | 0.008 | 6.3096E-14 | 50.77 |
| *PCDH1* | rs4645358 | C | T | 0.5747 | 0.081 | 0.007 | 1E-27 | 133.90 |
| *PCDH1* | rs4077811 | A | G | 0.1444 | 0.074 | 0.01 | 1E-12 | 54.76 |
| *PCDH1* | rs8176741 | G | A | 0.0612 | 0.231 | 0.015 | 3.9811E-51 | 237.16 |
| *PCDH1* | rs56278466 | T | G | 0.6613 | 0.054 | 0.008 | 3.1623E-12 | 45.56 |
| *PCDH1* | rs34937091 | G | GT | 0.3571 | 0.054 | 0.008 | 3.9811E-12 | 45.56 |
| *PCDH1* | rs58534292 | T | C | 0.6497 | -0.125 | 0.008 | 7.9433E-58 | 244.14 |
| *PCDH1* | rs2005652 | T | C | 0.6307 | -0.131 | 0.008 | 3.1623E-63 | 268.14 |
| *PCDH12* | rs11927405 | C | T | 0.4406 | -0.096 | 0.007 | 3.1623E-39 | 188.08 |
| *PCDH12* | rs73186030 | C | T | 0.1275 | 0.121 | 0.011 | 3.9811E-29 | 121.00 |
| *PCDH12* | rs164085 | G | A | 0.5097 | 0.327 | 0.008 | 1E-200 | 1670.77 |
| *PCDH12* | rs2519093 | C | T | 0.1843 | -0.271 | 0.009 | 2.512E-182 | 906.68 |
| *PCDH12* | rs186021206 | G | A | 0.0064 | 0.361 | 0.049 | 2.5119E-13 | 54.28 |
| *PCDH12* | rs1135062 | A | G | 0.2913 | -0.066 | 0.008 | 5.0119E-17 | 68.06 |
| *PCDH17* | rs6789547 | C | T | 0.1731 | -0.064 | 0.009 | 3.9811E-12 | 50.57 |
| *PCDH17* | rs35467127 | C | T | 0.2007 | 0.068 | 0.009 | 1E-14 | 57.09 |
| *PCDH17* | rs8176749 | C | T | 0.0611 | 0.649 | 0.015 | 1E-200 | 1872.00 |
| *PCDH17* | rs56278466 | T | G | 0.6615 | 0.112 | 0.007 | 1E-52 | 256.00 |
| *PCDH17* | rs11603123 | G | A | 0.0336 | 0.169 | 0.019 | 1.2589E-18 | 79.12 |
| *PCDH17* | rs3184504 | T | C | 0.5174 | -0.082 | 0.007 | 3.9811E-32 | 137.22 |
| *PCDH17* | rs11148472 | A | C | 0.3913 | 0.353 | 0.008 | 1E-200 | 1947.02 |
| *PCDH17* | rs186021206 | G | A | 0.0064 | 0.357 | 0.048 | 5.0119E-14 | 55.32 |
| *PCDH17* | rs5757683 | G | A | 0.7455 | -0.073 | 0.008 | 6.3096E-20 | 83.27 |
| *PCDH7* | rs646776 | C | T | 0.7778 | 0.099 | 0.009 | 2.5119E-28 | 121.00 |
| *PCDH7* | rs1374657 | C | T | 0.2835 | -0.116 | 0.008 | 1E-44 | 210.25 |
| *PCDH7* | rs61000956 | T | C | 0.482 | 0.06 | 0.008 | 5.0119E-15 | 56.25 |
| *PCDH7* | rs2731672 | T | C | 0.7438 | 0.066 | 0.008 | 1E-14 | 68.06 |
| *PCDH7* | rs445925 | G | A | 0.1095 | 0.09 | 0.012 | 3.9811E-14 | 56.25 |
| *PCDH9* | rs9874262 | C | T | 0.4828 | 0.073 | 0.007 | 2.5119E-24 | 108.76 |
| *PCDH9* | rs28601761 | C | G | 0.419 | -0.051 | 0.007 | 2.5119E-12 | 53.08 |
| *PCDH9* | rs10984506 | A | T | 0.3586 | -0.054 | 0.007 | 2.5119E-13 | 59.51 |
| *PCDH9* | rs111338191 | A | AAATT | 0.521 | -0.055 | 0.007 | 1.5849E-14 | 61.73 |
| *PCDH9* | rs7310409 | A | G | 0.6236 | 0.079 | 0.007 | 5.0119E-27 | 127.37 |
| *PCDH9* | rs73196184 | G | A | 0.1135 | 0.341 | 0.012 | 1.259E-185 | 807.51 |
| *PCDH9* | rs186021206 | G | A | 0.0065 | 0.655 | 0.049 | 3.1623E-41 | 178.69 |
| *PCDH9* | rs28745910 | TA | T | 0.4986 | -0.051 | 0.007 | 1.5849E-12 | 53.08 |
| *PCDHB15* | rs1622822 | T | C | 0.8551 | -0.103 | 0.01 | 1.9953E-24 | 106.09 |
| *PCDHB15* | rs13088318 | A | G | 0.3385 | -0.051 | 0.007 | 7.9433E-12 | 53.08 |
| *PCDHB15* | rs2740584 | C | T | 0.2339 | -0.409 | 0.009 | 1E-200 | 2065.20 |
| *PCDHB15* | rs28367646 | A | C | 0.2403 | 0.061 | 0.009 | 3.9811E-12 | 45.94 |
| *PCDHB15* | rs11599284 | G | A | 0.1267 | 0.115 | 0.011 | 1.2589E-25 | 109.30 |
| *PCDHB15* | rs260416 | A | G | 0.1019 | 0.081 | 0.012 | 2.5119E-12 | 45.56 |
| *PCNA* | rs371337007 | G | A | 0.6457 | -0.062 | 0.009 | 1.5849E-11 | 47.46 |
| *PCOLCE* | rs9801017 | G | A | 0.6237 | 0.172 | 0.012 | 6.3096E-47 | 205.44 |
| *PCSK7* | rs4406273 | G | A | 0.0921 | 0.128 | 0.013 | 3.9811E-23 | 96.95 |

| | | | | | | | | |
|---|---|---|---|---|---|---|---|---|
| *PCSK7* | rs171052 | C | G | 0.2261 | -0.379 | 0.009 | 1E-200 | 1773.35 |
| *PCSK9* | rs11591147 | G | T | 0.0177 | -1.153 | 0.028 | 1E-200 | 1695.67 |
| *PCSK9* | rs547186 | T | A | 0.4963 | 0.072 | 0.007 | 1.5849E-23 | 105.80 |
| *PCSK9* | rs1260326 | T | C | 0.6084 | -0.076 | 0.007 | 2.5119E-25 | 117.88 |
| *PCSK9* | rs2001945 | G | C | 0.5189 | -0.057 | 0.007 | 1.9953E-15 | 66.31 |
| *PCSK9* | rs174568 | C | T | 0.3439 | 0.079 | 0.007 | 5.0119E-26 | 127.37 |
| *PCSK9* | rs673335 | T | C | 0.1553 | -0.066 | 0.01 | 1.5849E-11 | 43.56 |
| *PCSK9* | rs261342 | G | C | 0.7772 | -0.066 | 0.009 | 3.1623E-14 | 53.78 |
| *PCSK9* | rs8106503 | T | C | 0.1188 | -0.094 | 0.011 | 1.5849E-17 | 73.02 |
| *PCSK9* | rs756350040 | TGACA | T | 0.0735 | -0.125 | 0.014 | 1.2589E-19 | 79.72 |
| *PCYT2* | rs1354034 | T | C | 0.6046 | 0.072 | 0.008 | 7.9433E-21 | 81.00 |
| *PCYT2* | rs6993770 | A | T | 0.2866 | -0.094 | 0.008 | 6.3096E-30 | 138.06 |
| *PDAP1* | rs1354034 | T | C | 0.6045 | 0.073 | 0.008 | 2.5119E-21 | 83.27 |
| *PDAP1* | rs342298 | C | T | 0.4545 | -0.054 | 0.008 | 1E-12 | 45.56 |
| *PDAP1* | rs60822569 | T | TTTCTC | 0.5538 | 0.052 | 0.008 | 6.3096E-12 | 42.25 |
| *PDCD1* | rs37471 | C | T | 0.3522 | 0.071 | 0.008 | 7.9433E-21 | 78.77 |
| *PDCD1* | rs2476601 | A | G | 0.8993 | -0.086 | 0.012 | 1.2589E-12 | 51.36 |
| *PDCD1* | rs376812565 | C | T | 0.6388 | -0.062 | 0.009 | 1.5849E-12 | 47.46 |
| *PDCD1* | rs11888817 | T | C | 0.642 | 0.054 | 0.008 | 3.9811E-12 | 45.56 |
| *PDCD1* | rs11571291 | T | C | 0.424 | -0.058 | 0.008 | 1.5849E-14 | 52.56 |
| *PDCD1* | rs9838308 | T | C | 0.3658 | -0.054 | 0.008 | 1.9953E-12 | 45.56 |
| *PDCD1* | rs3131622 | T | G | 0.4501 | 0.054 | 0.007 | 1E-13 | 59.51 |
| *PDCD1* | rs80318131 | CT | C | 0.3053 | 0.056 | 0.008 | 1.5849E-11 | 49.00 |
| *PDCD1* | rs12772559 | G | C | 0.2564 | 0.084 | 0.008 | 1E-23 | 110.25 |
| *PDCD1* | rs111338191 | A | AAATT | 0.5203 | -0.101 | 0.007 | 6.3096E-43 | 208.18 |
| *PDCD1* | rs3213566 | T | C | 0.528 | -0.05 | 0.007 | 7.9433E-12 | 51.02 |
| *PDCD1* | rs7161799 | C | T | 0.0765 | 0.129 | 0.014 | 6.3096E-21 | 84.90 |
| *PDCD1* | rs186021206 | G | A | 0.0064 | 0.616 | 0.05 | 1.2589E-34 | 151.78 |
| *PDCD1* | rs2659005 | C | T | 0.4417 | 0.065 | 0.007 | 1.2589E-18 | 86.22 |
| *PDCD1* | rs753324368 | C | CA | 0.7397 | -0.078 | 0.008 | 1.2589E-20 | 95.06 |
| *PDCD1LG2* | rs812490 | T | C | 0.4519 | -0.058 | 0.006 | 1E-20 | 93.44 |
| *PDCD1LG2* | rs10935473 | G | T | 0.4404 | -0.459 | 0.007 | 1E-200 | 4299.61 |
| *PDCD1LG2* | rs112125244 | T | TTTTG | 0.2848 | -0.05 | 0.007 | 3.1623E-13 | 51.02 |
| *PDCD1LG2* | rs2381282 | T | C | 0.4391 | 0.601 | 0.007 | 1E-200 | 7371.45 |
| *PDCD1LG2* | rs507666 | G | A | 0.1847 | -0.199 | 0.009 | 1.995E-101 | 488.90 |
| *PDCD1LG2* | rs4055121 | C | T | 0.1347 | -0.081 | 0.009 | 3.9811E-19 | 81.00 |
| *PDCD1LG2* | rs3184504 | T | C | 0.5168 | -0.053 | 0.006 | 3.1623E-18 | 78.03 |
| *PDCD1LG2* | rs186021206 | G | A | 0.0063 | 0.973 | 0.043 | 5.012E-115 | 512.02 |
| *PDCD5* | rs1354034 | T | C | 0.6042 | 0.065 | 0.007 | 3.9811E-19 | 86.22 |
| *PDCD5* | rs60822569 | T | TTTCTC | 0.5544 | 0.052 | 0.007 | 6.3096E-13 | 55.18 |
| *PDCD5* | rs10402931 | G | A | 0.6839 | 0.384 | 0.008 | 1E-200 | 2304.00 |
| *PDCD6* | rs676705 | G | A | 0.351 | -0.055 | 0.006 | 6.3096E-22 | 84.03 |
| *PDCD6* | rs9654451 | C | T | 0.1284 | -1.497 | 0.011 | 1E-200 | 18520.74 |
| *PDCD6* | rs9495061 | C | A | 0.0585 | -0.086 | 0.012 | 6.3096E-14 | 51.36 |
| *PDCD6* | rs11438680 | G | GA | 0.4605 | 0.042 | 0.005 | 1.2589E-14 | 70.56 |
| *PDCD6* | rs226379 | T | C | 0.3551 | -0.057 | 0.006 | 7.9433E-24 | 90.25 |
| *PDCD6* | rs112635299 | G | T | 0.0216 | 0.161 | 0.019 | 3.9811E-18 | 71.80 |
| *PDE5A* | rs1354034 | T | C | 0.6044 | 0.053 | 0.008 | 3.1623E-12 | 43.89 |
| *PDE5A* | rs58583086 | A | G | 0.6255 | -0.158 | 0.008 | 5.0119E-91 | 390.06 |
| *PDE5A* | rs7080386 | C | A | 0.412 | 0.064 | 0.008 | 6.3096E-17 | 64.00 |
| *PDGFA* | rs6671171 | G | A | 0.111 | 0.087 | 0.012 | 1.5849E-13 | 52.56 |
| *PDGFA* | rs3811444 | C | T | 0.3323 | -0.072 | 0.008 | 7.9433E-20 | 81.00 |
| *PDGFA* | rs7618405 | C | A | 0.2048 | -0.066 | 0.009 | 7.9433E-13 | 53.78 |
| *PDGFA* | rs1354034 | T | C | 0.6039 | -0.121 | 0.008 | 6.3096E-58 | 228.77 |
| *PDGFA* | rs2278668 | T | C | 0.5935 | -0.058 | 0.008 | 1.9953E-14 | 52.56 |
| *PDGFA* | rs4572884 | C | T | 0.3977 | -0.069 | 0.008 | 7.9433E-20 | 74.39 |
| *PDGFA* | rs10058074 | G | A | 0.4755 | -0.053 | 0.007 | 7.9433E-13 | 57.33 |
| *PDGFA* | rs2516471 | G | A | 0.0636 | -0.124 | 0.015 | 5.0119E-16 | 68.34 |
| *PDGFA* | rs146842698 | T | TAAATA | 0.3052 | -0.059 | 0.008 | 7.9433E-13 | 54.39 |
| *PDGFA* | rs2759391 | A | C | 0.3434 | -0.056 | 0.008 | 1E-12 | 49.00 |
| *PDGFA* | rs144168379 | C | T | 0.0033 | -0.446 | 0.066 | 1.5849E-11 | 45.66 |
| *PDGFA* | rs6961069 | C | T | 0.4026 | 0.069 | 0.008 | 1E-19 | 74.39 |
| *PDGFA* | rs6993770 | A | T | 0.2865 | -0.18 | 0.008 | 6.31E-106 | 506.25 |
| *PDGFA* | rs7080386 | C | A | 0.4115 | 0.118 | 0.008 | 3.1623E-55 | 217.56 |
| *PDGFA* | rs34377578 | A | C | 0.2529 | 0.082 | 0.009 | 1.5849E-21 | 83.01 |
| *PDGFA* | rs5796220 | C | CACT | 0.4893 | -0.051 | 0.007 | 1E-11 | 53.08 |
| *PDGFA* | rs59001897 | T | A | 0.1749 | 0.079 | 0.01 | 7.9433E-16 | 62.41 |
| *PDGFA* | rs12445050 | C | T | 0.1384 | 0.113 | 0.011 | 6.3096E-26 | 105.53 |

| | | | | | | | | |
|---|---|---|---|---|---|---|---|---|
| *PDGFA* | rs1654425 | T | C | 0.8343 | 0.169 | 0.01 | 1.5849E-63 | 285.61 |
| *PDGFA* | rs6081565 | G | A | 0.3523 | 0.073 | 0.008 | 3.9811E-21 | 83.27 |
| *PDGFB* | rs75699653 | C | T | 0.0236 | 0.173 | 0.025 | 1.9953E-12 | 47.89 |
| *PDGFB* | rs68066031 | T | C | 0.2298 | 0.229 | 0.009 | 2.512E-145 | 647.42 |
| *PDGFB* | rs1354034 | T | C | 0.6041 | -0.075 | 0.008 | 2.5119E-23 | 87.89 |
| *PDGFB* | rs4572884 | C | T | 0.3976 | -0.065 | 0.008 | 6.3096E-18 | 66.02 |
| *PDGFB* | rs274555 | C | T | 0.3986 | 0.053 | 0.008 | 6.3096E-12 | 43.89 |
| *PDGFB* | rs138614581 | A | AT | 0.08 | -0.107 | 0.014 | 6.3096E-15 | 58.41 |
| *PDGFB* | rs1917342 | A | G | 0.3036 | -0.06 | 0.008 | 1.2589E-13 | 56.25 |
| *PDGFB* | rs6961069 | C | T | 0.4025 | 0.066 | 0.008 | 3.1623E-18 | 68.06 |
| *PDGFB* | rs6993770 | A | T | 0.2867 | -0.14 | 0.008 | 5.0119E-65 | 306.25 |
| *PDGFB* | rs10820606 | A | C | 0.2298 | -0.077 | 0.009 | 1E-17 | 73.20 |
| *PDGFB* | rs10761737 | T | C | 0.4132 | 0.105 | 0.008 | 1.9953E-43 | 172.27 |
| *PDGFB* | rs34377578 | A | C | 0.2529 | 0.081 | 0.009 | 3.1623E-21 | 81.00 |
| *PDGFB* | rs1362213 | C | T | 0.4794 | -0.054 | 0.008 | 6.3096E-13 | 45.56 |
| *PDGFB* | rs60128101 | T | A | 0.1747 | 0.071 | 0.01 | 7.9433E-13 | 50.41 |
| *PDGFB* | rs11639051 | C | A | 0.2949 | 0.091 | 0.008 | 5.0119E-29 | 129.39 |
| *PDGFB* | rs12445050 | C | T | 0.138 | 0.105 | 0.011 | 2.5119E-22 | 91.12 |
| *PDGFB* | rs1654425 | T | C | 0.8344 | 0.165 | 0.01 | 5.0119E-60 | 272.25 |
| *PDGFB* | rs3790176 | G | A | 0.3426 | 0.078 | 0.008 | 1.9953E-23 | 95.06 |
| *PDGFC* | rs12499706 | C | G | 0.2747 | -0.091 | 0.009 | 2.5119E-26 | 102.23 |
| *PDGFC* | rs3775298 | A | G | 0.5133 | -0.086 | 0.008 | 3.9811E-29 | 115.56 |
| *PDGFC* | rs75077631 | G | GC | 0.742 | -0.061 | 0.009 | 3.9811E-12 | 45.94 |
| *PDGFC* | rs150611042 | C | A | 0.0719 | -0.472 | 0.015 | 1E-200 | 990.15 |
| *PDGFC* | rs28929474 | C | T | 0.0207 | 0.352 | 0.027 | 6.3096E-39 | 169.96 |
| *PDGFC* | rs149394327 | G | C | 0.0297 | 0.359 | 0.023 | 1.9953E-56 | 243.63 |
| *PDGFC* | rs1065853 | G | T | 0.0809 | 0.183 | 0.014 | 2.5119E-38 | 170.86 |
| *PDGFRA* | rs61801010 | A | G | 0.106 | 0.154 | 0.012 | 3.9811E-36 | 164.69 |
| *PDGFRA* | rs56033718 | A | G | 0.3709 | 0.082 | 0.007 | 1E-27 | 137.22 |
| *PDGFRA* | rs4558798 | A | G | 0.6725 | -0.053 | 0.008 | 3.9811E-12 | 43.89 |
| *PDGFRA* | rs35597368 | T | C | 0.1097 | -0.425 | 0.012 | 1E-200 | 1254.34 |
| *PDGFRA* | rs6913664 | C | T | 0.4729 | 0.056 | 0.007 | 7.9433E-15 | 64.00 |
| *PDGFRA* | rs3751198 | A | G | 0.6042 | 0.126 | 0.008 | 3.9811E-63 | 248.06 |
| *PDGFRA* | rs12148472 | T | C | 0.1303 | 0.076 | 0.011 | 1E-12 | 47.74 |
| *PDGFRA* | rs180915454 | C | T | 0.0045 | 0.406 | 0.059 | 5.0119E-12 | 47.35 |
| *PDGFRA* | rs2294915 | C | T | 0.2316 | 0.063 | 0.009 | 1.9953E-13 | 49.00 |
| *PDGFRB* | rs2304058 | C | G | 0.5575 | 1.179 | 0.008 | 1E-200 | 21719.39 |
| *PDGFRB* | rs68055275 | C | CT | 0.1448 | -0.067 | 0.007 | 2.5119E-22 | 91.61 |
| *PDGFRB* | rs186021206 | G | A | 0.0064 | 0.418 | 0.033 | 3.1623E-37 | 160.44 |
| *PDIA3* | rs59162344 | AT | A | 0.7633 | 0.065 | 0.009 | 1.5849E-12 | 52.16 |
| *PDIA4* | rs1354034 | T | C | 0.6046 | 0.067 | 0.008 | 3.1623E-18 | 70.14 |
| *PDIA4* | rs3804749 | C | T | 0.5926 | -0.053 | 0.008 | 5.0119E-12 | 43.89 |
| *PDIA4* | rs6953222 | G | T | 0.1744 | -0.117 | 0.01 | 2.5119E-32 | 136.89 |
| *PDIA4* | rs6993770 | A | T | 0.2863 | -0.057 | 0.008 | 6.3096E-12 | 50.77 |
| *PDIA5* | rs3804749 | C | T | 0.5931 | -0.249 | 0.008 | 1E-200 | 968.77 |
| *PDIA5* | rs2844530 | C | G | 0.1442 | 0.291 | 0.011 | 5.012E-156 | 699.84 |
| *PDIA5* | rs1646724 | T | G | 0.4815 | 0.073 | 0.008 | 1E-21 | 83.27 |
| *PDIA5* | rs6081565 | G | A | 0.3537 | 0.08 | 0.008 | 5.0119E-24 | 100.00 |
| *PDLIM5* | rs1354034 | T | C | 0.6043 | 0.055 | 0.008 | 2.5119E-13 | 47.27 |
| *PDLIM5* | rs342298 | C | T | 0.4549 | -0.078 | 0.007 | 2.5119E-25 | 124.16 |
| *PDLIM5* | rs6993770 | A | T | 0.2864 | -0.075 | 0.008 | 1.5849E-19 | 87.89 |
| *PDLIM5* | rs11553699 | A | G | 0.1354 | -0.08 | 0.011 | 1.5849E-12 | 52.89 |
| *PDLIM7* | rs1354034 | T | C | 0.6041 | 0.055 | 0.008 | 3.1623E-13 | 47.27 |
| *PDLIM7* | rs342296 | G | A | 0.4563 | -0.08 | 0.007 | 7.9433E-27 | 130.61 |
| *PDLIM7* | rs11553699 | A | G | 0.1354 | -0.084 | 0.011 | 1E-13 | 58.31 |
| *PDZD2* | rs183711 | T | G | 0.9197 | 0.394 | 0.014 | 7.943E-173 | 792.02 |
| *PDZD2* | rs342293 | C | G | 0.4598 | -0.066 | 0.008 | 2.5119E-18 | 68.06 |
| *PDZD2* | rs6580981 | G | A | 0.4581 | 0.076 | 0.008 | 6.3096E-23 | 90.25 |
| *PDZK1* | rs1471633 | A | C | 0.5339 | -0.111 | 0.008 | 6.3096E-49 | 192.52 |
| *PDZK1* | rs4917 | T | C | 0.6405 | 0.132 | 0.008 | 1.9953E-62 | 272.25 |
| *PDZK1* | rs7990 | C | A | 0.135 | -0.139 | 0.012 | 1.5849E-33 | 134.17 |
| *PEAR1* | rs4661012 | T | G | 0.362 | -0.213 | 0.008 | 1.585E-165 | 708.89 |
| *PEAR1* | rs1354034 | T | C | 0.6039 | -0.064 | 0.007 | 1.5849E-17 | 83.59 |
| *PEAR1* | rs13107325 | C | T | 0.0749 | 0.17 | 0.014 | 3.9811E-34 | 147.45 |
| *PEAR1* | rs9389269 | T | C | 0.2718 | 0.061 | 0.008 | 1.5849E-13 | 58.14 |
| *PEAR1* | rs10758659 | A | G | 0.5642 | 0.057 | 0.007 | 2.5119E-14 | 66.31 |
| *PEAR1* | rs8176741 | G | A | 0.0612 | 0.438 | 0.015 | 6.31E-176 | 852.64 |
| *PEAR1* | rs3967200 | C | T | 0.1346 | -0.078 | 0.011 | 3.9811E-13 | 50.28 |

| | | | | | | | | |
|---|---|---|---|---|---|---|---|---|
| *PEAR1* | rs370664797 | T | TAC | 0.2507 | -0.064 | 0.009 | 3.9811E-12 | 50.57 |
| *PEAR1* | rs35350651 | A | AC | 0.5037 | -0.076 | 0.007 | 3.9811E-25 | 117.88 |
| *PEAR1* | rs186021206 | G | A | 0.0064 | 0.758 | 0.05 | 3.9811E-51 | 229.83 |
| *PEBP1* | rs1354034 | T | C | 0.6041 | 0.059 | 0.007 | 5.0119E-15 | 71.04 |
| *PECAM1* | rs1354034 | T | C | 0.6041 | 0.073 | 0.007 | 7.9433E-24 | 108.76 |
| *PECAM1* | rs56083751 | T | A | 0.4776 | -0.051 | 0.007 | 7.9433E-13 | 53.08 |
| *PECAM1* | rs2519093 | C | T | 0.1843 | -0.587 | 0.01 | 1E-200 | 3445.69 |
| *PECAM1* | rs35166255 | G | A | 0.0336 | 0.324 | 0.02 | 3.9811E-60 | 262.44 |
| *PECAM1* | rs3184504 | T | C | 0.5175 | -0.054 | 0.007 | 1.5849E-14 | 59.51 |
| *PECAM1* | rs186021206 | G | A | 0.0064 | 0.611 | 0.049 | 6.3096E-36 | 155.49 |
| *PECR* | rs10197542 | A | C | 0.1044 | -0.454 | 0.013 | 1E-200 | 1219.62 |
| *PECR* | rs6993770 | A | T | 0.2866 | -0.059 | 0.008 | 2.5119E-12 | 54.39 |
| *PENK* | rs781656 | G | A | 0.1832 | 0.09 | 0.009 | 1E-21 | 100.00 |
| *PENK* | rs2870238 | C | T | 0.503 | 0.049 | 0.007 | 7.9433E-12 | 49.00 |
| *PENK* | rs11154902 | C | T | 0.4622 | -0.057 | 0.007 | 6.3096E-15 | 66.31 |
| *PENK* | rs2576575 | C | T | 0.4662 | -0.301 | 0.007 | 1E-200 | 1849.00 |
| *PEPD* | rs763228358 | AG | A | 0.245 | -0.068 | 0.009 | 2.5119E-15 | 57.09 |
| *PEPD* | rs1047891 | C | A | 0.3131 | -0.072 | 0.008 | 5.0119E-21 | 81.00 |
| *PEPD* | rs7032525 | T | C | 0.2002 | -0.132 | 0.009 | 6.3096E-50 | 215.11 |
| *PEPD* | rs35032070 | G | A | 0.364 | -0.078 | 0.007 | 2.5119E-26 | 124.16 |
| *PEPD* | rs2353483 | C | T | 0.7252 | 0.057 | 0.008 | 7.9433E-13 | 50.77 |
| *PEPD* | rs72731930 | C | T | 0.0717 | -0.21 | 0.014 | 1.2589E-52 | 225.00 |
| *PEPD* | rs17569 | G | A | 0.1566 | -0.493 | 0.01 | 1E-200 | 2430.49 |
| *PEPD* | rs6125961 | C | A | 0.2006 | -0.063 | 0.009 | 1E-12 | 49.00 |
| *PEPD* | rs2904552 | C | T | 0.1043 | 0.135 | 0.012 | 1E-29 | 126.56 |
| *PER3* | rs228647 | G | A | 0.5634 | 0.37 | 0.008 | 1E-200 | 2139.06 |
| *PER3* | rs1354034 | T | C | 0.6039 | 0.052 | 0.007 | 1E-12 | 55.18 |
| *PER3* | rs6440003 | G | A | 0.4426 | -0.05 | 0.007 | 1.9953E-12 | 51.02 |
| *PER3* | rs6993770 | A | T | 0.2866 | -0.133 | 0.008 | 1.5849E-63 | 276.39 |
| *PER3* | rs249177 | C | T | 0.1844 | -0.086 | 0.009 | 6.3096E-21 | 91.31 |
| *PF4* | rs13412535 | G | A | 0.2307 | -0.061 | 0.009 | 6.3096E-12 | 45.94 |
| *PF4* | rs1354034 | T | C | 0.6046 | -0.072 | 0.008 | 6.3096E-21 | 81.00 |
| *PF4* | rs11574452 | C | A | 0.0451 | -0.201 | 0.018 | 1E-27 | 124.69 |
| *PF4* | rs4698839 | C | T | 0.3983 | -0.058 | 0.008 | 3.9811E-14 | 52.56 |
| *PF4* | rs274555 | C | T | 0.3988 | 0.06 | 0.008 | 7.9433E-15 | 56.25 |
| *PF4* | rs2516471 | G | A | 0.0633 | -0.114 | 0.016 | 2.5119E-13 | 50.77 |
| *PF4* | rs6961069 | C | T | 0.403 | 0.063 | 0.008 | 1.2589E-16 | 62.02 |
| *PF4* | rs6993770 | A | T | 0.2865 | -0.115 | 0.008 | 6.3096E-44 | 206.64 |
| *PF4* | rs10820606 | A | C | 0.2297 | -0.066 | 0.009 | 2.5119E-13 | 53.78 |
| *PF4* | rs10761737 | T | C | 0.4128 | 0.121 | 0.008 | 1.9953E-56 | 228.77 |
| *PF4* | rs12445050 | C | T | 0.1384 | 0.087 | 0.011 | 1E-15 | 62.55 |
| *PF4* | rs1654425 | T | C | 0.8343 | 0.149 | 0.01 | 1.2589E-48 | 222.01 |
| *PF4* | rs6081569 | G | C | 0.3686 | 0.072 | 0.008 | 3.1623E-20 | 81.00 |
| *PFKFB2* | rs1560 | A | G | 0.4821 | -0.188 | 0.007 | 3.981E-142 | 721.31 |
| *PFKFB2* | rs1354034 | T | C | 0.6047 | 0.058 | 0.007 | 1E-14 | 68.65 |
| *PFKFB2* | rs6993770 | A | T | 0.2864 | -0.06 | 0.008 | 1.9953E-13 | 56.25 |
| *PFKFB2* | rs7080386 | C | A | 0.4115 | 0.054 | 0.007 | 7.9433E-13 | 59.51 |
| *PGA4* | rs3762954 | T | C | 0.3028 | 0.058 | 0.008 | 7.9433E-13 | 52.56 |
| *PGA4* | rs1639109 | G | A | 0.1541 | -0.093 | 0.01 | 6.3096E-20 | 86.49 |
| *PGA4* | rs34357864 | G | GA | 0.3177 | -0.055 | 0.008 | 2.5119E-12 | 47.27 |
| *PGA4* | rs117739035 | G | T | 0.0368 | 0.148 | 0.02 | 3.9811E-14 | 54.76 |
| *PGA4* | rs4414241 | C | T | 0.2435 | 0.078 | 0.01 | 5.0119E-16 | 60.84 |
| *PGA4* | rs112585477 | G | A | 0.041 | 0.129 | 0.019 | 3.9811E-12 | 46.10 |
| *PGD* | rs2229688 | G | A | 0.0171 | 0.867 | 0.029 | 1E-200 | 893.80 |
| *PGD* | rs1354034 | T | C | 0.6045 | 0.056 | 0.008 | 1.2589E-13 | 49.00 |
| *PGD* | rs142714816 | A | G | 0.0008 | 0.985 | 0.134 | 1.5849E-13 | 54.03 |
| *PGD* | rs6993770 | A | T | 0.2866 | -0.059 | 0.008 | 3.9811E-13 | 54.39 |
| *PGD* | rs3847326 | A | G | 0.4815 | -0.06 | 0.007 | 3.9811E-16 | 73.47 |
| *PGD* | rs4759076 | T | C | 0.4567 | 0.056 | 0.007 | 1.2589E-13 | 64.00 |
| *PGF* | rs9943922 | T | C | 0.5333 | 0.084 | 0.007 | 6.3096E-36 | 144.00 |
| *PGF* | rs4902651 | C | T | 0.5019 | -0.047 | 0.007 | 3.1623E-12 | 45.08 |
| *PGF* | rs704 | G | A | 0.4733 | -0.048 | 0.007 | 5.0119E-13 | 47.02 |
| *PGLYRP1* | rs3917932 | C | G | 0.5756 | -0.067 | 0.007 | 1.5849E-19 | 91.61 |
| *PGLYRP1* | rs113542380 | G | A | 0.074 | 0.138 | 0.014 | 1E-22 | 97.16 |
| *PGLYRP1* | rs6436124 | C | A | 0.6066 | 0.084 | 0.008 | 5.0119E-29 | 110.25 |
| *PGLYRP1* | rs2290547 | G | A | 0.1757 | 0.065 | 0.01 | 7.9433E-12 | 42.25 |
| *PGLYRP1* | rs41266485 | C | T | 0.2044 | 0.078 | 0.009 | 6.3096E-18 | 75.11 |
| *PGLYRP1* | rs218264 | A | T | 0.2472 | 0.066 | 0.009 | 1.2589E-14 | 53.78 |

| | | | | | | | | |
|---|---|---|---|---|---|---|---|---|
| *PGLYRP1* | rs141484466 | G | C | 0.0417 | 0.315 | 0.019 | 3.9811E-60 | 274.86 |
| *PGLYRP1* | rs11775560 | A | G | 0.1864 | 0.13 | 0.009 | 3.9811E-44 | 208.64 |
| *PGLYRP1* | rs4407843 | A | C | 0.5683 | -0.054 | 0.007 | 3.9811E-13 | 59.51 |
| *PGLYRP1* | rs2074038 | G | T | 0.1 | 0.117 | 0.012 | 1E-21 | 95.06 |
| *PGLYRP1* | rs2511241 | C | T | 0.9293 | 0.101 | 0.014 | 1E-12 | 52.05 |
| *PGLYRP1* | rs371147134 | G | GA | 0.5246 | -0.06 | 0.007 | 6.3096E-16 | 73.47 |
| *PGLYRP1* | rs7310615 | C | G | 0.5173 | -0.051 | 0.007 | 3.9811E-12 | 53.08 |
| *PGLYRP1* | rs2072563 | G | A | 0.3253 | -0.297 | 0.008 | 1E-200 | 1378.27 |
| *PGLYRP2* | rs59950280 | G | A | 0.3301 | 0.064 | 0.006 | 1E-28 | 113.78 |
| *PGLYRP2* | rs33951980 | C | T | 0.1287 | -0.072 | 0.008 | 1E-19 | 81.00 |
| *PGLYRP2* | rs2243458 | C | T | 0.3071 | -0.046 | 0.006 | 7.9433E-16 | 58.78 |
| *PGLYRP2* | rs34440547 | C | T | 0.2193 | -1.144 | 0.009 | 1E-200 | 16157.23 |
| *PGLYRP2* | rs2229742 | G | C | 0.1036 | 0.065 | 0.009 | 7.9433E-14 | 52.16 |
| *PGLYRP4* | rs111586430 | C | T | 0.0569 | 0.922 | 0.017 | 1E-200 | 2941.47 |
| *PGR* | rs139299944 | C | CT | 0.3437 | 0.098 | 0.008 | 5.0119E-34 | 150.06 |
| *PGR* | rs4813331 | C | T | 0.3814 | -0.149 | 0.008 | 2.5119E-78 | 346.89 |
| *PHACTR2* | rs1354034 | T | C | 0.6043 | 0.07 | 0.008 | 2.5119E-20 | 76.56 |
| *PHACTR2* | rs9484759 | A | G | 0.2042 | 0.248 | 0.009 | 6.31E-157 | 759.31 |
| *PHLDB1* | rs1354034 | T | C | 0.6046 | 0.061 | 0.008 | 1.2589E-15 | 58.14 |
| *PHLDB1* | rs342293 | C | G | 0.4598 | -0.075 | 0.007 | 1.2589E-23 | 114.80 |
| *PHOSPHO1* | rs543343625 | C | T | 0.0008 | -1.879 | 0.142 | 7.9433E-40 | 175.10 |
| *PHYKPL* | rs142181517 | T | A | 0.0074 | -2.038 | 0.044 | 1E-200 | 2145.37 |
| *PI16* | rs61821645 | A | G | 0.0272 | 0.173 | 0.023 | 5.0119E-14 | 56.58 |
| *PI16* | rs35004449 | G | T | 0.2592 | 0.343 | 0.008 | 1E-200 | 1838.27 |
| *PI16* | rs62406524 | G | C | 0.0964 | -0.43 | 0.013 | 1E-200 | 1094.08 |
| *PI16* | rs34931250 | C | T | 0.0606 | -0.166 | 0.015 | 3.1623E-29 | 122.47 |
| *PI16* | rs12975366 | T | C | 0.3956 | 0.073 | 0.007 | 3.9811E-24 | 108.76 |
| *PI3* | rs34155263 | C | T | 0.4377 | -0.059 | 0.007 | 1.2589E-16 | 71.04 |
| *PI3* | rs28550378 | C | T | 0.4653 | 0.05 | 0.007 | 6.3096E-13 | 51.02 |
| *PI3* | rs7088444 | C | T | 0.4783 | 0.06 | 0.007 | 1.2589E-17 | 73.47 |
| *PI3* | rs35770435 | C | T | 0.1804 | 0.353 | 0.009 | 1E-200 | 1538.38 |
| *PIBF1* | rs1354034 | T | C | 0.6043 | 0.094 | 0.008 | 1E-34 | 138.06 |
| *PIBF1* | rs342296 | G | A | 0.4567 | -0.058 | 0.007 | 7.9433E-15 | 68.65 |
| *PIBF1* | rs6993770 | A | T | 0.2864 | -0.061 | 0.008 | 2.5119E-13 | 58.14 |
| *PIBF1* | rs4759076 | T | C | 0.457 | 0.054 | 0.008 | 1.2589E-12 | 45.56 |
| *PIBF1* | rs61966211 | G | T | 0.3844 | -0.062 | 0.008 | 7.9433E-16 | 60.06 |
| *PIGR* | rs291102 | G | A | 0.024 | -0.508 | 0.025 | 5.0119E-95 | 412.90 |
| *PIGR* | rs1200427 | A | T | 0.2451 | 0.085 | 0.009 | 1.9953E-22 | 89.20 |
| *PIGR* | rs113810201 | A | G | 0.1148 | 0.101 | 0.012 | 7.9433E-18 | 70.84 |
| *PIGR* | rs61882275 | G | A | 0.3771 | -0.057 | 0.008 | 1.9953E-13 | 50.77 |
| *PIGR* | rs708686 | C | T | 0.2676 | 0.11 | 0.009 | 1.5849E-38 | 149.38 |
| *PIK3AP1* | rs11204523 | G | C | 0.7589 | -0.079 | 0.009 | 5.0119E-20 | 77.05 |
| *PIK3AP1* | rs1354034 | T | C | 0.6043 | 0.103 | 0.007 | 3.1623E-44 | 216.51 |
| *PIK3AP1* | rs7695151 | C | T | 0.4644 | -0.054 | 0.007 | 6.3096E-14 | 59.51 |
| *PIK3AP1* | rs78842280 | T | G | 0.274 | -0.063 | 0.008 | 1.9953E-14 | 62.02 |
| *PIK3AP1* | rs342293 | C | G | 0.4594 | -0.052 | 0.007 | 5.0119E-13 | 55.18 |
| *PIK3AP1* | rs41317268 | A | G | 0.1948 | -0.23 | 0.009 | 6.31E-136 | 653.09 |
| *PIK3IP1* | rs28640218 | G | T | 0.1914 | -0.074 | 0.009 | 1.2589E-16 | 67.60 |
| *PIK3IP1* | rs2413028 | T | C | 0.171 | -0.368 | 0.009 | 1E-200 | 1671.90 |
| *PIKFYVE* | rs1354034 | T | C | 0.6039 | 0.054 | 0.008 | 1.2589E-12 | 45.56 |
| *PIKFYVE* | rs34436714 | C | A | 0.2092 | 0.066 | 0.009 | 7.9433E-13 | 53.78 |
| *PILRA* | rs61747728 | C | T | 0.0379 | 0.1 | 0.015 | 1.5849E-11 | 44.44 |
| *PILRA* | rs377744692 | A | AAAAAG | 0.581 | 0.04 | 0.006 | 1.5849E-11 | 44.44 |
| *PILRA* | rs10085857 | C | T | 0.4804 | 0.189 | 0.008 | 6.31E-129 | 558.14 |
| *PILRA* | rs1859788 | A | G | 0.6809 | 1 | 0.008 | 1E-200 | 15625.00 |
| *PILRA* | rs3184504 | T | C | 0.5175 | -0.062 | 0.006 | 3.9811E-28 | 106.78 |
| *PILRA* | rs186021206 | G | A | 0.0064 | 0.556 | 0.039 | 6.3096E-46 | 203.25 |
| *PILRA* | rs140528980 | T | C | 0.3949 | -0.042 | 0.006 | 6.3096E-13 | 49.00 |
| *PILRA* | rs61698755 | T | C | 0.5669 | -0.05 | 0.006 | 2.5119E-18 | 69.44 |
| *PILRB* | rs10085857 | C | T | 0.4807 | 0.219 | 0.008 | 1E-168 | 749.39 |
| *PILRB* | rs1859788 | A | G | 0.6812 | 1.147 | 0.008 | 1E-200 | 20556.39 |
| *PILRB* | rs7310615 | C | G | 0.5178 | -0.057 | 0.005 | 1.9953E-28 | 129.96 |
| *PILRB* | rs186021206 | G | A | 0.0064 | 0.471 | 0.035 | 1.5849E-41 | 181.09 |
| *PILRB* | rs140528980 | T | C | 0.395 | -0.036 | 0.005 | 7.9433E-12 | 51.84 |
| *PILRB* | rs8077394 | A | G | 0.554 | -0.046 | 0.005 | 2.5119E-19 | 84.64 |
| *PINLYP* | rs80293268 | G | C | 0.0483 | -0.12 | 0.016 | 1.2589E-14 | 56.25 |
| *PINLYP* | rs36010924 | A | G | 0.1842 | -0.089 | 0.008 | 1.5849E-27 | 123.77 |
| *PINLYP* | rs4681529 | T | A | 0.6773 | -0.059 | 0.007 | 2.5119E-18 | 71.04 |

| | | | | | | | | |
|---|---|---|---|---|---|---|---|---|
| *PINLYP* | rs35457250 | C | T | 0.0088 | 0.634 | 0.034 | 1E-78 | 347.71 |
| *PINLYP* | rs2517801 | T | C | 0.6125 | 0.046 | 0.007 | 1.5849E-11 | 43.18 |
| *PINLYP* | rs16888292 | G | A | 0.1943 | -0.059 | 0.008 | 1.2589E-13 | 54.39 |
| *PINLYP* | rs7905367 | G | C | 0.7973 | 0.053 | 0.008 | 1E-11 | 43.89 |
| *PINLYP* | rs58589035 | T | C | 0.1198 | -0.067 | 0.01 | 6.3096E-12 | 44.89 |
| *PINLYP* | rs3803856 | T | C | 0.6483 | -0.047 | 0.007 | 3.1623E-12 | 45.08 |
| *PINLYP* | rs112001035 | G | A | 0.0614 | 0.354 | 0.014 | 1.995E-150 | 639.37 |
| *PINLYP* | rs12457893 | A | C | 0.4583 | -0.047 | 0.006 | 2.5119E-13 | 61.36 |
| *PINLYP* | rs3213276 | C | T | 0.5878 | 0.767 | 0.008 | 1E-200 | 9192.02 |
| *PKD1* | rs2728104 | T | C | 0.1108 | -0.098 | 0.011 | 6.3096E-20 | 79.37 |
| *PKD1* | rs8176749 | C | T | 0.0612 | 0.143 | 0.014 | 3.1623E-24 | 104.33 |
| *PKD1* | rs56278466 | T | G | 0.6612 | 0.109 | 0.007 | 3.9811E-52 | 242.47 |
| *PKD1* | rs186021206 | G | A | 0.0064 | 0.669 | 0.047 | 1.2589E-46 | 202.61 |
| *PKD1* | rs868875 | A | G | 0.3014 | -0.052 | 0.007 | 2.5119E-12 | 55.18 |
| *PKD1* | rs189448562 | G | C | 0.0113 | 0.545 | 0.033 | 3.1623E-62 | 272.75 |
| *PKD2* | rs1354034 | T | C | 0.6037 | 0.1 | 0.008 | 3.9811E-38 | 156.25 |
| *PKD2* | rs2728104 | T | C | 0.1109 | -0.137 | 0.012 | 6.3096E-30 | 130.34 |
| *PKD2* | rs7080536 | G | A | 0.0436 | -0.227 | 0.019 | 2.5119E-33 | 142.74 |
| *PKD2* | rs11604127 | C | T | 0.2304 | 0.064 | 0.009 | 1.9953E-12 | 50.57 |
| *PKD2* | rs12292693 | A | C | 0.2603 | -0.06 | 0.009 | 5.0119E-12 | 44.44 |
| *PKLR* | rs77350683 | T | C | 0.0678 | -0.364 | 0.015 | 2.512E-132 | 588.87 |
| *PKLR* | rs6592965 | G | A | 0.4536 | 0.06 | 0.007 | 1.9953E-16 | 73.47 |
| *PKLR* | rs2361710 | T | C | 0.4703 | -0.052 | 0.007 | 1.5849E-12 | 55.18 |
| *PKN3* | rs1354034 | T | C | 0.6041 | 0.079 | 0.008 | 2.5119E-24 | 97.52 |
| *PKN3* | rs13283282 | C | G | 0.1465 | 0.215 | 0.011 | 7.9433E-89 | 382.02 |
| *PLA2G10* | rs10889356 | G | A | 0.3482 | 0.058 | 0.007 | 6.3096E-15 | 68.65 |
| *PLA2G10* | rs6547409 | C | T | 0.0524 | 0.111 | 0.016 | 7.9433E-12 | 48.13 |
| *PLA2G10* | rs1260326 | T | C | 0.6084 | 0.05 | 0.007 | 7.9433E-12 | 51.02 |
| *PLA2G10* | rs149401529 | G | C | 0.0058 | -0.464 | 0.047 | 5.0119E-23 | 97.46 |
| *PLA2G10* | rs28367569 | T | C | 0.0447 | 0.571 | 0.018 | 1E-200 | 1006.30 |
| *PLA2G10* | rs78963197 | T | C | 0.0975 | 0.107 | 0.012 | 1E-18 | 79.51 |
| *PLA2G10* | rs2976388 | G | A | 0.4226 | -0.257 | 0.007 | 1E-200 | 1347.94 |
| *PLA2G10* | rs2519093 | C | T | 0.1843 | -0.112 | 0.009 | 5.0119E-34 | 154.86 |
| *PLA2G10* | rs34481144 | C | T | 0.5015 | -0.111 | 0.007 | 5.0119E-52 | 251.45 |
| *PLA2G10* | rs7232 | T | A | 0.3732 | 0.07 | 0.007 | 7.9433E-21 | 100.00 |
| *PLA2G10* | rs964184 | G | C | 0.8672 | 0.113 | 0.011 | 1.2589E-26 | 105.53 |
| *PLA2G10* | rs568086689 | T | C | 0.0046 | -1.393 | 0.056 | 3.981E-138 | 618.77 |
| *PLA2G10* | rs186021206 | G | A | 0.0064 | 0.379 | 0.049 | 1E-14 | 59.83 |
| *PLA2G10* | rs112001035 | G | A | 0.061 | 0.156 | 0.015 | 1.5849E-24 | 108.16 |
| *PLA2G10* | rs1065853 | G | T | 0.0811 | 0.212 | 0.013 | 3.1623E-58 | 265.94 |
| *PLA2G10* | rs492602 | A | G | 0.5096 | -0.075 | 0.007 | 1.9953E-25 | 114.80 |
| *PLA2G15* | rs2187668 | C | T | 0.1477 | 0.136 | 0.01 | 1.5849E-41 | 184.96 |
| *PLA2G15* | rs9346804 | G | A | 0.1369 | -0.081 | 0.011 | 1E-14 | 54.22 |
| *PLA2G15* | rs139130389 | C | CTA | 0.0726 | -0.094 | 0.014 | 1.5849E-11 | 45.08 |
| *PLA2G15* | rs10778152 | A | G | 0.2877 | -0.236 | 0.008 | 5.012E-184 | 870.25 |
| *PLA2G15* | rs7965471 | A | C | 0.4799 | 0.088 | 0.008 | 3.1623E-31 | 121.00 |
| *PLA2G15* | rs145078947 | G | T | 0.0025 | 1.26 | 0.072 | 1E-68 | 306.25 |
| *PLA2G15* | rs3743588 | G | A | 0.2864 | 0.062 | 0.008 | 6.3096E-14 | 60.06 |
| *PLA2G15* | rs7672 | C | G | 0.7147 | -0.245 | 0.008 | 2.512E-196 | 937.89 |
| *PLA2G15* | rs186021206 | G | A | 0.0064 | 0.46 | 0.05 | 1.5849E-20 | 84.64 |
| *PLA2G15* | rs58542926 | C | T | 0.0754 | 0.123 | 0.014 | 2.5119E-19 | 77.19 |
| *PLA2G1B* | rs1260326 | T | C | 0.6084 | 0.053 | 0.007 | 1.5849E-12 | 57.33 |
| *PLA2G1B* | rs17032925 | T | C | 0.0829 | -0.094 | 0.013 | 2.5119E-12 | 52.28 |
| *PLA2G1B* | rs376333472 | G | A | 0.0247 | 0.172 | 0.024 | 6.3096E-13 | 51.36 |
| *PLA2G1B* | rs1361108 | C | T | 0.4557 | -0.079 | 0.007 | 5.0119E-27 | 127.37 |
| *PLA2G1B* | rs7003060 | T | G | 0.6994 | -0.062 | 0.008 | 1E-14 | 60.06 |
| *PLA2G1B* | rs1372993 | G | A | 0.1162 | -0.164 | 0.012 | 6.3096E-46 | 186.78 |
| *PLA2G1B* | rs116842520 | G | A | 0.0361 | 0.149 | 0.02 | 1E-13 | 55.50 |
| *PLA2G1B* | rs2438988 | A | G | 0.6354 | -0.074 | 0.008 | 1.2589E-20 | 85.56 |
| *PLA2G1B* | rs4751995 | A | G | 0.5191 | -0.079 | 0.007 | 5.0119E-27 | 127.37 |
| *PLA2G1B* | rs61729512 | G | A | 0.1177 | 0.081 | 0.011 | 1E-12 | 54.22 |
| *PLA2G1B* | rs11065078 | C | T | 0.2081 | -0.068 | 0.009 | 3.9811E-14 | 57.09 |
| *PLA2G1B* | rs56994090 | T | C | 0.4171 | -0.051 | 0.007 | 7.9433E-12 | 53.08 |
| *PLA2G1B* | rs72802342 | C | A | 0.0755 | -0.203 | 0.014 | 1.2589E-46 | 210.25 |
| *PLA2G1B* | rs1673931 | T | C | 0.3783 | -0.135 | 0.008 | 1.5849E-71 | 284.77 |
| *PLA2G1B* | rs1468481 | C | T | 0.7678 | 0.062 | 0.009 | 1E-12 | 47.46 |
| *PLA2G1B* | rs62090604 | G | T | 0.077 | 0.094 | 0.014 | 6.3096E-12 | 45.08 |
| *PLA2G1B* | rs681343 | C | T | 0.5088 | 0.104 | 0.007 | 1E-45 | 220.73 |

| | | | | | | | | |
|---|---|---|---|---|---|---|---|---|
| *PLA2G1B* | rs71327329 | A | G | 0.1286 | -0.075 | 0.011 | 7.9433E-12 | 46.49 |
| *PLA2G2A* | rs11573156 | G | C | 0.2283 | 0.731 | 0.009 | 1E-200 | 6597.05 |
| *PLA2G2A* | rs6698842 | C | T | 0.6326 | 0.081 | 0.008 | 1.2589E-26 | 102.52 |
| *PLA2G2A* | rs61729512 | G | A | 0.118 | 0.084 | 0.01 | 2.5119E-16 | 70.56 |
| *PLA2G2A* | rs34931250 | C | T | 0.0604 | 0.189 | 0.014 | 2.5119E-42 | 182.25 |
| *PLA2G2A* | rs112896133 | G | A | 0.0133 | -0.201 | 0.029 | 2.5119E-12 | 48.04 |
| *PLA2G2A* | rs10405357 | T | C | 0.435 | -0.047 | 0.007 | 2.5119E-12 | 45.08 |
| *PLA2G4A* | rs556306120 | G | GCTACAGA. | 0.0832 | 0.13 | 0.014 | 1.9953E-21 | 86.22 |
| *PLA2G4A* | rs1354034 | T | C | 0.6041 | 0.066 | 0.008 | 5.0119E-18 | 68.06 |
| *PLA2G4A* | rs1149577 | T | C | 0.2131 | 0.067 | 0.009 | 1.9953E-13 | 55.42 |
| *PLA2G7* | rs11591147 | G | T | 0.0176 | -0.219 | 0.027 | 5.0119E-16 | 65.79 |
| *PLA2G7* | rs7528419 | A | G | 0.2228 | -0.099 | 0.009 | 5.0119E-31 | 121.00 |
| *PLA2G7* | rs6871667 | G | A | 0.4038 | 0.052 | 0.007 | 1E-12 | 55.18 |
| *PLA2G7* | rs574476364 | C | T | 0.0012 | -1.451 | 0.112 | 3.1623E-38 | 167.84 |
| *PLA2G7* | rs2119690 | G | A | 0.2875 | -0.055 | 0.008 | 2.5119E-12 | 47.27 |
| *PLA2G7* | rs28601761 | C | G | 0.4192 | -0.063 | 0.007 | 5.0119E-18 | 81.00 |
| *PLA2G7* | rs562028 | A | C | 0.6291 | 0.074 | 0.007 | 1.2589E-23 | 111.76 |
| *PLA2G7* | rs1065853 | G | T | 0.0815 | -0.481 | 0.013 | 1E-200 | 1369.00 |
| *PLAT* | rs7765526 | A | G | 0.5411 | 0.05 | 0.007 | 3.1623E-12 | 51.02 |
| *PLAT* | rs796629870 | T | TG | 0.4573 | -0.061 | 0.007 | 7.9433E-18 | 75.94 |
| *PLAT* | rs2020921 | G | A | 0.0252 | -0.28 | 0.023 | 1.9953E-35 | 148.20 |
| *PLAT* | rs4576860 | A | G | 0.5815 | -0.054 | 0.007 | 6.3096E-14 | 59.51 |
| *PLAT* | rs8112983 | T | C | 0.5365 | 0.069 | 0.007 | 1.9953E-22 | 97.16 |
| *PLAU* | rs10194846 | T | C | 0.6141 | 0.056 | 0.007 | 1.9953E-14 | 64.00 |
| *PLAU* | rs17231342 | T | G | 0.4595 | -0.085 | 0.007 | 1.5849E-32 | 147.45 |
| *PLAU* | rs2470750 | A | T | 0.3928 | -0.066 | 0.007 | 3.9811E-19 | 88.90 |
| *PLAU* | rs6858148 | T | C | 0.356 | 0.051 | 0.007 | 7.9433E-12 | 53.08 |
| *PLAU* | rs8176741 | G | A | 0.0611 | 0.172 | 0.015 | 1E-30 | 131.48 |
| *PLAU* | rs369705328 | G | C | 0.1284 | 0.084 | 0.011 | 7.9433E-15 | 58.31 |
| *PLAU* | rs55744193 | G | A | 0.0073 | -0.815 | 0.042 | 1E-83 | 376.54 |
| *PLAU* | rs4055121 | C | T | 0.1351 | -0.146 | 0.011 | 7.9433E-44 | 176.17 |
| *PLAU* | rs186021206 | G | A | 0.0064 | 1.269 | 0.049 | 2.512E-147 | 670.70 |
| *PLAU* | rs9948409 | A | G | 0.7035 | 0.111 | 0.008 | 3.9811E-45 | 192.52 |
| *PLAU* | rs36229204 | C | T | 0.0323 | -0.528 | 0.021 | 7.943E-145 | 632.16 |
| *PLAUR* | rs9378247 | G | A | 0.183 | -0.066 | 0.009 | 2.5119E-12 | 53.78 |
| *PLAUR* | rs8176741 | G | A | 0.0613 | 0.105 | 0.015 | 3.1623E-12 | 49.00 |
| *PLAUR* | rs2688623 | C | T | 0.5588 | 0.108 | 0.007 | 1.2589E-49 | 238.04 |
| *PLAUR* | rs2511241 | C | T | 0.9294 | 0.139 | 0.014 | 5.0119E-23 | 98.58 |
| *PLAUR* | rs7952602 | G | C | 0.1344 | -0.099 | 0.011 | 1.2589E-20 | 81.00 |
| *PLAUR* | rs7137828 | C | T | 0.5159 | -0.054 | 0.007 | 3.9811E-14 | 59.51 |
| *PLAUR* | rs186021206 | G | A | 0.0064 | 0.908 | 0.05 | 7.9433E-75 | 329.79 |
| *PLAUR* | rs2302524 | T | C | 0.166 | -0.177 | 0.01 | 1.5849E-74 | 313.29 |
| *PLB1* | rs80293268 | G | C | 0.0483 | -0.151 | 0.013 | 3.1623E-30 | 134.92 |
| *PLB1* | rs34119886 | A | G | 0.2015 | -0.049 | 0.007 | 3.9811E-13 | 49.00 |
| *PLB1* | rs6753929 | G | C | 0.2534 | -0.987 | 0.008 | 1E-200 | 15221.39 |
| *PLB1* | rs833270 | T | C | 0.6174 | 0.053 | 0.006 | 2.5119E-21 | 78.03 |
| *PLB1* | rs25914 | C | T | 0.1499 | -0.093 | 0.008 | 2.5119E-34 | 135.14 |
| *PLB1* | rs2436396 | A | G | 0.7014 | -0.049 | 0.006 | 1.9953E-16 | 66.69 |
| *PLB1* | rs144826968 | AG | A | 0.116 | 0.083 | 0.008 | 5.0119E-23 | 107.64 |
| *PLB1* | rs12214582 | G | T | 0.6586 | 0.049 | 0.006 | 7.9433E-18 | 66.69 |
| *PLB1* | rs7905367 | G | C | 0.7972 | 0.063 | 0.007 | 3.1623E-21 | 81.00 |
| *PLB1* | rs11220477 | C | T | 0.0416 | -0.113 | 0.013 | 2.5119E-17 | 75.56 |
| *PLB1* | rs7306778 | C | T | 0.171 | -0.065 | 0.007 | 7.9433E-20 | 86.22 |
| *PLB1* | rs11105795 | G | T | 0.2725 | 0.043 | 0.006 | 6.3096E-13 | 51.36 |
| *PLB1* | rs2649998 | T | C | 0.6414 | -0.07 | 0.006 | 7.9433E-36 | 136.11 |
| *PLB1* | rs12957090 | T | C | 0.4863 | 0.036 | 0.005 | 1.2589E-11 | 51.84 |
| *PLB1* | rs12457893 | A | C | 0.4572 | -0.048 | 0.005 | 1.9953E-18 | 92.16 |
| *PLB1* | rs35633871 | G | A | 0.0481 | -0.161 | 0.012 | 5.0119E-38 | 180.01 |
| *PLCB2* | rs1354034 | T | C | 0.6043 | 0.092 | 0.008 | 1.9953E-33 | 132.25 |
| *PLCB2* | rs11242109 | G | T | 0.481 | -0.052 | 0.007 | 2.5119E-12 | 55.18 |
| *PLCB2* | rs342298 | C | T | 0.4553 | -0.069 | 0.007 | 6.3096E-20 | 97.16 |
| *PLCB2* | rs6993770 | A | T | 0.2865 | -0.119 | 0.008 | 3.1623E-47 | 221.27 |
| *PLCB2* | rs4924450 | G | A | 0.7057 | -0.153 | 0.008 | 6.3096E-77 | 365.77 |
| *PLEKHO1* | rs72692857 | T | C | 0.1654 | 0.078 | 0.01 | 3.9811E-15 | 60.84 |
| *PLEKHO1* | rs1354034 | T | C | 0.6043 | 0.078 | 0.008 | 6.3096E-25 | 95.06 |
| *PLG* | rs1260326 | T | C | 0.6082 | -0.086 | 0.007 | 5.0119E-32 | 150.94 |
| *PLG* | rs9898 | C | T | 0.3286 | 0.074 | 0.008 | 1.9953E-22 | 85.56 |
| *PLG* | rs9274442 | G | C | 0.2828 | -0.067 | 0.009 | 6.3096E-13 | 55.42 |

| | | | | | | | | |
|---|---|---|---|---|---|---|---|---|
| *PLG* | rs73015965 | A | G | 0.0064 | -2.183 | 0.051 | 1E-200 | 1832.18 |
| *PLG* | rs17145738 | C | T | 0.1214 | -0.084 | 0.011 | 1.2589E-14 | 58.31 |
| *PLIN1* | rs150004289 | G | A | 0.0029 | -0.76 | 0.07 | 5.0119E-27 | 117.88 |
| *PLIN3* | rs6993770 | A | T | 0.2866 | -0.063 | 0.008 | 5.0119E-14 | 62.02 |
| *PLIN3* | rs262557 | C | T | 0.7574 | 0.074 | 0.009 | 1.2589E-16 | 67.60 |
| *PLPBP* | rs1354034 | T | C | 0.6037 | 0.067 | 0.008 | 7.9433E-19 | 70.14 |
| *PLPBP* | rs6993770 | A | T | 0.2866 | -0.062 | 0.008 | 7.9433E-14 | 60.06 |
| *PLSCR3* | rs7924036 | G | T | 0.5011 | 0.056 | 0.007 | 6.3096E-14 | 64.00 |
| *PLSCR3* | rs2277641 | G | A | 0.0216 | -0.224 | 0.026 | 1E-17 | 74.22 |
| *PLTP* | rs34211178 | G | A | 0.4399 | -0.078 | 0.007 | 3.1623E-28 | 124.16 |
| *PLTP* | rs3916027 | G | A | 0.2636 | 0.057 | 0.008 | 6.3096E-13 | 50.77 |
| *PLTP* | rs13284054 | T | C | 0.1244 | -0.075 | 0.011 | 3.9811E-12 | 46.49 |
| *PLTP* | rs174533 | G | A | 0.3463 | -0.058 | 0.007 | 1.9953E-15 | 68.65 |
| *PLTP* | rs78689694 | G | C | 0.1325 | -0.09 | 0.01 | 3.1623E-18 | 81.00 |
| *PLTP* | rs147233090 | C | T | 0.0235 | -0.174 | 0.023 | 5.0119E-14 | 57.23 |
| *PLTP* | rs35853021 | G | T | 0.3563 | 0.096 | 0.007 | 1.2589E-39 | 188.08 |
| *PLTP* | rs247617 | C | A | 0.3245 | 0.125 | 0.007 | 1.2589E-62 | 318.88 |
| *PLTP* | rs4986970 | A | T | 0.0352 | -0.132 | 0.019 | 3.1623E-12 | 48.27 |
| *PLTP* | rs186021206 | G | A | 0.0064 | 0.995 | 0.048 | 3.1623E-95 | 429.70 |
| *PLTP* | rs6073958 | T | C | 0.1987 | -0.502 | 0.009 | 1E-200 | 3111.16 |
| *PLXDC1* | rs3775298 | A | G | 0.5133 | -0.073 | 0.007 | 7.9433E-23 | 108.76 |
| *PLXDC1* | rs11386832 | T | TA | 0.7375 | 0.128 | 0.008 | 5.0119E-53 | 256.00 |
| *PLXDC1* | rs885122 | A | G | 0.3951 | -0.053 | 0.008 | 3.1623E-12 | 43.89 |
| *PLXDC1* | rs79428469 | T | C | 0.2236 | 0.063 | 0.009 | 2.5119E-12 | 49.00 |
| *PLXDC1* | rs142988850 | A | G | 0.005 | 0.488 | 0.052 | 1.2589E-20 | 88.07 |
| *PLXDC1* | rs62076601 | C | T | 0.0492 | -0.535 | 0.018 | 5.012E-198 | 883.41 |
| *PLXDC2* | rs9332635 | T | C | 0.271 | 0.093 | 0.008 | 3.9811E-29 | 135.14 |
| *PLXDC2* | rs10185914 | T | C | 0.0307 | -0.475 | 0.022 | 5.012E-108 | 466.17 |
| *PLXDC2* | rs1486236 | C | A | 0.3732 | 0.069 | 0.008 | 3.1623E-19 | 74.39 |
| *PLXDC2* | rs34928543 | G | C | 0.193 | -0.273 | 0.009 | 3.981E-182 | 920.11 |
| *PLXDC2* | rs2519093 | C | T | 0.1844 | -0.19 | 0.01 | 3.9811E-88 | 361.00 |
| *PLXDC2* | rs7919659 | G | C | 0.2008 | 0.086 | 0.009 | 1.5849E-20 | 91.31 |
| *PLXDC2* | rs186021206 | G | A | 0.0065 | 0.962 | 0.05 | 2.5119E-81 | 370.18 |
| *PLXDC2* | rs112001035 | G | A | 0.0612 | -0.147 | 0.016 | 1.2589E-20 | 84.41 |
| *PLXNA4* | rs1354034 | T | C | 0.6043 | 0.099 | 0.008 | 1E-37 | 153.14 |
| *PLXNA4* | rs11242109 | G | T | 0.4816 | -0.063 | 0.008 | 1E-16 | 62.02 |
| *PLXNA4* | rs6467443 | G | T | 0.2167 | 0.104 | 0.009 | 3.1623E-29 | 133.53 |
| *PLXNA4* | rs6993770 | A | T | 0.2869 | -0.107 | 0.008 | 7.9433E-38 | 178.89 |
| *PLXNA4* | rs892090 | T | G | 0.8337 | 0.081 | 0.01 | 1.5849E-15 | 65.61 |
| *PLXNB2* | rs7523508 | A | G | 0.2768 | -0.051 | 0.007 | 1.2589E-13 | 53.08 |
| *PLXNB2* | rs1260326 | T | C | 0.6086 | 0.05 | 0.006 | 3.1623E-15 | 69.44 |
| *PLXNB2* | rs1126735 | A | G | 0.3686 | 0.053 | 0.006 | 1.5849E-16 | 78.03 |
| *PLXNB2* | rs9269965 | A | G | 0.3937 | -0.055 | 0.007 | 3.1623E-15 | 61.73 |
| *PLXNB2* | rs2519093 | C | T | 0.1845 | -0.118 | 0.008 | 7.9433E-49 | 217.56 |
| *PLXNB2* | rs56278466 | T | G | 0.6613 | 0.189 | 0.007 | 6.31E-181 | 729.00 |
| *PLXNB2* | rs112635299 | G | T | 0.0217 | 0.146 | 0.021 | 6.3096E-12 | 48.34 |
| *PLXNB2* | rs186021206 | G | A | 0.0064 | 0.315 | 0.043 | 1.5849E-13 | 53.66 |
| *PLXNB2* | rs1801689 | A | C | 0.0302 | 0.145 | 0.018 | 1E-15 | 64.89 |
| *PLXNB2* | rs868876 | A | T | 0.3006 | -0.059 | 0.007 | 2.5119E-18 | 71.04 |
| *PLXNB2* | rs11547731 | T | C | 0.6528 | -0.713 | 0.008 | 1E-200 | 7943.27 |
| *PLXNB3* | rs9402686 | G | A | 0.2713 | 0.061 | 0.008 | 6.3096E-13 | 58.14 |
| *PLXNB3* | rs6993770 | A | T | 0.2867 | -0.058 | 0.008 | 2.5119E-12 | 52.56 |
| *PLXNB3* | rs409801 | T | C | 0.5073 | 0.059 | 0.008 | 3.9811E-15 | 54.39 |
| *PLXNB3* | rs4502008 | G | T | 0.5216 | -0.061 | 0.008 | 7.9433E-16 | 58.14 |
| *PLXNB3* | rs111338191 | A | AAATT | 0.5214 | -0.068 | 0.008 | 3.1623E-19 | 72.25 |
| *PLXNB3* | rs892090 | T | G | 0.8339 | 0.077 | 0.01 | 5.0119E-14 | 59.29 |
| *PM20D1* | rs760077 | A | T | 0.6052 | -0.074 | 0.008 | 1.9953E-21 | 85.56 |
| *PM20D1* | rs112268616 | T | C | 0.3398 | -0.84 | 0.008 | 1E-200 | 11025.00 |
| *PM20D1* | rs11680123 | A | G | 0.5929 | 0.124 | 0.006 | 5.0119E-97 | 427.11 |
| *PM20D1* | rs121908120 | T | A | 0.0276 | -0.121 | 0.018 | 1.5849E-11 | 45.19 |
| *PM20D1* | rs11707641 | T | C | 0.1621 | 0.057 | 0.008 | 2.5119E-13 | 50.77 |
| *PM20D1* | rs2519093 | C | T | 0.1844 | -0.057 | 0.007 | 3.1623E-14 | 66.31 |
| *PM20D1* | rs12764992 | G | A | 0.5817 | -0.046 | 0.006 | 1E-14 | 58.78 |
| *PM20D1* | rs34560261 | C | T | 0.1698 | -0.062 | 0.008 | 1.9953E-15 | 60.06 |
| *PMM2* | rs34258285 | A | C | 0.0255 | -0.68 | 0.024 | 5.012E-172 | 802.78 |
| *PMVK* | rs114951074 | C | T | 0.0074 | -0.658 | 0.047 | 2.5119E-45 | 196.00 |
| *PMVK* | rs1354034 | T | C | 0.6034 | 0.056 | 0.008 | 6.3096E-13 | 49.00 |
| *PMVK* | rs342298 | C | T | 0.4543 | -0.07 | 0.008 | 6.3096E-20 | 76.56 |

| | | | | | | | | |
|---|---|---|---|---|---|---|---|---|
| *PMVK* | rs6993770 | A | T | 0.2869 | -0.066 | 0.008 | 5.0119E-15 | 68.06 |
| *PMVK* | rs7896518 | A | G | 0.4244 | 0.054 | 0.008 | 3.9811E-12 | 45.56 |
| *PMVK* | rs7297265 | A | G | 0.4433 | 0.054 | 0.008 | 5.0119E-12 | 45.56 |
| *PNLIP* | rs333947 | G | A | 0.1487 | -0.078 | 0.01 | 3.9811E-14 | 60.84 |
| *PNLIP* | rs2487569 | A | T | 0.113 | -0.082 | 0.012 | 1.9953E-12 | 46.69 |
| *PNLIP* | rs17032925 | T | C | 0.0828 | -0.178 | 0.013 | 2.5119E-40 | 187.48 |
| *PNLIP* | rs66473811 | T | C | 0.1658 | 0.083 | 0.01 | 6.3096E-17 | 68.89 |
| *PNLIP* | rs10213531 | A | G | 0.3436 | -0.061 | 0.008 | 3.1623E-15 | 58.14 |
| *PNLIP* | rs2854008 | G | A | 0.3036 | -0.06 | 0.008 | 1E-13 | 56.25 |
| *PNLIP* | rs72280460 | AAAT | A | 0.6723 | -0.061 | 0.008 | 1.9953E-14 | 58.14 |
| *PNLIP* | rs6571015 | G | A | 0.7907 | 0.146 | 0.009 | 1.5849E-58 | 263.16 |
| *PNLIP* | rs9482772 | T | C | 0.4504 | 0.084 | 0.007 | 1E-29 | 144.00 |
| *PNLIP* | rs572393412 | C | CA | 0.3096 | 0.06 | 0.009 | 1.5849E-12 | 44.44 |
| *PNLIP* | rs4921967 | G | T | 0.6868 | -0.079 | 0.008 | 1.5849E-23 | 97.52 |
| *PNLIP* | rs4733612 | G | A | 0.7327 | 0.06 | 0.008 | 3.1623E-13 | 56.25 |
| *PNLIP* | rs8176749 | C | T | 0.0612 | 0.503 | 0.015 | 1E-200 | 1124.48 |
| *PNLIP* | rs529700 | T | C | 0.5147 | 0.075 | 0.009 | 2.5119E-18 | 69.44 |
| *PNLIP* | rs174548 | C | G | 0.3145 | 0.066 | 0.008 | 3.9811E-17 | 68.06 |
| *PNLIP* | rs61910478 | A | T | 0.2688 | -0.075 | 0.008 | 6.3096E-20 | 87.89 |
| *PNLIP* | rs72802342 | C | A | 0.0756 | -0.283 | 0.014 | 7.9433E-89 | 408.62 |
| *PNLIP* | rs1673931 | T | C | 0.378 | -0.123 | 0.008 | 3.1623E-59 | 236.39 |
| *PNLIP* | rs601338 | G | A | 0.5088 | 0.168 | 0.007 | 3.981E-114 | 576.00 |
| *PNLIPRP1* | rs35004807 | T | TGTAA | 0.3449 | 0.061 | 0.008 | 1.5849E-15 | 58.14 |
| *PNLIPRP1* | rs2027447 | T | C | 0.1111 | -0.095 | 0.012 | 1.9953E-16 | 62.67 |
| *PNLIPRP1* | rs11724862 | T | C | 0.2363 | -0.068 | 0.009 | 3.9811E-15 | 57.09 |
| *PNLIPRP1* | rs3215532 | G | GA | 0.2528 | 0.079 | 0.008 | 1E-21 | 97.52 |
| *PNLIPRP1* | rs35536837 | C | CT | 0.4519 | -0.079 | 0.007 | 1.2589E-27 | 127.37 |
| *PNLIPRP1* | rs7800611 | A | G | 0.9537 | 0.126 | 0.017 | 1.9953E-13 | 54.93 |
| *PNLIPRP1* | rs6966838 | T | C | 0.3677 | 0.059 | 0.007 | 1.9953E-15 | 71.04 |
| *PNLIPRP1* | rs3812304 | G | A | 0.2323 | -0.063 | 0.008 | 1.5849E-13 | 62.02 |
| *PNLIPRP1* | rs2131181 | C | T | 0.6822 | -0.08 | 0.008 | 3.9811E-25 | 100.00 |
| *PNLIPRP1* | rs505922 | T | C | 0.3173 | 0.074 | 0.008 | 1E-21 | 85.56 |
| *PNLIPRP1* | rs28456 | A | G | 0.3146 | 0.059 | 0.008 | 1.5849E-14 | 54.39 |
| *PNLIPRP1* | rs686056 | G | A | 0.3227 | -0.089 | 0.008 | 3.1623E-31 | 123.77 |
| *PNLIPRP1* | rs4883568 | A | G | 0.2312 | -0.064 | 0.009 | 7.9433E-14 | 50.57 |
| *PNLIPRP1* | rs9579128 | T | C | 0.4106 | -0.05 | 0.007 | 7.9433E-12 | 51.02 |
| *PNLIPRP1* | rs72802342 | C | A | 0.0757 | -0.663 | 0.014 | 1E-200 | 2242.70 |
| *PNLIPRP1* | rs1126464 | G | C | 0.2441 | -0.099 | 0.009 | 1E-30 | 121.00 |
| *PNLIPRP1* | rs17138478 | C | A | 0.1305 | 0.096 | 0.011 | 1.9953E-19 | 76.17 |
| *PNLIPRP1* | rs62090602 | A | T | 0.0769 | 0.097 | 0.013 | 5.0119E-13 | 55.67 |
| *PNLIPRP1* | rs2836255 | A | G | 0.5161 | 0.053 | 0.007 | 1.5849E-13 | 57.33 |
| *PNLIPRP1* | rs80184311 | G | C | 0.1057 | 0.08 | 0.012 | 3.9811E-12 | 44.44 |
| *PNLIPRP2* | rs8176741 | G | A | 0.0613 | 0.205 | 0.01 | 3.1623E-95 | 420.25 |
| *PNLIPRP2* | rs56278466 | T | G | 0.6616 | 0.057 | 0.008 | 1E-12 | 50.77 |
| *PNLIPRP2* | rs4751995 | A | G | 0.5188 | 1.136 | 0.008 | 1E-200 | 20164.00 |
| *PNLIPRP2* | rs72802342 | C | A | 0.0758 | -0.119 | 0.009 | 5.0119E-39 | 174.83 |
| *PNLIPRP2* | rs601338 | G | A | 0.5096 | 0.16 | 0.005 | 1E-200 | 1024.00 |
| *PNMA1* | rs342293 | C | G | 0.4595 | -0.055 | 0.008 | 5.0119E-13 | 47.27 |
| *PNMA1* | rs113315674 | G | A | 0.3647 | 0.057 | 0.008 | 6.3096E-13 | 50.77 |
| *PNMA1* | rs140960424 | G | GGAAAGAG | 0.7353 | 0.173 | 0.009 | 3.1623E-88 | 369.49 |
| *PNMA1* | rs11170874 | A | G | 0.3301 | 0.056 | 0.008 | 6.3096E-12 | 49.00 |
| *PNMA1* | rs35564315 | C | A | 0.1668 | 0.071 | 0.01 | 5.0119E-12 | 50.41 |
| *PNPT1* | rs9394070 | C | A | 0.0647 | -0.143 | 0.016 | 3.1623E-20 | 79.88 |
| *PNPT1* | rs6001857 | T | C | 0.1668 | -0.084 | 0.01 | 1.5849E-16 | 70.56 |
| *PODXL* | rs767298 | G | T | 0.6343 | -0.161 | 0.008 | 1E-97 | 405.02 |
| *PODXL* | rs3212298 | C | T | 0.0439 | -0.549 | 0.018 | 1E-200 | 930.25 |
| *PODXL* | rs590666 | T | C | 0.5191 | -0.098 | 0.008 | 1E-38 | 150.06 |
| *PODXL* | rs2519093 | C | T | 0.1845 | -0.549 | 0.01 | 1E-200 | 3014.01 |
| *PODXL* | rs34434834 | G | A | 0.0335 | 0.332 | 0.02 | 1.9953E-61 | 275.56 |
| *PODXL* | rs186021206 | G | A | 0.0063 | 0.863 | 0.05 | 1E-66 | 297.91 |
| *PODXL2* | rs35196180 | C | CG | 0.1368 | 0.078 | 0.011 | 7.9433E-13 | 50.28 |
| *PODXL2* | rs61747728 | C | T | 0.0379 | 0.129 | 0.019 | 1.5849E-11 | 46.10 |
| *PODXL2* | rs1047891 | C | A | 0.3127 | 0.055 | 0.008 | 2.5119E-12 | 47.27 |
| *PODXL2* | rs781656 | G | A | 0.1836 | 0.169 | 0.009 | 3.9811E-71 | 352.60 |
| *PODXL2* | rs1129187 | G | T | 0.4529 | -0.051 | 0.007 | 7.9433E-12 | 53.08 |
| *PODXL2* | rs635634 | C | T | 0.1841 | 0.073 | 0.01 | 2.5119E-14 | 53.29 |
| *PODXL2* | rs74841302 | G | A | 0.0171 | -0.198 | 0.028 | 1.5849E-12 | 50.01 |
| *PODXL2* | rs4781978 | C | G | 0.2121 | 0.117 | 0.009 | 3.9811E-38 | 169.00 |

| | | | | | | | | |
|---|---|---|---|---|---|---|---|---|
| *PODXL2* | rs11869940 | T | A | 0.4955 | 0.051 | 0.007 | 6.3096E-12 | 53.08 |
| *PODXL2* | rs6504649 | C | G | 0.388 | -0.117 | 0.008 | 2.5119E-55 | 213.89 |
| *PODXL2* | rs55639531 | C | G | 0.2855 | 0.058 | 0.008 | 7.9433E-13 | 52.56 |
| *PODXL2* | rs45512696 | C | T | 0.1757 | -0.073 | 0.01 | 3.1623E-14 | 53.29 |
| *POF1B* | rs62142192 | G | T | 0.202 | 0.083 | 0.01 | 2.5119E-18 | 68.89 |
| *POF1B* | rs11898628 | A | G | 0.1502 | 0.077 | 0.011 | 6.3096E-13 | 49.00 |
| *POF1B* | rs13208948 | C | T | 0.2636 | -0.065 | 0.009 | 1.2589E-12 | 52.16 |
| *POLR2F* | rs7137828 | C | T | 0.5157 | -0.054 | 0.007 | 2.5119E-13 | 59.51 |
| *POMC* | rs6680345 | C | T | 0.5076 | 0.123 | 0.007 | 1.5849E-63 | 308.76 |
| *POMC* | rs3754861 | A | C | 0.0708 | 0.145 | 0.014 | 1E-23 | 107.27 |
| *POMC* | rs2594950 | G | C | 0.3445 | 0.078 | 0.008 | 6.3096E-24 | 95.06 |
| *POMC* | rs9472040 | A | G | 0.3183 | -0.053 | 0.008 | 1E-11 | 43.89 |
| *POMC* | rs13199674 | G | A | 0.4428 | -0.051 | 0.007 | 2.5119E-12 | 53.08 |
| *POMC* | rs8045100 | C | T | 0.5432 | -0.18 | 0.007 | 1.995E-133 | 661.22 |
| *POMC* | rs877127 | T | A | 0.3223 | -0.065 | 0.008 | 6.3096E-17 | 66.02 |
| *POMC* | rs145530718 | G | C | 0.02 | 0.346 | 0.026 | 3.1623E-39 | 177.09 |
| *POMC* | rs4408777 | A | G | 0.438 | -0.087 | 0.007 | 1E-32 | 154.47 |
| *PON1* | rs6671730 | G | A | 0.4367 | 0.064 | 0.006 | 1.9953E-27 | 113.78 |
| *PON1* | rs705379 | G | A | 0.4774 | -0.757 | 0.008 | 1E-200 | 8953.89 |
| *PON1* | rs305 | A | G | 0.2344 | 0.052 | 0.007 | 6.3096E-14 | 55.18 |
| *PON1* | rs173539 | C | T | 0.3316 | 0.06 | 0.006 | 3.9811E-22 | 100.00 |
| *PON2* | rs79654026 | A | G | 0.181 | 0.081 | 0.008 | 7.9433E-24 | 102.52 |
| *PON2* | rs6961624 | T | C | 0.2384 | -0.903 | 0.009 | 1E-200 | 10066.78 |
| *PON2* | rs28929474 | C | T | 0.0208 | 0.206 | 0.021 | 1.5849E-22 | 96.23 |
| *PON2* | rs58542926 | C | T | 0.0756 | 0.154 | 0.011 | 5.0119E-42 | 196.00 |
| *PON3* | rs16840639 | T | C | 0.1947 | 0.085 | 0.009 | 1.9953E-22 | 89.20 |
| *PON3* | rs149867961 | T | C | 0.0264 | -1.051 | 0.023 | 1E-200 | 2088.09 |
| *PON3* | rs4240624 | G | A | 0.908 | 0.094 | 0.012 | 6.3096E-15 | 61.36 |
| *PON3* | rs3208305 | A | T | 0.2861 | 0.058 | 0.008 | 6.3096E-14 | 52.56 |
| *PON3* | rs1077835 | A | G | 0.222 | 0.057 | 0.008 | 1.2589E-11 | 50.77 |
| *PON3* | rs247617 | C | A | 0.3243 | 0.071 | 0.007 | 3.9811E-21 | 102.88 |
| *PON3* | rs149394327 | G | C | 0.0301 | 0.189 | 0.021 | 3.1623E-20 | 81.00 |
| *PON3* | rs34415062 | G | A | 0.1792 | 0.16 | 0.009 | 1.2589E-68 | 316.05 |
| *POSTN* | rs59410274 | G | C | 0.0064 | -0.317 | 0.045 | 2.5119E-12 | 49.62 |
| *POSTN* | rs74735511 | A | G | 0.0887 | -0.094 | 0.013 | 1.5849E-13 | 52.28 |
| *POSTN* | rs4705926 | C | T | 0.4638 | -0.058 | 0.007 | 1.2589E-15 | 68.65 |
| *POSTN* | rs2389858 | T | A | 0.4903 | -0.079 | 0.007 | 1.2589E-27 | 127.37 |
| *POSTN* | rs11380865 | T | TA | 0.3845 | -0.064 | 0.008 | 1.5849E-17 | 64.00 |
| *POSTN* | rs1609860 | C | A | 0.0815 | 0.41 | 0.013 | 1E-200 | 994.67 |
| *PPBP* | rs12086222 | G | C | 0.111 | 0.085 | 0.012 | 1.2589E-12 | 50.17 |
| *PPBP* | rs13412535 | G | A | 0.2307 | -0.072 | 0.009 | 5.0119E-16 | 64.00 |
| *PPBP* | rs7618405 | C | A | 0.205 | -0.065 | 0.009 | 3.9811E-12 | 52.16 |
| *PPBP* | rs1354034 | T | C | 0.6045 | -0.076 | 0.008 | 3.1623E-23 | 90.25 |
| *PPBP* | rs202224981 | C | T | 0.0017 | -1.18 | 0.092 | 1E-37 | 164.51 |
| *PPBP* | rs10016018 | A | T | 0.3973 | -0.069 | 0.008 | 1.9953E-19 | 74.39 |
| *PPBP* | rs2516471 | G | A | 0.0633 | -0.12 | 0.016 | 1E-14 | 56.25 |
| *PPBP* | rs6961069 | C | T | 0.4031 | 0.071 | 0.008 | 1.9953E-20 | 78.77 |
| *PPBP* | rs6993770 | A | T | 0.2865 | -0.128 | 0.008 | 5.0119E-54 | 256.00 |
| *PPBP* | rs10820606 | A | C | 0.2297 | -0.063 | 0.009 | 3.1623E-12 | 49.00 |
| *PPBP* | rs7080386 | C | A | 0.4117 | 0.127 | 0.008 | 5.0119E-62 | 252.02 |
| *PPBP* | rs12445050 | C | T | 0.1384 | 0.102 | 0.011 | 3.9811E-21 | 85.98 |
| *PPBP* | rs892090 | T | G | 0.8341 | 0.163 | 0.01 | 5.0119E-58 | 265.69 |
| *PPBP* | rs6081569 | G | C | 0.3686 | 0.072 | 0.008 | 1.2589E-20 | 81.00 |
| *PPCDC* | rs1354034 | T | C | 0.6042 | 0.051 | 0.007 | 3.1623E-12 | 53.08 |
| *PPCDC* | rs12591994 | A | G | 0.0645 | 0.67 | 0.015 | 1E-200 | 1995.11 |
| *PPIB* | rs1354034 | T | C | 0.6038 | 0.054 | 0.008 | 3.1623E-12 | 45.56 |
| *PPIB* | rs6993770 | A | T | 0.2866 | -0.076 | 0.008 | 6.3096E-20 | 90.25 |
| *PPIB* | rs892090 | T | G | 0.8341 | 0.074 | 0.01 | 3.1623E-13 | 54.76 |
| *PPIF* | rs6993770 | A | T | 0.2863 | -0.064 | 0.008 | 1.9953E-14 | 64.00 |
| *PPL* | rs151830 | G | A | 0.6059 | 0.056 | 0.008 | 7.9433E-13 | 49.00 |
| *PPL* | rs61499243 | C | T | 0.1115 | 0.137 | 0.012 | 1.2589E-28 | 130.34 |
| *PPM1B* | rs4548048 | C | T | 0.1033 | 0.12 | 0.013 | 1.2589E-19 | 85.21 |
| *PPM1F* | rs1354034 | T | C | 0.6045 | 0.059 | 0.008 | 1.2589E-14 | 54.39 |
| *PPM1F* | rs342298 | C | T | 0.4545 | -0.056 | 0.008 | 1.2589E-13 | 49.00 |
| *PPM1F* | rs6993770 | A | T | 0.2866 | -0.07 | 0.008 | 3.1623E-17 | 76.56 |
| *PPM1F* | rs5750281 | C | T | 0.043 | -0.126 | 0.018 | 7.9433E-12 | 49.00 |
| *PPME1* | rs3782123 | C | A | 0.7336 | 0.058 | 0.008 | 3.9811E-12 | 52.56 |
| *PPP1R12A* | rs68066031 | T | C | 0.2299 | 0.109 | 0.009 | 6.3096E-34 | 146.68 |

| | | | | | | | | |
|---|---|---|---|---|---|---|---|---|
| *PPP1R12A* | rs1354034 | T | C | 0.6041 | 0.055 | 0.008 | 7.9433E-13 | 47.27 |
| *PPP1R12A* | rs17622656 | G | A | 0.3883 | -0.052 | 0.008 | 1.5849E-11 | 42.25 |
| *PPP1R12A* | rs342298 | C | T | 0.4549 | -0.067 | 0.008 | 6.3096E-19 | 70.14 |
| *PPP1R12A* | rs6993770 | A | T | 0.2861 | -0.057 | 0.008 | 3.9811E-12 | 50.77 |
| *PPP1R12B* | rs68066031 | T | C | 0.2296 | 0.073 | 0.009 | 1.9953E-15 | 65.79 |
| *PPP1R14A* | rs342298 | C | T | 0.4549 | -0.064 | 0.008 | 1.5849E-17 | 64.00 |
| *PPP1R14A* | rs6993770 | A | T | 0.2864 | -0.056 | 0.008 | 1E-11 | 49.00 |
| *PPP1R14A* | rs71354995 | A | G | 0.2562 | -0.226 | 0.009 | 1.585E-151 | 630.57 |
| *PPP1R14D* | rs9271657 | T | C | 0.214 | 0.075 | 0.01 | 7.9433E-15 | 56.25 |
| *PPP1R14D* | rs11855641 | A | G | 0.3401 | 0.061 | 0.008 | 1.5849E-14 | 58.14 |
| *PPP1R2* | rs1354034 | T | C | 0.6037 | 0.079 | 0.008 | 3.1623E-25 | 97.52 |
| *PPP1R2* | rs7080386 | C | A | 0.4116 | 0.053 | 0.008 | 2.5119E-12 | 43.89 |
| *PPP1R2* | rs11502185 | T | C | 0.2595 | 0.074 | 0.01 | 1.9953E-14 | 54.76 |
| *PPP1R2* | rs892090 | T | G | 0.8338 | 0.069 | 0.01 | 6.3096E-12 | 47.61 |
| *PPP1R9B* | rs1354034 | T | C | 0.6041 | 0.068 | 0.007 | 1.9953E-19 | 94.37 |
| *PPP1R9B* | rs342296 | G | A | 0.4565 | -0.051 | 0.007 | 3.9811E-12 | 53.08 |
| *PPP2R5A* | rs351409 | A | G | 0.0208 | -0.301 | 0.026 | 2.5119E-30 | 134.03 |
| *PPP2R5A* | rs6993770 | A | T | 0.2866 | -0.064 | 0.008 | 7.9433E-15 | 64.00 |
| *PPP3R1* | rs188468174 | C | T | 0.0142 | -0.315 | 0.032 | 6.3096E-23 | 96.90 |
| *PPP3R1* | rs1354034 | T | C | 0.6041 | 0.054 | 0.008 | 1.2589E-12 | 45.56 |
| *PPP3R1* | rs55768089 | T | A | 0.3104 | 0.105 | 0.008 | 5.0119E-38 | 172.27 |
| *PPP3R1* | rs915651 | G | A | 0.1303 | -0.086 | 0.011 | 1.2589E-14 | 61.12 |
| *PPP3R1* | rs9414801 | G | A | 0.5334 | -0.064 | 0.008 | 1.9953E-17 | 64.00 |
| *PPY* | rs1493691 | G | A | 0.1083 | -0.106 | 0.012 | 2.5119E-19 | 78.03 |
| *PPY* | rs73174306 | A | T | 0.0415 | 0.415 | 0.018 | 1.259E-113 | 531.56 |
| *PPY* | rs6451770 | T | G | 0.3795 | -0.063 | 0.007 | 3.1623E-17 | 81.00 |
| *PPY* | rs9348441 | T | A | 0.2606 | -0.066 | 0.008 | 1.9953E-15 | 68.06 |
| *PPY* | rs9375435 | C | T | 0.453 | -0.067 | 0.007 | 3.9811E-20 | 91.61 |
| *PPY* | rs184163936 | T | C | 0.0066 | -0.366 | 0.047 | 5.0119E-15 | 60.64 |
| *PPY* | rs2438988 | A | G | 0.6351 | -0.217 | 0.008 | 3.162E-163 | 735.77 |
| *PPY* | rs375124982 | TTGTG | T | 0.3106 | -0.057 | 0.008 | 1E-12 | 50.77 |
| *PPY* | rs9320010 | A | G | 0.3994 | 0.052 | 0.007 | 3.1623E-12 | 55.18 |
| *PRAP1* | rs687339 | C | T | 0.7711 | 0.084 | 0.008 | 1.2589E-23 | 110.25 |
| *PRAP1* | rs782134971 | GAAACTGC | G | 0.2544 | -0.132 | 0.008 | 1.2589E-60 | 272.25 |
| *PRAP1* | rs143674922 | G | A | 0.0182 | -1.298 | 0.028 | 1E-200 | 2148.98 |
| *PRAP1* | rs681343 | C | T | 0.5093 | -0.119 | 0.007 | 1E-63 | 289.00 |
| *PRCP* | rs3010061 | C | T | 0.2001 | 0.061 | 0.009 | 7.9433E-12 | 45.94 |
| *PRCP* | rs11125916 | C | A | 0.1947 | 0.065 | 0.009 | 2.5119E-13 | 52.16 |
| *PRCP* | rs13111888 | G | A | 0.1776 | 0.064 | 0.009 | 1E-11 | 50.57 |
| *PRCP* | rs142032527 | A | AAT | 0.2374 | -0.061 | 0.008 | 5.0119E-13 | 58.14 |
| *PRCP* | rs7834466 | G | T | 0.6099 | -0.067 | 0.007 | 5.0119E-20 | 91.61 |
| *PRCP* | rs10104003 | C | T | 0.2103 | 0.166 | 0.009 | 3.9811E-81 | 340.20 |
| *PRCP* | rs10116966 | T | A | 0.3099 | -0.081 | 0.008 | 6.3096E-26 | 102.52 |
| *PRCP* | rs56278466 | T | G | 0.6615 | 0.141 | 0.007 | 1E-79 | 405.73 |
| *PRCP* | rs543233230 | C | A | 0.0019 | -1.544 | 0.082 | 7.9433E-80 | 354.54 |
| *PRCP* | rs540730 | T | C | 0.7507 | -0.077 | 0.008 | 1E-20 | 92.64 |
| *PRCP* | rs4764823 | G | A | 0.2883 | -0.22 | 0.008 | 7.943E-168 | 756.25 |
| *PRCP* | rs145078947 | G | T | 0.0026 | 1.052 | 0.07 | 3.1623E-51 | 225.86 |
| *PRDX1* | rs2356552 | T | C | 0.8982 | 0.109 | 0.012 | 3.1623E-18 | 82.51 |
| *PRDX1* | rs6993770 | A | T | 0.2862 | -0.066 | 0.008 | 1.5849E-15 | 68.06 |
| *PRDX2* | rs11759553 | A | T | 0.2728 | -0.063 | 0.008 | 5.0119E-15 | 62.02 |
| *PRDX3* | rs7358070 | G | C | 0.3246 | -0.065 | 0.008 | 2.5119E-15 | 66.02 |
| *PRDX5* | rs77269065 | G | A | 0.0054 | 0.98 | 0.053 | 3.1623E-77 | 341.90 |
| *PRDX6* | rs33951697 | C | T | 0.1849 | 0.174 | 0.01 | 6.3096E-74 | 302.76 |
| *PRELP* | rs41313926 | G | A | 0.0103 | -2.271 | 0.035 | 1E-200 | 4210.16 |
| *PRG2* | rs1260326 | T | C | 0.6083 | 0.052 | 0.008 | 7.9433E-12 | 42.25 |
| *PRG2* | rs1186706 | G | T | 0.4906 | -0.135 | 0.007 | 3.9811E-75 | 371.94 |
| *PRG2* | rs10208293 | G | A | 0.2801 | -0.057 | 0.008 | 3.1623E-12 | 50.77 |
| *PRG2* | rs11689257 | G | A | 0.069 | -0.102 | 0.015 | 5.0119E-12 | 46.24 |
| *PRG2* | rs1047891 | C | A | 0.3128 | 0.058 | 0.008 | 3.1623E-13 | 52.56 |
| *PRG2* | rs12619285 | A | G | 0.2655 | -0.063 | 0.008 | 2.5119E-14 | 62.02 |
| *PRG2* | rs900175 | T | G | 0.3441 | -0.065 | 0.008 | 7.9433E-17 | 66.02 |
| *PRG2* | rs559772038 | CCT | C | 0.1745 | 0.12 | 0.01 | 3.9811E-35 | 144.00 |
| *PRG2* | rs4328821 | A | G | 0.1069 | -0.244 | 0.012 | 1.9953E-94 | 413.44 |
| *PRG2* | rs11405616 | C | CA | 0.6058 | 0.052 | 0.008 | 5.0119E-12 | 42.25 |
| *PRG2* | rs76641566 | G | A | 0.0729 | -0.095 | 0.014 | 7.9433E-12 | 46.05 |
| *PRG2* | rs13256023 | C | T | 0.1855 | -0.108 | 0.009 | 7.9433E-31 | 144.00 |
| *PRG2* | rs7840212 | C | T | 0.3378 | -0.069 | 0.008 | 6.3096E-19 | 74.39 |

| | | | | | | | | |
|---|---|---|---|---|---|---|---|---|
| *PRG2* | rs1888909 | T | C | 0.7491 | -0.057 | 0.008 | 1E-11 | 50.77 |
| *PRG2* | rs10823305 | G | A | 0.8305 | 0.068 | 0.01 | 1.5849E-12 | 46.24 |
| *PRG2* | rs734095 | C | G | 0.1122 | -0.08 | 0.012 | 5.0119E-12 | 44.44 |
| *PRG2* | rs10896615 | T | C | 0.1238 | 0.146 | 0.011 | 1.5849E-39 | 176.17 |
| *PRG2* | rs10774625 | A | G | 0.5041 | -0.064 | 0.007 | 7.9433E-19 | 83.59 |
| *PRG2* | rs12431998 | G | A | 0.1866 | -0.085 | 0.009 | 1E-19 | 89.20 |
| *PRG2* | rs423135 | G | A | 0.5174 | 0.053 | 0.007 | 6.3096E-13 | 57.33 |
| *PRG2* | rs35617692 | C | A | 0.0112 | 0.416 | 0.035 | 3.1623E-33 | 141.27 |
| *PRG2* | rs4807601 | A | G | 0.5006 | -0.153 | 0.007 | 1.9953E-93 | 477.73 |
| *PRG2* | rs146776010 | G | C | 0.0059 | -0.53 | 0.048 | 1.5849E-28 | 121.92 |
| *PRG2* | rs2242892 | G | A | 0.0713 | -0.105 | 0.014 | 1.9953E-13 | 56.25 |
| *PRG3* | rs1434282 | C | T | 0.7265 | 0.077 | 0.008 | 1.5849E-20 | 92.64 |
| *PRG3* | rs7587389 | A | G | 0.5177 | 0.198 | 0.007 | 1.585E-158 | 800.08 |
| *PRG3* | rs1420101 | C | T | 0.3864 | 0.058 | 0.008 | 3.1623E-14 | 52.56 |
| *PRG3* | rs6740627 | A | G | 0.1575 | 0.077 | 0.01 | 3.1623E-14 | 59.29 |
| *PRG3* | rs12619285 | A | G | 0.2655 | -0.094 | 0.008 | 1.9953E-29 | 138.06 |
| *PRG3* | rs2596896 | T | G | 0.6752 | -0.067 | 0.008 | 1.9953E-17 | 70.14 |
| *PRG3* | rs13089722 | G | A | 0.1065 | -0.25 | 0.012 | 1E-98 | 434.03 |
| *PRG3* | rs7449456 | C | T | 0.1888 | 0.069 | 0.009 | 1.2589E-13 | 58.78 |
| *PRG3* | rs11405616 | C | CA | 0.6058 | 0.054 | 0.008 | 6.3096E-13 | 45.56 |
| *PRG3* | rs1136688 | G | A | 0.4295 | -0.057 | 0.007 | 2.5119E-14 | 66.31 |
| *PRG3* | rs76641566 | G | A | 0.0729 | -0.148 | 0.014 | 3.1623E-26 | 111.76 |
| *PRG3* | rs513973 | A | T | 0.2292 | 0.072 | 0.009 | 1.9953E-15 | 64.00 |
| *PRG3* | rs12539299 | C | T | 0.6159 | -0.055 | 0.008 | 2.5119E-13 | 47.27 |
| *PRG3* | rs13256023 | C | T | 0.1855 | -0.119 | 0.009 | 3.9811E-37 | 174.83 |
| *PRG3* | rs7840212 | C | T | 0.3378 | -0.09 | 0.008 | 1.5849E-31 | 126.56 |
| *PRG3* | rs6558372 | G | C | 0.4791 | -0.087 | 0.007 | 1.9953E-32 | 154.47 |
| *PRG3* | rs1888909 | T | C | 0.7491 | -0.076 | 0.008 | 1E-19 | 90.25 |
| *PRG3* | rs150813342 | C | T | 0.0085 | -0.279 | 0.041 | 1E-11 | 46.31 |
| *PRG3* | rs34108746 | A | G | 0.0793 | 0.226 | 0.014 | 1.9953E-62 | 260.59 |
| *PRG3* | rs10859274 | C | T | 0.5372 | -0.06 | 0.007 | 1.9953E-16 | 73.47 |
| *PRG3* | rs7137828 | C | T | 0.5162 | -0.067 | 0.007 | 3.1623E-20 | 91.61 |
| *PRG3* | rs9566645 | T | C | 0.1885 | -0.121 | 0.009 | 2.5119E-38 | 180.75 |
| *PRG3* | rs725613 | T | G | 0.3576 | -0.059 | 0.008 | 7.9433E-15 | 54.39 |
| *PRG3* | rs371029929 | A | C | 0.2461 | -0.1 | 0.009 | 7.9433E-32 | 123.46 |
| *PRG3* | rs35617692 | C | A | 0.0112 | 0.52 | 0.035 | 1E-50 | 220.73 |
| *PRG3* | rs146776010 | G | C | 0.0059 | -0.453 | 0.048 | 2.5119E-21 | 89.07 |
| *PRG3* | rs56117721 | T | A | 0.0717 | -0.1 | 0.014 | 1.9953E-12 | 51.02 |
| *PRKAB1* | rs1354034 | T | C | 0.6035 | 0.055 | 0.008 | 5.0119E-13 | 47.27 |
| *PRKAB1* | rs749140768 | AGGCACCT( | A | 0.0789 | 0.343 | 0.014 | 2.512E-129 | 600.25 |
| *PRKAR1A* | rs68066031 | T | C | 0.2298 | 0.221 | 0.009 | 2.512E-132 | 602.98 |
| *PRKAR1A* | rs1354034 | T | C | 0.6037 | 0.067 | 0.008 | 1.2589E-18 | 70.14 |
| *PRKAR1A* | rs342293 | C | G | 0.4597 | -0.056 | 0.008 | 1E-13 | 49.00 |
| *PRKAR1A* | rs7896518 | A | G | 0.4235 | 0.064 | 0.008 | 1.2589E-16 | 64.00 |
| *PRKAR1A* | rs1552950 | C | G | 0.2929 | 0.073 | 0.008 | 1E-18 | 83.27 |
| *PRKAR2A* | rs151304617 | A | G | 0.0013 | -1.667 | 0.115 | 1.2589E-47 | 210.12 |
| *PRKAR2A* | rs1354034 | T | C | 0.6044 | 0.053 | 0.008 | 1.9953E-12 | 43.89 |
| *PRKAR2A* | rs7896518 | A | G | 0.4236 | 0.055 | 0.008 | 3.1623E-13 | 47.27 |
| *PRKG1* | rs1354034 | T | C | 0.6044 | 0.06 | 0.008 | 5.0119E-15 | 56.25 |
| *PRKG1* | rs6993770 | A | T | 0.287 | -0.07 | 0.008 | 3.1623E-17 | 76.56 |
| *PRKG1* | rs1937682 | A | T | 0.2376 | -0.218 | 0.009 | 2.512E-133 | 586.72 |
| *PRKG1* | rs1863243 | G | A | 0.668 | 0.06 | 0.008 | 3.9811E-14 | 56.25 |
| *PRKRA* | rs1354034 | T | C | 0.6038 | 0.102 | 0.008 | 5.0119E-40 | 162.56 |
| *PRKRA* | rs11502185 | T | C | 0.2597 | 0.077 | 0.01 | 1.5849E-15 | 59.29 |
| *PRKRA* | rs60822569 | T | TTTCTC | 0.554 | 0.064 | 0.008 | 3.1623E-17 | 64.00 |
| *PRL* | rs3055719 | A | ATTTC | 0.7273 | -0.056 | 0.008 | 1.2589E-11 | 49.00 |
| *PRL* | rs7861599 | C | T | 0.4927 | -0.078 | 0.007 | 5.0119E-26 | 124.16 |
| *PRL* | rs11042976 | G | A | 0.482 | -0.051 | 0.007 | 3.1623E-12 | 53.08 |
| *PRND* | rs9657541 | C | T | 0.2055 | -0.121 | 0.009 | 5.0119E-41 | 180.75 |
| *PRND* | rs507666 | G | A | 0.1846 | -0.268 | 0.01 | 1.585E-174 | 718.24 |
| *PRND* | rs1015605 | G | A | 0.3636 | 0.068 | 0.008 | 6.3096E-19 | 72.25 |
| *PRND* | rs4760 | A | G | 0.1554 | 0.074 | 0.01 | 1.5849E-13 | 54.76 |
| *PRND* | rs741233 | C | T | 0.3116 | -0.077 | 0.008 | 5.0119E-23 | 92.64 |
| *PRND* | rs36029876 | C | A | 0.1041 | -0.381 | 0.012 | 1E-200 | 1008.06 |
| *PROC* | rs599839 | G | A | 0.7718 | 0.086 | 0.008 | 1.2589E-25 | 115.56 |
| *PROC* | rs1260326 | T | C | 0.6081 | -0.136 | 0.007 | 1.9953E-80 | 377.47 |
| *PROC* | rs1799810 | A | T | 0.4265 | -0.199 | 0.007 | 3.162E-172 | 808.18 |
| *PROC* | rs13108218 | A | G | 0.619 | -0.053 | 0.007 | 1.9953E-13 | 57.33 |

| | | | | | | | | |
|---|---|---|---|---|---|---|---|---|
| *PROC* | rs9275239 | G | A | 0.4795 | 0.054 | 0.007 | 1.2589E-14 | 59.51 |
| *PROC* | rs13226650 | A | G | 0.1967 | -0.076 | 0.009 | 1.5849E-18 | 71.31 |
| *PROC* | rs2393969 | A | C | 0.4687 | -0.05 | 0.007 | 5.0119E-13 | 51.02 |
| *PROC* | rs149394327 | G | C | 0.03 | 0.142 | 0.02 | 1.9953E-12 | 50.41 |
| *PROCR* | rs867186 | A | G | 0.0861 | 0.73 | 0.014 | 1E-200 | 2718.88 |
| *PROK1* | rs1857513 | G | A | 0.1587 | 0.479 | 0.008 | 1E-200 | 3585.02 |
| *PROK1* | rs7451690 | C | T | 0.5185 | -0.035 | 0.005 | 1E-11 | 49.00 |
| *PROK1* | rs2856694 | T | C | 0.1857 | -0.06 | 0.007 | 1.5849E-17 | 73.47 |
| *PROK1* | rs7497289 | G | A | 0.2832 | 0.04 | 0.006 | 1.2589E-12 | 44.44 |
| *PROS1* | rs1260326 | T | C | 0.6084 | -0.067 | 0.008 | 3.9811E-19 | 70.14 |
| *PROS1* | rs562281690 | G | T | 0.0027 | -1.093 | 0.075 | 1.2589E-48 | 212.38 |
| *PROS1* | rs528128538 | C | T | 0.003 | -0.995 | 0.068 | 1.5849E-48 | 214.11 |
| *PROS1* | rs10982156 | T | A | 0.0693 | -0.421 | 0.015 | 6.31E-170 | 787.74 |
| *PROS1* | rs11054397 | G | A | 0.4059 | -0.06 | 0.007 | 5.0119E-16 | 73.47 |
| *PROS1* | rs924135 | A | T | 0.6103 | 0.055 | 0.008 | 3.9811E-13 | 47.27 |
| *PRR4* | rs12032424 | G | A | 0.4126 | 0.067 | 0.007 | 1.2589E-24 | 91.61 |
| *PRR4* | rs10182233 | C | T | 0.6028 | 0.053 | 0.007 | 1E-15 | 57.33 |
| *PRR4* | rs17513198 | G | A | 0.0929 | 0.119 | 0.011 | 3.1623E-27 | 117.03 |
| *PRR4* | rs780477370 | CCA | C | 0.2684 | -0.076 | 0.007 | 5.0119E-26 | 117.88 |
| *PRR4* | rs56030777 | G | A | 0.2279 | -0.054 | 0.008 | 1.5849E-12 | 45.56 |
| *PRR4* | rs2981737 | C | G | 0.6926 | -0.052 | 0.007 | 1.5849E-13 | 55.18 |
| *PRR4* | rs1063193 | C | T | 0.5648 | -0.783 | 0.008 | 1E-200 | 9579.52 |
| *PRR4* | rs34239025 | C | T | 0.1859 | -0.152 | 0.008 | 2.5119E-75 | 361.00 |
| *PRRT3* | rs61747728 | C | T | 0.0381 | 0.17 | 0.019 | 1.5849E-18 | 80.06 |
| *PRRT3* | rs75166367 | G | A | 0.06 | -0.141 | 0.016 | 2.5119E-19 | 77.66 |
| *PRRT3* | rs55847233 | C | G | 0.2485 | -0.213 | 0.009 | 1.259E-132 | 560.11 |
| *PRSS2* | rs34296809 | CT | C | 0.2059 | 0.072 | 0.009 | 3.9811E-14 | 64.00 |
| *PRSS2* | rs1003582 | C | G | 0.5056 | 0.053 | 0.007 | 7.9433E-13 | 57.33 |
| *PRSS2* | rs9482772 | T | C | 0.451 | 0.078 | 0.007 | 1.2589E-25 | 124.16 |
| *PRSS2* | rs3752404 | A | G | 0.4161 | -0.139 | 0.008 | 6.3096E-76 | 301.89 |
| *PRSS2* | rs748208 | T | C | 0.7121 | -0.07 | 0.008 | 3.1623E-17 | 76.56 |
| *PRSS2* | rs4733612 | G | A | 0.7329 | 0.068 | 0.008 | 7.9433E-16 | 72.25 |
| *PRSS2* | rs174578 | T | A | 0.353 | 0.064 | 0.008 | 1.2589E-16 | 64.00 |
| *PRSS2* | rs686056 | G | A | 0.3226 | -0.078 | 0.008 | 6.3096E-23 | 95.06 |
| *PRSS2* | rs72802342 | C | A | 0.0754 | -0.264 | 0.014 | 1.2589E-75 | 355.59 |
| *PRSS2* | rs533406 | A | G | 0.3815 | -0.107 | 0.008 | 3.9811E-44 | 178.89 |
| *PRSS2* | rs17138478 | C | A | 0.1309 | 0.076 | 0.011 | 5.0119E-12 | 47.74 |
| *PRSS2* | rs7234846 | T | A | 0.0769 | 0.109 | 0.014 | 5.0119E-15 | 60.62 |
| *PRSS22* | rs6917212 | C | G | 0.4267 | -0.054 | 0.008 | 1.2589E-12 | 45.56 |
| *PRSS22* | rs56278466 | T | G | 0.6612 | 0.062 | 0.008 | 7.9433E-15 | 60.06 |
| *PRSS22* | rs148815576 | T | G | 0.0018 | 0.639 | 0.094 | 1.2589E-11 | 46.21 |
| *PRSS22* | rs11634947 | C | G | 0.3686 | -0.054 | 0.008 | 3.1623E-12 | 45.56 |
| *PRSS22* | rs8046218 | C | T | 0.3248 | 0.26 | 0.008 | 1E-200 | 1056.25 |
| *PRSS27* | rs2360962 | A | T | 0.738 | -0.056 | 0.008 | 3.9811E-13 | 49.00 |
| *PRSS27* | rs1354034 | T | C | 0.6038 | -0.049 | 0.007 | 1.2589E-12 | 49.00 |
| *PRSS27* | rs35225290 | C | T | 0.3436 | -0.091 | 0.007 | 5.0119E-37 | 169.00 |
| *PRSS27* | rs541115 | T | A | 0.3382 | 0.052 | 0.007 | 2.5119E-13 | 55.18 |
| *PRSS27* | rs968042 | G | A | 0.6818 | 0.054 | 0.007 | 1.2589E-13 | 59.51 |
| *PRSS27* | rs9501828 | T | C | 0.4669 | -0.157 | 0.007 | 5.012E-116 | 503.04 |
| *PRSS27* | rs10758189 | T | C | 0.3106 | -0.054 | 0.007 | 1.5849E-13 | 59.51 |
| *PRSS27* | rs141129381 | T | TG | 0.3627 | 0.079 | 0.007 | 7.9433E-29 | 127.37 |
| *PRSS27* | rs635634 | C | T | 0.1843 | -0.06 | 0.009 | 1.2589E-11 | 44.44 |
| *PRSS27* | rs112972631 | G | T | 0.2527 | -0.058 | 0.008 | 1E-13 | 52.56 |
| *PRSS27* | rs546202 | T | C | 0.5501 | 0.065 | 0.007 | 1E-20 | 86.22 |
| *PRSS27* | rs11220477 | C | T | 0.0417 | -0.527 | 0.017 | 1E-200 | 961.00 |
| *PRSS27* | rs7306988 | G | A | 0.171 | -0.067 | 0.009 | 1.2589E-13 | 55.42 |
| *PRSS27* | rs79977579 | C | A | 0.0989 | 0.078 | 0.011 | 1E-11 | 50.28 |
| *PRSS27* | rs71386687 | G | T | 0.0824 | 0.387 | 0.013 | 5.012E-193 | 886.21 |
| *PRSS27* | rs708686 | C | T | 0.2677 | 0.16 | 0.008 | 1.9953E-82 | 400.00 |
| *PRSS27* | rs681343 | C | T | 0.5092 | 0.553 | 0.007 | 1E-200 | 6241.00 |
| *PRSS27* | rs61658003 | C | A | 0.8425 | 0.077 | 0.01 | 3.9811E-14 | 59.29 |
| *PRSS27* | rs587935 | C | T | 0.6664 | 0.053 | 0.007 | 1.9953E-13 | 57.33 |
| *PRSS53* | rs903323 | C | T | 0.4796 | -0.06 | 0.006 | 2.5119E-26 | 100.00 |
| *PRSS53* | rs35487539 | C | T | 0.2054 | 0.051 | 0.007 | 3.1623E-13 | 53.08 |
| *PRSS53* | rs142160737 | G | A | 0.0761 | 0.082 | 0.011 | 3.1623E-14 | 55.57 |
| *PRSS53* | rs2286782 | C | A | 0.2188 | -0.048 | 0.007 | 5.0119E-12 | 47.02 |
| *PRSS53* | rs10743951 | A | C | 0.7743 | -0.174 | 0.007 | 2.512E-138 | 617.88 |
| *PRSS53* | rs28929474 | C | T | 0.0208 | 0.235 | 0.02 | 1.5849E-32 | 138.06 |

| | | | | | | | | |
|---|---|---|---|---|---|---|---|---|
| *PRSS53* | rs4468641 | A | C | 0.3851 | 0.885 | 0.008 | 1E-200 | 12237.89 |
| *PRSS53* | rs1801689 | A | C | 0.03 | 0.48 | 0.017 | 2.512E-183 | 797.23 |
| *PRSS8* | rs1260326 | T | C | 0.6087 | -0.065 | 0.007 | 5.0119E-20 | 86.22 |
| *PRSS8* | rs13229619 | G | A | 0.1277 | -0.106 | 0.01 | 1.9953E-24 | 112.36 |
| *PRSS8* | rs764066899 | AT | A | 0.1929 | 0.067 | 0.009 | 5.0119E-14 | 55.42 |
| *PRSS8* | rs62032958 | G | A | 0.0684 | -0.197 | 0.014 | 1.2589E-45 | 198.01 |
| *PRSS8* | rs889555 | C | T | 0.2822 | 0.115 | 0.008 | 6.3096E-49 | 206.64 |
| *PRSS8* | rs9912592 | G | A | 0.5576 | -0.06 | 0.007 | 1.9953E-17 | 73.47 |
| *PRSS8* | rs57822461 | C | T | 0.1822 | -0.094 | 0.009 | 3.9811E-25 | 109.09 |
| *PRTFDC1* | rs143356584 | C | T | 0.0137 | -0.858 | 0.033 | 1E-150 | 676.00 |
| *PRTFDC1* | rs7297265 | A | G | 0.4432 | 0.057 | 0.008 | 7.9433E-14 | 50.77 |
| *PRTG* | rs13107325 | C | T | 0.0748 | 0.096 | 0.013 | 5.0119E-13 | 54.53 |
| *PRTG* | rs78506734 | C | T | 0.1749 | 0.072 | 0.009 | 1.5849E-14 | 64.00 |
| *PRTG* | rs8176746 | G | T | 0.0614 | 0.299 | 0.015 | 2.5119E-94 | 397.34 |
| *PRTG* | rs10851591 | A | G | 0.693 | 0.449 | 0.008 | 1E-200 | 3150.02 |
| *PRTG* | rs7498431 | C | G | 0.6122 | 0.092 | 0.007 | 6.3096E-38 | 172.73 |
| *PRTG* | rs186021206 | G | A | 0.0064 | 0.559 | 0.048 | 1.9953E-31 | 135.63 |
| *PRTG* | rs704 | G | A | 0.4729 | -0.057 | 0.007 | 2.5119E-16 | 66.31 |
| *PRTG* | rs77542162 | A | G | 0.0229 | 0.19 | 0.023 | 3.1623E-16 | 68.24 |
| *PRTG* | rs6565921 | A | G | 0.6339 | -0.064 | 0.007 | 3.9811E-19 | 83.59 |
| *PRTN3* | rs3917932 | C | G | 0.5758 | -0.07 | 0.007 | 1.9953E-22 | 100.00 |
| *PRTN3* | rs768193814 | TACTTA | T | 0.2143 | 0.069 | 0.009 | 1.9953E-15 | 58.78 |
| *PRTN3* | rs6782228 | G | C | 0.2686 | -0.054 | 0.008 | 1.5849E-11 | 45.56 |
| *PRTN3* | rs41559422 | G | A | 0.0373 | 0.248 | 0.019 | 1.9953E-40 | 170.37 |
| *PRTN3* | rs915125 | C | T | 0.2778 | 0.088 | 0.008 | 1.9953E-28 | 121.00 |
| *PRTN3* | rs11155297 | G | T | 0.2638 | -0.055 | 0.008 | 6.3096E-12 | 47.27 |
| *PRTN3* | rs2977810 | T | C | 0.334 | 0.068 | 0.008 | 7.9433E-19 | 72.25 |
| *PRTN3* | rs13277237 | G | A | 0.5674 | -0.089 | 0.007 | 2.5119E-35 | 161.65 |
| *PRTN3* | rs118119317 | G | A | 0.0324 | -0.138 | 0.02 | 3.9811E-12 | 47.61 |
| *PRTN3* | rs73000965 | T | A | 0.3161 | 0.06 | 0.008 | 3.1623E-15 | 56.25 |
| *PRTN3* | rs16942887 | G | A | 0.117 | 0.075 | 0.011 | 1.2589E-11 | 46.49 |
| *PRTN3* | rs8078723 | T | C | 0.3934 | 0.091 | 0.007 | 1E-36 | 169.00 |
| *PRTN3* | rs56378716 | A | G | 0.0134 | 0.242 | 0.031 | 5.0119E-15 | 60.94 |
| *PRTN3* | rs10425544 | T | C | 0.713 | 0.499 | 0.009 | 1E-200 | 3074.09 |
| *PRUNE2* | rs11819066 | T | C | 0.0325 | 0.248 | 0.022 | 3.1623E-30 | 127.07 |
| *PSAP* | rs1736085 | T | C | 0.2604 | 0.094 | 0.008 | 2.5119E-30 | 138.06 |
| *PSAP* | rs6993770 | A | T | 0.2864 | -0.093 | 0.008 | 3.1623E-31 | 135.14 |
| *PSAP* | rs57681041 | C | T | 0.2215 | -0.249 | 0.009 | 6.31E-176 | 765.44 |
| *PSAP* | rs4883201 | A | G | 0.1042 | 0.093 | 0.012 | 5.0119E-15 | 60.06 |
| *PSAP* | rs7310104 | G | A | 0.3251 | 0.053 | 0.008 | 7.9433E-12 | 43.89 |
| *PSAP* | rs10778152 | A | G | 0.2875 | -0.079 | 0.008 | 1.2589E-22 | 97.52 |
| *PSAP* | rs111338191 | A | AAATT | 0.5212 | -0.06 | 0.007 | 2.5119E-16 | 73.47 |
| *PSAP* | rs145078947 | G | T | 0.0025 | 1.032 | 0.072 | 6.3096E-47 | 205.44 |
| *PSAP* | rs2289702 | C | T | 0.1066 | 0.133 | 0.012 | 1.2589E-29 | 122.84 |
| *PSAP* | rs58542926 | C | T | 0.0755 | 0.097 | 0.014 | 1.2589E-12 | 48.01 |
| *PSAPL1* | rs7537101 | T | C | 0.465 | 0.047 | 0.007 | 1E-12 | 45.08 |
| *PSAPL1* | rs1062964 | C | G | 0.0549 | -0.125 | 0.014 | 6.3096E-18 | 79.72 |
| *PSAPL1* | rs34790280 | CA | C | 0.7591 | -0.084 | 0.008 | 3.9811E-26 | 110.25 |
| *PSAPL1* | rs61816761 | G | A | 0.0234 | 0.529 | 0.023 | 1E-115 | 529.00 |
| *PSAPL1* | rs6757005 | T | C | 0.4856 | 0.05 | 0.007 | 1.9953E-14 | 51.02 |
| *PSAPL1* | rs1436662 | A | G | 0.5934 | 0.093 | 0.007 | 3.1623E-44 | 176.51 |
| *PSAPL1* | rs10014338 | C | A | 0.1845 | 0.63 | 0.009 | 1E-200 | 4900.00 |
| *PSAPL1* | rs9368728 | G | A | 0.2885 | 0.05 | 0.007 | 6.3096E-12 | 51.02 |
| *PSAPL1* | rs3132568 | A | G | 0.256 | -0.063 | 0.008 | 6.3096E-17 | 62.02 |
| *PSAPL1* | rs376534130 | C | T | 0.4136 | 0.058 | 0.007 | 1.5849E-17 | 68.65 |
| *PSAPL1* | rs11249885 | C | T | 0.2419 | 0.092 | 0.008 | 3.1623E-33 | 132.25 |
| *PSAPL1* | rs3779660 | G | T | 0.3392 | 0.049 | 0.007 | 1.2589E-12 | 49.00 |
| *PSAPL1* | rs3758294 | T | C | 0.204 | -0.088 | 0.008 | 1E-27 | 121.00 |
| *PSAPL1* | rs4883201 | A | G | 0.1042 | 0.074 | 0.011 | 6.3096E-12 | 45.26 |
| *PSAPL1* | rs11835758 | G | A | 0.4521 | 0.075 | 0.007 | 1E-28 | 114.80 |
| *PSAPL1* | rs59199058 | A | T | 0.3392 | 0.054 | 0.007 | 6.3096E-15 | 59.51 |
| *PSAPL1* | rs17658212 | C | T | 0.0599 | -0.281 | 0.014 | 3.1623E-87 | 402.86 |
| *PSAPL1* | rs889591 | C | G | 0.7387 | 0.051 | 0.007 | 7.9433E-12 | 53.08 |
| *PSAPL1* | rs181040167 | C | T | 0.0057 | -0.336 | 0.047 | 7.9433E-13 | 51.11 |
| *PSAPL1* | rs10686390 | A | ATC | 0.6089 | -0.058 | 0.007 | 1E-17 | 68.65 |
| *PSAPL1* | rs4078130 | C | T | 0.5516 | 0.086 | 0.007 | 3.9811E-39 | 150.94 |
| *PSAPL1* | rs6508781 | A | C | 0.1682 | -0.195 | 0.009 | 5.012E-109 | 469.44 |
| *PSAPL1* | rs181163991 | T | C | 0.0172 | -0.229 | 0.028 | 1.5849E-16 | 66.89 |

| | | | | | | | | |
|---|---|---|---|---|---|---|---|---|
| *PSCA* | rs2976388 | G | A | 0.4223 | 1.161 | 0.008 | 1E-200 | 21061.27 |
| *PSCA* | rs2519093 | C | T | 0.1842 | -0.146 | 0.006 | 1.259E-146 | 592.11 |
| *PSCA* | rs186021206 | G | A | 0.0065 | 0.228 | 0.03 | 1.9953E-14 | 57.76 |
| *PSCA* | rs2608894 | T | C | 0.7651 | -0.036 | 0.005 | 1.2589E-11 | 51.84 |
| *PSCA* | rs4760 | A | G | 0.1555 | 0.116 | 0.006 | 6.3096E-82 | 373.78 |
| *PSCA* | rs516246 | C | T | 0.5092 | -0.12 | 0.004 | 3.981E-164 | 900.00 |
| *PSG1* | rs7526027 | T | C | 0.292 | 0.048 | 0.007 | 5.0119E-13 | 47.02 |
| *PSG1* | rs78689694 | G | C | 0.1327 | -0.097 | 0.009 | 6.3096E-28 | 116.16 |
| *PSG1* | rs909222 | A | C | 0.4558 | 0.045 | 0.006 | 5.0119E-14 | 56.25 |
| *PSG1* | rs186021206 | G | A | 0.0064 | 0.318 | 0.041 | 7.9433E-15 | 60.16 |
| *PSG1* | rs12986075 | G | A | 0.1987 | -0.962 | 0.009 | 1E-200 | 11425.23 |
| *PSG1* | rs61740142 | T | C | 0.0383 | -0.29 | 0.016 | 3.9811E-77 | 328.52 |
| *PSG1* | rs6068155 | C | T | 0.6322 | 0.044 | 0.006 | 7.9433E-13 | 53.78 |
| *PSIP1* | rs1354034 | T | C | 0.6043 | 0.126 | 0.008 | 1.9953E-62 | 248.06 |
| *PSIP1* | rs7080536 | G | A | 0.0436 | -0.16 | 0.018 | 3.1623E-18 | 79.01 |
| *PSIP1* | rs11604127 | C | T | 0.2306 | 0.06 | 0.009 | 1E-11 | 44.44 |
| *PSIP1* | rs111338191 | A | AAATT | 0.5212 | -0.061 | 0.007 | 1.9953E-16 | 75.94 |
| *PSMD9* | rs80090724 | T | C | 0.2924 | -0.194 | 0.008 | 7.943E-129 | 588.06 |
| *PSME1* | rs1354034 | T | C | 0.6041 | 0.054 | 0.007 | 5.0119E-13 | 59.51 |
| *PSME1* | rs2236352 | T | C | 0.2559 | 0.094 | 0.008 | 1.2589E-28 | 138.06 |
| *PSME2* | rs2236352 | T | C | 0.2559 | 0.104 | 0.008 | 5.0119E-35 | 169.00 |
| *PSMG3* | rs9503502 | T | A | 0.8319 | -0.196 | 0.01 | 3.1623E-91 | 384.16 |
| *PSMG3* | rs34407549 | C | G | 0.0081 | -0.29 | 0.042 | 7.9433E-12 | 47.68 |
| *PSPN* | rs4381790 | C | A | 0.545 | -0.046 | 0.006 | 1.2589E-16 | 58.78 |
| *PSPN* | rs6542472 | A | C | 0.3571 | 0.041 | 0.006 | 1.2589E-12 | 46.69 |
| *PSPN* | rs2102843 | T | C | 0.0597 | -0.095 | 0.012 | 6.3096E-16 | 62.67 |
| *PSPN* | rs13111751 | C | G | 0.3374 | -0.046 | 0.006 | 7.9433E-15 | 58.78 |
| *PSPN* | rs60415382 | C | G | 0.1924 | 0.081 | 0.007 | 3.1623E-30 | 133.90 |
| *PSPN* | rs9543374 | C | T | 0.2643 | -0.071 | 0.006 | 2.5119E-29 | 140.03 |
| *PSPN* | rs138903858 | G | T | 0.0279 | -0.145 | 0.017 | 7.9433E-17 | 72.75 |
| *PSPN* | rs1863115 | C | A | 0.7423 | -0.046 | 0.007 | 3.9811E-12 | 43.18 |
| *PSPN* | rs749522465 | AG | A | 0.3088 | 0.046 | 0.006 | 3.9811E-14 | 58.78 |
| *PSPN* | rs58634535 | G | A | 0.1563 | -0.056 | 0.008 | 3.9811E-13 | 49.00 |
| *PSPN* | rs549768142 | G | GAAAAAAA | 0.7965 | 0.068 | 0.007 | 1.2589E-21 | 94.37 |
| *PSPN* | rs6517463 | C | T | 0.2451 | 0.05 | 0.006 | 7.9433E-15 | 69.44 |
| *PSRC1* | rs34863121 | C | T | 0.0129 | 0.725 | 0.034 | 6.31E-103 | 454.69 |
| *PSRC1* | rs10733789 | T | C | 0.3128 | 0.056 | 0.008 | 1.5849E-11 | 49.00 |
| *PSRC1* | rs709049 | T | G | 0.5536 | -0.066 | 0.008 | 7.9433E-17 | 68.06 |
| *PSRC1* | rs6070696 | A | G | 0.1852 | 0.094 | 0.01 | 1.2589E-21 | 88.36 |
| *PSTPIP2* | rs10822145 | C | T | 0.4721 | 0.059 | 0.008 | 2.5119E-14 | 54.39 |
| *PSTPIP2* | rs72642432 | G | A | 0.1318 | -0.226 | 0.011 | 1.9953E-87 | 422.12 |
| *PTGDS* | rs12934455 | C | T | 0.1664 | -0.067 | 0.01 | 2.5119E-12 | 44.89 |
| *PTGDS* | rs704 | G | A | 0.473 | 0.048 | 0.007 | 1.5849E-11 | 47.02 |
| *PTGES2* | rs13283456 | C | T | 0.2103 | -0.097 | 0.009 | 3.1623E-25 | 116.16 |
| *PTGR1* | rs2169387 | A | G | 0.8985 | -0.085 | 0.012 | 1.2589E-13 | 50.17 |
| *PTGR1* | rs112875651 | G | A | 0.3913 | -0.055 | 0.007 | 1.9953E-14 | 61.73 |
| *PTGR1* | rs7036102 | A | G | 0.0504 | 0.758 | 0.017 | 1E-200 | 1988.11 |
| *PTGR1* | rs1788783 | C | T | 0.5018 | -0.052 | 0.007 | 6.3096E-14 | 55.18 |
| *PTGR1* | rs12975366 | T | C | 0.3956 | -0.049 | 0.007 | 6.3096E-12 | 49.00 |
| *PTGR1* | rs738409 | C | G | 0.2172 | 0.061 | 0.008 | 3.1623E-13 | 58.14 |
| *PTH* | rs10051765 | T | C | 0.3312 | -0.074 | 0.008 | 1.2589E-20 | 85.56 |
| *PTH* | rs7800944 | T | C | 0.2861 | -0.07 | 0.008 | 3.1623E-17 | 76.56 |
| *PTH* | rs286 | A | T | 0.0856 | -0.096 | 0.013 | 7.9433E-13 | 54.53 |
| *PTH* | rs34954997 | C | CTTCG | 0.238 | 0.087 | 0.009 | 6.3096E-23 | 93.44 |
| *PTH* | rs35870583 | G | GT | 0.1871 | 0.122 | 0.01 | 6.3096E-35 | 148.84 |
| *PTH1R* | rs121434601 | C | T | 0.0027 | 2.259 | 0.074 | 1E-200 | 931.90 |
| *PTK7* | rs116450546 | A | T | 0.0361 | 0.156 | 0.021 | 6.3096E-14 | 55.18 |
| *PTN* | rs4252165 | C | A | 0.294 | 0.071 | 0.008 | 2.5119E-19 | 78.77 |
| *PTN* | rs1431092 | T | A | 0.4248 | 0.151 | 0.007 | 1.2589E-93 | 465.33 |
| *PTN* | rs4541868 | C | A | 0.2738 | -0.058 | 0.008 | 1E-12 | 52.56 |
| *PTN* | rs1654425 | T | C | 0.8341 | 0.071 | 0.01 | 1.9953E-13 | 50.41 |
| *PTP4A3* | rs2242197 | A | C | 0.3204 | -0.236 | 0.008 | 1.259E-183 | 870.25 |
| *PTPN1* | rs1354034 | T | C | 0.6041 | 0.084 | 0.008 | 3.9811E-28 | 110.25 |
| *PTPN1* | rs342293 | C | G | 0.459 | -0.052 | 0.007 | 5.0119E-12 | 55.18 |
| *PTPN1* | rs7080536 | G | A | 0.0436 | -0.194 | 0.019 | 2.5119E-25 | 104.25 |
| *PTPN6* | rs1354034 | T | C | 0.6039 | 0.061 | 0.008 | 3.1623E-15 | 58.14 |
| *PTPN6* | rs342298 | C | T | 0.4545 | -0.073 | 0.008 | 5.0119E-22 | 83.27 |
| *PTPRB* | rs4543739 | A | C | 0.2491 | -0.054 | 0.008 | 6.3096E-12 | 45.56 |

| | | | | | | | | |
|---|---|---|---|---|---|---|---|---|
| *PTPRB* | rs545002076 | C | CTT | 0.4448 | -0.057 | 0.007 | 3.1623E-15 | 66.31 |
| *PTPRB* | rs6825454 | T | C | 0.2573 | 0.085 | 0.008 | 2.5119E-27 | 112.89 |
| *PTPRB* | rs189209601 | G | A | 0.0566 | 0.116 | 0.016 | 7.9433E-13 | 52.56 |
| *PTPRB* | rs8176747 | C | G | 0.0612 | 0.541 | 0.014 | 1E-200 | 1493.27 |
| *PTPRB* | rs34434834 | G | A | 0.0333 | 0.256 | 0.019 | 1.2589E-40 | 181.54 |
| *PTPRB* | rs2465811 | T | C | 0.2865 | -0.367 | 0.008 | 1E-200 | 2104.52 |
| *PTPRB* | rs3184504 | T | C | 0.517 | 0.052 | 0.007 | 3.9811E-13 | 55.18 |
| *PTPRB* | rs2141247 | C | A | 0.6134 | 0.095 | 0.007 | 3.9811E-37 | 184.18 |
| *PTPRB* | rs6602909 | T | C | 0.3239 | 0.084 | 0.007 | 6.3096E-30 | 144.00 |
| *PTPRB* | rs653152 | G | A | 0.6777 | -0.081 | 0.007 | 3.1623E-28 | 133.90 |
| *PTPRC* | rs139888769 | G | GT | 0.0416 | 0.171 | 0.019 | 1E-19 | 81.00 |
| *PTPRC* | rs552839429 | T | C | 0.0168 | -0.806 | 0.029 | 7.943E-173 | 772.46 |
| *PTPRC* | rs34357864 | G | GA | 0.3178 | 0.059 | 0.008 | 1.5849E-13 | 54.39 |
| *PTPRC* | rs75071241 | G | A | 0.0596 | -0.154 | 0.016 | 3.9811E-23 | 92.64 |
| *PTPRC* | rs3184504 | T | C | 0.5174 | -0.136 | 0.007 | 1.2589E-76 | 377.47 |
| *PTPRC* | rs186021206 | G | A | 0.0064 | 1.325 | 0.051 | 6.31E-150 | 674.98 |
| *PTPRF* | rs2842194 | G | A | 0.225 | -0.089 | 0.009 | 3.1623E-25 | 97.79 |
| *PTPRF* | rs6531216 | G | A | 0.5297 | -0.062 | 0.007 | 6.3096E-18 | 78.45 |
| *PTPRF* | rs9873618 | G | A | 0.2887 | -0.064 | 0.008 | 3.9811E-16 | 64.00 |
| *PTPRF* | rs146996008 | A | AGT | 0.4168 | -0.049 | 0.007 | 1.2589E-11 | 49.00 |
| *PTPRF* | rs17039766 | C | A | 0.0563 | 0.292 | 0.016 | 1.5849E-77 | 333.06 |
| *PTPRF* | rs2308527 | G | T | 0.5861 | -0.059 | 0.007 | 5.0119E-16 | 71.04 |
| *PTPRF* | rs581080 | G | C | 0.8209 | 0.064 | 0.009 | 1E-11 | 50.57 |
| *PTPRF* | rs507666 | G | A | 0.1845 | -0.083 | 0.009 | 3.9811E-19 | 85.05 |
| *PTPRF* | rs10761741 | G | T | 0.4134 | 0.069 | 0.007 | 5.0119E-21 | 97.16 |
| *PTPRF* | rs9633740 | A | G | 0.7954 | 0.138 | 0.009 | 1.2589E-54 | 235.11 |
| *PTPRF* | rs1535 | A | G | 0.3467 | 0.055 | 0.007 | 2.5119E-13 | 61.73 |
| *PTPRF* | rs11045856 | T | G | 0.2403 | 0.097 | 0.008 | 1.5849E-30 | 147.02 |
| *PTPRF* | rs28929474 | C | T | 0.0208 | 0.334 | 0.025 | 3.1623E-40 | 178.49 |
| *PTPRF* | rs1801689 | A | C | 0.0302 | -0.217 | 0.021 | 3.9811E-25 | 106.78 |
| *PTPRF* | rs516246 | C | T | 0.5088 | -0.101 | 0.007 | 1.2589E-44 | 208.18 |
| *PTPRH* | rs35939242 | A | C | 0.3409 | 0.048 | 0.007 | 1.5849E-12 | 47.02 |
| *PTPRH* | rs13067380 | A | G | 0.1955 | -0.096 | 0.008 | 1.9953E-31 | 144.00 |
| *PTPRH* | rs7383388 | A | T | 0.4035 | -0.054 | 0.007 | 1E-15 | 59.51 |
| *PTPRH* | rs3731211 | T | A | 0.722 | -0.057 | 0.007 | 3.9811E-15 | 66.31 |
| *PTPRH* | rs2519093 | C | T | 0.1842 | -0.207 | 0.008 | 7.943E-132 | 669.52 |
| *PTPRH* | rs59889308 | C | T | 0.2372 | 0.061 | 0.008 | 1.5849E-15 | 58.14 |
| *PTPRH* | rs3862630 | C | T | 0.1224 | -0.145 | 0.01 | 7.9433E-45 | 210.25 |
| *PTPRH* | rs653178 | C | T | 0.5166 | -0.104 | 0.006 | 6.3096E-58 | 300.44 |
| *PTPRH* | rs753728 | G | A | 0.1146 | -0.089 | 0.01 | 2.5119E-18 | 79.21 |
| *PTPRH* | rs186021206 | G | A | 0.0065 | 0.442 | 0.045 | 3.1623E-23 | 96.48 |
| *PTPRH* | rs2288523 | C | A | 0.1307 | 0.907 | 0.011 | 1E-200 | 6798.75 |
| *PTPRK* | rs9321109 | G | A | 0.8289 | -0.136 | 0.01 | 1.2589E-41 | 184.96 |
| *PTPRK* | rs2519093 | C | T | 0.1842 | -0.167 | 0.01 | 5.0119E-65 | 278.89 |
| *PTPRK* | rs56278466 | T | G | 0.6608 | 0.089 | 0.008 | 3.9811E-29 | 123.77 |
| *PTPRM* | rs61804211 | G | T | 0.1046 | 0.096 | 0.011 | 7.9433E-20 | 76.17 |
| *PTPRM* | rs34579268 | C | T | 0.371 | 0.046 | 0.007 | 1E-11 | 43.18 |
| *PTPRM* | rs13107325 | C | T | 0.0748 | 0.117 | 0.012 | 2.5119E-21 | 95.06 |
| *PTPRM* | rs7768875 | C | T | 0.3281 | -0.051 | 0.007 | 3.9811E-13 | 53.08 |
| *PTPRM* | rs2519093 | C | T | 0.1842 | -0.699 | 0.01 | 1E-200 | 4886.01 |
| *PTPRM* | rs12359178 | G | A | 0.1445 | 0.079 | 0.009 | 5.0119E-18 | 77.05 |
| *PTPRM* | rs174537 | G | T | 0.3451 | 0.047 | 0.007 | 1.9953E-12 | 45.08 |
| *PTPRM* | rs11603123 | G | A | 0.0337 | 0.178 | 0.018 | 1.2589E-23 | 97.79 |
| *PTPRM* | rs186021206 | G | A | 0.0064 | 0.638 | 0.044 | 3.9811E-47 | 210.25 |
| *PTPRM* | rs72911294 | C | T | 0.271 | -0.08 | 0.008 | 6.3096E-25 | 100.00 |
| *PTPRM* | rs4429561 | A | G | 0.6876 | -0.058 | 0.007 | 3.9811E-17 | 68.65 |
| *PTPRN2* | rs3800860 | C | T | 0.3576 | -0.155 | 0.008 | 1.9953E-88 | 375.39 |
| *PTPRN2* | rs7203642 | A | G | 0.1921 | -0.064 | 0.009 | 7.9433E-12 | 50.57 |
| *PTPRR* | rs144705265 | G | A | 0.0053 | -1.154 | 0.051 | 1.585E-114 | 512.00 |
| *PTPRR* | rs77924615 | G | A | 0.1965 | -0.064 | 0.009 | 6.3096E-12 | 50.57 |
| *PTPRS* | rs1260326 | T | C | 0.6084 | 0.091 | 0.007 | 1.5849E-34 | 169.00 |
| *PTPRS* | rs545971 | C | T | 0.318 | 0.057 | 0.008 | 3.9811E-13 | 50.77 |
| *PTPRS* | rs144268704 | A | AGGTTTTG( | 0.337 | 0.08 | 0.008 | 3.9811E-25 | 100.00 |
| *PTPRS* | rs186021206 | G | A | 0.0064 | 0.433 | 0.05 | 5.0119E-18 | 75.00 |
| *PTPRS* | rs55763631 | C | A | 0.0481 | 0.659 | 0.018 | 1E-200 | 1340.37 |
| *PTPRZ1* | rs1984769 | G | A | 0.1604 | 0.086 | 0.01 | 1.5849E-18 | 73.96 |
| *PTPRZ1* | rs17422797 | C | T | 0.0744 | 0.095 | 0.013 | 6.3096E-14 | 53.40 |
| *PTPRZ1* | rs7797305 | C | T | 0.4866 | -0.169 | 0.007 | 6.31E-139 | 582.88 |

| | | | | | | | | |
|---|---|---|---|---|---|---|---|---|
| *PTPRZ1* | rs35281152 | A | AG | 0.5315 | 0.057 | 0.007 | 2.5119E-17 | 66.31 |
| *PTPRZ1* | rs10956755 | A | G | 0.2999 | -0.099 | 0.007 | 3.1623E-42 | 200.02 |
| *PTPRZ1* | rs56278466 | T | G | 0.6612 | 0.086 | 0.007 | 1.9953E-34 | 150.94 |
| *PTPRZ1* | rs548589580 | T | TA | 0.4841 | 0.075 | 0.007 | 1.9953E-29 | 114.80 |
| *PTPRZ1* | rs373909380 | A | T | 0.2278 | 0.112 | 0.008 | 2.5119E-40 | 196.00 |
| *PTPRZ1* | rs200489612 | G | A | 0.0052 | 0.811 | 0.05 | 6.3096E-59 | 263.09 |
| *PTPRZ1* | rs12451299 | G | C | 0.242 | 0.059 | 0.008 | 3.9811E-14 | 54.39 |
| *PTPRZ1* | rs601338 | G | A | 0.5092 | 0.099 | 0.007 | 3.9811E-49 | 200.02 |
| *PTRHD1* | rs138426814 | T | C | 0.0011 | -1.145 | 0.116 | 3.9811E-23 | 97.43 |
| *PTRHD1* | rs1354034 | T | C | 0.6044 | 0.071 | 0.008 | 1.5849E-20 | 78.77 |
| *PTRHD1* | rs7080386 | C | A | 0.412 | 0.062 | 0.008 | 7.9433E-16 | 60.06 |
| *PTS* | rs183616935 | A | C | 0.0058 | 0.48 | 0.049 | 1.5849E-22 | 95.96 |
| *PTS* | rs112875651 | G | A | 0.3908 | -0.054 | 0.008 | 1.9953E-12 | 45.56 |
| *PTS* | rs12796145 | G | C | 0.2328 | -0.236 | 0.009 | 3.981E-154 | 687.60 |
| *PTS* | rs738408 | C | T | 0.2167 | 0.086 | 0.009 | 1.5849E-21 | 91.31 |
| *PTX3* | rs678 | A | T | 0.3566 | -0.079 | 0.008 | 1E-23 | 97.52 |
| *PTX3* | rs56025932 | A | C | 0.0427 | -0.15 | 0.019 | 1.5849E-15 | 62.33 |
| *PTX3* | rs41341748 | G | A | 0.0111 | 0.337 | 0.035 | 1.5849E-21 | 92.71 |
| *PTX3* | rs7084817 | C | G | 0.0517 | -0.636 | 0.017 | 1E-200 | 1399.64 |
| *PVALB* | rs3811444 | C | T | 0.3323 | -0.067 | 0.007 | 7.9433E-24 | 91.61 |
| *PVALB* | rs10820606 | A | C | 0.2292 | 0.052 | 0.008 | 7.9433E-12 | 42.25 |
| *PVALB* | rs7896518 | A | G | 0.4237 | 0.059 | 0.006 | 1.5849E-20 | 96.69 |
| *PVALB* | rs6580981 | G | A | 0.4587 | 0.05 | 0.006 | 3.1623E-15 | 69.44 |
| *PVALB* | rs11553699 | A | G | 0.1351 | -0.066 | 0.01 | 6.3096E-12 | 43.56 |
| *PVALB* | rs4821544 | T | C | 0.3032 | -0.813 | 0.008 | 1E-200 | 10327.64 |
| *PVR* | rs13234131 | A | G | 0.127 | -0.074 | 0.009 | 6.3096E-16 | 67.60 |
| *PVR* | rs200489612 | G | A | 0.0052 | 0.314 | 0.046 | 7.9433E-12 | 46.60 |
| *PVR* | rs2301274 | T | C | 0.2406 | -0.786 | 0.009 | 1E-200 | 7627.11 |
| *PXDNL* | rs7833909 | A | G | 0.1235 | -0.345 | 0.012 | 3.162E-196 | 826.56 |
| *PXN* | rs3129754 | A | G | 0.4198 | 0.056 | 0.007 | 7.9433E-16 | 64.00 |
| *PXN* | rs4767884 | T | C | 0.7657 | 0.757 | 0.009 | 1E-200 | 7074.68 |
| *PXN* | rs4632248 | G | T | 0.2095 | 0.068 | 0.008 | 3.9811E-16 | 72.25 |
| *PYDC1* | rs61816761 | G | A | 0.0236 | 0.401 | 0.023 | 1E-68 | 303.97 |
| *PYDC1* | rs11417925 | A | AC | 0.5968 | 0.056 | 0.007 | 7.9433E-17 | 64.00 |
| *PYDC1* | rs1352846 | A | G | 0.2925 | -0.706 | 0.008 | 1E-200 | 7788.06 |
| *PYDC1* | rs6557841 | A | C | 0.6152 | -0.048 | 0.007 | 7.9433E-13 | 47.02 |
| *PYDC1* | rs10995328 | A | G | 0.5998 | -0.054 | 0.007 | 3.9811E-16 | 59.51 |
| *PYDC1* | rs17658212 | C | T | 0.0597 | -0.175 | 0.014 | 2.5119E-35 | 156.25 |
| *PYDC1* | rs4561481 | G | A | 0.6741 | 0.306 | 0.007 | 1E-200 | 1910.94 |
| *PYDC1* | rs761061860 | GGTC | G | 0.1682 | -0.134 | 0.009 | 3.1623E-52 | 221.68 |
| *PYDC1* | rs112710702 | G | A | 0.0425 | -0.138 | 0.016 | 1.5849E-17 | 74.39 |
| *PYDC1* | rs6070809 | C | G | 0.309 | 0.104 | 0.007 | 1.5849E-49 | 220.73 |
| *PYY* | rs56092644 | G | T | 0.2446 | 0.166 | 0.009 | 5.0119E-79 | 340.20 |
| *PYY* | rs750123962 | ATGTGCAA' | A | 0.6947 | -0.079 | 0.008 | 3.9811E-22 | 97.52 |
| *PYY* | rs2341378 | C | T | 0.6797 | 0.245 | 0.008 | 1E-199 | 937.89 |
| *PYY* | rs72943036 | C | T | 0.0847 | 0.103 | 0.013 | 2.5119E-14 | 62.78 |
| *PYY* | rs770560346 | CTCT | C | 0.2583 | 0.072 | 0.009 | 6.3096E-17 | 64.00 |
| *PYY* | rs516246 | C | T | 0.5089 | 0.069 | 0.008 | 3.9811E-20 | 74.39 |
| *PZP* | rs7082470 | G | A | 0.4681 | -0.058 | 0.005 | 1.9953E-29 | 134.56 |
| *PZP* | rs7311982 | T | C | 0.7108 | 0.333 | 0.006 | 1E-200 | 3080.25 |
| *PZP* | rs9905583 | C | T | 0.4856 | 0.037 | 0.005 | 3.1623E-13 | 54.76 |
| *PZP* | rs11666424 | A | G | 0.2738 | -0.039 | 0.006 | 7.9433E-12 | 42.25 |
| *QDPR* | rs1354034 | T | C | 0.6035 | 0.061 | 0.007 | 3.1623E-16 | 75.94 |
| *QPCT* | rs188468174 | C | T | 0.0143 | -0.398 | 0.031 | 1.9953E-37 | 164.83 |
| *QPCT* | rs4670696 | A | C | 0.0236 | -0.884 | 0.025 | 1E-200 | 1250.33 |
| *QPCT* | rs4917 | T | C | 0.6398 | 0.072 | 0.008 | 5.0119E-21 | 81.00 |
| *QPCT* | rs1130389 | C | T | 0.1473 | -0.086 | 0.011 | 1.9953E-14 | 61.12 |
| *QPCT* | rs73169678 | C | A | 0.1124 | 0.095 | 0.012 | 2.5119E-16 | 62.67 |
| *QPCT* | rs13277237 | G | A | 0.5673 | -0.07 | 0.007 | 2.5119E-21 | 100.00 |
| *QPCT* | rs56278466 | T | G | 0.6615 | 0.089 | 0.008 | 6.3096E-31 | 123.77 |
| *QPCT* | rs10896045 | A | G | 0.7022 | -0.15 | 0.008 | 3.9811E-78 | 351.56 |
| *QPCT* | rs3803800 | A | G | 0.7889 | -0.063 | 0.009 | 1.2589E-12 | 49.00 |
| *QPCT* | rs34562254 | G | A | 0.097 | 0.118 | 0.012 | 5.0119E-22 | 96.69 |
| *QPCT* | rs34003767 | A | G | 0.3687 | -0.055 | 0.008 | 3.1623E-13 | 47.27 |
| *QPCT* | rs2725405 | G | C | 0.4412 | 0.051 | 0.007 | 6.3096E-12 | 53.08 |
| *QSOX1* | rs12371 | A | G | 0.0876 | -1.086 | 0.013 | 1E-200 | 6978.67 |
| *QSOX1* | rs10817200 | A | G | 0.2648 | -0.056 | 0.008 | 3.9811E-13 | 49.00 |
| *QSOX1* | rs186021206 | G | A | 0.0065 | 0.346 | 0.046 | 1E-13 | 56.58 |

| | | | | | | | | |
|---|---|---|---|---|---|---|---|---|
| *RAB10* | rs1354034 | T | C | 0.6045 | 0.058 | 0.008 | 6.3096E-14 | 52.56 |
| *RAB10* | rs7916868 | A | T | 0.5016 | 0.055 | 0.008 | 1.9953E-13 | 47.27 |
| *RAB11FIP3* | rs7916868 | A | T | 0.5016 | 0.058 | 0.007 | 6.3096E-15 | 68.65 |
| *RAB11FIP3* | rs74847787 | T | C | 0.0307 | -0.165 | 0.022 | 3.1623E-14 | 56.25 |
| *RAB27B* | rs2631367 | C | G | 0.5189 | 0.057 | 0.008 | 2.5119E-14 | 50.77 |
| *RAB27B* | rs342299 | C | T | 0.4546 | -0.051 | 0.008 | 1E-11 | 40.64 |
| *RAB27B* | rs6993770 | A | T | 0.2866 | -0.099 | 0.008 | 1E-32 | 153.14 |
| *RAB27B* | rs7896518 | A | G | 0.424 | 0.067 | 0.008 | 5.0119E-18 | 70.14 |
| *RAB27B* | rs1362213 | C | T | 0.4792 | -0.052 | 0.008 | 7.9433E-12 | 42.25 |
| *RAB27B* | rs55656227 | G | A | 0.8116 | 0.069 | 0.01 | 1.2589E-12 | 47.61 |
| *RAB2B* | rs1354034 | T | C | 0.6044 | 0.065 | 0.008 | 2.5119E-17 | 66.02 |
| *RAB2B* | rs7916868 | A | T | 0.5014 | 0.056 | 0.007 | 5.0119E-14 | 64.00 |
| *RAB2B* | rs34855848 | C | CA | 0.1366 | -0.233 | 0.011 | 5.012E-100 | 448.67 |
| *RAB44* | rs143756287 | A | ACTCCTGA( | 0.2793 | -0.147 | 0.009 | 6.3096E-67 | 266.78 |
| *RAB44* | rs10424405 | A | G | 0.2118 | 0.077 | 0.009 | 6.3096E-17 | 73.20 |
| *RAB6A* | rs5030062 | A | C | 0.3728 | 0.196 | 0.007 | 1.585E-155 | 784.00 |
| *RAB6A* | rs4861708 | G | A | 0.5119 | 0.39 | 0.007 | 1E-200 | 3104.08 |
| *RAB6A* | rs1801020 | A | G | 0.745 | 0.209 | 0.008 | 1E-145 | 682.52 |
| *RAB6A* | rs35939600 | TA | T | 0.1492 | 0.073 | 0.01 | 7.9433E-13 | 53.29 |
| *RAB6A* | rs821840 | A | G | 0.3251 | 0.068 | 0.008 | 1.5849E-19 | 72.25 |
| *RABEP1* | rs1354034 | T | C | 0.6044 | 0.061 | 0.008 | 5.0119E-16 | 58.14 |
| *RABEP1* | rs342293 | C | G | 0.4596 | -0.051 | 0.007 | 1E-11 | 53.08 |
| *RABEP1* | rs6993770 | A | T | 0.287 | -0.059 | 0.008 | 6.3096E-13 | 54.39 |
| *RABEPK* | rs7858935 | A | T | 0.1472 | 0.207 | 0.011 | 5.0119E-80 | 354.12 |
| *RABGAP1L* | rs6661868 | A | G | 0.2014 | 0.092 | 0.009 | 1.2589E-22 | 104.49 |
| *RABGAP1L* | rs1354034 | T | C | 0.6035 | 0.057 | 0.008 | 6.3096E-14 | 50.77 |
| *RAD23B* | rs6993770 | A | T | 0.2869 | -0.061 | 0.008 | 6.3096E-13 | 58.14 |
| *RAD23B* | rs7035725 | C | G | 0.5498 | -0.064 | 0.008 | 5.0119E-17 | 64.00 |
| *RALB* | rs148102406 | T | TGTGA | 0.6835 | -0.117 | 0.008 | 1.5849E-44 | 213.89 |
| *RALY* | rs7080536 | G | A | 0.0437 | -0.162 | 0.019 | 6.3096E-18 | 72.70 |
| *RANBP1* | rs1354034 | T | C | 0.6045 | 0.06 | 0.008 | 3.9811E-15 | 56.25 |
| *RANBP1* | rs76889809 | C | A | 0.0547 | 0.123 | 0.017 | 1.2589E-13 | 52.35 |
| *RAP1A* | rs11571302 | G | T | 0.4753 | -0.061 | 0.008 | 2.5119E-15 | 58.14 |
| *RAP1A* | rs13263709 | T | C | 0.6487 | 0.055 | 0.008 | 6.3096E-12 | 47.27 |
| *RARRES1* | rs6441224 | T | C | 0.5249 | 0.457 | 0.008 | 1E-200 | 3263.27 |
| *RARRES2* | rs1260326 | T | C | 0.6085 | -0.054 | 0.008 | 7.9433E-13 | 45.56 |
| *RARRES2* | rs1265905 | A | G | 0.1299 | -0.094 | 0.011 | 6.3096E-18 | 73.02 |
| *RARRES2* | rs3735167 | C | T | 0.2565 | 0.216 | 0.008 | 3.162E-144 | 729.00 |
| *RASSF2* | rs2246735 | C | T | 0.5726 | 0.095 | 0.008 | 7.9433E-36 | 141.02 |
| *RBFOX3* | rs7080536 | G | A | 0.0435 | -0.122 | 0.017 | 1.2589E-12 | 51.50 |
| *RBKS* | rs140948699 | C | G | 0.0091 | -1.812 | 0.039 | 1E-200 | 2158.67 |
| *RBM17* | rs342293 | C | G | 0.4596 | -0.075 | 0.008 | 3.9811E-23 | 87.89 |
| *RBM17* | rs6993770 | A | T | 0.2864 | -0.079 | 0.008 | 1.5849E-21 | 97.52 |
| *RBM17* | rs11256771 | C | T | 0.0374 | -0.378 | 0.02 | 7.9433E-81 | 357.21 |
| *RBP1* | rs2071387 | A | G | 0.1824 | 0.137 | 0.01 | 1E-42 | 187.69 |
| *RBP1* | rs607929 | C | G | 0.4485 | -0.062 | 0.008 | 1.2589E-15 | 60.06 |
| *RBP2* | rs1375131 | T | C | 0.2655 | 0.059 | 0.009 | 1.2589E-11 | 42.98 |
| *RBP2* | rs295469 | G | A | 0.3785 | -0.068 | 0.008 | 1.5849E-18 | 72.25 |
| *RBP2* | rs6472539 | G | C | 0.515 | -0.062 | 0.008 | 1.5849E-16 | 60.06 |
| *RBP2* | rs17655730 | T | C | 0.2455 | 0.112 | 0.009 | 2.5119E-37 | 154.86 |
| *RBP5* | rs746441505 | CTCTCTT | C | 0.4129 | 0.07 | 0.007 | 2.5119E-22 | 100.00 |
| *RBP5* | rs112875651 | G | A | 0.3909 | -0.051 | 0.007 | 3.9811E-12 | 53.08 |
| *RBP5* | rs1110236 | G | A | 0.1348 | -0.09 | 0.01 | 5.0119E-18 | 81.00 |
| *RBP5* | rs11370710 | C | CA | 0.4936 | -0.053 | 0.007 | 7.9433E-14 | 57.33 |
| *RBP5* | rs76904513 | A | G | 0.0878 | -0.511 | 0.013 | 1E-200 | 1545.09 |
| *RBP7* | rs12990177 | A | T | 0.5188 | -0.049 | 0.007 | 1.2589E-11 | 49.00 |
| *RBP7* | rs9657541 | C | T | 0.2055 | -0.06 | 0.009 | 6.3096E-12 | 44.44 |
| *RBP7* | rs7296211 | T | A | 0.7253 | -0.056 | 0.008 | 1.2589E-12 | 49.00 |
| *RBP7* | rs34436714 | C | A | 0.2092 | 0.325 | 0.009 | 1E-200 | 1304.01 |
| *RBPMS* | rs3021304 | G | C | 0.4476 | -0.072 | 0.008 | 1.5849E-20 | 81.00 |
| *RBPMS2* | rs1354034 | T | C | 0.6045 | 0.1 | 0.008 | 3.1623E-39 | 156.25 |
| *RBPMS2* | rs342299 | C | T | 0.4549 | -0.055 | 0.008 | 2.5119E-13 | 47.27 |
| *RBPMS2* | rs6993770 | A | T | 0.2866 | -0.056 | 0.008 | 1.2589E-11 | 49.00 |
| *RBPMS2* | rs73396521 | G | A | 0.2449 | 0.061 | 0.009 | 3.9811E-12 | 45.94 |
| *RBPMS2* | rs10876550 | G | A | 0.5566 | 0.069 | 0.008 | 1E-19 | 74.39 |
| *RBPMS2* | rs28711906 | T | A | 0.0482 | 0.181 | 0.018 | 6.3096E-25 | 101.11 |
| *RCC1* | rs12881562 | C | G | 0.1458 | 0.077 | 0.011 | 7.9433E-12 | 49.00 |
| *RCOR1* | rs7214731 | C | T | 0.2772 | 0.062 | 0.009 | 5.0119E-13 | 47.46 |

| | | | | | | | | |
|---|---|---|---|---|---|---|---|---|
| *RECK* | rs13135092 | A | G | 0.0836 | 0.087 | 0.013 | 1.2589E-11 | 44.79 |
| *RECK* | rs71521299 | T | G | 0.0162 | -0.853 | 0.029 | 1.995E-190 | 865.17 |
| *RECK* | rs10793962 | A | T | 0.0611 | 0.134 | 0.015 | 3.1623E-19 | 79.80 |
| *RECK* | rs56278466 | T | G | 0.6614 | 0.091 | 0.007 | 2.5119E-34 | 169.00 |
| *RECK* | rs603424 | G | A | 0.1662 | -0.069 | 0.01 | 5.0119E-13 | 47.61 |
| *RECK* | rs186021206 | G | A | 0.0065 | 0.354 | 0.048 | 2.5119E-13 | 54.39 |
| *RECK* | rs4760 | A | G | 0.1559 | -0.07 | 0.01 | 6.3096E-13 | 49.00 |
| *REEP4* | rs1354034 | T | C | 0.6043 | 0.063 | 0.008 | 1.5849E-16 | 62.02 |
| *REEP4* | rs117291678 | G | C | 0.0178 | -0.597 | 0.028 | 1.995E-100 | 454.60 |
| *REEP4* | rs541453292 | T | G | 0.0026 | 1.703 | 0.076 | 5.012E-111 | 502.11 |
| *REG1A* | rs11126696 | A | G | 0.6181 | 0.251 | 0.008 | 1E-200 | 984.39 |
| *REG1A* | rs9274437 | C | T | 0.2523 | 0.065 | 0.009 | 2.5119E-14 | 52.16 |
| *REG1A* | rs529565 | T | C | 0.3174 | 0.139 | 0.008 | 1.5849E-73 | 301.89 |
| *REG1A* | rs4421693 | G | T | 0.5762 | 0.189 | 0.007 | 1.259E-146 | 729.00 |
| *REG1A* | rs708686 | C | T | 0.2677 | 0.106 | 0.008 | 2.5119E-39 | 175.56 |
| *REG1A* | rs679574 | C | G | 0.5095 | -0.186 | 0.007 | 7.943E-147 | 706.04 |
| *REG1B* | rs11126696 | A | G | 0.6187 | 0.345 | 0.008 | 1E-200 | 1859.77 |
| *REG1B* | rs529565 | T | C | 0.3175 | 0.156 | 0.008 | 2.5119E-94 | 380.25 |
| *REG1B* | rs4421693 | G | T | 0.5763 | 0.119 | 0.007 | 6.3096E-61 | 289.00 |
| *REG1B* | rs72802342 | C | A | 0.0755 | -0.189 | 0.014 | 7.9433E-44 | 182.25 |
| *REG1B* | rs708686 | C | T | 0.2678 | 0.09 | 0.008 | 1.5849E-29 | 126.56 |
| *REG1B* | rs492602 | A | G | 0.5089 | -0.183 | 0.007 | 1.995E-144 | 683.45 |
| *REG3A* | rs147616162 | T | A | 0.0589 | 0.407 | 0.015 | 1E-155 | 736.22 |
| *REG3A* | rs11707109 | A | G | 0.1435 | -0.079 | 0.01 | 1E-14 | 62.41 |
| *REG3A* | rs35846682 | C | T | 0.0923 | 0.127 | 0.012 | 7.9433E-25 | 112.01 |
| *REG3A* | rs12201703 | G | A | 0.0789 | -0.165 | 0.013 | 3.1623E-36 | 161.09 |
| *REG3A* | rs11761603 | T | C | 0.6996 | 0.056 | 0.008 | 1.9953E-12 | 49.00 |
| *REG3A* | rs13253974 | G | A | 0.3242 | -0.061 | 0.008 | 7.9433E-16 | 58.14 |
| *REG3A* | rs17338413 | C | T | 0.0775 | -0.353 | 0.013 | 7.943E-155 | 737.33 |
| *REG3A* | rs570805211 | C | CT | 0.6366 | -0.051 | 0.007 | 6.3096E-12 | 53.08 |
| *REG3G* | rs2861742 | A | G | 0.7466 | -0.287 | 0.009 | 1E-200 | 1016.90 |
| *REG4* | rs79795228 | C | A | 0.0173 | 0.633 | 0.028 | 2.512E-115 | 511.08 |
| *REG4* | rs587654685 | C | T | 0.0104 | 0.492 | 0.042 | 2.5119E-31 | 137.22 |
| *REG4* | rs137872268 | T | C | 0.0176 | 0.26 | 0.028 | 7.9433E-21 | 86.22 |
| *RELB* | rs228682 | T | C | 0.3954 | 0.09 | 0.008 | 2.5119E-30 | 126.56 |
| *RELT* | rs13146355 | G | A | 0.4566 | 0.047 | 0.007 | 7.9433E-13 | 45.08 |
| *RELT* | rs11235729 | G | A | 0.2066 | 0.399 | 0.008 | 1E-200 | 2487.52 |
| *RELT* | rs3184504 | T | C | 0.5175 | -0.059 | 0.007 | 1E-19 | 71.04 |
| *RELT* | rs77924615 | G | A | 0.1966 | -0.064 | 0.008 | 1.5849E-14 | 64.00 |
| *REN* | rs1288923 | A | G | 0.4435 | 0.058 | 0.007 | 1.2589E-15 | 68.65 |
| *REN* | rs193280350 | G | A | 0.0076 | 0.838 | 0.043 | 1E-85 | 379.80 |
| *REN* | rs5051 | C | T | 0.3989 | -0.05 | 0.007 | 7.9433E-12 | 51.02 |
| *REN* | rs116219813 | C | T | 0.1277 | 0.098 | 0.011 | 5.0119E-20 | 79.37 |
| *RET* | rs6747755 | G | A | 0.2206 | 0.06 | 0.008 | 1.2589E-12 | 56.25 |
| *RET* | rs6762552 | C | T | 0.6902 | -0.083 | 0.008 | 7.9433E-28 | 107.64 |
| *RET* | rs10819413 | T | C | 0.6076 | -0.049 | 0.007 | 7.9433E-12 | 49.00 |
| *RET* | rs189906184 | A | T | 0.0241 | 0.393 | 0.025 | 1.5849E-56 | 247.12 |
| *RET* | rs2744077 | C | T | 0.1747 | 0.471 | 0.01 | 1E-200 | 2218.41 |
| *RET* | rs35887873 | T | C | 0.2286 | 0.082 | 0.008 | 1E-22 | 105.06 |
| *RET* | rs186021206 | G | A | 0.0063 | 0.465 | 0.049 | 1E-21 | 90.06 |
| *RET* | rs9952412 | A | C | 0.5174 | 0.048 | 0.007 | 7.9433E-12 | 47.02 |
| *RETN* | rs3917932 | C | G | 0.5757 | -0.062 | 0.007 | 3.9811E-17 | 78.45 |
| *RETN* | rs17405635 | G | A | 0.2556 | 0.098 | 0.008 | 1.9953E-31 | 150.06 |
| *RETN* | rs2305637 | C | T | 0.1756 | 0.076 | 0.01 | 1.9953E-15 | 57.76 |
| *RETN* | rs9880192 | G | C | 0.4142 | -0.052 | 0.007 | 3.1623E-12 | 55.18 |
| *RETN* | rs218264 | A | T | 0.2473 | 0.057 | 0.008 | 1.2589E-11 | 50.77 |
| *RETN* | rs13107325 | C | T | 0.075 | 0.109 | 0.014 | 3.9811E-15 | 60.62 |
| *RETN* | rs149110519 | C | T | 0.0363 | 0.384 | 0.02 | 5.0119E-86 | 368.64 |
| *RETN* | rs445 | C | T | 0.0952 | -0.11 | 0.012 | 5.0119E-19 | 84.03 |
| *RETN* | rs1865223 | G | T | 0.5677 | -0.086 | 0.007 | 7.9433E-32 | 150.94 |
| *RETN* | rs2074038 | G | T | 0.1001 | 0.115 | 0.012 | 1.5849E-21 | 91.84 |
| *RETN* | rs11642657 | C | T | 0.0078 | 0.313 | 0.044 | 7.9433E-13 | 50.60 |
| *RETN* | rs6503533 | C | T | 0.6164 | -0.109 | 0.007 | 1.2589E-48 | 242.47 |
| *RETN* | rs3219175 | G | A | 0.0076 | 1.543 | 0.046 | 1E-200 | 1125.16 |
| *RETN* | rs6069597 | T | C | 0.5309 | -0.06 | 0.007 | 7.9433E-17 | 73.47 |
| *RETN* | rs243591 | A | C | 0.4793 | -0.06 | 0.007 | 2.5119E-16 | 73.47 |
| *RFC4* | rs200303599 | T | TA | 0.0053 | 0.8 | 0.053 | 3.1623E-51 | 227.84 |
| *RGCC* | rs3811444 | C | T | 0.3327 | -0.062 | 0.008 | 6.3096E-15 | 60.06 |

| | | | | | | | | |
|---|---|---|---|---|---|---|---|---|
| *RGCC* | rs12767683 | C | A | 0.3252 | 0.059 | 0.008 | 1E-13 | 54.39 |
| *RGCC* | rs4759076 | T | C | 0.457 | 0.065 | 0.008 | 7.9433E-18 | 66.02 |
| *RGMA* | rs147440039 | C | G | 0.0308 | -0.151 | 0.021 | 3.1623E-13 | 51.70 |
| *RGMA* | rs1260326 | T | C | 0.6085 | 0.08 | 0.007 | 1.2589E-28 | 130.61 |
| *RGMA* | rs6804368 | A | G | 0.4441 | -0.066 | 0.007 | 2.5119E-20 | 88.90 |
| *RGMA* | rs13147041 | C | T | 0.3614 | -0.055 | 0.008 | 2.5119E-13 | 47.27 |
| *RGMA* | rs11956061 | C | T | 0.4811 | 0.069 | 0.007 | 3.1623E-22 | 97.16 |
| *RGMA* | rs4841132 | A | G | 0.9081 | -0.085 | 0.012 | 5.0119E-12 | 50.17 |
| *RGMA* | rs1885372 | A | G | 0.407 | -0.283 | 0.007 | 1E-200 | 1634.47 |
| *RGMA* | rs657152 | C | A | 0.3407 | 0.055 | 0.008 | 3.1623E-13 | 47.27 |
| *RGMA* | rs705702 | A | G | 0.3369 | 0.059 | 0.007 | 3.9811E-15 | 71.04 |
| *RGMA* | rs68109100 | A | AG | 0.5042 | 0.049 | 0.007 | 7.9433E-12 | 49.00 |
| *RGMA* | rs4775282 | A | T | 0.354 | -0.058 | 0.008 | 1.5849E-14 | 52.56 |
| *RGMA* | rs56058910 | T | C | 0.1385 | 0.081 | 0.011 | 1.5849E-14 | 54.22 |
| *RGMA* | rs3752102 | C | A | 0.4688 | -0.227 | 0.007 | 1E-200 | 1051.61 |
| *RGMA* | rs117353460 | C | T | 0.0332 | 0.159 | 0.02 | 7.9433E-16 | 63.20 |
| *RGMA* | rs2904766 | A | G | 0.6699 | 0.06 | 0.008 | 1.9953E-15 | 56.25 |
| *RGMB* | rs61747728 | C | T | 0.0379 | 0.138 | 0.019 | 2.5119E-13 | 52.75 |
| *RGMB* | rs56870134 | C | T | 0.5478 | 0.171 | 0.007 | 1E-116 | 596.76 |
| *RGMB* | rs765574776 | C | CG | 0.2625 | 0.058 | 0.008 | 7.9433E-12 | 52.56 |
| *RGMB* | rs12359178 | G | A | 0.1445 | 0.083 | 0.01 | 1E-15 | 68.89 |
| *RGMB* | rs34262842 | A | G | 0.1663 | -0.084 | 0.01 | 6.3096E-18 | 70.56 |
| *RGS10* | rs17622656 | G | A | 0.3882 | -0.055 | 0.008 | 1E-12 | 47.27 |
| *RGS10* | rs6993770 | A | T | 0.2864 | -0.059 | 0.008 | 1E-12 | 54.39 |
| *RGS8* | rs34917868 | G | A | 0.1421 | 0.239 | 0.011 | 1.259E-101 | 472.07 |
| *RHOC* | rs1006298 | G | C | 0.4817 | 0.084 | 0.008 | 3.9811E-28 | 110.25 |
| *RHOC* | rs1354034 | T | C | 0.604 | 0.063 | 0.008 | 3.9811E-16 | 62.02 |
| *RHOC* | rs342293 | C | G | 0.4588 | -0.054 | 0.008 | 1.5849E-12 | 45.56 |
| *RIDA* | rs77539382 | T | TC | 0.3193 | -0.318 | 0.008 | 1E-200 | 1580.06 |
| *RIDA* | rs7041363 | C | G | 0.4899 | -0.06 | 0.007 | 1E-16 | 73.47 |
| *RIDA* | rs3747207 | G | A | 0.2151 | 0.072 | 0.009 | 1.9953E-16 | 64.00 |
| *RILP* | rs1354034 | T | C | 0.6042 | 0.079 | 0.007 | 3.9811E-26 | 127.37 |
| *RILP* | rs6993770 | A | T | 0.2864 | -0.07 | 0.008 | 1E-17 | 76.56 |
| *RILP* | rs11604127 | C | T | 0.2308 | 0.06 | 0.009 | 7.9433E-12 | 44.44 |
| *RILP* | rs183827902 | C | T | 0.0172 | 0.268 | 0.028 | 3.9811E-21 | 91.61 |
| *RILPL2* | rs736801 | C | T | 0.388 | -0.053 | 0.008 | 5.0119E-12 | 43.89 |
| *RILPL2* | rs6993770 | A | T | 0.2866 | -0.07 | 0.008 | 1.9953E-17 | 76.56 |
| *RILPL2* | rs186269301 | G | A | 0.0285 | 0.235 | 0.022 | 1E-25 | 114.10 |
| *RIPK4* | rs3829431 | G | A | 0.1608 | 0.115 | 0.011 | 1E-25 | 109.30 |
| *RLN2* | rs41264945 | C | T | 0.0562 | 0.192 | 0.016 | 5.0119E-33 | 144.00 |
| *RLN2* | rs68066031 | T | C | 0.2299 | -0.07 | 0.009 | 5.0119E-15 | 60.49 |
| *RLN2* | rs2273782 | C | T | 0.2263 | 0.158 | 0.009 | 3.9811E-70 | 308.20 |
| *RNASE1* | rs4859682 | C | A | 0.4564 | 0.05 | 0.007 | 2.5119E-12 | 51.02 |
| *RNASE1* | rs17254387 | G | A | 0.6943 | 0.217 | 0.008 | 1E-163 | 735.77 |
| *RNASE10* | rs7144945 | C | T | 0.5074 | -0.803 | 0.008 | 1E-200 | 10075.14 |
| *RNASE3* | rs28498283 | A | T | 0.2546 | 0.064 | 0.009 | 5.0119E-14 | 50.57 |
| *RNASE3* | rs75004383 | G | A | 0.0422 | 0.129 | 0.018 | 1.5849E-12 | 51.36 |
| *RNASE3* | rs147307766 | C | T | 0.0537 | 0.569 | 0.017 | 1E-200 | 1120.28 |
| *RNASE4* | rs780094 | T | C | 0.6201 | -0.056 | 0.007 | 3.1623E-14 | 64.00 |
| *RNASE4* | rs1047891 | C | A | 0.3132 | -0.057 | 0.008 | 1.5849E-13 | 50.77 |
| *RNASE4* | rs28817415 | C | T | 0.4567 | 0.052 | 0.007 | 2.5119E-13 | 55.18 |
| *RNASE4* | rs12588573 | C | T | 0.2177 | -0.365 | 0.009 | 1E-200 | 1644.75 |
| *RNASE6* | rs9272278 | C | G | 0.1558 | 0.101 | 0.009 | 1.9953E-26 | 125.94 |
| *RNASE6* | rs117566084 | C | T | 0.0099 | 0.276 | 0.035 | 1.2589E-15 | 62.18 |
| *RNASE6* | rs3184504 | T | C | 0.5173 | -0.063 | 0.007 | 2.5119E-20 | 81.00 |
| *RNASE6* | rs1045922 | G | A | 0.2344 | -0.445 | 0.009 | 1E-200 | 2444.75 |
| *RNASE6* | rs145078947 | G | T | 0.0025 | 0.647 | 0.072 | 3.9811E-19 | 80.75 |
| *RNASEH2A* | rs750931137 | ATTTTTATT | A | 0.0258 | -0.313 | 0.024 | 2.5119E-39 | 170.09 |
| *RNASEH2A* | rs7247284 | T | C | 0.0332 | -0.286 | 0.021 | 6.3096E-42 | 185.48 |
| *RNASET2* | rs61747728 | C | T | 0.0381 | 0.133 | 0.018 | 7.9433E-14 | 54.60 |
| *RNASET2* | rs751785721 | A | ATT | 0.3289 | -0.054 | 0.007 | 2.5119E-13 | 59.51 |
| *RNASET2* | rs56193304 | T | G | 0.1364 | 0.073 | 0.01 | 6.3096E-13 | 53.29 |
| *RNASET2* | rs13107325 | C | T | 0.075 | 0.122 | 0.013 | 1E-20 | 88.07 |
| *RNASET2* | rs6914950 | C | T | 0.1476 | 0.263 | 0.01 | 1E-141 | 691.69 |
| *RNASET2* | rs3756838 | G | A | 0.2153 | 0.464 | 0.009 | 1E-200 | 2657.98 |
| *RNASET2* | rs12359178 | G | A | 0.1448 | 0.071 | 0.01 | 2.5119E-13 | 50.41 |
| *RNASET2* | rs149871778 | G | GAAC | 0.1046 | 0.086 | 0.011 | 2.5119E-14 | 61.12 |
| *RNASET2* | rs11111026 | G | T | 0.181 | -0.174 | 0.009 | 3.1623E-84 | 373.78 |

| | | | | | | | | |
|---|---|---|---|---|---|---|---|---|
| *RNASET2* | rs111338191 | A | AAATT | 0.5211 | -0.063 | 0.007 | 1E-19 | 81.00 |
| *RNASET2* | rs145078947 | G | T | 0.0026 | 1.331 | 0.068 | 6.3096E-86 | 383.12 |
| *RNF149* | rs61747728 | C | T | 0.0381 | 0.137 | 0.019 | 7.9433E-13 | 51.99 |
| *RNF149* | rs12621632 | C | T | 0.4065 | 0.281 | 0.008 | 1E-200 | 1233.77 |
| *RNF149* | rs10131857 | T | C | 0.4417 | 0.111 | 0.008 | 6.3096E-49 | 192.52 |
| *RNF168* | rs74750549 | T | C | 0.7075 | 0.062 | 0.009 | 1.2589E-11 | 47.46 |
| *RNF43* | rs28929474 | C | T | 0.0207 | 0.202 | 0.027 | 5.0119E-14 | 55.97 |
| *RNF43* | rs34523089 | C | T | 0.1574 | 0.139 | 0.011 | 3.1623E-40 | 159.68 |
| *RNF43* | rs201630261 | GT | G | 0.8983 | 0.101 | 0.013 | 3.9811E-15 | 60.36 |
| *RNF5* | rs1354034 | T | C | 0.6045 | 0.098 | 0.008 | 1.2589E-36 | 150.06 |
| *RNF5* | rs11794772 | G | A | 0.258 | -0.08 | 0.009 | 3.1623E-20 | 79.01 |
| *RNF5* | rs1976848 | T | A | 0.2142 | 0.083 | 0.01 | 6.3096E-18 | 68.89 |
| *RNF5* | rs11231074 | A | G | 0.2853 | -0.106 | 0.008 | 3.1623E-36 | 175.56 |
| *RNF5* | rs11671654 | C | G | 0.4953 | 0.09 | 0.008 | 2.5119E-32 | 126.56 |
| *ROBO1* | rs12752838 | A | G | 0.5064 | 0.06 | 0.007 | 1.5849E-16 | 73.47 |
| *ROBO1* | rs3773244 | G | A | 0.1969 | 0.236 | 0.009 | 3.981E-143 | 687.60 |
| *ROBO1* | rs10935478 | C | T | 0.445 | -0.06 | 0.007 | 7.9433E-16 | 73.47 |
| *ROBO1* | rs3849768 | C | A | 0.2095 | 0.064 | 0.009 | 7.9433E-13 | 50.57 |
| *ROBO1* | rs116017457 | G | T | 0.3487 | -0.089 | 0.008 | 1.9953E-29 | 123.77 |
| *ROBO1* | rs9987289 | A | G | 0.9079 | -0.085 | 0.013 | 1.5849E-11 | 42.75 |
| *ROBO1* | rs56278466 | T | G | 0.6616 | 0.12 | 0.008 | 6.3096E-55 | 225.00 |
| *ROBO1* | rs111338191 | A | AAATT | 0.521 | -0.061 | 0.007 | 6.3096E-17 | 75.94 |
| *ROBO1* | rs186021206 | G | A | 0.0064 | 0.674 | 0.05 | 2.5119E-41 | 181.71 |
| *ROBO1* | rs77542162 | A | G | 0.0229 | 0.186 | 0.024 | 1.9953E-14 | 60.06 |
| *ROBO2* | rs775722 | T | C | 0.4463 | -0.103 | 0.007 | 2.5119E-47 | 216.51 |
| *ROBO2* | rs67370636 | C | T | 0.3345 | 0.078 | 0.008 | 3.9811E-22 | 95.06 |
| *ROBO2* | rs9987289 | A | G | 0.9079 | -0.112 | 0.012 | 2.5119E-20 | 87.11 |
| *ROBO2* | rs56278466 | T | G | 0.6611 | 0.06 | 0.007 | 5.0119E-16 | 73.47 |
| *ROBO2* | rs186021206 | G | A | 0.0064 | 0.487 | 0.048 | 3.1623E-24 | 102.94 |
| *ROBO4* | rs2519093 | C | T | 0.1842 | -0.541 | 0.01 | 1E-200 | 2926.81 |
| *ROBO4* | rs59648931 | T | C | 0.2308 | -0.104 | 0.009 | 7.9433E-33 | 133.53 |
| *ROBO4* | rs200489612 | G | A | 0.0053 | 0.383 | 0.055 | 1.9953E-12 | 48.49 |
| *ROR1* | rs7532110 | T | A | 0.8034 | 0.067 | 0.009 | 3.1623E-13 | 55.42 |
| *ROR1* | rs6588083 | T | C | 0.1704 | -0.394 | 0.01 | 1E-200 | 1552.36 |
| *ROR1* | rs150816167 | T | C | 0.0418 | 0.207 | 0.019 | 6.3096E-29 | 118.70 |
| *ROR1* | rs1260326 | T | C | 0.6083 | 0.063 | 0.007 | 1E-17 | 81.00 |
| *ROR1* | rs75166367 | G | A | 0.0604 | -0.107 | 0.015 | 1E-12 | 50.88 |
| *ROR1* | rs13107325 | C | T | 0.0751 | 0.103 | 0.014 | 3.1623E-14 | 54.13 |
| *ROR1* | rs2298475 | T | C | 0.0752 | -0.102 | 0.013 | 2.5119E-14 | 61.56 |
| *ROR1* | rs77924615 | G | A | 0.1969 | -0.077 | 0.009 | 1.2589E-17 | 73.20 |
| *ROR1* | rs186021206 | G | A | 0.0064 | 0.465 | 0.049 | 1.5849E-21 | 90.06 |
| *ROR1* | rs2659005 | C | T | 0.4411 | 0.066 | 0.007 | 1.9953E-20 | 88.90 |
| *ROR1* | rs33950747 | C | T | 0.075 | 0.133 | 0.013 | 7.9433E-23 | 104.67 |
| *RP2* | rs429358 | T | C | 0.1553 | 0.145 | 0.011 | 5.0119E-43 | 173.76 |
| *RPA2* | rs17185038 | C | G | 0.0666 | 0.204 | 0.015 | 6.3096E-40 | 184.96 |
| *RPE* | rs2723211 | G | A | 0.4469 | 0.057 | 0.008 | 7.9433E-14 | 50.77 |
| *RPE* | rs9271696 | T | C | 0.3802 | -0.057 | 0.008 | 5.0119E-13 | 50.77 |
| *RPL14* | rs148850234 | C | T | 0.0523 | -0.118 | 0.017 | 7.9433E-12 | 48.18 |
| *RPL14* | rs1354034 | T | C | 0.6045 | 0.064 | 0.008 | 1.9953E-16 | 64.00 |
| *RRAS* | rs342298 | C | T | 0.4547 | -0.058 | 0.008 | 3.9811E-14 | 52.56 |
| *RRM2* | rs111338191 | A | AAATT | 0.5208 | -0.057 | 0.008 | 2.5119E-13 | 50.77 |
| *RRM2* | rs7247412 | T | C | 0.2668 | -0.067 | 0.009 | 1.5849E-14 | 55.42 |
| *RRM2B* | rs1354034 | T | C | 0.6038 | 0.089 | 0.008 | 1E-30 | 123.77 |
| *RRM2B* | rs5893603 | C | CG | 0.0781 | 0.194 | 0.014 | 3.1623E-42 | 192.02 |
| *RRM2B* | rs11604127 | C | T | 0.2307 | 0.062 | 0.009 | 1.5849E-11 | 47.46 |
| *RSPO1* | rs36043533 | T | G | 0.0564 | 0.478 | 0.016 | 3.981E-194 | 892.52 |
| *RSPO1* | rs13412535 | G | A | 0.2312 | -0.06 | 0.009 | 3.1623E-12 | 44.44 |
| *RSPO1* | rs1354034 | T | C | 0.6041 | -0.055 | 0.007 | 1.5849E-13 | 61.73 |
| *RSPO1* | rs6993770 | A | T | 0.2865 | -0.089 | 0.008 | 3.1623E-28 | 123.77 |
| *RSPO1* | rs61469632 | T | C | 0.0633 | -0.109 | 0.015 | 3.9811E-13 | 52.80 |
| *RSPO1* | rs3184504 | T | C | 0.517 | -0.065 | 0.007 | 2.5119E-19 | 86.22 |
| *RSPO1* | rs12445050 | C | T | 0.1381 | 0.078 | 0.011 | 1.2589E-13 | 50.28 |
| *RSPO1* | rs892090 | T | G | 0.8341 | 0.106 | 0.01 | 3.1623E-27 | 112.36 |
| *RSPO1* | rs13651 | T | C | 0.5124 | -0.085 | 0.007 | 1.2589E-31 | 147.45 |
| *RSPO3* | rs1936800 | C | T | 0.5272 | 0.157 | 0.007 | 1E-111 | 503.04 |
| *RSPO3* | rs11751347 | C | T | 0.104 | -0.119 | 0.012 | 3.9811E-25 | 98.34 |
| *RSPO3* | rs6993770 | A | T | 0.2863 | -0.063 | 0.008 | 2.5119E-16 | 62.02 |
| *RSPO3* | rs8050794 | T | C | 0.7041 | 0.063 | 0.008 | 1E-16 | 62.02 |

| | | | | | | | | |
|---|---|---|---|---|---|---|---|---|
| *RSPO3* | rs77924615 | G | A | 0.1966 | -0.061 | 0.009 | 5.0119E-12 | 45.94 |
| *RSPO3* | rs892090 | T | G | 0.8342 | 0.076 | 0.009 | 3.9811E-16 | 71.31 |
| *RSPO3* | rs2743388 | A | C | 0.5129 | -0.058 | 0.007 | 6.3096E-17 | 68.65 |
| *RTBDN* | rs7687767 | A | G | 0.1838 | 0.07 | 0.01 | 2.5119E-13 | 49.00 |
| *RTBDN* | rs7255045 | G | A | 0.2623 | -0.171 | 0.008 | 1.2589E-91 | 456.89 |
| *RTN4IP1* | rs2054365 | G | A | 0.4103 | 0.093 | 0.008 | 3.1623E-34 | 135.14 |
| *RTN4IP1* | rs4759076 | T | C | 0.457 | 0.062 | 0.008 | 3.1623E-16 | 60.06 |
| *RTN4R* | rs6658843 | G | C | 0.6112 | -0.062 | 0.007 | 1E-17 | 78.45 |
| *RTN4R* | rs1260326 | T | C | 0.6084 | -0.058 | 0.007 | 1.5849E-15 | 68.65 |
| *RTN4R* | rs1047891 | C | A | 0.3128 | 0.062 | 0.008 | 3.1623E-16 | 60.06 |
| *RTN4R* | rs6779146 | T | C | 0.2308 | -0.058 | 0.008 | 3.9811E-12 | 52.56 |
| *RTN4R* | rs2126259 | T | C | 0.8973 | -0.082 | 0.012 | 1.9953E-12 | 46.69 |
| *RTN4R* | rs28929474 | C | T | 0.0208 | 0.311 | 0.025 | 3.9811E-36 | 154.75 |
| *RTN4R* | rs56332871 | C | A | 0.2704 | 0.054 | 0.008 | 1E-11 | 45.56 |
| *RTN4R* | rs1065853 | G | T | 0.0813 | 0.116 | 0.013 | 3.9811E-19 | 79.62 |
| *RTN4R* | rs663353 | G | A | 0.7478 | -0.298 | 0.009 | 1E-200 | 1096.35 |
| *RUVBL1* | rs1312 | T | C | 0.4112 | -0.239 | 0.008 | 1.995E-182 | 892.52 |
| *RWDD1* | rs1354034 | T | C | 0.6042 | 0.074 | 0.008 | 7.9433E-23 | 85.56 |
| *RWDD1* | rs147899431 | G | C | 0.0434 | 0.307 | 0.018 | 6.3096E-63 | 290.89 |
| *RWDD1* | rs60822569 | T | TTTCTC | 0.5544 | 0.055 | 0.008 | 3.1623E-13 | 47.27 |
| *S100A11* | rs2999553 | G | C | 0.0423 | 0.326 | 0.018 | 3.1623E-72 | 328.01 |
| *S100A11* | rs34436714 | C | A | 0.2087 | 0.229 | 0.009 | 2.512E-141 | 647.42 |
| *S100A12* | rs3014874 | G | A | 0.2647 | -0.237 | 0.008 | 5.012E-176 | 877.64 |
| *S100A12* | rs6984820 | C | T | 0.4193 | 0.051 | 0.007 | 5.0119E-12 | 53.08 |
| *S100A12* | rs10982156 | T | A | 0.0695 | 0.137 | 0.015 | 7.9433E-20 | 83.42 |
| *S100A12* | rs34436714 | C | A | 0.2091 | 0.168 | 0.009 | 3.9811E-77 | 348.44 |
| *S100A13* | rs114678888 | C | A | 0.0405 | -0.504 | 0.019 | 3.981E-158 | 703.65 |
| *S100A13* | rs61729512 | G | A | 0.1179 | 0.079 | 0.011 | 6.3096E-12 | 51.58 |
| *S100A13* | rs10405357 | T | C | 0.4352 | -0.066 | 0.007 | 3.9811E-19 | 88.90 |
| *S100A14* | rs11548102 | A | T | 0.4894 | -0.175 | 0.007 | 6.31E-121 | 625.00 |
| *S100A14* | rs35853577 | T | C | 0.4256 | -0.068 | 0.008 | 2.5119E-19 | 72.25 |
| *S100A14* | rs2971446 | G | C | 0.8405 | -0.07 | 0.01 | 7.9433E-12 | 49.00 |
| *S100A14* | rs2291428 | G | C | 0.2435 | -0.072 | 0.009 | 7.9433E-17 | 64.00 |
| *S100A14* | rs56398830 | G | A | 0.0111 | -0.608 | 0.035 | 7.9433E-66 | 301.77 |
| *S100A14* | rs35193388 | G | GC | 0.7356 | -0.065 | 0.009 | 1.5849E-14 | 52.16 |
| *S100A14* | rs12938714 | G | A | 0.33 | 0.126 | 0.008 | 7.9433E-57 | 248.06 |
| *S100A14* | rs2318997 | T | G | 0.4338 | -0.062 | 0.008 | 1.5849E-16 | 60.06 |
| *S100A14* | rs601338 | G | A | 0.5089 | -0.109 | 0.007 | 5.0119E-48 | 242.47 |
| *S100A14* | rs3746778 | G | A | 0.4204 | 0.052 | 0.008 | 1.2589E-11 | 42.25 |
| *S100A16* | rs9700928 | A | C | 0.4862 | -0.104 | 0.008 | 2.5119E-40 | 169.00 |
| *S100A16* | rs56398830 | G | A | 0.011 | -0.288 | 0.037 | 6.3096E-15 | 60.59 |
| *S100A16* | rs601338 | G | A | 0.5093 | -0.075 | 0.008 | 3.9811E-22 | 87.89 |
| *S100A4* | rs56350425 | T | C | 0.0507 | 0.361 | 0.017 | 3.162E-101 | 450.94 |
| *S100A4* | rs6993770 | A | T | 0.2861 | -0.058 | 0.008 | 1.2589E-12 | 52.56 |
| *S100A4* | rs150813342 | C | T | 0.0085 | -0.351 | 0.042 | 3.1623E-17 | 69.84 |
| *S100A4* | rs3817621 | G | C | 0.2402 | -0.061 | 0.009 | 3.1623E-12 | 45.94 |
| *S100G* | rs784504 | G | C | 0.8115 | 0.07 | 0.01 | 5.0119E-13 | 49.00 |
| *S100G* | rs1169288 | A | C | 0.3142 | -0.092 | 0.008 | 3.1623E-29 | 132.25 |
| *S100P* | rs12613605 | G | T | 0.2127 | 0.074 | 0.009 | 6.3096E-16 | 67.60 |
| *S100P* | rs7846314 | A | T | 0.1854 | 0.08 | 0.01 | 6.3096E-17 | 64.00 |
| *S100P* | rs6503533 | C | T | 0.6171 | -0.055 | 0.008 | 5.0119E-13 | 47.27 |
| *S100P* | rs7412 | C | T | 0.081 | 0.19 | 0.014 | 3.1623E-43 | 184.18 |
| *S100P* | rs4632248 | G | T | 0.2095 | 0.069 | 0.009 | 7.9433E-14 | 58.78 |
| *SAA4* | rs7117890 | G | A | 0.5239 | -0.364 | 0.008 | 1E-200 | 2070.25 |
| *SAMD9L* | rs1354034 | T | C | 0.6043 | 0.052 | 0.007 | 2.5119E-12 | 55.18 |
| *SAMD9L* | rs17165120 | A | C | 0.0689 | -0.567 | 0.015 | 1E-200 | 1428.84 |
| *SARG* | rs74441312 | T | G | 0.0186 | 0.263 | 0.028 | 1E-20 | 88.23 |
| *SARG* | rs2631367 | C | G | 0.5189 | 0.061 | 0.007 | 3.1623E-16 | 75.94 |
| *SARG* | rs139141690 | G | A | 0.0048 | -0.416 | 0.055 | 2.5119E-14 | 57.21 |
| *SARG* | rs10820606 | A | C | 0.2293 | -0.095 | 0.009 | 2.5119E-26 | 111.42 |
| *SARG* | rs11611693 | G | T | 0.429 | -0.052 | 0.008 | 6.3096E-12 | 42.25 |
| *SARG* | rs57171122 | C | T | 0.5222 | -0.074 | 0.007 | 5.0119E-23 | 111.76 |
| *SAT2* | rs13894 | G | A | 0.0727 | -1.014 | 0.015 | 1E-200 | 4569.76 |
| *SBSN* | rs61816761 | G | A | 0.0235 | 0.224 | 0.025 | 7.9433E-20 | 80.28 |
| *SBSN* | rs1260326 | T | C | 0.6085 | 0.054 | 0.007 | 7.9433E-14 | 59.51 |
| *SBSN* | rs141134546 | C | T | 0.0198 | 0.184 | 0.025 | 2.5119E-13 | 54.17 |
| *SBSN* | rs17082071 | T | G | 0.0203 | 0.193 | 0.025 | 7.9433E-15 | 59.60 |
| *SBSN* | rs138854007 | C | G | 0.0751 | -0.134 | 0.014 | 6.3096E-21 | 91.61 |

| | | | | | | | | |
|---|---|---|---|---|---|---|---|---|
| *SCAMP3* | rs1142287 | C | T | 0.2569 | -0.085 | 0.009 | 3.9811E-23 | 89.20 |
| *SCAMP3* | rs1354034 | T | C | 0.6046 | 0.082 | 0.008 | 3.1623E-27 | 105.06 |
| *SCAMP3* | rs342293 | C | G | 0.4591 | -0.058 | 0.007 | 1E-14 | 68.65 |
| *SCAMP3* | rs6993770 | A | T | 0.2864 | -0.061 | 0.008 | 1.5849E-13 | 58.14 |
| *SCARA5* | rs149340229 | T | TA | 0.0369 | 0.133 | 0.018 | 1.2589E-13 | 54.60 |
| *SCARA5* | rs2726951 | C | T | 0.2686 | -0.337 | 0.008 | 1E-200 | 1774.52 |
| *SCARA5* | rs8176719 | T | TC | 0.3405 | 0.103 | 0.007 | 6.3096E-47 | 216.51 |
| *SCARA5* | rs4055121 | C | T | 0.135 | -0.082 | 0.01 | 1.5849E-16 | 67.24 |
| *SCARA5* | rs10774624 | G | A | 0.5134 | 0.047 | 0.007 | 3.9811E-12 | 45.08 |
| *SCARA5* | rs186021206 | G | A | 0.0064 | 0.374 | 0.046 | 7.9433E-16 | 66.10 |
| *SCARB1* | rs113075502 | G | A | 0.2264 | 0.07 | 0.01 | 1.2589E-11 | 49.00 |
| *SCARB2* | rs10935473 | G | T | 0.4406 | -0.053 | 0.007 | 5.0119E-14 | 57.33 |
| *SCARB2* | rs28563976 | A | C | 0.146 | -0.315 | 0.01 | 1E-200 | 992.25 |
| *SCARB2* | rs1042133 | G | C | 0.1573 | 0.081 | 0.01 | 5.0119E-17 | 65.61 |
| *SCARB2* | rs13192569 | G | A | 0.1386 | -0.07 | 0.01 | 5.0119E-12 | 49.00 |
| *SCARB2* | rs62018815 | G | A | 0.0266 | -0.221 | 0.022 | 2.5119E-24 | 100.91 |
| *SCARF1* | rs3811444 | C | T | 0.3323 | -0.053 | 0.008 | 3.1623E-12 | 43.89 |
| *SCARF1* | rs342298 | C | T | 0.4547 | -0.053 | 0.007 | 3.1623E-13 | 57.33 |
| *SCARF1* | rs6993770 | A | T | 0.2865 | -0.123 | 0.008 | 1.9953E-53 | 236.39 |
| *SCARF1* | rs579459 | T | C | 0.2077 | -0.064 | 0.009 | 1E-12 | 50.57 |
| *SCARF1* | rs111338191 | A | AAATT | 0.5209 | -0.069 | 0.007 | 3.9811E-21 | 97.16 |
| *SCARF1* | rs2272011 | G | A | 0.5911 | -0.379 | 0.008 | 1E-200 | 2244.39 |
| *SCARF1* | rs892090 | T | G | 0.8342 | 0.086 | 0.01 | 1.5849E-18 | 73.96 |
| *SCARF2* | rs61747728 | C | T | 0.0379 | 0.125 | 0.017 | 6.3096E-14 | 54.07 |
| *SCARF2* | rs1260326 | T | C | 0.6085 | 0.044 | 0.007 | 1.5849E-11 | 39.51 |
| *SCARF2* | rs765335847 | AAAGT | A | 0.2519 | 0.069 | 0.007 | 1.9953E-20 | 97.16 |
| *SCARF2* | rs507666 | G | A | 0.1848 | -0.121 | 0.008 | 1.9953E-48 | 228.77 |
| *SCARF2* | rs4055121 | C | T | 0.135 | -0.073 | 0.009 | 6.3096E-15 | 65.79 |
| *SCARF2* | rs215225 | A | G | 0.4281 | 0.057 | 0.006 | 6.3096E-19 | 90.25 |
| *SCARF2* | rs77924615 | G | A | 0.1967 | -0.058 | 0.008 | 1E-12 | 52.56 |
| *SCARF2* | rs186021206 | G | A | 0.0064 | 0.61 | 0.044 | 2.5119E-44 | 192.20 |
| *SCARF2* | rs5763025 | A | C | 0.8022 | 0.374 | 0.008 | 1E-200 | 2185.56 |
| *SCG2* | rs1761879 | A | G | 0.6777 | 0.066 | 0.008 | 1.9953E-16 | 68.06 |
| *SCG2* | rs635634 | C | T | 0.1842 | -0.069 | 0.01 | 1E-12 | 47.61 |
| *SCG2* | rs77924615 | G | A | 0.1967 | -0.071 | 0.009 | 5.0119E-14 | 62.23 |
| *SCG2* | rs681343 | C | T | 0.5092 | 0.073 | 0.007 | 1.9953E-22 | 108.76 |
| *SCG3* | rs61801010 | A | G | 0.106 | 0.082 | 0.011 | 1.9953E-13 | 55.57 |
| *SCG3* | rs6798507 | G | A | 0.6445 | 0.052 | 0.007 | 2.5119E-14 | 55.18 |
| *SCG3* | rs781657 | A | G | 0.1839 | 0.108 | 0.008 | 1.5849E-37 | 182.25 |
| *SCG3* | rs806798 | T | C | 0.4682 | 0.052 | 0.007 | 1.5849E-15 | 55.18 |
| *SCG3* | rs3751198 | A | G | 0.6037 | 0.054 | 0.007 | 1.2589E-15 | 59.51 |
| *SCG3* | rs2606134 | G | C | 0.7567 | 0.708 | 0.009 | 1E-200 | 6188.44 |
| *SCG3* | rs186021206 | G | A | 0.0064 | 0.441 | 0.045 | 7.9433E-23 | 96.04 |
| *SCGB1A1* | rs761160638 | AAAAT | A | 0.0791 | -0.115 | 0.014 | 1E-16 | 67.47 |
| *SCGB1A1* | rs3856521 | G | A | 0.1511 | -0.069 | 0.01 | 1.5849E-12 | 47.61 |
| *SCGB1A1* | rs1515498 | A | G | 0.3625 | -0.106 | 0.007 | 1.9953E-47 | 229.31 |
| *SCGB1A1* | rs466502 | A | G | 0.4422 | -0.055 | 0.007 | 1.2589E-14 | 61.73 |
| *SCGB1A1* | rs10051184 | T | C | 0.4972 | -0.054 | 0.007 | 1E-14 | 59.51 |
| *SCGB1A1* | rs3210176 | T | C | 0.3718 | -0.057 | 0.007 | 1.2589E-14 | 66.31 |
| *SCGB1A1* | rs372203682 | TA | T | 0.5515 | -0.052 | 0.008 | 3.1623E-12 | 42.25 |
| *SCGB1A1* | rs7040029 | T | C | 0.7075 | -0.054 | 0.008 | 3.9811E-12 | 45.56 |
| *SCGB1A1* | rs7072547 | G | A | 0.1792 | -0.07 | 0.009 | 3.1623E-14 | 60.49 |
| *SCGB1A1* | rs10501135 | T | C | 0.4138 | -0.142 | 0.007 | 3.1623E-83 | 411.51 |
| *SCGB1A1* | rs3741240 | G | A | 0.3515 | -0.339 | 0.008 | 1E-200 | 1795.64 |
| *SCGB1A1* | rs7962469 | A | G | 0.646 | -0.087 | 0.007 | 3.1623E-32 | 154.47 |
| *SCGB1A1* | rs7963771 | C | T | 0.3351 | -0.055 | 0.008 | 5.0119E-13 | 47.27 |
| *SCGB1A1* | rs1766147 | T | C | 0.7023 | 0.06 | 0.008 | 1.2589E-14 | 56.25 |
| *SCGB1A1* | rs7182041 | A | C | 0.4403 | 0.083 | 0.007 | 3.1623E-31 | 140.59 |
| *SCGB1A1* | rs55826010 | G | A | 0.2381 | -0.072 | 0.008 | 3.1623E-18 | 81.00 |
| *SCGB1A1* | rs1997814 | T | G | 0.6817 | -0.059 | 0.008 | 1.9953E-14 | 54.39 |
| *SCGB3A1* | rs4971100 | G | A | 0.426 | 0.09 | 0.007 | 1E-34 | 165.31 |
| *SCGB3A1* | rs58235267 | C | G | 0.4886 | -0.067 | 0.007 | 5.0119E-20 | 91.61 |
| *SCGB3A1* | rs62279940 | G | A | 0.2647 | -0.057 | 0.008 | 6.3096E-12 | 50.77 |
| *SCGB3A1* | rs2432162 | T | C | 0.3685 | -0.171 | 0.008 | 1.995E-111 | 456.89 |
| *SCGB3A1* | rs307802 | T | C | 0.4059 | 0.118 | 0.007 | 3.9811E-56 | 284.16 |
| *SCGB3A1* | rs35795116 | G | A | 0.126 | -0.086 | 0.012 | 5.0119E-12 | 51.36 |
| *SCGB3A1* | rs141020575 | T | TA | 0.5386 | -0.082 | 0.007 | 2.5119E-29 | 137.22 |
| *SCGB3A1* | rs502857 | T | C | 0.7093 | -0.203 | 0.008 | 1.585E-137 | 643.89 |

| | | | | | | | | |
|---|---|---|---|---|---|---|---|---|
| *SCGB3A1* | rs3741240 | G | A | 0.3515 | 0.165 | 0.008 | 1.995E-100 | 425.39 |
| *SCGB3A1* | rs2238133 | A | C | 0.4186 | -0.059 | 0.007 | 2.5119E-15 | 71.04 |
| *SCGB3A1* | rs1927191 | A | G | 0.5716 | -0.056 | 0.007 | 2.5119E-14 | 64.00 |
| *SCGB3A1* | rs56156922 | T | C | 0.3246 | 0.087 | 0.008 | 7.9433E-29 | 118.27 |
| *SCGB3A1* | rs371181880 | A | G | 0.0513 | -0.118 | 0.017 | 1E-12 | 48.18 |
| *SCGB3A1* | rs139063750 | C | T | 0.0976 | 0.104 | 0.012 | 1.9953E-17 | 75.11 |
| *SCGB3A2* | rs72691756 | G | A | 0.1237 | 0.184 | 0.011 | 3.9811E-60 | 279.80 |
| *SCGB3A2* | rs34153848 | G | A | 0.3649 | 0.057 | 0.008 | 7.9433E-14 | 50.77 |
| *SCGB3A2* | rs58235267 | C | G | 0.4881 | -0.074 | 0.007 | 1.5849E-23 | 111.76 |
| *SCGB3A2* | rs1515498 | A | G | 0.3626 | -0.132 | 0.008 | 5.0119E-67 | 272.25 |
| *SCGB3A2* | rs17717320 | C | G | 0.1979 | -0.248 | 0.009 | 2.512E-154 | 759.31 |
| *SCGB3A2* | rs6905736 | C | A | 0.8488 | -0.086 | 0.01 | 1.2589E-16 | 73.96 |
| *SCGB3A2* | rs6927418 | C | T | 0.5568 | -0.052 | 0.007 | 3.1623E-12 | 55.18 |
| *SCGB3A2* | rs62405419 | G | T | 0.117 | -0.077 | 0.011 | 1.5849E-11 | 49.00 |
| *SCGB3A2* | rs12666406 | G | A | 0.5352 | -0.063 | 0.007 | 6.3096E-18 | 81.00 |
| *SCGB3A2* | rs4881548 | C | T | 0.4996 | -0.058 | 0.007 | 2.5119E-15 | 68.65 |
| *SCGB3A2* | rs7942701 | C | T | 0.4147 | 0.064 | 0.007 | 6.3096E-18 | 83.59 |
| *SCGB3A2* | rs3741240 | G | A | 0.3514 | 0.138 | 0.008 | 6.3096E-72 | 297.56 |
| *SCGB3A2* | rs7962469 | A | G | 0.646 | -0.093 | 0.008 | 1E-33 | 135.14 |
| *SCGB3A2* | rs2414000 | T | G | 0.4322 | 0.059 | 0.007 | 1.5849E-15 | 71.04 |
| *SCGB3A2* | rs173539 | C | T | 0.3316 | 0.067 | 0.008 | 5.0119E-18 | 70.14 |
| *SCGB3A2* | rs756719 | C | T | 0.2748 | -0.058 | 0.008 | 2.5119E-12 | 52.56 |
| *SCGB3A2* | rs12167717 | G | A | 0.2394 | -0.071 | 0.009 | 7.9433E-17 | 62.23 |
| *SCGN* | rs6859236 | G | A | 0.5145 | -0.104 | 0.008 | 1.2589E-43 | 169.00 |
| *SCGN* | rs10946788 | G | A | 0.2752 | -0.092 | 0.008 | 1E-27 | 132.25 |
| *SCGN* | rs9297994 | G | A | 0.6664 | 0.059 | 0.008 | 1.9953E-13 | 54.39 |
| *SCGN* | rs782134971 | GAAACTGC( | G | 0.2542 | -0.065 | 0.009 | 5.0119E-14 | 52.16 |
| *SCGN* | rs11043076 | C | T | 0.1336 | 0.09 | 0.011 | 7.9433E-16 | 66.94 |
| *SCGN* | rs601338 | G | A | 0.5092 | -0.136 | 0.008 | 7.9433E-72 | 289.00 |
| *SCLY* | rs62196005 | A | T | 0.2751 | 0.194 | 0.008 | 1.259E-127 | 588.06 |
| *SCLY* | rs4835265 | C | A | 0.1564 | 0.073 | 0.01 | 1E-13 | 53.29 |
| *SCLY* | rs10733608 | G | T | 0.4885 | -0.051 | 0.007 | 2.5119E-12 | 53.08 |
| *SCLY* | rs10883451 | T | C | 0.5018 | -0.052 | 0.007 | 5.0119E-13 | 55.18 |
| *SCLY* | rs58964858 | C | T | 0.1073 | -0.086 | 0.012 | 1.9953E-13 | 51.36 |
| *SCLY* | rs3184504 | T | C | 0.5166 | -0.05 | 0.007 | 1.5849E-12 | 51.02 |
| *SCLY* | rs112635299 | G | T | 0.0216 | 0.171 | 0.025 | 3.9811E-12 | 46.79 |
| *SCLY* | rs3747207 | G | A | 0.2155 | 0.074 | 0.009 | 2.5119E-17 | 67.60 |
| *SCN4B* | rs1260326 | T | C | 0.6084 | 0.05 | 0.007 | 1E-14 | 51.02 |
| *SCN4B* | rs9273757 | G | C | 0.2326 | 0.053 | 0.008 | 1E-11 | 43.89 |
| *SCN4B* | rs35632913 | G | GT | 0.6387 | -0.66 | 0.008 | 1E-200 | 6806.25 |
| *SCP2* | rs17200109 | C | T | 0.0567 | -0.118 | 0.017 | 3.1623E-12 | 48.18 |
| *SCPEP1* | rs6993770 | A | T | 0.2864 | -0.085 | 0.008 | 3.1623E-25 | 112.89 |
| *SCPEP1* | rs1031101 | A | G | 0.1418 | 0.081 | 0.011 | 2.5119E-14 | 54.22 |
| *SCPEP1* | rs10761756 | C | T | 0.4698 | 0.053 | 0.007 | 5.0119E-13 | 57.33 |
| *SCPEP1* | rs111338191 | A | AAATT | 0.5203 | -0.052 | 0.007 | 1.9953E-12 | 55.18 |
| *SCPEP1* | rs34449028 | A | T | 0.3008 | -0.365 | 0.009 | 1E-200 | 1644.75 |
| *SCRIB* | rs2621380 | C | T | 0.5539 | -0.053 | 0.008 | 3.9811E-12 | 43.89 |
| *SCRN1* | rs1434282 | C | T | 0.7261 | 0.095 | 0.008 | 5.0119E-29 | 141.02 |
| *SCRN1* | rs17324153 | T | C | 0.0428 | 0.267 | 0.019 | 7.9433E-47 | 197.48 |
| *SCT* | rs13264644 | A | G | 0.432 | 0.058 | 0.008 | 7.9433E-14 | 52.56 |
| *SDC1* | rs907866 | G | A | 0.4474 | 0.131 | 0.007 | 7.9433E-71 | 350.22 |
| *SDC1* | rs1260326 | T | C | 0.6083 | 0.073 | 0.007 | 7.9433E-23 | 108.76 |
| *SDC1* | rs13107325 | C | T | 0.0751 | 0.335 | 0.014 | 1.259E-127 | 572.58 |
| *SDC1* | rs28522260 | C | A | 0.1675 | -0.072 | 0.01 | 1.2589E-13 | 51.84 |
| *SDC1* | rs9987289 | A | G | 0.9076 | -0.11 | 0.013 | 1.2589E-18 | 71.60 |
| *SDC1* | rs532436 | G | A | 0.1848 | -0.105 | 0.009 | 1E-28 | 136.11 |
| *SDC1* | rs11438680 | G | GA | 0.4605 | 0.061 | 0.007 | 6.3096E-17 | 75.94 |
| *SDC1* | rs10881600 | T | C | 0.5525 | 0.049 | 0.007 | 1.2589E-11 | 49.00 |
| *SDC1* | rs186021206 | G | A | 0.0064 | 0.668 | 0.05 | 5.0119E-41 | 178.49 |
| *SDC1* | rs2204211 | G | A | 0.1025 | 0.088 | 0.012 | 2.5119E-13 | 53.78 |
| *SDC1* | rs620982 | T | C | 0.4562 | -0.075 | 0.007 | 6.3096E-25 | 114.80 |
| *SDC1* | rs601338 | G | A | 0.5091 | 0.076 | 0.007 | 2.5119E-25 | 117.88 |
| *SDC1* | rs2294915 | C | T | 0.2316 | 0.058 | 0.009 | 1.2589E-11 | 41.53 |
| *SDC4* | rs6662263 | A | G | 0.8039 | 0.072 | 0.009 | 1E-14 | 64.00 |
| *SDC4* | rs1354034 | T | C | 0.6037 | -0.083 | 0.007 | 1E-28 | 140.59 |
| *SDC4* | rs2735095 | C | A | 0.3202 | -0.088 | 0.008 | 5.0119E-29 | 121.00 |
| *SDC4* | rs778641135 | TTAC | T | 0.2579 | 0.061 | 0.008 | 5.0119E-13 | 58.14 |
| *SDC4* | rs61177759 | C | T | 0.2008 | -0.062 | 0.009 | 1.5849E-11 | 47.46 |

| | | | | | | | | |
|---|---|---|---|---|---|---|---|---|
| *SDC4* | rs11763765 | C | T | 0.1477 | -0.173 | 0.01 | 1.2589E-62 | 299.29 |
| *SDC4* | rs139141690 | G | A | 0.0048 | -0.448 | 0.054 | 1E-16 | 68.83 |
| *SDC4* | rs6993770 | A | T | 0.2866 | -0.121 | 0.008 | 1.2589E-50 | 228.77 |
| *SDC4* | rs385893 | T | C | 0.5237 | 0.087 | 0.007 | 1.9953E-32 | 154.47 |
| *SDC4* | rs61750929 | C | T | 0.055 | -0.112 | 0.016 | 3.9811E-12 | 49.00 |
| *SDC4* | rs8193003 | T | C | 0.6074 | -0.056 | 0.008 | 1.9953E-13 | 49.00 |
| *SDC4* | rs58641111 | C | CGT | 0.4657 | -0.083 | 0.007 | 6.3096E-29 | 140.59 |
| *SDC4* | rs12762934 | C | T | 0.3246 | 0.102 | 0.008 | 1E-38 | 162.56 |
| *SDC4* | rs33972805 | C | CCT | 0.527 | -0.055 | 0.007 | 1.2589E-13 | 61.73 |
| *SDC4* | rs11553699 | A | G | 0.1356 | 0.17 | 0.011 | 1.9953E-52 | 238.84 |
| *SDC4* | rs12881545 | G | C | 0.6684 | 0.057 | 0.008 | 2.5119E-13 | 50.77 |
| *SDC4* | rs34890846 | A | ATTG | 0.189 | -0.076 | 0.009 | 5.0119E-16 | 71.31 |
| *SDC4* | rs4783186 | T | C | 0.8742 | -0.076 | 0.011 | 3.9811E-12 | 47.74 |
| *SDC4* | rs1654425 | T | C | 0.834 | 0.076 | 0.01 | 1.5849E-14 | 57.76 |
| *SDC4* | rs4411786 | T | C | 0.2633 | -0.064 | 0.008 | 3.1623E-14 | 64.00 |
| *SDC4* | rs2251577 | G | C | 0.5129 | -0.223 | 0.007 | 7.943E-198 | 1014.88 |
| *SDC4* | rs2229742 | G | C | 0.1033 | 0.087 | 0.012 | 3.9811E-13 | 52.56 |
| *SDCCAG8* | rs953492 | A | G | 0.534 | 0.203 | 0.007 | 6.31E-165 | 841.00 |
| *SDCCAG8* | rs1354034 | T | C | 0.6044 | 0.086 | 0.007 | 1.2589E-30 | 150.94 |
| *SDHB* | rs4361977 | C | T | 0.4335 | -0.055 | 0.008 | 6.3096E-13 | 47.27 |
| *SDHB* | rs4835265 | C | A | 0.156 | 0.14 | 0.01 | 1.2589E-41 | 196.00 |
| *SDHB* | rs10075805 | A | G | 0.2757 | -0.06 | 0.008 | 1E-12 | 56.25 |
| *SDHB* | rs4581712 | C | A | 0.2716 | 0.058 | 0.008 | 6.3096E-12 | 52.56 |
| *SDHB* | rs4245267 | C | A | 0.7888 | -0.096 | 0.009 | 1.5849E-25 | 113.78 |
| *SDK2* | rs116522341 | C | G | 0.0328 | 0.209 | 0.021 | 5.0119E-24 | 99.05 |
| *SDK2* | rs653178 | C | T | 0.5159 | -0.066 | 0.007 | 5.0119E-21 | 88.90 |
| *SDK2* | rs186021206 | G | A | 0.0064 | 0.574 | 0.052 | 7.9433E-29 | 121.85 |
| *SDK2* | rs11655934 | G | A | 0.1494 | -0.674 | 0.011 | 1E-200 | 3754.35 |
| *SEC31A* | rs1354034 | T | C | 0.6045 | 0.083 | 0.008 | 5.0119E-27 | 107.64 |
| *SEC31A* | rs11541628 | C | A | 0.0253 | -0.31 | 0.024 | 2.5119E-37 | 166.84 |
| *SEC31A* | rs342293 | C | G | 0.4595 | -0.058 | 0.008 | 2.5119E-14 | 52.56 |
| *SEC31A* | rs6993770 | A | T | 0.2866 | -0.063 | 0.008 | 6.3096E-14 | 62.02 |
| *SEL1L* | rs11499034 | T | C | 0.0148 | -1.255 | 0.032 | 1E-200 | 1538.11 |
| *SEL1L* | rs28929474 | C | T | 0.0208 | 0.557 | 0.027 | 1.9953E-97 | 425.58 |
| *SELE* | rs599839 | G | A | 0.7718 | 0.07 | 0.007 | 5.0119E-21 | 100.00 |
| *SELE* | rs7538317 | C | G | 0.6555 | -0.095 | 0.007 | 6.3096E-47 | 184.18 |
| *SELE* | rs10935473 | G | T | 0.4399 | -0.088 | 0.006 | 5.0119E-45 | 215.11 |
| *SELE* | rs13135092 | A | G | 0.0835 | 0.08 | 0.011 | 1.2589E-12 | 52.89 |
| *SELE* | rs2519093 | C | T | 0.1843 | -0.978 | 0.01 | 1E-200 | 9564.84 |
| *SELE* | rs35166255 | G | A | 0.0338 | 0.324 | 0.017 | 1.5849E-78 | 363.24 |
| *SELE* | rs7979473 | A | G | 0.6165 | -0.057 | 0.006 | 1.2589E-18 | 90.25 |
| *SELE* | rs186021206 | G | A | 0.0064 | 0.587 | 0.043 | 7.9433E-43 | 186.35 |
| *SELE* | rs17855739 | C | T | 0.0411 | 0.179 | 0.016 | 2.5119E-30 | 125.16 |
| *SELENOP* | rs10038285 | G | A | 0.7327 | 0.103 | 0.009 | 6.3096E-34 | 130.98 |
| *SELL* | rs2223286 | T | C | 0.3107 | -0.407 | 0.008 | 1E-200 | 2588.27 |
| *SELL* | rs139015452 | C | CACTG | 0.2291 | 0.058 | 0.008 | 2.5119E-12 | 52.56 |
| *SELL* | rs9824474 | A | G | 0.3672 | -0.051 | 0.007 | 1E-12 | 53.08 |
| *SELL* | rs218265 | T | C | 0.1532 | 0.073 | 0.01 | 3.1623E-14 | 53.29 |
| *SELL* | rs6796 | T | C | 0.2791 | 0.077 | 0.008 | 1E-23 | 92.64 |
| *SELL* | rs612169 | A | G | 0.318 | 0.051 | 0.007 | 3.9811E-12 | 53.08 |
| *SELL* | rs1794089 | G | A | 0.2339 | 0.084 | 0.008 | 3.1623E-24 | 110.25 |
| *SELL* | rs7137828 | C | T | 0.516 | -0.107 | 0.007 | 1E-54 | 233.65 |
| *SELL* | rs76428106 | T | C | 0.0129 | 0.326 | 0.032 | 5.0119E-25 | 103.79 |
| *SELL* | rs186021206 | G | A | 0.0064 | 0.726 | 0.047 | 1.5849E-53 | 238.60 |
| *SELL* | rs12941811 | T | C | 0.5744 | -0.069 | 0.007 | 1.5849E-23 | 97.16 |
| *SELL* | rs7224668 | T | C | 0.5571 | -0.049 | 0.007 | 2.5119E-12 | 49.00 |
| *SELP* | rs6136 | T | G | 0.1136 | -0.403 | 0.012 | 1E-200 | 1127.84 |
| *SELP* | rs56043070 | G | A | 0.0733 | 0.114 | 0.014 | 1.9953E-15 | 66.31 |
| *SELP* | rs11242109 | G | T | 0.4814 | -0.062 | 0.007 | 3.9811E-17 | 78.45 |
| *SELP* | rs6961069 | C | T | 0.4025 | 0.056 | 0.007 | 1E-13 | 64.00 |
| *SELP* | rs6993770 | A | T | 0.2865 | -0.116 | 0.008 | 3.1623E-46 | 210.25 |
| *SELP* | rs2519093 | C | T | 0.184 | -0.219 | 0.01 | 1.995E-116 | 479.61 |
| *SELP* | rs7896518 | A | G | 0.424 | 0.074 | 0.007 | 3.1623E-23 | 111.76 |
| *SELP* | rs12445050 | C | T | 0.1384 | 0.099 | 0.011 | 7.9433E-21 | 81.00 |
| *SELP* | rs892090 | T | G | 0.8345 | 0.136 | 0.01 | 5.0119E-43 | 184.96 |
| *SELPLG* | rs343808 | C | T | 0.242 | -0.074 | 0.008 | 6.3096E-19 | 85.56 |
| *SELPLG* | rs2990223 | A | G | 0.6051 | 0.143 | 0.007 | 6.3096E-86 | 417.33 |
| *SELPLG* | rs10214273 | T | G | 0.2734 | -0.068 | 0.008 | 7.9433E-18 | 72.25 |

| | | | | | | | | |
|---|---|---|---|---|---|---|---|---|
| *SELPLG* | rs4248814 | G | A | 0.2022 | 0.071 | 0.009 | 6.3096E-16 | 62.23 |
| *SELPLG* | rs10093797 | C | A | 0.7517 | -0.058 | 0.008 | 1.2589E-12 | 52.56 |
| *SELPLG* | rs2142306 | T | C | 0.4117 | 0.049 | 0.007 | 1.2589E-11 | 49.00 |
| *SELPLG* | rs536644 | G | A | 0.5459 | 0.099 | 0.007 | 1.5849E-43 | 200.02 |
| *SELPLG* | rs10819317 | A | G | 0.1656 | 0.096 | 0.01 | 7.9433E-24 | 92.16 |
| *SELPLG* | rs7294337 | C | T | 0.016 | -1.974 | 0.03 | 1E-200 | 4329.64 |
| *SELPLG* | rs60005225 | G | GT | 0.3816 | -0.052 | 0.007 | 1.2589E-12 | 55.18 |
| *SEMA3F* | rs554375669 | C | CT | 0.2072 | -0.176 | 0.01 | 3.9811E-72 | 309.76 |
| *SEMA3F* | rs4679317 | G | A | 0.686 | -0.075 | 0.008 | 3.9811E-21 | 87.89 |
| *SEMA3F* | rs782134971 | GAAACTGC( | G | 0.2543 | -0.071 | 0.008 | 3.1623E-17 | 78.77 |
| *SEMA3F* | rs564699 | C | T | 0.5363 | -0.065 | 0.007 | 7.9433E-19 | 86.22 |
| *SEMA3F* | rs3184504 | T | C | 0.5174 | -0.062 | 0.007 | 1E-17 | 78.45 |
| *SEMA3F* | rs1037117 | G | A | 0.2522 | 0.065 | 0.008 | 1.9953E-14 | 66.02 |
| *SEMA3F* | rs8055162 | G | A | 0.5672 | -0.067 | 0.007 | 1.9953E-19 | 91.61 |
| *SEMA3G* | rs12740374 | G | T | 0.221 | -0.302 | 0.008 | 1E-200 | 1425.06 |
| *SEMA3G* | rs376333472 | G | A | 0.0245 | 0.182 | 0.023 | 7.9433E-16 | 62.62 |
| *SEMA3G* | rs2766588 | T | C | 0.607 | 0.458 | 0.008 | 1E-200 | 3277.56 |
| *SEMA3G* | rs4947530 | C | T | 0.954 | 0.111 | 0.016 | 1.5849E-11 | 48.13 |
| *SEMA3G* | rs2653414 | C | A | 0.0128 | 0.305 | 0.03 | 1E-23 | 103.36 |
| *SEMA3G* | rs117119759 | G | A | 0.0292 | 0.142 | 0.021 | 6.3096E-12 | 45.72 |
| *SEMA3G* | rs4747199 | C | T | 0.2883 | -0.092 | 0.008 | 5.0119E-34 | 132.25 |
| *SEMA3G* | rs72802342 | C | A | 0.0759 | -0.215 | 0.013 | 3.9811E-60 | 273.52 |
| *SEMA3G* | rs5848 | C | T | 0.2747 | -0.101 | 0.008 | 1.2589E-38 | 159.39 |
| *SEMA4C* | rs756513821 | A | G | 0.0011 | -1.044 | 0.126 | 1.2589E-16 | 68.65 |
| *SEMA4C* | rs537675308 | G | C | 0.001 | -0.834 | 0.122 | 1E-11 | 46.73 |
| *SEMA4C* | rs34794906 | T | C | 0.2643 | 0.062 | 0.008 | 1.5849E-13 | 60.06 |
| *SEMA4C* | rs564699 | C | T | 0.5361 | -0.056 | 0.007 | 3.1623E-14 | 64.00 |
| *SEMA4C* | rs186021206 | G | A | 0.0063 | 0.505 | 0.051 | 5.0119E-23 | 98.05 |
| *SEMA4C* | rs6010224 | A | C | 0.4431 | 0.05 | 0.007 | 1.2589E-11 | 51.02 |
| *SEMA4D* | rs1354034 | T | C | 0.6042 | 0.101 | 0.008 | 6.3096E-40 | 159.39 |
| *SEMA4D* | rs6993770 | A | T | 0.2865 | -0.056 | 0.008 | 1.5849E-11 | 49.00 |
| *SEMA4D* | rs36020742 | C | T | 0.1831 | 0.107 | 0.01 | 5.0119E-28 | 114.49 |
| *SEMA4D* | rs7080386 | C | A | 0.4122 | 0.054 | 0.008 | 1.9953E-12 | 45.56 |
| *SEMA4D* | rs7949566 | G | A | 0.4218 | 0.067 | 0.008 | 1.9953E-18 | 70.14 |
| *SEMA4D* | rs111338191 | A | AAATT | 0.5209 | -0.093 | 0.008 | 3.1623E-34 | 135.14 |
| *SEMA6C* | rs758673791 | TAGAA | T | 0.7807 | 0.111 | 0.009 | 5.0119E-33 | 152.11 |
| *SEMA7A* | rs61801010 | A | G | 0.1062 | 0.095 | 0.012 | 1.9953E-14 | 62.67 |
| *SEMA7A* | rs7606173 | G | C | 0.432 | -0.063 | 0.007 | 6.3096E-18 | 81.00 |
| *SEMA7A* | rs2009581 | G | A | 0.2726 | -0.057 | 0.008 | 3.9811E-12 | 50.77 |
| *SEMA7A* | rs3864106 | G | C | 0.5843 | -0.187 | 0.007 | 3.162E-139 | 713.65 |
| *SEMA7A* | rs28716466 | T | C | 0.2966 | 0.07 | 0.008 | 3.1623E-18 | 76.56 |
| *SEMA7A* | rs13135092 | A | G | 0.0834 | 0.094 | 0.013 | 1.2589E-12 | 52.28 |
| *SEMA7A* | rs9263850 | A | G | 0.2799 | 0.06 | 0.008 | 1.5849E-13 | 56.25 |
| *SEMA7A* | rs58217236 | T | C | 0.553 | -0.059 | 0.007 | 1.2589E-15 | 71.04 |
| *SEMA7A* | rs2347784 | C | G | 0.2715 | -0.083 | 0.008 | 1.9953E-24 | 107.64 |
| *SEMA7A* | rs12554596 | A | G | 0.2038 | -0.067 | 0.009 | 1E-13 | 55.42 |
| *SEMA7A* | rs10793962 | A | T | 0.0614 | 0.225 | 0.015 | 6.3096E-50 | 225.00 |
| *SEMA7A* | rs56278466 | T | G | 0.6614 | 0.162 | 0.008 | 3.9811E-98 | 410.06 |
| *SEMA7A* | rs73119306 | A | G | 0.2461 | 0.062 | 0.008 | 2.5119E-13 | 60.06 |
| *SEMA7A* | rs3184504 | T | C | 0.5168 | -0.115 | 0.007 | 6.3096E-57 | 269.90 |
| *SEMA7A* | rs608394 | C | T | 0.5059 | -0.073 | 0.007 | 1.5849E-23 | 108.76 |
| *SEMA7A* | rs78994380 | C | A | 0.1118 | -0.231 | 0.012 | 3.9811E-84 | 370.56 |
| *SEMA7A* | rs200489612 | G | A | 0.0052 | 0.705 | 0.055 | 3.9811E-38 | 164.31 |
| *SEMA7A* | rs34557412 | A | G | 0.0068 | 0.554 | 0.044 | 3.9811E-36 | 158.53 |
| *SEMA7A* | rs485186 | A | G | 0.5374 | 0.05 | 0.007 | 7.9433E-12 | 51.02 |
| *SEPTIN3* | rs115126145 | C | T | 0.0217 | 0.181 | 0.025 | 6.3096E-13 | 52.42 |
| *SEPTIN3* | rs3777182 | T | A | 0.2667 | 0.098 | 0.008 | 7.9433E-32 | 150.06 |
| *SEPTIN3* | rs686250 | A | G | 0.5681 | -0.084 | 0.008 | 3.1623E-28 | 110.25 |
| *SEPTIN3* | rs751503974 | ATG | A | 0.2585 | -0.067 | 0.009 | 5.0119E-14 | 55.42 |
| *SEPTIN3* | rs75430132 | C | T | 0.1624 | -0.069 | 0.01 | 7.9433E-12 | 47.61 |
| *SEPTIN3* | rs61896141 | A | C | 0.1915 | 0.067 | 0.009 | 1.5849E-12 | 55.42 |
| *SEPTIN3* | rs550897 | A | G | 0.4616 | 0.233 | 0.007 | 1E-200 | 1107.94 |
| *SEPTIN3* | rs117775520 | G | A | 0.0361 | -0.196 | 0.02 | 1.2589E-23 | 96.04 |
| *SEPTIN3* | rs34562254 | G | A | 0.0969 | 0.12 | 0.012 | 1.9953E-22 | 100.00 |
| *SEPTIN3* | rs9624334 | G | C | 0.151 | 0.09 | 0.01 | 3.9811E-18 | 81.00 |
| *SEPTIN8* | rs11204523 | G | C | 0.7585 | -0.07 | 0.009 | 3.1623E-16 | 60.49 |
| *SEPTIN8* | rs7665521 | T | C | 0.5768 | -0.071 | 0.007 | 6.3096E-22 | 102.88 |
| *SEPTIN8* | rs30512 | C | T | 0.1122 | -0.145 | 0.011 | 6.3096E-37 | 173.76 |

| | | | | | | | | |
|---|---|---|---|---|---|---|---|---|
| *SEPTIN9* | rs55645755 | A | G | 0.0155 | -0.355 | 0.031 | 1.2589E-30 | 131.14 |
| *SEPTIN9* | rs10432210 | C | T | 0.2087 | 0.065 | 0.009 | 2.5119E-12 | 52.16 |
| *SERPINA1* | rs17580 | T | A | 0.0496 | -0.374 | 0.018 | 3.981E-100 | 431.72 |
| *SERPINA11* | rs1260326 | T | C | 0.6084 | -0.066 | 0.006 | 1.9953E-29 | 121.00 |
| *SERPINA11* | rs4634107 | G | T | 0.5234 | 0.056 | 0.006 | 3.9811E-23 | 87.11 |
| *SERPINA11* | rs13108218 | A | G | 0.619 | -0.044 | 0.006 | 1.5849E-13 | 53.78 |
| *SERPINA11* | rs144861591 | C | T | 0.0768 | -0.076 | 0.011 | 1.9953E-12 | 47.74 |
| *SERPINA11* | rs10740131 | A | T | 0.4698 | 0.057 | 0.006 | 3.9811E-23 | 90.25 |
| *SERPINA11* | rs7139079 | G | A | 0.5946 | -0.044 | 0.006 | 3.1623E-14 | 53.78 |
| *SERPINA11* | rs12898137 | T | A | 0.4111 | -0.621 | 0.007 | 1E-200 | 7870.22 |
| *SERPINA11* | rs7188155 | C | G | 0.3104 | 0.043 | 0.006 | 3.9811E-12 | 51.36 |
| *SERPINA11* | rs12976652 | T | C | 0.1192 | -0.097 | 0.009 | 1.5849E-28 | 116.16 |
| *SERPINA12* | rs759945735 | A | AT | 0.2853 | -0.056 | 0.007 | 2.5119E-16 | 64.00 |
| *SERPINA12* | rs1495743 | G | C | 0.7779 | -0.051 | 0.007 | 7.9433E-12 | 53.08 |
| *SERPINA12* | rs174533 | G | A | 0.3463 | -0.046 | 0.006 | 1.2589E-12 | 58.78 |
| *SERPINA12* | rs17091005 | T | C | 0.2184 | 0.778 | 0.009 | 1E-200 | 7472.64 |
| *SERPINA12* | rs339969 | C | A | 0.6169 | -0.095 | 0.006 | 3.9811E-51 | 250.69 |
| *SERPINA12* | rs738408 | C | T | 0.2163 | 0.055 | 0.007 | 1.2589E-13 | 61.73 |
| *SERPINA3* | rs11208702 | A | C | 0.1952 | -0.073 | 0.009 | 5.0119E-15 | 65.79 |
| *SERPINA3* | rs28601761 | C | G | 0.4194 | 0.053 | 0.007 | 1E-12 | 57.33 |
| *SERPINA3* | rs61976127 | A | G | 0.1853 | -0.447 | 0.01 | 1E-200 | 1998.09 |
| *SERPINA4* | rs1260326 | T | C | 0.6082 | -0.076 | 0.007 | 7.9433E-28 | 117.88 |
| *SERPINA4* | rs146996008 | A | AGT | 0.4164 | 0.055 | 0.007 | 2.5119E-15 | 61.73 |
| *SERPINA4* | rs1230167 | A | T | 0.6717 | 0.065 | 0.007 | 6.3096E-19 | 86.22 |
| *SERPINA4* | rs1051921 | G | A | 0.196 | -0.067 | 0.009 | 7.9433E-15 | 55.42 |
| *SERPINA4* | rs2264782 | C | T | 0.3476 | 0.056 | 0.007 | 1E-14 | 64.00 |
| *SERPINA4* | rs5511 | A | T | 0.2204 | 0.617 | 0.009 | 1E-200 | 4699.86 |
| *SERPINA5* | rs1260326 | T | C | 0.6083 | -0.092 | 0.008 | 1.2589E-34 | 132.25 |
| *SERPINA5* | rs710446 | T | C | 0.409 | 0.06 | 0.007 | 1E-15 | 73.47 |
| *SERPINA5* | rs3733402 | G | A | 0.5123 | 0.18 | 0.007 | 5.012E-132 | 661.22 |
| *SERPINA5* | rs1801020 | A | G | 0.7448 | 0.193 | 0.008 | 6.31E-116 | 582.02 |
| *SERPINA5* | rs1130267 | G | A | 0.5988 | 0.201 | 0.008 | 3.162E-153 | 631.27 |
| *SERPINA5* | rs7212366 | C | T | 0.4918 | 0.054 | 0.008 | 6.3096E-12 | 45.56 |
| *SERPINA6* | rs550899198 | T | TG | 0.8278 | 0.067 | 0.009 | 3.9811E-14 | 55.42 |
| *SERPINA6* | rs5785581 | C | CA | 0.476 | 0.057 | 0.007 | 2.5119E-17 | 66.31 |
| *SERPINA6* | rs139097404 | T | C | 0.0226 | 0.177 | 0.023 | 7.9433E-15 | 59.22 |
| *SERPINA7* | rs60935030 | T | TAATTA | 0.7482 | -0.06 | 0.009 | 7.9433E-12 | 44.44 |
| *SERPINA7* | rs10095930 | C | T | 0.5836 | 0.06 | 0.008 | 3.9811E-15 | 56.25 |
| *SERPINA7* | rs1169288 | A | C | 0.3142 | 0.086 | 0.008 | 1.9953E-26 | 115.56 |
| *SERPINA9* | rs9264768 | T | C | 0.1022 | 0.075 | 0.011 | 1.2589E-11 | 46.49 |
| *SERPINA9* | rs10849448 | A | G | 0.7537 | -0.055 | 0.008 | 3.1623E-12 | 47.27 |
| *SERPINA9* | rs2038499 | C | T | 0.2823 | -0.498 | 0.008 | 1E-200 | 3875.06 |
| *SERPINB1* | rs6925835 | C | T | 0.1759 | 0.102 | 0.01 | 3.1623E-25 | 104.04 |
| *SERPINB1* | rs6993770 | A | T | 0.2863 | -0.082 | 0.008 | 2.5119E-23 | 105.06 |
| *SERPINB5* | rs148729762 | AAAAC | A | 0.6637 | 0.056 | 0.008 | 6.3096E-12 | 49.00 |
| *SERPINB6* | rs9405601 | G | A | 0.438 | 0.161 | 0.008 | 7.943E-101 | 405.02 |
| *SERPINB6* | rs6993770 | A | T | 0.2866 | -0.064 | 0.008 | 7.9433E-15 | 64.00 |
| *SERPINB8* | rs3826616 | A | G | 0.581 | -0.727 | 0.008 | 1E-200 | 8258.27 |
| *SERPINB8* | rs4632248 | G | T | 0.2096 | 0.099 | 0.007 | 3.9811E-40 | 200.02 |
| *SERPINB9* | rs7751676 | C | T | 0.0642 | 0.223 | 0.015 | 1E-47 | 221.02 |
| *SERPINB9* | rs6993770 | A | T | 0.2865 | -0.07 | 0.008 | 3.9811E-17 | 76.56 |
| *SERPINC1* | rs2227624 | A | T | 0.0034 | -0.871 | 0.063 | 1.2589E-43 | 191.14 |
| *SERPINC1* | rs1260326 | T | C | 0.6082 | -0.089 | 0.008 | 1.2589E-32 | 123.77 |
| *SERPINC1* | rs431204 | C | T | 0.1246 | -0.081 | 0.011 | 1.5849E-12 | 54.22 |
| *SERPINC1* | rs782470771 | AAG | A | 0.1277 | -0.087 | 0.011 | 3.1623E-15 | 62.55 |
| *SERPINC1* | rs7920036 | T | C | 0.475 | -0.067 | 0.007 | 5.0119E-20 | 91.61 |
| *SERPINC1* | rs123698 | G | C | 0.6049 | 0.058 | 0.007 | 7.9433E-15 | 68.65 |
| *SERPIND1* | rs1260326 | T | C | 0.6082 | -0.111 | 0.007 | 6.3096E-51 | 251.45 |
| *SERPIND1* | rs2856451 | A | G | 0.5142 | 0.055 | 0.007 | 3.1623E-14 | 61.73 |
| *SERPIND1* | rs36019311 | G | GTA | 0.1293 | -0.1 | 0.011 | 1.5849E-20 | 82.64 |
| *SERPIND1* | rs217184 | T | C | 0.1963 | 0.319 | 0.009 | 1E-200 | 1256.31 |
| *SERPIND1* | rs393665 | A | T | 0.7032 | 0.066 | 0.008 | 6.3096E-17 | 68.06 |
| *SERPIND1* | rs117254553 | G | T | 0.0027 | -0.872 | 0.076 | 1.5849E-30 | 131.65 |
| *SERPIND1* | rs738408 | C | T | 0.217 | -0.064 | 0.009 | 1.5849E-13 | 50.57 |
| *SERPINE1* | rs2274319 | T | C | 0.6531 | -0.058 | 0.008 | 7.9433E-14 | 52.56 |
| *SERPINE1* | rs13412535 | G | A | 0.231 | -0.061 | 0.009 | 3.9811E-12 | 45.94 |
| *SERPINE1* | rs78909033 | G | A | 0.1359 | -0.085 | 0.011 | 5.0119E-15 | 59.71 |
| *SERPINE1* | rs7618405 | C | A | 0.2048 | -0.064 | 0.009 | 2.5119E-12 | 50.57 |

| | | | | | | | | |
|---|---|---|---|---|---|---|---|---|
| *SERPINE1* | rs1354034 | T | C | 0.6039 | -0.121 | 0.008 | 1E-57 | 228.77 |
| *SERPINE1* | rs3792376 | T | G | 0.5938 | -0.052 | 0.008 | 5.0119E-12 | 42.25 |
| *SERPINE1* | rs4572884 | C | T | 0.3977 | -0.058 | 0.008 | 1.9953E-14 | 52.56 |
| *SERPINE1* | rs114694170 | T | C | 0.06 | 0.15 | 0.016 | 1.2589E-21 | 87.89 |
| *SERPINE1* | rs2631367 | C | G | 0.5186 | 0.057 | 0.007 | 1.9953E-14 | 66.31 |
| *SERPINE1* | rs35173808 | C | A | 0.0966 | -0.085 | 0.013 | 1.5849E-11 | 42.75 |
| *SERPINE1* | rs6961069 | C | T | 0.4026 | 0.066 | 0.008 | 3.9811E-18 | 68.06 |
| *SERPINE1* | rs2227674 | A | G | 0.2 | -0.119 | 0.009 | 6.3096E-37 | 174.83 |
| *SERPINE1* | rs6993770 | A | T | 0.2865 | -0.16 | 0.008 | 7.9433E-85 | 400.00 |
| *SERPINE1* | rs61469632 | T | C | 0.0632 | -0.138 | 0.015 | 1.5849E-19 | 84.64 |
| *SERPINE1* | rs34377578 | A | C | 0.2529 | 0.071 | 0.009 | 1E-16 | 62.23 |
| *SERPINE1* | rs6580981 | G | A | 0.4584 | 0.064 | 0.007 | 7.9433E-18 | 83.59 |
| *SERPINE1* | rs61978213 | G | A | 0.0417 | 0.127 | 0.018 | 7.9433E-12 | 49.78 |
| *SERPINE1* | rs60128101 | T | A | 0.1749 | 0.07 | 0.01 | 7.9433E-13 | 49.00 |
| *SERPINE1* | rs12445050 | C | T | 0.1384 | 0.099 | 0.011 | 1.9953E-20 | 81.00 |
| *SERPINE1* | rs1654425 | T | C | 0.8343 | 0.159 | 0.01 | 6.3096E-57 | 252.81 |
| *SERPINE1* | rs3790176 | G | A | 0.3424 | 0.07 | 0.008 | 3.9811E-19 | 76.56 |
| *SERPINE2* | rs12086222 | G | C | 0.111 | 0.078 | 0.011 | 1E-11 | 50.28 |
| *SERPINE2* | rs68066031 | T | C | 0.2299 | -0.632 | 0.009 | 1E-200 | 4931.16 |
| *SERPINE2* | rs7618405 | C | A | 0.2047 | -0.069 | 0.009 | 1E-14 | 58.78 |
| *SERPINE2* | rs1354034 | T | C | 0.6045 | -0.062 | 0.007 | 3.1623E-17 | 78.45 |
| *SERPINE2* | rs10016018 | A | T | 0.3975 | -0.054 | 0.007 | 2.5119E-13 | 59.51 |
| *SERPINE2* | rs2516471 | G | A | 0.0635 | -0.101 | 0.015 | 1.2589E-11 | 45.34 |
| *SERPINE2* | rs6961069 | C | T | 0.4023 | 0.052 | 0.007 | 1.9953E-12 | 55.18 |
| *SERPINE2* | rs6993770 | A | T | 0.2866 | -0.122 | 0.008 | 3.1623E-53 | 232.56 |
| *SERPINE2* | rs61469632 | T | C | 0.0633 | -0.119 | 0.015 | 1.2589E-15 | 62.94 |
| *SERPINE2* | rs7080386 | C | A | 0.412 | 0.085 | 0.007 | 7.9433E-31 | 147.45 |
| *SERPINE2* | rs12445050 | C | T | 0.1383 | 0.099 | 0.01 | 1.9953E-21 | 98.01 |
| *SERPINE2* | rs1654425 | T | C | 0.8344 | 0.142 | 0.01 | 3.9811E-48 | 201.64 |
| *SERPINE2* | rs6081569 | G | C | 0.3684 | 0.061 | 0.007 | 3.9811E-16 | 75.94 |
| *SERPINF1* | rs61747728 | C | T | 0.038 | 0.166 | 0.019 | 3.9811E-19 | 76.33 |
| *SERPINF1* | rs62088172 | C | T | 0.3533 | -0.283 | 0.008 | 1E-200 | 1251.39 |
| *SERPINF2* | rs140584594 | A | G | 0.7322 | 0.064 | 0.008 | 5.0119E-15 | 64.00 |
| *SERPINF2* | rs1260326 | T | C | 0.6081 | -0.09 | 0.007 | 3.1623E-33 | 165.31 |
| *SERPINF2* | rs451637 | C | T | 0.3098 | -0.06 | 0.008 | 1.9953E-13 | 56.25 |
| *SERPINF2* | rs11078597 | T | C | 0.1881 | -0.207 | 0.009 | 1E-107 | 529.00 |
| *SERPING1* | rs140584594 | A | G | 0.7322 | 0.063 | 0.008 | 5.0119E-15 | 62.02 |
| *SERPING1* | rs9427402 | G | C | 0.1154 | 0.094 | 0.011 | 5.0119E-17 | 73.02 |
| *SERPING1* | rs1260326 | T | C | 0.6082 | -0.105 | 0.007 | 3.9811E-46 | 225.00 |
| *SERPING1* | rs35332062 | G | A | 0.1265 | -0.084 | 0.011 | 7.9433E-15 | 58.31 |
| *SERPING1* | rs9987289 | A | G | 0.9079 | 0.091 | 0.012 | 2.5119E-13 | 57.51 |
| *SERPING1* | rs112055915 | A | G | 0.0691 | -0.096 | 0.014 | 1E-11 | 47.02 |
| *SERPING1* | rs150336831 | A | T | 0.0418 | 0.184 | 0.019 | 1E-22 | 93.78 |
| *SERPING1* | rs28362944 | T | C | 0.049 | -0.425 | 0.017 | 1.585E-140 | 625.00 |
| *SERPING1* | rs10849546 | G | A | 0.1353 | -0.184 | 0.01 | 5.0119E-69 | 338.56 |
| *SERPING1* | rs28929474 | C | T | 0.0209 | -0.268 | 0.025 | 1E-26 | 114.92 |
| *SERPINH1* | rs1354034 | T | C | 0.6041 | 0.061 | 0.008 | 1.9953E-15 | 58.14 |
| *SERPINH1* | rs9886239 | A | C | 0.3312 | 0.064 | 0.008 | 1E-15 | 64.00 |
| *SERPINH1* | rs7080386 | C | A | 0.4118 | 0.08 | 0.008 | 7.9433E-26 | 100.00 |
| *SERPINH1* | rs636418 | A | T | 0.5794 | 0.137 | 0.008 | 6.3096E-70 | 293.27 |
| *SERPINH1* | rs2006166 | T | C | 0.1179 | -0.12 | 0.013 | 5.0119E-20 | 85.21 |
| *SERPINI1* | rs11717419 | G | A | 0.221 | 0.462 | 0.009 | 1E-200 | 2635.11 |
| *SERPINI1* | rs4253328 | C | CTTT | 0.5132 | 0.148 | 0.007 | 1E-95 | 447.02 |
| *SERPINI1* | rs1801020 | A | G | 0.7446 | 0.138 | 0.008 | 3.9811E-65 | 297.56 |
| *SERPINI1* | rs56278466 | T | G | 0.6609 | 0.053 | 0.007 | 1E-12 | 57.33 |
| *SERPINI1* | rs10740118 | G | C | 0.4142 | 0.05 | 0.007 | 3.1623E-12 | 51.02 |
| *SERPINI1* | rs79629788 | C | T | 0.0235 | 0.163 | 0.023 | 2.5119E-12 | 50.22 |
| *SERPINI2* | rs145849315 | T | C | 0.0971 | -0.088 | 0.012 | 3.1623E-14 | 53.78 |
| *SERPINI2* | rs35004807 | T | TGTAA | 0.3441 | 0.067 | 0.007 | 3.1623E-23 | 91.61 |
| *SERPINI2* | rs2487583 | G | A | 0.1091 | -0.076 | 0.01 | 7.9433E-14 | 57.76 |
| *SERPINI2* | rs17032925 | T | C | 0.0825 | -0.142 | 0.012 | 1.5849E-34 | 140.03 |
| *SERPINI2* | rs376333472 | G | A | 0.0248 | 0.219 | 0.021 | 5.0119E-26 | 108.76 |
| *SERPINI2* | rs6444344 | G | A | 0.1238 | 0.509 | 0.01 | 1E-200 | 2590.81 |
| *SERPINI2* | rs3215532 | G | GA | 0.2529 | 0.05 | 0.007 | 5.0119E-12 | 51.02 |
| *SERPINI2* | rs9398804 | T | A | 0.439 | -0.071 | 0.006 | 3.1623E-28 | 140.03 |
| *SERPINI2* | rs572393412 | C | CA | 0.3097 | 0.06 | 0.007 | 3.9811E-16 | 73.47 |
| *SERPINI2* | rs13258924 | A | G | 0.7126 | -0.058 | 0.007 | 3.9811E-16 | 68.65 |
| *SERPINI2* | rs4733612 | G | A | 0.7329 | 0.054 | 0.007 | 6.3096E-14 | 59.51 |

| | | | | | | | | |
|---|---|---|---|---|---|---|---|---|
| *SERPINI2* | rs8176743 | C | T | 0.0615 | 1.146 | 0.014 | 1E-200 | 6700.59 |
| *SERPINI2* | rs174564 | A | G | 0.3492 | 0.059 | 0.007 | 3.9811E-19 | 71.04 |
| *SERPINI2* | rs74997273 | A | T | 0.2678 | -0.06 | 0.007 | 7.9433E-17 | 73.47 |
| *SERPINI2* | rs12426349 | C | T | 0.142 | -0.079 | 0.009 | 1.9953E-18 | 77.05 |
| *SERPINI2* | rs9579128 | T | C | 0.4103 | -0.065 | 0.006 | 7.9433E-24 | 117.36 |
| *SERPINI2* | rs28365941 | T | C | 0.136 | -0.138 | 0.01 | 1.9953E-45 | 190.44 |
| *SERPINI2* | rs72802342 | C | A | 0.0758 | -0.631 | 0.013 | 1E-200 | 2355.98 |
| *SERPINI2* | rs1126464 | G | C | 0.2443 | -0.074 | 0.008 | 3.1623E-22 | 85.56 |
| *SERPINI2* | rs186021206 | G | A | 0.0065 | 0.405 | 0.043 | 1E-20 | 88.71 |
| *SERPINI2* | rs17138478 | C | A | 0.1305 | 0.071 | 0.009 | 6.3096E-14 | 62.23 |
| *SERPINI2* | rs62090602 | A | T | 0.0769 | 0.098 | 0.012 | 1.9953E-16 | 66.69 |
| *SERPINI2* | rs601338 | G | A | 0.5092 | 0.332 | 0.007 | 1E-200 | 2249.47 |
| *SERPINI2* | rs2410017 | C | T | 0.6147 | 0.054 | 0.007 | 1.2589E-16 | 59.51 |
| *SESTD1* | rs10170520 | A | T | 0.3291 | -0.104 | 0.008 | 1.2589E-37 | 169.00 |
| *SESTD1* | rs1354034 | T | C | 0.6035 | 0.055 | 0.008 | 1E-12 | 47.27 |
| *SESTD1* | rs10733789 | T | C | 0.3133 | 0.064 | 0.008 | 5.0119E-15 | 64.00 |
| *SETMAR* | rs35083095 | T | C | 0.4934 | -0.216 | 0.007 | 1.259E-194 | 952.16 |
| *SEZ6* | rs186021206 | G | A | 0.0064 | 0.435 | 0.052 | 1E-16 | 69.98 |
| *SEZ6* | rs778315867 | A | AAACA | 0.1244 | -0.096 | 0.012 | 1.5849E-16 | 64.00 |
| *SEZ6L* | rs61801010 | A | G | 0.1061 | 0.103 | 0.012 | 3.9811E-17 | 73.67 |
| *SEZ6L* | rs781657 | A | G | 0.1836 | 0.064 | 0.009 | 3.9811E-12 | 50.57 |
| *SEZ6L* | rs9270747 | A | G | 0.6575 | 0.055 | 0.008 | 3.9811E-13 | 47.27 |
| *SEZ6L* | rs10838702 | G | T | 0.3857 | 0.056 | 0.007 | 3.9811E-14 | 64.00 |
| *SEZ6L* | rs60194243 | C | T | 0.0592 | -0.114 | 0.015 | 3.9811E-14 | 57.76 |
| *SEZ6L* | rs1070073 | T | G | 0.6729 | 0.057 | 0.008 | 5.0119E-14 | 50.77 |
| *SEZ6L* | rs186021206 | G | A | 0.0064 | 0.488 | 0.049 | 3.1623E-23 | 99.19 |
| *SEZ6L* | rs137203 | G | T | 0.0191 | -0.571 | 0.026 | 3.981E-105 | 482.31 |
| *SEZ6L2* | rs1260326 | T | C | 0.6081 | 0.047 | 0.007 | 1.5849E-11 | 45.08 |
| *SEZ6L2* | rs1961950 | C | T | 0.0849 | -0.182 | 0.012 | 1.9953E-50 | 230.03 |
| *SEZ6L2* | rs2523567 | C | G | 0.2319 | -0.075 | 0.008 | 1.5849E-20 | 87.89 |
| *SEZ6L2* | rs56278466 | T | G | 0.6611 | 0.057 | 0.007 | 1E-15 | 66.31 |
| *SEZ6L2* | rs12372718 | A | G | 0.4079 | -0.082 | 0.007 | 1E-32 | 137.22 |
| *SEZ6L2* | rs113443718 | G | A | 0.3083 | 0.162 | 0.007 | 1.585E-106 | 535.59 |
| *SEZ6L2* | rs186021206 | G | A | 0.0063 | 0.663 | 0.047 | 3.9811E-45 | 198.99 |
| *SF3B4* | rs12078573 | G | A | 0.0787 | -0.097 | 0.014 | 5.0119E-12 | 48.01 |
| *SF3B4* | rs10067881 | G | A | 0.129 | -0.109 | 0.011 | 3.9811E-22 | 98.19 |
| *SF3B4* | rs6993770 | A | T | 0.2864 | -0.063 | 0.008 | 2.5119E-14 | 62.02 |
| *SFRP1* | rs2318761 | A | G | 0.4677 | 0.055 | 0.007 | 1.5849E-14 | 61.73 |
| *SFRP1* | rs72643819 | G | T | 0.3867 | -0.189 | 0.007 | 2.512E-140 | 729.00 |
| *SFRP1* | rs6420185 | G | A | 0.3878 | -0.062 | 0.007 | 1.2589E-16 | 78.45 |
| *SFRP1* | rs13651 | T | C | 0.5122 | -0.061 | 0.007 | 6.3096E-18 | 75.94 |
| *SFRP4* | rs7526027 | T | C | 0.2922 | -0.062 | 0.008 | 3.9811E-14 | 60.06 |
| *SFRP4* | rs75207237 | T | A | 0.1283 | 0.296 | 0.011 | 1.995E-155 | 724.10 |
| *SFRP4* | rs35887873 | T | C | 0.2278 | 0.138 | 0.009 | 1.9953E-55 | 235.11 |
| *SFRP4* | rs215223 | G | A | 0.4004 | 0.102 | 0.008 | 7.9433E-42 | 162.56 |
| *SFTPA1* | rs1965708 | G | T | 0.1872 | -0.284 | 0.01 | 3.162E-181 | 806.56 |
| *SFTPA2* | rs11675149 | C | A | 0.5711 | -0.047 | 0.007 | 1.5849E-12 | 45.08 |
| *SFTPA2* | rs6883627 | T | C | 0.4585 | 0.054 | 0.007 | 3.1623E-16 | 59.51 |
| *SFTPA2* | rs17886395 | C | G | 0.1319 | 0.94 | 0.011 | 1E-200 | 7302.48 |
| *SFTPA2* | rs7962469 | A | G | 0.6455 | 0.05 | 0.007 | 5.0119E-13 | 51.02 |
| *SFTPA2* | rs2753532 | T | C | 0.8821 | -0.099 | 0.012 | 1E-17 | 68.06 |
| *SFTPA2* | rs117981338 | C | T | 0.0143 | 0.25 | 0.028 | 1E-18 | 79.72 |
| *SFTPD* | rs2075571 | C | T | 0.4244 | 0.042 | 0.006 | 1.9953E-13 | 49.00 |
| *SFTPD* | rs56218657 | G | A | 0.0383 | -0.115 | 0.015 | 7.9433E-15 | 58.78 |
| *SFTPD* | rs10031518 | T | A | 0.3407 | 0.056 | 0.006 | 5.0119E-21 | 87.11 |
| *SFTPD* | rs4262138 | A | T | 0.1427 | 0.065 | 0.008 | 1.2589E-15 | 66.02 |
| *SFTPD* | rs12660817 | A | G | 0.0554 | 0.694 | 0.013 | 1E-200 | 2849.92 |
| *SFTPD* | rs2451951 | T | C | 0.4948 | -0.113 | 0.006 | 1E-87 | 354.69 |
| *SFTPD* | rs721917 | A | G | 0.4179 | -0.758 | 0.007 | 1E-200 | 11725.80 |
| *SFTPD* | rs35705950 | G | T | 0.1104 | 0.066 | 0.009 | 3.1623E-13 | 53.78 |
| *SFTPD* | rs635804 | G | C | 0.285 | 0.043 | 0.006 | 6.3096E-12 | 51.36 |
| *SFTPD* | rs58462329 | G | C | 0.1447 | -0.214 | 0.008 | 1.585E-152 | 715.56 |
| *SFTPD* | rs369234167 | T | C | 0.2002 | -0.064 | 0.008 | 1.5849E-17 | 64.00 |
| *SGSH* | rs174557 | A | G | 0.3091 | 0.04 | 0.006 | 1.2589E-12 | 44.44 |
| *SGSH* | rs10778152 | A | G | 0.2878 | -0.219 | 0.006 | 1E-200 | 1332.25 |
| *SGSH* | rs145078947 | G | T | 0.0025 | 0.84 | 0.051 | 5.0119E-60 | 271.28 |
| *SGSH* | rs7503034 | C | T | 0.3314 | -0.886 | 0.007 | 1E-200 | 16020.33 |
| *SGSH* | rs58542926 | C | T | 0.0752 | 0.096 | 0.01 | 1.2589E-22 | 92.16 |

| | | | | | | | | |
|---|---|---|---|---|---|---|---|---|
| *SGSH* | rs429358 | T | C | 0.1551 | -0.051 | 0.007 | 1.5849E-12 | 53.08 |
| *SGSH* | rs738409 | C | G | 0.2169 | 0.055 | 0.006 | 1.9953E-18 | 84.03 |
| *SH2B3* | rs11950562 | A | C | 0.4784 | -0.055 | 0.008 | 5.0119E-13 | 47.27 |
| *SH2B3* | rs342293 | C | G | 0.4596 | -0.061 | 0.008 | 7.9433E-16 | 58.14 |
| *SH2B3* | rs6993770 | A | T | 0.2869 | -0.089 | 0.008 | 2.5119E-26 | 123.77 |
| *SH2B3* | rs409801 | T | C | 0.5073 | 0.053 | 0.008 | 2.5119E-12 | 43.89 |
| *SH2D1A* | rs3184504 | T | C | 0.5174 | -0.078 | 0.008 | 6.3096E-25 | 95.06 |
| *SH2D1A* | rs75763843 | A | C | 0.1265 | 0.128 | 0.011 | 1.9953E-29 | 135.40 |
| *SH3BP1* | rs12710562 | A | G | 0.1827 | 0.268 | 0.01 | 3.981E-161 | 718.24 |
| *SH3GLB2* | rs9866126 | A | G | 0.3398 | -0.094 | 0.008 | 1.5849E-34 | 138.06 |
| *SH3GLB2* | rs62585602 | G | A | 0.4754 | -0.273 | 0.007 | 1E-200 | 1521.00 |
| *SHBG* | rs114165349 | G | C | 0.0234 | -0.167 | 0.022 | 5.0119E-14 | 57.62 |
| *SHBG* | rs1730850 | C | T | 0.657 | -0.053 | 0.007 | 1E-13 | 57.33 |
| *SHBG* | rs1260326 | T | C | 0.6086 | 0.064 | 0.007 | 7.9433E-21 | 83.59 |
| *SHBG* | rs13108218 | A | G | 0.6191 | -0.051 | 0.007 | 2.5119E-13 | 53.08 |
| *SHBG* | rs11434868 | G | GA | 0.5788 | 0.048 | 0.007 | 1.9953E-12 | 47.02 |
| *SHBG* | rs7015 | A | G | 0.8152 | 0.068 | 0.009 | 3.1623E-15 | 57.09 |
| *SHBG* | rs7047907 | G | A | 0.7456 | -0.053 | 0.008 | 6.3096E-12 | 43.89 |
| *SHBG* | rs11045833 | G | A | 0.2102 | 0.058 | 0.008 | 1.2589E-12 | 52.56 |
| *SHBG* | rs28929474 | C | T | 0.0209 | 0.184 | 0.023 | 3.9811E-15 | 64.00 |
| *SHBG* | rs139974673 | T | C | 0.0243 | -0.147 | 0.022 | 1.2589E-11 | 44.65 |
| *SHBG* | rs56332871 | C | A | 0.2703 | 0.058 | 0.008 | 1E-14 | 52.56 |
| *SHBG* | rs858519 | T | C | 0.5582 | 0.274 | 0.007 | 1E-200 | 1532.16 |
| *SHBG* | rs530923403 | T | TTG | 0.0844 | 0.105 | 0.012 | 1.9953E-17 | 76.56 |
| *SHBG* | rs149751685 | G | A | 0.0248 | 0.18 | 0.024 | 5.0119E-14 | 56.25 |
| *SHD* | rs58534292 | T | C | 0.6507 | 0.068 | 0.008 | 5.0119E-17 | 72.25 |
| *SHD* | rs72481571 | G | A | 0.1061 | 0.214 | 0.014 | 1.5849E-52 | 233.65 |
| *SHH* | rs10225434 | T | C | 0.4187 | -0.057 | 0.008 | 5.0119E-13 | 50.77 |
| *SHISA5* | rs11706087 | T | C | 0.0247 | -0.392 | 0.023 | 1.5849E-63 | 290.48 |
| *SHISA5* | rs13146355 | G | A | 0.4562 | 0.048 | 0.007 | 6.3096E-12 | 47.02 |
| *SHISA5* | rs3184504 | T | C | 0.5174 | -0.049 | 0.007 | 5.0119E-12 | 49.00 |
| *SHISA5* | rs7203642 | A | G | 0.1923 | -0.067 | 0.009 | 1.5849E-13 | 55.42 |
| *SHMT1* | rs1354034 | T | C | 0.6039 | 0.057 | 0.007 | 5.0119E-18 | 66.31 |
| *SHMT1* | rs342296 | G | A | 0.4563 | -0.047 | 0.006 | 3.9811E-13 | 61.36 |
| *SHMT1* | rs6993770 | A | T | 0.2867 | -0.074 | 0.007 | 1.9953E-25 | 111.76 |
| *SHMT1* | rs669340 | G | C | 0.6339 | 0.719 | 0.008 | 1E-200 | 8077.52 |
| *SHPK* | rs224495 | G | A | 0.1007 | -0.132 | 0.013 | 6.3096E-25 | 103.10 |
| *SIAE* | rs114269697 | C | A | 0.0135 | -0.239 | 0.033 | 6.3096E-13 | 52.45 |
| *SIAE* | rs6993770 | A | T | 0.2862 | -0.067 | 0.008 | 2.5119E-16 | 70.14 |
| *SIAE* | rs56278466 | T | G | 0.6613 | 0.074 | 0.008 | 2.5119E-21 | 85.56 |
| *SIAE* | rs7896518 | A | G | 0.4241 | 0.094 | 0.008 | 3.9811E-35 | 138.06 |
| *SIAE* | rs78778622 | T | C | 0.0597 | -0.444 | 0.016 | 1E-174 | 770.06 |
| *SIAE* | rs2559850 | G | A | 0.5868 | 0.069 | 0.008 | 1.2589E-19 | 74.39 |
| *SIAE* | rs3184504 | T | C | 0.5174 | -0.052 | 0.007 | 2.5119E-12 | 55.18 |
| *SIAE* | rs72701845 | G | A | 0.0379 | -0.155 | 0.02 | 1E-14 | 60.06 |
| *SIAE* | rs892090 | T | G | 0.8344 | 0.095 | 0.01 | 1.9953E-21 | 90.25 |
| *SIGLEC1* | rs17849502 | G | T | 0.0525 | 0.111 | 0.015 | 7.9433E-13 | 54.76 |
| *SIGLEC1* | rs62165726 | C | A | 0.037 | -0.408 | 0.018 | 2.512E-108 | 513.78 |
| *SIGLEC1* | rs115216147 | T | A | 0.0666 | 0.107 | 0.014 | 5.0119E-14 | 58.41 |
| *SIGLEC1* | rs11185602 | A | G | 0.3247 | -0.064 | 0.007 | 3.9811E-18 | 83.59 |
| *SIGLEC1* | rs2031902 | C | T | 0.5875 | -0.072 | 0.007 | 3.1623E-24 | 105.80 |
| *SIGLEC1* | rs11220505 | C | T | 0.2223 | 0.099 | 0.008 | 5.0119E-32 | 153.14 |
| *SIGLEC1* | rs3184504 | T | C | 0.5173 | -0.068 | 0.007 | 3.9811E-23 | 94.37 |
| *SIGLEC1* | rs186021206 | G | A | 0.0064 | 1.325 | 0.048 | 2.512E-170 | 761.99 |
| *SIGLEC1* | rs11668950 | G | A | 0.2687 | 0.09 | 0.008 | 6.3096E-31 | 126.56 |
| *SIGLEC1* | rs189448562 | G | C | 0.0113 | -1.842 | 0.035 | 1E-200 | 2769.77 |
| *SIGLEC10* | rs1257169 | C | A | 0.4984 | -0.052 | 0.007 | 1.2589E-13 | 55.18 |
| *SIGLEC10* | rs4857410 | C | T | 0.1489 | -0.081 | 0.01 | 2.5119E-16 | 65.61 |
| *SIGLEC10* | rs634501 | A | G | 0.736 | -0.06 | 0.008 | 5.0119E-14 | 56.25 |
| *SIGLEC10* | rs562289 | C | T | 0.23 | 0.068 | 0.008 | 3.9811E-16 | 72.25 |
| *SIGLEC10* | rs3184504 | T | C | 0.5174 | -0.11 | 0.007 | 3.1623E-55 | 246.94 |
| *SIGLEC10* | rs186021206 | G | A | 0.0064 | 0.84 | 0.048 | 1.5849E-67 | 306.25 |
| *SIGLEC10* | rs34557412 | A | G | 0.0068 | 0.44 | 0.043 | 6.3096E-25 | 104.71 |
| *SIGLEC10* | rs148783636 | T | C | 0.0197 | -1.577 | 0.029 | 1E-200 | 2957.11 |
| *SIGLEC10* | rs738409 | C | G | 0.2175 | 0.061 | 0.008 | 7.9433E-13 | 58.14 |
| *SIGLEC5* | rs7137828 | C | T | 0.5164 | -0.058 | 0.006 | 1.5849E-23 | 93.44 |
| *SIGLEC5* | rs186021206 | G | A | 0.0064 | 0.445 | 0.04 | 2.5119E-29 | 123.77 |
| *SIGLEC5* | rs1106476 | T | A | 0.113 | -1.219 | 0.012 | 1E-200 | 10319.17 |

| | | | | | | | | |
|---|---|---|---|---|---|---|---|---|
| *SIGLEC6* | rs115427247 | C | T | 0.0299 | -0.156 | 0.02 | 3.1623E-15 | 60.84 |
| *SIGLEC6* | rs4601639 | T | C | 0.2661 | -0.065 | 0.008 | 1.2589E-17 | 66.02 |
| *SIGLEC6* | rs74227709 | G | A | 0.0736 | 0.141 | 0.013 | 2.5119E-28 | 117.64 |
| *SIGLEC6* | rs3111414 | C | G | 0.7984 | -0.119 | 0.008 | 1.5849E-45 | 221.27 |
| *SIGLEC6* | rs4384720 | C | G | 0.9198 | 0.116 | 0.012 | 2.5119E-21 | 93.44 |
| *SIGLEC6* | rs1516527 | T | C | 0.9505 | 0.223 | 0.015 | 6.3096E-47 | 221.02 |
| *SIGLEC6* | rs576666495 | A | G | 0.0033 | -0.437 | 0.059 | 1.2589E-13 | 54.86 |
| *SIGLEC6* | rs13107325 | C | T | 0.0748 | 0.116 | 0.013 | 1.2589E-19 | 79.62 |
| *SIGLEC6* | rs4863714 | A | G | 0.6413 | 0.055 | 0.007 | 2.5119E-15 | 61.73 |
| *SIGLEC6* | rs3131618 | A | G | 0.1425 | 0.124 | 0.01 | 3.9811E-38 | 153.76 |
| *SIGLEC6* | rs56278466 | T | G | 0.6612 | 0.049 | 0.007 | 2.5119E-12 | 49.00 |
| *SIGLEC6* | rs4937127 | A | G | 0.5221 | -0.103 | 0.007 | 7.9433E-53 | 216.51 |
| *SIGLEC6* | rs3184504 | T | C | 0.5166 | -0.071 | 0.007 | 1E-26 | 102.88 |
| *SIGLEC6* | rs146590204 | C | CGATTTTCA | 0.3464 | -0.07 | 0.007 | 1.5849E-22 | 100.00 |
| *SIGLEC6* | rs76428106 | T | C | 0.0129 | 0.491 | 0.031 | 3.1623E-57 | 250.86 |
| *SIGLEC6* | rs2745083 | C | A | 0.5132 | -0.114 | 0.007 | 2.5119E-63 | 265.22 |
| *SIGLEC6* | rs111164440 | A | G | 0.3683 | -0.054 | 0.007 | 2.5119E-14 | 59.51 |
| *SIGLEC6* | rs146261845 | C | T | 0.0069 | 0.434 | 0.043 | 1.2589E-23 | 101.87 |
| *SIGLEC6* | rs2659005 | C | T | 0.4416 | 0.046 | 0.007 | 7.9433E-12 | 43.18 |
| *SIGLEC6* | rs78744187 | C | T | 0.0817 | 0.15 | 0.013 | 5.0119E-29 | 133.14 |
| *SIGLEC6* | rs62617068 | C | A | 0.0123 | -1.758 | 0.034 | 1E-200 | 2673.50 |
| *SIGLEC7* | rs1257169 | C | A | 0.4992 | -0.059 | 0.007 | 1.2589E-16 | 71.04 |
| *SIGLEC7* | rs11721064 | G | T | 0.6051 | -0.056 | 0.007 | 1.5849E-14 | 64.00 |
| *SIGLEC7* | rs4393849 | A | G | 0.3083 | 0.056 | 0.008 | 3.1623E-13 | 49.00 |
| *SIGLEC7* | rs11220462 | G | A | 0.1323 | -0.084 | 0.01 | 1.5849E-15 | 70.56 |
| *SIGLEC7* | rs3184504 | T | C | 0.5174 | -0.068 | 0.007 | 1.2589E-21 | 94.37 |
| *SIGLEC7* | rs186021206 | G | A | 0.0064 | 0.962 | 0.049 | 1.9953E-86 | 385.44 |
| *SIGLEC7* | rs2160725 | A | C | 0.4854 | -0.083 | 0.007 | 1.2589E-30 | 140.59 |
| *SIGLEC7* | rs60202083 | T | A | 0.4223 | -0.066 | 0.007 | 1E-18 | 88.90 |
| *SIGLEC7* | rs140185670 | G | C | 0.0682 | -0.635 | 0.015 | 1E-200 | 1792.11 |
| *SIGLEC8* | rs3085908 | T | TAA | 0.3388 | 0.089 | 0.007 | 1.9953E-35 | 161.65 |
| *SIGLEC8* | rs77762937 | G | T | 0.2875 | 0.053 | 0.007 | 1E-12 | 57.33 |
| *SIGLEC8* | rs55921103 | G | T | 0.6526 | 0.1 | 0.007 | 5.0119E-45 | 204.08 |
| *SIGLEC8* | rs10032035 | T | C | 0.257 | 0.057 | 0.008 | 1.9953E-13 | 50.77 |
| *SIGLEC8* | rs2279587 | G | A | 0.059 | 0.132 | 0.014 | 2.5119E-20 | 88.90 |
| *SIGLEC8* | rs2256974 | C | A | 0.1738 | 0.065 | 0.009 | 1.9953E-13 | 52.16 |
| *SIGLEC8* | rs6458951 | T | C | 0.2625 | 0.068 | 0.008 | 7.9433E-19 | 72.25 |
| *SIGLEC8* | rs4876611 | A | G | 0.718 | 0.17 | 0.008 | 6.31E-112 | 451.56 |
| *SIGLEC8* | rs10796294 | A | G | 0.5692 | 0.078 | 0.007 | 5.0119E-30 | 124.16 |
| *SIGLEC8* | rs2159408 | A | C | 0.4894 | -0.076 | 0.007 | 1.5849E-29 | 117.88 |
| *SIGLEC8* | rs4766578 | T | A | 0.5039 | -0.062 | 0.007 | 3.1623E-20 | 78.45 |
| *SIGLEC8* | rs7485656 | A | G | 0.1398 | 0.066 | 0.01 | 1.5849E-11 | 43.56 |
| *SIGLEC8* | rs200489612 | G | A | 0.0053 | 0.455 | 0.05 | 1.2589E-19 | 82.81 |
| *SIGLEC8* | rs151278524 | C | T | 0.0068 | 0.28 | 0.041 | 1.2589E-11 | 46.64 |
| *SIGLEC8* | rs36518 | G | A | 0.1371 | 0.684 | 0.011 | 1E-200 | 3866.58 |
| *SIGLEC9* | rs61830291 | A | C | 0.0974 | -0.07 | 0.01 | 1.5849E-12 | 49.00 |
| *SIGLEC9* | rs2593704 | G | C | 0.7819 | -0.061 | 0.007 | 3.9811E-18 | 75.94 |
| *SIGLEC9* | rs10935473 | G | T | 0.44 | -0.631 | 0.007 | 1E-200 | 8125.73 |
| *SIGLEC9* | rs1468906 | A | G | 0.3441 | -0.049 | 0.007 | 3.9811E-12 | 49.00 |
| *SIGLEC9* | rs9260 | A | G | 0.8074 | -0.111 | 0.007 | 1E-49 | 251.45 |
| *SIGLEC9* | rs4841132 | A | G | 0.9079 | -0.096 | 0.01 | 3.9811E-21 | 92.16 |
| *SIGLEC9* | rs3967200 | C | T | 0.1346 | -0.196 | 0.009 | 7.943E-115 | 474.27 |
| *SIGLEC9* | rs186021206 | G | A | 0.0064 | 1.557 | 0.04 | 1E-200 | 1515.16 |
| *SIGLEC9* | rs2075803 | A | G | 0.5531 | -0.573 | 0.007 | 1E-200 | 6700.59 |
| *SIL1* | rs35457250 | C | T | 0.0089 | 0.444 | 0.041 | 2.5119E-27 | 117.27 |
| *SIL1* | rs2351905 | C | T | 0.4404 | 0.059 | 0.008 | 3.9811E-14 | 54.39 |
| *SIL1* | rs10843403 | T | C | 0.6095 | 0.084 | 0.008 | 7.9433E-26 | 110.25 |
| *SIL1* | rs28929474 | C | T | 0.0208 | 0.5 | 0.027 | 1.5849E-76 | 342.94 |
| *SIL1* | rs1801689 | A | C | 0.03 | 0.165 | 0.022 | 2.5119E-13 | 56.25 |
| *SIRPA* | rs61747728 | C | T | 0.0382 | 0.071 | 0.01 | 3.9811E-13 | 50.41 |
| *SIRPA* | rs1260326 | T | C | 0.6086 | 0.031 | 0.004 | 3.1623E-16 | 60.06 |
| *SIRPA* | rs34458031 | G | A | 0.0858 | 0.072 | 0.007 | 7.9433E-27 | 105.80 |
| *SIRPA* | rs9269915 | G | T | 0.7243 | -0.031 | 0.004 | 1E-12 | 60.06 |
| *SIRPA* | rs507666 | G | A | 0.1849 | -0.076 | 0.005 | 6.3096E-54 | 231.04 |
| *SIRPA* | rs186021206 | G | A | 0.0064 | 0.431 | 0.026 | 1.2589E-61 | 274.79 |
| *SIRPA* | rs2659005 | C | T | 0.4408 | 0.029 | 0.004 | 6.3096E-14 | 52.56 |
| *SIRPA* | rs6136377 | A | G | 0.3821 | -1.234 | 0.008 | 1E-200 | 23793.06 |
| *SIRPB1* | rs61747728 | C | T | 0.038 | 0.137 | 0.016 | 2.5119E-18 | 73.32 |

| | | | | | | | | |
|---|---|---|---|---|---|---|---|---|
| *SIRPB1* | rs1260326 | T | C | 0.6086 | 0.044 | 0.006 | 1.2589E-12 | 53.78 |
| *SIRPB1* | rs10053046 | A | G | 0.3254 | -0.05 | 0.006 | 1.2589E-14 | 69.44 |
| *SIRPB1* | rs13277237 | G | A | 0.5682 | -0.051 | 0.006 | 6.3096E-17 | 72.25 |
| *SIRPB1* | rs3184504 | T | C | 0.5172 | -0.079 | 0.006 | 1E-39 | 173.36 |
| *SIRPB1* | rs76428106 | T | C | 0.0129 | 0.279 | 0.028 | 6.3096E-24 | 99.29 |
| *SIRPB1* | rs777834943 | C | CT | 0.1835 | -0.055 | 0.008 | 5.0119E-12 | 47.27 |
| *SIRPB1* | rs186021206 | G | A | 0.0064 | 0.36 | 0.041 | 3.9811E-18 | 77.10 |
| *SIRPB1* | rs1045929 | C | T | 0.3885 | 0.052 | 0.006 | 2.5119E-17 | 75.11 |
| *SIRPB1* | rs2777895 | A | G | 0.5489 | 0.079 | 0.006 | 1.9953E-39 | 173.36 |
| *SIRPB1* | rs7224668 | T | C | 0.5579 | -0.05 | 0.006 | 2.5119E-16 | 69.44 |
| *SIRPB1* | rs79462675 | C | A | 0.1317 | -0.95 | 0.011 | 1E-200 | 7458.68 |
| *SIRT1* | rs28672722 | G | T | 0.2224 | 0.093 | 0.009 | 1.2589E-23 | 106.78 |
| *SIRT2* | rs1354034 | T | C | 0.6046 | 0.063 | 0.008 | 2.5119E-16 | 62.02 |
| *SIRT2* | rs342298 | C | T | 0.4545 | -0.065 | 0.008 | 3.9811E-18 | 66.02 |
| *SIRT2* | rs7896518 | A | G | 0.4236 | 0.063 | 0.008 | 3.1623E-16 | 62.02 |
| *SIRT2* | rs11502185 | T | C | 0.2596 | 0.066 | 0.01 | 5.0119E-12 | 43.56 |
| *SIRT2* | rs144373891 | C | T | 0.0092 | -0.716 | 0.039 | 6.3096E-74 | 337.05 |
| *SIT1* | rs3184504 | T | C | 0.5177 | -0.094 | 0.007 | 6.3096E-36 | 180.33 |
| *SIT1* | rs10468017 | C | T | 0.2967 | -0.066 | 0.008 | 1.5849E-15 | 68.06 |
| *SIT1* | rs71374020 | A | AT | 0.2992 | -0.062 | 0.009 | 1.2589E-12 | 47.46 |
| *SIT1* | rs12446515 | C | T | 0.3241 | -0.084 | 0.008 | 1.5849E-25 | 110.25 |
| *SIT1* | rs4613118 | A | G | 0.8594 | -0.081 | 0.011 | 6.3096E-14 | 54.22 |
| *SIT1* | rs7208422 | A | T | 0.477 | -0.084 | 0.008 | 1E-28 | 110.25 |
| *SIT1* | rs10469472 | T | G | 0.1192 | 0.099 | 0.012 | 2.5119E-17 | 68.06 |
| *SKAP1* | rs28414666 | G | A | 0.2095 | 0.068 | 0.009 | 1.2589E-13 | 57.09 |
| *SKAP1* | rs76438067 | C | T | 0.2332 | -0.092 | 0.009 | 2.5119E-25 | 104.49 |
| *SKAP1* | rs3176789 | C | T | 0.2787 | -0.058 | 0.008 | 3.9811E-12 | 52.56 |
| *SKAP1* | rs7310615 | C | G | 0.5175 | -0.074 | 0.008 | 1E-22 | 85.56 |
| *SKAP1* | rs71374020 | A | AT | 0.2993 | -0.062 | 0.009 | 1E-12 | 47.46 |
| *SKAP1* | rs5817082 | C | CA | 0.2664 | 0.06 | 0.009 | 1.5849E-12 | 44.44 |
| *SKAP1* | rs368876017 | AAAGCAAG | A | 0.3399 | 0.061 | 0.008 | 2.5119E-13 | 58.14 |
| *SKAP1* | rs7208422 | A | T | 0.4778 | -0.067 | 0.007 | 1.9953E-19 | 91.61 |
| *SKAP1* | rs74460666 | T | G | 0.1213 | 0.093 | 0.011 | 3.9811E-16 | 71.48 |
| *SKAP2* | rs1354034 | T | C | 0.6041 | 0.069 | 0.008 | 1.9953E-19 | 74.39 |
| *SKAP2* | rs17622656 | G | A | 0.3886 | -0.055 | 0.008 | 6.3096E-13 | 47.27 |
| *SKAP2* | rs6993770 | A | T | 0.2867 | -0.068 | 0.008 | 1.2589E-16 | 72.25 |
| *SLA2* | rs113102870 | G | A | 0.0067 | -0.764 | 0.047 | 2.5119E-60 | 264.24 |
| *SLAMF1* | rs37456 | C | T | 0.3524 | 0.061 | 0.008 | 1.9953E-14 | 58.14 |
| *SLAMF1* | rs7535367 | G | T | 0.1392 | -0.145 | 0.011 | 1.9953E-39 | 173.76 |
| *SLAMF1* | rs3821819 | G | A | 0.387 | -0.058 | 0.008 | 1E-13 | 52.56 |
| *SLAMF1* | rs9264669 | A | T | 0.5725 | 0.068 | 0.008 | 3.9811E-18 | 72.25 |
| *SLAMF1* | rs35350651 | A | AC | 0.5046 | -0.053 | 0.008 | 3.9811E-12 | 43.89 |
| *SLAMF1* | rs186021206 | G | A | 0.0064 | 0.777 | 0.052 | 1E-49 | 223.27 |
| *SLAMF6* | rs11265413 | T | A | 0.203 | -0.215 | 0.009 | 5.012E-114 | 570.68 |
| *SLAMF6* | rs3184504 | T | C | 0.517 | -0.065 | 0.008 | 5.0119E-18 | 66.02 |
| *SLAMF6* | rs186021206 | G | A | 0.0063 | 0.441 | 0.053 | 6.3096E-17 | 69.23 |
| *SLAMF7* | rs188468174 | C | T | 0.0143 | -0.545 | 0.032 | 6.3096E-65 | 290.06 |
| *SLAMF7* | rs11581248 | C | T | 0.1361 | -0.943 | 0.011 | 1E-200 | 7349.17 |
| *SLAMF7* | rs4972973 | G | A | 0.4686 | -0.056 | 0.007 | 1E-16 | 64.00 |
| *SLAMF7* | rs7621161 | C | A | 0.2803 | -0.135 | 0.008 | 5.0119E-71 | 284.77 |
| *SLAMF7* | rs3094087 | T | C | 0.1706 | -0.096 | 0.009 | 6.3096E-27 | 113.78 |
| *SLAMF7* | rs3780480 | G | T | 0.2543 | -0.054 | 0.008 | 1.9953E-12 | 45.56 |
| *SLAMF7* | rs111338191 | A | AAATT | 0.5212 | -0.056 | 0.007 | 2.5119E-16 | 64.00 |
| *SLAMF7* | rs186021206 | G | A | 0.0064 | 0.943 | 0.046 | 6.3096E-92 | 420.25 |
| *SLAMF7* | rs34562254 | G | A | 0.0971 | 0.096 | 0.011 | 1.5849E-17 | 76.17 |
| *SLAMF7* | rs8077394 | A | G | 0.5542 | -0.054 | 0.007 | 1.5849E-15 | 59.51 |
| *SLAMF8* | rs34687326 | G | A | 0.1222 | -1.343 | 0.012 | 1E-200 | 12525.34 |
| *SLAMF8* | rs2190504 | T | C | 0.4434 | 0.052 | 0.007 | 1E-13 | 55.18 |
| *SLAMF8* | rs12667978 | C | T | 0.5721 | 0.049 | 0.007 | 5.0119E-12 | 49.00 |
| *SLAMF8* | rs3184504 | T | C | 0.517 | -0.097 | 0.007 | 5.0119E-44 | 192.02 |
| *SLAMF8* | rs186021206 | G | A | 0.0063 | 0.469 | 0.049 | 5.0119E-22 | 91.61 |
| *SLC16A1* | rs6670866 | A | G | 0.5614 | -0.062 | 0.008 | 1.5849E-15 | 60.06 |
| *SLC16A1* | rs3811444 | C | T | 0.333 | -0.104 | 0.008 | 1.2589E-37 | 169.00 |
| *SLC16A1* | rs4835097 | A | G | 0.7085 | 0.069 | 0.008 | 3.9811E-16 | 74.39 |
| *SLC16A1* | rs633382 | T | C | 0.9644 | 0.256 | 0.021 | 6.3096E-35 | 148.61 |
| *SLC27A4* | rs1354034 | T | C | 0.6035 | 0.093 | 0.008 | 3.1623E-33 | 135.14 |
| *SLC27A4* | rs137948800 | T | TA | 0.1232 | 0.081 | 0.012 | 2.5119E-12 | 45.56 |
| *SLC27A4* | rs7080536 | G | A | 0.0435 | -0.19 | 0.019 | 7.9433E-24 | 100.00 |

| | | | | | | | | |
|---|---|---|---|---|---|---|---|---|
| *SLC39A14* | rs79624003 | A | G | 0.127 | -0.081 | 0.011 | 1.2589E-12 | 54.22 |
| *SLC39A14* | rs35486529 | C | T | 0.2549 | -0.107 | 0.009 | 3.9811E-34 | 141.35 |
| *SLC39A14* | rs28929474 | C | T | 0.0207 | 0.309 | 0.027 | 1.2589E-30 | 130.98 |
| *SLC39A5* | rs12065546 | C | T | 0.8516 | 0.078 | 0.01 | 3.1623E-14 | 60.84 |
| *SLC39A5* | rs9263789 | G | A | 0.1647 | -0.092 | 0.01 | 5.0119E-21 | 84.64 |
| *SLC39A5* | rs7786376 | A | G | 0.2774 | -0.067 | 0.008 | 7.9433E-17 | 70.14 |
| *SLC39A5* | rs2272662 | T | C | 0.5775 | -0.058 | 0.007 | 3.1623E-15 | 68.65 |
| *SLC39A5* | rs147572362 | T | C | 0.0075 | -0.696 | 0.042 | 1.9953E-61 | 274.61 |
| *SLC39A5* | rs111349624 | CAA | C | 0.0841 | 0.093 | 0.013 | 1.2589E-12 | 51.18 |
| *SLC39A5* | rs28929474 | C | T | 0.0207 | 0.338 | 0.025 | 3.1623E-40 | 182.79 |
| *SLC39A5* | rs11078597 | T | C | 0.1876 | 0.064 | 0.009 | 6.3096E-12 | 50.57 |
| *SLC39A5* | rs12609794 | G | A | 0.7484 | 0.056 | 0.008 | 1.2589E-11 | 49.00 |
| *SLC39A5* | rs55953905 | C | T | 0.2471 | 0.095 | 0.008 | 1.5849E-29 | 141.02 |
| *SLC4A1* | rs12734030 | C | T | 0.1482 | -0.071 | 0.01 | 1E-11 | 50.41 |
| *SLC4A1* | rs145919155 | G | A | 0.0499 | -0.138 | 0.017 | 6.3096E-16 | 65.90 |
| *SLC51B* | rs4414734 | T | A | 0.3353 | -0.108 | 0.008 | 1.9953E-42 | 182.25 |
| *SLC51B* | rs939885 | G | A | 0.4855 | -0.168 | 0.007 | 1.585E-110 | 576.00 |
| *SLC51B* | rs11239528 | A | G | 0.2434 | -0.096 | 0.009 | 2.5119E-28 | 113.78 |
| *SLC51B* | rs56398830 | G | A | 0.0112 | -0.687 | 0.035 | 1.9953E-83 | 385.28 |
| *SLC51B* | rs3813582 | T | C | 0.3145 | 0.055 | 0.008 | 6.3096E-12 | 47.27 |
| *SLC51B* | rs12938714 | G | A | 0.33 | 0.144 | 0.008 | 6.3096E-74 | 324.00 |
| *SLC51B* | rs11078916 | T | C | 0.7214 | -0.072 | 0.008 | 7.9433E-18 | 81.00 |
| *SLC51B* | rs681343 | C | T | 0.509 | -0.22 | 0.008 | 5.012E-185 | 756.25 |
| *SLC9A3R1* | rs342299 | C | T | 0.4549 | -0.058 | 0.008 | 1.9953E-14 | 52.56 |
| *SLC9A3R1* | rs6993770 | A | T | 0.287 | -0.057 | 0.008 | 6.3096E-12 | 50.77 |
| *SLC9A3R1* | rs60822569 | T | TTTCTC | 0.5542 | 0.056 | 0.008 | 3.9811E-13 | 49.00 |
| *SLC9A3R1* | rs745666 | G | C | 0.4388 | 0.092 | 0.008 | 6.3096E-31 | 132.25 |
| *SLC9A3R2* | rs782455289 | TAAGAC | T | 0.3169 | 0.07 | 0.008 | 1.5849E-18 | 76.56 |
| *SLC9A3R2* | rs7967845 | A | G | 0.5749 | 0.059 | 0.008 | 1.2589E-14 | 54.39 |
| *SLC9A3R2* | rs3211995 | G | A | 0.1594 | -0.147 | 0.01 | 1.9953E-46 | 216.09 |
| *SLC9A3R2* | rs3764261 | C | A | 0.325 | -0.059 | 0.008 | 1.5849E-13 | 54.39 |
| *SLC9A3R2* | rs35434379 | C | CA | 0.6008 | -0.089 | 0.008 | 3.9811E-31 | 123.77 |
| *SLIRP* | rs77333309 | G | A | 0.1059 | 0.101 | 0.014 | 1.2589E-12 | 52.05 |
| *SLIT2* | rs1185160 | G | T | 0.7005 | -0.061 | 0.008 | 3.1623E-13 | 58.14 |
| *SLITRK1* | rs13107325 | C | T | 0.0747 | 0.108 | 0.014 | 1E-14 | 59.51 |
| *SLITRK1* | rs3782473 | T | C | 0.237 | 0.083 | 0.009 | 1.2589E-21 | 85.05 |
| *SLITRK1* | rs2876803 | T | A | 0.1704 | 0.157 | 0.01 | 3.9811E-58 | 246.49 |
| *SLITRK1* | rs186021206 | G | A | 0.0063 | 0.503 | 0.051 | 2.5119E-23 | 97.27 |
| *SLITRK1* | rs33950747 | C | T | 0.075 | 0.101 | 0.014 | 3.1623E-13 | 52.05 |
| *SLITRK2* | rs61830291 | A | C | 0.0974 | -0.088 | 0.012 | 1E-12 | 53.78 |
| *SLITRK2* | rs62262391 | C | T | 0.2249 | 0.074 | 0.009 | 1.5849E-17 | 67.60 |
| *SLITRK2* | rs68119427 | C | G | 0.3489 | 0.052 | 0.008 | 1E-11 | 42.25 |
| *SLITRK2* | rs13107325 | C | T | 0.0748 | 0.263 | 0.014 | 2.5119E-79 | 352.90 |
| *SLITRK2* | rs78334147 | C | T | 0.1008 | 0.105 | 0.012 | 5.0119E-18 | 76.56 |
| *SLITRK2* | rs2811708 | G | T | 0.2646 | 0.068 | 0.008 | 1.5849E-16 | 72.25 |
| *SLITRK2* | rs1131773 | A | G | 0.1442 | 0.071 | 0.01 | 7.9433E-12 | 50.41 |
| *SLITRK2* | rs56278466 | T | G | 0.6612 | 0.073 | 0.008 | 2.5119E-21 | 83.27 |
| *SLITRK2* | rs10823396 | C | G | 0.465 | 0.051 | 0.007 | 1.9953E-12 | 53.08 |
| *SLITRK2* | rs12805381 | G | T | 0.4514 | 0.083 | 0.007 | 3.1623E-29 | 140.59 |
| *SLITRK2* | rs11105298 | T | C | 0.2424 | -0.091 | 0.009 | 3.1623E-26 | 102.23 |
| *SLITRK2* | rs186021206 | G | A | 0.0064 | 0.438 | 0.05 | 2.5119E-18 | 76.74 |
| *SLITRK6* | rs61747728 | C | T | 0.0381 | 0.143 | 0.019 | 6.3096E-14 | 56.65 |
| *SLITRK6* | rs1260326 | T | C | 0.6088 | 0.064 | 0.007 | 1.9953E-17 | 83.59 |
| *SLITRK6* | rs13107325 | C | T | 0.0747 | 0.104 | 0.014 | 1E-13 | 55.18 |
| *SLITRK6* | rs7712631 | G | A | 0.4204 | 0.087 | 0.007 | 6.3096E-32 | 154.47 |
| *SLITRK6* | rs587600964 | C | G | 0.1198 | -0.085 | 0.011 | 7.9433E-14 | 59.71 |
| *SLITRK6* | rs36115375 | C | A | 0.0627 | -0.151 | 0.015 | 2.5119E-23 | 101.34 |
| *SLITRK6* | rs56278466 | T | G | 0.661 | 0.066 | 0.008 | 1.5849E-17 | 68.06 |
| *SLITRK6* | rs12863734 | G | A | 0.0172 | -1.411 | 0.029 | 1E-200 | 2367.33 |
| *SLMAP* | rs1354034 | T | C | 0.6046 | 0.071 | 0.008 | 1E-20 | 78.77 |
| *SLURP1* | rs61816761 | G | A | 0.0235 | 0.239 | 0.025 | 1E-21 | 91.39 |
| *SLURP1* | rs10624245 | C | CTG | 0.5334 | 0.409 | 0.007 | 1E-200 | 3413.90 |
| *SLURP1* | rs2847501 | C | T | 0.0803 | 0.092 | 0.013 | 1.5849E-12 | 50.08 |
| *SLURP1* | rs10426 | G | A | 0.2149 | 0.064 | 0.009 | 2.5119E-13 | 50.57 |
| *SMAD1* | rs1354034 | T | C | 0.6038 | 0.053 | 0.008 | 5.0119E-12 | 43.89 |
| *SMAD1* | rs450269 | G | C | 0.45 | -0.062 | 0.008 | 3.9811E-16 | 60.06 |
| *SMAD1* | rs6993770 | A | T | 0.2869 | -0.074 | 0.008 | 1E-18 | 85.56 |
| *SMAD1* | rs964184 | G | C | 0.8667 | -0.08 | 0.011 | 7.9433E-13 | 52.89 |

| | | | | | | | | |
|---|---|---|---|---|---|---|---|---|
| *SMAD3* | rs3811444 | C | T | 0.3325 | -0.07 | 0.008 | 3.9811E-19 | 76.56 |
| *SMAD3* | rs342293 | C | G | 0.4598 | -0.102 | 0.007 | 1.9953E-43 | 212.33 |
| *SMAD3* | rs60822569 | T | TTTCTC | 0.5539 | 0.092 | 0.007 | 7.9433E-35 | 172.73 |
| *SMAD3* | rs34189958 | C | A | 0.2329 | 0.245 | 0.009 | 6.31E-169 | 741.05 |
| *SMAD5* | rs4643790 | G | A | 0.197 | -0.214 | 0.009 | 6.31E-117 | 565.38 |
| *SMAD5* | rs3129777 | C | A | 0.2152 | -0.081 | 0.01 | 5.0119E-17 | 65.61 |
| *SMNDC1* | rs4632248 | G | T | 0.2097 | 0.158 | 0.009 | 5.0119E-70 | 308.20 |
| *SMOC1* | rs2090034 | G | A | 0.4688 | -0.057 | 0.007 | 6.3096E-17 | 66.31 |
| *SMOC1* | rs35457250 | C | T | 0.0089 | 0.298 | 0.036 | 2.5119E-16 | 68.52 |
| *SMOC1* | rs11751024 | C | A | 0.3984 | 0.056 | 0.007 | 1.2589E-15 | 64.00 |
| *SMOC1* | rs2960834 | G | C | 0.4458 | 0.053 | 0.007 | 1.9953E-14 | 57.33 |
| *SMOC1* | rs705379 | G | A | 0.4774 | 0.077 | 0.007 | 1E-28 | 121.00 |
| *SMOC1* | rs1958078 | A | C | 0.8469 | 0.367 | 0.01 | 1E-200 | 1346.89 |
| *SMOC1* | rs28929474 | C | T | 0.0208 | 0.188 | 0.025 | 2.5119E-14 | 56.55 |
| *SMOC1* | rs77542162 | A | G | 0.0229 | 0.463 | 0.023 | 5.0119E-91 | 405.23 |
| *SMOC1* | rs584007 | A | G | 0.6417 | 0.058 | 0.007 | 2.5119E-16 | 68.65 |
| *SMOC1* | rs2267867 | A | G | 0.2426 | -0.061 | 0.008 | 2.5119E-14 | 58.14 |
| *SMOC1* | rs4239651 | T | C | 0.7921 | -0.071 | 0.008 | 3.9811E-17 | 78.77 |
| *SMOC2* | rs56296467 | T | C | 0.2775 | -0.395 | 0.008 | 1E-200 | 2437.89 |
| *SMOC2* | rs6984210 | C | G | 0.0541 | -0.111 | 0.015 | 5.0119E-14 | 54.76 |
| *SMOC2* | rs4759276 | G | A | 0.3911 | 0.052 | 0.007 | 1.2589E-14 | 55.18 |
| *SMOC2* | rs77924615 | G | A | 0.1967 | -0.057 | 0.008 | 7.9433E-12 | 50.77 |
| *SMOC2* | rs7247412 | T | C | 0.2669 | -0.119 | 0.008 | 3.9811E-57 | 221.27 |
| *SMPD1* | rs78444298 | G | A | 0.0195 | -0.205 | 0.025 | 2.5119E-16 | 67.24 |
| *SMPD1* | rs11735092 | T | C | 0.4378 | -0.049 | 0.007 | 1.2589E-12 | 49.00 |
| *SMPD1* | rs114816312 | C | T | 0.0077 | 0.345 | 0.039 | 1.5849E-18 | 78.25 |
| *SMPD1* | rs2389688 | A | C | 0.4418 | -0.05 | 0.007 | 6.3096E-13 | 51.02 |
| *SMPD1* | rs77945361 | G | A | 0.1481 | -0.08 | 0.01 | 1.9953E-16 | 64.00 |
| *SMPD1* | rs199922514 | A | AAGAAGAA | 0.9076 | 0.08 | 0.012 | 1.5849E-11 | 44.44 |
| *SMPD1* | rs11791806 | C | T | 0.1732 | 0.065 | 0.009 | 1E-12 | 52.16 |
| *SMPD1* | rs3841602 | A | AGGCGGC | 0.4685 | 0.084 | 0.007 | 2.5119E-34 | 144.00 |
| *SMPD1* | rs2862954 | T | C | 0.5015 | -0.056 | 0.007 | 1.5849E-16 | 64.00 |
| *SMPD1* | rs1050239 | G | A | 0.2431 | -0.438 | 0.008 | 1E-200 | 2997.56 |
| *SMPD1* | rs10778152 | A | G | 0.2877 | -0.347 | 0.008 | 1E-200 | 1881.39 |
| *SMPD1* | rs150641790 | T | TG | 0.0747 | 0.12 | 0.013 | 7.9433E-20 | 85.21 |
| *SMPD1* | rs71435622 | A | AT | 0.2871 | 0.067 | 0.008 | 3.9811E-17 | 70.14 |
| *SMPD1* | rs28929474 | C | T | 0.0208 | 0.35 | 0.024 | 3.9811E-48 | 212.67 |
| *SMPD1* | rs139974673 | T | C | 0.0244 | 0.168 | 0.022 | 3.9811E-14 | 58.31 |
| *SMPD1* | rs58542926 | C | T | 0.0753 | 0.157 | 0.013 | 1.9953E-33 | 145.85 |
| *SMPD1* | rs429358 | T | C | 0.1554 | -0.098 | 0.01 | 6.3096E-25 | 96.04 |
| *SMPD1* | rs11668882 | T | C | 0.4408 | 0.059 | 0.007 | 2.5119E-17 | 71.04 |
| *SMPD1* | rs738409 | C | G | 0.217 | 0.07 | 0.008 | 1.9953E-17 | 76.56 |
| *SMPD3* | rs139299944 | C | CT | 0.3437 | 0.079 | 0.008 | 1.2589E-22 | 97.52 |
| *SMPDL3A* | rs114165349 | G | C | 0.0233 | 0.177 | 0.02 | 1.2589E-18 | 78.32 |
| *SMPDL3A* | rs687339 | C | T | 0.7712 | 0.09 | 0.007 | 1.5849E-35 | 165.31 |
| *SMPDL3A* | rs114816312 | C | T | 0.0077 | 0.266 | 0.035 | 1.2589E-14 | 57.76 |
| *SMPDL3A* | rs17259257 | C | T | 0.0555 | -0.1 | 0.013 | 3.9811E-14 | 59.17 |
| *SMPDL3A* | rs41280463 | G | A | 0.1681 | -0.076 | 0.008 | 3.9811E-21 | 90.25 |
| *SMPDL3A* | rs2522062 | A | G | 0.1836 | 0.054 | 0.008 | 3.1623E-12 | 45.56 |
| *SMPDL3A* | rs567092558 | C | CT | 0.14 | -0.066 | 0.009 | 6.3096E-14 | 53.78 |
| *SMPDL3A* | rs28385609 | C | T | 0.1379 | -1.017 | 0.011 | 1E-200 | 8547.84 |
| *SMPDL3A* | rs9414801 | G | A | 0.5329 | -0.057 | 0.006 | 1E-20 | 90.25 |
| *SMPDL3A* | rs2862954 | T | C | 0.5015 | -0.063 | 0.006 | 7.9433E-26 | 110.25 |
| *SMPDL3A* | rs10787429 | T | C | 0.7273 | -0.058 | 0.007 | 1.2589E-17 | 68.65 |
| *SMPDL3A* | rs7964859 | C | G | 0.288 | -0.416 | 0.007 | 1E-200 | 3531.76 |
| *SMPDL3A* | rs150641790 | T | TG | 0.0748 | 0.124 | 0.012 | 6.3096E-27 | 106.78 |
| *SMPDL3A* | rs6571912 | G | C | 0.4605 | 0.063 | 0.006 | 3.1623E-25 | 110.25 |
| *SMPDL3A* | rs145078947 | G | T | 0.0025 | 0.805 | 0.06 | 2.5119E-41 | 180.01 |
| *SMPDL3A* | rs147233090 | C | T | 0.0235 | 0.171 | 0.02 | 1E-17 | 73.10 |
| *SMPDL3A* | rs339969 | C | A | 0.6169 | 0.05 | 0.006 | 1.2589E-15 | 69.44 |
| *SMPDL3A* | rs58542926 | C | T | 0.0756 | 0.189 | 0.011 | 3.1623E-61 | 295.21 |
| *SMPDL3A* | rs429358 | T | C | 0.1554 | -0.103 | 0.008 | 6.3096E-35 | 165.77 |
| *SMPDL3A* | rs626283 | G | C | 0.4418 | 0.057 | 0.006 | 1E-20 | 90.25 |
| *SMPDL3A* | rs738408 | C | T | 0.217 | 0.07 | 0.007 | 7.9433E-22 | 100.00 |
| *SMTN* | rs1354034 | T | C | 0.6044 | 0.072 | 0.007 | 7.9433E-22 | 105.80 |
| *SMTN* | rs114694170 | T | C | 0.0598 | 0.151 | 0.016 | 5.0119E-22 | 89.07 |
| *SMTN* | rs342293 | C | G | 0.4599 | -0.05 | 0.007 | 7.9433E-12 | 51.02 |
| *SMTN* | rs12292693 | A | C | 0.2598 | -0.075 | 0.008 | 7.9433E-19 | 87.89 |

| | | | | | | | | |
|---|---|---|---|---|---|---|---|---|
| *SNAP23* | rs1354034 | T | C | 0.6037 | 0.111 | 0.008 | 1E-47 | 192.52 |
| *SNAP23* | rs17622656 | G | A | 0.3884 | -0.052 | 0.008 | 1.5849E-11 | 42.25 |
| *SNAP23* | rs6993770 | A | T | 0.2866 | -0.059 | 0.008 | 1.2589E-12 | 54.39 |
| *SNAP23* | rs10820606 | A | C | 0.2297 | -0.084 | 0.009 | 1.9953E-20 | 87.11 |
| *SNAP23* | rs11502185 | T | C | 0.2595 | 0.084 | 0.01 | 1.9953E-18 | 70.56 |
| *SNAP25* | rs10908505 | T | A | 0.6517 | 0.056 | 0.008 | 1.2589E-13 | 49.00 |
| *SNAP25* | rs342293 | C | G | 0.4594 | -0.19 | 0.007 | 1E-151 | 736.73 |
| *SNAP25* | rs3780420 | T | C | 0.3642 | 0.059 | 0.008 | 6.3096E-15 | 54.39 |
| *SNAP25* | rs60757417 | C | G | 0.0607 | 0.111 | 0.015 | 3.1623E-13 | 54.76 |
| *SNAP25* | rs7896518 | A | G | 0.4236 | 0.059 | 0.007 | 1.9953E-15 | 71.04 |
| *SNAP25* | rs1477123 | T | A | 0.7261 | -0.069 | 0.008 | 1.9953E-17 | 74.39 |
| *SNAP25* | rs429358 | T | C | 0.1557 | 0.625 | 0.01 | 1E-200 | 3906.25 |
| *SNAP25* | rs362562 | G | A | 0.4136 | -0.103 | 0.008 | 1.5849E-42 | 165.77 |
| *SNAP29* | rs1354034 | T | C | 0.6041 | 0.078 | 0.008 | 3.1623E-24 | 95.06 |
| *SNAP29* | rs11242109 | G | T | 0.4811 | -0.058 | 0.008 | 1.5849E-14 | 52.56 |
| *SNAP29* | rs6993770 | A | T | 0.2861 | -0.078 | 0.008 | 7.9433E-21 | 95.06 |
| *SNAP29* | rs892090 | T | G | 0.8343 | 0.078 | 0.01 | 2.5119E-14 | 60.84 |
| *SNAP29* | rs5759874 | A | G | 0.086 | -0.13 | 0.014 | 1.9953E-21 | 86.22 |
| *SNCA* | rs3811444 | C | T | 0.3325 | -0.07 | 0.008 | 3.9811E-18 | 76.56 |
| *SNCA* | rs1354034 | T | C | 0.6044 | 0.078 | 0.008 | 1.9953E-24 | 95.06 |
| *SNCA* | rs55730499 | C | T | 0.082 | 0.103 | 0.014 | 6.3096E-14 | 54.13 |
| *SNCA* | rs342296 | G | A | 0.4567 | -0.064 | 0.008 | 1.5849E-17 | 64.00 |
| *SNCA* | rs2393967 | A | C | 0.314 | 0.071 | 0.008 | 5.0119E-18 | 78.77 |
| *SNCG* | rs873110 | C | T | 0.2451 | -1.061 | 0.009 | 1E-200 | 13897.79 |
| *SNED1* | rs2074822 | G | A | 0.4929 | -0.125 | 0.008 | 7.9433E-60 | 244.14 |
| *SNX15* | rs1354034 | T | C | 0.6044 | 0.056 | 0.007 | 1E-14 | 64.00 |
| *SNX15* | rs6993770 | A | T | 0.287 | -0.06 | 0.008 | 1.5849E-14 | 56.25 |
| *SNX15* | rs2393967 | A | C | 0.314 | 0.052 | 0.008 | 1.2589E-11 | 42.25 |
| *SNX15* | rs2670910 | G | A | 0.0864 | -0.781 | 0.013 | 1E-200 | 3609.24 |
| *SNX18* | rs2548611 | T | C | 0.8123 | -0.096 | 0.01 | 1.5849E-23 | 92.16 |
| *SNX2* | rs27740 | A | T | 0.5528 | 0.096 | 0.008 | 2.5119E-36 | 144.00 |
| *SNX2* | rs28891437 | G | A | 0.212 | -0.08 | 0.01 | 1.2589E-15 | 64.00 |
| *SNX2* | rs55648536 | T | G | 0.295 | -0.081 | 0.009 | 1.9953E-20 | 81.00 |
| *SNX5* | rs1354034 | T | C | 0.6045 | 0.055 | 0.008 | 1.9953E-12 | 47.27 |
| *SNX5* | rs13172043 | G | A | 0.5561 | 0.141 | 0.008 | 1.9953E-74 | 310.64 |
| *SNX9* | rs688181 | T | C | 0.7167 | -0.089 | 0.008 | 3.9811E-27 | 123.77 |
| *SNX9* | rs342298 | C | T | 0.4549 | -0.073 | 0.007 | 1.9953E-22 | 108.76 |
| *SOD1* | rs6993770 | A | T | 0.286 | -0.109 | 0.008 | 3.9811E-40 | 185.64 |
| *SOD1* | rs7282332 | G | A | 0.103 | 0.098 | 0.012 | 1.9953E-15 | 66.69 |
| *SOD2* | rs1570805 | C | T | 0.1119 | 0.083 | 0.012 | 5.0119E-13 | 47.84 |
| *SOD2* | rs915651 | G | A | 0.1305 | -0.073 | 0.011 | 1.5849E-11 | 44.04 |
| *SOD2* | rs5746105 | A | G | 0.299 | 0.158 | 0.008 | 2.5119E-87 | 390.06 |
| *SOD2* | rs4541868 | C | A | 0.2737 | -0.057 | 0.008 | 5.0119E-12 | 50.77 |
| *SOD2* | rs117692263 | T | C | 0.0848 | 0.16 | 0.013 | 1.9953E-34 | 151.48 |
| *SOD2* | rs2072694 | C | G | 0.5182 | 0.056 | 0.007 | 1.5849E-14 | 64.00 |
| *SOD2* | rs12975366 | T | C | 0.3954 | -0.158 | 0.007 | 2.512E-99 | 509.47 |
| *SOD3* | rs3796735 | C | T | 0.6664 | -0.114 | 0.007 | 3.9811E-54 | 265.22 |
| *SOD3* | rs1799895 | C | G | 0.0119 | 2.301 | 0.032 | 1E-200 | 5170.51 |
| *SOD3* | rs13107325 | C | T | 0.0748 | 0.159 | 0.013 | 1.5849E-33 | 149.59 |
| *SOD3* | rs56278466 | T | G | 0.6608 | 0.061 | 0.007 | 7.9433E-18 | 75.94 |
| *SOD3* | rs597808 | A | G | 0.5158 | 0.054 | 0.007 | 6.3096E-16 | 59.51 |
| *SOD3* | rs184458383 | C | T | 0.0761 | -0.126 | 0.013 | 1.9953E-23 | 93.94 |
| *SOD3* | rs77542162 | A | G | 0.0228 | 0.167 | 0.022 | 7.9433E-14 | 57.62 |
| *SORCS2* | rs4234798 | T | G | 0.6173 | -0.457 | 0.008 | 1E-200 | 3263.27 |
| *SORCS2* | rs4253238 | C | T | 0.5128 | -0.074 | 0.007 | 3.1623E-24 | 111.76 |
| *SORCS2* | rs8181784 | A | G | 0.2327 | 0.057 | 0.008 | 2.5119E-12 | 50.77 |
| *SORCS2* | rs7203642 | A | G | 0.1919 | -0.065 | 0.009 | 6.3096E-14 | 52.16 |
| *SORD* | rs4131305 | G | C | 0.0344 | -0.262 | 0.02 | 6.3096E-39 | 171.61 |
| *SORD* | rs56060952 | T | C | 0.1282 | 0.205 | 0.011 | 1E-76 | 347.31 |
| *SORD* | rs3747207 | G | A | 0.2155 | 0.072 | 0.009 | 7.9433E-16 | 64.00 |
| *SORT1* | rs61394658 | G | A | 0.2266 | -0.187 | 0.009 | 1.5849E-97 | 431.72 |
| *SORT1* | rs562529886 | T | TA | 0.4709 | -0.059 | 0.008 | 7.9433E-13 | 54.39 |
| *SORT1* | rs13303 | T | C | 0.5602 | 0.066 | 0.007 | 7.9433E-19 | 88.90 |
| *SORT1* | rs61244449 | T | TCTC | 0.3714 | 0.064 | 0.008 | 7.9433E-17 | 64.00 |
| *SORT1* | rs139596721 | C | CT | 0.0322 | 0.16 | 0.021 | 2.5119E-14 | 58.05 |
| *SORT1* | rs4703589 | T | C | 0.5304 | 0.05 | 0.007 | 1.2589E-11 | 51.02 |
| *SORT1* | rs4098923 | A | G | 0.4477 | -0.055 | 0.008 | 1.5849E-12 | 47.27 |
| *SORT1* | rs139141690 | G | A | 0.0048 | 0.65 | 0.054 | 3.9811E-33 | 144.89 |

| | | | | | | | | |
|---|---|---|---|---|---|---|---|---|
| *SORT1* | rs6993770 | A | T | 0.2866 | -0.179 | 0.008 | 3.981E-106 | 500.64 |
| *SORT1* | rs385893 | T | C | 0.5238 | 0.057 | 0.007 | 7.9433E-15 | 66.31 |
| *SORT1* | rs141129381 | T | TG | 0.3628 | 0.077 | 0.008 | 1.2589E-23 | 92.64 |
| *SORT1* | rs2519093 | C | T | 0.1844 | -0.089 | 0.01 | 1.5849E-20 | 79.21 |
| *SORT1* | rs774510679 | ACTTTGCC | A | 0.412 | 0.091 | 0.008 | 1.5849E-33 | 129.39 |
| *SORT1* | rs17490626 | G | C | 0.1296 | 0.166 | 0.011 | 3.1623E-51 | 227.74 |
| *SORT1* | rs3184504 | T | C | 0.5175 | -0.074 | 0.007 | 3.1623E-24 | 111.76 |
| *SORT1* | rs892090 | T | G | 0.8338 | 0.127 | 0.01 | 1.5849E-37 | 161.29 |
| *SOST* | rs4973180 | T | C | 0.1776 | -0.08 | 0.009 | 1.5849E-18 | 79.01 |
| *SOST* | rs17138646 | T | G | 0.1155 | 0.116 | 0.011 | 1.5849E-26 | 111.21 |
| *SOST* | rs66642874 | T | C | 0.2244 | -0.066 | 0.008 | 1.2589E-15 | 68.06 |
| *SOST* | rs34115816 | G | A | 0.3212 | 0.082 | 0.007 | 7.9433E-29 | 137.22 |
| *SOST* | rs61591132 | G | A | 0.1128 | 0.089 | 0.011 | 3.1623E-16 | 65.46 |
| *SOST* | rs11995824 | C | G | 0.5435 | -0.104 | 0.007 | 3.9811E-51 | 220.73 |
| *SOST* | rs505429 | A | G | 0.3138 | 0.055 | 0.007 | 7.9433E-14 | 61.73 |
| *SOST* | rs215226 | A | G | 0.4007 | -0.197 | 0.007 | 2.512E-168 | 792.02 |
| *SOST* | rs34876302 | C | CA | 0.2157 | 0.062 | 0.009 | 1.9953E-12 | 47.46 |
| *SOST* | rs34001253 | A | G | 0.0329 | 0.189 | 0.02 | 5.0119E-22 | 89.30 |
| *SOST* | rs34369065 | C | CTT | 0.5085 | -0.06 | 0.007 | 1E-17 | 73.47 |
| *SOST* | rs1513671 | G | C | 0.6391 | -0.061 | 0.007 | 1.9953E-17 | 75.94 |
| *SOST* | rs884205 | A | C | 0.7469 | 0.072 | 0.008 | 3.1623E-19 | 81.00 |
| *SOST* | rs34952318 | G | A | 0.0504 | -0.135 | 0.016 | 1.9953E-17 | 71.19 |
| *SOST* | rs6127099 | A | T | 0.2782 | -0.056 | 0.008 | 1.5849E-12 | 49.00 |
| *SPAG1* | rs2439461 | G | A | 0.5678 | -0.715 | 0.008 | 1E-200 | 7987.89 |
| *SPARC* | rs13412535 | G | A | 0.2311 | -0.065 | 0.009 | 2.5119E-13 | 52.16 |
| *SPARC* | rs7618405 | C | A | 0.2047 | -0.063 | 0.009 | 1.2589E-11 | 49.00 |
| *SPARC* | rs1354034 | T | C | 0.6041 | -0.103 | 0.008 | 1.2589E-41 | 165.77 |
| *SPARC* | rs755492124 | TAAAG | T | 0.3938 | -0.063 | 0.008 | 1.2589E-16 | 62.02 |
| *SPARC* | rs576037440 | G | A | 0.0038 | -0.698 | 0.064 | 2.5119E-27 | 118.95 |
| *SPARC* | rs138614581 | A | AT | 0.08 | -0.099 | 0.014 | 7.9433E-13 | 50.01 |
| *SPARC* | rs6961069 | C | T | 0.4023 | 0.066 | 0.008 | 1E-17 | 68.06 |
| *SPARC* | rs6993770 | A | T | 0.2861 | -0.116 | 0.008 | 1.5849E-44 | 210.25 |
| *SPARC* | rs12767683 | C | A | 0.3249 | 0.096 | 0.008 | 3.1623E-33 | 144.00 |
| *SPARC* | rs35785237 | C | CA | 0.4311 | 0.066 | 0.008 | 3.9811E-17 | 68.06 |
| *SPARC* | rs59001897 | T | A | 0.1749 | 0.072 | 0.01 | 6.3096E-13 | 51.84 |
| *SPARC* | rs28734133 | A | G | 0.1797 | 0.083 | 0.01 | 1.2589E-17 | 68.89 |
| *SPARC* | rs892090 | T | G | 0.8343 | 0.162 | 0.01 | 2.5119E-57 | 262.44 |
| *SPARC* | rs3790176 | G | A | 0.3427 | 0.054 | 0.008 | 5.0119E-12 | 45.56 |
| *SPARCL1* | rs56273319 | A | G | 0.3534 | 0.714 | 0.008 | 1E-200 | 7965.56 |
| *SPARCL1* | rs12291877 | C | T | 0.247 | 0.05 | 0.007 | 7.9433E-12 | 51.02 |
| *SPARCL1* | rs217184 | T | C | 0.1963 | -0.15 | 0.008 | 1.9953E-78 | 351.56 |
| *SPARCL1* | rs77542162 | A | G | 0.0229 | 0.216 | 0.021 | 1.9953E-24 | 105.80 |
| *SPARCL1* | rs55639531 | C | G | 0.2857 | 0.047 | 0.007 | 1.5849E-11 | 45.08 |
| *SPART* | rs342299 | C | T | 0.4549 | -0.053 | 0.008 | 2.5119E-12 | 43.89 |
| *SPART* | rs6993770 | A | T | 0.287 | -0.078 | 0.008 | 2.5119E-21 | 95.06 |
| *SPESP1* | rs6433469 | G | A | 0.6246 | -0.089 | 0.007 | 6.3096E-36 | 161.65 |
| *SPESP1* | rs3750228 | T | C | 0.1237 | 0.09 | 0.01 | 5.0119E-18 | 81.00 |
| *SPESP1* | rs55813566 | A | C | 0.3798 | 0.063 | 0.007 | 6.3096E-19 | 81.00 |
| *SPESP1* | rs8038764 | A | C | 0.4665 | -0.105 | 0.007 | 1.9953E-51 | 225.00 |
| *SPESP1* | rs72752337 | T | C | 0.007 | 1.354 | 0.043 | 1E-200 | 991.52 |
| *SPESP1* | rs12609754 | T | C | 0.5728 | 0.061 | 0.007 | 1.5849E-18 | 75.94 |
| *SPINK1* | rs28817415 | C | T | 0.457 | 0.053 | 0.007 | 1.2589E-13 | 57.33 |
| *SPINK1* | rs7725017 | C | A | 0.6043 | -0.405 | 0.008 | 1E-200 | 2562.89 |
| *SPINK1* | rs77924615 | G | A | 0.1966 | -0.065 | 0.009 | 6.3096E-13 | 52.16 |
| *SPINK2* | rs188468174 | C | T | 0.0142 | 0.37 | 0.026 | 7.9433E-45 | 202.51 |
| *SPINK2* | rs28656303 | A | G | 0.0205 | 1.46 | 0.024 | 1E-200 | 3700.69 |
| *SPINK2* | rs2522050 | C | T | 0.1899 | 0.07 | 0.008 | 3.1623E-19 | 76.56 |
| *SPINK2* | rs56353819 | C | T | 0.3511 | -0.047 | 0.006 | 3.1623E-13 | 61.36 |
| *SPINK2* | rs78740585 | G | A | 0.1137 | 0.168 | 0.01 | 6.3096E-68 | 282.24 |
| *SPINK2* | rs10982164 | G | A | 0.0672 | 0.328 | 0.013 | 7.943E-137 | 636.59 |
| *SPINK2* | rs28449958 | G | A | 0.3439 | 0.048 | 0.006 | 7.9433E-14 | 64.00 |
| *SPINK2* | rs77542162 | A | G | 0.0229 | 0.247 | 0.021 | 1.9953E-33 | 138.34 |
| *SPINK2* | rs7247412 | T | C | 0.2657 | -0.048 | 0.007 | 7.9433E-12 | 47.02 |
| *SPINK4* | rs808096 | G | T | 0.5264 | -0.056 | 0.006 | 7.9433E-21 | 87.11 |
| *SPINK4* | rs369940 | A | T | 0.6786 | -0.068 | 0.006 | 1.2589E-25 | 128.44 |
| *SPINK4* | rs4879679 | C | T | 0.8428 | -1.114 | 0.01 | 1E-200 | 12409.96 |
| *SPINK4* | rs12804877 | A | G | 0.2244 | 0.052 | 0.007 | 7.9433E-13 | 55.18 |
| *SPINK4* | rs28529068 | A | G | 0.2425 | 0.115 | 0.007 | 2.5119E-60 | 269.90 |

| | | | | | | | | |
|---|---|---|---|---|---|---|---|---|
| *SPINK4* | rs2834350 | C | T | 0.3773 | -0.045 | 0.006 | 3.9811E-13 | 56.25 |
| *SPINK5* | rs1260326 | T | C | 0.6084 | 0.063 | 0.007 | 1E-18 | 81.00 |
| *SPINK5* | rs11384422 | A | AG | 0.5832 | 0.051 | 0.007 | 1E-12 | 53.08 |
| *SPINK5* | rs6883868 | C | T | 0.6236 | 0.41 | 0.008 | 1E-200 | 2626.56 |
| *SPINK5* | rs587600964 | C | G | 0.1193 | -0.094 | 0.011 | 1E-17 | 73.02 |
| *SPINK5* | rs11220477 | C | T | 0.0416 | -0.133 | 0.017 | 3.1623E-14 | 61.21 |
| *SPINK5* | rs186021206 | G | A | 0.0064 | 0.865 | 0.048 | 1.2589E-72 | 324.75 |
| *SPINK5* | rs681343 | C | T | 0.5096 | 0.198 | 0.007 | 1.259E-171 | 800.08 |
| *SPINK6* | rs12717962 | A | G | 0.5537 | -0.359 | 0.007 | 1E-200 | 2630.22 |
| *SPINK8* | rs78159520 | T | C | 0.3144 | -0.461 | 0.008 | 1E-200 | 3320.64 |
| *SPINT1* | rs4671378 | T | C | 0.5588 | 0.063 | 0.007 | 1.9953E-17 | 81.00 |
| *SPINT1* | rs13080057 | G | A | 0.0856 | 0.13 | 0.013 | 1E-22 | 100.00 |
| *SPINT1* | rs538174489 | C | T | 0.3078 | -0.079 | 0.008 | 5.0119E-23 | 97.52 |
| *SPINT1* | rs56278466 | T | G | 0.6612 | 0.089 | 0.008 | 2.5119E-30 | 123.77 |
| *SPINT1* | rs73415720 | T | C | 0.4738 | -0.05 | 0.007 | 1.2589E-11 | 51.02 |
| *SPINT1* | rs17658212 | C | T | 0.0599 | -0.676 | 0.016 | 1E-200 | 1785.06 |
| *SPINT1* | rs186021206 | G | A | 0.0064 | 0.84 | 0.051 | 1E-61 | 271.28 |
| *SPINT1* | rs708686 | C | T | 0.268 | 0.104 | 0.008 | 2.5119E-35 | 169.00 |
| *SPINT1* | rs584768 | G | A | 0.5339 | 0.052 | 0.007 | 2.5119E-12 | 55.18 |
| *SPINT2* | rs13412535 | G | A | 0.231 | -0.069 | 0.009 | 1.9953E-15 | 58.78 |
| *SPINT2* | rs1354034 | T | C | 0.6041 | -0.095 | 0.007 | 5.0119E-37 | 184.18 |
| *SPINT2* | rs5030062 | A | C | 0.3731 | -0.068 | 0.008 | 7.9433E-19 | 72.25 |
| *SPINT2* | rs10016018 | A | T | 0.3971 | -0.06 | 0.008 | 1.9953E-15 | 56.25 |
| *SPINT2* | rs139596721 | C | CT | 0.0321 | 0.145 | 0.021 | 6.3096E-12 | 47.68 |
| *SPINT2* | rs66530140 | T | C | 0.5136 | -0.153 | 0.007 | 1.5849E-94 | 477.73 |
| *SPINT2* | rs10058074 | G | A | 0.4757 | -0.056 | 0.007 | 1.9953E-14 | 64.00 |
| *SPINT2* | rs2545801 | T | C | 0.7444 | -0.072 | 0.008 | 6.3096E-18 | 81.00 |
| *SPINT2* | rs35173808 | C | A | 0.0966 | -0.098 | 0.012 | 3.9811E-15 | 66.69 |
| *SPINT2* | rs6961069 | C | T | 0.4025 | 0.071 | 0.008 | 3.1623E-21 | 78.77 |
| *SPINT2* | rs139141690 | G | A | 0.0048 | 0.439 | 0.054 | 3.9811E-16 | 66.09 |
| *SPINT2* | rs6993770 | A | T | 0.2867 | -0.127 | 0.008 | 2.5119E-55 | 252.02 |
| *SPINT2* | rs10761737 | T | C | 0.4132 | 0.091 | 0.007 | 3.1623E-34 | 169.00 |
| *SPINT2* | rs11591571 | G | A | 0.3409 | 0.053 | 0.008 | 7.9433E-12 | 43.89 |
| *SPINT2* | rs111338191 | A | AAATT | 0.5211 | -0.059 | 0.007 | 1.5849E-15 | 71.04 |
| *SPINT2* | rs12445050 | C | T | 0.138 | 0.09 | 0.011 | 2.5119E-17 | 66.94 |
| *SPINT2* | rs45437199 | G | A | 0.1832 | -0.191 | 0.009 | 6.3096E-90 | 450.38 |
| *SPINT2* | rs892090 | T | G | 0.8342 | 0.133 | 0.01 | 1E-40 | 176.89 |
| *SPINT3* | rs6073773 | G | C | 0.5333 | 0.091 | 0.005 | 3.1623E-81 | 331.24 |
| *SPOCK1* | rs34546909 | G | GT | 0.804 | 0.149 | 0.009 | 1.9953E-58 | 274.09 |
| *SPOCK1* | rs4655581 | C | T | 0.0942 | -0.094 | 0.013 | 7.9433E-14 | 52.28 |
| *SPOCK1* | rs77683480 | G | A | 0.3108 | -0.055 | 0.008 | 6.3096E-12 | 47.27 |
| *SPOCK1* | rs62155324 | C | A | 0.0352 | -0.186 | 0.02 | 5.0119E-20 | 86.49 |
| *SPOCK1* | rs4835737 | A | G | 0.3651 | -0.131 | 0.008 | 5.0119E-66 | 268.14 |
| *SPOCK1* | rs7756288 | T | C | 0.2564 | -0.098 | 0.008 | 5.0119E-31 | 150.06 |
| *SPOCK1* | rs3129777 | C | A | 0.2151 | -0.09 | 0.01 | 7.9433E-21 | 81.00 |
| *SPOCK1* | rs2505507 | C | T | 0.275 | 0.061 | 0.008 | 1.5849E-13 | 58.14 |
| *SPOCK1* | rs703987 | G | C | 0.6115 | -0.056 | 0.008 | 6.3096E-14 | 49.00 |
| *SPOCK1* | rs149440923 | A | G | 0.0154 | 0.227 | 0.03 | 3.9811E-14 | 57.25 |
| *SPOCK1* | rs36100715 | T | G | 0.1135 | -0.113 | 0.012 | 1.5849E-22 | 88.67 |
| *SPOCK1* | rs3916033 | C | T | 0.5549 | 0.052 | 0.007 | 1.5849E-12 | 55.18 |
| *SPON1* | rs1270942 | A | G | 0.1319 | -0.097 | 0.01 | 1.5849E-21 | 94.09 |
| *SPON1* | rs8176741 | G | A | 0.0612 | 0.159 | 0.014 | 2.5119E-28 | 128.98 |
| *SPON1* | rs10832164 | C | T | 0.5196 | -0.291 | 0.007 | 1E-200 | 1728.18 |
| *SPON1* | rs4939113 | A | G | 0.1969 | 0.076 | 0.009 | 1.9953E-17 | 71.31 |
| *SPON1* | rs77542162 | A | G | 0.0229 | 0.276 | 0.023 | 5.0119E-33 | 144.00 |
| *SPON1* | rs516316 | G | C | 0.5097 | -0.074 | 0.007 | 6.3096E-27 | 111.76 |
| *SPON2* | rs28855974 | A | G | 0.1178 | 0.315 | 0.012 | 5.012E-163 | 689.06 |
| *SPP1* | rs35085476 | T | C | 0.0232 | -0.167 | 0.024 | 3.9811E-12 | 48.42 |
| *SPP1* | rs559637118 | G | GT | 0.4033 | -0.053 | 0.007 | 3.1623E-13 | 57.33 |
| *SPP1* | rs13303 | T | C | 0.56 | 0.05 | 0.007 | 5.0119E-12 | 51.02 |
| *SPP1* | rs56254643 | T | C | 0.1917 | -0.114 | 0.009 | 7.9433E-36 | 160.44 |
| *SPP1* | rs142201367 | T | TTTTCTAAC | 0.4954 | -0.06 | 0.007 | 1.9953E-16 | 73.47 |
| *SPP1* | rs11995824 | C | G | 0.5434 | 0.063 | 0.007 | 2.5119E-18 | 81.00 |
| *SPP1* | rs548589580 | T | TA | 0.4834 | 0.065 | 0.007 | 3.1623E-19 | 86.22 |
| *SPP1* | rs117068593 | C | T | 0.1871 | -0.086 | 0.009 | 1.2589E-20 | 91.31 |
| *SPP1* | rs537848612 | T | TCGG | 0.1173 | 0.076 | 0.011 | 1.5849E-11 | 47.74 |
| *SPP1* | rs738408 | C | T | 0.217 | 0.063 | 0.009 | 6.3096E-13 | 49.00 |
| *SPRED2* | rs7569084 | C | T | 0.5856 | -0.072 | 0.008 | 7.9433E-21 | 81.00 |

| | | | | | | | | |
|---|---|---|---|---|---|---|---|---|
| *SPRING1* | rs71534539 | G | A | 0.464 | -0.071 | 0.009 | 1.2589E-16 | 62.23 |
| *SPRR1B* | rs9272544 | G | A | 0.4953 | -0.055 | 0.008 | 7.9433E-13 | 47.27 |
| *SPRR3* | rs61811427 | C | T | 0.5515 | 0.24 | 0.007 | 1E-200 | 1175.51 |
| *SPRR3* | rs7959052 | T | C | 0.1802 | -0.072 | 0.009 | 7.9433E-16 | 64.00 |
| *SPRR3* | rs35887622 | A | G | 0.0147 | 0.302 | 0.028 | 1.5849E-26 | 116.33 |
| *SPRR3* | rs12970138 | G | T | 0.4083 | 0.052 | 0.007 | 1E-13 | 55.18 |
| *SPRY2* | rs1354034 | T | C | 0.6035 | 0.1 | 0.008 | 2.5119E-38 | 156.25 |
| *SPRY2* | rs6866614 | A | G | 0.5736 | 0.052 | 0.008 | 1.5849E-11 | 42.25 |
| *SPRY2* | rs342298 | C | T | 0.4542 | -0.067 | 0.008 | 1.5849E-18 | 70.14 |
| *SPRY2* | rs6993770 | A | T | 0.2872 | -0.08 | 0.008 | 6.3096E-22 | 100.00 |
| *SPRY2* | rs11502185 | T | C | 0.2594 | 0.072 | 0.01 | 1.5849E-13 | 51.84 |
| *SPRY2* | rs6580981 | G | A | 0.4595 | 0.079 | 0.008 | 7.9433E-25 | 97.52 |
| *SPRY2* | rs504122 | G | A | 0.3698 | -0.082 | 0.008 | 1.2589E-25 | 105.06 |
| *SPTLC1* | rs45461899 | C | A | 0.024 | -0.55 | 0.025 | 7.943E-108 | 484.00 |
| *SPTLC1* | rs2272587 | C | G | 0.5779 | -0.078 | 0.008 | 3.1623E-24 | 95.06 |
| *SRC* | rs17622656 | G | A | 0.3884 | -0.054 | 0.008 | 2.5119E-12 | 45.56 |
| *SRP14* | rs1354034 | T | C | 0.6046 | 0.108 | 0.008 | 5.0119E-47 | 182.25 |
| *SRPK2* | rs1354034 | T | C | 0.6043 | 0.108 | 0.008 | 7.9433E-46 | 182.25 |
| *SRPK2* | rs342298 | C | T | 0.4546 | -0.053 | 0.007 | 7.9433E-13 | 57.33 |
| *SRPK2* | rs7080386 | C | A | 0.4121 | 0.064 | 0.008 | 3.9811E-17 | 64.00 |
| *SRPK2* | rs11502185 | T | C | 0.2598 | 0.065 | 0.01 | 7.9433E-12 | 42.25 |
| *SRPK2* | rs60822569 | T | TTTCTC | 0.5547 | 0.052 | 0.008 | 6.3096E-12 | 42.25 |
| *SRPX* | rs28929474 | C | T | 0.0209 | 0.209 | 0.024 | 1E-17 | 75.84 |
| *SRPX* | rs77542162 | A | G | 0.0229 | 0.434 | 0.023 | 2.5119E-77 | 356.06 |
| *SSC4D* | rs12140070 | A | G | 0.3076 | -0.154 | 0.007 | 5.0119E-95 | 484.00 |
| *SSC4D* | rs469773 | C | T | 0.1974 | -0.07 | 0.009 | 5.0119E-16 | 60.49 |
| *SSC4D* | rs6908943 | C | T | 0.1768 | -0.067 | 0.009 | 3.9811E-13 | 55.42 |
| *SSC4D* | rs12540573 | A | C | 0.0615 | 0.687 | 0.015 | 1E-200 | 2097.64 |
| *SSC4D* | rs11183272 | C | A | 0.4894 | 0.053 | 0.007 | 7.9433E-15 | 57.33 |
| *SSC4D* | rs112635299 | G | T | 0.0216 | 0.184 | 0.024 | 5.0119E-15 | 58.78 |
| *SSC4D* | rs2498786 | C | G | 0.6189 | -0.103 | 0.007 | 7.9433E-48 | 216.51 |
| *SSC4D* | rs339969 | C | A | 0.617 | 0.058 | 0.007 | 1.2589E-16 | 68.65 |
| *SSC4D* | rs35523349 | C | CA | 0.4598 | 0.052 | 0.007 | 2.5119E-13 | 55.18 |
| *SSC4D* | rs4803852 | G | T | 0.8228 | 0.065 | 0.009 | 3.9811E-13 | 52.16 |
| *SSC4D* | rs1800961 | C | T | 0.0323 | -0.157 | 0.019 | 3.1623E-16 | 68.28 |
| *SSC4D* | rs2739344 | A | C | 0.5803 | -0.055 | 0.007 | 1.9953E-15 | 61.73 |
| *SSC5D* | rs56210800 | C | G | 0.1306 | -0.092 | 0.01 | 1.5849E-19 | 84.64 |
| *SSC5D* | rs13107325 | C | T | 0.075 | 0.15 | 0.013 | 1.9953E-30 | 133.14 |
| *SSC5D* | rs2142306 | T | C | 0.4118 | 0.09 | 0.007 | 6.3096E-38 | 165.31 |
| *SSC5D* | rs7358384 | T | A | 0.383 | 0.107 | 0.007 | 2.5119E-51 | 233.65 |
| *SSC5D* | rs964184 | G | C | 0.8668 | -0.075 | 0.01 | 1.5849E-13 | 56.25 |
| *SSC5D* | rs10778118 | C | T | 0.7893 | 0.062 | 0.008 | 1.9953E-13 | 60.06 |
| *SSC5D* | rs200489612 | G | A | 0.0052 | 0.685 | 0.052 | 2.5119E-40 | 173.53 |
| *SSC5D* | rs55799523 | C | A | 0.2652 | 0.539 | 0.008 | 1E-200 | 4539.39 |
| *SSC5D* | rs181210490 | G | A | 0.0118 | 0.85 | 0.033 | 3.162E-150 | 663.45 |
| *SSH3* | rs9275576 | C | T | 0.144 | -0.101 | 0.011 | 1E-19 | 84.31 |
| *SSNA1* | rs1354034 | T | C | 0.6046 | 0.101 | 0.008 | 7.9433E-40 | 159.39 |
| *SSNA1* | rs342298 | C | T | 0.455 | -0.055 | 0.008 | 1.9953E-13 | 47.27 |
| *SSNA1* | rs535590282 | G | T | 0.0014 | -1.026 | 0.108 | 1.9953E-21 | 90.25 |
| *SSNA1* | rs79755767 | G | A | 0.0989 | 0.095 | 0.013 | 6.3096E-14 | 53.40 |
| *ST13* | rs1354034 | T | C | 0.6044 | 0.068 | 0.008 | 1.2589E-19 | 72.25 |
| *ST13* | rs1005402 | T | C | 0.5334 | -0.054 | 0.007 | 3.1623E-13 | 59.51 |
| *ST3GAL1* | rs12740374 | G | T | 0.2212 | -0.104 | 0.009 | 3.9811E-33 | 133.53 |
| *ST3GAL1* | rs539657009 | C | T | 0.1294 | 0.08 | 0.011 | 5.0119E-13 | 52.89 |
| *ST3GAL1* | rs4672497 | C | G | 0.2218 | 0.18 | 0.009 | 1E-94 | 400.00 |
| *ST3GAL1* | rs9643300 | C | T | 0.565 | -0.237 | 0.007 | 1E-200 | 1146.31 |
| *ST3GAL1* | rs782134971 | GAAACTGC( | G | 0.2543 | 0.139 | 0.008 | 1.9953E-62 | 301.89 |
| *ST3GAL1* | rs56278466 | T | G | 0.6611 | 0.053 | 0.008 | 1.9953E-12 | 43.89 |
| *ST3GAL1* | rs28929474 | C | T | 0.0207 | 0.244 | 0.025 | 5.0119E-22 | 95.26 |
| *ST3GAL1* | rs28873836 | G | C | 0.2658 | -0.101 | 0.008 | 5.0119E-35 | 159.39 |
| *ST6GAL1* | rs193294272 | G | T | 0.0087 | 0.306 | 0.042 | 3.9811E-13 | 53.08 |
| *ST6GAL1* | rs4686837 | G | A | 0.3128 | -0.18 | 0.008 | 3.162E-113 | 506.25 |
| *ST6GAL1* | rs6993770 | A | T | 0.2865 | -0.057 | 0.008 | 1.5849E-12 | 50.77 |
| *ST6GAL1* | rs142744740 | A | C | 0.8114 | -0.076 | 0.01 | 5.0119E-13 | 57.76 |
| *ST6GAL1* | rs71454159 | A | T | 0.7165 | 0.059 | 0.008 | 5.0119E-13 | 54.39 |
| *ST6GAL1* | rs2695293 | C | A | 0.5766 | 0.054 | 0.007 | 5.0119E-13 | 59.51 |
| *ST6GAL1* | rs28929474 | C | T | 0.0208 | 0.209 | 0.026 | 3.1623E-16 | 64.62 |
| *ST6GAL1* | rs61658003 | C | A | 0.8428 | 0.072 | 0.01 | 1.9953E-12 | 51.84 |

| | | | | | | | | |
|---|---|---|---|---|---|---|---|---|
| *ST8SIA1* | rs28636516 | A | G | 0.1282 | 0.088 | 0.012 | 3.1623E-14 | 53.78 |
| *STAB2* | rs61801010 | A | G | 0.1063 | -0.128 | 0.012 | 1E-28 | 113.78 |
| *STAB2* | rs10935473 | G | T | 0.4405 | -0.097 | 0.007 | 5.0119E-46 | 192.02 |
| *STAB2* | rs200663261 | C | CTACA | 0.2612 | -0.079 | 0.008 | 1.2589E-24 | 97.52 |
| *STAB2* | rs4896219 | C | A | 0.5167 | -0.053 | 0.007 | 6.3096E-15 | 57.33 |
| *STAB2* | rs1969976 | A | G | 0.3059 | 0.074 | 0.008 | 6.3096E-22 | 85.56 |
| *STAB2* | rs4055121 | C | T | 0.1347 | -0.082 | 0.01 | 1.9953E-16 | 67.24 |
| *STAB2* | rs215225 | A | G | 0.4282 | -0.115 | 0.007 | 2.5119E-56 | 269.90 |
| *STAB2* | rs115840458 | A | G | 0.0672 | 0.8 | 0.014 | 1E-200 | 3265.31 |
| *STAB2* | rs12920182 | A | G | 0.3319 | -0.059 | 0.007 | 1.2589E-16 | 71.04 |
| *STAB2* | rs186021206 | G | A | 0.0064 | 0.673 | 0.047 | 3.1623E-47 | 205.04 |
| *STAB2* | rs77542162 | A | G | 0.0229 | 0.192 | 0.023 | 3.1623E-17 | 69.69 |
| *STAB2* | rs12979429 | G | A | 0.2839 | 0.063 | 0.008 | 1.2589E-16 | 62.02 |
| *STAB2* | rs11672900 | A | G | 0.5445 | 0.052 | 0.007 | 1.9953E-14 | 55.18 |
| *STAB2* | rs145144275 | A | G | 0.0089 | -0.323 | 0.036 | 3.9811E-19 | 80.50 |
| *STAMBP* | rs559935910 | G | A | 0.0016 | -0.899 | 0.108 | 7.9433E-17 | 69.29 |
| *STAMBP* | rs1354034 | T | C | 0.6039 | 0.058 | 0.008 | 3.9811E-14 | 52.56 |
| *STAMBP* | rs10733789 | T | C | 0.3128 | 0.055 | 0.008 | 7.9433E-12 | 47.27 |
| *STAT2* | rs1354034 | T | C | 0.6039 | 0.076 | 0.008 | 3.9811E-23 | 90.25 |
| *STAT2* | rs61754171 | A | G | 0.0187 | -0.558 | 0.029 | 1.5849E-83 | 370.23 |
| *STAT5B* | rs342298 | C | T | 0.4545 | -0.062 | 0.008 | 1.9953E-16 | 60.06 |
| *STAT5B* | rs6993770 | A | T | 0.2864 | -0.068 | 0.008 | 2.5119E-16 | 72.25 |
| *STAU1* | rs13412535 | G | A | 0.2306 | -0.116 | 0.009 | 1.5849E-36 | 166.12 |
| *STAU1* | rs71534539 | G | A | 0.464 | -0.079 | 0.009 | 2.5119E-20 | 77.05 |
| *STAU1* | rs34200987 | T | G | 0.2328 | 0.106 | 0.01 | 6.3096E-25 | 112.36 |
| *STAU1* | rs11631612 | C | T | 0.2924 | -0.059 | 0.009 | 6.3096E-12 | 42.98 |
| *STC1* | rs57247992 | G | A | 0.0765 | 0.121 | 0.014 | 1.2589E-18 | 74.70 |
| *STC1* | rs2519093 | C | T | 0.1846 | -0.08 | 0.009 | 3.1623E-17 | 79.01 |
| *STC1* | rs113185690 | C | T | 0.0766 | 0.113 | 0.014 | 5.0119E-16 | 65.15 |
| *STC1* | rs35451048 | C | CT | 0.39 | -0.06 | 0.008 | 1.2589E-14 | 56.25 |
| *STC1* | rs9529913 | C | T | 0.604 | 0.079 | 0.007 | 7.9433E-26 | 127.37 |
| *STC1* | rs12460997 | C | A | 0.4845 | -0.114 | 0.007 | 2.5119E-55 | 265.22 |
| *STC2* | rs113239047 | A | AAG | 0.2861 | 0.058 | 0.008 | 1.2589E-12 | 52.56 |
| *STC2* | rs2894602 | A | G | 0.7658 | 0.061 | 0.009 | 1.2589E-12 | 45.94 |
| *STC2* | rs11103374 | C | T | 0.132 | 0.075 | 0.011 | 3.1623E-12 | 46.49 |
| *STC2* | rs11362821 | G | A | 0.9753 | -0.164 | 0.024 | 3.1623E-12 | 46.69 |
| *STC2* | rs3184504 | T | C | 0.5175 | -0.057 | 0.007 | 1.9953E-15 | 66.31 |
| *STC2* | rs528207197 | C | CT | 0.1367 | 0.074 | 0.011 | 2.5119E-12 | 45.26 |
| *STC2* | rs1126464 | G | C | 0.2438 | -0.068 | 0.008 | 6.3096E-16 | 72.25 |
| *STC2* | rs12986064 | T | C | 0.5076 | -0.069 | 0.007 | 2.5119E-20 | 97.16 |
| *STIP1* | rs1354034 | T | C | 0.604 | 0.064 | 0.008 | 2.5119E-17 | 64.00 |
| *STIP1* | rs10739067 | A | G | 0.3429 | 0.077 | 0.008 | 7.9433E-23 | 92.64 |
| *STK11* | rs1354034 | T | C | 0.6038 | 0.073 | 0.008 | 1.2589E-21 | 83.27 |
| *STK11* | rs342293 | C | G | 0.4597 | -0.052 | 0.008 | 6.3096E-12 | 42.25 |
| *STK11* | rs6993770 | A | T | 0.2867 | -0.072 | 0.008 | 6.3096E-18 | 81.00 |
| *STK11* | rs7924036 | G | T | 0.5014 | 0.052 | 0.007 | 3.9811E-12 | 55.18 |
| *STK11* | rs892090 | T | G | 0.8339 | 0.068 | 0.01 | 1.5849E-11 | 46.24 |
| *STK24* | rs6993770 | A | T | 0.2869 | -0.059 | 0.008 | 1.9953E-12 | 54.39 |
| *STK4* | rs1354034 | T | C | 0.6037 | 0.073 | 0.008 | 1.2589E-21 | 83.27 |
| *STK4* | rs6993770 | A | T | 0.2866 | -0.077 | 0.008 | 2.5119E-20 | 92.64 |
| *STK4* | rs2299978 | T | C | 0.4741 | 0.053 | 0.008 | 1.9953E-12 | 43.89 |
| *STX16* | rs218476 | G | A | 0.3796 | 0.062 | 0.008 | 3.9811E-15 | 60.06 |
| *STX3* | rs10191964 | G | A | 0.1596 | -0.253 | 0.01 | 3.162E-132 | 640.09 |
| *STX3* | rs9271377 | T | G | 0.3663 | -0.059 | 0.008 | 6.3096E-14 | 54.39 |
| *STX4* | rs138182020 | C | CT | 0.2625 | -0.074 | 0.009 | 1.5849E-17 | 67.60 |
| *STX7* | rs3811444 | C | T | 0.3324 | -0.081 | 0.007 | 1.2589E-30 | 133.90 |
| *STX7* | rs1318638 | C | T | 0.0634 | 0.105 | 0.015 | 3.9811E-12 | 49.00 |
| *STX7* | rs3813356 | C | T | 0.4513 | -0.59 | 0.008 | 1E-200 | 5439.06 |
| *STX8* | rs1354034 | T | C | 0.6043 | 0.054 | 0.008 | 1.5849E-12 | 45.56 |
| *STX8* | rs6993770 | A | T | 0.2869 | -0.06 | 0.008 | 2.5119E-13 | 56.25 |
| *STX8* | rs8077500 | G | A | 0.2441 | 0.102 | 0.009 | 2.5119E-31 | 128.44 |
| *STXBP1* | rs1354034 | T | C | 0.6043 | 0.058 | 0.008 | 3.1623E-14 | 52.56 |
| *STXBP1* | rs7896518 | A | G | 0.4237 | 0.083 | 0.008 | 7.9433E-27 | 107.64 |
| *SUGP1* | rs17751061 | C | T | 0.1595 | 0.325 | 0.01 | 1E-200 | 1056.25 |
| *SUGT1* | rs1354034 | T | C | 0.6038 | 0.063 | 0.008 | 3.9811E-16 | 62.02 |
| *SULT1A1* | rs148788997 | G | C | 0.1261 | 0.15 | 0.012 | 1E-38 | 156.25 |
| *SULT2A1* | rs4861708 | G | A | 0.5119 | 0.077 | 0.007 | 1.5849E-25 | 121.00 |
| *SULT2A1* | rs6894249 | A | G | 0.3868 | -0.052 | 0.008 | 6.3096E-12 | 42.25 |

| | | | | | | | | |
|---|---|---|---|---|---|---|---|---|
| *SULT2A1* | rs62129966 | C | A | 0.1644 | -0.378 | 0.01 | 1E-200 | 1428.84 |
| *SULT2A1* | rs3747207 | G | A | 0.216 | 0.073 | 0.009 | 3.1623E-16 | 65.79 |
| *SUMF1* | rs10494172 | G | C | 0.4943 | -0.11 | 0.008 | 6.3096E-46 | 189.06 |
| *SUMF2* | rs1289395 | T | C | 0.4627 | -0.048 | 0.007 | 1E-11 | 47.02 |
| *SUMF2* | rs9852409 | T | A | 0.8215 | -0.189 | 0.009 | 3.1623E-91 | 441.00 |
| *SUMF2* | rs114082534 | G | A | 0.0555 | 0.115 | 0.016 | 1.2589E-13 | 51.66 |
| *SUMF2* | rs62396264 | C | G | 0.0361 | 0.255 | 0.019 | 3.9811E-41 | 180.12 |
| *SUMF2* | rs6796 | T | C | 0.2789 | 0.072 | 0.008 | 3.9811E-18 | 81.00 |
| *SUMF2* | rs35384521 | G | A | 0.019 | -1.533 | 0.027 | 1E-200 | 3223.72 |
| *SUMF2* | rs556893622 | G | A | 0.0083 | -0.533 | 0.046 | 1E-31 | 134.26 |
| *SUMF2* | rs79492875 | C | A | 0.0688 | -0.192 | 0.015 | 5.0119E-38 | 163.84 |
| *SUMF2* | rs199922514 | A | AAGAAGAA | 0.9076 | -0.224 | 0.012 | 1.2589E-73 | 348.44 |
| *SUMF2* | rs11999525 | G | A | 0.2591 | -0.061 | 0.008 | 7.9433E-14 | 58.14 |
| *SUMF2* | rs477838 | A | G | 0.2874 | -0.061 | 0.008 | 5.0119E-15 | 58.14 |
| *SUMF2* | rs1169288 | A | C | 0.3139 | -0.054 | 0.008 | 2.5119E-12 | 45.56 |
| *SUMF2* | rs2291256 | C | T | 0.0862 | -0.099 | 0.013 | 5.0119E-15 | 57.99 |
| *SUMF2* | rs438811 | C | T | 0.2376 | 0.063 | 0.008 | 6.3096E-14 | 62.02 |
| *SUMF2* | rs4806498 | C | T | 0.4311 | -0.071 | 0.007 | 3.1623E-23 | 102.88 |
| *SUOX* | rs6674486 | A | T | 0.3468 | 0.053 | 0.008 | 7.9433E-12 | 43.89 |
| *SUOX* | rs13874 | C | T | 0.4371 | 0.055 | 0.007 | 1.2589E-13 | 61.73 |
| *SUOX* | rs9272212 | G | A | 0.3879 | -0.055 | 0.008 | 7.9433E-13 | 47.27 |
| *SUOX* | rs75773116 | A | G | 0.0136 | 1.179 | 0.032 | 1E-200 | 1357.46 |
| *SUOX* | rs141735896 | C | T | 0.0059 | -0.393 | 0.048 | 1.9953E-16 | 67.04 |
| *SUOX* | rs112635299 | G | T | 0.0217 | 0.173 | 0.025 | 6.3096E-12 | 47.89 |
| *SUOX* | rs3747207 | G | A | 0.2157 | 0.075 | 0.009 | 3.9811E-17 | 69.44 |
| *SUSD1* | rs9820435 | C | T | 0.5932 | -0.055 | 0.008 | 1.9953E-12 | 47.27 |
| *SUSD1* | rs2631360 | G | A | 0.5165 | 0.06 | 0.008 | 2.5119E-15 | 56.25 |
| *SUSD1* | rs146879704 | C | T | 0.0032 | -1.267 | 0.068 | 7.9433E-77 | 347.16 |
| *SUSD1* | rs61744929 | T | C | 0.0191 | -0.441 | 0.028 | 1.2589E-56 | 248.06 |
| *SUSD2* | rs62292583 | C | T | 0.1285 | -0.084 | 0.01 | 1E-16 | 70.56 |
| *SUSD2* | rs9269502 | A | G | 0.359 | -0.062 | 0.008 | 1E-13 | 60.06 |
| *SUSD2* | rs200489612 | G | A | 0.0053 | 0.578 | 0.05 | 1.9953E-30 | 133.63 |
| *SUSD2* | rs117587385 | G | A | 0.0375 | 1.205 | 0.019 | 1E-200 | 4022.23 |
| *SUSD4* | rs1418152 | T | G | 0.4014 | -0.38 | 0.008 | 1E-200 | 2256.25 |
| *SUSD4* | rs28929474 | C | T | 0.0208 | 0.314 | 0.025 | 1.5849E-35 | 157.75 |
| *SUSD5* | rs150816167 | T | C | 0.0419 | 0.12 | 0.016 | 1.2589E-14 | 56.25 |
| *SUSD5* | rs1260326 | T | C | 0.6081 | 0.056 | 0.006 | 6.3096E-19 | 87.11 |
| *SUSD5* | rs75166367 | G | A | 0.0602 | -0.098 | 0.013 | 3.9811E-14 | 56.83 |
| *SUSD5* | rs62621812 | G | A | 0.0211 | 0.224 | 0.022 | 2.5119E-24 | 103.67 |
| *SUSD5* | rs4752550 | C | T | 0.4155 | 0.043 | 0.006 | 5.0119E-12 | 51.36 |
| *SUSD5* | rs36060036 | C | T | 0.1676 | -0.061 | 0.008 | 1.2589E-13 | 58.14 |
| *SV2A* | rs68144650 | A | C | 0.0802 | -0.296 | 0.014 | 3.9811E-98 | 447.02 |
| *SWAP70* | rs7123629 | C | G | 0.9385 | -0.29 | 0.016 | 1.5849E-75 | 328.52 |
| *SYAP1* | rs1354034 | T | C | 0.6044 | 0.079 | 0.008 | 6.3096E-25 | 97.52 |
| *SYAP1* | rs342299 | C | T | 0.4549 | -0.055 | 0.008 | 5.0119E-13 | 47.27 |
| *SYAP1* | rs6993770 | A | T | 0.287 | -0.061 | 0.008 | 3.1623E-13 | 58.14 |
| *SYAP1* | rs7080386 | C | A | 0.412 | 0.06 | 0.008 | 3.9811E-15 | 56.25 |
| *SYTL4* | rs11242109 | G | T | 0.4813 | -0.051 | 0.008 | 1E-11 | 40.64 |
| *TAB2* | rs1354034 | T | C | 0.6041 | 0.053 | 0.008 | 3.9811E-12 | 43.89 |
| *TACC3* | rs745543985 | CCGTCCCC/ | C | 0.4674 | 0.205 | 0.008 | 3.162E-161 | 656.64 |
| *TACC3* | rs6993770 | A | T | 0.2864 | -0.067 | 0.008 | 5.0119E-16 | 70.14 |
| *TACC3* | rs7080386 | C | A | 0.4117 | 0.084 | 0.008 | 3.1623E-28 | 110.25 |
| *TACC3* | rs11865642 | A | C | 0.1889 | 0.071 | 0.01 | 1.9953E-13 | 50.41 |
| *TACC3* | rs13381663 | T | C | 0.2089 | 0.088 | 0.009 | 3.9811E-22 | 95.60 |
| *TACSTD2* | rs7333 | C | T | 0.1643 | -0.907 | 0.011 | 1E-200 | 6798.75 |
| *TACSTD2* | rs12740374 | G | T | 0.2215 | -0.147 | 0.009 | 3.1623E-54 | 266.78 |
| *TACSTD2* | rs34070949 | C | A | 0.4723 | -0.053 | 0.007 | 1.9953E-15 | 57.33 |
| *TACSTD2* | rs73745607 | T | C | 0.082 | 0.102 | 0.013 | 1.5849E-15 | 61.56 |
| *TACSTD2* | rs7824174 | T | C | 0.4791 | 0.259 | 0.007 | 1E-200 | 1369.00 |
| *TACSTD2* | rs8176693 | C | T | 0.0612 | 0.277 | 0.014 | 1.2589E-88 | 391.47 |
| *TACSTD2* | rs529700 | T | C | 0.514 | 0.071 | 0.008 | 1.9953E-19 | 78.77 |
| *TACSTD2* | rs261342 | G | C | 0.7774 | -0.07 | 0.008 | 5.0119E-18 | 76.56 |
| *TACSTD2* | rs56228609 | C | T | 0.3152 | 0.073 | 0.007 | 3.9811E-24 | 108.76 |
| *TACSTD2* | rs72802342 | C | A | 0.0762 | -0.1 | 0.013 | 3.9811E-15 | 59.17 |
| *TACSTD2* | rs186021206 | G | A | 0.0065 | 0.659 | 0.045 | 7.9433E-48 | 214.46 |
| *TACSTD2* | rs72835428 | C | T | 0.1234 | 0.116 | 0.01 | 3.9811E-30 | 134.56 |
| *TACSTD2* | rs601338 | G | A | 0.5097 | 0.066 | 0.007 | 7.9433E-23 | 88.90 |
| *TACSTD2* | rs2868346 | C | T | 0.76 | 0.06 | 0.008 | 5.0119E-15 | 56.25 |

| | | | | | | | | |
|---|---|---|---|---|---|---|---|---|
| *TADA3* | rs1354034 | T | C | 0.6044 | 0.057 | 0.008 | 1E-13 | 50.77 |
| *TAFA5* | rs2870238 | C | T | 0.5033 | 0.061 | 0.007 | 7.9433E-17 | 75.94 |
| *TAFA5* | rs113878851 | C | CT | 0.1719 | -0.081 | 0.01 | 5.0119E-16 | 65.61 |
| *TAFA5* | rs79072 | G | T | 0.7146 | 0.182 | 0.009 | 3.1623E-98 | 408.94 |
| *TALDO1* | rs1354034 | T | C | 0.6046 | 0.062 | 0.007 | 1.5849E-16 | 78.45 |
| *TALDO1* | rs6993770 | A | T | 0.2864 | -0.085 | 0.008 | 1.5849E-25 | 112.89 |
| *TALDO1* | rs150770244 | G | A | 0.0072 | 0.506 | 0.047 | 1.5849E-26 | 115.91 |
| *TALDO1* | rs6580981 | G | A | 0.4581 | 0.059 | 0.007 | 1.9953E-15 | 71.04 |
| *TARBP2* | rs1354034 | T | C | 0.6035 | 0.073 | 0.008 | 2.5119E-21 | 83.27 |
| *TARBP2* | rs342293 | C | G | 0.4592 | -0.055 | 0.008 | 3.1623E-13 | 47.27 |
| *TAX1BP1* | rs1354034 | T | C | 0.6043 | 0.099 | 0.008 | 3.9811E-39 | 153.14 |
| *TAX1BP1* | rs6993770 | A | T | 0.2865 | -0.057 | 0.008 | 1.9953E-12 | 50.77 |
| *TAX1BP1* | rs11604127 | C | T | 0.2307 | 0.064 | 0.009 | 5.0119E-13 | 50.57 |
| *TBC1D17* | rs7248744 | A | G | 0.4253 | 0.414 | 0.007 | 1E-200 | 3497.88 |
| *TBC1D23* | rs1036770 | C | T | 0.5847 | -0.154 | 0.008 | 2.5119E-90 | 370.56 |
| *TBC1D23* | rs342298 | C | T | 0.4545 | -0.051 | 0.007 | 1E-11 | 53.08 |
| *TBC1D23* | rs6993770 | A | T | 0.2864 | -0.066 | 0.008 | 1.9953E-15 | 68.06 |
| *TBC1D23* | rs7090758 | T | C | 0.4765 | -0.051 | 0.007 | 6.3096E-12 | 53.08 |
| *TBC1D5* | rs1354034 | T | C | 0.6043 | 0.059 | 0.008 | 5.0119E-15 | 54.39 |
| *TBC1D5* | rs9971407 | C | T | 0.2424 | -0.067 | 0.009 | 1E-14 | 55.42 |
| *TBCA* | rs1354034 | T | C | 0.6044 | 0.07 | 0.008 | 5.0119E-20 | 76.56 |
| *TBCA* | rs254415 | G | T | 0.3049 | 0.14 | 0.008 | 5.0119E-65 | 306.25 |
| *TBCB* | rs1354034 | T | C | 0.6041 | 0.063 | 0.008 | 1.5849E-16 | 62.02 |
| *TBCB* | rs342298 | C | T | 0.4544 | -0.054 | 0.008 | 6.3096E-13 | 45.56 |
| *TBCB* | rs6993770 | A | T | 0.2865 | -0.063 | 0.008 | 3.9811E-14 | 62.02 |
| *TBCB* | rs7896518 | A | G | 0.4238 | 0.06 | 0.008 | 7.9433E-15 | 56.25 |
| *TBCC* | rs1354034 | T | C | 0.6041 | 0.061 | 0.008 | 1.9953E-15 | 58.14 |
| *TBCC* | rs2234026 | A | G | 0.9869 | 0.577 | 0.033 | 7.9433E-69 | 305.72 |
| *TBCC* | rs342298 | C | T | 0.4544 | -0.057 | 0.008 | 6.3096E-14 | 50.77 |
| *TBCC* | rs10822145 | C | T | 0.472 | 0.052 | 0.008 | 6.3096E-12 | 42.25 |
| *TBCC* | rs892090 | T | G | 0.8341 | 0.069 | 0.01 | 1.5849E-11 | 47.61 |
| *TBL1X* | rs1354034 | T | C | 0.6038 | -0.124 | 0.008 | 1.2589E-57 | 240.25 |
| *TBL1X* | rs11950562 | A | C | 0.4784 | -0.052 | 0.008 | 7.9433E-12 | 42.25 |
| *TBL1X* | rs139141690 | G | A | 0.0048 | -0.441 | 0.056 | 2.5119E-15 | 62.02 |
| *TBL1X* | rs6993770 | A | T | 0.2869 | -0.072 | 0.008 | 1E-17 | 81.00 |
| *TBL1X* | rs12767683 | C | A | 0.3249 | 0.081 | 0.008 | 7.9433E-24 | 102.52 |
| *TBL1X* | rs7297265 | A | G | 0.4426 | 0.067 | 0.008 | 3.9811E-18 | 70.14 |
| *TCL1A* | rs301816 | G | A | 0.5901 | -0.052 | 0.007 | 3.1623E-12 | 55.18 |
| *TCL1A* | rs17361097 | G | A | 0.4378 | 0.052 | 0.007 | 7.9433E-13 | 55.18 |
| *TCL1A* | rs10689049 | C | CCT | 0.3091 | 0.064 | 0.008 | 2.5119E-16 | 64.00 |
| *TCL1A* | rs77018303 | G | T | 0.0182 | -0.215 | 0.027 | 1.5849E-15 | 63.41 |
| *TCL1A* | rs17056278 | C | G | 0.0763 | -0.2 | 0.014 | 1.5849E-48 | 204.08 |
| *TCL1A* | rs34907473 | A | G | 0.277 | -0.078 | 0.008 | 1.5849E-21 | 95.06 |
| *TCL1A* | rs652951 | C | A | 0.4923 | -0.051 | 0.007 | 2.5119E-12 | 53.08 |
| *TCL1A* | rs9483788 | T | C | 0.257 | -0.074 | 0.008 | 6.3096E-19 | 85.56 |
| *TCL1A* | rs116968179 | G | A | 0.0331 | -0.146 | 0.021 | 1.2589E-12 | 48.34 |
| *TCL1A* | rs1433577 | G | A | 0.2742 | -0.084 | 0.008 | 5.0119E-25 | 110.25 |
| *TCL1A* | rs4948492 | C | T | 0.6591 | -0.074 | 0.008 | 3.1623E-22 | 85.56 |
| *TCL1A* | rs12784975 | T | C | 0.1918 | -0.078 | 0.009 | 2.5119E-17 | 75.11 |
| *TCL1A* | rs76428106 | T | C | 0.0129 | 0.229 | 0.033 | 5.0119E-12 | 48.16 |
| *TCL1A* | rs78986913 | G | A | 0.0432 | -0.714 | 0.018 | 1E-200 | 1573.44 |
| *TCL1A* | rs11853271 | G | A | 0.3221 | 0.06 | 0.008 | 5.0119E-15 | 56.25 |
| *TCL1A* | rs13330176 | T | A | 0.2389 | 0.088 | 0.009 | 5.0119E-25 | 95.60 |
| *TCL1A* | rs12947321 | A | G | 0.4927 | -0.062 | 0.007 | 1.2589E-17 | 78.45 |
| *TCL1A* | rs35222145 | T | G | 0.4921 | -0.102 | 0.007 | 6.3096E-44 | 212.33 |
| *TCL1A* | rs6065926 | A | G | 0.753 | -0.057 | 0.008 | 1.5849E-11 | 50.77 |
| *TCL1A* | rs75743846 | G | A | 0.0177 | -0.189 | 0.028 | 7.9433E-12 | 45.56 |
| *TCN1* | rs10208011 | T | C | 0.1324 | 0.082 | 0.01 | 5.0119E-17 | 67.24 |
| *TCN1* | rs4857909 | A | G | 0.8815 | 0.102 | 0.01 | 7.9433E-23 | 104.04 |
| *TCN1* | rs72667759 | C | G | 0.1927 | -0.058 | 0.008 | 6.3096E-12 | 52.56 |
| *TCN1* | rs849867 | A | G | 0.5177 | -0.055 | 0.007 | 1.5849E-16 | 61.73 |
| *TCN1* | rs11757137 | C | T | 0.4404 | 0.058 | 0.007 | 1E-16 | 68.65 |
| *TCN1* | rs9294380 | G | A | 0.5696 | -0.051 | 0.007 | 5.0119E-14 | 53.08 |
| *TCN1* | rs6796 | T | C | 0.2789 | 0.085 | 0.007 | 1.2589E-30 | 147.45 |
| *TCN1* | rs34324219 | C | A | 0.107 | -0.754 | 0.012 | 1E-200 | 3948.03 |
| *TCN1* | rs3967200 | C | T | 0.1349 | -0.108 | 0.011 | 1.5849E-23 | 96.40 |
| *TCN1* | rs8015478 | A | C | 0.7359 | 0.062 | 0.008 | 3.1623E-16 | 60.06 |
| *TCN1* | rs186021206 | G | A | 0.0064 | 0.659 | 0.046 | 5.0119E-47 | 205.24 |

| | | | | | | | | |
|---|---|---|---|---|---|---|---|---|
| *TCN1* | rs12941811 | T | C | 0.5741 | -0.061 | 0.007 | 7.9433E-20 | 75.94 |
| *TCN1* | rs17855739 | C | T | 0.0412 | 0.233 | 0.017 | 1.2589E-40 | 187.85 |
| *TCN1* | rs679574 | C | G | 0.5089 | 0.395 | 0.007 | 1E-200 | 3184.18 |
| *TCN2* | rs61729512 | G | A | 0.1177 | 0.064 | 0.01 | 1.5849E-11 | 40.96 |
| *TCN2* | rs150384171 | ACTC | A | 0.0135 | 1.239 | 0.027 | 1E-200 | 2105.79 |
| *TCN2* | rs12986064 | T | C | 0.5074 | -0.071 | 0.006 | 1E-27 | 140.03 |
| *TCN2* | rs740234 | A | G | 0.2041 | 0.633 | 0.009 | 1E-200 | 4946.78 |
| *TCOF1* | rs56180593 | C | T | 0.0051 | -0.549 | 0.052 | 1E-25 | 111.46 |
| *TCOF1* | rs7080536 | G | A | 0.0437 | -0.16 | 0.019 | 5.0119E-18 | 70.91 |
| *TCTN3* | rs78444298 | G | A | 0.0194 | 0.193 | 0.023 | 1.9953E-17 | 70.41 |
| *TCTN3* | rs7646106 | C | T | 0.1746 | 0.058 | 0.008 | 1.9953E-12 | 52.56 |
| *TCTN3* | rs35457250 | C | T | 0.0089 | 0.486 | 0.033 | 7.9433E-49 | 216.89 |
| *TCTN3* | rs28601761 | C | G | 0.4197 | -0.052 | 0.006 | 2.5119E-16 | 75.11 |
| *TCTN3* | rs61871796 | A | C | 0.3487 | 0.631 | 0.008 | 1E-200 | 6221.27 |
| *TCTN3* | rs774882452 | CT | C | 0.3446 | 0.053 | 0.007 | 3.1623E-16 | 57.33 |
| *TCTN3* | rs10843403 | T | C | 0.6097 | 0.053 | 0.006 | 1E-16 | 78.03 |
| *TCTN3* | rs71308015 | A | AG | 0.4111 | -0.123 | 0.006 | 7.9433E-84 | 420.25 |
| *TCTN3* | rs28929474 | C | T | 0.0209 | 0.272 | 0.022 | 3.9811E-36 | 152.86 |
| *TCTN3* | rs56990244 | C | G | 0.5885 | -0.077 | 0.006 | 3.9811E-34 | 164.69 |
| *TCTN3* | rs12599863 | A | G | 0.3089 | 0.061 | 0.007 | 1.9953E-19 | 75.94 |
| *TCTN3* | rs186021206 | G | A | 0.0064 | 0.524 | 0.043 | 1.9953E-34 | 148.50 |
| *TCTN3* | rs1801689 | A | C | 0.0301 | 0.278 | 0.018 | 5.0119E-53 | 238.53 |
| *TCTN3* | rs112001035 | G | A | 0.061 | 0.169 | 0.013 | 3.9811E-37 | 169.00 |
| *TCTN3* | rs429358 | T | C | 0.1553 | -0.09 | 0.009 | 7.9433E-26 | 100.00 |
| *TDGF1* | rs112481213 | T | A | 0.2403 | 1.319 | 0.009 | 1E-200 | 21478.53 |
| *TDGF1* | rs113354603 | G | A | 0.0699 | -0.07 | 0.01 | 6.3096E-12 | 49.00 |
| *TDGF1* | rs507666 | G | A | 0.184 | -0.052 | 0.006 | 3.9811E-16 | 75.11 |
| *TDP1* | rs9268833 | C | T | 0.3172 | 0.068 | 0.008 | 7.9433E-17 | 72.25 |
| *TDP1* | rs79027981 | A | G | 0.0173 | 0.307 | 0.029 | 1.9953E-26 | 112.07 |
| *TDP1* | rs10147250 | T | C | 0.4474 | -0.056 | 0.008 | 1.9953E-12 | 49.00 |
| *TDRKH* | rs4845556 | G | A | 0.4812 | 0.314 | 0.008 | 1E-200 | 1540.56 |
| *TDRKH* | rs1354034 | T | C | 0.6035 | 0.057 | 0.008 | 6.3096E-14 | 50.77 |
| *TDRKH* | rs6993770 | A | T | 0.2869 | -0.069 | 0.008 | 3.9811E-17 | 74.39 |
| *TDRKH* | rs7896518 | A | G | 0.4244 | 0.055 | 0.008 | 2.5119E-13 | 47.27 |
| *TEF* | rs4822027 | G | A | 0.2098 | 0.084 | 0.01 | 6.3096E-19 | 70.56 |
| *TEK* | rs61801010 | A | G | 0.1061 | 0.08 | 0.011 | 6.3096E-13 | 52.89 |
| *TEK* | rs10935473 | G | T | 0.44 | -0.072 | 0.007 | 6.3096E-28 | 105.80 |
| *TEK* | rs13107325 | C | T | 0.0749 | 0.117 | 0.012 | 3.1623E-21 | 95.06 |
| *TEK* | rs1050518 | T | A | 0.298 | -0.223 | 0.007 | 7.943E-197 | 1014.88 |
| *TEK* | rs2134964 | A | G | 0.7001 | -0.105 | 0.007 | 1.9953E-49 | 225.00 |
| *TEK* | rs682632 | A | C | 0.9602 | -1.045 | 0.019 | 1E-200 | 3025.00 |
| *TEK* | rs8176759 | G | A | 0.0613 | 0.811 | 0.015 | 1E-200 | 2923.20 |
| *TEK* | rs174530 | A | G | 0.3728 | 0.045 | 0.007 | 1.5849E-11 | 41.33 |
| *TEK* | rs35166255 | G | A | 0.0338 | 0.286 | 0.018 | 5.0119E-57 | 252.46 |
| *TEK* | rs1042704 | G | A | 0.2155 | -0.097 | 0.008 | 1.2589E-34 | 147.02 |
| *TEK* | rs186021206 | G | A | 0.0064 | 0.479 | 0.045 | 6.3096E-27 | 113.30 |
| *TET2* | rs2516886 | C | T | 0.8585 | -0.078 | 0.011 | 1.9953E-12 | 50.28 |
| *TEX101* | rs35033974 | G | T | 0.1285 | -0.332 | 0.008 | 1E-200 | 1722.25 |
| *TF* | rs13008704 | C | T | 0.5055 | 0.054 | 0.007 | 7.9433E-14 | 59.51 |
| *TF* | rs8177247 | T | G | 0.3422 | 0.329 | 0.008 | 1E-200 | 1691.27 |
| *TF* | rs79220007 | T | C | 0.0772 | -0.435 | 0.014 | 1E-200 | 965.43 |
| *TF* | rs174576 | C | A | 0.3524 | 0.051 | 0.008 | 1.2589E-11 | 40.64 |
| *TFF1* | rs760077 | A | T | 0.6052 | -0.095 | 0.007 | 5.0119E-39 | 184.18 |
| *TFF1* | rs62133344 | C | A | 0.056 | 0.409 | 0.015 | 2.512E-154 | 743.47 |
| *TFF1* | rs2075842 | G | C | 0.4163 | 0.072 | 0.007 | 5.0119E-24 | 105.80 |
| *TFF1* | rs536255 | C | T | 0.1912 | -0.081 | 0.009 | 1.2589E-18 | 81.00 |
| *TFF1* | rs3761376 | G | A | 0.2355 | -0.304 | 0.009 | 1E-200 | 1140.94 |
| *TFF2* | rs760077 | A | T | 0.6049 | -0.099 | 0.007 | 2.5119E-41 | 200.02 |
| *TFF2* | rs11379990 | A | AT | 0.5019 | -0.087 | 0.007 | 5.0119E-34 | 154.47 |
| *TFF2* | rs2978981 | C | T | 0.4407 | -0.052 | 0.007 | 3.1623E-13 | 55.18 |
| *TFF2* | rs2075842 | G | C | 0.4163 | 0.114 | 0.007 | 1.9953E-54 | 265.22 |
| *TFF2* | rs2217142 | C | T | 0.5823 | -0.075 | 0.007 | 1.5849E-24 | 114.80 |
| *TFF2* | rs186021206 | G | A | 0.0064 | 0.715 | 0.049 | 7.9433E-48 | 212.92 |
| *TFF2* | rs601338 | G | A | 0.5097 | -0.05 | 0.007 | 3.9811E-12 | 51.02 |
| *TFF2* | rs1534081 | A | G | 0.7476 | 0.07 | 0.008 | 5.0119E-17 | 76.56 |
| *TFF2* | rs225344 | G | A | 0.7265 | 0.125 | 0.008 | 3.1623E-54 | 244.14 |
| *TFF3* | rs10023335 | T | C | 0.4108 | 0.05 | 0.007 | 5.0119E-12 | 51.02 |
| *TFF3* | rs2524277 | G | A | 0.0586 | 0.614 | 0.016 | 1E-200 | 1472.64 |

| | | | | | | | | |
|---|---|---|---|---|---|---|---|---|
| *TFF3* | rs118095917 | C | T | 0.0062 | -1.277 | 0.046 | 1.259E-172 | 770.67 |
| *TFPI* | rs12127364 | C | A | 0.2711 | 0.145 | 0.008 | 1.5849E-74 | 328.52 |
| *TFPI* | rs1260326 | T | C | 0.6082 | -0.054 | 0.007 | 2.5119E-13 | 59.51 |
| *TFPI* | rs7576066 | G | A | 0.2941 | 0.347 | 0.008 | 1E-200 | 1881.39 |
| *TFPI* | rs562281690 | G | T | 0.0027 | -0.486 | 0.071 | 6.3096E-12 | 46.85 |
| *TFPI* | rs528128538 | C | T | 0.003 | -0.449 | 0.065 | 3.1623E-12 | 47.72 |
| *TFPI* | rs28601761 | C | G | 0.4192 | -0.06 | 0.007 | 5.0119E-17 | 73.47 |
| *TFPI* | rs1111796 | A | T | 0.0755 | -0.172 | 0.014 | 1.5849E-35 | 150.94 |
| *TFPI* | rs7412 | C | T | 0.0816 | -0.153 | 0.013 | 1E-32 | 138.51 |
| *TFPI2* | rs61804208 | G | T | 0.1048 | 0.137 | 0.012 | 5.0119E-32 | 130.34 |
| *TFPI2* | rs68066031 | T | C | 0.2299 | 0.07 | 0.008 | 1.9953E-16 | 76.56 |
| *TFPI2* | rs6768289 | G | A | 0.253 | -0.141 | 0.008 | 5.0119E-67 | 310.64 |
| *TFPI2* | rs5879368 | T | TA | 0.3258 | -0.057 | 0.008 | 1.9953E-13 | 50.77 |
| *TFPI2* | rs6924387 | A | G | 0.4062 | -0.06 | 0.007 | 1.5849E-16 | 73.47 |
| *TFPI2* | rs62466699 | T | C | 0.058 | 0.242 | 0.015 | 5.0119E-55 | 260.28 |
| *TFPI2* | rs10112582 | A | G | 0.2011 | 0.094 | 0.009 | 1.5849E-26 | 109.09 |
| *TFPI2* | rs35719208 | C | T | 0.1885 | 0.096 | 0.009 | 3.9811E-26 | 113.78 |
| *TFPI2* | rs5791751 | T | TA | 0.6732 | -0.085 | 0.008 | 6.3096E-29 | 112.89 |
| *TFPI2* | rs10842452 | C | A | 0.8828 | -0.138 | 0.011 | 1.5849E-34 | 157.39 |
| *TFPI2* | rs2160725 | A | C | 0.4847 | -0.058 | 0.007 | 3.1623E-16 | 68.65 |
| *TFRC* | rs766000481 | AAAC | A | 0.3558 | -0.054 | 0.007 | 2.5119E-13 | 59.51 |
| *TFRC* | rs3811647 | G | A | 0.3423 | -0.055 | 0.008 | 2.5119E-12 | 47.27 |
| *TFRC* | rs3817672 | C | T | 0.5633 | -0.524 | 0.008 | 1E-200 | 4290.25 |
| *TFRC* | rs252152 | A | G | 0.662 | 0.081 | 0.007 | 1.5849E-28 | 133.90 |
| *TFRC* | rs1800562 | G | A | 0.0778 | -0.277 | 0.013 | 1E-100 | 454.02 |
| *TFRC* | rs6592965 | G | A | 0.4543 | -0.063 | 0.007 | 7.9433E-20 | 81.00 |
| *TFRC* | rs3211931 | C | T | 0.4328 | -0.05 | 0.007 | 6.3096E-13 | 51.02 |
| *TFRC* | rs2075672 | A | G | 0.6232 | -0.067 | 0.007 | 3.1623E-21 | 91.61 |
| *TFRC* | rs550057 | C | T | 0.2552 | -0.102 | 0.008 | 5.0119E-38 | 162.56 |
| *TFRC* | rs560046267 | G | GT | 0.3109 | 0.054 | 0.007 | 5.0119E-13 | 59.51 |
| *TFRC* | rs652455 | C | T | 0.8514 | -0.193 | 0.01 | 6.3096E-86 | 372.49 |
| *TFRC* | rs74035509 | C | T | 0.0779 | 0.093 | 0.013 | 1E-12 | 51.18 |
| *TFRC* | rs59435462 | T | TA | 0.7583 | -0.062 | 0.008 | 1.2589E-14 | 60.06 |
| *TFRC* | rs2072860 | G | A | 0.534 | -0.139 | 0.007 | 5.0119E-89 | 394.31 |
| *TG* | rs334721 | A | C | 0.9603 | -0.171 | 0.019 | 1.9953E-19 | 81.00 |
| *TG* | rs17020146 | A | C | 0.0884 | 0.157 | 0.013 | 1.5849E-33 | 145.85 |
| *TG* | rs1690789 | C | T | 0.4865 | 0.084 | 0.007 | 5.0119E-30 | 144.00 |
| *TG* | rs3748916 | G | A | 0.4718 | 0.07 | 0.007 | 2.5119E-21 | 100.00 |
| *TG* | rs13015993 | G | A | 0.7324 | -0.07 | 0.008 | 7.9433E-17 | 76.56 |
| *TG* | rs4678938 | G | C | 0.4692 | -0.058 | 0.007 | 3.1623E-15 | 68.65 |
| *TG* | rs10519980 | C | T | 0.2039 | -0.064 | 0.009 | 1.2589E-12 | 50.57 |
| *TG* | rs66760320 | C | T | 0.2497 | 0.063 | 0.008 | 1.5849E-13 | 62.02 |
| *TG* | rs9497965 | C | T | 0.4089 | 0.085 | 0.007 | 6.3096E-30 | 147.45 |
| *TG* | rs17719705 | A | T | 0.4541 | 0.055 | 0.007 | 6.3096E-14 | 61.73 |
| *TG* | rs75143612 | G | C | 0.0282 | 0.246 | 0.022 | 1.5849E-28 | 125.03 |
| *TG* | rs10814915 | T | C | 0.5857 | 0.052 | 0.007 | 5.0119E-12 | 55.18 |
| *TG* | rs976575 | A | G | 0.3602 | -0.08 | 0.008 | 3.9811E-25 | 100.00 |
| *TG* | rs7128207 | G | T | 0.5596 | -0.067 | 0.007 | 2.5119E-19 | 91.61 |
| *TG* | rs11830537 | C | T | 0.0764 | 0.116 | 0.014 | 1.2589E-16 | 68.65 |
| *TG* | rs9507285 | C | G | 0.3477 | -0.054 | 0.008 | 3.9811E-12 | 45.56 |
| *TG* | rs4338740 | T | C | 0.2592 | 0.095 | 0.008 | 7.9433E-30 | 141.02 |
| *TG* | rs12148331 | C | T | 0.3204 | -0.054 | 0.008 | 1E-11 | 45.56 |
| *TG* | rs17689024 | G | C | 0.3134 | 0.095 | 0.008 | 6.3096E-33 | 141.02 |
| *TG* | rs1203948 | G | A | 0.7849 | -0.121 | 0.009 | 1.2589E-41 | 180.75 |
| *TGFA* | rs17639251 | T | C | 0.1193 | 0.175 | 0.012 | 3.1623E-51 | 212.67 |
| *TGFA* | rs7310615 | C | G | 0.5177 | -0.075 | 0.007 | 2.5119E-23 | 114.80 |
| *TGFA* | rs746056915 | TTC | T | 0.3927 | 0.086 | 0.008 | 3.9811E-29 | 115.56 |
| *TGFB1* | rs3804749 | C | T | 0.5928 | -0.072 | 0.008 | 2.5119E-21 | 81.00 |
| *TGFB1* | rs373385028 | A | T | 0.5516 | -0.087 | 0.007 | 3.9811E-32 | 154.47 |
| *TGFB1* | rs6796 | T | C | 0.2792 | 0.06 | 0.008 | 2.5119E-13 | 56.25 |
| *TGFB1* | rs2544221 | G | C | 0.4624 | 0.066 | 0.007 | 6.3096E-19 | 88.90 |
| *TGFB1* | rs6993770 | A | T | 0.2866 | -0.139 | 0.008 | 2.5119E-65 | 301.89 |
| *TGFB1* | rs2519093 | C | T | 0.1844 | -0.11 | 0.01 | 1E-30 | 121.00 |
| *TGFB1* | rs7136223 | A | G | 0.2754 | 0.079 | 0.008 | 7.9433E-22 | 97.52 |
| *TGFB1* | rs73045269 | C | T | 0.1715 | 0.288 | 0.01 | 3.162E-187 | 829.44 |
| *TGFB1* | rs892090 | T | G | 0.8342 | 0.117 | 0.01 | 1.2589E-31 | 136.89 |
| *TGFB2* | rs7538934 | C | T | 0.1187 | 0.197 | 0.012 | 3.9811E-62 | 269.51 |
| *TGFBI* | rs2493134 | T | C | 0.3993 | -0.257 | 0.007 | 1E-200 | 1347.94 |

| | | | | | | | | |
|---|---|---|---|---|---|---|---|---|
| *TGFBI* | rs11127048 | G | A | 0.6177 | -0.064 | 0.007 | 6.3096E-20 | 83.59 |
| *TGFBI* | rs13159365 | C | T | 0.5103 | -0.546 | 0.007 | 1E-200 | 6084.00 |
| *TGFBI* | rs9268812 | A | G | 0.6583 | 0.069 | 0.007 | 3.1623E-22 | 97.16 |
| *TGFBI* | rs10645152 | G | GTT | 0.745 | -0.114 | 0.008 | 7.9433E-50 | 203.06 |
| *TGFBI* | rs28601761 | C | G | 0.419 | -0.061 | 0.007 | 5.0119E-19 | 75.94 |
| *TGFBI* | rs10982156 | T | A | 0.0695 | -0.445 | 0.014 | 1E-200 | 1010.33 |
| *TGFBI* | rs56278466 | T | G | 0.6609 | 0.056 | 0.007 | 1E-15 | 64.00 |
| *TGFBI* | rs28929474 | C | T | 0.0209 | 0.192 | 0.023 | 1.9953E-16 | 69.69 |
| *TGFBI* | rs116683462 | A | T | 0.5425 | 0.074 | 0.007 | 1E-28 | 111.76 |
| *TGFBI* | rs12975366 | T | C | 0.3948 | -0.064 | 0.007 | 5.0119E-21 | 83.59 |
| *TGFBR2* | rs61747728 | C | T | 0.0379 | 0.192 | 0.019 | 5.0119E-25 | 102.12 |
| *TGFBR2* | rs1260326 | T | C | 0.6087 | 0.056 | 0.007 | 3.1623E-14 | 64.00 |
| *TGFBR2* | rs75166367 | G | A | 0.0604 | -0.103 | 0.015 | 6.3096E-12 | 47.15 |
| *TGFBR2* | rs56388355 | A | AG | 0.4025 | 0.051 | 0.008 | 1.2589E-11 | 40.64 |
| *TGFBR2* | rs3129887 | G | A | 0.1883 | 0.089 | 0.009 | 1.5849E-22 | 97.79 |
| *TGFBR2* | rs10265221 | T | C | 0.287 | 0.053 | 0.008 | 1E-11 | 43.89 |
| *TGFBR2* | rs507666 | G | A | 0.1844 | -0.17 | 0.009 | 3.9811E-75 | 356.79 |
| *TGFBR2* | rs2837988 | C | A | 0.3616 | 0.051 | 0.007 | 7.9433E-12 | 53.08 |
| *TGFBR3* | rs11207423 | G | C | 0.2766 | 0.1 | 0.008 | 2.5119E-33 | 156.25 |
| *TGFBR3* | rs2634023 | C | T | 0.0802 | -0.231 | 0.014 | 1.9953E-64 | 272.25 |
| *TGFBR3* | rs150816167 | T | C | 0.0419 | 0.144 | 0.019 | 1.2589E-14 | 57.44 |
| *TGFBR3* | rs71640034 | G | A | 0.5139 | -0.076 | 0.007 | 6.3096E-25 | 117.88 |
| *TGFBR3* | rs181242111 | G | A | 0.118 | 0.098 | 0.012 | 1.2589E-15 | 66.69 |
| *TGFBR3* | rs77924615 | G | A | 0.1969 | -0.072 | 0.009 | 1.5849E-14 | 64.00 |
| *TGFBR3* | rs1236213 | G | T | 0.593 | -0.159 | 0.008 | 3.1623E-97 | 395.02 |
| *TGM2* | rs9272445 | C | T | 0.1309 | -0.116 | 0.011 | 3.1623E-25 | 111.21 |
| *TGM2* | rs6592965 | G | A | 0.4538 | 0.062 | 0.007 | 1.2589E-17 | 78.45 |
| *TGM2* | rs771133280 | TC | T | 0.202 | 0.073 | 0.009 | 1.5849E-15 | 65.79 |
| *TGM2* | rs2076382 | C | T | 0.6295 | 0.169 | 0.007 | 3.162E-113 | 582.88 |
| *TGOLN2* | rs629301 | G | T | 0.7777 | -0.061 | 0.008 | 3.9811E-15 | 58.14 |
| *TGOLN2* | rs61747728 | C | T | 0.038 | 0.182 | 0.017 | 1.2589E-27 | 114.62 |
| *TGOLN2* | rs4247303 | G | A | 0.4821 | -0.715 | 0.007 | 1E-200 | 10433.16 |
| *TGOLN2* | rs169504 | C | A | 0.1855 | 0.062 | 0.008 | 1.2589E-13 | 60.06 |
| *TGOLN2* | rs4997081 | G | C | 0.1925 | -0.066 | 0.008 | 7.9433E-16 | 68.06 |
| *TGOLN2* | rs3814995 | C | T | 0.3149 | 0.057 | 0.007 | 2.5119E-16 | 66.31 |
| *THAP12* | rs2516980 | A | C | 0.7802 | 0.125 | 0.011 | 3.1623E-32 | 129.13 |
| *THBD* | rs1337526 | A | G | 0.8032 | 0.067 | 0.009 | 7.9433E-14 | 55.42 |
| *THBD* | rs61747728 | C | T | 0.038 | 0.173 | 0.019 | 1E-20 | 82.91 |
| *THBD* | rs1260326 | T | C | 0.6083 | 0.052 | 0.007 | 1.2589E-12 | 55.18 |
| *THBD* | rs75166367 | G | A | 0.0604 | -0.113 | 0.015 | 3.1623E-14 | 56.75 |
| *THBD* | rs638333 | T | C | 0.29 | 0.071 | 0.008 | 7.9433E-19 | 78.77 |
| *THBD* | rs9264666 | G | C | 0.5545 | -0.061 | 0.007 | 1.2589E-17 | 75.94 |
| *THBD* | rs8176743 | C | T | 0.0612 | 0.329 | 0.015 | 6.31E-108 | 481.07 |
| *THBD* | rs34434834 | G | A | 0.0334 | 0.253 | 0.02 | 3.9811E-37 | 160.02 |
| *THBD* | rs8038694 | G | A | 0.2195 | 0.063 | 0.009 | 3.1623E-13 | 49.00 |
| *THBD* | rs77924615 | G | A | 0.1969 | -0.077 | 0.009 | 2.5119E-17 | 73.20 |
| *THBD* | rs186021206 | G | A | 0.0064 | 0.398 | 0.049 | 3.1623E-16 | 65.97 |
| *THBD* | rs2659005 | C | T | 0.4411 | 0.068 | 0.007 | 3.9811E-21 | 94.37 |
| *THBD* | rs33950747 | C | T | 0.075 | 0.144 | 0.014 | 1.5849E-26 | 105.80 |
| *THBD* | rs1042579 | G | A | 0.1915 | 0.201 | 0.009 | 3.981E-107 | 498.78 |
| *THBS2* | rs2271708 | T | C | 0.006 | -0.343 | 0.046 | 7.9433E-14 | 55.60 |
| *THBS2* | rs78109196 | C | A | 0.0972 | 0.661 | 0.013 | 1E-200 | 2585.33 |
| *THBS2* | rs3967200 | C | T | 0.1351 | -0.118 | 0.01 | 1.5849E-29 | 139.24 |
| *THBS2* | rs531745895 | G | GT | 0.3409 | -0.055 | 0.007 | 1.2589E-13 | 61.73 |
| *THBS2* | rs186021206 | G | A | 0.0064 | 0.56 | 0.049 | 1.5849E-30 | 130.61 |
| *THBS2* | rs146385050 | C | A | 0.1835 | 0.233 | 0.009 | 1.259E-139 | 670.23 |
| *THBS2* | rs2111504 | T | A | 0.1724 | 0.144 | 0.009 | 1.9953E-53 | 256.00 |
| *THBS2* | rs738409 | C | G | 0.2168 | 0.07 | 0.009 | 3.9811E-16 | 60.49 |
| *THBS4* | rs2438632 | G | T | 0.3983 | 0.244 | 0.008 | 1E-200 | 930.25 |
| *THBS4* | rs2881756 | C | T | 0.4105 | 0.074 | 0.007 | 6.3096E-23 | 111.76 |
| *THBS4* | rs8176719 | T | TC | 0.34 | 0.053 | 0.008 | 3.1623E-12 | 43.89 |
| *THBS4* | rs56278466 | T | G | 0.661 | 0.093 | 0.008 | 1.2589E-33 | 135.14 |
| *THBS4* | rs112771035 | C | G | 0.0713 | -0.133 | 0.014 | 6.3096E-21 | 90.25 |
| *THBS4* | rs186021206 | G | A | 0.0064 | 1.326 | 0.05 | 3.981E-154 | 703.31 |
| *THOP1* | rs2741990 | C | T | 0.2932 | 0.268 | 0.008 | 1E-200 | 1122.25 |
| *THPO* | rs540210247 | C | CA | 0.5265 | -0.063 | 0.008 | 1.2589E-15 | 62.02 |
| *THPO* | rs113926195 | G | GCC | 0.2304 | -0.067 | 0.009 | 1E-13 | 55.42 |
| *THPO* | rs3792366 | G | A | 0.593 | -0.052 | 0.008 | 1.2589E-11 | 42.25 |

| | | | | | | | | |
|---|---|---|---|---|---|---|---|---|
| *THPO* | rs6141 | C | T | 0.532 | 0.062 | 0.008 | 2.5119E-16 | 60.06 |
| *THPO* | rs11242109 | G | T | 0.4815 | -0.06 | 0.008 | 1.5849E-15 | 56.25 |
| *THPO* | rs56255857 | T | C | 0.2449 | -0.123 | 0.009 | 5.0119E-45 | 186.78 |
| *THPO* | rs6993770 | A | T | 0.2867 | -0.128 | 0.008 | 3.1623E-53 | 256.00 |
| *THPO* | rs774510679 | ACTTTGCC | A | 0.4121 | 0.078 | 0.008 | 3.1623E-24 | 95.06 |
| *THPO* | rs10762489 | A | G | 0.2233 | 0.068 | 0.009 | 3.9811E-14 | 57.09 |
| *THPO* | rs56385468 | T | G | 0.0867 | -0.212 | 0.013 | 1E-56 | 265.94 |
| *THPO* | rs1362213 | C | T | 0.4795 | -0.065 | 0.008 | 1.9953E-17 | 66.02 |
| *THPO* | rs2015599 | G | A | 0.459 | -0.053 | 0.008 | 1.9953E-12 | 43.89 |
| *THPO* | rs6580981 | G | A | 0.4584 | 0.058 | 0.008 | 3.1623E-14 | 52.56 |
| *THPO* | rs28929474 | C | T | 0.0208 | 0.36 | 0.026 | 1.9953E-42 | 191.72 |
| *THPO* | rs151234 | G | C | 0.1291 | 0.083 | 0.011 | 1.5849E-13 | 56.93 |
| *THPO* | rs12445050 | C | T | 0.138 | 0.076 | 0.011 | 3.9811E-12 | 47.74 |
| *THPO* | rs149394327 | G | C | 0.0301 | 0.157 | 0.022 | 1E-12 | 50.93 |
| *THPO* | rs1654425 | T | C | 0.8341 | 0.102 | 0.01 | 1E-23 | 104.04 |
| *THSD1* | rs2519093 | C | T | 0.1847 | -0.202 | 0.01 | 1.5849E-89 | 408.04 |
| *THSD1* | rs149590732 | A | G | 0.0038 | 1.111 | 0.063 | 1.2589E-70 | 310.99 |
| *THTPA* | rs1354034 | T | C | 0.6044 | 0.063 | 0.007 | 6.3096E-17 | 81.00 |
| *THTPA* | rs342298 | C | T | 0.4552 | -0.057 | 0.007 | 1.5849E-14 | 66.31 |
| *THTPA* | rs28403575 | C | T | 0.0885 | -0.75 | 0.013 | 1E-200 | 3328.40 |
| *THY1* | rs603424 | G | A | 0.1665 | -0.081 | 0.01 | 7.9433E-17 | 65.61 |
| *THY1* | rs55933700 | G | T | 0.0247 | -1.094 | 0.025 | 1E-200 | 1914.94 |
| *THY1* | rs186021206 | G | A | 0.0064 | 0.35 | 0.049 | 1.5849E-12 | 51.02 |
| *TIA1* | rs1354034 | T | C | 0.604 | 0.085 | 0.008 | 5.0119E-28 | 112.89 |
| *TIA1* | rs11502185 | T | C | 0.2599 | 0.076 | 0.01 | 7.9433E-15 | 57.76 |
| *TIA1* | rs60822569 | T | TTTCTC | 0.5545 | 0.058 | 0.008 | 5.0119E-14 | 52.56 |
| *TIE1* | rs4660729 | C | G | 0.6088 | 0.191 | 0.007 | 5.012E-154 | 744.51 |
| *TIE1* | rs10935473 | G | T | 0.4399 | -0.222 | 0.007 | 1E-200 | 1005.80 |
| *TIE1* | rs2962006 | G | C | 0.3873 | -0.051 | 0.007 | 1.9953E-12 | 53.08 |
| *TIE1* | rs9271325 | C | G | 0.4718 | -0.054 | 0.007 | 3.1623E-14 | 59.51 |
| *TIE1* | rs4841133 | A | G | 0.908 | -0.096 | 0.012 | 2.5119E-15 | 64.00 |
| *TIE1* | rs8176743 | C | T | 0.0614 | 0.897 | 0.015 | 1E-200 | 3576.04 |
| *TIE1* | rs56278466 | T | G | 0.6614 | 0.065 | 0.007 | 1.2589E-18 | 86.22 |
| *TIE1* | rs174564 | A | G | 0.3487 | 0.059 | 0.007 | 7.9433E-16 | 71.04 |
| *TIE1* | rs34434834 | G | A | 0.0334 | 0.164 | 0.02 | 6.3096E-17 | 67.24 |
| *TIE1* | rs878381 | C | T | 0.8576 | -0.143 | 0.01 | 1E-45 | 204.49 |
| *TIE1* | rs186021206 | G | A | 0.0064 | 0.52 | 0.048 | 5.0119E-27 | 117.36 |
| *TIE1* | rs4015 | C | T | 0.1955 | -0.067 | 0.009 | 1.9953E-14 | 55.42 |
| *TIGAR* | rs3217860 | A | G | 0.2477 | -0.066 | 0.009 | 1.2589E-13 | 53.78 |
| *TIGAR* | rs4632248 | G | T | 0.2095 | 0.075 | 0.01 | 3.9811E-15 | 56.25 |
| *TIGIT* | rs6792290 | C | G | 0.2895 | 0.095 | 0.008 | 2.5119E-29 | 141.02 |
| *TIGIT* | rs111338191 | A | AAATT | 0.5206 | -0.056 | 0.008 | 3.9811E-13 | 49.00 |
| *TIMD4* | rs1060622 | G | A | 0.6214 | 0.07 | 0.007 | 6.3096E-22 | 100.00 |
| *TIMD4* | rs1878051 | T | A | 0.3313 | -0.055 | 0.008 | 3.9811E-13 | 47.27 |
| *TIMD4* | rs10189685 | G | A | 0.2962 | 0.123 | 0.008 | 1.5849E-56 | 236.39 |
| *TIMD4* | rs11395592 | A | AT | 0.4271 | -0.05 | 0.007 | 2.5119E-12 | 51.02 |
| *TIMD4* | rs371216559 | C | CT | 0.2659 | -0.068 | 0.008 | 2.5119E-17 | 72.25 |
| *TIMD4* | rs115216147 | T | A | 0.0665 | 0.11 | 0.015 | 7.9433E-14 | 53.78 |
| *TIMD4* | rs38029 | C | G | 0.5606 | -0.065 | 0.007 | 1.5849E-19 | 86.22 |
| *TIMD4* | rs4704826 | C | A | 0.6384 | -0.232 | 0.007 | 1E-200 | 1098.45 |
| *TIMD4* | rs9270270 | T | C | 0.8398 | -0.149 | 0.01 | 1.2589E-49 | 222.01 |
| *TIMD4* | rs10456852 | C | T | 0.1307 | 0.105 | 0.011 | 5.0119E-23 | 91.12 |
| *TIMD4* | rs2737245 | G | T | 0.2821 | 0.056 | 0.008 | 1.9953E-12 | 49.00 |
| *TIMD4* | rs2142306 | T | C | 0.4118 | 0.063 | 0.007 | 2.5119E-18 | 81.00 |
| *TIMD4* | rs550057 | C | T | 0.2552 | 0.097 | 0.008 | 7.9433E-33 | 147.02 |
| *TIMD4* | rs2068888 | G | A | 0.4496 | 0.061 | 0.007 | 1.2589E-17 | 75.94 |
| *TIMD4* | rs72823014 | G | A | 0.1269 | 0.083 | 0.011 | 5.0119E-15 | 56.93 |
| *TIMD4* | rs10769256 | C | T | 0.4002 | -0.063 | 0.007 | 1.5849E-17 | 81.00 |
| *TIMD4* | rs112771035 | C | G | 0.0714 | 0.495 | 0.014 | 1E-200 | 1250.13 |
| *TIMD4* | rs34623398 | C | CAACT | 0.2757 | 0.083 | 0.008 | 2.5119E-25 | 107.64 |
| *TIMD4* | rs10778118 | C | T | 0.7893 | -0.072 | 0.009 | 1.5849E-16 | 64.00 |
| *TIMD4* | rs3764261 | C | A | 0.325 | 0.055 | 0.008 | 3.9811E-13 | 47.27 |
| *TIMD4* | rs34417180 | C | A | 0.1683 | -0.065 | 0.009 | 5.0119E-12 | 52.16 |
| *TIMD4* | rs4843861 | A | T | 0.2695 | -0.074 | 0.008 | 3.9811E-20 | 85.56 |
| *TIMD4* | rs186021206 | G | A | 0.0064 | 1.131 | 0.049 | 3.162E-119 | 532.76 |
| *TIMD4* | rs601338 | G | A | 0.5094 | 0.049 | 0.007 | 3.1623E-12 | 49.00 |
| *TIMD4* | rs2143876 | T | C | 0.1364 | 0.09 | 0.01 | 3.9811E-18 | 81.00 |
| *TIMD4* | rs66817580 | G | T | 0.1527 | 0.158 | 0.01 | 6.3096E-58 | 249.64 |

| | | | | | | | | |
|---|---|---|---|---|---|---|---|---|
| *TIMM10* | rs1354034 | T | C | 0.6045 | 0.063 | 0.008 | 2.5119E-16 | 62.02 |
| *TIMM10* | rs2848626 | C | T | 0.7672 | -0.215 | 0.009 | 7.943E-128 | 570.68 |
| *TIMM8A* | rs6993770 | A | T | 0.2865 | -0.059 | 0.008 | 1E-12 | 54.39 |
| *TIMP1* | rs1775815 | G | T | 0.5713 | 0.066 | 0.008 | 1.5849E-18 | 68.06 |
| *TIMP1* | rs12407843 | G | A | 0.1102 | 0.098 | 0.012 | 1.2589E-16 | 66.69 |
| *TIMP1* | rs9380143 | C | T | 0.2842 | -0.096 | 0.008 | 1.5849E-31 | 144.00 |
| *TIMP1* | rs6993770 | A | T | 0.2866 | -0.115 | 0.008 | 5.0119E-45 | 206.64 |
| *TIMP1* | rs1654425 | T | C | 0.8344 | 0.113 | 0.01 | 1.5849E-29 | 127.69 |
| *TIMP2* | rs76685449 | T | G | 0.0227 | 0.18 | 0.025 | 1E-12 | 51.84 |
| *TIMP2* | rs75314265 | T | C | 0.0131 | 0.508 | 0.033 | 1.9953E-52 | 236.97 |
| *TIMP2* | rs8066695 | T | C | 0.4546 | 0.093 | 0.008 | 1E-34 | 135.14 |
| *TIMP3* | rs2274319 | T | C | 0.6532 | -0.063 | 0.007 | 7.9433E-19 | 81.00 |
| *TIMP3* | rs13412535 | G | A | 0.231 | -0.057 | 0.008 | 7.9433E-13 | 50.77 |
| *TIMP3* | rs1354034 | T | C | 0.6041 | -0.047 | 0.007 | 6.3096E-12 | 45.08 |
| *TIMP3* | rs11734099 | G | A | 0.1742 | 0.077 | 0.009 | 3.1623E-18 | 73.20 |
| *TIMP3* | rs755492124 | TAAAG | T | 0.3935 | -0.05 | 0.007 | 2.5119E-13 | 51.02 |
| *TIMP3* | rs114694170 | T | C | 0.0601 | 0.173 | 0.014 | 7.9433E-34 | 152.70 |
| *TIMP3* | rs11953411 | G | A | 0.6055 | -0.061 | 0.007 | 1.5849E-18 | 75.94 |
| *TIMP3* | rs142985623 | G | GT | 0.1959 | -0.06 | 0.009 | 2.5119E-12 | 44.44 |
| *TIMP3* | rs6961069 | C | T | 0.4025 | 0.049 | 0.007 | 1.5849E-12 | 49.00 |
| *TIMP3* | rs61469632 | T | C | 0.0634 | -0.123 | 0.014 | 6.3096E-19 | 77.19 |
| *TIMP3* | rs7896518 | A | G | 0.4241 | 0.082 | 0.007 | 1.2589E-32 | 137.22 |
| *TIMP3* | rs17655730 | T | C | 0.2454 | 0.056 | 0.008 | 7.9433E-13 | 49.00 |
| *TIMP3* | rs1362213 | C | T | 0.4794 | -0.046 | 0.007 | 1.5849E-11 | 43.18 |
| *TIMP3* | rs12445050 | C | T | 0.138 | 0.086 | 0.01 | 6.3096E-19 | 73.96 |
| *TIMP3* | rs74035509 | C | T | 0.0779 | 0.09 | 0.013 | 1.2589E-12 | 47.93 |
| *TIMP3* | rs892090 | T | G | 0.8342 | 0.136 | 0.009 | 7.9433E-51 | 228.35 |
| *TIMP3* | rs6081569 | G | C | 0.3685 | 0.05 | 0.007 | 7.9433E-13 | 51.02 |
| *TIMP3* | rs5749504 | C | T | 0.7262 | -0.652 | 0.008 | 1E-200 | 6642.25 |
| *TIMP4* | rs78058190 | G | A | 0.0515 | -0.119 | 0.018 | 1.2589E-11 | 43.71 |
| *TIMP4* | rs2943649 | C | G | 0.6488 | -0.06 | 0.007 | 7.9433E-17 | 73.47 |
| *TIMP4* | rs184262 | A | G | 0.8281 | 0.447 | 0.01 | 1E-200 | 1998.09 |
| *TIMP4* | rs34579624 | G | A | 0.1456 | 0.088 | 0.011 | 6.3096E-17 | 64.00 |
| *TIMP4* | rs1358980 | C | T | 0.4802 | -0.058 | 0.007 | 5.0119E-17 | 68.65 |
| *TIMP4* | rs1716403 | T | C | 0.6798 | -0.053 | 0.007 | 6.3096E-13 | 57.33 |
| *TIMP4* | rs10405357 | T | C | 0.4357 | -0.048 | 0.007 | 1E-11 | 47.02 |
| *TINAGL1* | rs78265569 | C | A | 0.0888 | -0.089 | 0.013 | 7.9433E-12 | 46.87 |
| *TINAGL1* | rs2229474 | G | A | 0.0141 | 0.618 | 0.031 | 1E-87 | 397.42 |
| *TINAGL1* | rs945214 | A | G | 0.6137 | -0.085 | 0.008 | 2.5119E-28 | 112.89 |
| *TINAGL1* | rs7648959 | A | G | 0.2975 | 0.088 | 0.008 | 2.5119E-28 | 121.00 |
| *TINAGL1* | rs56278466 | T | G | 0.6613 | 0.062 | 0.008 | 6.3096E-16 | 60.06 |
| *TINAGL1* | rs7137828 | C | T | 0.5164 | -0.054 | 0.007 | 6.3096E-14 | 59.51 |
| *TINAGL1* | rs9583474 | A | C | 0.2105 | -0.077 | 0.009 | 6.3096E-18 | 73.20 |
| *TINAGL1* | rs8124907 | A | G | 0.2414 | -0.066 | 0.009 | 1E-14 | 53.78 |
| *TJAP1* | rs1354034 | T | C | 0.6041 | 0.062 | 0.007 | 1.2589E-16 | 78.45 |
| *TJAP1* | rs549884121 | C | CTA | 0.0063 | -0.324 | 0.047 | 3.9811E-12 | 47.52 |
| *TK1* | rs9697210 | G | A | 0.1464 | 0.091 | 0.011 | 7.9433E-17 | 68.44 |
| *TK1* | rs144204502 | C | T | 0.0128 | -0.35 | 0.035 | 3.9811E-24 | 100.00 |
| *TLR1* | rs5743618 | C | A | 0.2244 | 0.503 | 0.009 | 1E-200 | 3123.57 |
| *TLR1* | rs56278466 | T | G | 0.6612 | 0.068 | 0.008 | 1E-18 | 72.25 |
| *TLR3* | rs61804206 | A | G | 0.1051 | 0.041 | 0.006 | 1.5849E-11 | 46.69 |
| *TLR3* | rs3775291 | C | T | 0.2972 | -1.151 | 0.008 | 1E-200 | 20700.02 |
| *TLR3* | rs635634 | C | T | 0.1842 | -0.042 | 0.005 | 3.1623E-18 | 70.56 |
| *TLR3* | rs56278466 | T | G | 0.6613 | 0.042 | 0.004 | 1E-26 | 110.25 |
| *TLR3* | rs16940186 | T | C | 0.1486 | -0.036 | 0.005 | 6.3096E-12 | 51.84 |
| *TLR3* | rs4760 | A | G | 0.156 | 0.086 | 0.005 | 1.9953E-63 | 295.84 |
| *TLR3* | rs738409 | C | G | 0.2172 | 0.056 | 0.005 | 1.9953E-35 | 125.44 |
| *TLR4* | rs4986790 | A | G | 0.0597 | -0.327 | 0.016 | 7.9433E-90 | 417.69 |
| *TMCO5A* | rs516246 | C | T | 0.5086 | 0.055 | 0.008 | 1E-12 | 47.27 |
| *TMED8* | rs1354034 | T | C | 0.6046 | 0.052 | 0.008 | 1E-11 | 42.25 |
| *TMED8* | rs6993770 | A | T | 0.2866 | -0.057 | 0.008 | 5.0119E-12 | 50.77 |
| *TMED8* | rs11620921 | A | G | 0.6027 | -0.066 | 0.008 | 5.0119E-18 | 68.06 |
| *TMEM106A* | rs4950771 | T | G | 0.9343 | -0.198 | 0.015 | 3.1623E-39 | 174.24 |
| *TMEM106A* | rs9857570 | A | G | 0.3236 | -0.099 | 0.008 | 3.9811E-35 | 153.14 |
| *TMEM106A* | rs62443342 | A | T | 0.2485 | -0.083 | 0.009 | 5.0119E-22 | 85.05 |
| *TMEM106A* | rs1293303 | G | C | 0.4458 | -0.113 | 0.008 | 5.0119E-50 | 199.52 |
| *TMEM106A* | rs6993770 | A | T | 0.2864 | -0.075 | 0.008 | 1.2589E-19 | 87.89 |
| *TMEM106A* | rs10812628 | G | A | 0.2638 | -0.08 | 0.009 | 1E-20 | 79.01 |

| | | | | | | | | |
|---|---|---|---|---|---|---|---|---|
| *TMEM106A* | rs7080386 | C | A | 0.412 | 0.123 | 0.008 | 2.5119E-58 | 236.39 |
| *TMEM106A* | rs72701845 | G | A | 0.0377 | -0.384 | 0.02 | 1.5849E-79 | 368.64 |
| *TMEM106A* | rs34687590 | T | TG | 0.3854 | -0.107 | 0.008 | 1E-40 | 178.89 |
| *TMEM106A* | rs892090 | T | G | 0.8342 | 0.091 | 0.01 | 3.1623E-19 | 82.81 |
| *TMEM106A* | rs142598052 | T | C | 0.0064 | -0.367 | 0.048 | 2.5119E-14 | 58.46 |
| *TMEM132A* | rs11379371 | A | AG | 0.4011 | 0.613 | 0.008 | 1E-200 | 5871.39 |
| *TMEM25* | rs45558732 | G | T | 0.0337 | -1.02 | 0.021 | 1E-200 | 2359.18 |
| *TMEM25* | rs186021206 | G | A | 0.0064 | 0.382 | 0.051 | 5.0119E-14 | 56.10 |
| *TMPRSS11D* | rs148060334 | A | G | 0.0356 | -0.342 | 0.02 | 3.1623E-65 | 292.41 |
| *TMPRSS11D* | rs143325653 | A | AAAAGAAG | 0.4932 | -0.05 | 0.007 | 1.5849E-11 | 51.02 |
| *TMPRSS15* | rs11134475 | A | G | 0.6389 | 0.076 | 0.008 | 1.2589E-22 | 90.25 |
| *TMPRSS15* | rs113981974 | C | G | 0.1106 | 0.097 | 0.012 | 5.0119E-16 | 65.34 |
| *TMPRSS15* | rs550057 | C | T | 0.2553 | -0.208 | 0.009 | 2.512E-129 | 534.12 |
| *TMPRSS15* | rs708686 | C | T | 0.2673 | 0.114 | 0.008 | 2.5119E-41 | 203.06 |
| *TMPRSS15* | rs438811 | C | T | 0.2372 | 0.059 | 0.009 | 1.5849E-11 | 42.98 |
| *TMPRSS15* | rs492602 | A | G | 0.5088 | -0.182 | 0.008 | 6.31E-129 | 517.56 |
| *TMPRSS15* | rs2824805 | G | C | 0.1731 | 0.126 | 0.01 | 1E-37 | 158.76 |
| *TMPRSS5* | rs35383942 | C | T | 0.0618 | 0.132 | 0.013 | 1.9953E-25 | 103.10 |
| *TMPRSS5* | rs12024555 | G | A | 0.1946 | 0.054 | 0.008 | 1.9953E-12 | 45.56 |
| *TMPRSS5* | rs199927946 | TGTG | T | 0.6207 | 0.044 | 0.006 | 7.9433E-12 | 53.78 |
| *TMPRSS5* | rs1082409 | G | T | 0.3376 | -0.051 | 0.006 | 2.5119E-15 | 72.25 |
| *TMPRSS5* | rs67670470 | C | A | 0.3359 | 0.049 | 0.006 | 3.1623E-14 | 66.69 |
| *TMPRSS5* | rs4686658 | C | T | 0.7757 | -0.052 | 0.007 | 1.5849E-12 | 55.18 |
| *TMPRSS5* | rs60028714 | C | T | 0.6219 | -0.06 | 0.006 | 3.9811E-21 | 100.00 |
| *TMPRSS5* | rs9386298 | A | T | 0.4973 | 0.048 | 0.006 | 7.9433E-15 | 64.00 |
| *TMPRSS5* | rs116923669 | G | A | 0.0659 | -0.148 | 0.013 | 5.0119E-32 | 129.61 |
| *TMPRSS5* | rs56278466 | T | G | 0.6613 | 0.067 | 0.006 | 2.5119E-25 | 124.69 |
| *TMPRSS5* | rs10887793 | G | T | 0.4493 | 0.052 | 0.006 | 1.9953E-17 | 75.11 |
| *TMPRSS5* | rs7114195 | A | C | 0.6391 | 0.785 | 0.008 | 1E-200 | 9628.52 |
| *TMPRSS5* | rs68055275 | C | CT | 0.1448 | -0.086 | 0.011 | 2.5119E-15 | 61.12 |
| *TMPRSS5* | rs28404455 | T | C | 0.0929 | 0.084 | 0.01 | 1.2589E-15 | 70.56 |
| *TMPRSS5* | rs72681869 | G | C | 0.0111 | 0.209 | 0.029 | 7.9433E-13 | 51.94 |
| *TMPRSS5* | rs3742780 | G | C | 0.4759 | -0.052 | 0.006 | 1.9953E-17 | 75.11 |
| *TMPRSS5* | rs186021206 | G | A | 0.0064 | 0.555 | 0.042 | 3.9811E-40 | 174.62 |
| *TMPRSS5* | rs504549 | C | T | 0.3786 | -0.046 | 0.006 | 2.5119E-13 | 58.78 |
| *TMPRSS5* | rs2836683 | T | C | 0.338 | 0.045 | 0.006 | 1.9953E-12 | 56.25 |
| *TMSB10* | rs1354034 | T | C | 0.6041 | 0.055 | 0.008 | 5.0119E-13 | 47.27 |
| *TMSB10* | rs342293 | C | G | 0.4592 | -0.057 | 0.008 | 2.5119E-14 | 50.77 |
| *TMSB10* | rs6993770 | A | T | 0.2864 | -0.06 | 0.008 | 5.0119E-13 | 56.25 |
| *TMSB10* | rs11502185 | T | C | 0.2595 | 0.073 | 0.01 | 2.5119E-14 | 53.29 |
| *TMSB10* | rs10774624 | G | A | 0.5137 | -0.06 | 0.008 | 1.5849E-15 | 56.25 |
| *TMSB10* | rs4632248 | G | T | 0.209 | 0.13 | 0.009 | 3.1623E-44 | 208.64 |
| *TNC* | rs1892534 | C | T | 0.3745 | -0.051 | 0.007 | 1E-12 | 53.08 |
| *TNC* | rs2646260 | A | G | 0.2442 | -0.06 | 0.008 | 1.9953E-13 | 56.25 |
| *TNC* | rs4686846 | C | T | 0.2325 | 0.067 | 0.008 | 5.0119E-16 | 70.14 |
| *TNC* | rs13159365 | C | T | 0.5103 | 0.194 | 0.007 | 2.512E-167 | 768.08 |
| *TNC* | rs113760175 | G | A | 0.0707 | 0.104 | 0.014 | 3.9811E-14 | 55.18 |
| *TNC* | rs1990790 | C | T | 0.513 | -0.052 | 0.007 | 1.9953E-13 | 55.18 |
| *TNC* | rs2685412 | C | G | 0.2701 | 0.38 | 0.008 | 1E-200 | 2256.25 |
| *TNC* | rs4263799 | C | T | 0.4796 | 0.082 | 0.007 | 6.3096E-29 | 137.22 |
| *TNC* | rs112286501 | C | T | 0.0442 | 0.149 | 0.018 | 6.3096E-17 | 68.52 |
| *TNC* | rs7021057 | T | C | 0.2935 | -0.244 | 0.008 | 1E-200 | 930.25 |
| *TNC* | rs7949566 | G | A | 0.4214 | 0.08 | 0.007 | 1.5849E-29 | 130.61 |
| *TNC* | rs9668810 | T | C | 0.7351 | -0.076 | 0.008 | 5.0119E-22 | 90.25 |
| *TNC* | rs760715 | T | C | 0.2471 | 0.104 | 0.009 | 1.5849E-34 | 133.53 |
| *TNF* | rs4645843 | C | T | 0.0011 | -1.446 | 0.112 | 3.9811E-38 | 166.69 |
| *TNF* | rs7161799 | C | T | 0.0765 | 0.237 | 0.014 | 5.0119E-63 | 286.58 |
| *TNF* | rs3184504 | T | C | 0.517 | -0.106 | 0.007 | 7.9433E-46 | 229.31 |
| *TNFAIP2* | rs342293 | C | G | 0.4598 | -0.086 | 0.007 | 7.9433E-31 | 150.94 |
| *TNFAIP2* | rs12342201 | G | A | 0.4763 | -0.055 | 0.007 | 1.2589E-13 | 61.73 |
| *TNFAIP2* | rs576859558 | T | G | 0.0038 | -0.596 | 0.063 | 3.1623E-21 | 89.50 |
| *TNFAIP8* | rs1035376 | G | A | 0.8825 | 0.123 | 0.012 | 6.3096E-25 | 105.06 |
| *TNFAIP8L2* | rs4970991 | T | C | 0.793 | 0.066 | 0.009 | 1.2589E-12 | 53.78 |
| *TNFAIP8L2* | rs1354034 | T | C | 0.6044 | 0.065 | 0.008 | 2.5119E-17 | 66.02 |
| *TNFAIP8L2* | rs342299 | C | T | 0.4549 | -0.062 | 0.008 | 6.3096E-16 | 60.06 |
| *TNFRSF10A* | rs79287178 | G | A | 0.0301 | 0.249 | 0.02 | 6.3096E-35 | 155.00 |
| *TNFRSF10A* | rs1632984 | T | A | 0.8539 | 0.069 | 0.009 | 1.5849E-13 | 58.78 |
| *TNFRSF10A* | rs13278062 | G | T | 0.502 | -0.462 | 0.007 | 1E-200 | 4356.00 |

| | | | | | | | | |
|---|---|---|---|---|---|---|---|---|
| *TNFRSF10A* | rs1801689 | A | C | 0.0301 | 0.129 | 0.019 | 1.5849E-11 | 46.10 |
| *TNFRSF10A* | rs3072826 | GGAAA | G | 0.3639 | -0.048 | 0.007 | 1.5849E-12 | 47.02 |
| *TNFRSF10B* | rs231996 | C | A | 0.7601 | -0.06 | 0.008 | 1.2589E-14 | 56.25 |
| *TNFRSF10B* | rs1105944 | A | G | 0.8957 | 0.538 | 0.011 | 1E-200 | 2392.10 |
| *TNFRSF10B* | rs3184504 | T | C | 0.517 | -0.05 | 0.007 | 7.9433E-14 | 51.02 |
| *TNFRSF10C* | rs3917932 | C | G | 0.5756 | -0.045 | 0.006 | 3.1623E-14 | 56.25 |
| *TNFRSF10C* | rs3014874 | G | A | 0.2638 | -0.045 | 0.007 | 1.2589E-11 | 41.33 |
| *TNFRSF10C* | rs5030738 | G | T | 0.0171 | 0.2 | 0.023 | 1E-18 | 75.61 |
| *TNFRSF10C* | rs6723921 | A | G | 0.6132 | 0.047 | 0.006 | 5.0119E-15 | 61.36 |
| *TNFRSF10C* | rs10182318 | T | C | 0.8976 | -0.088 | 0.01 | 7.9433E-20 | 77.44 |
| *TNFRSF10C* | rs12497115 | C | T | 0.1754 | 0.053 | 0.008 | 5.0119E-12 | 43.89 |
| *TNFRSF10C* | rs3774315 | A | G | 0.2728 | 0.08 | 0.007 | 5.0119E-34 | 130.61 |
| *TNFRSF10C* | rs2249741 | A | C | 0.5038 | 0.056 | 0.006 | 5.0119E-22 | 87.11 |
| *TNFRSF10C* | rs149110519 | C | T | 0.0364 | 0.134 | 0.016 | 1.5849E-17 | 70.14 |
| *TNFRSF10C* | rs1010366 | T | C | 0.3174 | 0.063 | 0.006 | 5.0119E-24 | 110.25 |
| *TNFRSF10C* | rs445 | C | T | 0.0954 | -0.091 | 0.01 | 5.0119E-20 | 82.81 |
| *TNFRSF10C* | rs7846314 | A | T | 0.186 | 0.059 | 0.008 | 1.9953E-14 | 54.39 |
| *TNFRSF10C* | rs3124753 | G | A | 0.4724 | -0.068 | 0.006 | 1.5849E-31 | 128.44 |
| *TNFRSF10C* | rs2241244 | G | C | 0.537 | -0.091 | 0.006 | 6.3096E-55 | 230.03 |
| *TNFRSF10C* | rs4760 | A | G | 0.1563 | -1.185 | 0.01 | 1E-200 | 14042.25 |
| *TNFRSF11A* | rs550677346 | A | AT | 0.2007 | 0.058 | 0.008 | 3.1623E-12 | 52.56 |
| *TNFRSF11A* | rs9275587 | T | C | 0.4727 | -0.046 | 0.007 | 6.3096E-12 | 43.18 |
| *TNFRSF11A* | rs342293 | C | G | 0.4594 | 0.053 | 0.007 | 6.3096E-16 | 57.33 |
| *TNFRSF11A* | rs141129381 | T | TG | 0.3626 | 0.047 | 0.007 | 6.3096E-12 | 45.08 |
| *TNFRSF11A* | rs7080386 | C | A | 0.412 | 0.053 | 0.007 | 3.1623E-15 | 57.33 |
| *TNFRSF11A* | rs12946745 | G | C | 0.665 | -0.049 | 0.007 | 3.9811E-12 | 49.00 |
| *TNFRSF11A* | rs113339733 | G | A | 0.1927 | -0.598 | 0.009 | 1E-200 | 4414.86 |
| *TNFRSF11A* | rs601338 | G | A | 0.5089 | -0.122 | 0.007 | 2.5119E-74 | 303.76 |
| *TNFRSF11B* | rs114165349 | G | C | 0.0232 | -0.234 | 0.023 | 2.5119E-25 | 103.51 |
| *TNFRSF11B* | rs79287178 | G | A | 0.0301 | 0.205 | 0.021 | 6.3096E-23 | 95.29 |
| *TNFRSF11B* | rs748443516 | CT | C | 0.4996 | 0.2 | 0.007 | 3.981E-185 | 816.33 |
| *TNFRSF11B* | rs1013495 | C | T | 0.4549 | 0.056 | 0.007 | 3.1623E-16 | 64.00 |
| *TNFRSF11B* | rs7927191 | T | C | 0.2655 | -0.06 | 0.008 | 3.1623E-15 | 56.25 |
| *TNFRSF11B* | rs3825568 | C | T | 0.4601 | -0.047 | 0.007 | 2.5119E-12 | 45.08 |
| *TNFRSF11B* | rs28929474 | C | T | 0.0208 | 0.161 | 0.024 | 1E-11 | 45.00 |
| *TNFRSF11B* | rs200489612 | G | A | 0.0052 | 0.358 | 0.051 | 2.5119E-12 | 49.27 |
| *TNFRSF11B* | rs704 | G | A | 0.4734 | -0.198 | 0.007 | 3.981E-186 | 800.08 |
| *TNFRSF11B* | rs77542162 | A | G | 0.0229 | 0.157 | 0.023 | 6.3096E-12 | 46.60 |
| *TNFRSF11B* | rs3747207 | G | A | 0.2154 | 0.075 | 0.008 | 6.3096E-20 | 87.89 |
| *TNFRSF12A* | rs139130389 | C | CTA | 0.0726 | -0.283 | 0.014 | 7.9433E-97 | 408.62 |
| *TNFRSF12A* | rs964184 | G | C | 0.8667 | -0.078 | 0.01 | 6.3096E-14 | 60.84 |
| *TNFRSF12A* | rs74758164 | C | T | 0.0393 | 0.159 | 0.018 | 2.5119E-18 | 78.03 |
| *TNFRSF12A* | rs77542162 | A | G | 0.0228 | 0.221 | 0.024 | 6.3096E-21 | 84.79 |
| *TNFRSF13B* | rs1793891 | C | T | 0.2681 | -0.059 | 0.008 | 1.2589E-12 | 54.39 |
| *TNFRSF13B* | rs10806425 | C | A | 0.4135 | -0.053 | 0.007 | 1.9953E-12 | 57.33 |
| *TNFRSF13B* | rs3804329 | A | G | 0.2037 | 0.089 | 0.009 | 3.1623E-22 | 97.79 |
| *TNFRSF13B* | rs73169672 | A | T | 0.1141 | 0.089 | 0.012 | 1.2589E-14 | 55.01 |
| *TNFRSF13B* | rs1483572 | A | C | 0.6889 | -0.054 | 0.008 | 7.9433E-12 | 45.56 |
| *TNFRSF13B* | rs75587749 | C | T | 0.0854 | 0.1 | 0.013 | 2.5119E-14 | 59.17 |
| *TNFRSF13B* | rs3184504 | T | C | 0.5173 | -0.071 | 0.007 | 3.9811E-22 | 102.88 |
| *TNFRSF13B* | rs374039502 | T | A | 0.0205 | 0.714 | 0.028 | 2.512E-145 | 650.25 |
| *TNFRSF13B* | rs9989163 | G | A | 0.4748 | -0.054 | 0.007 | 1.9953E-13 | 59.51 |
| *TNFRSF13B* | rs34562254 | G | A | 0.0972 | -0.174 | 0.012 | 1.2589E-44 | 210.25 |
| *TNFRSF13B* | rs12975366 | T | C | 0.3952 | -0.061 | 0.008 | 5.0119E-16 | 58.14 |
| *TNFRSF13B* | rs6053024 | G | T | 0.3905 | 0.056 | 0.008 | 1.5849E-13 | 49.00 |
| *TNFRSF13B* | rs73165110 | G | A | 0.079 | -0.189 | 0.014 | 3.9811E-43 | 182.25 |
| *TNFRSF13C* | rs17056298 | C | G | 0.0753 | -0.13 | 0.014 | 1E-21 | 86.22 |
| *TNFRSF13C* | rs17198888 | A | G | 0.1912 | 0.072 | 0.009 | 1.9953E-15 | 64.00 |
| *TNFRSF13C* | rs2395904 | T | G | 0.2746 | -0.054 | 0.008 | 1.2589E-11 | 45.56 |
| *TNFRSF13C* | rs12784975 | T | C | 0.1914 | -0.073 | 0.009 | 6.3096E-16 | 65.79 |
| *TNFRSF13C* | rs774868556 | A | ACA | 0.3073 | -0.115 | 0.008 | 2.5119E-44 | 206.64 |
| *TNFRSF13C* | rs374039502 | T | A | 0.0206 | 0.217 | 0.027 | 3.9811E-16 | 64.59 |
| *TNFRSF13C* | rs13330176 | T | A | 0.2389 | 0.059 | 0.008 | 1.9953E-12 | 54.39 |
| *TNFRSF13C* | rs34557412 | A | G | 0.0068 | 0.667 | 0.043 | 3.9811E-53 | 240.61 |
| *TNFRSF13C* | rs35222145 | T | G | 0.4919 | -0.073 | 0.007 | 3.9811E-24 | 108.76 |
| *TNFRSF13C* | rs17676919 | A | G | 0.0974 | -0.082 | 0.012 | 6.3096E-12 | 46.69 |
| *TNFRSF13C* | rs1967330 | C | T | 0.5675 | 0.05 | 0.007 | 2.5119E-12 | 51.02 |
| *TNFRSF13C* | rs28823595 | G | A | 0.0874 | 0.088 | 0.013 | 3.1623E-12 | 45.82 |

| | | | | | | | | |
|---|---|---|---|---|---|---|---|---|
| *TNFRSF13C* | rs763882049 | AAT | A | 0.0947 | -0.793 | 0.013 | 1E-200 | 3721.00 |
| *TNFRSF14* | rs2495366 | G | A | 0.4937 | 0.115 | 0.007 | 3.9811E-54 | 269.90 |
| *TNFRSF14* | rs61747728 | C | T | 0.0379 | 0.134 | 0.019 | 3.9811E-12 | 49.74 |
| *TNFRSF14* | rs11926707 | T | C | 0.6218 | -0.053 | 0.008 | 6.3096E-12 | 43.89 |
| *TNFRSF14* | rs342293 | C | G | 0.4593 | -0.054 | 0.007 | 3.9811E-13 | 59.51 |
| *TNFRSF14* | rs6993770 | A | T | 0.2867 | -0.063 | 0.008 | 1.2589E-14 | 62.02 |
| *TNFRSF14* | rs10774624 | G | A | 0.5134 | -0.056 | 0.007 | 5.0119E-14 | 64.00 |
| *TNFRSF17* | rs6759 | C | T | 0.5149 | 0.059 | 0.007 | 5.0119E-15 | 71.04 |
| *TNFRSF17* | rs9799792 | T | C | 0.5568 | -0.065 | 0.008 | 2.5119E-17 | 66.02 |
| *TNFRSF17* | rs142714816 | A | G | 0.0008 | -1.193 | 0.135 | 1E-18 | 78.09 |
| *TNFRSF17* | rs2647074 | C | T | 0.3273 | -0.08 | 0.008 | 1.9953E-23 | 100.00 |
| *TNFRSF17* | rs61839660 | C | T | 0.097 | 0.085 | 0.013 | 1.2589E-11 | 42.75 |
| *TNFRSF17* | rs1265847 | G | A | 0.2616 | -0.058 | 0.009 | 1.2589E-11 | 41.53 |
| *TNFRSF17* | rs3803286 | A | G | 0.666 | -0.087 | 0.008 | 7.9433E-28 | 118.27 |
| *TNFRSF17* | rs71389075 | C | CT | 0.3371 | -0.144 | 0.008 | 7.9433E-73 | 324.00 |
| *TNFRSF17* | rs34562254 | G | A | 0.0972 | 0.232 | 0.013 | 7.9433E-76 | 318.49 |
| *TNFRSF17* | rs9957535 | G | A | 0.0844 | -0.091 | 0.013 | 1.2589E-11 | 49.00 |
| *TNFRSF17* | rs5754102 | C | A | 0.1809 | 0.07 | 0.01 | 7.9433E-13 | 49.00 |
| *TNFRSF19* | rs6605317 | T | A | 0.2265 | 0.064 | 0.008 | 1.2589E-14 | 64.00 |
| *TNFRSF19* | rs13146355 | G | A | 0.4567 | 0.049 | 0.007 | 1.5849E-12 | 49.00 |
| *TNFRSF19* | rs61947047 | C | T | 0.3095 | 0.259 | 0.008 | 1E-200 | 1048.14 |
| *TNFRSF19* | rs12434499 | C | T | 0.161 | 0.089 | 0.01 | 2.5119E-19 | 79.21 |
| *TNFRSF19* | rs35830321 | C | CA | 0.1667 | -0.078 | 0.009 | 7.9433E-17 | 75.11 |
| *TNFRSF19* | rs2252576 | C | T | 0.2436 | 0.147 | 0.008 | 1.5849E-71 | 337.64 |
| *TNFRSF1A* | rs61747728 | C | T | 0.0379 | 0.165 | 0.019 | 1.5849E-18 | 75.42 |
| *TNFRSF1A* | rs13019008 | G | A | 0.5143 | 0.073 | 0.007 | 1.2589E-23 | 108.76 |
| *TNFRSF1A* | rs553340154 | T | G | 0.3343 | 0.069 | 0.009 | 3.1623E-14 | 58.78 |
| *TNFRSF1A* | rs10982156 | T | A | 0.0692 | 0.176 | 0.015 | 2.5119E-32 | 137.67 |
| *TNFRSF1A* | rs8176672 | C | T | 0.0612 | 0.123 | 0.015 | 2.5119E-16 | 67.24 |
| *TNFRSF1A* | rs4149584 | C | T | 0.0167 | -0.303 | 0.028 | 6.3096E-27 | 117.10 |
| *TNFRSF1A* | rs28640218 | G | T | 0.1914 | -0.075 | 0.009 | 3.1623E-16 | 69.44 |
| *TNFRSF1A* | rs516316 | G | C | 0.5097 | -0.055 | 0.007 | 6.3096E-14 | 61.73 |
| *TNFRSF1B* | rs5746026 | G | A | 0.0398 | -0.392 | 0.019 | 1.2589E-96 | 425.66 |
| *TNFRSF1B* | rs17162190 | G | A | 0.1992 | 0.062 | 0.009 | 1.5849E-11 | 47.46 |
| *TNFRSF1B* | rs61747728 | C | T | 0.0379 | 0.183 | 0.019 | 1E-21 | 92.77 |
| *TNFRSF1B* | rs2239527 | C | G | 0.3598 | 0.086 | 0.008 | 5.0119E-30 | 115.56 |
| *TNFRSF1B* | rs3184504 | T | C | 0.5175 | -0.075 | 0.007 | 1.9953E-25 | 114.80 |
| *TNFRSF1B* | rs77924615 | G | A | 0.1966 | -0.065 | 0.009 | 1.9953E-12 | 52.16 |
| *TNFRSF1B* | rs3814995 | C | T | 0.3153 | 0.056 | 0.008 | 6.3096E-13 | 49.00 |
| *TNFRSF21* | rs7528419 | A | G | 0.2227 | -0.073 | 0.009 | 1E-16 | 65.79 |
| *TNFRSF21* | rs150816167 | T | C | 0.0416 | 0.184 | 0.019 | 3.1623E-23 | 93.78 |
| *TNFRSF21* | rs1260326 | T | C | 0.6085 | 0.058 | 0.007 | 1.2589E-14 | 68.65 |
| *TNFRSF21* | rs75166367 | G | A | 0.0603 | -0.107 | 0.015 | 3.1623E-12 | 50.88 |
| *TNFRSF21* | rs13107325 | C | T | 0.0747 | 0.114 | 0.014 | 2.5119E-16 | 66.31 |
| *TNFRSF21* | rs17613643 | A | G | 0.2179 | 0.076 | 0.009 | 3.9811E-16 | 71.31 |
| *TNFRSF21* | rs148598211 | C | T | 0.0042 | -1.874 | 0.057 | 1E-200 | 1080.91 |
| *TNFRSF21* | rs2298475 | T | C | 0.0753 | -0.183 | 0.014 | 6.3096E-40 | 170.86 |
| *TNFRSF21* | rs77924615 | G | A | 0.1967 | -0.07 | 0.009 | 3.9811E-14 | 60.49 |
| *TNFRSF21* | rs186021206 | G | A | 0.0064 | 1.139 | 0.05 | 5.012E-114 | 518.93 |
| *TNFRSF21* | rs10445407 | C | A | 0.4787 | -0.053 | 0.007 | 5.0119E-13 | 57.33 |
| *TNFRSF4* | rs2524137 | C | T | 0.6919 | -0.125 | 0.008 | 1.5849E-55 | 244.14 |
| *TNFRSF4* | rs4946811 | A | C | 0.3612 | 0.054 | 0.008 | 1.5849E-12 | 45.56 |
| *TNFRSF4* | rs7137828 | C | T | 0.5162 | -0.124 | 0.007 | 7.9433E-66 | 313.80 |
| *TNFRSF4* | rs72778736 | C | T | 0.2564 | 0.057 | 0.008 | 1.2589E-11 | 50.77 |
| *TNFRSF6B* | rs200607887 | G | GT | 0.0169 | 0.219 | 0.028 | 1.2589E-14 | 61.17 |
| *TNFRSF6B* | rs7137828 | C | T | 0.5158 | -0.072 | 0.007 | 6.3096E-24 | 105.80 |
| *TNFRSF6B* | rs77542162 | A | G | 0.023 | 0.204 | 0.024 | 1.2589E-17 | 72.25 |
| *TNFRSF6B* | rs6062497 | C | T | 0.6658 | -0.309 | 0.008 | 1E-200 | 1491.89 |
| *TNFRSF8* | rs35249183 | A | G | 0.1 | 0.348 | 0.013 | 3.162E-168 | 716.59 |
| *TNFRSF8* | rs3094005 | G | T | 0.1411 | 0.137 | 0.011 | 5.0119E-38 | 155.12 |
| *TNFRSF8* | rs10817685 | C | T | 0.4293 | -0.114 | 0.007 | 5.0119E-53 | 265.22 |
| *TNFRSF8* | rs3184504 | T | C | 0.5171 | -0.079 | 0.007 | 2.5119E-27 | 127.37 |
| *TNFRSF8* | rs76428106 | T | C | 0.0129 | 0.231 | 0.034 | 7.9433E-12 | 46.16 |
| *TNFRSF8* | rs2695167 | A | G | 0.0173 | 0.369 | 0.028 | 3.9811E-39 | 173.67 |
| *TNFRSF8* | rs186021206 | G | A | 0.0064 | 0.388 | 0.05 | 1.5849E-14 | 60.22 |
| *TNFRSF8* | rs3814995 | C | T | 0.3147 | 0.054 | 0.008 | 1.2589E-11 | 45.56 |
| *TNFRSF9* | rs2493214 | A | G | 0.1904 | -0.194 | 0.009 | 5.0119E-97 | 464.64 |
| *TNFRSF9* | rs3117574 | G | A | 0.1326 | 0.124 | 0.011 | 6.3096E-31 | 127.07 |

| | | | | | | | | |
|---|---|---|---|---|---|---|---|---|
| *TNFRSF9* | rs111338191 | A | AAATT | 0.5209 | -0.079 | 0.007 | 2.5119E-27 | 127.37 |
| *TNFRSF9* | rs374039502 | T | A | 0.0206 | 0.198 | 0.027 | 2.5119E-13 | 53.78 |
| *TNFRSF9* | rs34557412 | A | G | 0.0068 | 0.495 | 0.044 | 1.9953E-29 | 126.56 |
| *TNFRSF9* | rs61750000 | G | C | 0.0047 | -0.395 | 0.053 | 7.9433E-14 | 55.54 |
| *TNFSF10* | rs7519758 | C | T | 0.1942 | 0.133 | 0.009 | 3.1623E-51 | 218.38 |
| *TNFSF10* | rs79287178 | G | A | 0.0302 | -0.537 | 0.022 | 6.31E-135 | 595.80 |
| *TNFSF10* | rs756671612 | GAATT | G | 0.1083 | -0.143 | 0.011 | 1.9953E-35 | 169.00 |
| *TNFSF10* | rs10502 | G | C | 0.3809 | 0.05 | 0.007 | 3.1623E-12 | 51.02 |
| *TNFSF10* | rs174547 | T | C | 0.3446 | 0.069 | 0.007 | 6.3096E-21 | 97.16 |
| *TNFSF10* | rs1027315 | T | C | 0.4295 | -0.058 | 0.007 | 3.1623E-16 | 68.65 |
| *TNFSF10* | rs28929474 | C | T | 0.0208 | 0.843 | 0.025 | 1E-200 | 1137.04 |
| *TNFSF10* | rs4788460 | C | T | 0.2944 | 0.093 | 0.008 | 3.1623E-33 | 135.14 |
| *TNFSF10* | rs8178824 | C | T | 0.03 | 0.513 | 0.021 | 1.995E-134 | 596.76 |
| *TNFSF10* | rs680321 | T | C | 0.4558 | -0.109 | 0.007 | 1.9953E-53 | 242.47 |
| *TNFSF10* | rs4760 | A | G | 0.156 | -0.48 | 0.01 | 1E-200 | 2304.00 |
| *TNFSF11* | rs552693039 | T | TC | 0.1831 | 0.066 | 0.009 | 3.1623E-12 | 53.78 |
| *TNFSF11* | rs79287178 | G | A | 0.03 | -0.661 | 0.023 | 3.981E-187 | 825.94 |
| *TNFSF11* | rs7011570 | T | C | 0.7467 | 0.177 | 0.009 | 1.5849E-94 | 386.78 |
| *TNFSF11* | rs2737212 | C | T | 0.5499 | -0.054 | 0.007 | 3.9811E-13 | 59.51 |
| *TNFSF11* | rs1156545 | G | A | 0.4584 | -0.222 | 0.008 | 7.943E-192 | 770.06 |
| *TNFSF11* | rs7137828 | C | T | 0.5163 | -0.058 | 0.007 | 1.9953E-15 | 68.65 |
| *TNFSF11* | rs2062305 | G | A | 0.5316 | -0.112 | 0.007 | 7.9433E-52 | 256.00 |
| *TNFSF11* | rs7146217 | T | C | 0.5311 | 0.051 | 0.007 | 5.0119E-12 | 53.08 |
| *TNFSF11* | rs2207132 | G | A | 0.0326 | -0.149 | 0.021 | 5.0119E-13 | 50.34 |
| *TNFSF12* | rs2228099 | C | G | 0.3673 | -0.063 | 0.007 | 1.5849E-17 | 81.00 |
| *TNFSF12* | rs1354034 | T | C | 0.6041 | -0.052 | 0.007 | 1.5849E-12 | 55.18 |
| *TNFSF12* | rs828596 | C | G | 0.4142 | -0.066 | 0.007 | 1.5849E-19 | 88.90 |
| *TNFSF12* | rs9842051 | G | C | 0.7311 | 0.071 | 0.008 | 2.5119E-18 | 78.77 |
| *TNFSF12* | rs13135092 | A | G | 0.0835 | 0.136 | 0.013 | 1.2589E-25 | 109.44 |
| *TNFSF12* | rs4098923 | A | G | 0.4478 | -0.057 | 0.008 | 6.3096E-14 | 50.77 |
| *TNFSF12* | rs139141690 | G | A | 0.0048 | 0.358 | 0.053 | 1E-11 | 45.63 |
| *TNFSF12* | rs6993770 | A | T | 0.2866 | -0.071 | 0.008 | 5.0119E-19 | 78.77 |
| *TNFSF12* | rs10107388 | T | C | 0.3659 | 0.076 | 0.007 | 3.1623E-24 | 117.88 |
| *TNFSF12* | rs10761741 | G | T | 0.4139 | 0.098 | 0.007 | 1.9953E-41 | 196.00 |
| *TNFSF12* | rs745680717 | CCTG | C | 0.4041 | -0.065 | 0.007 | 1.2589E-18 | 86.22 |
| *TNFSF12* | rs11158588 | G | A | 0.7931 | -0.074 | 0.009 | 7.9433E-17 | 67.60 |
| *TNFSF12* | rs151233 | C | T | 0.1298 | 0.074 | 0.011 | 3.1623E-12 | 45.26 |
| *TNFSF12* | rs80067372 | G | A | 0.2885 | 0.372 | 0.008 | 1E-200 | 2162.25 |
| *TNFSF12* | rs77542162 | A | G | 0.0229 | 0.181 | 0.025 | 2.5119E-13 | 52.42 |
| *TNFSF12* | rs892090 | T | G | 0.8341 | 0.104 | 0.01 | 6.3096E-27 | 108.16 |
| *TNFSF13* | rs12740374 | G | T | 0.2214 | -0.237 | 0.008 | 1.995E-175 | 877.64 |
| *TNFSF13* | rs1354034 | T | C | 0.6041 | -0.053 | 0.007 | 1E-13 | 57.33 |
| *TNFSF13* | rs35173808 | C | A | 0.0966 | -0.099 | 0.012 | 3.9811E-17 | 68.06 |
| *TNFSF13* | rs11570136 | T | A | 0.3268 | 0.067 | 0.007 | 1.9953E-19 | 91.61 |
| *TNFSF13* | rs3803800 | A | G | 0.7887 | -0.325 | 0.009 | 1E-200 | 1304.01 |
| *TNFSF13* | rs16961828 | G | C | 0.1021 | -0.099 | 0.012 | 5.0119E-17 | 68.06 |
| *TNFSF13* | rs77542162 | A | G | 0.0229 | 0.177 | 0.024 | 1E-13 | 54.39 |
| *TNFSF13* | rs892090 | T | G | 0.8342 | 0.076 | 0.009 | 3.1623E-16 | 71.31 |
| *TNFSF13B* | rs35675148 | C | CAT | 0.7745 | -0.077 | 0.008 | 3.9811E-20 | 92.64 |
| *TNFSF13B* | rs3129755 | A | G | 0.5685 | 0.096 | 0.007 | 1.9953E-40 | 188.08 |
| *TNFSF13B* | rs6488394 | T | C | 0.9113 | -0.15 | 0.012 | 3.1623E-34 | 156.25 |
| *TNFSF13B* | rs374039502 | T | A | 0.0206 | 0.746 | 0.027 | 2.512E-174 | 763.40 |
| *TNFSF13B* | rs79874727 | G | A | 0.1059 | 0.089 | 0.013 | 3.9811E-12 | 46.87 |
| *TNFSF13B* | rs34557412 | A | G | 0.0068 | 0.598 | 0.043 | 6.3096E-45 | 193.40 |
| *TNFSF13B* | rs12986064 | T | C | 0.5074 | -0.118 | 0.007 | 1.5849E-59 | 284.16 |
| *TNFSF13B* | rs763882049 | AAT | A | 0.0948 | 0.395 | 0.013 | 1E-200 | 923.22 |
| *TNFSF13B* | rs74510325 | C | G | 0.0347 | 0.203 | 0.019 | 6.3096E-26 | 114.15 |
| *TNFSF14* | rs2631367 | C | G | 0.5182 | 0.06 | 0.007 | 7.9433E-16 | 73.47 |
| *TNFSF14* | rs6993770 | A | T | 0.2865 | -0.055 | 0.008 | 1.5849E-11 | 47.27 |
| *TNFSF14* | rs12411442 | A | G | 0.4289 | -0.051 | 0.007 | 6.3096E-12 | 53.08 |
| *TNFSF14* | rs344560 | T | C | 0.9454 | 0.554 | 0.017 | 1E-200 | 1061.99 |
| *TNFSF14* | rs892090 | T | G | 0.8341 | 0.107 | 0.01 | 1.5849E-25 | 114.49 |
| *TNFSF8* | rs37453 | A | G | 0.3522 | 0.15 | 0.008 | 1.5849E-85 | 351.56 |
| *TNFSF8* | rs113280910 | T | TTGTC | 0.7717 | -0.063 | 0.009 | 1E-12 | 49.00 |
| *TNFSF8* | rs34479374 | G | A | 0.0836 | 0.154 | 0.013 | 2.5119E-31 | 140.33 |
| *TNFSF8* | rs9824474 | A | G | 0.3672 | -0.108 | 0.008 | 6.3096E-45 | 182.25 |
| *TNFSF8* | rs2523996 | A | G | 0.557 | -0.062 | 0.008 | 1E-15 | 60.06 |
| *TNFSF8* | rs2126259 | T | C | 0.8973 | -0.097 | 0.012 | 1E-15 | 65.34 |

| | | | | | | | | |
|---|---|---|---|---|---|---|---|---|
| *TNFSF8* | rs10081728 | T | C | 0.4069 | -0.236 | 0.008 | 1E-200 | 870.25 |
| *TNFSF8* | rs550057 | C | T | 0.2554 | 0.103 | 0.008 | 7.9433E-34 | 165.77 |
| *TNFSF8* | rs7937564 | G | A | 0.3204 | -0.055 | 0.008 | 2.5119E-12 | 47.27 |
| *TNFSF8* | rs78689694 | G | C | 0.1326 | -0.146 | 0.011 | 1.9953E-41 | 176.17 |
| *TNFSF8* | rs10774625 | A | G | 0.5041 | -0.099 | 0.007 | 2.5119E-42 | 200.02 |
| *TNFSF8* | rs186021206 | G | A | 0.0064 | 1.816 | 0.051 | 1E-200 | 1267.92 |
| *TNFSF8* | rs2659005 | C | T | 0.4413 | 0.065 | 0.007 | 3.1623E-18 | 86.22 |
| *TNIP1* | rs1354034 | T | C | 0.6043 | 0.074 | 0.007 | 3.9811E-23 | 111.76 |
| *TNIP1* | rs1585213 | C | T | 0.3989 | 0.051 | 0.007 | 1.2589E-11 | 53.08 |
| *TNIP1* | rs342293 | C | G | 0.46 | -0.053 | 0.007 | 7.9433E-13 | 57.33 |
| *TNIP1* | rs4759076 | T | C | 0.4571 | 0.065 | 0.007 | 1.5849E-18 | 86.22 |
| *TNN* | rs535777 | G | C | 0.1432 | -0.078 | 0.008 | 1.5849E-24 | 95.06 |
| *TNN* | rs4841132 | A | G | 0.908 | -0.073 | 0.009 | 2.5119E-15 | 65.79 |
| *TNN* | rs2685412 | C | G | 0.2704 | 0.146 | 0.006 | 3.162E-132 | 592.11 |
| *TNN* | rs2068888 | G | A | 0.4507 | -0.038 | 0.005 | 5.0119E-13 | 57.76 |
| *TNN* | rs174561 | T | C | 0.3097 | 0.046 | 0.006 | 5.0119E-16 | 58.78 |
| *TNN* | rs215224 | T | C | 0.4008 | -0.048 | 0.005 | 1.9953E-19 | 92.16 |
| *TNR* | rs2235256 | A | G | 0.4456 | -0.375 | 0.007 | 1E-200 | 2869.90 |
| *TNR* | rs2169305 | T | C | 0.3612 | 0.078 | 0.007 | 5.0119E-27 | 124.16 |
| *TNR* | rs2507997 | A | G | 0.306 | -0.071 | 0.008 | 7.9433E-21 | 78.77 |
| *TNR* | rs1111796 | A | G | 0.6036 | 0.138 | 0.007 | 1.5849E-79 | 388.65 |
| *TNR* | rs11594905 | G | A | 0.1359 | 0.08 | 0.01 | 3.9811E-15 | 64.00 |
| *TNR* | rs1727 | C | A | 0.7241 | -0.057 | 0.008 | 3.9811E-13 | 50.77 |
| *TNR* | rs7120175 | G | C | 0.4519 | -0.052 | 0.007 | 1.5849E-13 | 55.18 |
| *TNR* | rs11603123 | G | A | 0.0336 | 0.342 | 0.019 | 5.0119E-70 | 324.00 |
| *TNR* | rs4981022 | G | A | 0.6808 | 0.081 | 0.008 | 2.5119E-26 | 102.52 |
| *TNR* | rs3184504 | T | C | 0.517 | 0.063 | 0.007 | 1.9953E-19 | 81.00 |
| *TNR* | rs4767280 | C | T | 0.4006 | -0.114 | 0.007 | 2.5119E-57 | 265.22 |
| *TNR* | rs12879626 | T | G | 0.6103 | 0.133 | 0.007 | 3.9811E-75 | 361.00 |
| *TNR* | rs10658607 | A | AAAAT | 0.3235 | -0.055 | 0.008 | 2.5119E-13 | 47.27 |
| *TNR* | rs2439430 | G | T | 0.4994 | 0.067 | 0.007 | 1.9953E-21 | 91.61 |
| *TNR* | rs4886410 | G | C | 0.682 | 0.056 | 0.008 | 1E-13 | 49.00 |
| *TNR* | rs3817428 | C | G | 0.2663 | 0.152 | 0.008 | 5.0119E-82 | 361.00 |
| *TNR* | rs186021206 | G | A | 0.0064 | 0.59 | 0.048 | 1.5849E-34 | 151.09 |
| *TNXB* | rs147650812 | T | C | 0.0081 | -0.272 | 0.04 | 6.3096E-12 | 46.24 |
| *TNXB* | rs9267797 | C | T | 0.0147 | 1.661 | 0.031 | 1E-200 | 2870.89 |
| *TNXB* | rs181242111 | G | A | 0.118 | 0.089 | 0.011 | 3.9811E-15 | 65.46 |
| *TNXB* | rs78689694 | G | C | 0.133 | -0.15 | 0.01 | 3.9811E-50 | 225.00 |
| *TNXB* | rs186021206 | G | A | 0.0064 | 0.808 | 0.047 | 6.3096E-67 | 295.55 |
| *TNXB* | rs1236213 | G | T | 0.593 | -0.05 | 0.007 | 7.9433E-13 | 51.02 |
| *TOMM20* | rs1354034 | T | C | 0.6045 | 0.073 | 0.008 | 1.9953E-21 | 83.27 |
| *TOMM20* | rs342293 | C | G | 0.4592 | -0.06 | 0.008 | 1.2589E-15 | 56.25 |
| *TOMM20* | rs6993770 | A | T | 0.2866 | -0.065 | 0.008 | 6.3096E-15 | 66.02 |
| *TOMM20* | rs7896518 | A | G | 0.424 | 0.063 | 0.008 | 3.1623E-16 | 62.02 |
| *TOMM20* | rs73396521 | G | A | 0.245 | 0.06 | 0.009 | 7.9433E-12 | 44.44 |
| *TOMM20* | rs35277580 | G | A | 0.4922 | -0.052 | 0.008 | 7.9433E-12 | 42.25 |
| *TOP1* | rs760077 | A | T | 0.6058 | -0.074 | 0.008 | 7.9433E-22 | 85.56 |
| *TOP1* | rs112268616 | T | C | 0.3401 | -0.745 | 0.008 | 1E-200 | 8672.27 |
| *TOP1* | rs11680123 | A | G | 0.5928 | 0.115 | 0.006 | 1.9953E-79 | 367.36 |
| *TOP1* | rs121908120 | T | A | 0.0276 | -0.134 | 0.019 | 5.0119E-13 | 49.74 |
| *TOP1* | rs11707641 | T | C | 0.1625 | 0.059 | 0.008 | 2.5119E-13 | 54.39 |
| *TOP1* | rs34211178 | G | A | 0.4407 | -0.044 | 0.006 | 1.9953E-13 | 53.78 |
| *TOP1* | rs36034702 | C | T | 0.1686 | -0.07 | 0.008 | 6.3096E-18 | 76.56 |
| *TOP2B* | rs3806631 | A | T | 0.1998 | 0.098 | 0.009 | 5.0119E-25 | 118.57 |
| *TOP2B* | rs1354034 | T | C | 0.6039 | 0.194 | 0.008 | 1E-139 | 588.06 |
| *TOP2B* | rs6993770 | A | T | 0.2866 | -0.057 | 0.008 | 1E-11 | 50.77 |
| *TOP2B* | rs11605246 | C | G | 0.229 | 0.082 | 0.009 | 1.5849E-19 | 83.01 |
| *TOP2B* | rs6580981 | G | A | 0.4585 | 0.084 | 0.008 | 1.9953E-28 | 110.25 |
| *TOR1AIP1* | rs486906 | G | T | 0.6163 | 0.709 | 0.008 | 1E-200 | 7854.39 |
| *TOR1AIP1* | rs61000956 | T | C | 0.4824 | -0.056 | 0.007 | 2.5119E-16 | 64.00 |
| *TOR1AIP1* | rs1801020 | A | G | 0.7445 | -0.055 | 0.008 | 7.9433E-13 | 47.27 |
| *TOR1AIP1* | rs34436714 | C | A | 0.2087 | 0.081 | 0.008 | 1E-22 | 102.52 |
| *TP53* | rs1641549 | C | T | 0.2551 | 0.066 | 0.009 | 1.9953E-13 | 53.78 |
| *TP53BP1* | rs78037977 | A | G | 0.1244 | -0.081 | 0.012 | 7.9433E-12 | 45.56 |
| *TP53BP1* | rs3087243 | G | A | 0.4519 | -0.054 | 0.008 | 2.5119E-12 | 45.56 |
| *TP53BP1* | rs148451580 | G | A | 0.3924 | -0.122 | 0.008 | 1.9953E-49 | 232.56 |
| *TP53BP1* | rs10957978 | G | T | 0.6481 | 0.063 | 0.008 | 2.5119E-15 | 62.02 |
| *TP53I3* | rs1134516 | G | A | 0.1586 | -0.596 | 0.01 | 1E-200 | 3552.16 |

| | | | | | | | | |
|---|---|---|---|---|---|---|---|---|
| *TP53I3* | rs1354034 | T | C | 0.6042 | 0.069 | 0.007 | 1E-20 | 97.16 |
| *TP53I3* | rs6993770 | A | T | 0.287 | -0.079 | 0.008 | 1E-22 | 97.52 |
| *TP53INP1* | rs58534292 | T | C | 0.6498 | -0.057 | 0.008 | 2.5119E-12 | 50.77 |
| *TP53INP1* | rs56366203 | C | G | 0.4982 | -0.064 | 0.008 | 3.1623E-16 | 64.00 |
| *TP53INP1* | rs8192297 | T | C | 0.1135 | -0.153 | 0.012 | 2.5119E-36 | 162.56 |
| *TPBGL* | rs58175074 | A | G | 0.1969 | 0.074 | 0.01 | 2.5119E-14 | 54.76 |
| *TPD52L2* | rs1354034 | T | C | 0.6044 | 0.077 | 0.008 | 5.0119E-23 | 92.64 |
| *TPD52L2* | rs342298 | C | T | 0.4549 | -0.07 | 0.008 | 7.9433E-20 | 76.56 |
| *TPD52L2* | rs6993770 | A | T | 0.287 | -0.067 | 0.008 | 1.2589E-15 | 70.14 |
| *TPK1* | rs61747728 | C | T | 0.0379 | 0.14 | 0.019 | 5.0119E-14 | 54.29 |
| *TPK1* | rs10261806 | G | A | 0.4028 | -0.334 | 0.008 | 1E-200 | 1743.06 |
| *TPK1* | rs4879666 | G | A | 0.5104 | 0.062 | 0.007 | 6.3096E-18 | 78.45 |
| *TPK1* | rs12342201 | G | A | 0.4764 | -0.061 | 0.007 | 1E-17 | 75.94 |
| *TPK1* | rs56278466 | T | G | 0.6612 | 0.102 | 0.008 | 6.3096E-42 | 162.56 |
| *TPK1* | rs7213540 | C | T | 0.5389 | -0.049 | 0.007 | 6.3096E-12 | 49.00 |
| *TPK1* | rs33950747 | C | T | 0.075 | 0.112 | 0.013 | 7.9433E-17 | 74.22 |
| *TPM3* | rs4478991 | A | G | 0.5719 | 0.058 | 0.008 | 1E-13 | 52.56 |
| *TPMT* | rs115306927 | A | T | 0.0502 | -1.095 | 0.017 | 1E-200 | 4148.88 |
| *TPMT* | rs6580981 | G | A | 0.4585 | 0.05 | 0.007 | 7.9433E-12 | 51.02 |
| *TPP1* | rs77074394 | A | G | 0.0316 | 0.156 | 0.021 | 1.5849E-13 | 55.18 |
| *TPP1* | rs10455861 | G | A | 0.1371 | -0.099 | 0.011 | 3.9811E-20 | 81.00 |
| *TPP1* | rs17154155 | G | T | 0.4071 | 0.058 | 0.007 | 5.0119E-15 | 68.65 |
| *TPP1* | rs6993770 | A | T | 0.2868 | -0.113 | 0.008 | 5.0119E-44 | 199.52 |
| *TPP1* | rs9414801 | G | A | 0.5331 | -0.077 | 0.007 | 3.1623E-25 | 121.00 |
| *TPP1* | rs140726254 | G | A | 0.0022 | 2.89 | 0.079 | 1E-200 | 1338.26 |
| *TPP1* | rs149871778 | G | GAAC | 0.1048 | 0.183 | 0.012 | 1.5849E-51 | 232.56 |
| *TPP1* | rs10778152 | A | G | 0.288 | -0.131 | 0.008 | 3.1623E-57 | 268.14 |
| *TPP1* | rs3184504 | T | C | 0.5174 | -0.065 | 0.007 | 7.9433E-19 | 86.22 |
| *TPP1* | rs145078947 | G | T | 0.0026 | 0.876 | 0.072 | 7.9433E-34 | 148.03 |
| *TPP1* | rs1045693 | G | A | 0.5325 | -0.081 | 0.007 | 2.5119E-27 | 133.90 |
| *TPP1* | rs8064959 | A | T | 0.3105 | -0.054 | 0.008 | 7.9433E-12 | 45.56 |
| *TPP1* | rs200210321 | A | AG | 0.0742 | 0.124 | 0.014 | 1.2589E-18 | 78.45 |
| *TPP1* | rs1654425 | T | C | 0.8344 | 0.094 | 0.01 | 2.5119E-21 | 88.36 |
| *TPPP3* | rs13334364 | T | C | 0.0751 | -0.117 | 0.015 | 1.2589E-15 | 60.84 |
| *TPR* | rs115482159 | C | T | 0.0058 | 0.42 | 0.051 | 3.1623E-16 | 67.82 |
| *TPSAB1* | rs115427247 | C | T | 0.0299 | -0.155 | 0.015 | 2.5119E-25 | 106.78 |
| *TPSAB1* | rs16856110 | A | G | 0.23 | -0.057 | 0.006 | 3.9811E-21 | 90.25 |
| *TPSAB1* | rs2884425 | C | T | 0.4929 | -0.038 | 0.005 | 7.9433E-14 | 57.76 |
| *TPSAB1* | rs56043070 | G | A | 0.0732 | 0.094 | 0.01 | 2.5119E-22 | 88.36 |
| *TPSAB1* | rs62252239 | G | T | 0.2121 | 0.045 | 0.006 | 1.9953E-13 | 56.25 |
| *TPSAB1* | rs586320 | G | A | 0.4123 | 0.048 | 0.005 | 1E-20 | 92.16 |
| *TPSAB1* | rs28455789 | G | A | 0.1732 | -0.048 | 0.007 | 1E-12 | 47.02 |
| *TPSAB1* | rs876606 | G | A | 0.2563 | 0.048 | 0.006 | 3.1623E-16 | 64.00 |
| *TPSAB1* | rs56278466 | T | G | 0.6613 | 0.039 | 0.005 | 3.1623E-13 | 60.84 |
| *TPSAB1* | rs7127546 | G | A | 0.3742 | 0.052 | 0.005 | 3.9811E-23 | 108.16 |
| *TPSAB1* | rs8051930 | T | A | 0.478 | 0.832 | 0.008 | 1E-200 | 10816.00 |
| *TPSAB1* | rs17758695 | C | T | 0.0292 | -0.124 | 0.015 | 1E-16 | 68.34 |
| *TPSAB1* | rs78744187 | C | T | 0.0815 | 0.186 | 0.009 | 5.0119E-91 | 427.11 |
| *TPSAB1* | rs111402121 | C | CTCTT | 0.3768 | 0.07 | 0.005 | 1.2589E-40 | 196.00 |
| *TPSD1* | rs34072109 | C | CTCTA | 0.3769 | 0.054 | 0.008 | 7.9433E-13 | 45.56 |
| *TPSD1* | rs2745083 | C | A | 0.5131 | 0.267 | 0.008 | 1E-200 | 1113.89 |
| *TPSD1* | rs78744187 | C | T | 0.0816 | 0.14 | 0.013 | 1.5849E-25 | 115.98 |
| *TPSD1* | rs111402121 | C | CTCTT | 0.377 | 0.059 | 0.008 | 7.9433E-15 | 54.39 |
| *TRAF2* | rs1354034 | T | C | 0.6043 | 0.07 | 0.007 | 5.0119E-21 | 100.00 |
| *TRAF2* | rs532086 | T | C | 0.1616 | -0.069 | 0.01 | 3.9811E-12 | 47.61 |
| *TRAF2* | rs4239702 | T | C | 0.7225 | -0.061 | 0.008 | 1E-13 | 58.14 |
| *TRAF3* | rs6929057 | A | C | 0.249 | 0.096 | 0.009 | 1.2589E-27 | 113.78 |
| *TRAF3* | rs13263709 | T | C | 0.6487 | 0.056 | 0.008 | 2.5119E-12 | 49.00 |
| *TRDMT1* | rs1354034 | T | C | 0.6041 | 0.078 | 0.008 | 3.9811E-24 | 95.06 |
| *TRDMT1* | rs6602180 | T | C | 0.4629 | -0.147 | 0.008 | 3.1623E-82 | 337.64 |
| *TREH* | rs114165349 | G | C | 0.0233 | 0.122 | 0.018 | 3.9811E-12 | 45.94 |
| *TREH* | rs2484697 | A | G | 0.5237 | -0.037 | 0.005 | 6.3096E-12 | 54.76 |
| *TREH* | rs9492442 | G | A | 0.319 | -0.039 | 0.006 | 7.9433E-12 | 42.25 |
| *TREH* | rs13240065 | G | A | 0.1273 | 0.064 | 0.008 | 1E-15 | 64.00 |
| *TREH* | rs11455968 | G | GA | 0.3254 | 0.884 | 0.008 | 1E-200 | 12210.25 |
| *TREH* | rs33921258 | G | A | 0.1075 | 0.066 | 0.009 | 7.9433E-15 | 53.78 |
| *TREH* | rs1169292 | C | T | 0.3088 | 0.043 | 0.006 | 1E-13 | 51.36 |
| *TREH* | rs5023078 | T | C | 0.5242 | 0.039 | 0.005 | 2.5119E-13 | 60.84 |

| | | | | | | | | |
|---|---|---|---|---|---|---|---|---|
| *TREH* | rs8000528 | T | G | 0.5451 | 0.039 | 0.005 | 1.5849E-13 | 60.84 |
| *TREH* | rs55707100 | C | T | 0.0242 | 0.166 | 0.017 | 3.9811E-22 | 95.35 |
| *TREH* | rs339969 | C | A | 0.6164 | 0.061 | 0.005 | 7.9433E-29 | 148.84 |
| *TREH* | rs5841058 | C | CT | 0.4223 | -0.045 | 0.005 | 1.9953E-16 | 81.00 |
| *TREM2* | rs10919543 | A | G | 0.3161 | 0.05 | 0.007 | 7.9433E-12 | 51.02 |
| *TREM2* | rs2274700 | G | A | 0.4033 | -0.057 | 0.007 | 5.0119E-17 | 66.31 |
| *TREM2* | rs531950386 | G | T | 0.4823 | -0.066 | 0.008 | 3.1623E-18 | 68.06 |
| *TREM2* | rs143332484 | C | T | 0.0099 | -1.483 | 0.034 | 1E-200 | 1902.50 |
| *TREM2* | rs913914 | C | G | 0.3685 | -0.052 | 0.007 | 7.9433E-14 | 55.18 |
| *TREM2* | rs7232 | T | A | 0.3735 | 0.401 | 0.007 | 1E-200 | 3281.65 |
| *TREM2* | rs11057840 | A | C | 0.1383 | 0.102 | 0.01 | 1E-25 | 104.04 |
| *TREM2* | rs261290 | T | C | 0.6551 | -0.048 | 0.007 | 1E-11 | 47.02 |
| *TREM2* | rs738409 | C | G | 0.2169 | 0.065 | 0.008 | 6.3096E-16 | 66.02 |
| *TREML1* | rs4698839 | C | T | 0.3983 | -0.065 | 0.008 | 3.1623E-17 | 66.02 |
| *TREML1* | rs10058074 | G | A | 0.4752 | -0.057 | 0.008 | 3.1623E-14 | 50.77 |
| *TREML1* | rs6961069 | C | T | 0.4027 | 0.058 | 0.008 | 3.1623E-14 | 52.56 |
| *TREML1* | rs6993770 | A | T | 0.2866 | -0.105 | 0.008 | 1.2589E-36 | 172.27 |
| *TREML1* | rs7896518 | A | G | 0.4239 | 0.094 | 0.008 | 2.5119E-34 | 138.06 |
| *TREML1* | rs12445050 | C | T | 0.1385 | 0.088 | 0.011 | 7.9433E-16 | 64.00 |
| *TREML1* | rs1654425 | T | C | 0.8341 | 0.138 | 0.01 | 1E-41 | 190.44 |
| *TREML1* | rs6112308 | C | T | 0.3538 | 0.061 | 0.008 | 6.3096E-15 | 58.14 |
| *TREML2* | rs142421172 | G | A | 0.0399 | 0.135 | 0.019 | 1.9953E-12 | 50.48 |
| *TREML2* | rs11759347 | C | A | 0.2384 | 0.307 | 0.009 | 1E-200 | 1163.57 |
| *TREML2* | rs56293029 | C | A | 0.2554 | 0.083 | 0.009 | 1.2589E-21 | 85.05 |
| *TREML2* | rs409801 | T | C | 0.5069 | 0.061 | 0.007 | 5.0119E-17 | 75.94 |
| *TREML2* | rs7080386 | C | A | 0.4124 | 0.06 | 0.007 | 3.9811E-16 | 73.47 |
| *TREML2* | rs12576289 | A | G | 0.2193 | -0.065 | 0.009 | 3.9811E-13 | 52.16 |
| *TREML2* | rs111338191 | A | AAATT | 0.5212 | -0.12 | 0.007 | 3.1623E-59 | 293.88 |
| *TREML2* | rs186021206 | G | A | 0.0064 | 0.465 | 0.05 | 1.2589E-20 | 86.49 |
| *TRIAP1* | rs148212596 | A | G | 0.0209 | -0.434 | 0.026 | 5.0119E-62 | 278.63 |
| *TRIAP1* | rs3755115 | C | A | 0.0244 | 0.163 | 0.024 | 1.5849E-11 | 46.13 |
| *TRIAP1* | rs9856296 | T | C | 0.2958 | -0.079 | 0.008 | 1.2589E-21 | 97.52 |
| *TRIAP1* | rs1354034 | T | C | 0.6041 | 0.085 | 0.008 | 2.5119E-29 | 112.89 |
| *TRIAP1* | rs62508038 | C | G | 0.0018 | -1.029 | 0.096 | 1E-26 | 114.89 |
| *TRIAP1* | rs7896518 | A | G | 0.4239 | 0.055 | 0.008 | 5.0119E-13 | 47.27 |
| *TRIAP1* | rs60822569 | T | TTTCTC | 0.5546 | 0.081 | 0.008 | 1.2589E-26 | 102.52 |
| *TRIAP1* | rs10418046 | T | G | 0.2115 | 0.094 | 0.009 | 1.2589E-24 | 109.09 |
| *TRIM21* | rs915652 | G | A | 0.1306 | -0.09 | 0.011 | 1.9953E-16 | 66.94 |
| *TRIM21* | rs199881122 | A | T | 0.0023 | -1.139 | 0.077 | 1E-49 | 218.81 |
| *TRIM24* | rs1354034 | T | C | 0.6041 | 0.091 | 0.008 | 1.2589E-32 | 129.39 |
| *TRIM24* | rs11502185 | T | C | 0.2596 | 0.069 | 0.01 | 1E-12 | 47.61 |
| *TRIM25* | rs2525992 | C | T | 0.7321 | -0.1 | 0.008 | 6.3096E-33 | 156.25 |
| *TRIM40* | rs3815079 | A | T | 0.2334 | -0.275 | 0.009 | 1E-200 | 933.64 |
| *TRIM5* | rs11821656 | G | C | 0.1401 | -0.408 | 0.011 | 1E-200 | 1375.74 |
| *TRIM58* | rs11204523 | G | C | 0.7585 | 0.226 | 0.009 | 1.585E-142 | 630.57 |
| *TRIM58* | rs342293 | C | G | 0.4598 | -0.051 | 0.007 | 6.3096E-12 | 53.08 |
| *TRIM58* | rs7896518 | A | G | 0.4239 | 0.054 | 0.008 | 1E-12 | 45.56 |
| *TSC1* | rs4548048 | C | T | 0.1033 | 0.123 | 0.013 | 6.3096E-21 | 89.52 |
| *TSC22D1* | rs1354034 | T | C | 0.6043 | 0.053 | 0.008 | 3.9811E-12 | 43.89 |
| *TSC22D1* | rs6866614 | A | G | 0.5736 | 0.052 | 0.008 | 7.9433E-12 | 42.25 |
| *TSC22D1* | rs11502185 | T | C | 0.2598 | 0.065 | 0.01 | 1.2589E-11 | 42.25 |
| *TSC22D1* | rs963569 | C | T | 0.7814 | -0.069 | 0.009 | 5.0119E-14 | 58.78 |
| *TSHB* | rs6426808 | G | A | 0.5121 | 0.088 | 0.008 | 3.1623E-31 | 121.00 |
| *TSHB* | rs334713 | A | C | 0.9558 | 0.141 | 0.018 | 1.9953E-14 | 61.36 |
| *TSHB* | rs141134245 | C | CA | 0.0839 | 0.096 | 0.014 | 1.9953E-12 | 47.02 |
| *TSHB* | rs2712172 | G | A | 0.2601 | -0.076 | 0.009 | 1.2589E-18 | 71.31 |
| *TSHB* | rs10028213 | C | G | 0.204 | -0.109 | 0.009 | 6.3096E-32 | 146.68 |
| *TSHB* | rs2046045 | T | G | 0.3948 | 0.15 | 0.008 | 7.9433E-84 | 351.56 |
| *TSHB* | rs11751601 | C | T | 0.3917 | 0.061 | 0.008 | 3.1623E-14 | 58.14 |
| *TSHB* | rs2396084 | A | G | 0.6892 | 0.077 | 0.008 | 5.0119E-21 | 92.64 |
| *TSHB* | rs2983511 | G | C | 0.3067 | -0.12 | 0.008 | 6.3096E-49 | 225.00 |
| *TSHB* | rs7032019 | G | A | 0.6703 | 0.056 | 0.008 | 3.1623E-12 | 49.00 |
| *TSHB* | rs7128207 | G | T | 0.5601 | 0.074 | 0.008 | 1.5849E-22 | 85.56 |
| *TSHB* | rs61938844 | G | A | 0.0259 | 0.169 | 0.023 | 7.9433E-13 | 53.99 |
| *TSHB* | rs116909374 | C | T | 0.0388 | -0.183 | 0.019 | 5.0119E-21 | 92.77 |
| *TSHB* | rs10519226 | T | C | 0.2453 | -0.071 | 0.009 | 2.5119E-16 | 62.23 |
| *TSHB* | rs200293726 | A | T | 0.3063 | -0.081 | 0.008 | 3.1623E-23 | 102.52 |
| *TSPAN1* | rs34463133 | G | A | 0.0033 | -0.449 | 0.061 | 1.2589E-13 | 54.18 |

| | | | | | | | | |
|---|---|---|---|---|---|---|---|---|
| *TSPAN1* | rs2268535 | G | A | 0.2367 | 0.061 | 0.008 | 1E-13 | 58.14 |
| *TSPAN1* | rs708686 | C | T | 0.2677 | 0.06 | 0.008 | 3.9811E-14 | 56.25 |
| *TSPAN15* | rs1227967 | T | C | 0.5011 | -0.056 | 0.008 | 6.3096E-13 | 49.00 |
| *TSPAN7* | rs9271176 | A | G | 0.6735 | -0.064 | 0.008 | 7.9433E-15 | 64.00 |
| *TSPAN8* | rs4263162 | C | G | 0.3336 | -0.125 | 0.007 | 1E-66 | 318.88 |
| *TSPAN8* | rs4144796 | T | C | 0.4178 | -0.048 | 0.007 | 6.3096E-12 | 47.02 |
| *TSPAN8* | rs1727952 | C | T | 0.852 | -0.073 | 0.01 | 5.0119E-14 | 53.29 |
| *TSPAN8* | rs939885 | G | A | 0.4858 | 0.067 | 0.007 | 1.5849E-22 | 91.61 |
| *TSPAN8* | rs9478784 | C | T | 0.0957 | 0.085 | 0.012 | 3.1623E-13 | 50.17 |
| *TSPAN8* | rs8176759 | G | A | 0.0615 | 0.13 | 0.014 | 6.3096E-20 | 86.22 |
| *TSPAN8* | rs11178654 | T | C | 0.4087 | 0.476 | 0.007 | 1E-200 | 4624.00 |
| *TSPAN8* | rs56398830 | G | A | 0.0111 | -0.479 | 0.033 | 1E-48 | 210.69 |
| *TSPAN8* | rs2297066 | C | G | 0.2417 | 0.056 | 0.008 | 3.9811E-12 | 49.00 |
| *TSPAN8* | rs1968109 | C | G | 0.3151 | 0.09 | 0.007 | 1.9953E-34 | 165.31 |
| *TSPAN8* | rs34353378 | A | C | 0.3299 | 0.12 | 0.007 | 3.9811E-61 | 293.88 |
| *TSPAN8* | rs708686 | C | T | 0.2677 | 0.08 | 0.008 | 5.0119E-25 | 100.00 |
| *TSPAN8* | rs681343 | C | T | 0.5092 | -0.18 | 0.007 | 7.943E-149 | 661.22 |
| *TSPYL1* | rs61746508 | C | A | 0.0166 | -0.412 | 0.03 | 1.5849E-43 | 188.60 |
| *TSPYL1* | rs7080386 | C | A | 0.4124 | 0.056 | 0.008 | 5.0119E-13 | 49.00 |
| *TST* | rs4821544 | T | C | 0.3033 | -0.108 | 0.008 | 2.5119E-38 | 182.25 |
| *TTF2* | rs75261221 | A | G | 0.0522 | 0.784 | 0.017 | 1E-200 | 2126.84 |
| *TTN* | rs534819727 | C | T | 0.2559 | 0.079 | 0.009 | 1E-16 | 77.05 |
| *TTR* | rs140584594 | A | G | 0.7324 | 0.069 | 0.008 | 6.3096E-18 | 74.39 |
| *TTR* | rs1260326 | T | C | 0.6082 | -0.053 | 0.007 | 1.9953E-13 | 57.33 |
| *TTR* | rs4841133 | A | G | 0.9081 | 0.145 | 0.012 | 3.1623E-32 | 146.01 |
| *TTR* | rs3999089 | A | G | 0.4694 | -0.059 | 0.007 | 7.9433E-17 | 71.04 |
| *TTR* | rs4575545 | G | A | 0.3053 | -0.106 | 0.008 | 7.9433E-43 | 175.56 |
| *TTR* | rs1667229 | T | C | 0.54 | 0.118 | 0.007 | 1E-60 | 284.16 |
| *TTR* | rs1883711 | G | C | 0.0309 | -0.27 | 0.021 | 1.9953E-38 | 165.31 |
| *TTR* | rs3747207 | G | A | 0.2153 | -0.06 | 0.009 | 1.9953E-12 | 44.44 |
| *TWF2* | rs7622851 | C | G | 0.4978 | 0.057 | 0.007 | 3.9811E-14 | 66.31 |
| *TWF2* | rs1354034 | T | C | 0.6046 | 0.058 | 0.008 | 3.9811E-14 | 52.56 |
| *TWF2* | rs6993770 | A | T | 0.2863 | -0.062 | 0.008 | 7.9433E-14 | 60.06 |
| *TXK* | rs6457621 | A | T | 0.1974 | -0.074 | 0.01 | 7.9433E-13 | 54.76 |
| *TXLNA* | rs1354034 | T | C | 0.6041 | 0.102 | 0.008 | 6.3096E-41 | 162.56 |
| *TXLNA* | rs342293 | C | G | 0.459 | -0.058 | 0.007 | 5.0119E-15 | 68.65 |
| *TXLNA* | rs11502185 | T | C | 0.2597 | 0.067 | 0.01 | 1.9953E-12 | 44.89 |
| *TXN* | rs146018216 | G | A | 0.0016 | -1.152 | 0.095 | 6.3096E-34 | 147.05 |
| *TXNDC15* | rs188468174 | C | T | 0.0143 | -0.517 | 0.027 | 3.9811E-82 | 366.65 |
| *TXNDC15* | rs115276619 | T | A | 0.0166 | 0.208 | 0.025 | 3.1623E-16 | 69.22 |
| *TXNDC15* | rs6054 | C | T | 0.0059 | 0.311 | 0.042 | 1.9953E-13 | 54.83 |
| *TXNDC15* | rs6876611 | A | T | 0.482 | -0.053 | 0.008 | 1.5849E-12 | 43.89 |
| *TXNDC15* | rs3733897 | A | G | 0.135 | 1.17 | 0.011 | 1E-200 | 11313.22 |
| *TXNDC15* | rs2524096 | G | T | 0.42 | -0.171 | 0.006 | 1.995E-154 | 812.25 |
| *TXNDC15* | rs73015965 | A | G | 0.0065 | 0.337 | 0.043 | 3.1623E-15 | 61.42 |
| *TXNDC15* | rs139130389 | C | CTA | 0.0726 | -0.19 | 0.012 | 1E-55 | 250.69 |
| *TXNDC15* | rs28929474 | C | T | 0.0206 | 0.233 | 0.022 | 3.9811E-26 | 112.17 |
| *TXNDC15* | rs4781137 | T | C | 0.4899 | -0.044 | 0.006 | 2.5119E-12 | 53.78 |
| *TXNDC15* | rs9302635 | T | C | 0.182 | -0.086 | 0.008 | 6.3096E-26 | 115.56 |
| *TXNDC15* | rs1801689 | A | C | 0.0302 | 0.509 | 0.018 | 3.162E-167 | 799.63 |
| *TXNDC5* | rs1354034 | T | C | 0.6042 | 0.072 | 0.008 | 5.0119E-21 | 81.00 |
| *TXNDC5* | rs7764128 | G | A | 0.1083 | -0.158 | 0.012 | 1E-39 | 173.36 |
| *TXNDC5* | rs6993770 | A | T | 0.2865 | -0.068 | 0.008 | 1E-16 | 72.25 |
| *TXNDC5* | rs4759076 | T | C | 0.4572 | 0.059 | 0.008 | 5.0119E-15 | 54.39 |
| *TXNDC9* | rs147732775 | G | A | 0.0111 | 0.251 | 0.037 | 7.9433E-12 | 46.02 |
| *TXNDC9* | rs1354034 | T | C | 0.6043 | 0.074 | 0.008 | 6.3096E-22 | 85.56 |
| *TXNL1* | rs71534539 | G | A | 0.4644 | -0.107 | 0.009 | 6.3096E-36 | 141.35 |
| *TXNRD1* | rs1354034 | T | C | 0.6041 | 0.054 | 0.008 | 1E-12 | 45.56 |
| *TXNRD1* | rs6993770 | A | T | 0.2864 | -0.068 | 0.008 | 1.2589E-16 | 72.25 |
| *TXNRD1* | rs10743940 | A | T | 0.9728 | -0.175 | 0.023 | 2.5119E-14 | 57.89 |
| *TXNRD1* | rs60822569 | T | TTTCTC | 0.5545 | 0.055 | 0.008 | 3.9811E-13 | 47.27 |
| *TXNRD1* | rs201402862 | G | A | 0.0008 | -1.167 | 0.144 | 5.0119E-16 | 65.68 |
| *TXNRD1* | rs12975366 | T | C | 0.3954 | -0.061 | 0.008 | 1.2589E-15 | 58.14 |
| *TYMP* | rs10774624 | G | A | 0.5137 | -0.056 | 0.008 | 1.9953E-13 | 49.00 |
| *TYMP* | rs131805 | T | C | 0.7827 | 0.225 | 0.009 | 3.162E-132 | 625.00 |
| *TYRO3* | rs10935473 | G | T | 0.4403 | -0.23 | 0.007 | 1E-200 | 1079.59 |
| *TYRO3* | rs2519093 | C | T | 0.1843 | -0.225 | 0.008 | 3.162E-158 | 791.02 |
| *TYRO3* | rs174530 | A | G | 0.3727 | 0.049 | 0.007 | 1.9953E-13 | 49.00 |

| | | | | | | | | |
|---|---|---|---|---|---|---|---|---|
| *TYRO3* | rs3967200 | C | T | 0.1344 | -0.074 | 0.009 | 5.0119E-15 | 67.60 |
| *TYRO3* | rs8024626 | G | A | 0.3018 | -0.646 | 0.008 | 1E-200 | 6520.56 |
| *TYRO3* | rs186021206 | G | A | 0.0064 | 0.96 | 0.044 | 1.995E-105 | 476.03 |
| *TYRO3* | rs2008174 | C | T | 0.7395 | -0.088 | 0.007 | 6.3096E-32 | 158.04 |
| *TYRP1* | rs1801274 | A | G | 0.5346 | -0.065 | 0.008 | 3.9811E-17 | 66.02 |
| *TYRP1* | rs28746784 | C | T | 0.1996 | 0.101 | 0.011 | 1.2589E-20 | 84.31 |
| *TYRP1* | rs10960751 | T | C | 0.6295 | -0.081 | 0.008 | 3.1623E-24 | 102.52 |
| *TYRP1* | rs7153844 | T | G | 0.2519 | -0.078 | 0.009 | 2.5119E-17 | 75.11 |
| *UBAC1* | rs112505519 | G | C | 0.0017 | -0.988 | 0.096 | 7.9433E-25 | 105.92 |
| *UBE2L6* | rs1354034 | T | C | 0.6045 | 0.081 | 0.007 | 3.9811E-27 | 133.90 |
| *UBE2L6* | rs6993770 | A | T | 0.2866 | -0.062 | 0.008 | 2.5119E-14 | 60.06 |
| *UBE2L6* | rs150230220 | A | AT | 0.3114 | 0.062 | 0.008 | 1.5849E-14 | 60.06 |
| *UBE2L6* | rs28362950 | G | A | 0.2674 | 0.254 | 0.008 | 1.995E-199 | 1008.06 |
| *UBE2L6* | rs10774624 | G | A | 0.5131 | -0.063 | 0.007 | 1.2589E-17 | 81.00 |
| *UBE2Z* | rs62404112 | T | C | 0.0564 | 0.119 | 0.017 | 1.2589E-12 | 49.00 |
| *UBXN1* | rs1354034 | T | C | 0.6044 | 0.071 | 0.008 | 1E-20 | 78.77 |
| *UBXN1* | rs342298 | C | T | 0.4549 | -0.056 | 0.008 | 1E-13 | 49.00 |
| *UBXN1* | rs142782985 | G | C | 0.0022 | 0.556 | 0.08 | 3.9811E-12 | 48.30 |
| *UFD1* | rs342298 | C | T | 0.455 | -0.055 | 0.008 | 2.5119E-13 | 47.27 |
| *UGDH* | rs607929 | C | G | 0.449 | -0.066 | 0.008 | 3.1623E-17 | 68.06 |
| *ULBP2* | rs4703854 | C | T | 0.7627 | 0.069 | 0.008 | 6.3096E-18 | 74.39 |
| *ULBP2* | rs6924387 | A | G | 0.4063 | -0.057 | 0.008 | 5.0119E-14 | 50.77 |
| *ULBP2* | rs60340208 | A | AG | 0.1492 | -0.676 | 0.011 | 1E-200 | 3776.66 |
| *ULBP2* | rs10100209 | C | T | 0.6093 | -0.07 | 0.007 | 3.1623E-23 | 100.00 |
| *ULBP2* | rs10104997 | C | T | 0.2096 | 0.116 | 0.008 | 5.0119E-44 | 210.25 |
| *ULBP2* | rs677355 | G | A | 0.3178 | 0.14 | 0.007 | 6.3096E-81 | 400.00 |
| *ULBP2* | rs34774456 | C | CAA | 0.7637 | 0.054 | 0.008 | 1.5849E-11 | 45.56 |
| *ULBP2* | rs4804181 | C | A | 0.7769 | -0.115 | 0.008 | 5.0119E-44 | 206.64 |
| *UMOD* | rs4846826 | C | T | 0.6288 | -0.041 | 0.006 | 5.0119E-12 | 46.69 |
| *UMOD* | rs12465018 | G | A | 0.1715 | -0.057 | 0.008 | 7.9433E-14 | 50.77 |
| *UMOD* | rs72819488 | G | A | 0.1558 | -0.062 | 0.008 | 5.0119E-15 | 60.06 |
| *UMOD* | rs6542126 | C | A | 0.1628 | 0.054 | 0.008 | 2.5119E-12 | 45.56 |
| *UMOD* | rs6793835 | G | A | 0.2661 | 0.046 | 0.006 | 1E-12 | 58.78 |
| *UMOD* | rs56012466 | G | A | 0.2724 | 0.068 | 0.006 | 3.1623E-26 | 128.44 |
| *UMOD* | rs13288823 | C | T | 0.0534 | -0.094 | 0.013 | 1.5849E-13 | 52.28 |
| *UMOD* | rs774882452 | CT | C | 0.3443 | -0.043 | 0.006 | 5.0119E-13 | 51.36 |
| *UMOD* | rs77713116 | C | G | 0.3419 | -0.084 | 0.006 | 1.9953E-43 | 196.00 |
| *UMOD* | rs12368376 | C | G | 0.1707 | -0.064 | 0.008 | 3.9811E-17 | 64.00 |
| *UMOD* | rs11172134 | T | A | 0.2055 | 0.052 | 0.007 | 1.5849E-13 | 55.18 |
| *UMOD* | rs10851885 | A | G | 0.257 | -0.05 | 0.007 | 3.1623E-14 | 51.02 |
| *UMOD* | rs12922822 | C | T | 0.1817 | -0.969 | 0.009 | 1E-200 | 11592.11 |
| *UMOD* | rs186021206 | G | A | 0.0064 | 0.295 | 0.04 | 1.5849E-13 | 54.39 |
| *UMOD* | rs2934951 | A | G | 0.6829 | -0.043 | 0.006 | 6.3096E-12 | 51.36 |
| *UMOD* | rs72835417 | G | A | 0.1098 | 0.247 | 0.009 | 3.162E-156 | 753.20 |
| *UMOD* | rs896642 | C | T | 0.2224 | 0.047 | 0.007 | 7.9433E-12 | 45.08 |
| *UMOD* | rs1997596 | C | T | 0.3421 | 0.046 | 0.006 | 6.3096E-14 | 58.78 |
| *UNC5D* | rs13260977 | A | G | 0.4179 | -0.064 | 0.008 | 2.5119E-16 | 64.00 |
| *UNC79* | rs953492 | A | G | 0.5337 | 0.126 | 0.008 | 1E-59 | 248.06 |
| *UPB1* | rs1260326 | T | C | 0.6086 | -0.054 | 0.008 | 6.3096E-13 | 45.56 |
| *UPB1* | rs10119644 | T | A | 0.4968 | -0.061 | 0.007 | 3.1623E-16 | 75.94 |
| *UPB1* | rs143493067 | G | A | 0.0017 | -0.859 | 0.09 | 1.2589E-21 | 91.10 |
| *UPB1* | rs3747207 | G | A | 0.2157 | 0.089 | 0.009 | 5.0119E-23 | 97.79 |
| *UROD* | rs12749939 | C | T | 0.2583 | -0.477 | 0.008 | 1E-200 | 3555.14 |
| *UROD* | rs75214378 | G | A | 0.0417 | 0.155 | 0.018 | 7.9433E-18 | 74.15 |
| *UROD* | rs78564552 | C | T | 0.0037 | 0.47 | 0.059 | 1.2589E-15 | 63.46 |
| *UROD* | rs1354034 | T | C | 0.6044 | 0.078 | 0.007 | 1E-26 | 124.16 |
| *UROD* | rs2393967 | A | C | 0.314 | 0.055 | 0.008 | 1E-12 | 47.27 |
| *UROS* | rs57467915 | G | A | 0.015 | 0.275 | 0.031 | 1.2589E-18 | 78.69 |
| *UROS* | rs10794023 | A | C | 0.7938 | -0.138 | 0.009 | 3.1623E-48 | 235.11 |
| *USO1* | rs1354034 | T | C | 0.6046 | 0.062 | 0.008 | 3.1623E-16 | 60.06 |
| *USO1* | rs342298 | C | T | 0.4545 | -0.055 | 0.008 | 1.9953E-13 | 47.27 |
| *USO1* | rs6993770 | A | T | 0.2864 | -0.069 | 0.008 | 1E-16 | 74.39 |
| *USP25* | rs1354034 | T | C | 0.6044 | 0.08 | 0.008 | 1.9953E-26 | 100.00 |
| *USP25* | rs342298 | C | T | 0.455 | -0.05 | 0.007 | 1.2589E-11 | 51.02 |
| *USP28* | rs4288784 | C | G | 0.077 | -0.153 | 0.014 | 1.5849E-26 | 119.43 |
| *USP47* | rs5751547 | T | C | 0.5916 | 0.093 | 0.008 | 1.2589E-31 | 135.14 |
| *USP8* | rs78909033 | G | A | 0.1361 | -0.187 | 0.011 | 3.1623E-65 | 289.00 |
| *USP8* | rs1354034 | T | C | 0.6038 | 0.068 | 0.008 | 6.3096E-19 | 72.25 |

| | | | | | | | | |
|---|---|---|---|---|---|---|---|---|
| *USP8* | rs342298 | C | T | 0.455 | -0.057 | 0.008 | 2.5119E-14 | 50.77 |
| *USP8* | rs4380013 | G | A | 0.1964 | 0.091 | 0.009 | 6.3096E-22 | 102.23 |
| *UXS1* | rs660240 | T | C | 0.7854 | 0.062 | 0.009 | 1.2589E-11 | 47.46 |
| *UXS1* | rs138588539 | C | CT | 0.0781 | 0.175 | 0.014 | 1.5849E-35 | 156.25 |
| *UXS1* | rs68066031 | T | C | 0.2293 | -0.153 | 0.009 | 3.9811E-64 | 289.00 |
| *UXS1* | rs710446 | T | C | 0.4091 | -0.055 | 0.008 | 6.3096E-13 | 47.27 |
| *UXS1* | rs71640036 | G | T | 0.5057 | -0.127 | 0.008 | 1E-61 | 252.02 |
| *UXS1* | rs2731673 | T | C | 0.7444 | -0.068 | 0.009 | 3.1623E-15 | 57.09 |
| *UXS1* | rs1149607 | C | A | 0.1821 | -0.238 | 0.01 | 3.162E-129 | 566.44 |
| *VAMP5* | rs6547623 | T | A | 0.2995 | 0.084 | 0.008 | 1E-23 | 110.25 |
| *VAMP5* | rs505922 | T | C | 0.3165 | 0.062 | 0.008 | 3.1623E-14 | 60.06 |
| *VAMP8* | rs3731828 | C | T | 0.2831 | -0.173 | 0.008 | 1E-93 | 467.64 |
| *VASH1* | rs1354034 | T | C | 0.6043 | 0.103 | 0.008 | 6.3096E-41 | 165.77 |
| *VASH1* | rs114694170 | T | C | 0.0598 | 0.13 | 0.016 | 5.0119E-16 | 66.02 |
| *VASH1* | rs11502185 | T | C | 0.2598 | 0.071 | 0.01 | 2.5119E-13 | 50.41 |
| *VASH1* | rs11159226 | T | G | 0.0597 | 0.21 | 0.016 | 1.2589E-39 | 172.27 |
| *VASN* | rs150816167 | T | C | 0.0421 | 0.129 | 0.018 | 1E-12 | 51.36 |
| *VASN* | rs1260326 | T | C | 0.6085 | 0.064 | 0.007 | 1.9953E-18 | 83.59 |
| *VASN* | rs4841132 | A | G | 0.9082 | -0.097 | 0.012 | 5.0119E-15 | 65.34 |
| *VASN* | rs7896518 | A | G | 0.4242 | 0.053 | 0.007 | 3.1623E-13 | 57.33 |
| *VASN* | rs35713275 | G | A | 0.0631 | -0.904 | 0.016 | 1E-200 | 3192.25 |
| *VASN* | rs77542162 | A | G | 0.0228 | 0.187 | 0.024 | 5.0119E-15 | 60.71 |
| *VASP* | rs1354034 | T | C | 0.6042 | 0.057 | 0.008 | 6.3096E-14 | 50.77 |
| *VASP* | rs342299 | C | T | 0.4549 | -0.064 | 0.008 | 1.9953E-17 | 64.00 |
| *VAT1* | rs1047891 | C | A | 0.3125 | -0.065 | 0.008 | 7.9433E-16 | 66.02 |
| *VAT1* | rs11787335 | C | T | 0.3568 | 0.054 | 0.008 | 6.3096E-12 | 45.56 |
| *VAT1* | rs11379524 | A | AC | 0.6304 | 0.076 | 0.008 | 2.5119E-22 | 90.25 |
| *VAT1* | rs4149307 | C | T | 0.1532 | 0.095 | 0.01 | 5.0119E-20 | 90.25 |
| *VAT1* | rs190543502 | T | C | 0.0231 | -0.22 | 0.025 | 1.5849E-18 | 77.44 |
| *VAT1* | rs173539 | C | T | 0.3316 | 0.057 | 0.008 | 1.2589E-12 | 50.77 |
| *VAT1* | rs200561116 | A | AT | 0.116 | 0.103 | 0.012 | 5.0119E-18 | 73.67 |
| *VAT1* | rs7210098 | C | T | 0.3354 | -0.141 | 0.008 | 7.9433E-70 | 310.64 |
| *VAT1* | rs2868346 | C | T | 0.7595 | -0.152 | 0.009 | 1.9953E-68 | 285.23 |
| *VAV3* | rs345308 | G | A | 0.7909 | -0.065 | 0.009 | 3.1623E-12 | 52.16 |
| *VAV3* | rs1354034 | T | C | 0.6045 | 0.053 | 0.008 | 6.3096E-12 | 43.89 |
| *VCAM1* | rs139561173 | A | C | 0.2113 | -0.105 | 0.009 | 6.3096E-31 | 136.11 |
| *VCAM1* | rs61801010 | A | G | 0.1063 | 0.126 | 0.013 | 7.9433E-24 | 93.94 |
| *VCAM1* | rs2009581 | G | A | 0.2723 | -0.059 | 0.008 | 1E-12 | 54.39 |
| *VCAM1* | rs1257169 | C | A | 0.4987 | -0.11 | 0.007 | 6.3096E-50 | 246.94 |
| *VCAM1* | rs35723031 | G | GT | 0.5579 | -0.057 | 0.007 | 1.2589E-14 | 66.31 |
| *VCAM1* | rs28418426 | T | C | 0.5336 | 0.063 | 0.008 | 1E-14 | 62.02 |
| *VCAM1* | rs8176693 | C | T | 0.0614 | 0.146 | 0.015 | 7.9433E-22 | 94.74 |
| *VCAM1* | rs1070073 | T | G | 0.6721 | 0.072 | 0.008 | 5.0119E-20 | 81.00 |
| *VCAM1* | rs3184504 | T | C | 0.5172 | -0.141 | 0.007 | 5.0119E-82 | 405.73 |
| *VCAM1* | rs186021206 | G | A | 0.0064 | 0.858 | 0.05 | 7.9433E-65 | 294.47 |
| *VCAM1* | rs34557412 | A | G | 0.0068 | 0.328 | 0.045 | 1.9953E-13 | 53.13 |
| *VCAM1* | rs2092501 | G | A | 0.1518 | 0.07 | 0.01 | 1E-11 | 49.00 |
| *VCAN* | rs3754053 | C | G | 0.5759 | 0.052 | 0.007 | 1E-14 | 55.18 |
| *VCAN* | rs34552760 | A | G | 0.6237 | -0.094 | 0.007 | 3.9811E-44 | 180.33 |
| *VCAN* | rs12491835 | T | C | 0.2358 | -0.053 | 0.008 | 5.0119E-12 | 43.89 |
| *VCAN* | rs1229984 | T | C | 0.9774 | -0.268 | 0.022 | 3.1623E-34 | 148.40 |
| *VCAN* | rs13107325 | C | T | 0.0747 | 0.153 | 0.012 | 1E-34 | 162.56 |
| *VCAN* | rs309559 | A | G | 0.4813 | -0.649 | 0.007 | 1E-200 | 8595.94 |
| *VCAN* | rs1917368 | G | T | 0.3923 | -0.051 | 0.007 | 1E-14 | 53.08 |
| *VCAN* | rs11369732 | G | GT | 0.5173 | 0.062 | 0.007 | 5.0119E-21 | 78.45 |
| *VCAN* | rs12676683 | C | A | 0.0932 | -0.21 | 0.011 | 3.9811E-78 | 364.46 |
| *VCAN* | rs2945779 | C | T | 0.3704 | 0.05 | 0.007 | 1.9953E-13 | 51.02 |
| *VCAN* | rs2435381 | C | T | 0.2747 | 0.088 | 0.007 | 6.3096E-34 | 158.04 |
| *VCAN* | rs3138189 | T | C | 0.0633 | -0.095 | 0.013 | 5.0119E-13 | 53.40 |
| *VCAN* | rs112680434 | A | AGAATGT | 0.3651 | -0.058 | 0.007 | 6.3096E-18 | 68.65 |
| *VCAN* | rs200489612 | G | A | 0.0052 | 0.388 | 0.049 | 1.9953E-15 | 62.70 |
| *VCAN* | rs2665397 | C | A | 0.5441 | 0.056 | 0.006 | 3.9811E-18 | 87.11 |
| *VCAN* | rs55639531 | C | G | 0.2854 | 0.059 | 0.007 | 2.5119E-16 | 71.04 |
| *VCPKMT* | rs142528336 | A | C | 0.0097 | -0.395 | 0.04 | 1E-22 | 97.52 |
| *VCPKMT* | rs3814952 | G | A | 0.2263 | -0.069 | 0.01 | 1.5849E-11 | 47.61 |
| *VEGFA* | rs2274319 | T | C | 0.6529 | -0.055 | 0.007 | 3.9811E-16 | 61.73 |
| *VEGFA* | rs114694170 | T | C | 0.06 | 0.167 | 0.014 | 6.3096E-35 | 142.29 |
| *VEGFA* | rs6921438 | G | A | 0.4904 | -0.688 | 0.007 | 1E-200 | 9660.08 |

| | | | | | | | | |
|---|---|---|---|---|---|---|---|---|
| *VEGFA* | rs13236689 | T | G | 0.4072 | 0.05 | 0.006 | 1.2589E-14 | 69.44 |
| *VEGFA* | rs6993770 | A | T | 0.2866 | -0.224 | 0.007 | 1E-200 | 1024.00 |
| *VEGFA* | rs2375981 | C | G | 0.4438 | -0.19 | 0.007 | 6.31E-186 | 736.73 |
| *VEGFA* | rs10761731 | A | T | 0.4145 | 0.123 | 0.006 | 1.5849E-79 | 420.25 |
| *VEGFA* | rs3184504 | T | C | 0.5173 | -0.046 | 0.006 | 3.1623E-13 | 58.78 |
| *VEGFA* | rs12445050 | C | T | 0.1381 | 0.071 | 0.009 | 1E-14 | 62.23 |
| *VEGFA* | rs34416903 | T | C | 0.2796 | 0.084 | 0.007 | 6.3096E-31 | 144.00 |
| *VEGFA* | rs892090 | T | G | 0.8342 | 0.104 | 0.009 | 2.5119E-33 | 133.53 |
| *VEGFA* | rs3790176 | G | A | 0.3425 | 0.054 | 0.007 | 7.9433E-16 | 59.51 |
| *VEGFB* | rs143543800 | C | T | 0.003 | 0.959 | 0.068 | 1.5849E-44 | 198.89 |
| *VEGFB* | rs188468174 | C | T | 0.0141 | -0.538 | 0.032 | 3.1623E-62 | 282.66 |
| *VEGFB* | rs2305669 | G | A | 0.5076 | -0.072 | 0.008 | 1.9953E-21 | 81.00 |
| *VEGFB* | rs7040440 | C | T | 0.0809 | -0.146 | 0.015 | 1.2589E-23 | 94.74 |
| *VEGFB* | rs757081 | C | G | 0.3322 | -0.068 | 0.008 | 2.5119E-17 | 72.25 |
| *VEGFB* | rs660442 | G | A | 0.1996 | 0.115 | 0.009 | 3.9811E-34 | 163.27 |
| *VEGFC* | rs7618405 | C | A | 0.2047 | -0.067 | 0.009 | 6.3096E-13 | 55.42 |
| *VEGFC* | rs1354034 | T | C | 0.6041 | -0.071 | 0.008 | 6.3096E-21 | 78.77 |
| *VEGFC* | rs10016018 | A | T | 0.3974 | -0.064 | 0.008 | 3.1623E-17 | 64.00 |
| *VEGFC* | rs35029317 | A | G | 0.006 | -0.797 | 0.049 | 7.9433E-59 | 264.56 |
| *VEGFC* | rs11242109 | G | T | 0.4811 | -0.052 | 0.007 | 3.1623E-12 | 55.18 |
| *VEGFC* | rs35173808 | C | A | 0.0964 | -0.098 | 0.013 | 1.2589E-14 | 56.83 |
| *VEGFC* | rs6961069 | C | T | 0.4023 | 0.066 | 0.008 | 6.3096E-18 | 68.06 |
| *VEGFC* | rs6993770 | A | T | 0.2861 | -0.088 | 0.008 | 1.5849E-26 | 121.00 |
| *VEGFC* | rs71474568 | T | TTG | 0.3401 | 0.08 | 0.008 | 6.3096E-24 | 100.00 |
| *VEGFC* | rs73000929 | G | A | 0.0373 | -0.138 | 0.02 | 2.5119E-12 | 47.61 |
| *VEGFC* | rs59001897 | T | A | 0.1749 | 0.071 | 0.01 | 1E-12 | 50.41 |
| *VEGFC* | rs12445050 | C | T | 0.1384 | 0.105 | 0.011 | 1.9953E-22 | 91.12 |
| *VEGFC* | rs1654425 | T | C | 0.8345 | 0.163 | 0.01 | 1E-58 | 265.69 |
| *VEGFC* | rs6081555 | G | T | 0.3434 | 0.07 | 0.008 | 7.9433E-19 | 76.56 |
| *VEGFD* | rs710446 | T | C | 0.4089 | 0.06 | 0.008 | 1.2589E-15 | 56.25 |
| *VEGFD* | rs12331618 | A | G | 0.5123 | 0.142 | 0.007 | 5.0119E-80 | 411.51 |
| *VEGFD* | rs3217225 | TCTC | T | 0.1663 | -0.08 | 0.01 | 1E-15 | 64.00 |
| *VEGFD* | rs75077631 | G | GC | 0.7422 | 0.083 | 0.008 | 7.9433E-23 | 107.64 |
| *VEGFD* | rs10982164 | G | A | 0.0671 | -0.123 | 0.016 | 1.2589E-14 | 59.10 |
| *VEGFD* | rs56278466 | T | G | 0.6613 | 0.069 | 0.008 | 1.2589E-18 | 74.39 |
| *VEGFD* | rs1037117 | G | A | 0.2523 | -0.081 | 0.009 | 2.5119E-21 | 81.00 |
| *VGF* | rs773642372 | ACTCTCCTC | A | 0.0732 | -0.125 | 0.015 | 2.5119E-17 | 69.44 |
| *VIT* | rs4233367 | T | C | 0.6092 | -0.136 | 0.006 | 3.162E-115 | 513.78 |
| *VIT* | rs1468810 | C | A | 0.5862 | -0.432 | 0.007 | 1E-200 | 3808.65 |
| *VIT* | rs2577831 | C | A | 0.4787 | -0.045 | 0.006 | 7.9433E-15 | 56.25 |
| *VIT* | rs13107325 | C | T | 0.0751 | 0.113 | 0.011 | 1.9953E-24 | 105.53 |
| *VIT* | rs72083329 | C | CT | 0.2891 | -0.049 | 0.007 | 5.0119E-14 | 49.00 |
| *VIT* | rs2142306 | T | C | 0.4122 | 0.105 | 0.006 | 6.3096E-70 | 306.25 |
| *VIT* | rs1920719 | C | T | 0.0642 | 0.106 | 0.012 | 2.5119E-19 | 78.03 |
| *VIT* | rs186021206 | G | A | 0.0064 | 0.612 | 0.04 | 5.0119E-53 | 234.09 |
| *VIT* | rs704 | G | A | 0.4733 | -0.05 | 0.006 | 6.3096E-18 | 69.44 |
| *VIT* | rs2267372 | A | G | 0.5999 | 0.044 | 0.006 | 5.0119E-14 | 53.78 |
| *VMO1* | rs117779442 | C | T | 0.0743 | -0.073 | 0.011 | 1.5849E-11 | 44.04 |
| *VMO1* | rs112875651 | G | A | 0.3909 | -0.043 | 0.006 | 1.9953E-13 | 51.36 |
| *VMO1* | rs964184 | G | C | 0.8668 | -0.086 | 0.008 | 1.5849E-24 | 115.56 |
| *VMO1* | rs112597211 | T | C | 0.1364 | -0.11 | 0.008 | 1.2589E-39 | 189.06 |
| *VMO1* | rs261290 | T | C | 0.6557 | -0.127 | 0.006 | 6.3096E-97 | 448.03 |
| *VMO1* | rs8078118 | C | A | 0.4167 | 0.755 | 0.008 | 1E-200 | 8906.64 |
| *VMO1* | rs3826688 | T | C | 0.6408 | 0.047 | 0.006 | 3.9811E-15 | 61.36 |
| *VNN1* | rs1260326 | T | C | 0.6086 | -0.054 | 0.005 | 7.9433E-24 | 116.64 |
| *VNN1* | rs199918445 | TA | T | 0.7044 | 0.051 | 0.006 | 5.0119E-19 | 72.25 |
| *VNN1* | rs146101841 | C | CATAGACA' | 0.2566 | -1.001 | 0.009 | 1E-200 | 12370.38 |
| *VNN1* | rs13240065 | G | A | 0.1271 | 0.054 | 0.008 | 5.0119E-12 | 45.56 |
| *VNN1* | rs4240624 | G | A | 0.908 | -0.082 | 0.009 | 2.5119E-19 | 83.01 |
| *VNN1* | rs9414801 | G | A | 0.5332 | -0.038 | 0.005 | 3.9811E-13 | 57.76 |
| *VNN1* | rs174544 | C | A | 0.3087 | 0.048 | 0.006 | 3.9811E-17 | 64.00 |
| *VNN1* | rs147233090 | C | T | 0.0236 | 0.14 | 0.017 | 6.3096E-16 | 67.82 |
| *VNN1* | rs174418 | T | C | 0.5976 | 0.059 | 0.005 | 3.9811E-28 | 139.24 |
| *VNN1* | rs429358 | T | C | 0.1557 | -0.081 | 0.007 | 1E-28 | 133.90 |
| *VNN1* | rs11668882 | T | C | 0.4412 | 0.049 | 0.005 | 1.5849E-20 | 96.04 |
| *VNN2* | rs7583027 | A | C | 0.6434 | 0.046 | 0.007 | 5.0119E-12 | 43.18 |
| *VNN2* | rs9493425 | A | G | 0.3231 | -0.697 | 0.008 | 1E-200 | 7590.77 |
| *VNN2* | rs56278466 | T | G | 0.6611 | 0.067 | 0.007 | 3.9811E-23 | 91.61 |

| | | | | | | | | |
|---|---|---|---|---|---|---|---|---|
| *VNN2* | rs603424 | G | A | 0.1663 | -0.063 | 0.009 | 3.9811E-13 | 49.00 |
| *VNN2* | rs2511241 | C | T | 0.9297 | 0.177 | 0.013 | 1E-44 | 185.38 |
| *VNN2* | rs73017399 | G | C | 0.2391 | 0.074 | 0.008 | 1.2589E-22 | 85.56 |
| *VNN2* | rs111338191 | A | AAATT | 0.5208 | -0.045 | 0.007 | 7.9433E-12 | 41.33 |
| *VNN2* | rs146261845 | C | T | 0.0068 | 0.296 | 0.042 | 1.9953E-12 | 49.67 |
| *VNN2* | rs7208815 | C | T | 0.3922 | 0.101 | 0.007 | 1.9953E-52 | 208.18 |
| *VNN2* | rs4760 | A | G | 0.1559 | -0.184 | 0.009 | 1.5849E-95 | 417.98 |
| *VPS28* | rs72797306 | G | C | 0.4296 | -0.055 | 0.008 | 6.3096E-13 | 47.27 |
| *VPS28* | rs4756952 | C | T | 0.3354 | 0.085 | 0.008 | 1.5849E-26 | 112.89 |
| *VPS37A* | rs1354034 | T | C | 0.6046 | 0.053 | 0.008 | 3.9811E-12 | 43.89 |
| *VPS37A* | rs964184 | G | C | 0.8668 | -0.08 | 0.011 | 3.1623E-13 | 52.89 |
| *VPS4B* | rs1354034 | T | C | 0.6044 | 0.059 | 0.008 | 1.9953E-14 | 54.39 |
| *VPS4B* | rs7916868 | A | T | 0.5014 | 0.055 | 0.008 | 1.9953E-13 | 47.27 |
| *VPS53* | rs1354034 | T | C | 0.6046 | 0.055 | 0.008 | 3.9811E-13 | 47.27 |
| *VPS53* | rs342296 | G | A | 0.4563 | -0.053 | 0.007 | 1.9953E-12 | 57.33 |
| *VSIG10* | rs2519093 | C | T | 0.1844 | -0.129 | 0.008 | 3.9811E-54 | 260.02 |
| *VSIG10* | rs7307331 | G | A | 0.2965 | 0.779 | 0.008 | 1E-200 | 9481.89 |
| *VSIG10* | rs200489612 | G | A | 0.0053 | 0.394 | 0.048 | 1.9953E-16 | 67.38 |
| *VSIG10L* | rs1035454 | T | C | 0.5759 | 0.208 | 0.008 | 1E-154 | 676.00 |
| *VSIG2* | rs4556017 | C | T | 0.8535 | -0.088 | 0.01 | 7.9433E-18 | 77.44 |
| *VSIG2* | rs8176741 | G | A | 0.0612 | 0.21 | 0.015 | 2.5119E-44 | 196.00 |
| *VSIG2* | rs6591649 | A | G | 0.3447 | 0.137 | 0.008 | 5.0119E-71 | 293.27 |
| *VSIG2* | rs11219769 | G | T | 0.2559 | 0.213 | 0.008 | 1.259E-142 | 708.89 |
| *VSIG2* | rs708686 | C | T | 0.2676 | 0.126 | 0.008 | 1.5849E-54 | 248.06 |
| *VSIG2* | rs516246 | C | T | 0.509 | -0.083 | 0.007 | 3.9811E-30 | 140.59 |
| *VSIG4* | rs551397 | C | T | 0.2298 | -0.066 | 0.009 | 1.5849E-14 | 53.78 |
| *VSIG4* | rs11130306 | A | G | 0.4811 | 0.049 | 0.007 | 1.5849E-11 | 49.00 |
| *VSIG4* | rs12667978 | C | T | 0.571 | 0.049 | 0.007 | 1.2589E-11 | 49.00 |
| *VSIG4* | rs10769256 | C | T | 0.4002 | -0.102 | 0.007 | 1.5849E-43 | 212.33 |
| *VSIG4* | rs552431601 | G | A | 0.0088 | 0.284 | 0.041 | 6.3096E-12 | 47.98 |
| *VSIG4* | rs28640218 | G | T | 0.1914 | -0.064 | 0.009 | 3.9811E-12 | 50.57 |
| *VSIG4* | rs565238977 | CAAAAAAA | C | 0.8317 | -0.069 | 0.01 | 1.2589E-12 | 47.61 |
| *VSIG4* | rs2230199 | G | C | 0.2166 | 0.067 | 0.009 | 5.0119E-14 | 55.42 |
| *VSIR* | rs13108400 | A | G | 0.3974 | -0.054 | 0.008 | 1E-12 | 45.56 |
| *VSIR* | rs11242109 | G | T | 0.4815 | -0.06 | 0.007 | 3.9811E-16 | 73.47 |
| *VSIR* | rs6961069 | C | T | 0.4024 | 0.057 | 0.008 | 5.0119E-14 | 50.77 |
| *VSIR* | rs342299 | C | T | 0.455 | -0.058 | 0.007 | 3.1623E-15 | 68.65 |
| *VSIR* | rs6993770 | A | T | 0.2866 | -0.122 | 0.008 | 1E-50 | 232.56 |
| *VSIR* | rs10999992 | T | C | 0.1469 | 0.44 | 0.011 | 1E-200 | 1600.00 |
| *VSIR* | rs1362213 | C | T | 0.4795 | -0.05 | 0.007 | 1.5849E-11 | 51.02 |
| *VSIR* | rs1654425 | T | C | 0.834 | 0.104 | 0.01 | 1.5849E-25 | 108.16 |
| *VSNL1* | rs446947 | A | T | 0.2764 | 0.313 | 0.008 | 1E-200 | 1530.77 |
| *VSNL1* | rs5952 | A | G | 0.0051 | 1.357 | 0.048 | 3.162E-176 | 799.24 |
| *VSNL1* | rs112875651 | G | A | 0.3912 | -0.047 | 0.007 | 6.3096E-12 | 45.08 |
| *VSNL1* | rs113609637 | A | AT | 0.1416 | -0.069 | 0.01 | 3.9811E-13 | 47.61 |
| *VSNL1* | rs4926 | G | A | 0.274 | -0.06 | 0.007 | 3.9811E-16 | 73.47 |
| *VSNL1* | rs2289125 | A | C | 0.7891 | 0.061 | 0.008 | 5.0119E-14 | 58.14 |
| *VSNL1* | rs10405357 | T | C | 0.4355 | -0.05 | 0.007 | 7.9433E-14 | 51.02 |
| *VSTM1* | rs60816814 | C | A | 0.0446 | 0.183 | 0.016 | 1.5849E-31 | 130.82 |
| *VSTM1* | rs2433724 | T | C | 0.4291 | -0.768 | 0.008 | 1E-200 | 9216.00 |
| *VSTM1* | rs189448562 | G | C | 0.0111 | 0.225 | 0.029 | 6.3096E-15 | 60.20 |
| *VSTM2L* | rs646776 | C | T | 0.7777 | 0.209 | 0.009 | 1.585E-113 | 539.27 |
| *VSTM2L* | rs2273349 | C | A | 0.1836 | -0.146 | 0.01 | 1.9953E-48 | 213.16 |
| *VTA1* | rs1354034 | T | C | 0.6042 | 0.075 | 0.008 | 1E-22 | 87.89 |
| *VTA1* | rs10900809 | G | A | 0.3885 | -0.058 | 0.008 | 3.9811E-14 | 52.56 |
| *VTA1* | rs10733789 | T | C | 0.3133 | 0.058 | 0.008 | 7.9433E-13 | 52.56 |
| *VTA1* | rs60822569 | T | TTTCTC | 0.5544 | 0.055 | 0.008 | 3.9811E-13 | 47.27 |
| *VTCN1* | rs56174814 | T | A | 0.1567 | 0.108 | 0.011 | 6.3096E-24 | 96.40 |
| *VTCN1* | rs516316 | G | C | 0.509 | 0.078 | 0.008 | 1E-23 | 95.06 |
| *VTI1A* | rs1354034 | T | C | 0.6044 | 0.082 | 0.008 | 5.0119E-27 | 105.06 |
| *VTI1A* | rs342299 | C | T | 0.4549 | -0.053 | 0.008 | 1.5849E-12 | 43.89 |
| *VTI1A* | rs6993770 | A | T | 0.287 | -0.073 | 0.008 | 1.5849E-18 | 83.27 |
| *VTI1A* | rs7896518 | A | G | 0.4239 | 0.056 | 0.008 | 3.1623E-13 | 49.00 |
| *VTI1A* | rs79671623 | G | A | 0.1183 | -0.086 | 0.012 | 1.2589E-13 | 51.36 |
| *VWA1* | rs115503338 | G | A | 0.0137 | -1.494 | 0.033 | 1E-200 | 2049.62 |
| *VWA1* | rs35457250 | C | T | 0.0089 | 0.525 | 0.039 | 2.5119E-41 | 181.21 |
| *VWA1* | rs6054 | C | T | 0.0059 | 0.398 | 0.049 | 5.0119E-16 | 65.97 |
| *VWA1* | rs28929474 | C | T | 0.0207 | 0.24 | 0.026 | 7.9433E-21 | 85.21 |

| | | | | | | | | |
|---|---|---|---|---|---|---|---|---|
| *VWA1* | rs1801689 | A | C | 0.03 | 0.265 | 0.021 | 3.9811E-35 | 159.24 |
| *VWA1* | rs7256200 | G | T | 0.1277 | -0.085 | 0.011 | 1.2589E-14 | 59.71 |
| *VWC2* | rs61747728 | C | T | 0.0379 | 0.129 | 0.017 | 1.9953E-13 | 57.58 |
| *VWC2* | rs141154056 | C | T | 0.0018 | 1.802 | 0.08 | 1.585E-112 | 507.38 |
| *VWC2* | rs769604 | C | G | 0.4593 | -0.458 | 0.007 | 1E-200 | 4280.90 |
| *VWC2L* | rs10932542 | G | A | 0.2412 | 0.099 | 0.009 | 1.9953E-30 | 121.00 |
| *VWF* | rs9386182 | A | T | 0.5581 | 0.065 | 0.007 | 3.9811E-18 | 86.22 |
| *VWF* | rs2726953 | G | A | 0.2953 | 0.058 | 0.008 | 7.9433E-13 | 52.56 |
| *VWF* | rs505922 | T | C | 0.3169 | 0.318 | 0.008 | 1E-200 | 1580.06 |
| *VWF* | rs68120218 | G | A | 0.3449 | 0.098 | 0.008 | 3.9811E-36 | 150.06 |
| *VWF* | rs3751198 | A | G | 0.6039 | 0.06 | 0.008 | 1.5849E-15 | 56.25 |
| *WARS* | rs7137828 | C | T | 0.5164 | -0.075 | 0.007 | 6.3096E-24 | 114.80 |
| *WARS* | rs2273804 | C | G | 0.2599 | -0.258 | 0.009 | 1.995E-199 | 821.78 |
| *WAS* | rs5030082 | A | G | 0.4049 | 0.062 | 0.008 | 1.2589E-15 | 60.06 |
| *WAS* | rs4861708 | G | A | 0.5118 | 0.113 | 0.008 | 2.5119E-49 | 199.52 |
| *WAS* | rs1801020 | A | G | 0.7447 | 0.087 | 0.009 | 2.5119E-23 | 93.44 |
| *WASF1* | rs1980532 | A | G | 0.4454 | -0.287 | 0.008 | 1E-200 | 1287.02 |
| *WASF1* | rs7916868 | A | T | 0.5016 | 0.05 | 0.007 | 1.5849E-11 | 51.02 |
| *WASF3* | rs9512298 | C | G | 0.5555 | 0.133 | 0.008 | 5.0119E-66 | 276.39 |
| *WASHC3* | rs1354034 | T | C | 0.6043 | 0.055 | 0.008 | 5.0119E-13 | 47.27 |
| *WASHC3* | rs342293 | C | G | 0.46 | -0.058 | 0.007 | 7.9433E-15 | 68.65 |
| *WASHC3* | rs10860846 | T | G | 0.2174 | -0.099 | 0.009 | 1E-27 | 121.00 |
| *WASHC3* | rs79948267 | G | T | 0.0246 | -0.245 | 0.024 | 2.5119E-24 | 104.21 |
| *WASHC3* | rs707234 | T | A | 0.3439 | -0.083 | 0.008 | 1E-26 | 107.64 |
| *WASL* | rs3087243 | G | A | 0.4519 | -0.064 | 0.008 | 1.5849E-16 | 64.00 |
| *WASL* | rs9274167 | T | G | 0.3888 | -0.102 | 0.008 | 1E-34 | 162.56 |
| *WFDC1* | rs68066031 | T | C | 0.2303 | -0.202 | 0.009 | 2.512E-110 | 503.75 |
| *WFDC1* | rs400345 | C | T | 0.1845 | -0.357 | 0.01 | 1E-200 | 1274.49 |
| *WFDC12* | rs61816761 | G | A | 0.0237 | 0.73 | 0.021 | 1E-200 | 1208.39 |
| *WFDC12* | rs11264533 | G | C | 0.3347 | 0.054 | 0.006 | 2.5119E-17 | 81.00 |
| *WFDC12* | rs17350445 | C | T | 0.0638 | -0.12 | 0.012 | 2.5119E-22 | 100.00 |
| *WFDC12* | rs9845881 | T | C | 0.5905 | 0.073 | 0.006 | 6.3096E-33 | 148.03 |
| *WFDC12* | rs10979132 | T | A | 0.1682 | -0.054 | 0.008 | 1.2589E-11 | 45.56 |
| *WFDC12* | rs12816349 | G | A | 0.2397 | -0.065 | 0.007 | 6.3096E-21 | 86.22 |
| *WFDC12* | rs10483947 | C | T | 0.0931 | 0.079 | 0.01 | 1.9953E-14 | 62.41 |
| *WFDC12* | rs8038032 | A | G | 0.3229 | -0.084 | 0.006 | 3.9811E-39 | 196.00 |
| *WFDC12* | rs35673728 | T | C | 0.063 | -0.177 | 0.012 | 1E-47 | 217.56 |
| *WFDC12* | rs562177063 | C | T | 0.0031 | -0.648 | 0.058 | 1.9953E-29 | 124.82 |
| *WFDC12* | rs6508781 | A | C | 0.1686 | -0.134 | 0.008 | 5.0119E-63 | 280.56 |
| *WFDC12* | rs75002084 | C | G | 0.431 | -0.425 | 0.007 | 1E-200 | 3686.22 |
| *WFDC12* | rs2283638 | C | A | 0.3266 | -0.061 | 0.006 | 5.0119E-22 | 103.36 |
| *WFDC2* | rs77924615 | G | A | 0.1968 | -0.074 | 0.009 | 1E-16 | 67.60 |
| *WFDC2* | rs28894750 | A | T | 0.534 | 0.062 | 0.007 | 1.9953E-18 | 78.45 |
| *WFDC2* | rs973446 | C | G | 0.0784 | -0.139 | 0.013 | 1.9953E-26 | 114.33 |
| *WFIKKN1* | rs72654647 | G | A | 0.2445 | 0.071 | 0.008 | 3.9811E-18 | 78.77 |
| *WFIKKN1* | rs11676298 | C | G | 0.1919 | -0.082 | 0.009 | 2.5119E-20 | 83.01 |
| *WFIKKN1* | rs79431137 | A | G | 0.2195 | 0.071 | 0.008 | 5.0119E-17 | 78.77 |
| *WFIKKN1* | rs34250475 | C | T | 0.4539 | 0.048 | 0.007 | 1.2589E-11 | 47.02 |
| *WFIKKN1* | rs27659 | A | G | 0.5513 | -0.048 | 0.007 | 1E-11 | 47.02 |
| *WFIKKN1* | rs2647074 | C | T | 0.3271 | 0.091 | 0.007 | 5.0119E-34 | 169.00 |
| *WFIKKN1* | rs3731211 | T | A | 0.7215 | -0.082 | 0.008 | 7.9433E-26 | 105.06 |
| *WFIKKN1* | rs183805019 | T | C | 0.0036 | -0.457 | 0.06 | 1.5849E-14 | 58.01 |
| *WFIKKN1* | rs4937333 | T | C | 0.5257 | 0.052 | 0.007 | 3.1623E-13 | 55.18 |
| *WFIKKN1* | rs7955466 | C | T | 0.3621 | 0.054 | 0.007 | 1.9953E-13 | 59.51 |
| *WFIKKN1* | rs11513729 | C | T | 0.4139 | 0.072 | 0.007 | 6.3096E-24 | 105.80 |
| *WFIKKN1* | rs55798945 | T | G | 0.3044 | 0.278 | 0.008 | 1E-200 | 1207.56 |
| *WFIKKN1* | rs34557412 | A | G | 0.0069 | 0.392 | 0.042 | 2.5119E-20 | 87.11 |
| *WFIKKN1* | rs1883932 | A | T | 0.5089 | 0.112 | 0.007 | 2.5119E-58 | 256.00 |
| *WFIKKN2* | rs6664906 | T | C | 0.4642 | -0.047 | 0.007 | 2.5119E-12 | 45.08 |
| *WFIKKN2* | rs4546329 | C | T | 0.4972 | -0.05 | 0.007 | 5.0119E-14 | 51.02 |
| *WFIKKN2* | rs4591185 | T | A | 0.6577 | -0.68 | 0.008 | 1E-200 | 7225.00 |
| *WFIKKN2* | rs33950747 | C | T | 0.0751 | 0.14 | 0.013 | 1E-28 | 115.98 |
| *WIF1* | rs1414660 | C | T | 0.1942 | 0.084 | 0.009 | 2.5119E-20 | 87.11 |
| *WIF1* | rs6768977 | A | G | 0.5335 | 0.053 | 0.007 | 1.5849E-13 | 57.33 |
| *WIF1* | rs17613549 | G | C | 0.2248 | -0.069 | 0.009 | 1.9953E-14 | 58.78 |
| *WIF1* | rs10104997 | C | T | 0.2097 | -0.102 | 0.009 | 5.0119E-31 | 128.44 |
| *WIF1* | rs10963680 | G | A | 0.2426 | 0.057 | 0.008 | 1.5849E-11 | 50.77 |
| *WIF1* | rs7896518 | A | G | 0.4239 | 0.061 | 0.007 | 7.9433E-17 | 75.94 |

| | | | | | | | | |
|---|---|---|---|---|---|---|---|---|
| *WIF1* | rs10741178 | T | C | 0.7779 | 0.062 | 0.009 | 1.2589E-12 | 47.46 |
| *WIF1* | rs7949566 | G | A | 0.4217 | 0.069 | 0.007 | 1.2589E-21 | 97.16 |
| *WIF1* | rs6489548 | A | G | 0.5989 | -0.19 | 0.007 | 3.981E-143 | 736.73 |
| *WIF1* | rs462010 | G | A | 0.4453 | -0.192 | 0.007 | 3.981E-150 | 752.33 |
| *WIF1* | rs12894709 | G | A | 0.4742 | -0.055 | 0.007 | 1.9953E-14 | 61.73 |
| *WIF1* | rs117068593 | C | T | 0.1872 | -0.078 | 0.009 | 3.9811E-17 | 75.11 |
| *WIF1* | rs186021206 | G | A | 0.0064 | 0.488 | 0.049 | 6.3096E-23 | 99.19 |
| *WIF1* | rs2145943 | A | G | 0.4075 | 0.098 | 0.007 | 1.2589E-40 | 196.00 |
| *WNT9A* | rs4653881 | C | T | 0.3775 | -0.129 | 0.007 | 6.3096E-72 | 339.61 |
| *WNT9A* | rs9274045 | T | C | 0.1673 | -0.069 | 0.009 | 1.5849E-13 | 58.78 |
| *WWP2* | rs1354034 | T | C | 0.6041 | 0.082 | 0.008 | 7.9433E-28 | 105.06 |
| *WWP2* | rs342293 | C | G | 0.459 | -0.058 | 0.007 | 6.3096E-15 | 68.65 |
| *WWP2* | rs72783174 | C | T | 0.0784 | -0.304 | 0.014 | 1.995E-107 | 471.51 |
| *XCL1* | rs4656599 | C | T | 0.1905 | 0.833 | 0.01 | 1E-200 | 6938.89 |
| *XCL1* | rs2009581 | G | A | 0.2728 | -0.148 | 0.007 | 3.1623E-94 | 447.02 |
| *XCL1* | rs13063578 | T | A | 0.3981 | -0.089 | 0.007 | 3.1623E-42 | 161.65 |
| *XCL1* | rs2548993 | G | A | 0.7224 | 0.067 | 0.007 | 1.2589E-20 | 91.61 |
| *XCL1* | rs9265813 | A | T | 0.1696 | 0.074 | 0.009 | 1.5849E-17 | 67.60 |
| *XCL1* | rs705379 | G | A | 0.4774 | 0.063 | 0.006 | 1E-22 | 110.25 |
| *XCL1* | rs8177651 | T | A | 0.3237 | 0.051 | 0.007 | 1.9953E-13 | 53.08 |
| *XCL1* | rs3184504 | T | C | 0.5166 | -0.102 | 0.006 | 3.9811E-58 | 289.00 |
| *XCL1* | rs77542162 | A | G | 0.0229 | 0.268 | 0.021 | 5.0119E-36 | 162.87 |
| *XCL1* | rs547843340 | G | GT | 0.1265 | -0.068 | 0.01 | 1.5849E-12 | 46.24 |
| *XCL1* | rs6031304 | T | C | 0.7259 | 0.049 | 0.007 | 7.9433E-12 | 49.00 |
| *XG* | rs11178645 | T | A | 0.4085 | -0.055 | 0.006 | 7.9433E-22 | 84.03 |
| *XIAP* | rs557657296 | C | CA | 0.1912 | 0.079 | 0.01 | 6.3096E-16 | 62.41 |
| *XIAP* | rs1354034 | T | C | 0.6039 | 0.08 | 0.008 | 1.5849E-25 | 100.00 |
| *XPNPEP2* | rs114165349 | G | C | 0.0233 | 0.42 | 0.025 | 7.9433E-65 | 282.24 |
| *XPNPEP2* | rs1260326 | T | C | 0.6087 | -0.07 | 0.008 | 2.5119E-20 | 76.56 |
| *XPNPEP2* | rs1584300 | A | C | 0.1493 | 0.072 | 0.01 | 3.1623E-12 | 51.84 |
| *XPNPEP2* | rs687339 | C | T | 0.7711 | 0.061 | 0.009 | 5.0119E-12 | 45.94 |
| *XPNPEP2* | rs2851270 | G | A | 0.6633 | -0.08 | 0.008 | 1.9953E-24 | 100.00 |
| *XPNPEP2* | rs10822143 | C | T | 0.5014 | -0.061 | 0.007 | 2.5119E-16 | 75.94 |
| *XPNPEP2* | rs11838867 | G | A | 0.0245 | -0.164 | 0.024 | 7.9433E-12 | 46.69 |
| *XPNPEP2* | rs11621792 | C | T | 0.4525 | 0.076 | 0.007 | 1.9953E-24 | 117.88 |
| *XPNPEP2* | rs2498786 | C | G | 0.6192 | -0.083 | 0.008 | 1.5849E-27 | 107.64 |
| *XPNPEP2* | rs339969 | C | A | 0.6164 | 0.094 | 0.008 | 6.3096E-35 | 138.06 |
| *XPNPEP2* | rs60018147 | A | G | 0.1193 | -0.092 | 0.012 | 1.5849E-14 | 58.78 |
| *XPNPEP2* | rs2207132 | G | A | 0.0327 | -0.219 | 0.021 | 1.2589E-25 | 108.76 |
| *XPNPEP2* | rs1800961 | C | T | 0.0321 | -0.227 | 0.021 | 1.9953E-27 | 116.85 |
| *XRCC4* | rs1805377 | G | A | 0.107 | 0.104 | 0.012 | 6.3096E-17 | 75.11 |
| *YAP1* | rs542212699 | G | A | 0.0088 | -0.622 | 0.037 | 3.9811E-63 | 282.60 |
| *YAP1* | rs26515 | C | T | 0.548 | 0.142 | 0.007 | 1.5849E-93 | 411.51 |
| *YAP1* | rs10791554 | G | A | 0.4531 | 0.062 | 0.007 | 1.5849E-19 | 78.45 |
| *YAP1* | rs77924615 | G | A | 0.1965 | -0.061 | 0.009 | 2.5119E-12 | 45.94 |
| *YARS1* | rs1354034 | T | C | 0.6046 | 0.061 | 0.008 | 1E-15 | 58.14 |
| *YARS1* | rs342298 | C | T | 0.455 | -0.059 | 0.008 | 6.3096E-15 | 54.39 |
| *YARS1* | rs6993770 | A | T | 0.2866 | -0.067 | 0.008 | 6.3096E-16 | 70.14 |
| *YES1* | rs1354034 | T | C | 0.6046 | 0.059 | 0.008 | 7.9433E-15 | 54.39 |
| *YES1* | rs2741189 | G | C | 0.2287 | 0.085 | 0.009 | 7.9433E-22 | 89.20 |
| *YOD1* | rs1044145 | T | C | 0.5372 | 0.106 | 0.007 | 1.5849E-46 | 229.31 |
| *YOD1* | rs3181215 | A | G | 0.2032 | -0.076 | 0.009 | 6.3096E-17 | 71.31 |
| *YTHDF3* | rs1354034 | T | C | 0.6035 | 0.086 | 0.008 | 6.3096E-29 | 115.56 |
| *YTHDF3* | rs11502185 | T | C | 0.2594 | 0.073 | 0.01 | 7.9433E-14 | 53.29 |
| *YWHAQ* | rs1354034 | T | C | 0.6044 | 0.067 | 0.008 | 1.9953E-18 | 70.14 |
| *YWHAQ* | rs7080386 | C | A | 0.412 | 0.06 | 0.008 | 5.0119E-15 | 56.25 |
| *ZBP1* | rs45521631 | C | T | 0.0267 | -0.909 | 0.024 | 1E-200 | 1434.52 |
| *ZBTB16* | rs1354034 | T | C | 0.6038 | 0.064 | 0.007 | 1.9953E-18 | 83.59 |
| *ZBTB16* | rs1265905 | A | G | 0.1296 | -0.077 | 0.011 | 6.3096E-13 | 49.00 |
| *ZBTB16* | rs342296 | G | A | 0.4566 | -0.059 | 0.007 | 6.3096E-16 | 71.04 |
| *ZBTB16* | rs6993770 | A | T | 0.2869 | -0.06 | 0.008 | 6.3096E-14 | 56.25 |
| *ZBTB16* | rs12767683 | C | A | 0.3249 | 0.079 | 0.008 | 1.9953E-24 | 97.52 |
| *ZBTB16* | rs73000929 | G | A | 0.0372 | 0.852 | 0.02 | 1E-200 | 1814.76 |
| *ZBTB16* | rs7297265 | A | G | 0.4426 | 0.08 | 0.007 | 1.5849E-27 | 130.61 |
| *ZBTB17* | rs7080536 | G | A | 0.0437 | -0.161 | 0.019 | 7.9433E-18 | 71.80 |
| *ZBTB17* | rs7260293 | C | T | 0.6207 | 0.073 | 0.008 | 1.2589E-20 | 83.27 |
| *ZFYVE19* | rs1354034 | T | C | 0.6046 | 0.056 | 0.007 | 7.9433E-14 | 64.00 |
| *ZFYVE19* | rs342298 | C | T | 0.455 | -0.064 | 0.007 | 5.0119E-18 | 83.59 |

| | | | | | | | | |
|---|---|---|---|---|---|---|---|---|
| *ZFYVE19* | rs7896518 | A | G | 0.4237 | 0.084 | 0.007 | 6.3096E-29 | 144.00 |
| *ZFYVE19* | rs11858010 | T | C | 0.3547 | -0.374 | 0.008 | 1E-200 | 2185.56 |
| *ZNF830* | rs617119 | C | T | 0.5133 | 0.06 | 0.008 | 2.5119E-15 | 56.25 |
| *ZNF830* | rs3018449 | T | C | 0.2644 | -0.06 | 0.009 | 1.9953E-12 | 44.44 |
| *ZNRD2* | rs3844257 | A | G | 0.2704 | -0.064 | 0.008 | 2.5119E-14 | 64.00 |
| *ZNRD2* | rs1354034 | T | C | 0.6046 | 0.077 | 0.008 | 3.1623E-24 | 92.64 |
| *ZNRD2* | rs35523137 | C | CA | 0.3598 | -0.053 | 0.008 | 5.0119E-12 | 43.89 |
| *ZNRD2* | rs204893 | T | C | 0.4312 | 0.089 | 0.007 | 1.9953E-32 | 161.65 |
| *ZNRD2* | rs342293 | C | G | 0.4598 | -0.062 | 0.007 | 3.9811E-17 | 78.45 |
| *ZNRD2* | rs35923643 | A | G | 0.1962 | -0.081 | 0.009 | 6.3096E-18 | 81.00 |
| *ZNRD2* | rs6497185 | C | A | 0.7263 | -0.072 | 0.008 | 5.0119E-18 | 81.00 |
| *ZNRF4* | rs10914144 | T | C | 0.8033 | 0.222 | 0.01 | 7.943E-119 | 492.84 |
| *ZNRF4* | rs10822145 | C | T | 0.472 | 0.056 | 0.008 | 2.5119E-13 | 49.00 |
| *ZP3* | rs17718444 | C | T | 0.3189 | 0.053 | 0.005 | 5.0119E-27 | 112.36 |
| *ZP3* | rs9275559 | A | C | 0.559 | -0.034 | 0.005 | 3.1623E-13 | 46.24 |
| *ZP3* | rs10281089 | C | T | 0.4305 | 1.105 | 0.008 | 1E-200 | 19078.52 |
| *ZP3* | rs1265831 | C | G | 0.2632 | -0.106 | 0.005 | 7.9433E-91 | 449.44 |
| *ZP3* | rs3747481 | C | T | 0.2582 | -0.036 | 0.005 | 1E-11 | 51.84 |
| *ZP3* | rs8073937 | G | A | 0.6524 | -0.051 | 0.005 | 7.9433E-26 | 104.04 |
| *ZP3* | rs4807125 | C | T | 0.1506 | -0.069 | 0.006 | 1E-26 | 132.25 |

**Supplementary Table 2: Exposure dataset in East Asian populations**

| Protein-encoding gene | SNP | Other allele | Effect allele | Effect allele frequency | Beta | SE | p-value | F-statistic |
|---|---|---|---|---|---|---|---|---|
| *ABO* | rs8176719 | T | TC | 0.3482 | 0.979 | 0.097 | 5.01E-24 | 101.86 |
| *ACP6* | rs4950465 | C | T | 0.6998 | 0.984 | 0.09 | 5.01E-28 | 119.54 |
| *AFAP1* | rs35617438 | C | T | 0.0579 | 1.196 | 0.157 | 2E-14 | 58.03 |
| *AHSG* | rs6444149 | G | A | 0.134 | -0.389 | 0.042 | 9.68E-21 | 85.78 |
| *AHSG* | rs2077119 | G | T | 0.376 | -0.232 | 0.031 | 6.75E-14 | 56.01 |
| *AHSG* | rs116975397 | G | A | 0.054 | -0.347 | 0.063 | 3.09E-08 | 30.34 |
| *AHSG* | rs2070632 | C | A | 0.098 | -0.382 | 0.049 | 7.2E-15 | 60.78 |
| *ALPP* | rs12478529 | C | T | 0.4674 | 0.75 | 0.098 | 1.58E-14 | 58.57 |
| *APOA1* | rs5081 | A | T | 0.073 | -0.494 | 0.055 | 1.99E-19 | 80.67 |
| *APOA1* | rs5132 | C | T | 0.074 | -0.682 | 0.055 | 2.59E-35 | 153.76 |
| *APOA2* | rs59612622 | C | T | 0.375 | 0.175 | 0.026 | 3.04E-11 | 45.30 |
| *APOA4* | rs117341026 | G | A | 0.082 | 0.283 | 0.049 | 9.13E-09 | 33.36 |
| *APOA4* | rs7350481 | C | T | 0.25 | -0.336 | 0.03 | 1.57E-28 | 125.44 |
| *APOA4* | rs645901 | C | T | 0.425 | -0.538 | 0.027 | 2.46E-86 | 397.04 |
| *APOA4* | rs2513094 | T | C | 0.255 | 0.296 | 0.031 | 1.19E-21 | 91.17 |
| *APOA4* | rs1242127 | G | A | 0.402 | 0.305 | 0.051 | 2.84E-09 | 35.77 |
| *APOA4* | rs2510446 | T | C | 0.238 | 0.191 | 0.032 | 1.46E-09 | 35.63 |
| *APOB* | rs72654473 | C | A | 0.09 | -0.345 | 0.048 | 9.73E-13 | 51.66 |
| *APOB* | rs7412 | C | T | 0.087 | -0.328 | 0.048 | 9.42E-12 | 46.69 |
| *APOB* | rs190712692 | G | A | 0.086 | -0.365 | 0.05 | 2.27E-13 | 53.29 |
| *APOB* | rs769446 | T | C | 0.092 | -0.354 | 0.047 | 8.37E-14 | 56.73 |
| *APOB* | rs1469513 | T | C | 0.127 | -0.22 | 0.036 | 8.51E-10 | 37.35 |
| *APOB* | rs520354 | G | A | 0.159 | -0.333 | 0.039 | 7.3E-18 | 72.91 |
| *APOC1* | rs5112 | G | C | 0.352 | -0.169 | 0.03 | 1.87E-08 | 31.73 |
| *APOC2* | rs405697 | A | G | 0.445 | 0.196 | 0.029 | 1.32E-11 | 45.68 |
| *APOC2* | rs5112 | G | C | 0.352 | -0.201 | 0.03 | 2.39E-11 | 44.89 |
| *APOC4* | rs111809714 | T | C | 0.493 | 0.145 | 0.022 | 4.84E-11 | 43.44 |
| *APOC4* | rs5167 | G | T | 0.485 | -0.103 | 0.019 | 2.93E-08 | 29.39 |
| *APOE* | rs769446 | T | C | 0.092 | 0.613 | 0.048 | 5.94E-37 | 163.09 |
| *APOE* | rs1065853 | G | T | 0.087 | 0.54 | 0.049 | 2.21E-28 | 121.45 |
| *APOE* | rs5112 | G | C | 0.352 | -0.168 | 0.03 | 2.06E-08 | 31.36 |
| *APOE* | rs405509 | T | G | 0.347 | 0.181 | 0.029 | 6.64E-10 | 38.95 |
| *APOE* | rs61679753 | T | A | 0.092 | 0.554 | 0.048 | 5.04E-31 | 133.21 |
| *APOE* | rs66626994 | G | A | 0.098 | -0.658 | 0.065 | 5.93E-24 | 102.48 |
| *APOE* | rs7254892 | G | A | 0.077 | 0.496 | 0.053 | 1.46E-20 | 87.58 |
| *APOE* | rs7412 | C | T | 0.087 | 0.579 | 0.049 | 6.69E-32 | 139.63 |
| *APOH* | rs11651658 | T | C | 0.058 | -0.848 | 0.094 | 1.51E-19 | 81.38 |
| *APOM* | rs2736182 | G | A | 0.198 | -0.348 | 0.04 | 2.44E-18 | 75.69 |
| *APOM* | rs6913309 | T | A | 0.123 | 0.243 | 0.043 | 1.47E-08 | 31.94 |
| *APOM* | rs805258 | C | T | 0.198 | -0.366 | 0.037 | 4.58E-23 | 97.85 |
| *ARSA* | rs28871051 | T | G | 0.1437 | 1.573 | 0.155 | 3.98E-24 | 102.99 |
| *ASAH2* | rs10508921 | C | T | 0.3225 | -1.03 | 0.109 | 3.98E-21 | 89.29 |
| *BCL2* | rs1800477 | C | T | 0.0949 | -1.751 | 0.209 | 5.01E-17 | 70.19 |
| *BST1* | rs4263397 | T | G | 0.5655 | -0.669 | 0.084 | 1.58E-15 | 63.43 |
| *C4A* | rs6457302 | T | C | 0.472 | -0.168 | 0.029 | 4.11E-09 | 33.56 |
| *C4A* | rs2507978 | G | A | 0.383 | -0.235 | 0.03 | 2.52E-15 | 61.36 |
| *C4A* | rs2736163 | C | T | 0.197 | 0.32 | 0.045 | 8.51E-13 | 50.57 |
| *C4A* | rs187764 | C | T | 0.067 | -0.371 | 0.058 | 1.23E-10 | 40.92 |
| *C4A* | rs2244027 | A | G | 0.428 | 0.191 | 0.029 | 6.83E-11 | 43.38 |
| *C4A* | rs707938 | A | G | 0.363 | 0.262 | 0.032 | 2.16E-16 | 67.04 |
| *C4A* | rs9268541 | T | C | 0.154 | -0.245 | 0.04 | 9.11E-10 | 37.52 |
| *C4A* | rs443198 | G | A | 0.432 | -0.199 | 0.027 | 2.87E-13 | 54.32 |
| *C4A* | rs511027 | A | T | 0.089 | -0.282 | 0.047 | 2.26E-09 | 36.00 |
| *C4B* | rs2523559 | A | C | 0.137 | 0.236 | 0.041 | 9.83E-09 | 33.13 |
| *C4B* | rs389883 | T | G | 0.303 | -0.211 | 0.031 | 1.51E-11 | 46.33 |
| *C4B* | rs9268909 | T | C | 0.14 | -0.234 | 0.041 | 1.37E-08 | 32.57 |
| *C4B* | rs1077394 | T | C | 0.327 | 0.223 | 0.031 | 5.62E-13 | 51.75 |
| *C4B* | rs2072633 | A | G | 0.442 | -0.174 | 0.028 | 9.27E-10 | 38.62 |
| *C4B* | rs9268541 | T | C | 0.154 | -0.231 | 0.04 | 5.53E-09 | 33.35 |
| *C4B* | rs3778640 | G | C | 0.146 | 0.23 | 0.037 | 4.87E-10 | 38.64 |
| *C4B* | rs2853961 | G | A | 0.161 | 0.293 | 0.036 | 2.98E-16 | 66.24 |
| *C4B* | rs1051488 | T | C | 0.417 | 0.3 | 0.027 | 7.23E-29 | 123.46 |
| *C4B* | rs28752476 | T | C | 0.184 | 0.206 | 0.034 | 9.35E-10 | 36.71 |
| *C4B* | rs2230365 | C | T | 0.185 | 0.242 | 0.034 | 1.14E-12 | 50.66 |

| | | | | | | | | |
|---|---|---|---|---|---|---|---|---|
| *C4B* | rs494620 | G | A | 0.363 | 0.444 | 0.027 | 6.04E-59 | 270.42 |
| *C4B* | rs139347611 | G | A | 0.094 | 0.348 | 0.045 | 1.68E-14 | 59.80 |
| *C4B* | rs2021783 | C | T | 0.226 | -0.526 | 0.032 | 3.05E-61 | 270.19 |
| *C4B* | rs115461385 | C | T | 0.087 | 0.522 | 0.045 | 1.15E-30 | 134.56 |
| *C4B* | rs9469084 | C | T | 0.083 | 0.619 | 0.048 | 1.48E-37 | 166.30 |
| *C4B* | rs377763 | C | A | 0.151 | 0.461 | 0.038 | 1.21E-34 | 147.18 |
| *C4B* | rs2858308 | G | T | 0.151 | 0.396 | 0.037 | 2.38E-27 | 114.55 |
| *C4B* | rs241437 | G | A | 0.458 | -0.153 | 0.026 | 6.99E-09 | 34.63 |
| *C4B* | rs17220479 | C | T | 0.063 | -0.324 | 0.054 | 2.09E-09 | 36.00 |
| *C4BPA* | rs28368251 | T | C | 0.117 | 0.104 | 0.018 | 4.4E-09 | 33.38 |
| *C4BPA* | rs2808470 | C | T | 0.177 | 0.221 | 0.037 | 2.73E-09 | 35.68 |
| *C4BPA* | rs2808468 | A | G | 0.175 | 0.291 | 0.037 | 2.09E-15 | 61.86 |
| *C4BPA* | rs77660718 | A | T | 0.177 | -0.438 | 0.037 | 1.17E-32 | 140.13 |
| *C7* | rs2034492 | A | G | 0.443 | -0.19 | 0.029 | 8.11E-11 | 42.93 |
| *C7* | rs9687064 | G | T | 0.34 | 0.141 | 0.023 | 9.62E-10 | 37.58 |
| *C7* | rs75467805 | G | C | 0.127 | -0.187 | 0.033 | 1.81E-08 | 32.11 |
| *C7* | rs367660347 | A | G | 0.224 | 0.17 | 0.026 | 1.48E-10 | 42.75 |
| *C7* | rs2034491 | C | G | 0.101 | 0.395 | 0.039 | 3.11E-24 | 102.58 |
| *C7* | rs74837349 | T | C | 0.086 | 0.28 | 0.05 | 2.09E-08 | 31.36 |
| *C7* | rs1450657 | G | T | 0.27 | -0.211 | 0.032 | 3.03E-11 | 43.48 |
| *C7* | rs1349371 | G | T | 0.419 | 0.166 | 0.029 | 1.08E-08 | 32.77 |
| *C7* | rs62358580 | T | C | 0.163 | -0.256 | 0.039 | 3.3E-11 | 43.09 |
| *C7* | rs79241415 | A | C | 0.095 | 0.307 | 0.05 | 1.11E-09 | 37.70 |
| *C7* | rs2329433 | C | T | 0.478 | 0.215 | 0.029 | 1.3E-13 | 54.96 |
| *C7* | rs445118 | A | G | 0.46 | -0.167 | 0.029 | 8.2E-09 | 33.16 |
| *C8B* | rs684216 | G | C | 0.445 | -0.16 | 0.029 | 2.98E-08 | 30.44 |
| *C8B* | rs684844 | G | A | 0.445 | -0.182 | 0.028 | 8.61E-11 | 42.25 |
| *C8B* | rs3768214 | G | C | 0.384 | -0.189 | 0.028 | 2.89E-11 | 45.56 |
| *C8B* | rs713452 | C | T | 0.462 | -0.157 | 0.028 | 2.92E-08 | 31.44 |
| *C8G* | rs7868669 | A | G | 0.164 | 0.303 | 0.039 | 4.74E-15 | 60.36 |
| *C8G* | rs4880178 | A | T | 0.157 | 0.266 | 0.039 | 9.09E-12 | 46.52 |
| *C8G* | rs885070 | G | A | 0.164 | 0.358 | 0.056 | 2.33E-10 | 40.87 |
| *C8G* | rs10870152 | A | T | 0.183 | 0.222 | 0.037 | 2.12E-09 | 36.00 |
| *C8G* | rs908837 | A | G | 0.144 | 0.24 | 0.042 | 1.11E-08 | 32.65 |
| *C9orf131* | rs2075799 | C | T | 0.235 | 0.268 | 0.033 | 3.52E-16 | 65.95 |
| *C9orf131* | rs77633711 | C | T | 0.057 | -0.47 | 0.058 | 5.91E-16 | 65.67 |
| *CA6* | rs3765963 | A | G | 0.7358 | 0.84 | 0.101 | 7.94E-17 | 69.17 |
| *CAPG* | rs6419687 | A | G | 0.5077 | -0.699 | 0.077 | 1E-19 | 82.41 |
| *CCL15* | rs41508645 | T | G | 0.2878 | 1.486 | 0.131 | 7.94E-30 | 128.68 |
| *CCL8* | rs11654149 | T | C | 0.2946 | -1.106 | 0.111 | 1.58E-23 | 99.28 |
| *CD209* | rs505922 | T | C | 0.3459 | 0.91 | 0.117 | 7.94E-15 | 60.49 |
| *CD276* | rs10083681 | G | A | 0.1067 | -1.028 | 0.137 | 6.31E-14 | 56.30 |
| *CD300C* | rs7217136 | C | T | 0.3289 | 0.809 | 0.107 | 3.16E-14 | 57.16 |
| *CD300E* | rs2655606 | A | G | 0.5603 | -0.695 | 0.098 | 1E-12 | 50.29 |
| *CD300LF* | rs35489971 | A | G | 0.7269 | 1.059 | 0.088 | 3.98E-33 | 144.82 |
| *CD70* | rs7254538 | G | C | 0.3294 | 0.545 | 0.077 | 1.58E-12 | 50.10 |
| *CDH15* | rs190099280 | G | C | 0.1314 | 1.538 | 0.171 | 3.16E-19 | 80.89 |
| *CDH5* | rs8176730 | T | C | 0.1953 | 1.239 | 0.137 | 2E-19 | 81.79 |
| *CEACAM21* | rs723902 | G | C | 0.9056 | -1.222 | 0.145 | 3.98E-17 | 71.02 |
| *CFB* | rs444921 | C | T | 0.073 | 0.413 | 0.056 | 1.05E-13 | 54.39 |
| *CFB* | rs429608 | G | A | 0.07 | -0.805 | 0.061 | 5.7E-40 | 174.15 |
| *CFB* | rs8283 | A | G | 0.336 | -0.181 | 0.03 | 1.64E-09 | 36.40 |
| *CFB* | rs3129949 | C | A | 0.09 | -0.302 | 0.052 | 4.58E-09 | 33.73 |
| *CFB* | rs2071551 | T | C | 0.097 | -0.32 | 0.048 | 3.32E-11 | 44.44 |
| *CFH* | rs116877015 | A | G | 0.1 | -0.247 | 0.036 | 5.98E-12 | 47.07 |
| *CFH* | rs146340148 | C | T | 0.434 | 0.255 | 0.023 | 5.67E-29 | 122.92 |
| *CFH* | rs77747198 | T | C | 0.055 | 0.288 | 0.047 | 6.56E-10 | 37.55 |
| *CFH* | rs1009188 | T | C | 0.143 | -0.17 | 0.031 | 2.83E-08 | 30.07 |
| *CFH* | rs140300240 | A | G | 0.083 | -0.38 | 0.049 | 1.16E-14 | 60.14 |
| *CFH* | rs6693594 | A | C | 0.455 | -0.732 | 0.039 | 1.86E-78 | 352.28 |
| *CFH* | rs149859066 | T | C | 0.064 | -0.501 | 0.055 | 6.92E-20 | 82.98 |
| *CFH* | rs190577253 | G | A | 0.056 | -0.462 | 0.059 | 3.12E-15 | 61.32 |
| *CFH* | rs117485108 | G | A | 0.098 | 0.331 | 0.047 | 1.17E-12 | 49.60 |
| *CFH* | rs10754220 | G | A | 0.15 | 0.216 | 0.039 | 2.98E-08 | 30.67 |
| *CFH* | rs10733086 | T | A | 0.051 | -0.472 | 0.065 | 4.99E-13 | 52.73 |
| *CFHR1* | rs60642321 | A | G | 0.053 | -0.968 | 0.063 | 6.16E-54 | 236.09 |
| *CFHR1* | rs10733086 | T | A | 0.051 | 0.367 | 0.064 | 8.78E-09 | 32.88 |
| *CFHR1* | rs380733 | A | G | 0.059 | 0.335 | 0.048 | 5.03E-12 | 48.71 |

| | | | | | | | | |
|---|---|---|---|---|---|---|---|---|
| *CFHR1* | rs12240143 | T | C | 0.182 | -0.36 | 0.048 | 8.4E-14 | 56.25 |
| *CFHR1* | rs117485108 | G | A | 0.098 | 0.394 | 0.038 | 1.15E-24 | 107.50 |
| *CFHR1* | rs7542235 | A | G | 0.055 | -0.845 | 0.085 | 1.68E-23 | 98.83 |
| *CFHR2* | rs12029494 | T | G | 0.427 | -0.152 | 0.027 | 1.31E-08 | 31.69 |
| *CFHR2* | rs78531813 | C | T | 0.146 | 0.743 | 0.054 | 8.29E-44 | 189.32 |
| *CFHR2* | rs6428379 | C | T | 0.083 | 0.746 | 0.048 | 6.6E-55 | 241.54 |
| *CFHR2* | rs1923759 | G | A | 0.321 | 0.161 | 0.028 | 1.37E-08 | 33.06 |
| *CFHR2* | rs199615054 | C | T | 0.379 | -0.207 | 0.037 | 1.98E-08 | 31.30 |
| *CFHR2* | rs1359594 | G | T | 0.084 | 0.722 | 0.045 | 1.36E-58 | 257.42 |
| *CFHR2* | rs78684793 | A | G | 0.053 | 0.484 | 0.07 | 3.44E-12 | 47.81 |
| *CFHR2* | rs7526780 | A | T | 0.067 | 0.368 | 0.049 | 5.31E-14 | 56.40 |
| *CFHR2* | rs10922215 | G | A | 0.22 | 0.165 | 0.03 | 3.5E-08 | 30.25 |
| *CFHR2* | rs12029246 | C | T | 0.384 | -0.146 | 0.026 | 2.99E-08 | 31.53 |
| *CFHR2* | rs6667243 | C | T | 0.088 | 0.755 | 0.049 | 9.84E-54 | 237.41 |
| *CFHR2* | rs113605888 | G | A | 0.218 | 0.206 | 0.035 | 5.78E-09 | 34.64 |
| *CFHR2* | rs3790414 | T | A | 0.7157 | -0.868 | 0.081 | 5.01E-27 | 114.83 |
| *CFHR4* | rs6667243 | T | C | 0.8738 | -0.926 | 0.1 | 2E-20 | 85.75 |
| *CFI* | rs116867327 | C | A | 0.077 | -0.478 | 0.056 | 1.17E-17 | 72.86 |
| *CFI* | rs74817407 | C | T | 0.132 | 0.329 | 0.042 | 8.33E-15 | 61.36 |
| *CFI* | rs5030556 | T | C | 0.086 | -0.282 | 0.051 | 4.53E-08 | 30.57 |
| *CFI* | rs13104777 | T | C | 0.171 | 0.289 | 0.038 | 3.96E-14 | 57.84 |
| *CFI* | rs78298715 | C | G | 0.241 | 0.214 | 0.034 | 2.27E-10 | 39.62 |
| *CHIT1* | rs150192398 | C | CAGACCAT( | 0.5405 | -0.954 | 0.096 | 2.51E-23 | 98.75 |
| *CLEC4C* | rs7302014 | T | C | 0.3179 | 0.763 | 0.1 | 2.51E-14 | 58.22 |
| *CLEC4M* | rs505922 | T | C | 0.3516 | 0.842 | 0.116 | 3.16E-13 | 52.69 |
| *CLGN* | rs56041599 | G | A | 0.2106 | 1.061 | 0.122 | 3.98E-18 | 75.63 |
| *CNTN2* | rs79852451 | G | A | 0.0596 | 1.444 | 0.166 | 3.98E-18 | 75.67 |
| *COL28A1* | rs7810134 | G | A | 0.2379 | 1.031 | 0.127 | 6.31E-16 | 65.90 |
| *CPB2* | rs7318648 | T | C | 0.234 | 0.305 | 0.042 | 2.74E-13 | 52.74 |
| *CPB2* | rs34813434 | C | G | 0.148 | 0.244 | 0.04 | 1.05E-09 | 37.21 |
| *CPB2* | rs9534305 | G | A | 0.236 | 0.27 | 0.034 | 2.32E-15 | 63.06 |
| *CPB2* | rs1581846 | A | G | 0.209 | -0.199 | 0.034 | 2.82E-09 | 34.26 |
| *CPB2* | rs1326398 | G | T | 0.147 | 0.229 | 0.039 | 3.22E-09 | 34.48 |
| *CPN1* | rs12267374 | G | A | 0.227 | -0.211 | 0.034 | 3.57E-10 | 38.51 |
| *CPN1* | rs2862922 | A | T | 0.217 | -0.199 | 0.035 | 8.76E-09 | 32.33 |
| *CPN2* | rs11711157 | C | T | 0.306 | -0.225 | 0.031 | 5.01E-13 | 52.68 |
| *CPN2* | rs11080055 | C | A | 0.273 | 0.216 | 0.03 | 1.15E-12 | 51.84 |
| *CPPED1* | rs8050485 | A | G | 0.4164 | 0.752 | 0.092 | 2.51E-16 | 66.81 |
| *CRISP2* | rs555247 | G | A | 0.5148 | 0.603 | 0.082 | 2E-13 | 54.08 |
| *CSF2RB* | rs5756415 | C | G | 0.7796 | -1.18 | 0.114 | 5.01E-25 | 107.14 |
| *CST5* | rs4239744 | A | G | 0.503 | 0.792 | 0.093 | 1.26E-17 | 72.52 |
| *CSTB* | rs35285321 | G | A | 0.6325 | -0.878 | 0.093 | 5.01E-21 | 89.13 |
| *CTSH* | rs28699989 | T | C | 0.7735 | 1.05 | 0.118 | 5.01E-19 | 79.18 |
| *DBH* | rs1611115 | T | C | 0.7945 | 1.106 | 0.104 | 2E-26 | 113.10 |
| *DKKL1* | rs12983800 | G | A | 0.3488 | -1.228 | 0.101 | 7.94E-34 | 147.83 |
| *DNAJC25* | rs2507978 | G | A | 0.383 | -0.217 | 0.029 | 8.29E-14 | 55.99 |
| *DNAJC25* | rs77633711 | C | T | 0.057 | -0.465 | 0.06 | 1.33E-14 | 60.06 |
| *DPEP1* | rs1126464 | G | C | 0.3642 | -1.097 | 0.109 | 1.26E-23 | 101.29 |
| *DYNLT1* | rs4925749 | G | A | 0.1268 | 1.045 | 0.144 | 5.01E-13 | 52.66 |
| *ECM1* | rs1694385 | G | A | 0.212 | -0.354 | 0.032 | 1.3E-27 | 122.38 |
| *ECM1* | rs77257036 | C | T | 0.25 | -0.32 | 0.031 | 1.85E-25 | 106.56 |
| *ENPP5* | rs16874322 | T | A | 0.2365 | 0.719 | 0.098 | 1.58E-13 | 53.83 |
| *ENPP7* | rs11871061 | T | C | 0.5119 | 0.956 | 0.096 | 2.51E-23 | 99.17 |
| *ERP44* | rs78937374 | C | G | 0.2301 | 0.836 | 0.124 | 1.58E-11 | 45.45 |
| *F12* | rs72648808 | G | A | 0.248 | 0.245 | 0.034 | 4.62E-13 | 51.92 |
| *F12* | rs10866705 | A | C | 0.264 | 0.404 | 0.033 | 3.09E-34 | 149.88 |
| *F12* | rs12517919 | A | G | 0.197 | -0.251 | 0.035 | 1.24E-12 | 51.43 |
| *F12* | rs179809 | T | G | 0.191 | 0.233 | 0.036 | 1.87E-10 | 41.89 |
| *F12* | rs4976685 | A | G | 0.152 | 0.208 | 0.03 | 9.67E-12 | 48.07 |
| *F12* | rs1801020 | A | G | 0.194 | 0.79 | 0.053 | 3.55E-50 | 222.18 |
| *F12* | rs6556314 | C | T | 0.176 | 0.621 | 0.037 | 5.98E-63 | 281.70 |
| *F12* | rs200028730 | G | C | 0.317 | 0.195 | 0.031 | 3.54E-10 | 39.57 |
| *F12* | rs2545801 | T | C | 0.2422 | 1.11 | 0.098 | 6.31E-30 | 128.29 |
| *F13B* | rs4915363 | C | T | 0.252 | 0.187 | 0.033 | 1.04E-08 | 32.11 |
| *F2* | rs78579936 | T | C | 0.165 | -0.341 | 0.035 | 1.82E-22 | 94.92 |
| *F2* | rs10769199 | T | G | 0.195 | 0.243 | 0.033 | 1.85E-13 | 54.22 |
| *F2* | rs11038860 | C | T | 0.244 | 0.703 | 0.03 | 5.03E-122 | 549.12 |
| *F2* | rs80015663 | T | C | 0.082 | -0.457 | 0.056 | 1.91E-16 | 66.60 |

| | | | | | | | | |
|---|---|---|---|---|---|---|---|---|
| *F2* | rs12417149 | G | A | 0.092 | -0.443 | 0.052 | 1.38E-17 | 72.58 |
| *F2* | rs2596397 | G | A | 0.279 | 0.648 | 0.03 | 5.18E-101 | 466.56 |
| *F2* | rs7129948 | C | A | 0.048 | 0.327 | 0.06 | 4.11E-08 | 29.70 |
| *F5* | rs3753305 | G | C | 0.127 | 0.234 | 0.042 | 3.48E-08 | 31.04 |
| *FAM3D* | rs9411378 | C | A | 0.1318 | 0.96 | 0.141 | 1E-11 | 46.36 |
| *FAP* | rs10165813 | C | T | 0.126 | 1.16 | 0.169 | 6.31E-12 | 47.11 |
| *FCAR* | rs56216868 | C | T | 0.2854 | 0.985 | 0.116 | 2E-17 | 72.10 |
| *FCGR3B* | rs200688856 | G | C | 0.6056 | -0.782 | 0.1 | 3.98E-15 | 61.15 |
| *FCN3* | rs3813800 | G | C | 0.179 | -0.699 | 0.053 | 2.57E-40 | 173.94 |
| *FCN3* | rs181353661 | T | A | 0.16 | -0.385 | 0.052 | 8.42E-14 | 54.82 |
| *FCN3* | rs3813795 | A | G | 0.25 | 0.184 | 0.033 | 2.8E-08 | 31.09 |
| *FCN3* | rs58337722 | G | T | 0.184 | -0.171 | 0.024 | 5.17E-13 | 50.77 |
| *FCRL3* | rs7522061 | T | C | 0.3952 | 0.996 | 0.106 | 6.31E-21 | 88.29 |
| *FGF5* | rs10857147 | A | T | 0.4184 | 0.679 | 0.082 | 7.94E-17 | 68.57 |
| *FGFR4* | rs351855 | G | A | 0.4567 | 1.136 | 0.104 | 1E-27 | 119.31 |
| *FLT4* | rs10935473 | G | T | 0.4901 | -0.711 | 0.102 | 3.16E-12 | 48.59 |
| *FMN2* | rs117095836 | G | A | 0.054 | -0.627 | 0.061 | 1.03E-24 | 105.65 |
| *FOLR3* | rs139130389 | C | CTA | 0.0516 | 1.567 | 0.195 | 1E-15 | 64.58 |
| *FUT3_FUT5* | rs10409772 | C | A | 0.2756 | -0.934 | 0.109 | 7.94E-18 | 73.42 |
| *FUT8* | rs2127870 | G | C | 0.8035 | -0.865 | 0.112 | 1E-14 | 59.65 |
| *GBP4* | rs674903 | A | G | 0.4515 | -0.626 | 0.091 | 5.01E-12 | 47.32 |
| *GC* | rs6822876 | A | G | 0.441 | -0.346 | 0.027 | 3.36E-37 | 164.22 |
| *GC* | rs6845925 | T | A | 0.241 | 0.778 | 0.032 | 1.6E-132 | 591.10 |
| *GC* | rs222040 | G | A | 0.365 | -0.344 | 0.028 | 4.98E-34 | 150.94 |
| *GC* | rs66811958 | G | A | 0.389 | 0.385 | 0.028 | 2.46E-42 | 189.06 |
| *GC* | rs67325168 | A | C | 0.239 | 0.217 | 0.032 | 1.82E-11 | 45.99 |
| *GC* | rs9790741 | T | G | 0.297 | -0.162 | 0.03 | 4.79E-08 | 29.16 |
| *GC* | rs2365698 | C | T | 0.113 | 0.267 | 0.044 | 1.1E-09 | 36.82 |
| *GC* | rs4588 | G | T | 0.2442 | 1.235 | 0.113 | 7.94E-28 | 119.45 |
| *GCK* | rs2244027 | A | G | 0.426 | 0.469 | 0.026 | 1.5E-72 | 325.39 |
| *GGACT* | rs3825501 | C | T | 0.1504 | 1.249 | 0.149 | 5.01E-17 | 70.27 |
| *GGH* | rs750672688 | CATA | C | 0.3411 | 0.757 | 0.105 | 6.31E-13 | 51.98 |
| *GP2* | rs8176725 | G | A | 0.2437 | 0.801 | 0.109 | 1.58E-13 | 54.00 |
| *GPC5* | rs342706 | T | C | 0.2405 | -0.825 | 0.107 | 1E-14 | 59.45 |
| *GPR37* | rs11772423 | G | A | 0.306 | -0.817 | 0.103 | 1.58E-15 | 62.92 |
| *GSTT2B* | rs5751777 | C | T | 0.4961 | -0.916 | 0.089 | 7.94E-25 | 105.93 |
| *HBZ* | rs11431189 | C | CA | 0.6673 | -1.085 | 0.118 | 3.98E-20 | 84.55 |
| *HDDC2* | rs987166 | A | G | 0.1534 | -0.922 | 0.131 | 2E-12 | 49.54 |
| *HDGF* | rs4501833 | G | A | 0.1128 | 1.153 | 0.119 | 5.01E-22 | 93.88 |
| *HLA-A* | rs1611627 | T | C | 0.4241 | 0.682 | 0.086 | 2E-15 | 62.89 |
| *HP* | rs8053498 | T | C | 0.269 | -0.234 | 0.033 | 8.94E-13 | 50.28 |
| *HP* | rs11648557 | C | T | 0.05 | -0.757 | 0.066 | 3.85E-30 | 131.55 |
| *HP* | rs77303550 | C | T | 0.297 | 0.407 | 0.032 | 1.5E-37 | 161.77 |
| *HP* | rs16973266 | G | A | 0.217 | -0.205 | 0.035 | 8.21E-09 | 34.31 |
| *HP* | rs62053845 | G | A | 0.319 | 0.265 | 0.034 | 6.67E-15 | 60.75 |
| *HP* | rs12918905 | T | C | 0.234 | -0.35 | 0.035 | 4.1E-24 | 100.00 |
| *HP* | rs35484594 | A | C | 0.302 | 0.202 | 0.032 | 4.39E-10 | 39.85 |
| *HP* | rs75357168 | A | G | 0.256 | -0.194 | 0.033 | 5.56E-09 | 34.56 |
| *HP* | rs34042070 | C | G | 0.255 | -0.389 | 0.032 | 1.36E-33 | 147.77 |
| *HP* | rs116894978 | G | A | 0.368 | 0.169 | 0.03 | 1.52E-08 | 31.73 |
| *HP* | rs9930543 | A | G | 0.26 | -0.217 | 0.033 | 6.33E-11 | 43.24 |
| *HP* | rs28731397 | T | C | 0.234 | -0.366 | 0.046 | 1.66E-15 | 63.31 |
| *HRG* | rs41268617 | G | T | 0.476 | -0.398 | 0.029 | 1.49E-43 | 188.35 |
| *HRG* | rs9879899 | C | T | 0.291 | -0.202 | 0.032 | 2.12E-10 | 39.85 |
| *HRG* | rs6775048 | G | C | 0.125 | 0.272 | 0.044 | 5.84E-10 | 38.21 |
| *HRG* | rs1042464 | T | A | 0.187 | -0.375 | 0.037 | 9.2E-24 | 102.72 |
| *HRG* | rs55636108 | T | C | 0.454 | 0.182 | 0.031 | 4.97E-09 | 34.47 |
| *HRG* | rs60531302 | G | A | 0.189 | -0.301 | 0.052 | 5.03E-09 | 33.51 |
| *HRG* | rs114232169 | T | G | 0.407 | -0.437 | 0.03 | 1.04E-46 | 212.19 |
| *HRG* | rs3733159 | G | T | 0.474 | -0.316 | 0.033 | 1.18E-21 | 91.70 |
| *HRG* | rs28390219 | T | C | 0.203 | -0.236 | 0.035 | 2.75E-11 | 45.47 |
| *ICAM2* | rs10935473 | G | T | 0.4785 | -0.715 | 0.089 | 1.26E-15 | 64.54 |
| *ICAM5* | rs281439 | G | C | 0.4812 | 0.872 | 0.091 | 1E-21 | 91.82 |
| *IFNGR2* | rs9808753 | A | G | 0.4199 | 1.348 | 0.111 | 3.98E-34 | 147.48 |
| *IGHA1* | rs1407 | C | G | 0.228 | -0.404 | 0.033 | 1.1E-33 | 149.88 |
| *IGHG2* | rs1407 | C | G | 0.228 | -0.258 | 0.033 | 4.11E-15 | 61.12 |
| *IGHG3* | rs1407 | C | G | 0.227 | -0.527 | 0.034 | 1.58E-55 | 240.25 |
| *IGHG3* | rs115161557 | A | G | 0.24 | -0.437 | 0.033 | 1.48E-40 | 175.36 |

| | | | | | | | | |
|---|---|---|---|---|---|---|---|---|
| *IGHG3* | rs1134590 | T | C | 0.207 | -0.489 | 0.037 | 4.19E-39 | 174.67 |
| *IGHG4* | rs11574630 | T | C | 0.356 | 0.217 | 0.029 | 1.03E-13 | 55.99 |
| *IGHM* | rs2516994 | A | G | 0.255 | -0.307 | 0.035 | 1.31E-18 | 76.94 |
| *IGHM* | rs12434905 | T | A | 0.321 | -0.156 | 0.028 | 3.08E-08 | 31.04 |
| *IGHM* | rs192568966 | G | A | 0.105 | -0.335 | 0.043 | 7.36E-15 | 60.69 |
| *IGHM* | rs12433228 | G | A | 0.219 | 0.457 | 0.032 | 2.06E-46 | 203.95 |
| *IGHM* | rs2753538 | G | C | 0.114 | -0.418 | 0.042 | 1.97E-23 | 99.05 |
| *IGHM* | rs9671884 | G | C | 0.09 | 0.246 | 0.045 | 3.75E-08 | 29.88 |
| *IGHV3-38* | rs11627639 | T | C | 0.17 | -0.53 | 0.039 | 5.55E-43 | 184.68 |
| *IGHV3-38* | rs75453434 | C | A | 0.087 | -0.402 | 0.051 | 3.01E-15 | 62.13 |
| *IGHV3-38* | rs112394895 | A | G | 0.134 | -0.466 | 0.043 | 1.11E-27 | 117.45 |
| *IGKV1-9* | rs1102739 | C | G | 0.388 | 0.424 | 0.026 | 3.87E-58 | 265.94 |
| *IGKV1-9* | rs859750 | T | C | 0.064 | 0.295 | 0.053 | 2.27E-08 | 30.98 |
| *IGKV1-9* | rs11901621 | A | G | 0.398 | 0.415 | 0.026 | 3.55E-56 | 254.77 |
| *IGKV2-24* | rs11901621 | A | G | 0.399 | 0.283 | 0.04 | 2.76E-12 | 50.06 |
| *IL16* | rs3926278 | C | G | 0.2024 | -0.99 | 0.117 | 2.51E-17 | 71.60 |
| *IL17RA* | rs2241047 | C | G | 0.7876 | 0.737 | 0.081 | 6.31E-20 | 82.79 |
| *IL17RB* | rs768947265 | CCA | C | 0.6892 | -0.774 | 0.098 | 2.51E-15 | 62.38 |
| *IL18R1* | rs1861246 | T | C | 0.5781 | -0.699 | 0.087 | 1.26E-15 | 64.55 |
| *IL1RL1* | rs953934 | C | T | 0.561 | -0.666 | 0.083 | 1E-15 | 64.39 |
| *IL5RA* | rs17886680 | G | A | 0.3897 | 0.711 | 0.094 | 3.16E-14 | 57.21 |
| *IL6R* | rs12403159 | G | C | 0.3429 | 1.205 | 0.105 | 2E-30 | 131.70 |
| *IL7R* | rs6897932 | C | T | 0.1744 | -0.979 | 0.122 | 1E-15 | 64.39 |
| *ITIH3* | rs2276816 | C | T | 0.057 | 0.39 | 0.06 | 8.94E-11 | 42.25 |
| *ITIH3* | rs1060330 | T | C | 0.352 | 0.18 | 0.03 | 2.53E-09 | 36.00 |
| *ITIH3* | rs77017530 | C | T | 0.09 | -0.331 | 0.05 | 3.94E-11 | 43.82 |
| *ITIH3* | rs2286797 | G | A | 0.109 | -0.287 | 0.045 | 1.96E-10 | 40.68 |
| *ITIH3* | rs736408 | C | T | 0.321 | 0.142 | 0.024 | 4.47E-09 | 35.01 |
| *ITPA* | rs67002563 | G | A | 0.1633 | -1.407 | 0.148 | 2E-21 | 90.38 |
| *KIR2DS4* | rs17173106 | T | C | 0.4522 | 1.105 | 0.098 | 2E-29 | 127.14 |
| *KIR3DL1* | rs598452 | C | T | 0.4147 | -0.809 | 0.097 | 5.01E-17 | 69.56 |
| *KLB* | rs6531719 | A | G | 0.3592 | -0.754 | 0.097 | 7.94E-15 | 60.42 |
| *KLK12* | rs10635364 | T | TCC | 0.6605 | -1.249 | 0.108 | 6.31E-31 | 133.74 |
| *KLKB1* | rs4253278 | A | G | 0.373 | 0.171 | 0.029 | 3.02E-09 | 34.77 |
| *KLKB1* | rs10004932 | A | G | 0.448 | 0.167 | 0.031 | 4.99E-08 | 29.02 |
| *KNG1* | rs2689197 | C | T | 0.43 | -0.192 | 0.031 | 8.94E-10 | 38.36 |
| *KNG1* | rs2889832 | C | G | 0.389 | -0.17 | 0.029 | 7.57E-09 | 34.36 |
| *LAIR1* | rs74463408 | G | C | 0.2341 | -0.706 | 0.104 | 1E-11 | 46.08 |
| *LAIR2* | rs75571884 | C | G | 0.26 | 0.84 | 0.118 | 1E-12 | 50.68 |
| *LCP1* | rs4942487 | T | C | 0.9229 | -1.778 | 0.205 | 5.01E-18 | 75.22 |
| *LECT2* | rs248167 | A | G | 0.66 | -0.912 | 0.1 | 1E-19 | 83.17 |
| *LEFTY2* | rs360078 | G | A | 0.1596 | -0.841 | 0.121 | 3.98E-12 | 48.31 |
| *LEG1* | rs17855739 | C | T | 0.1222 | 1.176 | 0.162 | 3.98E-13 | 52.70 |
| *LILRA3* | rs1645788 | A | G | 0.5433 | -1.152 | 0.101 | 5.01E-30 | 130.10 |
| *LILRA6* | rs635711 | G | T | 0.6741 | -1.227 | 0.121 | 3.98E-24 | 102.83 |
| *LILRB2* | rs386056 | C | T | 0.5709 | -0.948 | 0.085 | 5.01E-29 | 124.39 |
| *LILRB5* | rs12975366 | T | C | 0.1191 | -0.9 | 0.101 | 3.98E-19 | 79.40 |
| *LPA* | rs7770628 | T | C | 0.103 | 0.401 | 0.049 | 4.06E-16 | 66.97 |
| *LPA* | rs4708876 | G | C | 0.104 | 0.346 | 0.042 | 7.76E-17 | 67.87 |
| *LPA* | rs578560 | A | G | 0.067 | 0.256 | 0.047 | 4.94E-08 | 29.67 |
| *LRG1* | rs4806987 | A | G | 0.107 | 0.415 | 0.059 | 2.73E-12 | 49.48 |
| *LRG1* | rs4807603 | T | G | 0.134 | 0.287 | 0.042 | 9.22E-12 | 46.69 |
| *LRIG1* | rs2306272 | T | C | 0.7058 | -1.069 | 0.107 | 1.26E-23 | 99.81 |
| *LRRC37A2* | rs374623679 | C | CA | 0.573 | -0.619 | 0.074 | 7.94E-17 | 69.97 |
| *LUM* | rs7135740 | A | T | 0.289 | -0.197 | 0.032 | 4.08E-10 | 37.90 |
| *LUM* | rs17018712 | T | C | 0.293 | -0.185 | 0.029 | 2.85E-10 | 40.70 |
| *LY75* | rs1397707 | C | T | 0.2945 | 0.723 | 0.096 | 3.98E-14 | 56.72 |
| *MAN2B2* | rs2301794 | G | A | 0.4364 | 0.802 | 0.091 | 7.94E-19 | 77.67 |
| *MANSC4* | rs11049143 | C | A | 0.1795 | 1.225 | 0.129 | 3.16E-21 | 90.18 |
| *MARK1* | rs12589136 | G | T | 0.388 | -0.237 | 0.035 | 1.43E-11 | 45.85 |
| *MARK1* | rs709932 | C | T | 0.233 | -0.503 | 0.066 | 2E-14 | 58.08 |
| *MBL2* | rs7899547 | T | G | 0.643 | 1.082 | 0.105 | 5.01E-25 | 106.19 |
| *MDGA1* | rs6937280 | T | G | 0.5859 | -0.994 | 0.09 | 1.58E-28 | 121.98 |
| *MEP1B* | rs680321 | T | C | 0.2513 | -1.067 | 0.111 | 5.01E-22 | 92.40 |
| *MIA* | rs2249854 | T | C | 0.1998 | 1.147 | 0.119 | 3.98E-22 | 92.90 |
| *MICB_MICA* | rs9348866 | A | T | 0.6178 | -1.019 | 0.102 | 1.26E-23 | 99.80 |
| *MRPS34* | rs760293 | C | T | 0.206 | 0.296 | 0.04 | 1.28E-13 | 54.76 |
| *MRPS34* | rs77633711 | C | T | 0.057 | -0.459 | 0.059 | 5.94E-15 | 60.52 |

| | | | | | | | | |
|---|---|---|---|---|---|---|---|---|
| *MSLN* | rs2235503 | C | A | 0.3506 | 0.907 | 0.109 | 1E-16 | 69.24 |
| *MSMB* | rs7911198 | G | A | 0.4914 | 0.917 | 0.092 | 3.16E-23 | 99.35 |
| *MST1* | rs11718165 | A | G | 0.0709 | -1.349 | 0.161 | 5.01E-17 | 70.21 |
| *MUC2* | rs8176719 | T | TC | 0.3482 | 0.742 | 0.108 | 5.01E-12 | 47.20 |
| *MYRIP* | rs79669707 | G | A | 0.068 | 0.336 | 0.039 | 9.35E-18 | 74.22 |
| *NOTCH2* | rs7551884 | C | T | 0.1267 | 1.667 | 0.2 | 7.94E-17 | 69.47 |
| *OBP2B* | rs10793959 | G | A | 0.6123 | 1.143 | 0.125 | 7.94E-20 | 83.61 |
| *PDCD6* | rs1971229 | A | G | 0.0634 | -1.413 | 0.172 | 2E-16 | 67.49 |
| *PDGFRB* | rs3816018 | C | T | 0.4805 | 1.298 | 0.104 | 7.94E-36 | 155.77 |
| *PF4* | rs3756074 | G | C | 0.142 | -0.27 | 0.039 | 3.37E-12 | 47.93 |
| *PGLYRP2* | rs6512033 | C | A | 0.286 | 0.569 | 0.04 | 1.28E-45 | 202.35 |
| *PGLYRP2* | rs10854145 | T | C | 0.117 | -0.217 | 0.034 | 2.44E-10 | 40.73 |
| *PGLYRP2* | rs57669263 | T | C | 0.124 | -0.229 | 0.039 | 4.88E-09 | 34.48 |
| *PGLYRP2* | rs2060262 | A | G | 0.09 | 0.252 | 0.04 | 3.15E-10 | 39.69 |
| *PGLYRP2* | rs919791 | G | A | 0.2645 | -1.138 | 0.108 | 6.31E-26 | 111.03 |
| *PILRA* | rs1859788 | A | G | 0.4353 | 0.93 | 0.089 | 2E-25 | 109.19 |
| *PILRB* | rs1859788 | A | G | 0.434 | 1.086 | 0.094 | 6.31E-31 | 133.48 |
| *PLA2G7* | rs76863441 | C | A | 0.0527 | -1.954 | 0.247 | 2E-15 | 62.58 |
| *PLB1* | rs6744005 | A | C | 0.7684 | 1.046 | 0.109 | 1E-21 | 92.09 |
| *PM20D1* | rs112268616 | T | C | 0.6875 | -1.23 | 0.122 | 1E-23 | 101.65 |
| *PNLIPRP2* | rs2286779 | G | C | 0.2846 | 1.273 | 0.116 | 7.94E-28 | 120.43 |
| *PON1* | rs62467349 | C | T | 0.078 | 0.394 | 0.053 | 9.45E-14 | 55.26 |
| *PON1* | rs13236941 | C | T | 0.366 | 0.314 | 0.03 | 2.06E-26 | 109.55 |
| *PON1* | rs2057682 | G | C | 0.231 | 0.312 | 0.034 | 6.3E-20 | 84.21 |
| *PON1* | rs10243325 | T | C | 0.442 | -0.169 | 0.029 | 5.25E-09 | 33.96 |
| *PON1* | rs705379 | G | A | 0.473 | -0.284 | 0.028 | 8.43E-24 | 102.88 |
| *PON1* | rs2237583 | T | C | 0.337 | 0.166 | 0.03 | 2.92E-08 | 30.62 |
| *PON2* | rs6961773 | T | G | 0.2019 | -0.917 | 0.12 | 2.51E-14 | 58.40 |
| *PROZ* | rs559054 | T | C | 0.481 | 0.377 | 0.023 | 7.09E-61 | 268.67 |
| *PROZ* | rs76645207 | A | G | 0.086 | -0.231 | 0.041 | 1.65E-08 | 31.74 |
| *PROZ* | rs7330624 | G | C | 0.467 | -0.179 | 0.023 | 1.59E-14 | 60.57 |
| *PROZ* | rs9549380 | G | A | 0.473 | -0.173 | 0.022 | 1.26E-14 | 61.84 |
| *PRPH2* | rs13343526 | G | C | 0.076 | -0.749 | 0.079 | 2.48E-21 | 89.89 |
| *PRPH2* | rs10407616 | C | G | 0.081 | -0.761 | 0.078 | 3.12E-22 | 95.19 |
| *PRR4* | rs3863320 | G | A | 0.6939 | -0.797 | 0.104 | 1.58E-14 | 58.73 |
| *PRSS53* | rs11150606 | T | C | 0.7168 | 0.71 | 0.101 | 2E-12 | 49.42 |
| *PSCA* | rs2976397 | G | T | 0.2251 | 1.072 | 0.099 | 3.98E-27 | 117.25 |
| *PTPRH* | rs12463268 | G | A | 0.2585 | 1.05 | 0.12 | 1.58E-18 | 76.56 |
| *PTPRM* | rs8176693 | C | T | 0.1868 | 1.072 | 0.13 | 2E-16 | 68.00 |
| *PVR* | rs7260482 | A | C | 0.4612 | -1.003 | 0.113 | 5.01E-19 | 78.79 |
| *PYDC1* | rs2298850 | G | C | 0.2346 | -0.923 | 0.127 | 3.98E-13 | 52.82 |
| *PZP* | rs2277413 | G | A | 0.487 | 0.247 | 0.025 | 1.98E-23 | 97.61 |
| *PZP* | rs11049897 | T | C | 0.483 | 0.143 | 0.025 | 9.5E-09 | 32.72 |
| *PZP* | rs11049351 | T | G | 0.422 | -0.195 | 0.02 | 5.79E-22 | 95.06 |
| *PZP* | rs12426061 | C | T | 0.332 | 0.261 | 0.023 | 5.76E-31 | 128.77 |
| *PZP* | rs7137576 | G | C | 0.143 | -0.284 | 0.03 | 4.67E-21 | 89.62 |
| *PZP* | rs139223232 | G | T | 0.182 | -0.225 | 0.027 | 6.67E-17 | 69.44 |
| *PZP* | rs1017301 | A | C | 0.41 | -0.218 | 0.021 | 2.24E-24 | 107.76 |
| *PZP* | rs10843050 | T | C | 0.448 | -0.287 | 0.023 | 1.35E-34 | 155.71 |
| *PZP* | rs78315493 | A | G | 0.104 | -0.33 | 0.048 | 5.69E-12 | 47.27 |
| *PZP* | rs888118 | A | C | 0.493 | 0.205 | 0.023 | 1.75E-19 | 79.44 |
| *PZP* | rs2377744 | C | T | 0.176 | -0.288 | 0.034 | 3.16E-17 | 71.75 |
| *PZP* | rs79910264 | T | C | 0.067 | -0.252 | 0.044 | 1.45E-08 | 32.80 |
| *PZP* | rs11048023 | G | A | 0.369 | 0.128 | 0.023 | 1.96E-08 | 30.97 |
| *PZP* | rs7311982 | C | T | 0.486 | -0.338 | 0.022 | 2.88E-52 | 236.04 |
| *PZP* | rs1017302 | G | A | 0.113 | -0.384 | 0.044 | 1.3E-18 | 76.17 |
| *QPCT* | rs10189564 | G | A | 0.3199 | -1.267 | 0.116 | 1E-27 | 119.30 |
| *RNASE10* | rs7144945 | C | T | 0.3878 | -0.699 | 0.08 | 3.16E-18 | 76.34 |
| *RPIA* | rs200567660 | C | G | 0.228 | -0.283 | 0.03 | 2.4E-21 | 88.99 |
| *RPIA* | rs1407 | C | G | 0.228 | -0.27 | 0.03 | 3.5E-19 | 81.00 |
| *SAA1* | rs75846249 | C | T | 0.072 | 0.255 | 0.043 | 2.99E-09 | 35.17 |
| *SAA1* | rs11024588 | A | G | 0.489 | 0.33 | 0.028 | 3.79E-32 | 138.90 |
| *SAA1* | rs12271810 | A | G | 0.179 | -0.161 | 0.029 | 3.8E-08 | 30.82 |
| *SAA2* | rs1520886 | C | T | 0.438 | -0.145 | 0.024 | 1.48E-09 | 36.50 |
| *SAA2* | rs11024588 | A | G | 0.489 | 0.439 | 0.027 | 3.31E-58 | 264.36 |
| *SAA2* | rs12271810 | A | G | 0.178 | -0.224 | 0.031 | 3.38E-13 | 52.21 |
| *SAA2* | rs4150674 | A | G | 0.126 | -0.293 | 0.036 | 5.38E-16 | 66.24 |
| *SAA2* | rs141771998 | C | G | 0.063 | 0.364 | 0.048 | 4.54E-14 | 57.51 |

| | | | | | | | | |
|---|---|---|---|---|---|---|---|---|
| *SAA2* | rs79669707 | G | A | 0.068 | 0.343 | 0.049 | 2.52E-12 | 49.00 |
| *SAA2* | rs7945581 | T | G | 0.259 | -0.27 | 0.027 | 9.34E-24 | 100.00 |
| *SAA2* | rs7925813 | G | A | 0.476 | -0.376 | 0.027 | 5.91E-44 | 193.93 |
| *SAA4* | rs2468789 | G | T | 0.112 | 0.298 | 0.047 | 1.74E-10 | 40.20 |
| *SAA4* | rs2956632 | A | G | 0.112 | 0.354 | 0.051 | 5.33E-12 | 48.18 |
| *SERPINA1* | rs1980618 | T | A | 0.357 | -0.242 | 0.03 | 1.05E-15 | 65.07 |
| *SERPINA1* | rs709932 | C | T | 0.233 | -0.794 | 0.046 | 1.52E-67 | 297.94 |
| *SERPINA11* | rs11624472 | G | A | 0.1896 | -0.876 | 0.119 | 1.58E-13 | 54.19 |
| *SERPINA12* | rs74644698 | C | T | 0.0412 | 2.059 | 0.243 | 2.51E-17 | 71.80 |
| *SERPINA4* | rs10142033 | A | T | 0.227 | 0.201 | 0.034 | 5.37E-09 | 34.95 |
| *SERPINA4* | rs8005151 | G | A | 0.29 | 0.674 | 0.098 | 6.31E-12 | 47.30 |
| *SERPINB8* | rs3826616 | A | G | 0.5465 | -0.732 | 0.089 | 3.16E-16 | 67.65 |
| *SERPIND1* | rs77303550 | C | T | 0.297 | 0.219 | 0.032 | 9.21E-12 | 46.84 |
| *SERPINF1* | rs34665863 | G | A | 0.483 | -0.159 | 0.028 | 2.22E-08 | 32.25 |
| *SERPINF1* | rs62088172 | C | T | 0.462 | -0.173 | 0.029 | 2.28E-09 | 35.59 |
| *SERPINF1* | rs8074840 | T | C | 0.368 | -0.202 | 0.03 | 7.46E-12 | 45.34 |
| *SERPINF2* | rs4790816 | A | T | 0.185 | -0.54 | 0.036 | 1.68E-50 | 225.00 |
| *SERPING1* | rs12292224 | A | G | 0.054 | -0.623 | 0.073 | 2.11E-17 | 72.83 |
| *SERPING1* | rs714942 | T | C | 0.076 | -0.484 | 0.085 | 1.47E-08 | 32.42 |
| *SERPINI2* | rs75179845 | T | C | 0.1905 | 1.081 | 0.134 | 7.94E-16 | 65.08 |
| *SFTPA2* | rs17886395 | C | G | 0.25 | 1.102 | 0.135 | 2.51E-16 | 66.63 |
| *SFTPD* | rs721917 | A | G | 0.6621 | -1.098 | 0.115 | 1.58E-21 | 91.16 |
| *SGSH* | rs7503034 | C | T | 0.3996 | -0.85 | 0.088 | 6.31E-22 | 93.30 |
| *SHMT1* | rs921985 | T | C | 0.5411 | 0.653 | 0.094 | 3.98E-12 | 48.26 |
| *SIGLEC5* | rs3829655 | G | C | 0.5027 | -0.838 | 0.106 | 3.16E-15 | 62.50 |
| *SIGLEC9* | rs273691 | C | T | 0.5365 | -0.619 | 0.085 | 3.98E-13 | 53.03 |
| *SIRPA* | rs1135192 | G | A | 0.6786 | -1.132 | 0.092 | 1.58E-34 | 151.40 |
| *SPAG1* | rs2088199 | A | T | 0.6442 | -0.81 | 0.11 | 2E-13 | 54.22 |
| *SPINK2* | rs1708768 | G | A | 0.0669 | 1.554 | 0.207 | 6.31E-14 | 56.36 |
| *SPINK4* | rs477706 | A | C | 0.1552 | 1.178 | 0.145 | 3.98E-16 | 66.00 |
| *SPINK5* | rs11405253 | G | GT | 0.4014 | -0.709 | 0.096 | 1.58E-13 | 54.54 |
| *TDGF1* | rs6769392 | G | A | 0.1562 | 1.214 | 0.121 | 7.94E-24 | 100.66 |
| *TF* | rs6794370 | C | A | 0.297 | -0.225 | 0.031 | 6.54E-13 | 52.68 |
| *TF* | rs8177204 | C | T | 0.408 | 0.168 | 0.029 | 1.01E-08 | 33.56 |
| *TF* | rs1405022 | C | T | 0.469 | 0.172 | 0.029 | 1.94E-09 | 35.18 |
| *TF* | rs1358021 | A | C | 0.464 | 0.196 | 0.031 | 3.5E-10 | 39.98 |
| *TF* | rs16840612 | C | A | 0.503 | 0.156 | 0.028 | 3.63E-08 | 31.04 |
| *TF* | rs1049296 | C | T | 0.24 | -0.346 | 0.05 | 3.11E-12 | 47.89 |
| *TF* | rs202128708 | C | T | 0.155 | 0.329 | 0.033 | 1.1E-23 | 99.39 |
| *TF* | rs6797713 | T | C | 0.426 | -0.464 | 0.024 | 1.8E-84 | 373.78 |
| *TF* | rs59486895 | T | A | 0.323 | 0.784 | 0.025 | 1E-200 | 983.45 |
| *TF* | rs1106170 | G | A | 0.057 | 0.528 | 0.051 | 2.56E-25 | 107.18 |
| *TF* | rs8177215 | G | A | 0.278 | -0.215 | 0.032 | 2.68E-11 | 45.14 |
| *TF* | rs2718796 | C | G | 0.263 | -0.193 | 0.033 | 2.89E-09 | 34.20 |
| *TF* | rs3811658 | C | T | 0.409 | 0.171 | 0.031 | 2.64E-08 | 30.43 |
| *TF* | rs6785365 | G | A | 0.07 | -0.348 | 0.055 | 2.4E-10 | 40.03 |
| *TF* | rs13080154 | T | C | 0.49 | 0.275 | 0.028 | 1.49E-22 | 96.46 |
| *TF* | rs3811657 | G | A | 0.172 | -0.93 | 0.036 | 2.23E-144 | 667.36 |
| *TF* | rs6781444 | A | C | 0.095 | -0.873 | 0.058 | 2.48E-50 | 226.55 |
| *TF* | rs116974632 | C | T | 0.114 | 0.325 | 0.046 | 1.17E-12 | 49.92 |
| *TF* | rs7616492 | G | A | 0.25 | -0.193 | 0.033 | 4.41E-09 | 34.20 |
| *TF* | rs6794361 | A | G | 0.463 | 0.19 | 0.031 | 1.35E-09 | 37.57 |
| *TF* | rs8177197 | G | A | 0.408 | 0.361 | 0.042 | 5.63E-18 | 73.88 |
| *TF* | rs6806081 | G | A | 0.053 | -0.492 | 0.073 | 1.31E-11 | 45.42 |
| *TF* | rs143071821 | T | C | 0.401 | 0.178 | 0.029 | 8.79E-10 | 37.67 |
| *TGFBI* | rs1990201 | T | C | 0.5931 | 0.721 | 0.094 | 2E-14 | 58.83 |
| *TIMP3* | rs1079734 | G | A | 0.5996 | -0.728 | 0.103 | 1.58E-12 | 49.96 |
| *TLR3* | rs3775291 | C | T | 0.312 | -1.021 | 0.098 | 1.26E-25 | 108.54 |
| *TNN* | rs77382942 | T | A | 0.5082 | -1.088 | 0.103 | 3.98E-26 | 111.58 |
| *TNXB* | rs9267785 | C | T | 0.0562 | 1.636 | 0.207 | 2.51E-15 | 62.46 |
| *TOP1* | rs112268616 | T | C | 0.6913 | -0.928 | 0.125 | 1E-13 | 55.12 |
| *TREH* | rs34808635 | A | ACT | 0.6795 | -0.929 | 0.091 | 2.51E-24 | 104.22 |
| *TXNDC15* | rs3733897 | A | G | 0.2381 | 1.218 | 0.131 | 1.26E-20 | 86.45 |
| *UGT2B11* | rs117095836 | G | A | 0.054 | -0.661 | 0.1 | 3.38E-11 | 43.69 |
| *VCAN* | rs188686 | T | A | 0.3946 | -0.678 | 0.096 | 2E-12 | 49.88 |
| *VMO1* | rs9892407 | C | G | 0.4781 | 0.756 | 0.092 | 2E-16 | 67.53 |
| *VNN1* | rs3798800 | A | C | 0.3348 | -1.136 | 0.108 | 7.94E-26 | 110.64 |
| *VNN2* | rs6935500 | G | A | 0.2646 | -0.815 | 0.116 | 2E-12 | 49.36 |

| | | | | | | | | |
|---|---|---|---|---|---|---|---|---|
| *VSIG10* | rs9669474 | T | C | 0.2456 | 0.664 | 0.098 | 1.26E-11 | 45.91 |
| *VWC2* | rs481076 | T | G | 0.4173 | -0.714 | 0.089 | 7.94E-16 | 64.36 |
| *VWF* | rs687621 | A | G | 0.38 | 0.324 | 0.033 | 2.01E-22 | 96.40 |
| *VWF* | rs687289 | G | A | 0.372 | 0.289 | 0.026 | 2.84E-28 | 123.55 |
| *VWF* | rs4962113 | T | C | 0.343 | -0.186 | 0.026 | 6.28E-13 | 51.18 |
| *VWF* | rs8176645 | T | A | 0.372 | 0.278 | 0.028 | 2E-23 | 98.58 |
| *XCL1* | rs12042332 | A | G | 0.635 | 0.914 | 0.103 | 5.01E-19 | 78.74 |
| *ZP3* | rs10261314 | T | C | 0.5819 | 0.849 | 0.091 | 7.94E-21 | 87.04 |

**Supplementary Table 3: Two-sample MR results in European populations**

| Protein-encoding gene | No. of SNPs | Method | Beta | SE | p-value | FDR | OR | Lower 95% CI | Upper 95% CI |
|---|---|---|---|---|---|---|---|---|---|
| *AKT3* | 1 | Wald ratio | 0.751 | 0.180 | 3.03E-05 | 1.09E-02 | 2.12 | 1.49 | 3.01 |
| *BST1* | 3 | Weighted median | 0.039 | 0.010 | 6.01E-05 | 1.67E-02 | 1.04 | 1.02 | 1.06 |
| *BST1* | 3 | Inverse variance weighted | 0.039 | 0.010 | 5.48E-05 | 1.59E-02 | 1.04 | 1.02 | 1.06 |
| *C2CD2L* | 1 | Wald ratio | 0.895 | 0.180 | 7.03E-07 | 3.31E-04 | 2.45 | 1.72 | 3.48 |
| *C5* | 1 | Wald ratio | -0.246 | 0.066 | 2.11E-04 | 4.81E-02 | 0.78 | 0.69 | 0.89 |
| *CCN5* | 1 | Wald ratio | 0.804 | 0.198 | 4.81E-05 | 1.45E-02 | 2.24 | 1.52 | 3.29 |
| *CD160* | 10 | Weighted median | -0.112 | 0.024 | 2.54E-06 | 1.06E-03 | 0.89 | 0.85 | 0.94 |
| *CD160* | 10 | Inverse variance weighted | -0.094 | 0.024 | 6.33E-05 | 1.70E-02 | 0.91 | 0.87 | 0.95 |
| *CDHR1* | 3 | Inverse variance weighted | 0.407 | 0.085 | 1.44E-06 | 6.38E-04 | 1.50 | 1.27 | 1.77 |
| *CTSB* | 5 | Weighted median | -0.144 | 0.023 | 3.57E-10 | 3.84E-07 | 0.87 | 0.83 | 0.91 |
| *EFCAB14* | 1 | Wald ratio | 1.587 | 0.209 | 2.94E-14 | 1.02E-10 | 4.89 | 3.25 | 7.36 |
| *EFNA1* | 4 | Weighted median | 0.148 | 0.029 | 5.08E-07 | 2.73E-04 | 1.16 | 1.09 | 1.23 |
| *ENTPD2* | 1 | Wald ratio | 1.084 | 0.190 | 1.23E-08 | 1.16E-05 | 2.96 | 2.04 | 4.29 |
| *FCGR2A* | 4 | Weighted median | 0.051 | 0.010 | 2.65E-07 | 1.81E-04 | 1.05 | 1.03 | 1.07 |
| *FCGR2A* | 4 | Inverse variance weighted | 0.051 | 0.010 | 2.96E-07 | 1.84E-04 | 1.05 | 1.03 | 1.07 |
| *FCGR2B* | 2 | Inverse variance weighted | -0.141 | 0.021 | 9.40E-12 | 1.41E-08 | 0.87 | 0.83 | 0.90 |
| *HIP1R* | 1 | Wald ratio | 1.556 | 0.121 | 8.36E-38 | 6.29E-34 | 4.74 | 3.74 | 6.01 |
| *HLA-DRA* | 3 | Weighted median | -0.225 | 0.036 | 3.36E-10 | 3.84E-07 | 0.80 | 0.74 | 0.86 |
| *HRC* | 4 | Weighted median | 0.375 | 0.096 | 9.95E-05 | 2.50E-02 | 1.45 | 1.20 | 1.76 |
| *IDUA* | 10 | Weighted median | -0.088 | 0.017 | 3.18E-07 | 1.84E-04 | 0.92 | 0.89 | 0.95 |
| *IDUA* | 10 | Inverse variance weighted | -0.084 | 0.016 | 1.65E-07 | 1.24E-04 | 0.92 | 0.89 | 0.95 |
| *INPP5J* | 1 | Wald ratio | 0.850 | 0.156 | 4.96E-08 | 4.14E-05 | 2.34 | 1.72 | 3.18 |
| *LAMP3* | 8 | Weighted median | 0.170 | 0.041 | 3.66E-05 | 1.20E-02 | 1.19 | 1.09 | 1.28 |
| *MYLPF* | 1 | Wald ratio | -0.669 | 0.180 | 1.98E-04 | 4.65E-02 | 0.51 | 0.36 | 0.73 |
| *PRSS53* | 7 | Weighted median | -0.086 | 0.011 | 4.06E-14 | 1.02E-10 | 0.92 | 0.90 | 0.94 |
| *PRSS53* | 7 | Inverse variance weighted | -0.085 | 0.012 | 3.32E-13 | 6.25E-10 | 0.92 | 0.90 | 0.94 |
| *TEF* | 1 | Wald ratio | -0.539 | 0.137 | 8.18E-05 | 2.12E-02 | 0.58 | 0.45 | 0.76 |
| *TPM3* | 1 | Wald ratio | -0.636 | 0.169 | 1.66E-04 | 4.04E-02 | 0.53 | 0.38 | 0.74 |
| *TRIM40* | 1 | Wald ratio | 0.213 | 0.043 | 5.73E-07 | 2.87E-04 | 1.24 | 1.14 | 1.34 |
| *TSPAN7* | 1 | Wald ratio | 0.847 | 0.188 | 6.28E-06 | 2.49E-03 | 2.33 | 1.62 | 3.37 |
| *TXNDC15* | 6 | Weighted median | -0.054 | 0.012 | 6.94E-06 | 2.61E-03 | 0.95 | 0.93 | 0.97 |
| *TXNDC15* | 6 | Inverse variance weighted | -0.053 | 0.013 | 3.47E-05 | 1.19E-02 | 0.95 | 0.92 | 0.97 |
| *TYRO3* | 6 | Weighted median | 0.062 | 0.015 | 4.43E-05 | 1.39E-02 | 1.06 | 1.03 | 1.10 |

**Supplementary Table 4: Two-sample MR results in East Asian populations**

| Protein-encoding gene | No. of SNPs | Method | Beta | SE | p-value | FDR | OR | Lower 95% CI | Upper 95% CI |
|---|---|---|---|---|---|---|---|---|---|
| *APOM* | 1 | Wald ratio | 0.264 | 0.086 | 2.18E-03 | 0.048 | 1.30 | 1.10 | 1.54 |
| *BST1* | 1 | Wald ratio | 0.168 | 0.035 | 1.50E-06 | 2.34E-04 | 1.18 | 1.10 | 1.27 |
| *C4A* | 3 | Inverse variance weighted | -0.179 | 0.056 | 1.53E-03 | 0.045 | 0.84 | 0.75 | 0.93 |
| *C9orf131* | 2 | Inverse variance weighted | -0.228 | 0.073 | 1.74E-03 | 0.045 | 0.80 | 0.69 | 0.92 |
| *HDGF* | 1 | Wald ratio | 0.108 | 0.033 | 1.12E-03 | 0.044 | 1.11 | 1.04 | 1.19 |
| *MRPS34* | 2 | Inverse variance weighted | -0.220 | 0.073 | 2.46E-03 | 0.048 | 0.80 | 0.70 | 0.93 |
| *PM20D1* | 1 | Wald ratio | -0.082 | 0.020 | 5.88E-05 | 3.06E-03 | 0.92 | 0.88 | 0.96 |
| *TOP1* | 1 | Wald ratio | -0.109 | 0.027 | 5.88E-05 | 3.06E-03 | 0.90 | 0.85 | 0.95 |

**Supplementary Table 5: Two-sample MR results for proteins shared between European and East Asian populations**

| Protein-encoding gene | Ancestry | No. of SNPs | Method | Beta | SE | p-value | FDR | OR | Lower 95% CI | Upper 95% CI |
|---|---|---|---|---|---|---|---|---|---|---|
| *BST1* | European | 3 | MR Egger | 0.043 | 0.016 | 0.227 | 0.991 | 1.04 | 1.01 | 1.08 |
| *BST1* | European | 3 | Inverse variance weighted | 0.039 | 0.010 | 5.48E-05 | 6.04E-03 | 1.04 | 1.02 | 1.06 |
| *BST1* | East Asian | 1 | Wald ratio | 0.168 | 0.035 | 1.50E-06 | 1.87E-04 | 1.18 | 1.10 | 1.27 |
| *HDGF* | European | 2 | Inverse variance weighted | 0.009 | 0.011 | 0.420 | 0.991 | 1.01 | 0.99 | 1.03 |
| *HDGF* | East Asian | 1 | Wald ratio | 0.108 | 0.033 | 1.12E-03 | 0.035 | 1.11 | 1.04 | 1.19 |
| *PM20D1* | European | 6 | MR Egger | -0.028 | 0.182 | 0.884 | 0.991 | 0.97 | 0.68 | 1.39 |
| *PM20D1* | European | 6 | Inverse variance weighted | -0.015 | 0.065 | 0.819 | 0.991 | 0.99 | 0.87 | 1.12 |
| *PM20D1* | East Asian | 1 | Wald ratio | -0.082 | 0.020 | 5.88E-05 | 2.45E-03 | 0.92 | 0.88 | 0.96 |
| *PRSS53* | European | 7 | MR Egger | -0.091 | 0.016 | 2.28E-03 | 0.126 | 0.91 | 0.89 | 0.94 |
| *PRSS53* | European | 7 | Inverse variance weighted | -0.085 | 0.012 | 3.32E-13 | 1.01E-10 | 0.92 | 0.90 | 0.94 |
| *PRSS53* | East Asian | 1 | Wald ratio | 0.070 | 0.040 | 0.081 | 0.727 | 1.07 | 0.99 | 1.16 |
| *TOP1* | European | 5 | MR Egger | -0.271 | 0.189 | 0.247 | 0.991 | 0.76 | 0.53 | 1.10 |
| *TOP1* | European | 5 | Inverse variance weighted | 0.043 | 0.088 | 0.624 | 0.991 | 1.04 | 0.88 | 1.24 |
| *TOP1* | East Asian | 1 | Wald ratio | -0.109 | 0.027 | 5.88E-05 | 2.45E-03 | 0.90 | 0.85 | 0.95 |
| *TXNDC15* | European | 6 | MR Egger | -0.064 | 0.015 | 0.011 | 0.462 | 0.94 | 0.91 | 0.96 |
| *TXNDC15* | European | 6 | Inverse variance weighted | -0.053 | 0.013 | 3.47E-05 | 5.27E-03 | 0.95 | 0.92 | 0.97 |
| *TXNDC15* | East Asian | 1 | Wald ratio | -0.064 | 0.023 | 4.96E-03 | 0.089 | 0.94 | 0.90 | 0.98 |

**Supplementary Table 6: Heterogeneity test results in European populations**

| Outcome | Exposure | Method | Q | Q_df |
|---|---|---|---|---|
| pd | *CD160* | Inverse variance weighted | 11.202 | 9 |
| pd | *CD160* | MR Egger | 7.111 | 8 |
| pd | *FCGR2A* | Inverse variance weighted | 1.210 | 3 |
| pd | *FCGR2A* | MR Egger | 1.155 | 2 |
| pd | *IDUA* | Inverse variance weighted | 7.027 | 9 |
| pd | *IDUA* | MR Egger | 4.939 | 8 |
| pd | *PRSS53* | Inverse variance weighted | 6.987 | 6 |
| pd | *PRSS53* | MR Egger | 6.549 | 5 |
| pd | *TXNDC15* | Inverse variance weighted | 5.840 | 5 |
| pd | *TXNDC15* | MR Egger | 3.999 | 4 |

| Q_pval |
| --- |
| 0.322 |
| 0.406 |
| 0.262 |
| 0.525 |
| 0.634 |
| 0.764 |
| 0.322 |
| 0.256 |
| 0.751 |
| 0.561 |

**Supplementary Table 7: MR-Egger sensitivity analysis in European populations**

| Outcome | Exposure | Egger_intercept | SE | p-value |
|---|---|---|---|---|
| pd | *CD160* | 0.012 | 0.006 | 0.078 |
| pd | *FCGR2A* | -0.002 | 0.009 | 0.837 |
| pd | *IDUA* | 0.008 | 0.006 | 0.186 |
| pd | *PRSS53* | 0.004 | 0.007 | 0.588 |
| pd | *TXNDC15* | 0.011 | 0.008 | 0.246 |

**Supplementary Table 8: Drug triangulation of proteins identified with causal impact on PD**

| Protein-encoding gene | Protein | Known Function/Gene Ontology annotations | Drug name | Clinical phase | Indications |
|---|---|---|---|---|---|
| *BST1* | bone marrow stromal cell antigen 1 | 1. Functions as an enzyme catalysing both the formation of cyclic ADP-beta-D-ribose (cADPR) from the oxidised form of nicotinamide adenine dinucleotide (NAD+) and its breakdown into ADP-D-ribose (ADPR).<br>2. Exhibits NAD+ nucleosidase and transferase activity.<br>3. The amino acid sequence shares 33% similarity with *CD38* . | nicotinamide | Approved, investigational | Adjunct treatment for gastrointestinal and hepatic insufficiencies, macrocytic anaemia, and secondary anaemia. |
| | | | nicotinamide riboside | Investigational (NCT03568968) | NA |
| *HDGF* | hepatoma derived growth factor | 1. Serves as a transcriptional repressor and possesses mitogenic effects on fibroblasts.<br>2. Binds to heparin and has RNA-binding and growth factor activity. | copper | Approved, investigational | Used in total parenteral nutrition supplementation. |
| *PM20D1* | Peptidase M20 Domain Containing 1 | 1. A secreted enzyme involved in regulating N-fatty acyl amino acids (NAAs) levels in tissues and circulation. It facilitates both the synthesis and hydrolysis of NAAs, which impact oxidative metabolism through mitochondrial uncoupling, thereby influencing overall energy expenditure.<br>2. Circulates in association with low-density lipoprotein (LDL) and high-density lipoprotein (HDL) particles.<br>3. Genetically altered *PM20D1* expression modulates Alzheimer's disease pathology, with increased expression providing neuroprotection.<br>4. Exhibits hydrolase, peptidase, and amidase activities. | cannabidiol | Approved, investigational | Adjunctive treatment in multiple sclerosis-related spasticity, neuropathic pain, and analgesia for advanced cancer pain unresponsive to high-dose opioids. |
| | | | fostamatinib | Approved, investigational | Treatment of chronic immune thrombocytopenia in patients with inadequate response to prior therapies. |
| | | | JNJ-42165279 | Investigational (NCT02498392) | NA |
| *PRSS53* | Serine Protease 53 | 1. Capable of degrading the fibrinogen alpha chain and pro-urokinase-type plasminogen activator in vitro.<br>2. Predicted to function as a serine-type endopeptidase involved in proteolysis and located in extracellular regions. | warfarin | Approved | Prevention and treatment of venous thromboembolism. |
| *TOP1* | DNA Topoisomerase 1 | 1. Alleviates supercoiling and torsional stress in DNA during replication and transcription by creating temporary breaks in one strand of the DNA double helix, which are then rejoined. Facilitates single-strand cleavage via transesterification, enabling strand rotation to relieve supercoils before re-ligation.<br>2. Modulates the alternative splicing of F3 pre-mRNA in endothelial cells and plays a role in circadian transcription by modifying chromatin structure near *BMAL1* promoter sites.<br>3. Demonstrates ATP-binding activity. | irinotecan | Approved, investigational | metastatic colorectal carcinoma |
| | | | trastuzumab deruxtecan | Approved, investigational | Treatment of certain types of HER2-positive breast cancer that are unresectable or metastatic. |
| *TXNDC15* | Thioredoxin Domain-Containing Protein 15 | 1. Positively regulates ciliary hedgehog signalling and contributes to ciliogenesis. | None | NA | NA |

**Supplementary Table 9: Comparison of MR estimates with previous studies across tissue and QTL types**

| Protein-encoding gene | Data source | Population | Tissue | QTL type | No. of SNPs | Method | Beta | SE | p-value | Adjusted p-value | OR (95% CI) | Direction | Reference |
|---|---|---|---|---|---|---|---|---|---|---|---|---|---|
| *ACP1* | **Sun 2023** | **European** | **blood** | **pQTL** | **1** | **Wald ratio** | **-0.084** | **0.026** | **1.22E-03** | **0.191** | **0.92 (0.87, 0.97)** | **-** | **current study** |
| | PPMI | European | CSF | pQTL | 15 | IVW | 0.040 | NR | NR | 0.011 | NR | + | Kaiser 2023 |
| *ADAMTS4* | **Sun 2023** | **European** | **blood** | **pQTL** | **2** | **IVW** | 0.165 | 0.074 | 0.026 | 1.000 | **1.18 (1.02, 1.36)** | **+** | **current study** |
| | PPMI | European | CSF | pQTL | 8 | IVW | -0.060 | NR | NR | 0.020 | NR | - | Kaiser 2023 |
| *BST1* | **Xu 2023** | **East Asian** | **blood** | **pQTL** | **1** | **Wald ratio** | **0.168** | **0.035** | **1.50E-06** | **2.34E-04** | **1.18 (1.10, 1.54)** | **+** | **current study** |
| | **Sun 2023** | **European** | **blood** | **pQTL** | **3** | **IVW** | **0.039** | **0.010** | **5.48E-05** | **0.016** | **1.04 (1.02, 1.06)** | **+** | **current study** |
| | Sun 2018 | European | blood | pQTL | 1 | Wald ratio | 0.059 | 0.016 | 3.46E-04 | 0.001 | 1.06 (1.03, 1.10) | + | Storm 2021 |
| | Suhre 2017 | European | blood | pQTL | 1 | Wald ratio | 0.052 | 0.019 | 7.17E-03 | 0.010 | NR | + | Storm 2021 |
| | PPMI | European | CSF | pQTL | 10 | IVW | 0.120 | NR | NR | 4.00E-05 | NR | + | Kaiser 2023 |
| | eQTLGen, PsychENCODE | European | blood | eQTL | 2 | IVW | -0.147 | 0.042 | 4.78E-04 | 0.001 | 0.86 (0.79, 0.94) | - | Storm 2021 |
| *C4A* | **Xu 2023** | **East Asian** | **blood** | **pQTL** | **3** | **IVW** | **-0.179** | **0.056** | **1.53E-03** | **0.045** | **0.84 (0.75, 0.93)** | **-** | **current study** |
| | PPMI | European | CSF | pQTL | 101 | IVW | 0.020 | NR | NR | 0.002 | NR | + | Kaiser 2023 |
| *CD38* | **Sun 2023** | **European** | **blood** | **pQTL** | **2** | **IVW** | **-0.015** | **0.032** | **0.650** | **1.000** | 0.99 (0.93, 1.05) | **-** | **current study** |
| | Hillary 2019 | European | blood | pQTL | 1 | Wald ratio | 0.044 | 0.048 | 0.362 | 0.362 | 1.04 (0.95, 1.15) | + | Storm 2021 |
| | ROSMAP | European | brain | pQTL | 1 | Wald ratio | NR | NR | NR | 6.99E-14 | 0.32 (0.24, 0.43) | - | Gu 2023 |
| | PPMI | European | CSF | pQTL | 1 | Wald ratio | -0.530 | NR | NR | 2.45E-09 | NR | - | Kaiser 2023 |
| | eQTLGen,PsychENCODE | European | brain | eQTL | 1 | IVW | -0.537 | 0.216 | 0.013 | 0.014 | 0.58 (0.38, 0.89) | - | Storm 2021 |
| *CTSB* | **Sun 2023** | **European** | **blood** | **pQTL** | **5** | **IVW** | **-0.135** | **0.042** | **1.41E-03** | **0.207** | 0.87 (0.80, 0.95) | **-** | **current study** |
| | Emilsson 2018 | European | blood | pQTL | 1 | Wald ratio | -0.154 | 0.031 | 8.59E-07 | 2.00E-06 | 0.86 (0.81, 0.91) | - | Storm 2021 |
| | Suhre 2017 | European | blood | pQTL | 1 | Wald ratio | -0.180 | 0.036 | 6.23E-07 | 2.00E-06 | 0.84 (0.78, 0.90) | - | Storm 2021 |
| | ROSMAP | European | brain | pQTL | 1 | Wald ratio | NR | NR | NR | 4.39E-05 | 0.20 (0.09, 0.43) | - | Gu 2023 |
| | Yang 2021 | European | CSF | pQTL | 1 | Wald ratio | NR | NR | NR | 2.19E-05 | 0.19 (0.09, 0.41) | - | Gu 2023 |
| | PPMI | European | CSF | pQTL | 12 | IVW | -0.110 | NR | NR | 2.63E-10 | NR | - | Kaiser 2023 |
| | eQTLGen,PsychENCODE | European | brain | eQTL | 2 | IVW | -0.361 | 0.111 | 1.12E-03 | 0.001 | 0.70 (0.56, 0.87) | - | Storm 2021 |
| | Nalls 2019 | European | blood | eQTL | 33 | not stated | -0.150 | 0.040 | 4.37E-09 | 4.50E-07 | NR | - | Nalls 2019 |
| *FCGR2A* | **Sun 2023** | **European** | **blood** | **pQTL** | **4** | **IVW** | **0.051** | **0.010** | **2.96E-07** | **1.84E-04** | **1.05 (1.03, 1.07)** | **+** | **current study** |
| | Sun 2018 | European | blood | pQTL | 3 | IVW | NR | NR | NR | 1.76E-05 | 1.06 (1.03, 1.09) | + | Gu 2023 |
| | PPMI | European | CSF | pQTL | 18 | IVW | 0.070 | NR | NR | 2.70E-12 | NR | + | Kaiser 2023 |
| *FCGR2A* | **Sun 2023** | **European** | **blood** | **pQTL** | **2** | **IVW** | **-0.141** | **0.021** | **9.40E-12** | **1.41E-08** | **0.87 (0.83, 0.90)** | **-** | **current study** |
| | PPMI | European | CSF | pQTL | 17 | IVW | 0.070 | NR | NR | 5.11E-14 | NR | + | Kaiser 2023 |
| *GPNMB* | **Sun 2023** | **European** | **blood** | **pQTL** | **3** | **IVW** | **-0.033** | **0.051** | **0.512** | **1.000** | 0.97 (0.88, 1.07) | **-** | **current study** |
| | Sun 2018 | European | blood | pQTL | 4 | IVW | 0.209 | 0.132 | 0.113 | 0.141 | 1.23 (0.95, 1.60) | + | Storm 2021 |
| | Sun 2018 | European | blood | pQTL | 1 | Wald ratio | NR | NR | NR | 0.000 | 1.64 (1.37, 1.97) | + | Gu 2023 |
| | Emilsson 2018 | European | blood | pQTL | 1 | Wald ratio | 0.428 | 0.064 | 2.45E-11 | 1.71E-10 | NR | + | Storm 2021 |
| | Suhre 2017 | European | blood | pQTL | 2 | IVW | 0.153 | 0.159 | 0.335 | 0.362 | 1.17 (0.85, 1.59) | + | Storm 2021 |
| | ROSMAP | European | brain | pQTL | 1 | Wald ratio | NR | NR | NR | 2.48E-08 | 1.47 (1.28, 1.68) | + | Gu 2023 |
| | Yang 2021 | European | CSF | pQTL | 1 | Wald ratio | NR | NR | NR | 3.95E-07 | 2.37 (1.17, 3.31) | + | Gu 2023 |
| | PPMI | European | CSF | pQTL | 11 | IVW | 0.140 | NR | NR | 1.17E-17 | NR | + | Kaiser 2023 |
| | eQTLGen,PsychENCODE | European | blood | eQTL | 1 | Wald ratio | 0.281 | 0.041 | 7.51E-12 | 5.26E-11 | 1.33 (1.22, 1.44) | + | Storm 2021 |
| | Nalls 2019 | European | blood | eQTL | 74 | not stated | 0.090 | 0.010 | 2.73E-21 | 2.81E-19 | NR | + | Nalls 2019 |
| *IDUA* | **Sun 2023** | **European** | **blood** | **pQTL** | **10** | **IVW** | **-0.084** | **0.016** | **1.65E-07** | **1.24E-04** | **0.91 (0.89, 0.95)** | **-** | **current study** |
| | PPMI | European | CSF | pQTL | 3 | IVW | 0.100 | NR | NR | 0.038 | NR | + | Kaiser 2023 |
| *LGALS3* | **Sun 2023** | **European** | **blood** | **pQTL** | **2** | **IVW** | **0.018** | **0.088** | **0.837** | **1.000** | **1.02 (0.86, 1.21)** | **+** | **current study** |
| | Emilsson 2018 | European | blood | pQTL | 1 | Wald ratio | 0.121 | 0.050 | 0.016 | 0.016 | 1.13 (1.02, 1.25) | + | Storm 2021 |
| | Suhre 2017 | European | blood | pQTL | 1 | Wald ratio | 0.122 | 0.047 | 0.010 | 0.012 | 1.13 (1.03, 1.24) | + | Storm 2021 |
| | PPMI | European | CSF | pQTL | 8 | IVW | 0.070 | NR | NR | 7.11E-04 | NR | + | Kaiser 2023 |
| | eQTLGen, PsychENCODE | European | brain | eQTL | 1 | Wald ratio | 0.727 | 0.191 | 1.44E-04 | 2.80E-04 | 2.07 (1.42, 3.01) | + | Storm 2021 |
| *PAM* | **Sun 2023** | **European** | **blood** | **pQTL** | **2** | **IVW** | **0.068** | **0.026** | **7.78E-03** | **0.557** | 1.07 (1.02, 1.13) | **+** | **current study** |
| | PPMI | European | CSF | pQTL | 12 | IVW | 0.090 | NR | NR | 9.63E-04 | NR | + | Kaiser 2023 |
| | Nalls 2019 | European | blood | eQTL | 679 | NR | -0.031 | 0.010 | 2.08E-06 | 4.36E-04 | NR | - | Nalls 2019 |
| *SIGLEC9* | **Sun 2023** | **European** | **blood** | **pQTL** | **7** | **IVW** | **0.032** | **0.012** | **5.23E-03** | **0.433** | 1.03 (1.01, 1.06) | **+** | **current study** |
| | PPMI | European | CSF | pQTL | 19 | IVW | 0.030 | NR | NR | 0.020 | NR | + | Kaiser 2023 |
